# The role of children in the spread of COVID-19: Using household data from Bnei Brak, Israel, to estimate the relative susceptibility and infectivity of children

**DOI:** 10.1101/2020.06.03.20121145

**Authors:** Itai Dattner, Yair Goldberg, Guy Katriel, Rami Yaari, Nurit Gal, Yoav Miron, Arnona Ziv, Rivka Sheffer, Yoram Hamo, Amit Huppert

## Abstract

One of the significant unanswered questions about COVID-19 epidemiology relates to the role of children in transmission. This study uses data on infections within households in order to estimate the susceptibility and infectivity of children compared to those of adults. The data were collected from households in the city of Bnei Brak, Israel, in which all household members were tested for COVID-19 using PCR (637 households, average household size of 5.3). In addition, serological tests were performed on a subset of the individuals in the study. Inspection of the PCR data shows that children are less likely to be tested positive compared to adults (25% of children positive over all households, 44% of adults positive over all households, excluding index cases), and the chance of being positive increases with age. Analysis of joint PCR/serological data shows that there is under-detection of infections in the PCR testing, which is more substantial in children. However, the differences in detection rates are not sufficient to account for the differences in PCR positive rates in the two age groups. To estimate relative transmission parameters, we employ a discrete stochastic model of the spread of infection within a household, allowing for susceptibility and infectivity parameters to differ among children and adults. The model is fitted to the household data using a simulated maximum likelihood approach. To adjust parameter estimates for under-detection of infections in the PCR results, we employ a multiple imputation procedure using estimates of under-detection in children and adults, based on the available serological data. We estimate that the susceptibility of children (under 20 years old) is 43% (95% CI: [31%, 55%]) of the susceptibility of adults. The infectivity of children was estimated to be 63% (95% CI: [37%, 88%]) relative to that of adults.

**Author Summary:** One of the significant unanswered questions about COVID-19 epidemiology relates to the role of children in transmission. In this study we estimate susceptibility and infectivity of children compared to those of adults using households data. The data were collected from households in the city of Bnei Brak, Israel, in which all household members were tested for COVID-19 using PCR. In addition, serological tests were performed on a subset of the individuals. Using a mathematical model to fit the data, we estimate that children are about half as susceptible to infection as adults, and are somewhat less prone to infect others compared to adults. In addition, using the serological data we find that under-detection of children, compared to that of adults, is more severe, given the PCR testing policy employed. Thus, both low susceptability and under-detection of children may explain the world-wide observation that the percentage of young children within confirmed COVID-19 cases is low compared to other age groups. However, the role of children in the spread of COVID-19 is also affected by different contact patterns and hygienic habits outside the household, so that more intense contact and mixing among children, for example in schools, could offset the effect of reduced susceptibility and infectivity.

## 1 Introduction

The COVID-19 pandemic, which emerged in Wuhan, China during December 2019, has now spread globally. Extreme measures have been taken worldwide in response to the outbreaks, among them, extended school and workplace closures. Guiding public health policies crucially depends on understanding the effect of age structure on the epidemic dynamics. In particular, susceptibility and infectivity are two critical aspects to consider when studying population heterogeneity in the context of infectious diseases. At this stage of the pandemic, it has become clear that the clinical characteristics of the disease among children are different from those in adults, with children having considerably lower risk of severe symptoms [1, 2]. In addition, studies report a markedly lower percentage of children diagnosed relative to their share in the population [3, 4, 5].

A key question is whether the above-noted difference between children and adults in rates of identified cases is the result of lower susceptibility of children to infection, or perhaps is due to the milder (or no) symptoms displayed by infected children, which, based on common testing policy, leads to under-detection [1]. These explanations are non-exclusive. Another crucial knowledge gap relates to the ability of those children who are already infected to infect others. As suggested by Kelvin et al. [6], the fact that children frequently do not display notable disease symptoms, raises the possibility that children could be facilitators of viral transmission. Deeper understanding of these issues is essential in order to assess the role of children in the transmission and spread of COVID-19, and has the potential to affect future policies to optimally mitigate the outbreaks.

Regarding the susceptibility of children to infection, available research yields mixed conclusions, but with some significant indications of lower susceptibility of children. Several random survey studies - not symptom based - have shown lower infection rates in children. In an icelandic study [7], using a PCR survey with random sampling, no children up to age 10 were found to be infected with SARS-CoV-2 as compared with 0.8% of persons over age 10. A 5 serological survey in Geneva [8] found that children aged 5-9 had a significantly lower risk of being seropositive compared to individuals aged 25-49, with a relative risk of 0.32. A random serological survey in Spain found seroprevalence by immunoassay of 3.8% in age group 0-19 compared to 4.6% in the population overall. These studies indicate that the lower rates of detected cases in children are not due only to under-detection because of milder symptoms, though they do not provide unequivocal evidence of lower susceptibility in children, since the lower rates of infection might also be affected by the isolation of children during a period of school closures. Another approach to examining susceptibility to infection is using contact tracing studies, in which contacts of known cases are isolated and tested. While a study of Bi et al. [9] concludes that children were as likely to be infected as adults, Zhang et al. [10] conclude that children are less likely to be infected compared to adults by about 60%. A study based on fitting an age-structured epidemic model to population and contact tracing data estimates that the susceptibility individuals under the age of 20 is approximately half that of those over 20, and also that the rates of manifestation of clinical symptoms are strongly age-dependent [11]. A systematic review and meta-analysis by Viner et al. [12] of contact tracing studies and population-level studies concluded that there is preliminary evidence that children and young people have lower susceptibility to SARS-CoV-2 - the pooled estimate over contact tracing studies of the odds ratio of being an infected contact in children compared to adults was 0.56 (95% CI: [0.37, 0.85]), with substantial heterogeneity over studies. Findings from population-level screening studies using PCR or serological testing were heterogeneous, with some studies showing a considerably lower rate of infection among children and others showing no such difference [12].

Regarding infectivity of children carriers, existing evidence is scarce. Cai et al’s [13] analysis of 10 children diagnosed with COVID-19, states that one cannot neglect the potential risk of transmission from the infected child to adult contacts, based on one patient. A study from New South Wales schools in Australia [14] based on both virus and antibody testing, suggests that children are not the primary drivers of COVID-19 spread in schools or in the community. According to Zimmerman et al. [15] the importance of children in transmitting the virus remains uncertain. Preliminary results from an ongoing research of the National Institute for Public Health and the Environment in the Netherlands (RIVM) [16] show no indications that children younger than 12 years were the first to be infected within the household, and suggest that patients under 20 years play a much smaller role in the spread than adults and the elderly.

In view of the crucial importance of understanding the role of children in the epidemiology of COVID-19, and of the considerable uncertainties that still remain, it is imperative to perform further studies collecting and analyzing relevant data.

In this study we aim to estimate the relative susceptibility and infectivity of children by analyzing a data set which is unique in both size and quality, collected from households in the city of Bnei Brak, Israel, in which all household members were tested for COVID-19 using PCR (637 households, 3,353 people of which 1,510 tested COVID-19 positive, average household size of 5.3). In a subset of these households, serological testing was also carried out. The PCR testing data reveals a clear dependence of positive rates on age. In order to gain understanding of the epidemiological mechanisms behind these age-related differences, we estimate susceptibility and infectivity parameters for children and for adults, by fitting a household transmission model to the data, using a simulated maximum likelihood approach. Since the PCR testing may miss some infections due to insufficiently frequent testing or false negatives, we also use the available serological data to assess the extent of this under-detection among children and adults. Specifically, we examine the effect of correcting for this under-detection on the estimates of the susceptibility and infectivity parameters, using a multiple imputation procedure.

## 2 Methods

### 2.1 Sources of data

This study is based on data collected from the city of Bnei Brak (population 213,046) which is the most densely populated city in Israel. Most of its residents are ultra-orthodox Jews, with large households and a young population (approximately 51% under the age of 20) [17]. Additional external data sources were used for estimating relevant quantities as described in the sequel.

#### 2.1.1 PCR testing

We used the results of real-time reverse transcriptase-polymerase chain reaction (RT-PCR) tests for SARS-CoV-2 and the epidemiological investigations from the Israeli COVID-19 database, performed in Bnei Brak between March 17 and May 3, 2020. During most of this period, the state of Israel was under lock-down. The city of Bnei Brak had the highest per capita infection rate in Israel and was the only city in Israel that was declared a “restricted area” [18].

Until the end of May, the general policy in Israel was to approve testing for people who had been in close contact with someone who had tested positive for COVID-19 or who had returned from abroad and, in both cases, had at least one of the symptoms in a list. However, in the case of the city of Bnei-Brak, approval was granted by the district physician to test all members of a household in which a suspected case occurred.

Testing was done for diagnostic purposes, by paramedics visiting the households. Most households in the data had several visits during the relevant period. Initial testing in a household was always due to a symptomatic individual reporting to the healthcare provider. Subsequent visits were prompted either by the need to re-test a household member who had already tested positive, or by a report of a newly symptomatic member of the household. Testing in a household ended when all positive cases had been confirmed as negative (by two subsequent tests) and no further reports of symptoms were made (in some households, testing continued after May 3, 2020, the last date of our data set). The decision as to which household members to test on each visit (in addition to the member who prompted the visit) was made on the spot by the medical team, and did not follow a systematic protocol. Therefore the number of times that an individual was tested and the gaps in time between consecutive tests vary among individuals, a fact which raises some concerns about the possibility of missed infections, which we will address in the following using information provided by serological tests.

For each individual, the data collected includes age, reports of symptoms, the dates on which the samples were taken and the results of the PCR testing. Symptoms and their dates of onset were self-reported: as part of the epidemiological investigations, those who tested positive were asked whether they had symptoms, and the date on which these symptoms appeared. The list of symptoms included fever, cough, shortness of breath, abdominal pains, headache, diarrhea, chills, sore-throat, muscle-pain, vomiting, other respiratory difficulties and additional symptoms, such as smell or taste problems, weakness, etc.

In order to map individuals to households, we used the municipality database of Bnei Brak residents born before May 25, 2020. The inclusion criteria, met by 637 households, were households with at least 2 members, in which all household members were tested and at least one member tested positive to COVID-19. The 637 households comprise a total of 3,353 individuals, of whom 1,510 COVID-19 tested positive, with mean household size of 5.3 (standard deviation 3.1). Histograms of household sizes in the data and the number of positives per household size are displayed in Figure 1.

**Figure 1:**
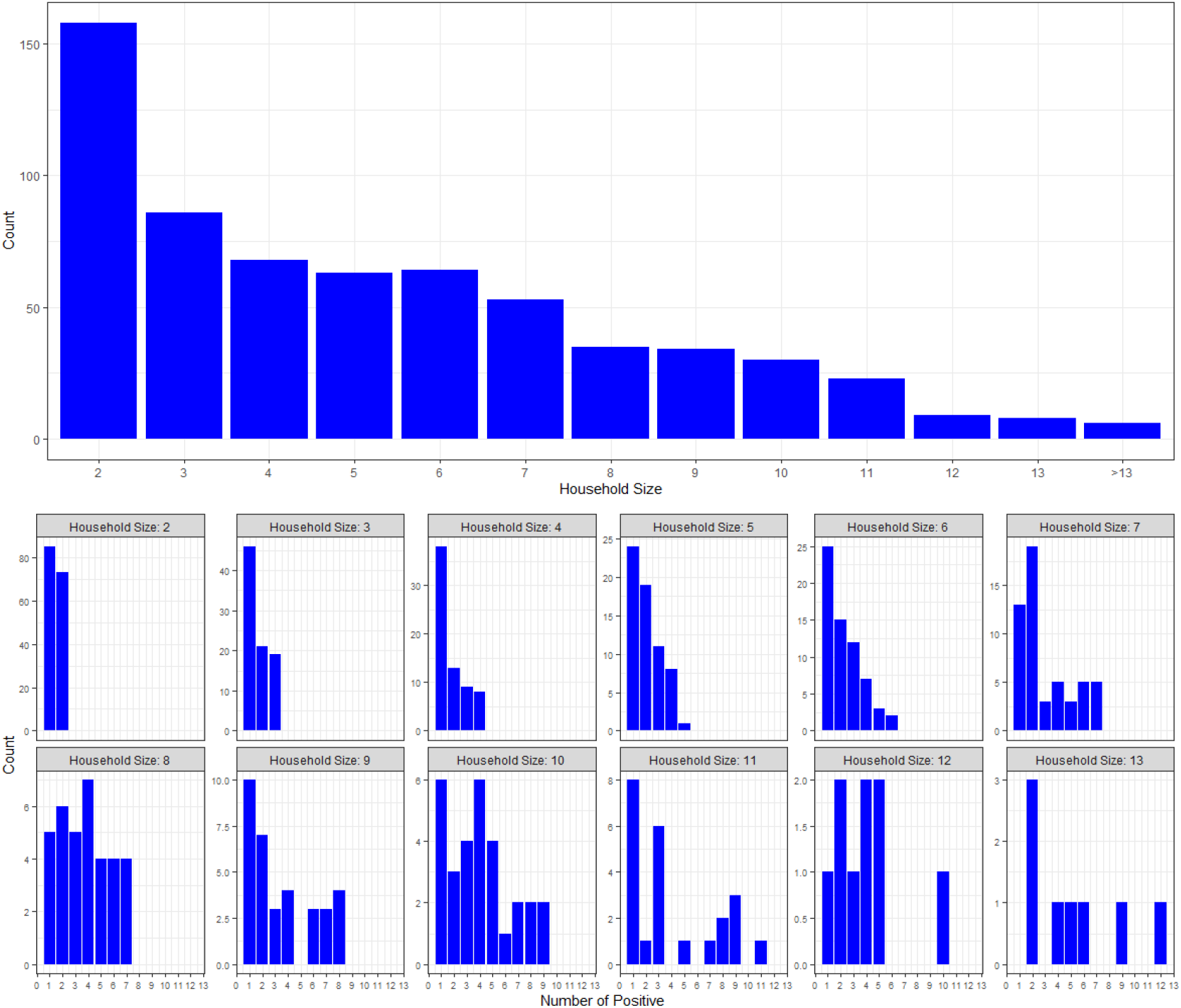
Top panel: histogram of household sizes in the Bnei Brak data set. Bottom panel: histogram of the number of positives per household size in the Bnei Brak data set (households of size greater than 13 are not shown due to their small sample size).

For each of the households, we have generated a plot which enables us to visualize the timeline of testing in the household. See Figure 2 for examples of two such plots, and its caption for explanations. The time-line plots for all households in the study are provided in the Supplementary Material.

**Figure 2:**
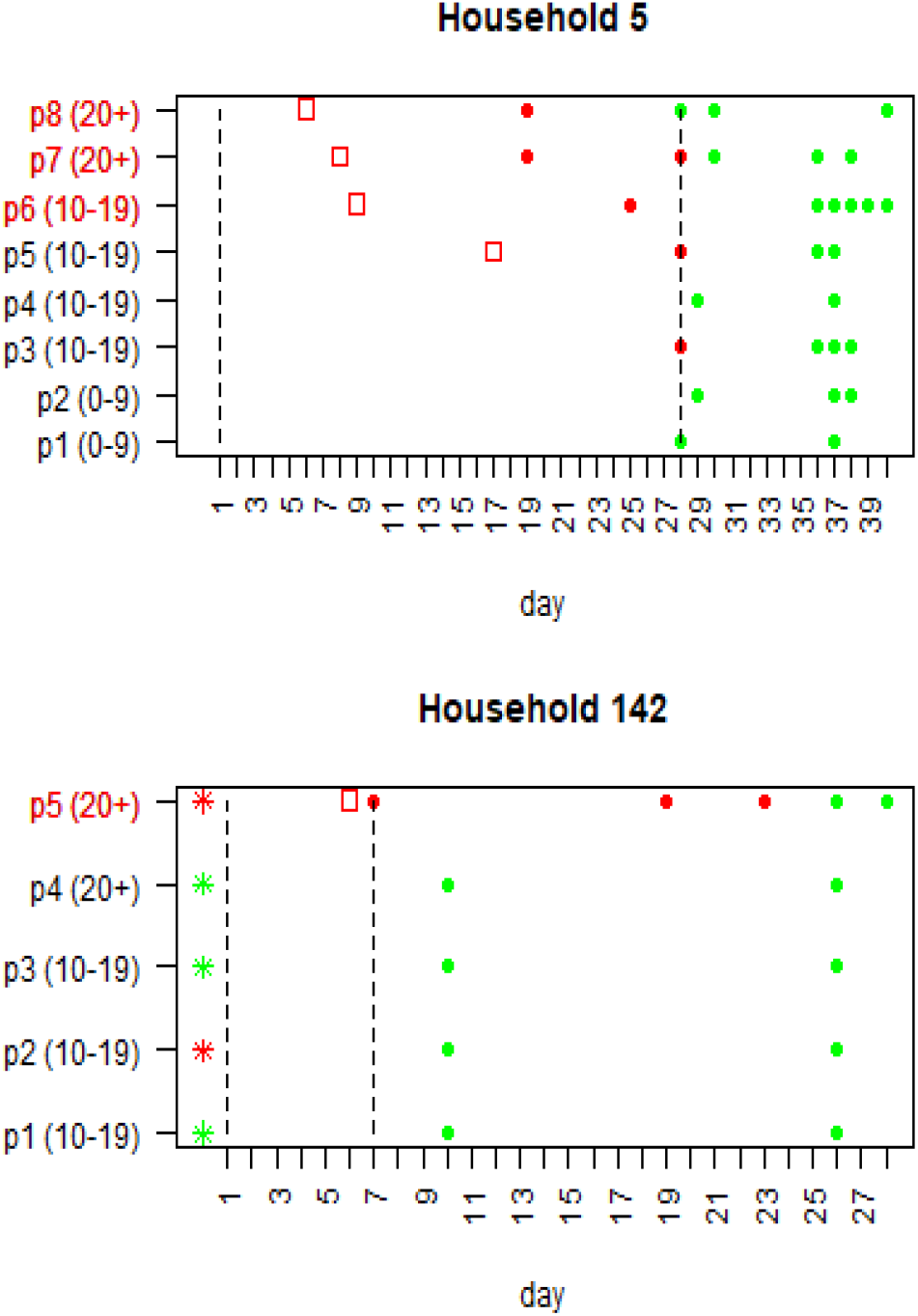
Example of two household timelines. Each row represents the timeline of an individual in the household. The age group of each member is given in parenthesis (precise ages are not supplied, due to privacy concerns). In these examples, Household 5 includes 8 individuals and Household 142 includes 5 individuals. Red circles denote positive PCR tests while green circles denote negative PCR tests. Red squares denote days of reported symptoms onset. The period between the two vertical lines denote the observed time-period used in the model fitting, which was set, for each household, according to the rules described in Section 2.2 of the Supplementary Material. Individuals whose label is colored in red are the suspected index cases, as determined by the criteria given in Section 2.3 of the Supplementary Material. In Household 5, there are three suspected index cases (p6-p8), whereas in Household 142 only one (p5). Members of Household 142 were also tested using serology and the results are shown using the asterisks - red for a positive result and green for a negative. As can be seen, member p5 of Household 142 was found positive both by PCR and serology, while member p2 was found positive using serology but not using PCR. The rest of the household members were found to be negative using both PCR and serology. The Supplementary Material contains similar timelines for all households in this study (excluding households with more than 10 individuals, which were removed for privacy considerations).

#### 2.1.2 Serological testing

A serological survey was conducted in Bnei Brak during June 2020. The survey was performed using a kit of the Abbott SARS-CoV-2 IgG, whose specificity was estimated as ∼100% and whose sensitivity at ≥21 days was estimated as ∼85% [19, 20]. As part of this survey, a subset of the households in our data set could be serologically tested. The criteria defined for selecting these households were: households with up to four adult (20+) members, at least two members who were negative using PCR, one of whom is an adult and the other a child (7-19), with preference for families with at least two members who were found positive using PCR. The rationale for these criteria was to include a sufficiently large sample of PCR negative cases to which the serological results can be compared. Altogether, 130 households out of the 637 household data set discussed above were serologically tested. In these selected households, tested between June 3 and June 21, 2020, all members were typically tested, except for children under the age of seven, who were not tested at all in the serological survey. Overall, 714 of the 3,353 individuals in the data set had serological tests. Nine of these individuals were found to be positive in PCR testing after May 3, 2020 (the last date used for the PCR testing data set) and before June 2020 (the period of serological testing) and have been removed from further analysis, leaving us with 705 serological test results. We note that the gap of one month between the end of the PCR testing in our data set and the beginning of serological testing is sufficiently long for seroconversion. Furthermore, during this period the COVID-19 outbreak in Israel, and in Bnei-Brak in particular, was at its lowest (see Figure 3), so that the probability that individuals in our database were infected during this period is low.

**Figure 3:**
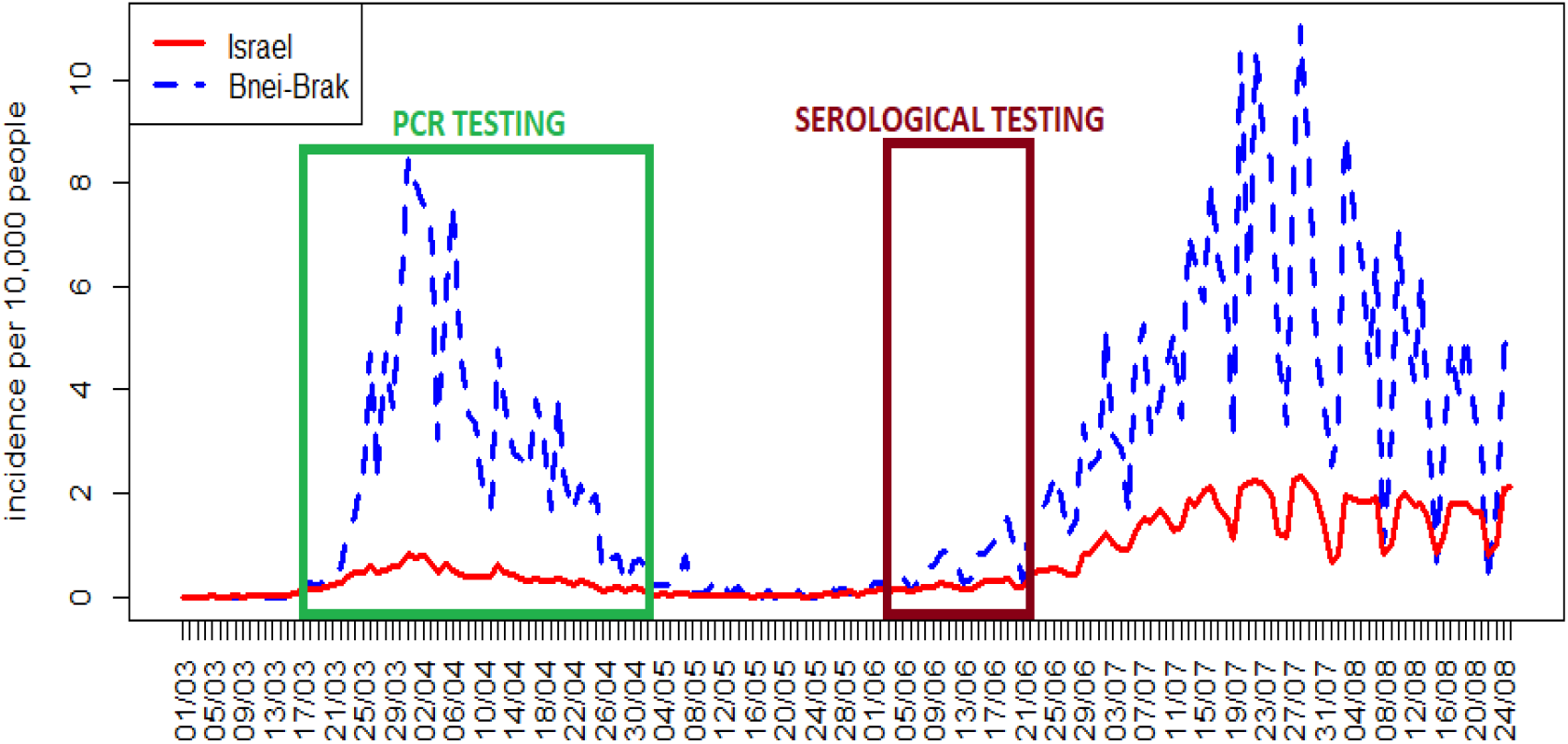
The number of new daily confirmed COVID-19 cases per 10,000 people in all of Israel and in the city of Bnei-Brak. The windows mark the periods of PCR and serological testing employed in our data set.

### 2.2 Estimating susceptibility and infectivity of children and adults through modelling

In order to assess differences in susceptibility and infectivity among children and adults, we use a dynamic stochastic mathematical model of a household outbreak allowing for these differences, and fit it to the observational data on infections in the households. Our data does not include information about who infected whom, nor dates of infection - the onset of symptoms and testing dates in our data are used only to identify the index case and set the observed duration, as described below. We use only aggregate numbers of infected individuals in the two age groups in the different households. The key point is that these data on outcomes of the many “household outbreaks” contain valuable information concerning the infectivity and susceptibility parameters, which can be extracted by a model-fitting approach: only certain ranges of values of these parameters will generate outcomes which are consistent with those observed in the data. Inference for household models is a classic issue in the epidemiological study of infectious diseases [21],[22]. We note that while for a complete household outbreak there exist elegant analytical expressions for the probabilities of outcomes, so that a likelihood function can be computed in closed form [23], in this work we cannot assume that the entire period of the household epidemic is observed in our data. Our outcomes relate to a certain duration of time for which PCR testing was conducted in a given household, and in this case we do not have closed expressions for the likelihood, which motivates the “simulated maximum likelihood” approach we take (see explanation below). For other approaches to dealing with the problem of ‘right-censoring’ involved in analyzing ongoing household epidemics see [24].

#### 2.2.1 The model

We use a stochastic dynamic model for a household outbreak. Time is indexed by the discrete variable *t* (in days). We denote by *S*_*a*_(*t*) and *S*_*c*_(*t*) the number of adults and children who are still susceptible on day *t*, respectively. The symbols *i*_*a*_(*t*) and *i*_*c*_(*t*) stand for the number of adults, and children who become infected on day *t*, respectively.

The dynamic equations are

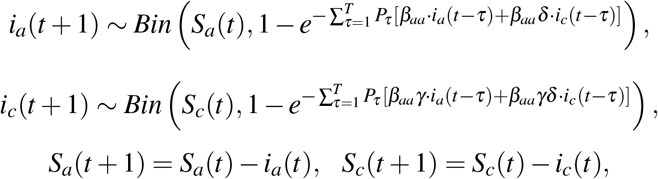

where *P*_*τ*_, *τ* ∈{1, …, *T*} is the generation-time distribution, set to be a discretized version of a gamma distribution with a mean of 4.5 days, and a standard deviation of 2.5 days. This mean generation time is based on the mean intervals between symptom onset obtained from ∼ 4500 pairs of known infector-infectee in the data set of confirmed cases in Israel (see Figure S2 in the Supplementary Material) and is also compatible with findings from other studies [25, 26, 27]. The parameter *β*_*aa*_ is the transmission rate among adults, *γ* is the susceptibility of children relative to that of adults, and *δ* is the infectivity of children relative to that of adults. In words, the relative susceptibility of two individuals is defined as the ratio of their probabilities of being infected per unit time, when exposed to the same infectious factor. The relative infectivity of two individuals is defined as the ratio of the probabilities per unit time that these individuals generate infection, when making contact with individuals who have identical susceptibilities. See Supplementary Material for a detailed description of the model.

#### 2.2.2 Fitting the model to data using simulated maximum-likelihood

We use the model to fit the data regarding the number of positive adults and children in each household. Given the number of individuals of each age group in a household, the age group to which the index case belongs, values of transmission parameters and a duration of observation, one can generate simulations of a household outbreak for the specified duration. For each such simulation, we record outcome: the number of individuals of each age group who were infected during the time period considered. Since the model is stochastic, different realizations of such a simulation will lead to different outcomes. The probability distribution over the finite set of possible outcomes is approximated for each of the households, by running 1000 simulations for each set of transmission parameter values in a specified range over a grid, with a resolution of 0.05. These probability distributions, which are dependent on the transmission parameters, enable us to compute the likelihood function corresponding to the outcome in each of the households in our empirical data. The total likelihood is then the product of the likelihoods for all households. This likelihood is a function of the transmission parameters (*β*_*aa*_, *γ, δ*) and is used to estimate the transmission parameters using maximum likelihood. Note that since our likelihood function is computed using simulation, our estimation method is what is known as “simulated maximum likelihood” [28].

Fitting the model to the available data requires to set the duration of the observed period for each household, which is used in the simulations providing the likelihood calculations. This is important in order to deal with potential right-censoring of household data: if we take a duration longer than that for which we have data, we implicitly assume that no infections occurred after the last observed one, an assumption which could be invalid. Note that since we do not exclude the possibility that further infections occurred after the observed time period - the observed period does not necessarily correspond to the full household outbreak. For details on the likelihood calculation and the procedures employed to set the observed period and the age group of the index case for each household see Section 2 of the Supplementary Material.

To test the ability of our estimation procedure to identify parameters, we carried out a simulation study in which household outbreaks with known parameters were generated in a collection of households of the same type as those in the data set, and our method was used to estimate the parameters. A similar procedure was used to obtain parametric bootstrap confidence intervals, by generating 1000 simulated data sets using the parameter estimated from the real data, and re-estimating the parameters (see Section 4 and Figure S4 in the Supplementary Material). An R software package applying our methodology in a computationally efficient way is available online [29], allowing other researchers to estimate relative susceptibility and infectivity of children and adults given an appropriate data set.

#### 2.2.3 Accounting for misclassified cases using multiple imputation

In the procedure described above, we have assumed that the data regarding infections in a household is complete for the period designated as the observed period in the household. In particular, it has been assumed that

a. All individuals who tested positive were indeed infected (no false positive).
b. All individuals who tested negative were not infected during the observed period (no false negatives).

We believe that assumption (a) is justified, in view of the high specificity of PCR tests, as well as the fact that most positive cases were symptomatic and tested positive more than once. Assumption (b) is, however, overly optimistic, for two reasons: the first is that PCR tests have a non-negligible false negative rate [30], so that an infected person might test negative. Even more importantly, while the inclusion criteria for households in our study stipulated that all household members be tested at least once, and indeed many of the subjects were tested multiple times, the frequency of testing of many of the subjects is not sufficiently high to exclude the possibility that some of them were infected but their infection was missed by the testing. This could occur if a person became infected during the period defined as the observed period but subsequent to the last date on which a test was taken from this individual, or if an individual had become infected prior to the observation period, or following one of the tests, but became negative before a subsequent test was taken.

With the availability of the serological data for a subset of the individuals, we could replace assumption (b) to take into account the possibility of cases misclassified as negative, by applying a multiple imputation procedure as follows:

1. We assume that all individuals who tested positive with PCR were indeed infected (even if a serological test was negative - since the serological tests have a 15% false negative rate [20]).
2. We classify those who have been tested negative with PCR as “questionable”, except for those who had serological tests, in which case we used the result of the serological test to determine their status.
3. We assume that a person classified as “questionable” has a certain probability of having been infected and not detected. To estimate this probability we exploit the serological data. Restricting ourselves to individuals for which a serological test is available, we calculate, for each age group (children and adults), the proportion of individuals who tested positive on the serological test out of those who tested negative on the PCR test. This proportion serves as our estimate of the probability that a “questionable” individual of the corresponding age group was in fact infected.
4. We generate 20 data sets in each of which the status (positive or negative) of each of the individuals classified as “questionable” is randomly assigned according to the probability estimated using the serological data (see] (3) above). In addition, the questionable cases who are imputed as positive were treated as suspected index cases. For each of these data sets, the model is re-fitted using the likelihood method described in Section 2.2.2 to obtain estimates for the parameters, and standard errors for these estimates are calculated using 100 parametric bootstrap simulations. Using Rubin’s rules [31], we then obtain confidence intervals for the parameters taking into account the multiple imputation procedure that was performed. We thus obtain parameter estimates which are adjusted for misclassified cases.

## 3 Results

### 3.1 PCR testing

Inspection of the data reveals that chances of becoming infected increase with age, up to around age 20, and remains approximately constant thereafter (Figure 4 top panel).

**Figure 4:**
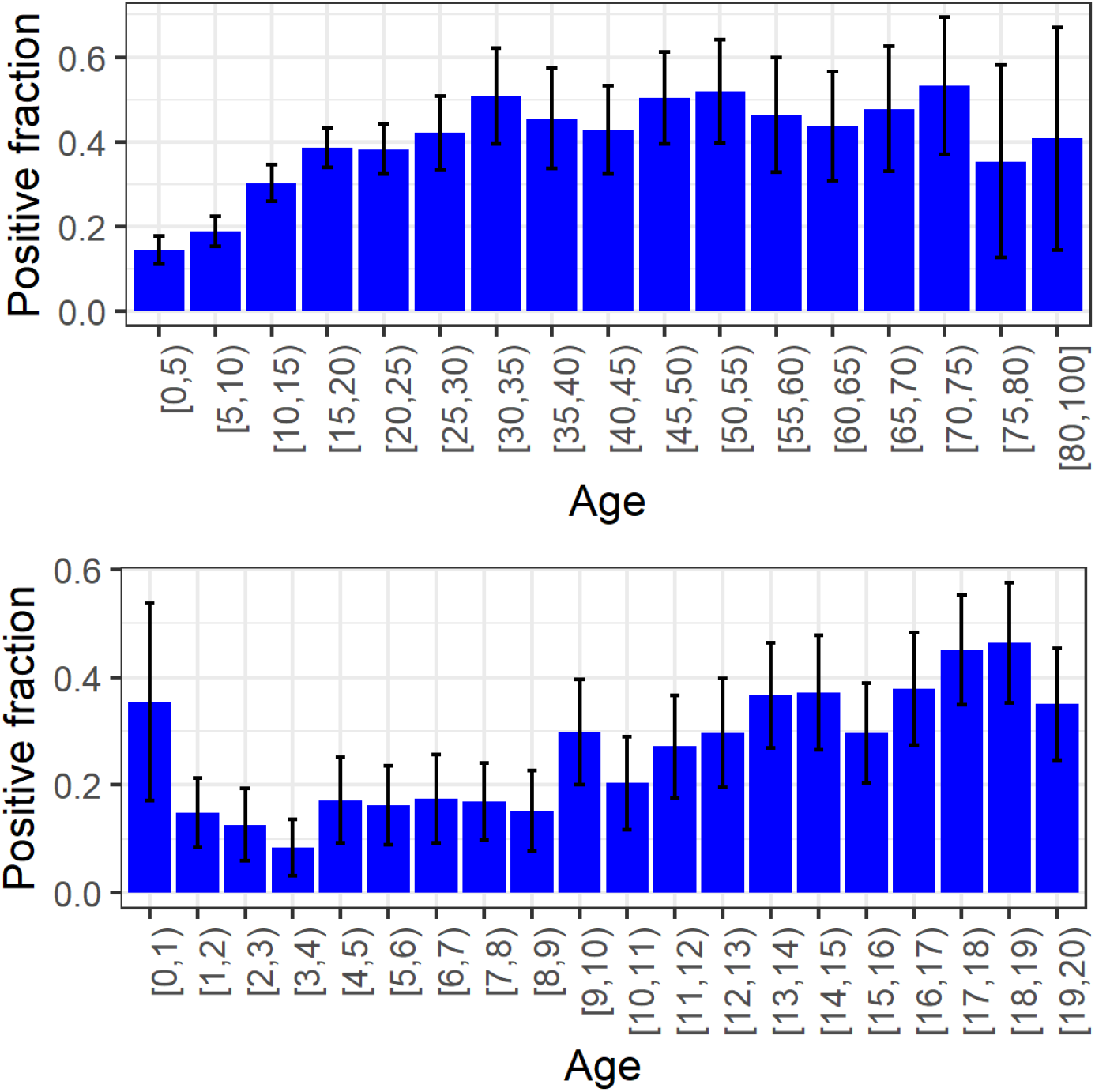
Top panel: Fraction of positives by age-group in the data set, excluding index cases. Bottom panel: Fraction of positives by age in children, excluding index cases. Binomial confidence intervals in both figures were calculated using the normal approximation.

We divide the population into a children’s group (0-19 including) and an adult group (20+). Table 1 summarizes the information regarding PCR testing and symptomatic cases in individuals belonging to the data set, using this cutoff. We obtain a positive rate of 65% among adults compared to 28% among children. Excluding index cases, which in most cases were adults (see Table S1 in the Supplementary Material), 44% of adults were infected compared to 25% of the children. Interestingly, children under the age of one seem to be more likely to be infected than children between one and four (Figure 4 bottom panel). Table 2 summarizes the information regarding the households in the data set and the results of the PCR testing in these households.

**Table 1:**
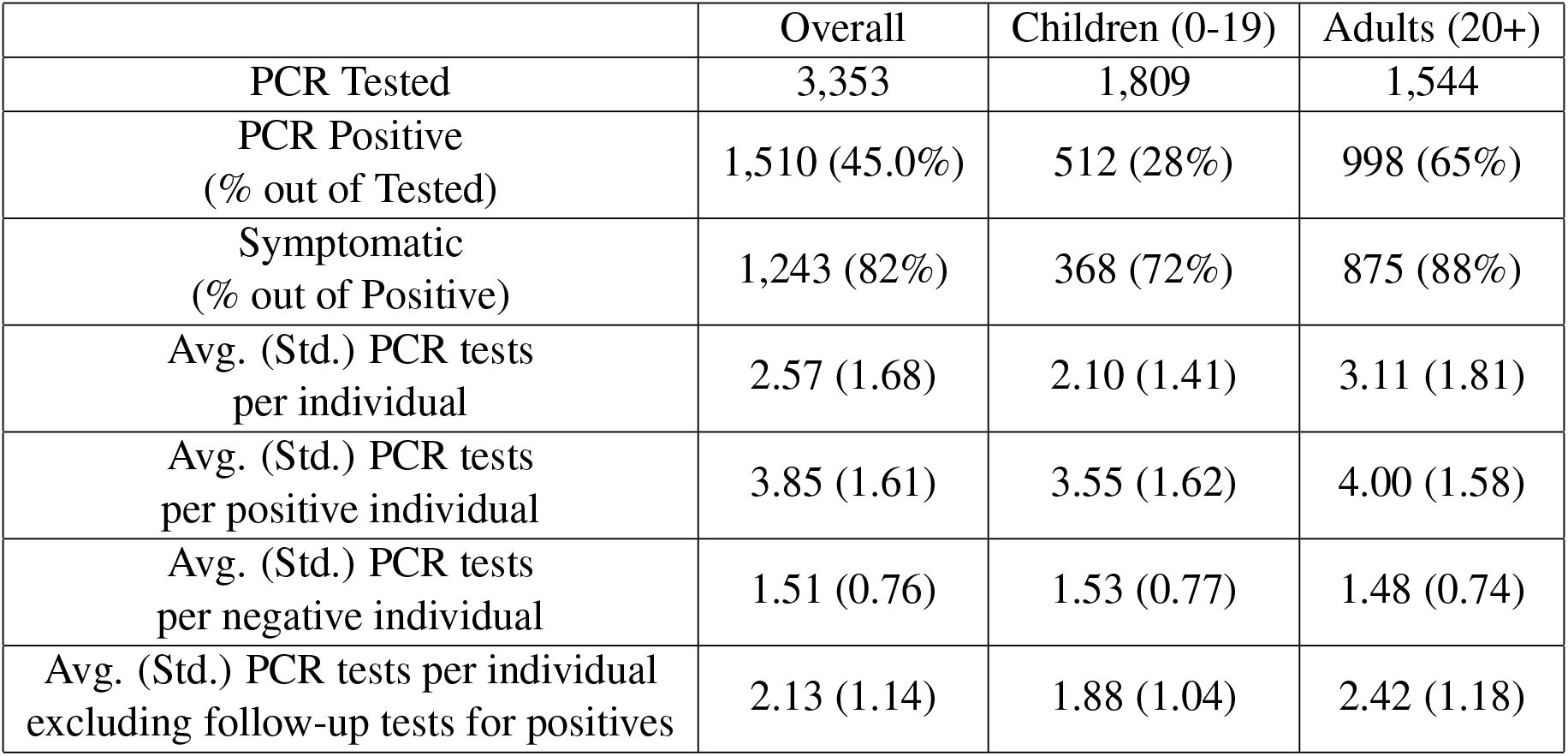
Summary of PCR testing results for individuals in the data set

|  | Overall | Children (0-19) | Adults (20+) |
| --- | --- | --- | --- |
| PCR Tested | 3,353 | 1,809 | 1,544 |
| PCR Positive<br>(% out of Tested) | 1,510 (45.0%) | 512 (28%) | 998 (65%) |
| Symptomatic<br>(% out of Positive) | 1,243 (82%) | 368 (72%) | 875 (88%) |
| Avg. (Std.) PCR tests<br>per individual | 2.57 (1.68) | 2.10 (1.41) | 3.11 (1.81) |
| Avg. (Std.) PCR tests<br>per positive individual | 3.85 (1.61) | 3.55 (1.62) | 4.00 (1.58) |
| Avg. (Std.) PCR tests<br>per negative individual | 1.51 (0.76) | 1.53 (0.77) | 1.48 (0.74) |
| Avg. (Std.) PCR tests per individual<br>excluding follow-up tests for positives | 2.13 (1.14) | 1.88 (1.04) | 2.42 (1.18) |

**Table 2:** Summary statistics for households in the data set.

|  |  |
| --- | --- |
| Number of Households | 637 |
| Household Size<br>Avg. / Std. / Min. / Max.) | 5.26 / 3.07 / 2 / 15 |
| Number of Adults 20+ per Household<br>(Avg. / Std. / Min. / Max.) | 2.42 / 1.04 / 1 / 8 |
| Number of Children 0-19 per Household<br>(Avg. / Std. / Min. / Max.) | 2.84 / 2.85 / 0 / 12 |
| Number of PCR Positives per Household<br>(Avg. / Std. / Min. / Max.) | 2.37 / 1.85 / 1 / 12 |
| Number of PCR Positive Adults 20+ per Household<br>(Avg. / Std. / Min. / Max.) | 1.57 / 0.88 / 0 / 7 |
| Number of PCR Positive Children 0-19 per Household<br>(Avg. / Std. / Min. / Max.) | 0.80 / 1.43 / 0 / 9 |

Figure 5 shows a boxplot of the observed duration (as defined in section 2.2 in the supplementary material) in households of different sizes. These durations give lower bounds to the durations of the household outbreaks. It demonstrates that a household outbreak can typically last 2-4 weeks, with a weak dependence on the household size, so that large households experience somewhat longer epidemics. This point should be relevant when considering the duration of lock-downs intended to curtail the epidemic in the population.

**Figure 5:**
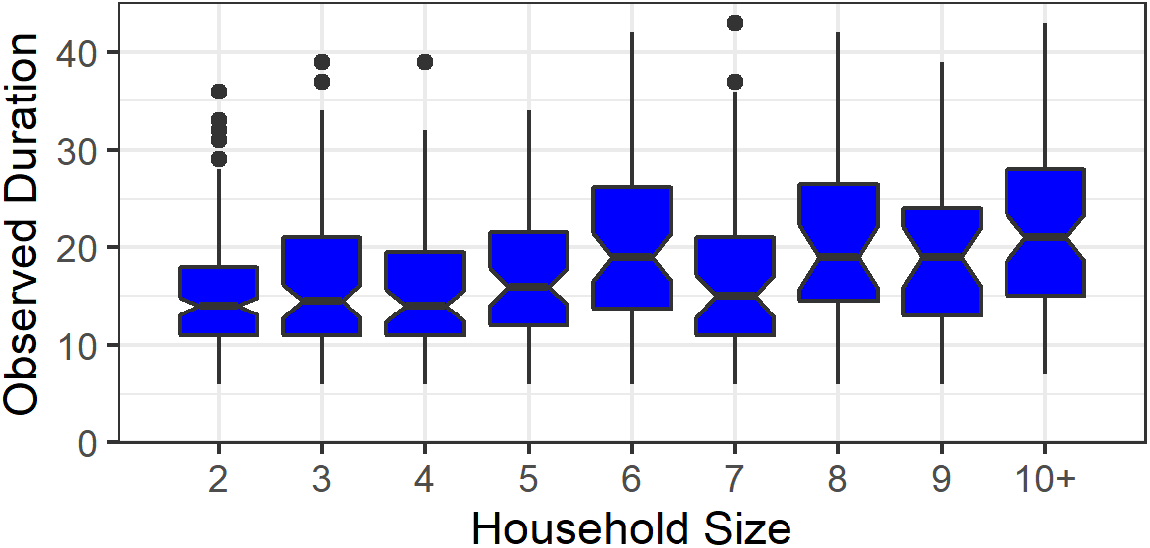
A boxplot of the observed duration (in days) for different household sizes in the Bnei-Brak data set.

### 3.2 Results of model fitting to PCR data

We now present the estimates obtained by our modelling approach (described in Methods Section 2.2.2), using the PCR testing data alone. In Section 3.4 below we present somewhat modified estimates obtained when adjustment is made to account for under-detection of cases, as estimated from the serological data.

We estimated that the relative susceptibility of children (*γ*) is 35% (95% CI: [30%, 40%]). The relative infectivity (*δ*) of children was estimated to be 70% (95% CI: [55%, 90%]). The adult-adult transmission parameter *β*_*aa*_ was estimated as 0.4 (95% CI: [0.35, 0.45]). The ranges reported are based on parametric bootstrap confidence intervals (with a precision limited by the grid resolution of 0.05). Figure 6 displays level curves of the negative log-likelihood as a function of the susceptibility and infectivity parameters, for three values of the adult-adult transmission parameter *β*_*aa*_. Figure 7 shows the model fit to the observations within households of different sizes. The fit obtained using the model is much better than the fit obtained using a naïve model that ignores secondary infections within households (see Figure S3 in the Supplementary Material).

**Figure 6:**
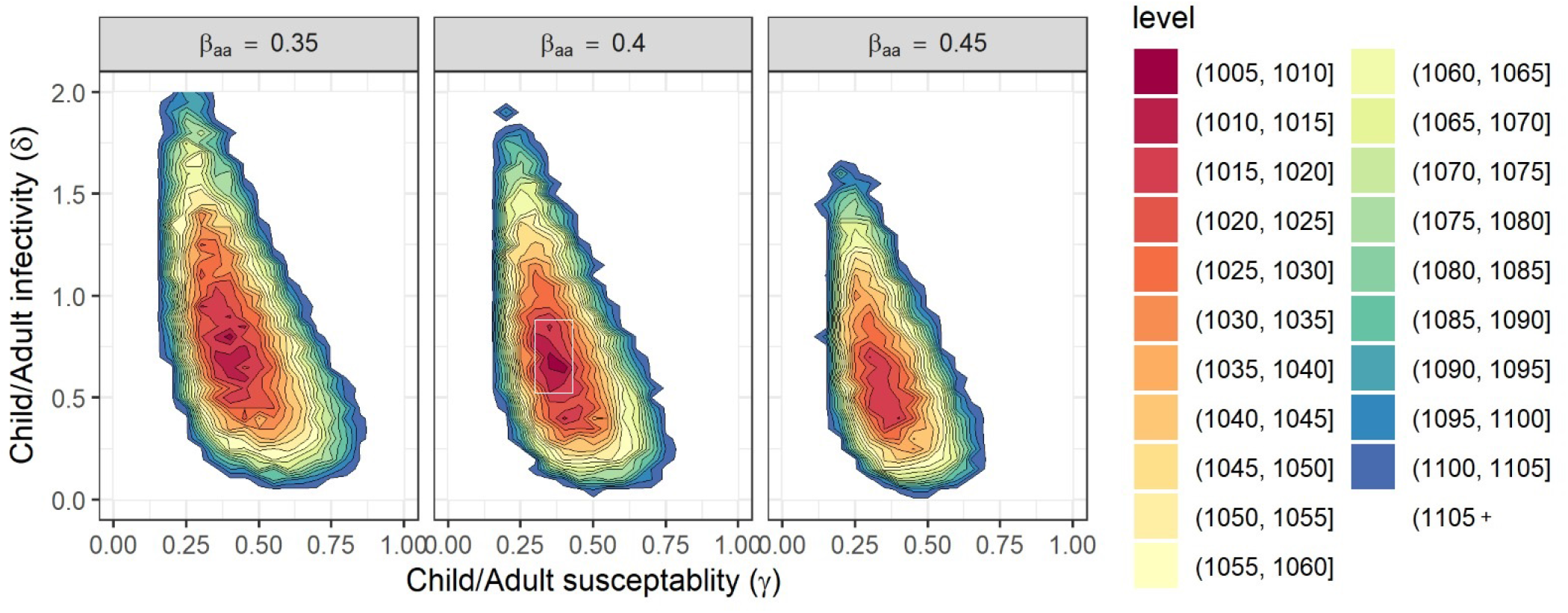
Negative log-likelihood level curves for the parameters *γ, δ*, and three values of *β*_*aa*_. For smaller values of *β*_*aa*_ the maximal-likelihood estimates of parameters *γ* and *δ* are larger. The white rectangle in the middle figure shows the 95% confidence region obtained using the bootstrap simulation.

**Figure 7:**
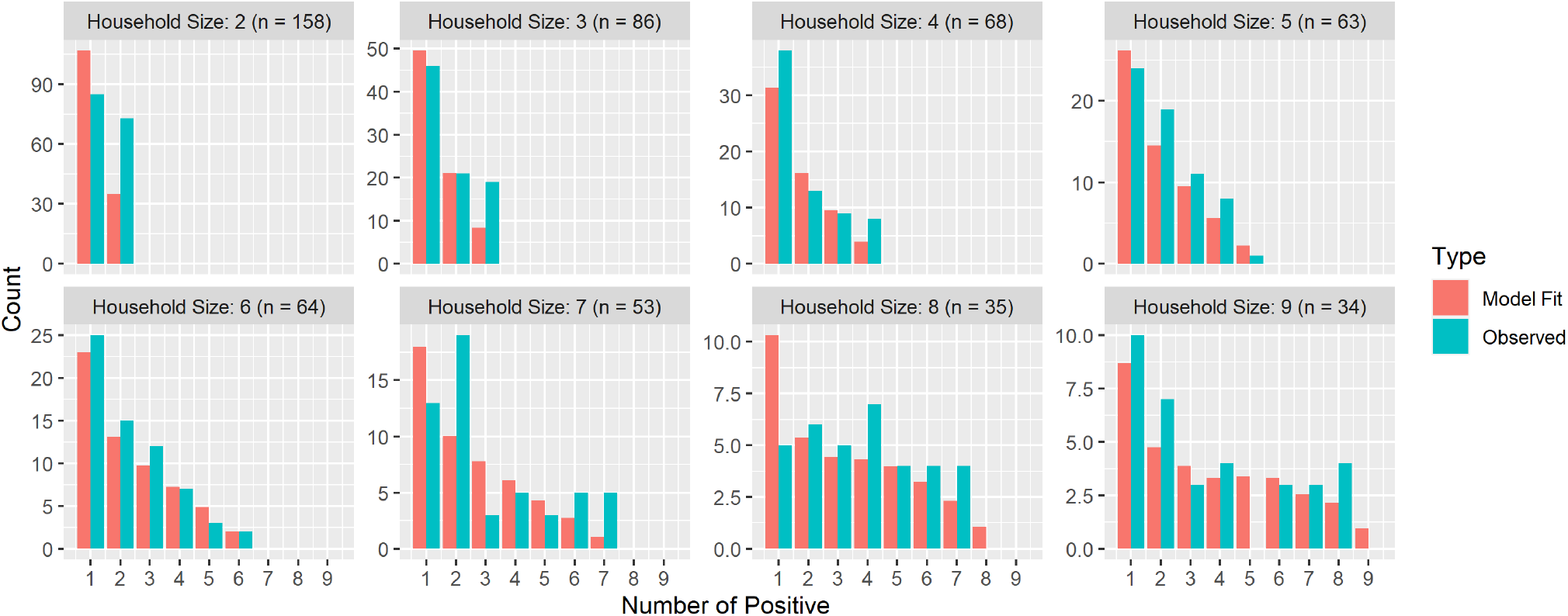
Best model fit to the PCR testing data in the Bnei Brak data set, aggregated according to the household size (fit to households of size greater than or equal to 10 are not shown due to their small sample size). The histograms show the distribution of the total number of infected individuals (children+adults) in households of different sizes, together with the corresponding distributions as predicted by the model with the fitted parameters. While the fitting was done to the number of infected children and adults separately, as described in the text, the figure shows the summary results in order to allow a visual examination of the goodness-of-fit.

We performed sensitivity analyses to examine the effect of various assumptions on these results. These included sensitivity to the assumed generation-time distribution, the assumed age-group of the index case in households in which there was doubt regarding the index case’s age-group, and to the assumed observed duration of the transmission in the households. In general, our results seem to be robust to reasonable variations in all of these attributes. Full description of the sensitivity analyses appears in Section 5 of the Supplementary Material.

### 3.3 Serological testing

Of the 705 individuals in our data set for whom serological test results were available, 417 were children (7-19 years old) and 288 were adults (20+). 34% of these children and 48% of the adults tested serologically positive.

Table 4 compares the serological and PCR results for those individuals in our database who were also serologically tested. Note that since the serological sampling targeted households with at least two negative cases (see Section 2.1.2), the positive rates of PCR testing in this sample were lower than those observed in the entire data set: 22% in children (compared to 28% in the the entire data set) and 45% in adults (compared to 65% in the entire data set).

**Table 3:**
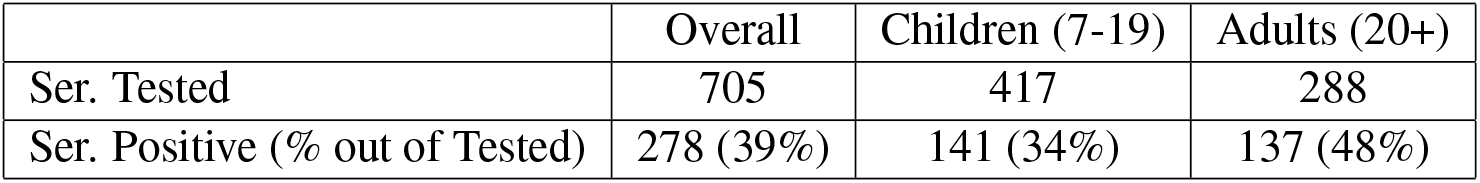
Results of serological testing on individuals from the Bnei-Brak data set

**Table 4:** Comparison of PCR and serological results for individuals in the Bnei-Brak data set tested by both.

|  | Overall |  | Children (7-19) |  | Adults (20+) |  |
| --- | --- | --- | --- | --- | --- | --- |
| PCR Ser. | Negative | Positive | Negative | Positive | Negative | Positive |
| Negative | 381 | 46 | 252 | 24 | 129 | 22 |
| Positive | 102 | 176 | 74 | 67 | 28 | 109 |

A crucial observation afforded by Table 4 is that among children who were confirmed positive in serological testing, only 48% (95% CI: [37%,56%]) were detected positive by the PCR testing, in comparison with a corresponding figure of 80% (95% CI: [72%,86%]) for adults. This indicates that children who were infected were considerably less likely to be detected by the PCR testing than adults who are positive. A likely explanation for this finding is that children who are positive tend to display less symptoms (see Table 1), leading to lower probability of detection. As we have noted in Section 2.2.3, although all individuals in our study have been PCR tested at least once, it is possible for infections to be missed if the timing of the testing is not appropriate. Since household visits were mostly prompted by the reporting of symptoms, if adults tend to display symptoms more than children we expect that positive adults would be detected at higher rates than children. In addition, PCR tests have a non-negligible false-negative rate, and it is possible that this rate is correlated with level of symptoms, leading to higher false-negative rate in children. Table 5 displays a similar comparison between serological and PCR testing data as in Table 4, but separated for those individuals who were PCR tested only once and those tested at least twice. As expected, for individuals tested more than once the probability of detecting a positive using PCR is higher - 77% for children (95% CI: [66%,86%]) and 88% for adults (95% CI: [81%,94%]).

**Table 5:** Comparison of PCR and serological results for individuals who were tested only once by PCR and individuals who were tested at least twice by PCR.

| Results for individuals who were tested only once by PCR |  |  |  |  |  |  |
| --- | --- | --- | --- | --- | --- | --- |
|  | Overall |  | Children (7-19) |  | Adults (20+) |  |
| PCR Ser. | Negative | Positive | Negative | Positive | Negative | Positive |
| Negative | 247 | 3 | 137 | 2 | 49 | 1 |
| Positive | 71 | 12 | 57 | 10 | 14 | 2 |

| Results for individuals who were tested at least twice by PCR |  |  |  |  |  |  |
| --- | --- | --- | --- | --- | --- | --- |
|  | Overall |  | Children (7-19) |  | Adults (20+) |  |
| PCR Ser. | Negative | Positive | Negative | Positive | Negative | Positive |
| Negative | 134 | 43 | 85 | 22 | 80 | 21 |
| Positive | 31 | 164 | 17 | 57 | 14 | 107 |

The finding that positive children were less likely to be detected than adults in the PCR testing data should be taken into account when studying the age dependence of transmission - otherwise a bias is introduced into the analysis. To do so we use the joint PCR/serological data to estimate the probability that an individual classified as negative using PCR is actually positive. Of those tested negative in PCR, 21% (95% CI: [18%,25%]) tested positive using serology - 23% (95% CI: [18%,28%]) in children and 18% (95% CI: [12%,25%]) in adults. We note that the fact that the serological tests were not performed on children under the age of seven might lead to some bias in the misclassification probability calculated for children.

We can adjust the PCR positivity rates observed in the entire data set (28% among children and 65% among adults) using the above estimates for under-detection. If we add the estimated number of undetected infections, we obtain a positive rate of 28+(100-28)*0.23=45% for children and 65+(100-65)*0.18=71% for adults. Thus, even following this correction, the positive rates of adults is considerably higher than that of children. This consideration implies that the lower rate of detection in children does not fully explain the difference in infection rates in the two age groups. We therefore conclude that differences in transmission characteristics among age groups still must play a role in explaining these differences in rates.

### 3.4 Results of model fitting to adjusted PCR data using multiple imputation

To assess the differences in transmission characteristics among age groups while adjusting for the under-detection of infection among children, we employ the multiple imputation procedure described in Section 2.3 using the estimates for under-detection derived from Table 4: the probability of having been infected despite not having been detected in the PCR testing is estimated at 23% for children and 18% for adults.

We estimated that the relative susceptibility of children (*γ*) is 43% (95% CI: [31%, 55%]). The relative infectivity (*δ*) of children was estimated to be 63% (95% CI: [37%, 88%]). The adult-adult transmission parameter *β*_*aa*_ was estimated as 0.47 (95% CI: [0.36, 0.57]). Compared to the estimates made without adjusting for misclassified cases (Section 3.2), the adjusted estimates give a somewhat higher value for the relative susceptibility of children and a somewhat lower value for their relative infectivity.

The confidence intervals derived from our imputation procedure might underestimate uncertainty, since the number of imputations employed was relatively small. In addition, we did not account for the uncertainty in the misclassification rates (due to sampling error) used in the imputation procedure. These limitations are due to the high computational burden of running the multiple imputation procedure.

## 4 Discussion

Currently, one of the most significant unanswered questions about COVID-19 transmission relates to the role of children in the spread of infection. As we have noted in the Introduction, the fact that the fraction of children among the confirmed cases has been found to be low in many countries can be accounted for by two (nonexclusive) hypotheses: (1) Children display milder symptoms than adults when infected, so are less likely to be tested in a typical testing policy triggered by symptoms, (2) Children are less susceptible to infection than adults.

Our analysis of the data obtained in this study lends support to *both* hypotheses, and indicates that both have a role in explaining the observed epidemiological patterns.

First consider hypothesis (1). Examination of the joint PCR/serological testing data, for the subset of individuals for which it was available, revealed that children who were infected (as evidenced by serological testing) were less likely to be detected using PCR compared to adults who were infected. A likely explanation for this difference is that children tend to be less symptomatic than adults (Table 1), and are therefore not tested as intensively, as evidenced by their lower average number of tests per individual when excluding follow up tests for positive cases (Table 1). In our data, all visits to a household were triggered by a report of a symptomatic household member, and during these visits additional tests within a household were conducted, even if no symptoms were observed. Thus, although all household members were eventually tested, the testing policy was symptom-biased, in the sense that those with symptoms were more likely to be tested during the time window in which viral load was sufficiently high for detection. Since children are more likely to present milder or no symptoms, we would expect a higher rate of under-detection in children compared to adults, as we see in the data, which is the effect described by hypothesis (1).

However, our analysis indicates that hypothesis (1), *by itself* is not sufficient to explain the lower proportion of positive cases within children compared to that in adults. Indeed, even after adjusting the rates of infection obtained from the PCR data, using estimates of under-detection rates derived from the joint PCR/serological data, we obtain lower infection rates in children compared to adults in our data. This indicates that differences in positive rates between children and adults are not fully explained by different detection rates. In order to account for the differences observed in the data one needs to posit age-related characteristics of transmission, and not only of detection.

To explore what differences in transmission characteristics could account for the data, we used a dynamic stochastic model for transmission in a household, allowing for different susceptibility and infectivity of children and adults, and fitted it to the available data on outcomes in the households of our data set. To account for under-detection, we also performed this analysis combined with multiple imputation, using the under-detection estimates from the joint PCR/serological data. This correction somewhat shifted our estimated parameters, though the change was not a major one.

The estimation results indicate that the role of children in the transmission of infection is less prominent than that of adults: children are less susceptible than adults (relative susceptibility 43% [31%, 45%]), and their infectivity is somewhat lower as well (relative infectivity 63% [37%, 88%]). The data were more informative regarding the relative susceptibility of children than regarding their relative infectivity, as indicated by much wider confidence intervals for the relative infectivity in comparison to those for the relative susceptibility. Data containing more index cases in the children’s group, as well as data providing more detailed information about the timeline of infections within the household, would provide more information about children’s infectivity.

Our result concerning the lower susceptibility of children is in agreement with the result of Davies et al. [11] who estimated that children under the age of 20 have a level of susceptibility half that of adults, based on fitting a population-level model to data on clinically-reported cases, together with data from contact-tracing studies. This result raises the question of possible biological mechanisms that could account for such an effect. One possible explanation that has been raised relates to lower expression in children of angiotensin-converting enzyme 2 (ACE2), the receptor that SARS-CoV-2 uses for host entry [32]. Another hypothesis relates to recent studies which found evidence suggesting the presence of some residual immunity in people not previously exposed to SARS-CoV-2, in the form of SARS-CoV-2-reactive CD4+ T cells, attributed to circulating “common cold” coronaviruses [33, 34]. It is possible that this form of partial protection is more common in children since infection rates with seasonal coronaviruses are higher in children [35]. The fact that in our data set, children under the age of one have higher rates of infection with SARS-CoV-2 compared to children between one and four, is consistent with the hypothesis that partial immunity to SARS-CoV-2 could be related to past exposure to seasonal coronaviruses. However, a recent study could not find evidence for a protective effect of prior infection with seasonal coronaviruses against SARS-CoV-2 infection in children [36].

While our estimates of children’s susceptibility and infectivity are lower than those of adults within a household, it is important to bear in mind that their role in the spread of COVID-19 is also affected by different contact patterns and hygienic habits outside the household, so that more intense contact and mixing among children and adolescents compared to adults, for example in schools, may offset the effect of reduced susceptibility. In Israel, a second wave of the epidemic started shortly after re-opening of schools at the beginning of May (see Figure 3) and included major outbreaks in several high-schools [37, 38]. At the moment of this writing, Israel has closed its schools again (and went into lock-down), after the re-opening of schools on September 1st, following the summer holiday, has generated another surge of cases, with particularly high rates among the 15-19 age group (see Figure S8 in Supplementary Material).

As in nearly all studies of disease transmission, the available data in this study is partial and imperfect. Our data is partial in the sense that it does not provide us with a full picture of the transmission in the household (who infected whom), and imperfect in that testing was not according to a systematic protocol, and some cases were not detected, as revealed by the partial serological data. Extracting insights into the underlying processes from the data requires analysis and modelling, which must be based on simplifying assumptions. We have used a model with two age groups, due both to computational complexity considerations and data limitations, though it is likely that susceptibility and infectivity would vary in a graded way with age. Specifically, more recent population-level data (as discussed above regarding the results of school opening) may suggest that individuals of age group 15-19 would be more appropriately classified as adults with respect to transmission characteristics. We have used imputation to account for the under-detection of infections, and our imputation process assumes that missed cases occur as independent random events, whose probability depends only on the age group, as estimated using the serological data. Violation of this assumption could lead to bias in our estimates. However, the sensitivity tests have shown that our general conclusions are robust to various variations in modelling assumptions.

We would suggest, in conclusion, the importance of carrying out further studies in households and in other settings, in order to refine our understanding of heterogeneity in transmission, whether related to age or to other factors, which is crucial for guiding public health policy. In particular, improving the resolution of data collection by frequent and regular testing of all individuals would allow to track the course of transmission in a more detailed way, enabling inference based on timing of detection and not only on final outcomes, and preventing under-detection and the need to adjust for it. Such studies, with enhanced data collection, have the potential to allow inference for more detailed models, as well as to reduce uncertainty in parameter estimates.

## Ethics statement

An exemption from institutional review board approval was given by the Israeli Ministry of Health as part of an active epidemiological investigation, based on use of anonymous data only and no medical intervention.

## Supporting information

Household timelines

## Data Availability

The household data used for fitting the model and the computer code (in the R language) for the data analysis can
be downloaded from https://github.com/yairgoldy/sl4hm.

## Data sharing

The household data used for fitting the model and the computer code (in the R language) for the data analysis can be downloaded from [29].

## Acknowledgments

The authors would like to thank Ofra Amir, Edward Goldstein, Boaz Lev and Baruch Velan for their support, encouragement, and fruitful discussions, and to the epidemiology team of the Tel Aviv health district.

