## Supplementary Material for "The role of children in the spread of COVID-19: Using household data from Bnei Brak, Israel, to estimate the relative susceptibility and infectivity of children"

#### 1 Household outbreak dynamic model

Assume a household has $N_a$ adults and $N_c$ children, $N = N_a + N_c$. The dynamic model we use for describing an outbreak in a household is a stochastic age-of-infection model, similar to the chain-binomial models traditionally used for household outbreaks ([1], ch. 14-15, [2], ch. 10), but somewhat more realistic in taking into account the generation-time distribution. The model is described below. The main parameters of the model are

$$\beta = \begin{bmatrix} \beta_{aa} & \beta_{ac} \\ \beta_{ca} & \beta_{cc} \end{bmatrix},$$

where $\beta_{ij}$ $(i, j = a, c)$ are transmission coefficients from individuals of type $j$ to individuals of type $i$. These parameters will be fitted using simulated maximum likelihood. In addition, our approach requires to estimate the generation-time distribution $P_\tau$ $(1 \le \tau \le T)$ based on previous data.

Infections of adults (children) occur according to a Poisson process with rate $\lambda_a(t)$ $(\lambda_c(t))$. These rates (also called forces of infection) are given by

$$\lambda_a(t) = \sum_{\tau=1}^{T} P_\tau \left[\beta_{aa} i_a(t-\tau) + \beta_{ac} i_c(t-\tau)\right],$$

$$\lambda_c(t) = \sum_{\tau=1}^{T} P_\tau \left[\beta_{ca} i_a(t-\tau) + \beta_{cc} i_c(t-\tau)\right].$$

The probability that a susceptible adult (child) becomes infected on day $t$ is therefore

$$p_a(t) = 1 - e^{-\lambda_a(t)}, \;\; p_c(t) = 1 - e^{-\lambda_c(t)}.$$

We denote by $S_a(t)$ and $S_c(t)$ the number of adults and children who are susceptible on day $t$, respectively. Further, denote by $i_a(t)$ and $i_c(t)$ the number of adults and children who become infected on day $t$, respectively. The dynamics is described by

$$i_a(t) \sim Bin\left(S_a(t), p_a(t)\right),$$

$$i_c(t) \sim Bin\left(S_c(t), p_c(t)\right),$$

$$S_a(t+1) = S_a(t) - i_a(t), \quad S_c(t+1) = S_c(t) - i_c(t).$$

The initial conditions are:

$$i_a(t) = i_c(t) = 0, \quad t < 0,$$

and, if the index case is an adult,

$$i_a(0) = 1, \;\; i_c(0) = 0,$$

$$S_a(0) = N_a - 1, \;\; S_c(0) = N_c,$$

while if the index case is a child:

$$i_a(0) = 0, \;\; i_c(0) = 1,$$

$$S_a(0) = N_a, \;\; S_c(0) = N_c - 1.$$

The household outbreak ends at time $t^*$ when no further infectious can occur in a household.

The total number of infected among adults and children in the household is

$$I_a = N_a - S_a(t^*), \;\; I_c = N_c - S_c(t^*).$$

Under the assumption that adults and children differ in susceptibility to being infected and in degree of infectivity, we can write

$$\begin{aligned} \beta_{ac} &= \beta_{aa} \cdot \delta, \\ \beta_{ca} &= \beta_{aa} \cdot \gamma, \\ \beta_{cc} &= \beta_{aa} \cdot \gamma \cdot \delta, \end{aligned} \tag{1}$$

where $\gamma$ is the susceptibility of children relative to that of adults, and $\delta$ is the infectivity of children relative to that of adults. Note that this parametrization restricts the structure of the $\beta$-matrix, since only three parameters are involved, rather than four.

An example of the use of the dynamic model is presented in Figure S1, where 10,000 epidemics were simulated using the maximum-likelihood (ML) parameter estimates ($\beta_{aa} = 0.40, \gamma = 0.35, \delta = 0.70$) for a household of 2 adults and 4 children and an index case that is an adult (left) or a child (right). One can see the mean daily number of infected individuals (children and adults), excluding the index case, along the duration of the epidemic.

### 2 Computing the likelihood and fitting the model

#### 2.1 Serial-interval and detection-time distributions

The model uses a generation-time distribution with a mean of 4.5 days. This mean generation time is based on the mean serial-interval distribution, representing intervals between symptoms onset of known couples of infected-infectee, obtained from epidemiological investigations in Israel during March and April 2020. To set the observed time period for a household (see below) we employ the observed mean detection time (interval between infection and confirmation using PCR), obtained from the same epidemiological investigations reference above. Figure S2 plots these two distributions.

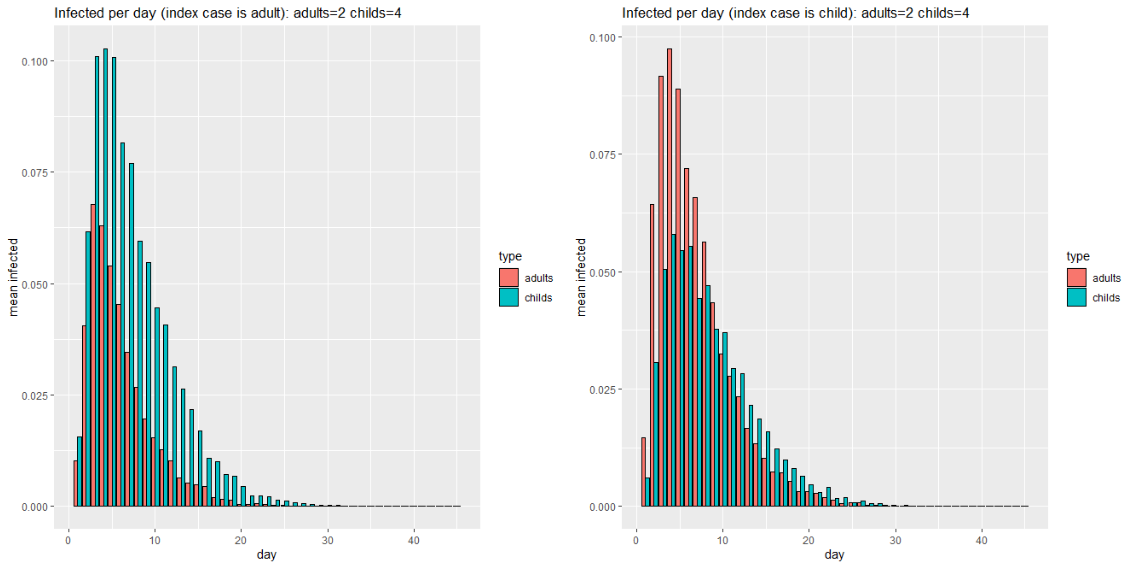


Figure S1: Results of 10,000 simulations of the dynamic model.

#### 2.2 Determining the observed time period

Fitting the model to the available data requires to determine the duration of the observed period for each household, which is used in the simulations providing the likelihood calculations below. We restrict the observed time period as follows:

(a) To set the start date of the observed period in a given household, we first identify the earliest indication of infection in the household, which is either onset of symptoms of a positive case or a first positive test. While testing a household was prompted by reporting of symptoms, in $\sim 10\%$ of the households, the earliest indication was a positive test, which could be attributed either to missing reports of symptoms or to cases in which the symptomatic individual who prompted the initial testing was found to be negative (our data set does not include reports of symptoms for negative cases). In addition, in cases in which the reported symptoms onset was over four weeks prior to the first test, the onset date of symptoms was discarded (12 cases).

(b) If the earliest indication is an onset of symptoms, then the start date is set to be five days prior to this onset (the mean incubation period [3]). Otherwise the start date is set as 10 days prior to the first date of a positive test (the mean time from infection to detection, during March-April 2020, as estimated from Israeli epidemiological investigation data, see Figure S2 below). This determination of the initial date reflects the approximate timing of infection of the index case.

(c) The end date of the observed period was set as the latest date on which some household member was tested positive for the first time.

#### 2.3 Determining the age group of the index case

The reported onset dates of symptoms and test dates were also used to discern the index case in each household. Positive household members whose first indicative date (onset of symptoms or first positive test) was five or less days from the earliest indicative date of all positive household members were considered suspected index cases with equal probability. Thus, for example, if there were three positive household members whose first indicative date was within five days of the first indicative date in the household, each of the three were given a probability of being an index case of 1/3 (and the rest of the household were given probability 0). For each household, the probability that the index case is an adult (child) was obtained by summing the probability of being an index case for all the adults (children) in that household. See Figure 2 in the main text for examples of the determination of the observed period and the suspected index cases in two households, using the above criteria. Table

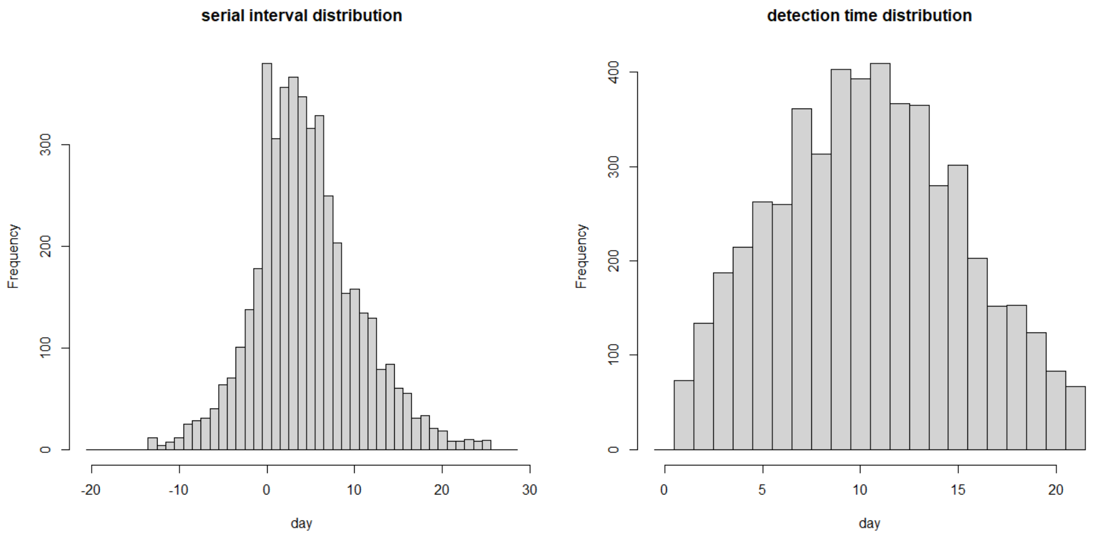


Figure S2: Serial-interval distribution with mean of 4.5 days and Detection-time distribution with mean of $\sim$ 10 days.

S1 presents the number of households with a given probability assigned for the index case to be an adult. Note that in most cases this probability is either 1 or 0.

| # households | 34 | 1 | 1 | 2 | 2 | 1 | 10 | 2 |
|---|---|---|---|---|---|---|---|---|
| P(index case is adult) | 0 | 0.17 | 0.18 | 0.20 | 0.25 | 0.29 | 0.33 | 0.40 |

| # households | 1 | 32 | 1 | 2 | 17 | 3 | 528 |
|---|---|---|---|---|---|---|---|
| P(index case is adult) | 0.43 | 0.50 | 0.55 | 0.60 | 0.66 | 0.80 | 1 |

Table S1: Number of households with a given probability assigned for the index case to be an adult.

#### 2.4 The likelihood function

Assume that we have information on a set of households indexed by $h$. For each household we have the data:

$$(N_a(h), N_c(h), \rho(h), I_a(h), I_c(h), D(h)),$$

where $N_a(h), N_c(h)$ are the number of adults and children in the household, $\rho(h)$ indicates the probability that the index case is an adult.

The variables $I_a(h)$ and $I_c(h)$ stand for the number of adults and of children who were infected, while $D(h)$ is the duration of the observed outbreak, determined as described in the main text.

Consider a household $h$, and assume that the index case $i$ is of type $ind(h)$ (where $ind = a$ for an adult or $ind = c$ for a child index case). Our goal is to compute the probabilities

$$P_h(I_a, I_c \mid ind, \beta)$$

of the possibles outcomes $I_a$ and $I_c$, which are the total infected adults and total infected children in such a household, respectively, given values of the parameters $\beta$. In order to compute $P_h$ we run $S = 1000$ realizations of the simulation of the household model, for a household with $N_a(h)$ adults, $N_c(h)$ children and with index case of type $ind$, with the parameters $\beta$, for a duration of $D(h)$ days. We generate a table of the fraction of the simulations in which each of the possible outcomes was obtained, thus obtaining estimates for the above probabilities.

Given the actual outcome $(I_a(h), I_c(h))$ in a household the likelihood corresponding to this household, taking into account the uncertainty regarding the identity of the index case, is

$$L_h(\beta) = \rho(h)P(I_a(h), I_c(h) \mid a, \beta) + (1 - \rho(h))P(I_a(h), I_c(h) \mid c, \beta).$$

The likelihood corresponding to the entire data set is given by the product of the likelihoods corresponding to each of the households:

$$L(\beta) = \prod_h L_h(\beta),$$

and the corresponding log-likelihood is

$$LL(\beta) = \sum_h \log L_h(\beta).$$

To estimate the parameters $\beta$ we maximize $LL(\beta)$ over the parameters $\beta$. Specifically, we consider the special structure of the $\beta$ matrix given by (1).

### 3 Comparison with an alternative model

We compared our model with an alternative, naive model fit to the data. We considered a simpler model in which in each household, all the infected members were infected by the index case, with no secondary infections within the household. We assume that in a household with an adult index case, the probability that a child will be infected is $p_{ca}$ and the probability that an adult will be infected is $p_{aa}$. Similarly, in a household with a child index case, the probability that a child will be infected is $p_{cc}$ and the probability that an adult will be infected is $p_{ac}$. In this simple model, in a household with $N_a$ adults and $N_c$ children and an adult index case, the number of children infected in the household will be binomially distributed with $n = N_c$, $p = p_{ca}$, and the number of adults infected (excluding the index case) will be binomially distributed with $n = N_a - 1$ and $p = p_{aa}$, and similarly for a household with a child index case. We therefore refer to this model as the binomial model. The parameters in this binomial model are easily estimated as the ratio of the number of individuals infected to the number of individuals at risk. The estimators are

$$\hat{p}_{aa} = 0.45, \quad \hat{p}_{ca} = 0.24, \quad \hat{p}_{ac} = 0.49, \quad \hat{p}_{cc} = 0.31.$$

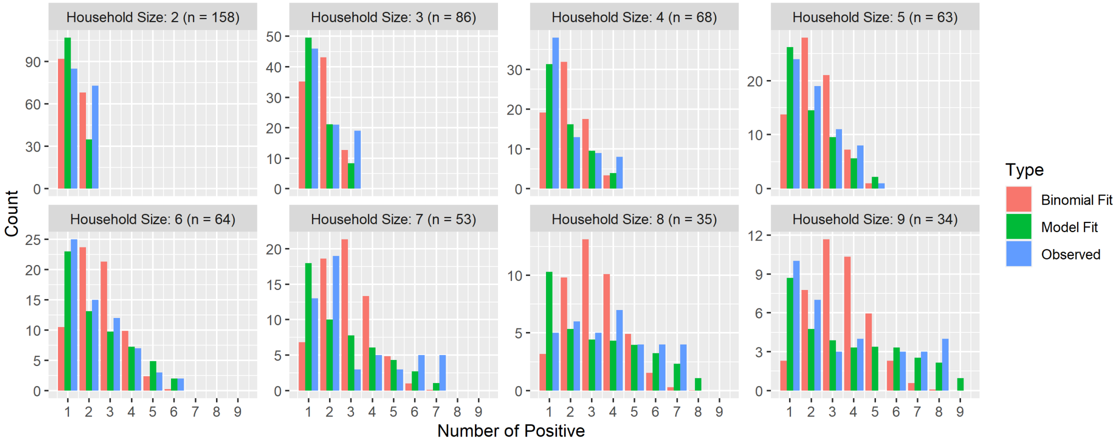


Figure S3: Comparison of the dynamic model and the naive binomial model fit to the data.

Figure S3 suggests that the dynamic model fits the data considerably better than the naive binomial alternative. Note that the dynamic model has 3 parameters compared to 4 of the binomial, hence is a more parsimonious model.

### 4 Bootstrap simulations

In order to obtain parametric bootstrap confidence intervals for the ML parameter estimates we ran 1000 simulations of the dynamic model using the ML parameter estimates and employing the exact same households as in the Bnei-Brak data, and generated the number of infected children and adults in each household. We then re-estimated the parameters for these simulated data sets. The results are shown in Figure S4. The obtained 95% confidence intervals are $[0.35-0.45]$ for $\beta_{aa}$, $[0.30-0.40]$ for $\gamma$ and $[0.55-0.90]$ for $\delta$.

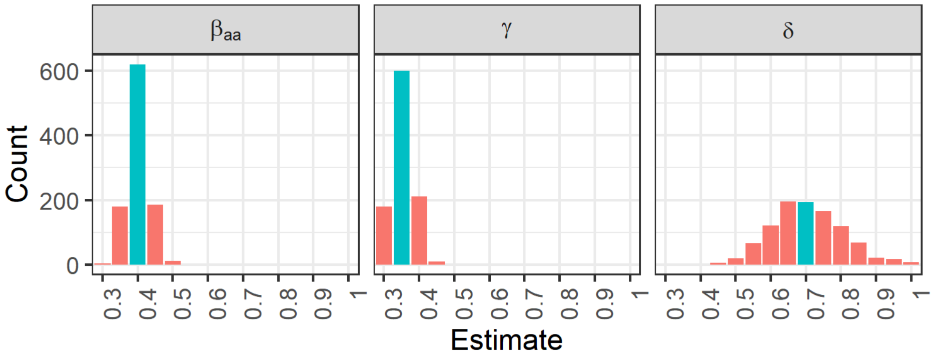


Figure S4: Results of 1000 bootstrap simulations. $\beta_{aa}$ - the transmission parameter among adults, $\gamma$ - relative susceptibility of children, $\delta$ - relative infectivity of children.

### 5 Sensitivity analysis

Sensitivity analysis was performed to examine the stability of the results to variations in the assumptions made when fitting the data.

#### 5.1 Sensitivity to observed epidemic duration

We examined the sensitivity to variations in the observed epidemic time-period of the households in the data. We modified the duration of the observed epidemic in each household in the data set as follows: If a household's first indicative date was some member's symptom onset date, then instead of using the mean incubation time to determine the start time of the epidemic, we sampled an incubation period from the incubation time distribution in the general COVID-19 cases data in Israel. If a household's first indicative date was some member's first test date, then instead of using the mean detection time to determine the start time of the epidemic, we sampled an detection period from the detection time distribution in the general COVID-19 cases data in Israel. We refitted the model to this data set to estimate the model parameters. This was repeated 100 times. The results of this test are shown in Figure S5. Blue color indicates the ML parameter estimates obtained using the duration set as in the main text. In most cases the inferred parameter values were identical or very similar to the original estimates, demonstrating small sensitivity of the results to the exact number of days the epidemic was observed in each household.

#### 5.2 Sensitivity to uncertainty regarding the index case age-group

We examined the sensitivity of the results to the fact that in some cases the age-group to which the index case belongs is uncertain. In cases where the index case age-group was not certain (in 75 of the 637 households - see Table S1), we randomly selected an age-group for the index case according to the probability of the index case being in each of the age-groups (as described in the Methods section). The model parameters were re-estimated by fitting the model to this data set. This was

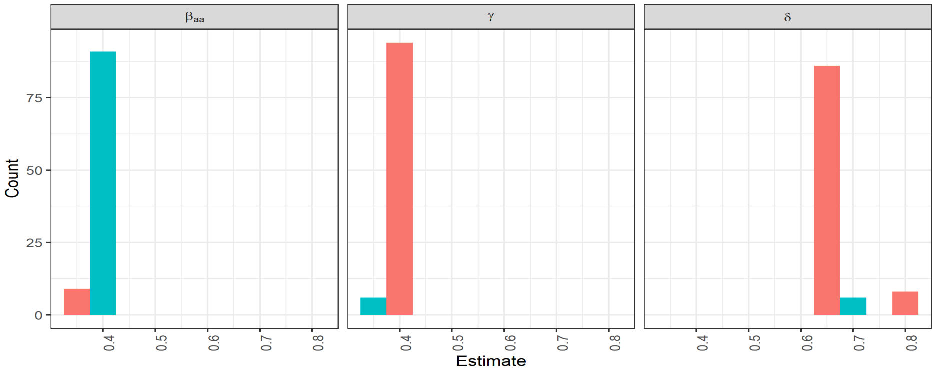


Figure S5: Sensitivity to observed epidemic duration. $\beta_{aa}$ - the transmission parameter among adults, $\gamma$ - relative susceptability of children, $\delta$ - relative infectivity of children.

repeated 100 times. The results of this test are shown in Figure S6. Blue color indicates the ML parameter estimates obtained using the original data set. In most cases the inferred parameter values were identical or very similar to the original estimates, demonstrating small sensitivity of the results to the uncertainty regarding the index case age-group.

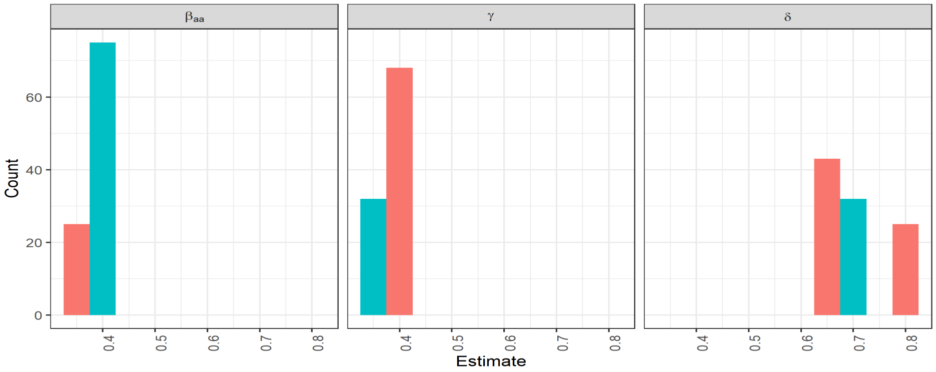


Figure S6: Sensitivity to uncertainty regarding the index case age-group. $\beta_{aa}$ - the transmission parameter among adults, $\gamma$ - relative susceptability of children, $\delta$ - relative infectivity of children.

#### 5.3 Sensitivity to the mean generation-time

This analysis is aimed for testing the sensitivity of the estimation results to the mean of the generation-time distribution. Recall that in the main text the results were obtained using a generation-time distribution with a mean value of 4.5. Here, we generated 100 simulated data sets with the exact household types as in the Bnei-Brak data set, while employing the ML parameter estimates and a generation-time distribution with mean of 4 days. Then we fitted a model to this data set assuming a mean generation time of 4.5 days, as used in Section Results of the main text. The results are presented in the top row of Figure S7. It shows that the obtained estimates are not too sensitive to these deviations from the actual mean generation time. We repeated this type of analysis, now generating 100 data sets with a mean generation time of 5 days, and fitting again a model to this data set assuming a mean generation time of 4.5 days. The results are presented in the bottom row of Figure S6 and here as well, the estimates are not too sensitive to such deviations.

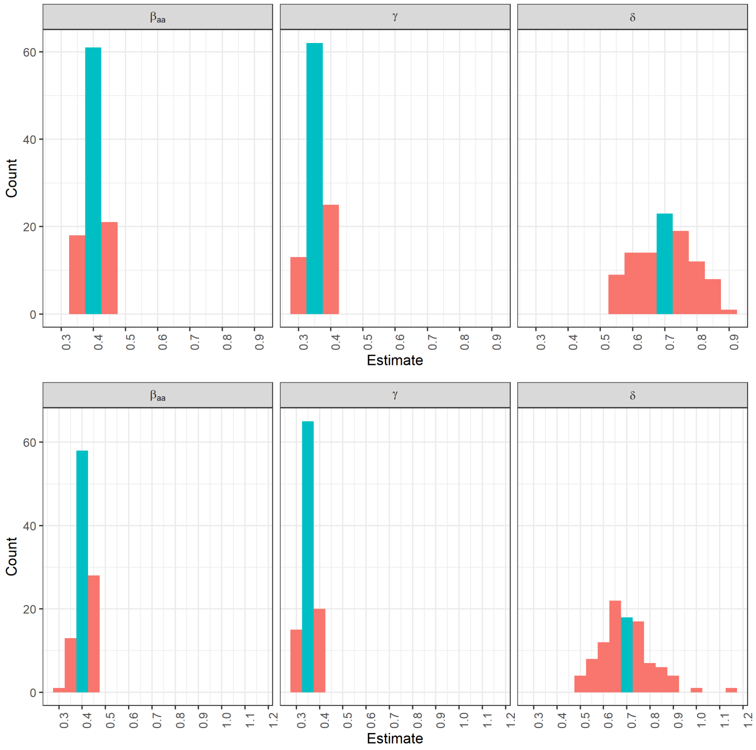


Figure S7: Results for data sets with mean generation-time of 4 days (top row) and mean generation-time of 5 days (bottom row). $\beta_{aa}$ - the transmission parameter among adults, $\gamma$ - relative susceptability of children, $\delta$ - relative infectivity of children.

#### 6 Positive rates per 10,000 in different age groups

Figure S8 shows the positive rates per 10,000 in different age groups through time. Since the re-opening of schools on September 1st the infection rates in children aged 15-19 has more than doubled, and seem to have triggered growing infection rates in other age groups as well.

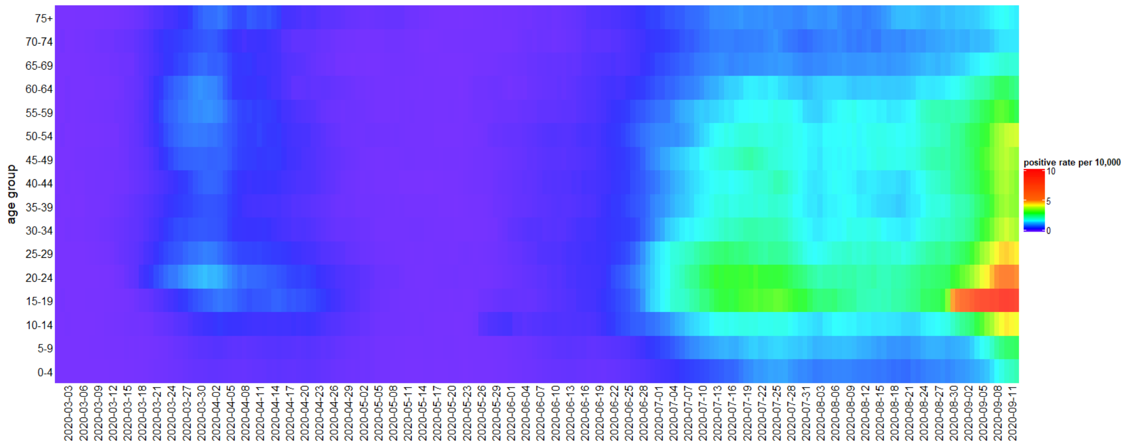


Figure S8: Positive rates per 10,000 per age group through time.
