## Supplementary material for "The role of children in the spread of COVID-19: Using household data from Bnei Brak, Israel, to estimate the relative susceptibility and infectivity of children": Household timelines

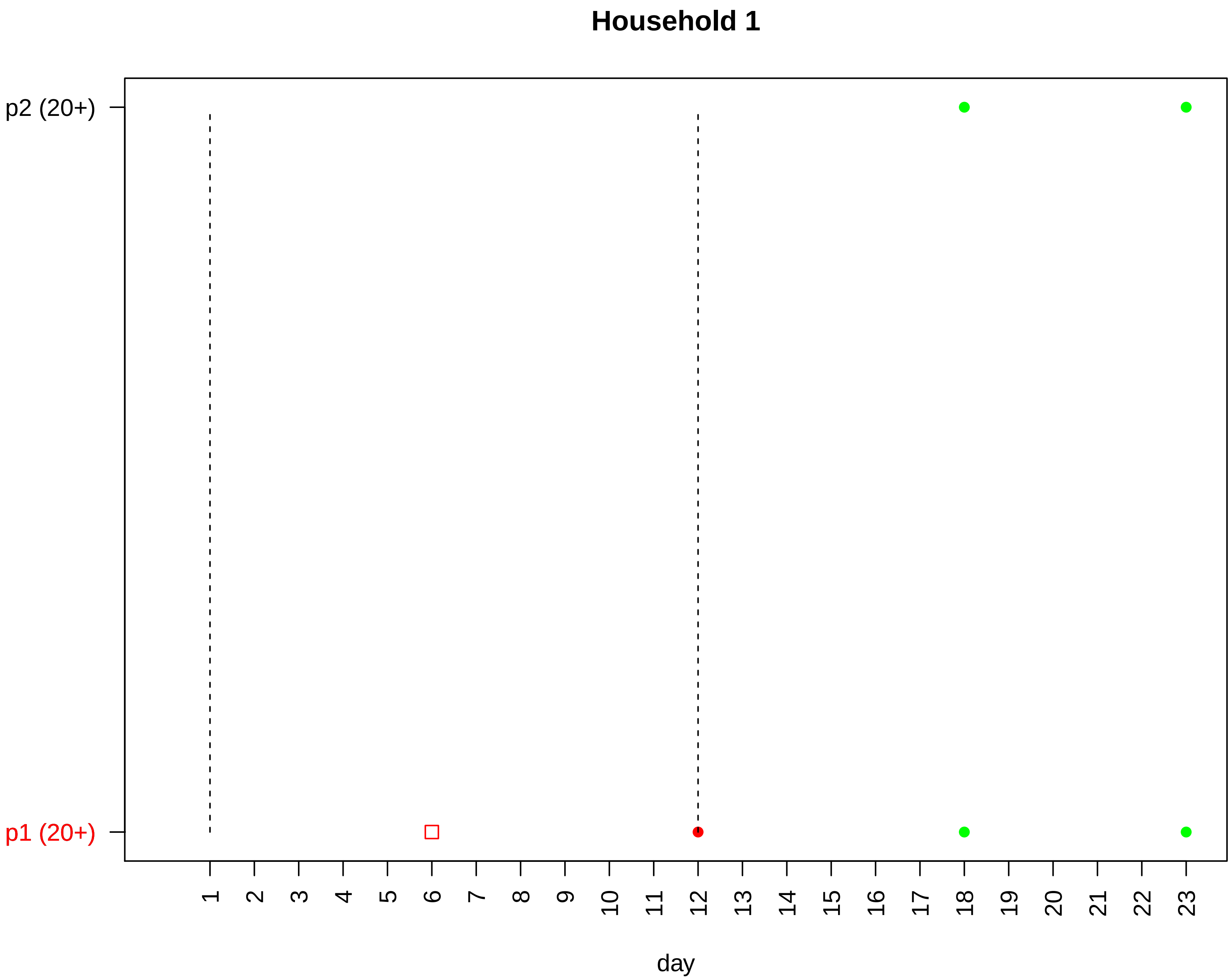

Household 1
p2 (20+)
p1 (20+)
day

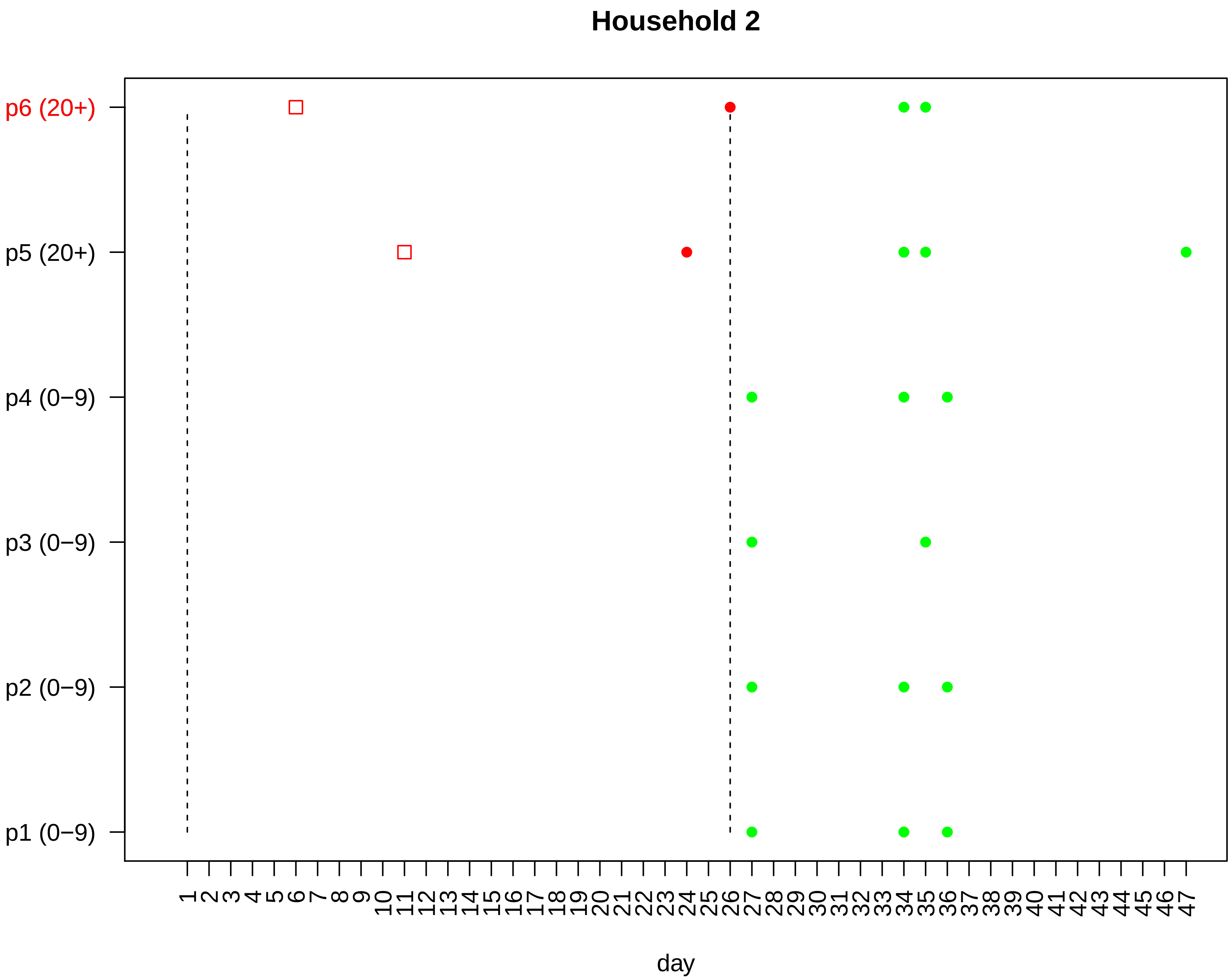
Household 2
p6 (20+)
p5 (20+)
p4 (0−9)
p3 (0−9)
p2 (0−9)
p1 (0−9)
1 2 3 4 5 6 7 8 9 10 11 12 13 14 15 16 17 18 19 20 21 22 23 24 25 26 27 28 29 30 31 32 33 34 35 36 37 38 39 40 41 42 43 44 45 46 47
day

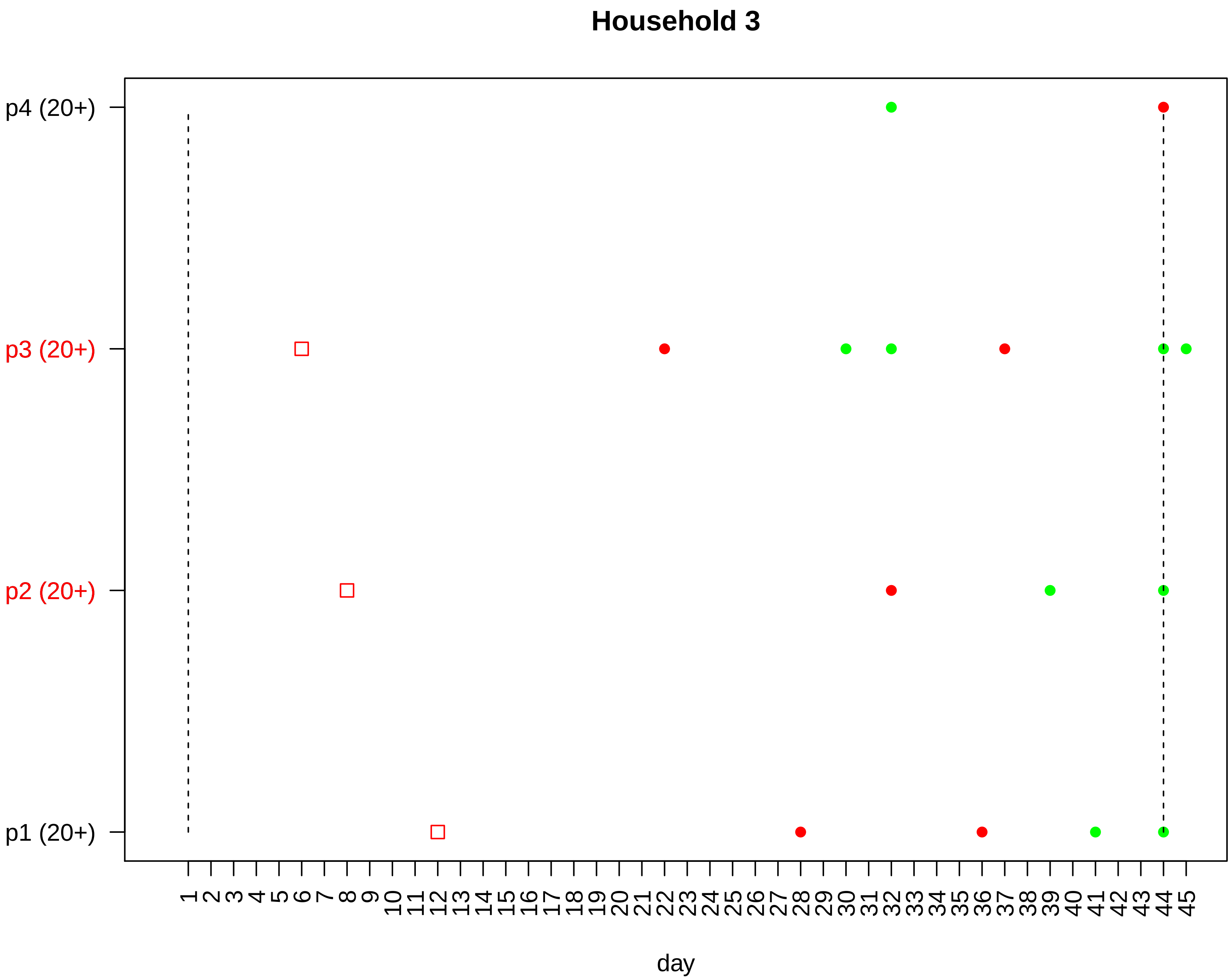

Household 3
p4 (20+)
p3 (20+)
p2 (20+)
p1 (20+)
1 2 3 4 5 6 7 8 9 10 11 12 13 14 15 16 17 18 19 20 21 22 23 24 25 26 27 28 29 30 31 32 33 34 35 36 37 38 39 40 41 42 43 44 45
day

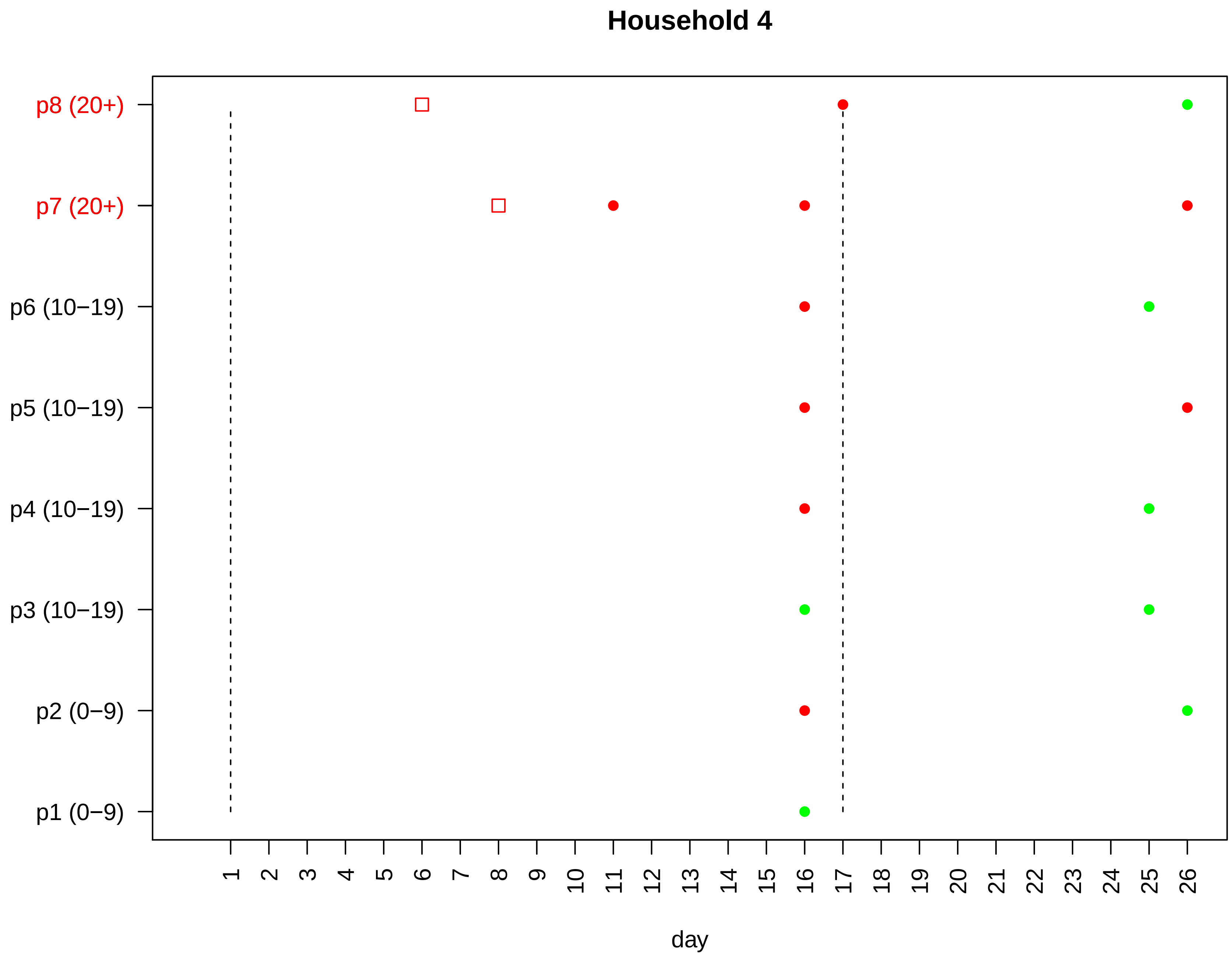

Household 4
p8 (20+)
p7 (20+)
p6 (10−19)
p5 (10−19)
p4 (10−19)
p3 (10−19)
p2 (0−9)
p1 (0−9)
1 2 3 4 5 6 7 8 9 10 11 12 13 14 15 16 17 18 19 20 21 22 23 24 25 26
day

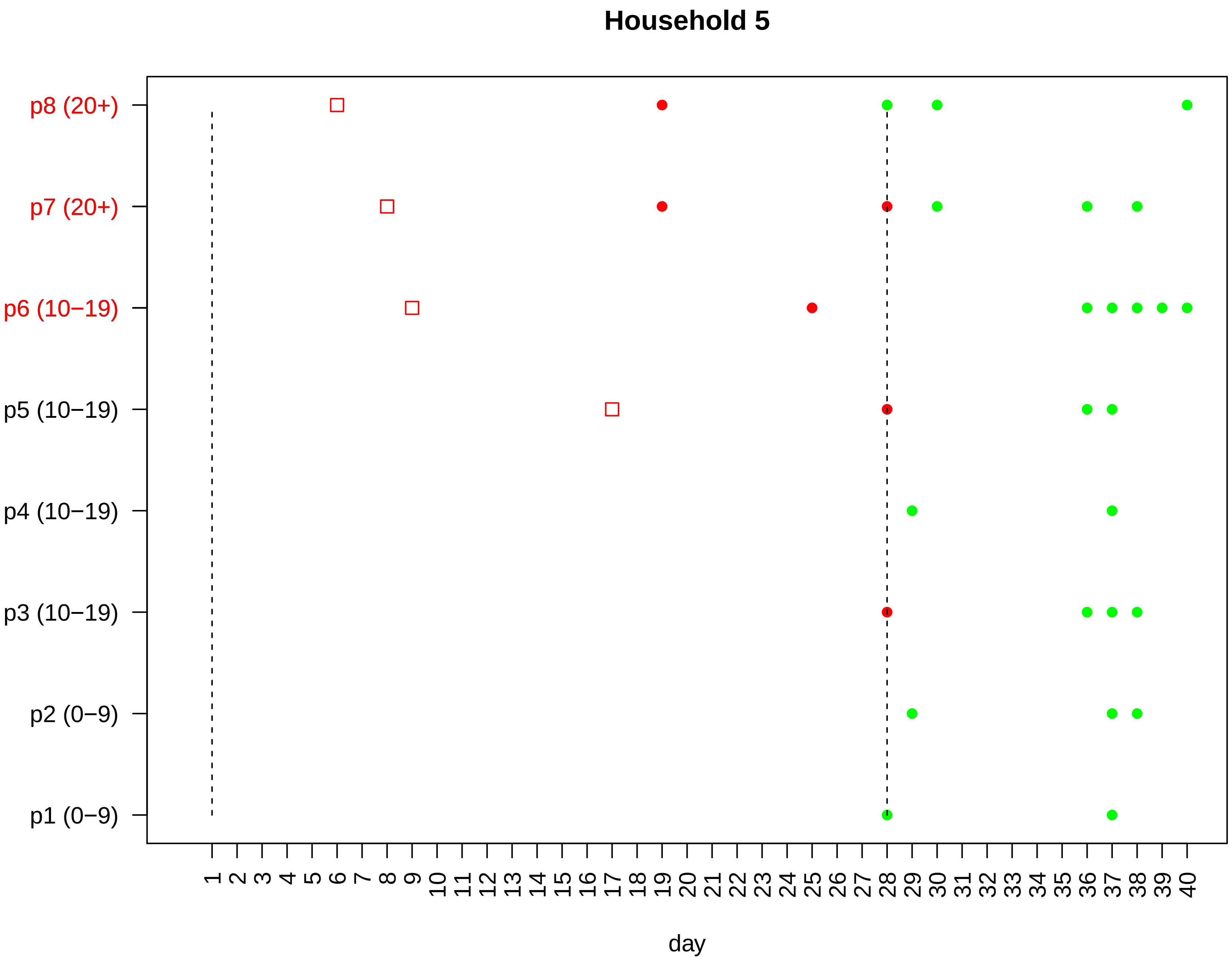

Household 5
p8 (20+)
p7 (20+)
p6 (10−19)
p5 (10−19)
p4 (10−19)
p3 (10−19)
p2 (0−9)
p1 (0−9)
1 2 3 4 5 6 7 8 9 10 11 12 13 14 15 16 17 18 19 20 21 22 23 24 25 26 27 28 29 30 31 32 33 34 35 36 37 38 39 40
day

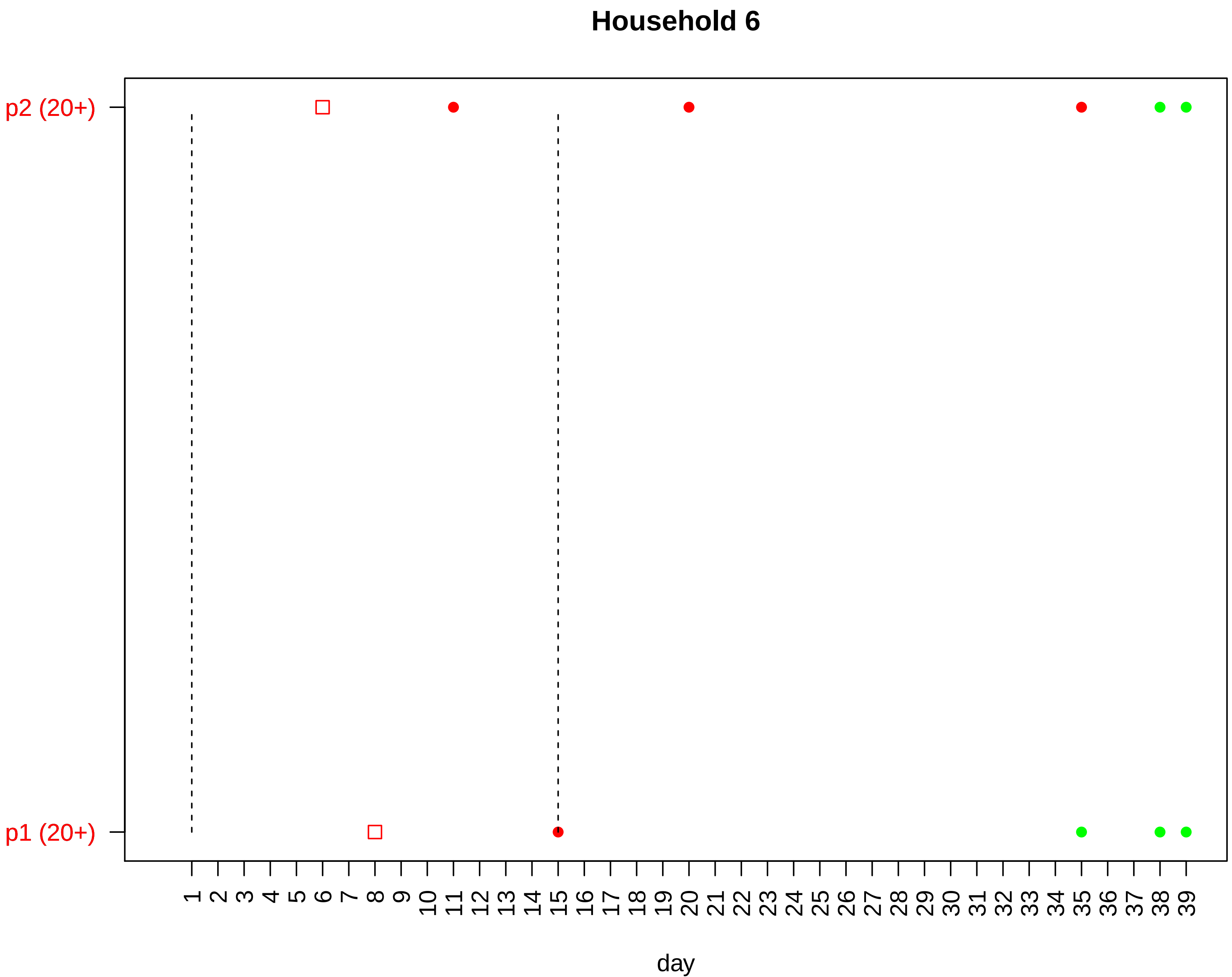
Household 6
p2 (20+)
p1 (20+)
day

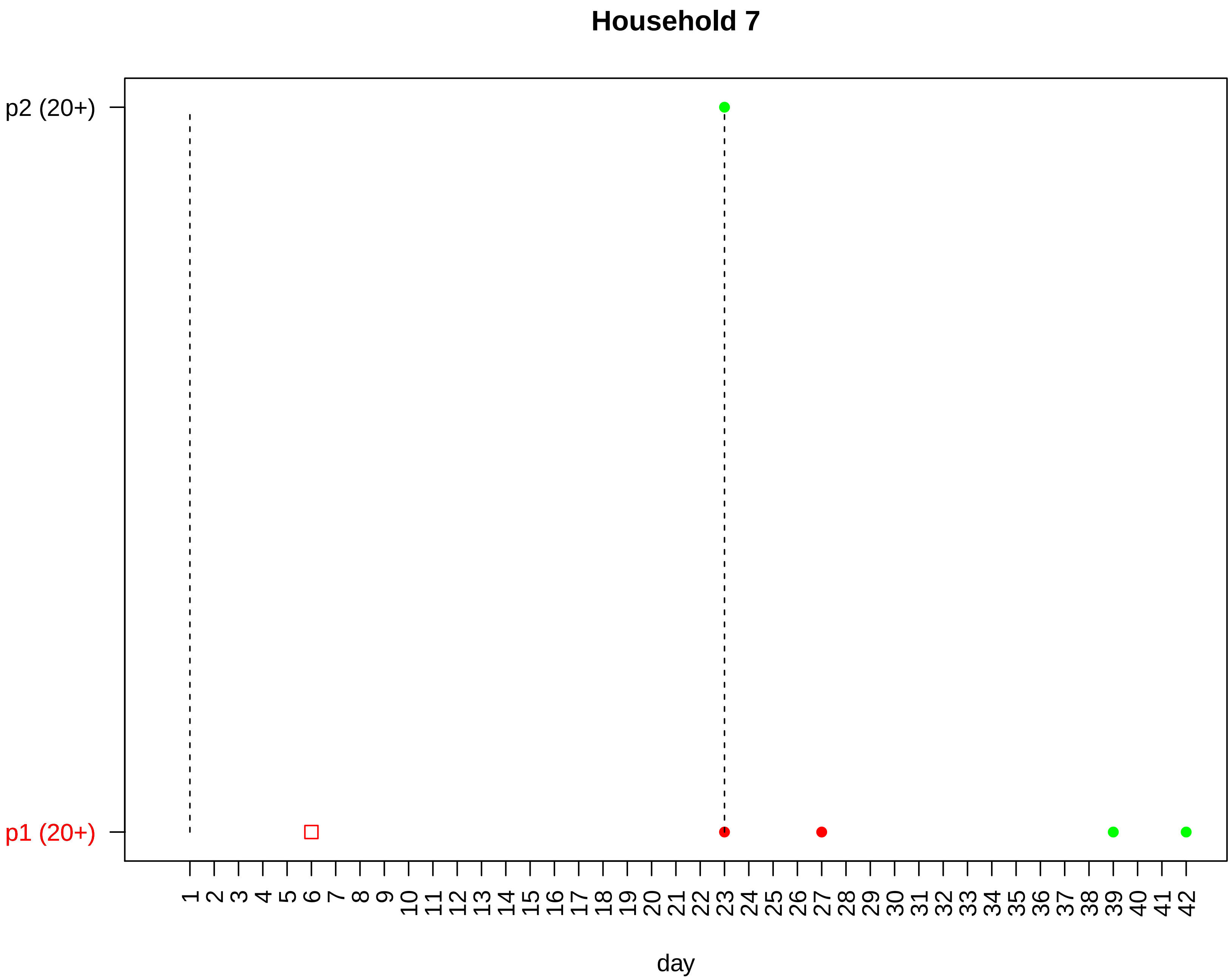

Household 7
p2 (20+)
p1 (20+)
1 2 3 4 5 6 7 8 9 10 11 12 13 14 15 16 17 18 19 20 21 22 23 24 25 26 27 28 29 30 31 32 33 34 35 36 37 38 39 40 41 42
day

#### Household 8

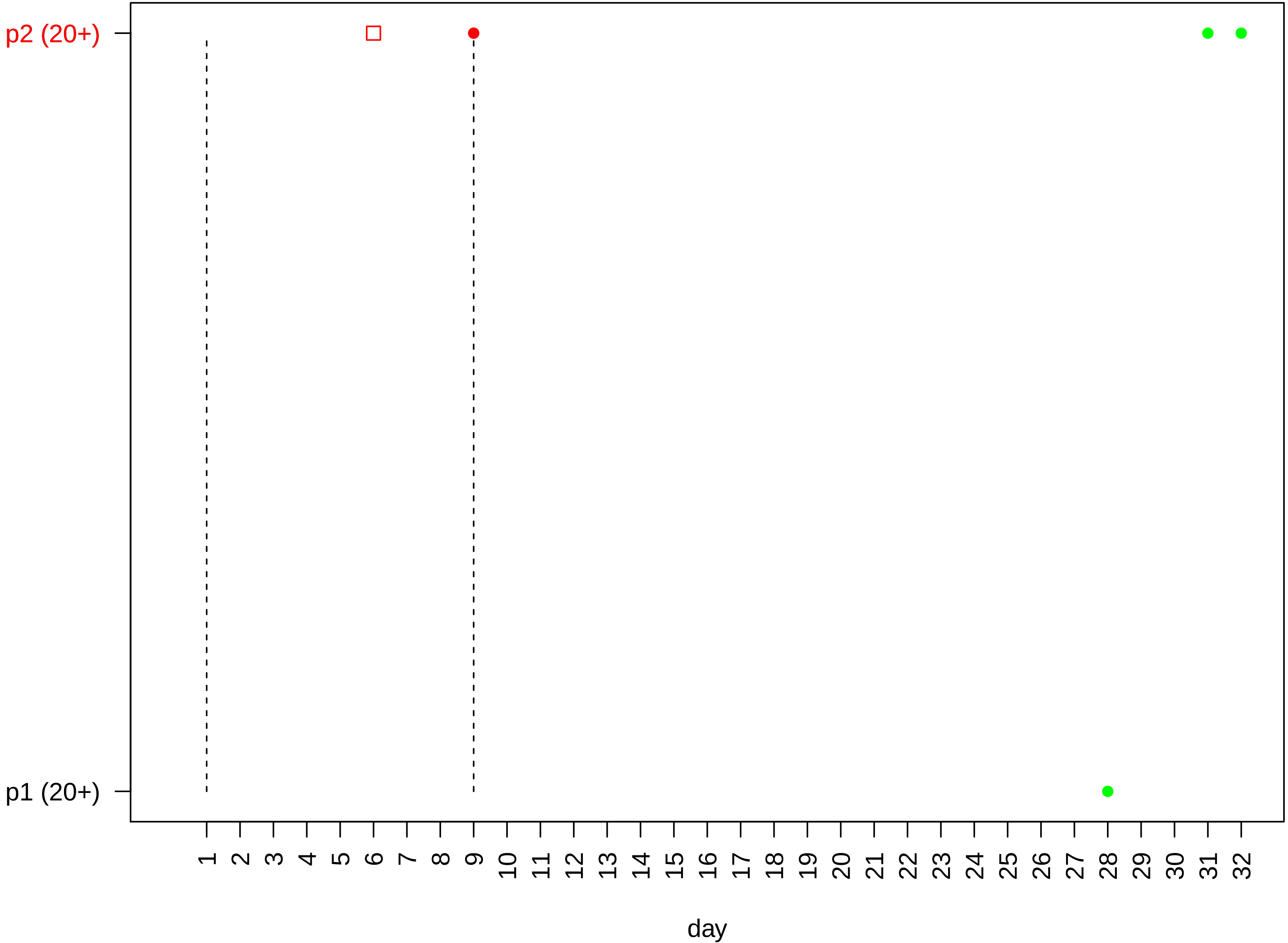

p2 (20+)
p1 (20+)
1 2 3 4 5 6 7 8 9 10 11 12 13 14 15 16 17 18 19 20 21 22 23 24 25 26 27 28 29 30 31 32
day

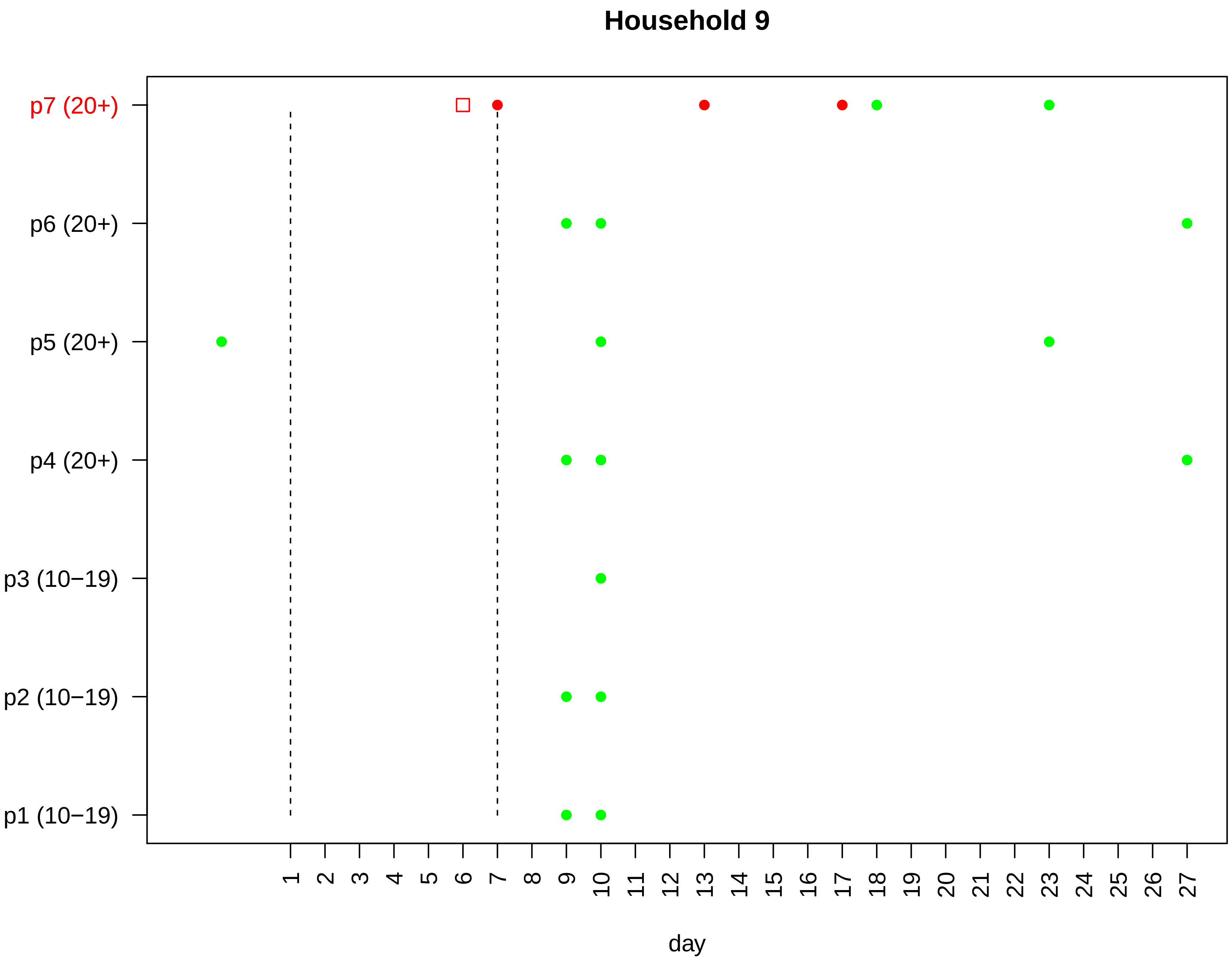

Household 9
p7 (20+)
p6 (20+)
p5 (20+)
p4 (20+)
p3 (10−19)
p2 (10−19)
p1 (10−19)
1 2 3 4 5 6 7 8 9 10 11 12 13 14 15 16 17 18 19 20 21 22 23 24 25 26 27
day

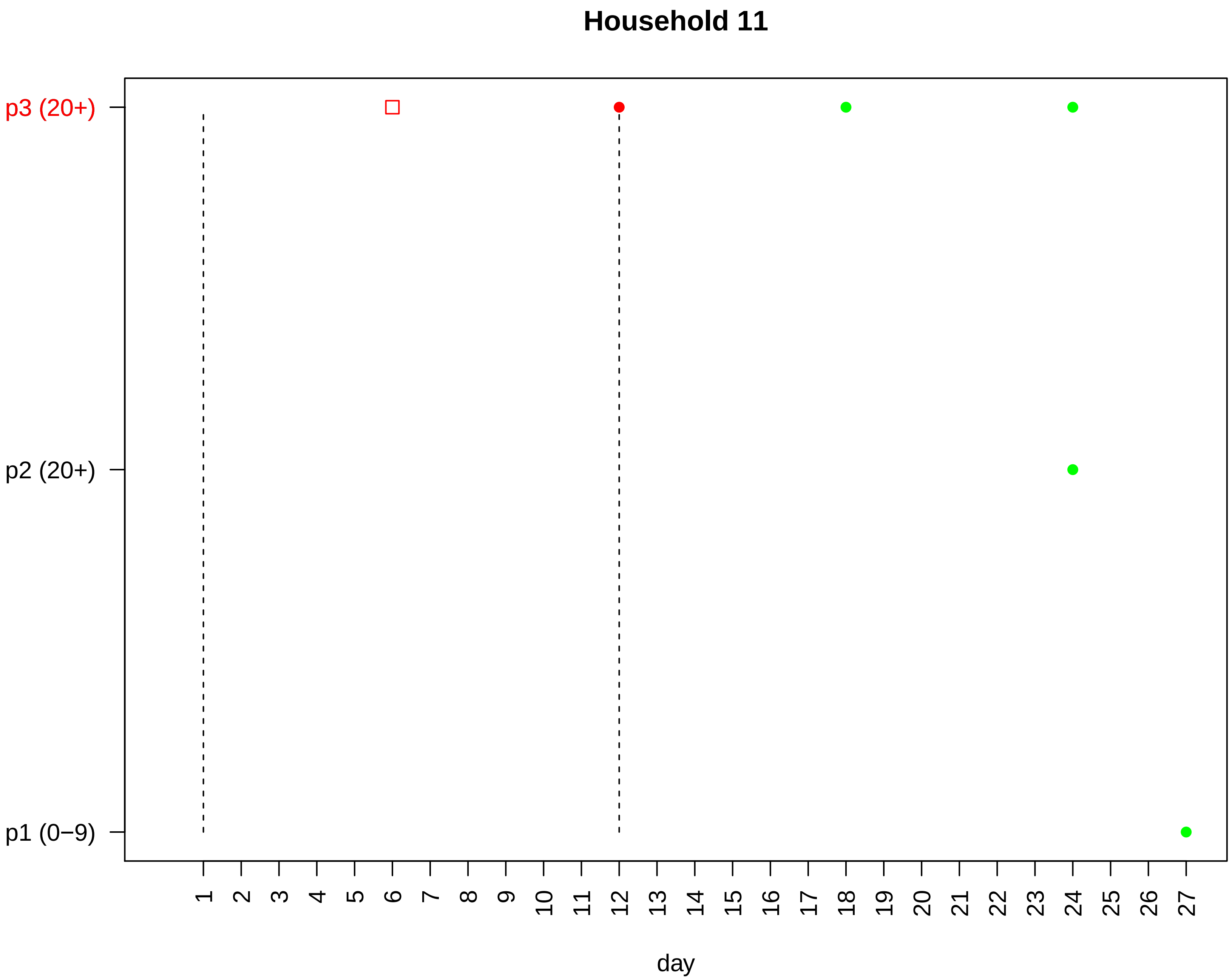

Household 11
p3 (20+)
p2 (20+)
p1 (0−9)
day

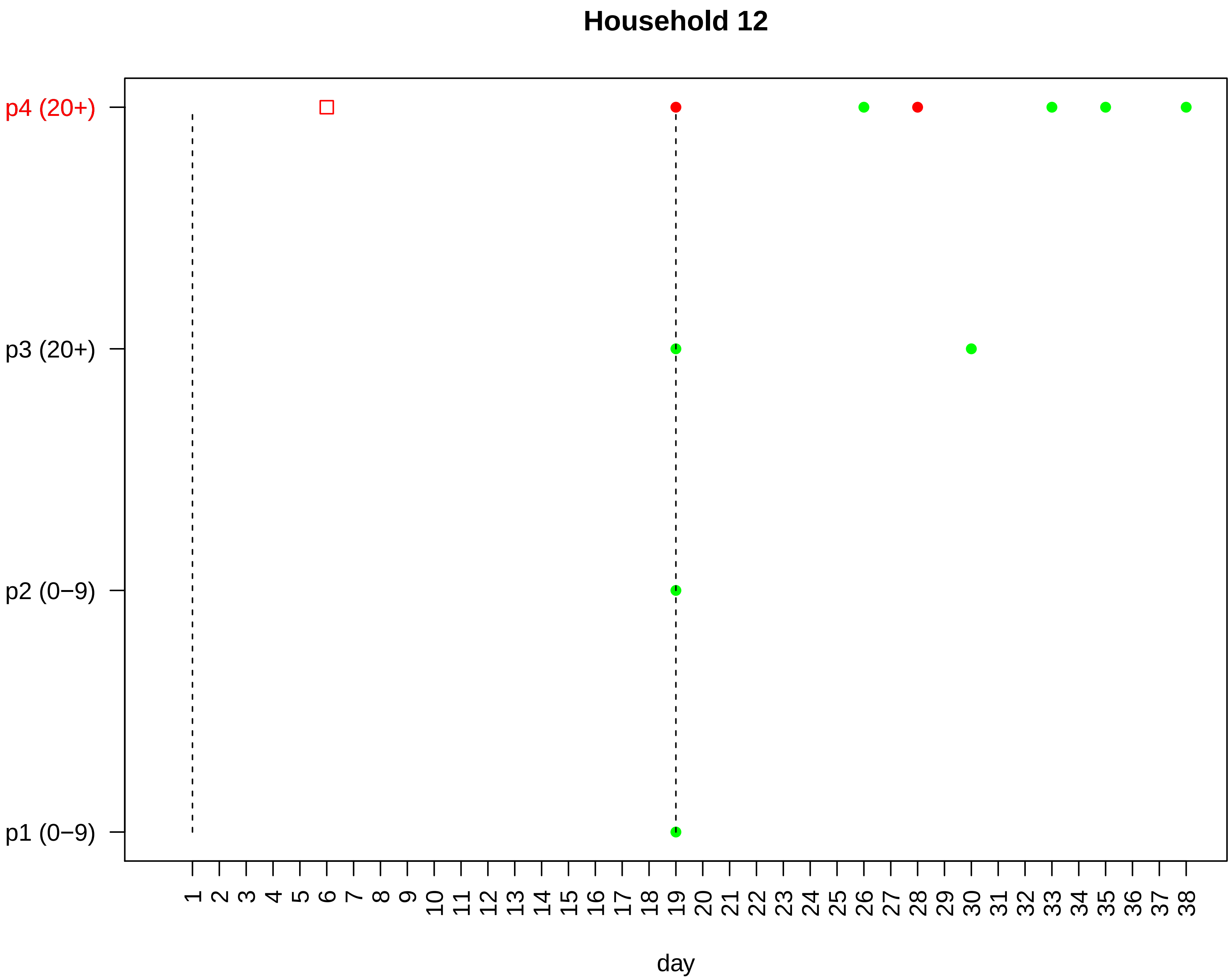

Household 12
p4 (20+)
p3 (20+)
p2 (0−9)
p1 (0−9)
1 2 3 4 5 6 7 8 9 10 11 12 13 14 15 16 17 18 19 20 21 22 23 24 25 26 27 28 29 30 31 32 33 34 35 36 37 38
day

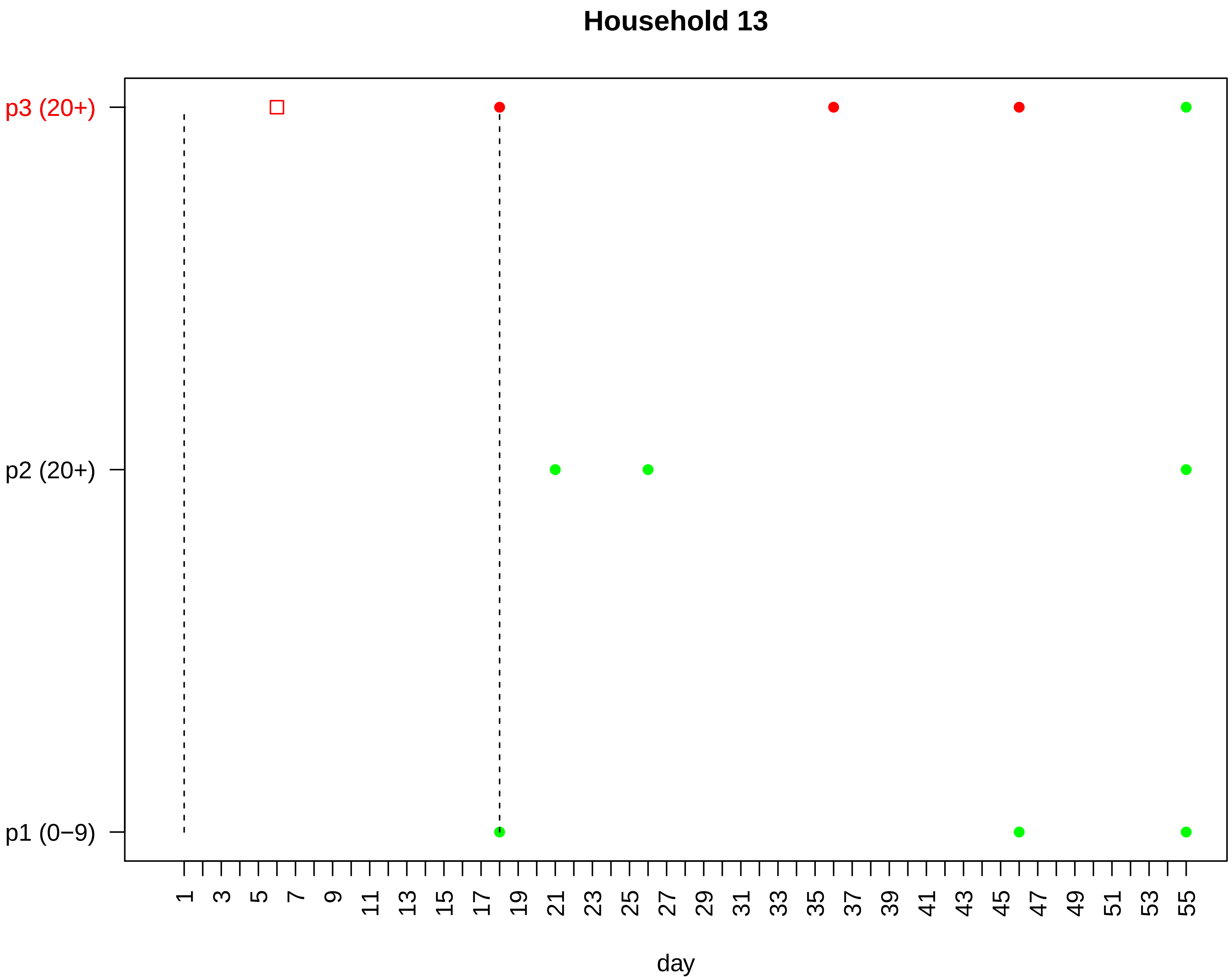
Household 13
p3 (20+)
p2 (20+)
p1 (0−9)
1
3
5
7
9
11
13
15
17
19
21
23
25
27
29
31
33
35
37
39
41
43
45
47
49
51
53
55
day

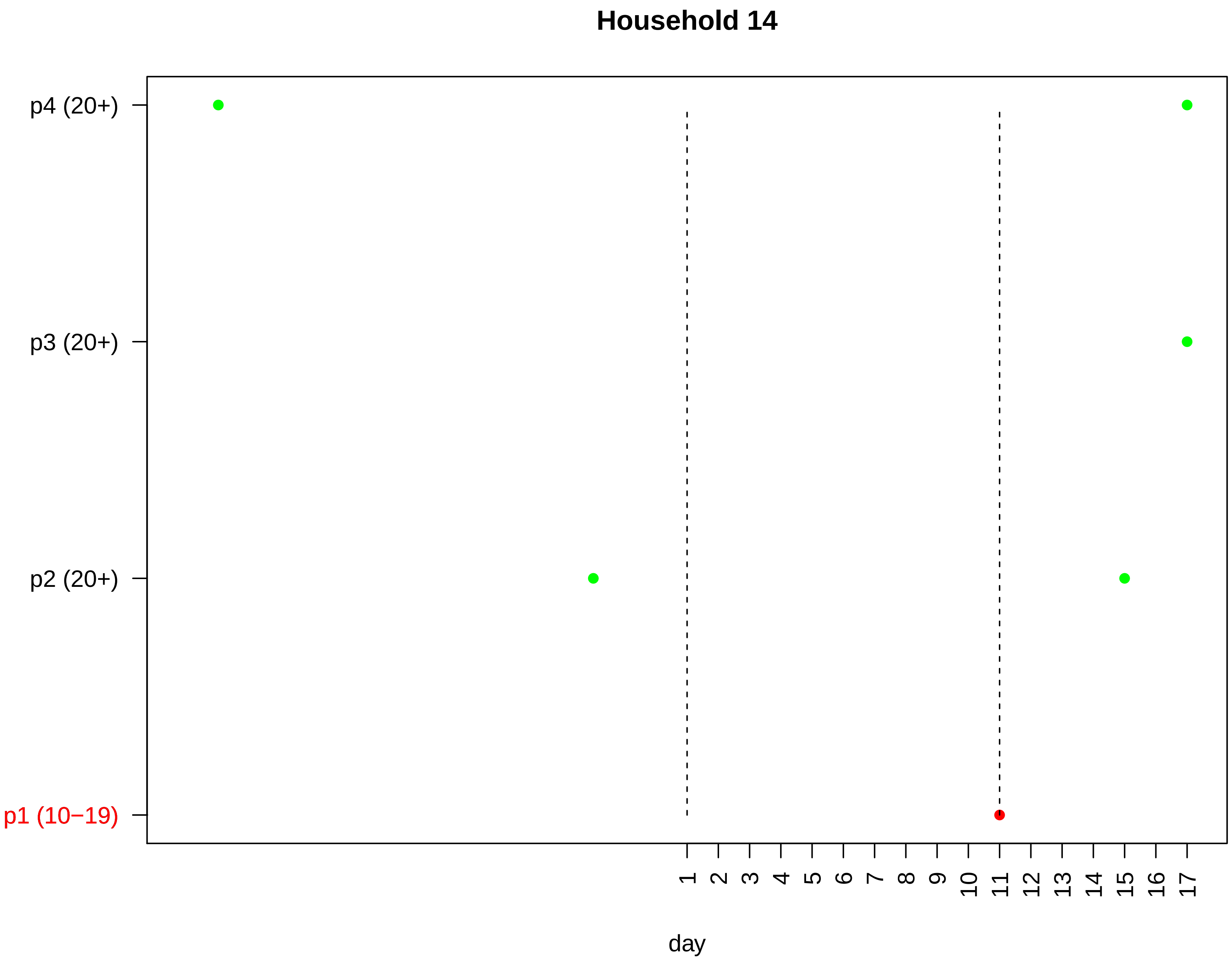

Household 14
p4 (20+)
p3 (20+)
p2 (20+)
p1 (10−19)
1 2 3 4 5 6 7 8 9 10 11 12 13 14 15 16 17
day

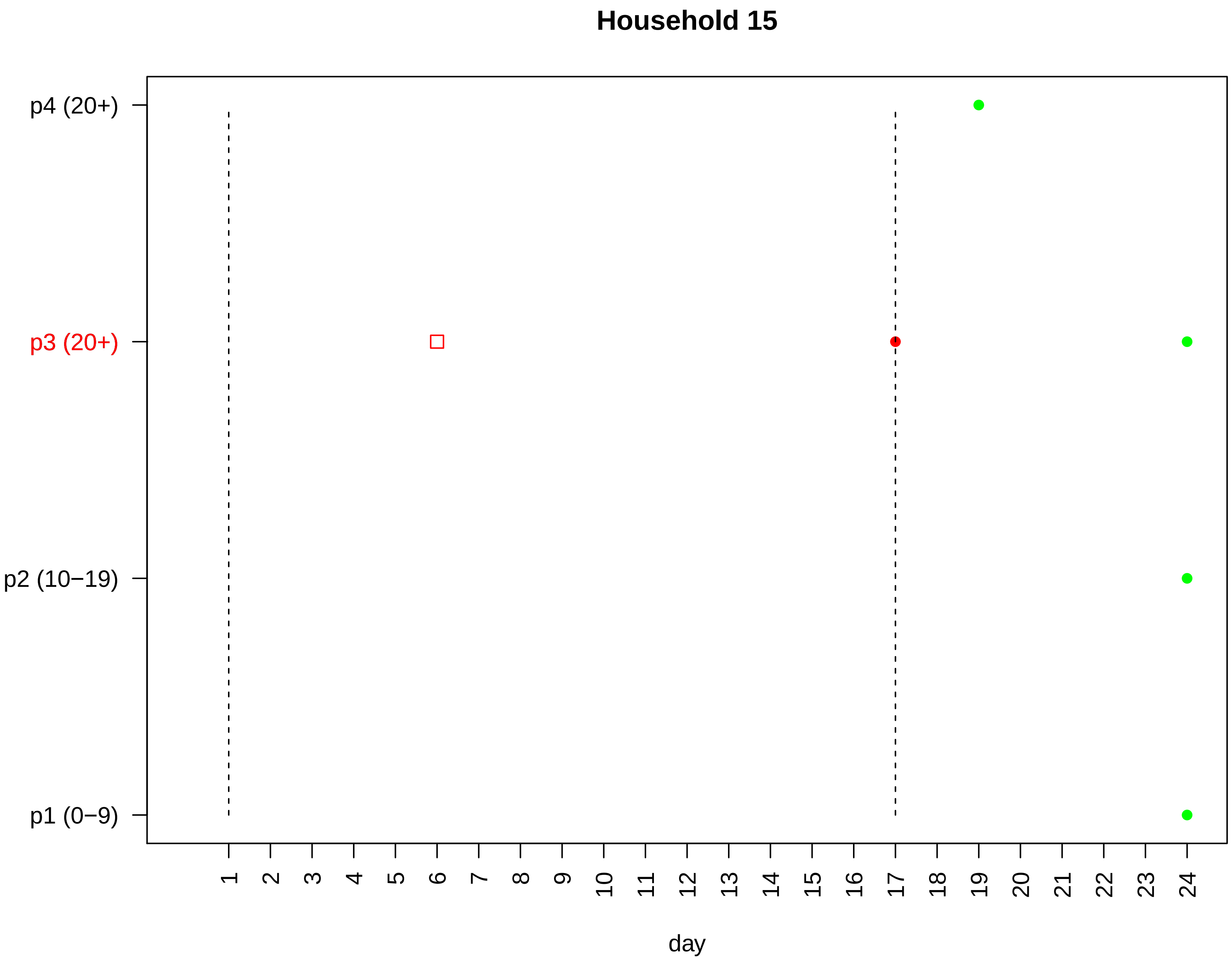

Household 15
p4 (20+)
p3 (20+)
p2 (10−19)
p1 (0−9)
day

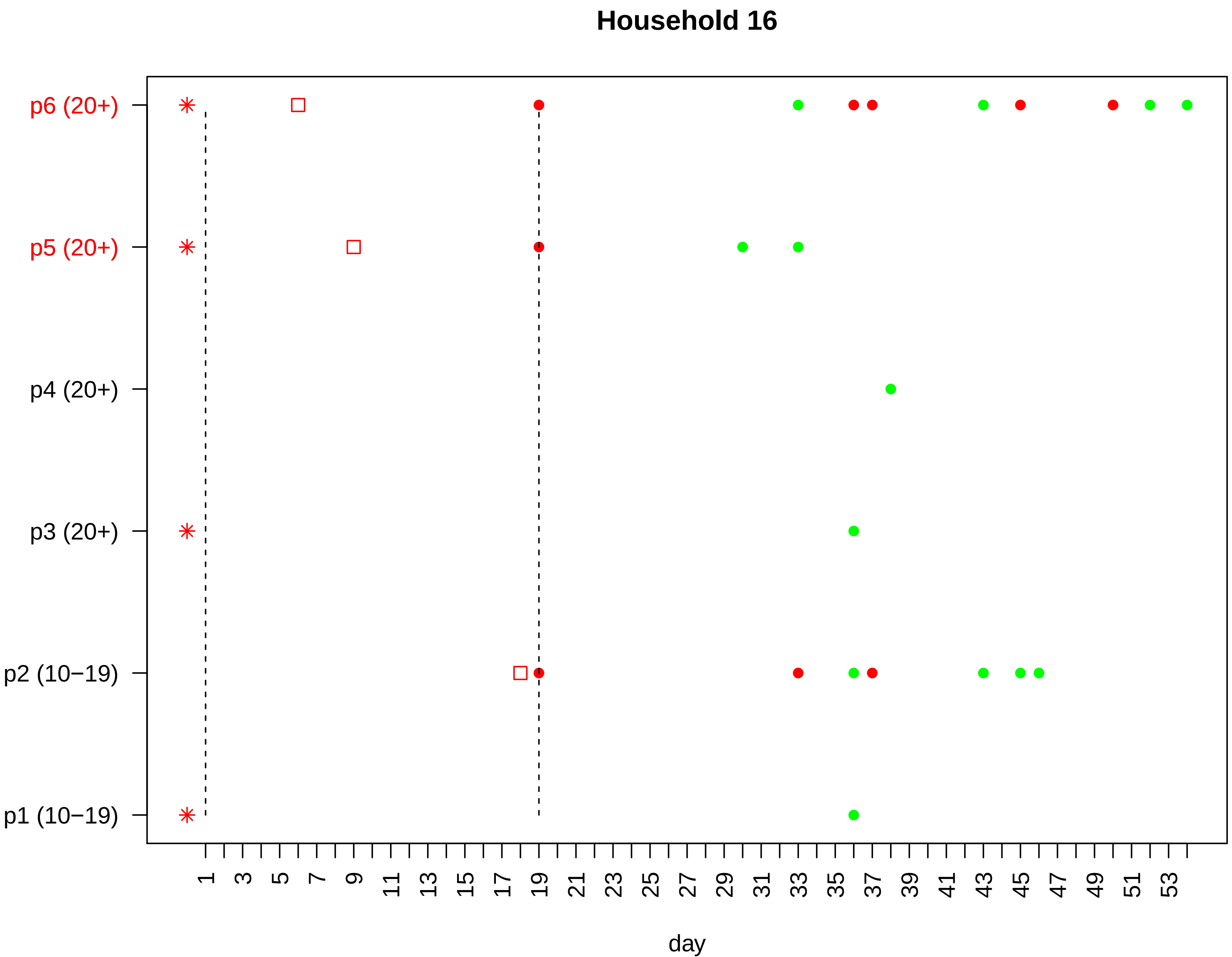

Household 16
p6 (20+)
p5 (20+)
p4 (20+)
p3 (20+)
p2 (10−19)
p1 (10−19)
1 3 5 7 9 11 13 15 17 19 21 23 25 27 29 31 33 35 37 39 41 43 45 47 49 51 53
day

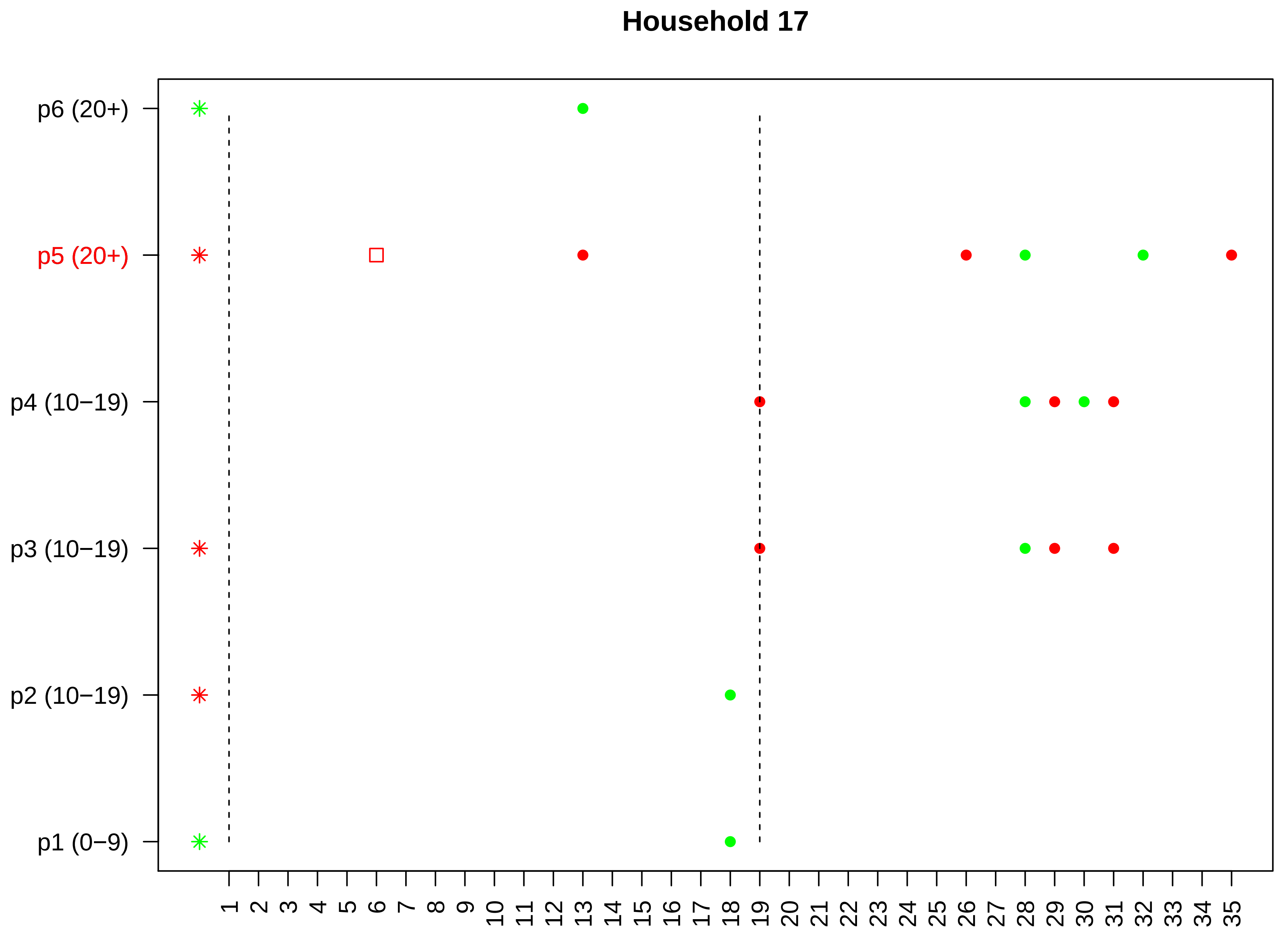

Household 17
p6 (20+)
p5 (20+)
p4 (10−19)
p3 (10−19)
p2 (10−19)
p1 (0−9)
1 2 3 4 5 6 7 8 9 10 11 12 13 14 15 16 17 18 19 20 21 22 23 24 25 26 27 28 29 30 31 32 33 34 35
day

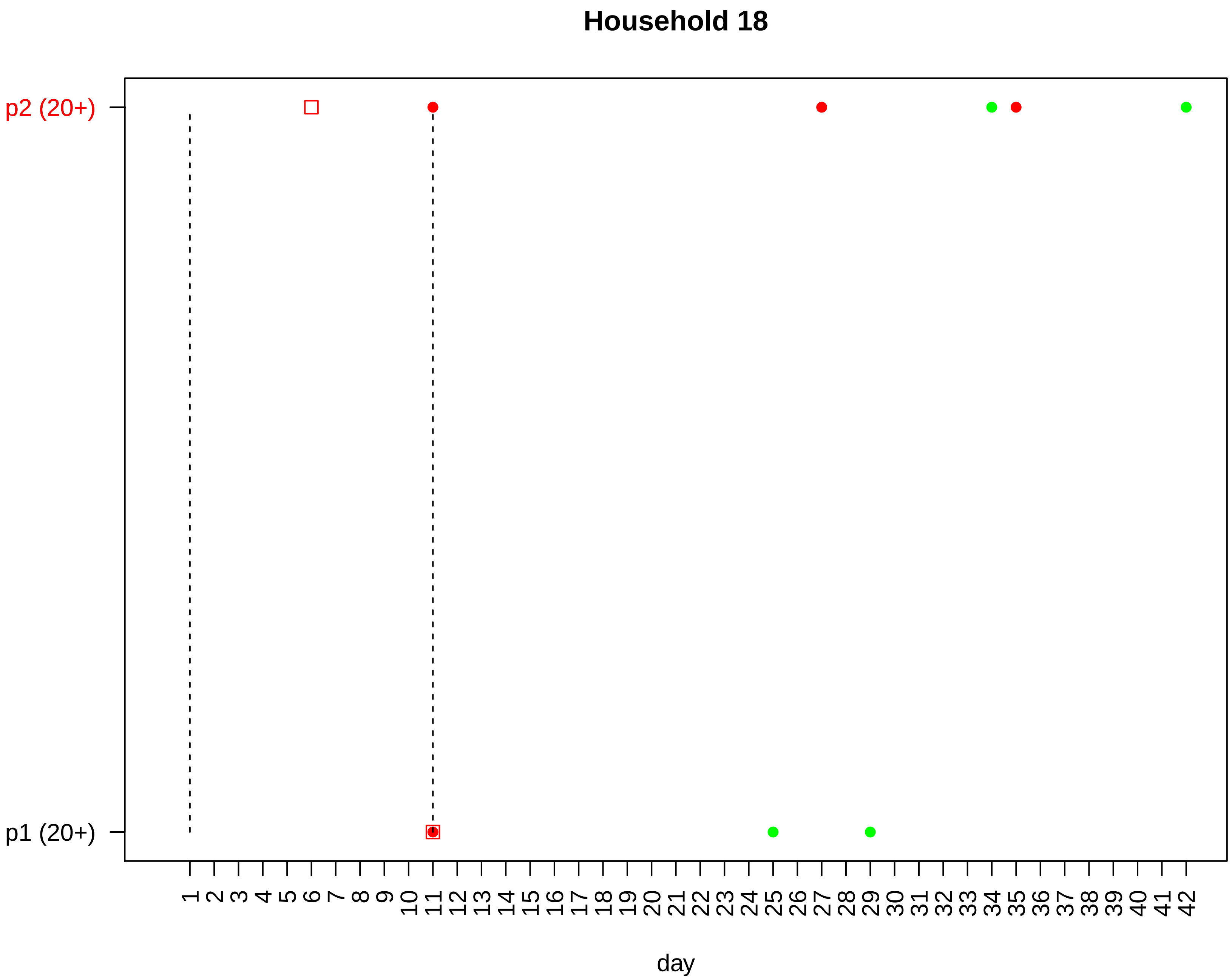

Household 18
p2 (20+)
p1 (20+)
1 2 3 4 5 6 7 8 9 10 11 12 13 14 15 16 17 18 19 20 21 22 23 24 25 26 27 28 29 30 31 32 33 34 35 36 37 38 39 40 41 42
day

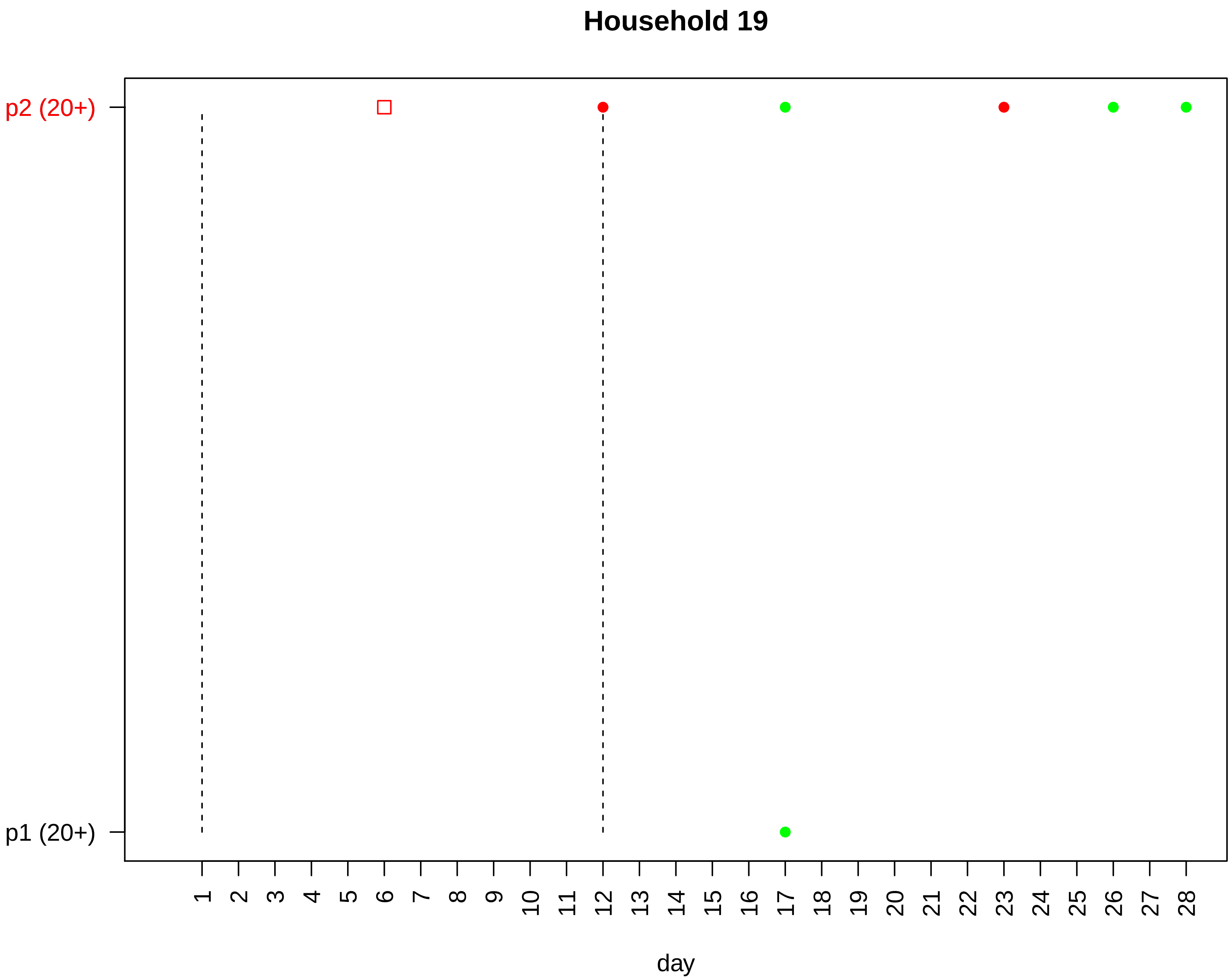
Household 19
p2 (20+)
p1 (20+)
day

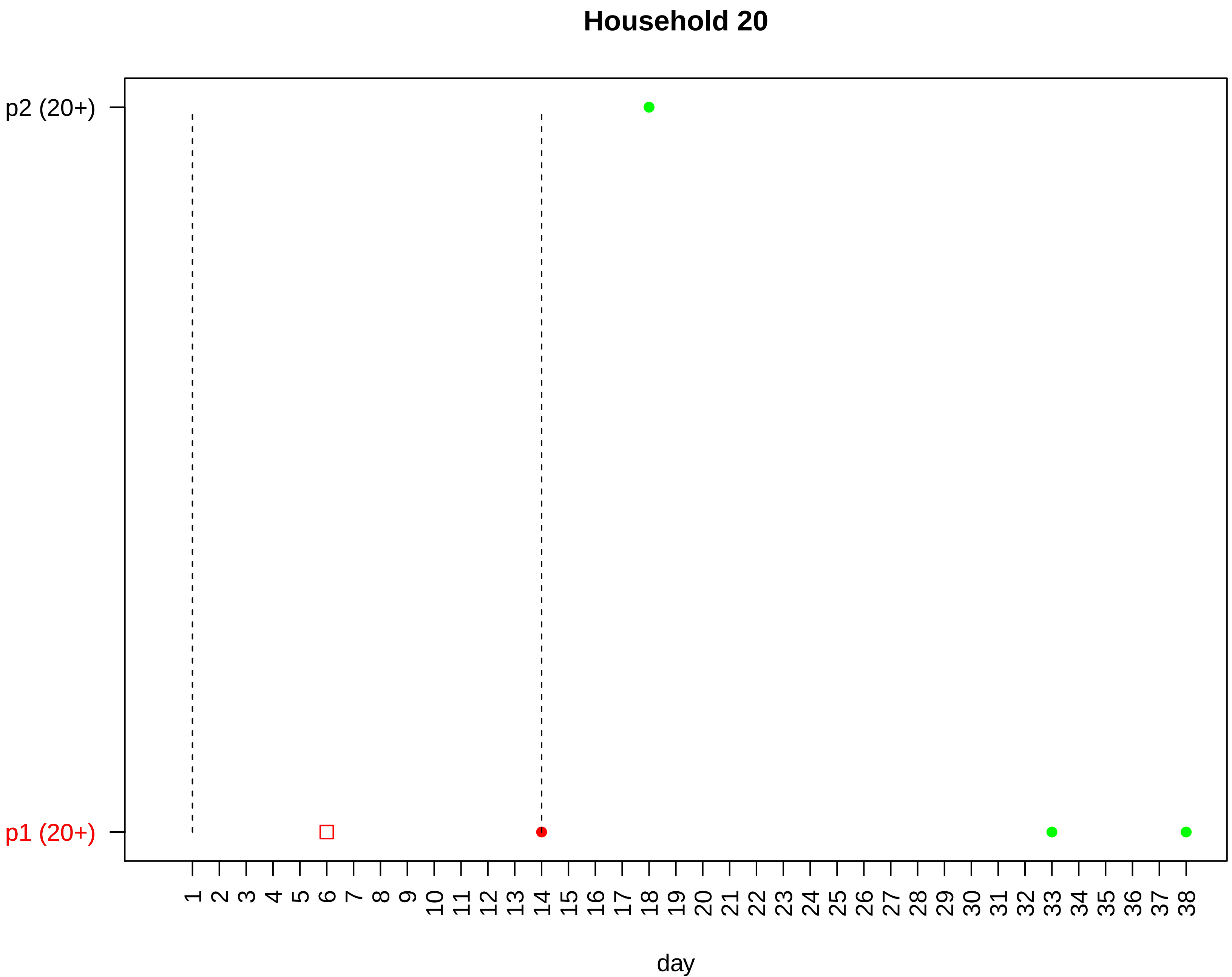
Household 20
p2 (20+)
p1 (20+)
1 2 3 4 5 6 7 8 9 10 11 12 13 14 15 16 17 18 19 20 21 22 23 24 25 26 27 28 29 30 31 32 33 34 35 36 37 38
day

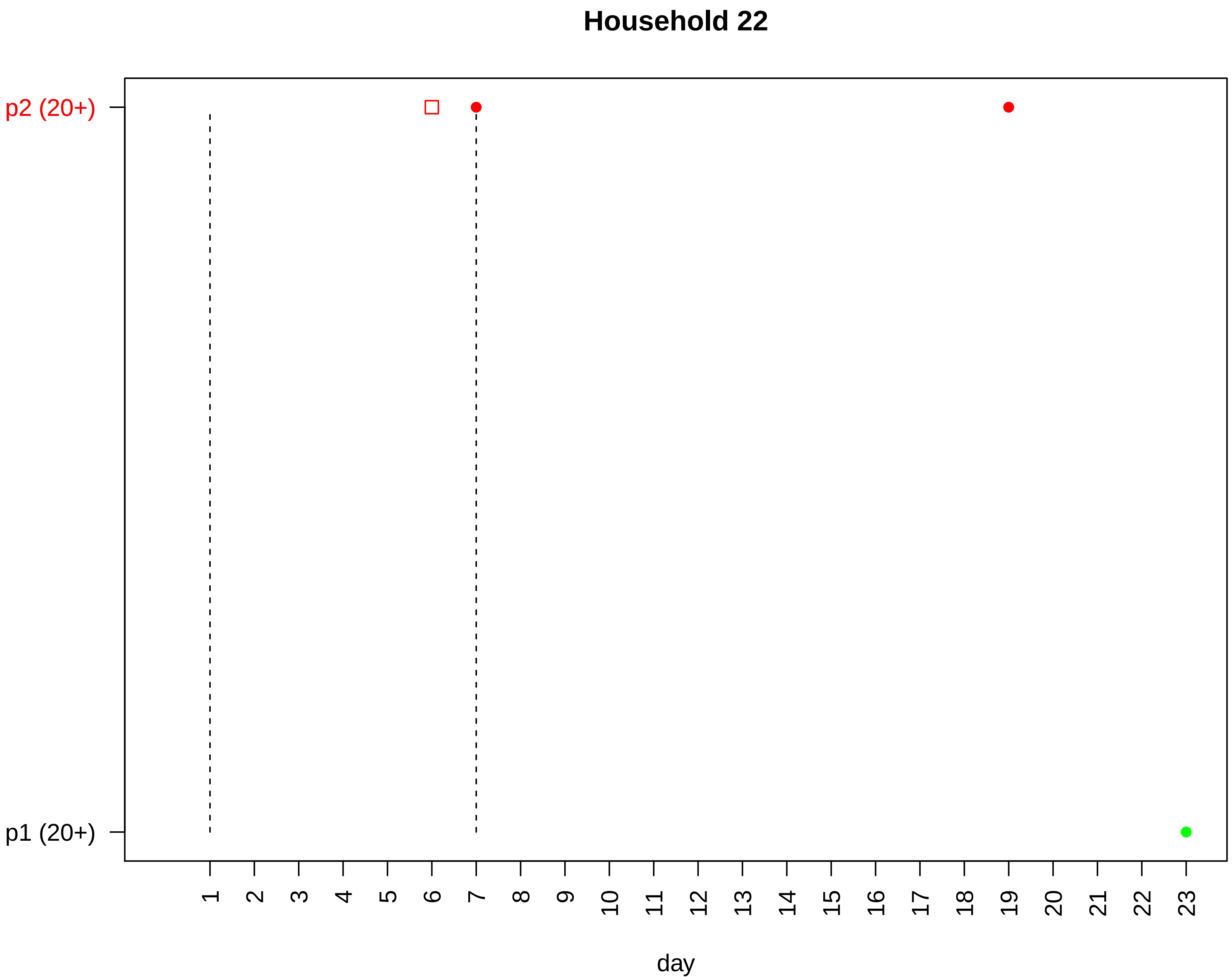

Household 22
p2 (20+)
p1 (20+)
day

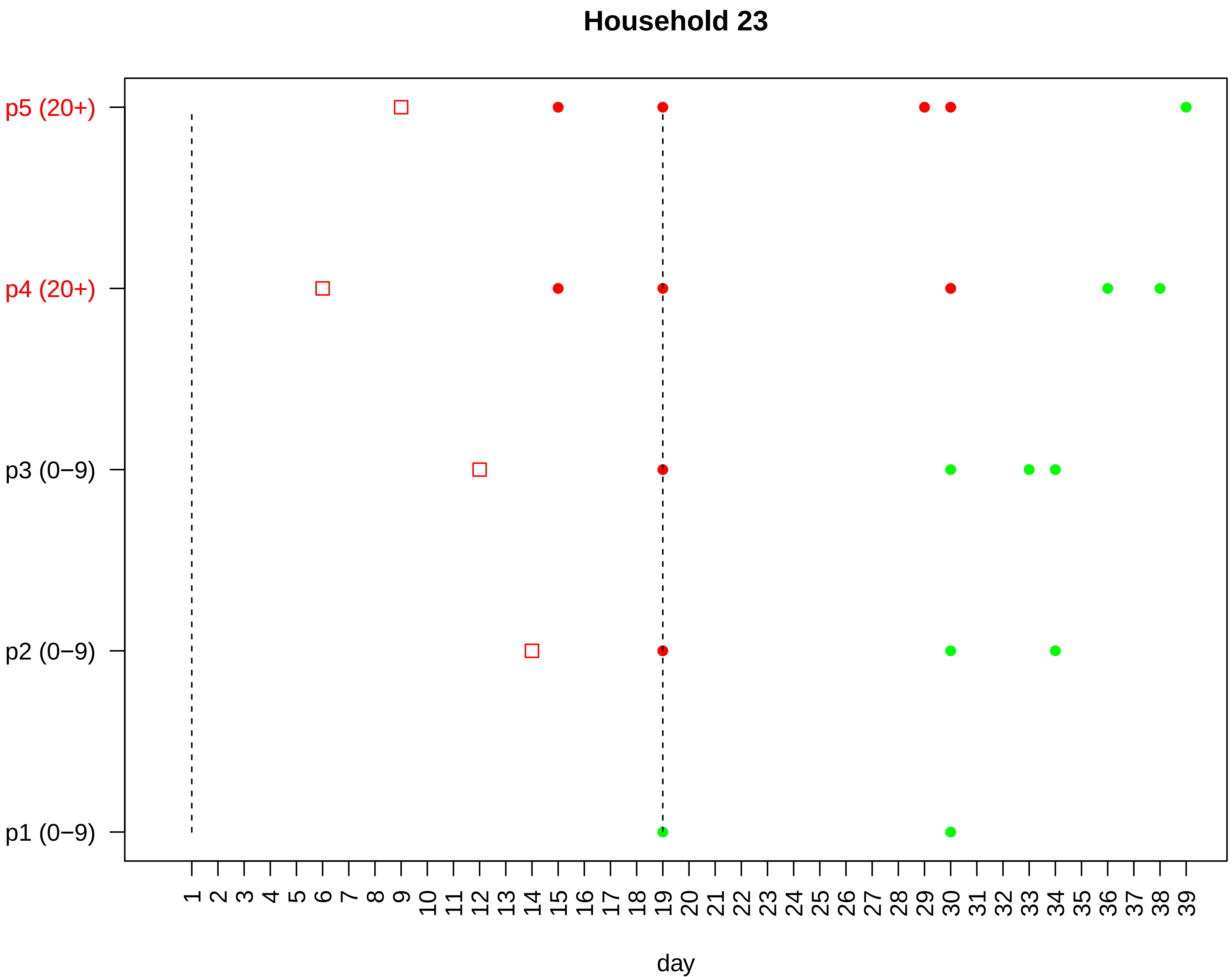

Household 23
p5 (20+)
p4 (20+)
p3 (0−9)
p2 (0−9)
p1 (0−9)
1 2 3 4 5 6 7 8 9 10 11 12 13 14 15 16 17 18 19 20 21 22 23 24 25 26 27 28 29 30 31 32 33 34 35 36 37 38 39
day

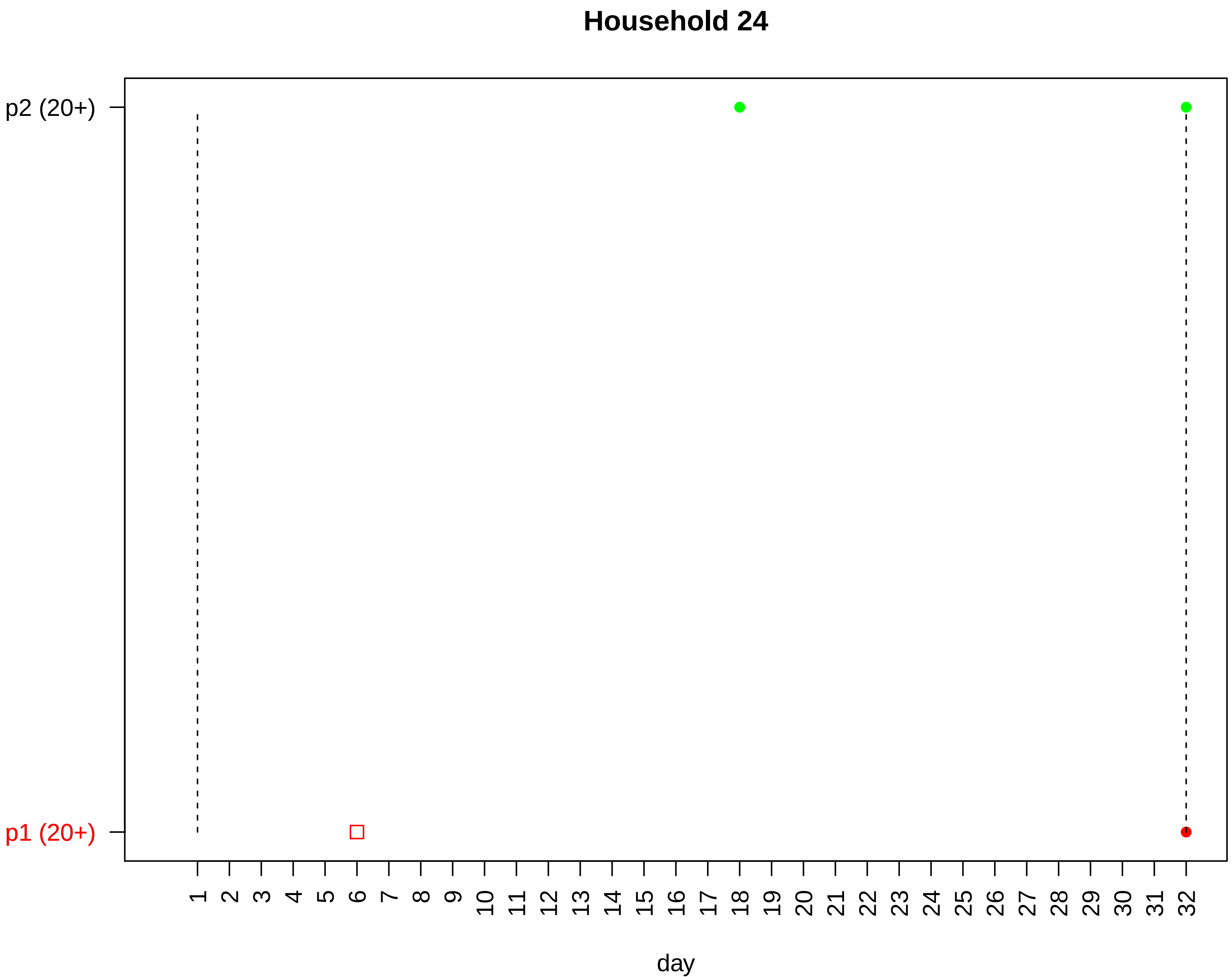
Household 24
p2 (20+)
p1 (20+)
1 2 3 4 5 6 7 8 9 10 11 12 13 14 15 16 17 18 19 20 21 22 23 24 25 26 27 28 29 30 31 32
day

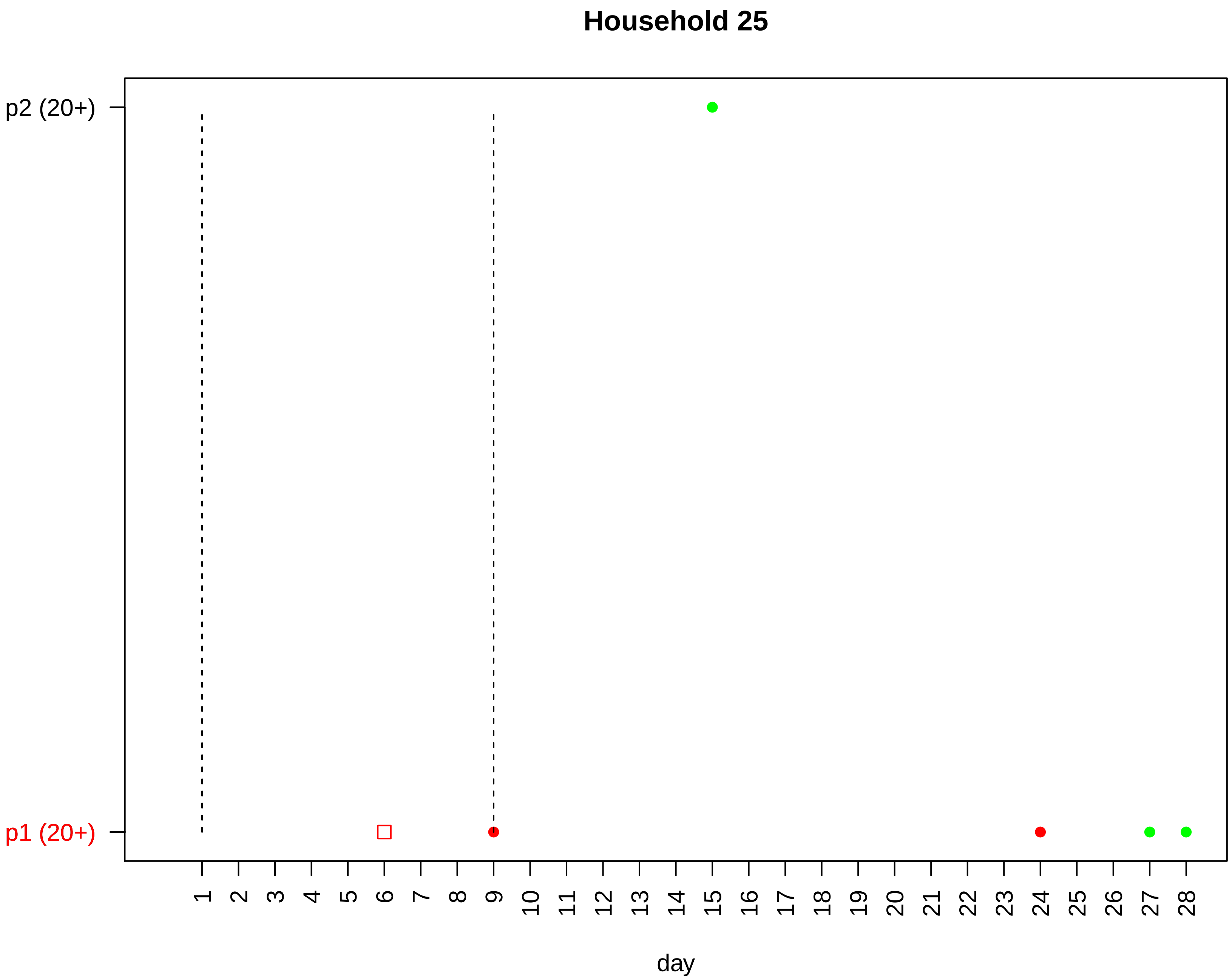
Household 25
p2 (20+)
p1 (20+)
1 2 3 4 5 6 7 8 9 10 11 12 13 14 15 16 17 18 19 20 21 22 23 24 25 26 27 28
day

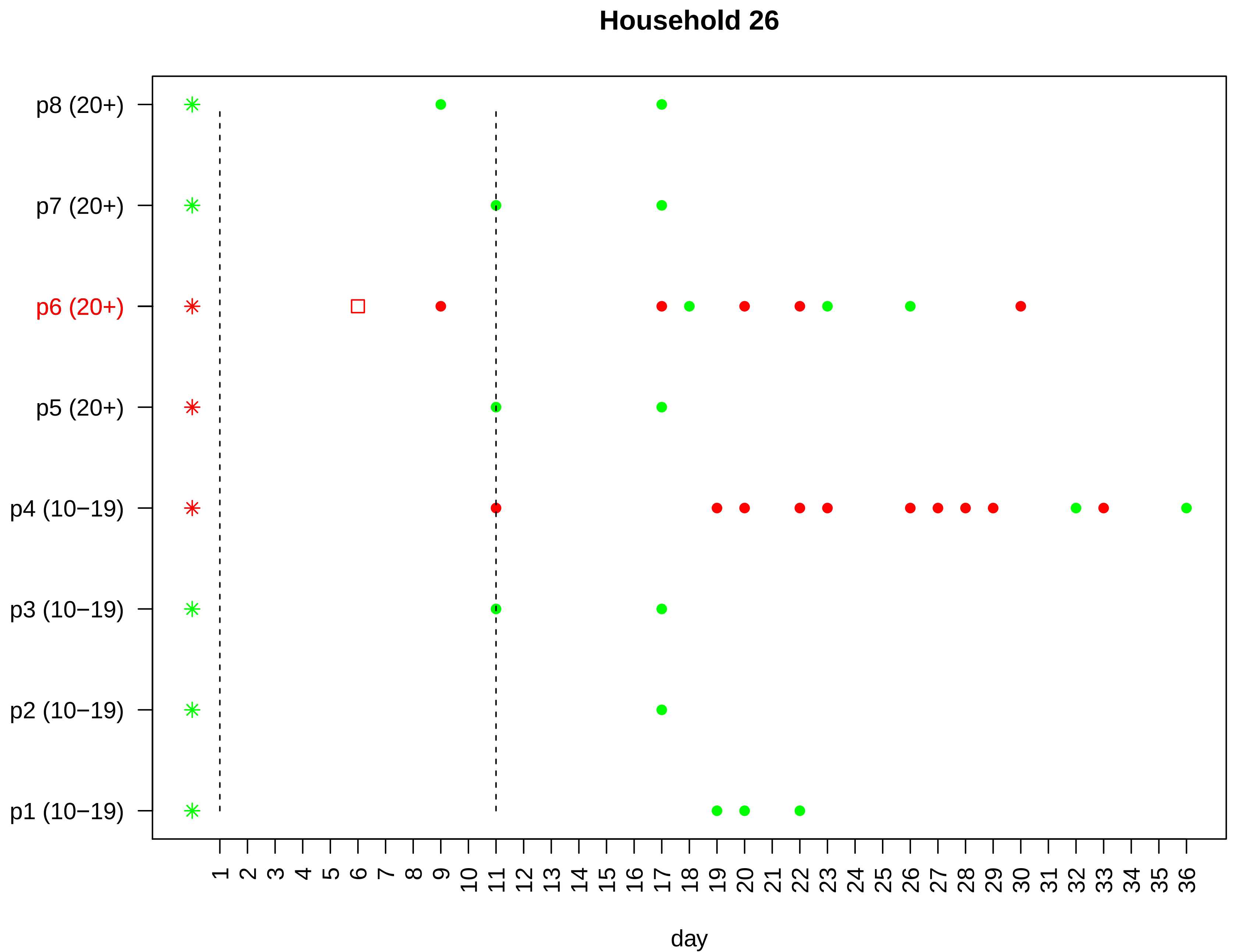

Household 26
p8 (20+)
p7 (20+)
p6 (20+)
p5 (20+)
p4 (10−19)
p3 (10−19)
p2 (10−19)
p1 (10−19)
1 2 3 4 5 6 7 8 9 10 11 12 13 14 15 16 17 18 19 20 21 22 23 24 25 26 27 28 29 30 31 32 33 34 35 36
day

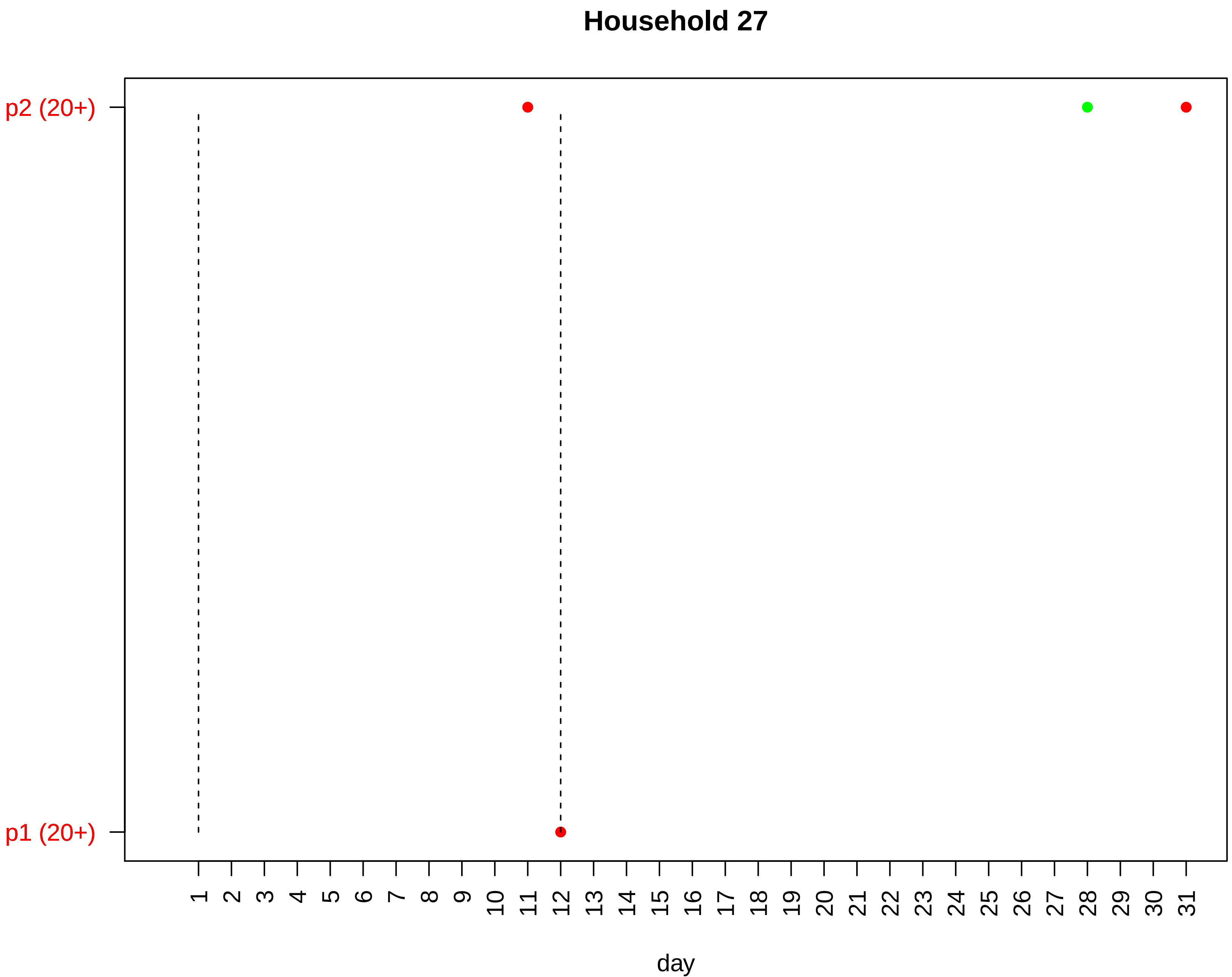

Household 27
p2 (20+)
p1 (20+)
day

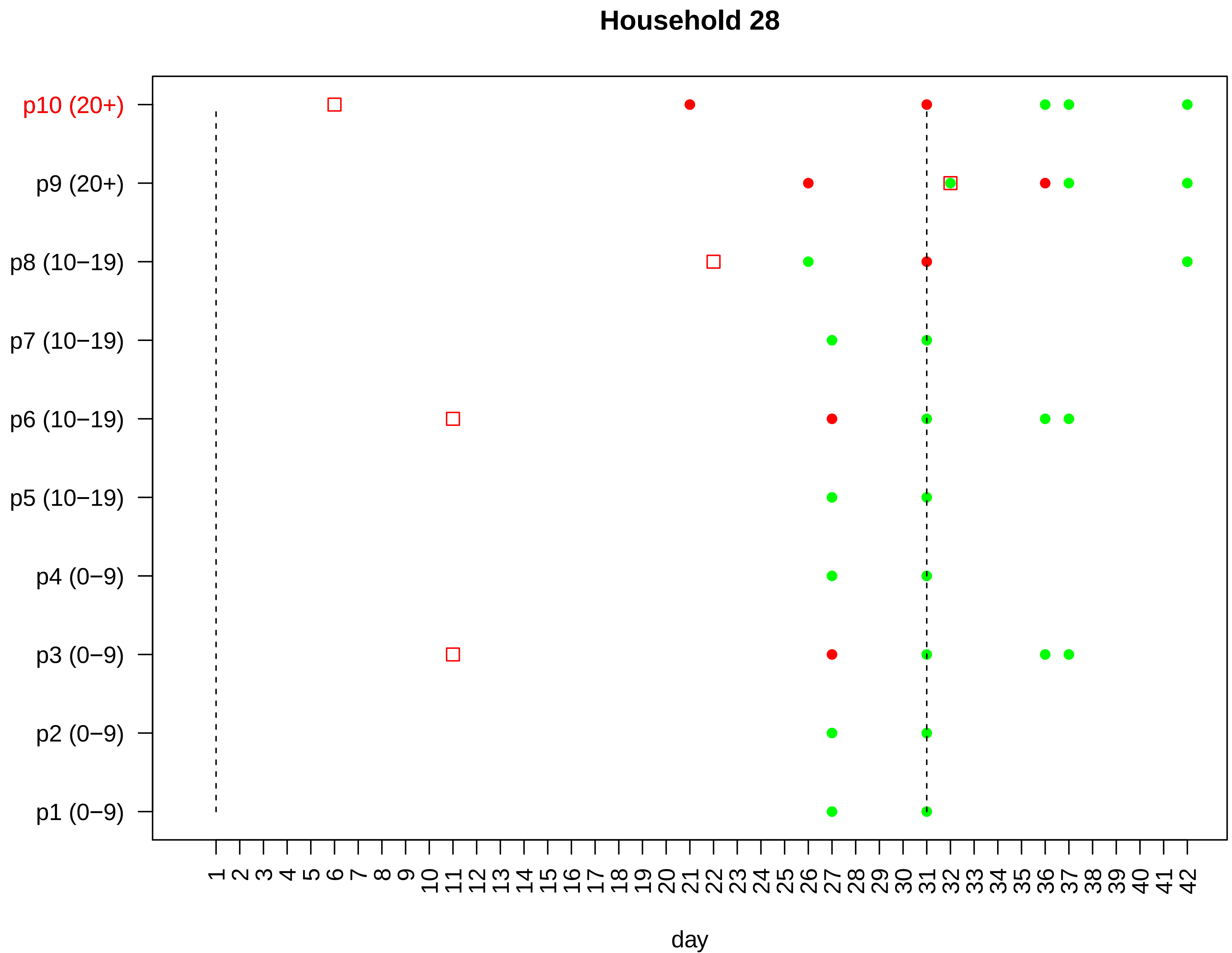

Household 28
p10 (20+)
p9 (20+)
p8 (10−19)
p7 (10−19)
p6 (10−19)
p5 (10−19)
p4 (0−9)
p3 (0−9)
p2 (0−9)
p1 (0−9)
1 2 3 4 5 6 7 8 9 10 11 12 13 14 15 16 17 18 19 20 21 22 23 24 25 26 27 28 29 30 31 32 33 34 35 36 37 38 39 40 41 42
day

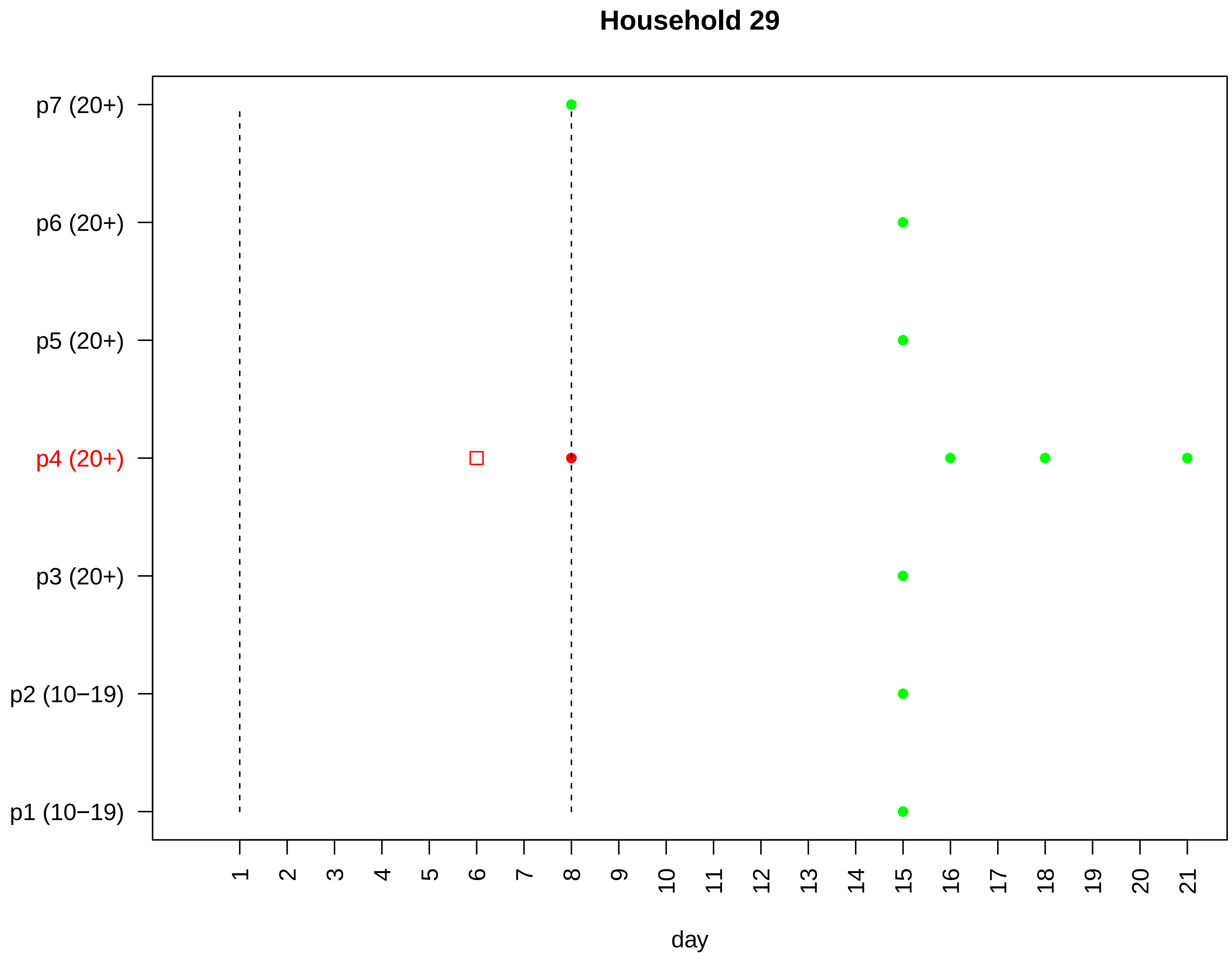
Household 29
p7 (20+)
p6 (20+)
p5 (20+)
p4 (20+)
p3 (20+)
p2 (10−19)
p1 (10−19)
day

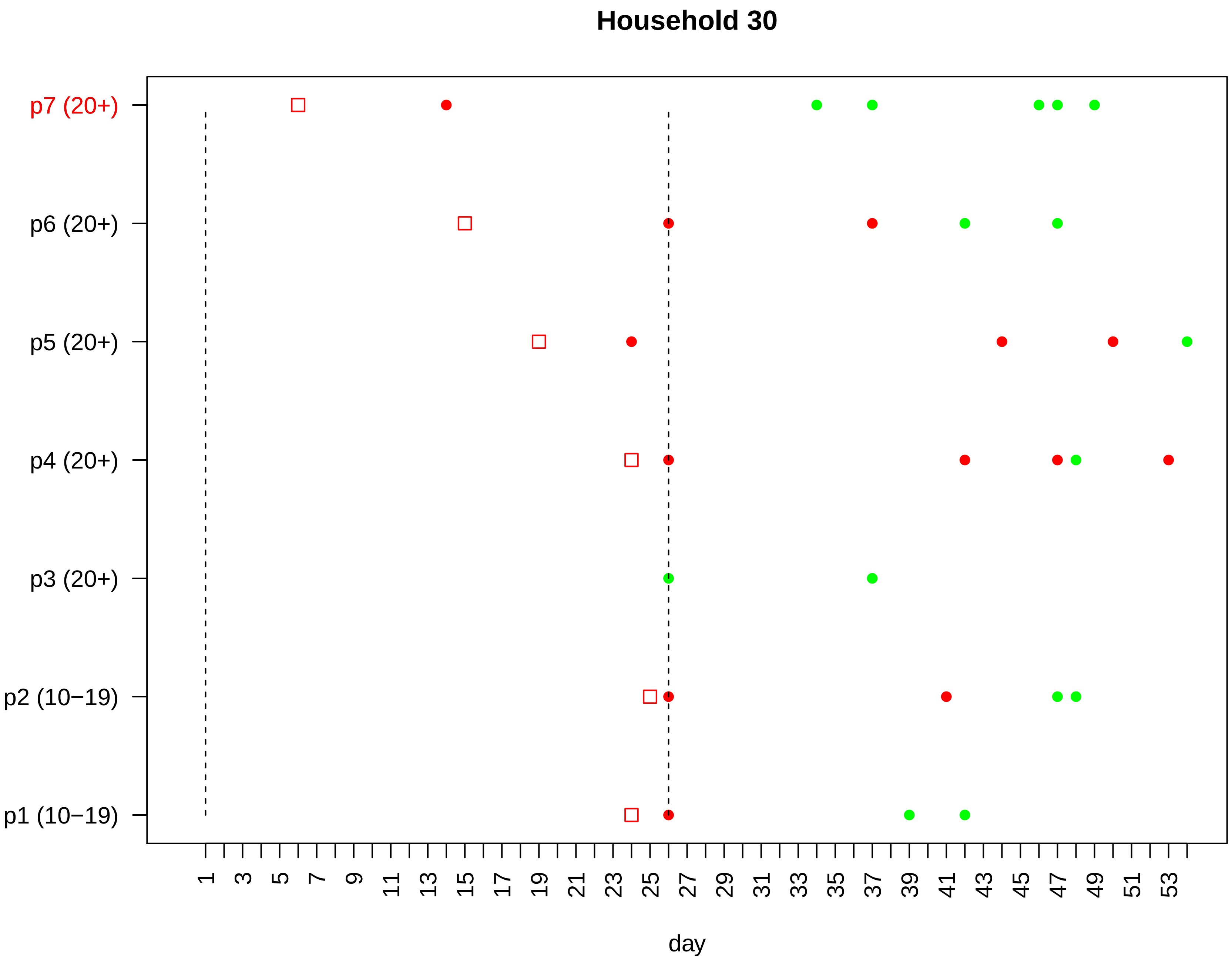
Household 30
p7 (20+)
p6 (20+)
p5 (20+)
p4 (20+)
p3 (20+)
p2 (10−19)
p1 (10−19)
1 3 5 7 9 11 13 15 17 19 21 23 25 27 29 31 33 35 37 39 41 43 45 47 49 51 53
day

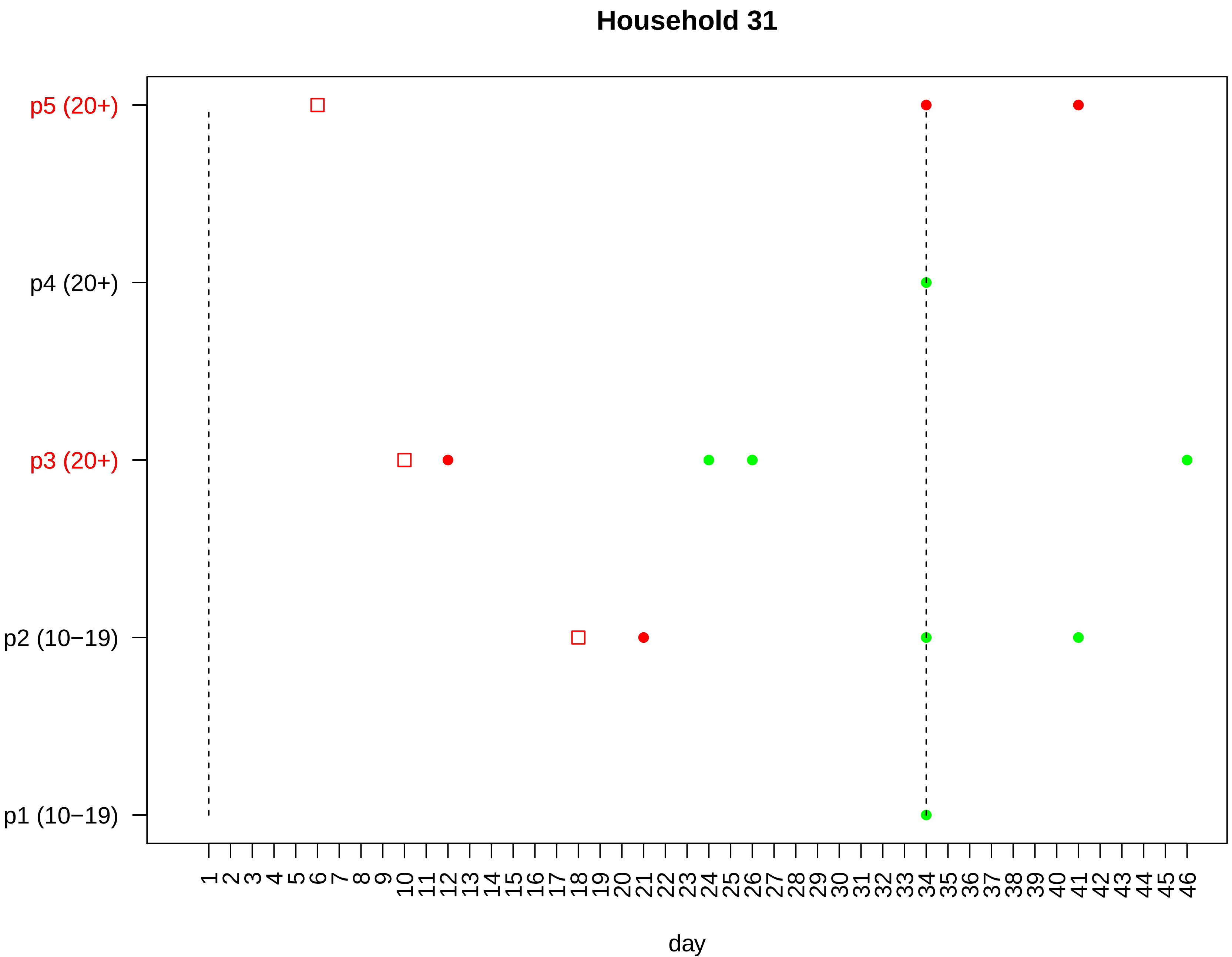

Household 31
p5 (20+)
p4 (20+)
p3 (20+)
p2 (10−19)
p1 (10−19)
1 2 3 4 5 6 7 8 9 10 11 12 13 14 15 16 17 18 19 20 21 22 23 24 25 26 27 28 29 30 31 32 33 34 35 36 37 38 39 40 41 42 43 44 45 46
day

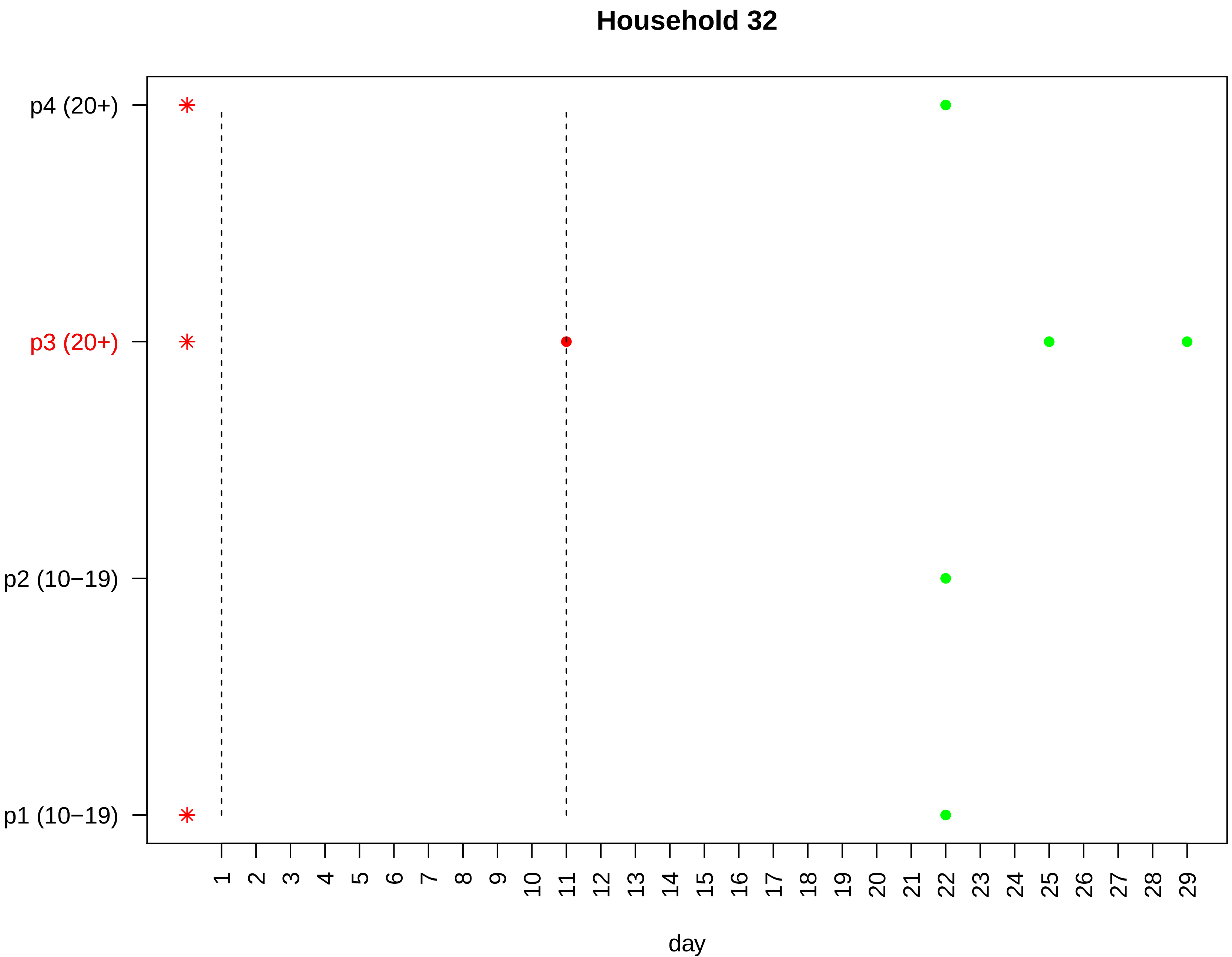

Household 32
p4 (20+)
p3 (20+)
p2 (10−19)
p1 (10−19)
day

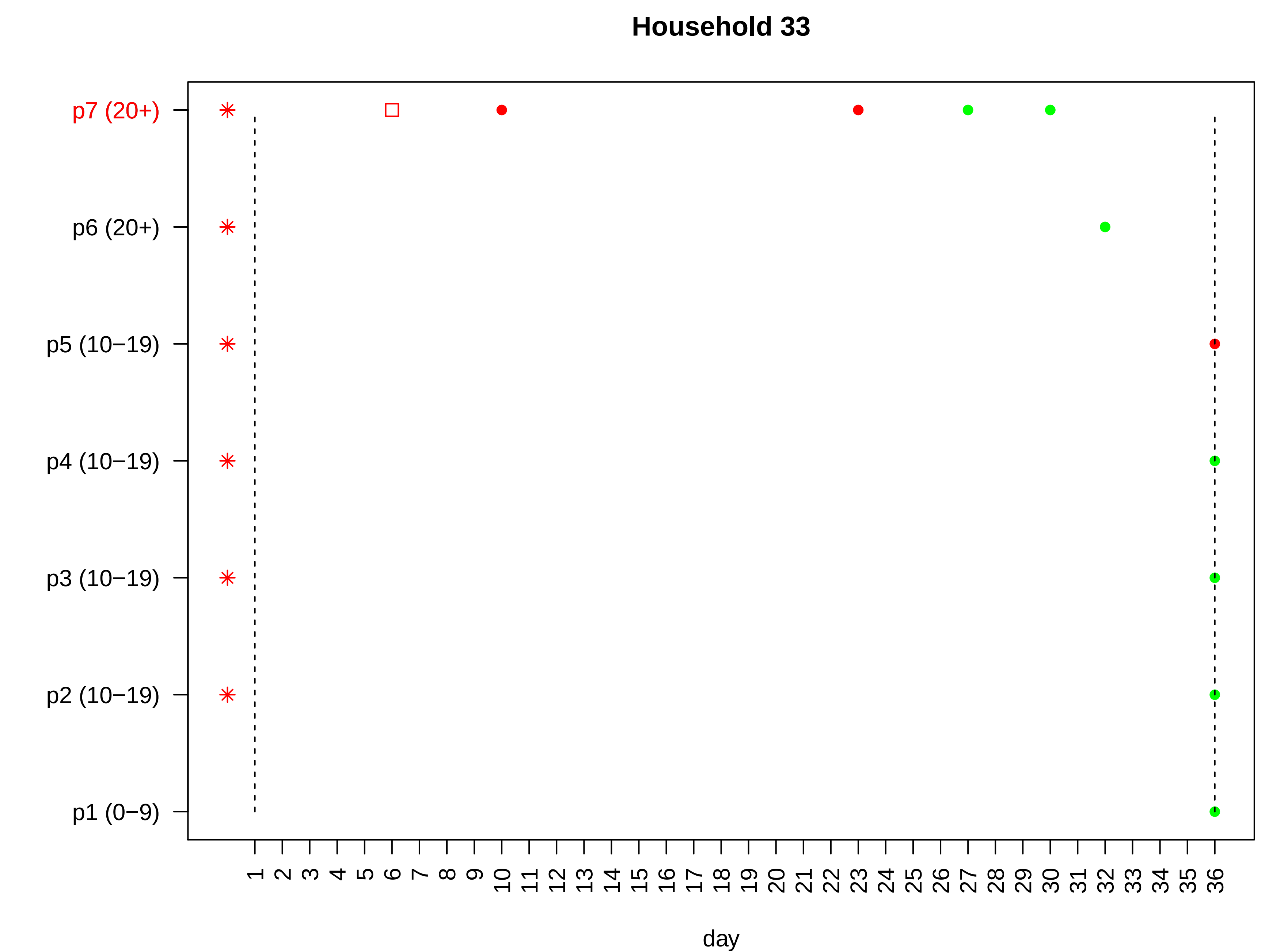

Household 33
p7 (20+)
p6 (20+)
p5 (10−19)
p4 (10−19)
p3 (10−19)
p2 (10−19)
p1 (0−9)
1 2 3 4 5 6 7 8 9 10 11 12 13 14 15 16 17 18 19 20 21 22 23 24 25 26 27 28 29 30 31 32 33 34 35 36
day

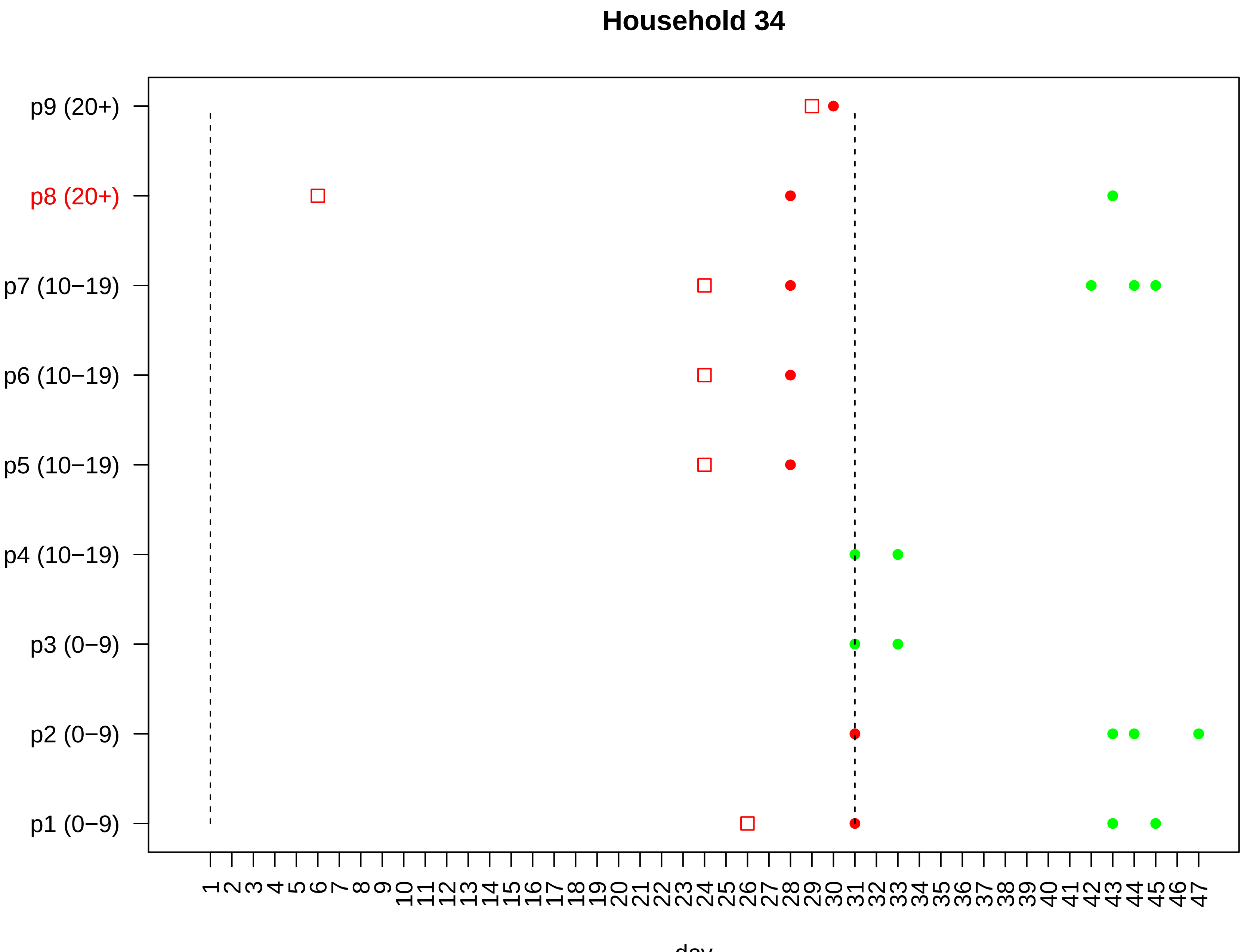

Household 34
p9 (20+)
p8 (20+)
p7 (10−19)
p6 (10−19)
p5 (10−19)
p4 (10−19)
p3 (0−9)
p2 (0−9)
p1 (0−9)
1 2 3 4 5 6 7 8 9 10 11 12 13 14 15 16 17 18 19 20 21 22 23 24 25 26 27 28 29 30 31 32 33 34 35 36 37 38 39 40 41 42 43 44 45 46 47
day

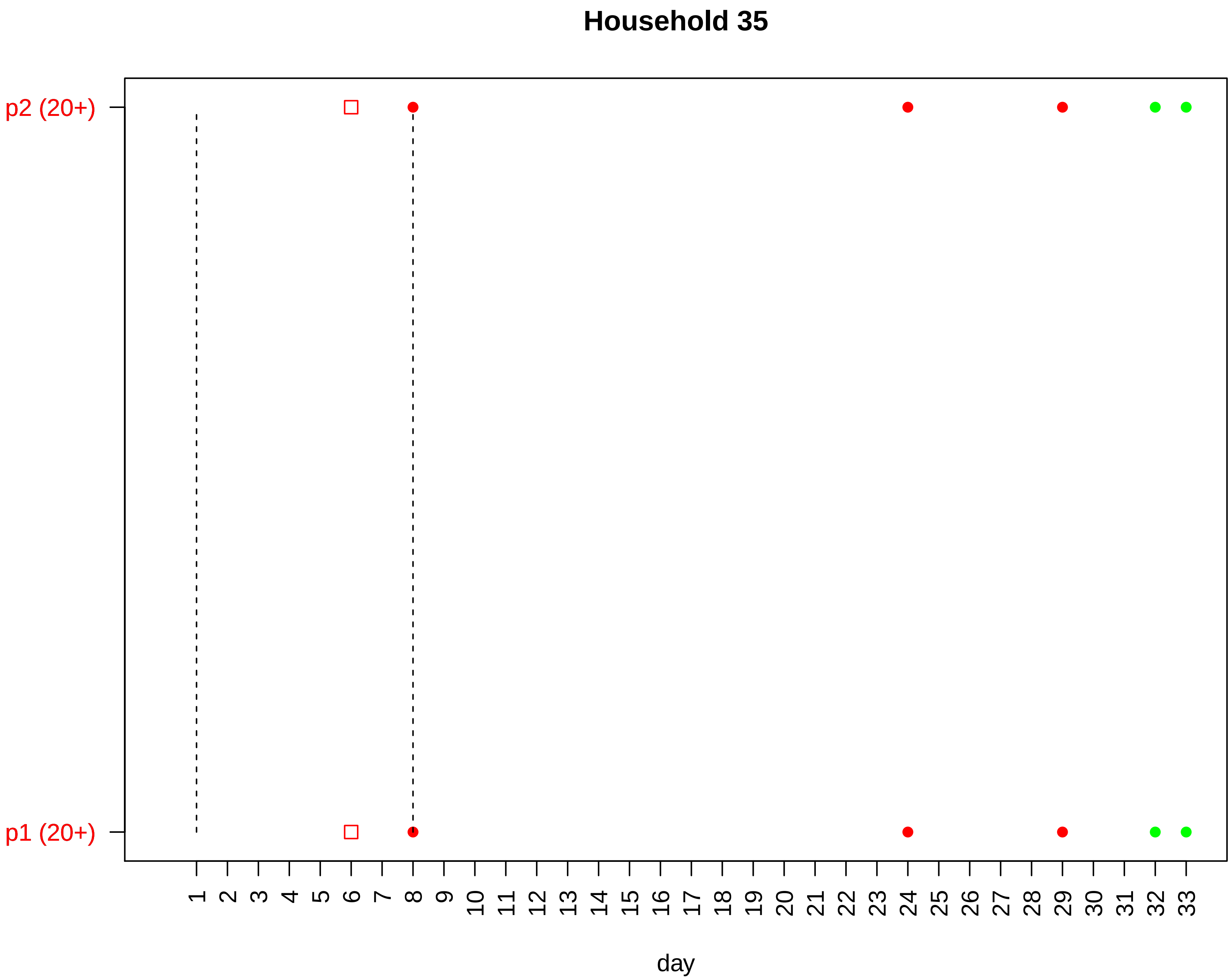

Household 35
p2 (20+)
p1 (20+)
day

#### Household 36

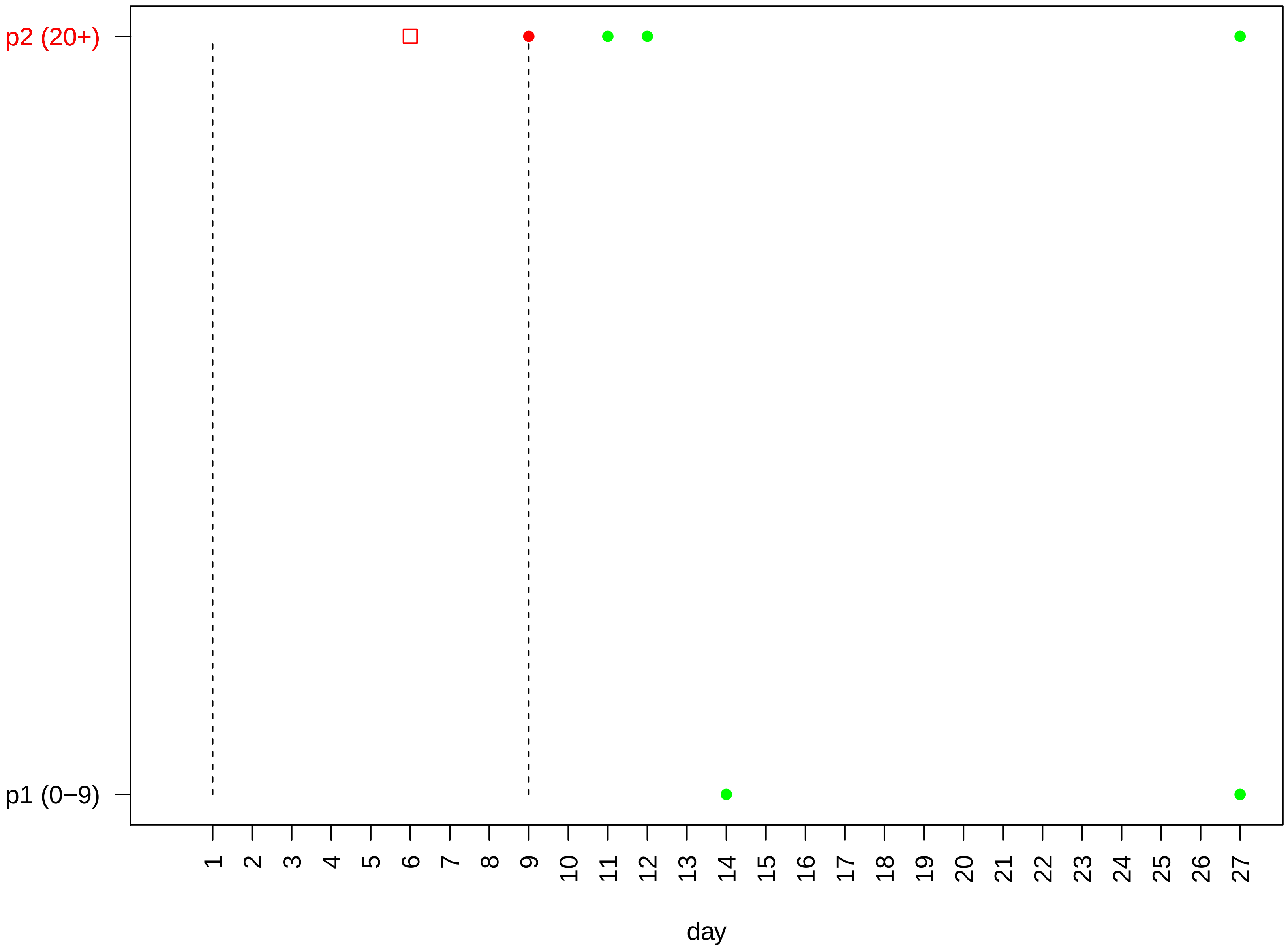

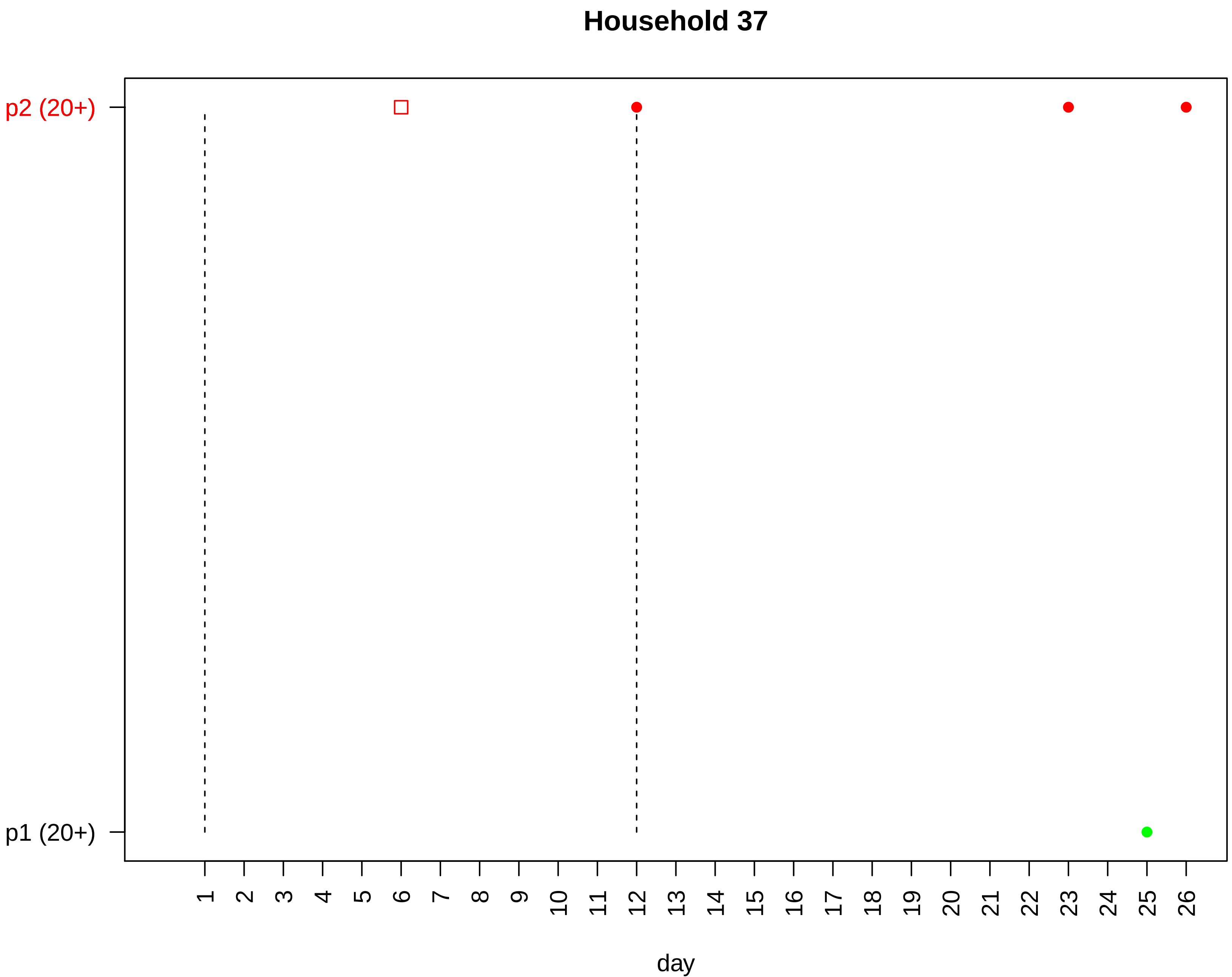
Household 37
p2 (20+)
p1 (20+)
day

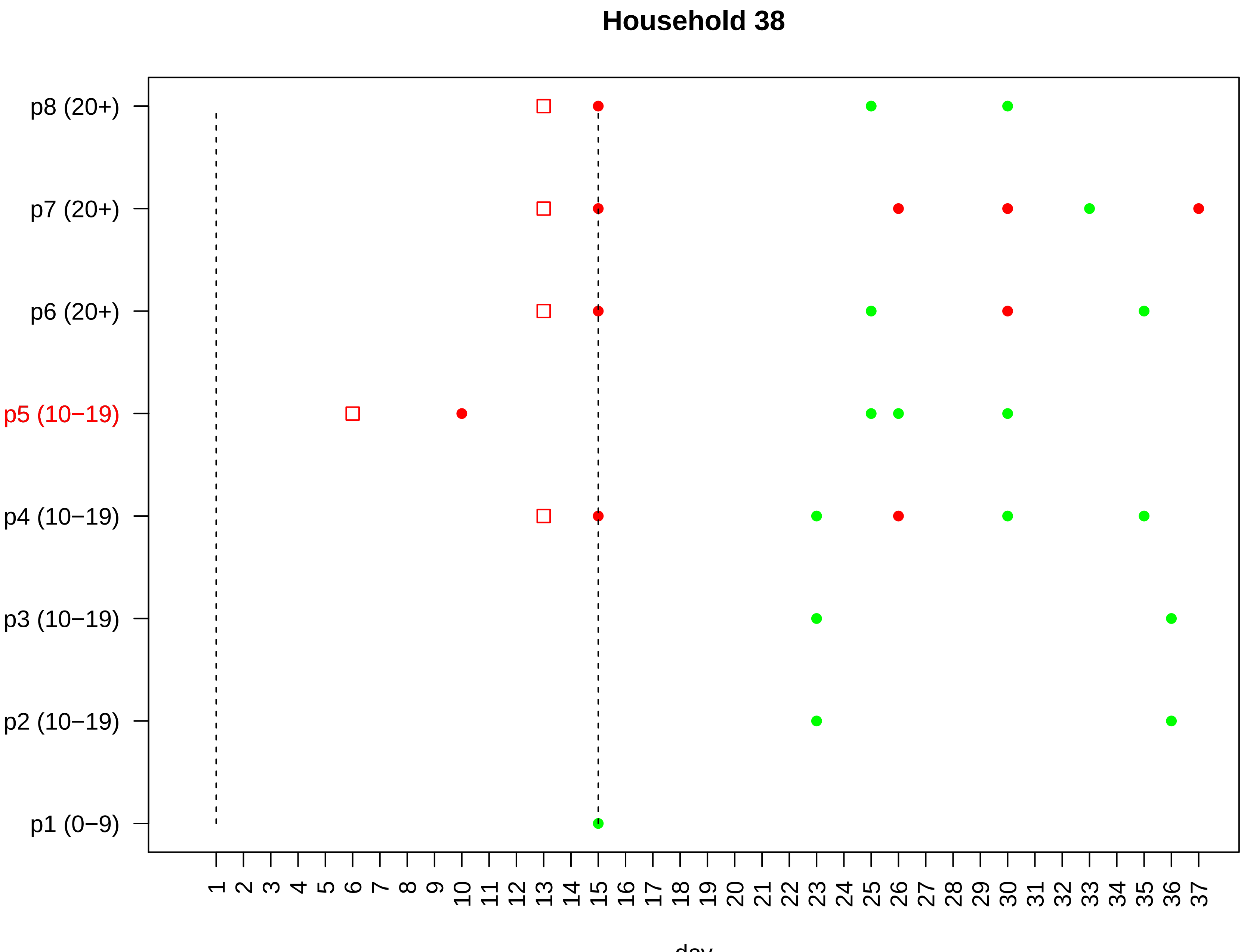

Household 38
p8 (20+)
p7 (20+)
p6 (20+)
p5 (10−19)
p4 (10−19)
p3 (10−19)
p2 (10−19)
p1 (0−9)
1 2 3 4 5 6 7 8 9 10 11 12 13 14 15 16 17 18 19 20 21 22 23 24 25 26 27 28 29 30 31 32 33 34 35 36 37
day

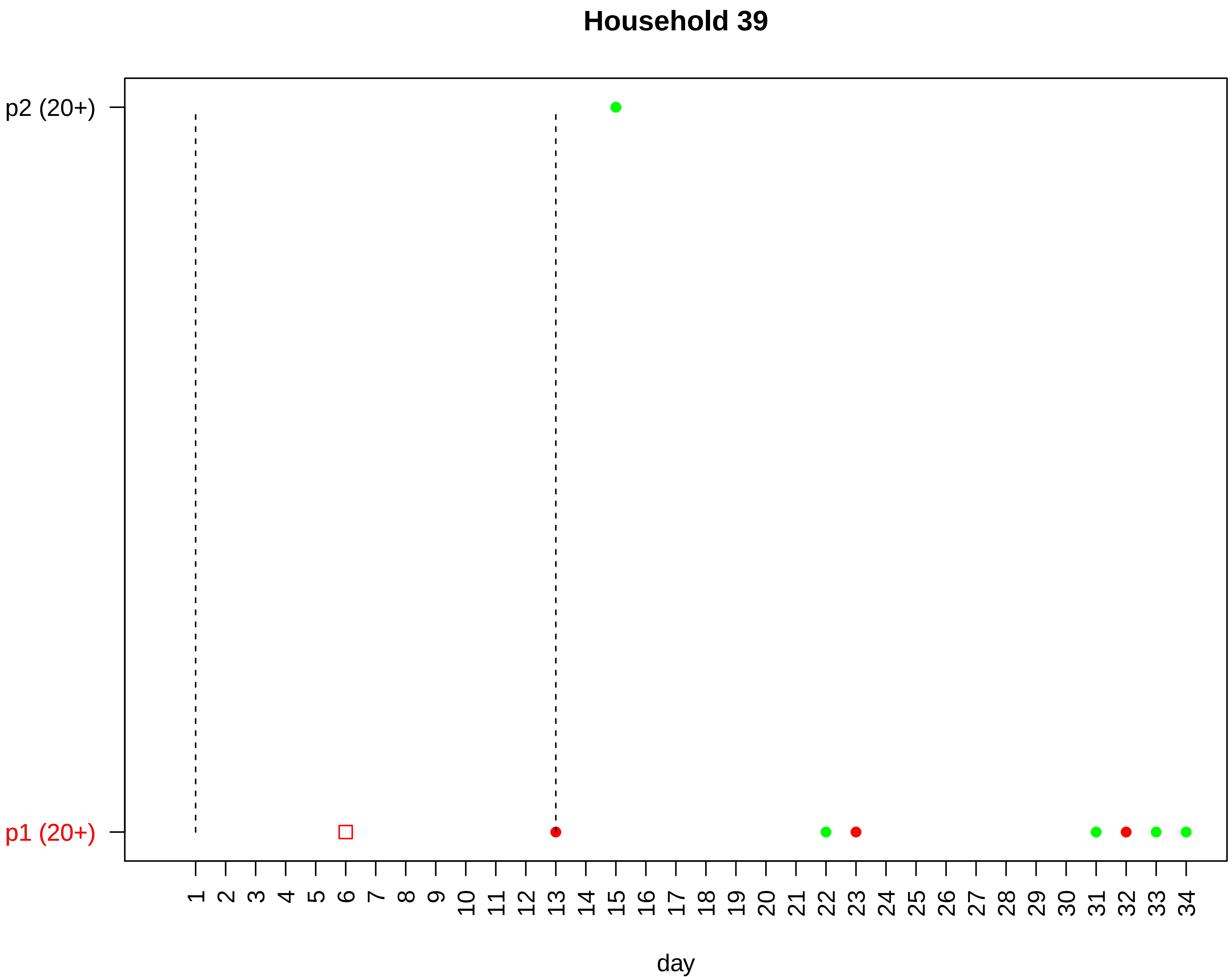

Household 39
p2 (20+)
p1 (20+)
day

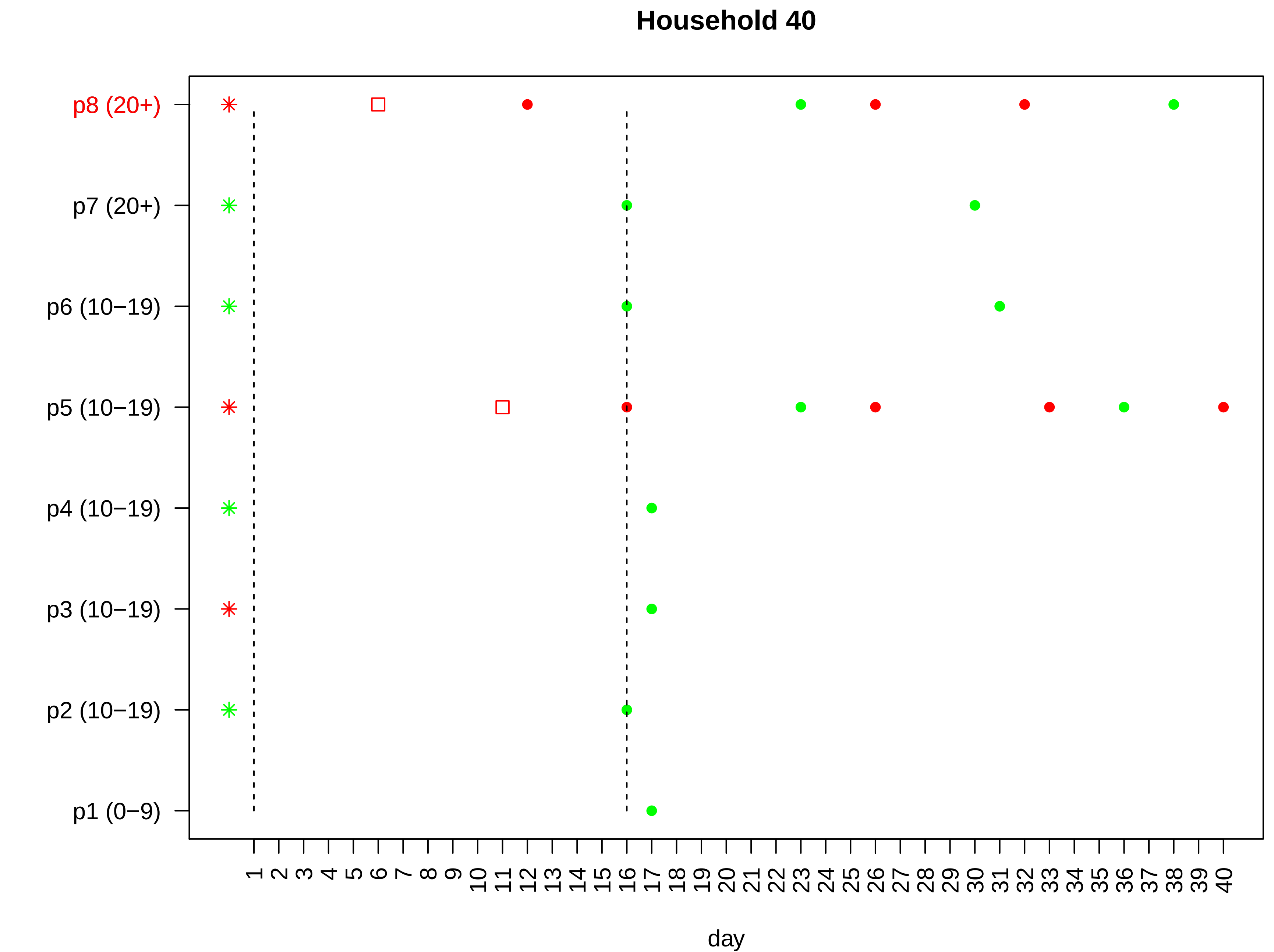

Household 40
p8 (20+)
p7 (20+)
p6 (10−19)
p5 (10−19)
p4 (10−19)
p3 (10−19)
p2 (10−19)
p1 (0−9)
1 2 3 4 5 6 7 8 9 10 11 12 13 14 15 16 17 18 19 20 21 22 23 24 25 26 27 28 29 30 31 32 33 34 35 36 37 38 39 40
day

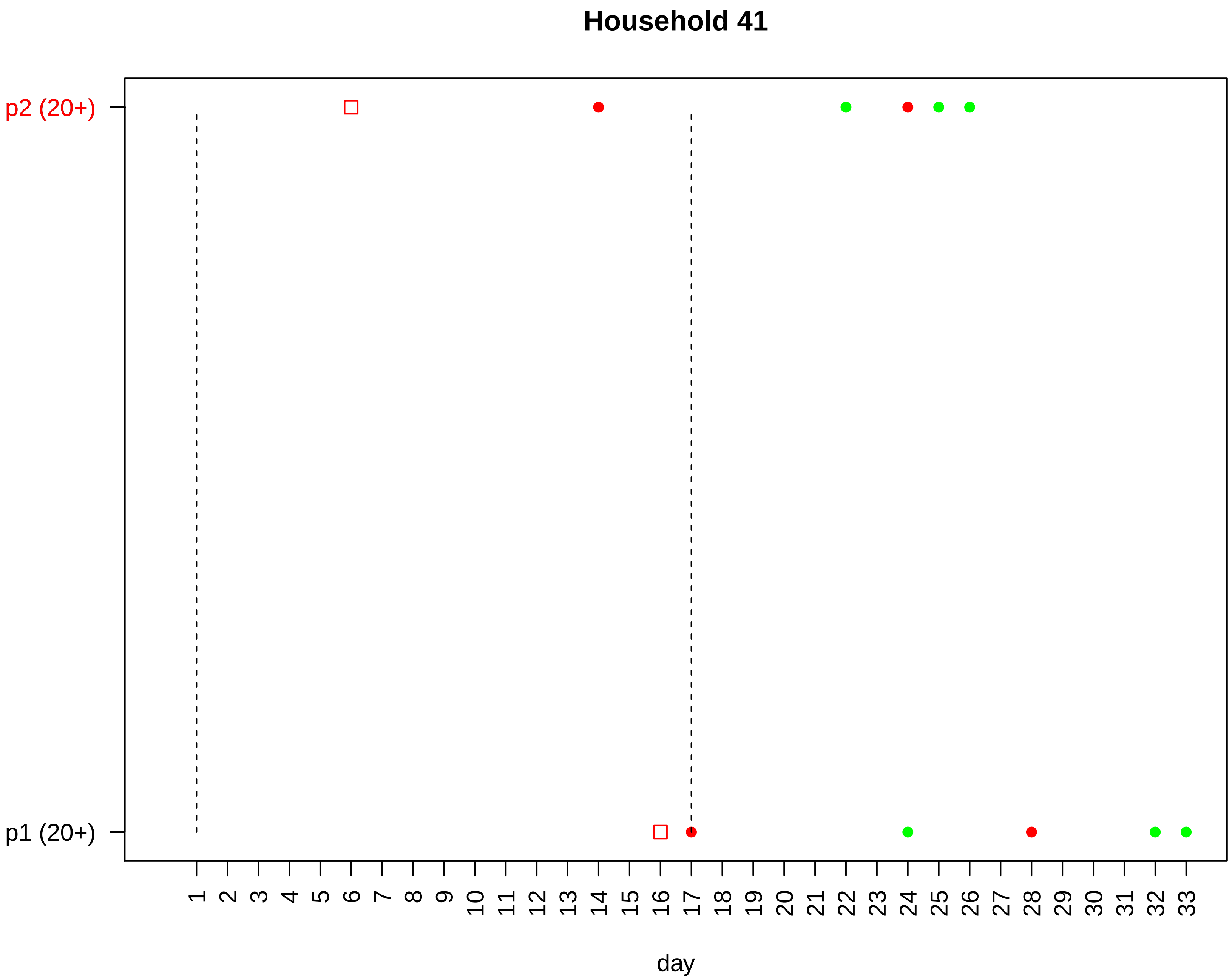

Household 41
p2 (20+)
p1 (20+)
day

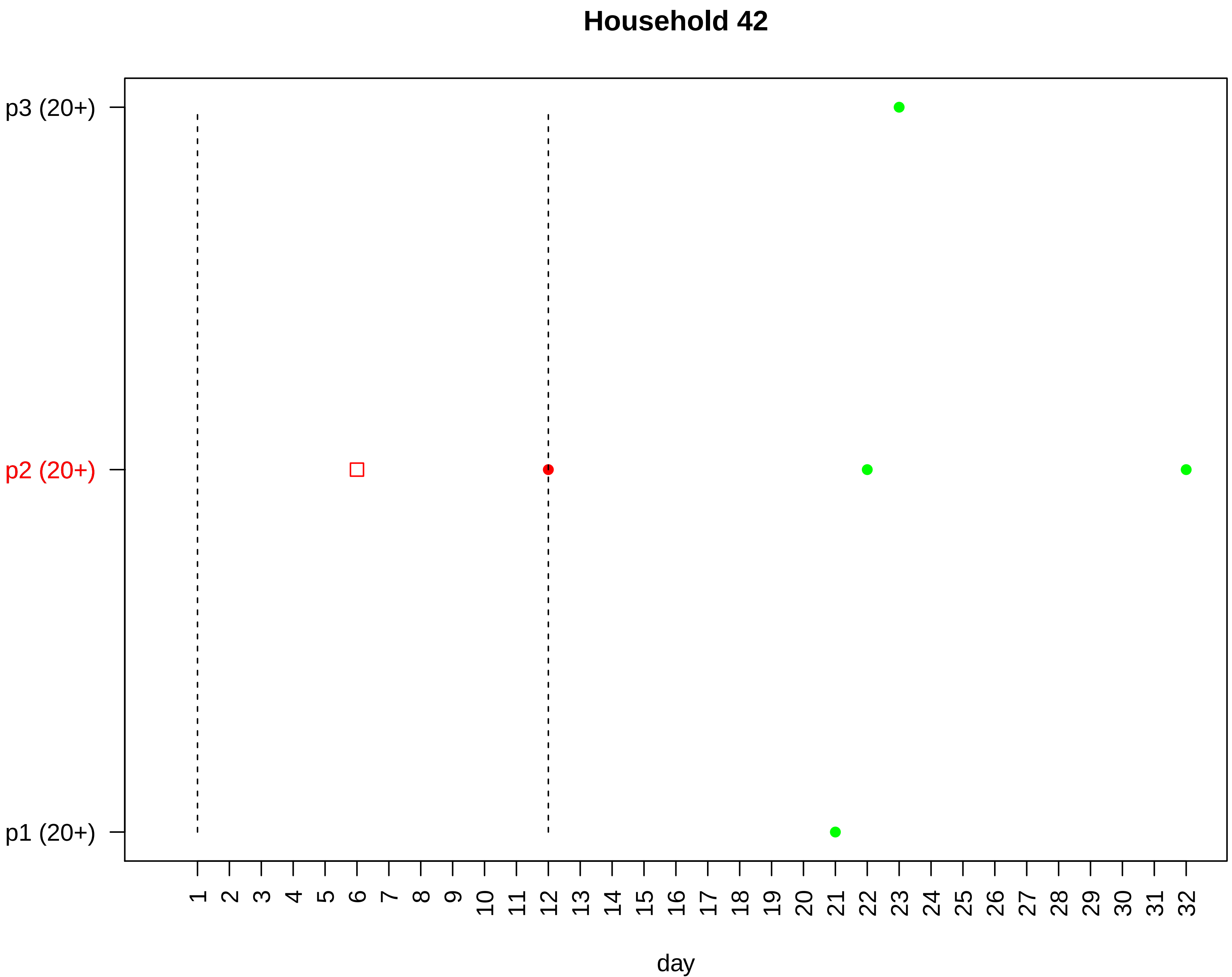

Household 42
p3 (20+)
p2 (20+)
p1 (20+)
1 2 3 4 5 6 7 8 9 10 11 12 13 14 15 16 17 18 19 20 21 22 23 24 25 26 27 28 29 30 31 32
day

#### Household 43

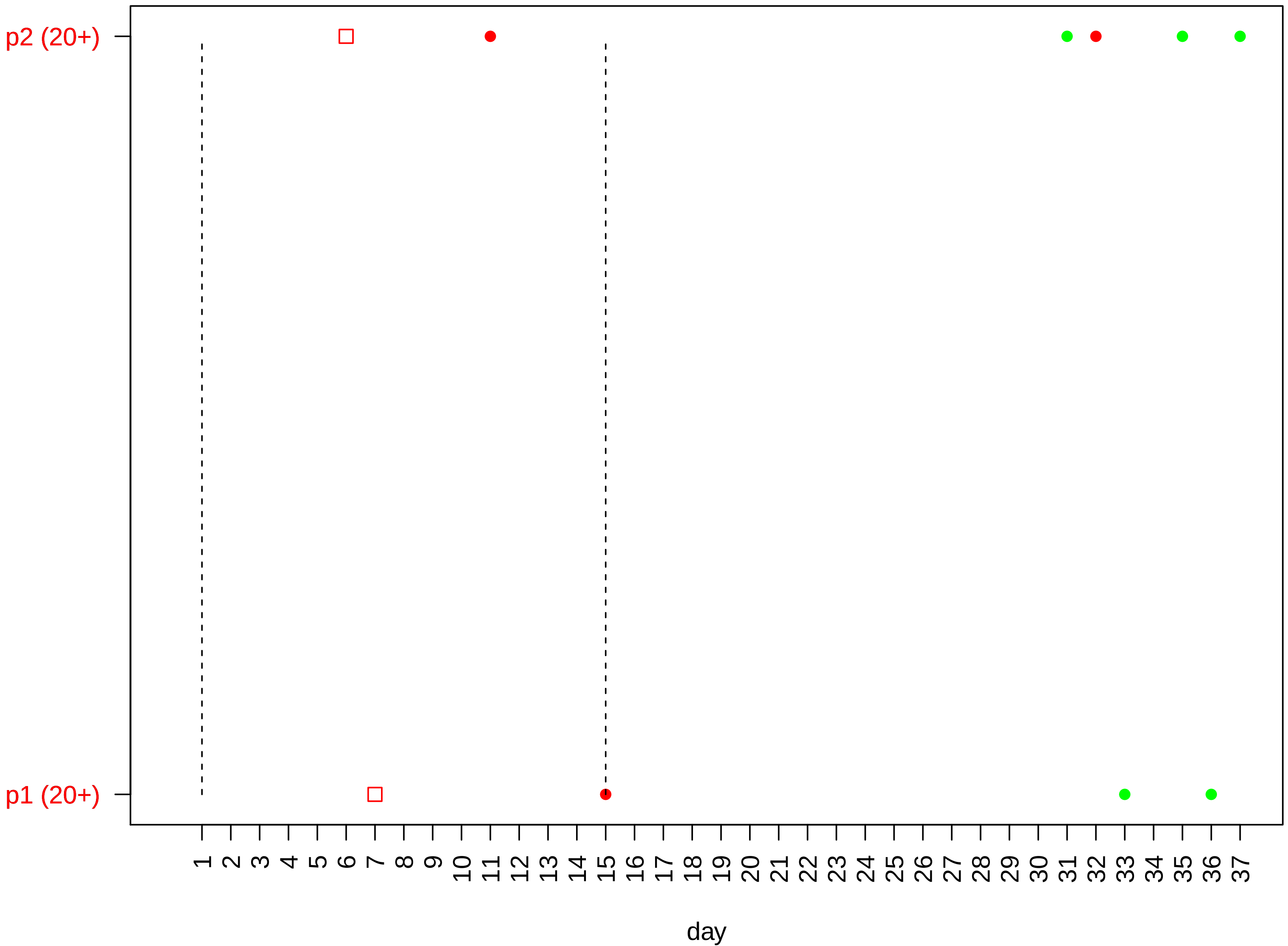

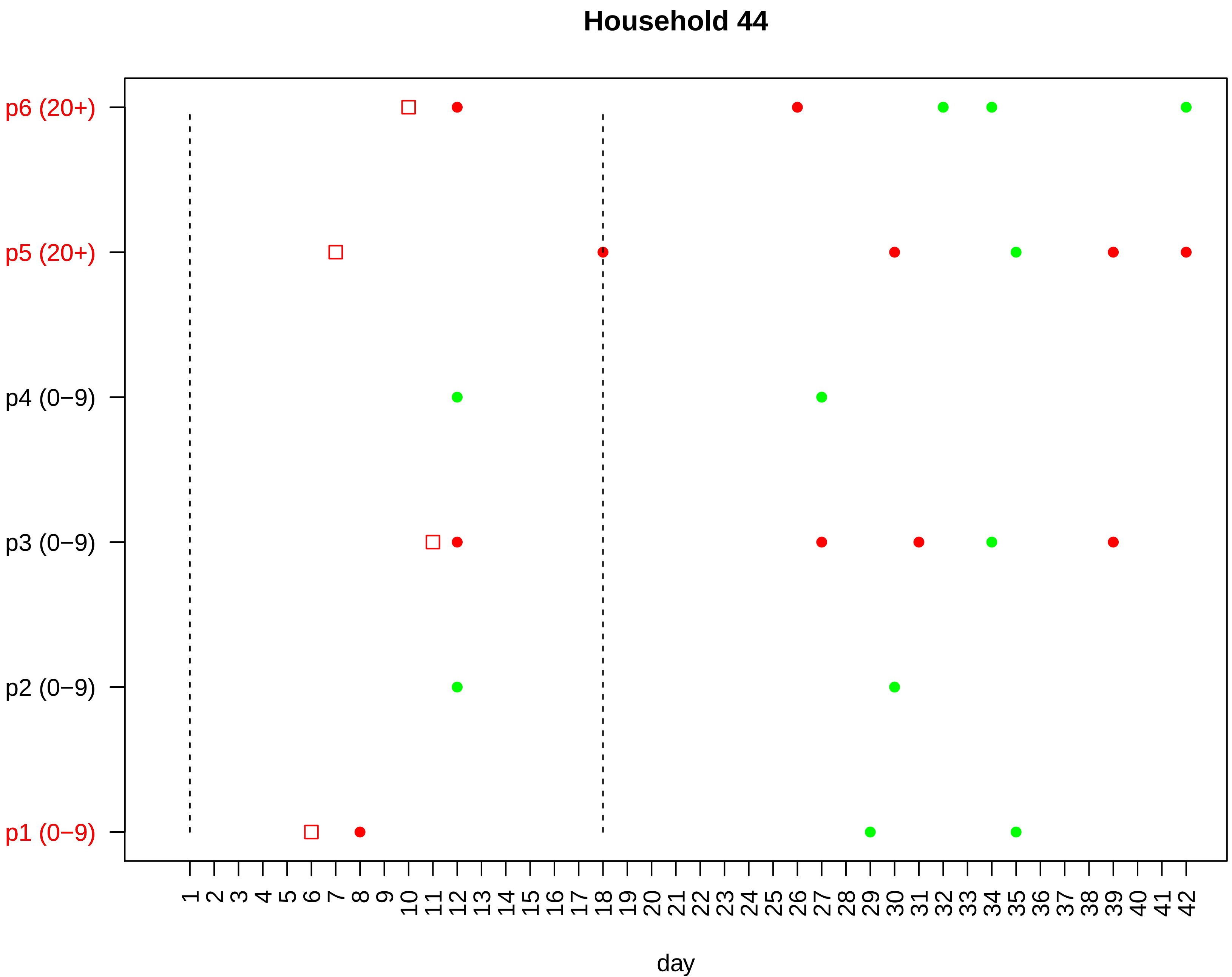
Household 44
p6 (20+)
p5 (20+)
p4 (0−9)
p3 (0−9)
p2 (0−9)
p1 (0−9)
1 2 3 4 5 6 7 8 9 10 11 12 13 14 15 16 17 18 19 20 21 22 23 24 25 26 27 28 29 30 31 32 33 34 35 36 37 38 39 40 41 42
day

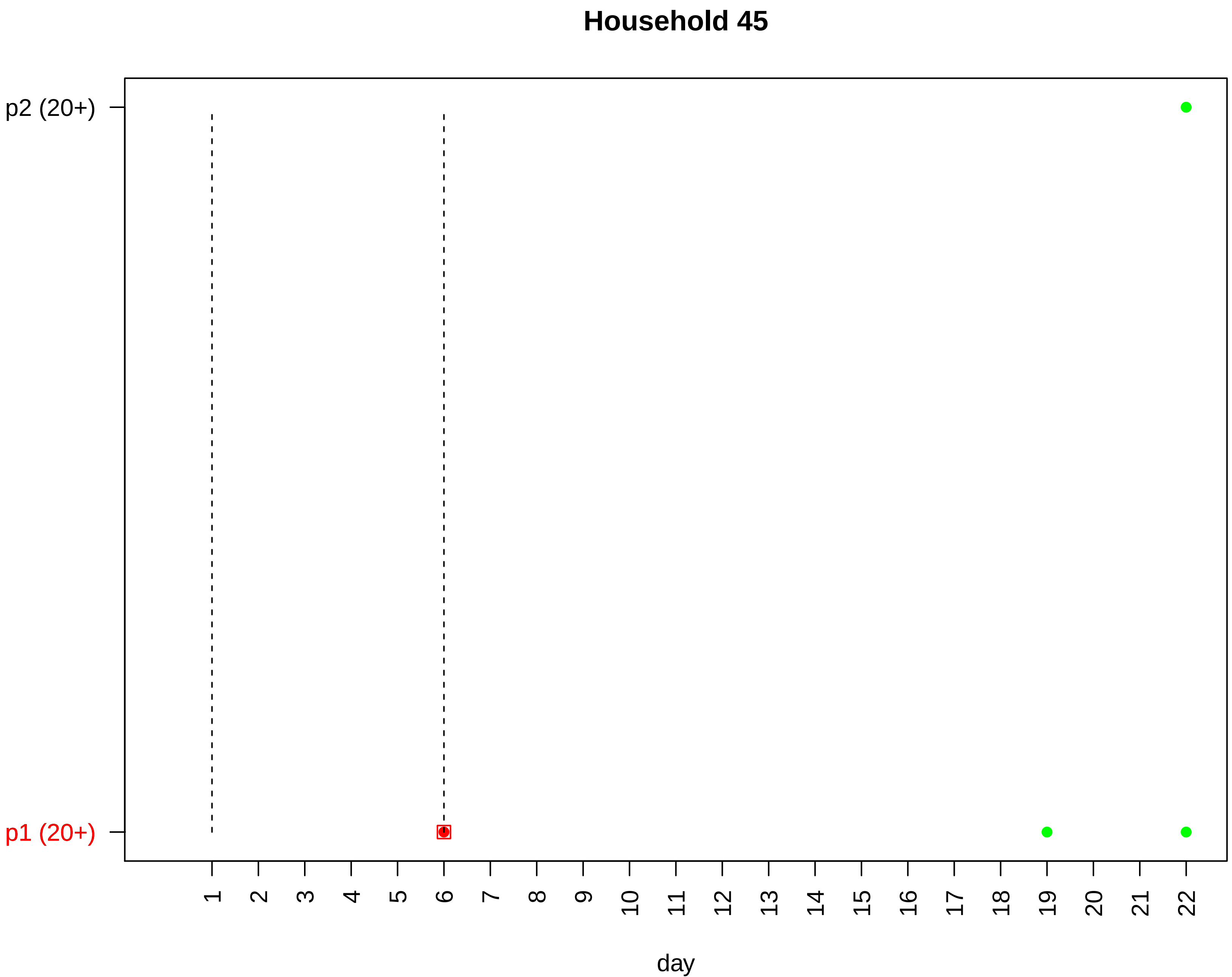

Household 45
p2 (20+)
p1 (20+)
day

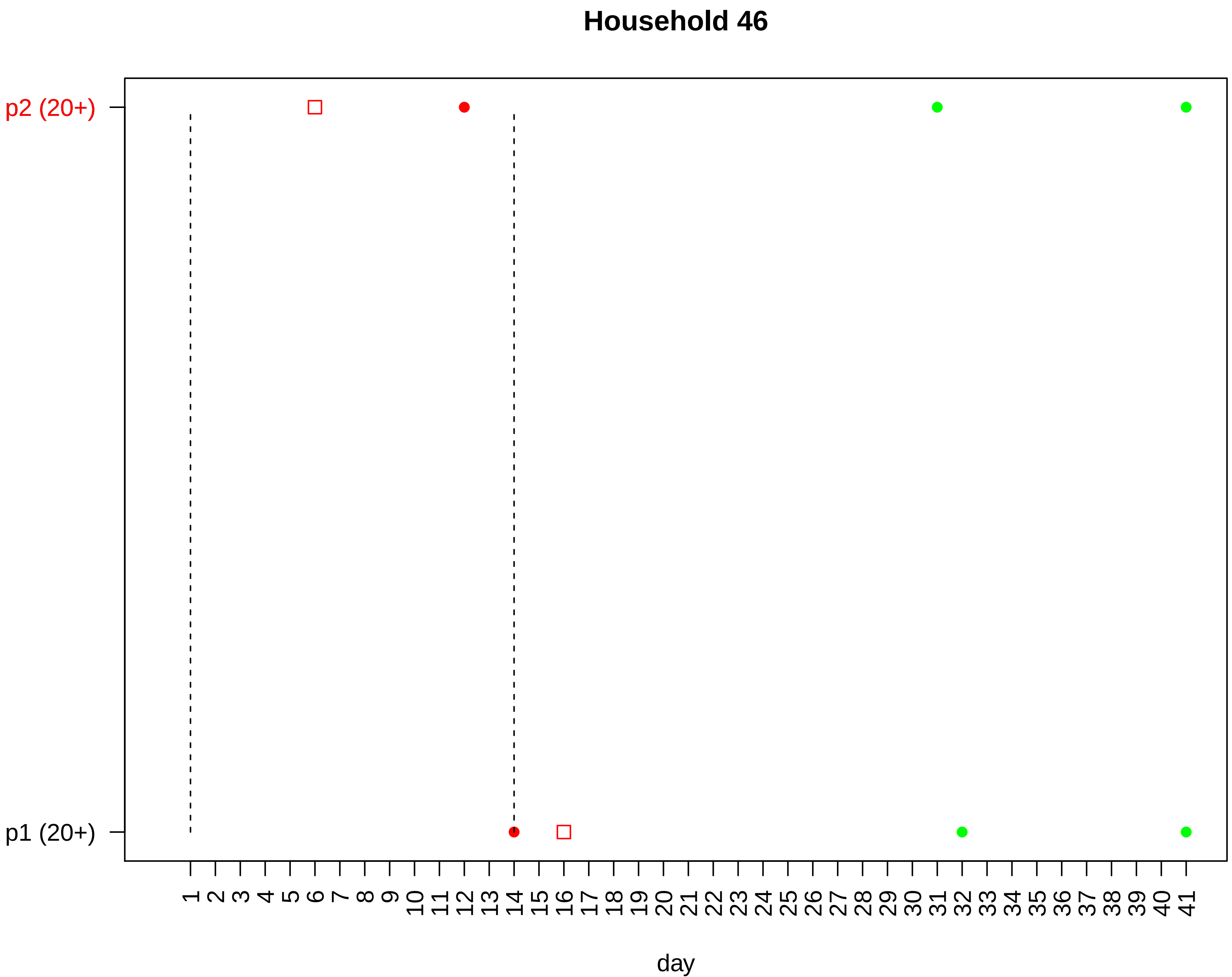
Household 46
p2 (20+)
p1 (20+)
1 2 3 4 5 6 7 8 9 10 11 12 13 14 15 16 17 18 19 20 21 22 23 24 25 26 27 28 29 30 31 32 33 34 35 36 37 38 39 40 41
day

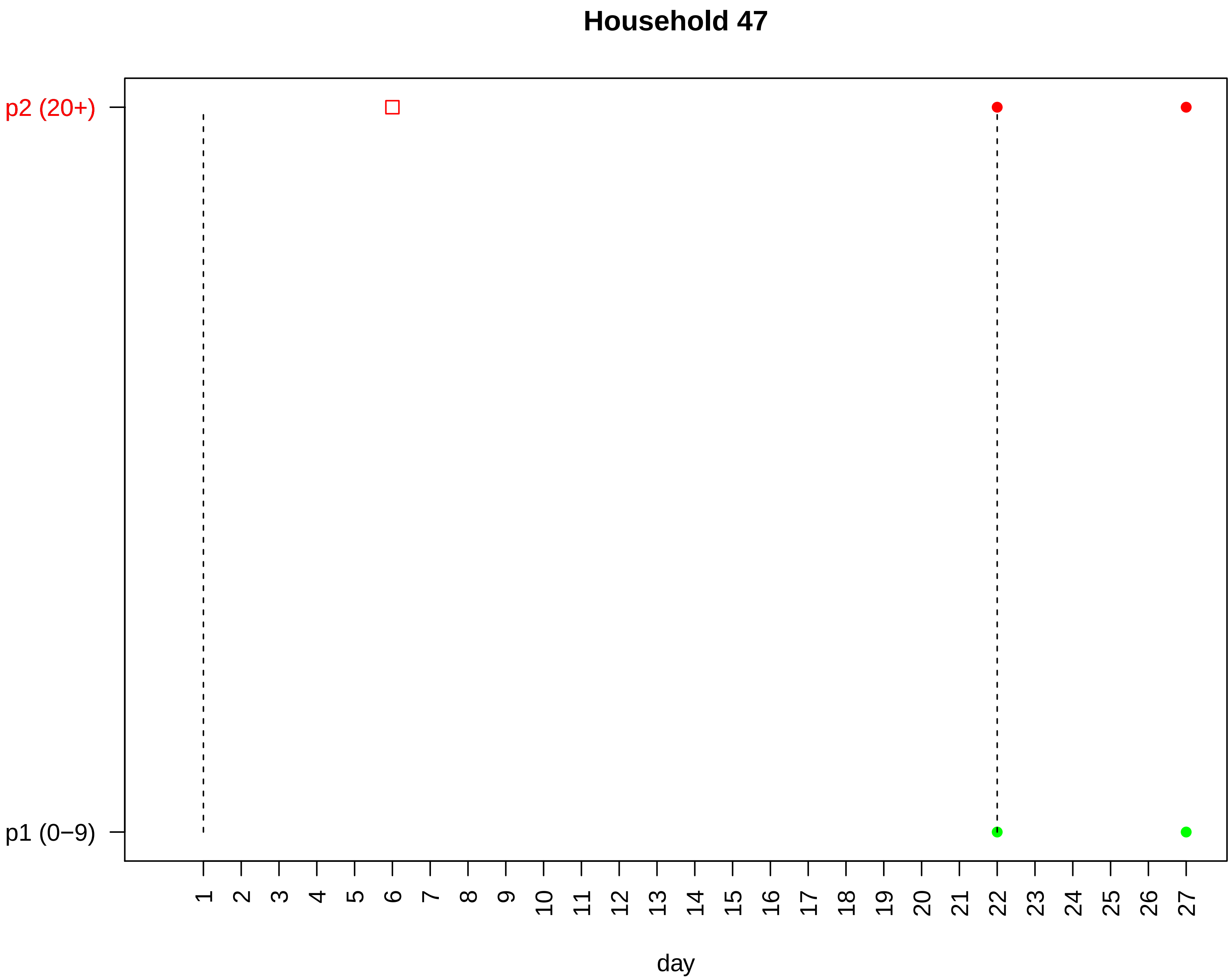

Household 47
p2 (20+)
p1 (0−9)
day

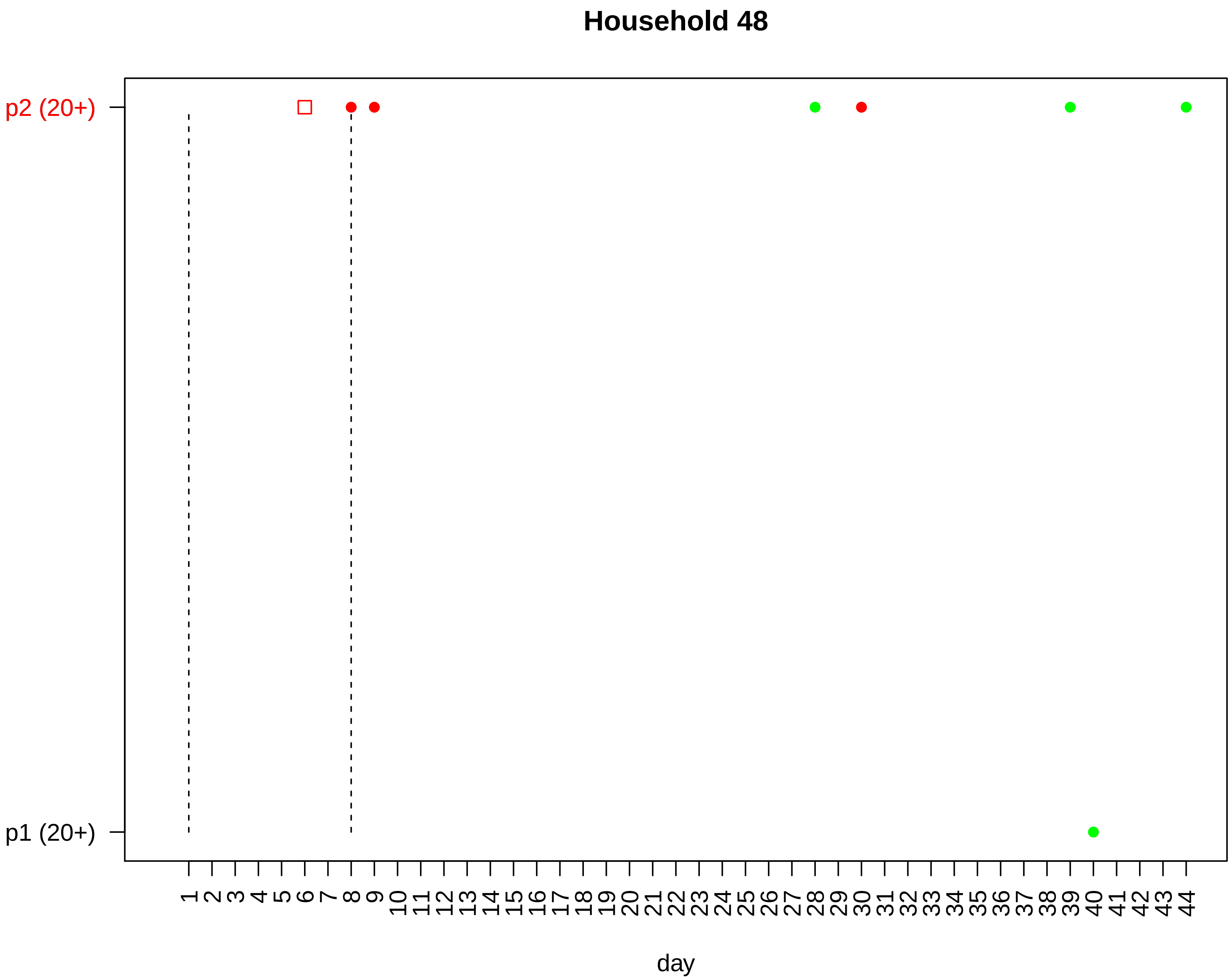

Household 48
p2 (20+)
p1 (20+)
1 2 3 4 5 6 7 8 9 10 11 12 13 14 15 16 17 18 19 20 21 22 23 24 25 26 27 28 29 30 31 32 33 34 35 36 37 38 39 40 41 42 43 44
day

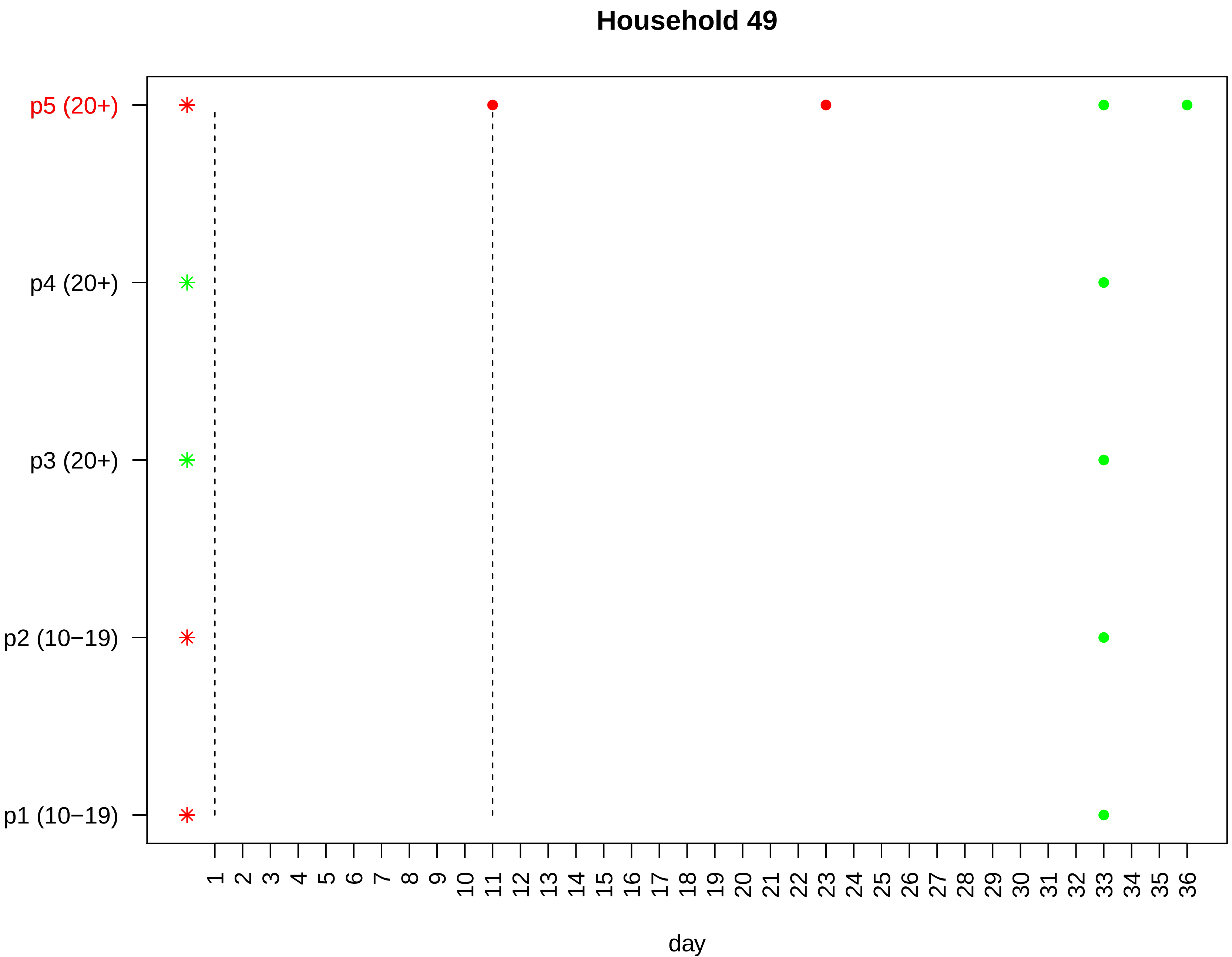
Household 49
p5 (20+)
p4 (20+)
p3 (20+)
p2 (10−19)
p1 (10−19)
1 2 3 4 5 6 7 8 9 10 11 12 13 14 15 16 17 18 19 20 21 22 23 24 25 26 27 28 29 30 31 32 33 34 35 36
day

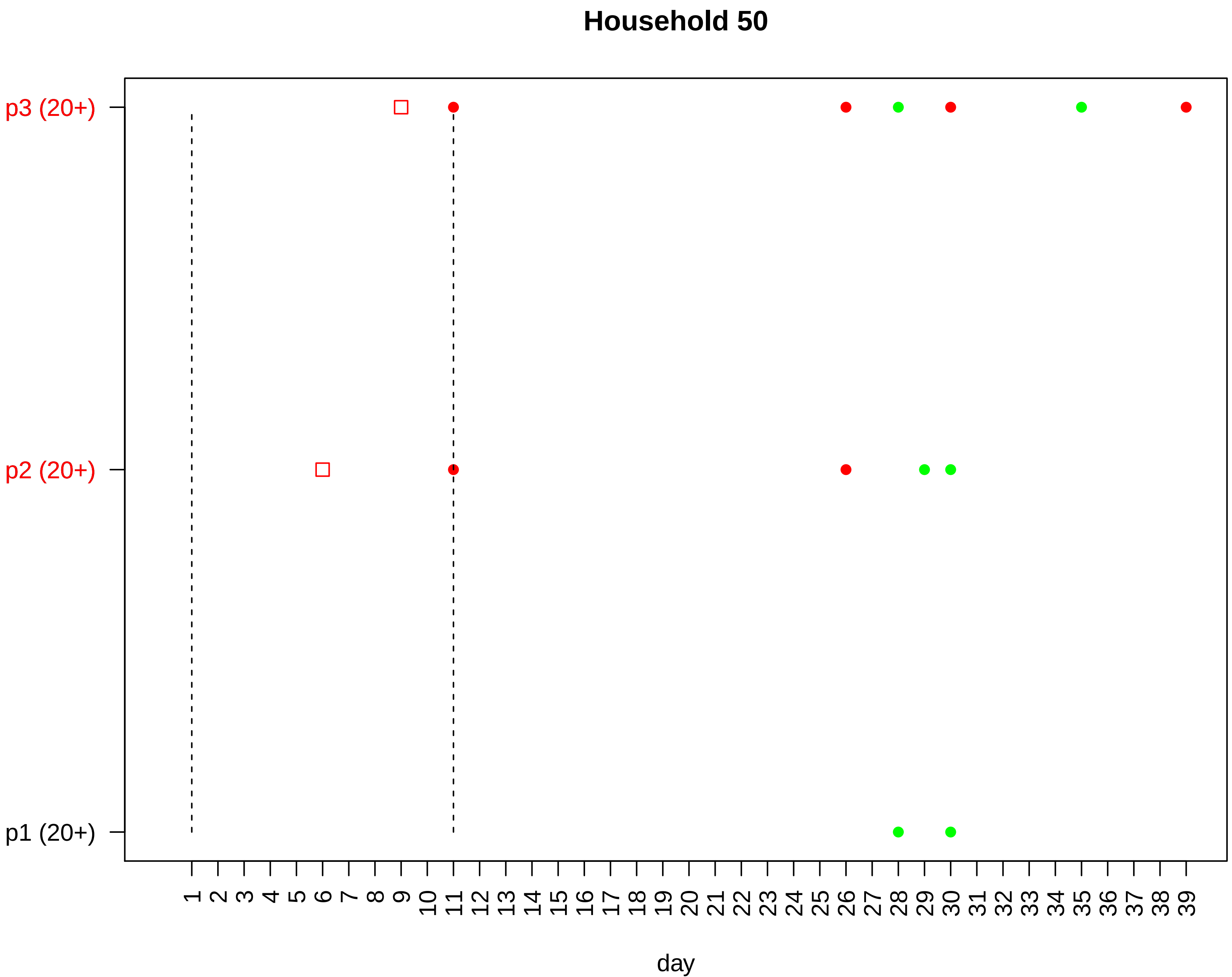

Household 50
p3 (20+)
p2 (20+)
p1 (20+)
1 2 3 4 5 6 7 8 9 10 11 12 13 14 15 16 17 18 19 20 21 22 23 24 25 26 27 28 29 30 31 32 33 34 35 36 37 38 39
day

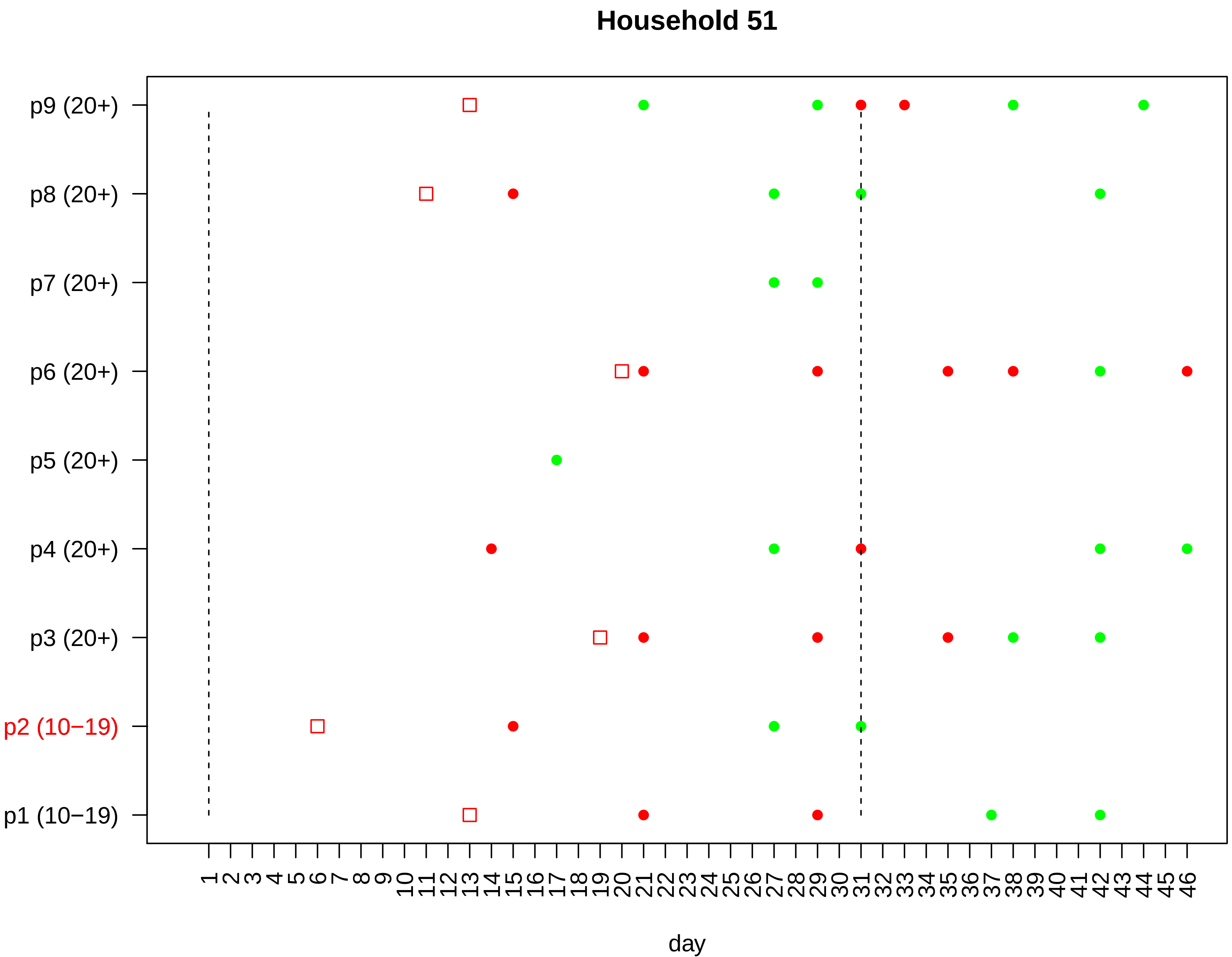

Household 51
p9 (20+)
p8 (20+)
p7 (20+)
p6 (20+)
p5 (20+)
p4 (20+)
p3 (20+)
p2 (10−19)
p1 (10−19)
1 2 3 4 5 6 7 8 9 10 11 12 13 14 15 16 17 18 19 20 21 22 23 24 25 26 27 28 29 30 31 32 33 34 35 36 37 38 39 40 41 42 43 44 45 46
day

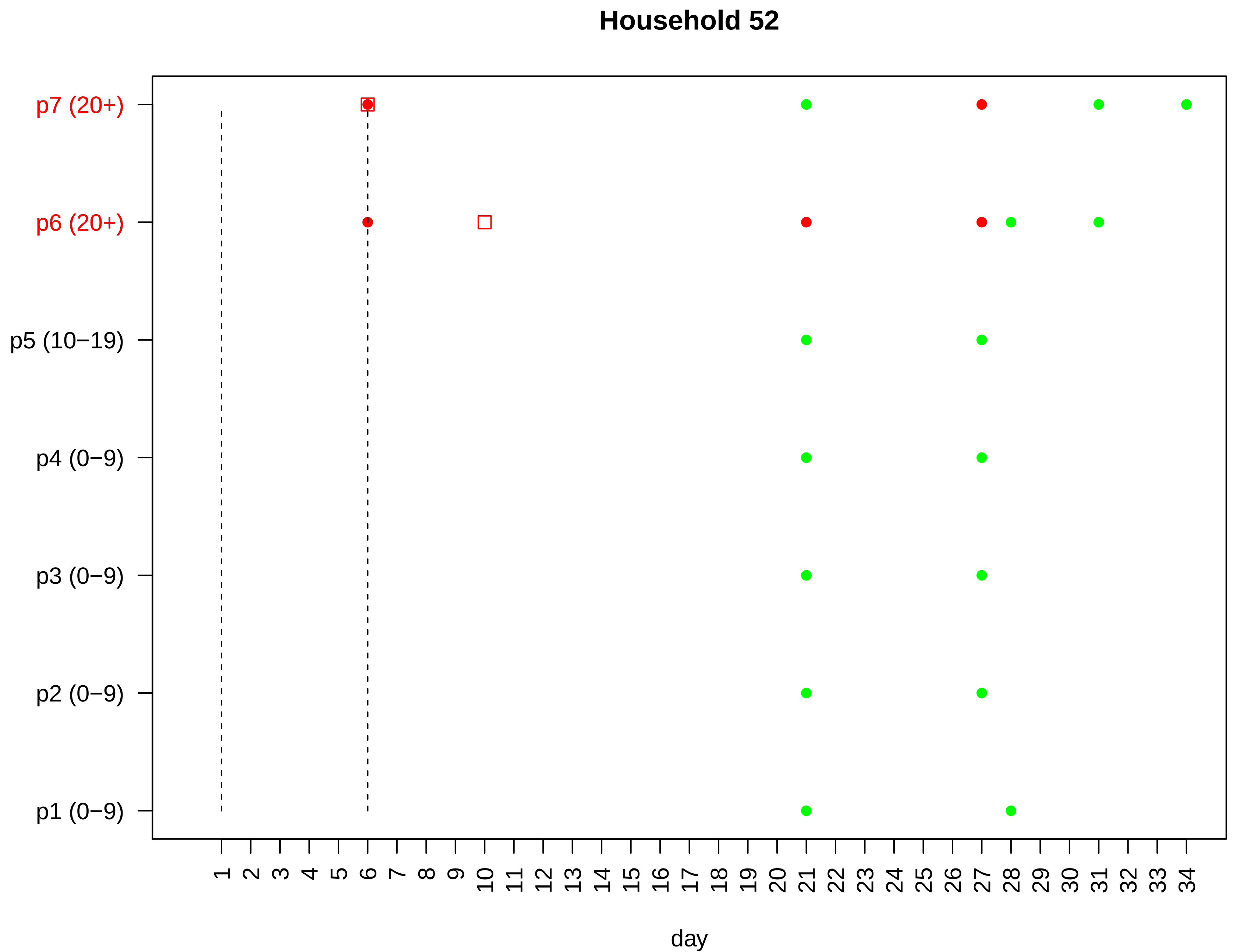

Household 52
p7 (20+)
p6 (20+)
p5 (10−19)
p4 (0−9)
p3 (0−9)
p2 (0−9)
p1 (0−9)
1 2 3 4 5 6 7 8 9 10 11 12 13 14 15 16 17 18 19 20 21 22 23 24 25 26 27 28 29 30 31 32 33 34
day

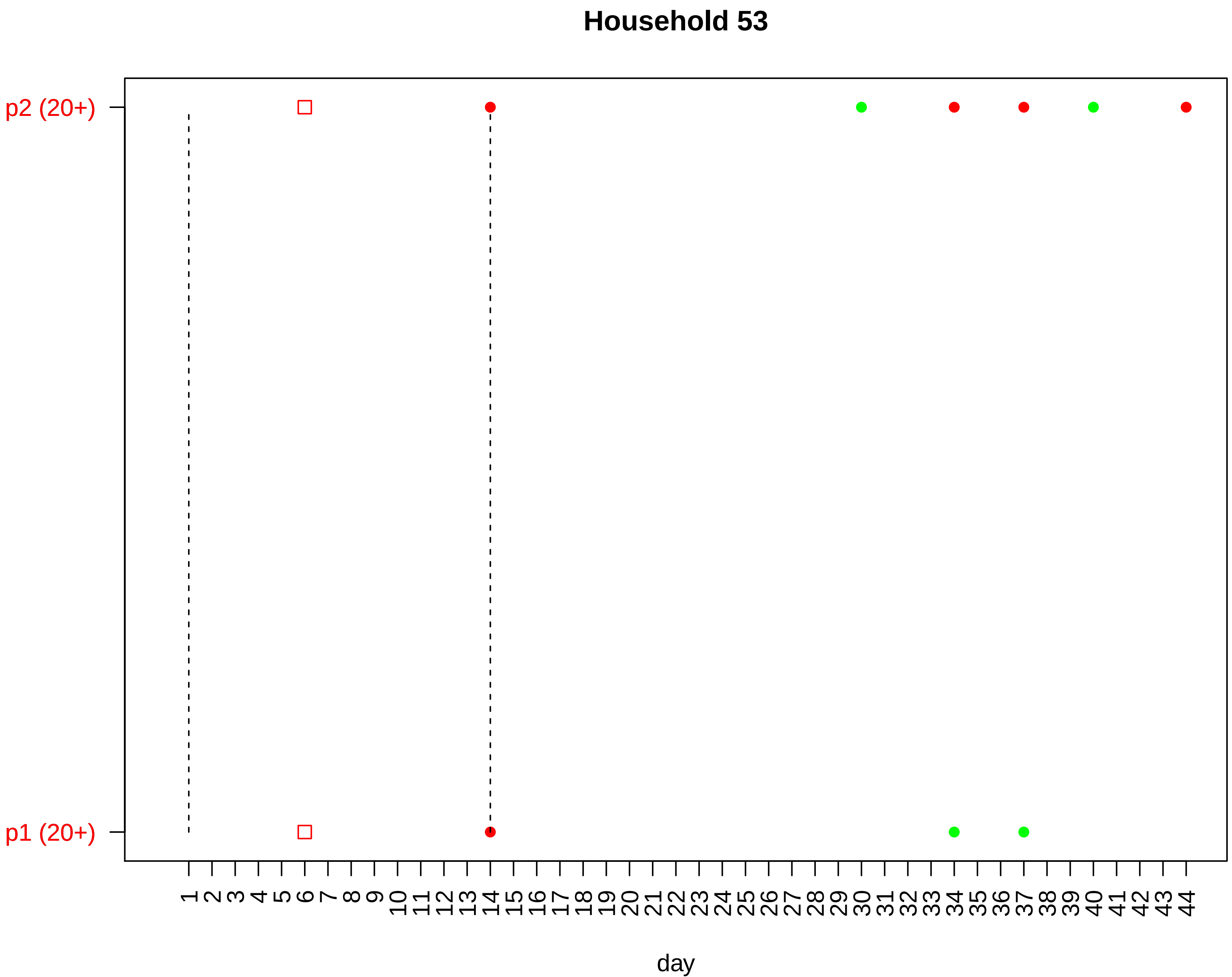

Household 53
p2 (20+)
p1 (20+)
day

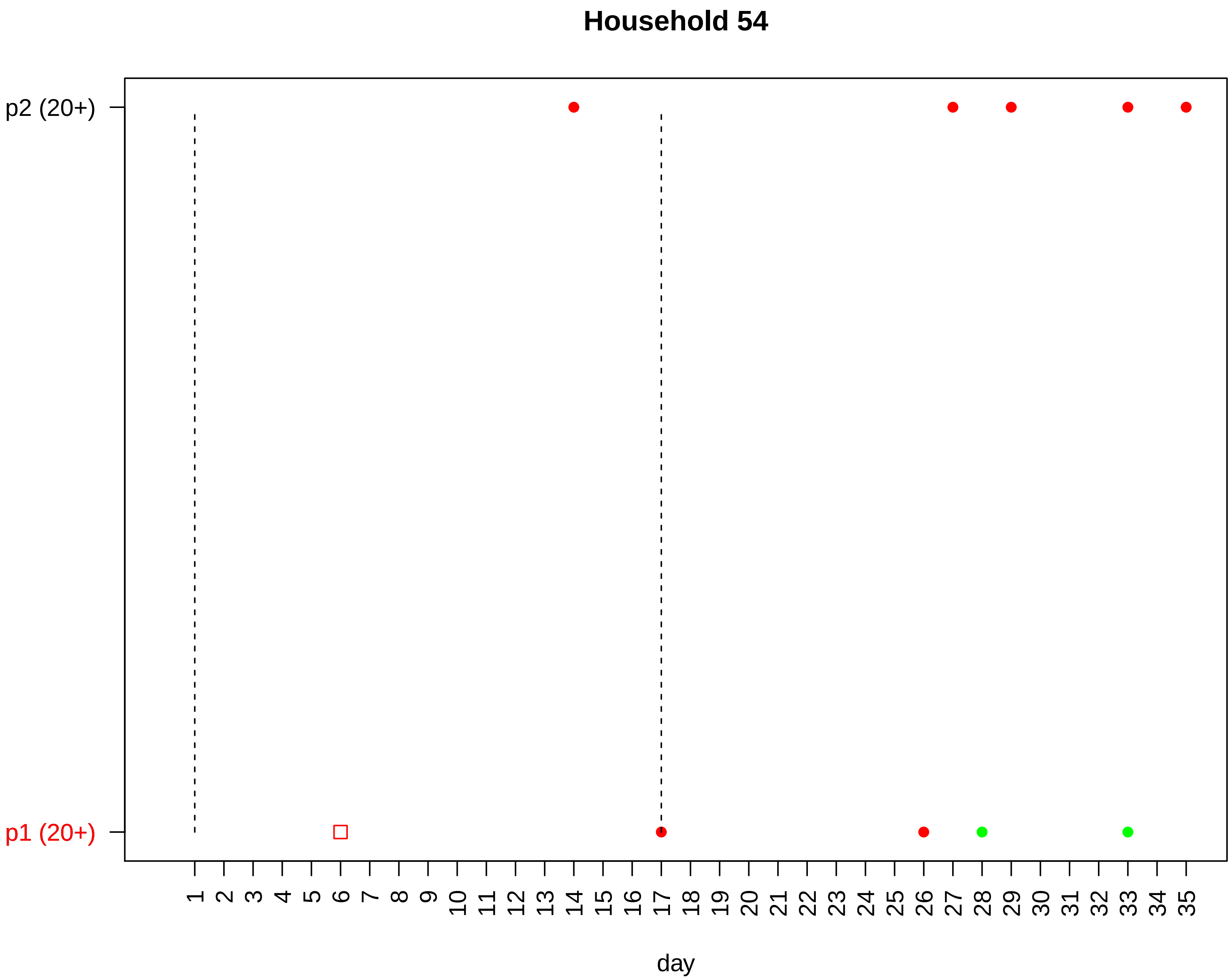

Household 54
p2 (20+)
p1 (20+)
day

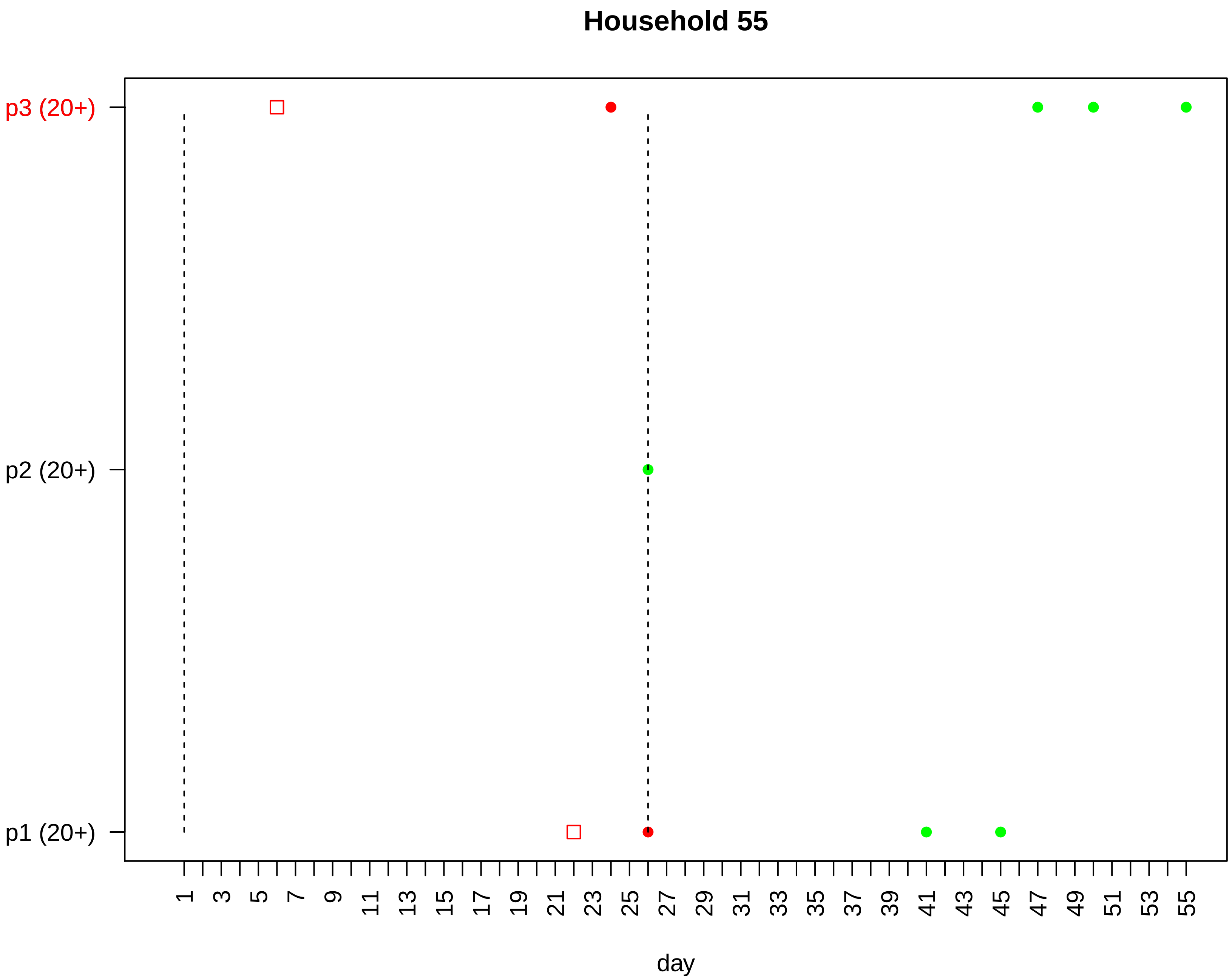

Household 55
p3 (20+)
p2 (20+)
p1 (20+)
1
3
5
7
9
11
13
15
17
19
21
23
25
27
29
31
33
35
37
39
41
43
45
47
49
51
53
55
day

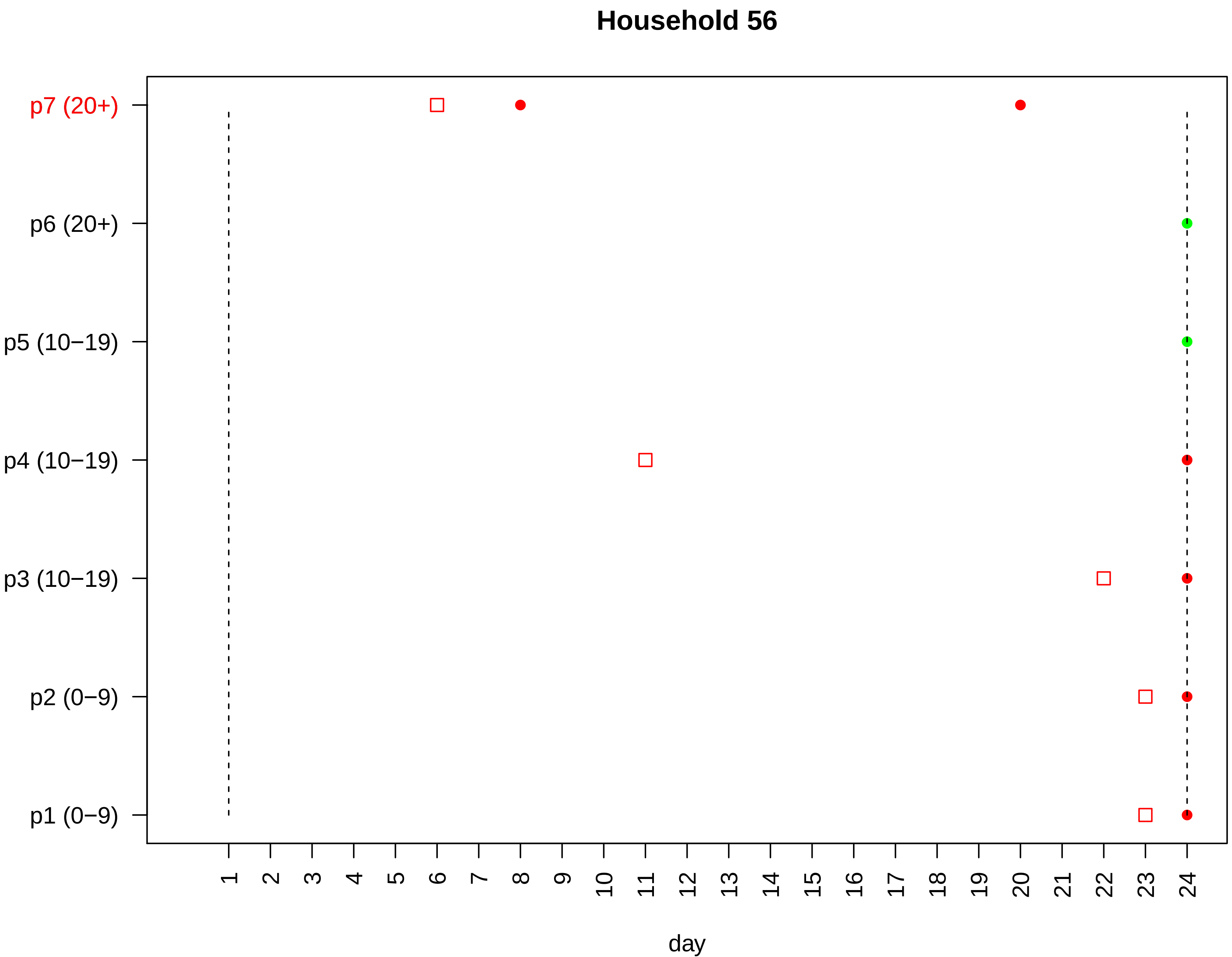

Household 56
p7 (20+)
p6 (20+)
p5 (10−19)
p4 (10−19)
p3 (10−19)
p2 (0−9)
p1 (0−9)
1 2 3 4 5 6 7 8 9 10 11 12 13 14 15 16 17 18 19 20 21 22 23 24
day

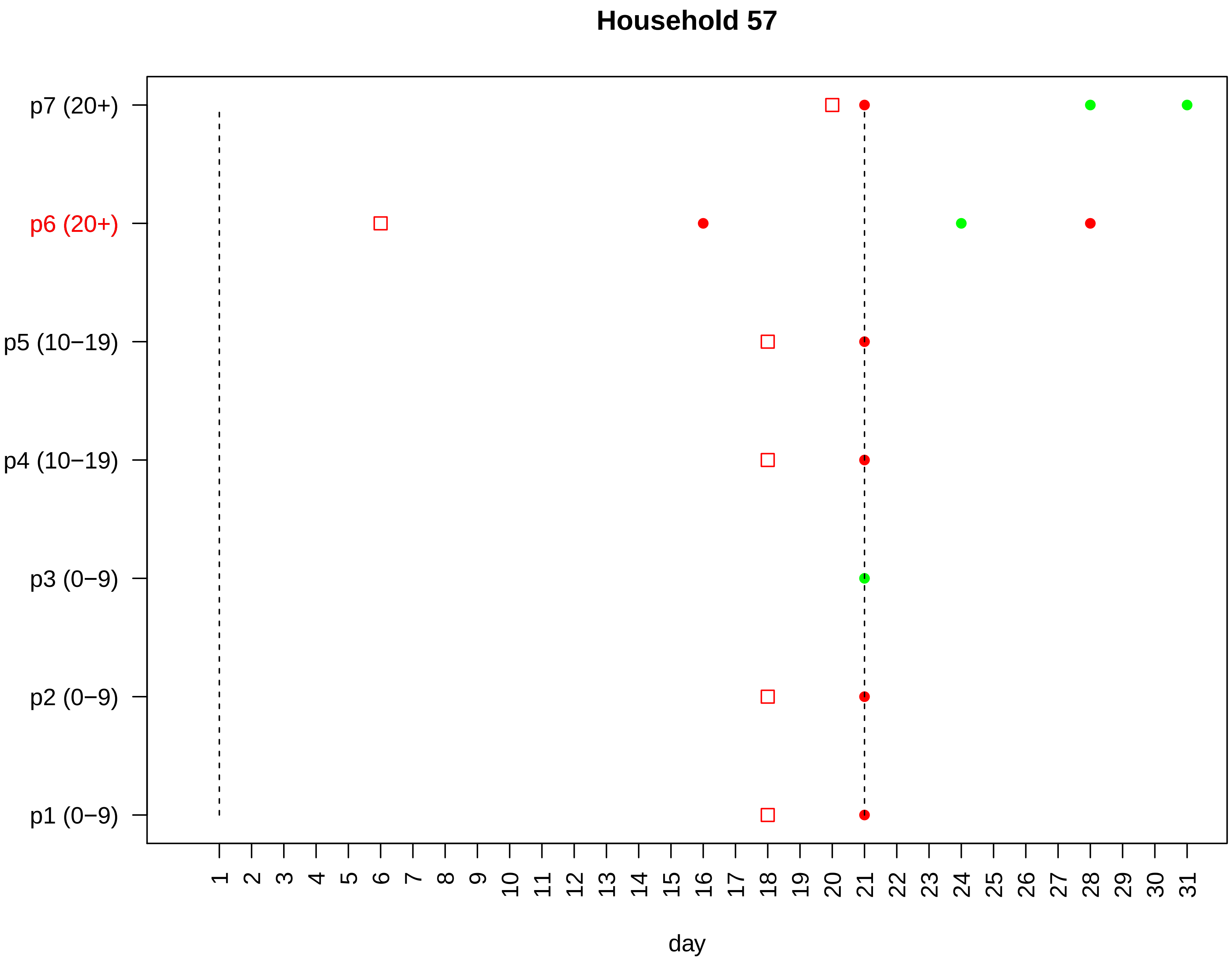

Household 57
p7 (20+)
p6 (20+)
p5 (10−19)
p4 (10−19)
p3 (0−9)
p2 (0−9)
p1 (0−9)
day

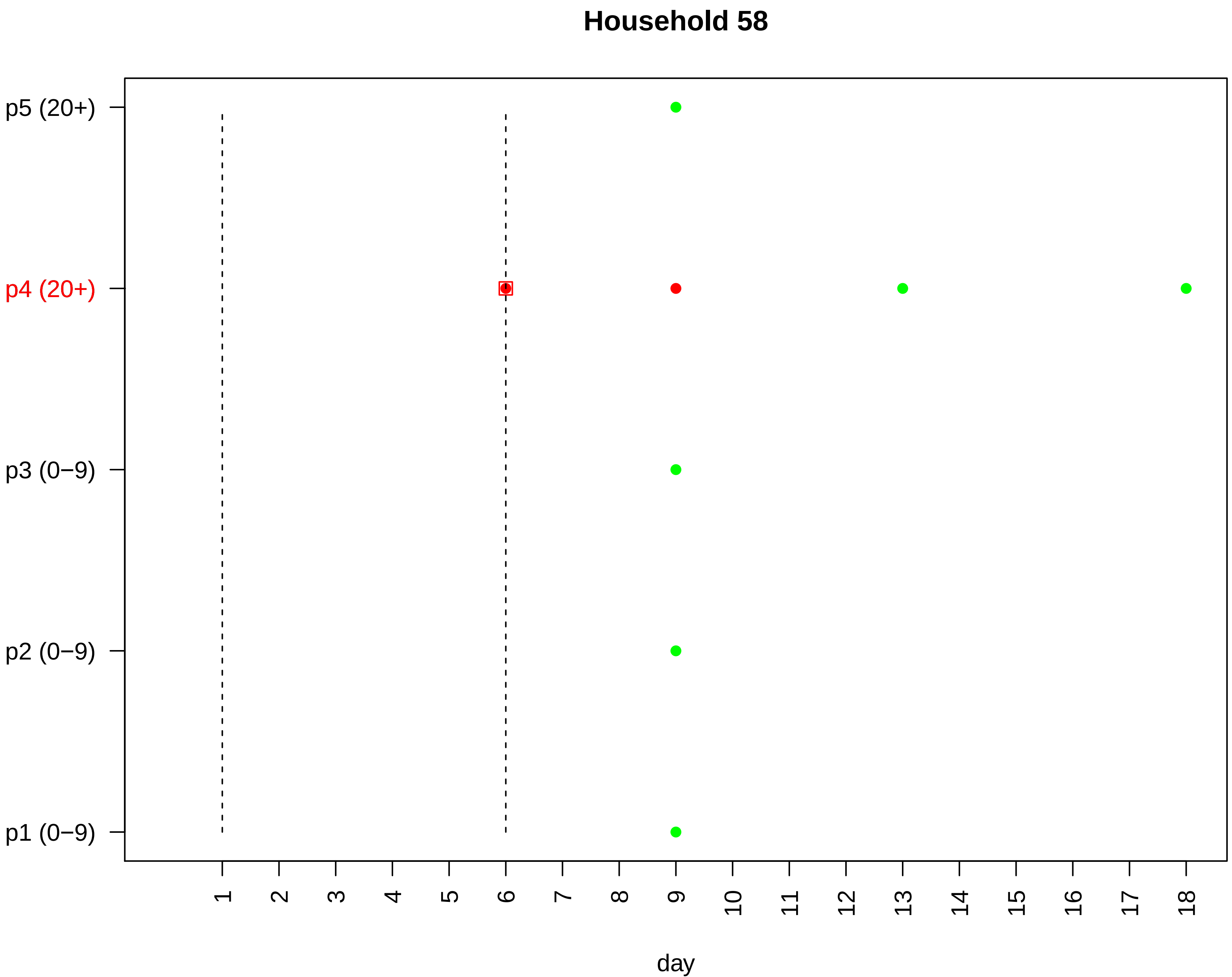
Household 58
p5 (20+)
p4 (20+)
p3 (0−9)
p2 (0−9)
p1 (0−9)
day

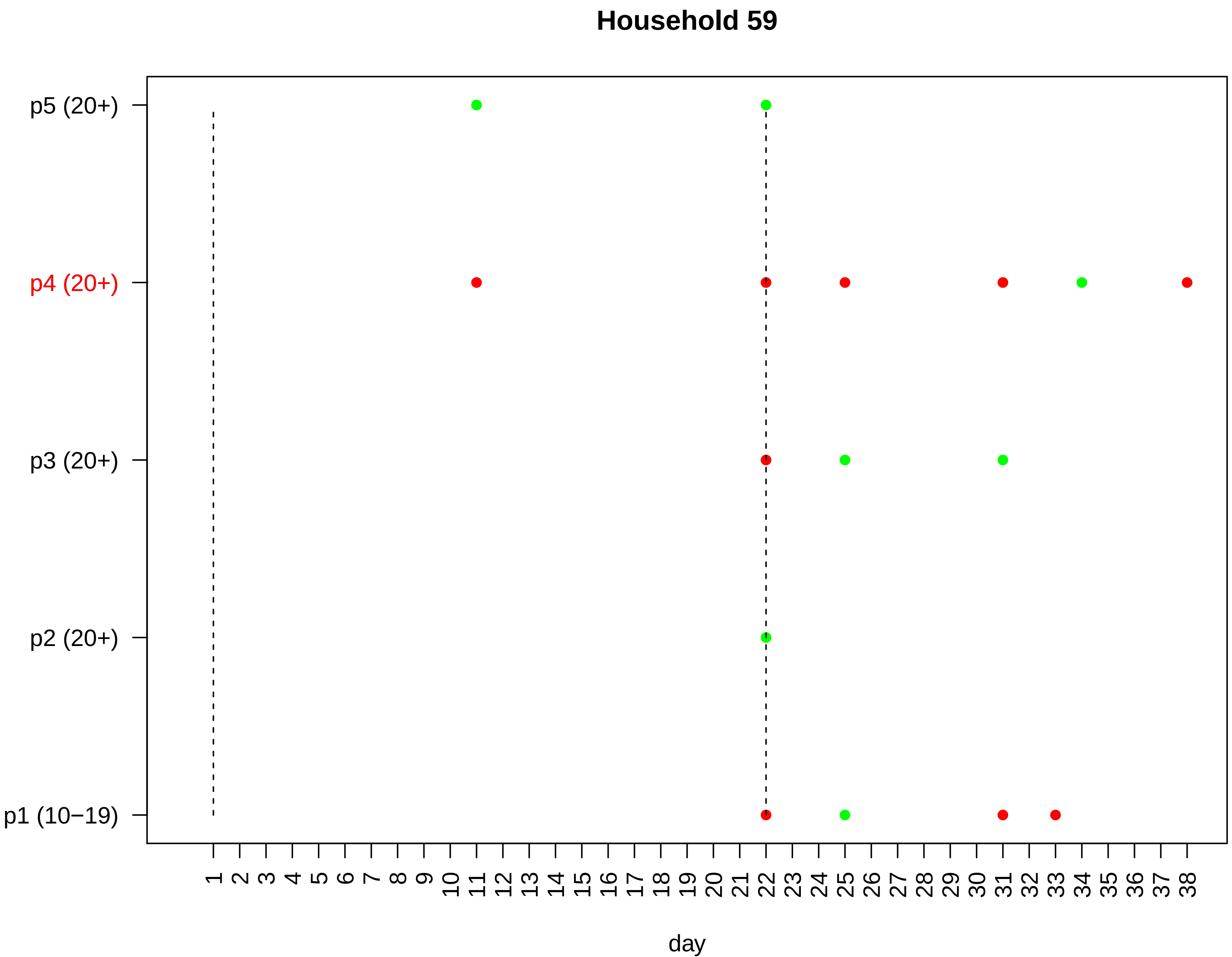

Household 59
p5 (20+)
p4 (20+)
p3 (20+)
p2 (20+)
p1 (10−19)
1 2 3 4 5 6 7 8 9 10 11 12 13 14 15 16 17 18 19 20 21 22 23 24 25 26 27 28 29 30 31 32 33 34 35 36 37 38
day

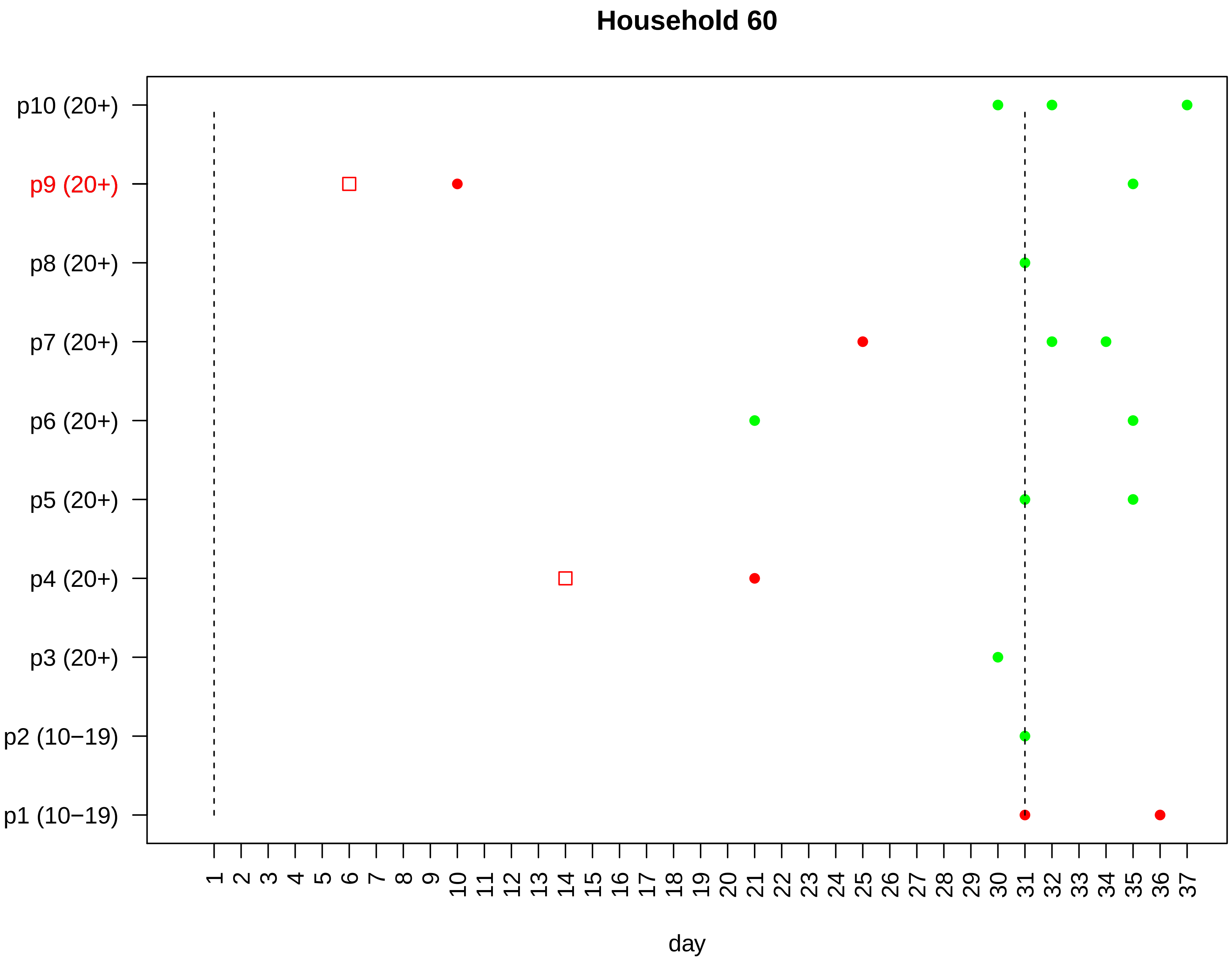

Household 60
p10 (20+)
p9 (20+)
p8 (20+)
p7 (20+)
p6 (20+)
p5 (20+)
p4 (20+)
p3 (20+)
p2 (10−19)
p1 (10−19)
1 2 3 4 5 6 7 8 9 10 11 12 13 14 15 16 17 18 19 20 21 22 23 24 25 26 27 28 29 30 31 32 33 34 35 36 37
day

#### Household 61

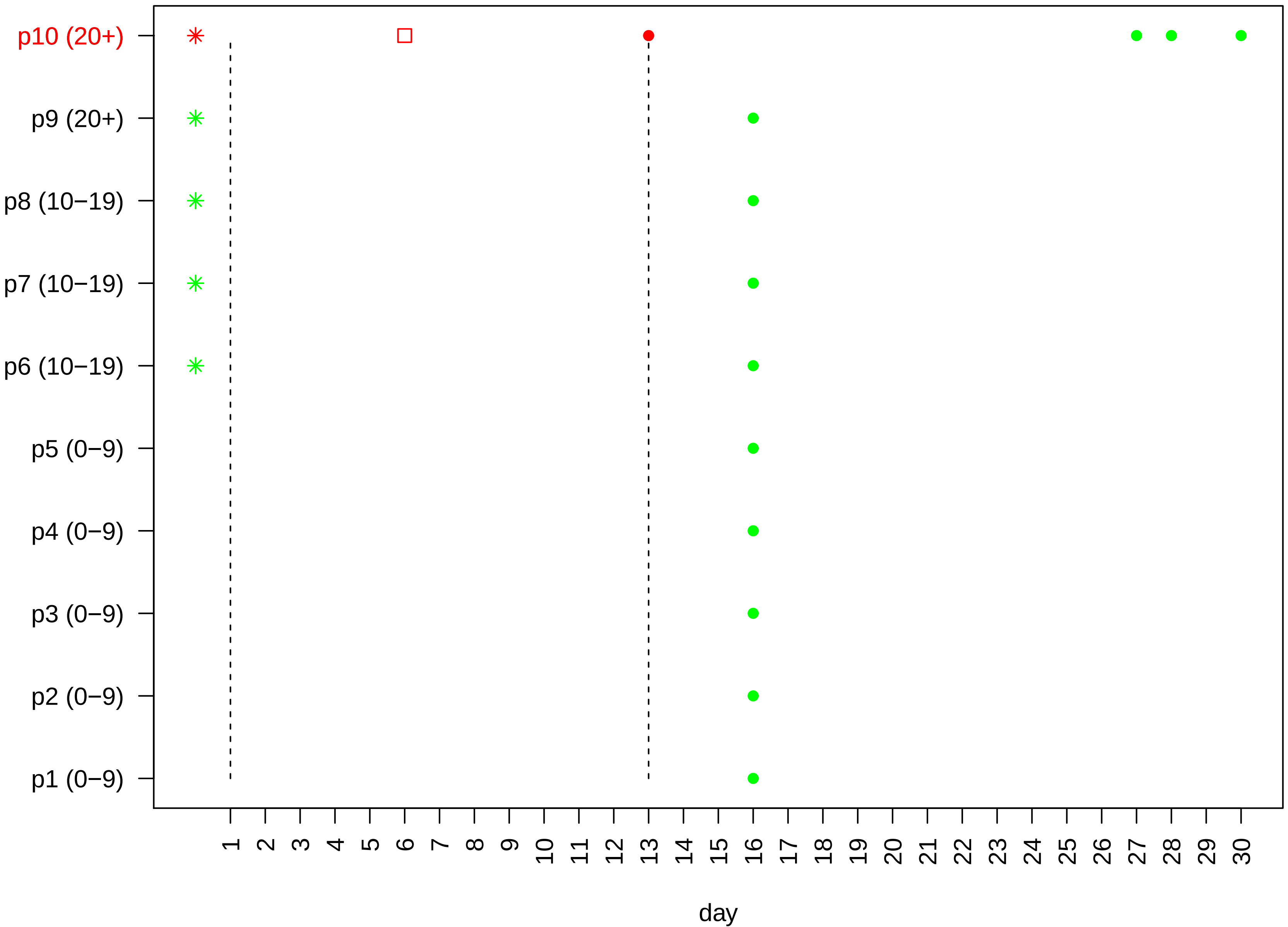

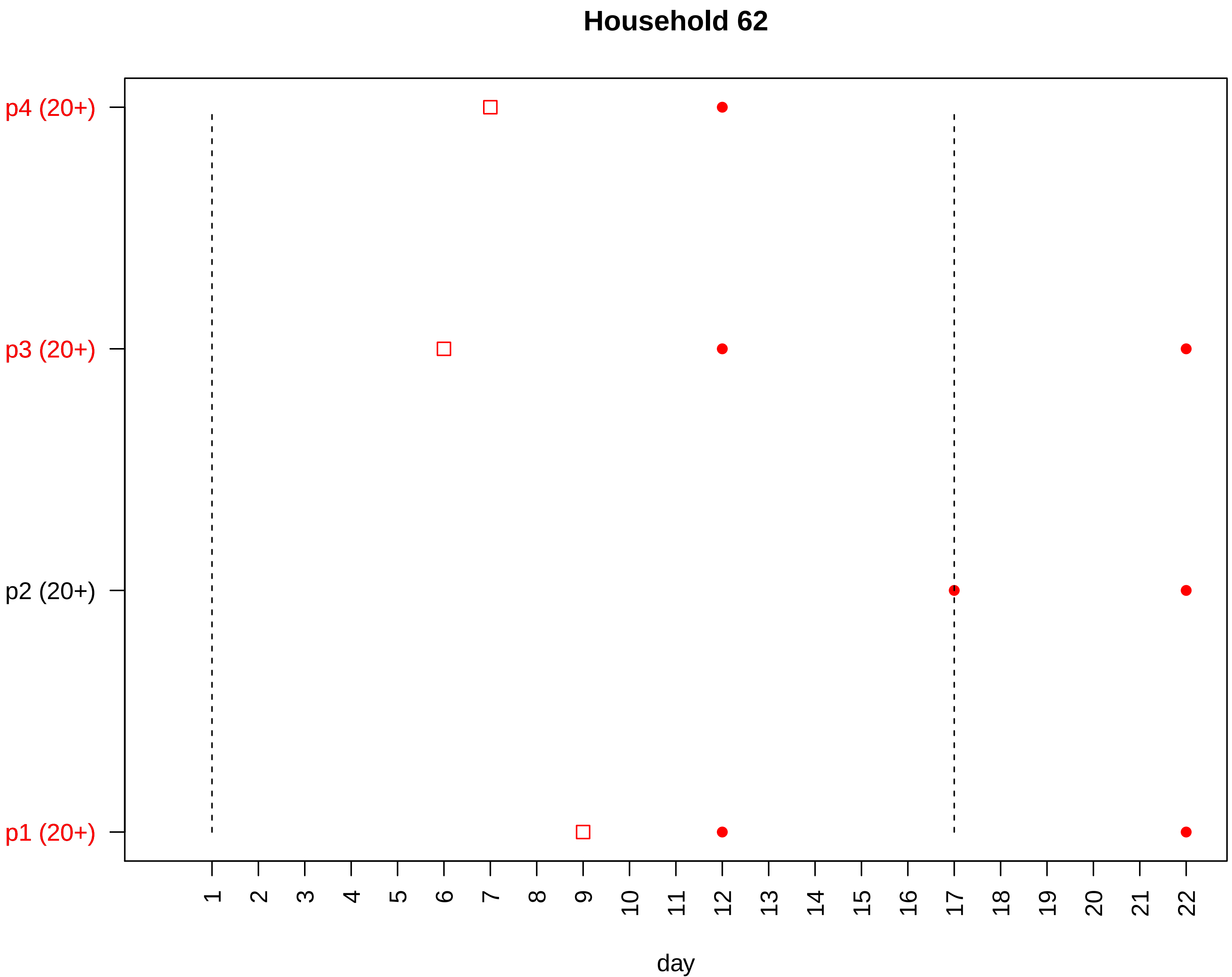

Household 62
p4 (20+)
p3 (20+)
p2 (20+)
p1 (20+)
day

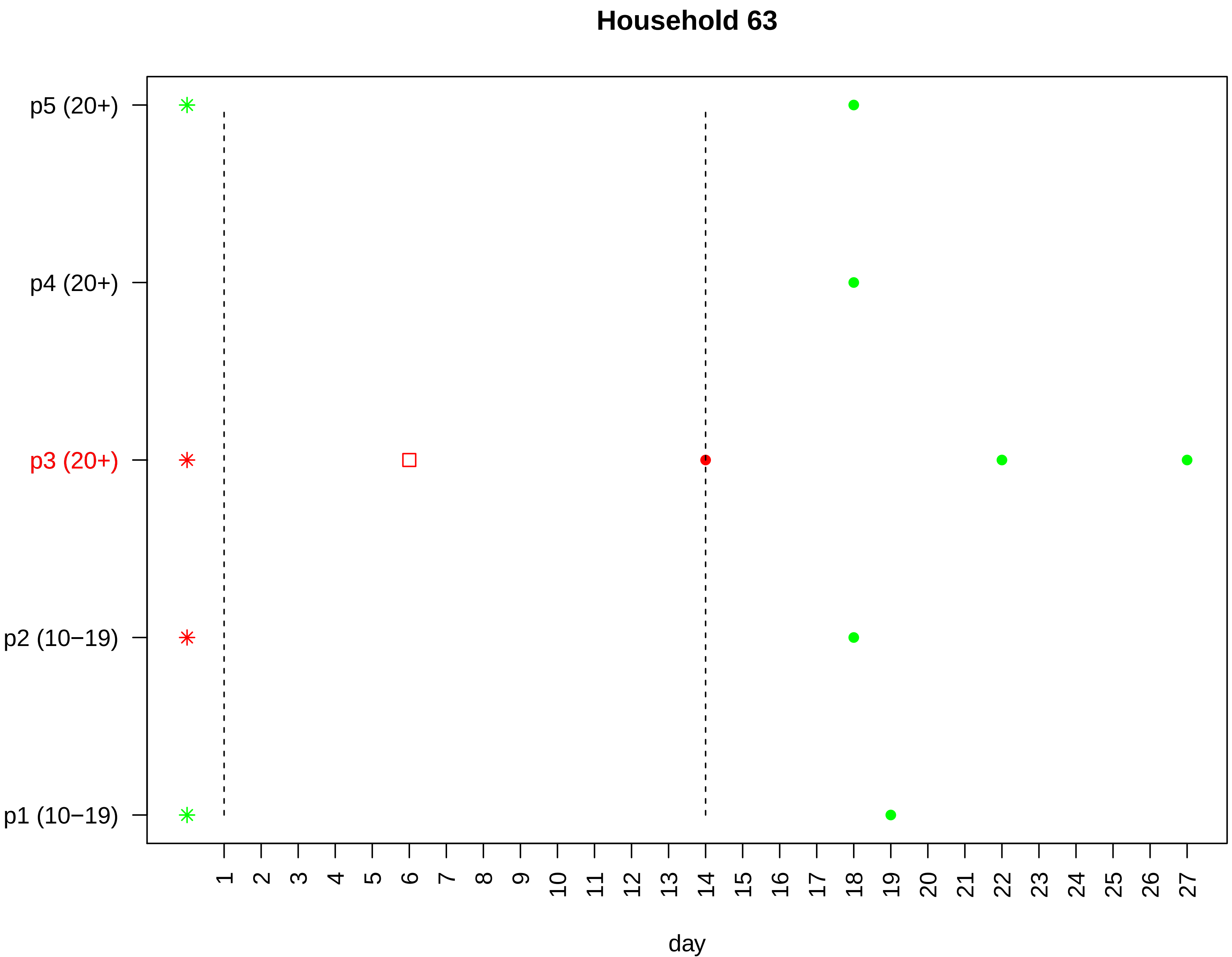
Household 63
p5 (20+)
p4 (20+)
p3 (20+)
p2 (10−19)
p1 (10−19)
day

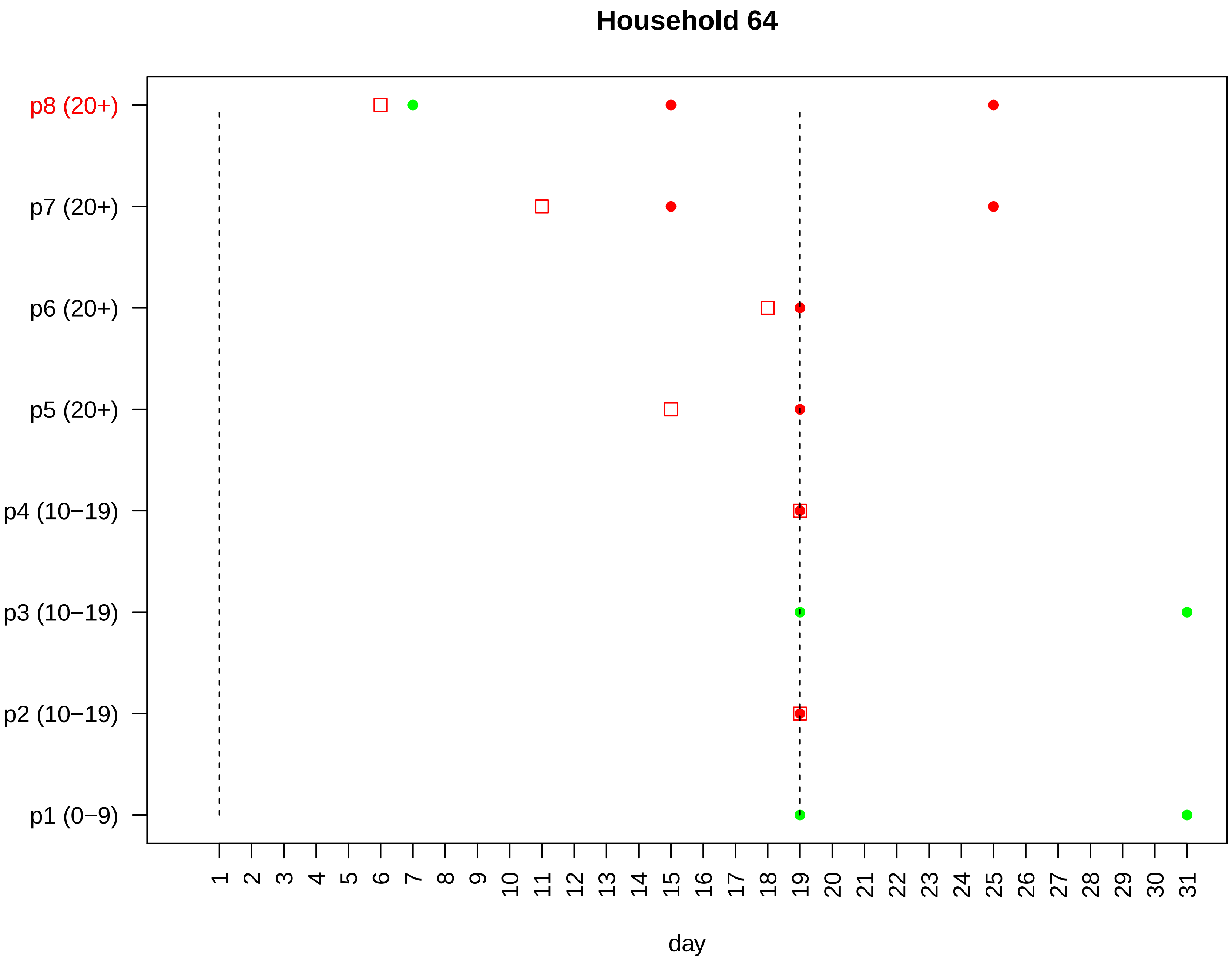

Household 64
p8 (20+)
p7 (20+)
p6 (20+)
p5 (20+)
p4 (10−19)
p3 (10−19)
p2 (10−19)
p1 (0−9)
1 2 3 4 5 6 7 8 9 10 11 12 13 14 15 16 17 18 19 20 21 22 23 24 25 26 27 28 29 30 31
day

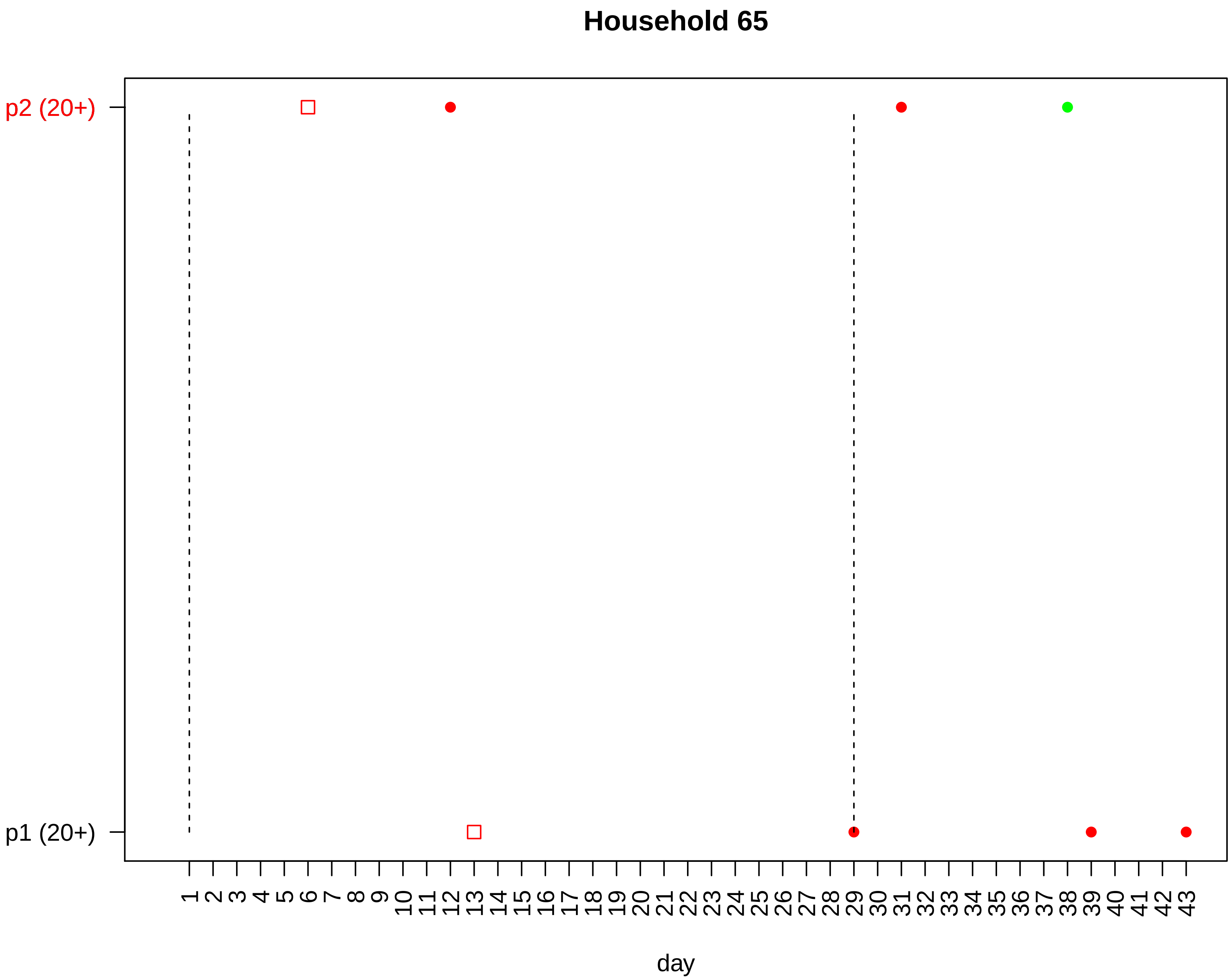

Household 65
p2 (20+)
p1 (20+)
1 2 3 4 5 6 7 8 9 10 11 12 13 14 15 16 17 18 19 20 21 22 23 24 25 26 27 28 29 30 31 32 33 34 35 36 37 38 39 40 41 42 43
day

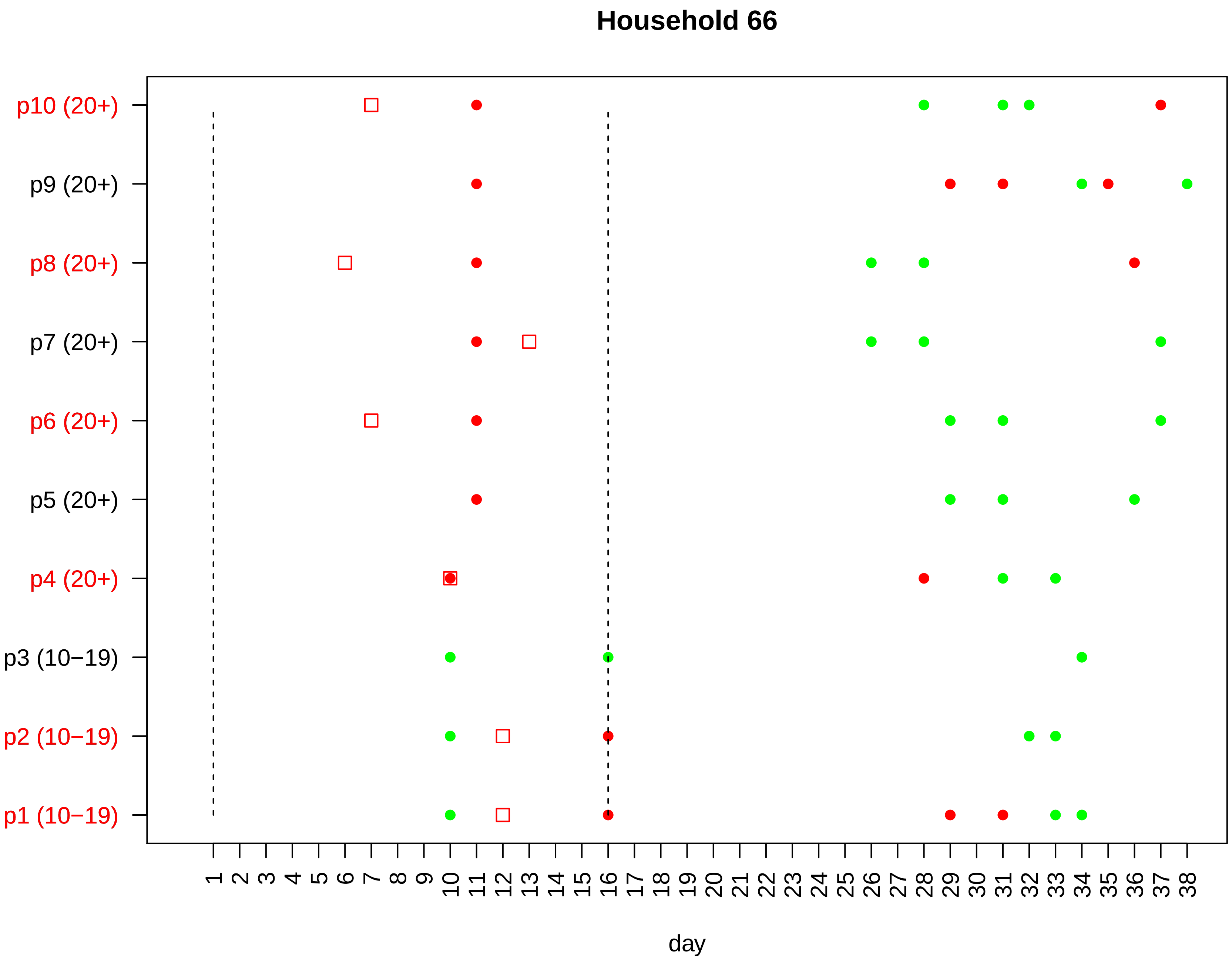

Household 66
p10 (20+)
p9 (20+)
p8 (20+)
p7 (20+)
p6 (20+)
p5 (20+)
p4 (20+)
p3 (10−19)
p2 (10−19)
p1 (10−19)
1 2 3 4 5 6 7 8 9 10 11 12 13 14 15 16 17 18 19 20 21 22 23 24 25 26 27 28 29 30 31 32 33 34 35 36 37 38
day

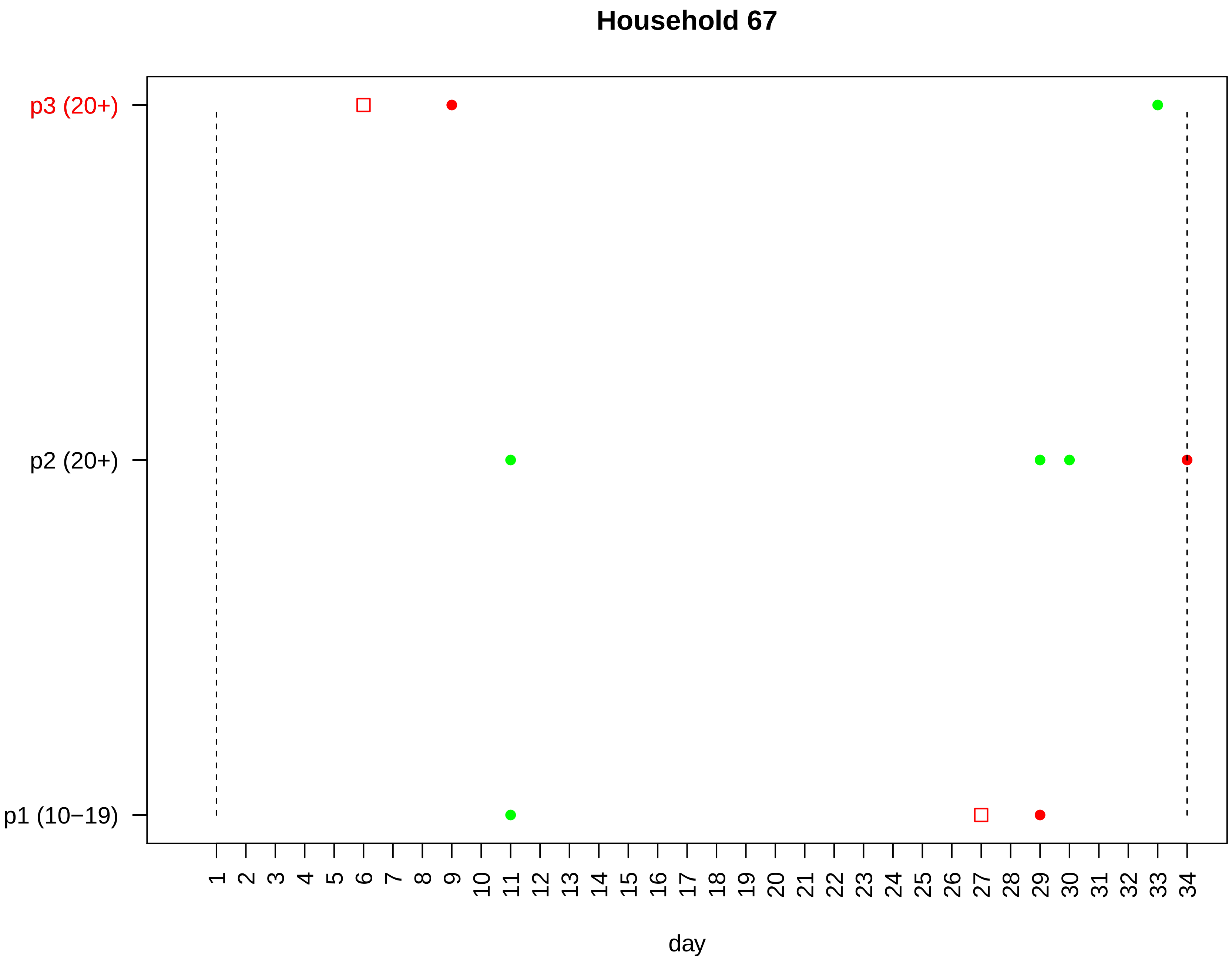

Household 67
p3 (20+)
p2 (20+)
p1 (10−19)
day

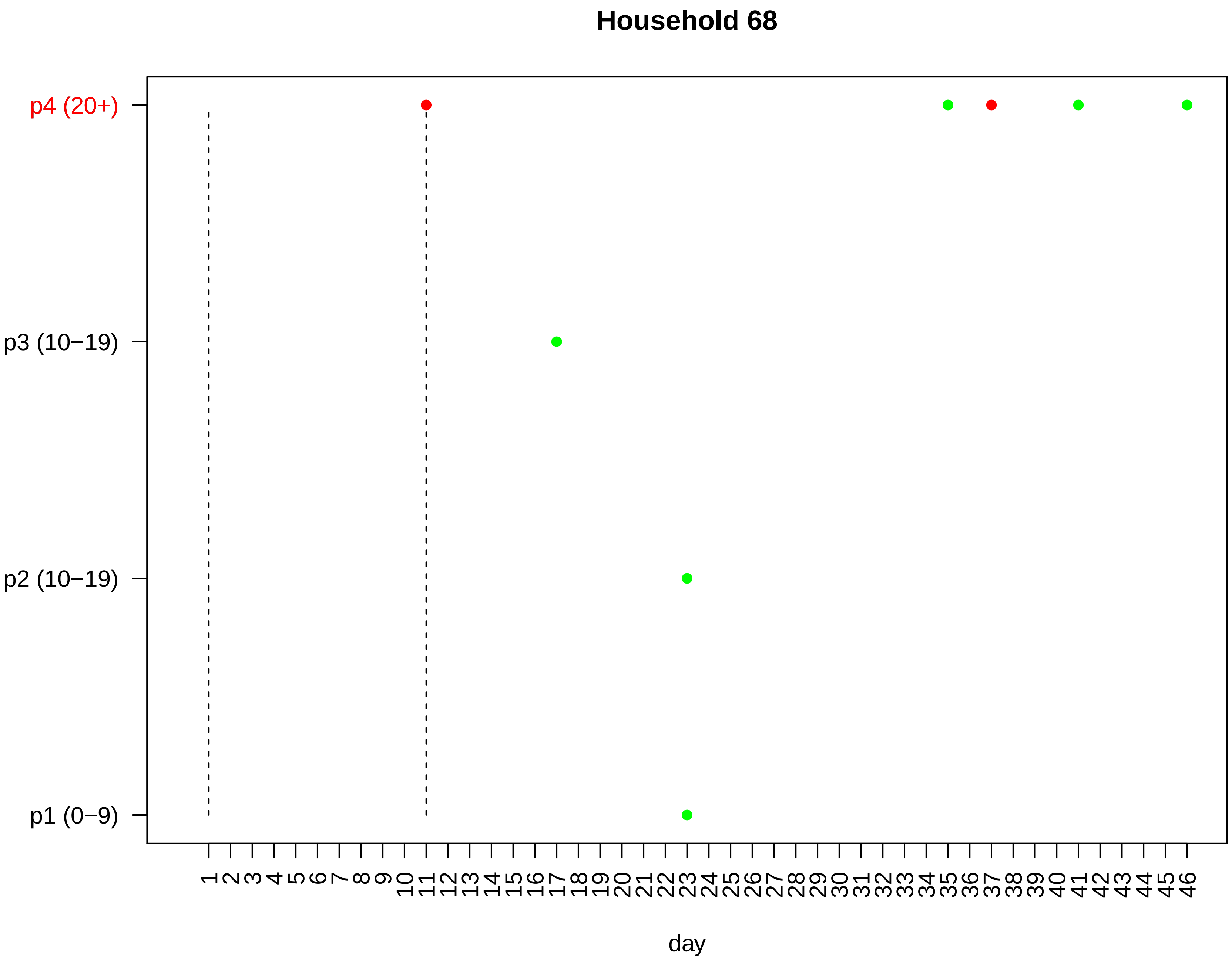

Household 68
p4 (20+)
p3 (10−19)
p2 (10−19)
p1 (0−9)
1 2 3 4 5 6 7 8 9 10 11 12 13 14 15 16 17 18 19 20 21 22 23 24 25 26 27 28 29 30 31 32 33 34 35 36 37 38 39 40 41 42 43 44 45 46
day

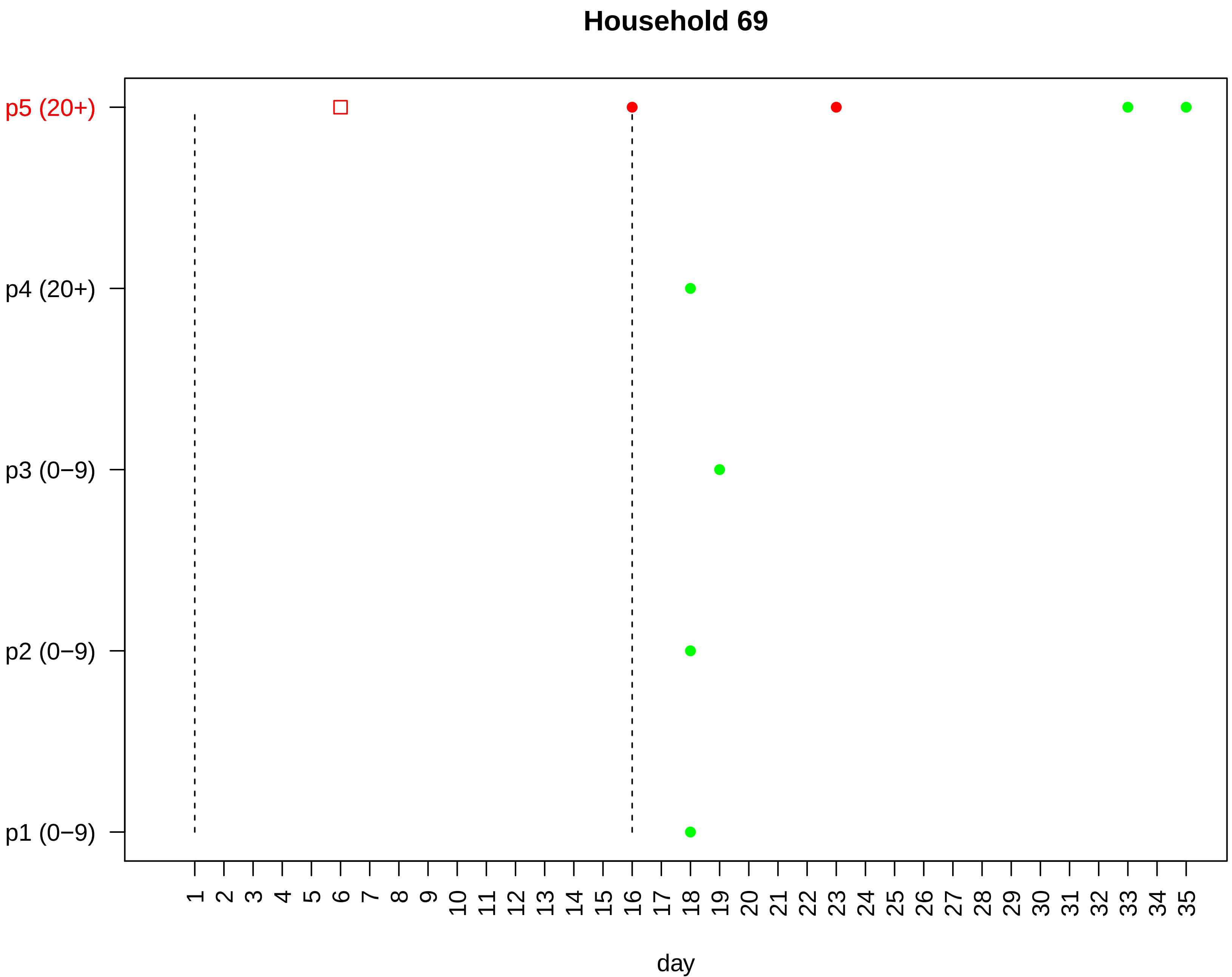
Household 69
p5 (20+)
p4 (20+)
p3 (0−9)
p2 (0−9)
p1 (0−9)
1 2 3 4 5 6 7 8 9 10 11 12 13 14 15 16 17 18 19 20 21 22 23 24 25 26 27 28 29 30 31 32 33 34 35
day

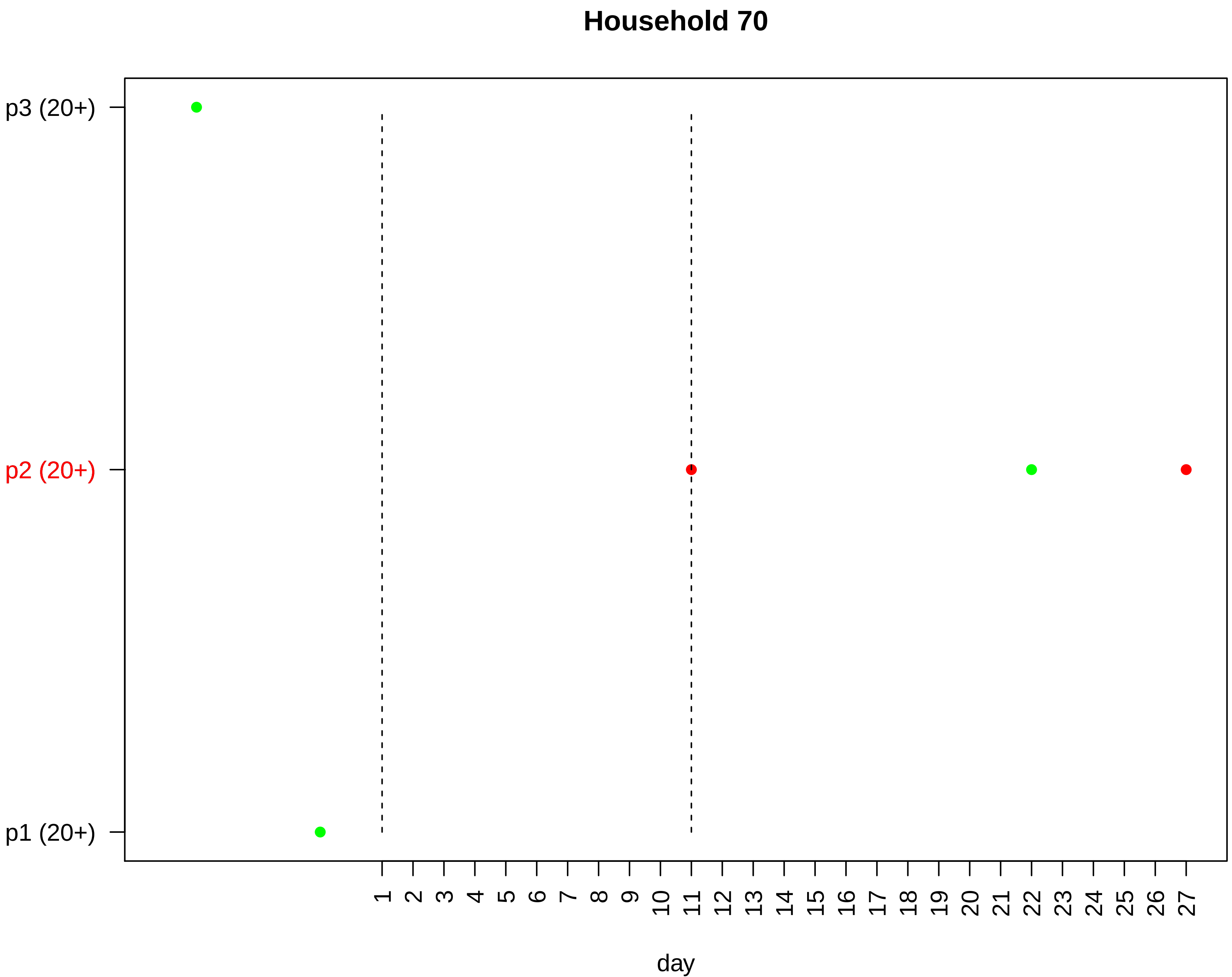

Household 70
p3 (20+)
p2 (20+)
p1 (20+)
day

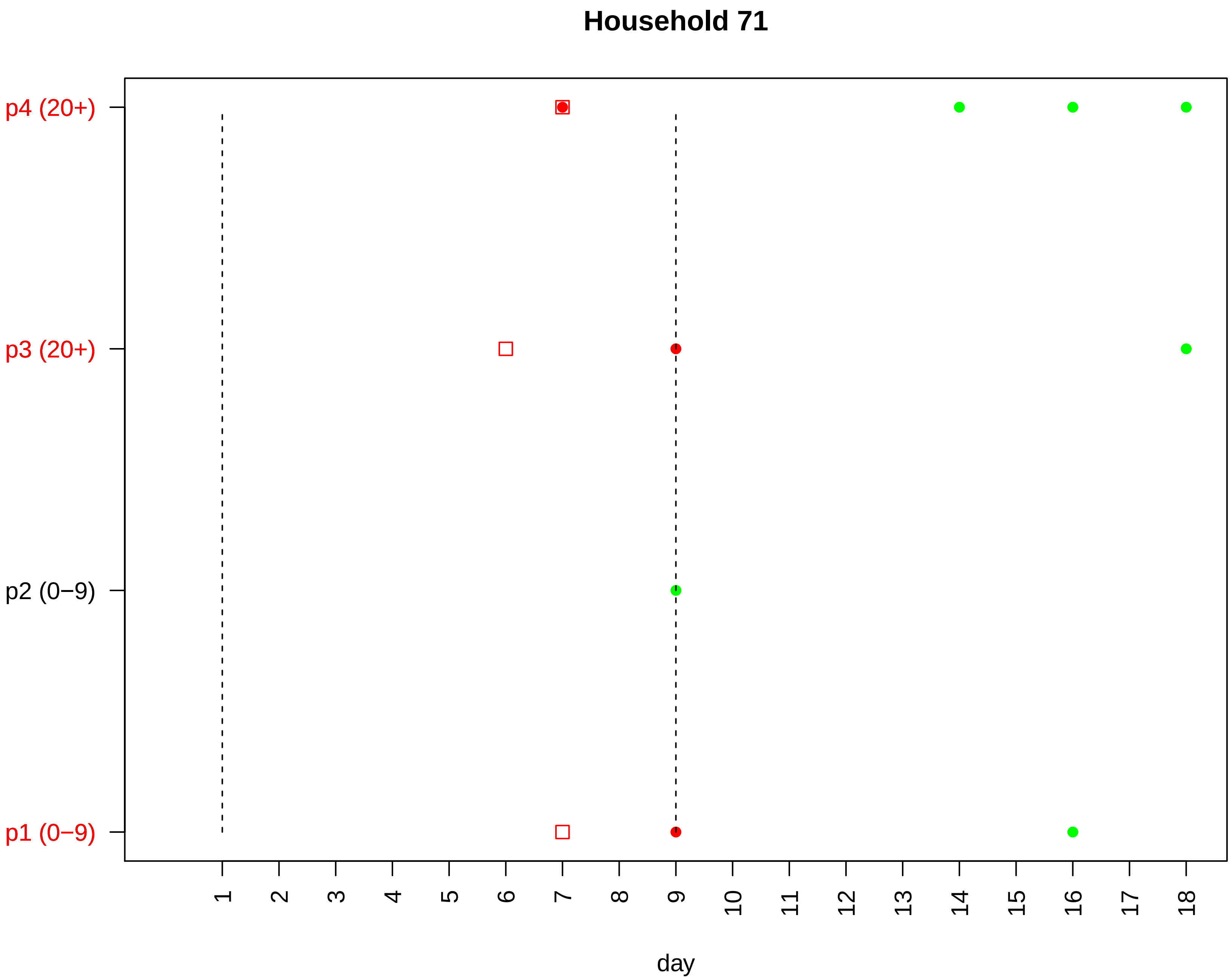

Household 71
p4 (20+)
p3 (20+)
p2 (0−9)
p1 (0−9)
day

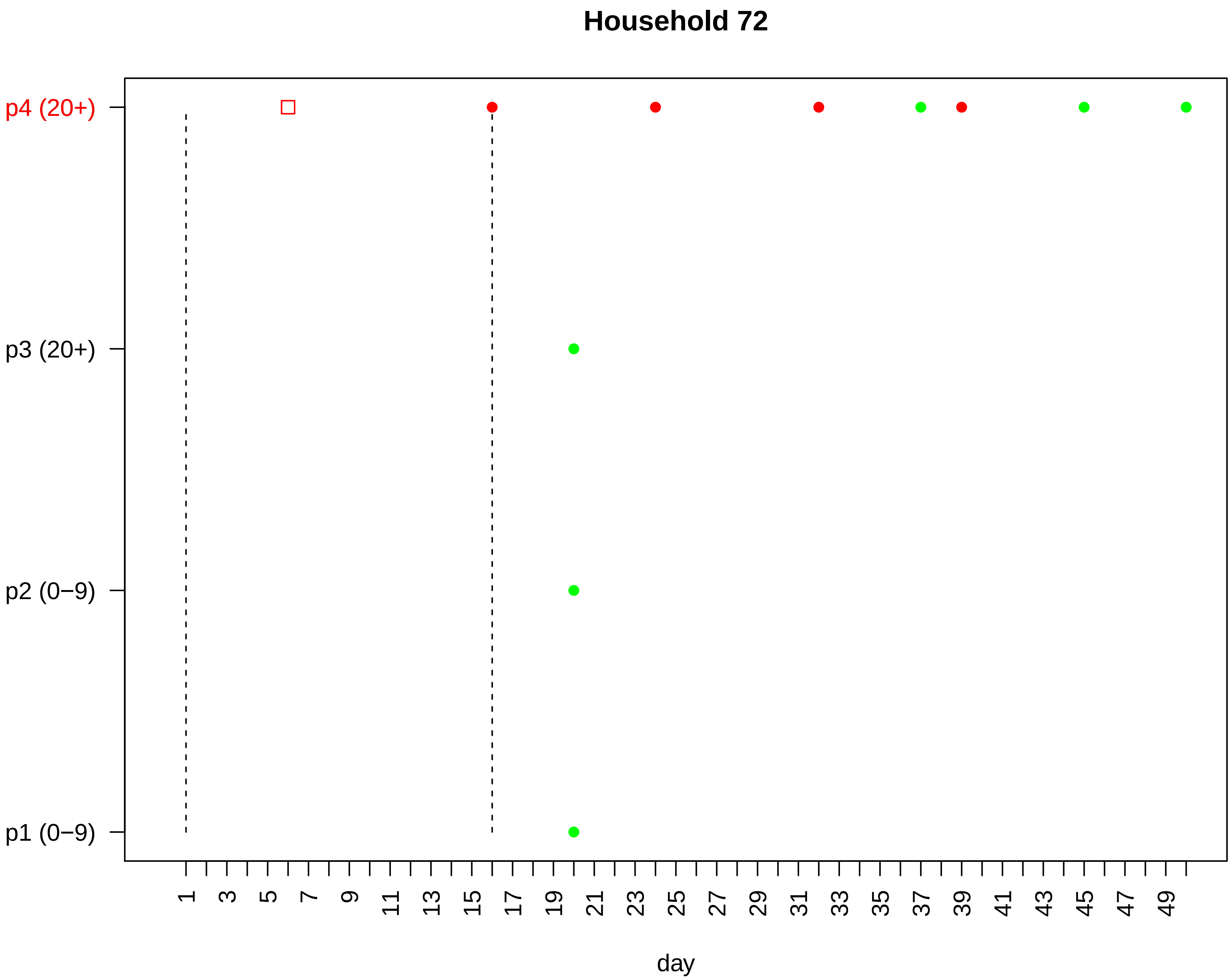
Household 72
p4 (20+)
p3 (20+)
p2 (0−9)
p1 (0−9)
1
3
5
7
9
11
13
15
17
19
21
23
25
27
29
31
33
35
37
39
41
43
45
47
49
day

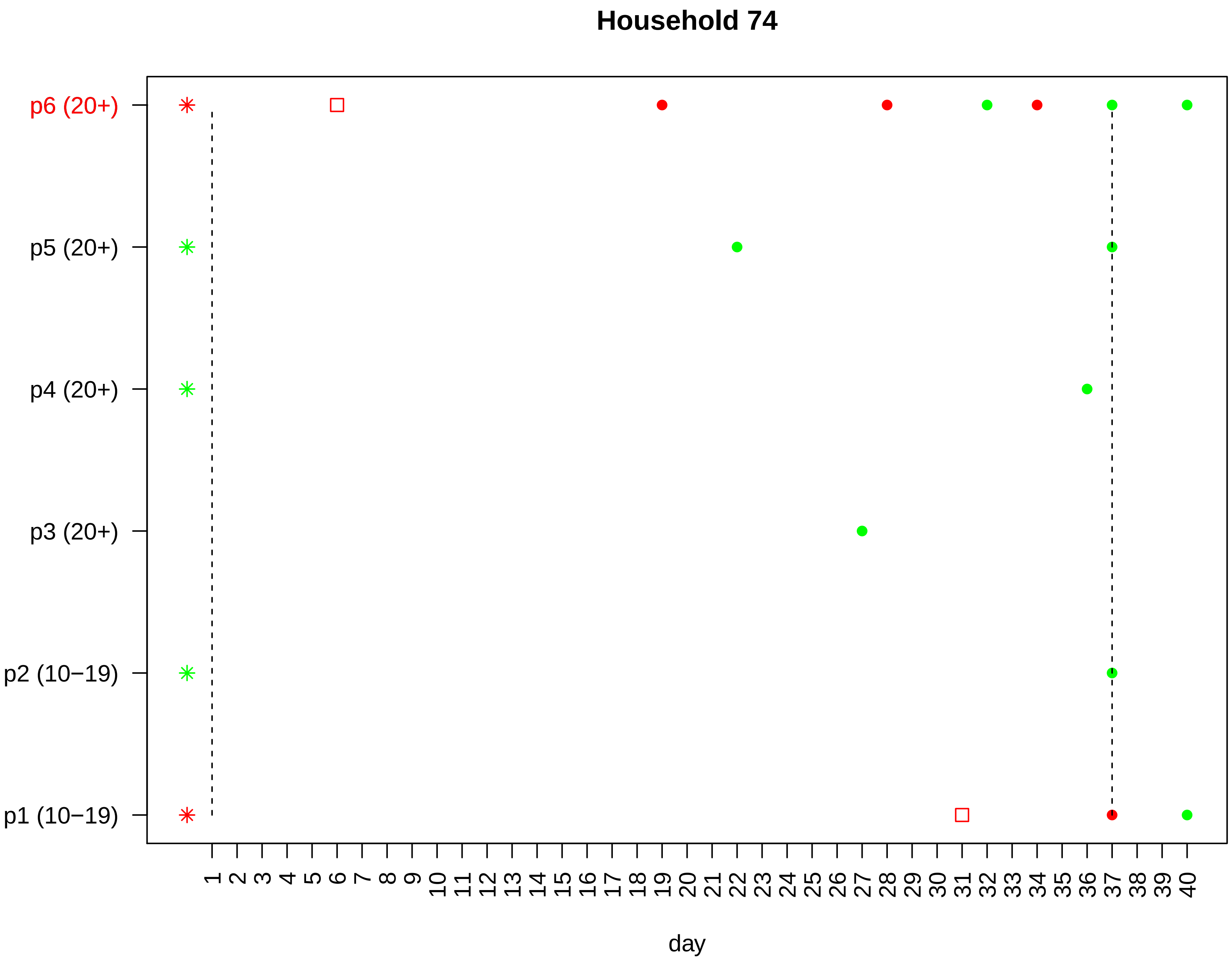

Household 74
p6 (20+)
p5 (20+)
p4 (20+)
p3 (20+)
p2 (10−19)
p1 (10−19)
1 2 3 4 5 6 7 8 9 10 11 12 13 14 15 16 17 18 19 20 21 22 23 24 25 26 27 28 29 30 31 32 33 34 35 36 37 38 39 40
day

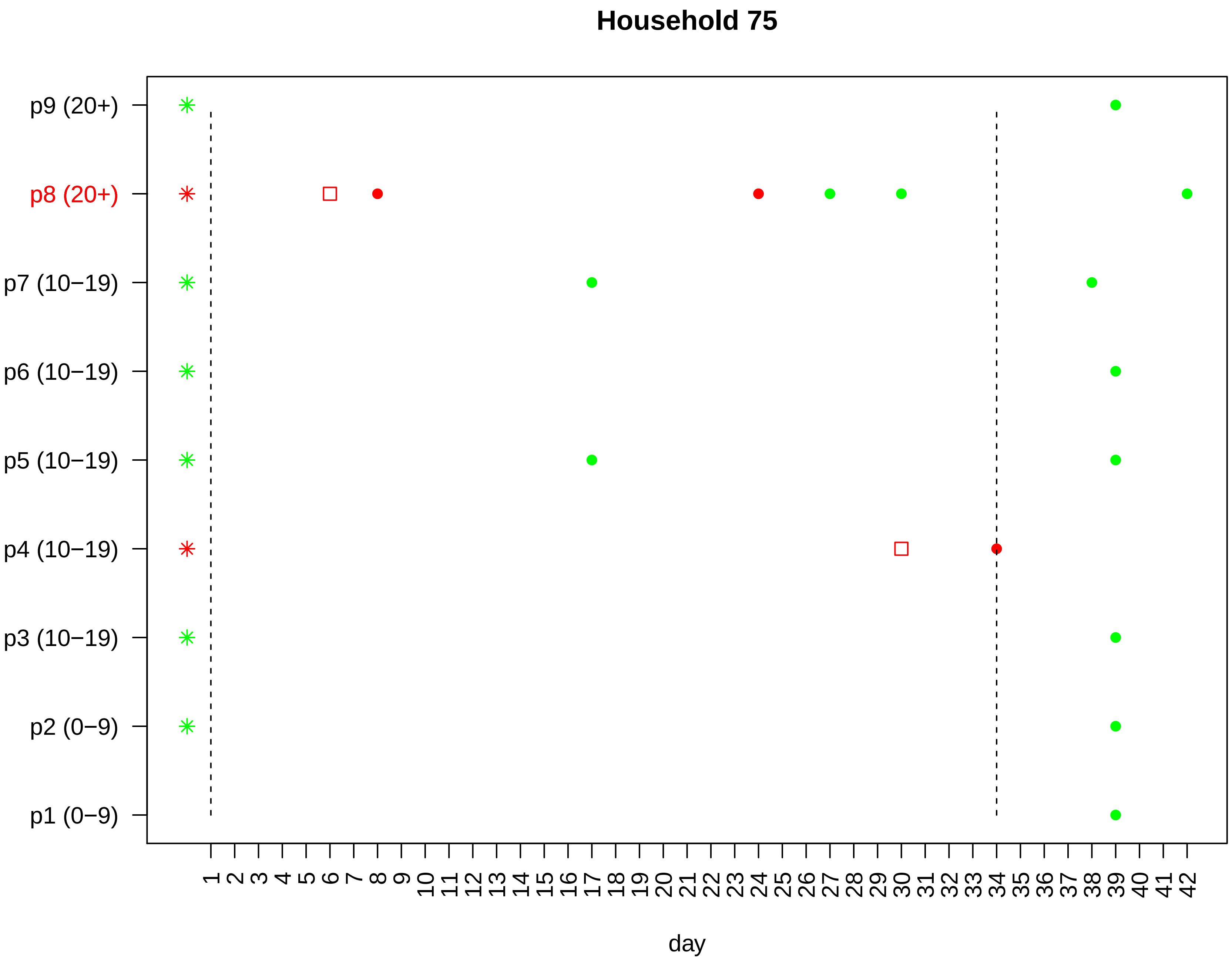

Household 75
p9 (20+)
p8 (20+)
p7 (10−19)
p6 (10−19)
p5 (10−19)
p4 (10−19)
p3 (10−19)
p2 (0−9)
p1 (0−9)
1 2 3 4 5 6 7 8 9 10 11 12 13 14 15 16 17 18 19 20 21 22 23 24 25 26 27 28 29 30 31 32 33 34 35 36 37 38 39 40 41 42
day

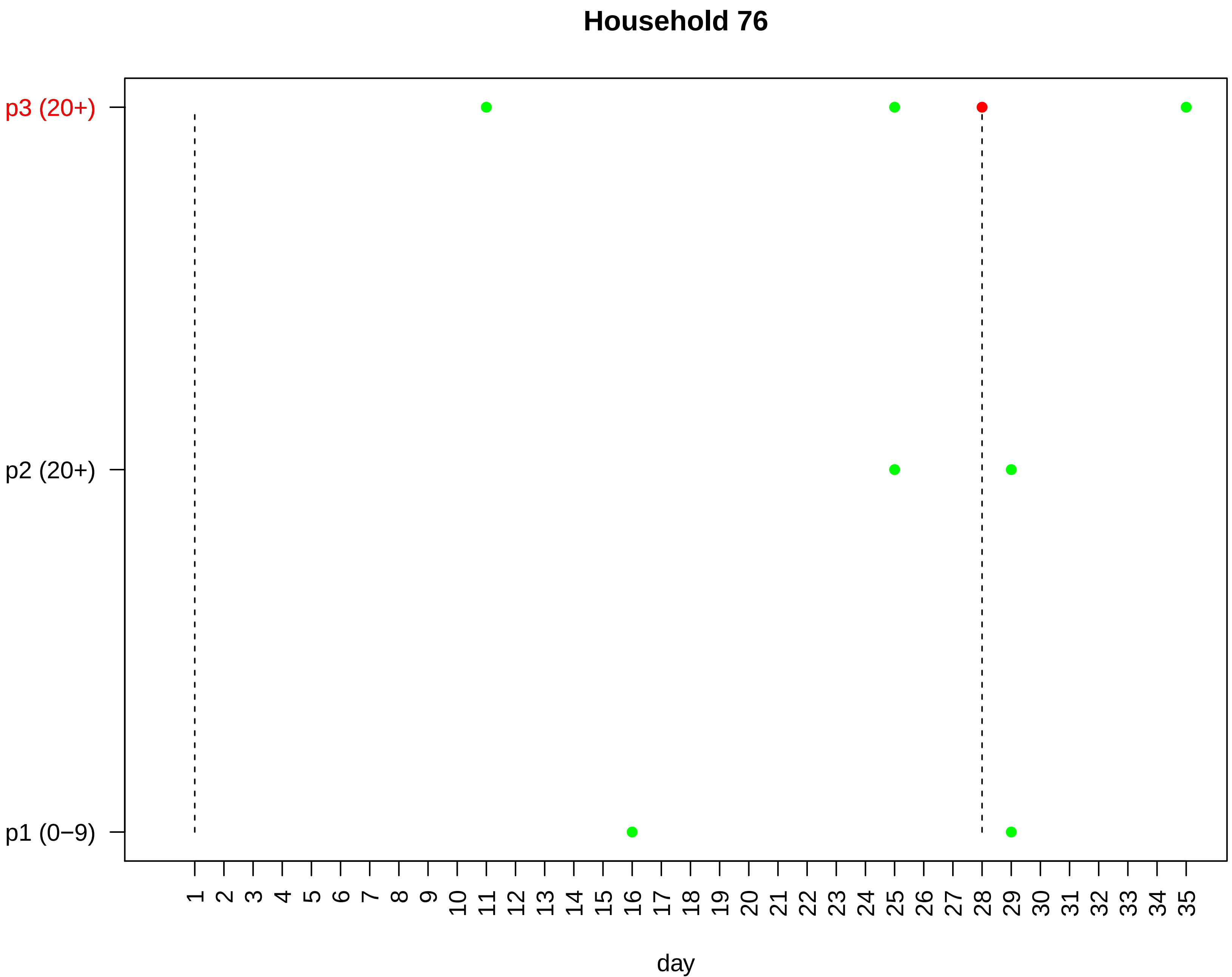
Household 76
p3 (20+)
p2 (20+)
p1 (0−9)
day

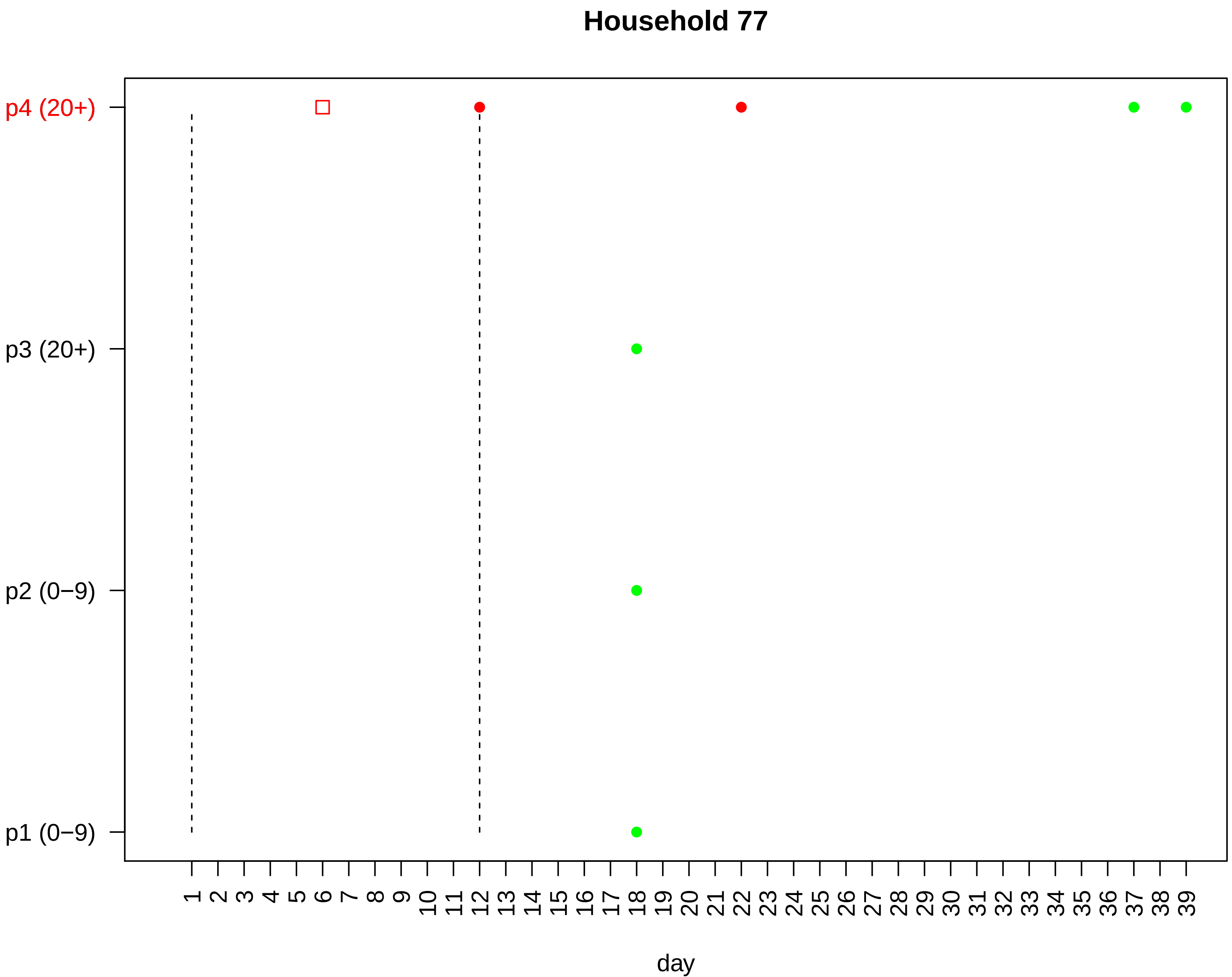
Household 77
p4 (20+)
p3 (20+)
p2 (0−9)
p1 (0−9)
1 2 3 4 5 6 7 8 9 10 11 12 13 14 15 16 17 18 19 20 21 22 23 24 25 26 27 28 29 30 31 32 33 34 35 36 37 38 39
day

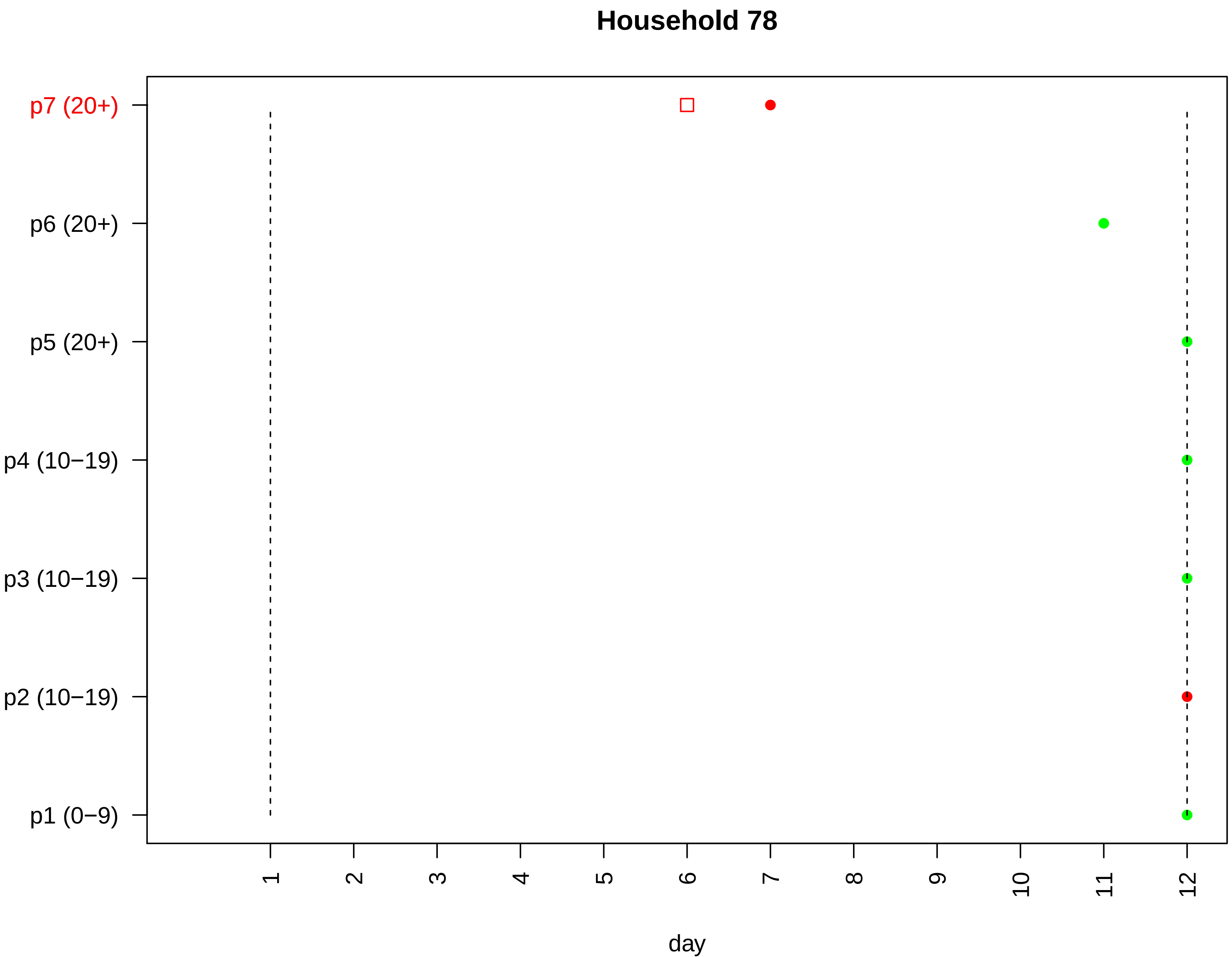

Household 78
p7 (20+)
p6 (20+)
p5 (20+)
p4 (10−19)
p3 (10−19)
p2 (10−19)
p1 (0−9)
1
2
3
4
5
6
7
8
9
10
11
12
day

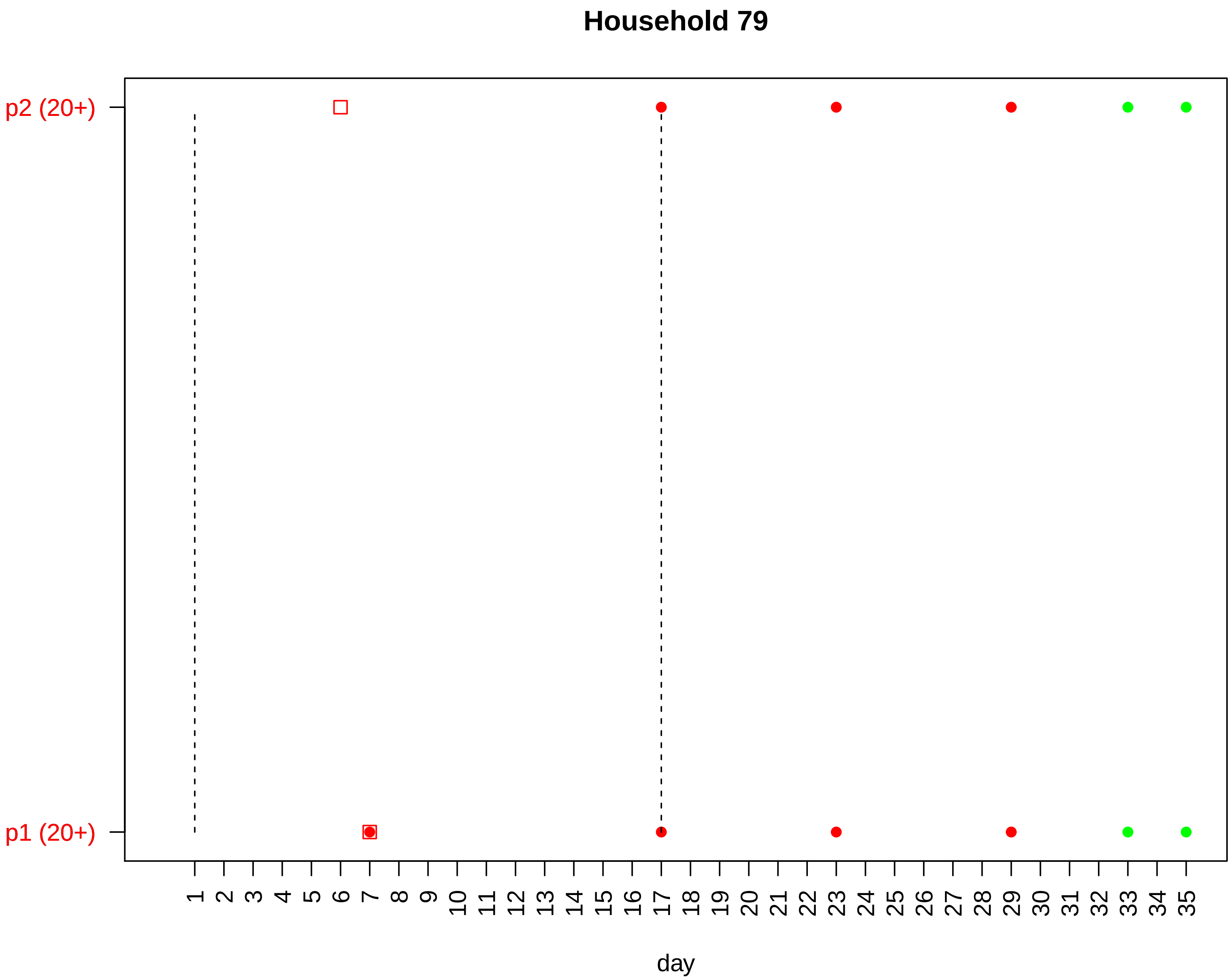

Household 79
p2 (20+)
p1 (20+)
day

#### Household 80

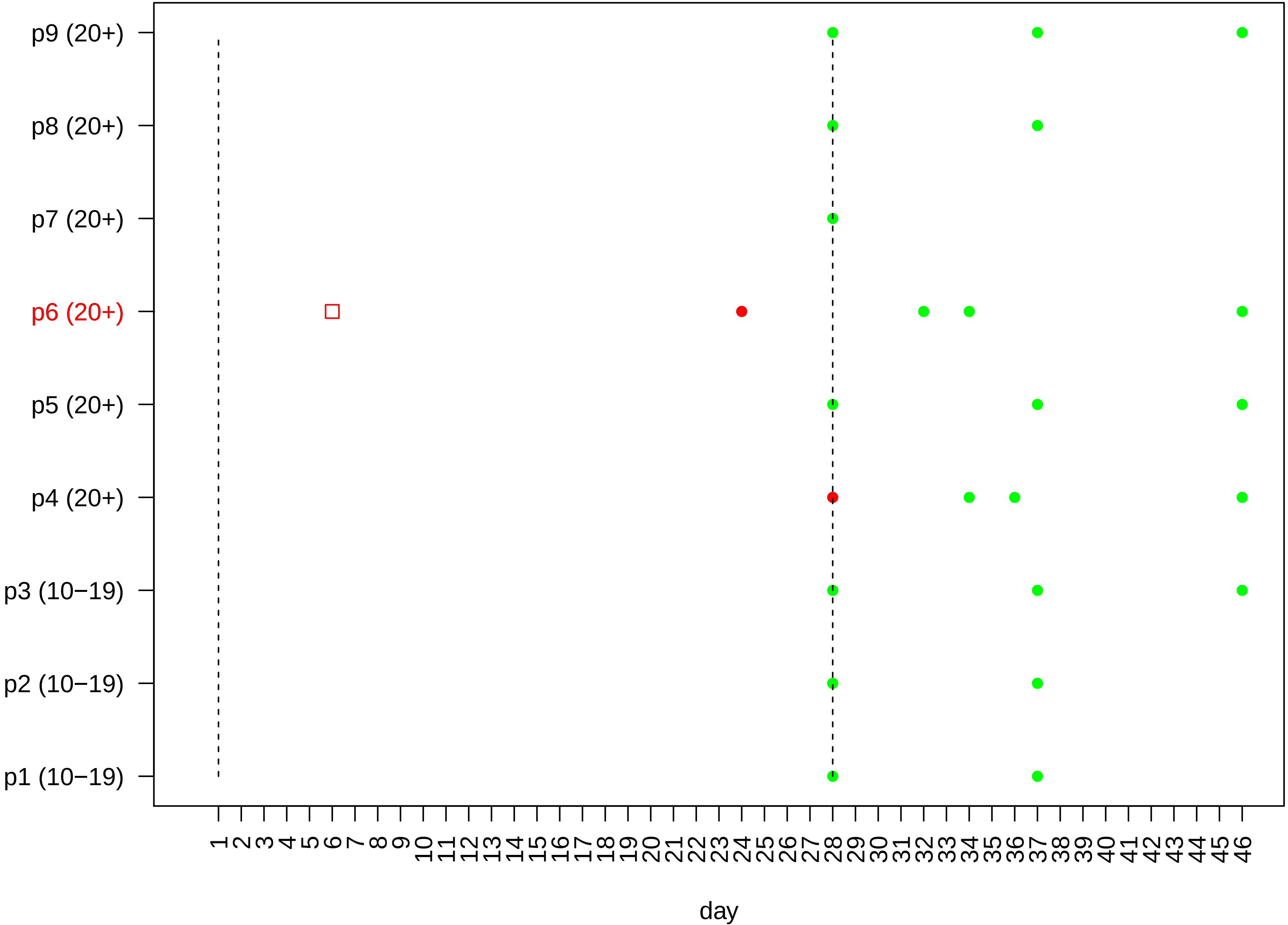

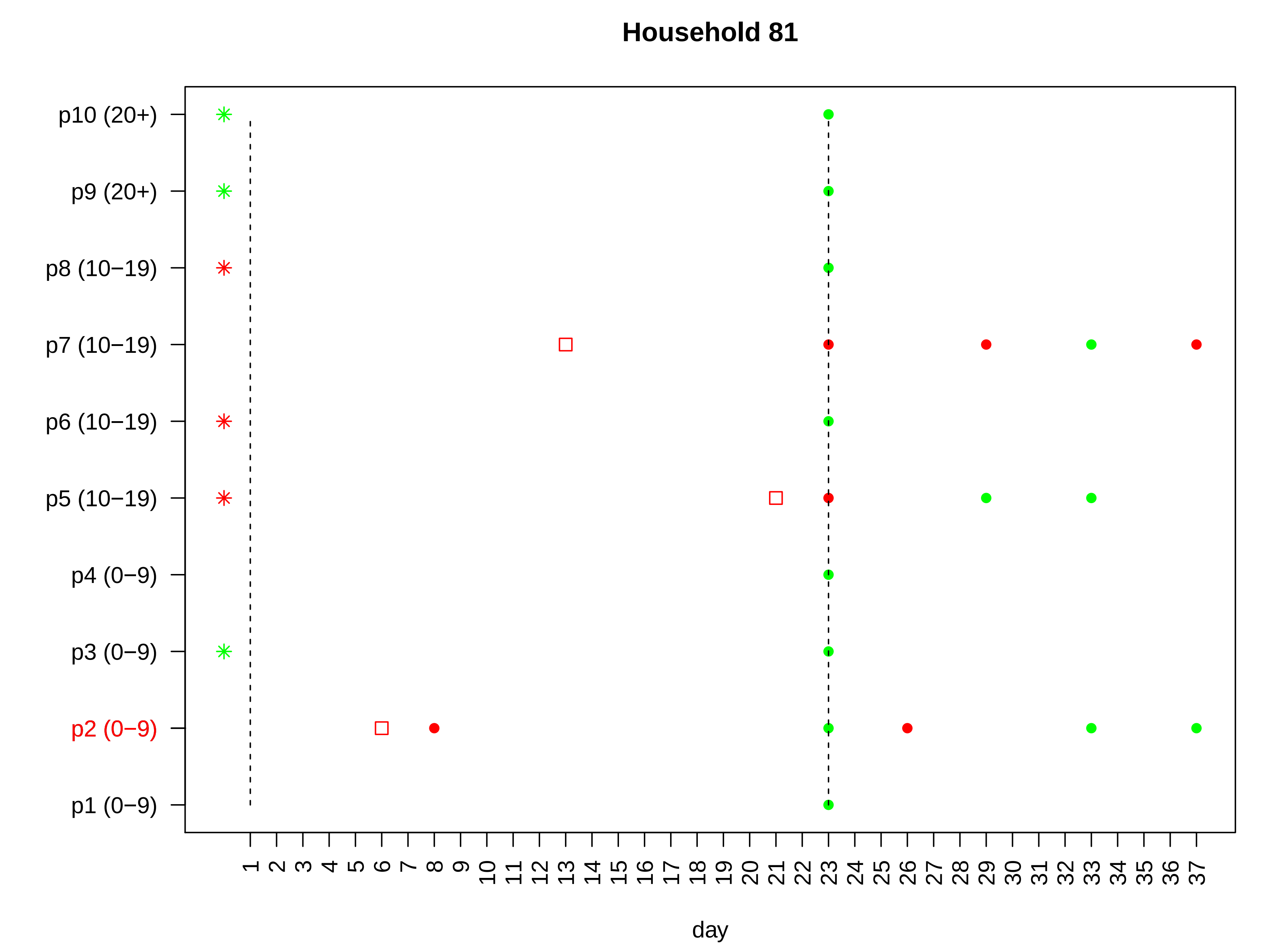
Household 81
p10 (20+)
p9 (20+)
p8 (10−19)
p7 (10−19)
p6 (10−19)
p5 (10−19)
p4 (0−9)
p3 (0−9)
p2 (0−9)
p1 (0−9)
1 2 3 4 5 6 7 8 9 10 11 12 13 14 15 16 17 18 19 20 21 22 23 24 25 26 27 28 29 30 31 32 33 34 35 36 37
day

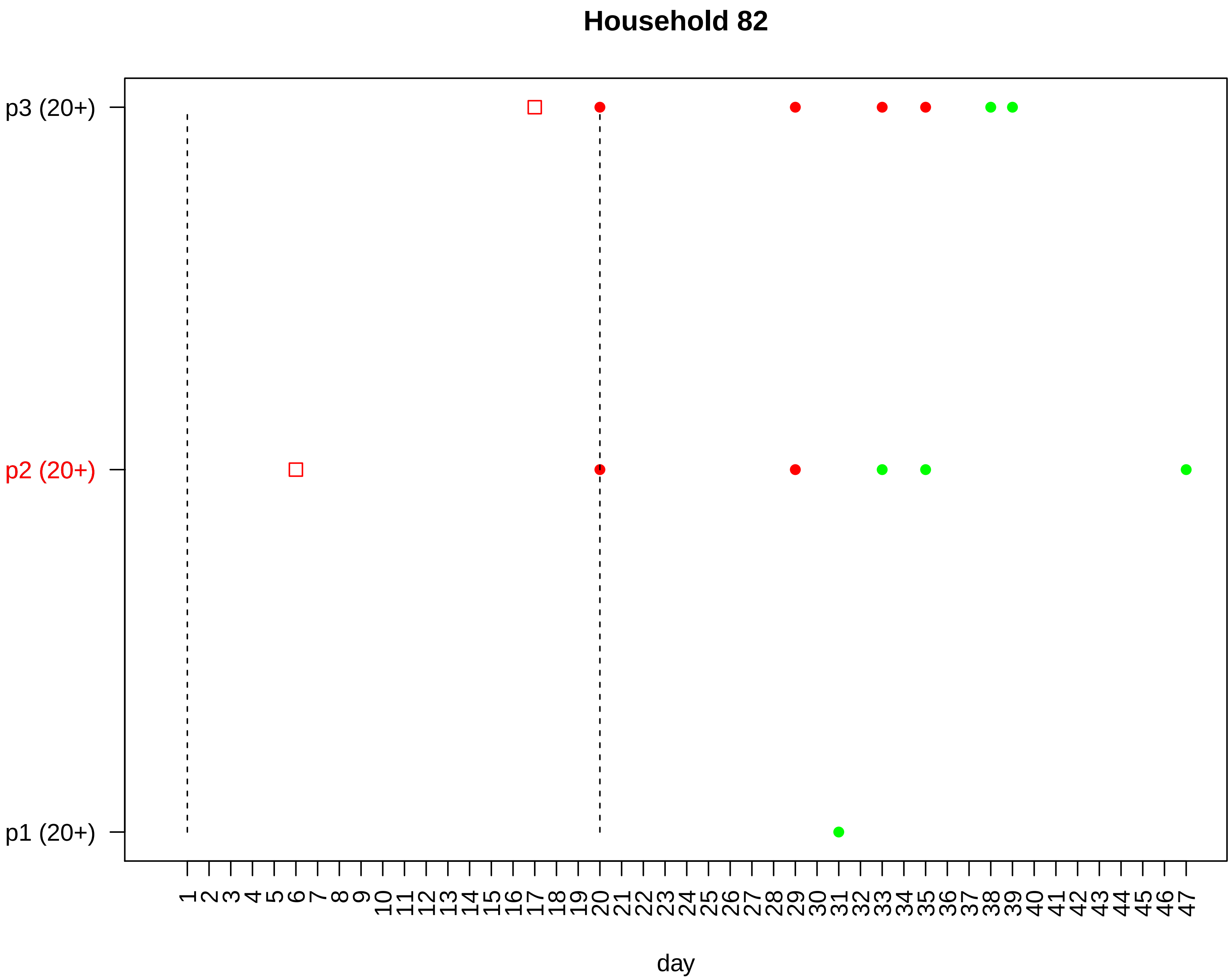

Household 82
p3 (20+)
p2 (20+)
p1 (20+)
day

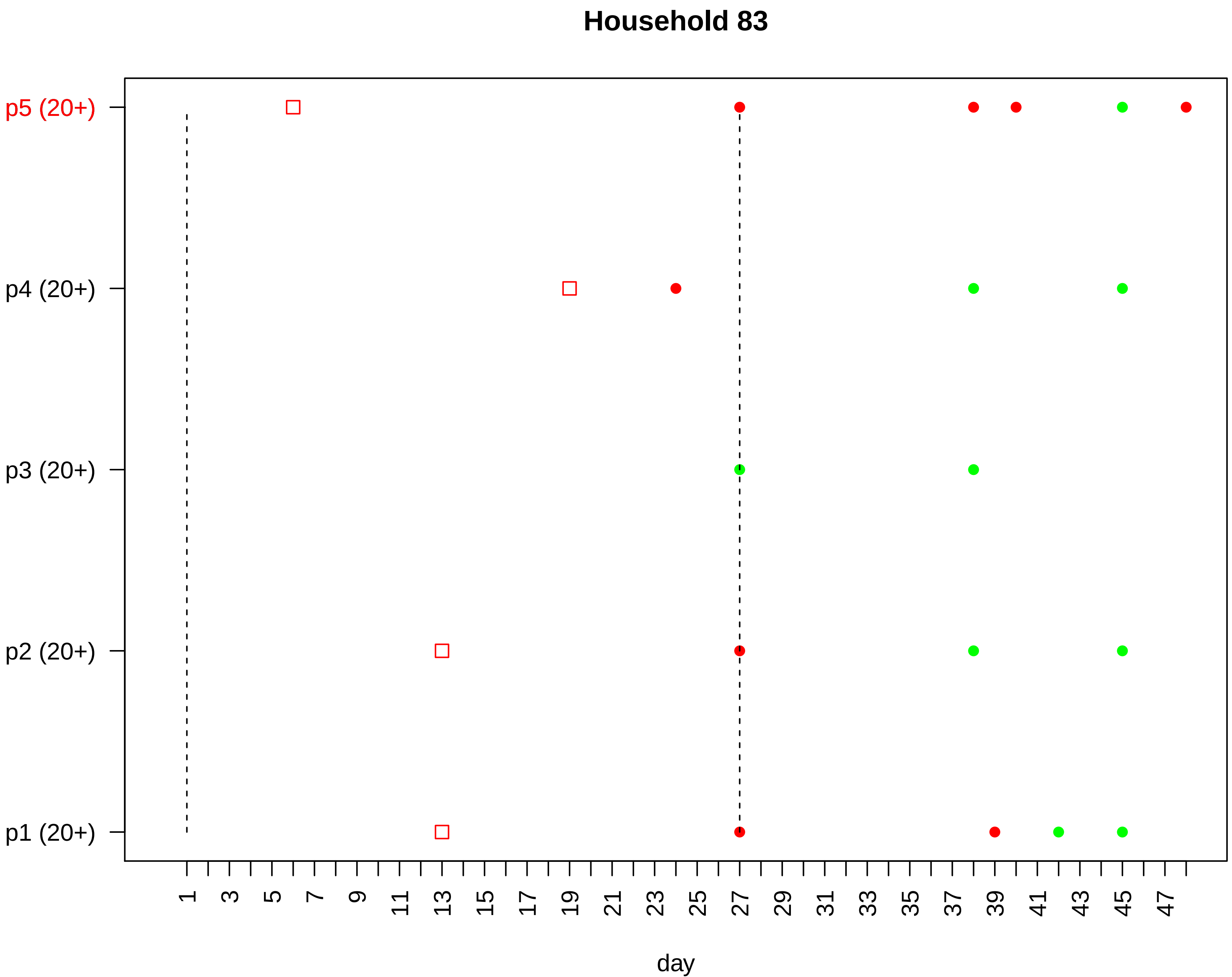

Household 83
p5 (20+)
p4 (20+)
p3 (20+)
p2 (20+)
p1 (20+)
1
3
5
7
9
11
13
15
17
19
21
23
25
27
29
31
33
35
37
39
41
43
45
47
day

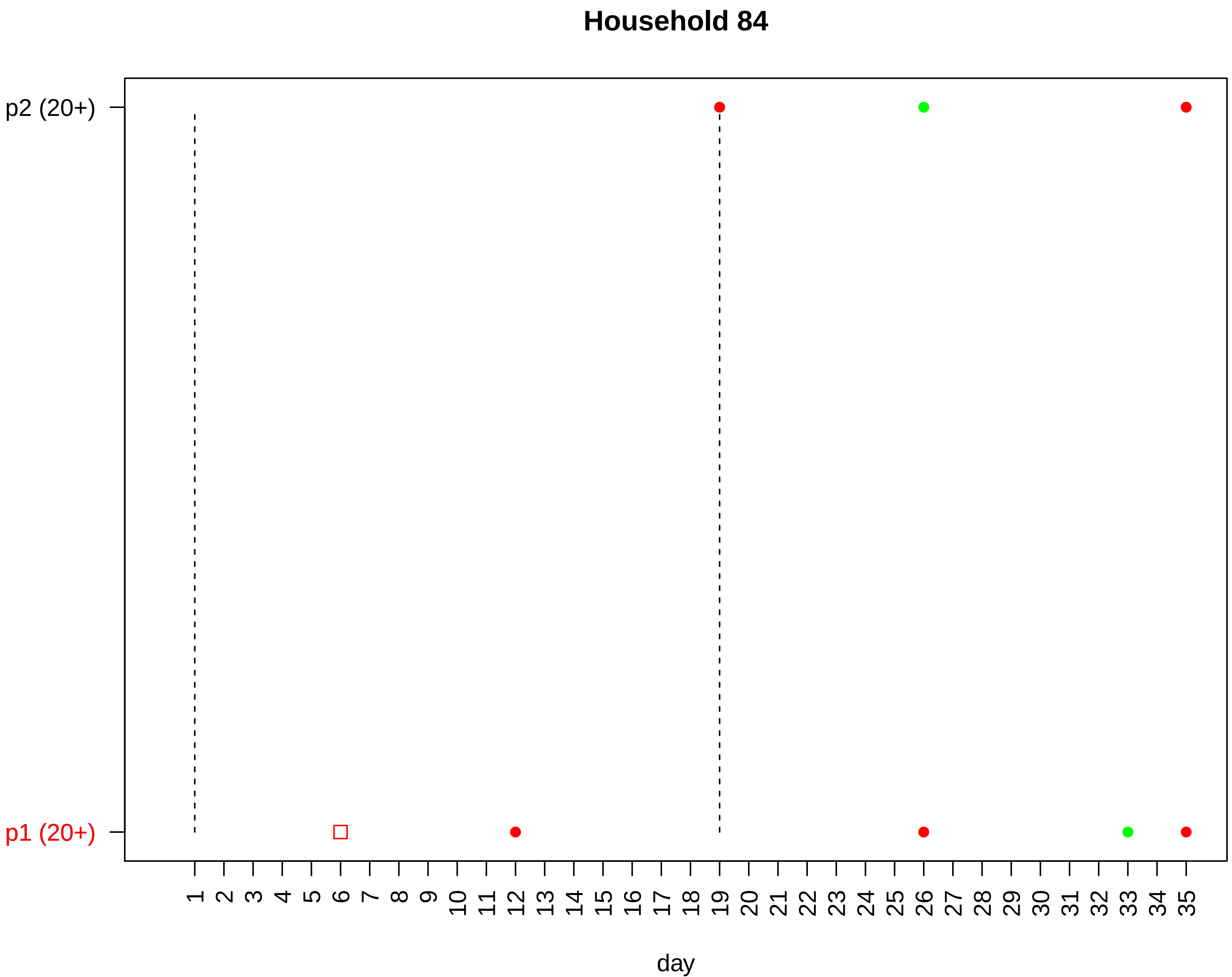
Household 84
p2 (20+)
p1 (20+)
1 2 3 4 5 6 7 8 9 10 11 12 13 14 15 16 17 18 19 20 21 22 23 24 25 26 27 28 29 30 31 32 33 34 35
day

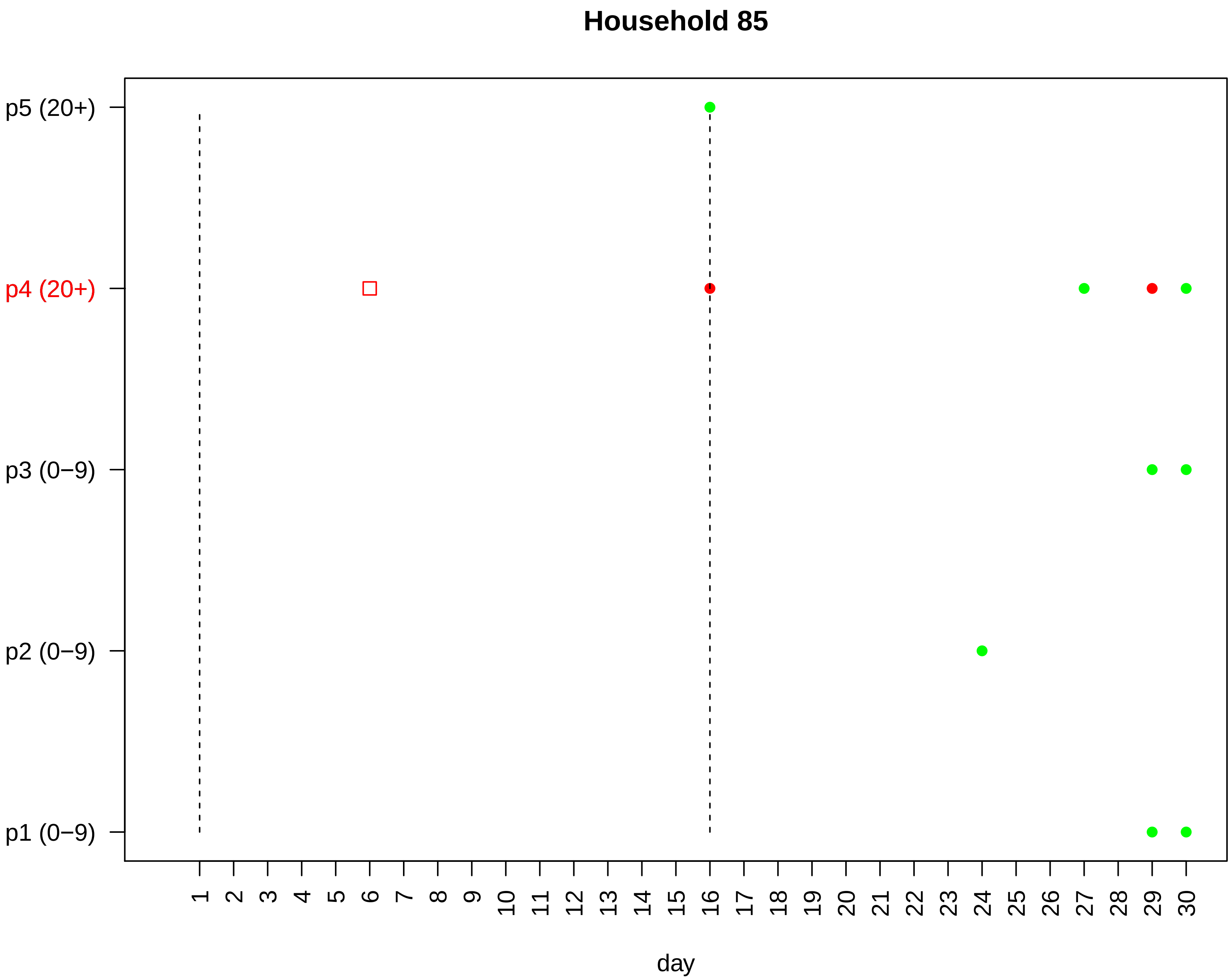

Household 85
p5 (20+)
p4 (20+)
p3 (0−9)
p2 (0−9)
p1 (0−9)
day

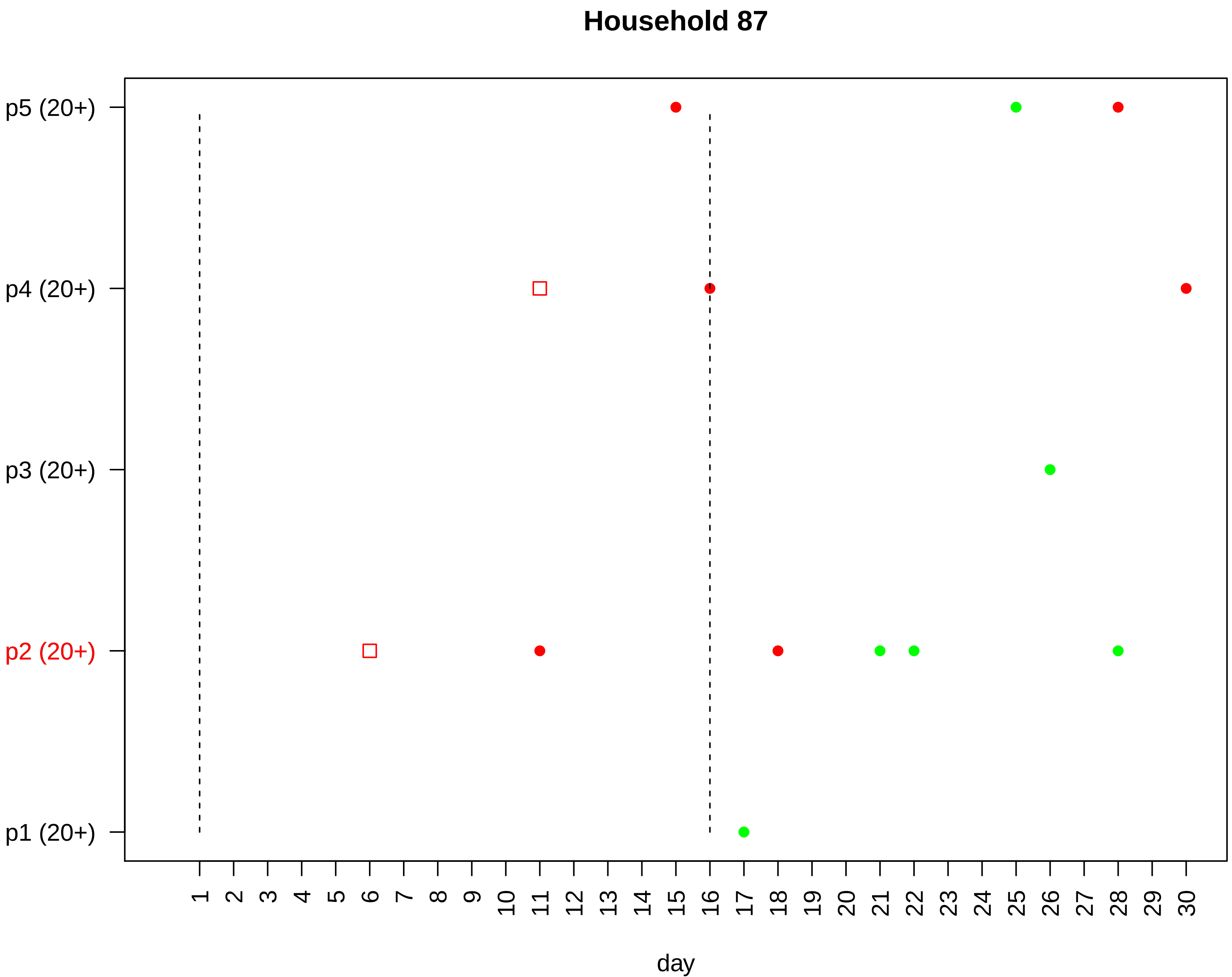

Household 87
p5 (20+)
p4 (20+)
p3 (20+)
p2 (20+)
p1 (20+)
1 2 3 4 5 6 7 8 9 10 11 12 13 14 15 16 17 18 19 20 21 22 23 24 25 26 27 28 29 30
day

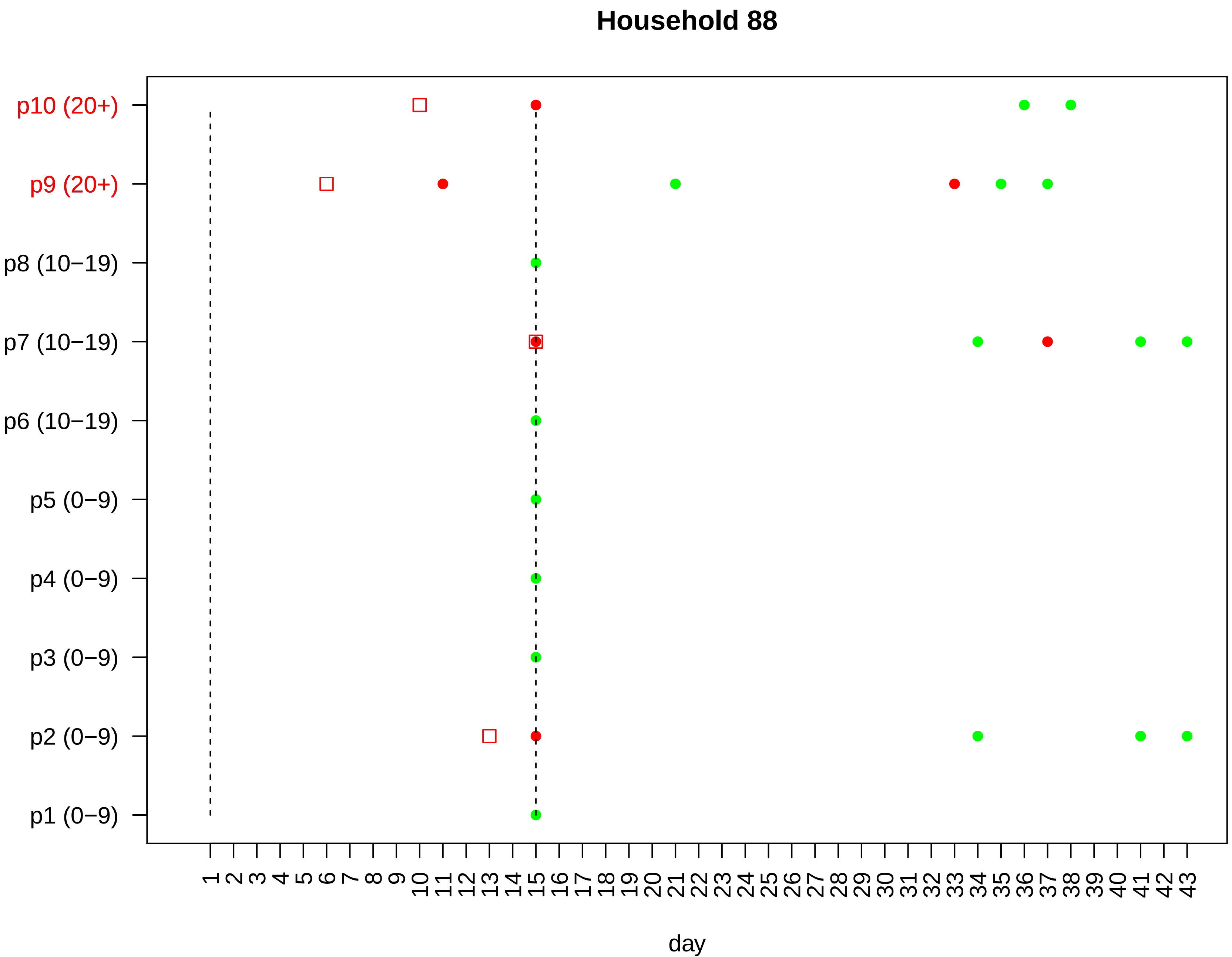

Household 88
p10 (20+)
p9 (20+)
p8 (10−19)
p7 (10−19)
p6 (10−19)
p5 (0−9)
p4 (0−9)
p3 (0−9)
p2 (0−9)
p1 (0−9)
1 2 3 4 5 6 7 8 9 10 11 12 13 14 15 16 17 18 19 20 21 22 23 24 25 26 27 28 29 30 31 32 33 34 35 36 37 38 39 40 41 42 43
day

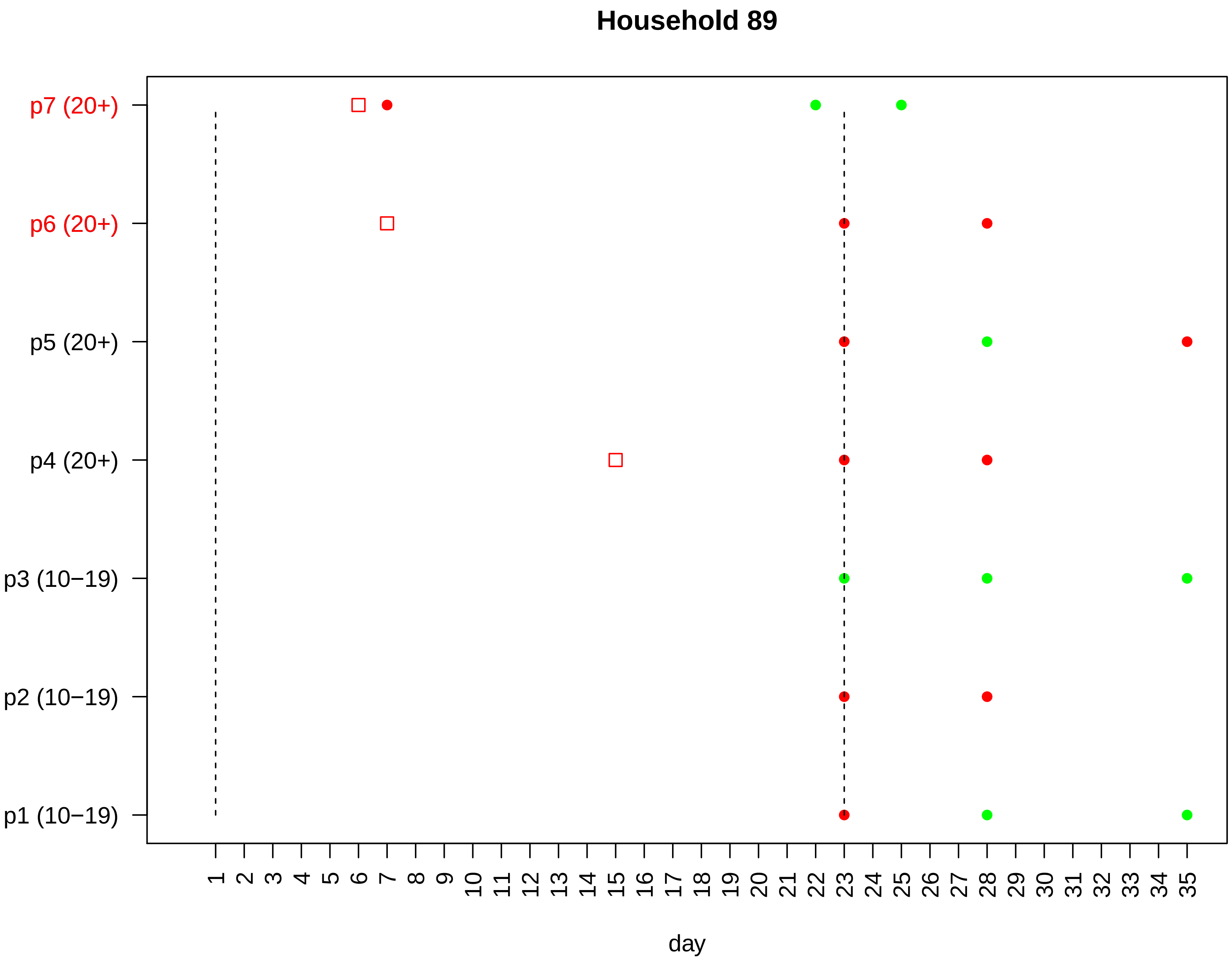
Household 89
p7 (20+)
p6 (20+)
p5 (20+)
p4 (20+)
p3 (10−19)
p2 (10−19)
p1 (10−19)
1 2 3 4 5 6 7 8 9 10 11 12 13 14 15 16 17 18 19 20 21 22 23 24 25 26 27 28 29 30 31 32 33 34 35
day

### Household 90

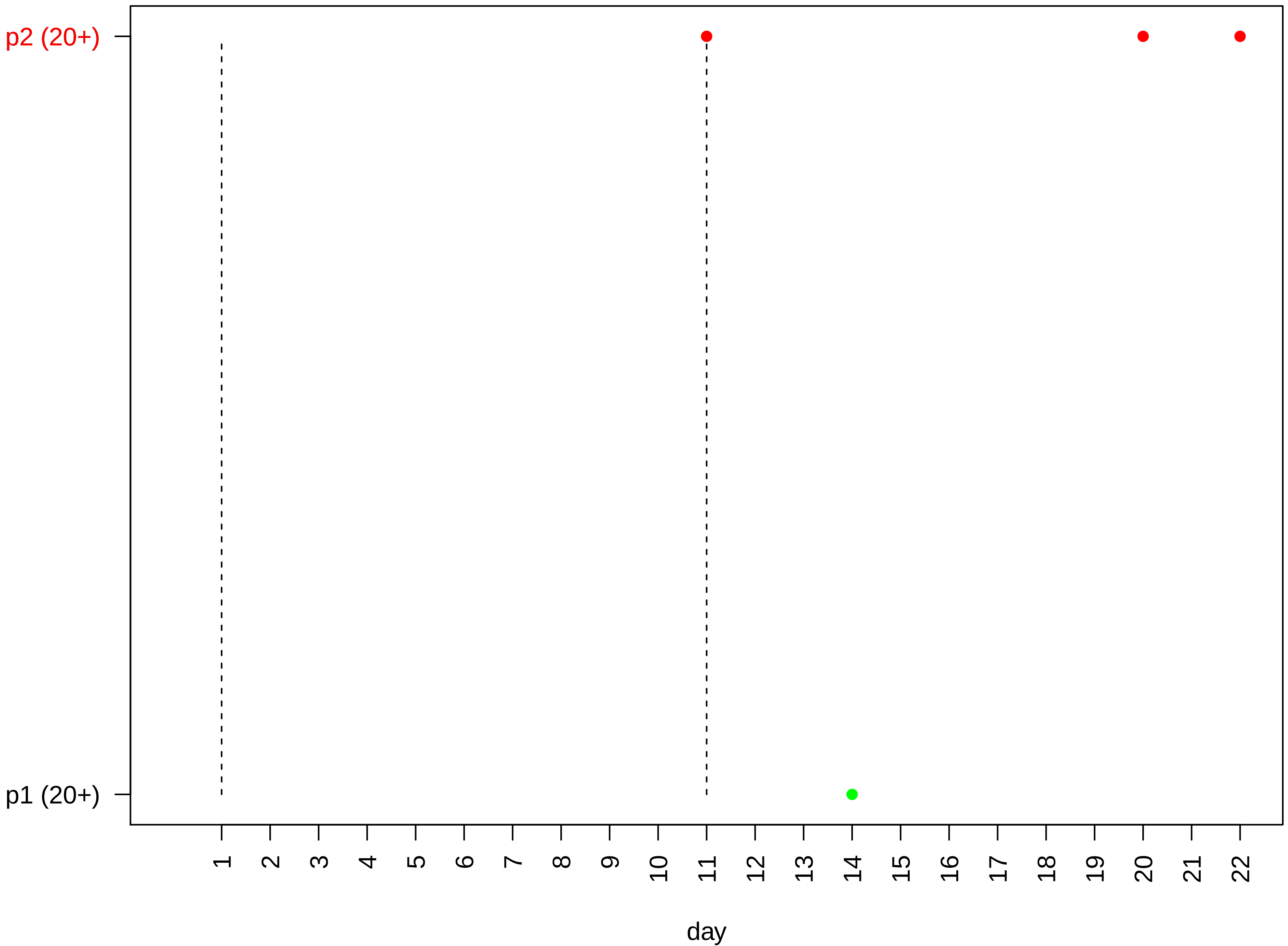

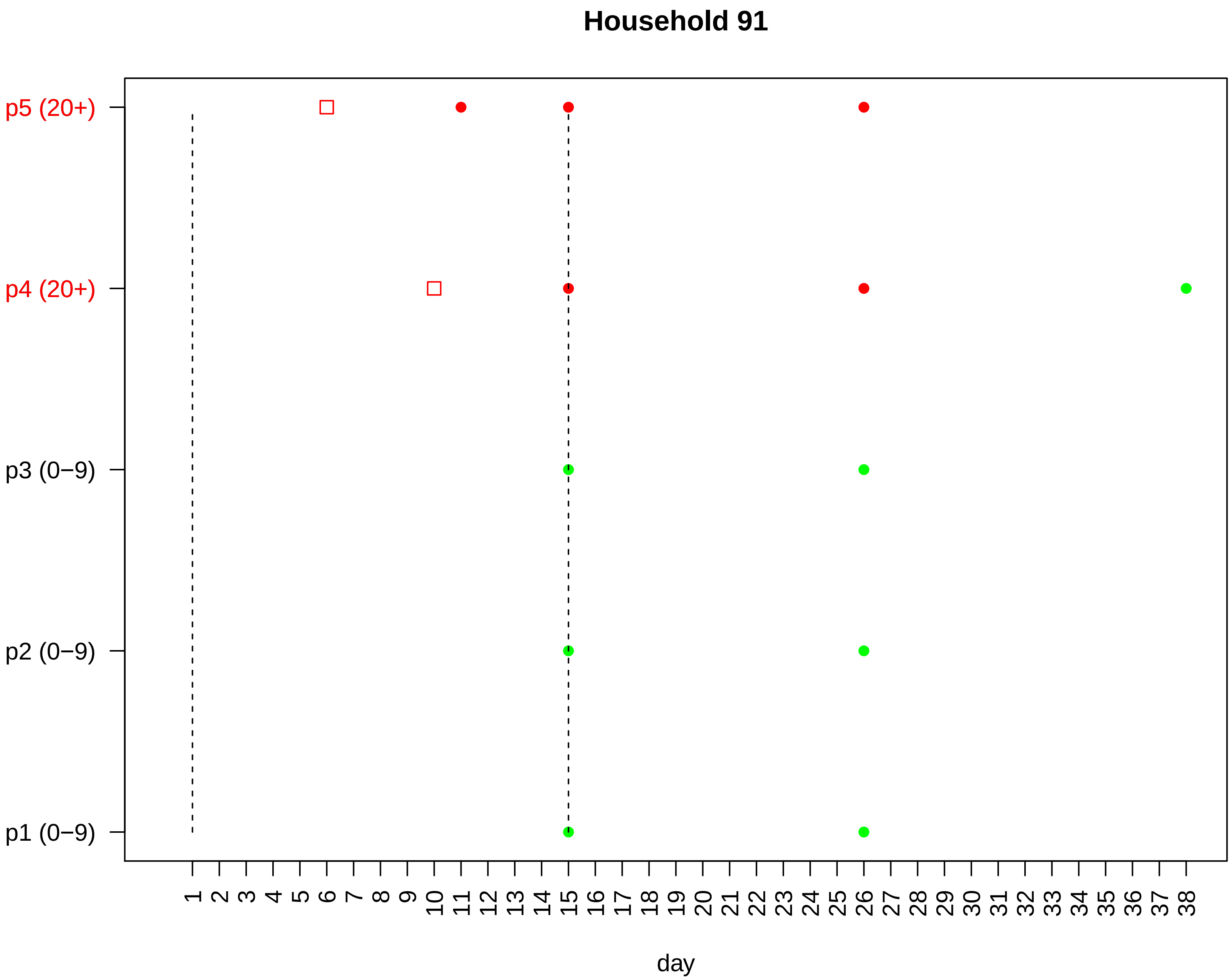

Household 91
p5 (20+)
p4 (20+)
p3 (0−9)
p2 (0−9)
p1 (0−9)
1 2 3 4 5 6 7 8 9 10 11 12 13 14 15 16 17 18 19 20 21 22 23 24 25 26 27 28 29 30 31 32 33 34 35 36 37 38
day

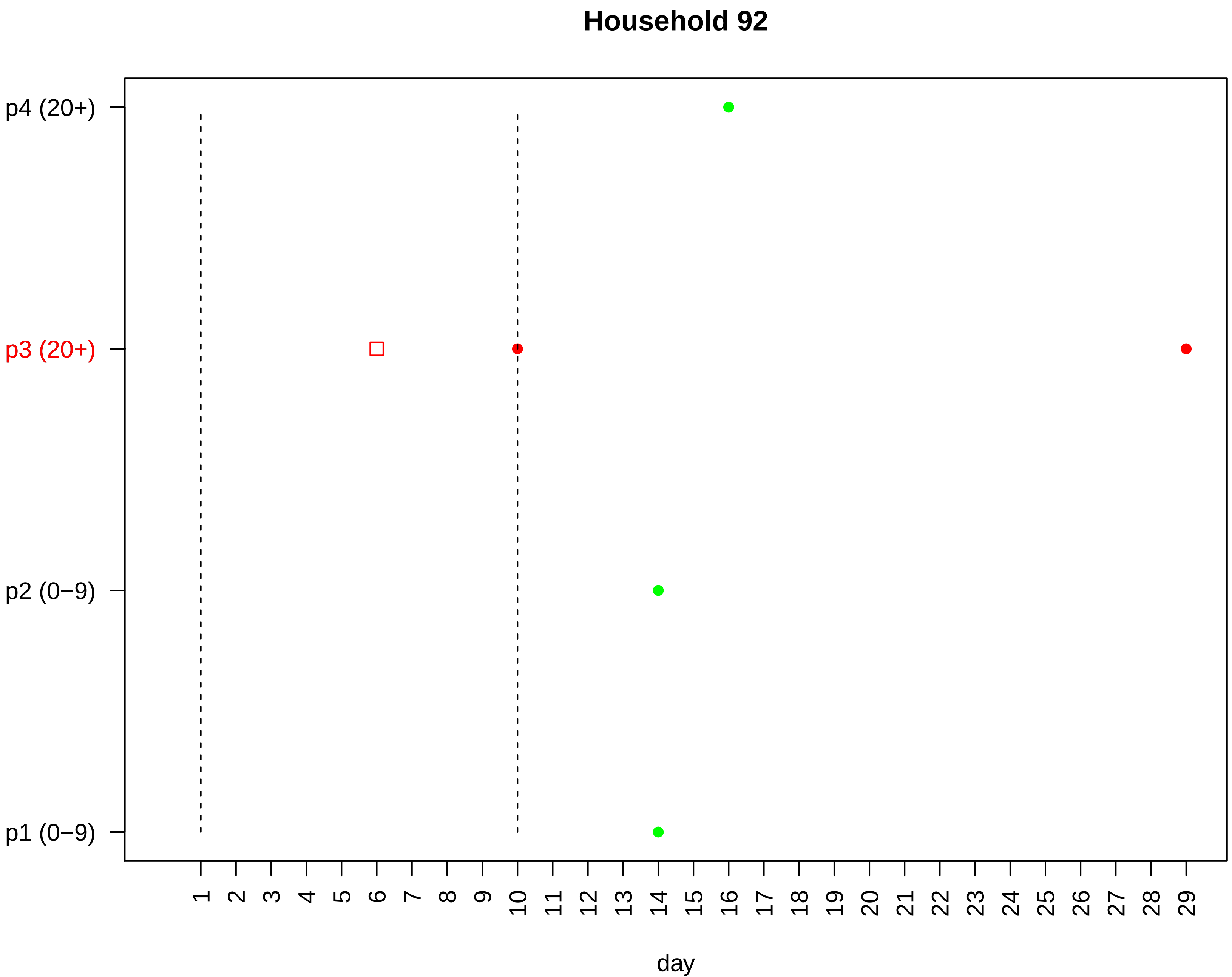
Household 92
p4 (20+)
p3 (20+)
p2 (0−9)
p1 (0−9)
1 2 3 4 5 6 7 8 9 10 11 12 13 14 15 16 17 18 19 20 21 22 23 24 25 26 27 28 29
day

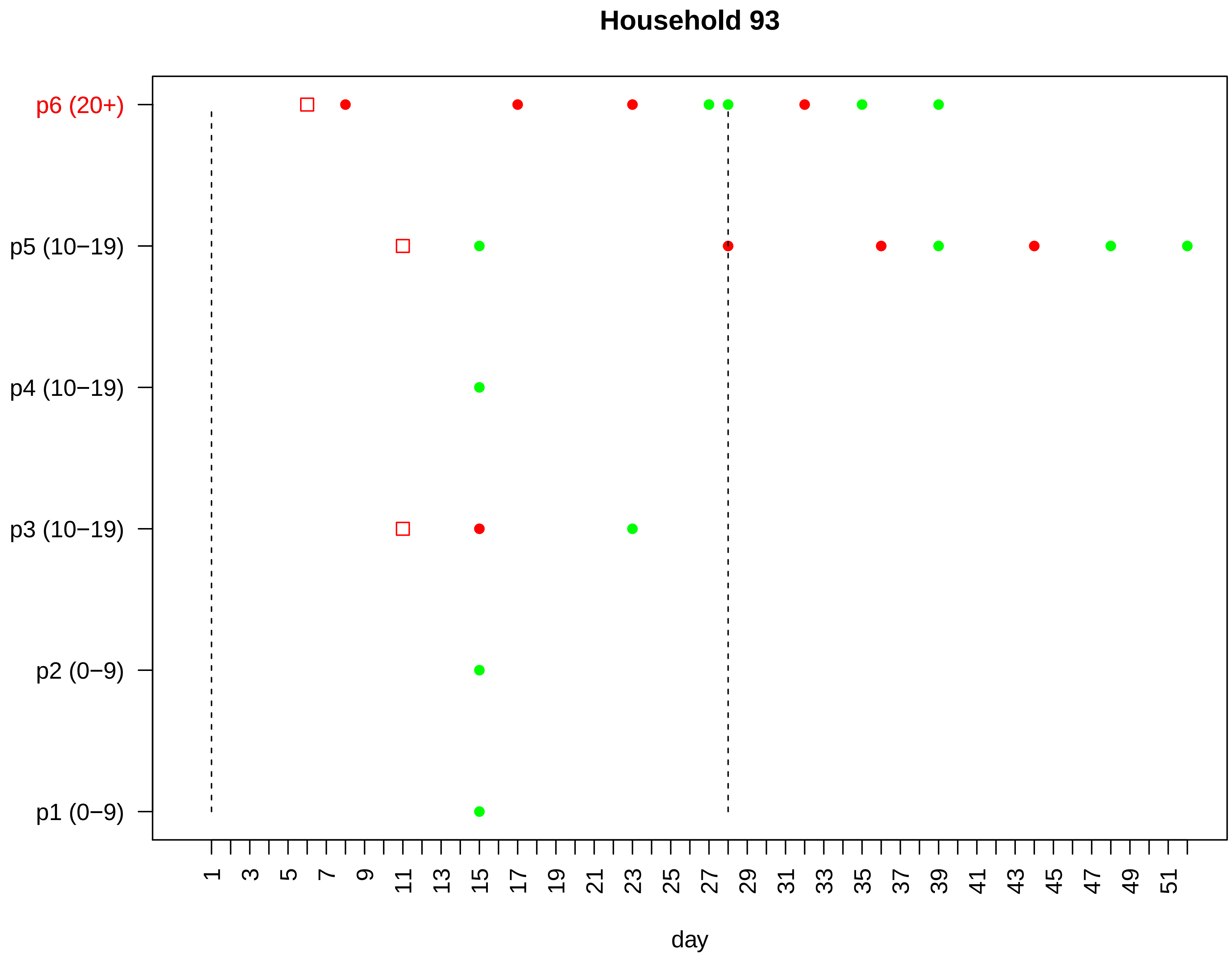

Household 93
p6 (20+)
p5 (10−19)
p4 (10−19)
p3 (10−19)
p2 (0−9)
p1 (0−9)
1
3
5
7
9
11
13
15
17
19
21
23
25
27
29
31
33
35
37
39
41
43
45
47
49
51
day

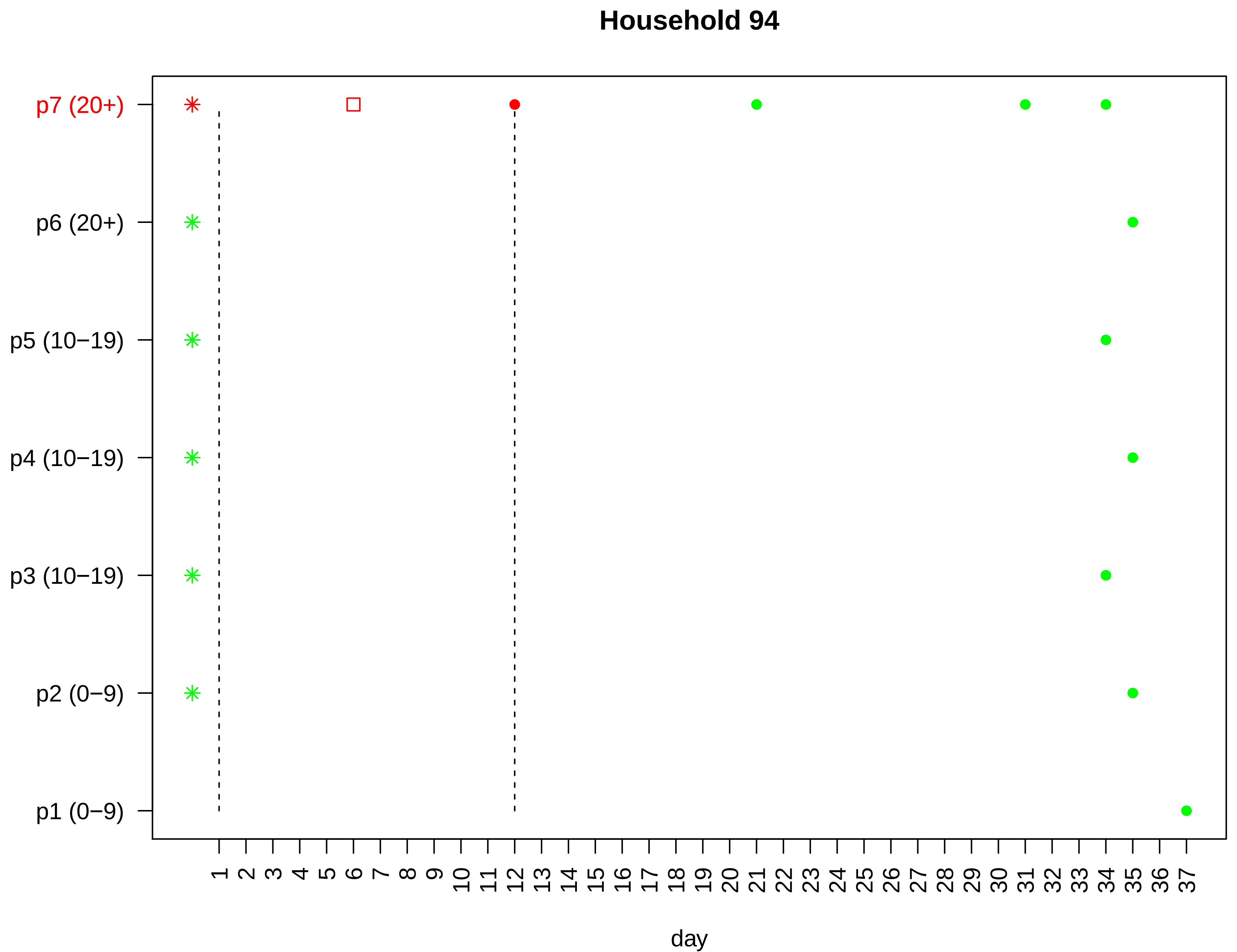

Household 94
p7 (20+)
p6 (20+)
p5 (10−19)
p4 (10−19)
p3 (10−19)
p2 (0−9)
p1 (0−9)
1 2 3 4 5 6 7 8 9 10 11 12 13 14 15 16 17 18 19 20 21 22 23 24 25 26 27 28 29 30 31 32 33 34 35 36 37
day

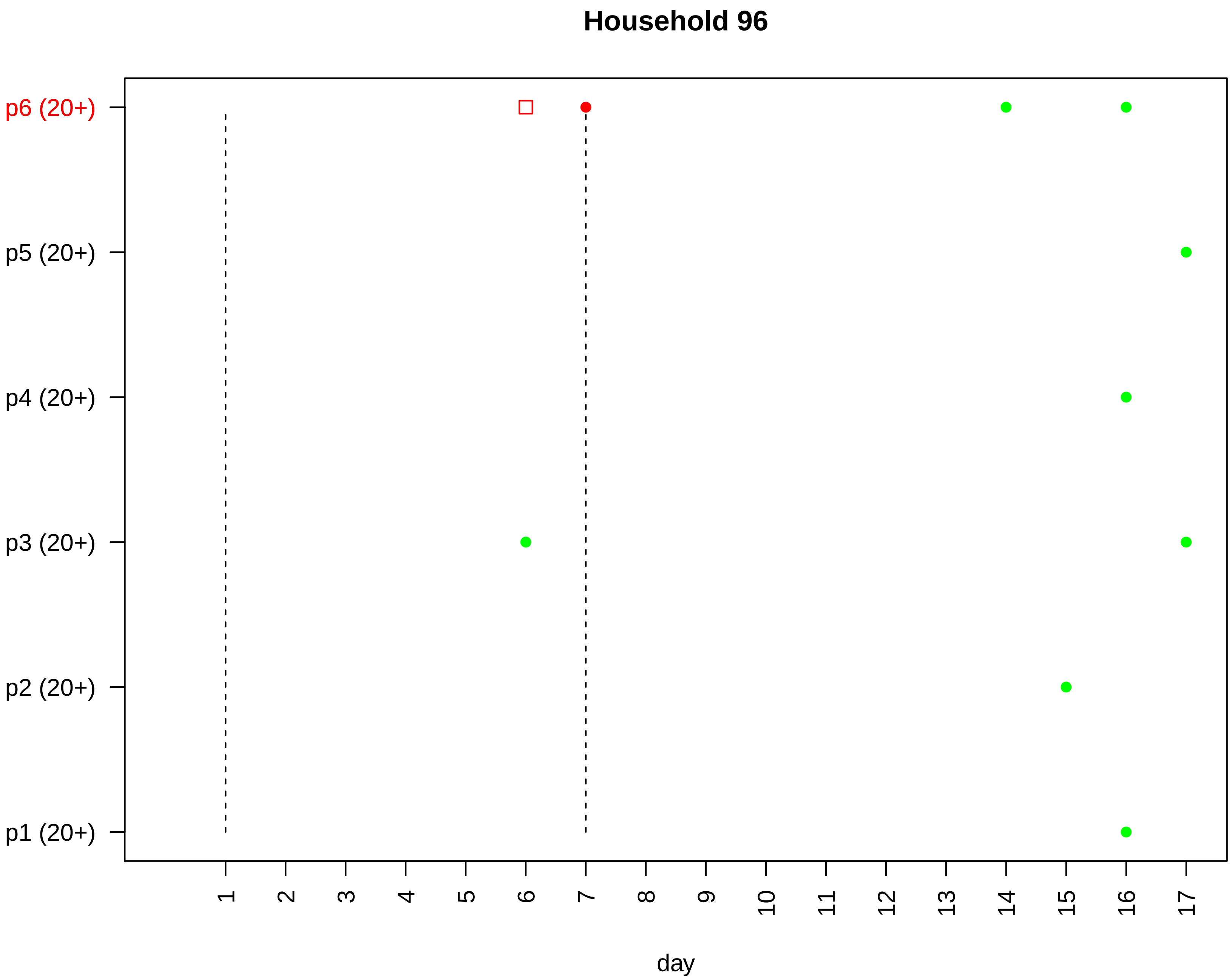

Household 96
p6 (20+)
p5 (20+)
p4 (20+)
p3 (20+)
p2 (20+)
p1 (20+)
day

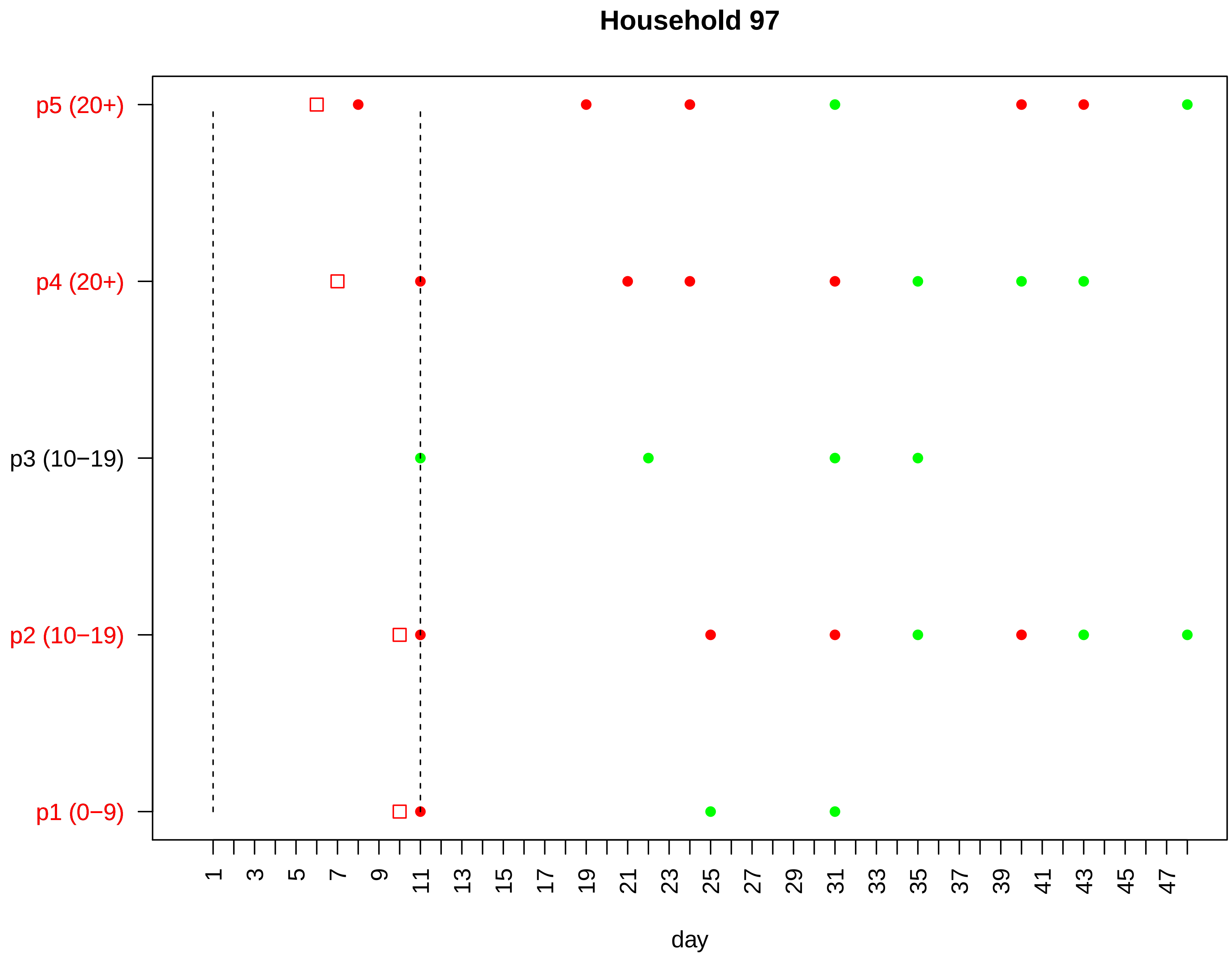

Household 97
p5 (20+)
p4 (20+)
p3 (10−19)
p2 (10−19)
p1 (0−9)
1
3
5
7
9
11
13
15
17
19
21
23
25
27
29
31
33
35
37
39
41
43
45
47
day

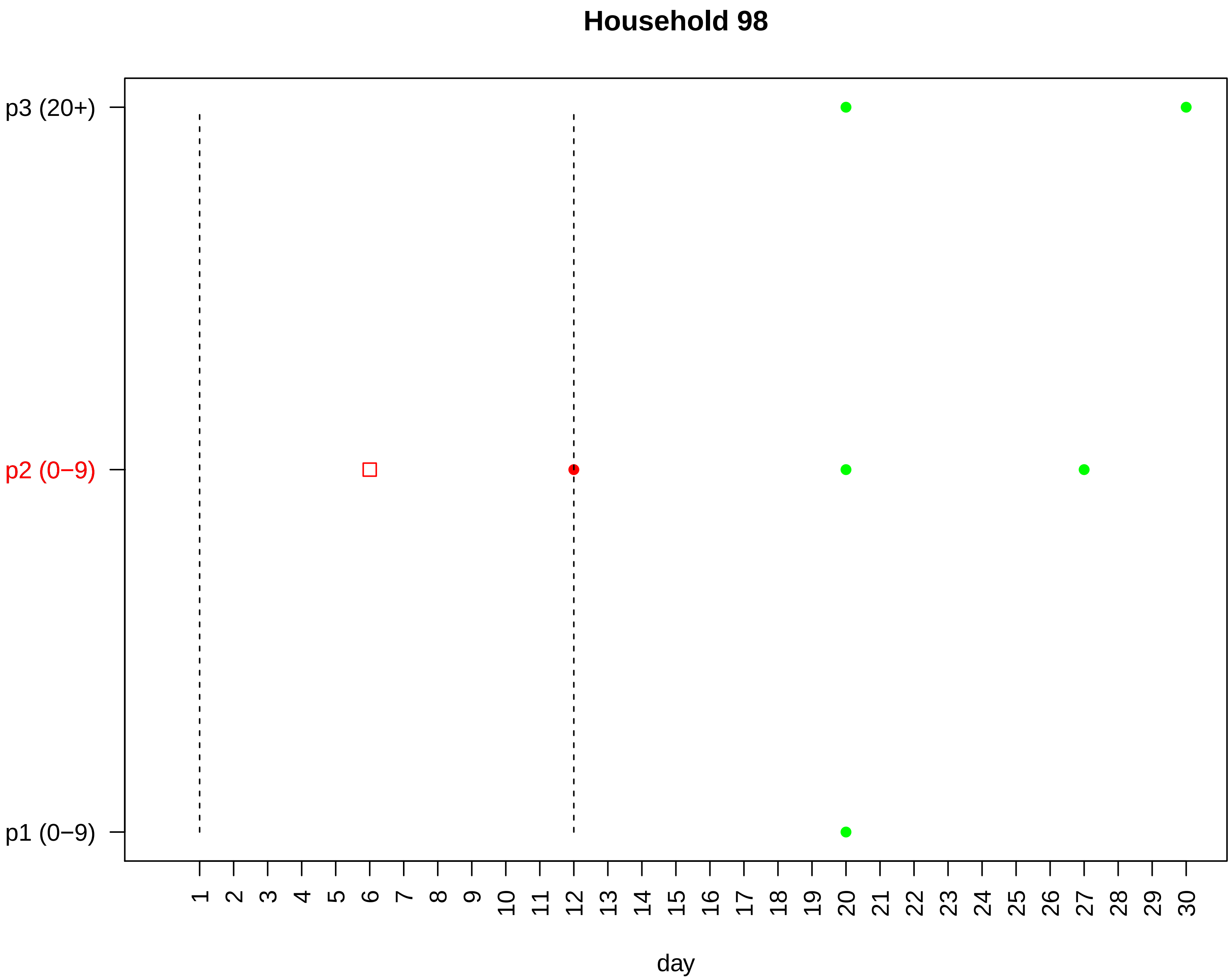

Household 98
p3 (20+)
p2 (0−9)
p1 (0−9)
1 2 3 4 5 6 7 8 9 10 11 12 13 14 15 16 17 18 19 20 21 22 23 24 25 26 27 28 29 30
day

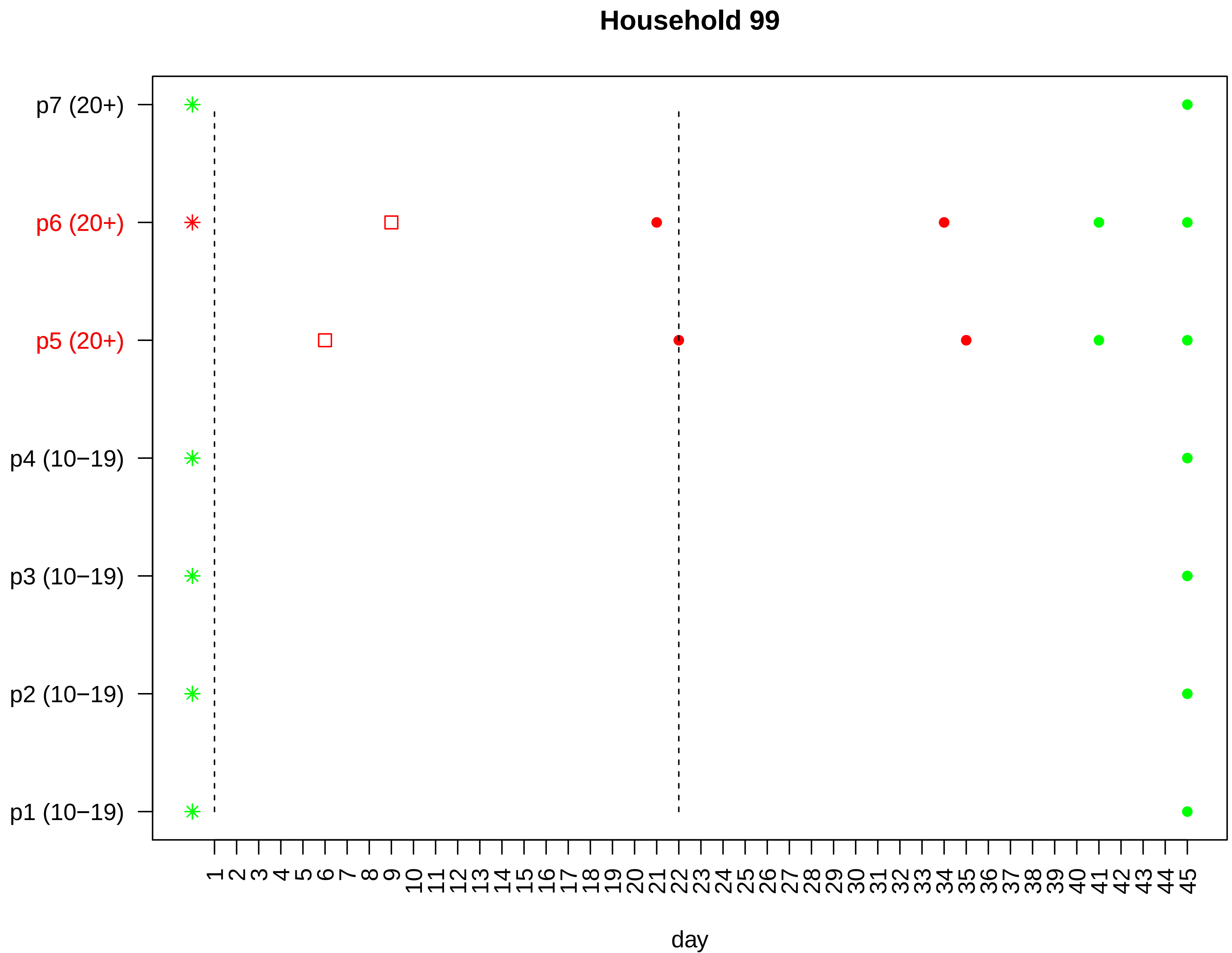

Household 99
p7 (20+)
p6 (20+)
p5 (20+)
p4 (10−19)
p3 (10−19)
p2 (10−19)
p1 (10−19)
1 2 3 4 5 6 7 8 9 10 11 12 13 14 15 16 17 18 19 20 21 22 23 24 25 26 27 28 29 30 31 32 33 34 35 36 37 38 39 40 41 42 43 44 45
day

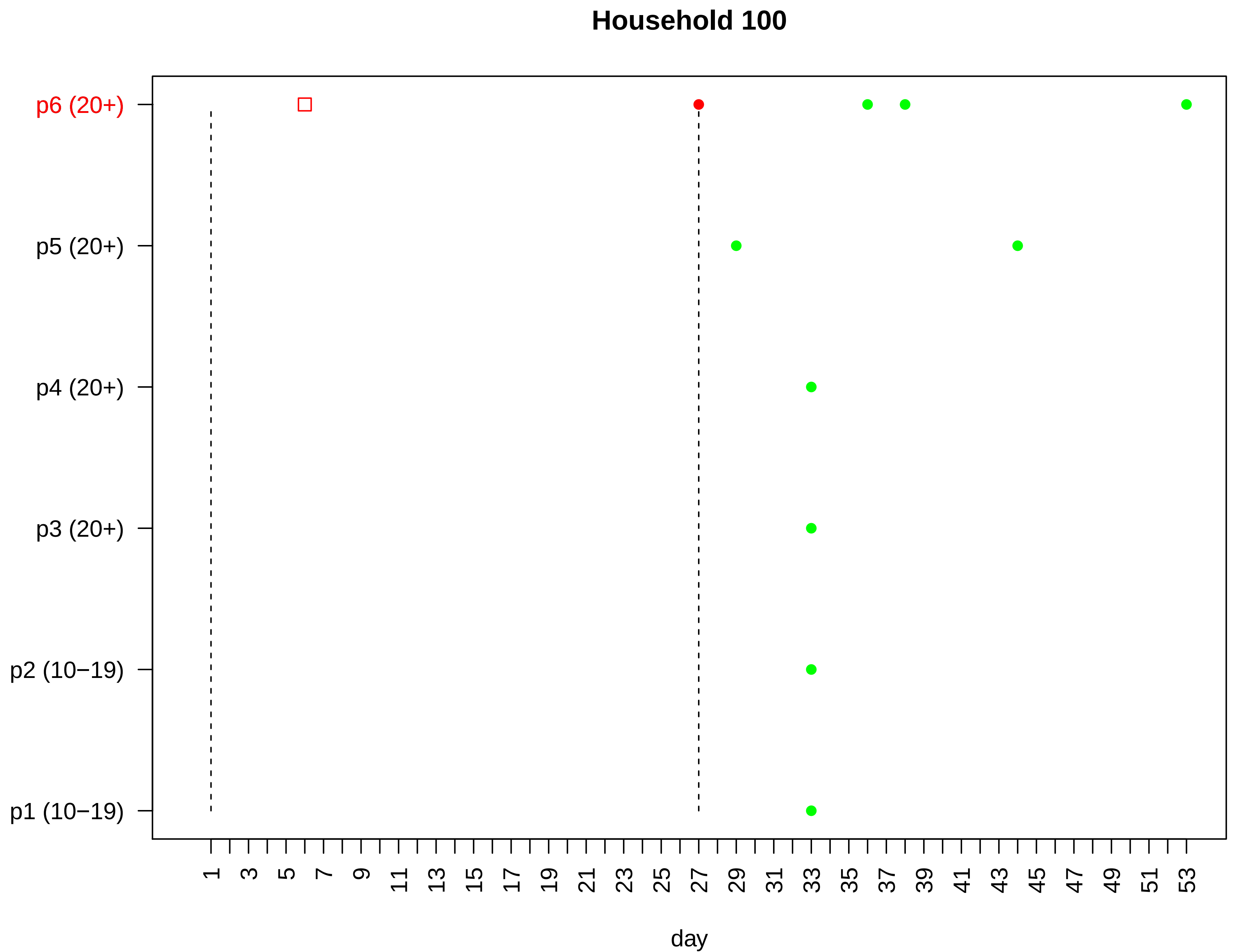

Household 100
p6 (20+)
p5 (20+)
p4 (20+)
p3 (20+)
p2 (10−19)
p1 (10−19)
1 3 5 7 9 11 13 15 17 19 21 23 25 27 29 31 33 35 37 39 41 43 45 47 49 51 53
day

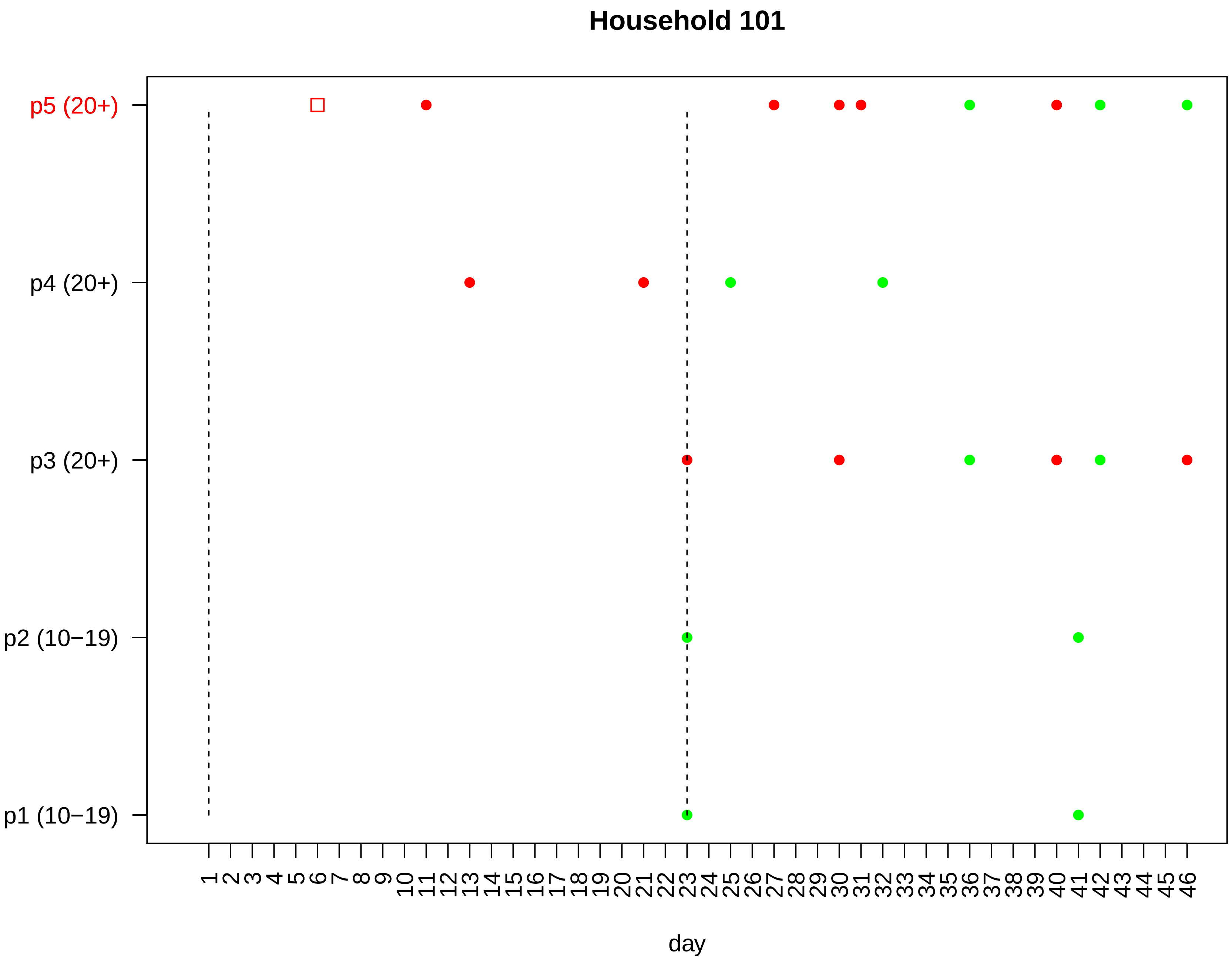
Household 101
p5 (20+)
p4 (20+)
p3 (20+)
p2 (10−19)
p1 (10−19)
1 2 3 4 5 6 7 8 9 10 11 12 13 14 15 16 17 18 19 20 21 22 23 24 25 26 27 28 29 30 31 32 33 34 35 36 37 38 39 40 41 42 43 44 45 46
day

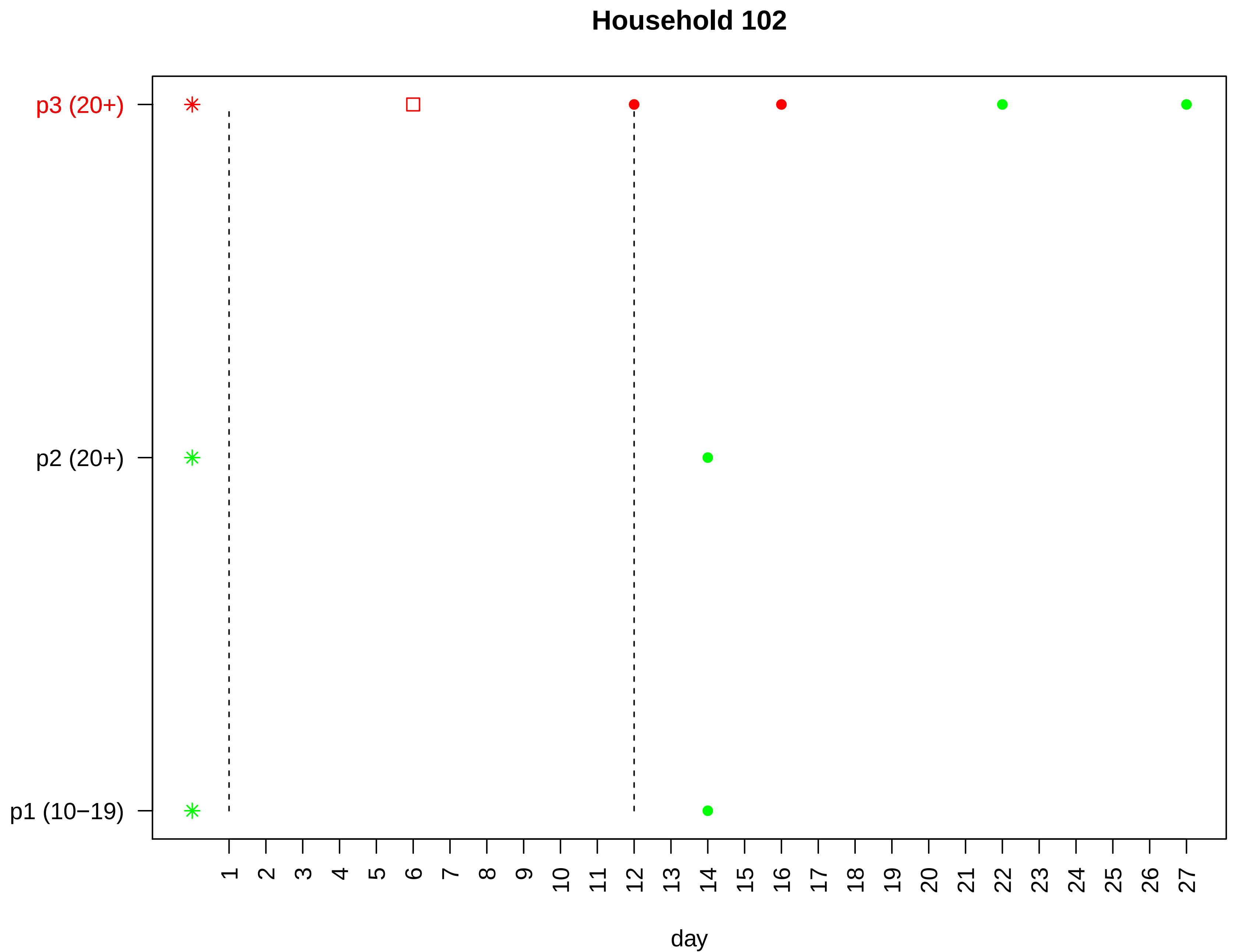

Household 102
p3 (20+)
p2 (20+)
p1 (10−19)
day

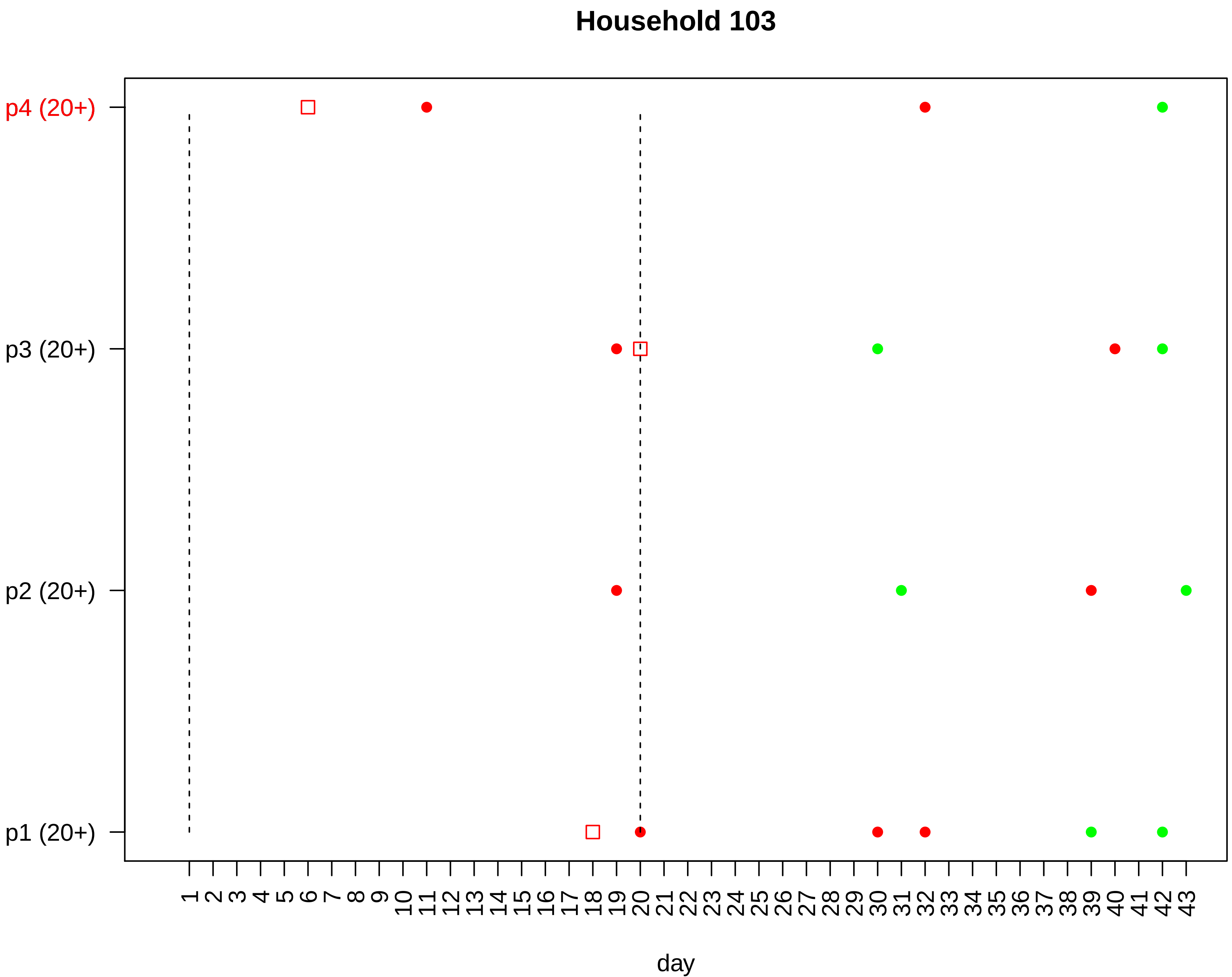

Household 103
p4 (20+)
p3 (20+)
p2 (20+)
p1 (20+)
1 2 3 4 5 6 7 8 9 10 11 12 13 14 15 16 17 18 19 20 21 22 23 24 25 26 27 28 29 30 31 32 33 34 35 36 37 38 39 40 41 42 43
day

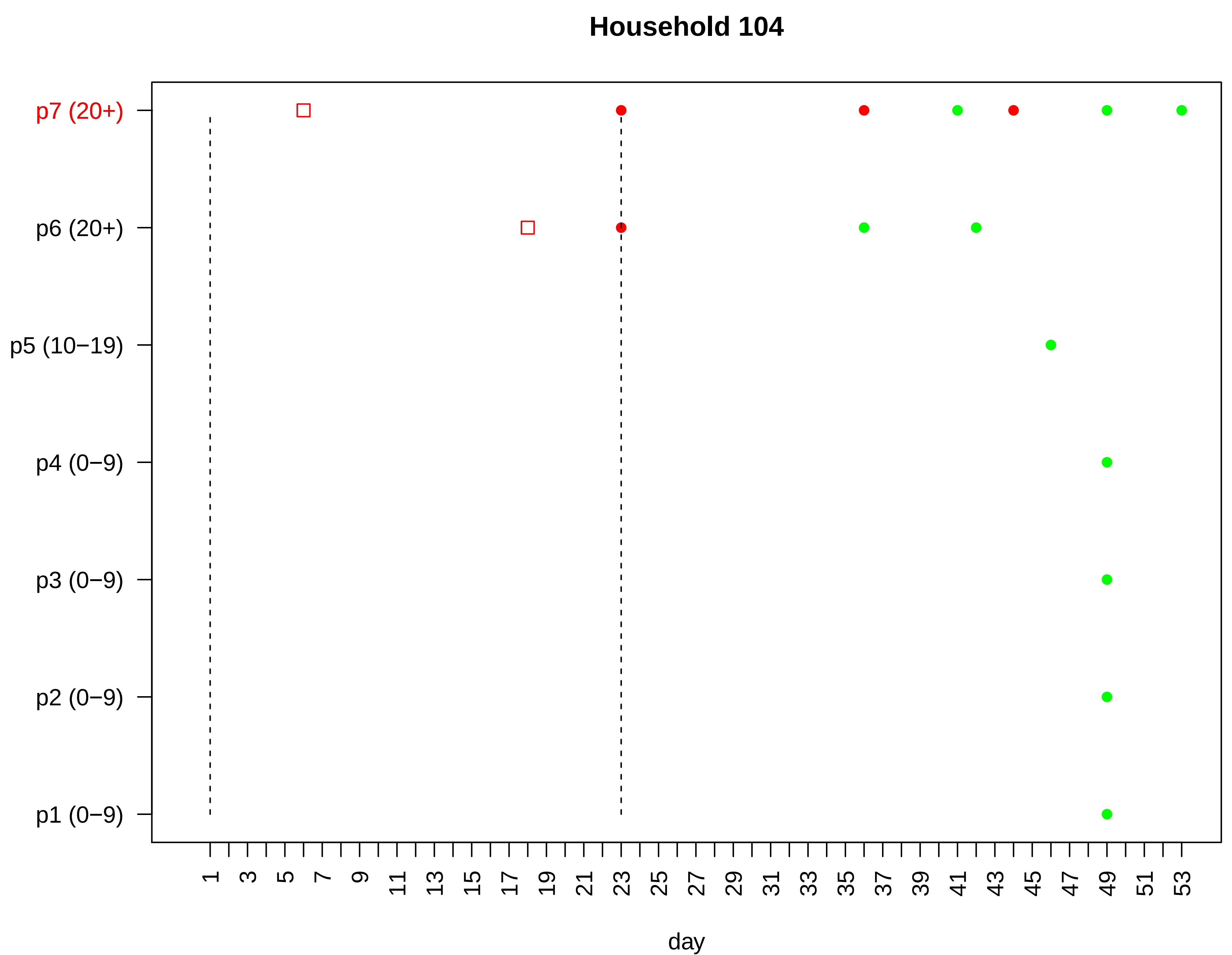

Household 104
p7 (20+)
p6 (20+)
p5 (10−19)
p4 (0−9)
p3 (0−9)
p2 (0−9)
p1 (0−9)
1
3
5
7
9
11
13
15
17
19
21
23
25
27
29
31
33
35
37
39
41
43
45
47
49
51
53
day

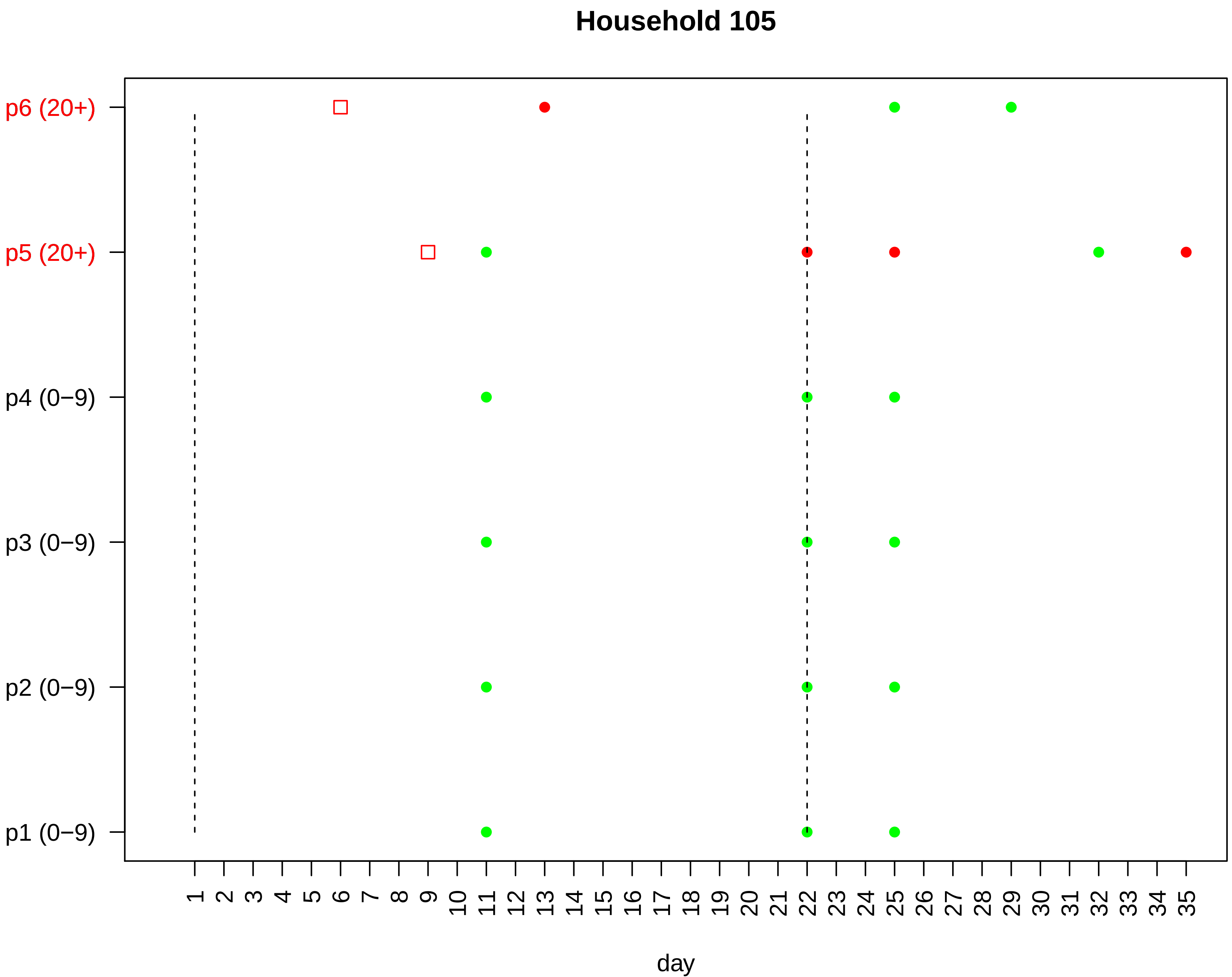

Household 105
p6 (20+)
p5 (20+)
p4 (0−9)
p3 (0−9)
p2 (0−9)
p1 (0−9)
1 2 3 4 5 6 7 8 9 10 11 12 13 14 15 16 17 18 19 20 21 22 23 24 25 26 27 28 29 30 31 32 33 34 35
day

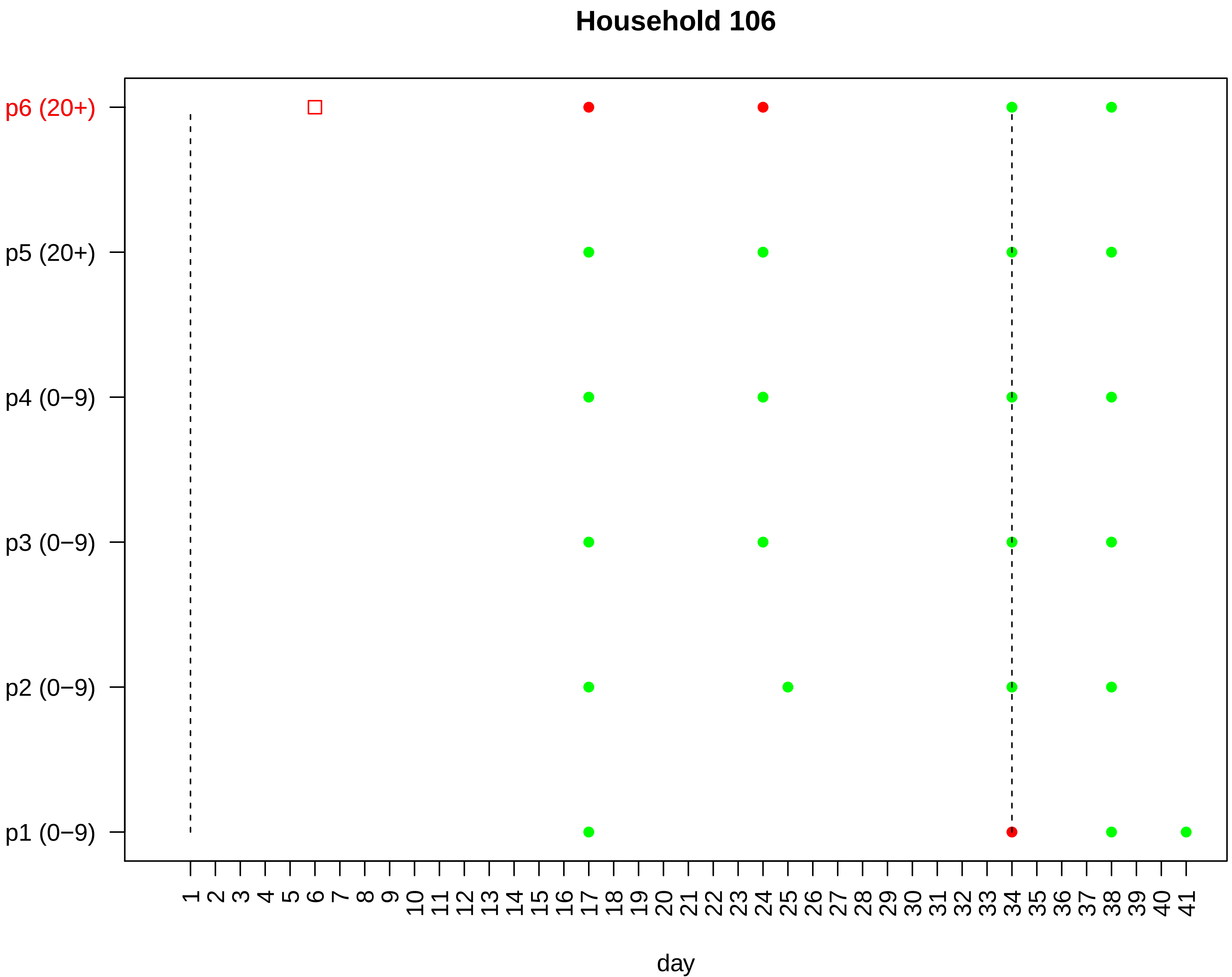

Household 106
p6 (20+)
p5 (20+)
p4 (0−9)
p3 (0−9)
p2 (0−9)
p1 (0−9)
1 2 3 4 5 6 7 8 9 10 11 12 13 14 15 16 17 18 19 20 21 22 23 24 25 26 27 28 29 30 31 32 33 34 35 36 37 38 39 40 41
day

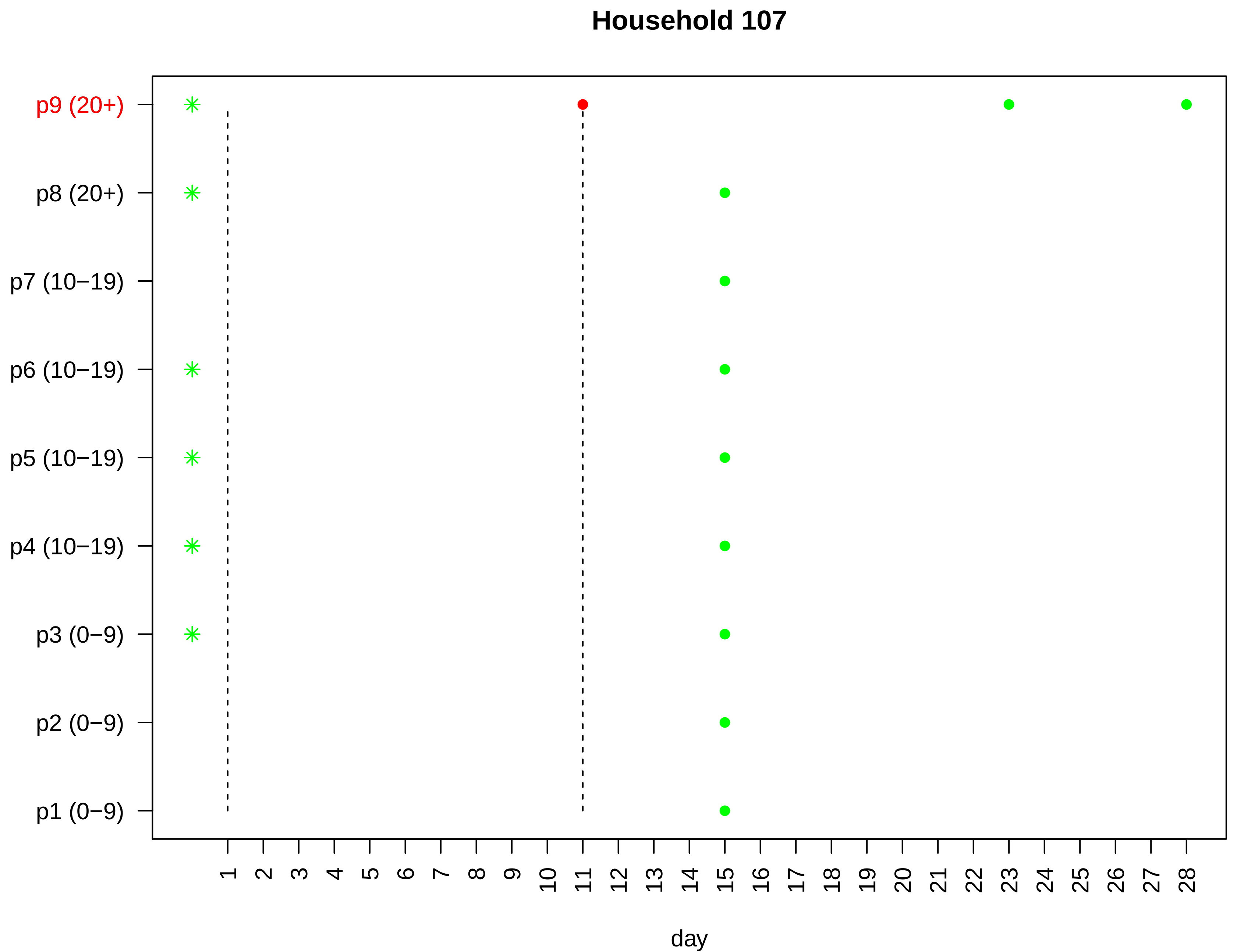
Household 107
p9 (20+)
p8 (20+)
p7 (10−19)
p6 (10−19)
p5 (10−19)
p4 (10−19)
p3 (0−9)
p2 (0−9)
p1 (0−9)
1 2 3 4 5 6 7 8 9 10 11 12 13 14 15 16 17 18 19 20 21 22 23 24 25 26 27 28
day

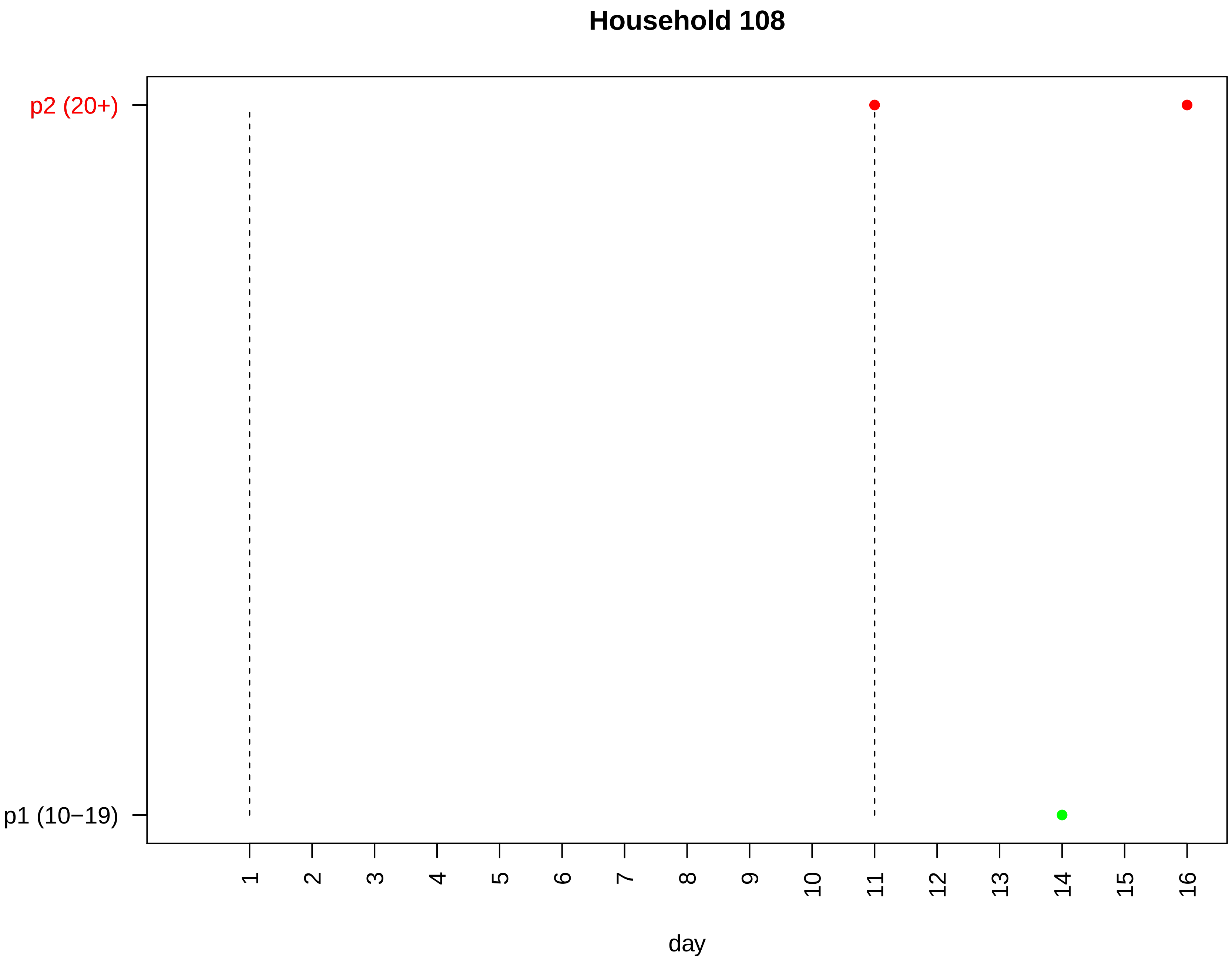

Household 108
p2 (20+)
p1 (10−19)
day

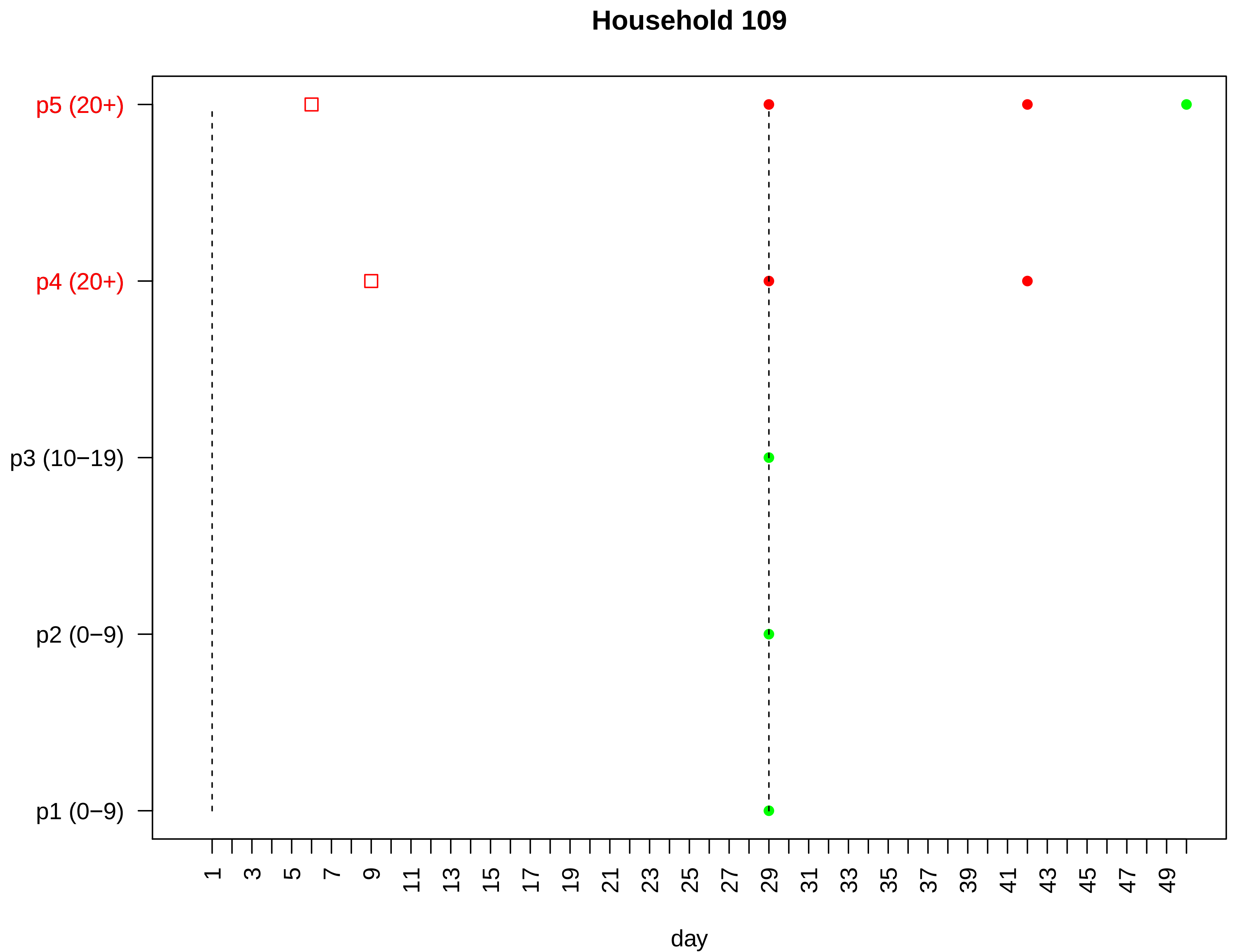

Household 109
p5 (20+)
p4 (20+)
p3 (10−19)
p2 (0−9)
p1 (0−9)
1
3
5
7
9
11
13
15
17
19
21
23
25
27
29
31
33
35
37
39
41
43
45
47
49
day

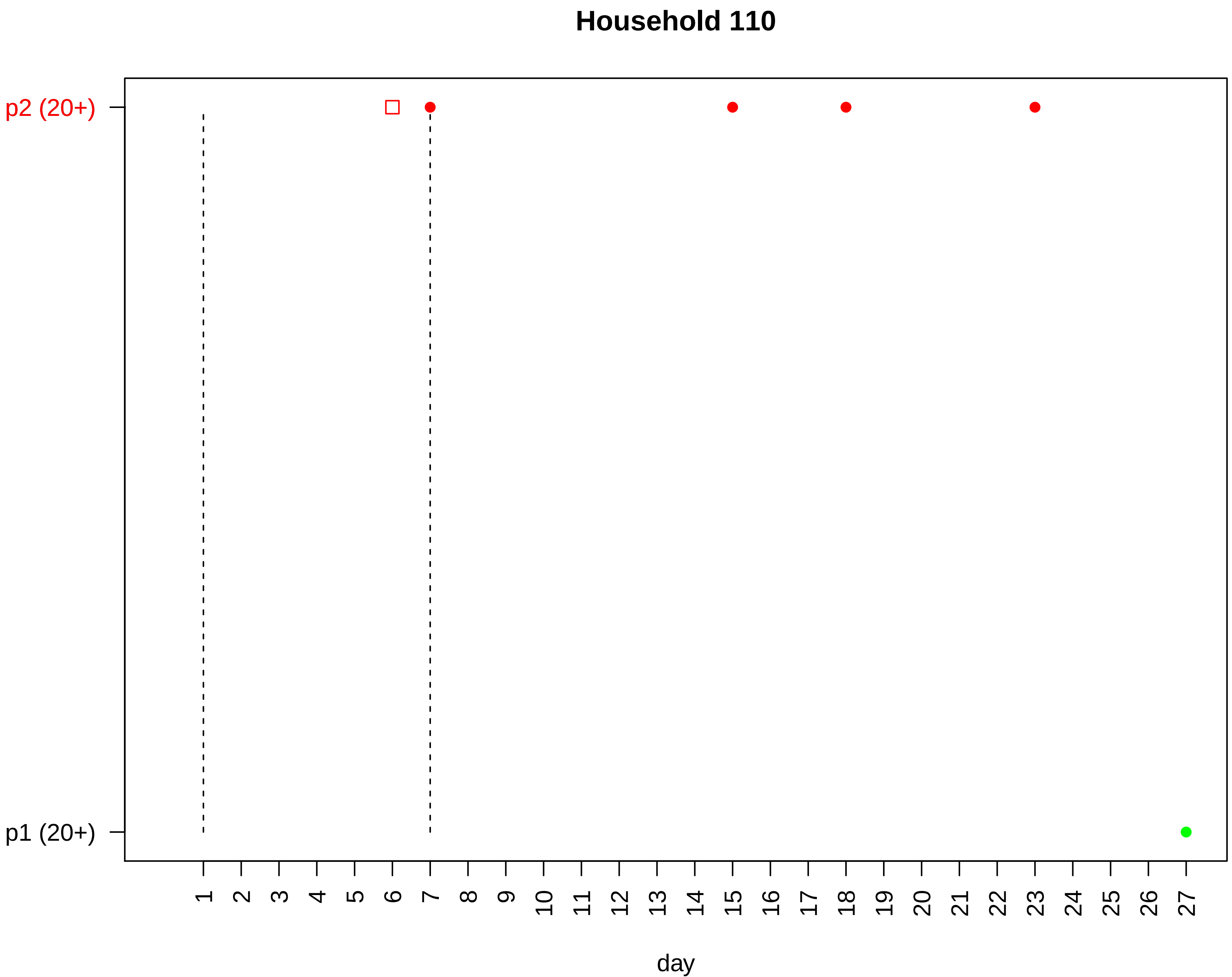

Household 110
p2 (20+)
p1 (20+)
day

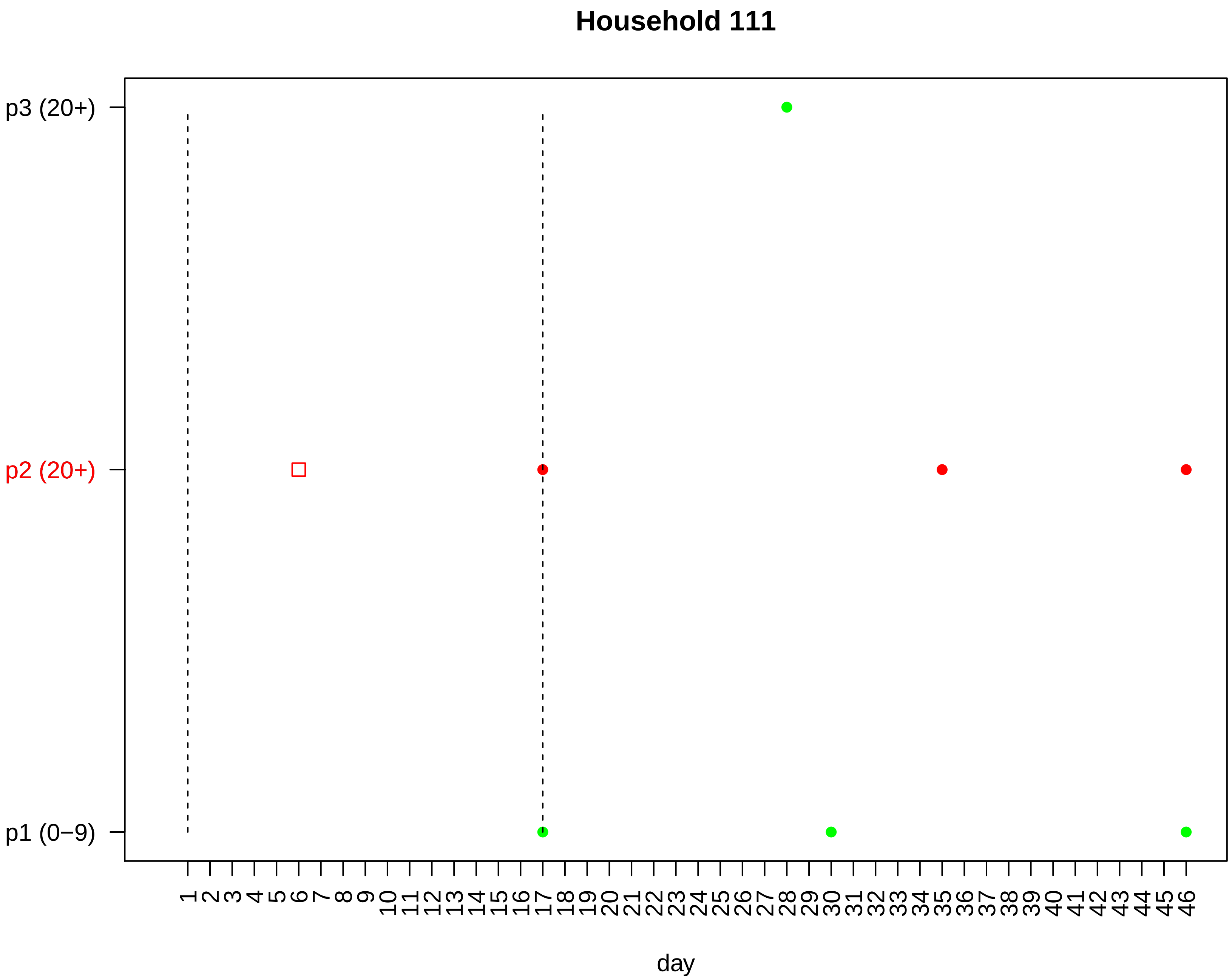

Household 111
p3 (20+)
p2 (20+)
p1 (0−9)
1 2 3 4 5 6 7 8 9 10 11 12 13 14 15 16 17 18 19 20 21 22 23 24 25 26 27 28 29 30 31 32 33 34 35 36 37 38 39 40 41 42 43 44 45 46
day

### Household 112

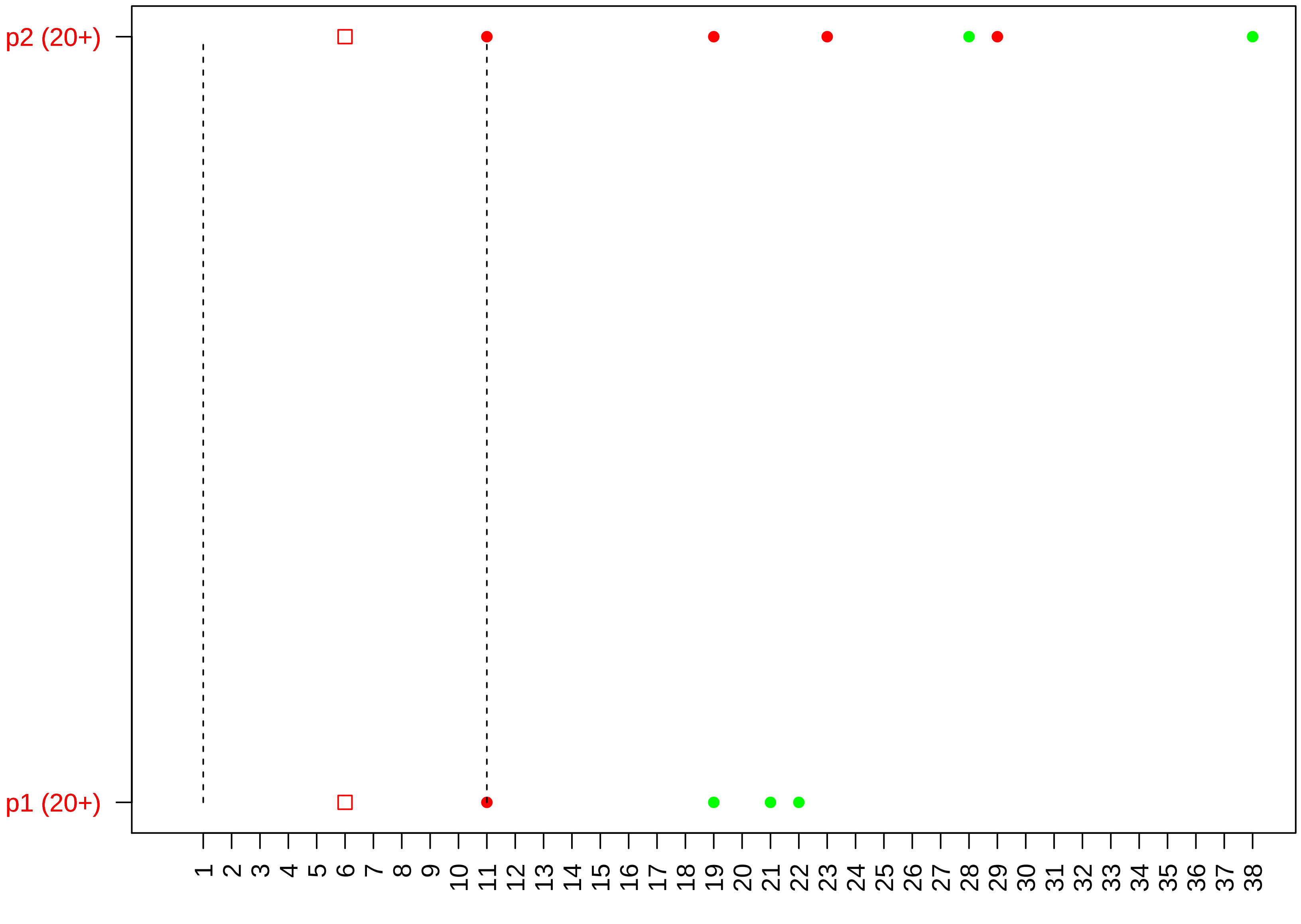

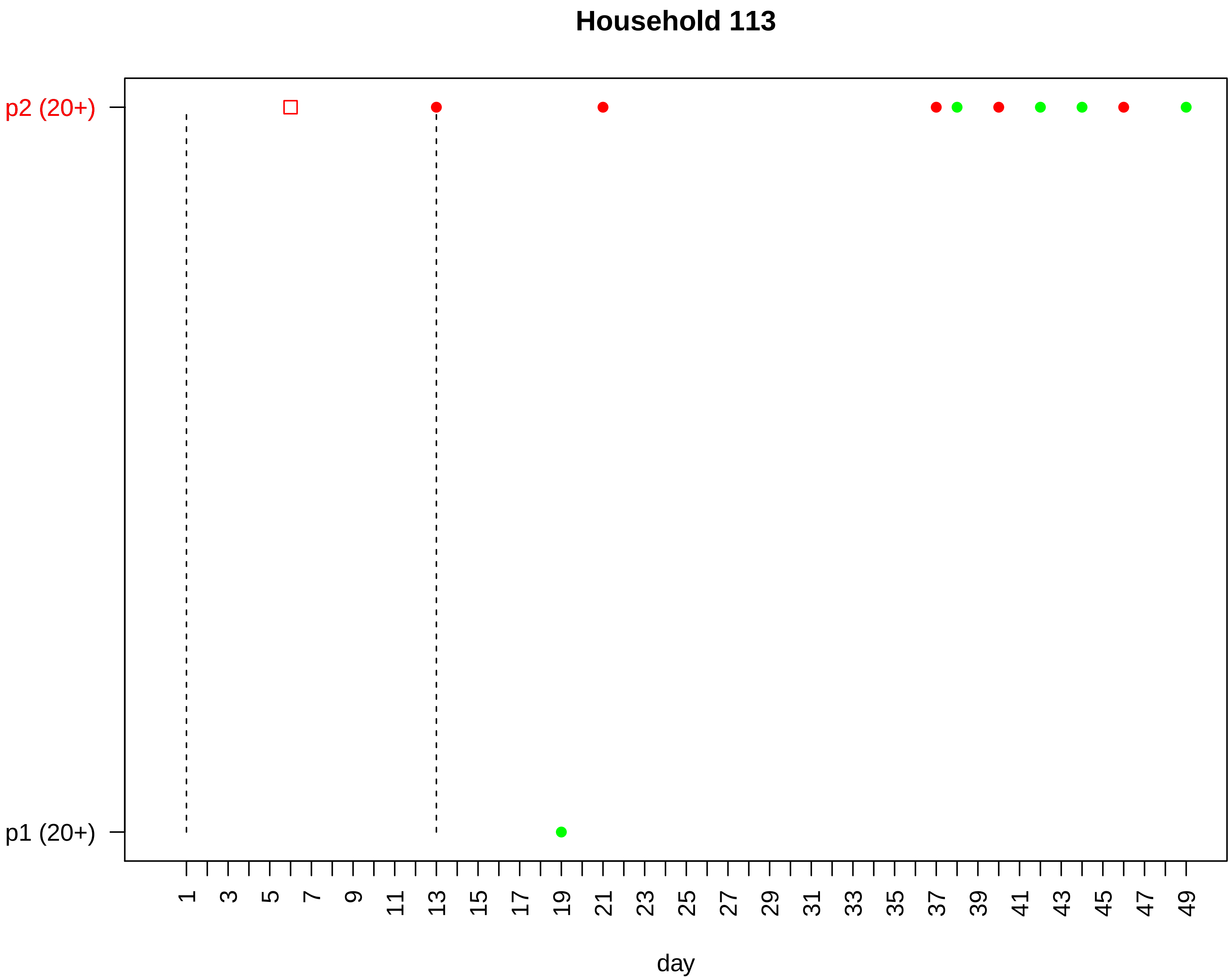
Household 113
p2 (20+)
p1 (20+)
1
3
5
7
9
11
13
15
17
19
21
23
25
27
29
31
33
35
37
39
41
43
45
47
49
day

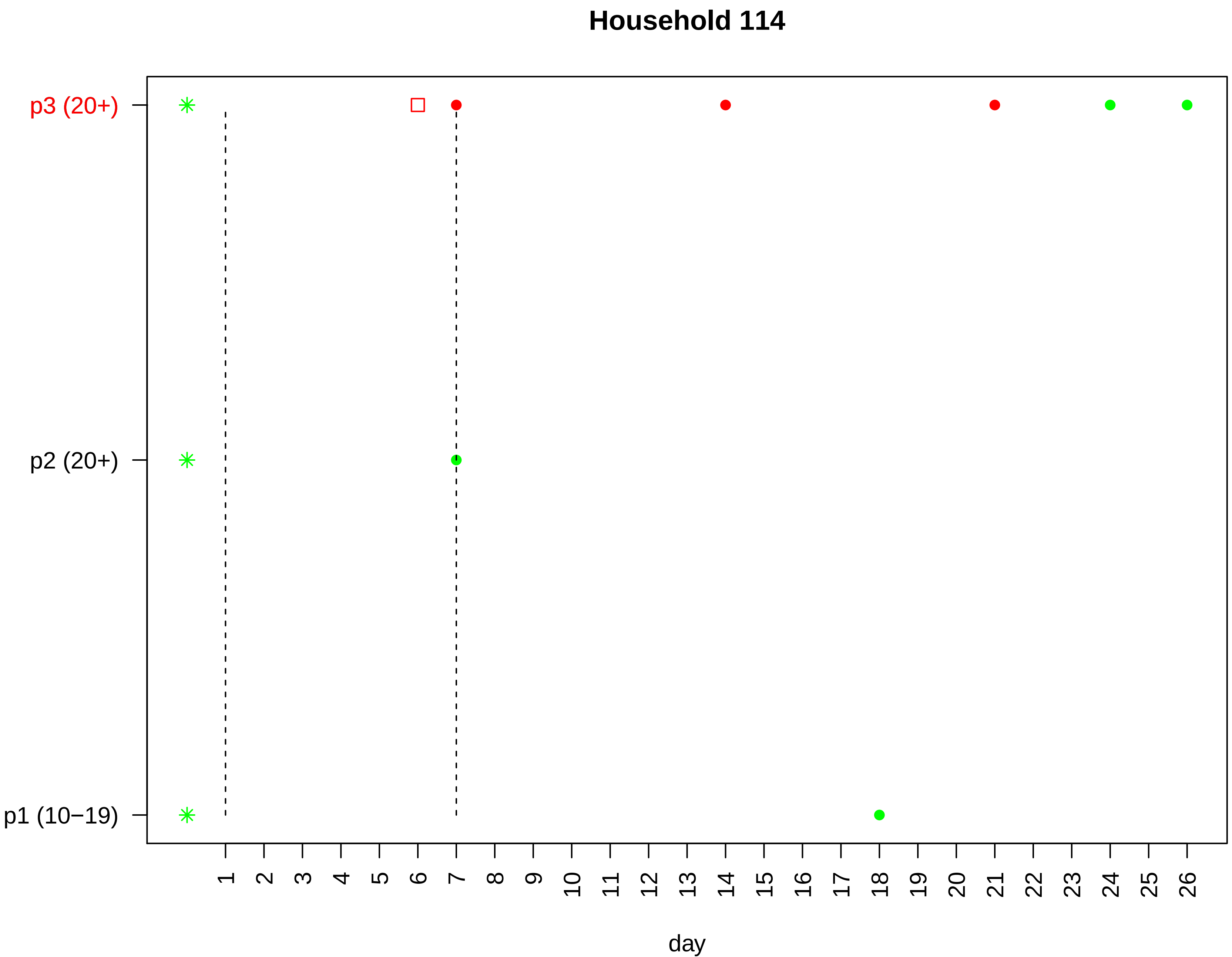

Household 114
p3 (20+)
p2 (20+)
p1 (10−19)
day

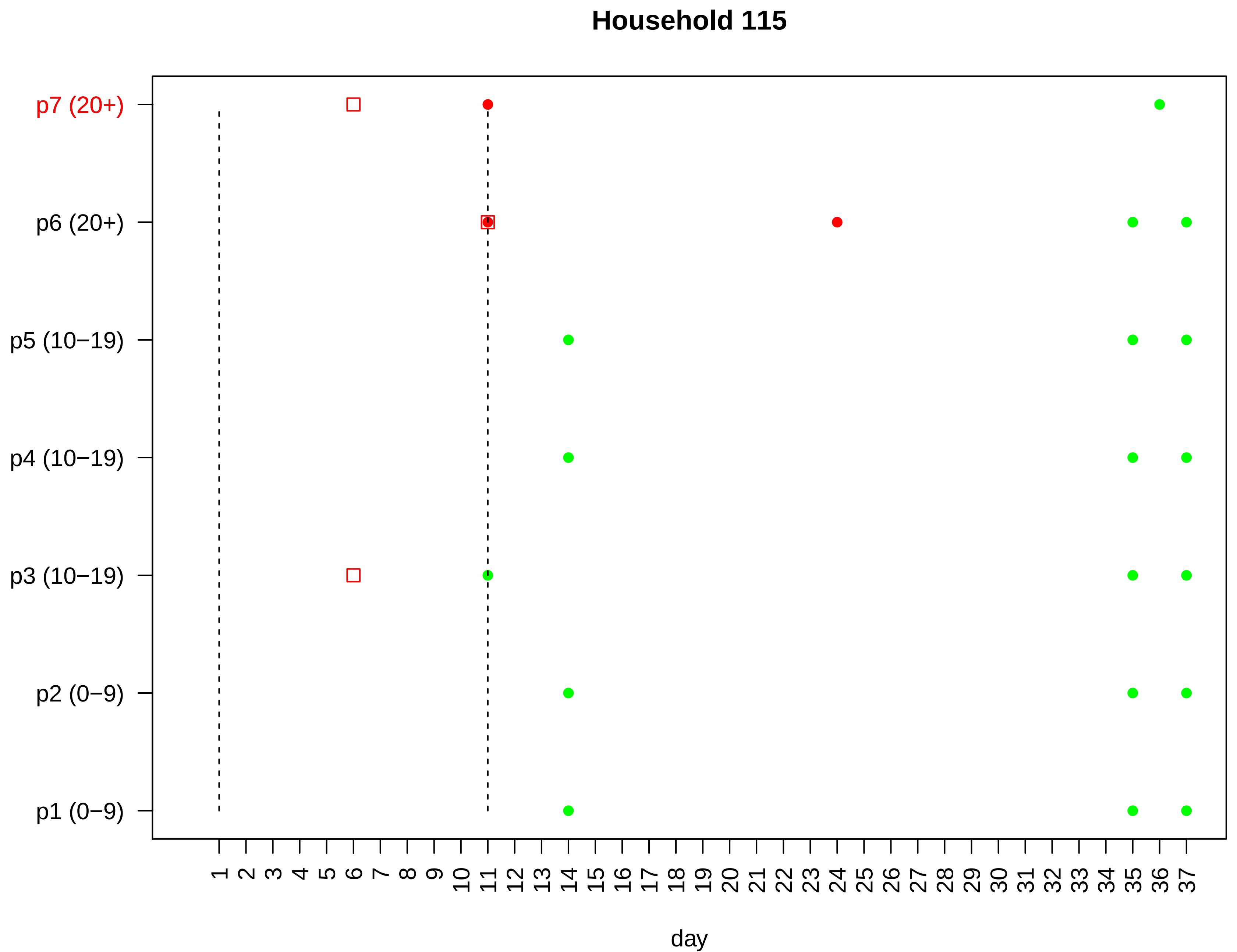

Household 115
p7 (20+)
p6 (20+)
p5 (10−19)
p4 (10−19)
p3 (10−19)
p2 (0−9)
p1 (0−9)
1 2 3 4 5 6 7 8 9 10 11 12 13 14 15 16 17 18 19 20 21 22 23 24 25 26 27 28 29 30 31 32 33 34 35 36 37
day

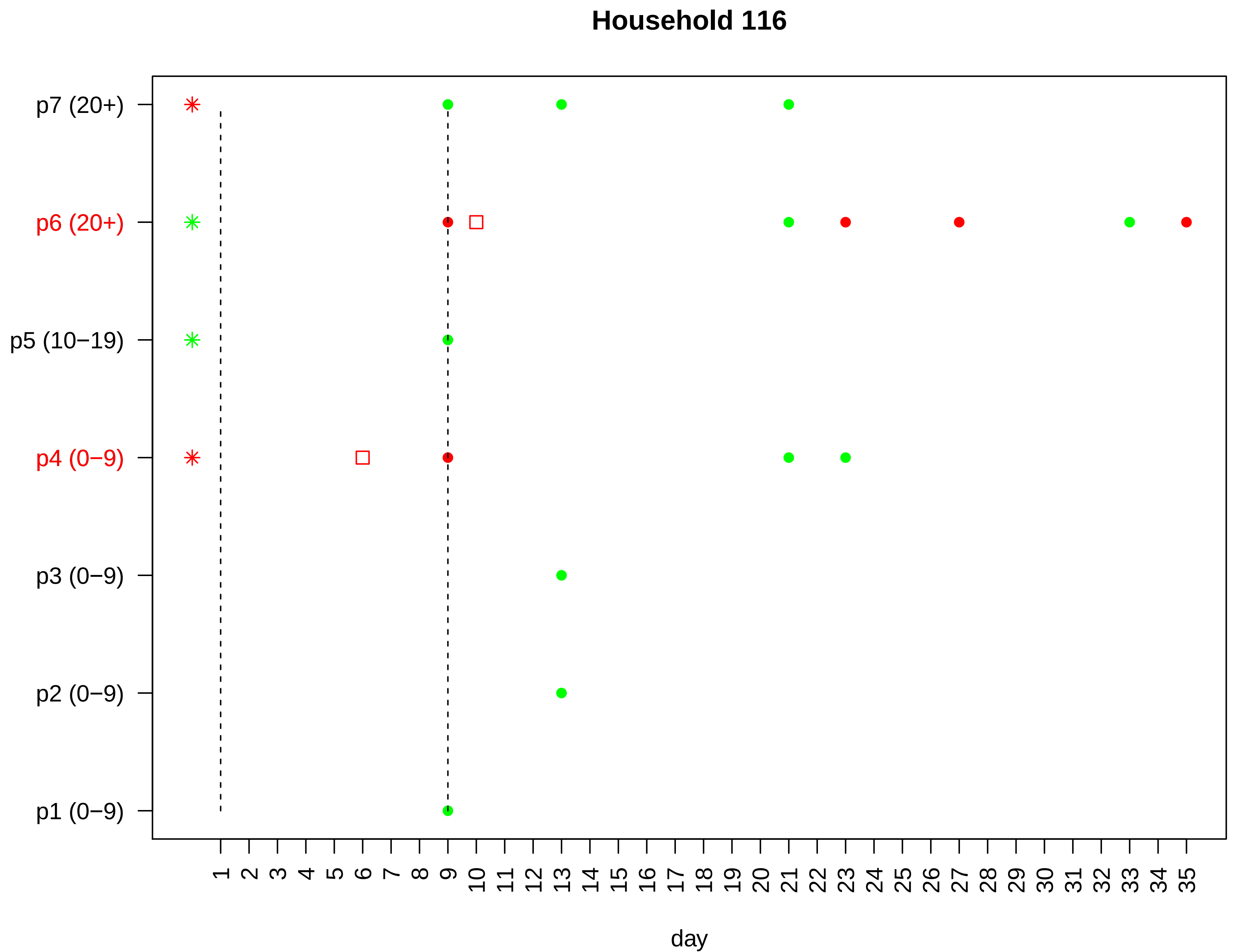

Household 116
p7 (20+)
p6 (20+)
p5 (10−19)
p4 (0−9)
p3 (0−9)
p2 (0−9)
p1 (0−9)
1 2 3 4 5 6 7 8 9 10 11 12 13 14 15 16 17 18 19 20 21 22 23 24 25 26 27 28 29 30 31 32 33 34 35
day

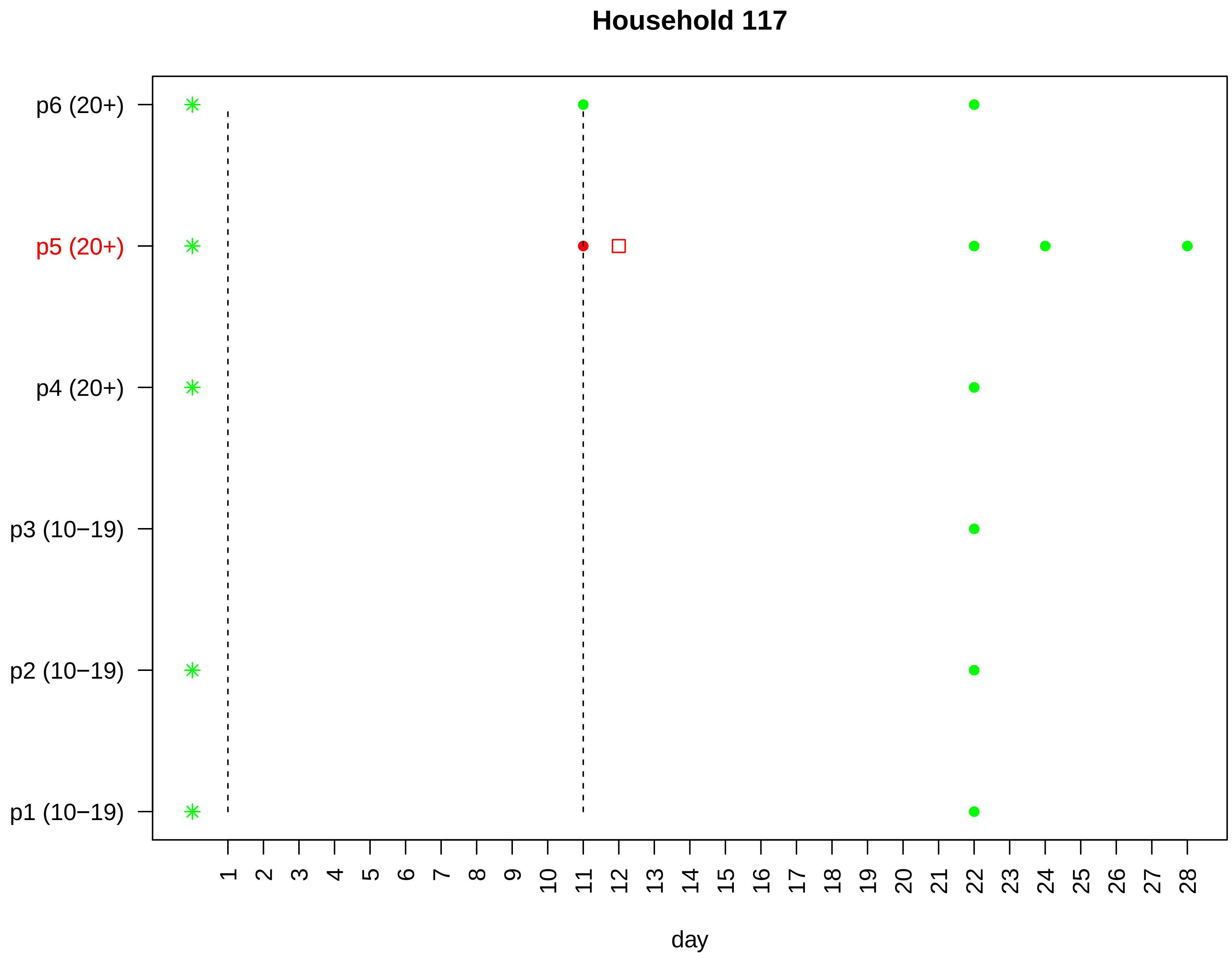

Household 117
p6 (20+)
p5 (20+)
p4 (20+)
p3 (10−19)
p2 (10−19)
p1 (10−19)
day

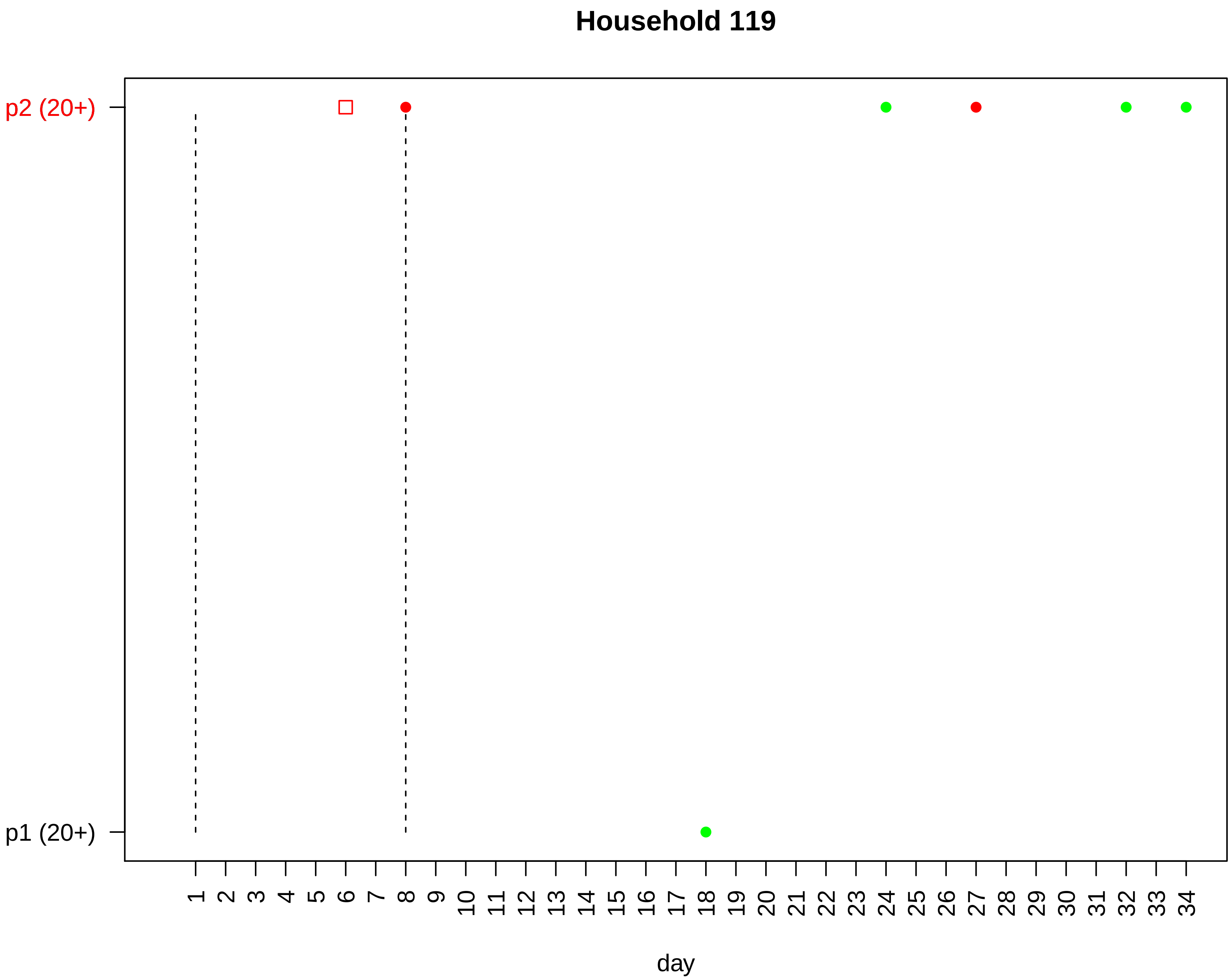

Household 119
p2 (20+)
p1 (20+)
day

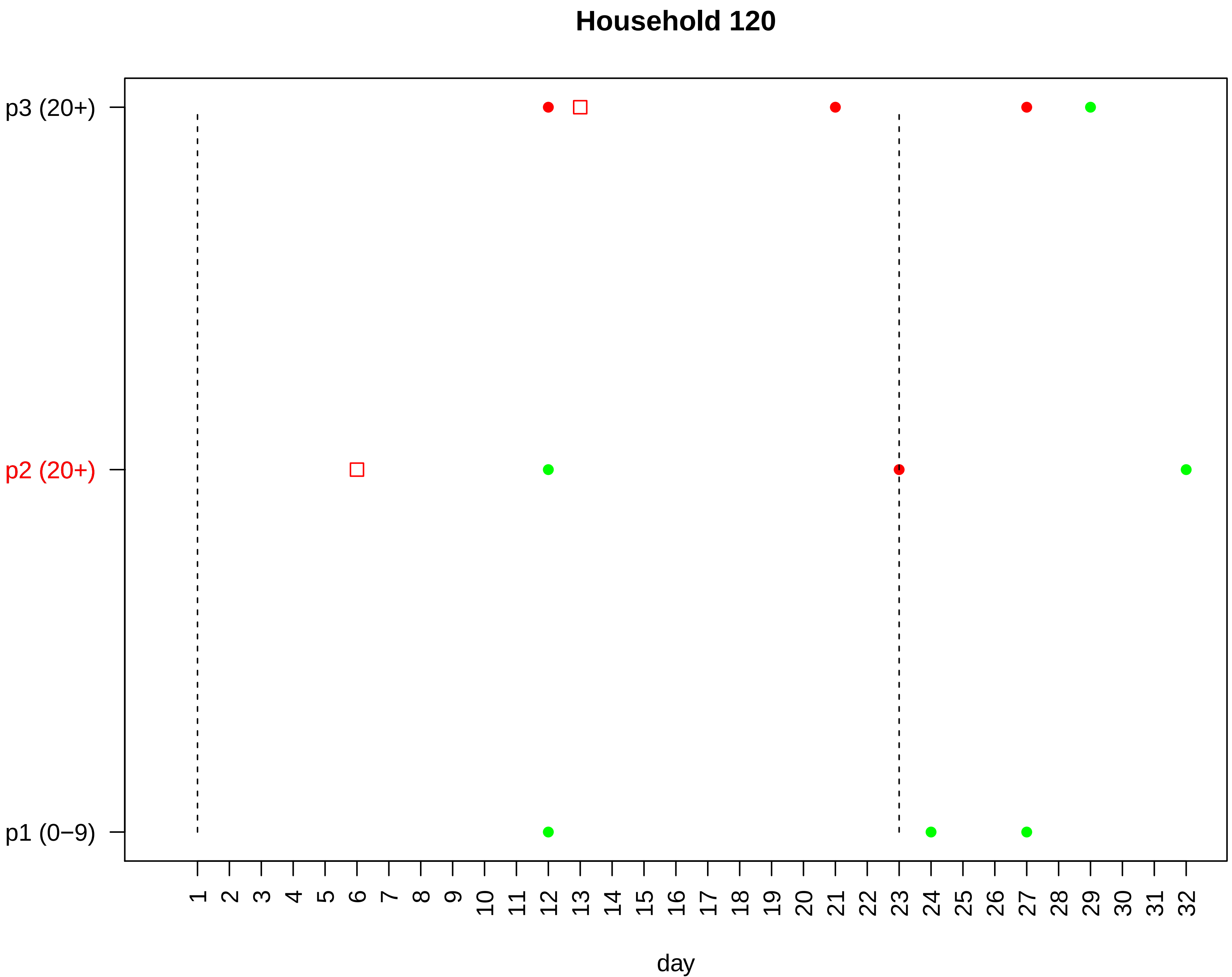

Household 120
p3 (20+)
p2 (20+)
p1 (0−9)
1 2 3 4 5 6 7 8 9 10 11 12 13 14 15 16 17 18 19 20 21 22 23 24 25 26 27 28 29 30 31 32
day

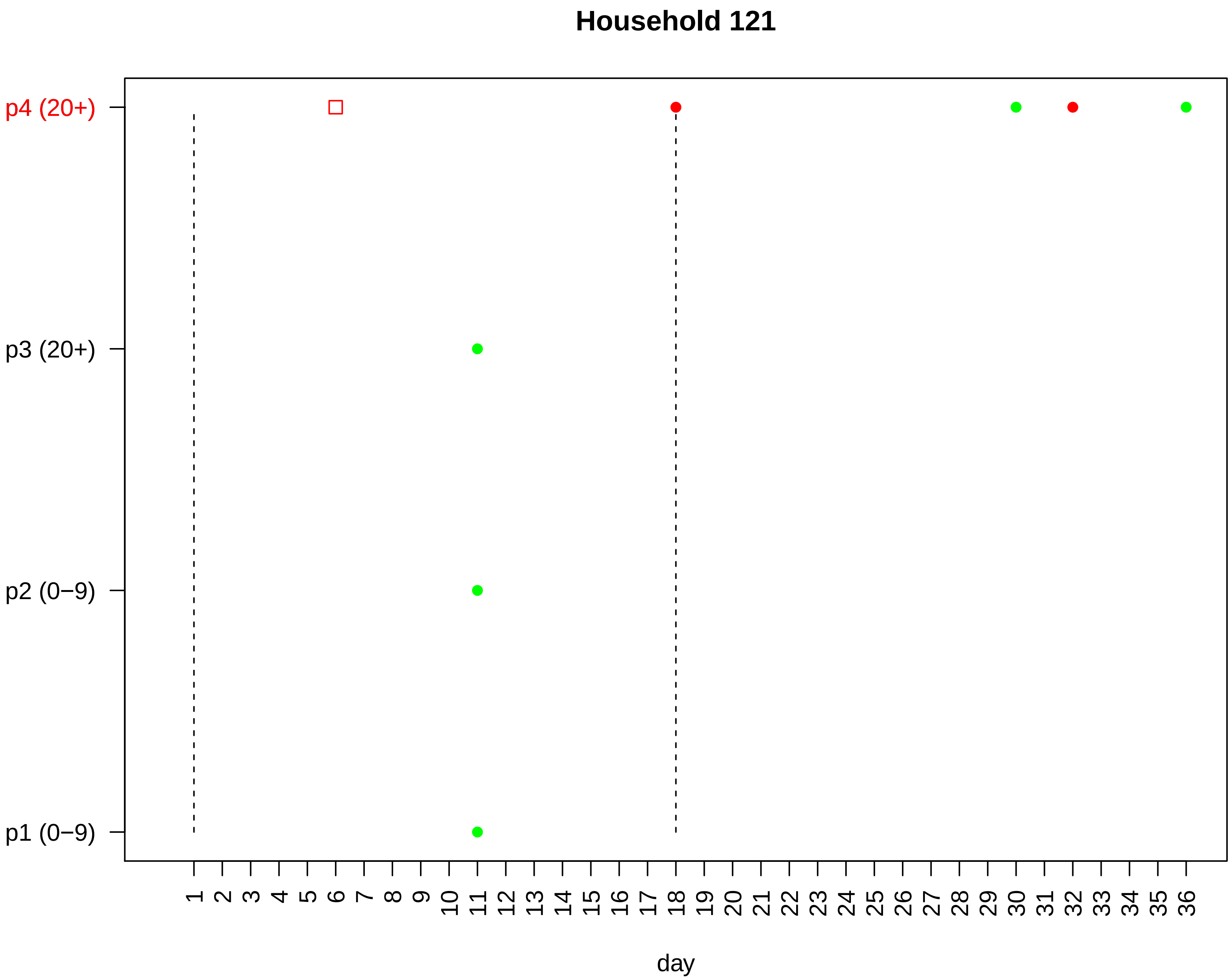

Household 121
p4 (20+)
p3 (20+)
p2 (0−9)
p1 (0−9)
1 2 3 4 5 6 7 8 9 10 11 12 13 14 15 16 17 18 19 20 21 22 23 24 25 26 27 28 29 30 31 32 33 34 35 36
day

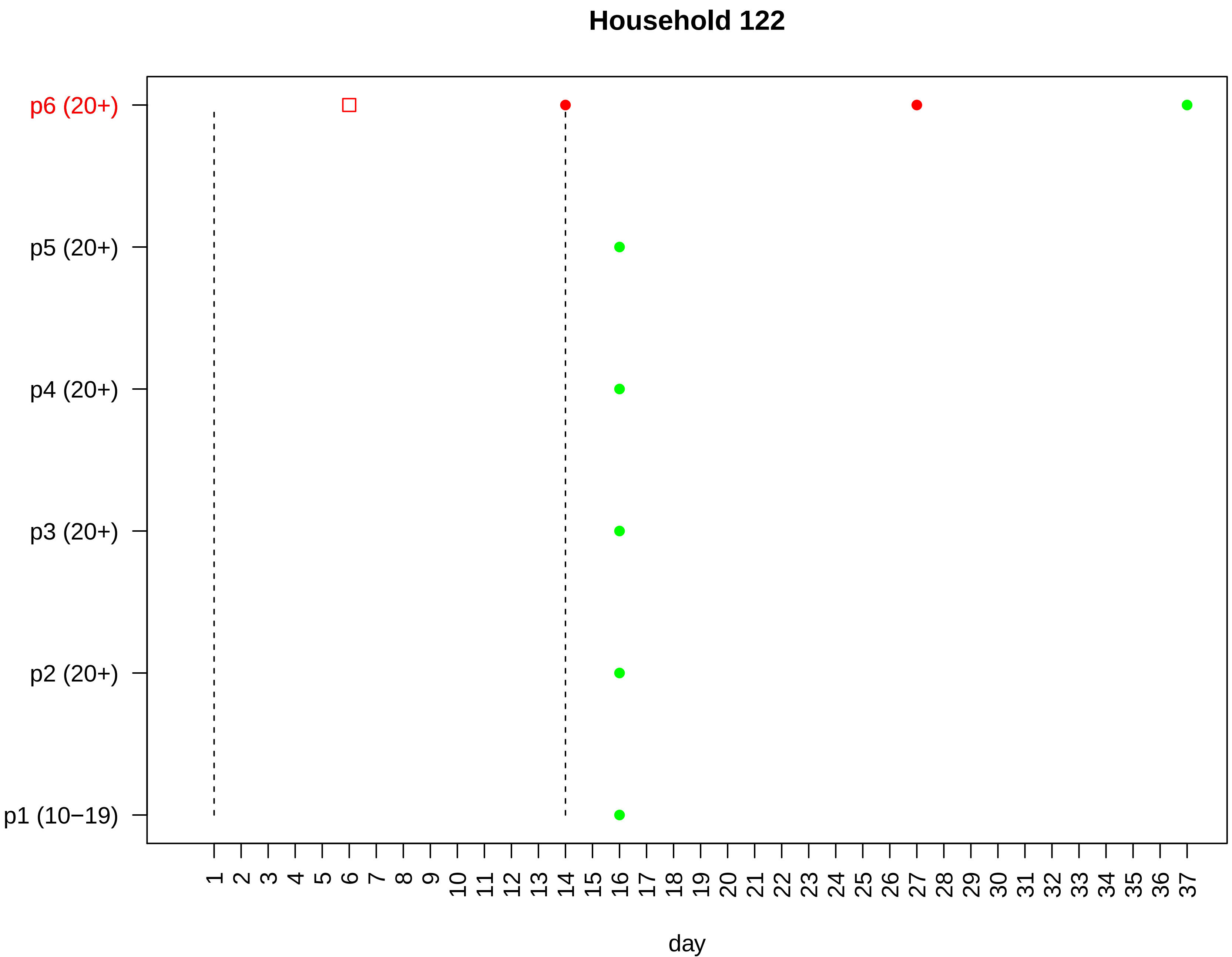

Household 122
p6 (20+)
p5 (20+)
p4 (20+)
p3 (20+)
p2 (20+)
p1 (10−19)
1 2 3 4 5 6 7 8 9 10 11 12 13 14 15 16 17 18 19 20 21 22 23 24 25 26 27 28 29 30 31 32 33 34 35 36 37
day

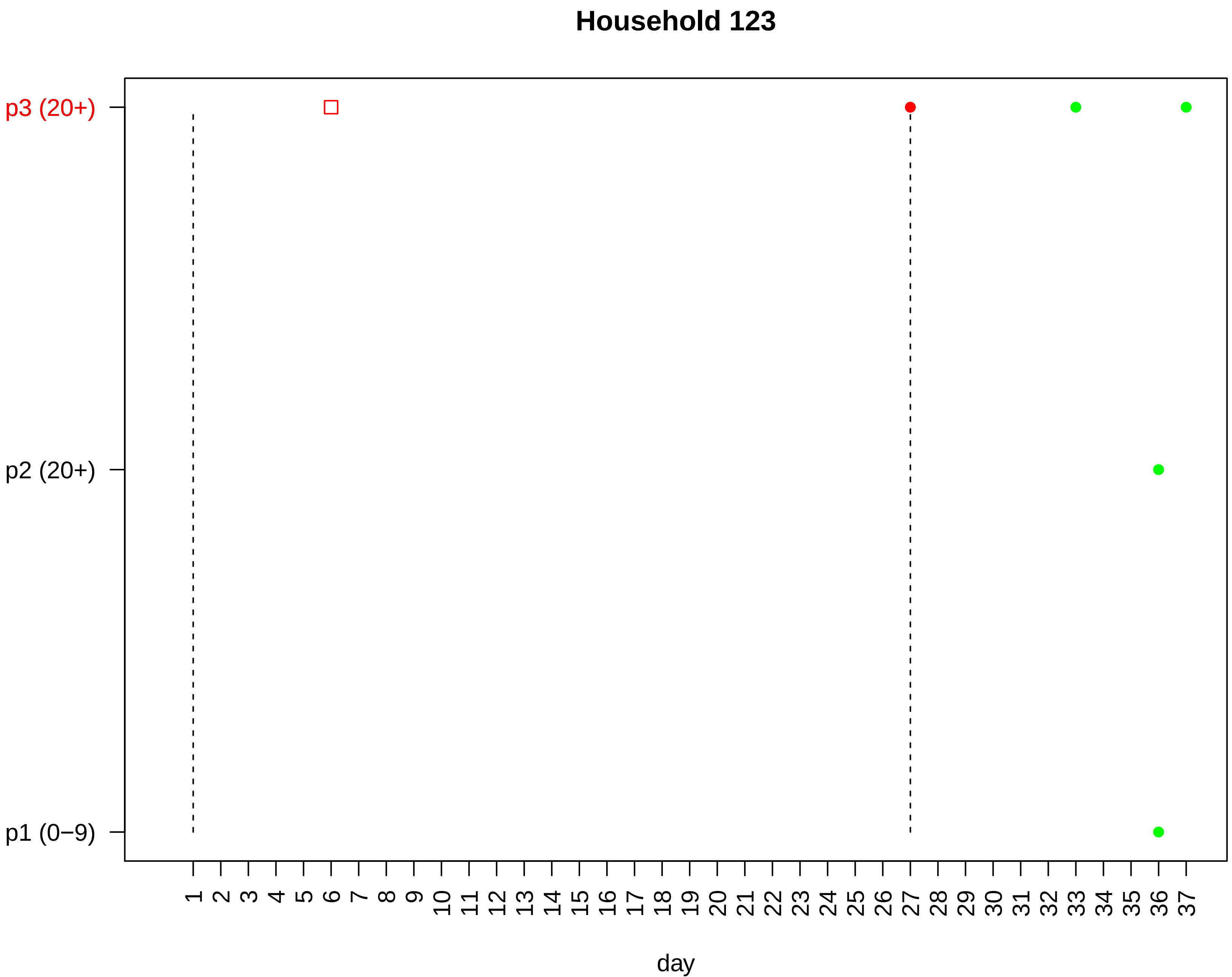
Household 123
p3 (20+)
p2 (20+)
p1 (0−9)
day

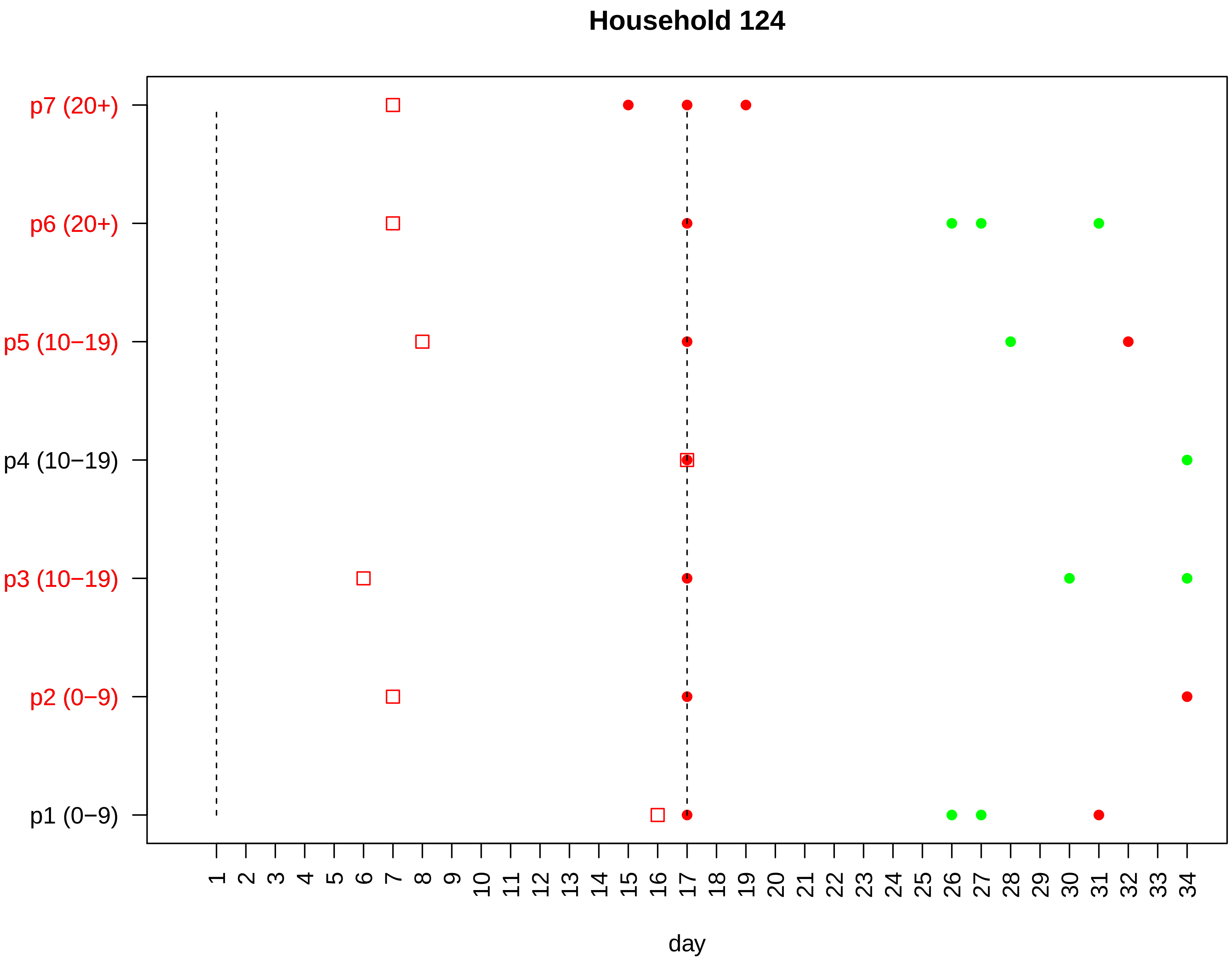
Household 124
p7 (20+)
p6 (20+)
p5 (10−19)
p4 (10−19)
p3 (10−19)
p2 (0−9)
p1 (0−9)
1 2 3 4 5 6 7 8 9 10 11 12 13 14 15 16 17 18 19 20 21 22 23 24 25 26 27 28 29 30 31 32 33 34
day

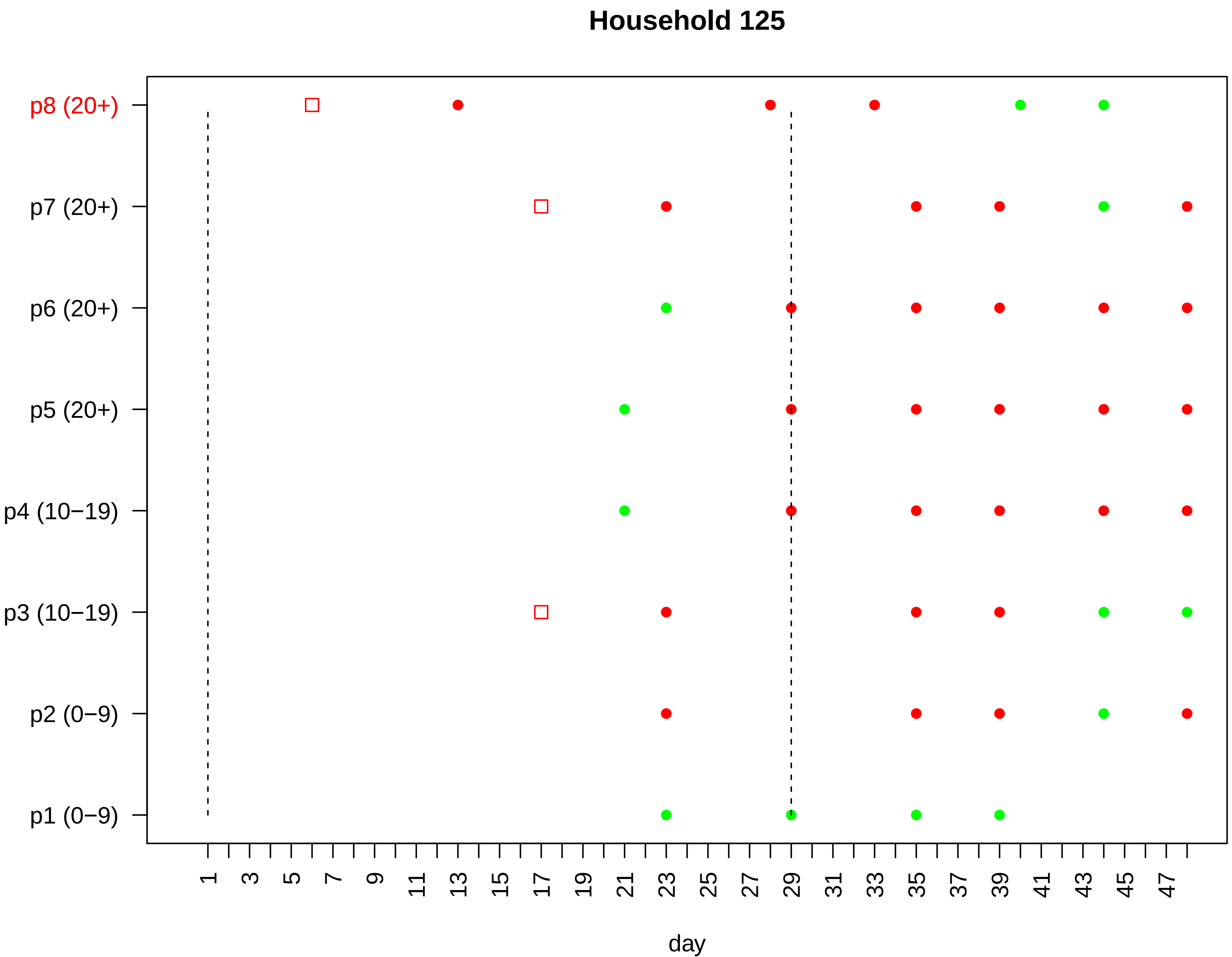

Household 125
p8 (20+)
p7 (20+)
p6 (20+)
p5 (20+)
p4 (10−19)
p3 (10−19)
p2 (0−9)
p1 (0−9)
1 3 5 7 9 11 13 15 17 19 21 23 25 27 29 31 33 35 37 39 41 43 45 47
day

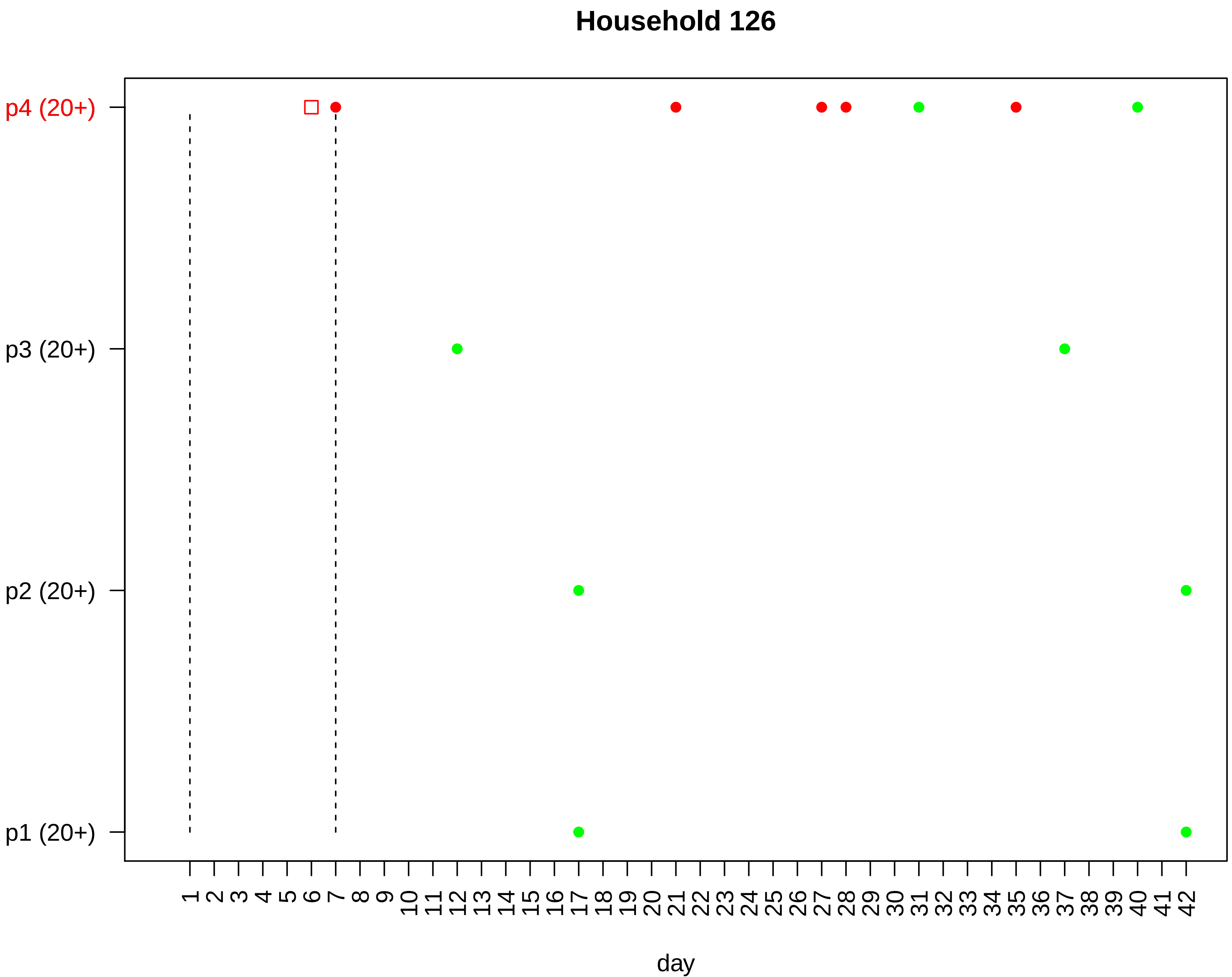
Household 126
p4 (20+)
p3 (20+)
p2 (20+)
p1 (20+)
1 2 3 4 5 6 7 8 9 10 11 12 13 14 15 16 17 18 19 20 21 22 23 24 25 26 27 28 29 30 31 32 33 34 35 36 37 38 39 40 41 42
day

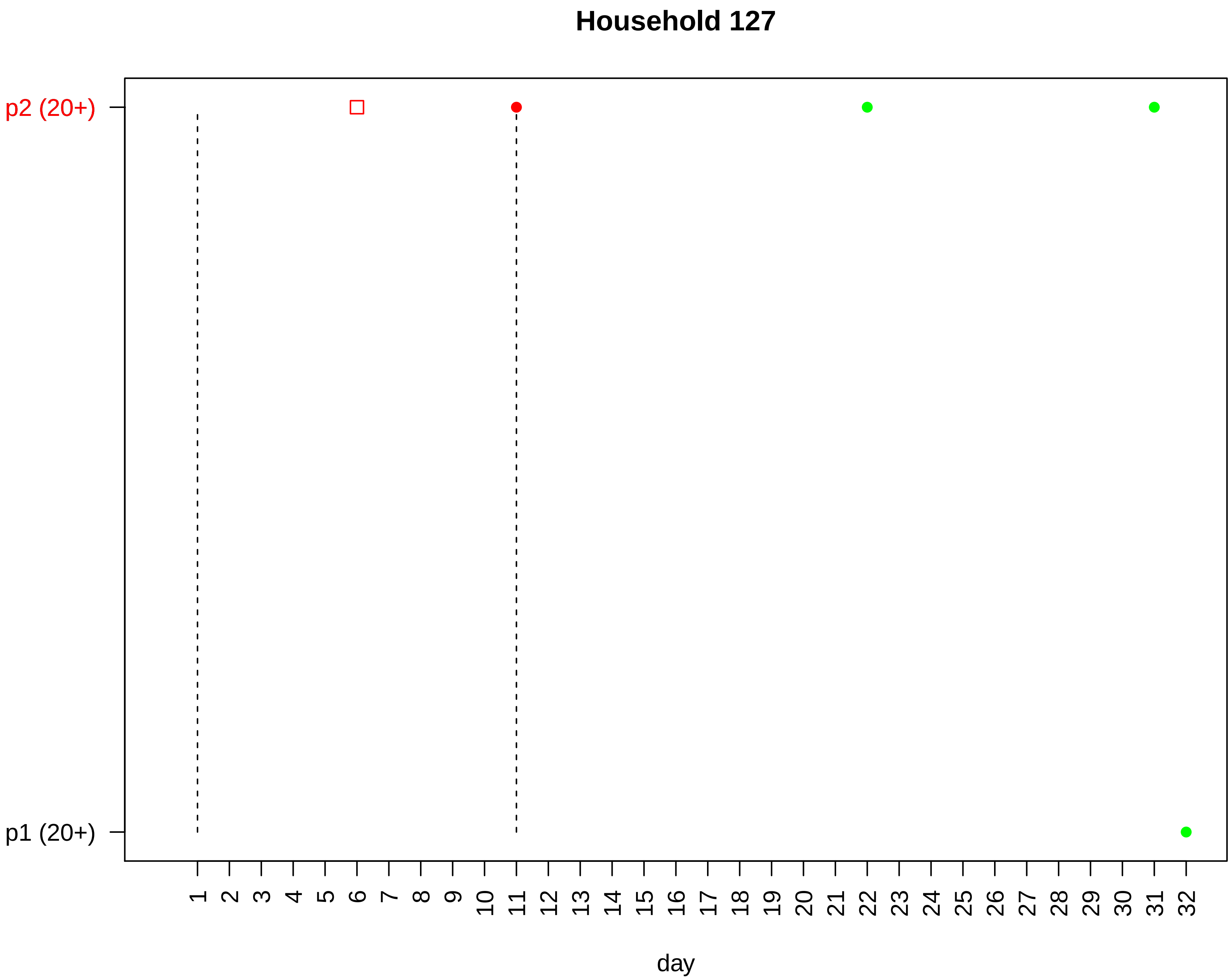

Household 127
p2 (20+)
p1 (20+)
day

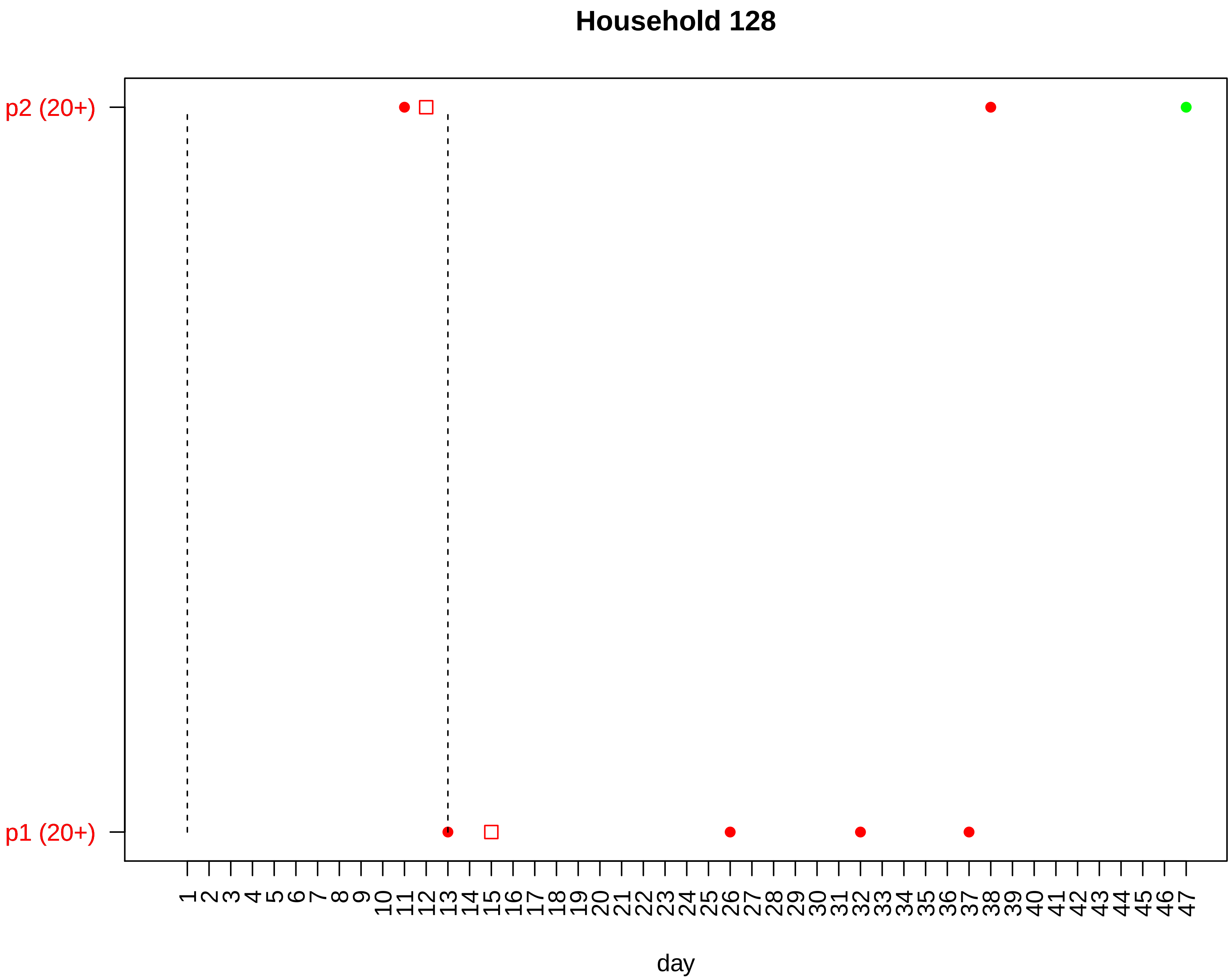
Household 128
p2 (20+)
p1 (20+)
1 2 3 4 5 6 7 8 9 10 11 12 13 14 15 16 17 18 19 20 21 22 23 24 25 26 27 28 29 30 31 32 33 34 35 36 37 38 39 40 41 42 43 44 45 46 47
day

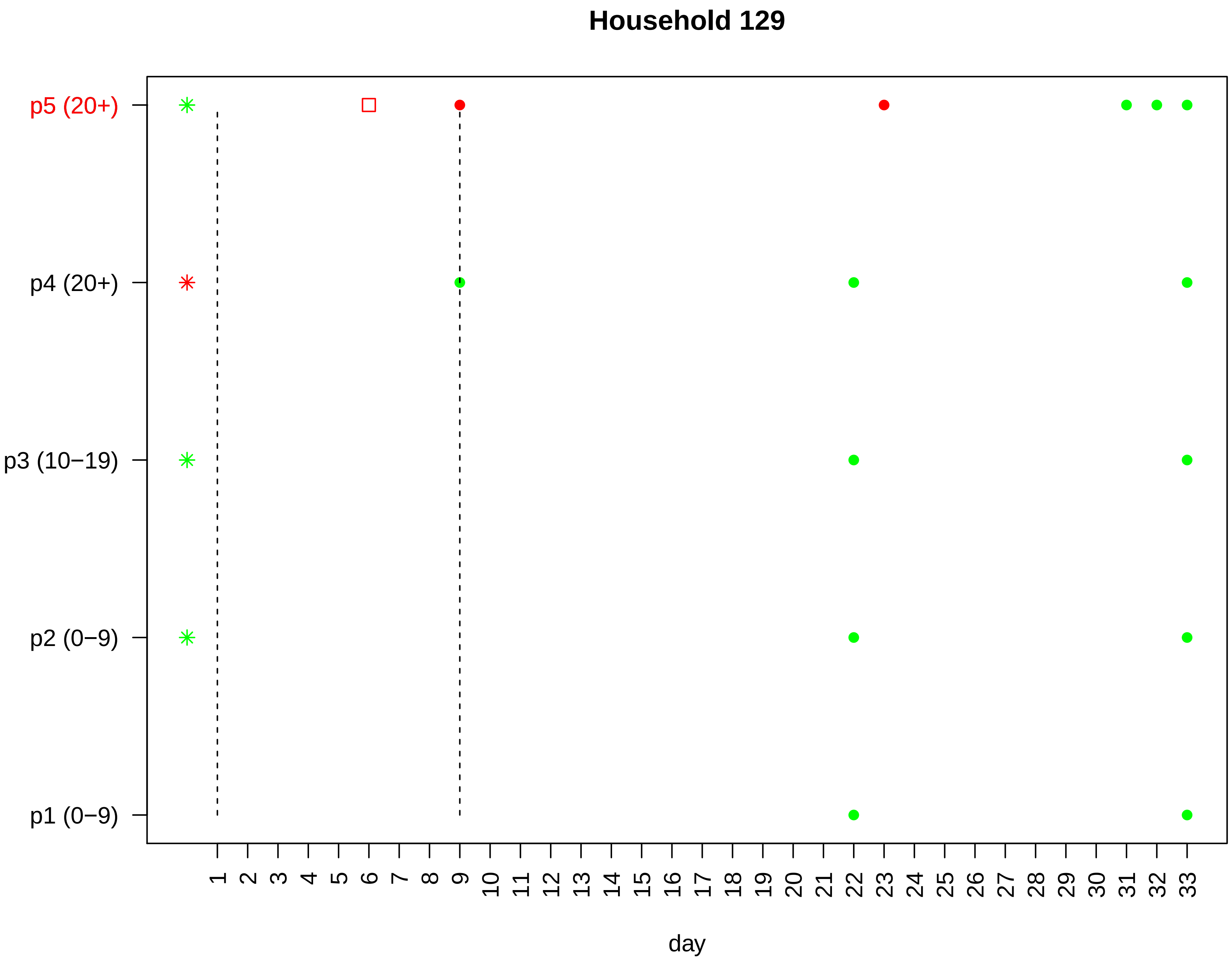

Household 129
p5 (20+)
p4 (20+)
p3 (10−19)
p2 (0−9)
p1 (0−9)
day

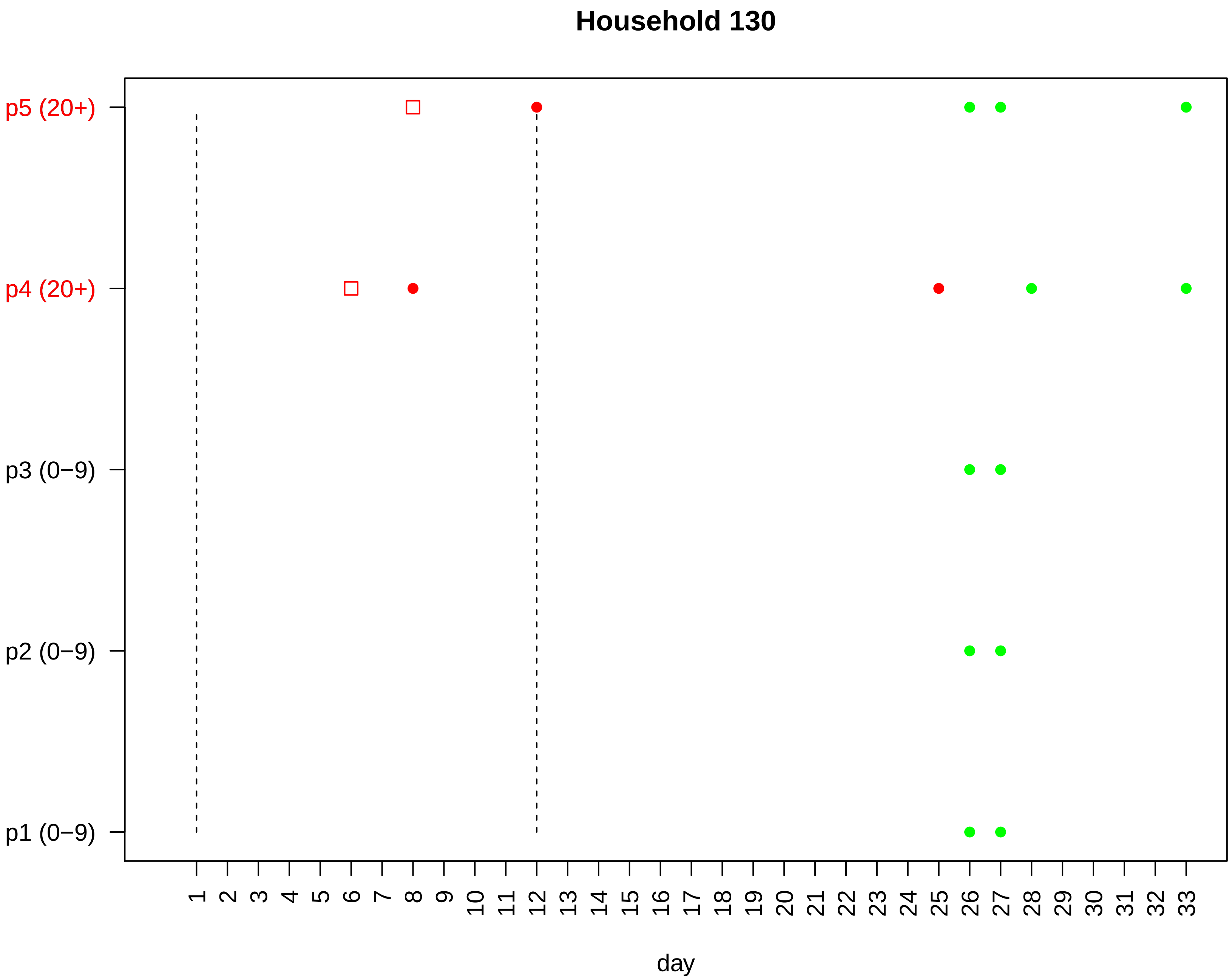

Household 130
p5 (20+)
p4 (20+)
p3 (0−9)
p2 (0−9)
p1 (0−9)
1 2 3 4 5 6 7 8 9 10 11 12 13 14 15 16 17 18 19 20 21 22 23 24 25 26 27 28 29 30 31 32 33
day

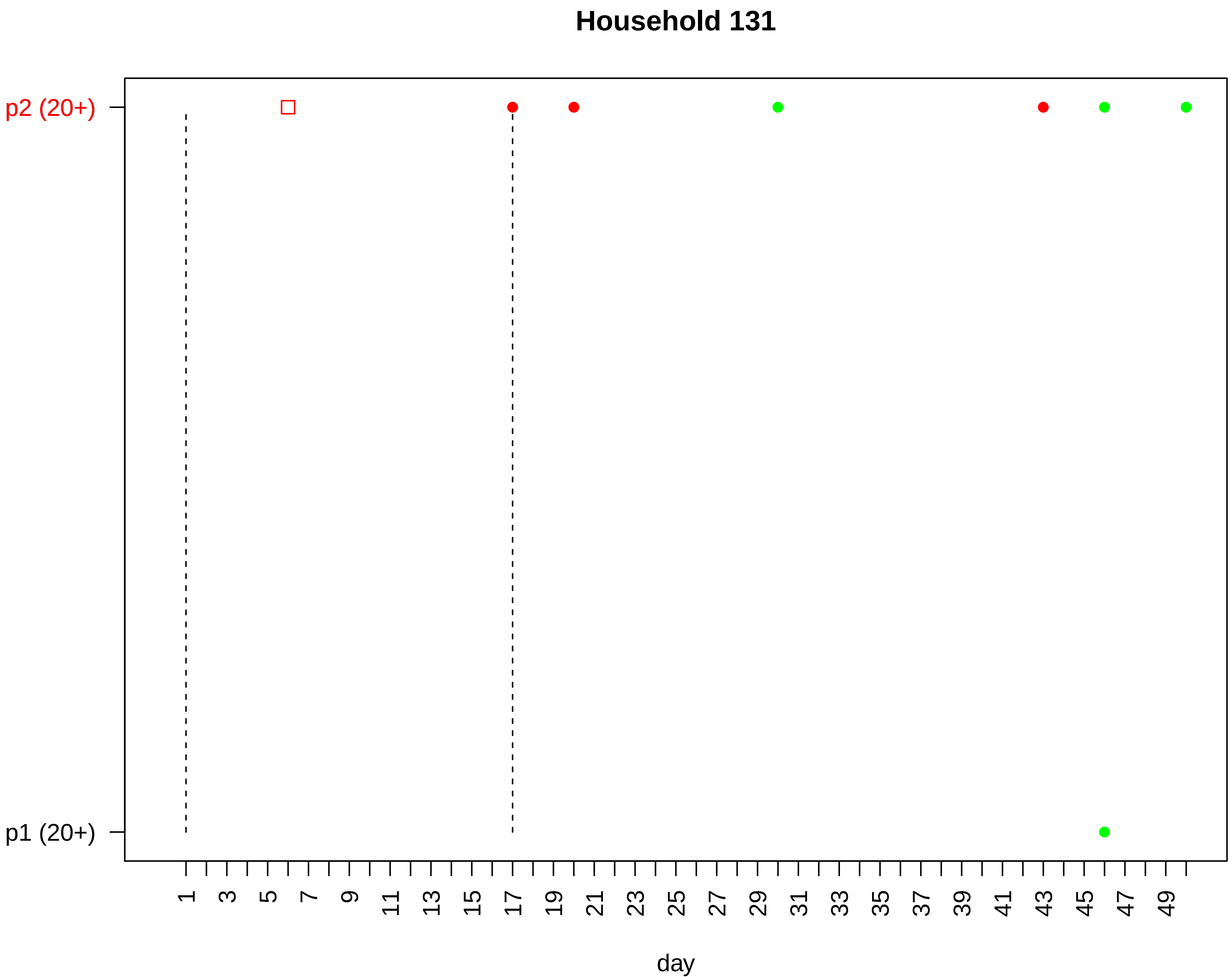

Household 131
p2 (20+)
p1 (20+)
1
3
5
7
9
11
13
15
17
19
21
23
25
27
29
31
33
35
37
39
41
43
45
47
49
day

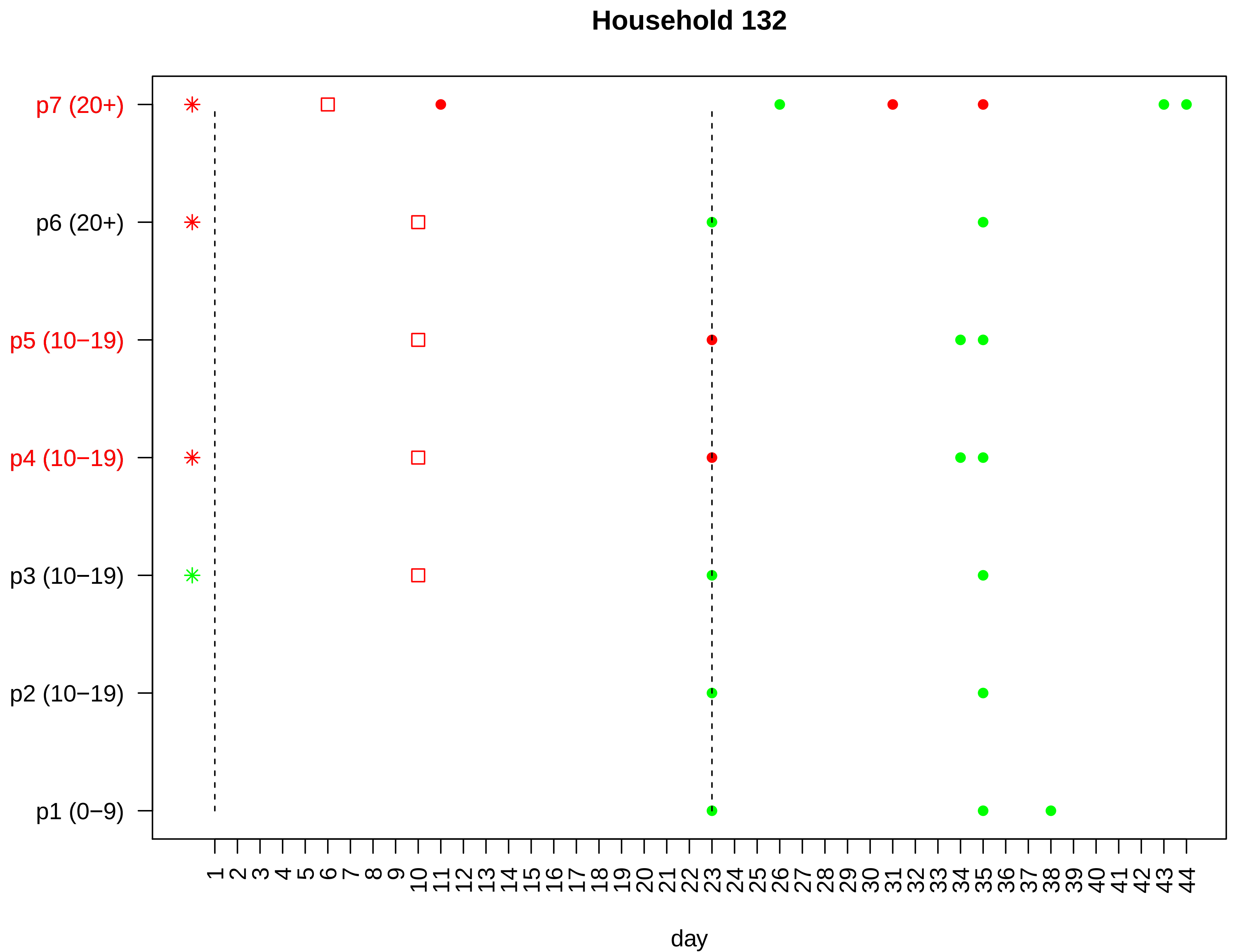
Household 132
p7 (20+)
p6 (20+)
p5 (10−19)
p4 (10−19)
p3 (10−19)
p2 (10−19)
p1 (0−9)
1 2 3 4 5 6 7 8 9 10 11 12 13 14 15 16 17 18 19 20 21 22 23 24 25 26 27 28 29 30 31 32 33 34 35 36 37 38 39 40 41 42 43 44
day

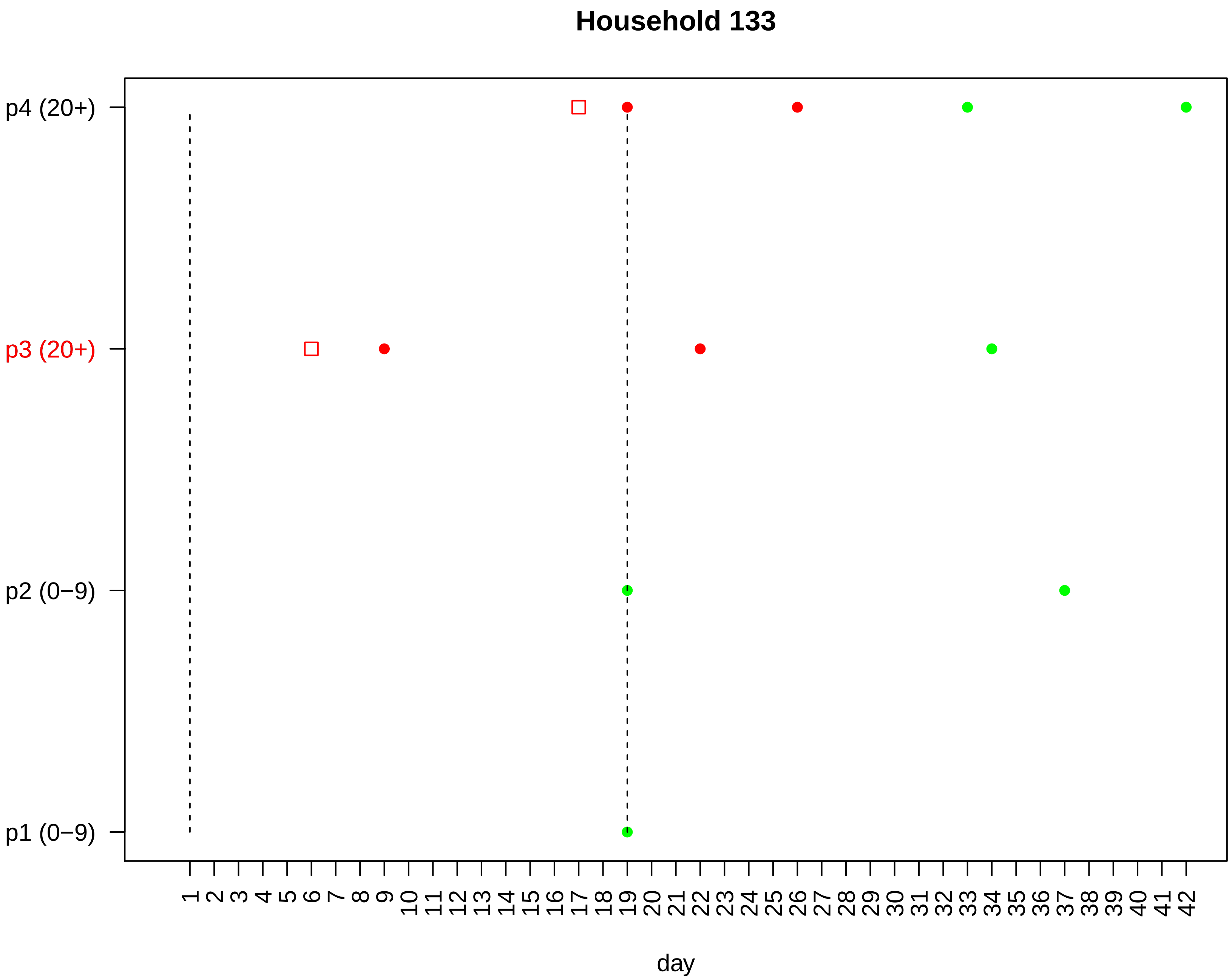

Household 133
p4 (20+)
p3 (20+)
p2 (0−9)
p1 (0−9)
1 2 3 4 5 6 7 8 9 10 11 12 13 14 15 16 17 18 19 20 21 22 23 24 25 26 27 28 29 30 31 32 33 34 35 36 37 38 39 40 41 42
day

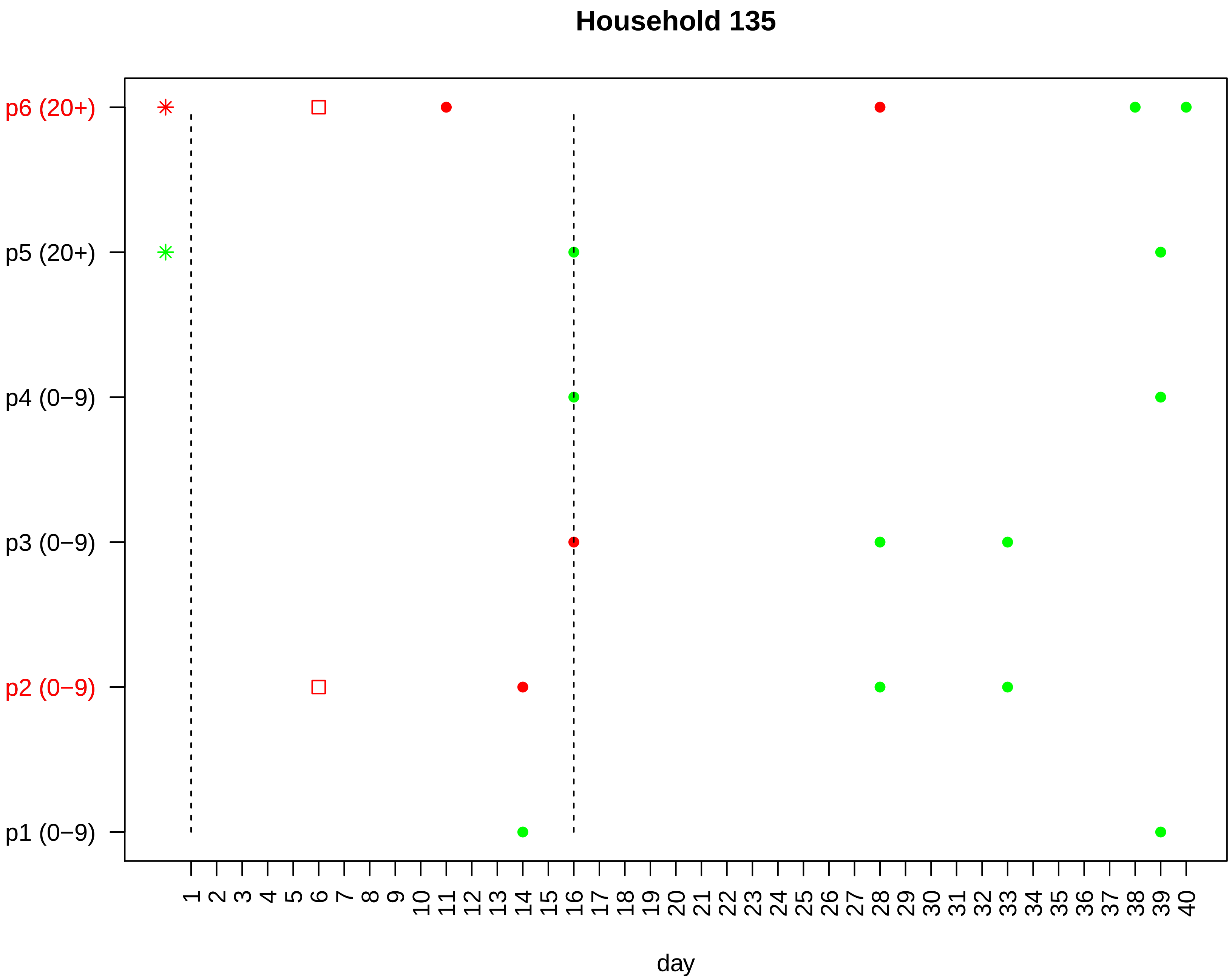

Household 135
p6 (20+)
p5 (20+)
p4 (0−9)
p3 (0−9)
p2 (0−9)
p1 (0−9)
1 2 3 4 5 6 7 8 9 10 11 12 13 14 15 16 17 18 19 20 21 22 23 24 25 26 27 28 29 30 31 32 33 34 35 36 37 38 39 40
day

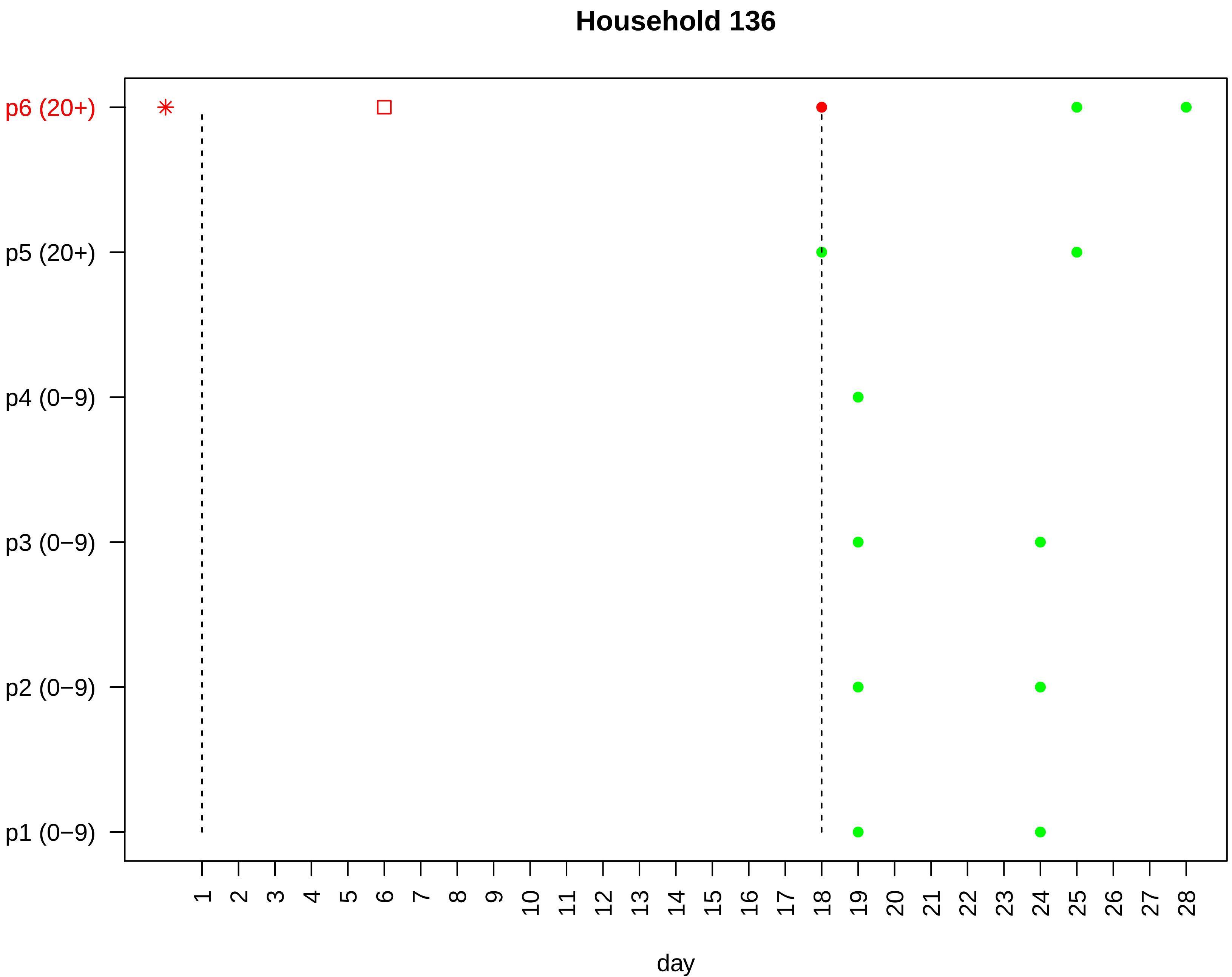
Household 136
p6 (20+)
p5 (20+)
p4 (0−9)
p3 (0−9)
p2 (0−9)
p1 (0−9)
1 2 3 4 5 6 7 8 9 10 11 12 13 14 15 16 17 18 19 20 21 22 23 24 25 26 27 28
day

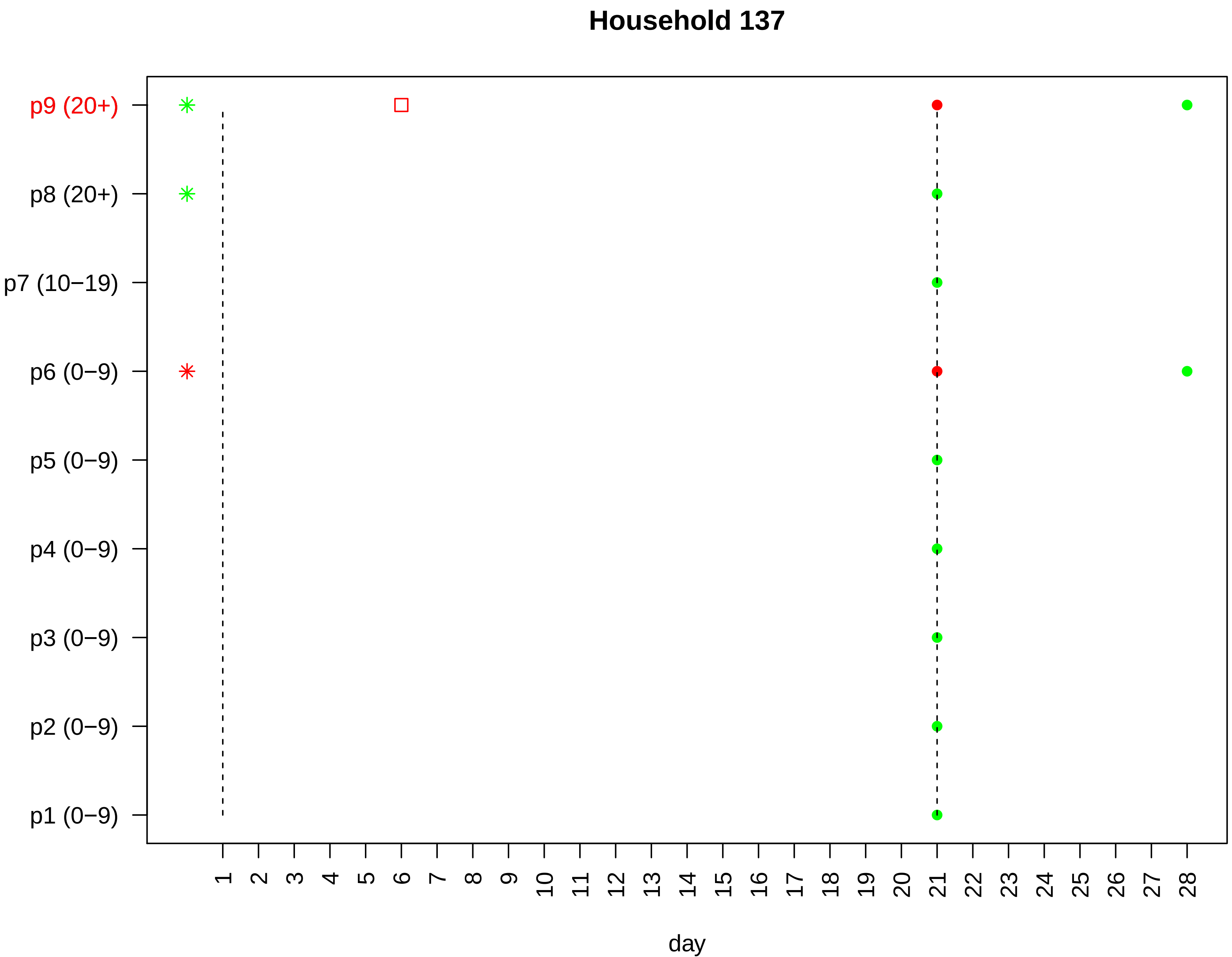

Household 137
p9 (20+)
p8 (20+)
p7 (10−19)
p6 (0−9)
p5 (0−9)
p4 (0−9)
p3 (0−9)
p2 (0−9)
p1 (0−9)
day

#### Household 138

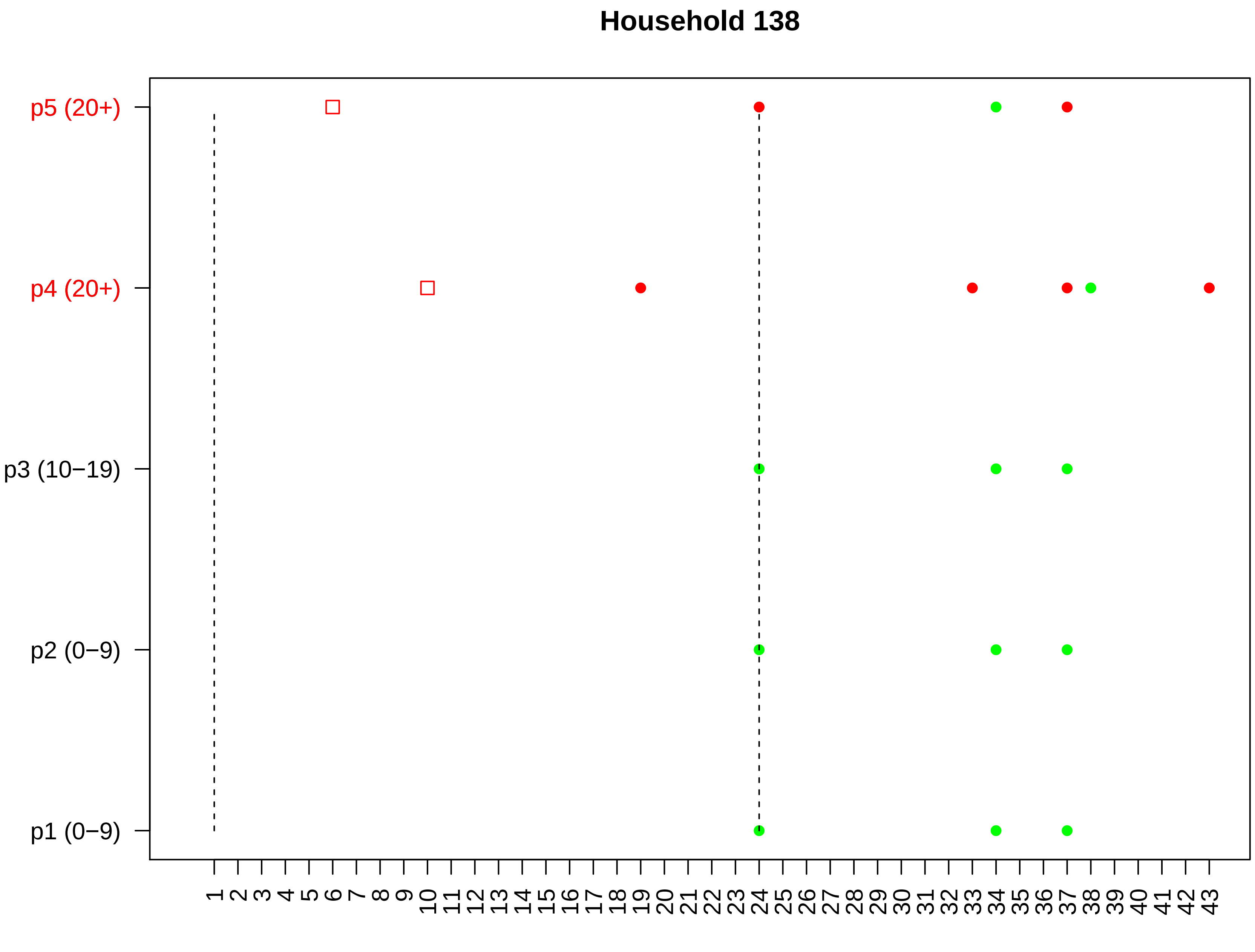

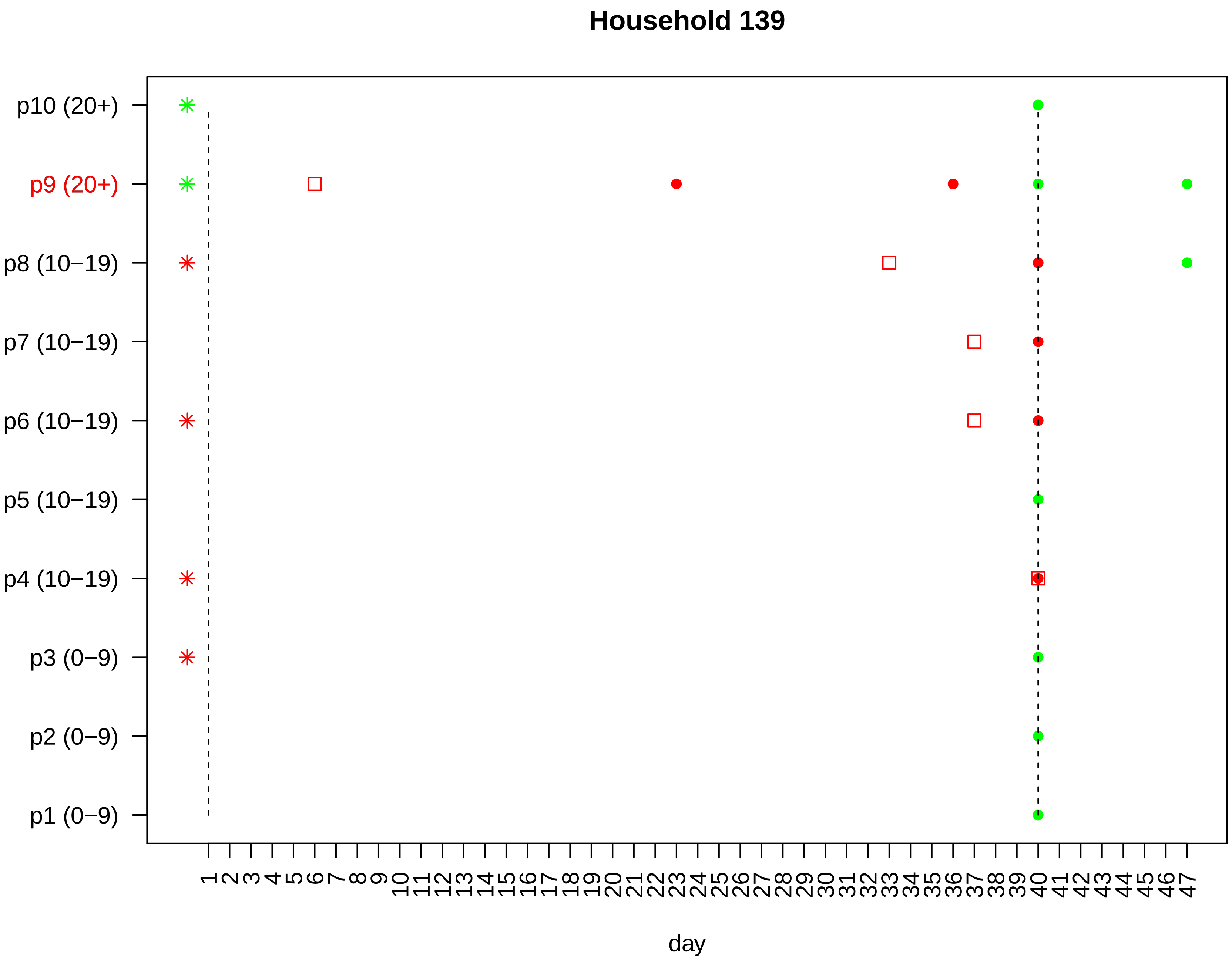
Household 139
p10 (20+)
p9 (20+)
p8 (10−19)
p7 (10−19)
p6 (10−19)
p5 (10−19)
p4 (10−19)
p3 (0−9)
p2 (0−9)
p1 (0−9)
1 2 3 4 5 6 7 8 9 10 11 12 13 14 15 16 17 18 19 20 21 22 23 24 25 26 27 28 29 30 31 32 33 34 35 36 37 38 39 40 41 42 43 44 45 46 47
day

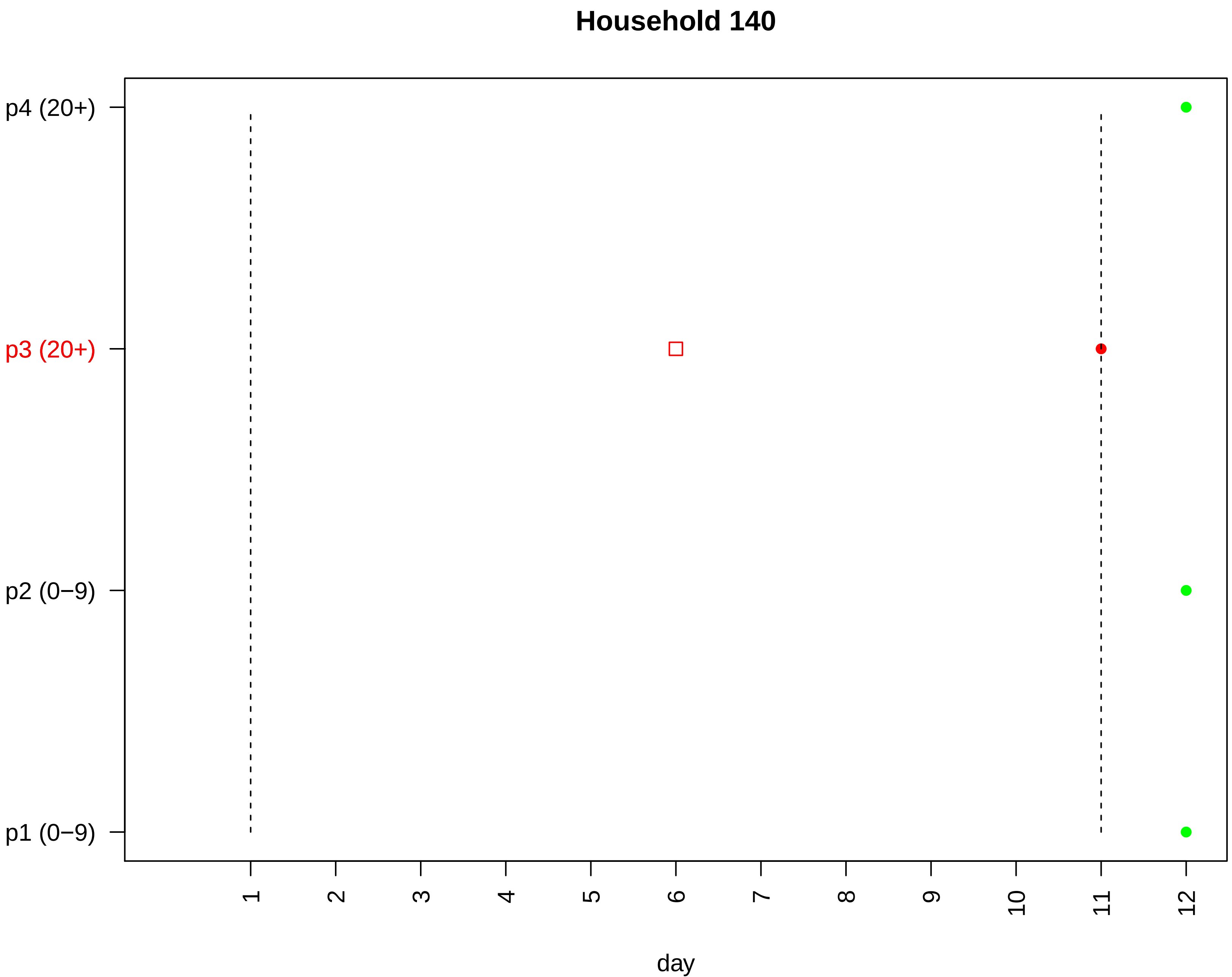
Household 140
p4 (20+)
p3 (20+)
p2 (0−9)
p1 (0−9)
1
2
3
4
5
6
7
8
9
10
11
12
day

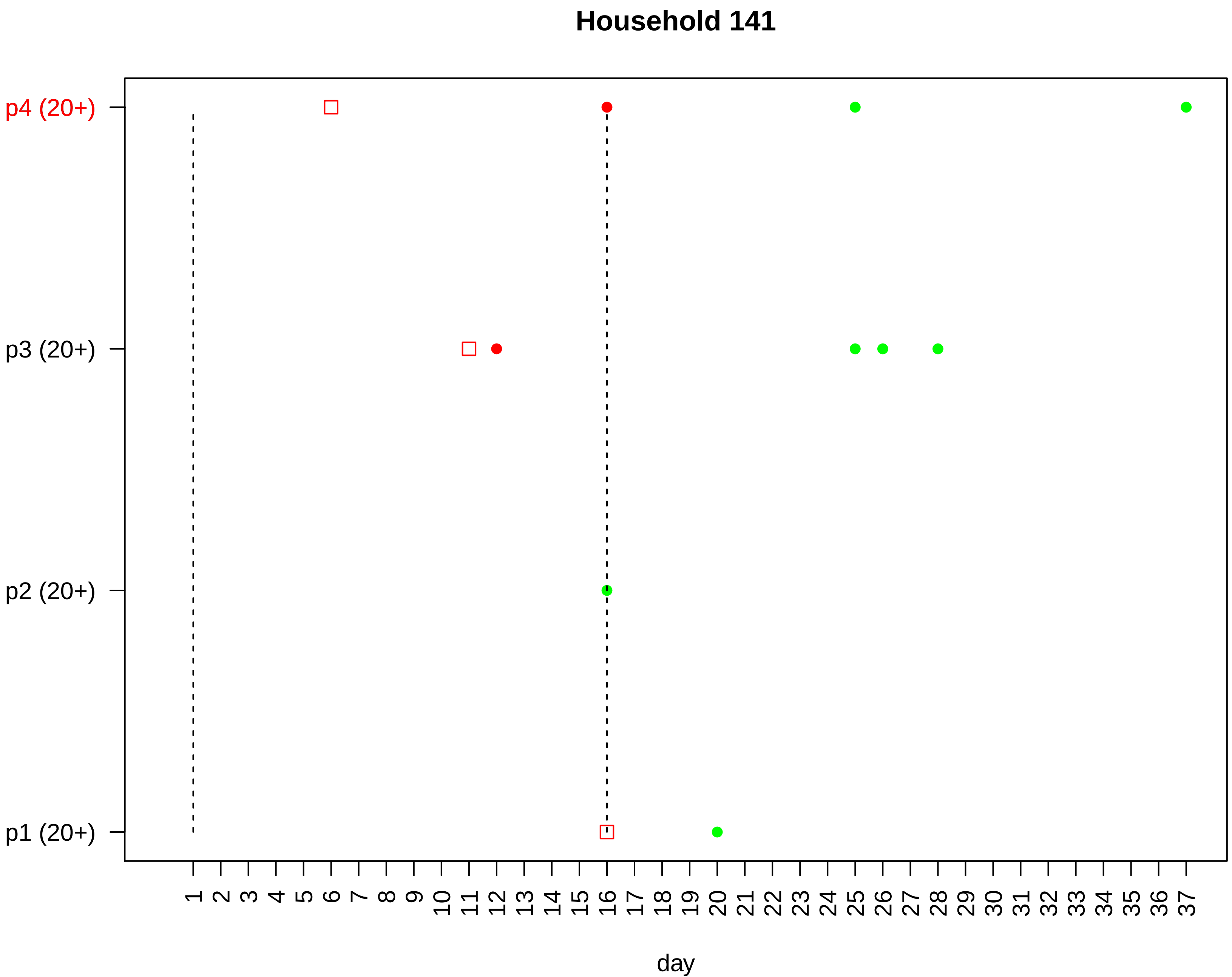
Household 141
p4 (20+)
p3 (20+)
p2 (20+)
p1 (20+)
day

### Household 142

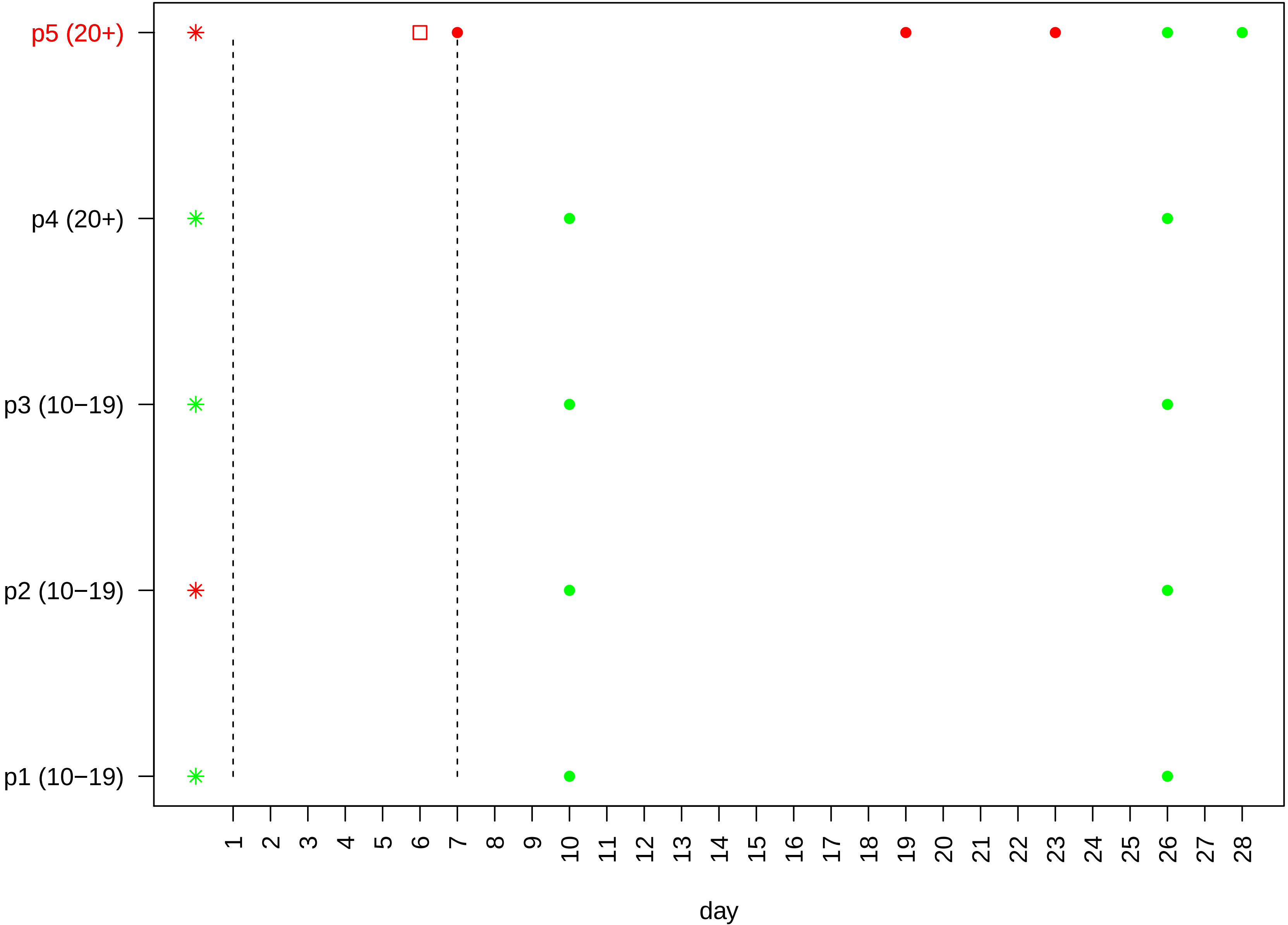

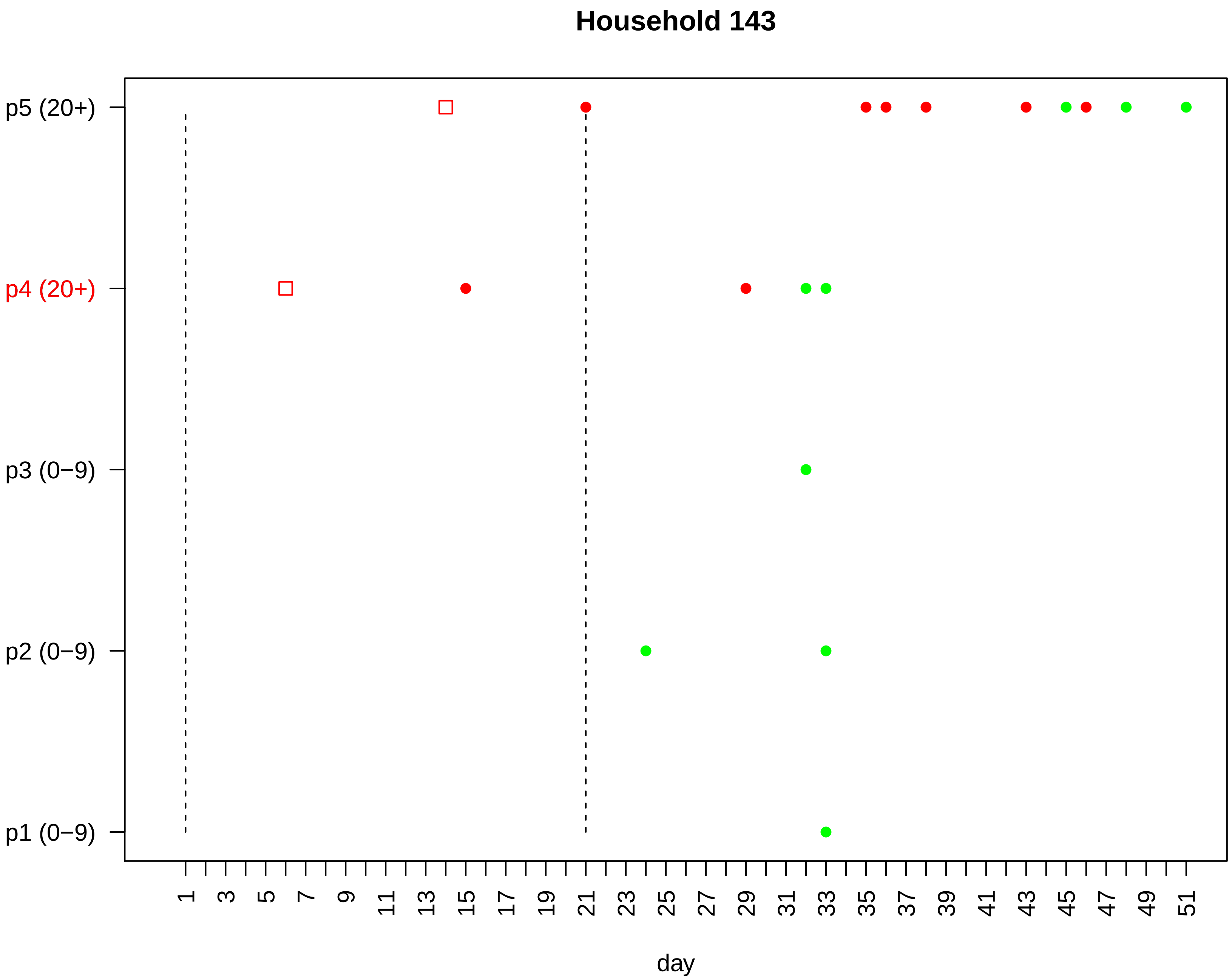

Household 143
p5 (20+)
p4 (20+)
p3 (0−9)
p2 (0−9)
p1 (0−9)
1
3
5
7
9
11
13
15
17
19
21
23
25
27
29
31
33
35
37
39
41
43
45
47
49
51
day

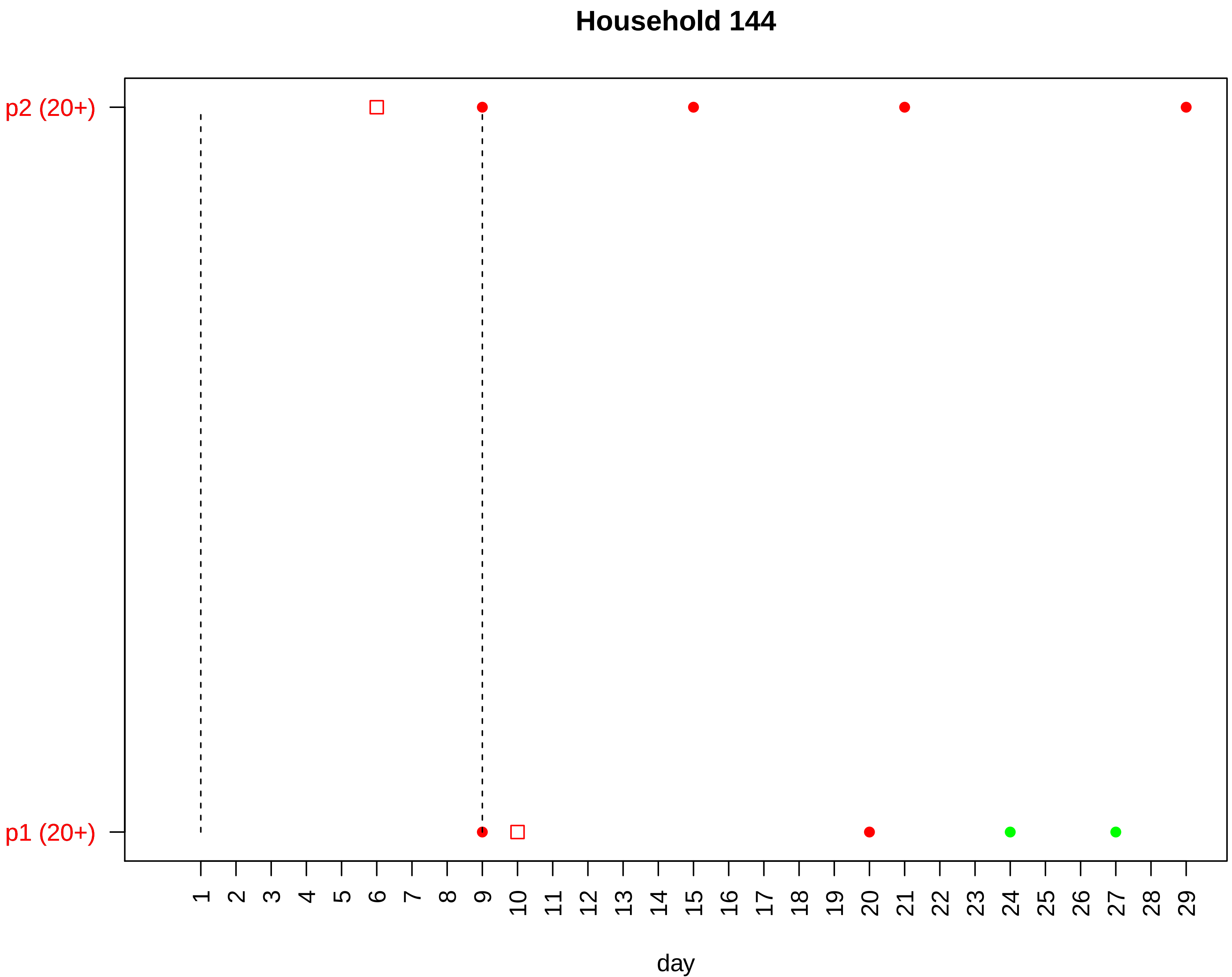

Household 144
p2 (20+)
p1 (20+)
day

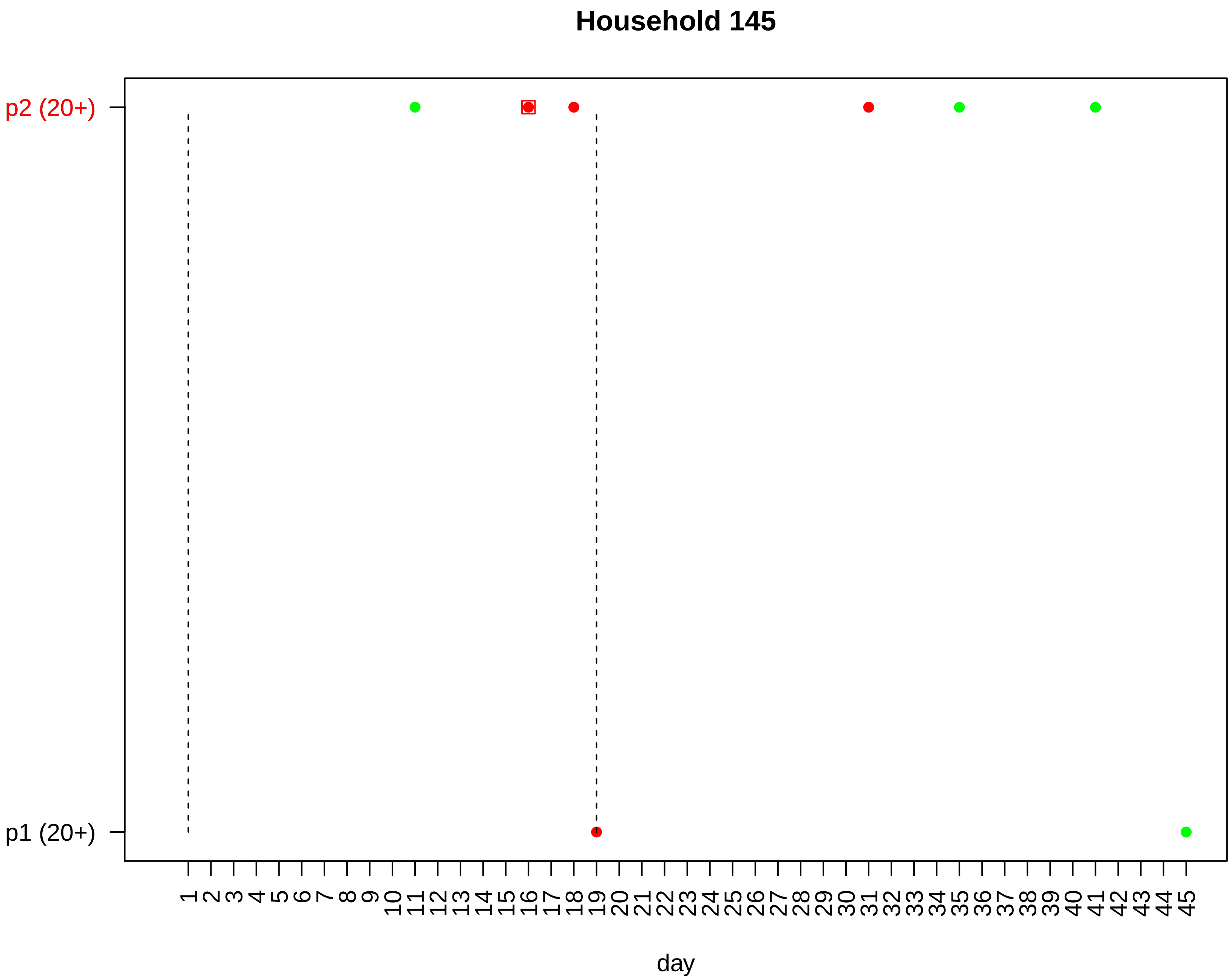
Household 145
p2 (20+)
p1 (20+)
1 2 3 4 5 6 7 8 9 10 11 12 13 14 15 16 17 18 19 20 21 22 23 24 25 26 27 28 29 30 31 32 33 34 35 36 37 38 39 40 41 42 43 44 45
day

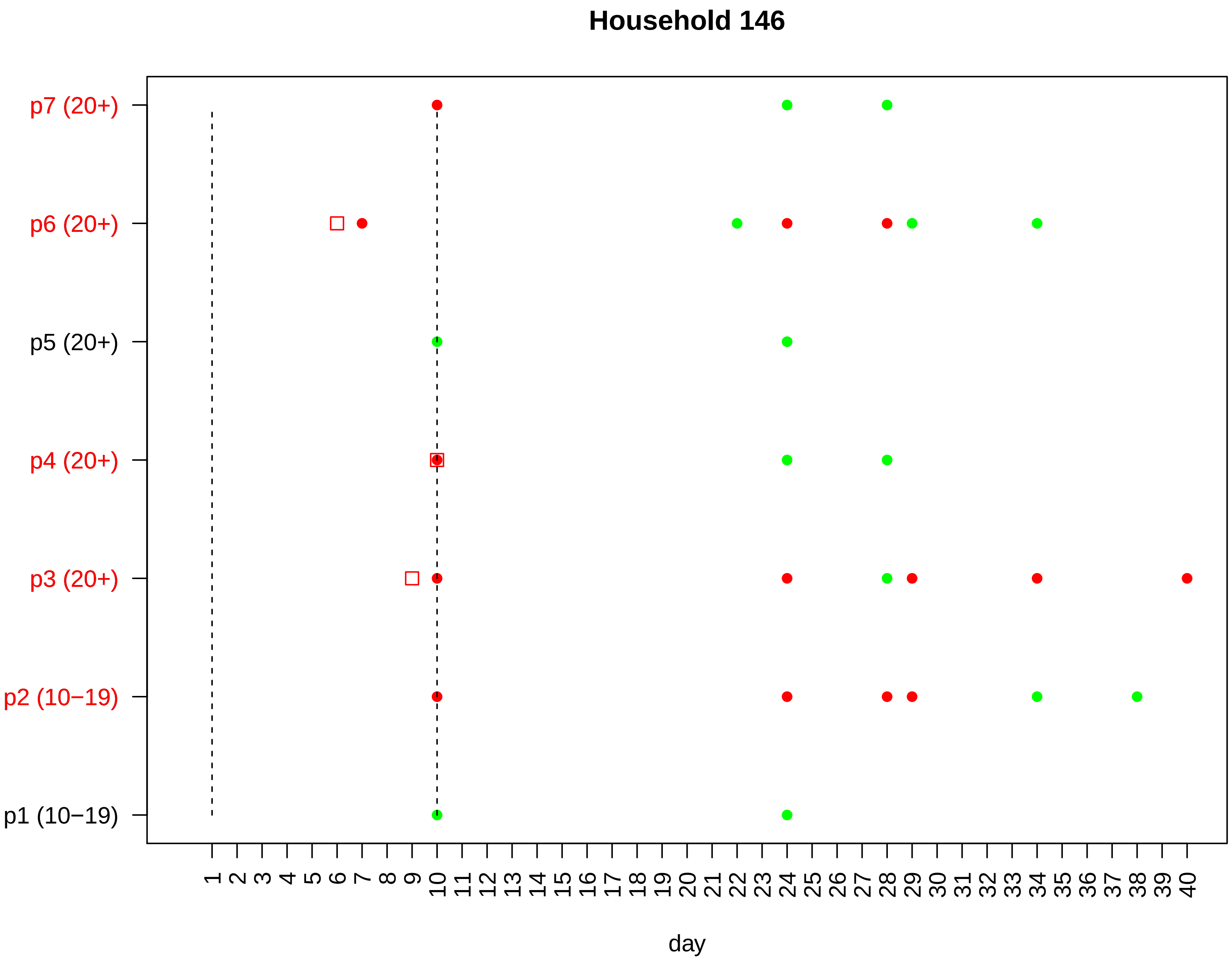

Household 146
p7 (20+)
p6 (20+)
p5 (20+)
p4 (20+)
p3 (20+)
p2 (10−19)
p1 (10−19)
1 2 3 4 5 6 7 8 9 10 11 12 13 14 15 16 17 18 19 20 21 22 23 24 25 26 27 28 29 30 31 32 33 34 35 36 37 38 39 40
day

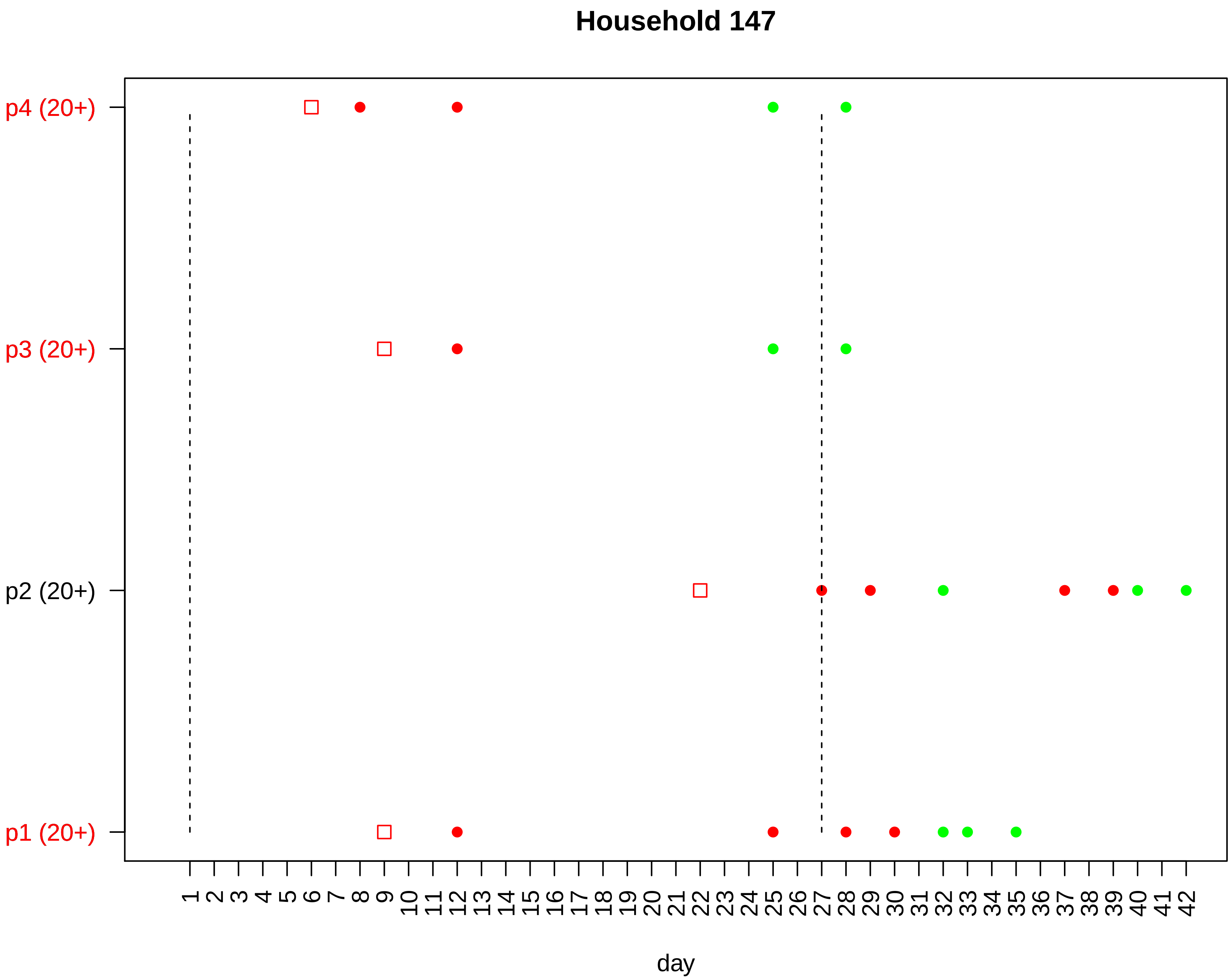

Household 147
p4 (20+)
p3 (20+)
p2 (20+)
p1 (20+)
1 2 3 4 5 6 7 8 9 10 11 12 13 14 15 16 17 18 19 20 21 22 23 24 25 26 27 28 29 30 31 32 33 34 35 36 37 38 39 40 41 42
day

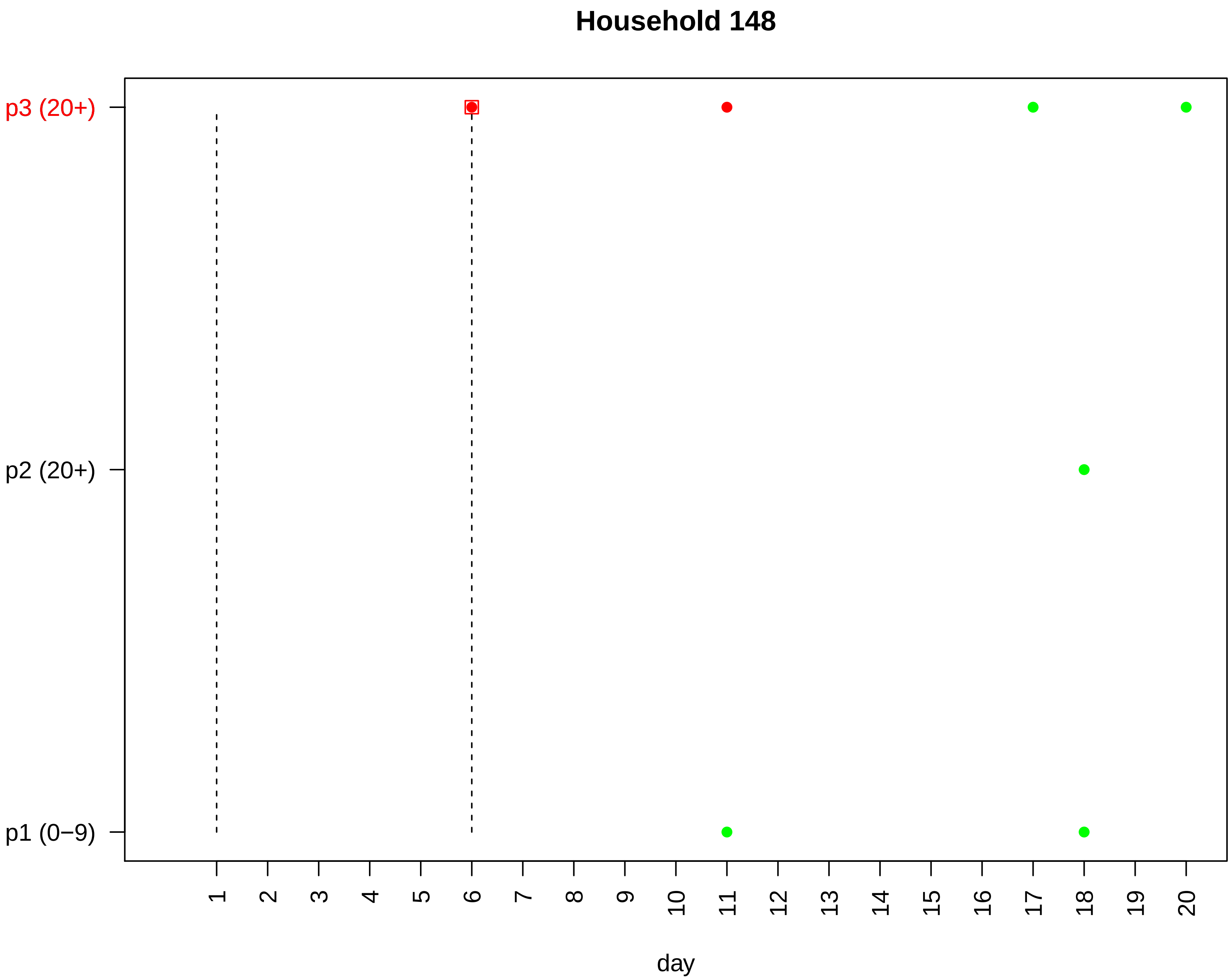
Household 148
p3 (20+)
p2 (20+)
p1 (0−9)
day

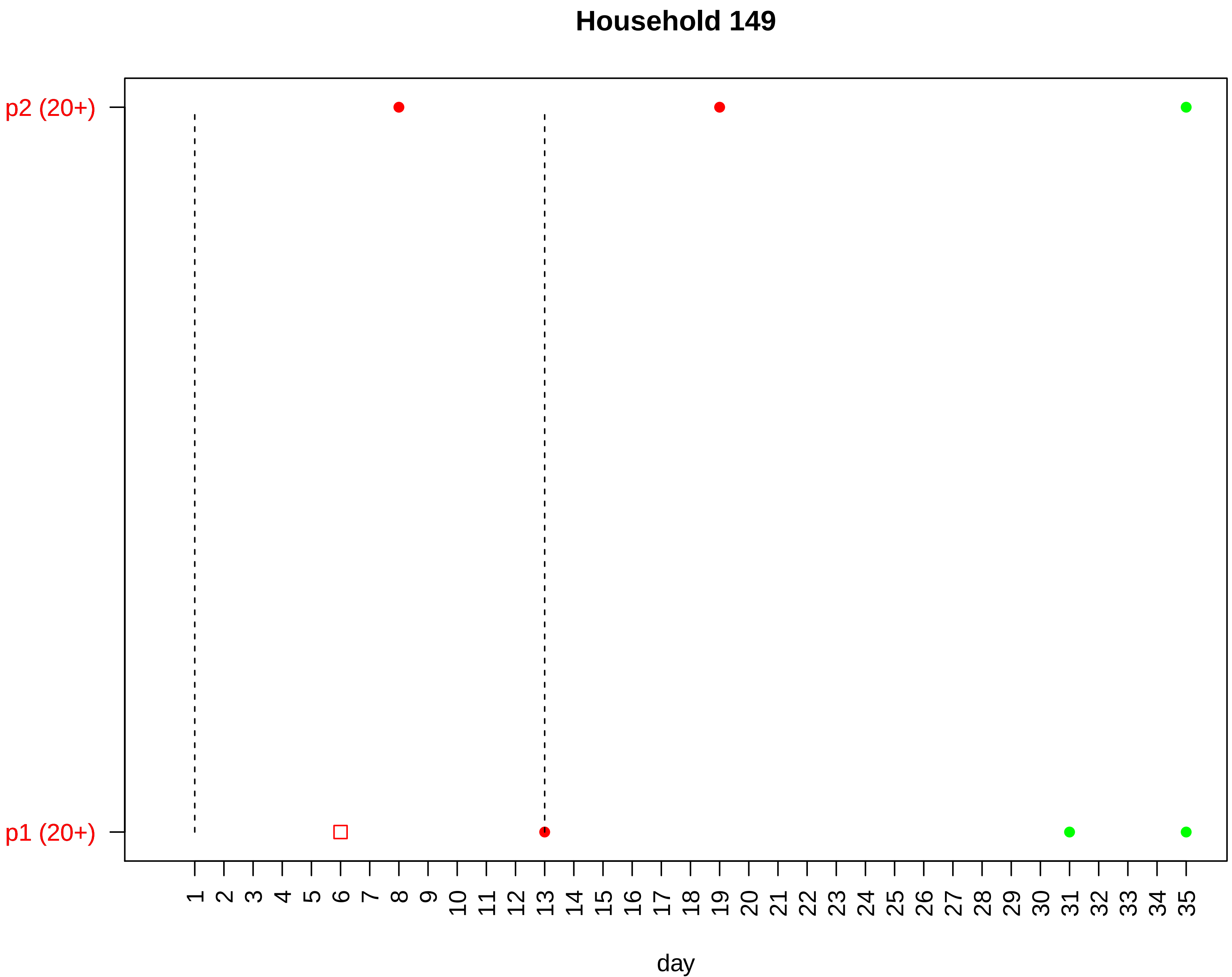

Household 149
p2 (20+)
p1 (20+)
1 2 3 4 5 6 7 8 9 10 11 12 13 14 15 16 17 18 19 20 21 22 23 24 25 26 27 28 29 30 31 32 33 34 35
day

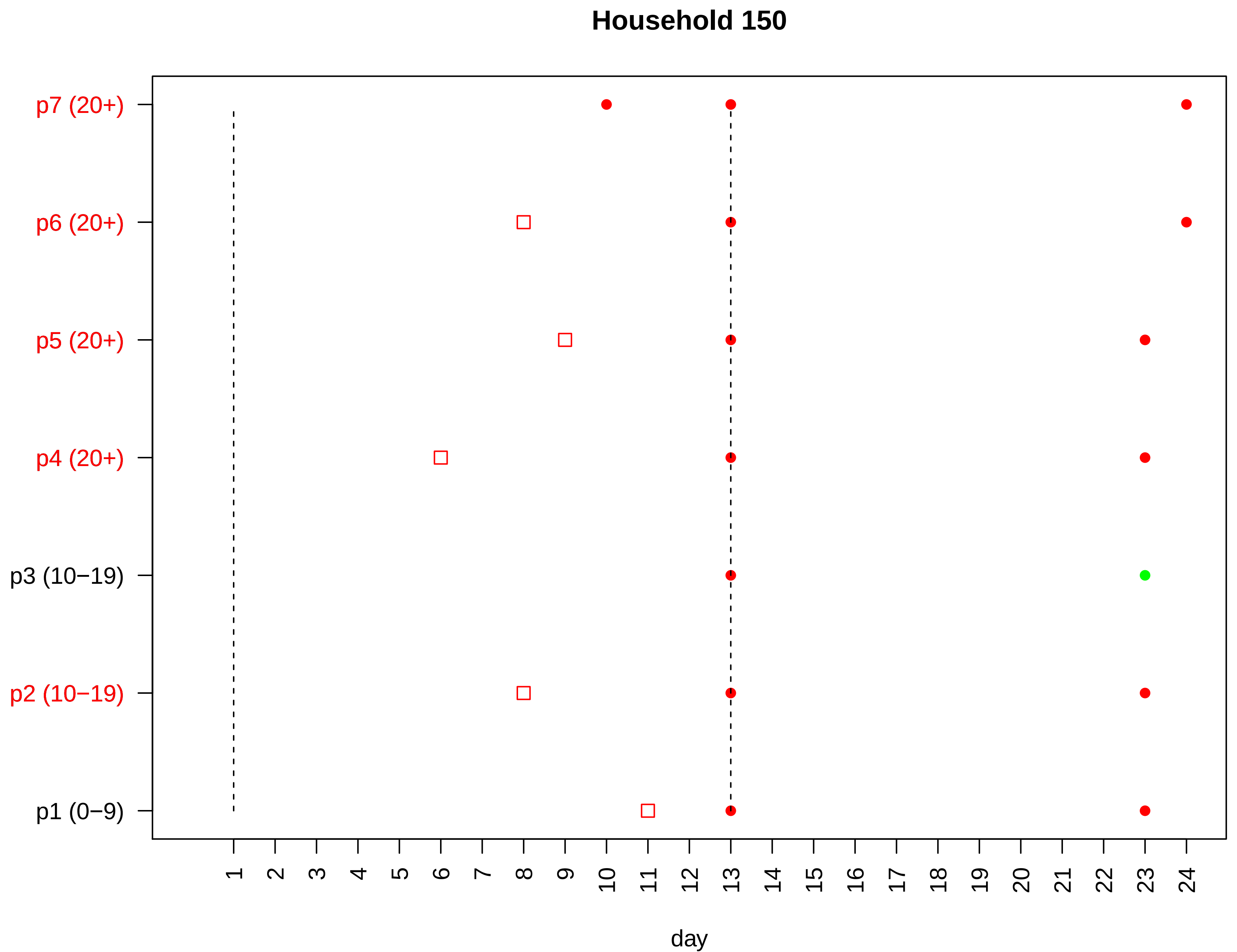
Household 150
p7 (20+)
p6 (20+)
p5 (20+)
p4 (20+)
p3 (10−19)
p2 (10−19)
p1 (0−9)
1 2 3 4 5 6 7 8 9 10 11 12 13 14 15 16 17 18 19 20 21 22 23 24
day

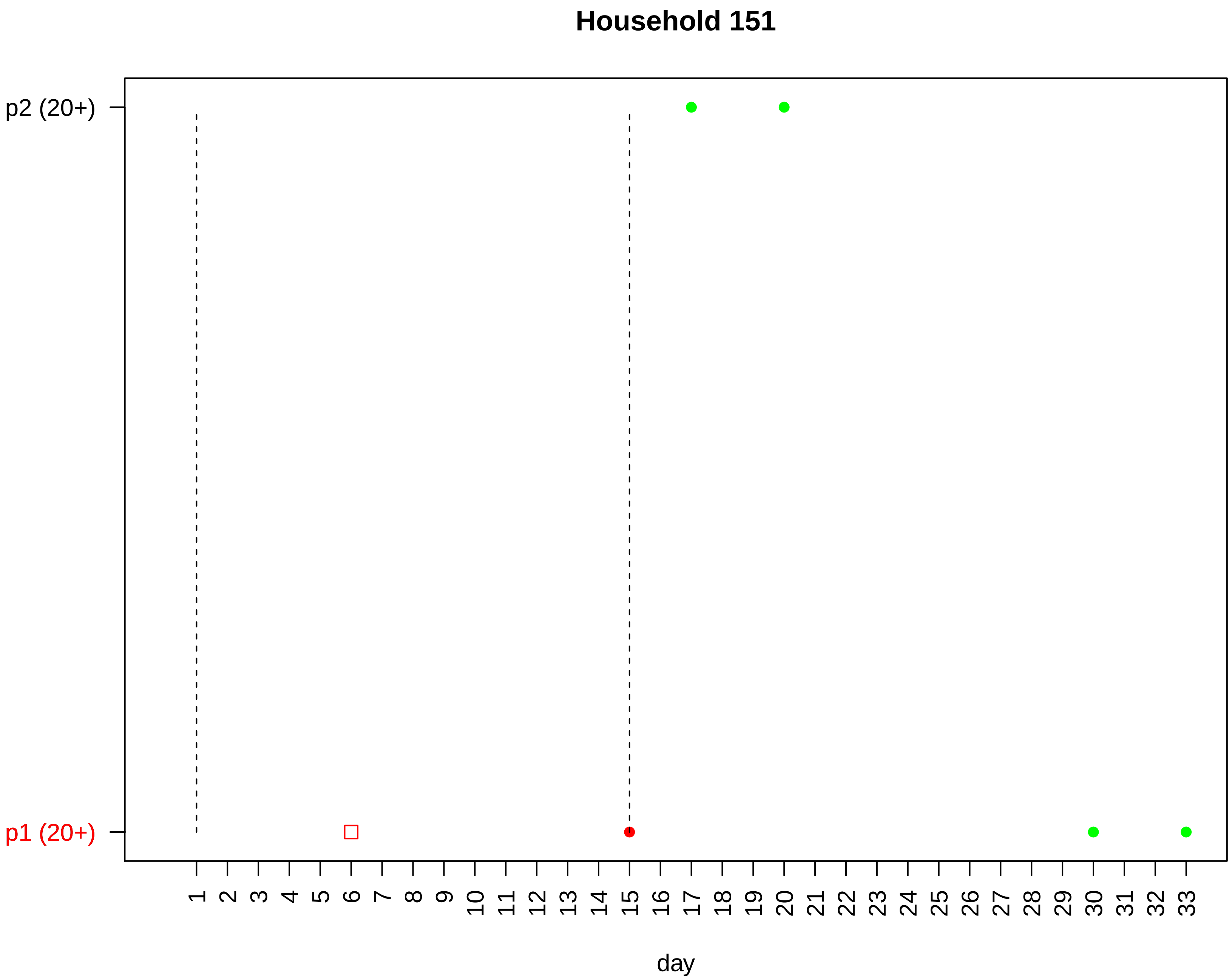

Household 151
p2 (20+)
p1 (20+)
day

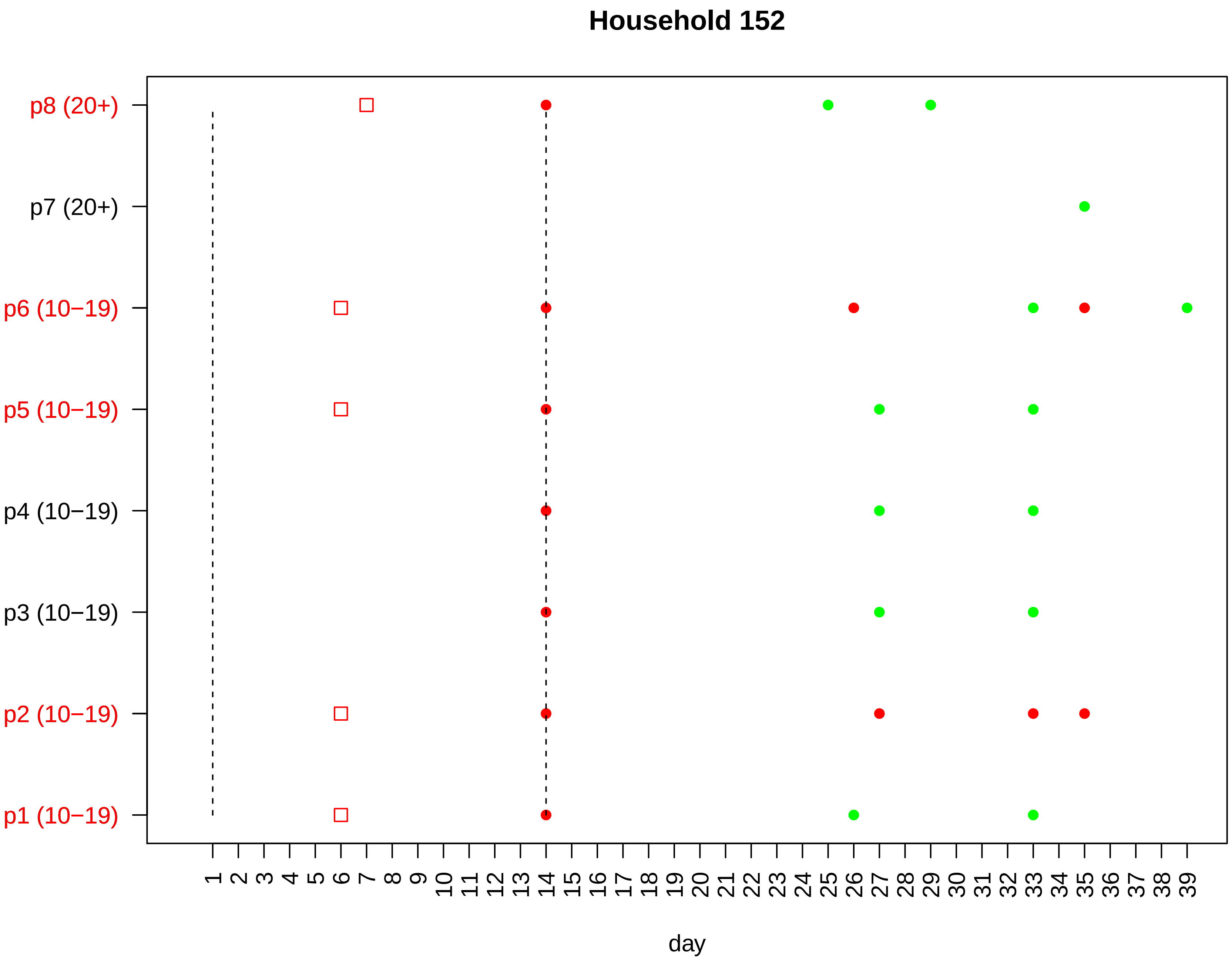

Household 152
p8 (20+)
p7 (20+)
p6 (10−19)
p5 (10−19)
p4 (10−19)
p3 (10−19)
p2 (10−19)
p1 (10−19)
1 2 3 4 5 6 7 8 9 10 11 12 13 14 15 16 17 18 19 20 21 22 23 24 25 26 27 28 29 30 31 32 33 34 35 36 37 38 39
day

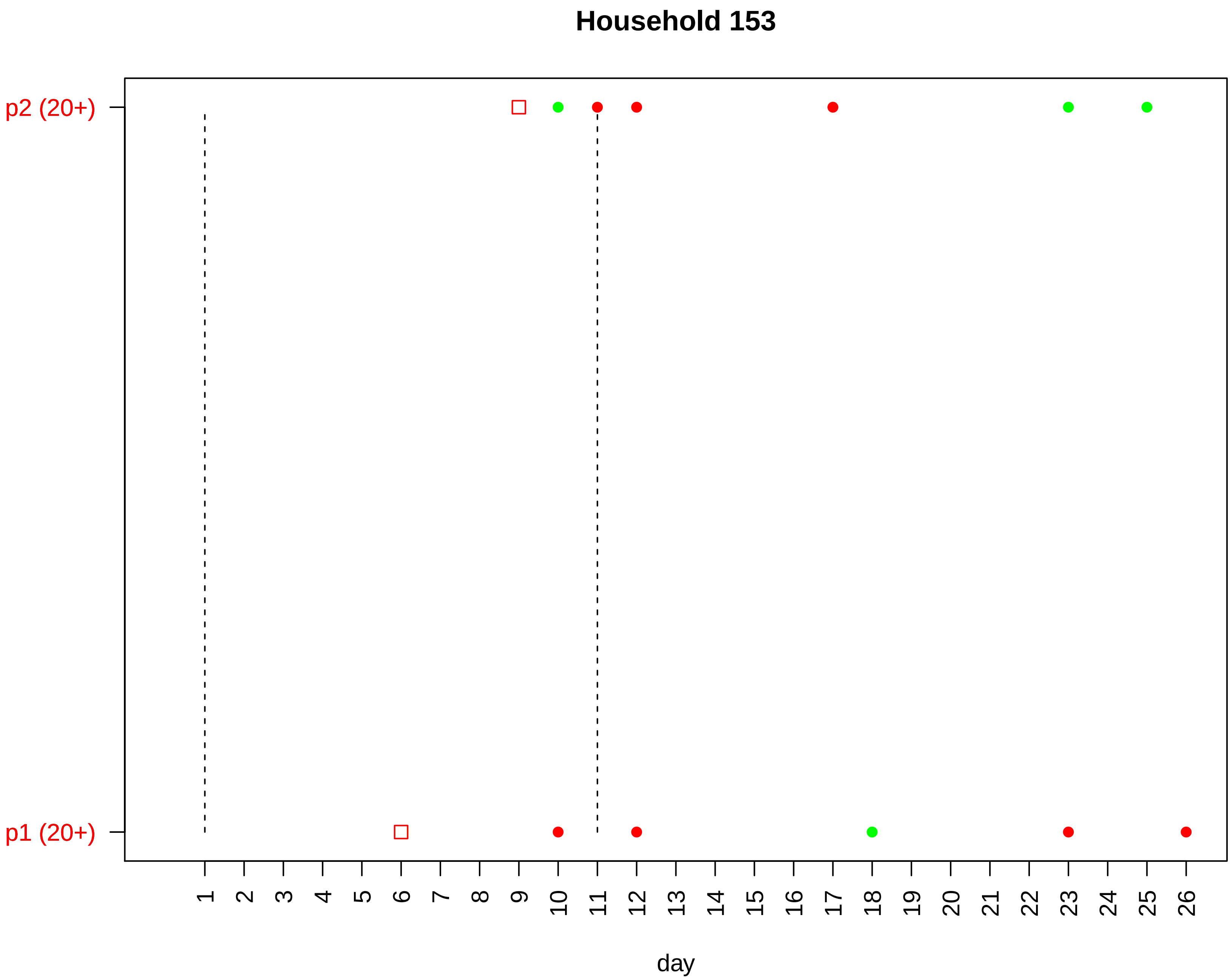

Household 153
p2 (20+)
p1 (20+)
day

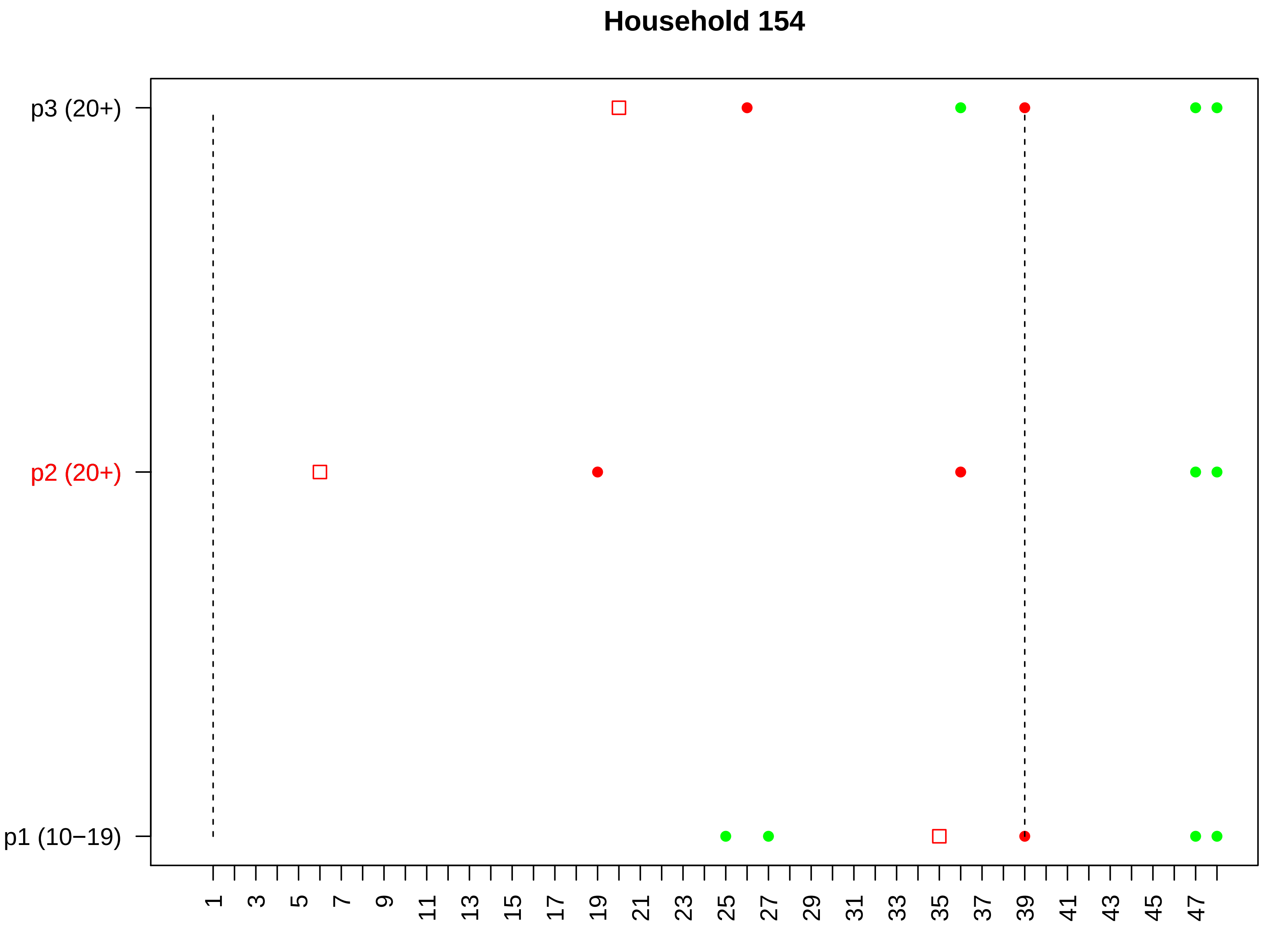
Household 154
p3 (20+)
p2 (20+)
p1 (10−19)
1
3
5
7
9
11
13
15
17
19
21
23
25
27
29
31
33
35
37
39
41
43
45
47
day

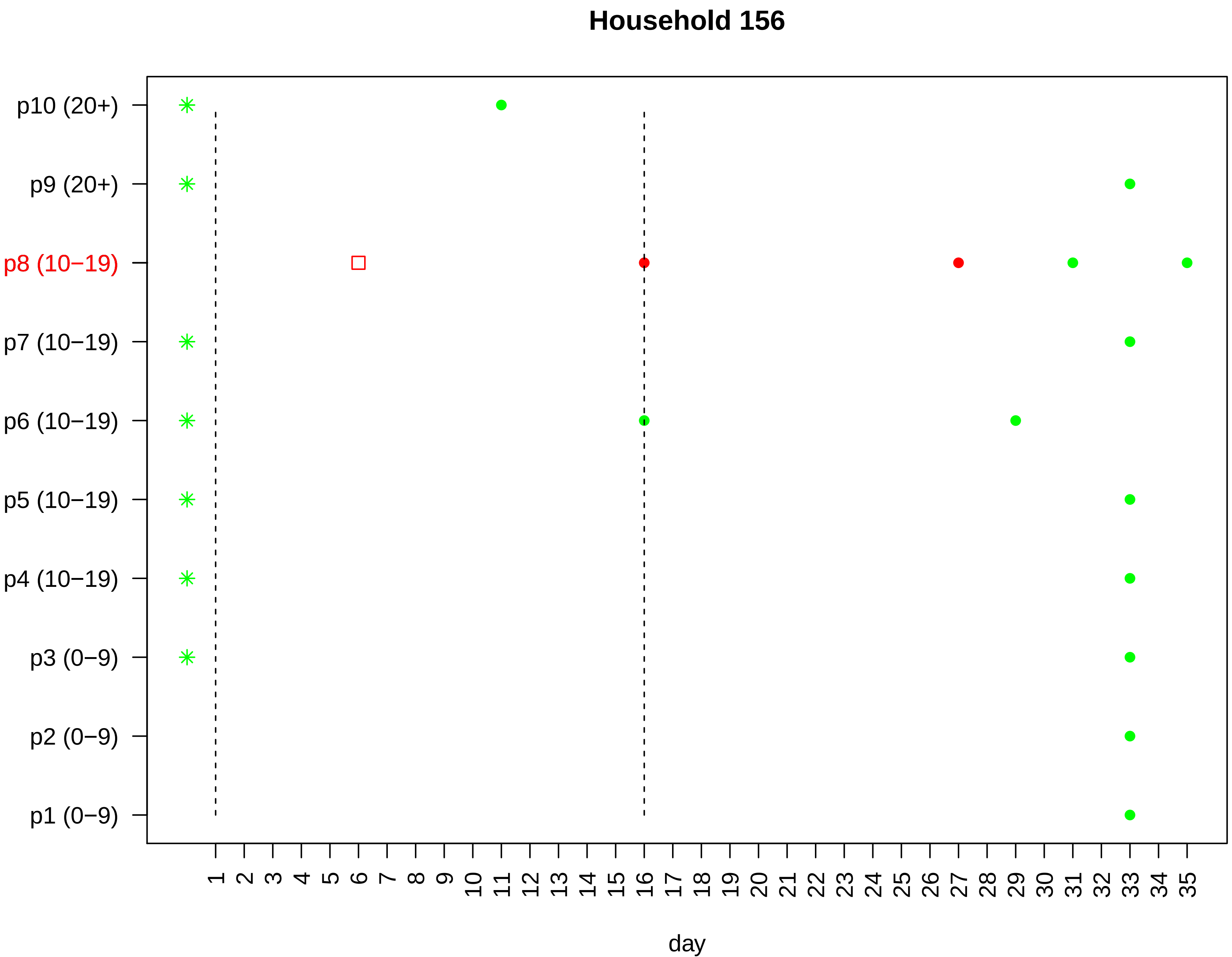

Household 156
p10 (20+)
p9 (20+)
p8 (10−19)
p7 (10−19)
p6 (10−19)
p5 (10−19)
p4 (10−19)
p3 (0−9)
p2 (0−9)
p1 (0−9)
1 2 3 4 5 6 7 8 9 10 11 12 13 14 15 16 17 18 19 20 21 22 23 24 25 26 27 28 29 30 31 32 33 34 35
day

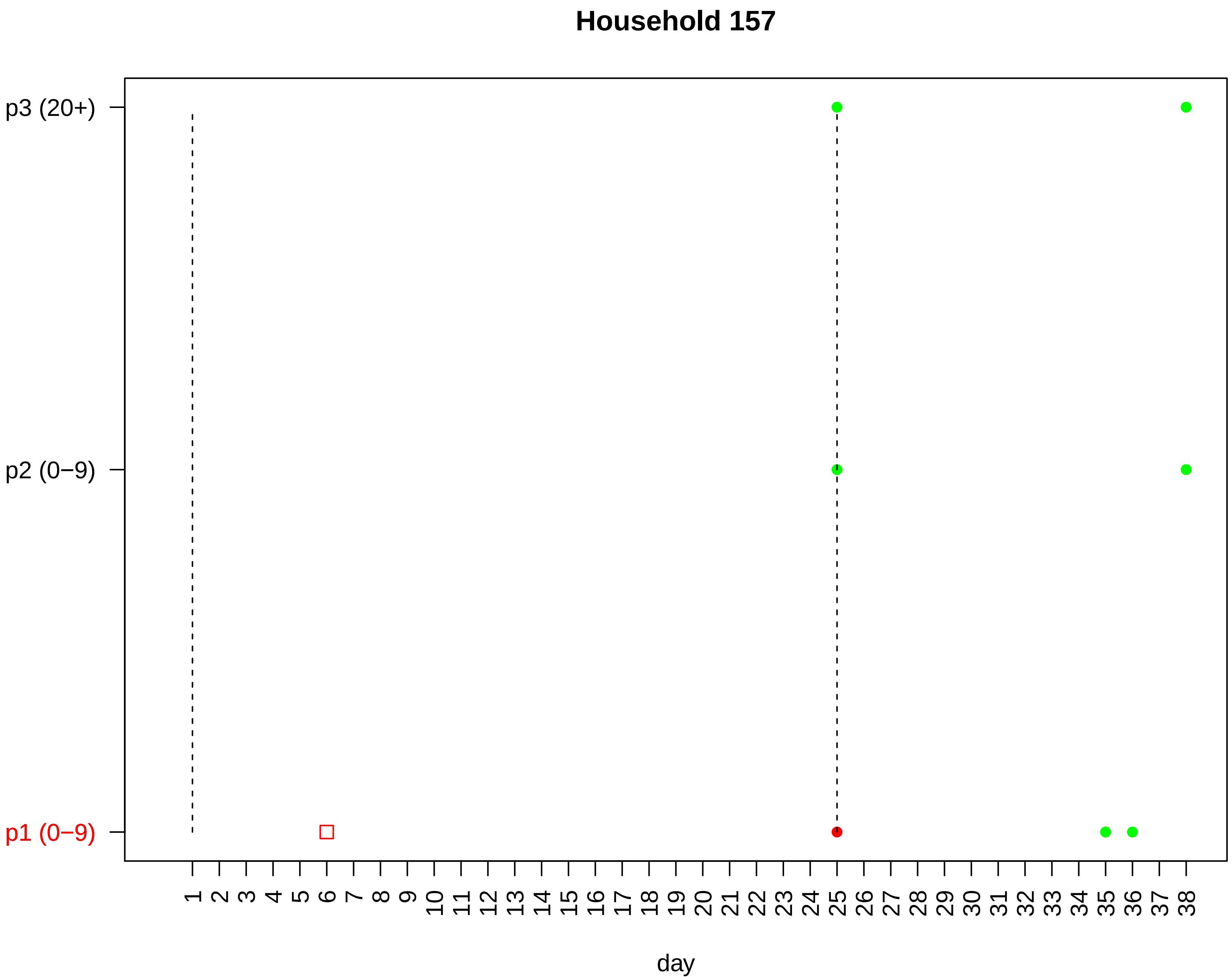
Household 157
p3 (20+)
p2 (0−9)
p1 (0−9)
1 2 3 4 5 6 7 8 9 10 11 12 13 14 15 16 17 18 19 20 21 22 23 24 25 26 27 28 29 30 31 32 33 34 35 36 37 38
day

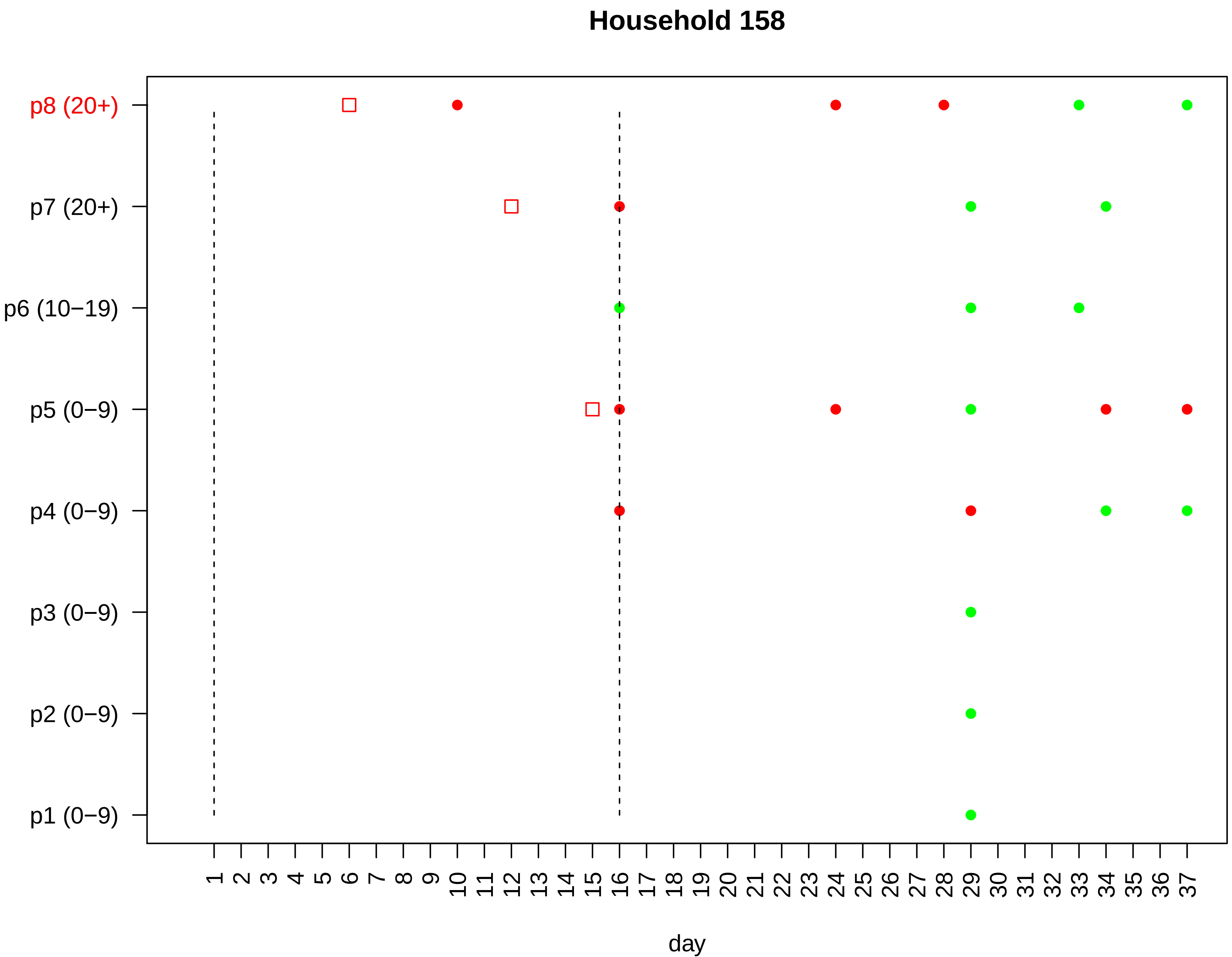
Household 158
p8 (20+)
p7 (20+)
p6 (10−19)
p5 (0−9)
p4 (0−9)
p3 (0−9)
p2 (0−9)
p1 (0−9)
1 2 3 4 5 6 7 8 9 10 11 12 13 14 15 16 17 18 19 20 21 22 23 24 25 26 27 28 29 30 31 32 33 34 35 36 37
day

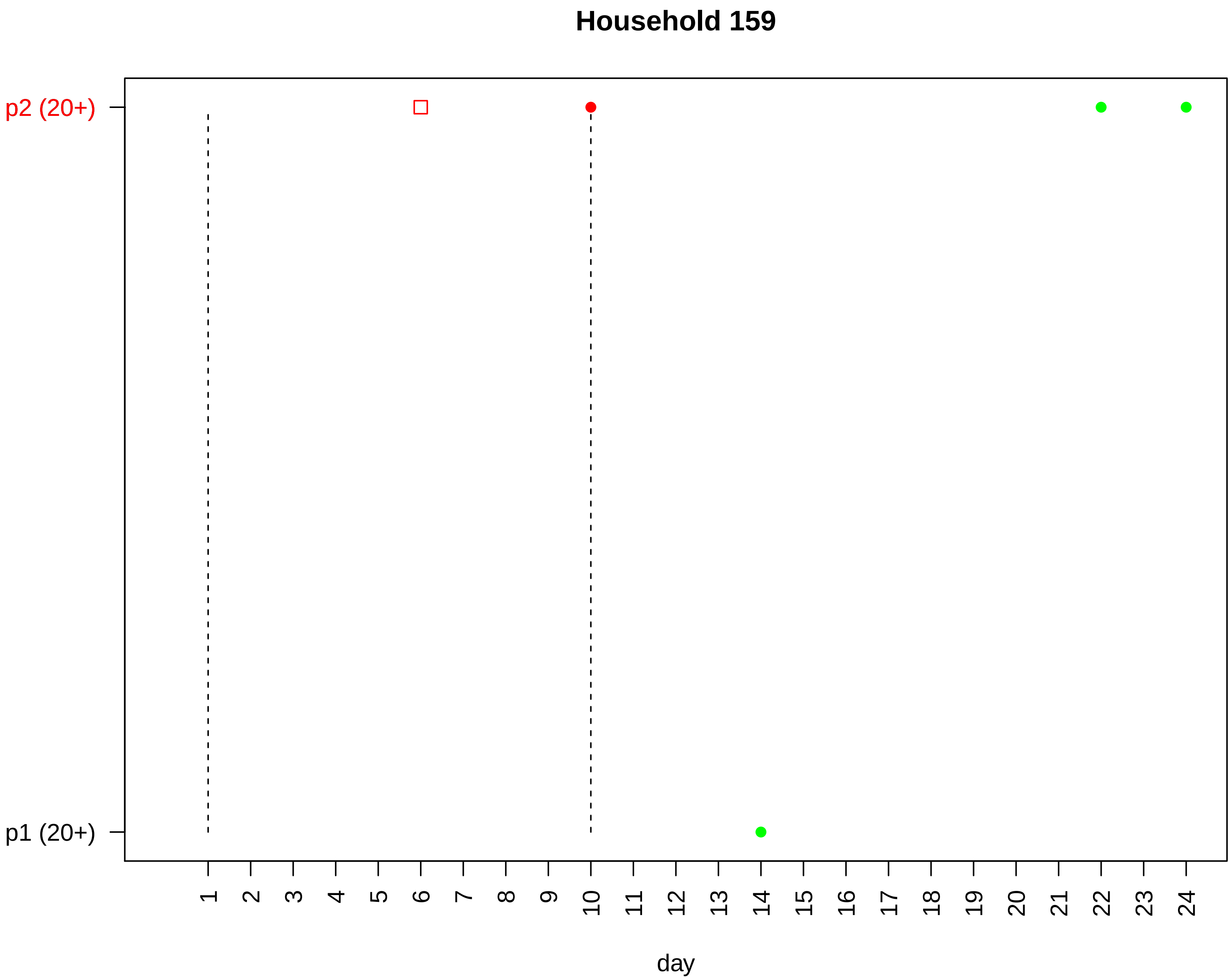

Household 159
p2 (20+)
p1 (20+)
day

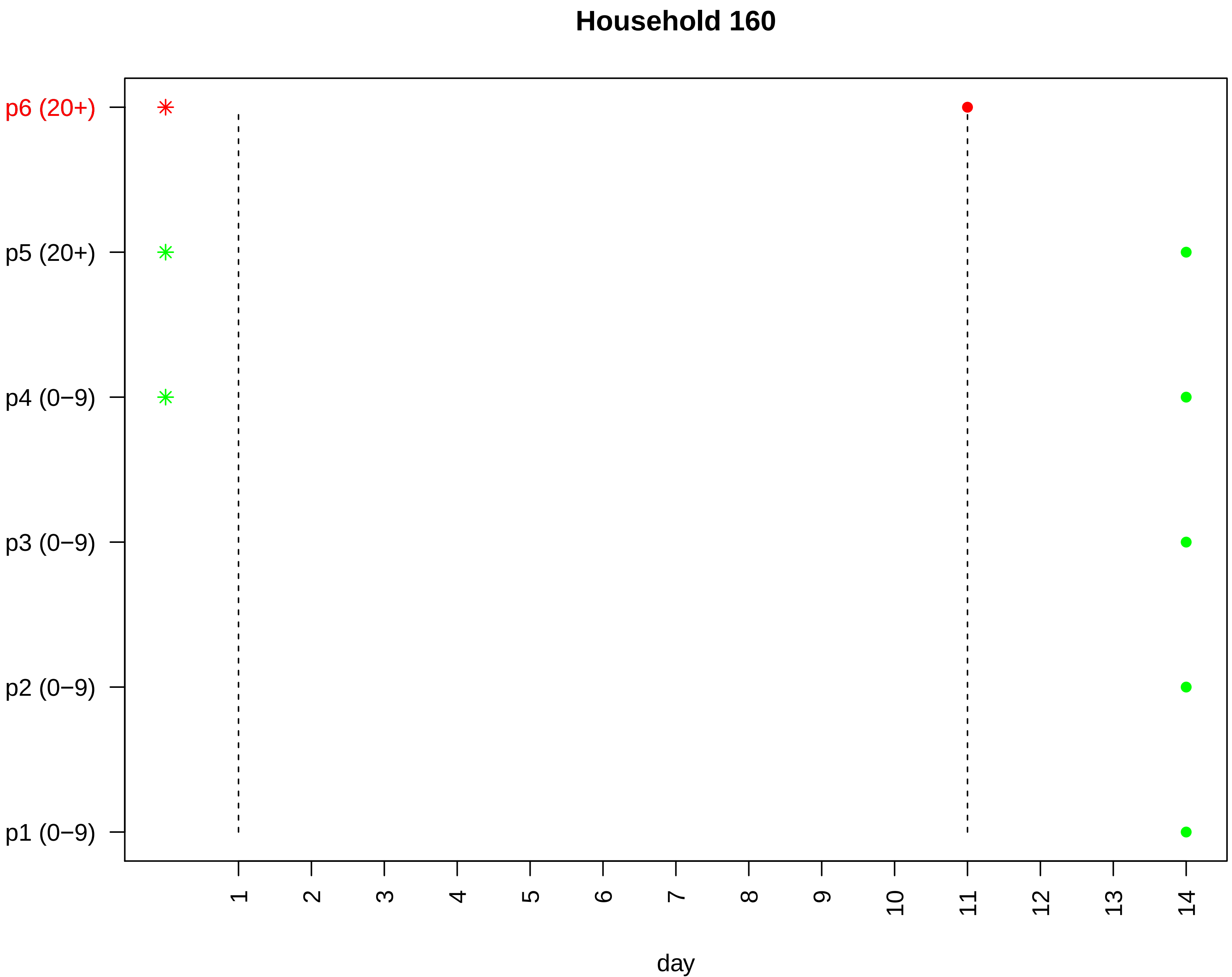

Household 160
p6 (20+)
p5 (20+)
p4 (0−9)
p3 (0−9)
p2 (0−9)
p1 (0−9)
1
2
3
4
5
6
7
8
9
10
11
12
13
14
day

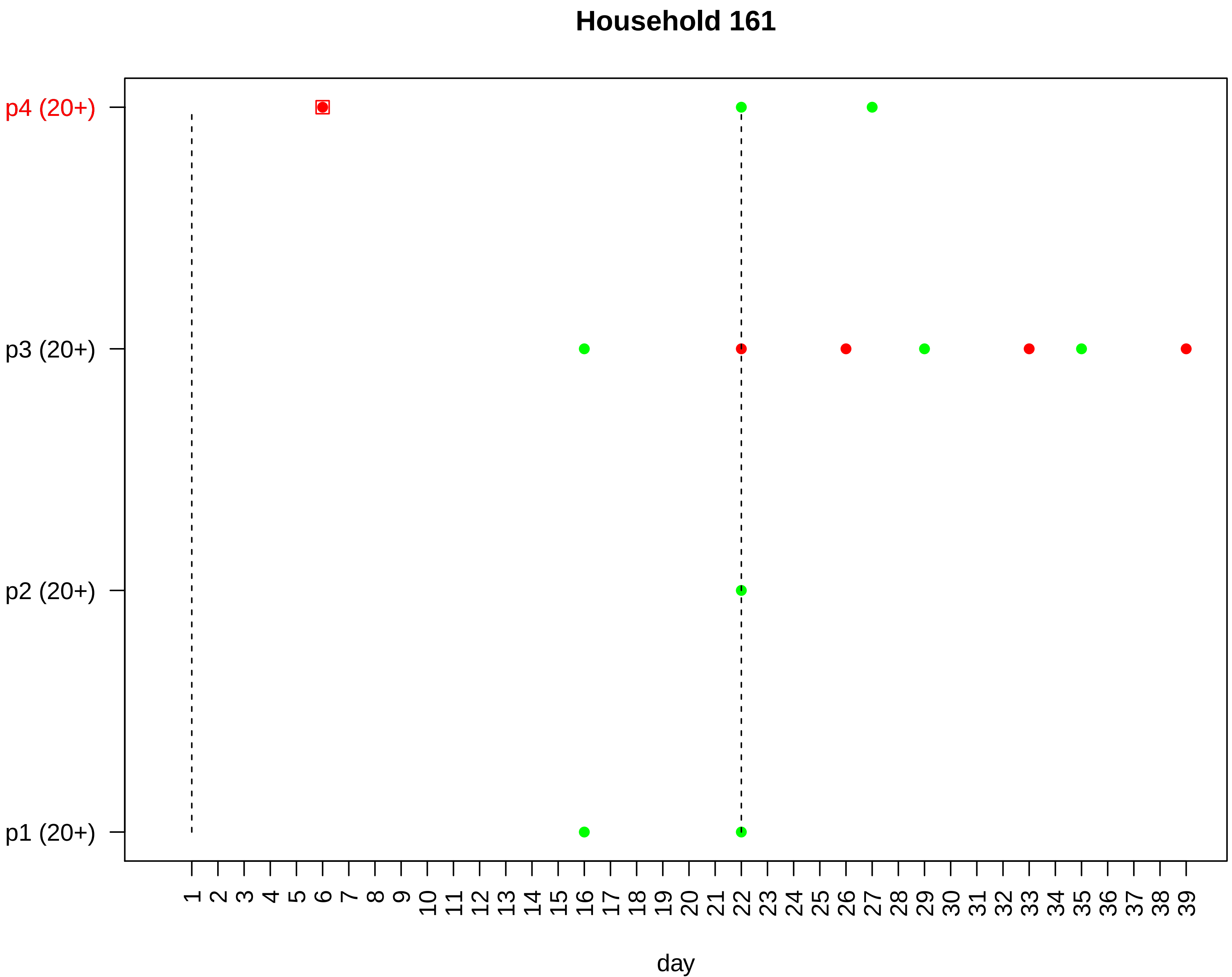

Household 161
p4 (20+)
p3 (20+)
p2 (20+)
p1 (20+)
day

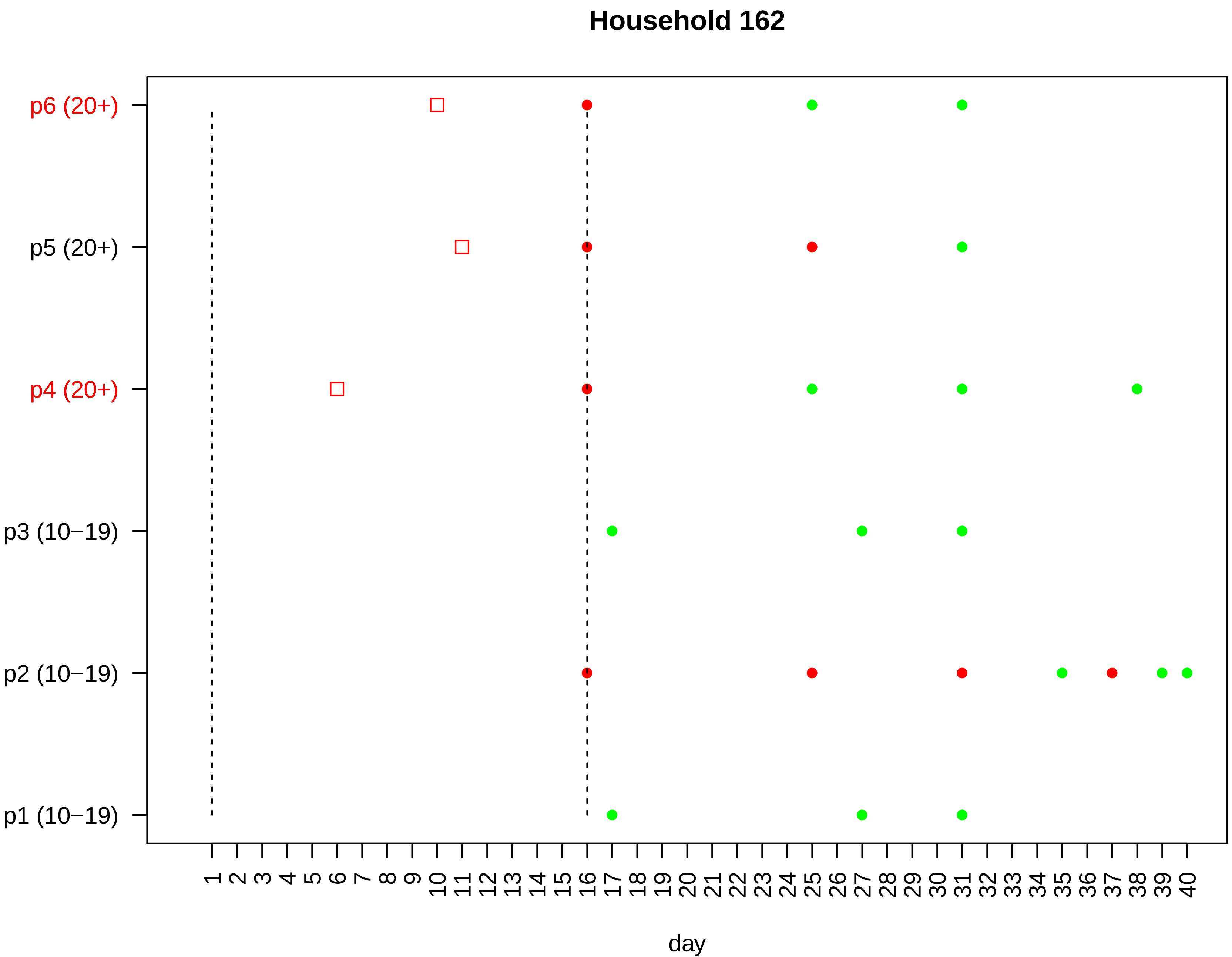
Household 162
p6 (20+)
p5 (20+)
p4 (20+)
p3 (10−19)
p2 (10−19)
p1 (10−19)
1 2 3 4 5 6 7 8 9 10 11 12 13 14 15 16 17 18 19 20 21 22 23 24 25 26 27 28 29 30 31 32 33 34 35 36 37 38 39 40
day

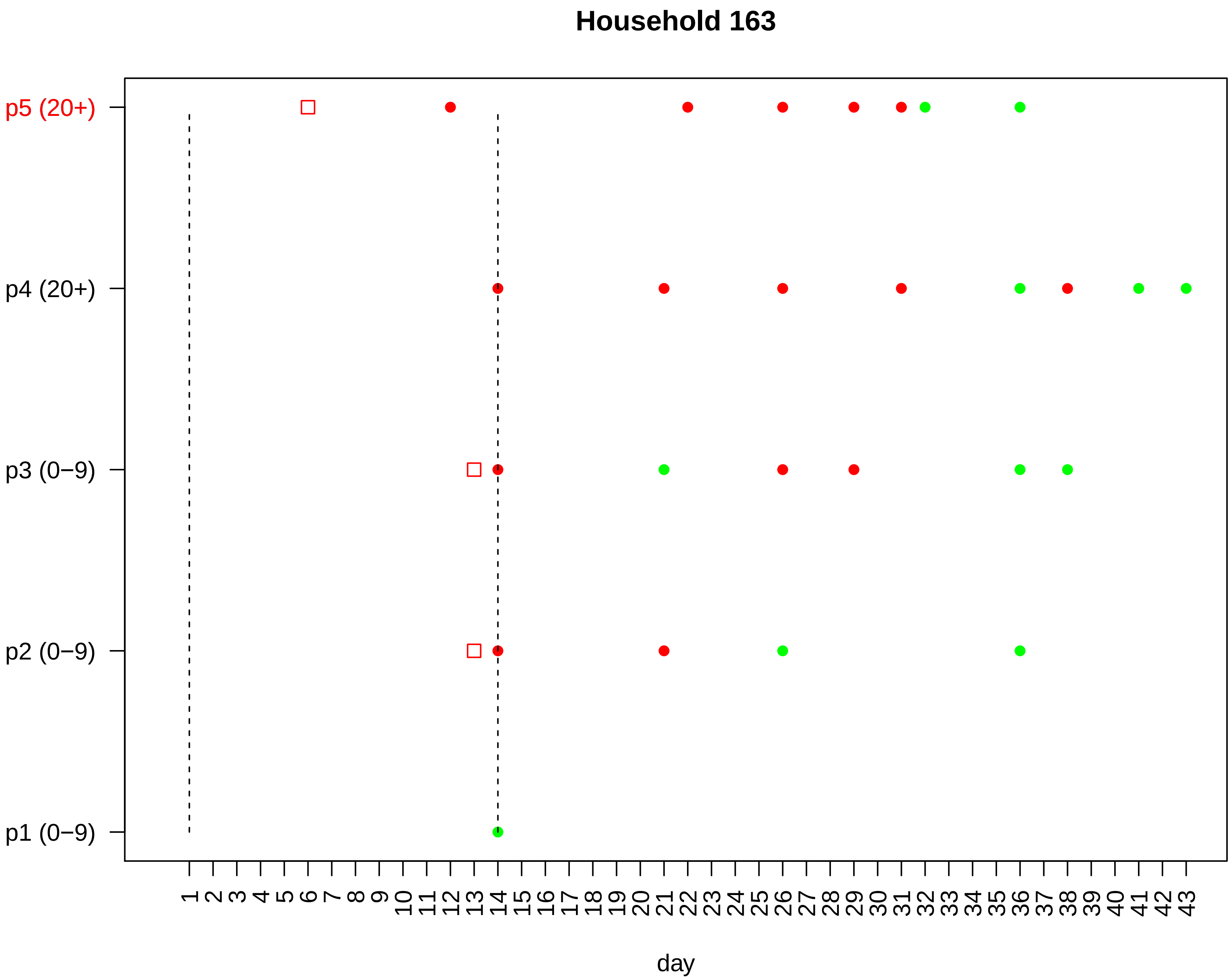

Household 163
p5 (20+)
p4 (20+)
p3 (0−9)
p2 (0−9)
p1 (0−9)
1 2 3 4 5 6 7 8 9 10 11 12 13 14 15 16 17 18 19 20 21 22 23 24 25 26 27 28 29 30 31 32 33 34 35 36 37 38 39 40 41 42 43
day

#### Household 164

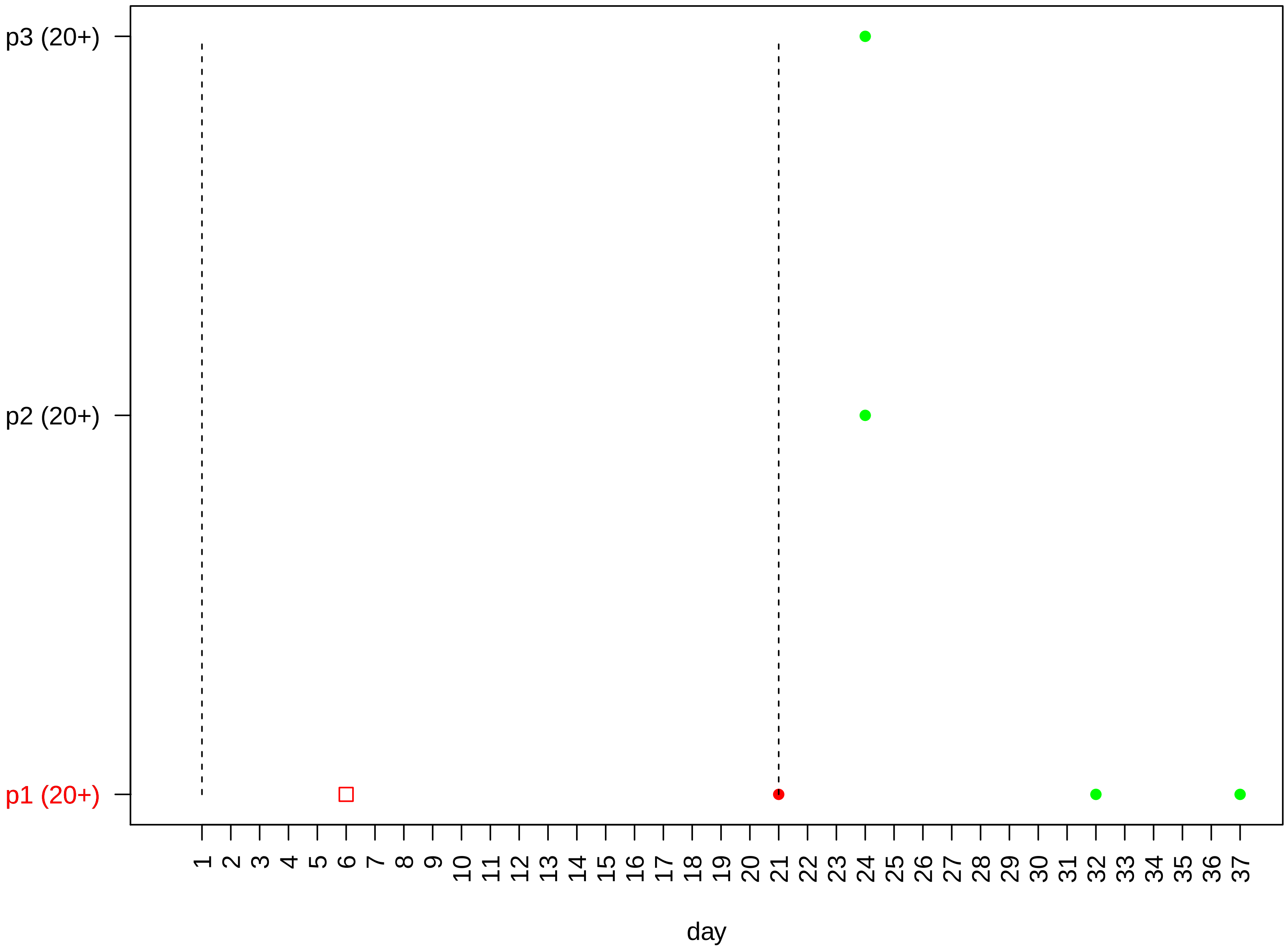

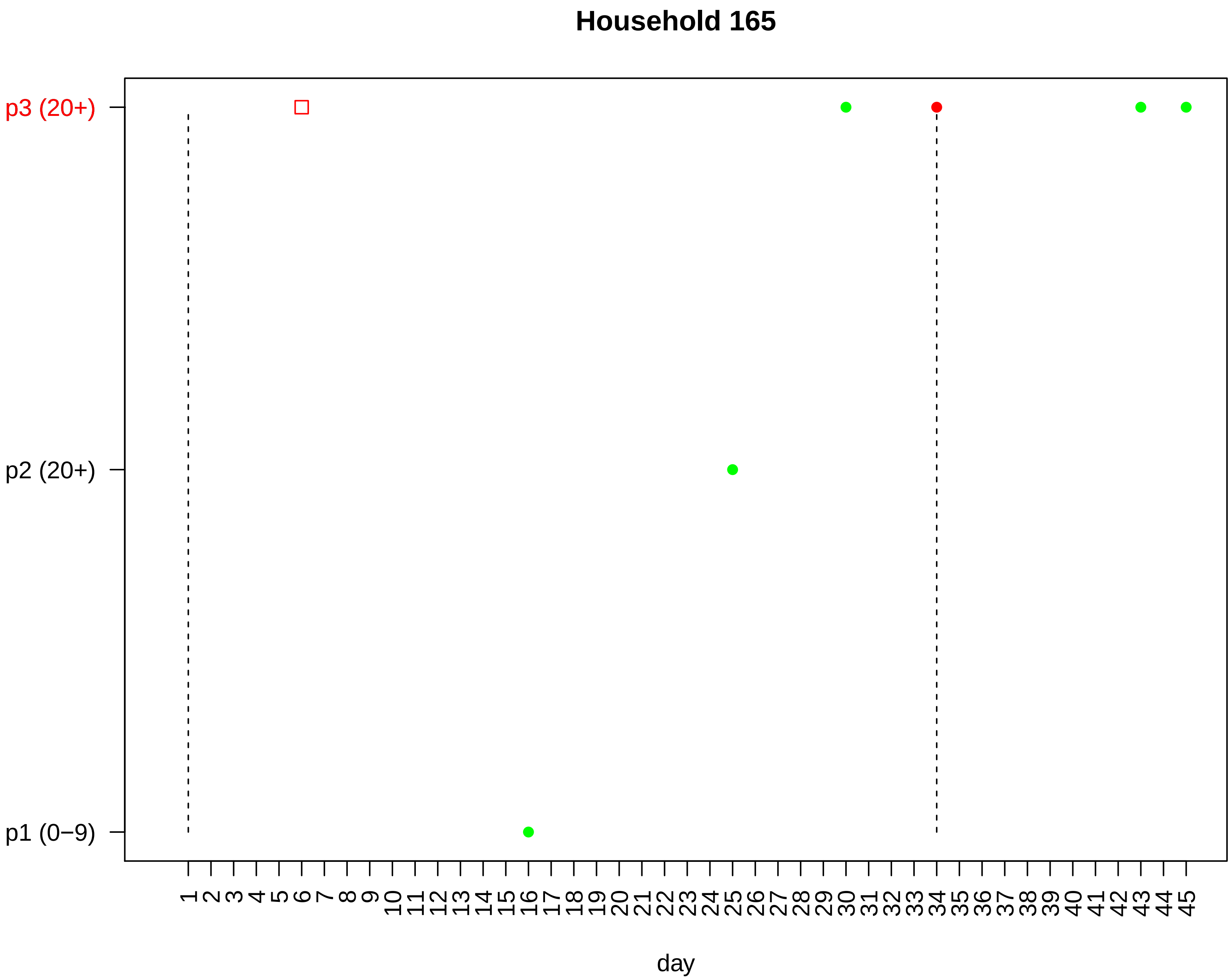

Household 165
p3 (20+)
p2 (20+)
p1 (0−9)
1 2 3 4 5 6 7 8 9 10 11 12 13 14 15 16 17 18 19 20 21 22 23 24 25 26 27 28 29 30 31 32 33 34 35 36 37 38 39 40 41 42 43 44 45
day

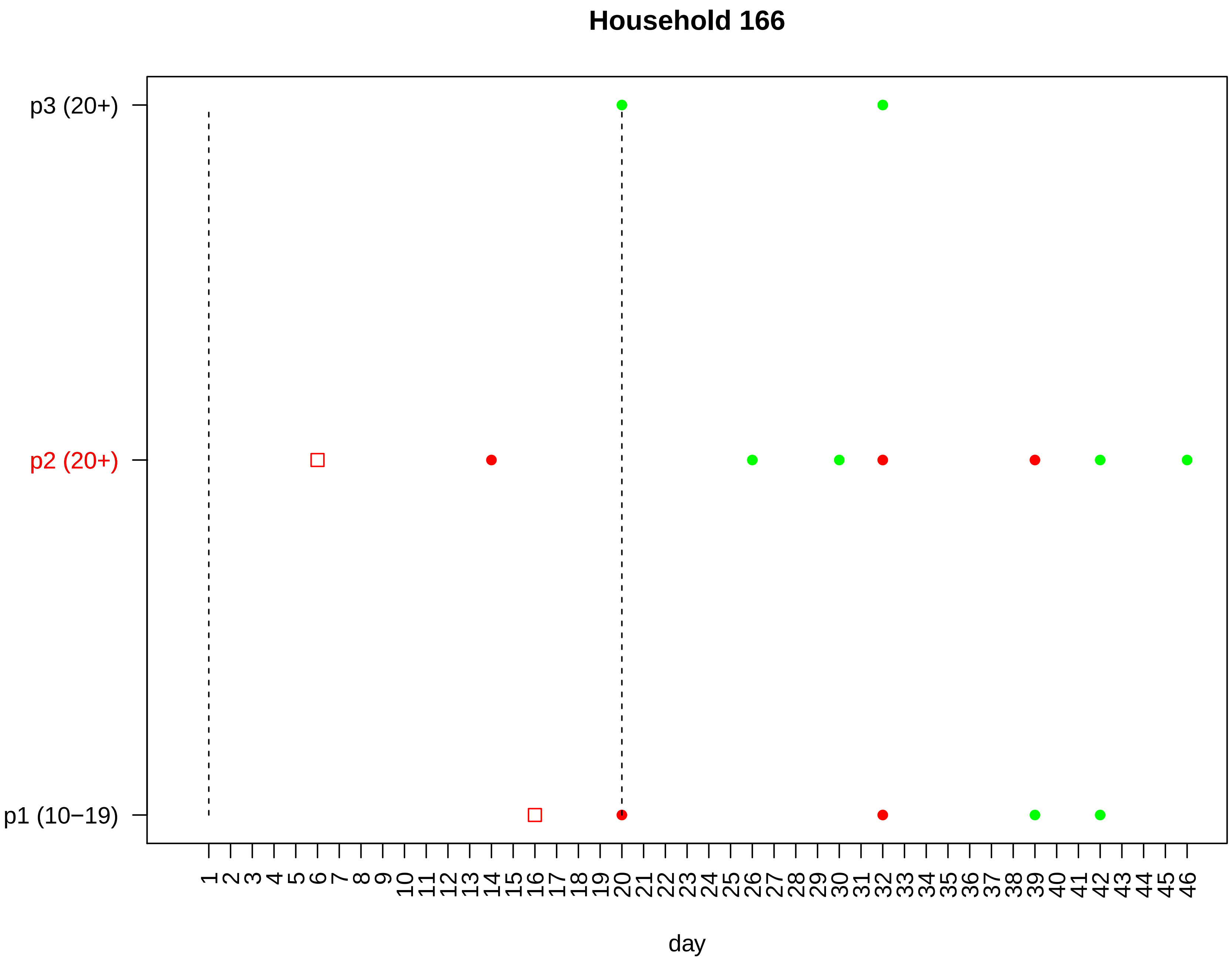

Household 166
p3 (20+)
p2 (20+)
p1 (10−19)
1 2 3 4 5 6 7 8 9 10 11 12 13 14 15 16 17 18 19 20 21 22 23 24 25 26 27 28 29 30 31 32 33 34 35 36 37 38 39 40 41 42 43 44 45 46
day

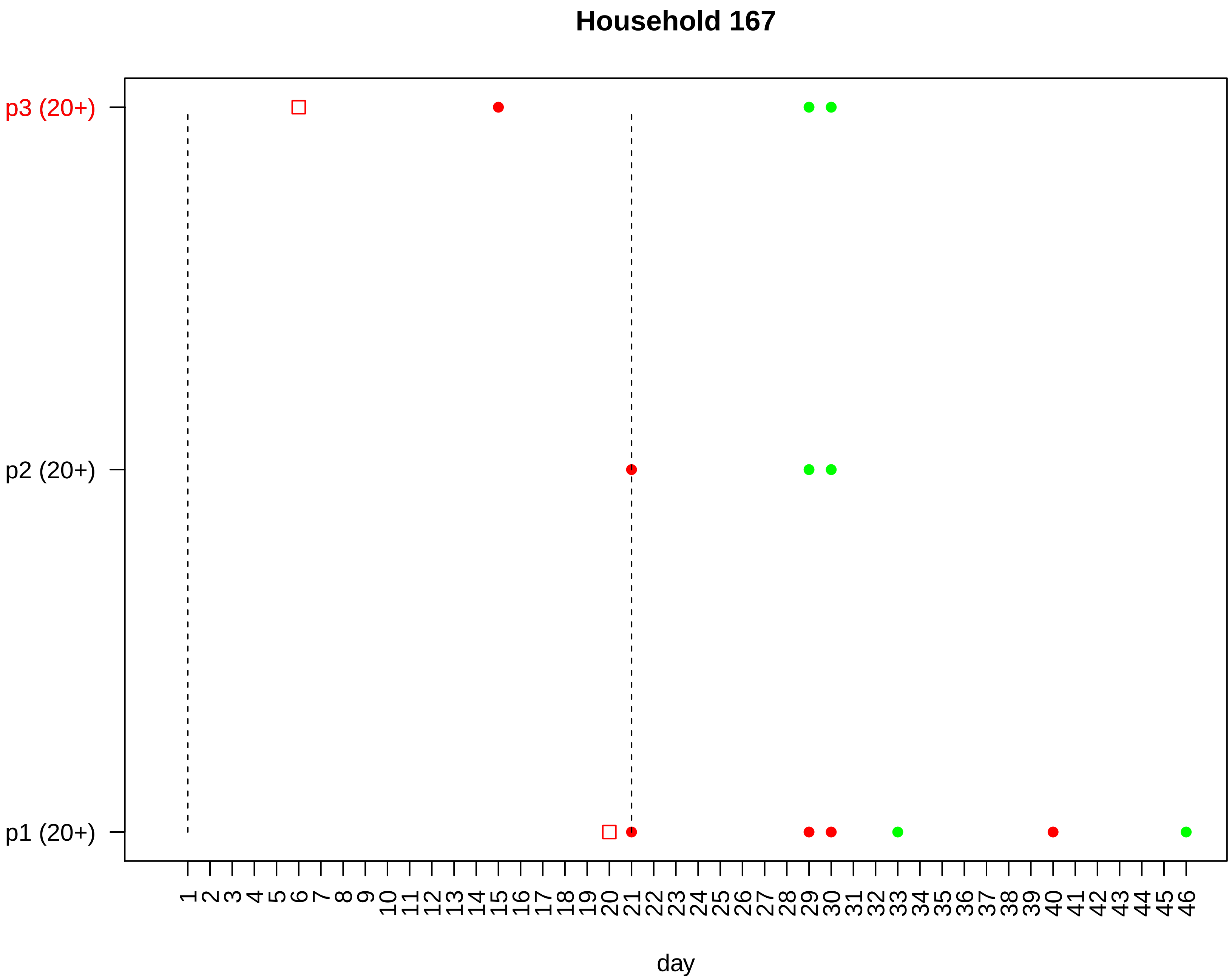
Household 167
p3 (20+)
p2 (20+)
p1 (20+)
1 2 3 4 5 6 7 8 9 10 11 12 13 14 15 16 17 18 19 20 21 22 23 24 25 26 27 28 29 30 31 32 33 34 35 36 37 38 39 40 41 42 43 44 45 46
day

#### Household 168

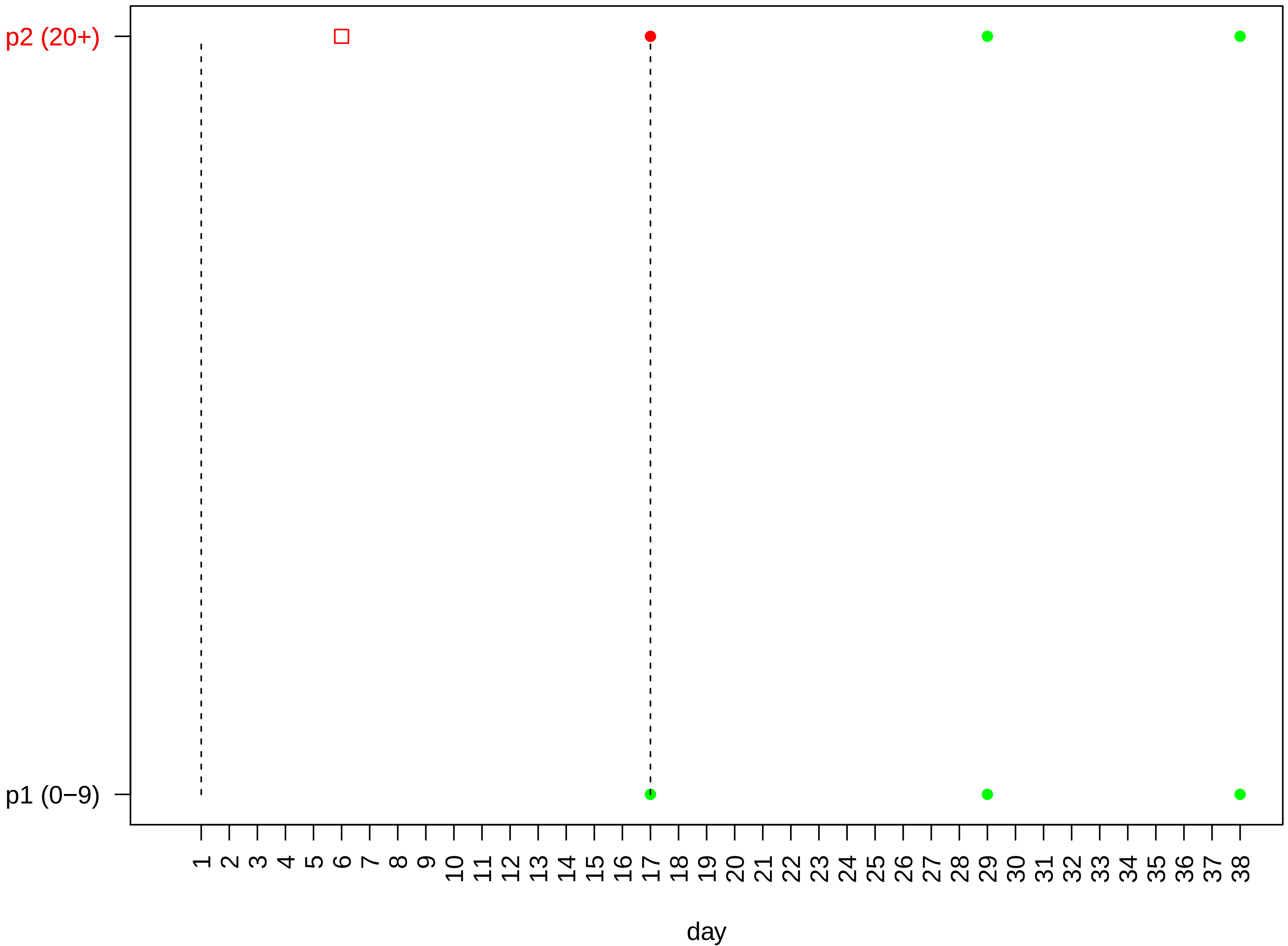

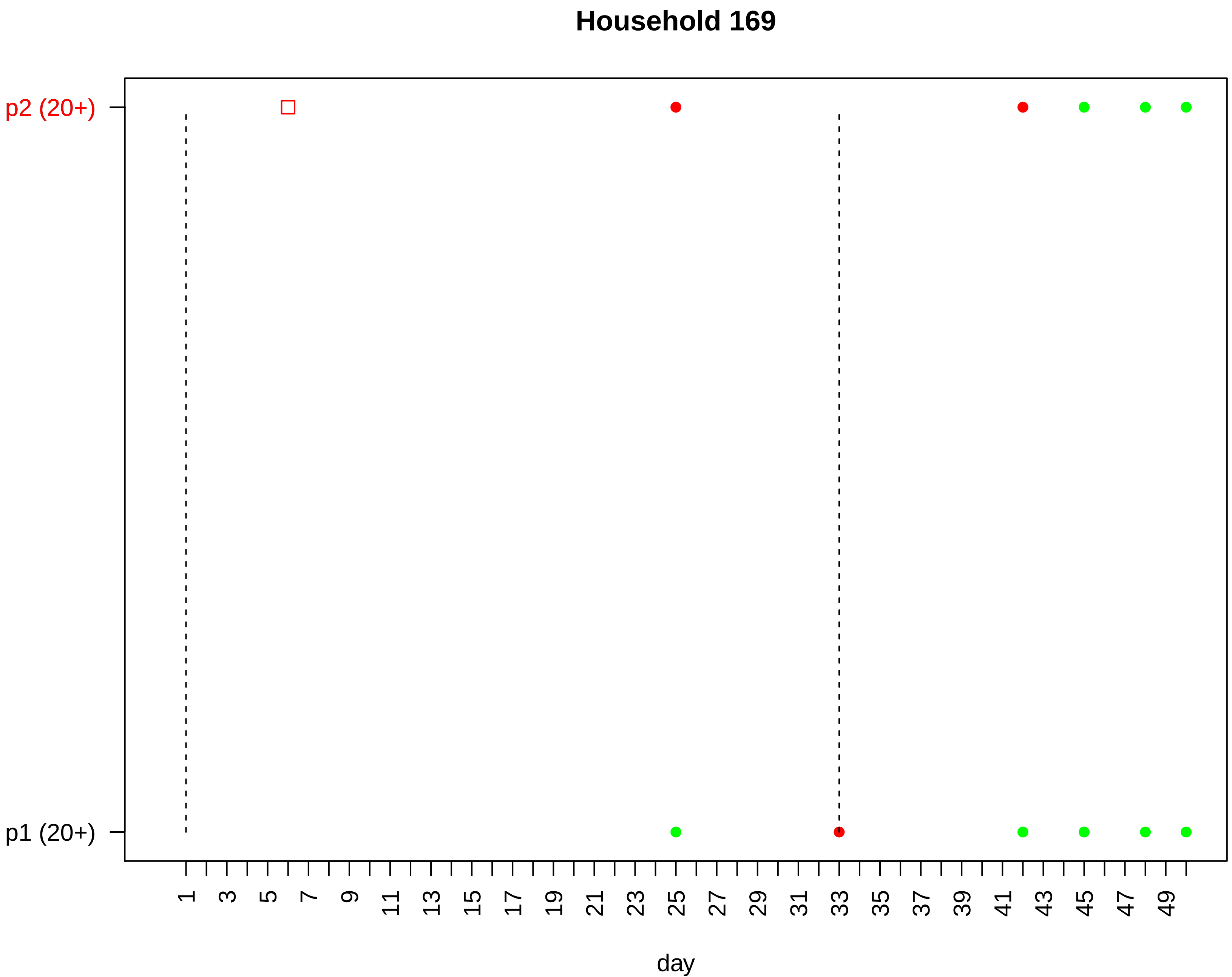
Household 169
p2 (20+)
p1 (20+)
1
3
5
7
9
11
13
15
17
19
21
23
25
27
29
31
33
35
37
39
41
43
45
47
49
day

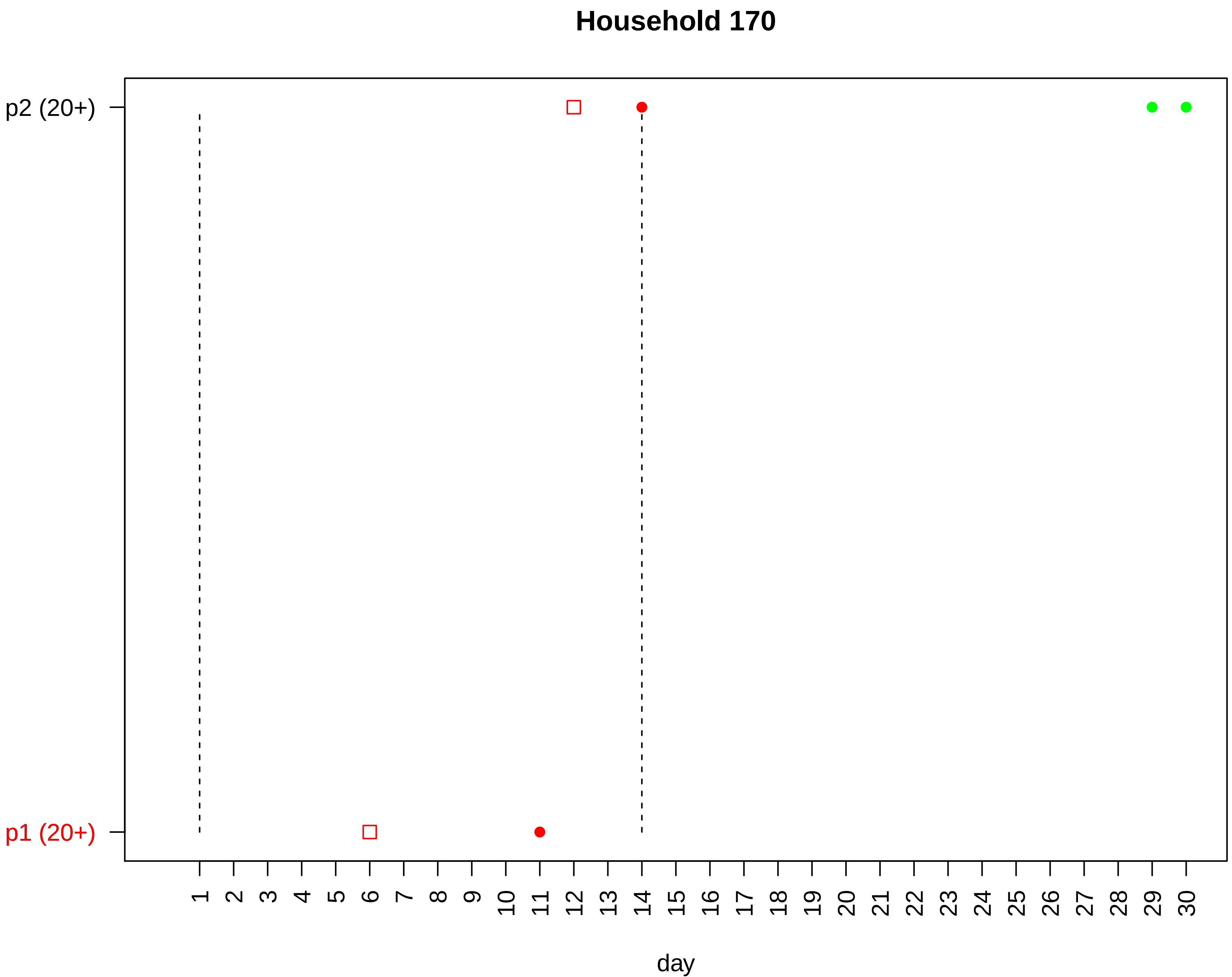

Household 170
p2 (20+)
p1 (20+)
day

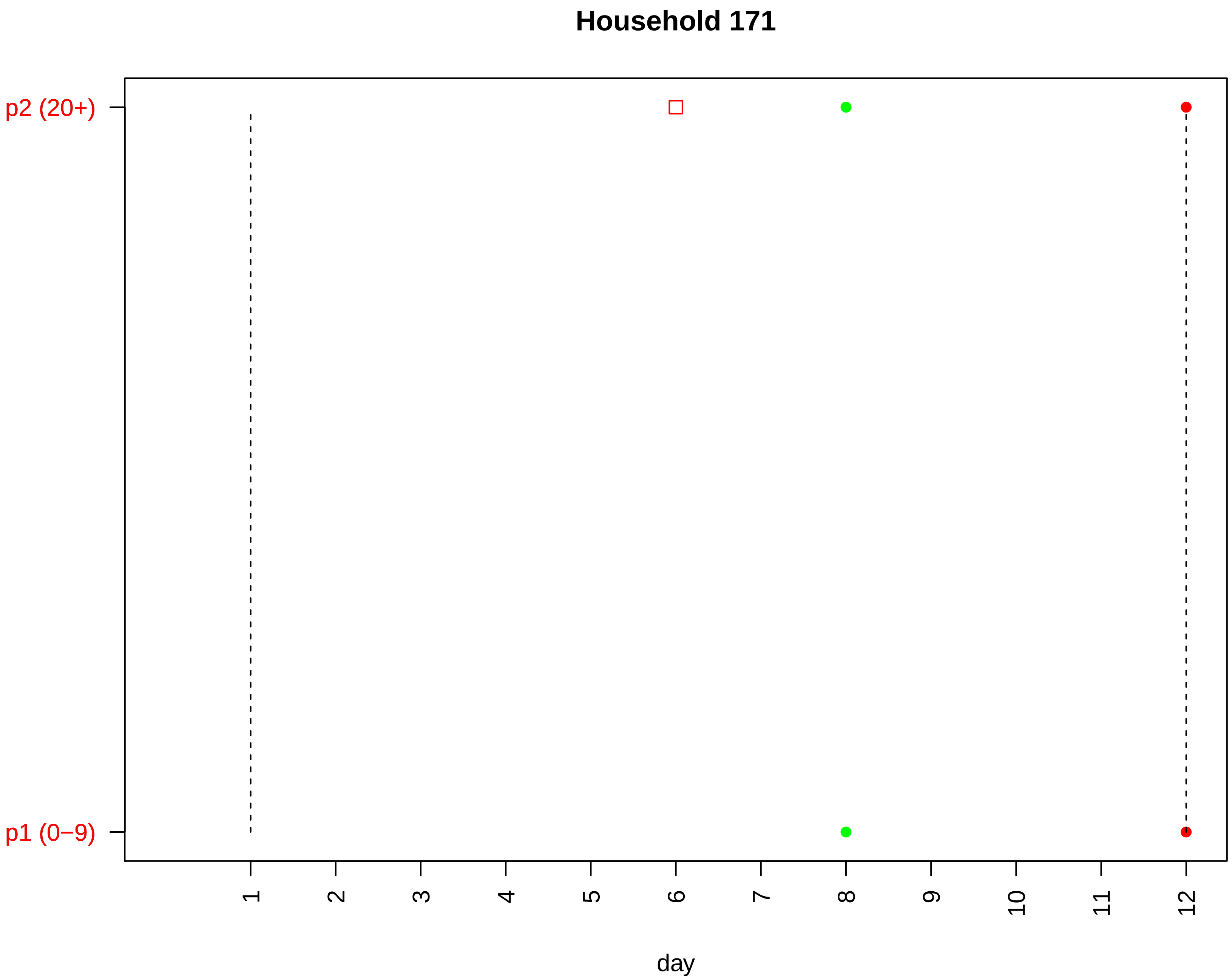

Household 171
p2 (20+)
p1 (0−9)
1
2
3
4
5
6
7
8
9
10
11
12
day

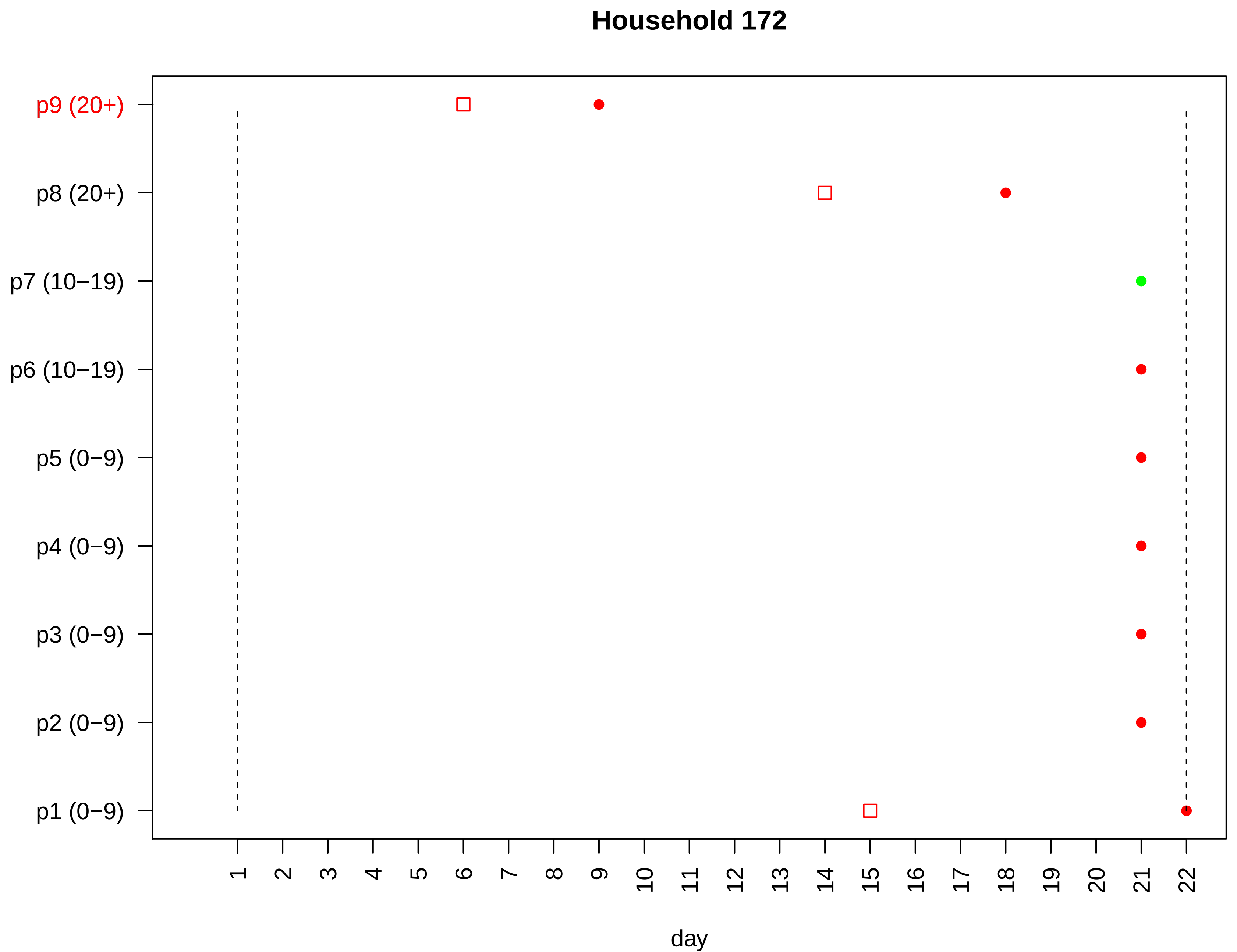

Household 172
p9 (20+)
p8 (20+)
p7 (10−19)
p6 (10−19)
p5 (0−9)
p4 (0−9)
p3 (0−9)
p2 (0−9)
p1 (0−9)
day

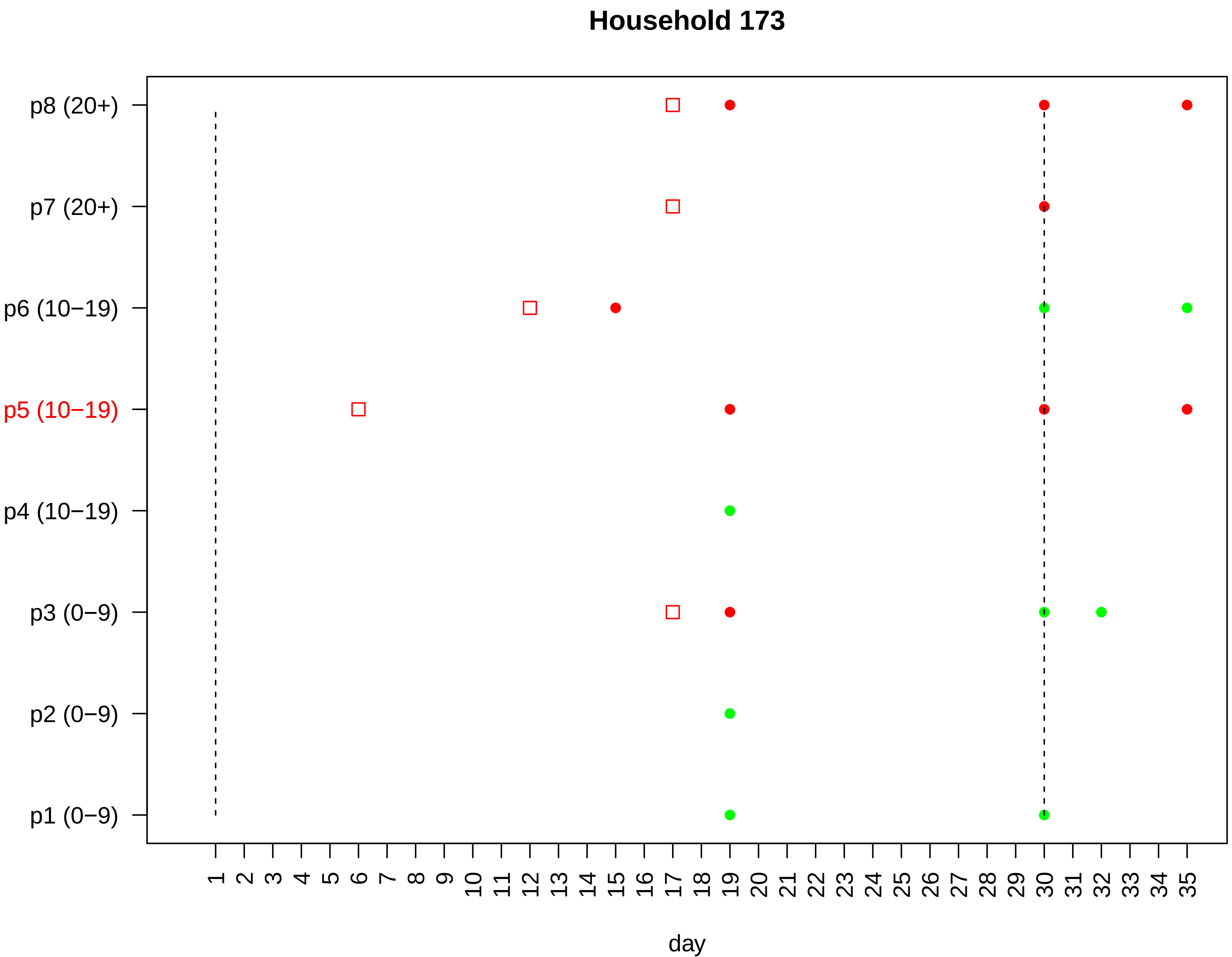

Household 173
p8 (20+)
p7 (20+)
p6 (10−19)
p5 (10−19)
p4 (10−19)
p3 (0−9)
p2 (0−9)
p1 (0−9)
1 2 3 4 5 6 7 8 9 10 11 12 13 14 15 16 17 18 19 20 21 22 23 24 25 26 27 28 29 30 31 32 33 34 35
day

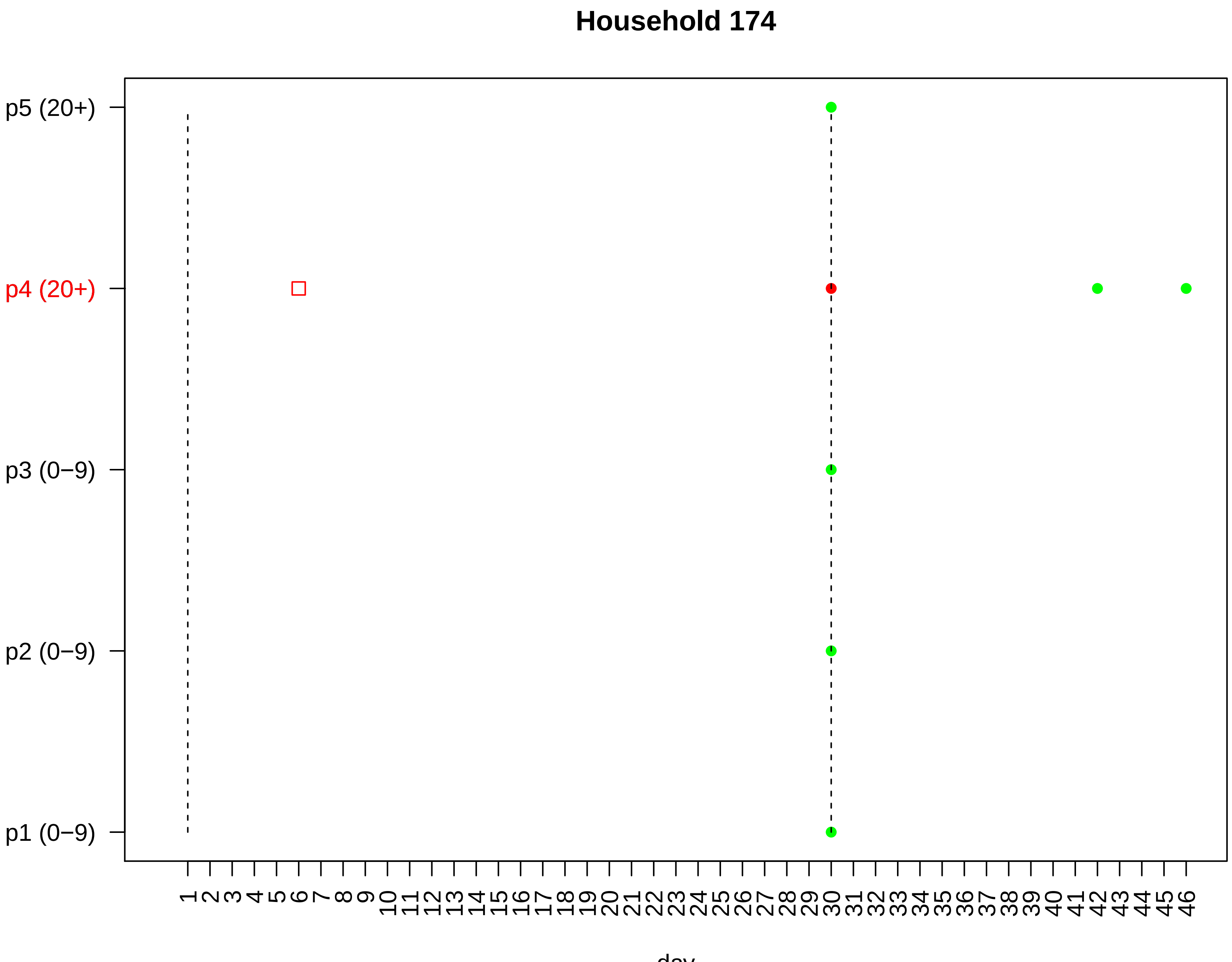

Household 174
p5 (20+)
p4 (20+)
p3 (0−9)
p2 (0−9)
p1 (0−9)
1 2 3 4 5 6 7 8 9 10 11 12 13 14 15 16 17 18 19 20 21 22 23 24 25 26 27 28 29 30 31 32 33 34 35 36 37 38 39 40 41 42 43 44 45 46
day

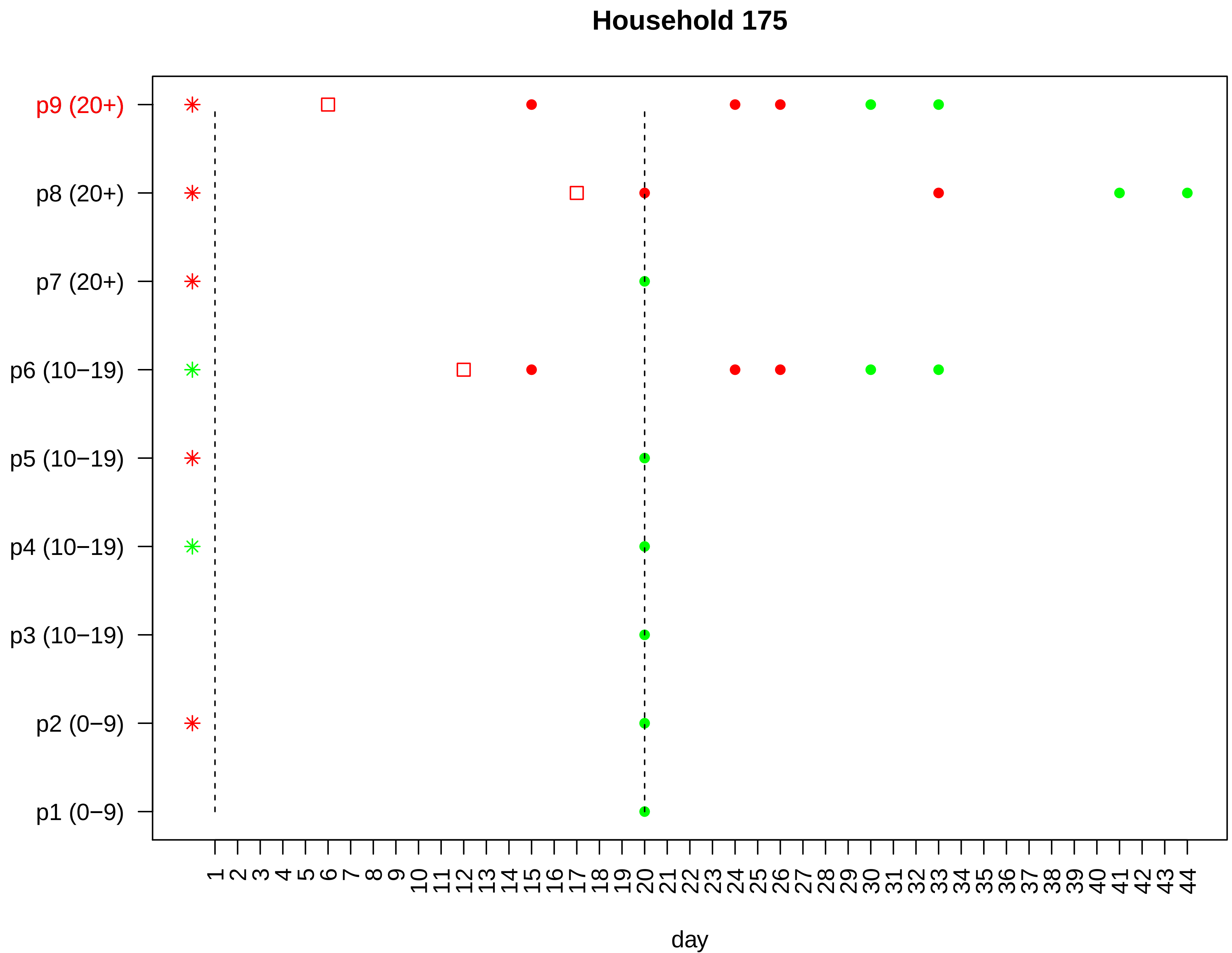

Household 175
p9 (20+)
p8 (20+)
p7 (20+)
p6 (10−19)
p5 (10−19)
p4 (10−19)
p3 (10−19)
p2 (0−9)
p1 (0−9)
1 2 3 4 5 6 7 8 9 10 11 12 13 14 15 16 17 18 19 20 21 22 23 24 25 26 27 28 29 30 31 32 33 34 35 36 37 38 39 40 41 42 43 44
day

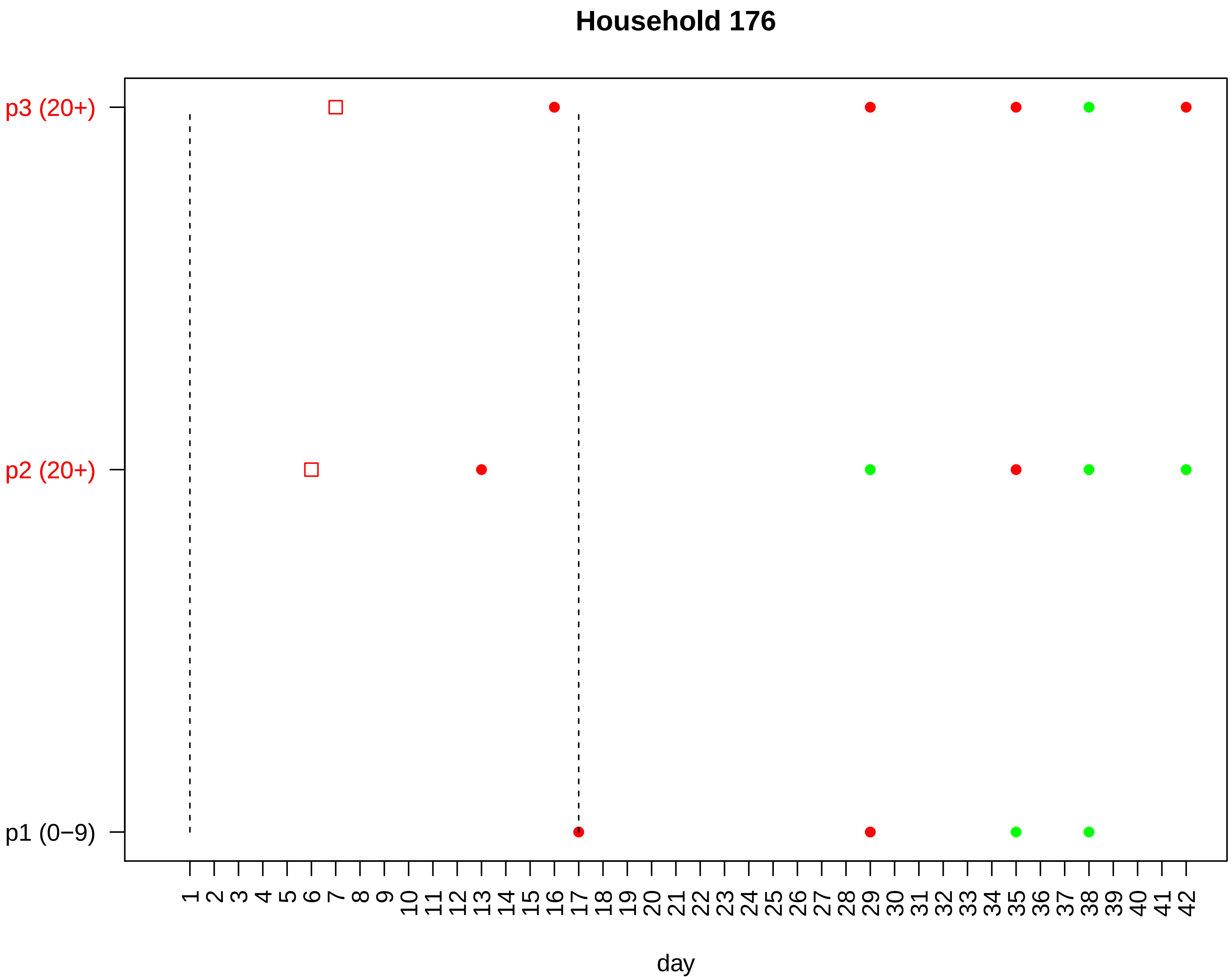

Household 176
p3 (20+)
p2 (20+)
p1 (0−9)
1 2 3 4 5 6 7 8 9 10 11 12 13 14 15 16 17 18 19 20 21 22 23 24 25 26 27 28 29 30 31 32 33 34 35 36 37 38 39 40 41 42
day

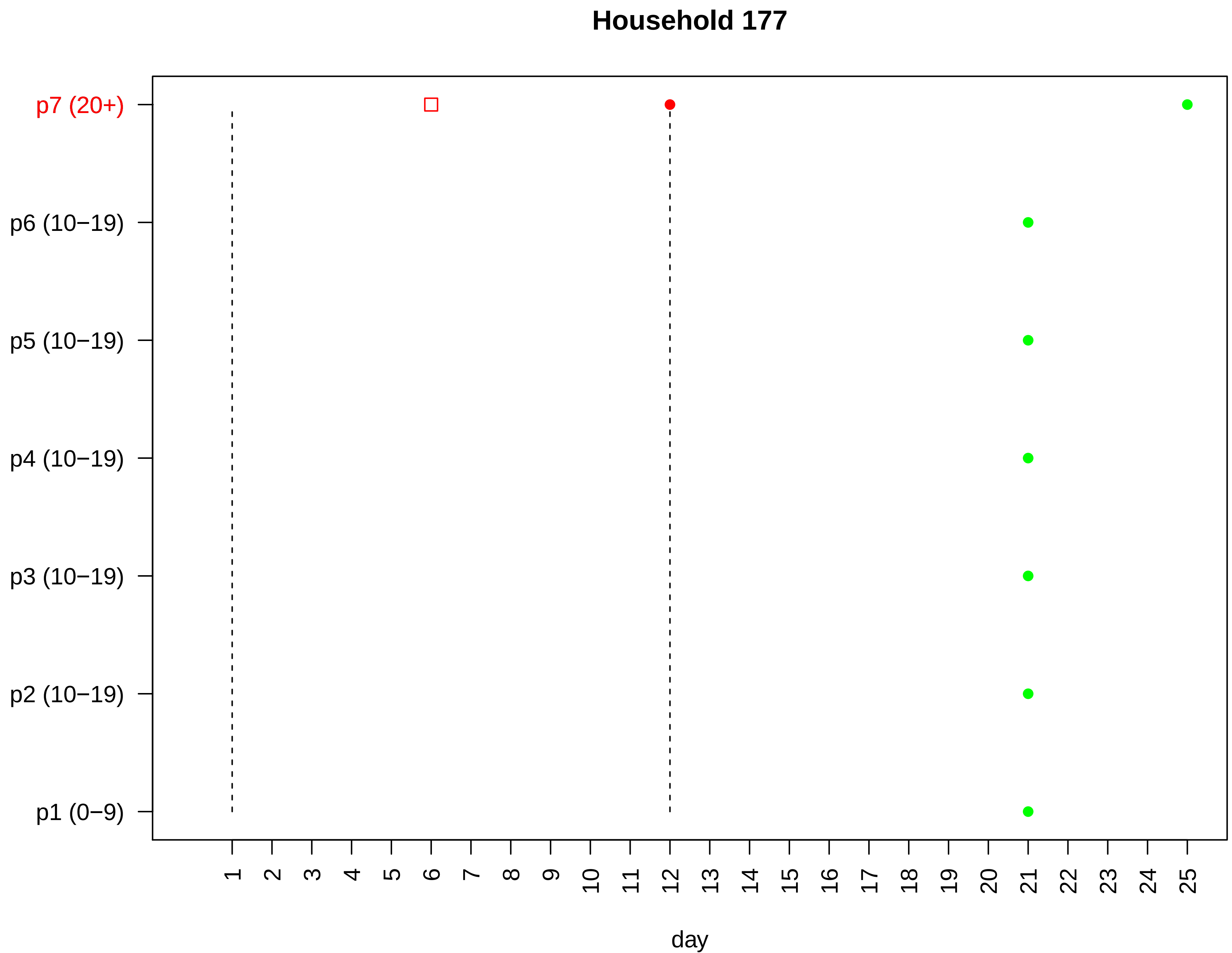

Household 177
p7 (20+)
p6 (10−19)
p5 (10−19)
p4 (10−19)
p3 (10−19)
p2 (10−19)
p1 (0−9)
day

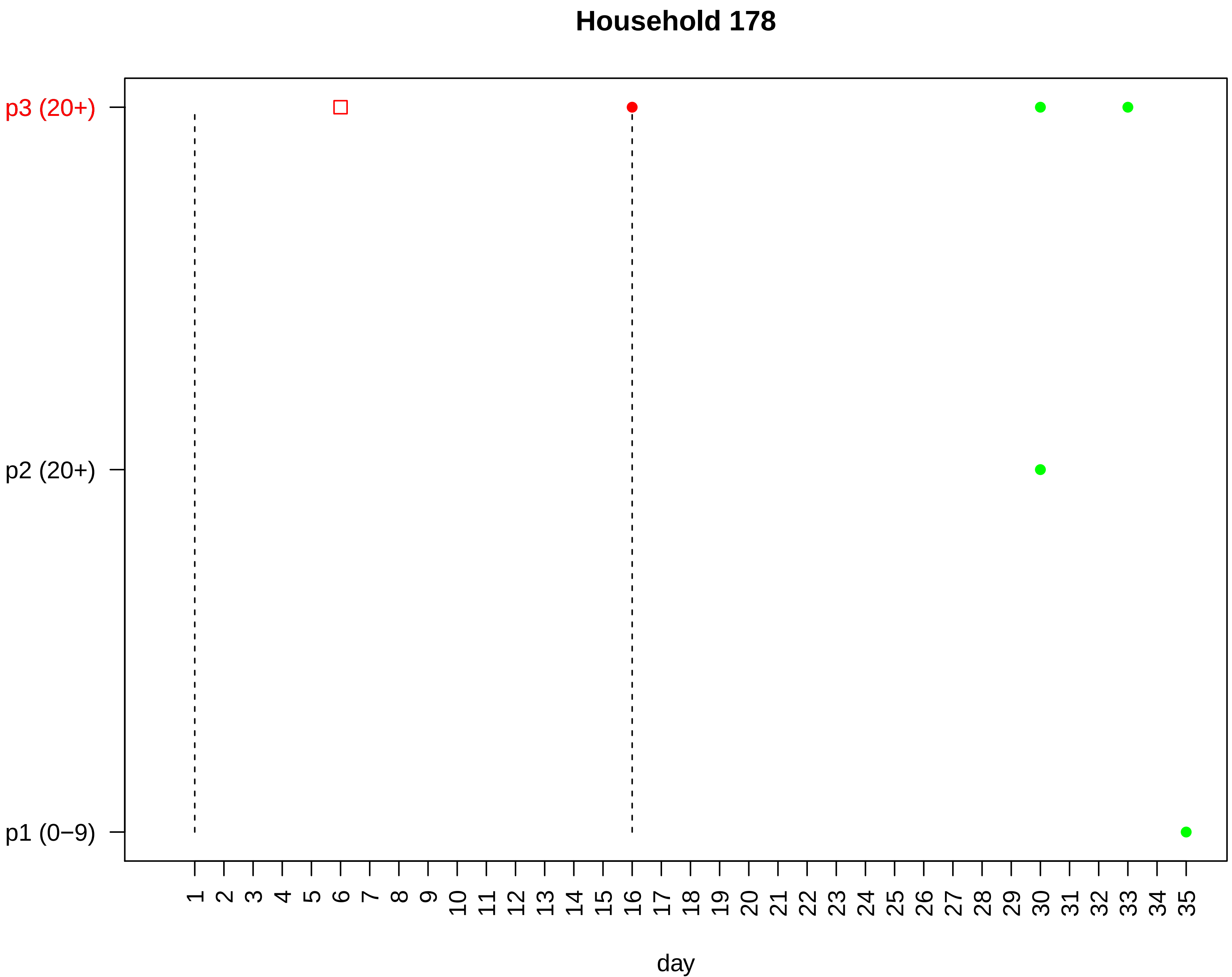
Household 178
p3 (20+)
p2 (20+)
p1 (0−9)
1 2 3 4 5 6 7 8 9 10 11 12 13 14 15 16 17 18 19 20 21 22 23 24 25 26 27 28 29 30 31 32 33 34 35
day

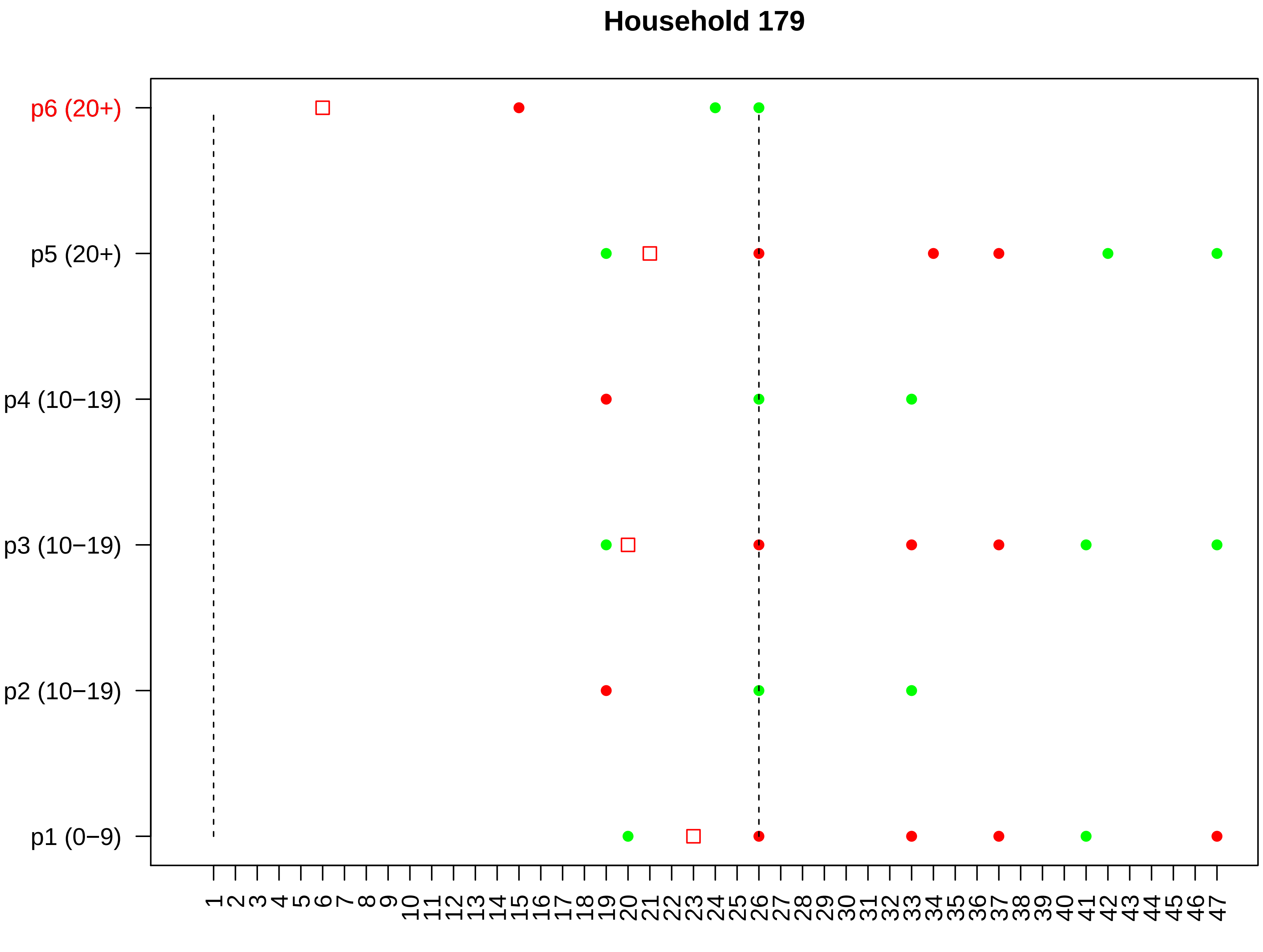

Household 179
p6 (20+)
p5 (20+)
p4 (10−19)
p3 (10−19)
p2 (10−19)
p1 (0−9)
1 2 3 4 5 6 7 8 9 10 11 12 13 14 15 16 17 18 19 20 21 22 23 24 25 26 27 28 29 30 31 32 33 34 35 36 37 38 39 40 41 42 43 44 45 46 47
day

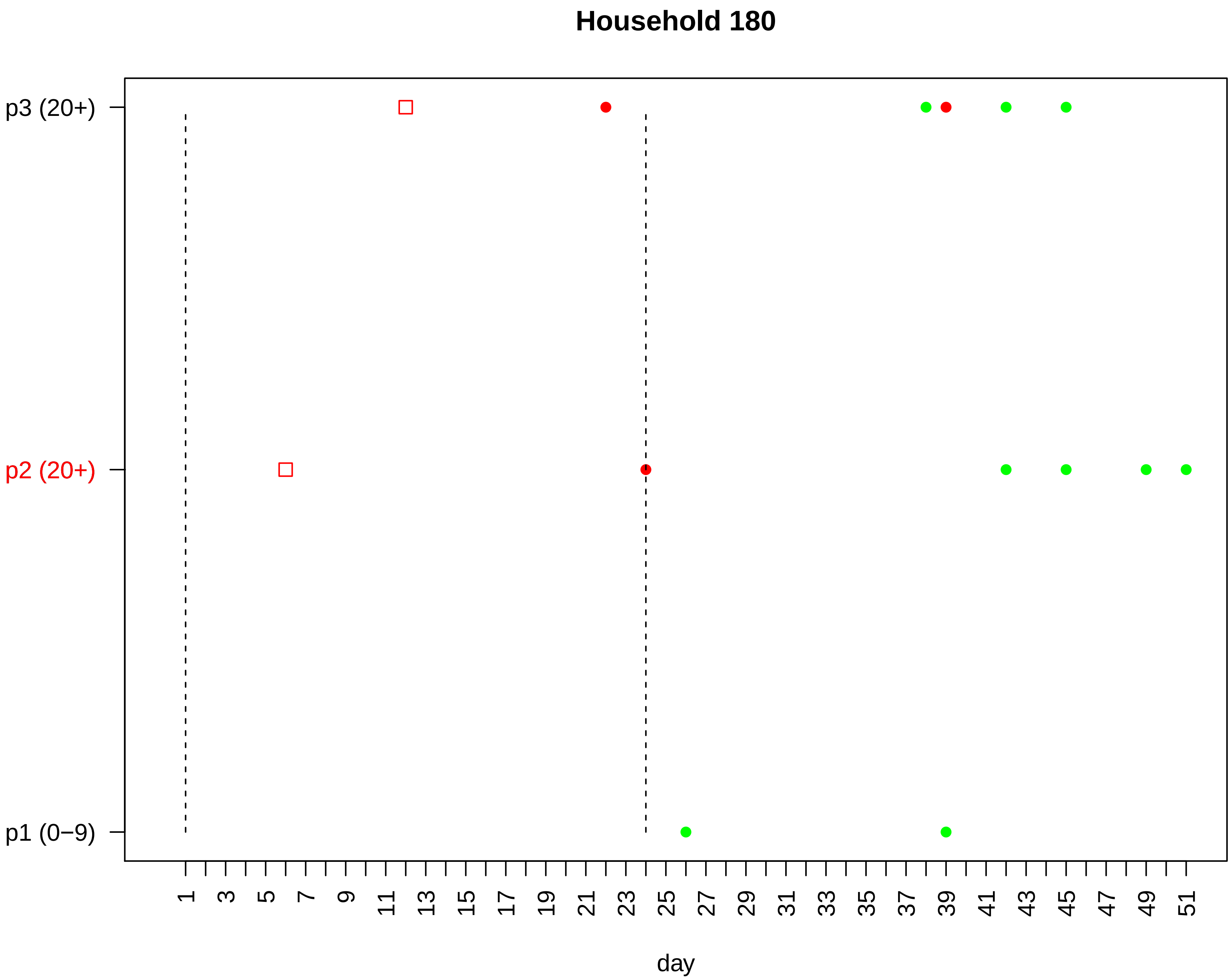

Household 180
p3 (20+)
p2 (20+)
p1 (0−9)
1 3 5 7 9 11 13 15 17 19 21 23 25 27 29 31 33 35 37 39 41 43 45 47 49 51
day

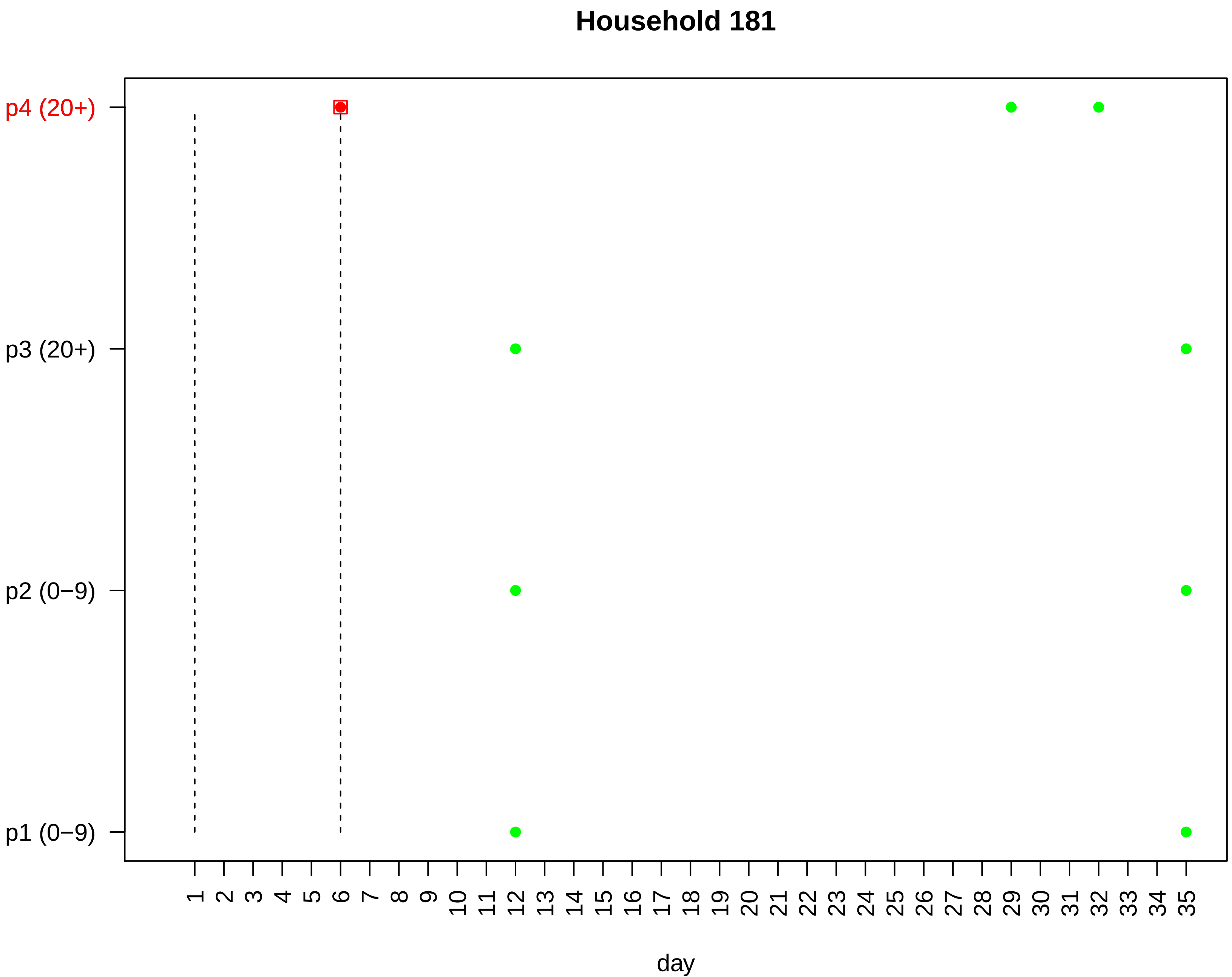

Household 181
p4 (20+)
p3 (20+)
p2 (0−9)
p1 (0−9)
1 2 3 4 5 6 7 8 9 10 11 12 13 14 15 16 17 18 19 20 21 22 23 24 25 26 27 28 29 30 31 32 33 34 35
day

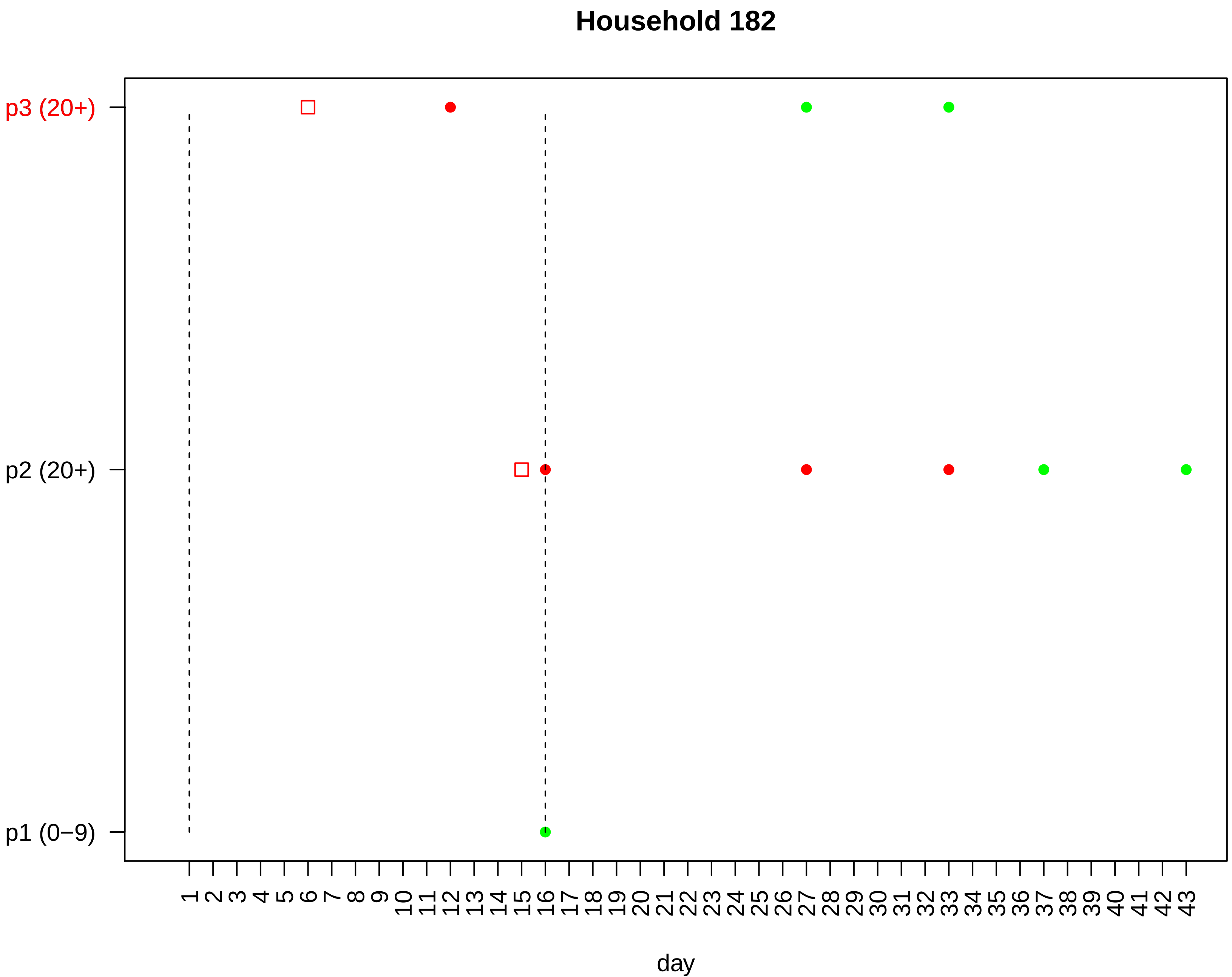

Household 182
p3 (20+)
p2 (20+)
p1 (0−9)
1 2 3 4 5 6 7 8 9 10 11 12 13 14 15 16 17 18 19 20 21 22 23 24 25 26 27 28 29 30 31 32 33 34 35 36 37 38 39 40 41 42 43
day

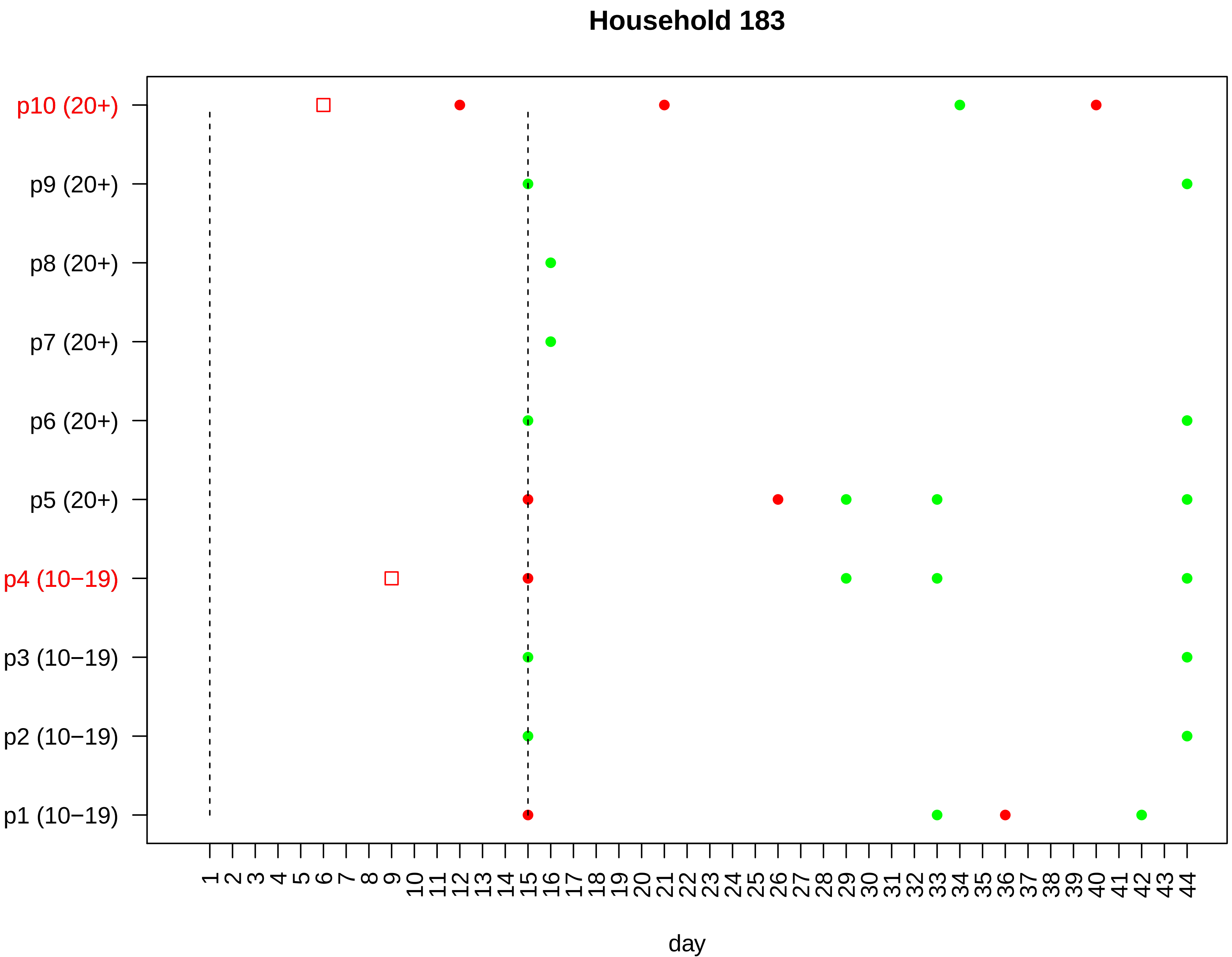

Household 183
p10 (20+)
p9 (20+)
p8 (20+)
p7 (20+)
p6 (20+)
p5 (20+)
p4 (10−19)
p3 (10−19)
p2 (10−19)
p1 (10−19)
1 2 3 4 5 6 7 8 9 10 11 12 13 14 15 16 17 18 19 20 21 22 23 24 25 26 27 28 29 30 31 32 33 34 35 36 37 38 39 40 41 42 43 44
day

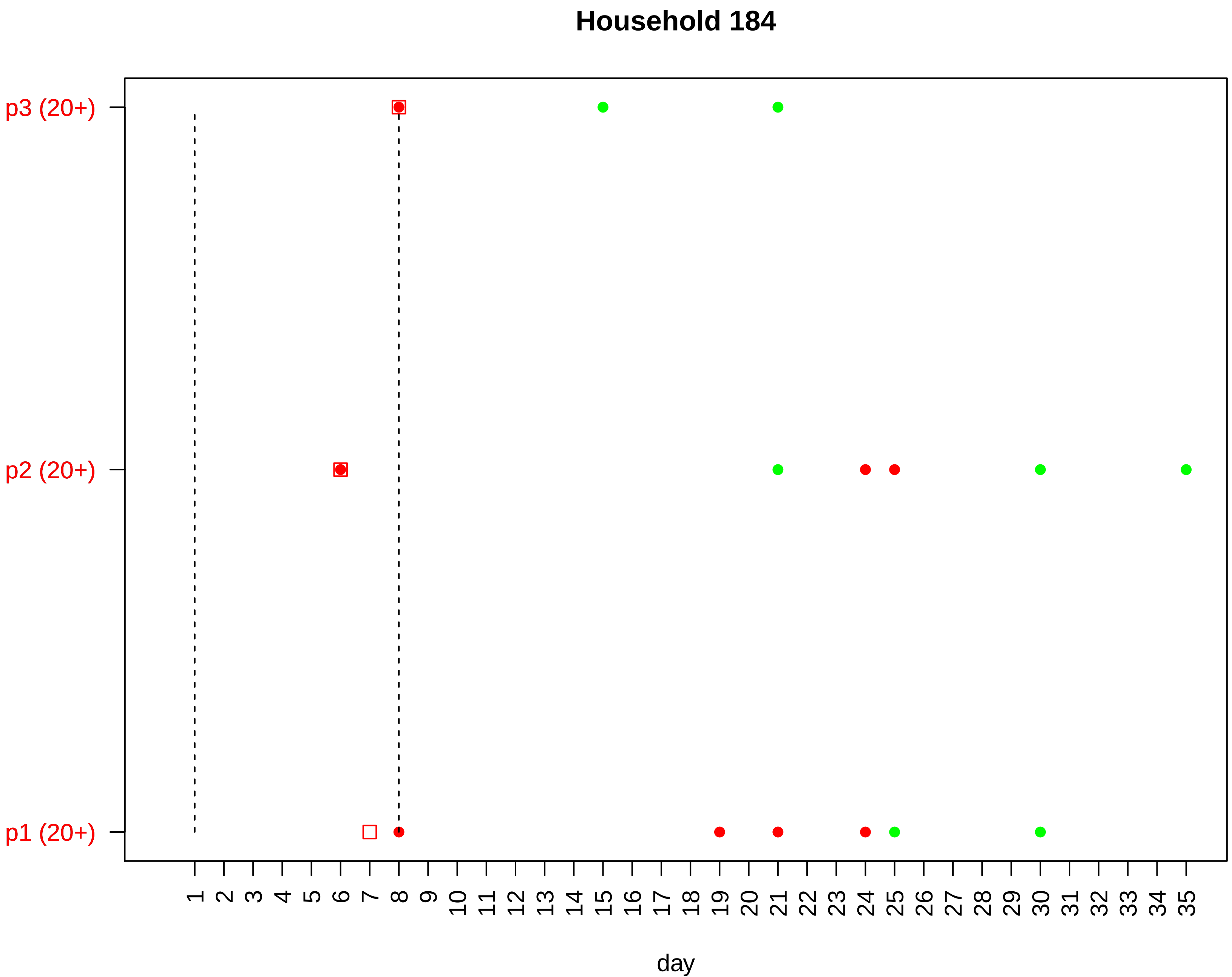

Household 184
p3 (20+)
p2 (20+)
p1 (20+)
1 2 3 4 5 6 7 8 9 10 11 12 13 14 15 16 17 18 19 20 21 22 23 24 25 26 27 28 29 30 31 32 33 34 35
day

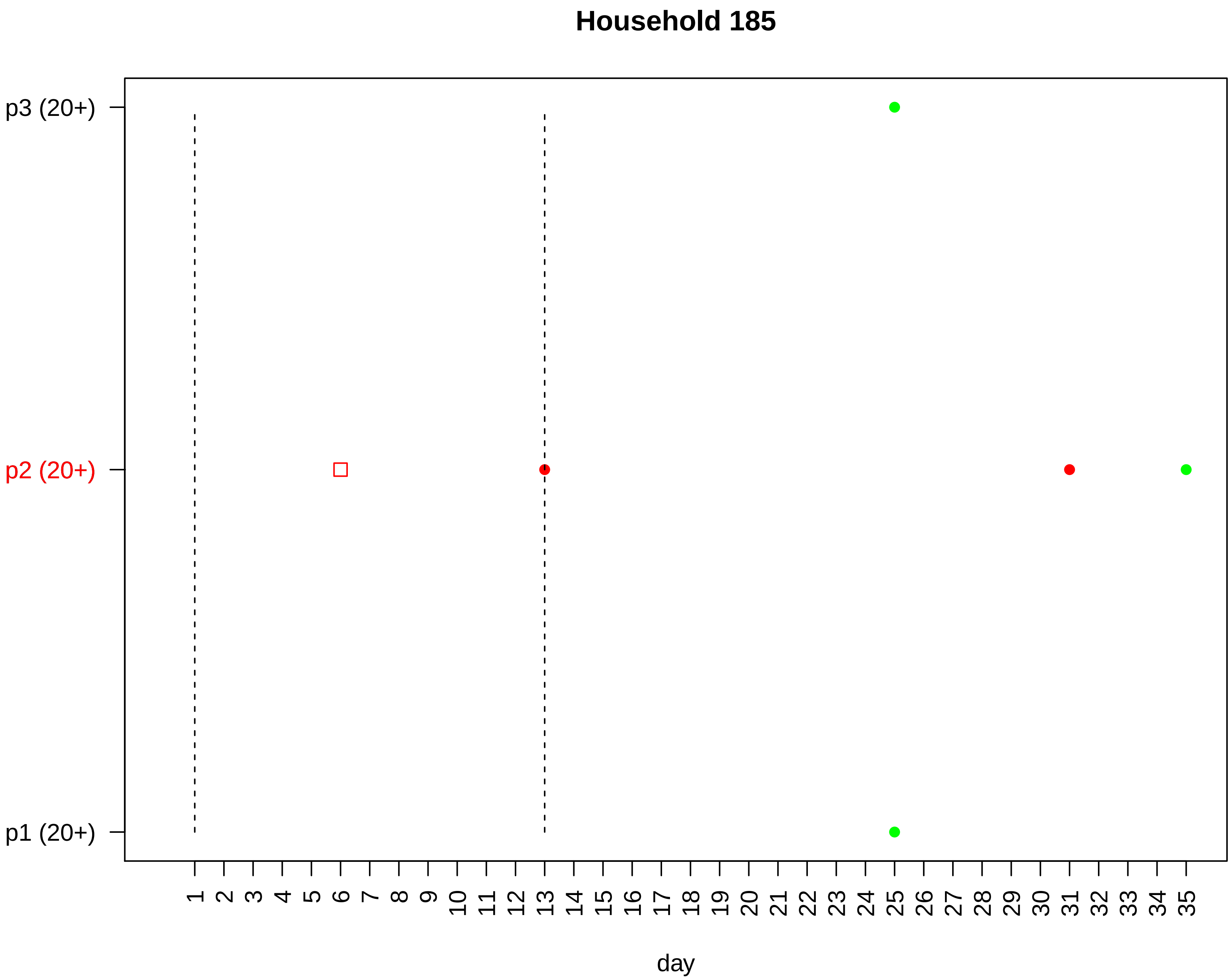

Household 185
p3 (20+)
p2 (20+)
p1 (20+)
1 2 3 4 5 6 7 8 9 10 11 12 13 14 15 16 17 18 19 20 21 22 23 24 25 26 27 28 29 30 31 32 33 34 35
day

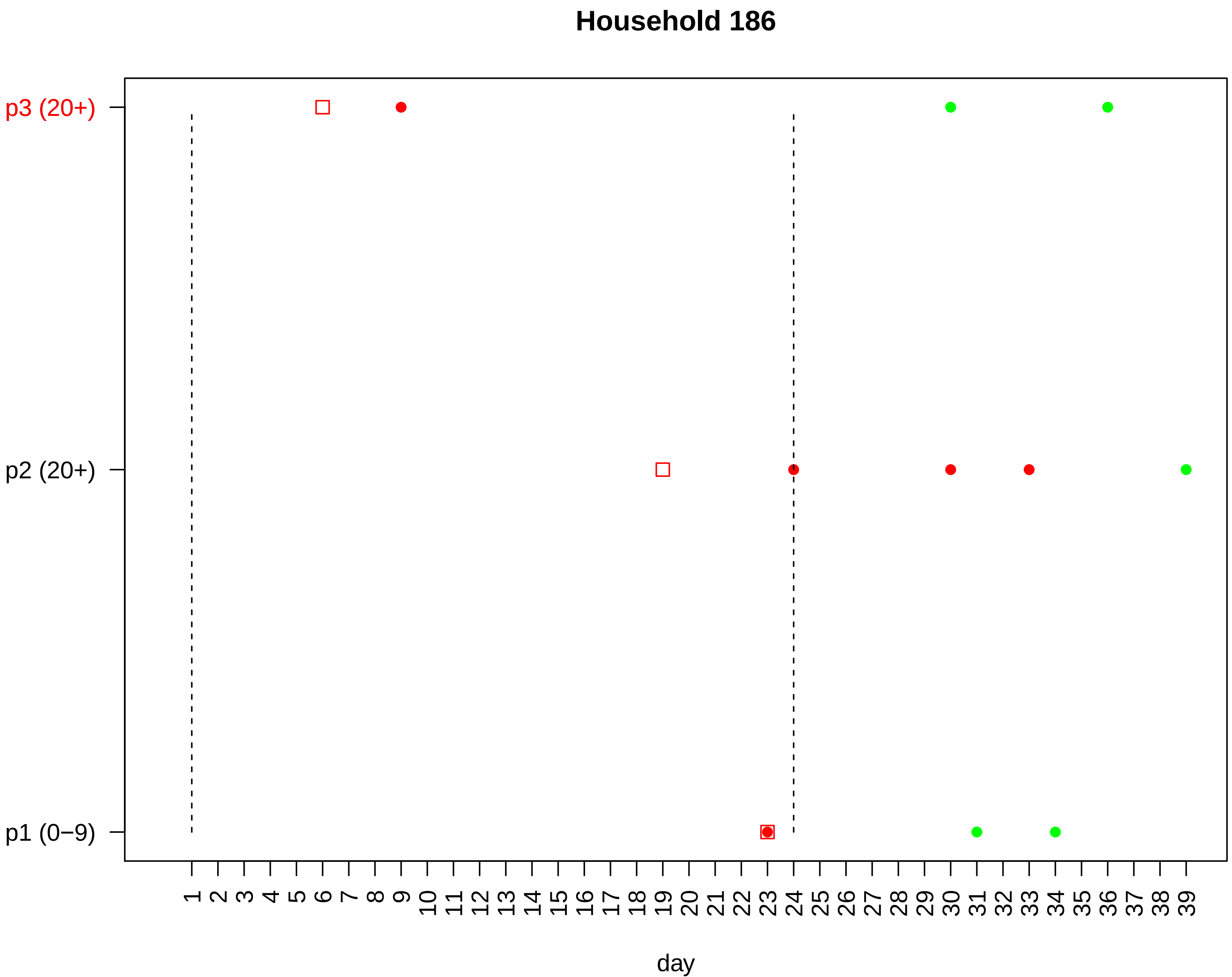

Household 186
p3 (20+)
p2 (20+)
p1 (0−9)
1 2 3 4 5 6 7 8 9 10 11 12 13 14 15 16 17 18 19 20 21 22 23 24 25 26 27 28 29 30 31 32 33 34 35 36 37 38 39
day

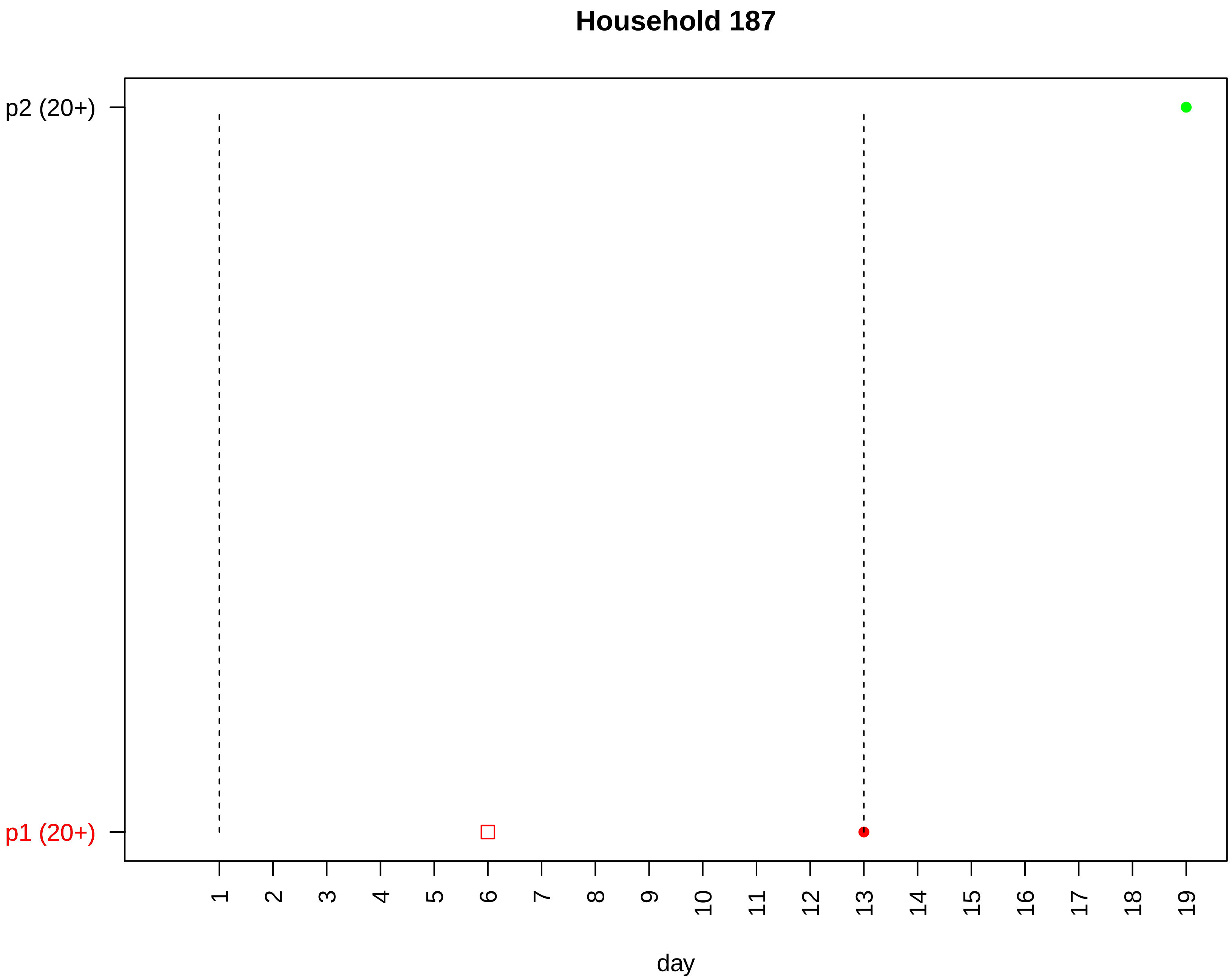
Household 187
p2 (20+)
p1 (20+)
day

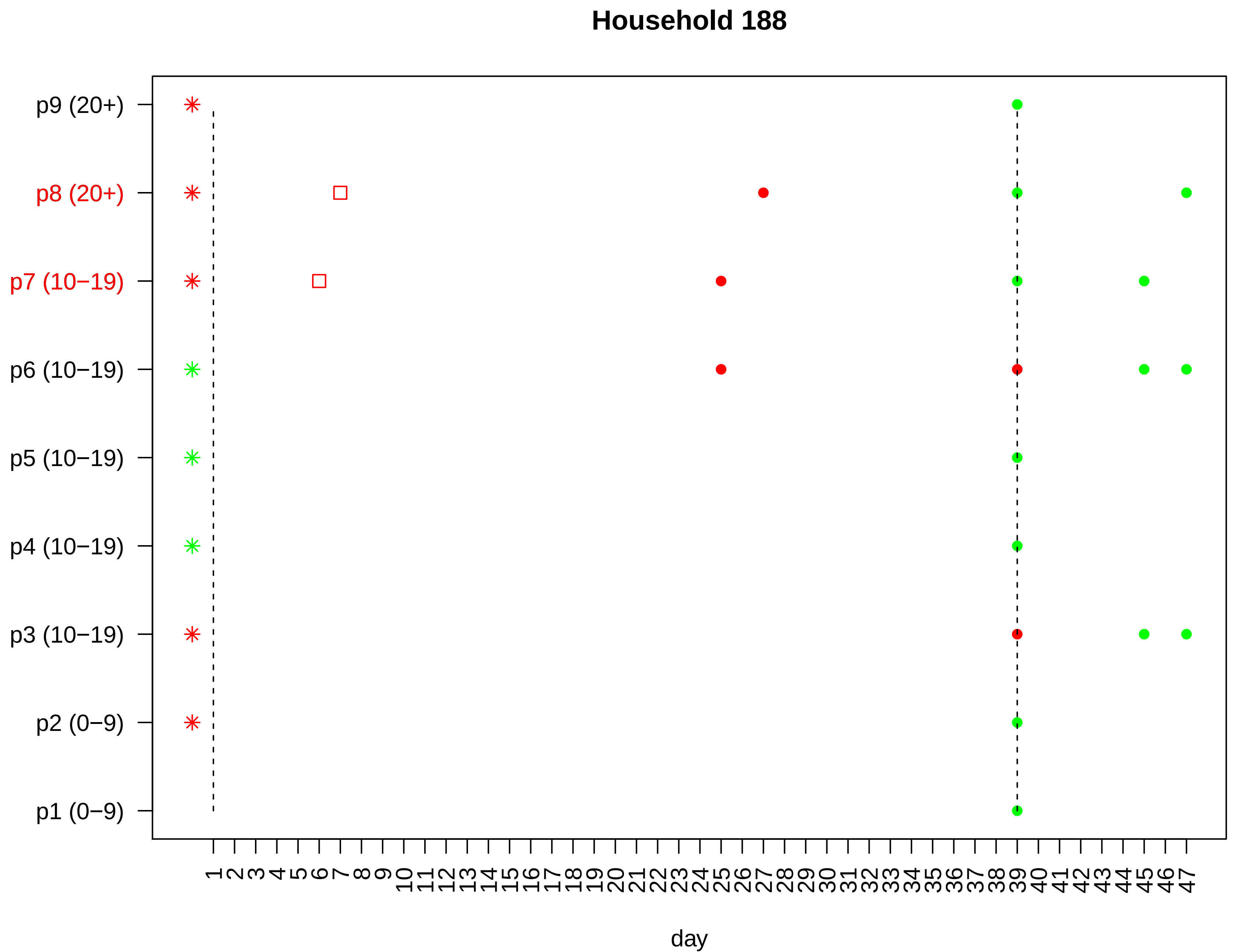

Household 188
p9 (20+)
p8 (20+)
p7 (10−19)
p6 (10−19)
p5 (10−19)
p4 (10−19)
p3 (10−19)
p2 (0−9)
p1 (0−9)
1 2 3 4 5 6 7 8 9 10 11 12 13 14 15 16 17 18 19 20 21 22 23 24 25 26 27 28 29 30 31 32 33 34 35 36 37 38 39 40 41 42 43 44 45 46 47
day

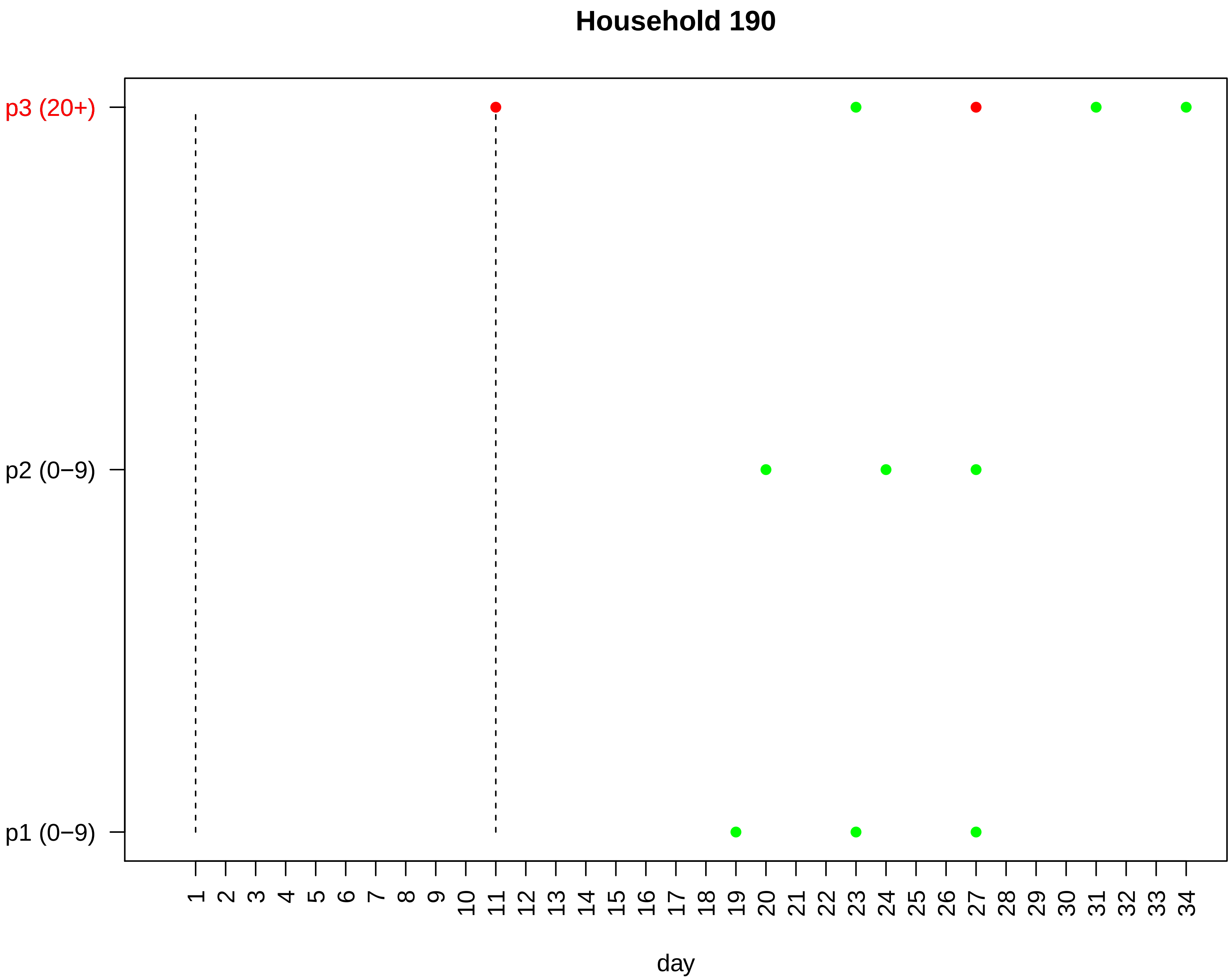

Household 190
p3 (20+)
p2 (0−9)
p1 (0−9)
day

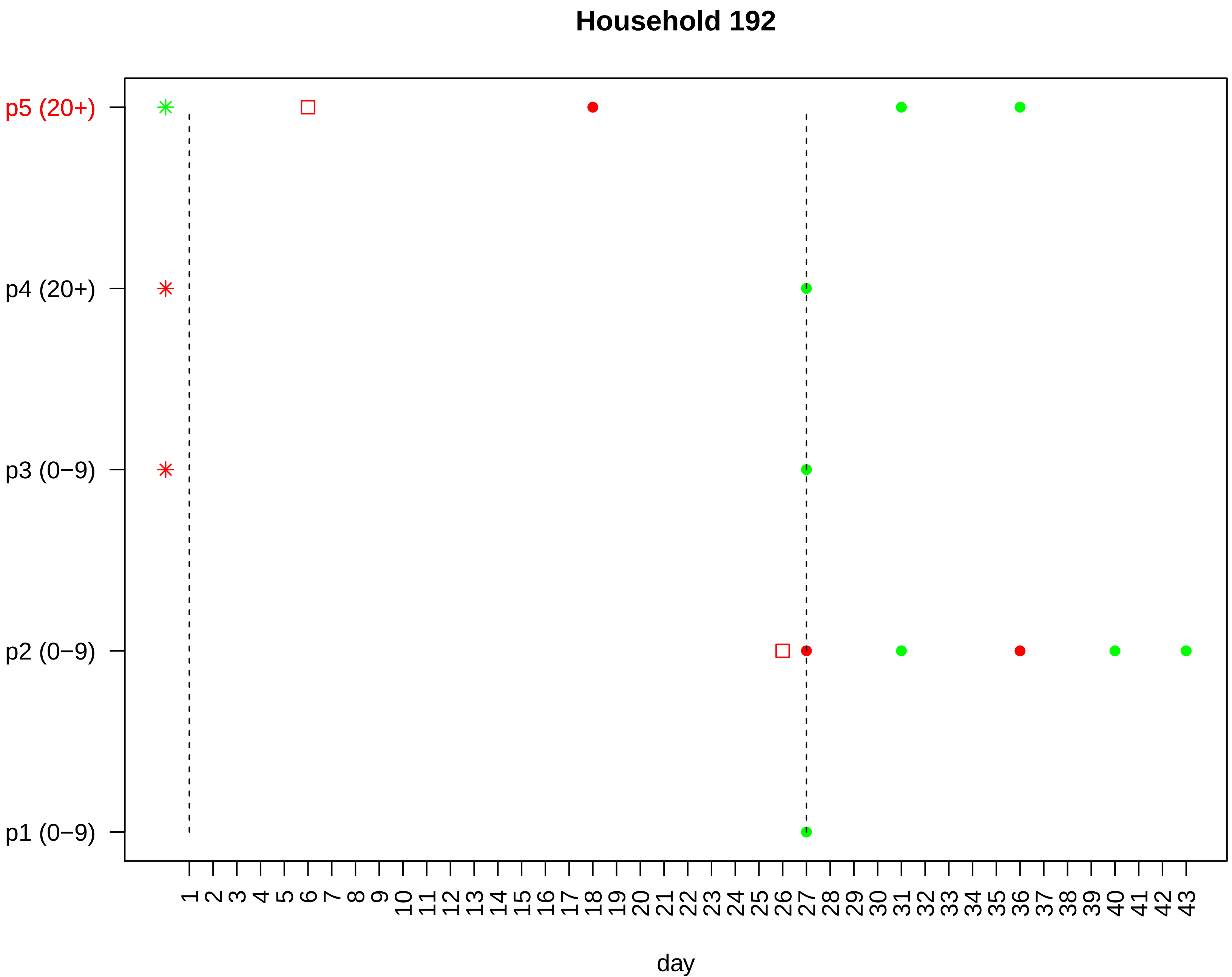
Household 192
p5 (20+)
p4 (20+)
p3 (0−9)
p2 (0−9)
p1 (0−9)
1 2 3 4 5 6 7 8 9 10 11 12 13 14 15 16 17 18 19 20 21 22 23 24 25 26 27 28 29 30 31 32 33 34 35 36 37 38 39 40 41 42 43
day

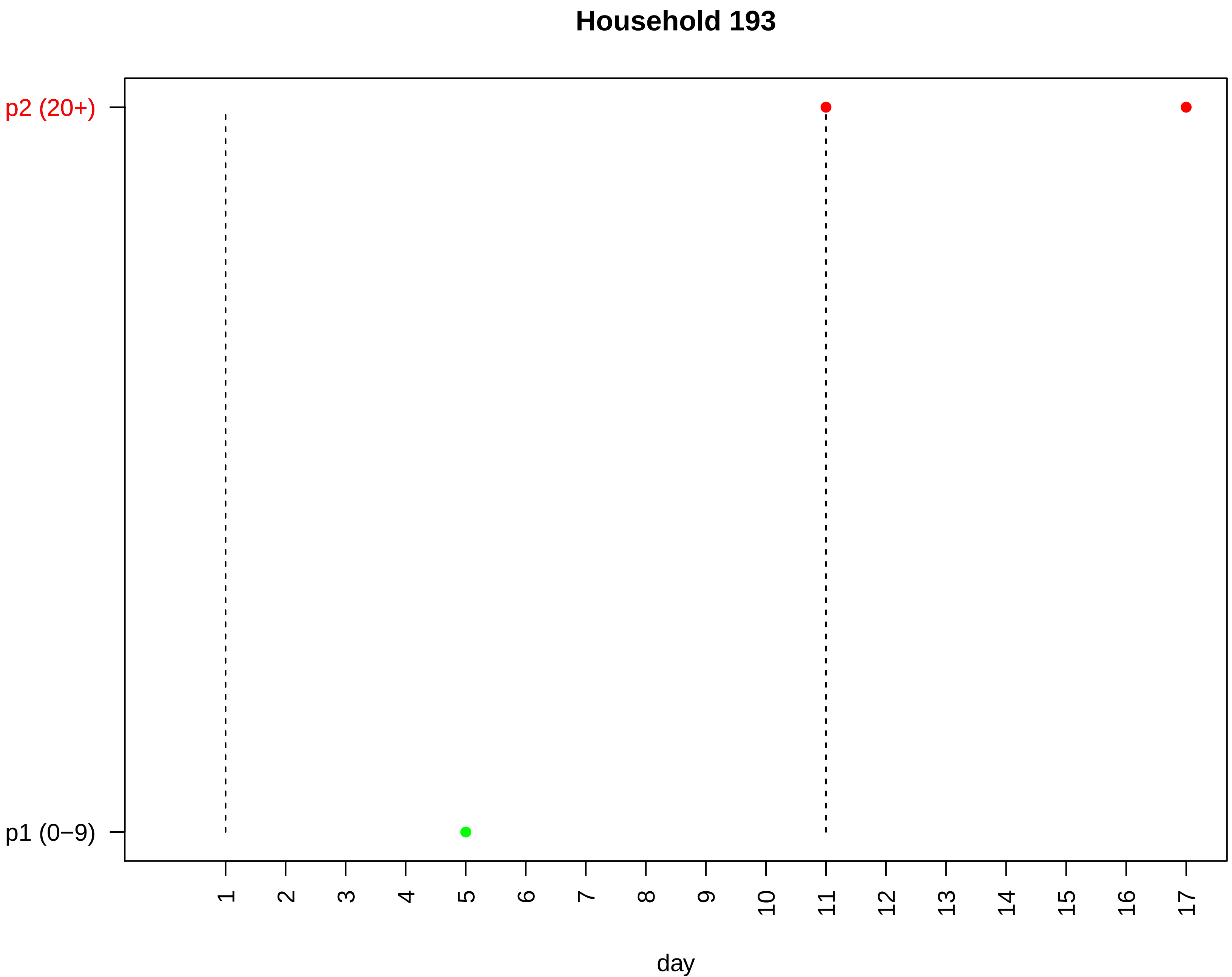
Household 193
p2 (20+)
p1 (0−9)
day

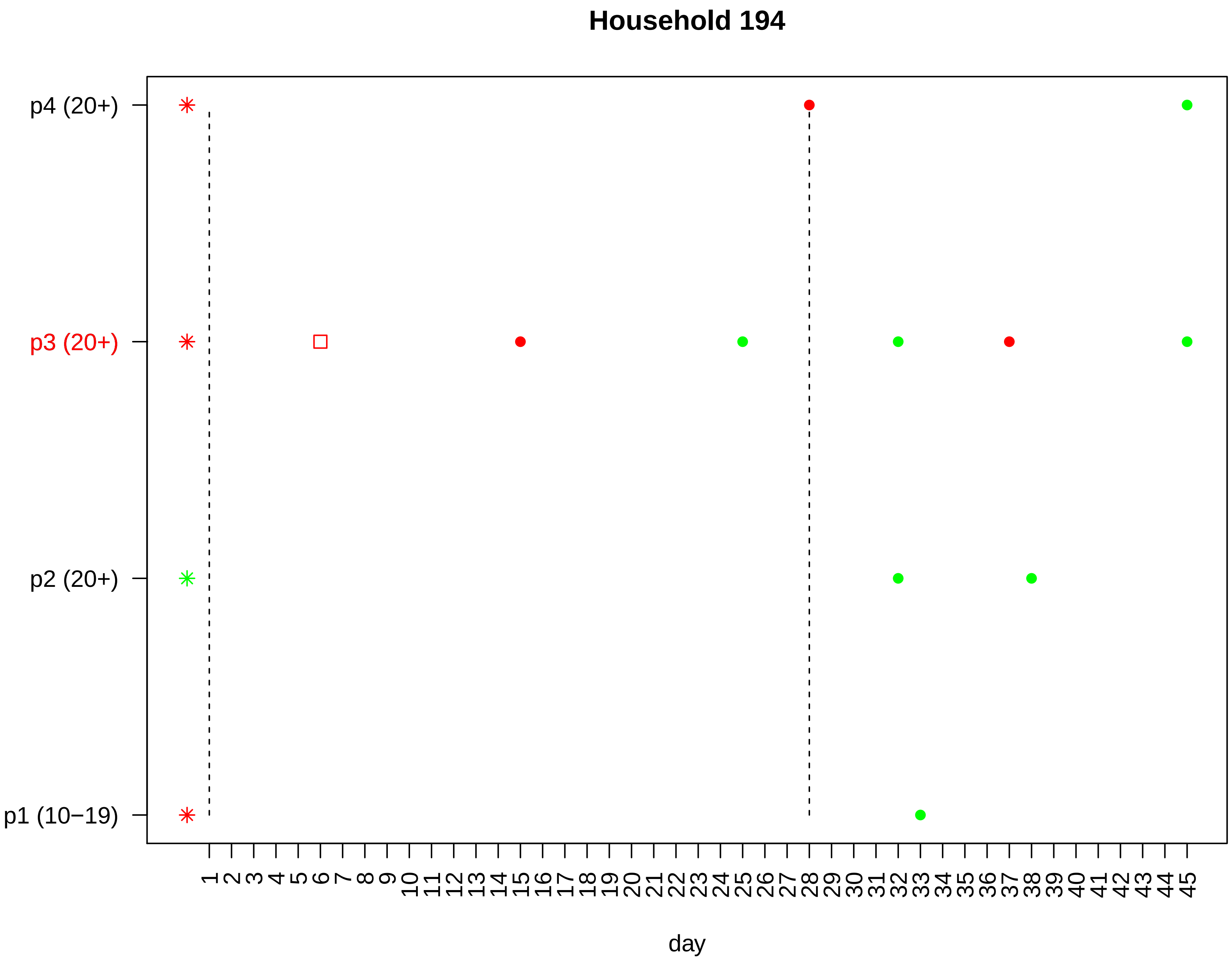

Household 194
p4 (20+)
p3 (20+)
p2 (20+)
p1 (10−19)
1 2 3 4 5 6 7 8 9 10 11 12 13 14 15 16 17 18 19 20 21 22 23 24 25 26 27 28 29 30 31 32 33 34 35 36 37 38 39 40 41 42 43 44 45
day

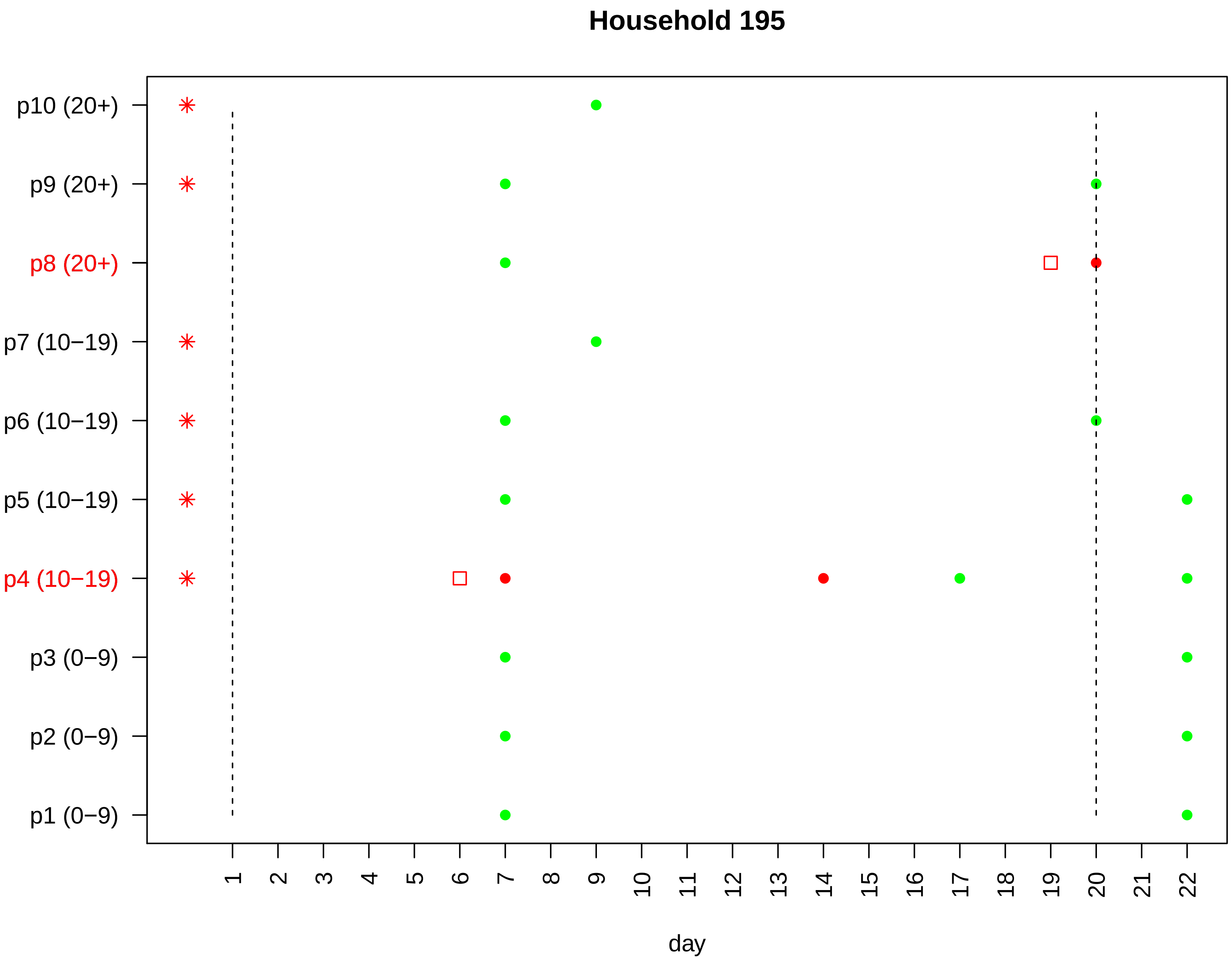

Household 195
p10 (20+)
p9 (20+)
p8 (20+)
p7 (10−19)
p6 (10−19)
p5 (10−19)
p4 (10−19)
p3 (0−9)
p2 (0−9)
p1 (0−9)
1 2 3 4 5 6 7 8 9 10 11 12 13 14 15 16 17 18 19 20 21 22
day

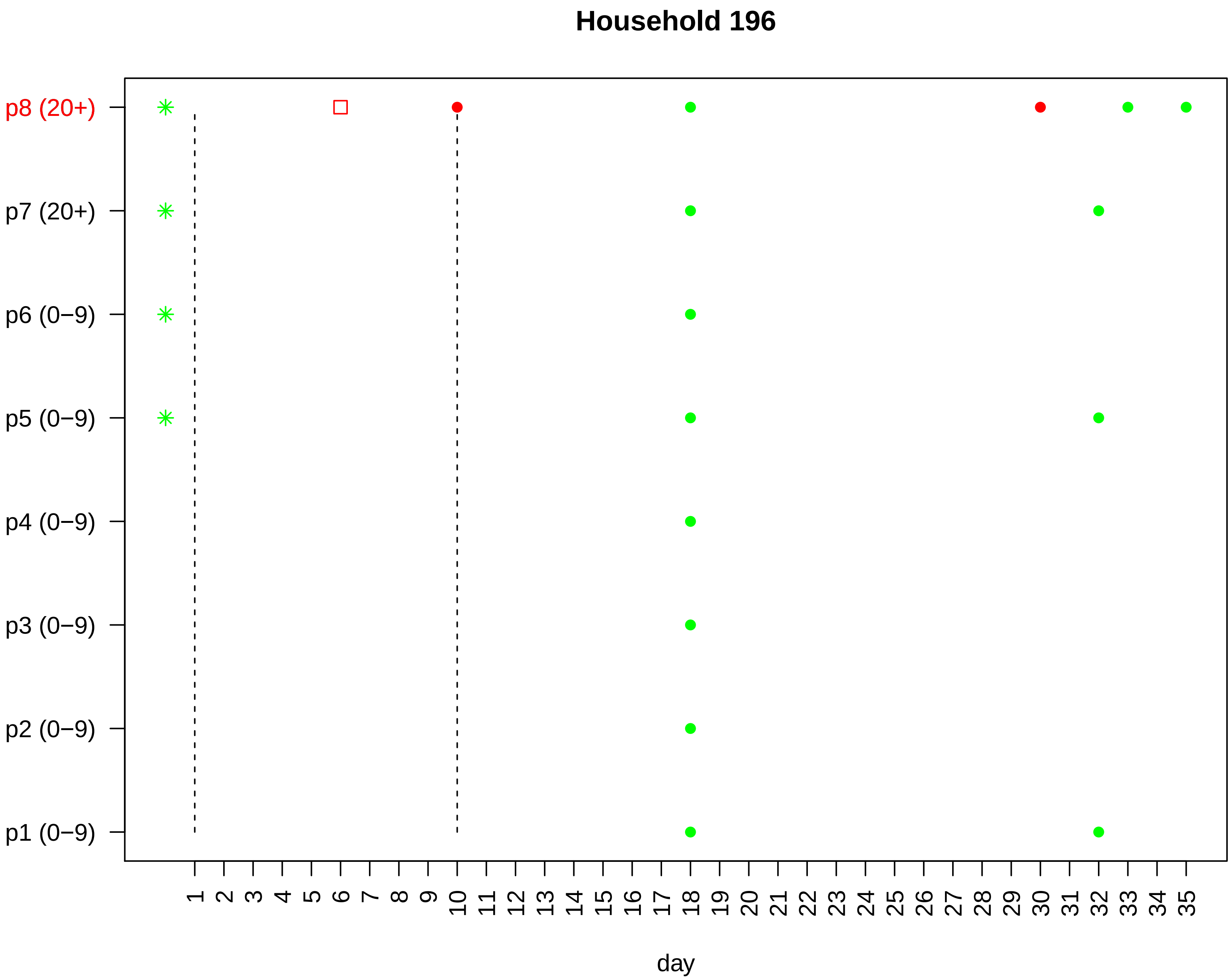

Household 196
p8 (20+)
p7 (20+)
p6 (0−9)
p5 (0−9)
p4 (0−9)
p3 (0−9)
p2 (0−9)
p1 (0−9)
1 2 3 4 5 6 7 8 9 10 11 12 13 14 15 16 17 18 19 20 21 22 23 24 25 26 27 28 29 30 31 32 33 34 35
day

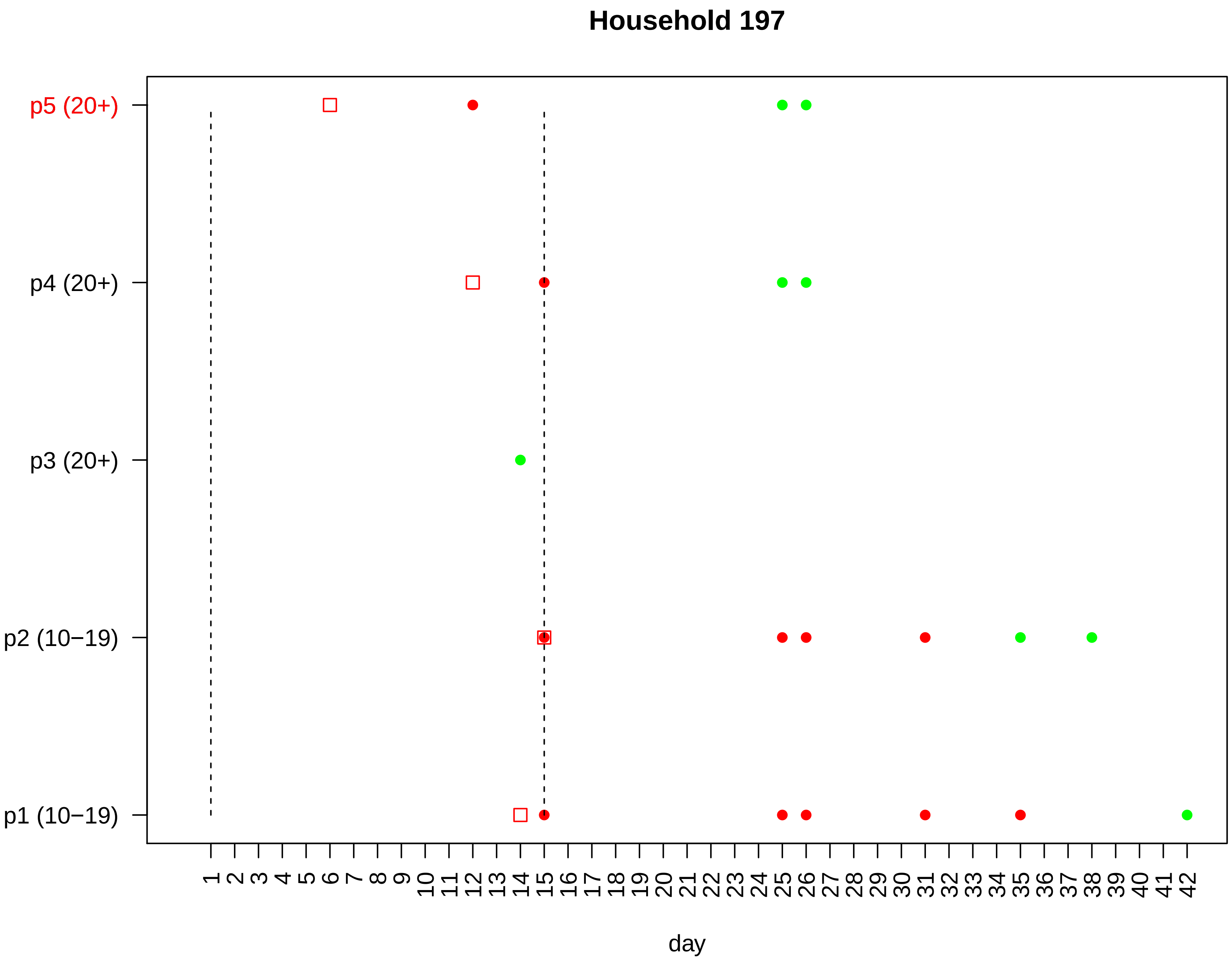

Household 197
p5 (20+)
p4 (20+)
p3 (20+)
p2 (10−19)
p1 (10−19)
1 2 3 4 5 6 7 8 9 10 11 12 13 14 15 16 17 18 19 20 21 22 23 24 25 26 27 28 29 30 31 32 33 34 35 36 37 38 39 40 41 42
day

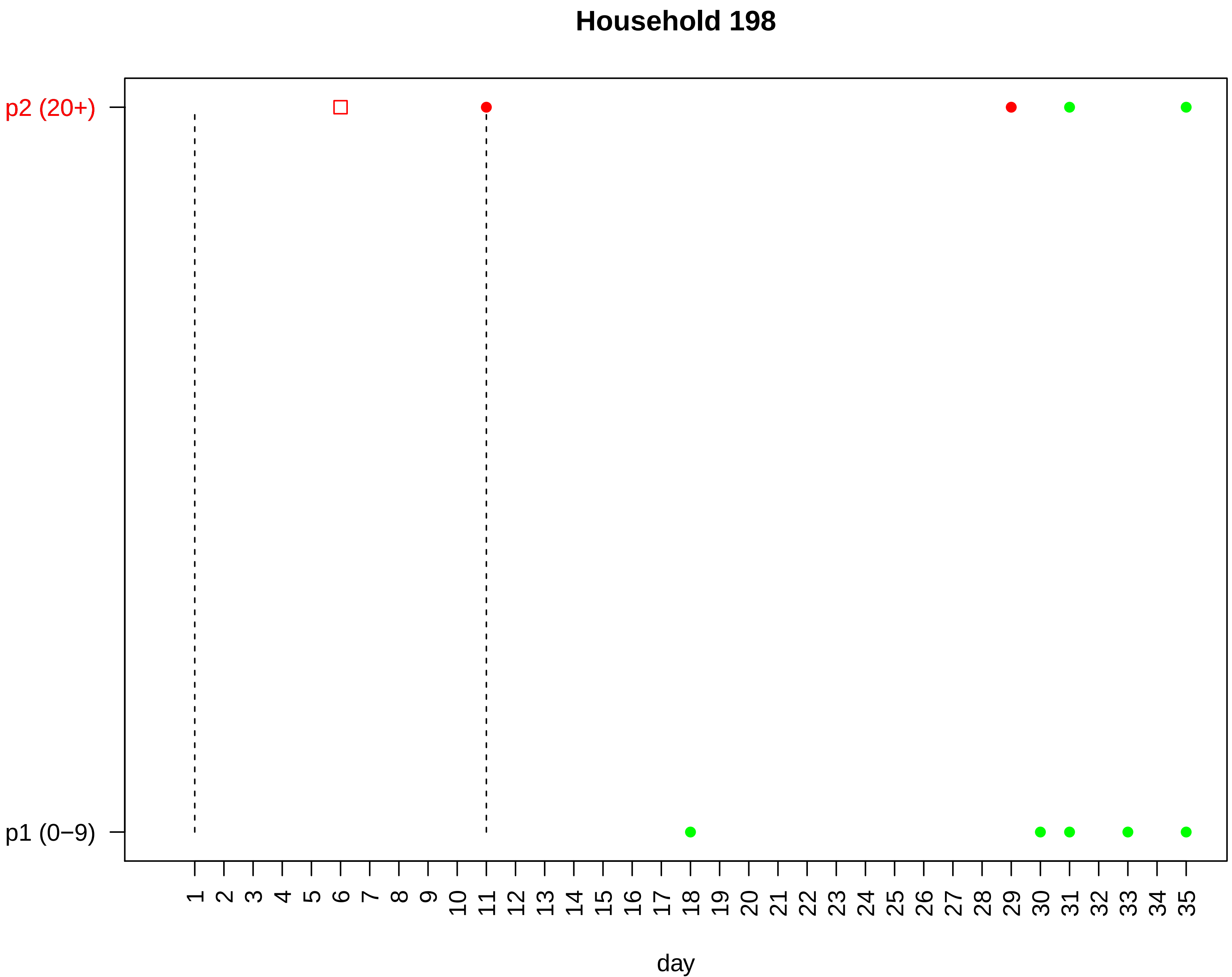

Household 198
p2 (20+)
p1 (0−9)
1 2 3 4 5 6 7 8 9 10 11 12 13 14 15 16 17 18 19 20 21 22 23 24 25 26 27 28 29 30 31 32 33 34 35
day

#### Household 199

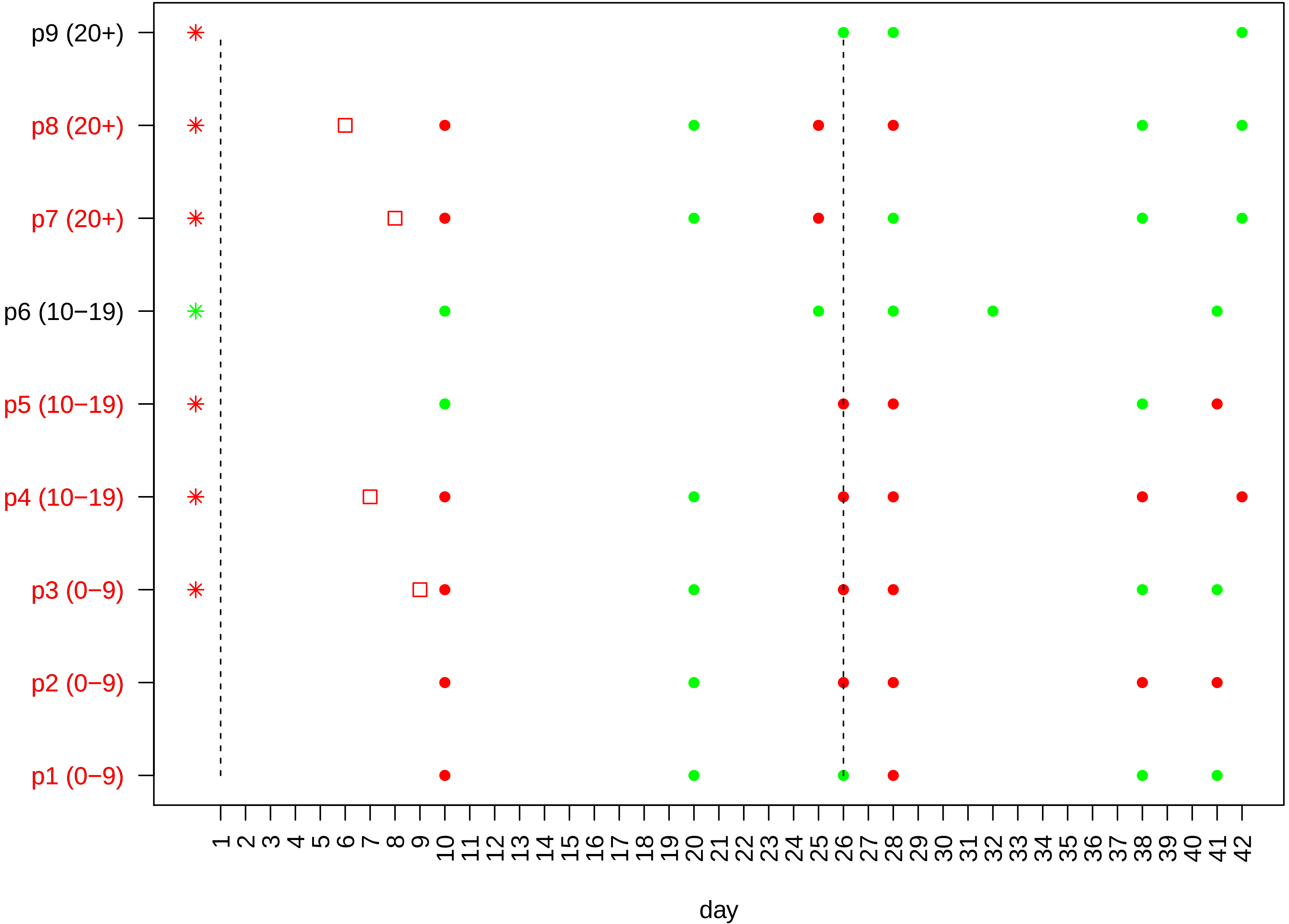

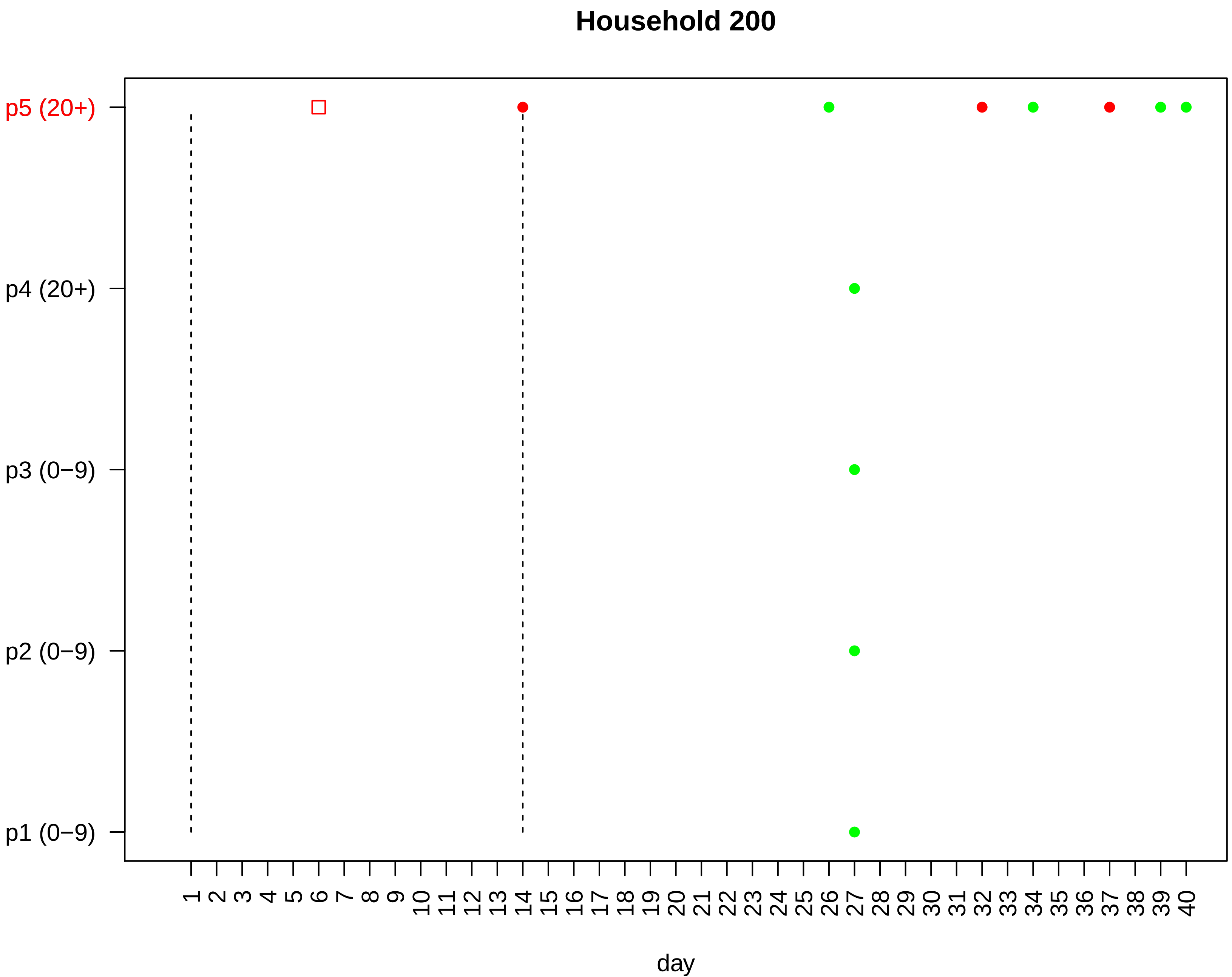

Household 200
p5 (20+)
p4 (20+)
p3 (0−9)
p2 (0−9)
p1 (0−9)
1 2 3 4 5 6 7 8 9 10 11 12 13 14 15 16 17 18 19 20 21 22 23 24 25 26 27 28 29 30 31 32 33 34 35 36 37 38 39 40
day

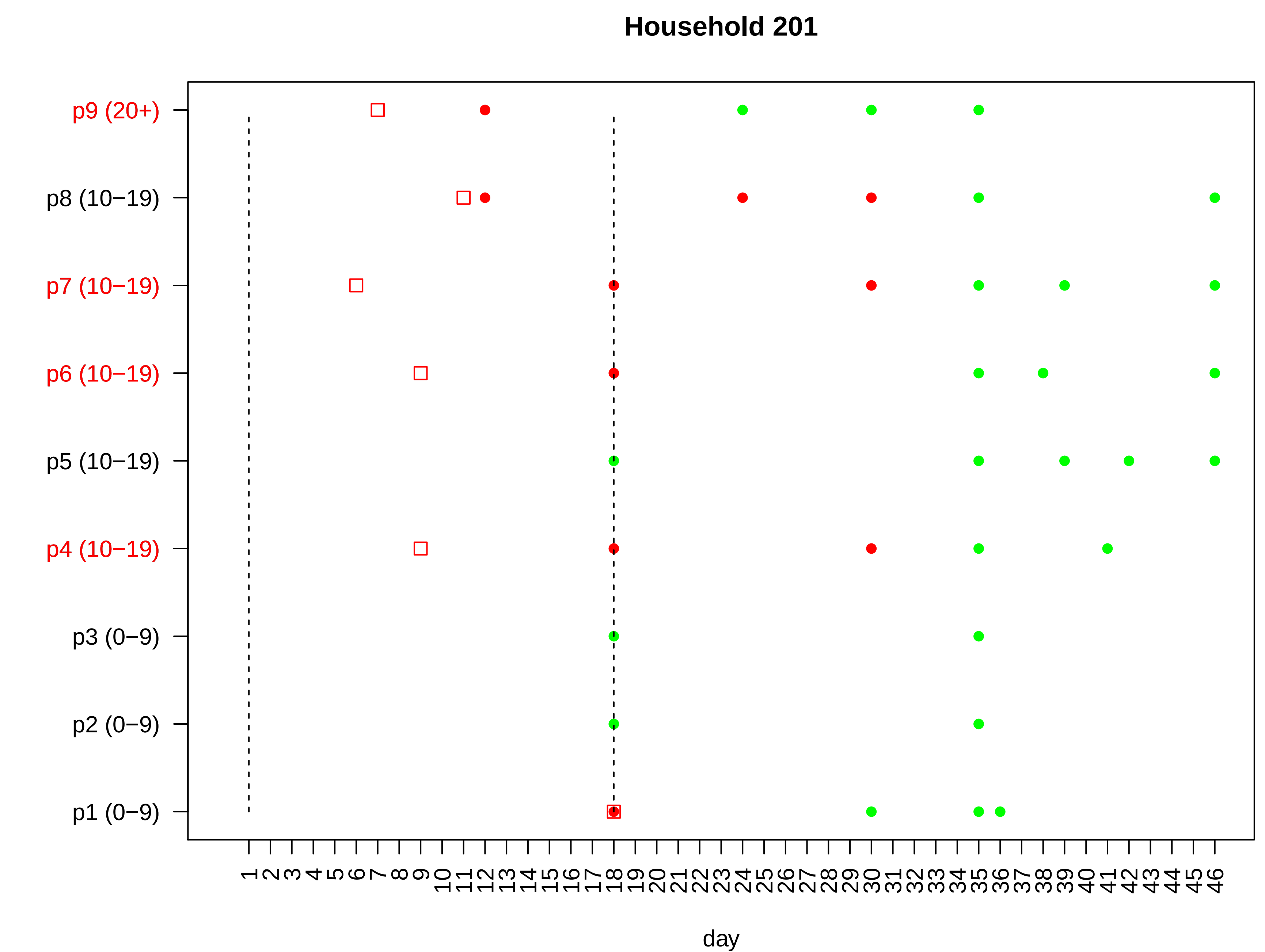

Household 201
p9 (20+)
p8 (10−19)
p7 (10−19)
p6 (10−19)
p5 (10−19)
p4 (10−19)
p3 (0−9)
p2 (0−9)
p1 (0−9)
1 2 3 4 5 6 7 8 9 10 11 12 13 14 15 16 17 18 19 20 21 22 23 24 25 26 27 28 29 30 31 32 33 34 35 36 37 38 39 40 41 42 43 44 45 46
day

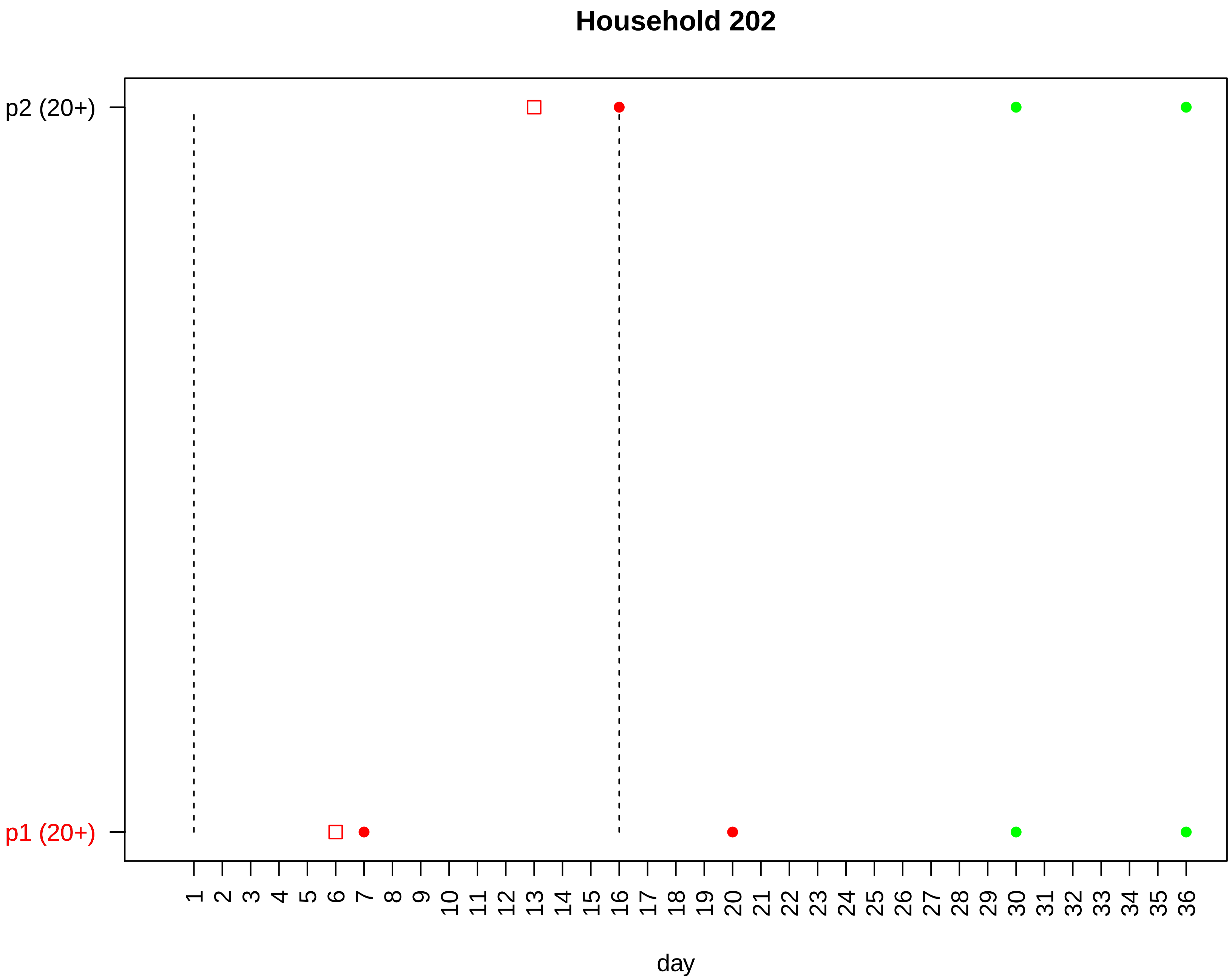

Household 202
p2 (20+)
p1 (20+)
day

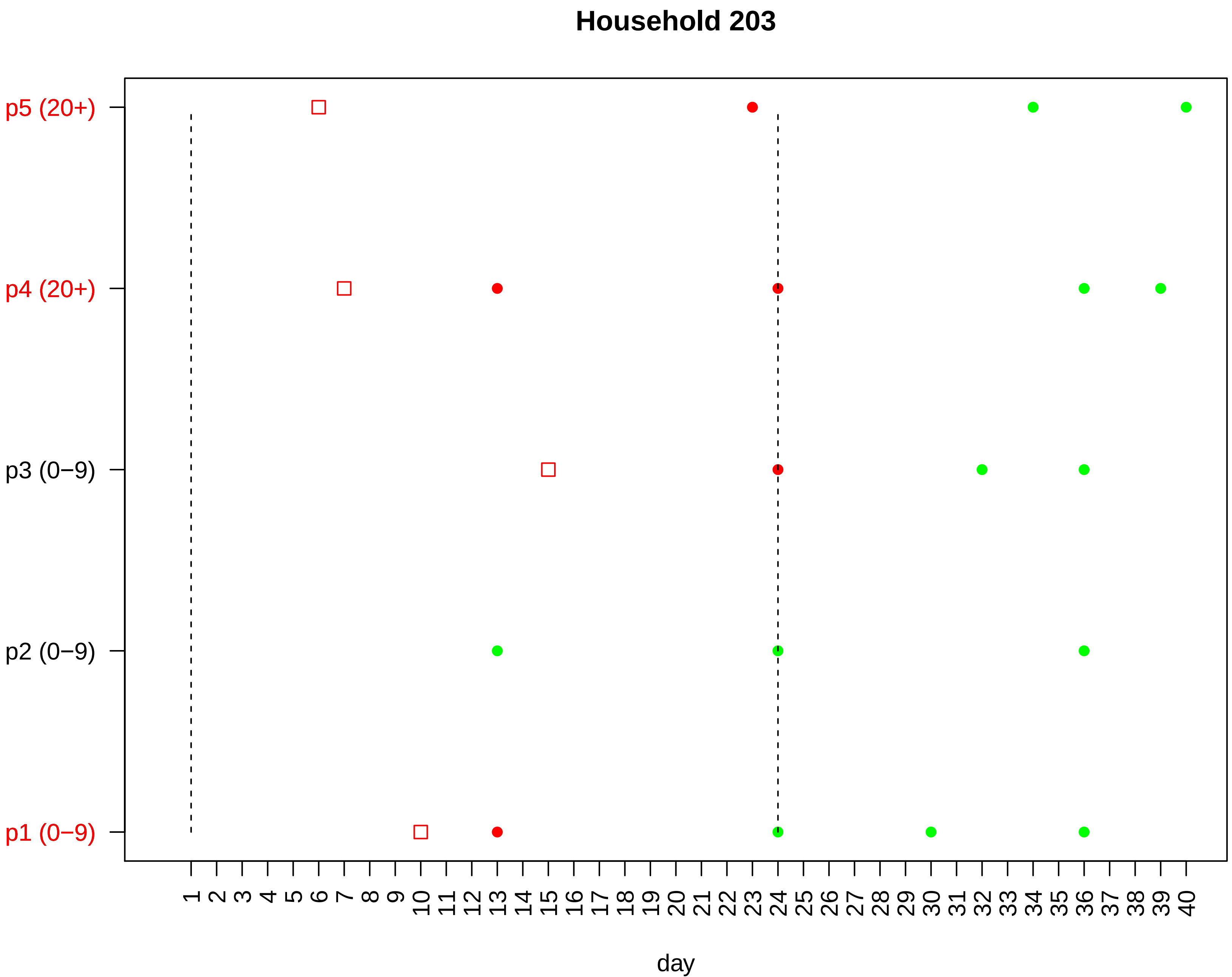

Household 203
p5 (20+)
p4 (20+)
p3 (0−9)
p2 (0−9)
p1 (0−9)
1 2 3 4 5 6 7 8 9 10 11 12 13 14 15 16 17 18 19 20 21 22 23 24 25 26 27 28 29 30 31 32 33 34 35 36 37 38 39 40
day

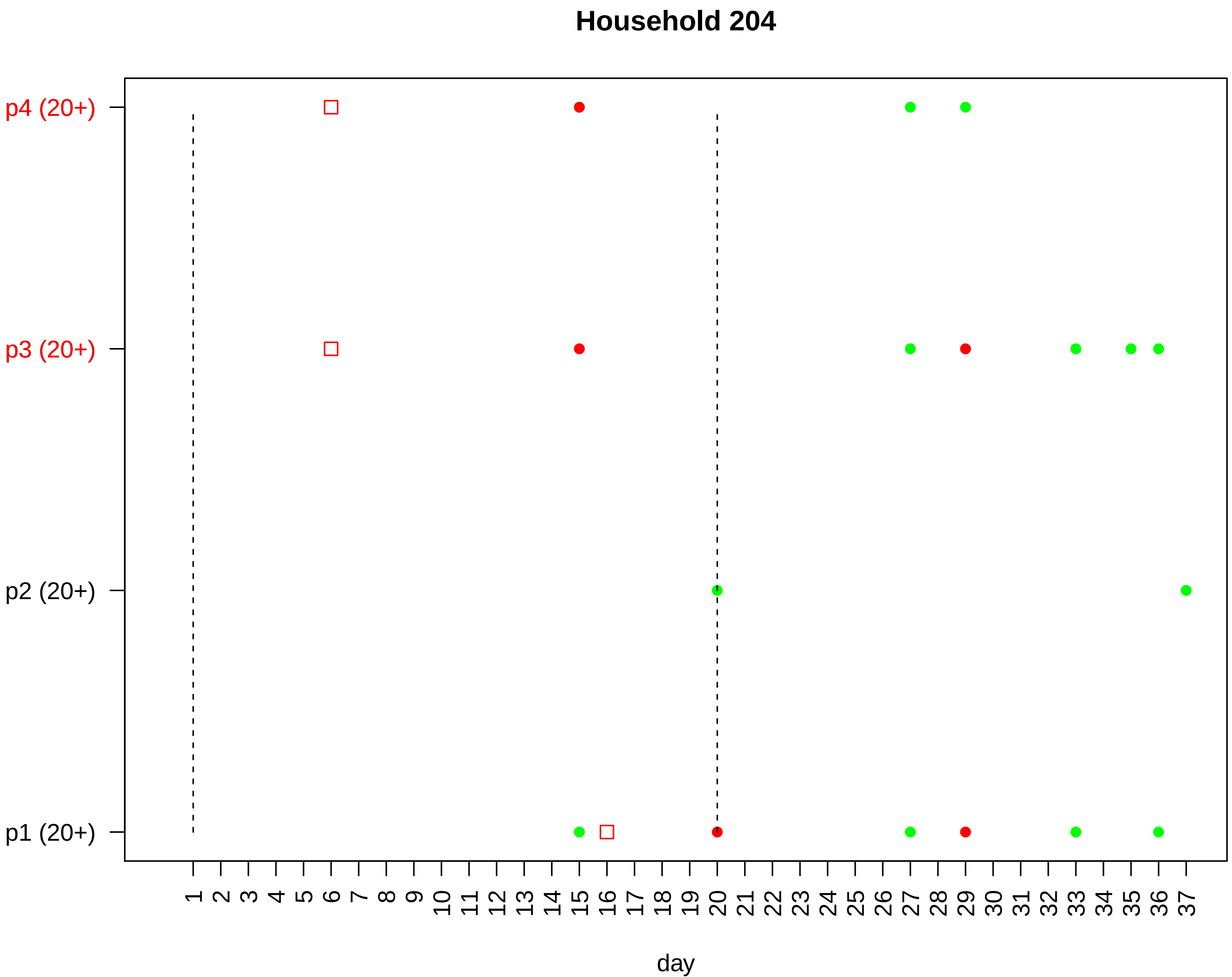
Household 204
p4 (20+)
p3 (20+)
p2 (20+)
p1 (20+)
1 2 3 4 5 6 7 8 9 10 11 12 13 14 15 16 17 18 19 20 21 22 23 24 25 26 27 28 29 30 31 32 33 34 35 36 37
day

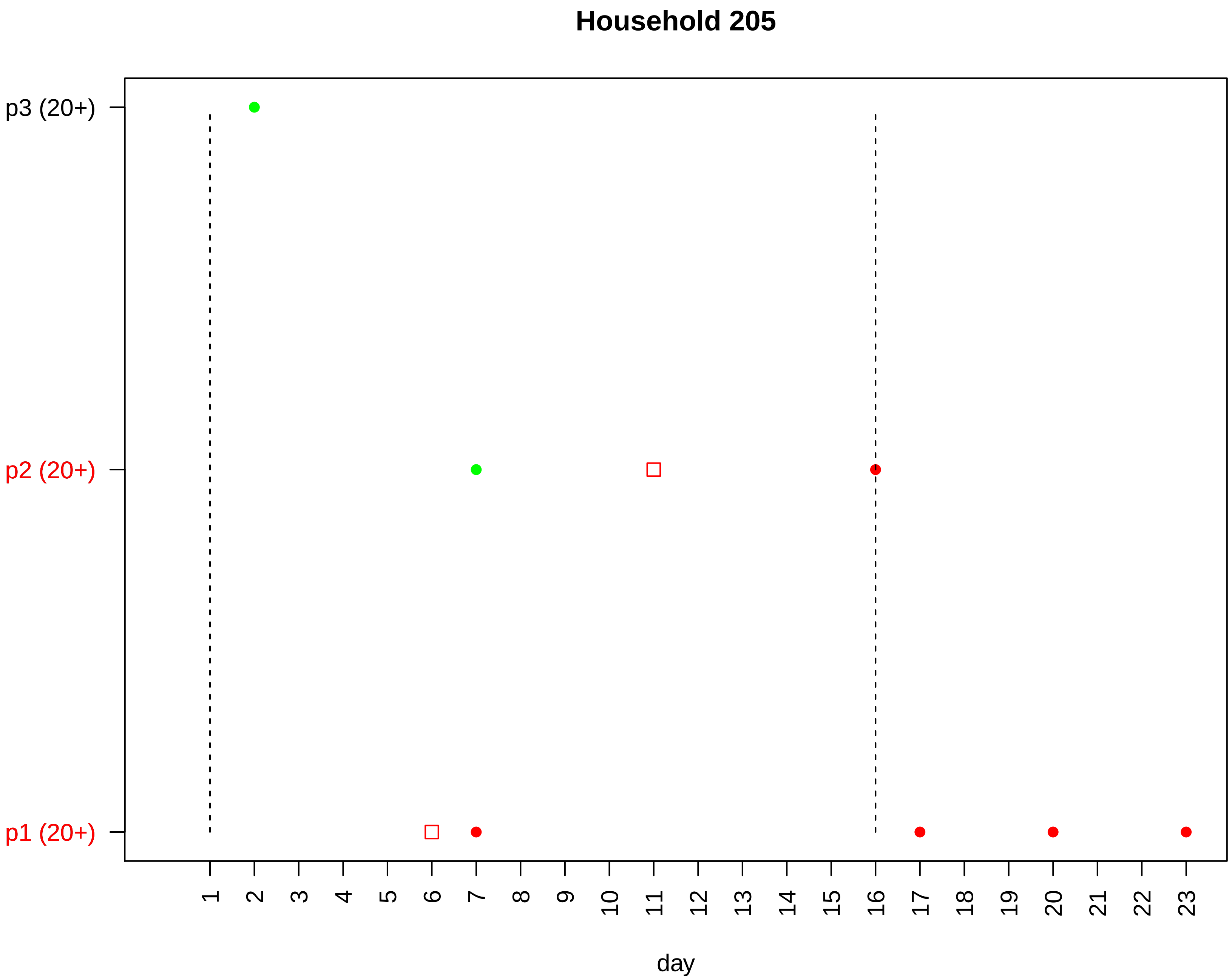

Household 205
p3 (20+)
p2 (20+)
p1 (20+)
day

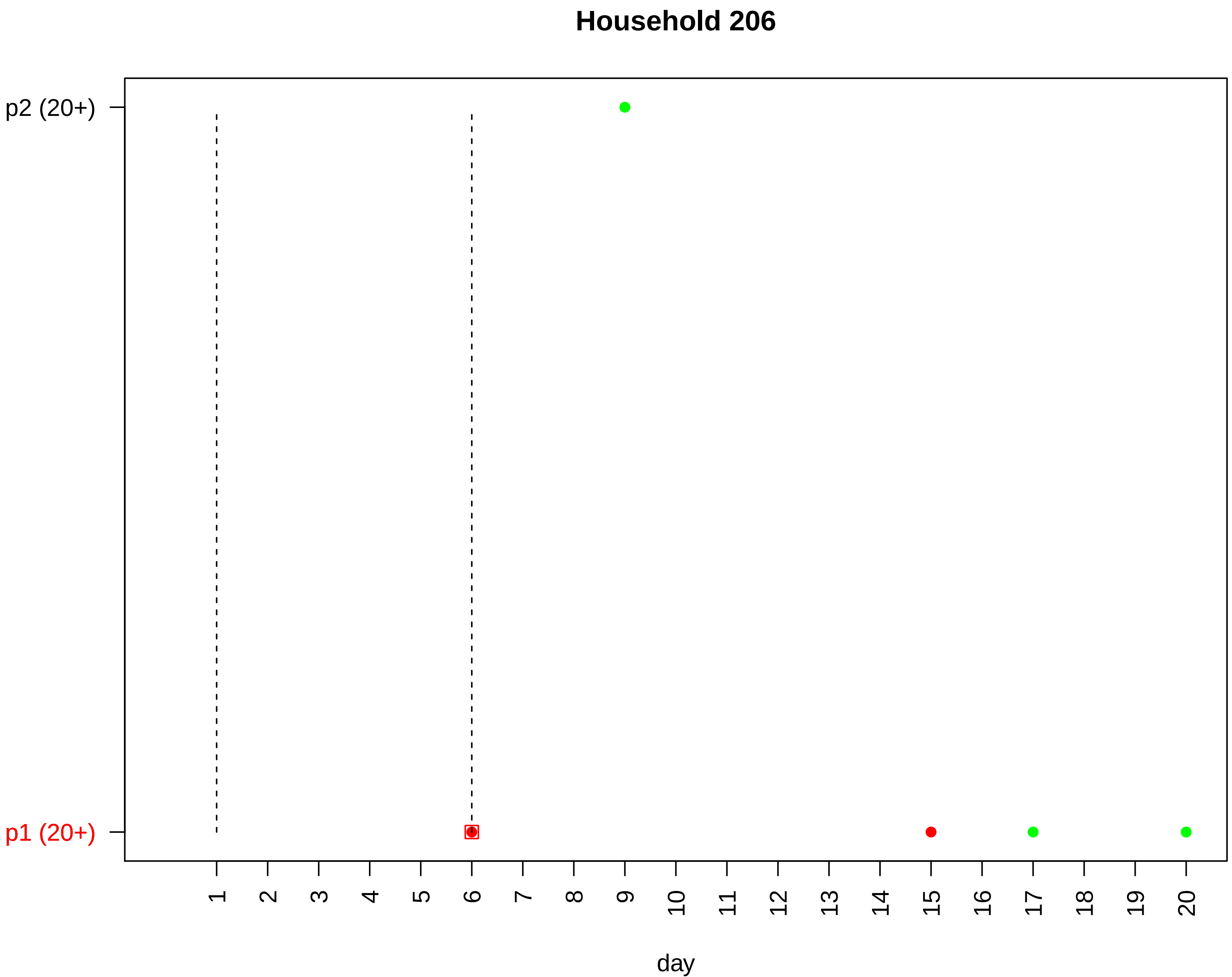
Household 206
p2 (20+)
p1 (20+)
day

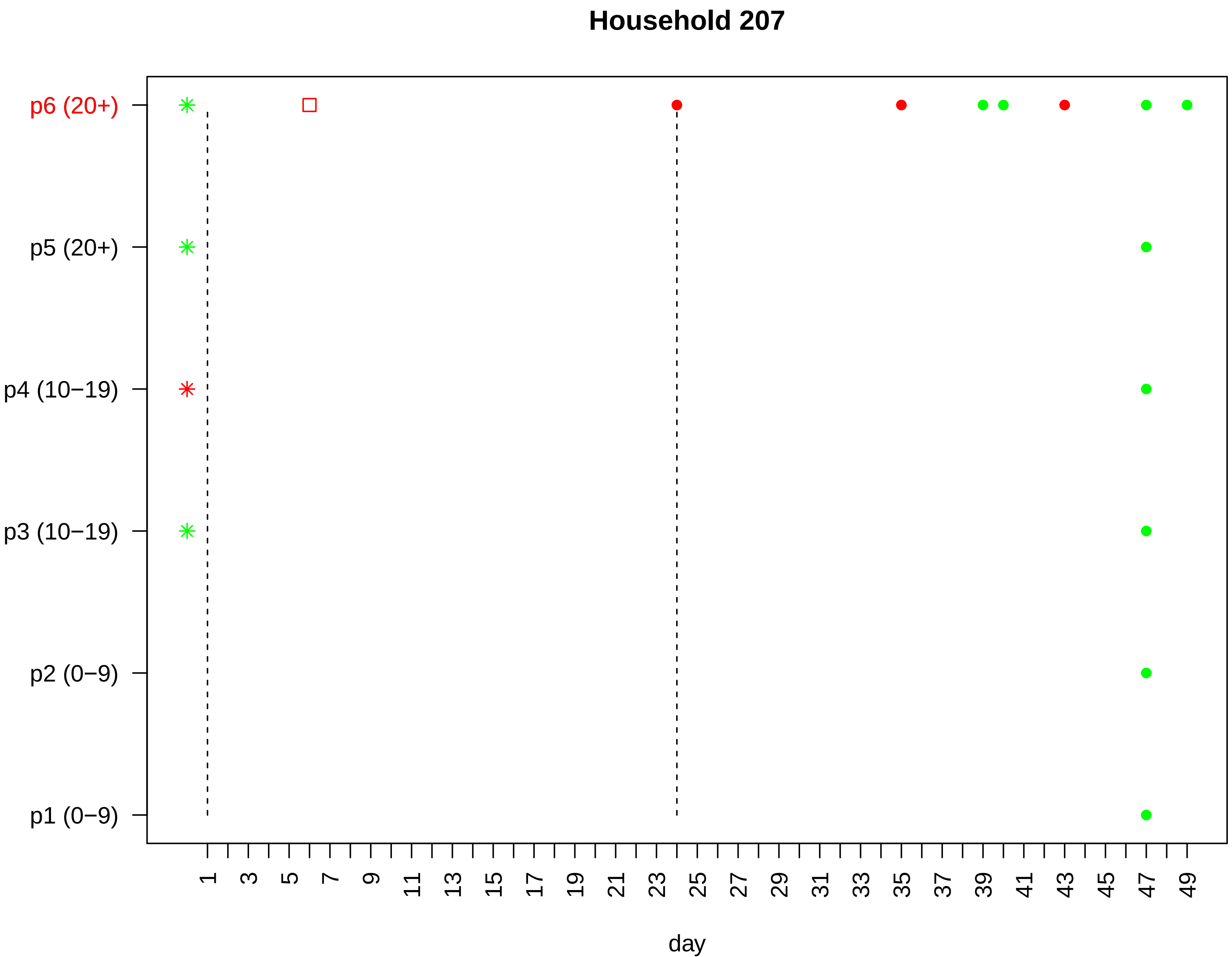

Household 207
p6 (20+)
p5 (20+)
p4 (10−19)
p3 (10−19)
p2 (0−9)
p1 (0−9)
1
3
5
7
9
11
13
15
17
19
21
23
25
27
29
31
33
35
37
39
41
43
45
47
49
day

Household 208

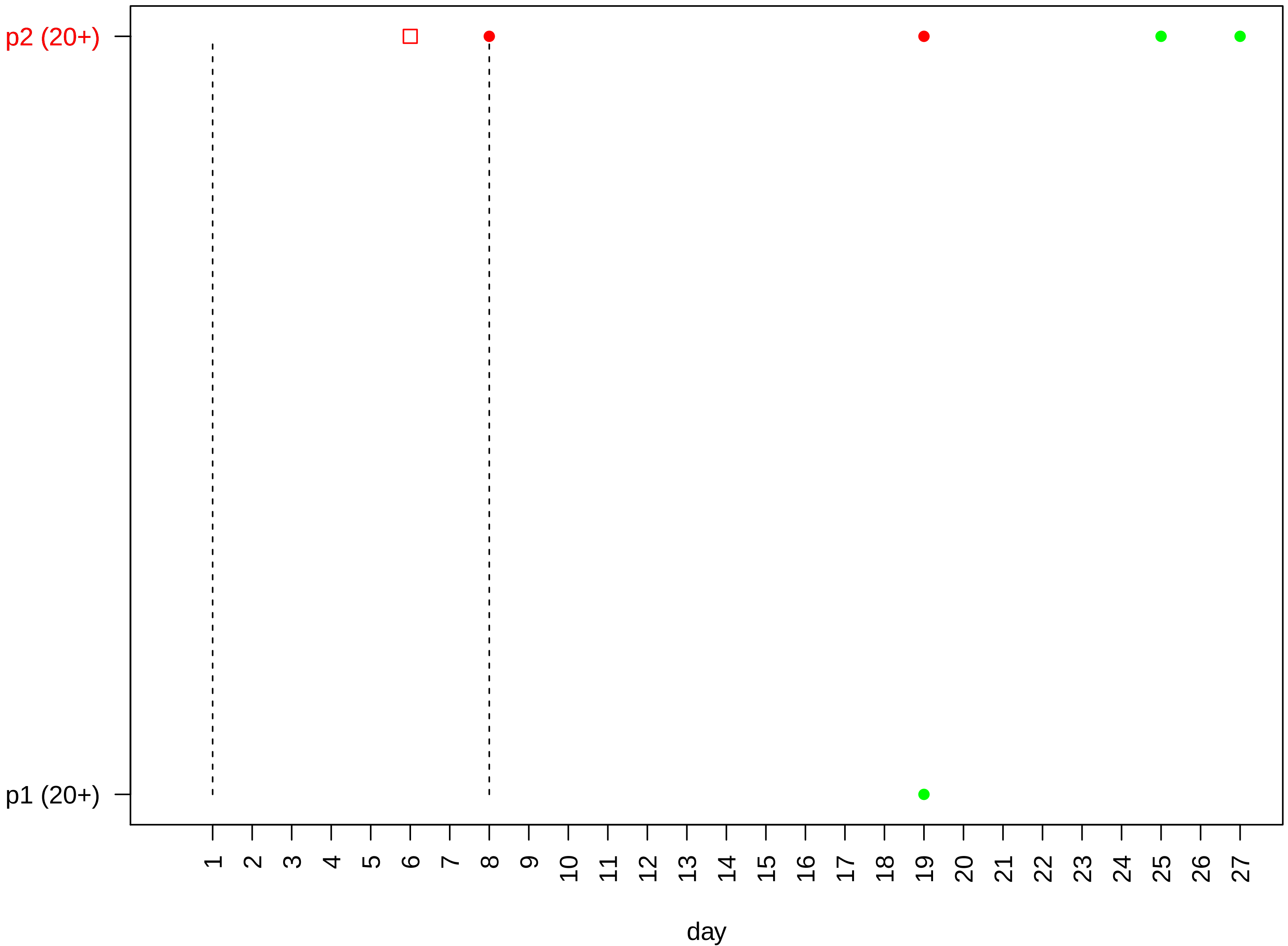

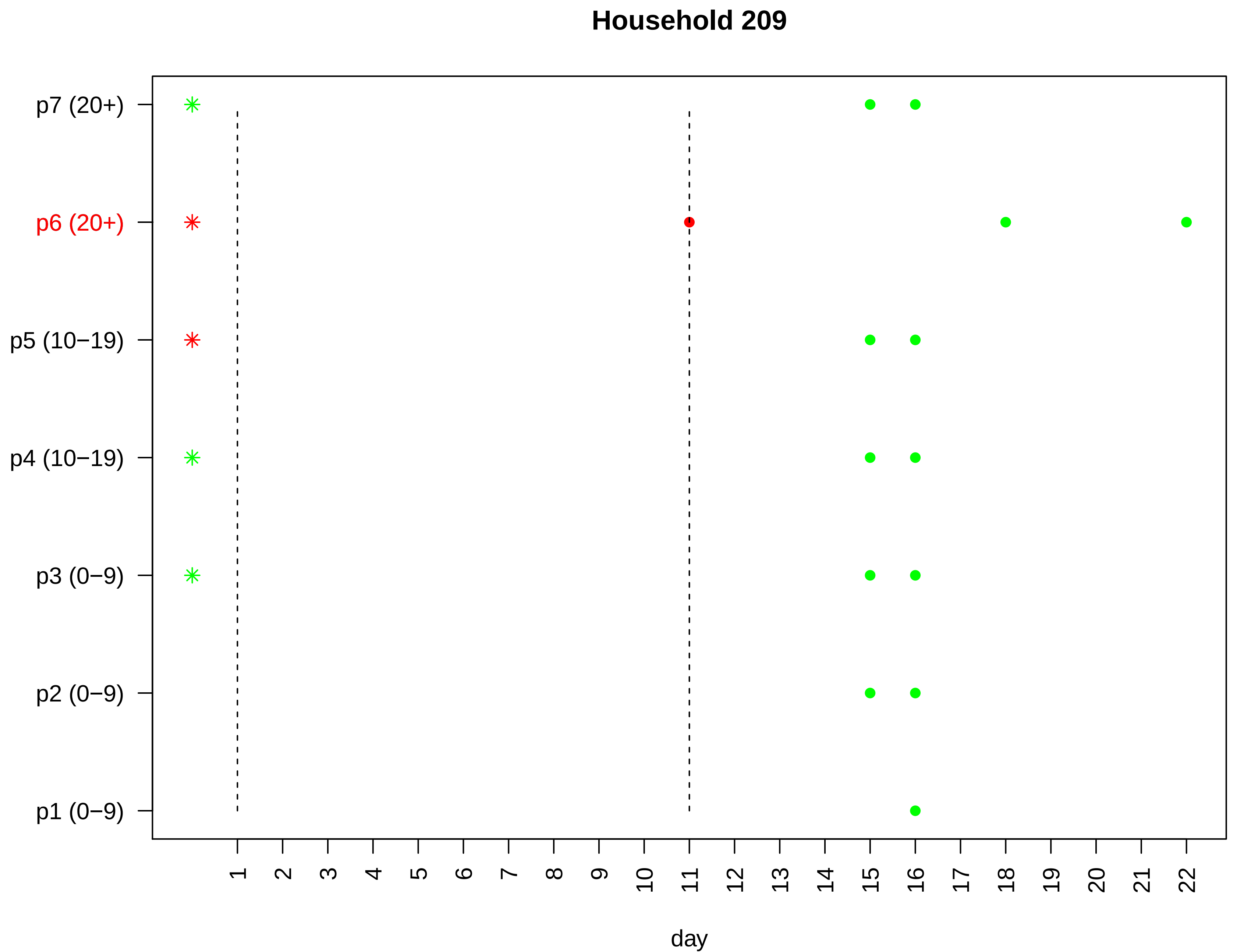
Household 209
p7 (20+)
p6 (20+)
p5 (10−19)
p4 (10−19)
p3 (0−9)
p2 (0−9)
p1 (0−9)
1 2 3 4 5 6 7 8 9 10 11 12 13 14 15 16 17 18 19 20 21 22
day

### Household 210

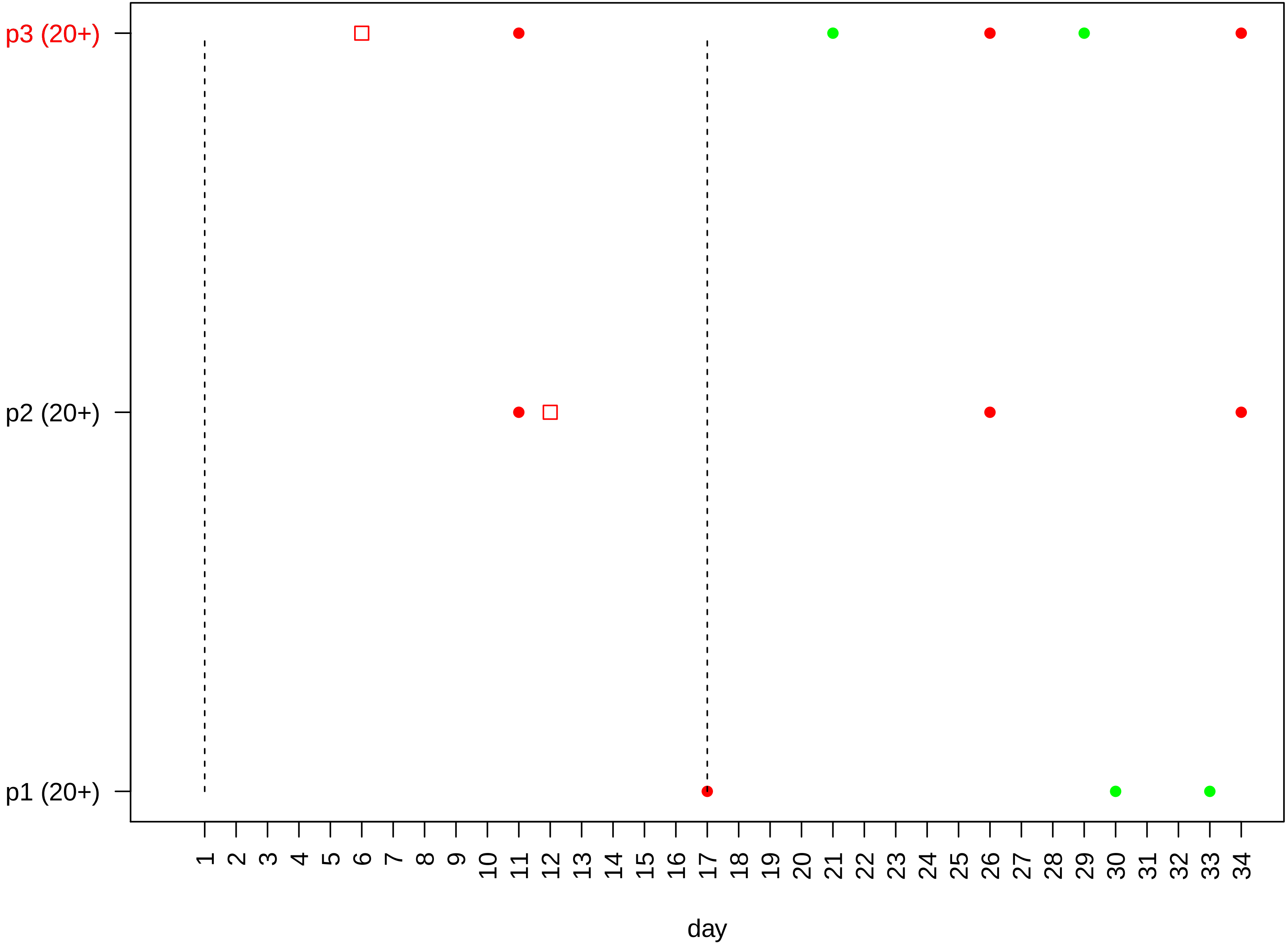

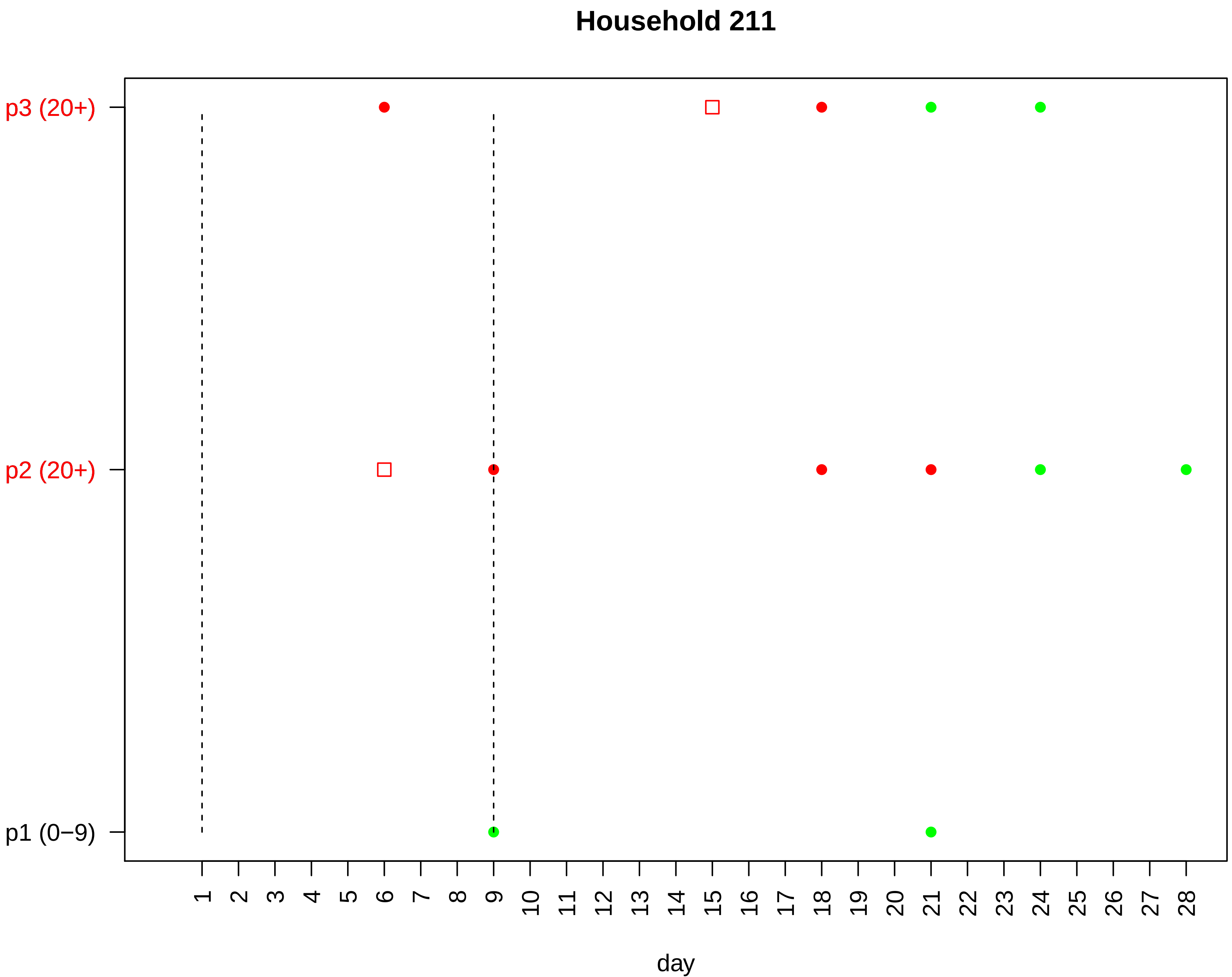

Household 211
p3 (20+)
p2 (20+)
p1 (0−9)
1 2 3 4 5 6 7 8 9 10 11 12 13 14 15 16 17 18 19 20 21 22 23 24 25 26 27 28
day

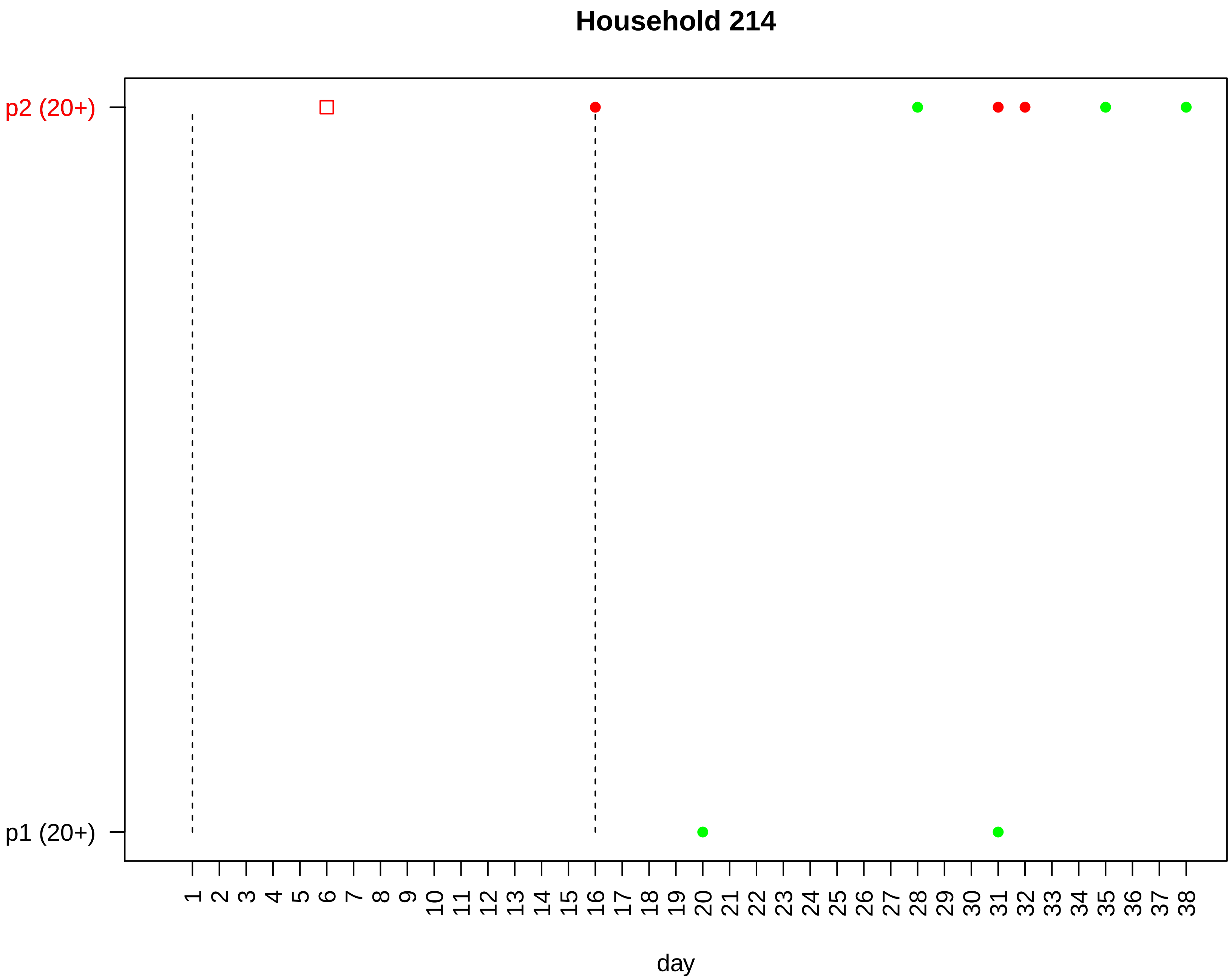
Household 214
p2 (20+)
p1 (20+)
1 2 3 4 5 6 7 8 9 10 11 12 13 14 15 16 17 18 19 20 21 22 23 24 25 26 27 28 29 30 31 32 33 34 35 36 37 38
day

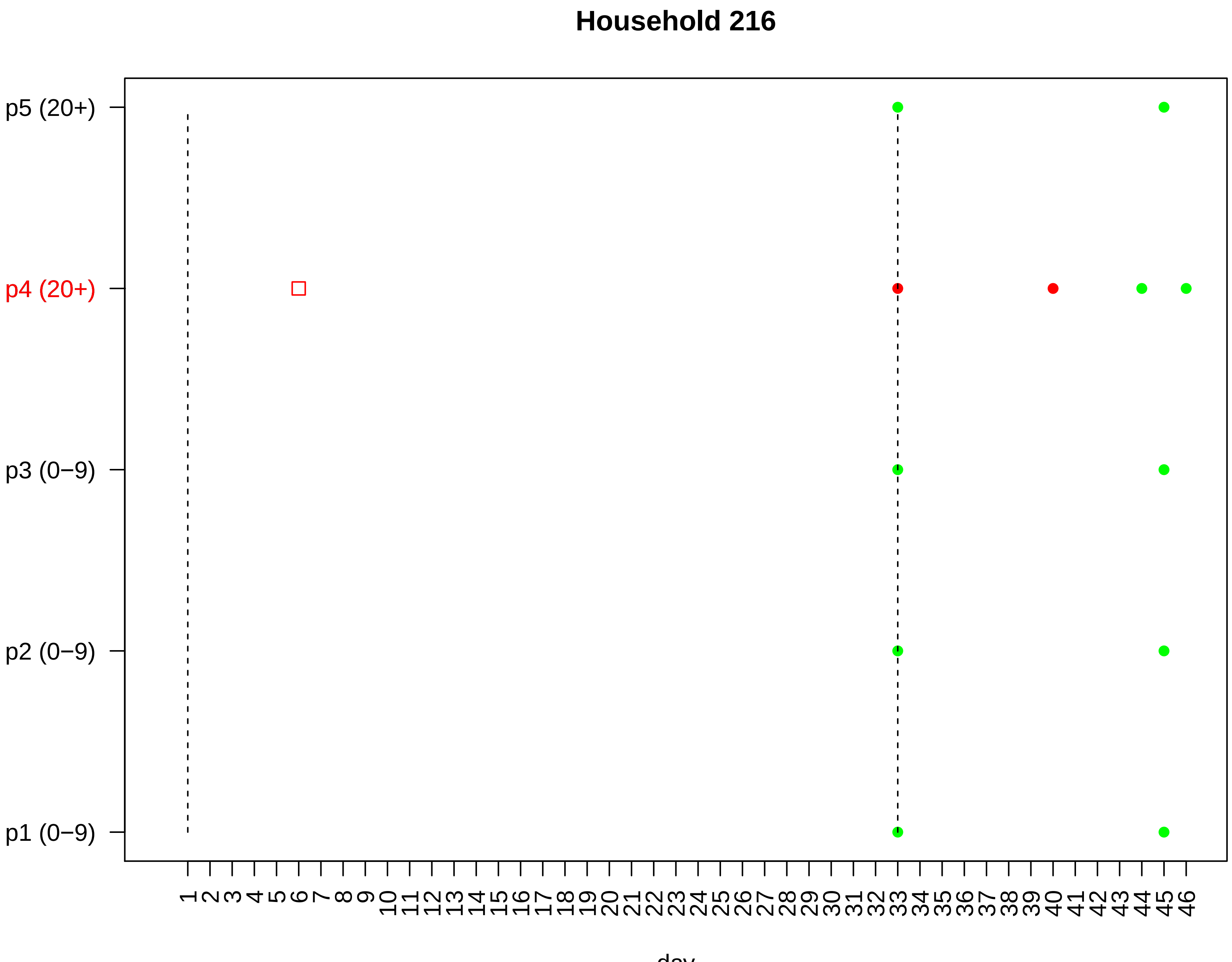
Household 216
p5 (20+)
p4 (20+)
p3 (0−9)
p2 (0−9)
p1 (0−9)
1 2 3 4 5 6 7 8 9 10 11 12 13 14 15 16 17 18 19 20 21 22 23 24 25 26 27 28 29 30 31 32 33 34 35 36 37 38 39 40 41 42 43 44 45 46
day

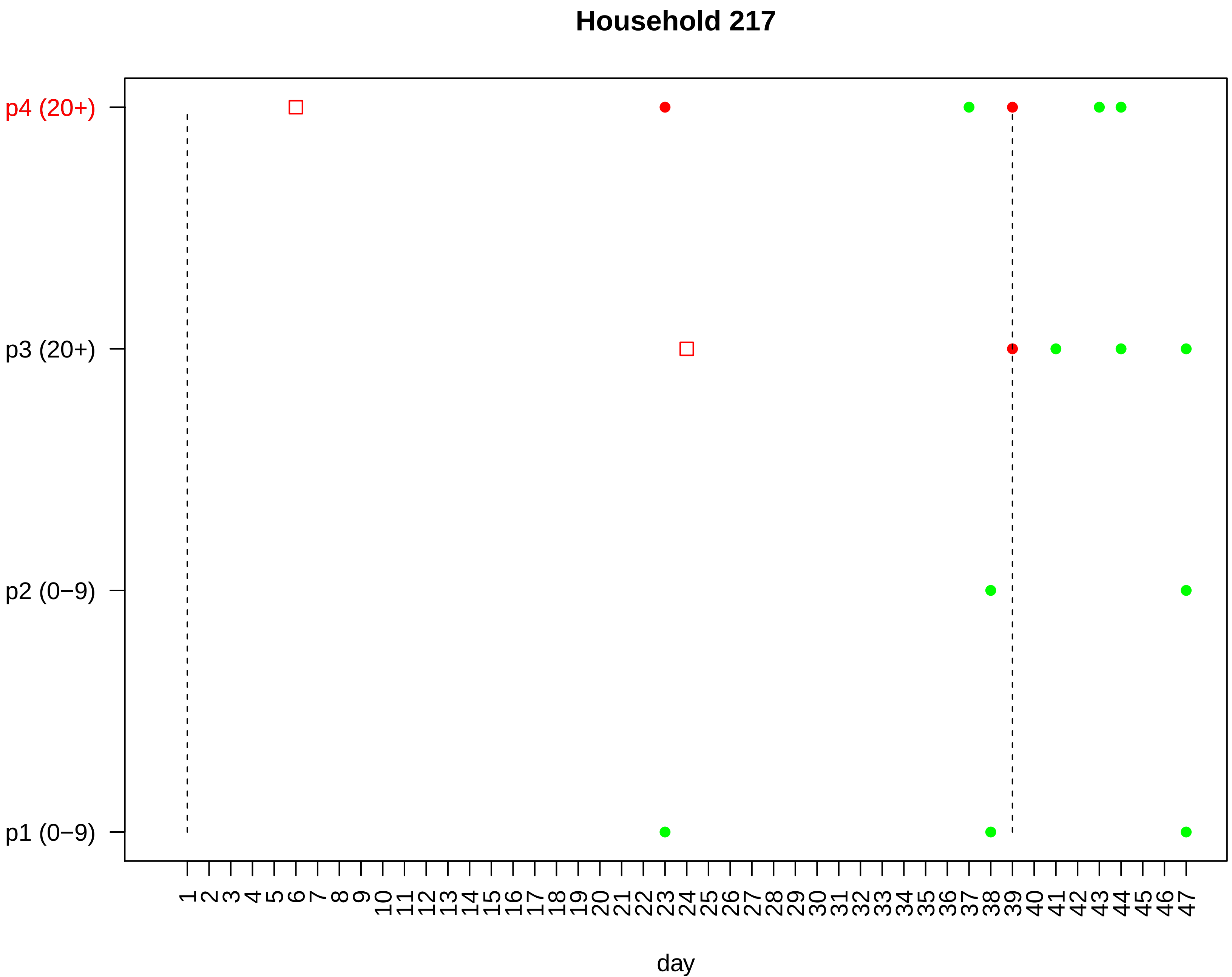

Household 217
p4 (20+)
p3 (20+)
p2 (0−9)
p1 (0−9)
1 2 3 4 5 6 7 8 9 10 11 12 13 14 15 16 17 18 19 20 21 22 23 24 25 26 27 28 29 30 31 32 33 34 35 36 37 38 39 40 41 42 43 44 45 46 47
day

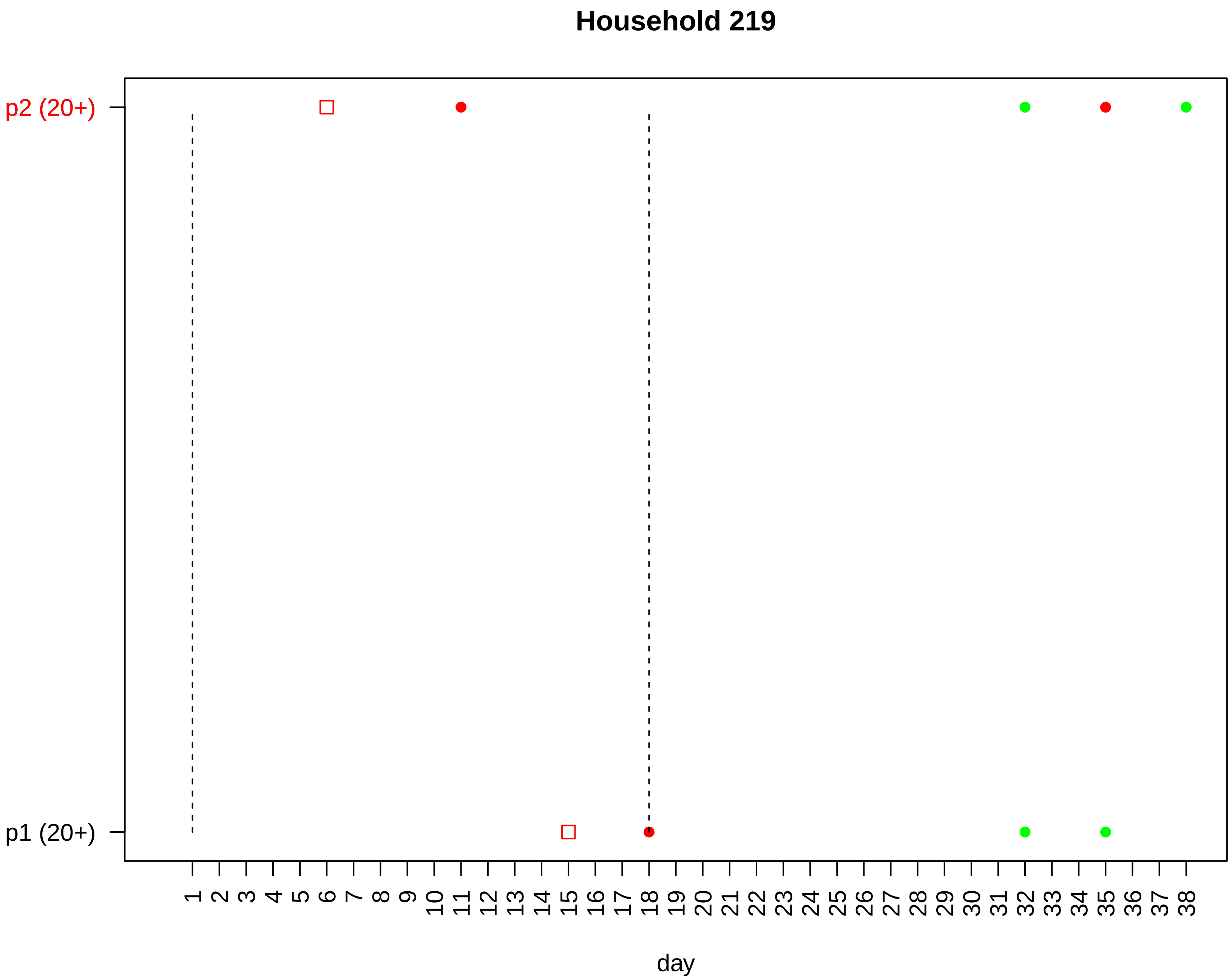
Household 219
p2 (20+)
p1 (20+)
1 2 3 4 5 6 7 8 9 10 11 12 13 14 15 16 17 18 19 20 21 22 23 24 25 26 27 28 29 30 31 32 33 34 35 36 37 38
day

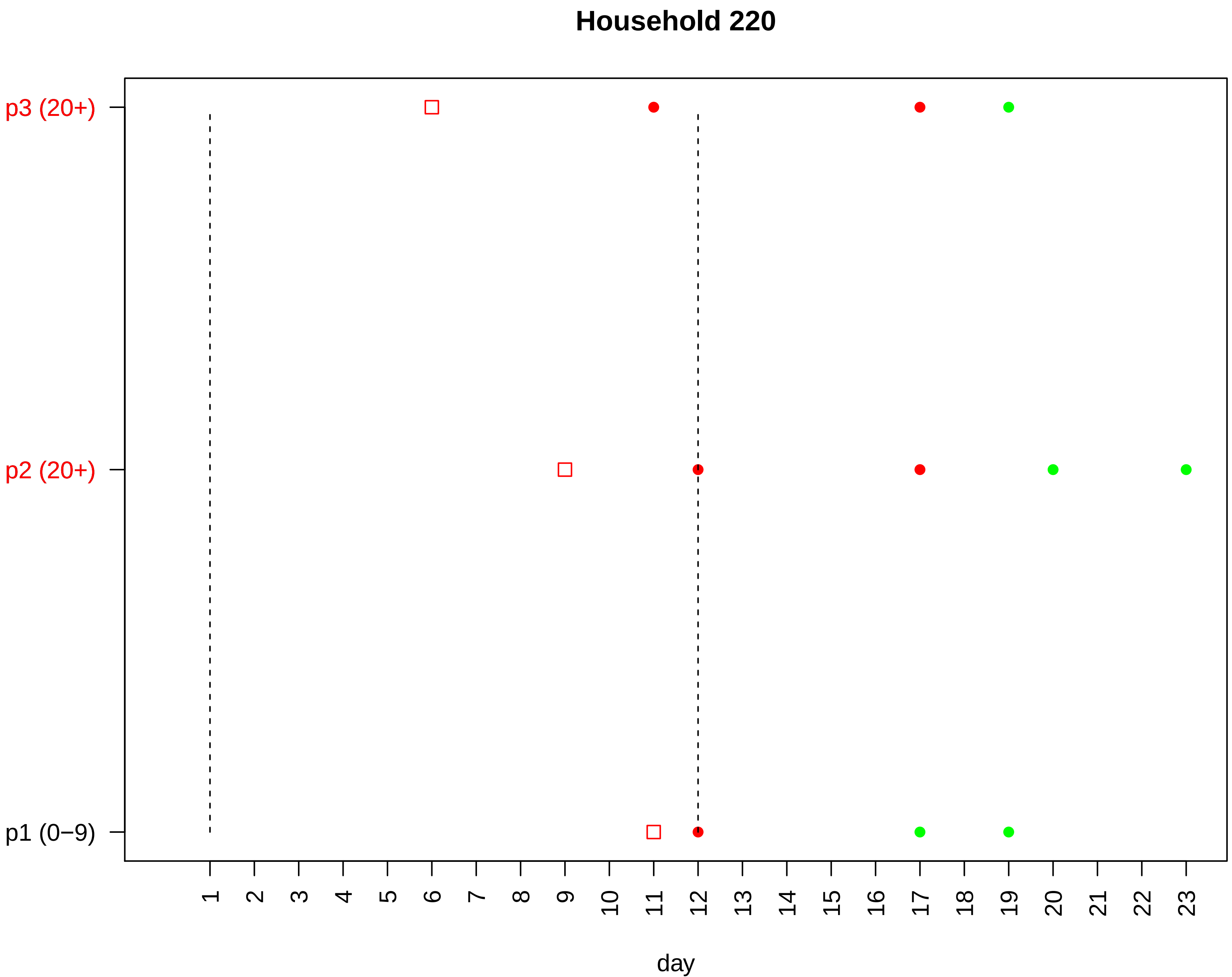
Household 220
p3 (20+)
p2 (20+)
p1 (0−9)
1 2 3 4 5 6 7 8 9 10 11 12 13 14 15 16 17 18 19 20 21 22 23
day

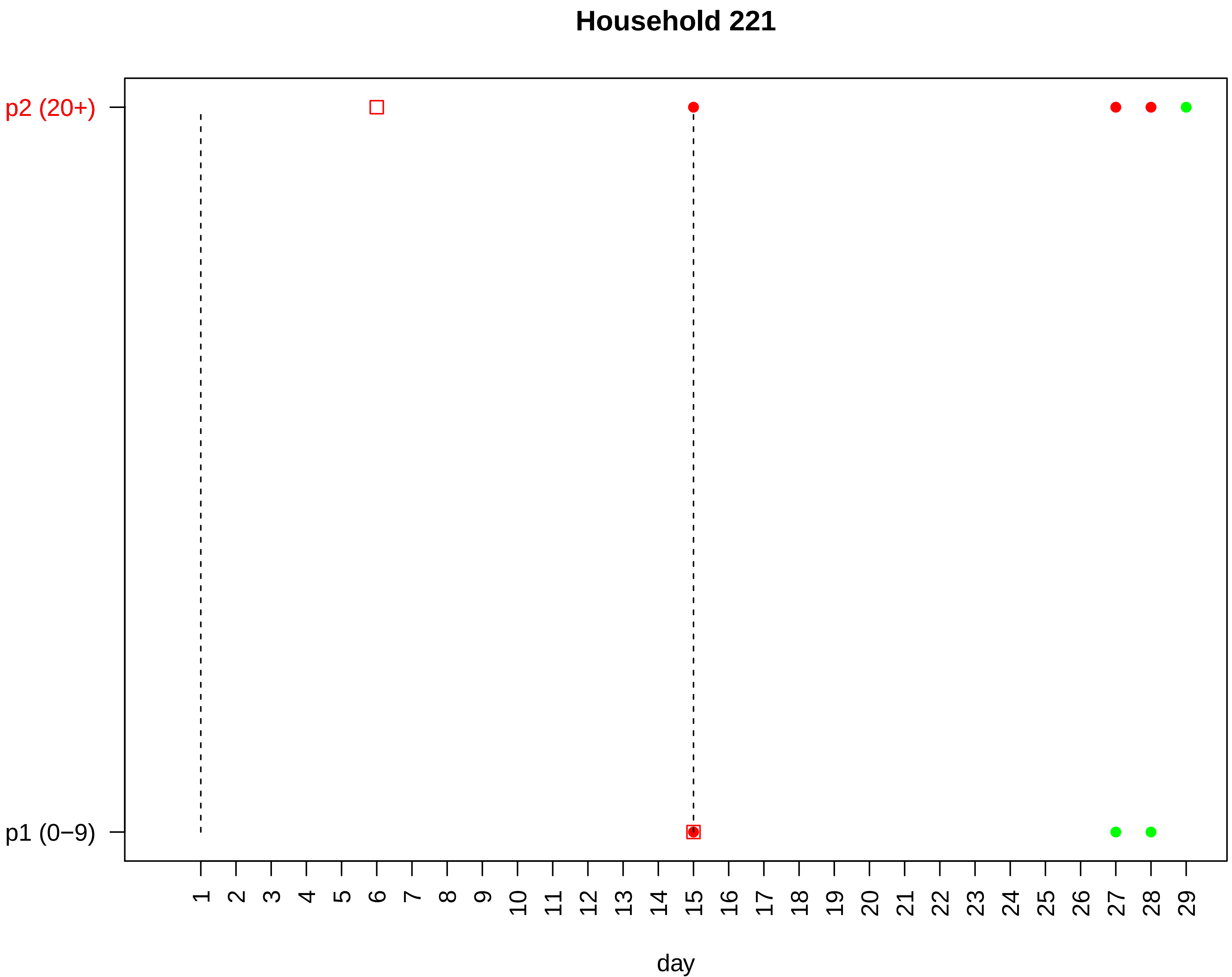
Household 221
p2 (20+)
p1 (0−9)
day

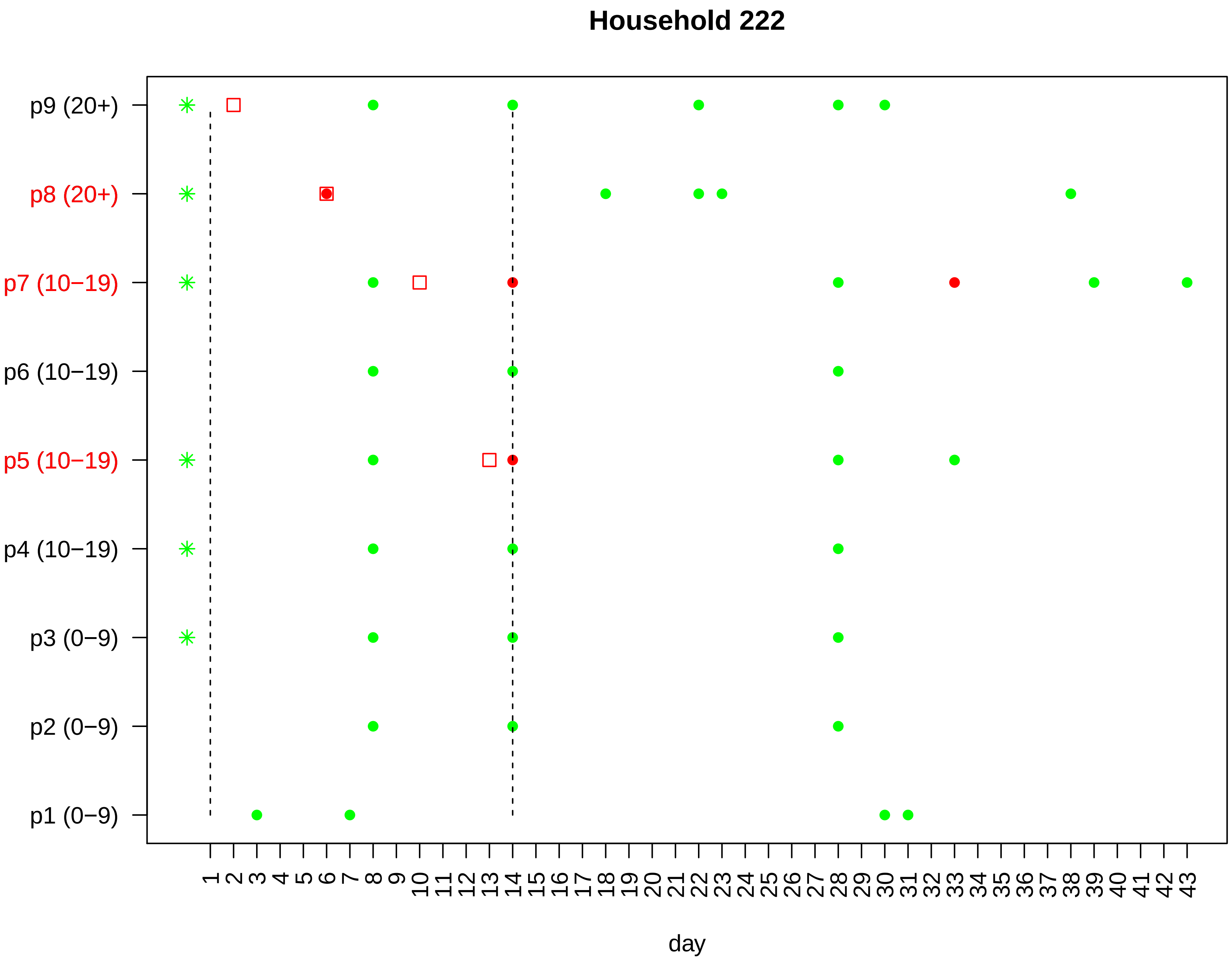

Household 222
p9 (20+)
p8 (20+)
p7 (10−19)
p6 (10−19)
p5 (10−19)
p4 (10−19)
p3 (0−9)
p2 (0−9)
p1 (0−9)
1 2 3 4 5 6 7 8 9 10 11 12 13 14 15 16 17 18 19 20 21 22 23 24 25 26 27 28 29 30 31 32 33 34 35 36 37 38 39 40 41 42 43
day

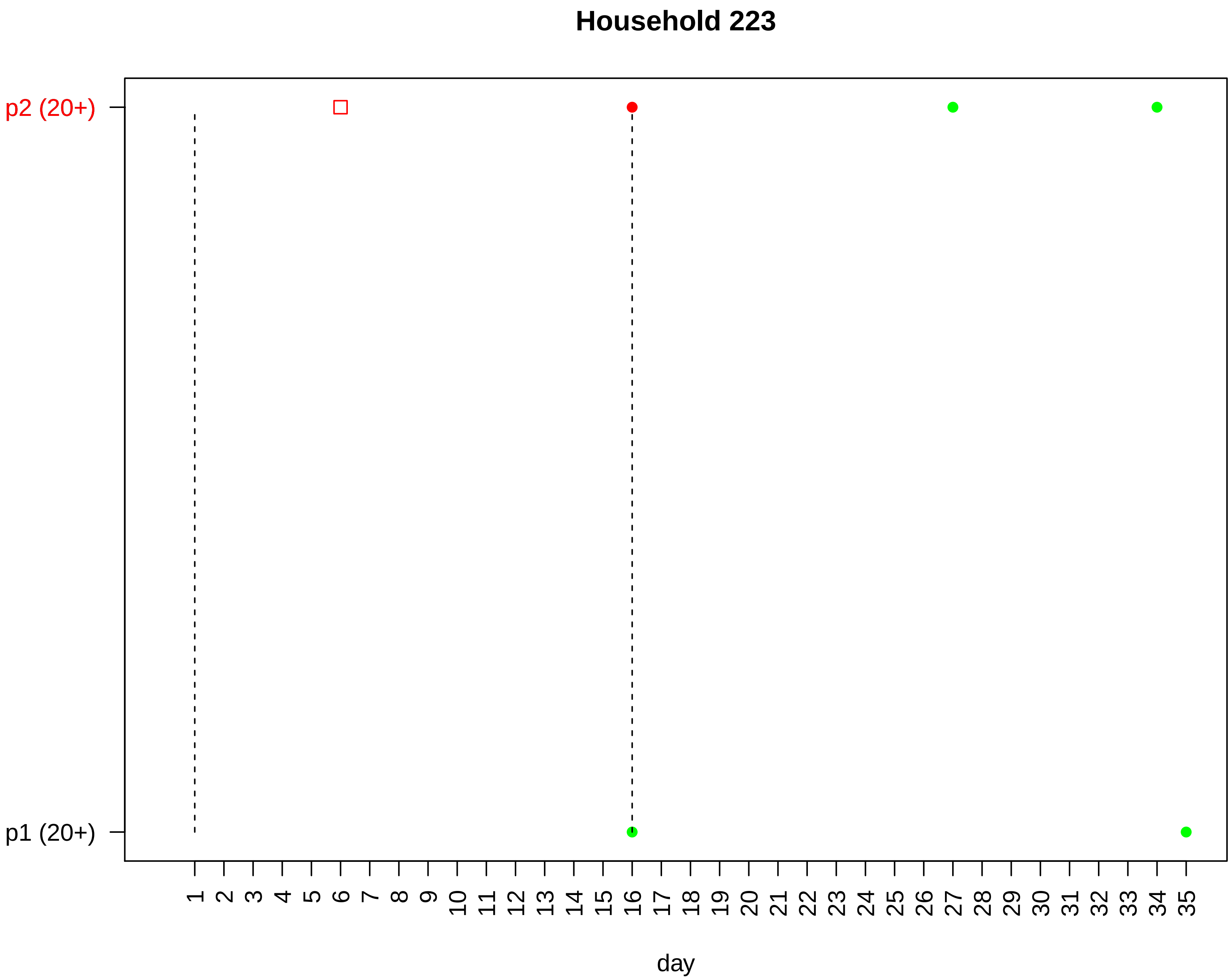
Household 223
p2 (20+)
p1 (20+)
1 2 3 4 5 6 7 8 9 10 11 12 13 14 15 16 17 18 19 20 21 22 23 24 25 26 27 28 29 30 31 32 33 34 35
day

#### Household 224

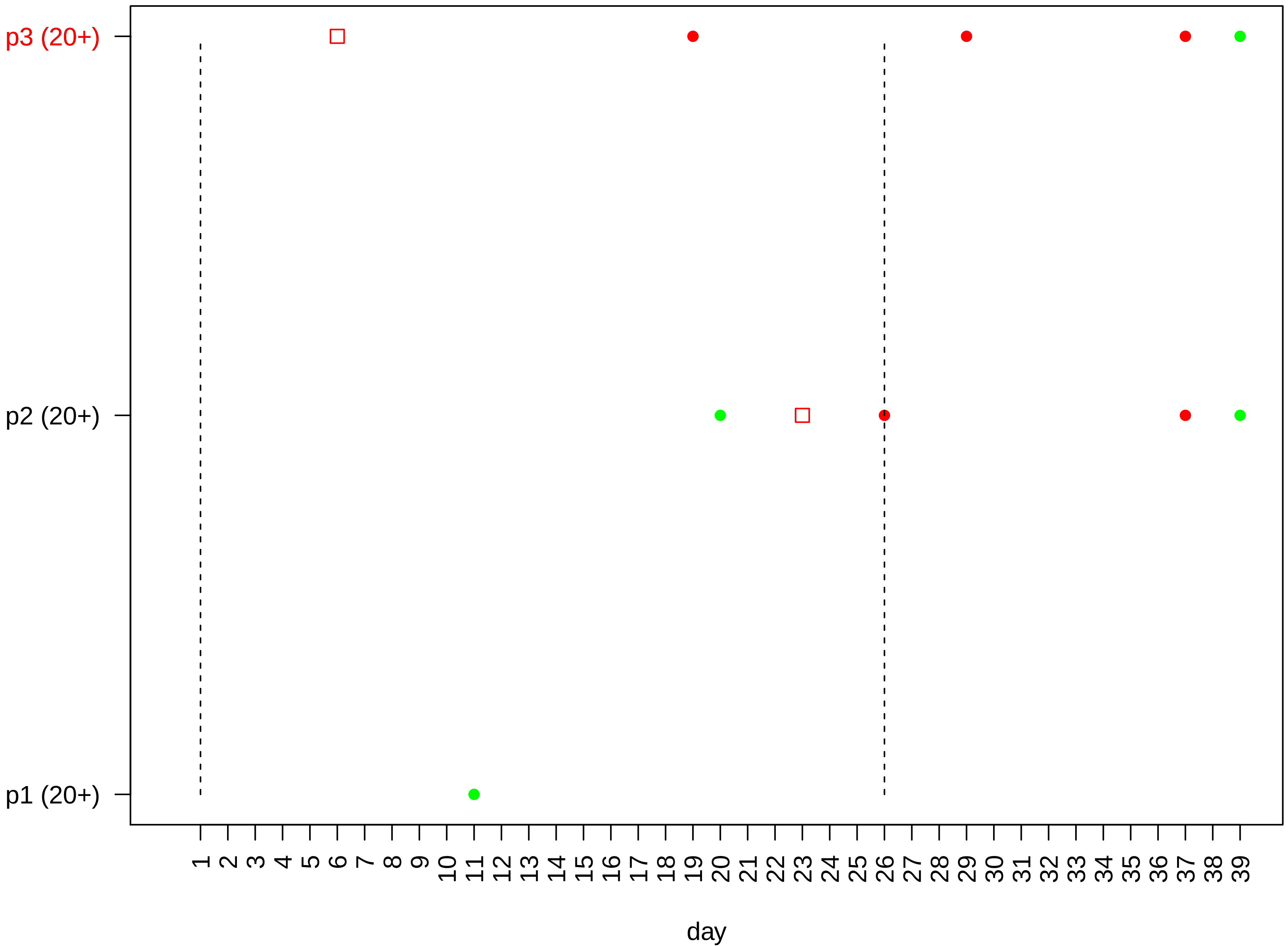

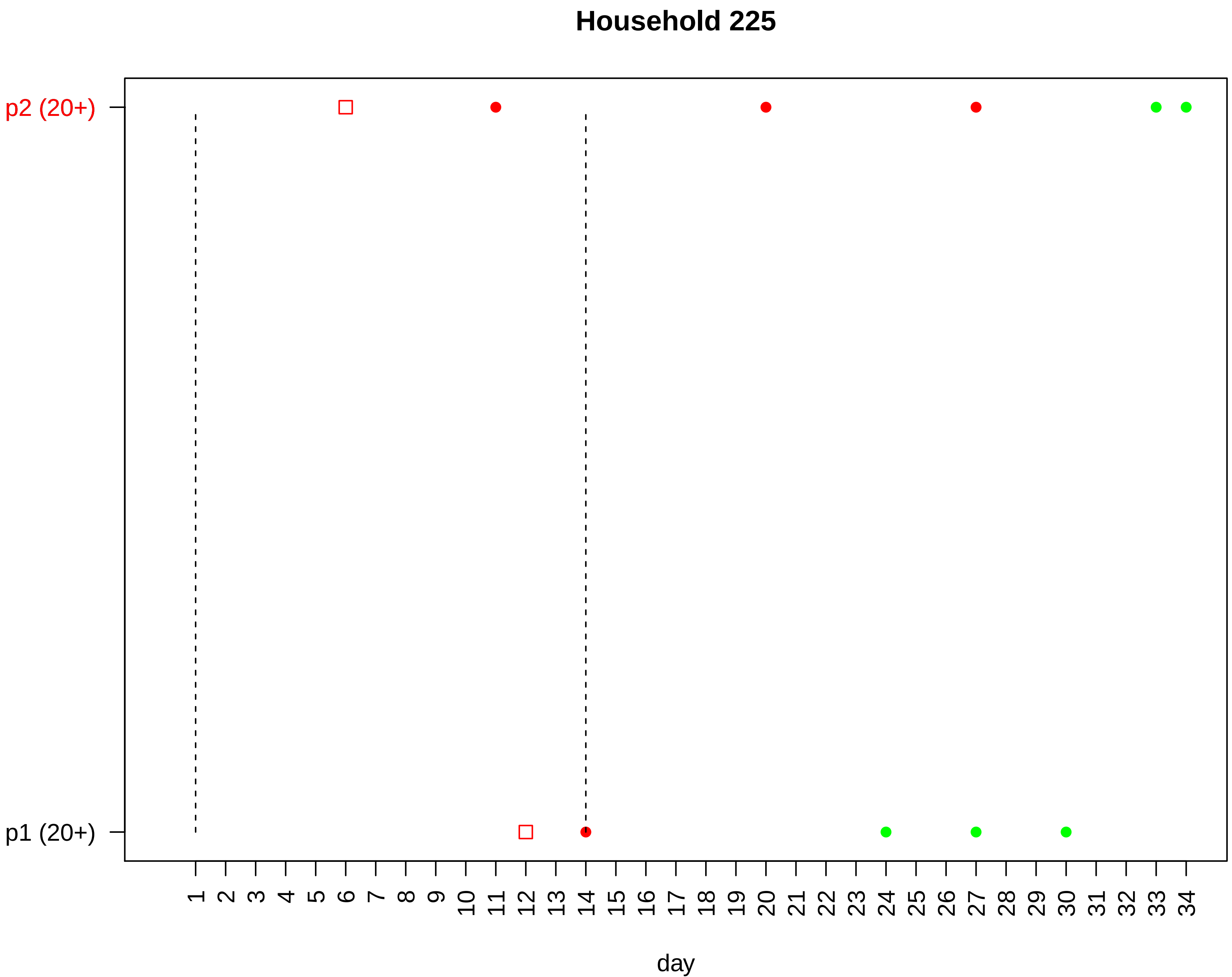

Household 225
p2 (20+)
p1 (20+)
day

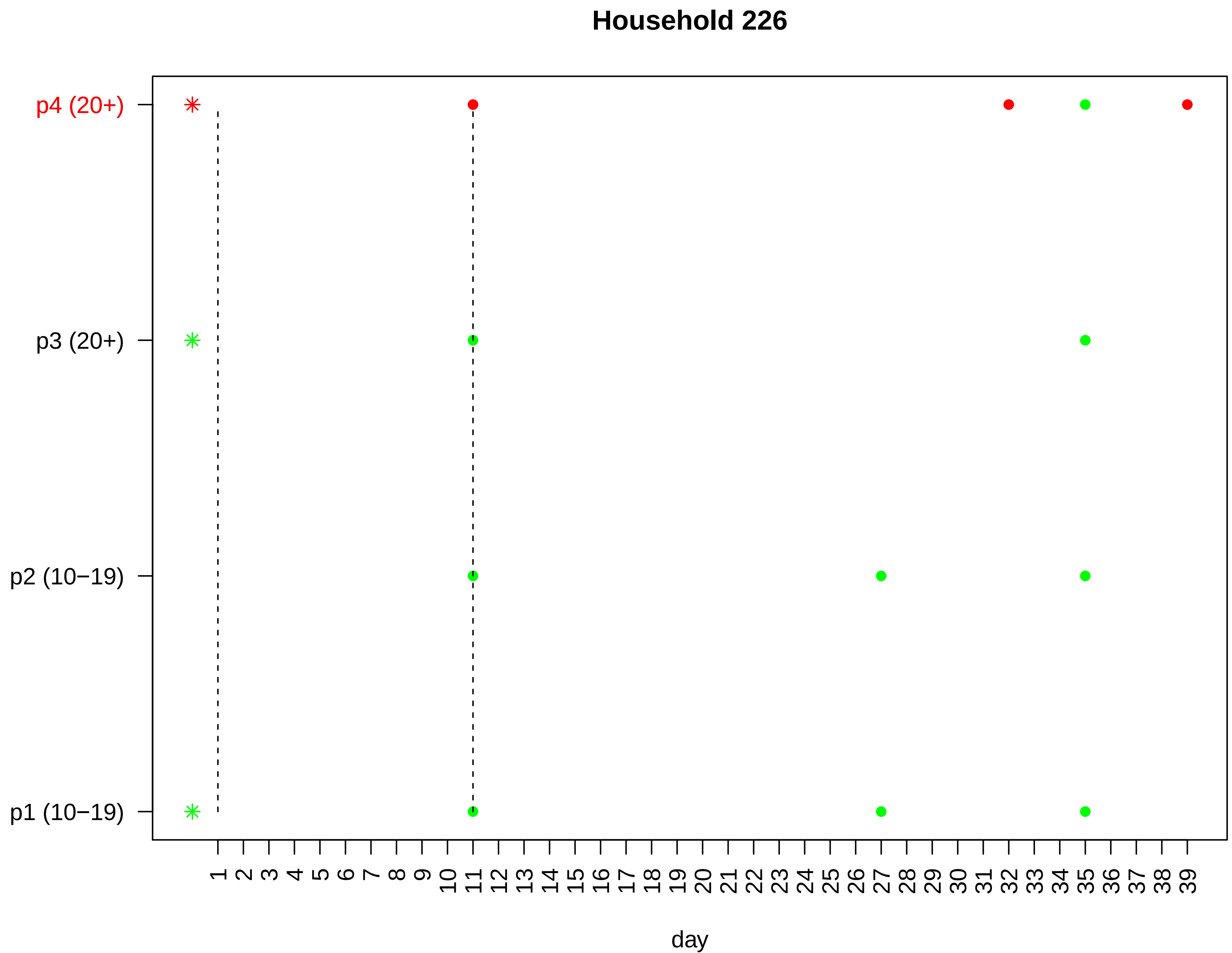
Household 226
p4 (20+)
p3 (20+)
p2 (10−19)
p1 (10−19)
1 2 3 4 5 6 7 8 9 10 11 12 13 14 15 16 17 18 19 20 21 22 23 24 25 26 27 28 29 30 31 32 33 34 35 36 37 38 39
day

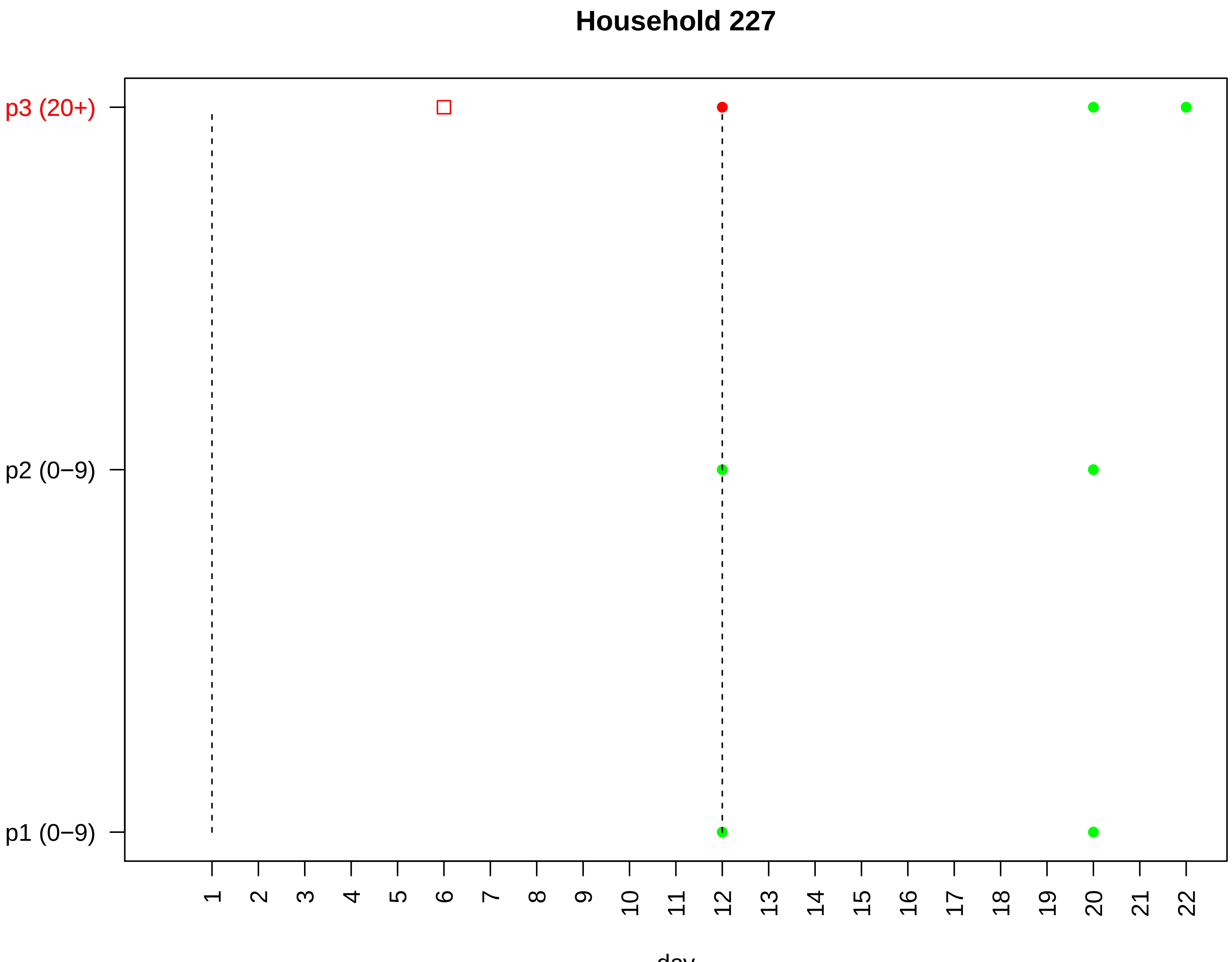

Household 227
p3 (20+)
p2 (0−9)
p1 (0−9)
day

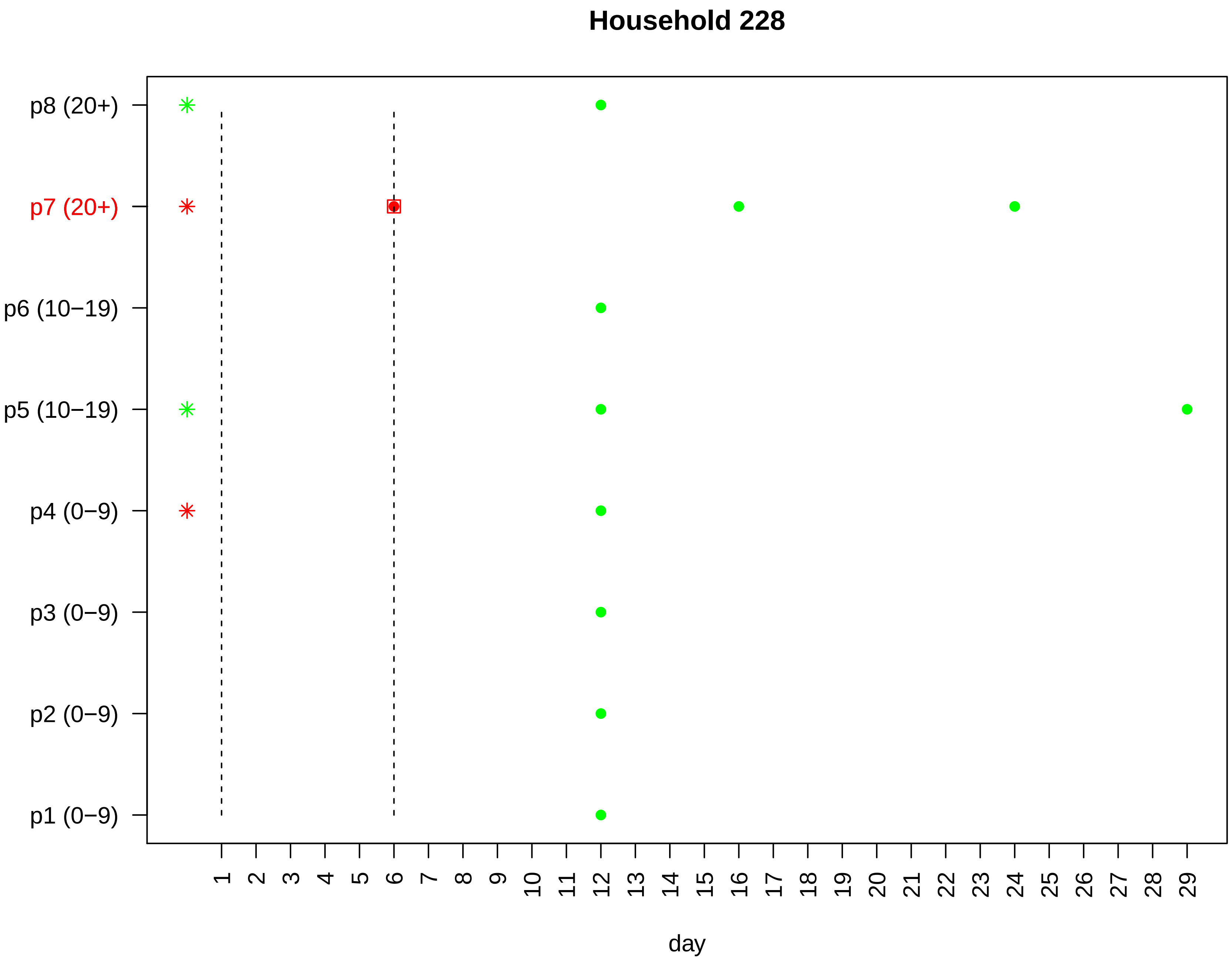
Household 228
p8 (20+)
p7 (20+)
p6 (10−19)
p5 (10−19)
p4 (0−9)
p3 (0−9)
p2 (0−9)
p1 (0−9)
1 2 3 4 5 6 7 8 9 10 11 12 13 14 15 16 17 18 19 20 21 22 23 24 25 26 27 28 29
day

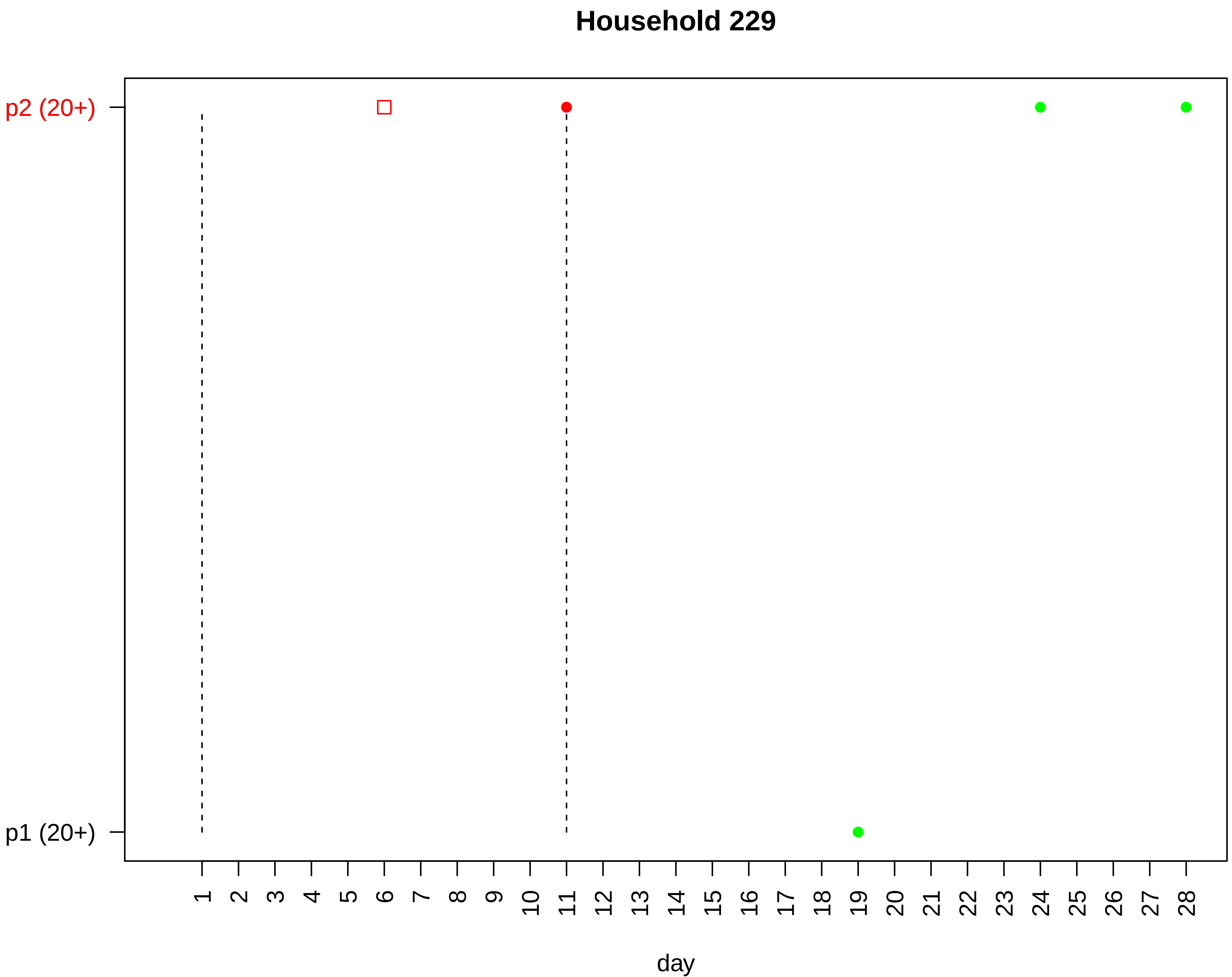
Household 229
p2 (20+)
p1 (20+)
day

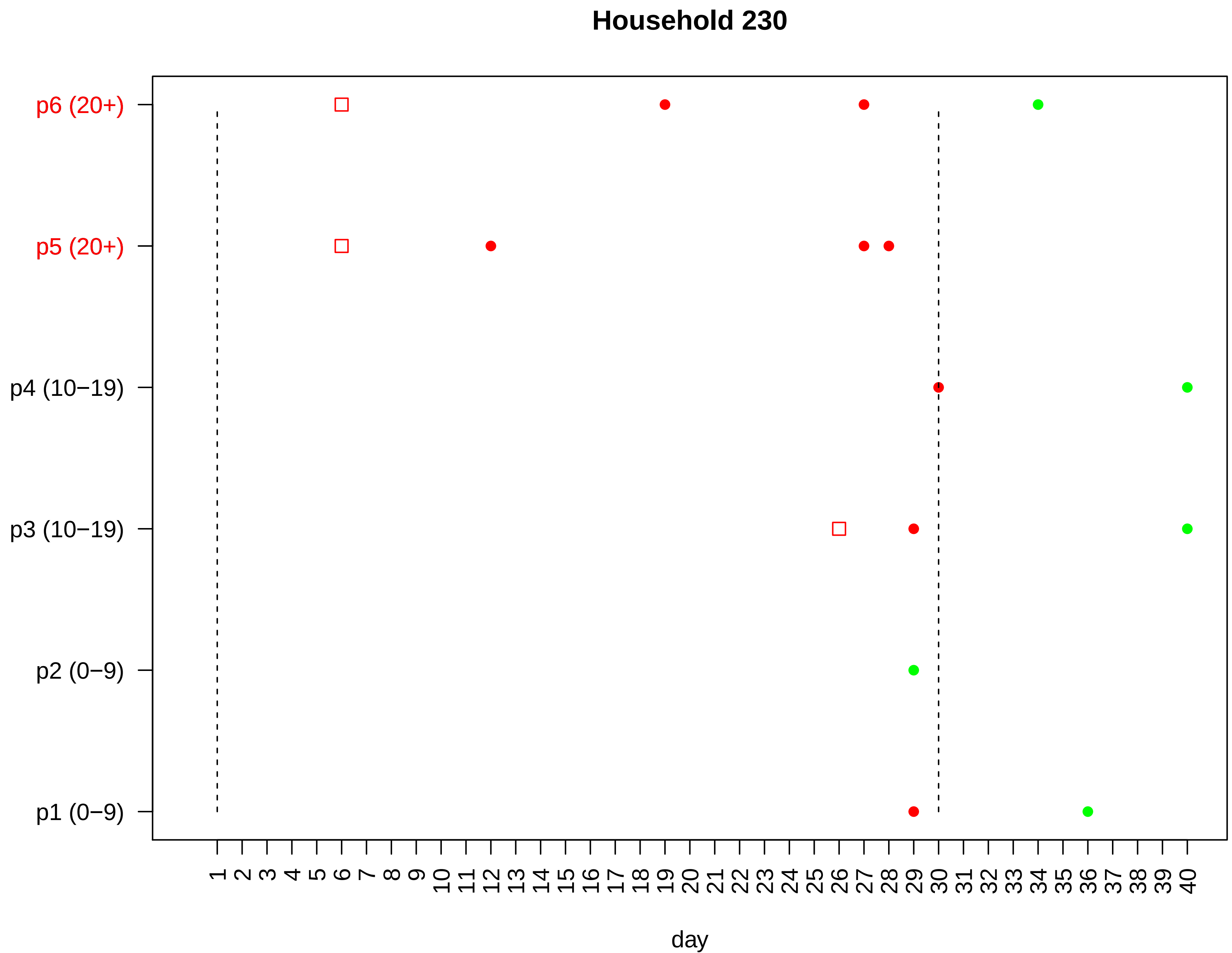

Household 230
p6 (20+)
p5 (20+)
p4 (10−19)
p3 (10−19)
p2 (0−9)
p1 (0−9)
1 2 3 4 5 6 7 8 9 10 11 12 13 14 15 16 17 18 19 20 21 22 23 24 25 26 27 28 29 30 31 32 33 34 35 36 37 38 39 40
day

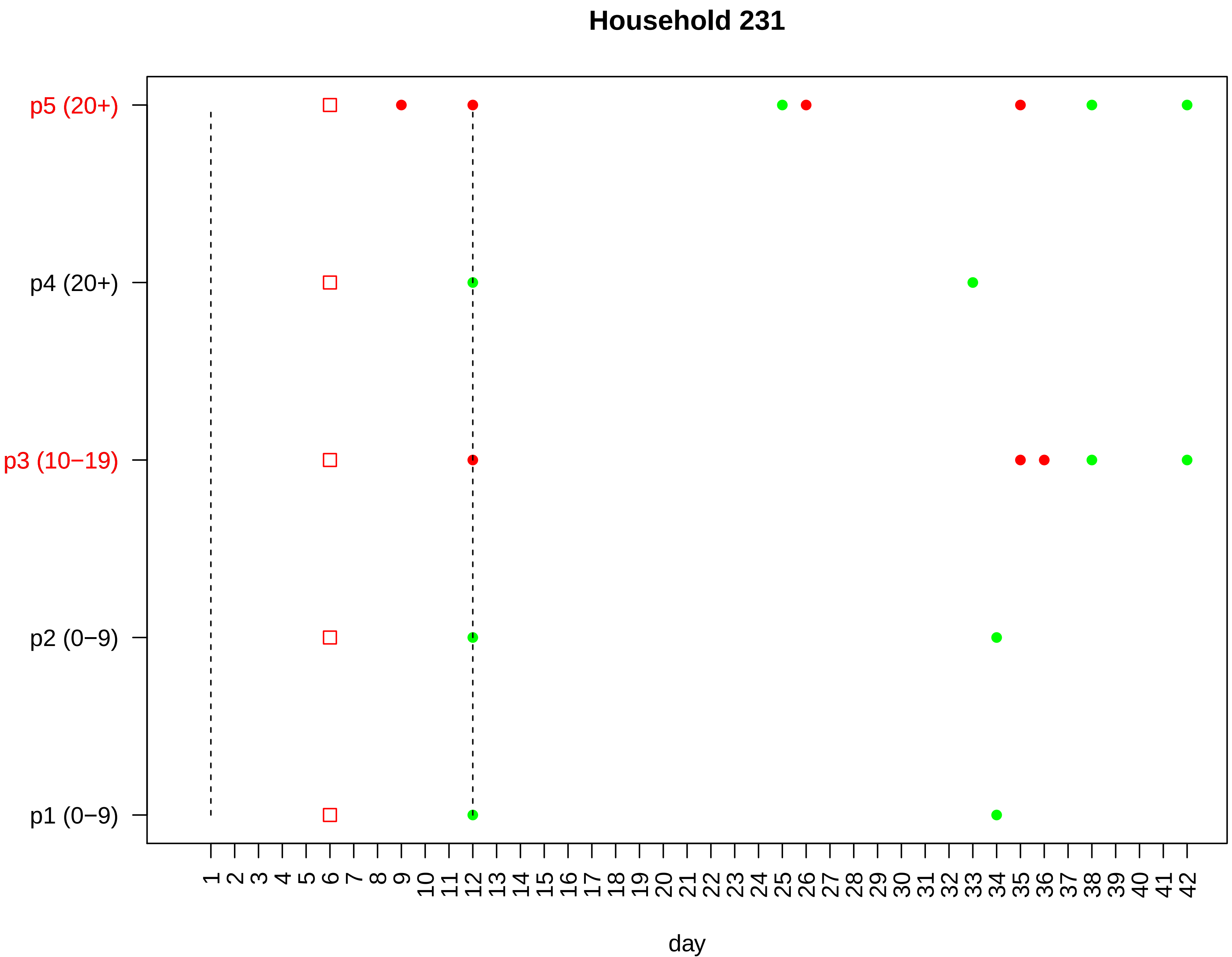

Household 231
p5 (20+)
p4 (20+)
p3 (10−19)
p2 (0−9)
p1 (0−9)
1 2 3 4 5 6 7 8 9 10 11 12 13 14 15 16 17 18 19 20 21 22 23 24 25 26 27 28 29 30 31 32 33 34 35 36 37 38 39 40 41 42
day

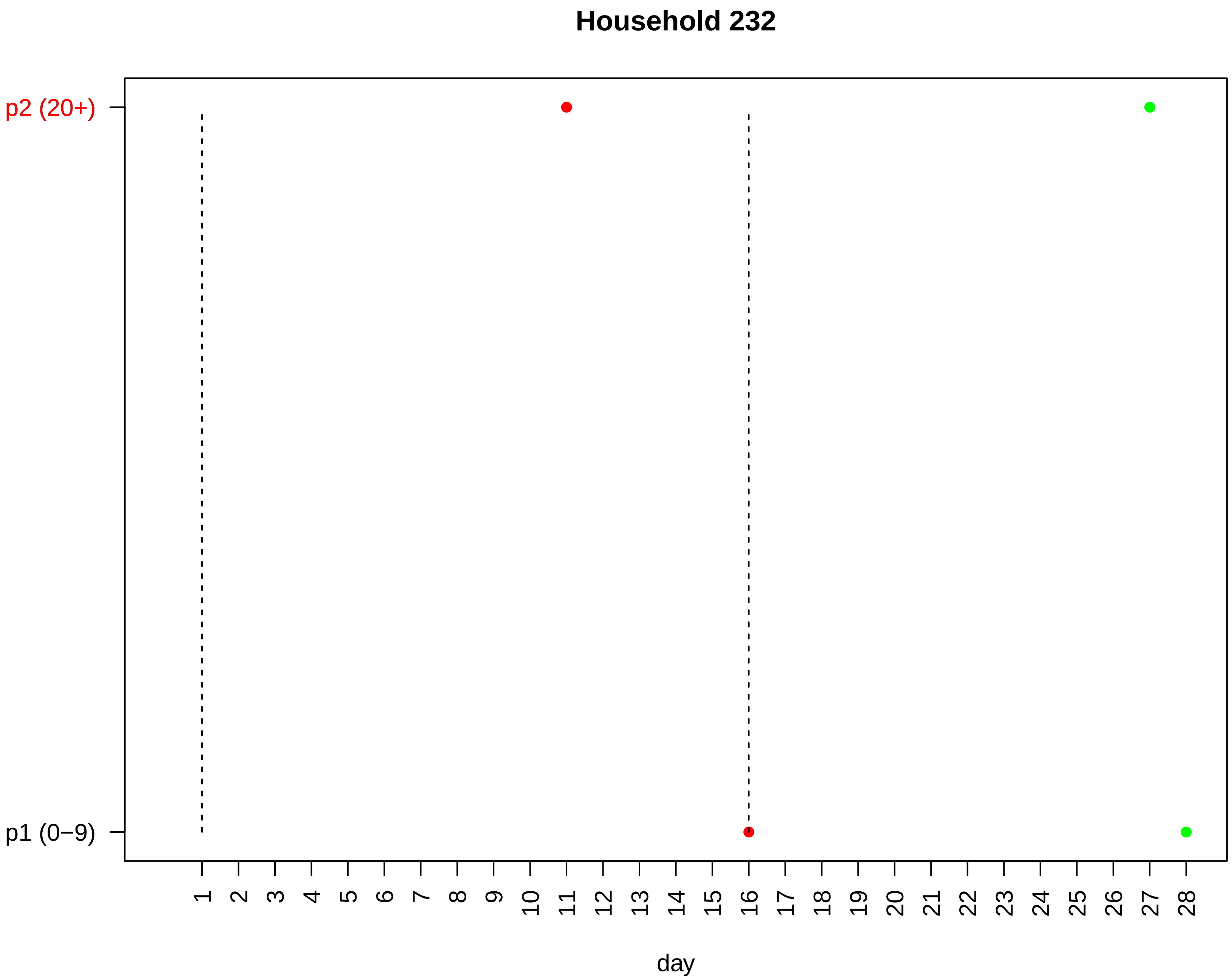
Household 232
p2 (20+)
p1 (0−9)
day

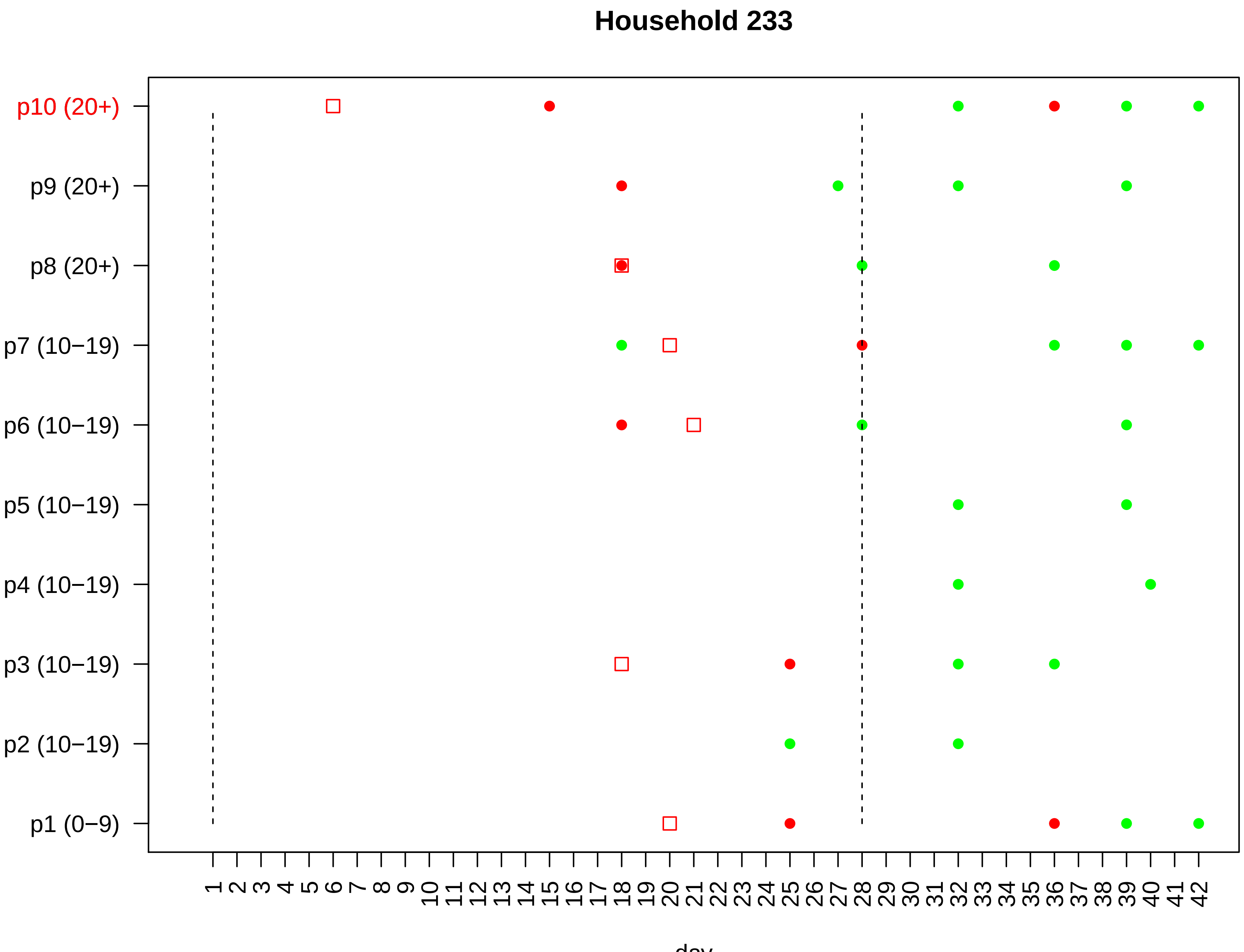

Household 233
p10 (20+)
p9 (20+)
p8 (20+)
p7 (10−19)
p6 (10−19)
p5 (10−19)
p4 (10−19)
p3 (10−19)
p2 (10−19)
p1 (0−9)
1 2 3 4 5 6 7 8 9 10 11 12 13 14 15 16 17 18 19 20 21 22 23 24 25 26 27 28 29 30 31 32 33 34 35 36 37 38 39 40 41 42
day

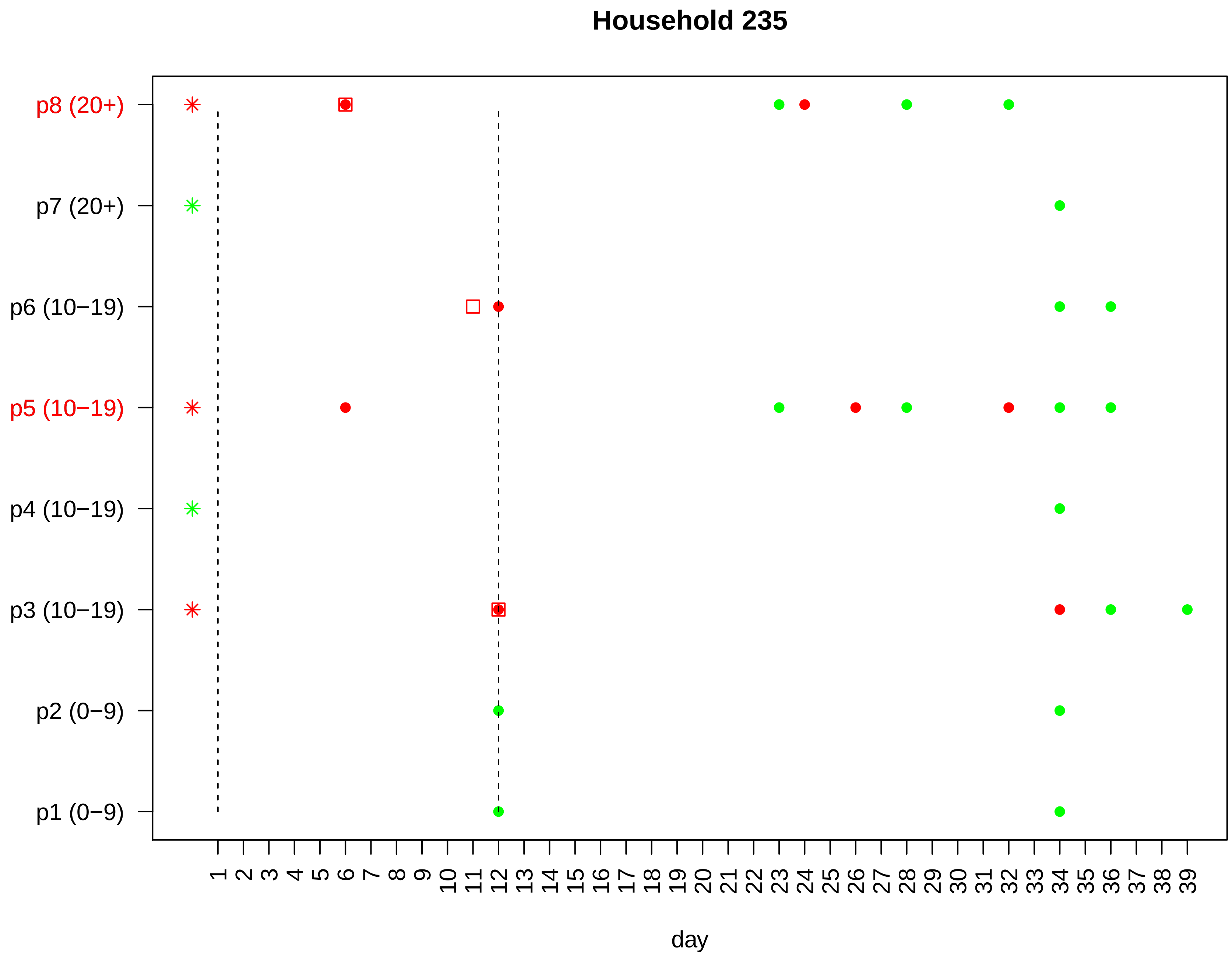

Household 235
p8 (20+)
p7 (20+)
p6 (10−19)
p5 (10−19)
p4 (10−19)
p3 (10−19)
p2 (0−9)
p1 (0−9)
1 2 3 4 5 6 7 8 9 10 11 12 13 14 15 16 17 18 19 20 21 22 23 24 25 26 27 28 29 30 31 32 33 34 35 36 37 38 39
day

#### Household 236

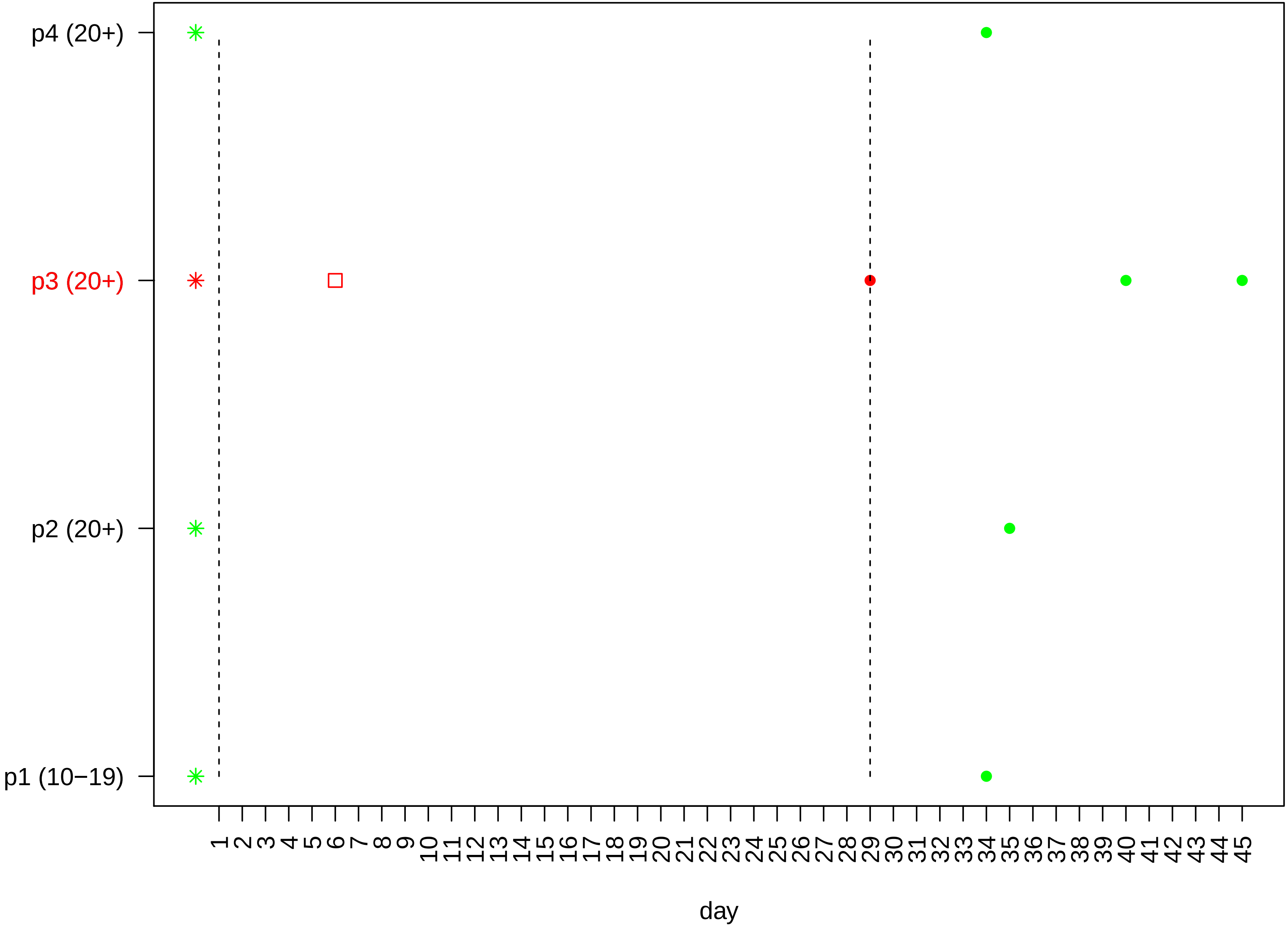

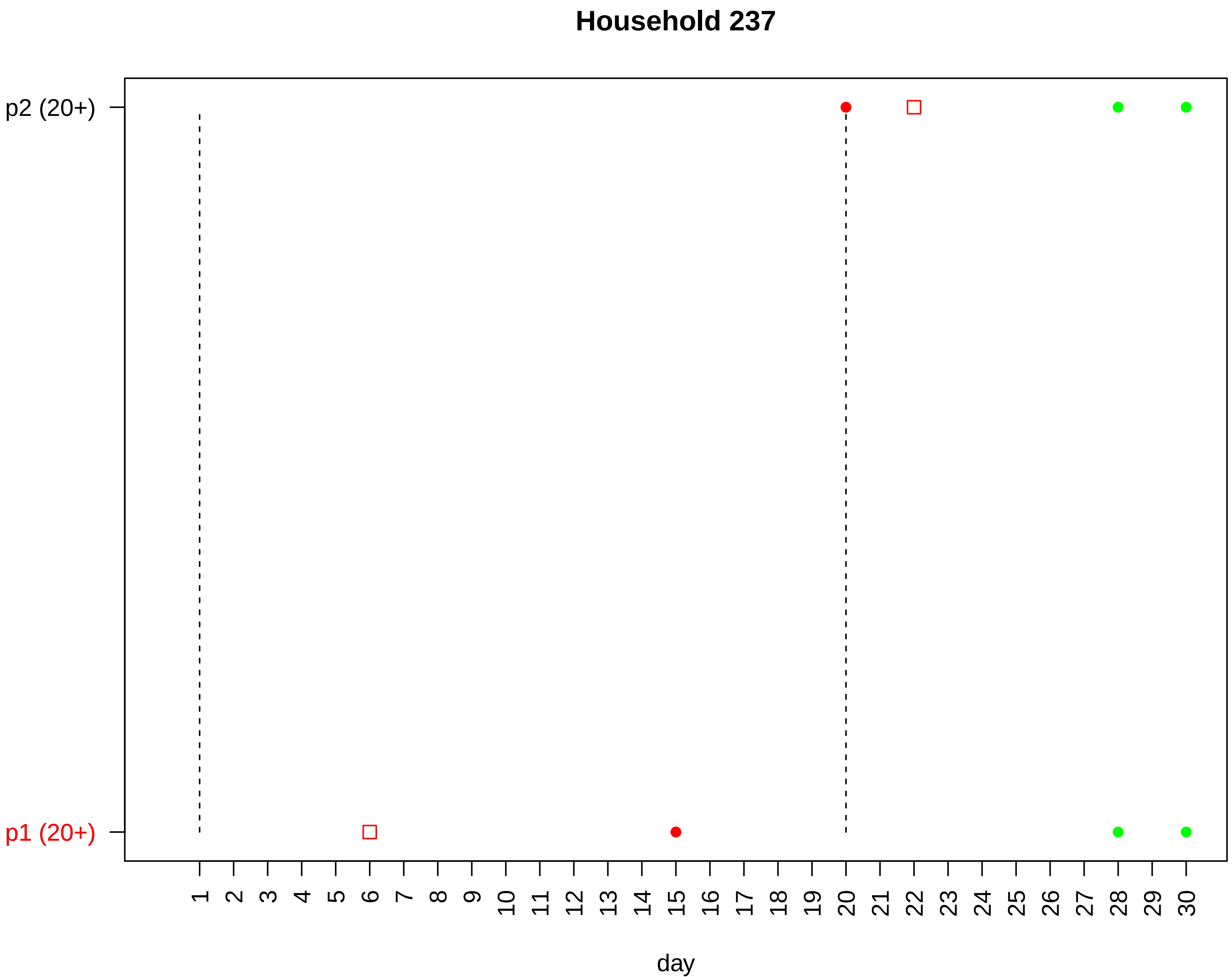

Household 237
p2 (20+)
p1 (20+)
day

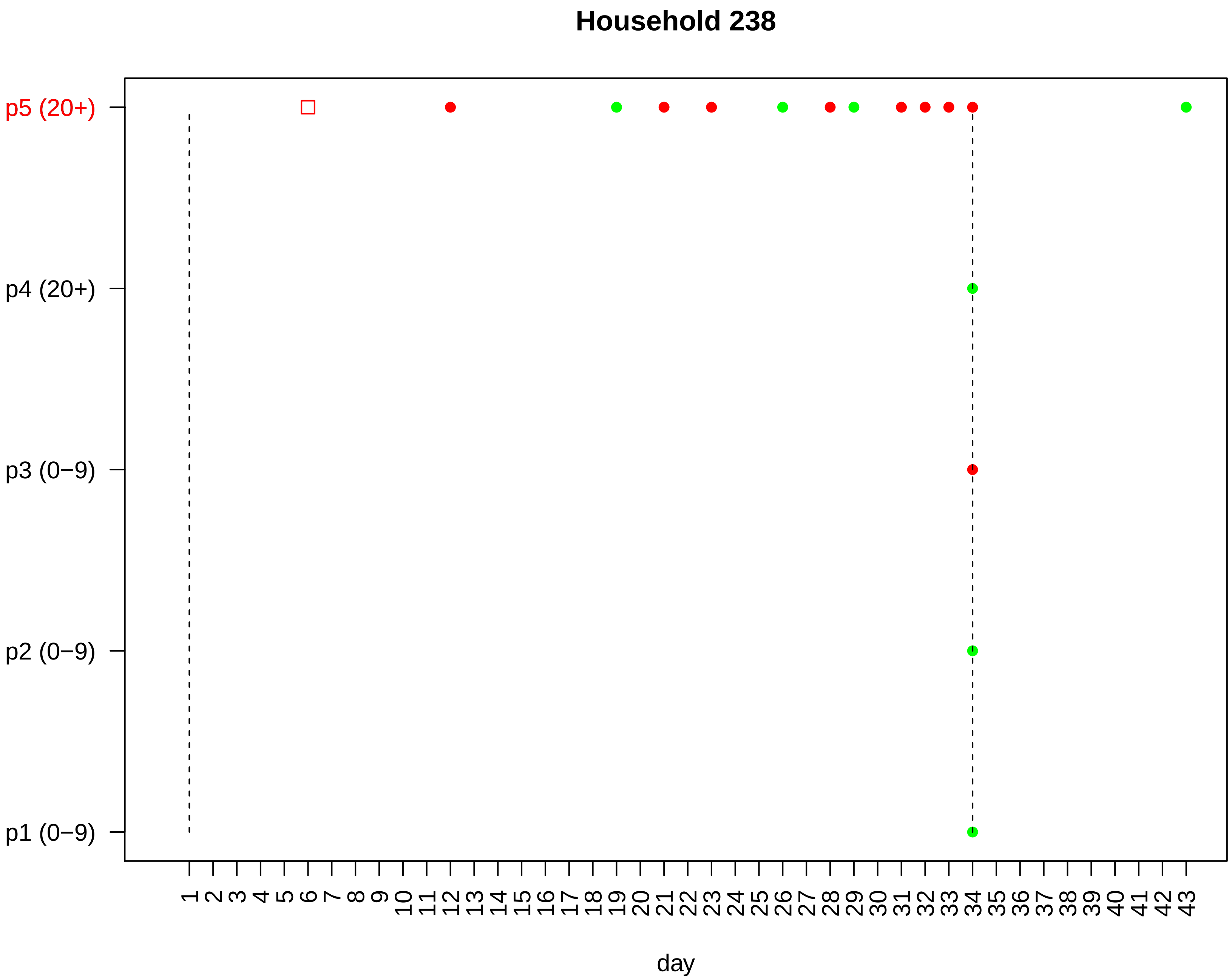

Household 238
p5 (20+)
p4 (20+)
p3 (0−9)
p2 (0−9)
p1 (0−9)
1 2 3 4 5 6 7 8 9 10 11 12 13 14 15 16 17 18 19 20 21 22 23 24 25 26 27 28 29 30 31 32 33 34 35 36 37 38 39 40 41 42 43
day

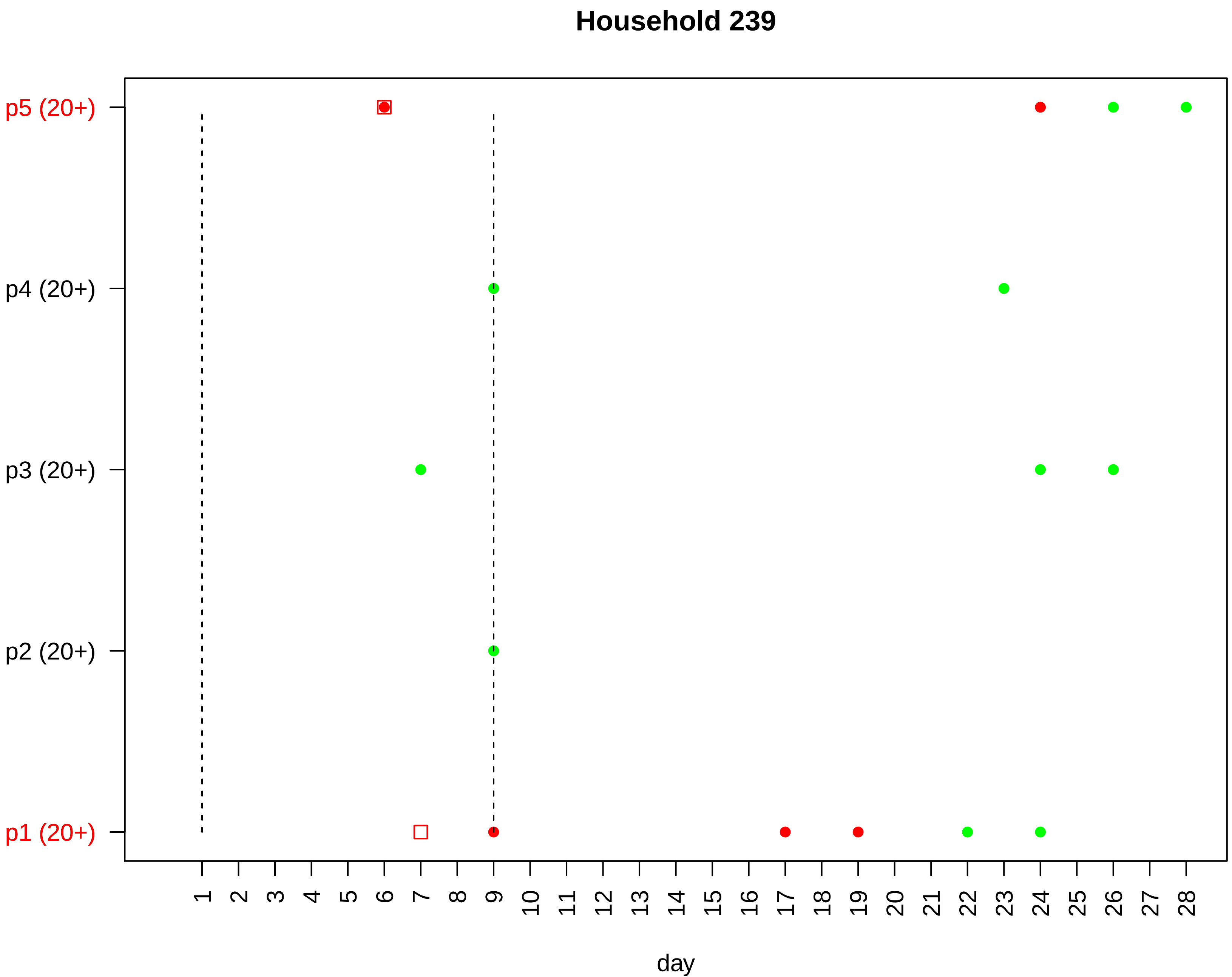

Household 239
p5 (20+)
p4 (20+)
p3 (20+)
p2 (20+)
p1 (20+)
1 2 3 4 5 6 7 8 9 10 11 12 13 14 15 16 17 18 19 20 21 22 23 24 25 26 27 28
day

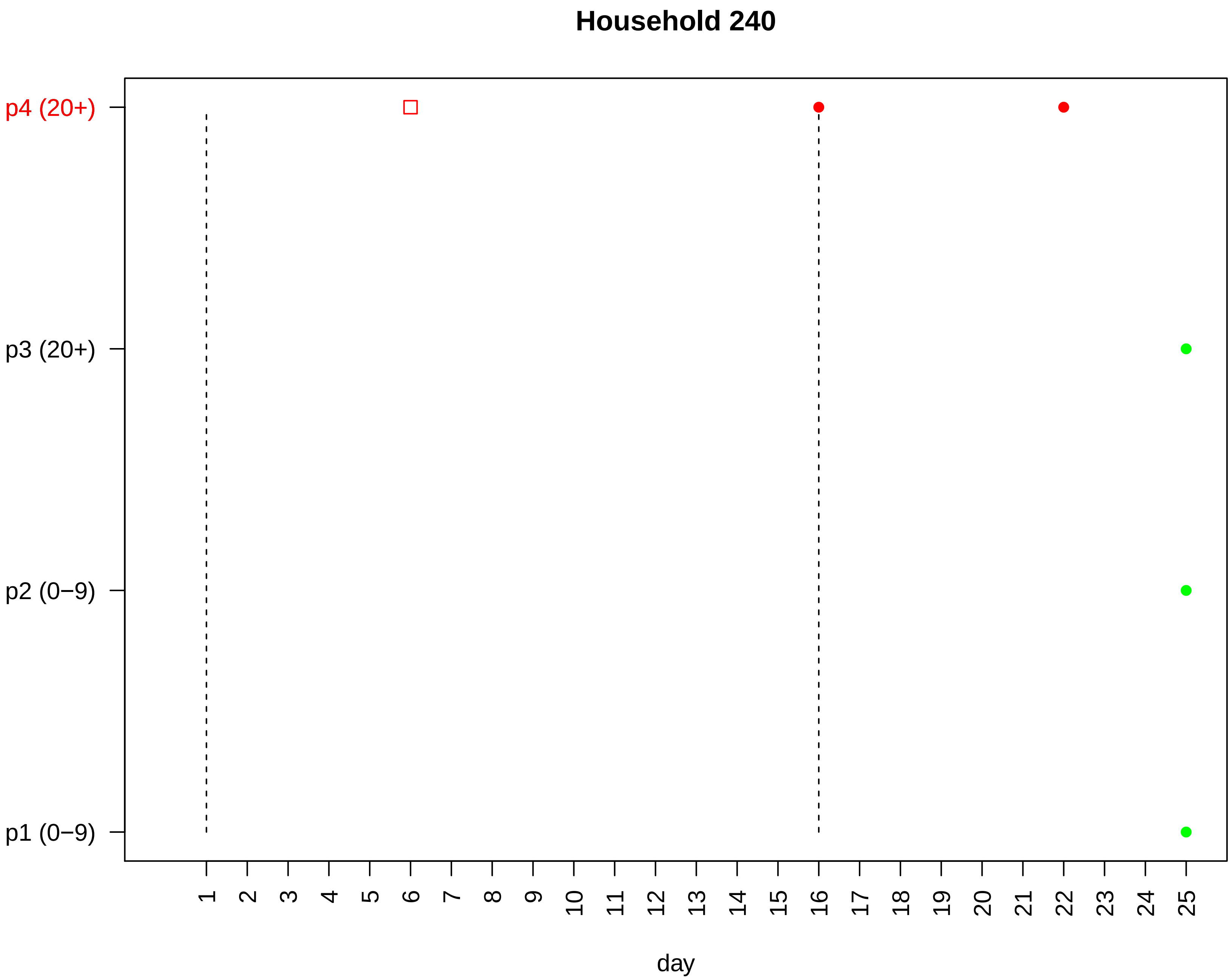

Household 240
p4 (20+)
p3 (20+)
p2 (0−9)
p1 (0−9)
day

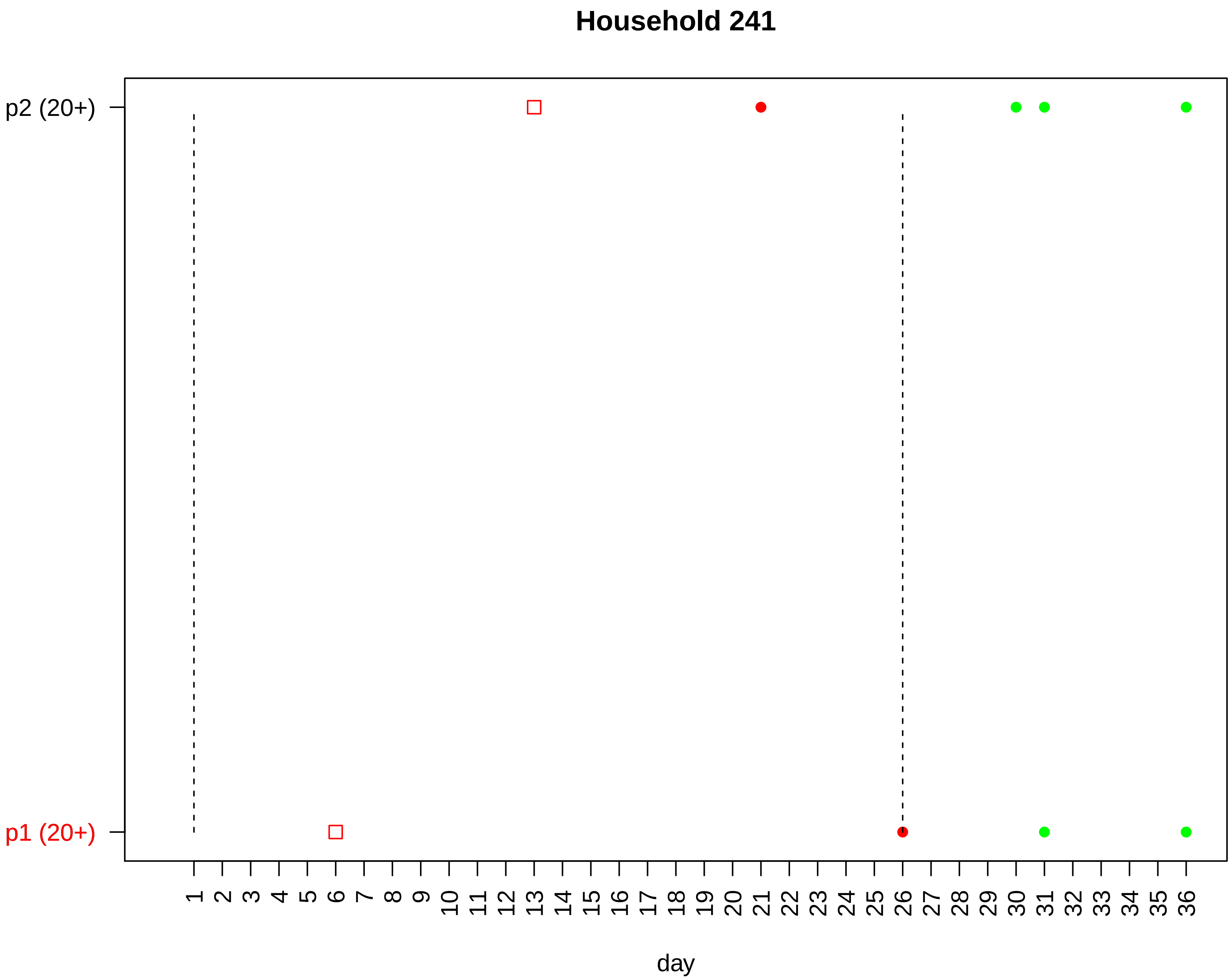
Household 241
p2 (20+)
p1 (20+)
day

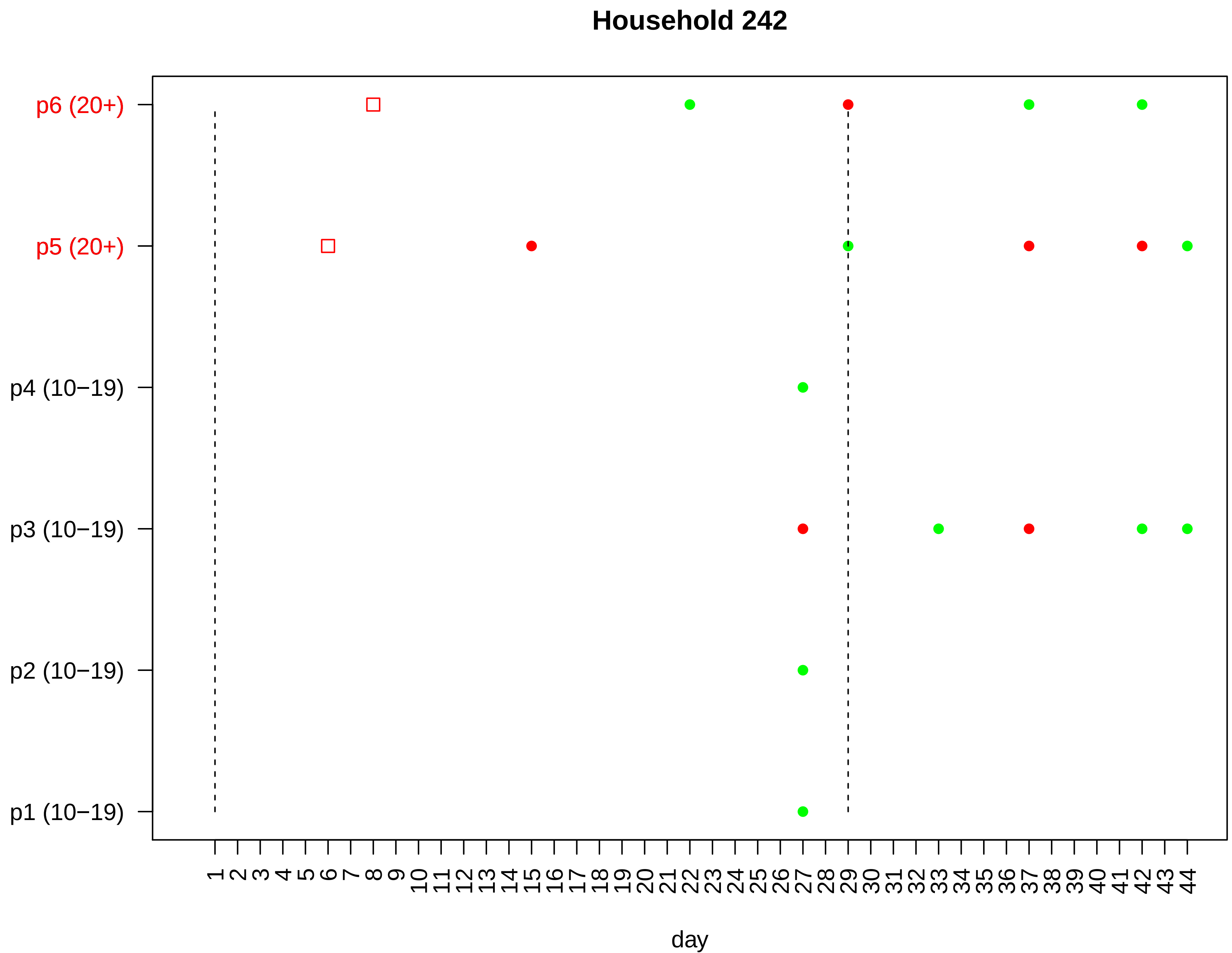
Household 242
p6 (20+)
p5 (20+)
p4 (10−19)
p3 (10−19)
p2 (10−19)
p1 (10−19)
1 2 3 4 5 6 7 8 9 10 11 12 13 14 15 16 17 18 19 20 21 22 23 24 25 26 27 28 29 30 31 32 33 34 35 36 37 38 39 40 41 42 43 44
day

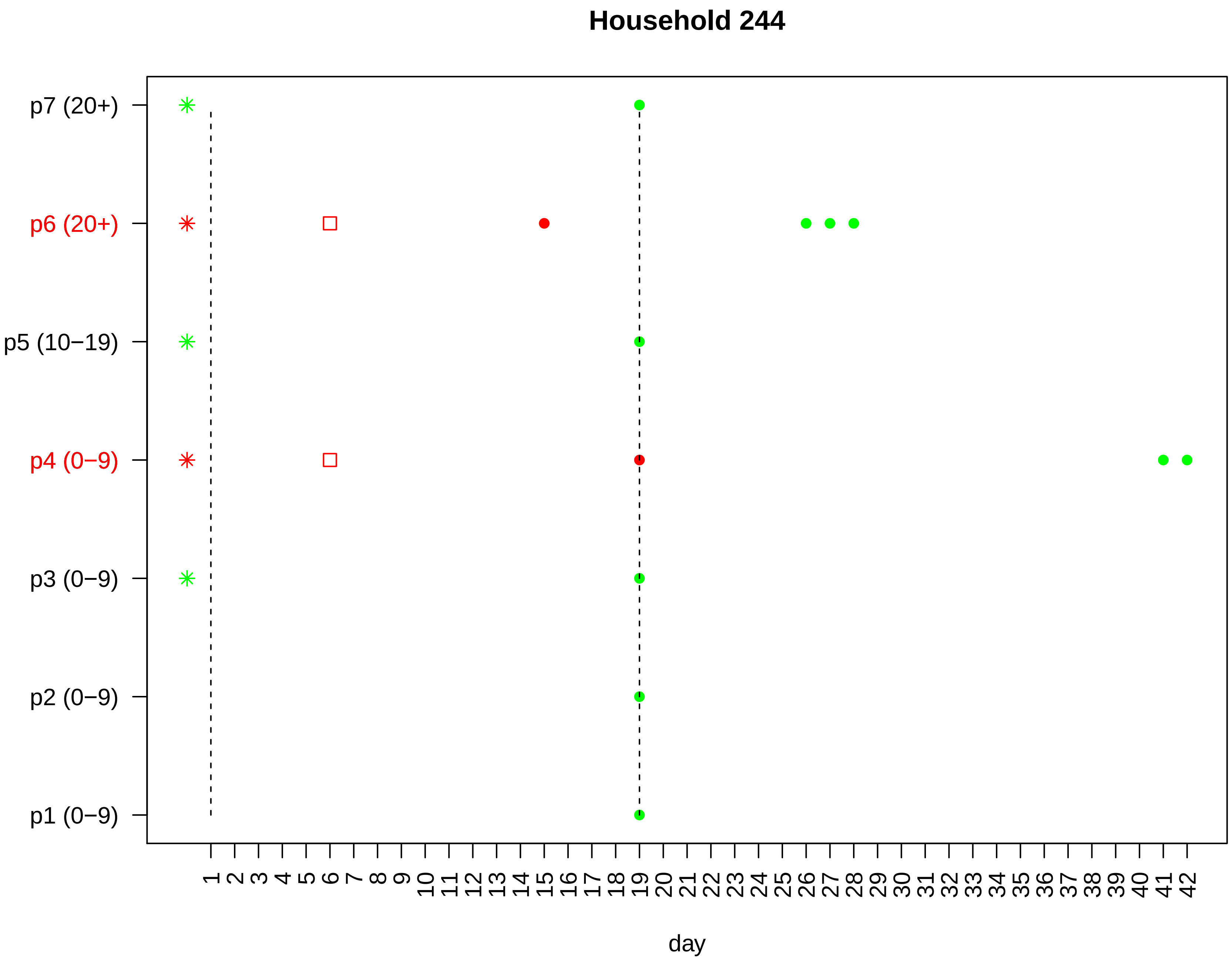

Household 244
p7 (20+)
p6 (20+)
p5 (10−19)
p4 (0−9)
p3 (0−9)
p2 (0−9)
p1 (0−9)
1 2 3 4 5 6 7 8 9 10 11 12 13 14 15 16 17 18 19 20 21 22 23 24 25 26 27 28 29 30 31 32 33 34 35 36 37 38 39 40 41 42
day

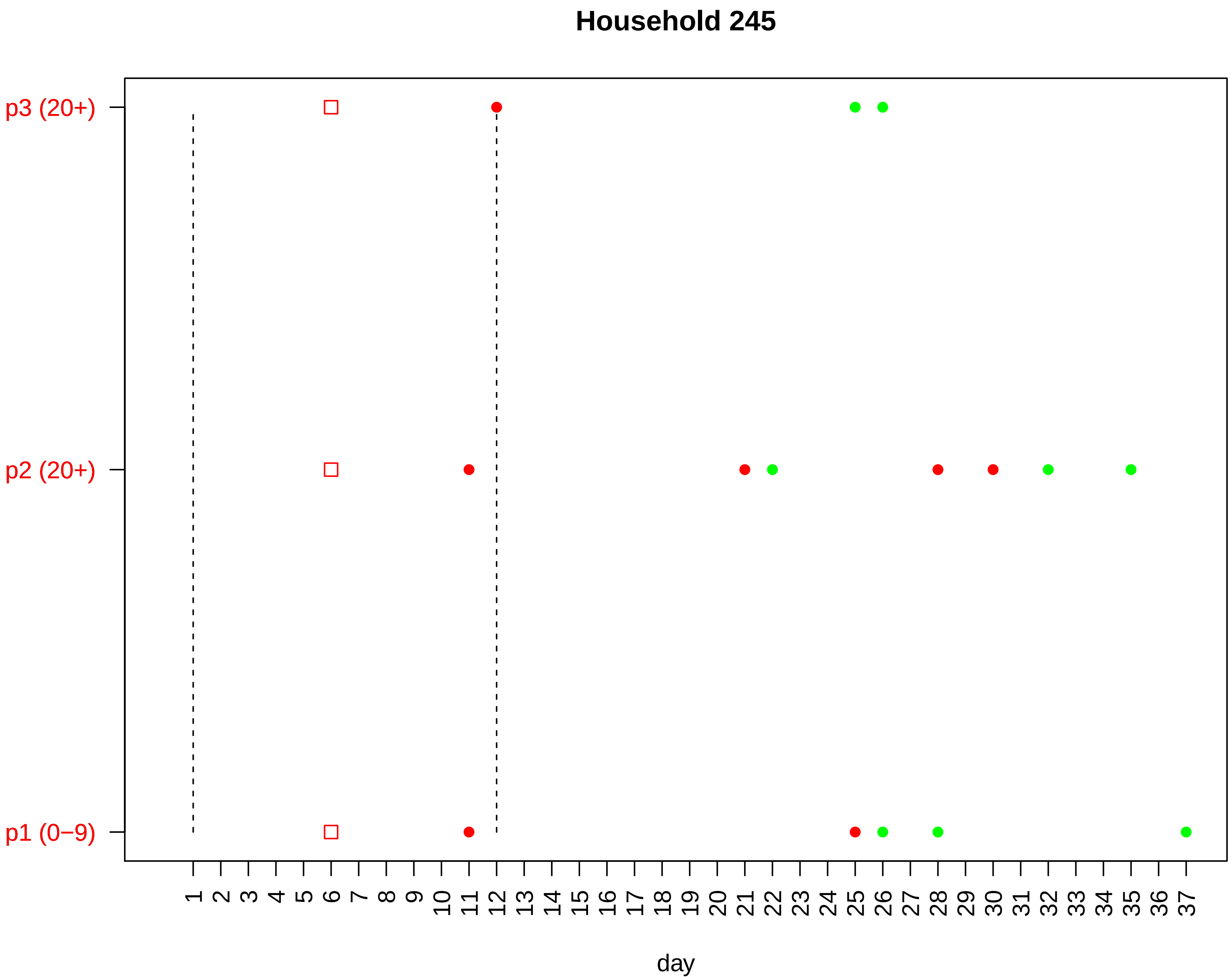
Household 245
p3 (20+)
p2 (20+)
p1 (0−9)
1 2 3 4 5 6 7 8 9 10 11 12 13 14 15 16 17 18 19 20 21 22 23 24 25 26 27 28 29 30 31 32 33 34 35 36 37
day

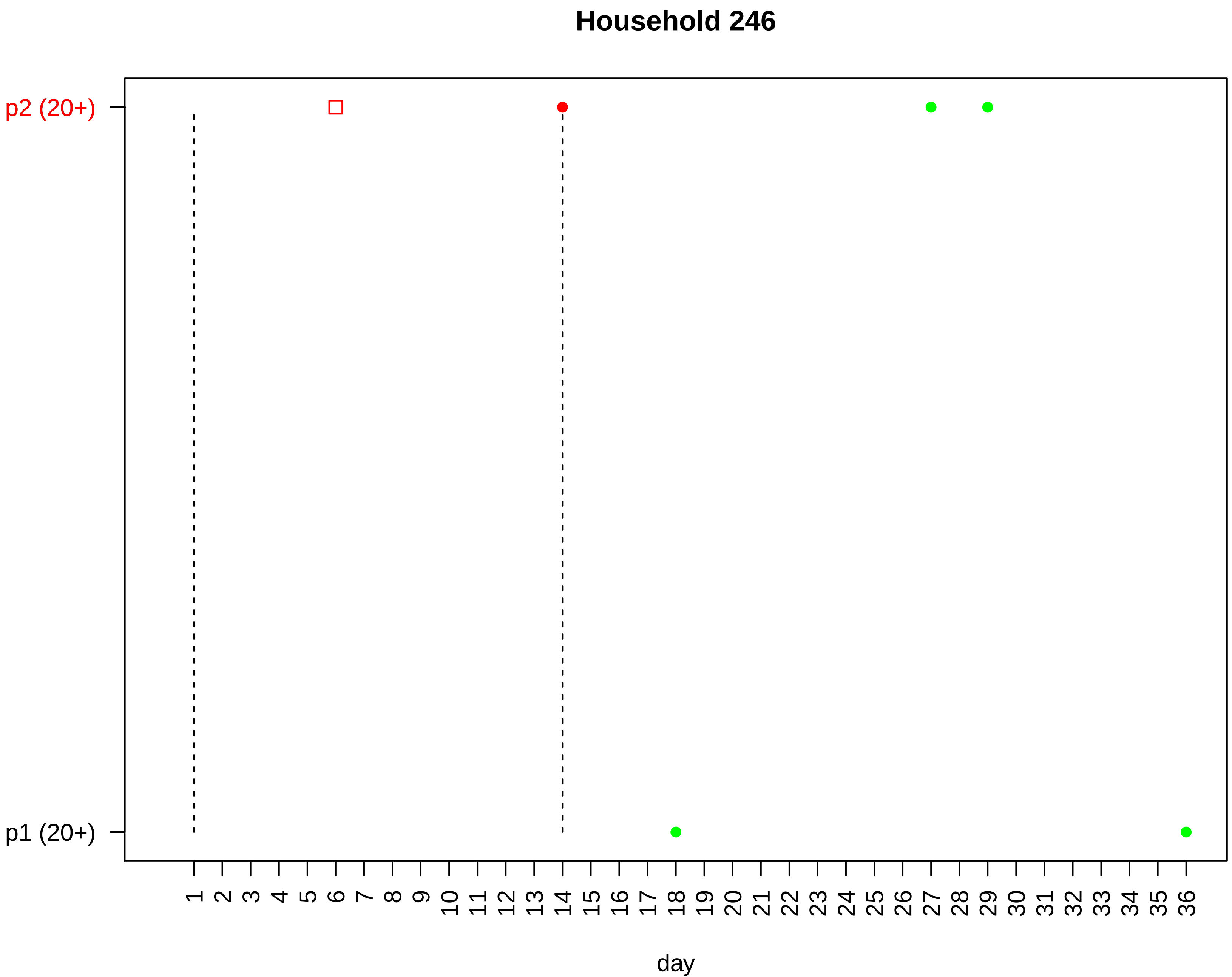

Household 246
p2 (20+)
p1 (20+)
1 2 3 4 5 6 7 8 9 10 11 12 13 14 15 16 17 18 19 20 21 22 23 24 25 26 27 28 29 30 31 32 33 34 35 36
day

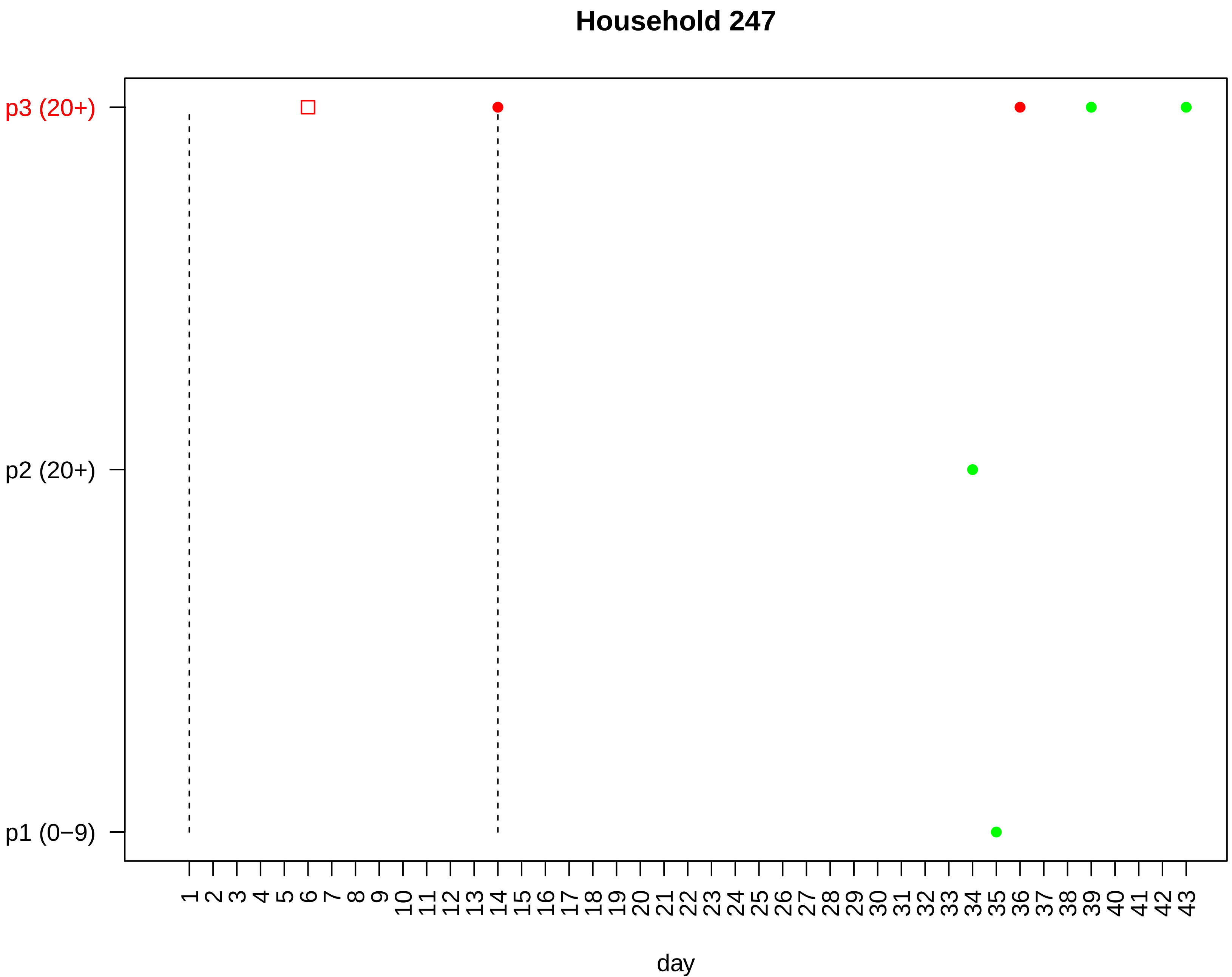

Household 247
p3 (20+)
p2 (20+)
p1 (0−9)
1 2 3 4 5 6 7 8 9 10 11 12 13 14 15 16 17 18 19 20 21 22 23 24 25 26 27 28 29 30 31 32 33 34 35 36 37 38 39 40 41 42 43
day

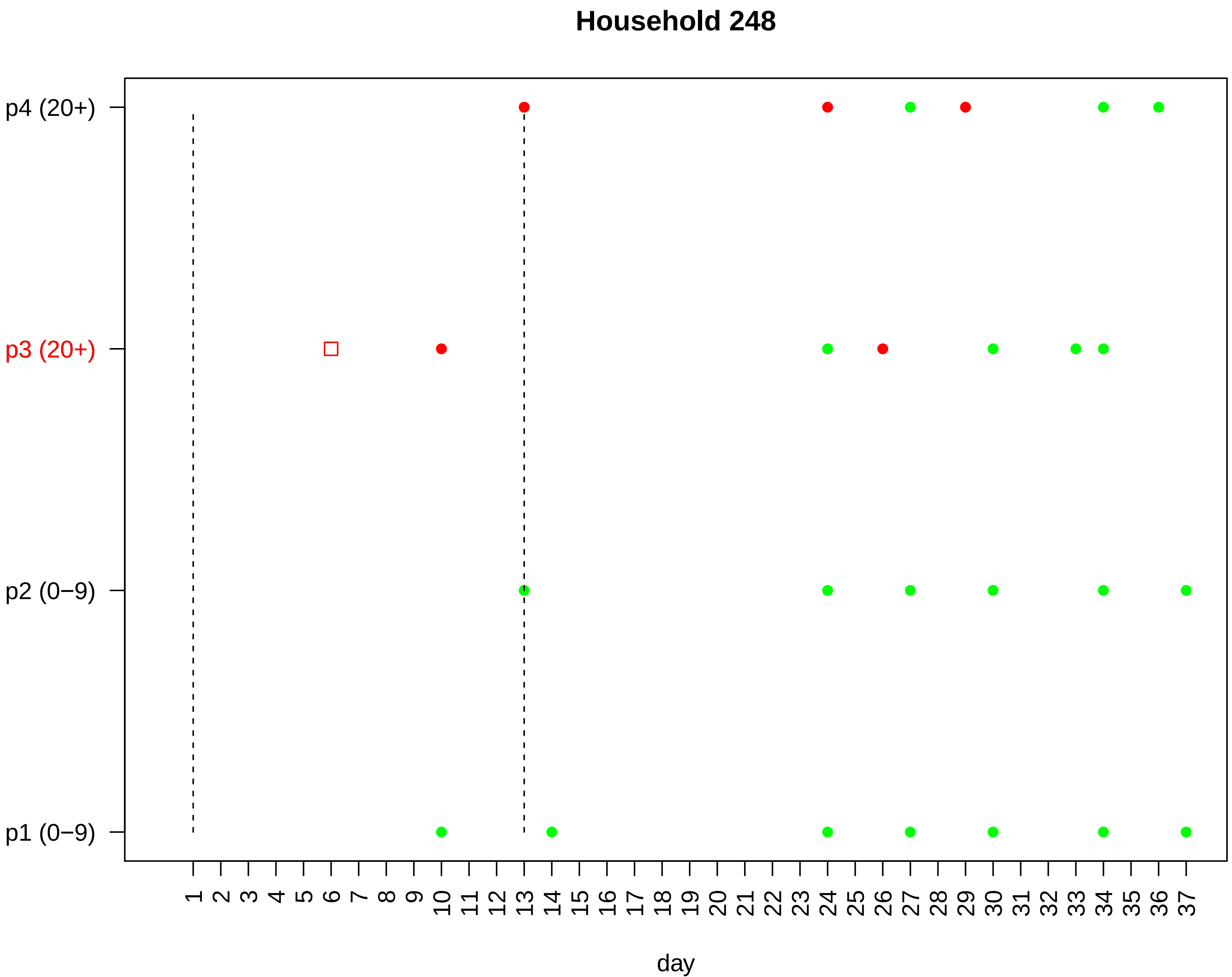

Household 248
p4 (20+)
p3 (20+)
p2 (0−9)
p1 (0−9)
1 2 3 4 5 6 7 8 9 10 11 12 13 14 15 16 17 18 19 20 21 22 23 24 25 26 27 28 29 30 31 32 33 34 35 36 37
day

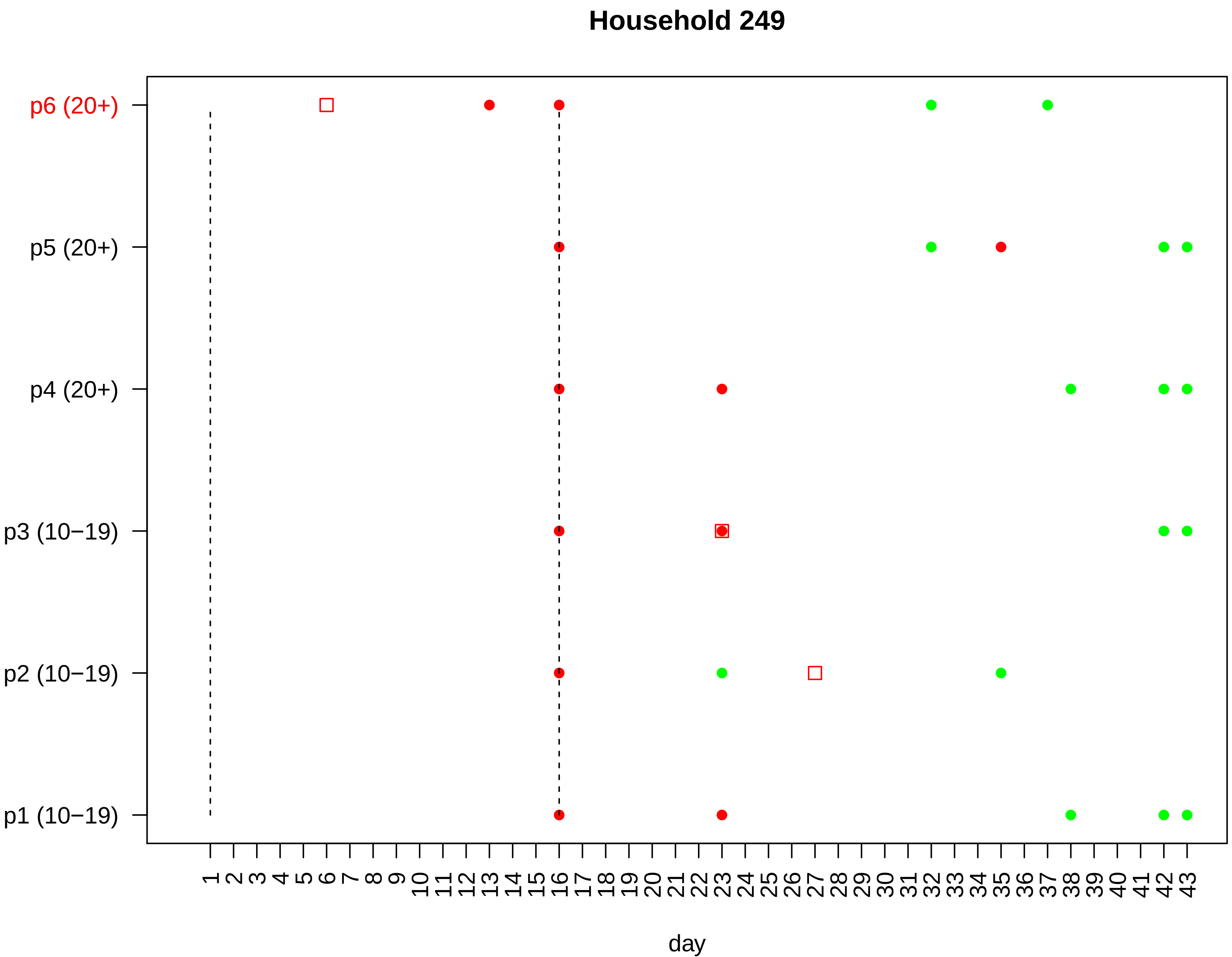

Household 249
p6 (20+)
p5 (20+)
p4 (20+)
p3 (10−19)
p2 (10−19)
p1 (10−19)
1 2 3 4 5 6 7 8 9 10 11 12 13 14 15 16 17 18 19 20 21 22 23 24 25 26 27 28 29 30 31 32 33 34 35 36 37 38 39 40 41 42 43
day

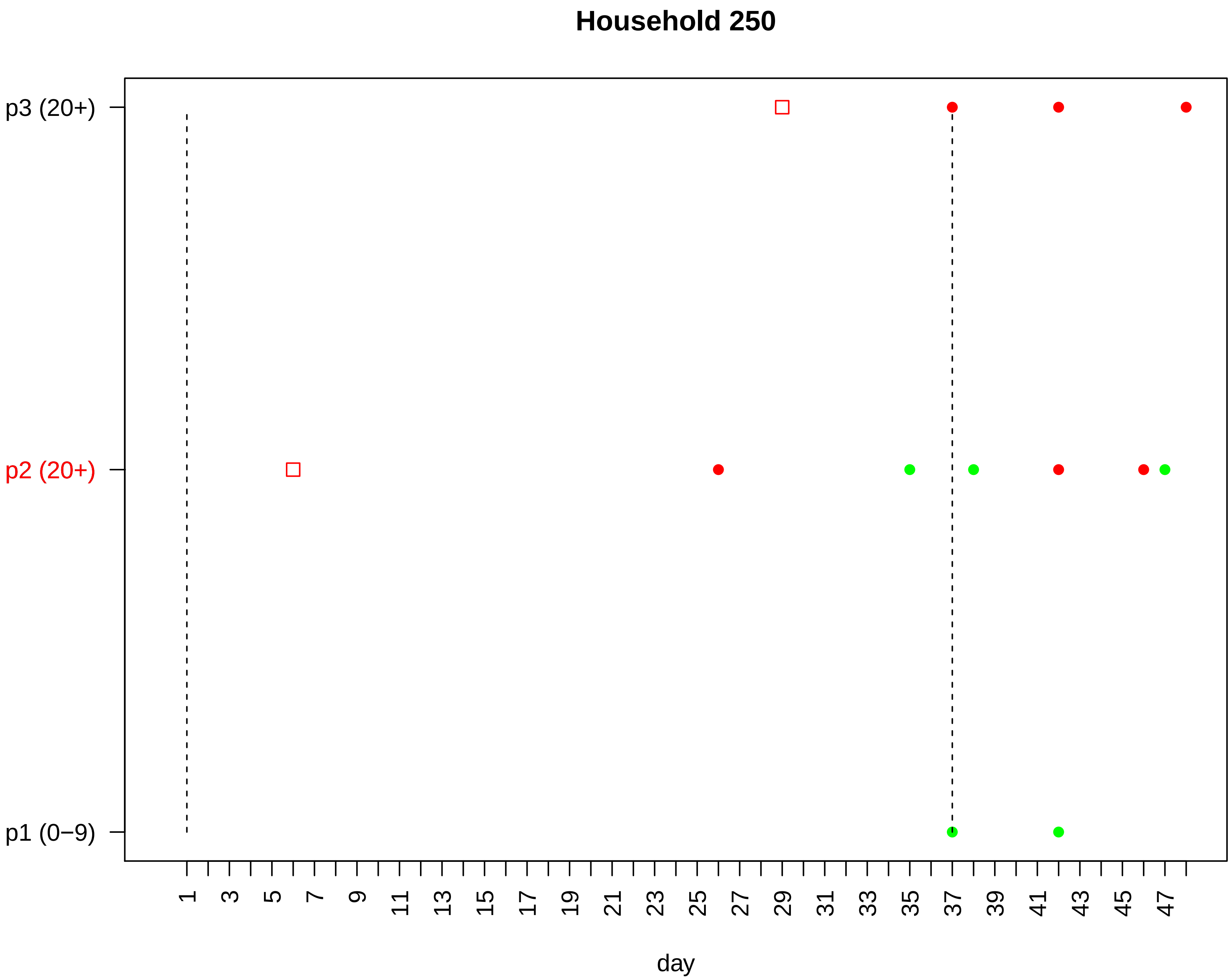

Household 250
p3 (20+)
p2 (20+)
p1 (0−9)
1
3
5
7
9
11
13
15
17
19
21
23
25
27
29
31
33
35
37
39
41
43
45
47
day

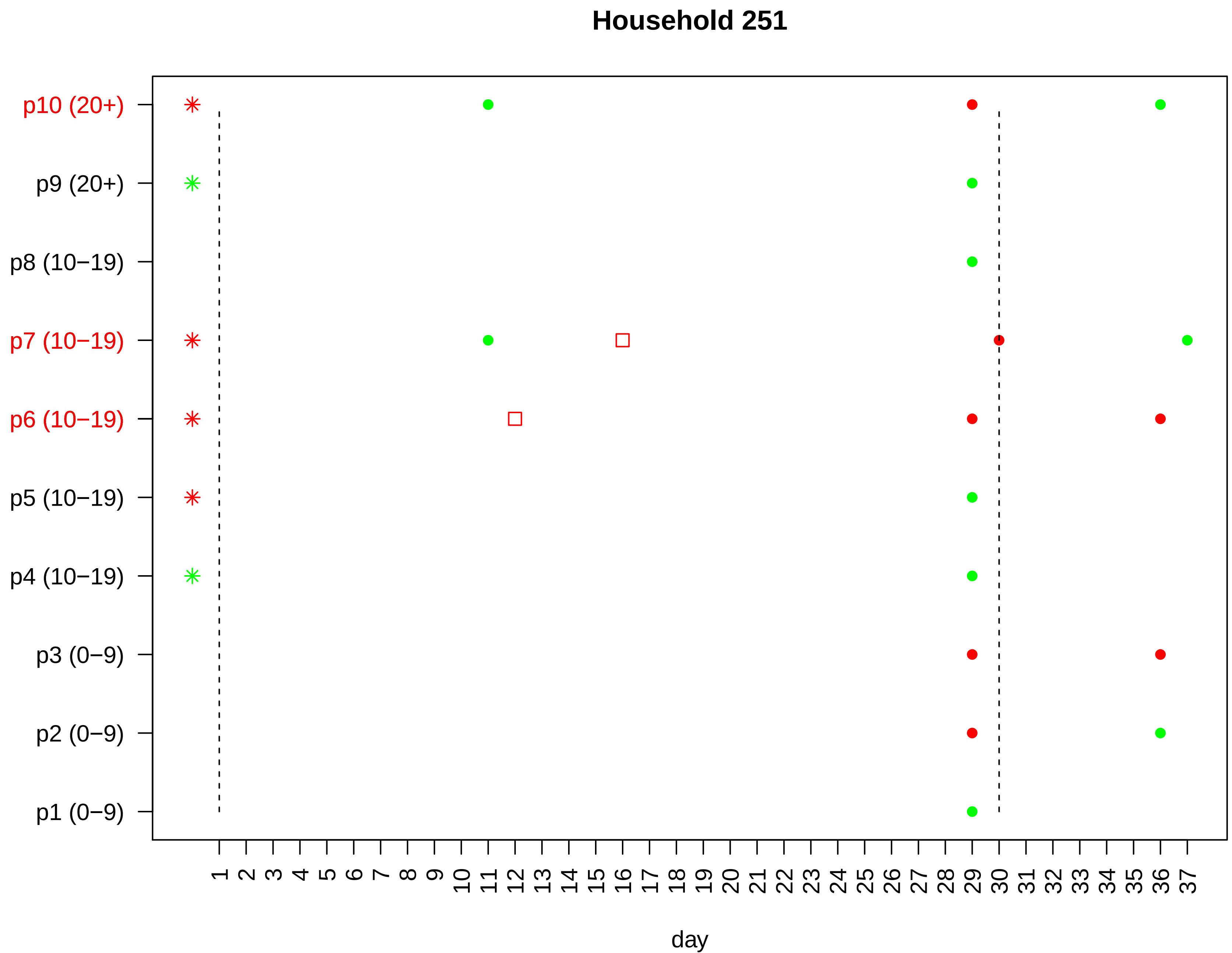

Household 251
p10 (20+)
p9 (20+)
p8 (10−19)
p7 (10−19)
p6 (10−19)
p5 (10−19)
p4 (10−19)
p3 (0−9)
p2 (0−9)
p1 (0−9)
1 2 3 4 5 6 7 8 9 10 11 12 13 14 15 16 17 18 19 20 21 22 23 24 25 26 27 28 29 30 31 32 33 34 35 36 37
day

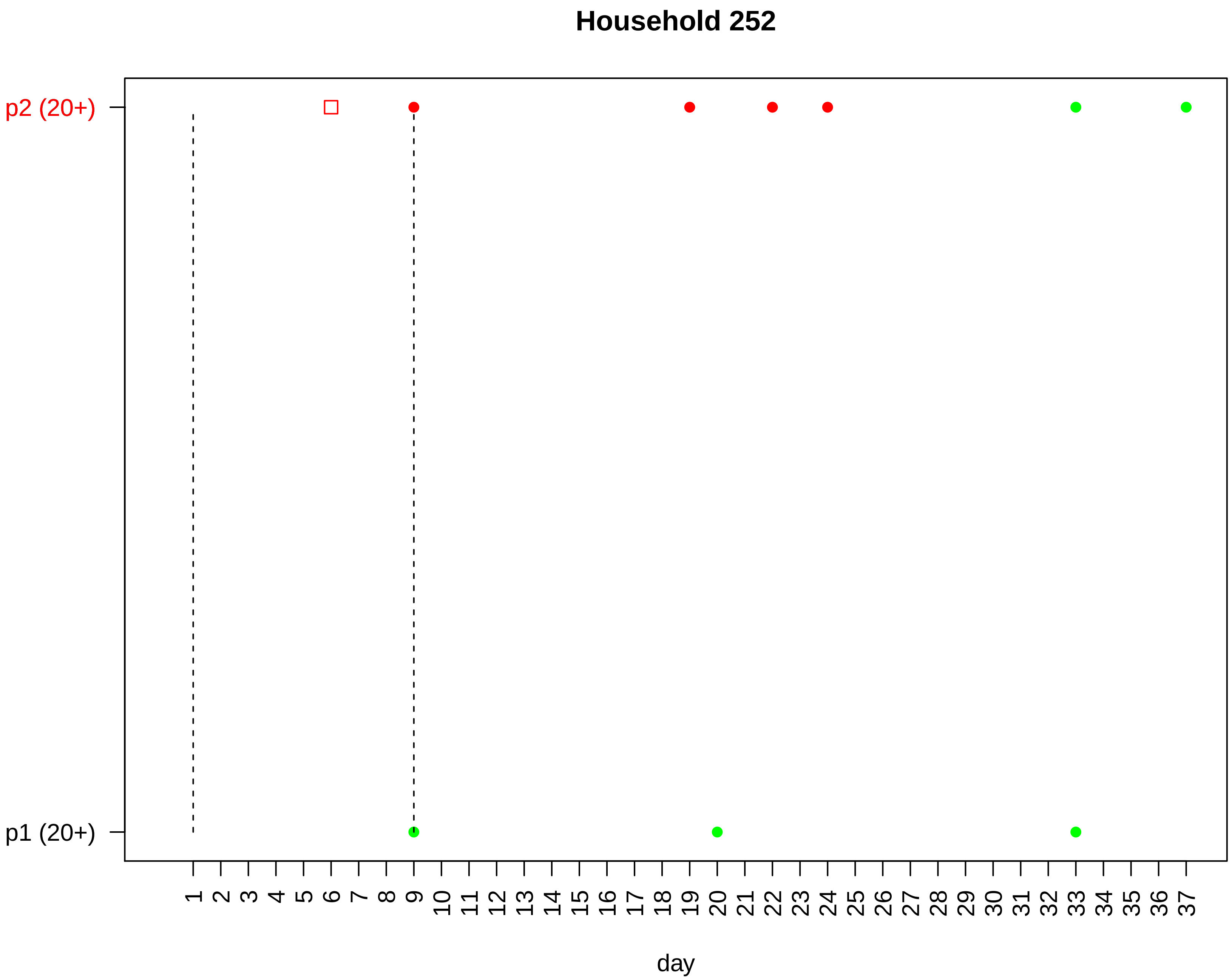
Household 252
p2 (20+)
p1 (20+)
1 2 3 4 5 6 7 8 9 10 11 12 13 14 15 16 17 18 19 20 21 22 23 24 25 26 27 28 29 30 31 32 33 34 35 36 37
day

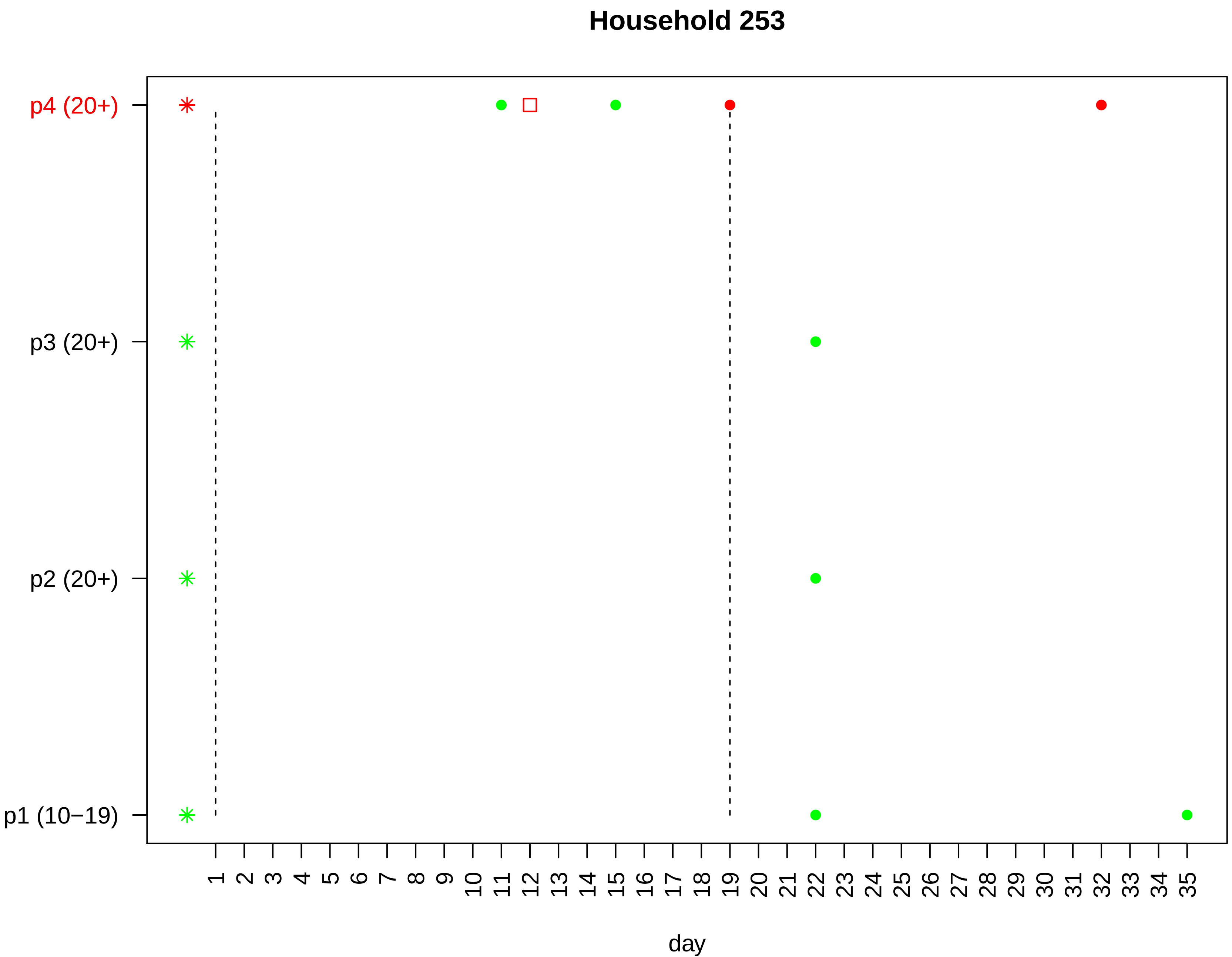

Household 253
p4 (20+)
p3 (20+)
p2 (20+)
p1 (10−19)
1 2 3 4 5 6 7 8 9 10 11 12 13 14 15 16 17 18 19 20 21 22 23 24 25 26 27 28 29 30 31 32 33 34 35
day

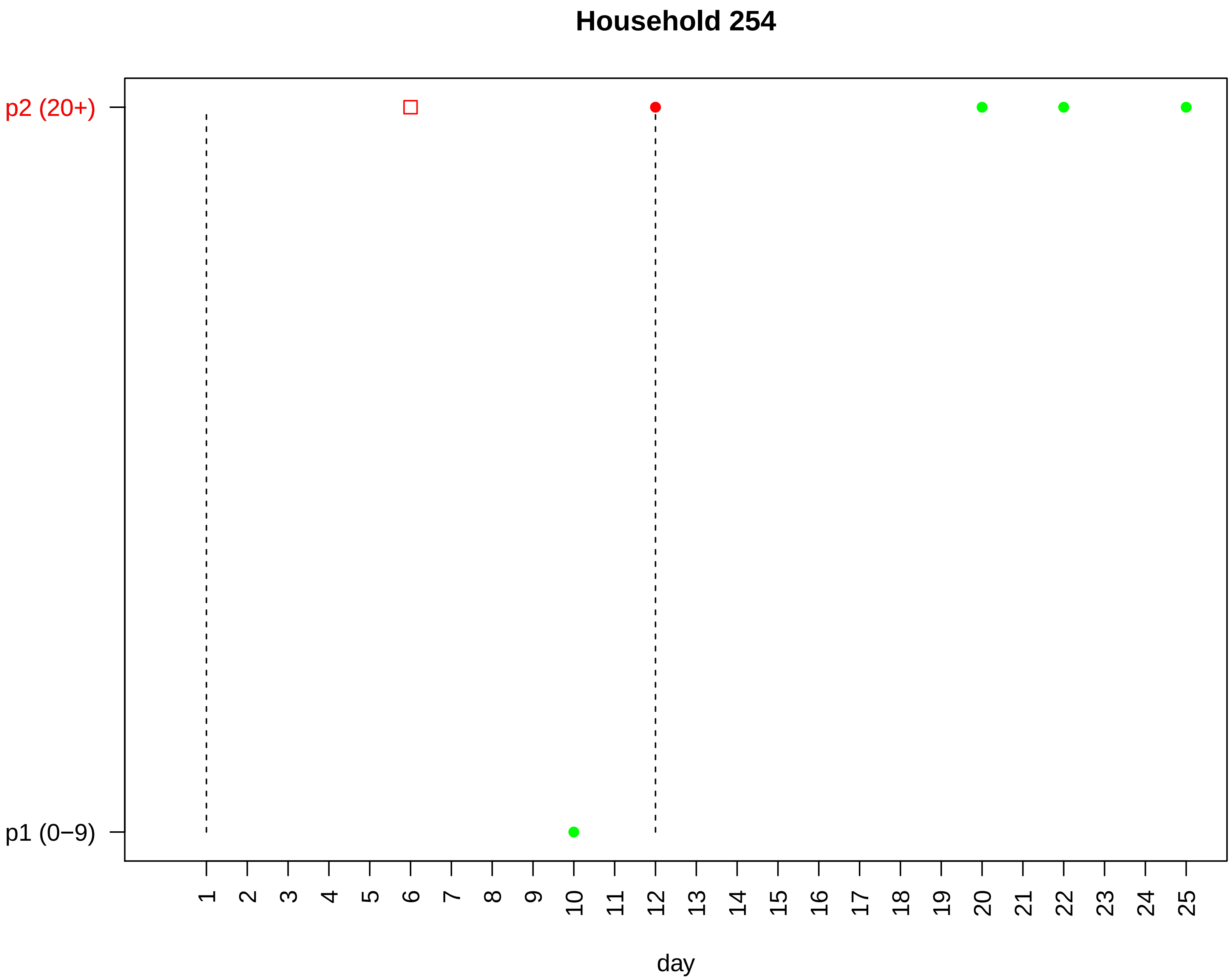
Household 254
p2 (20+)
p1 (0−9)
day

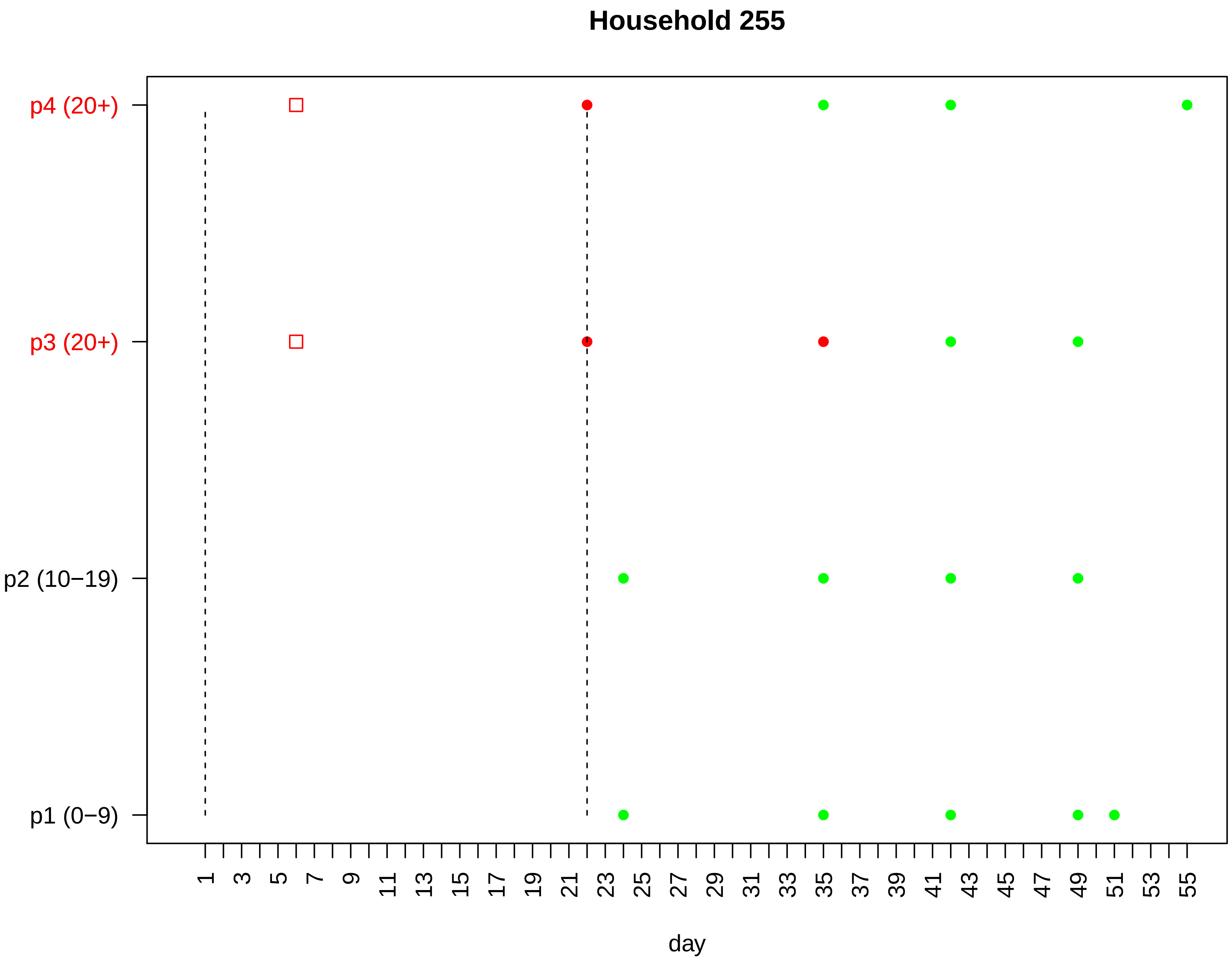

Household 255
p4 (20+)
p3 (20+)
p2 (10−19)
p1 (0−9)
1 3 5 7 9 11 13 15 17 19 21 23 25 27 29 31 33 35 37 39 41 43 45 47 49 51 53 55
day

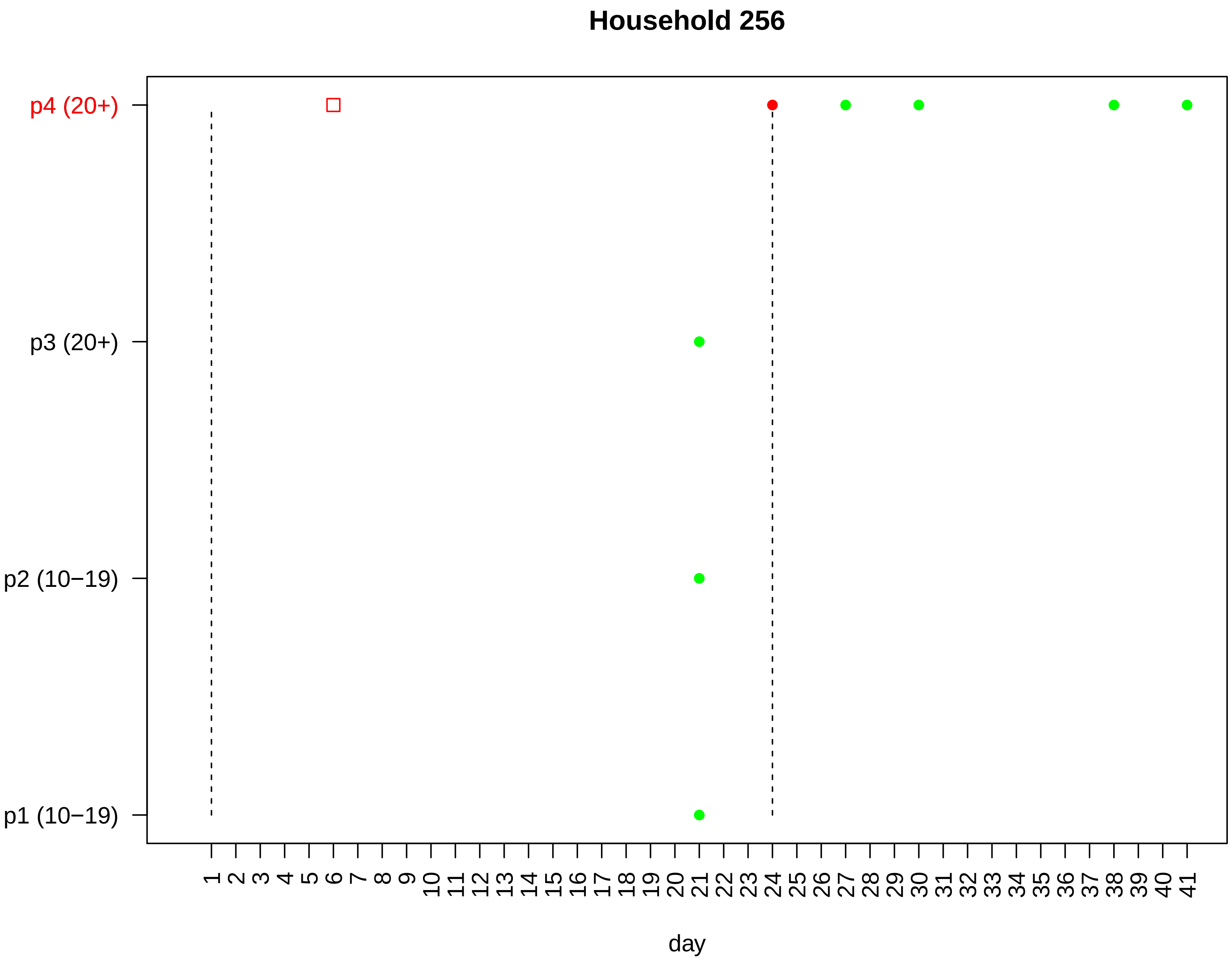

Household 256
p4 (20+)
p3 (20+)
p2 (10−19)
p1 (10−19)
1 2 3 4 5 6 7 8 9 10 11 12 13 14 15 16 17 18 19 20 21 22 23 24 25 26 27 28 29 30 31 32 33 34 35 36 37 38 39 40 41
day

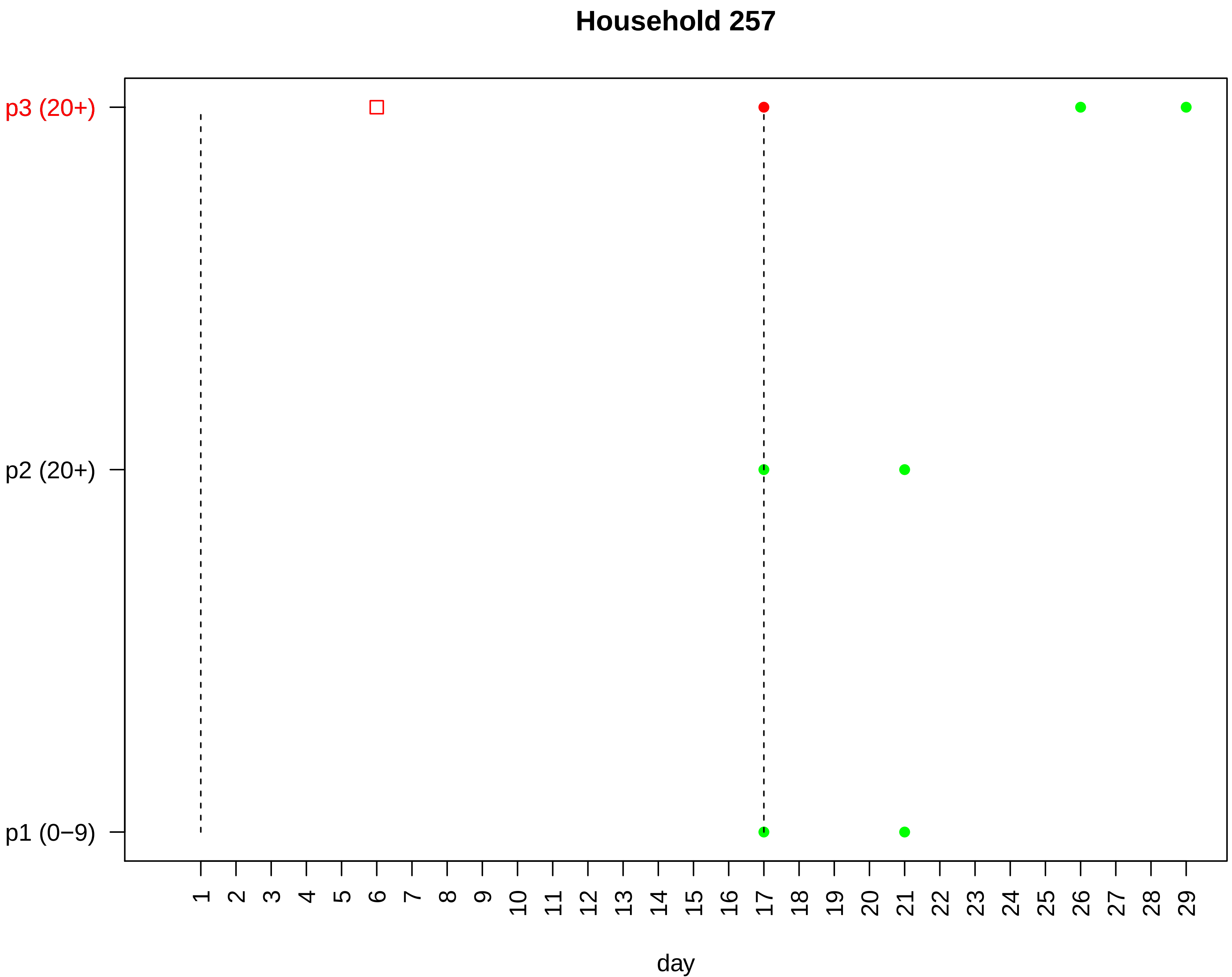

Household 257
p3 (20+)
p2 (20+)
p1 (0−9)
1 2 3 4 5 6 7 8 9 10 11 12 13 14 15 16 17 18 19 20 21 22 23 24 25 26 27 28 29
day

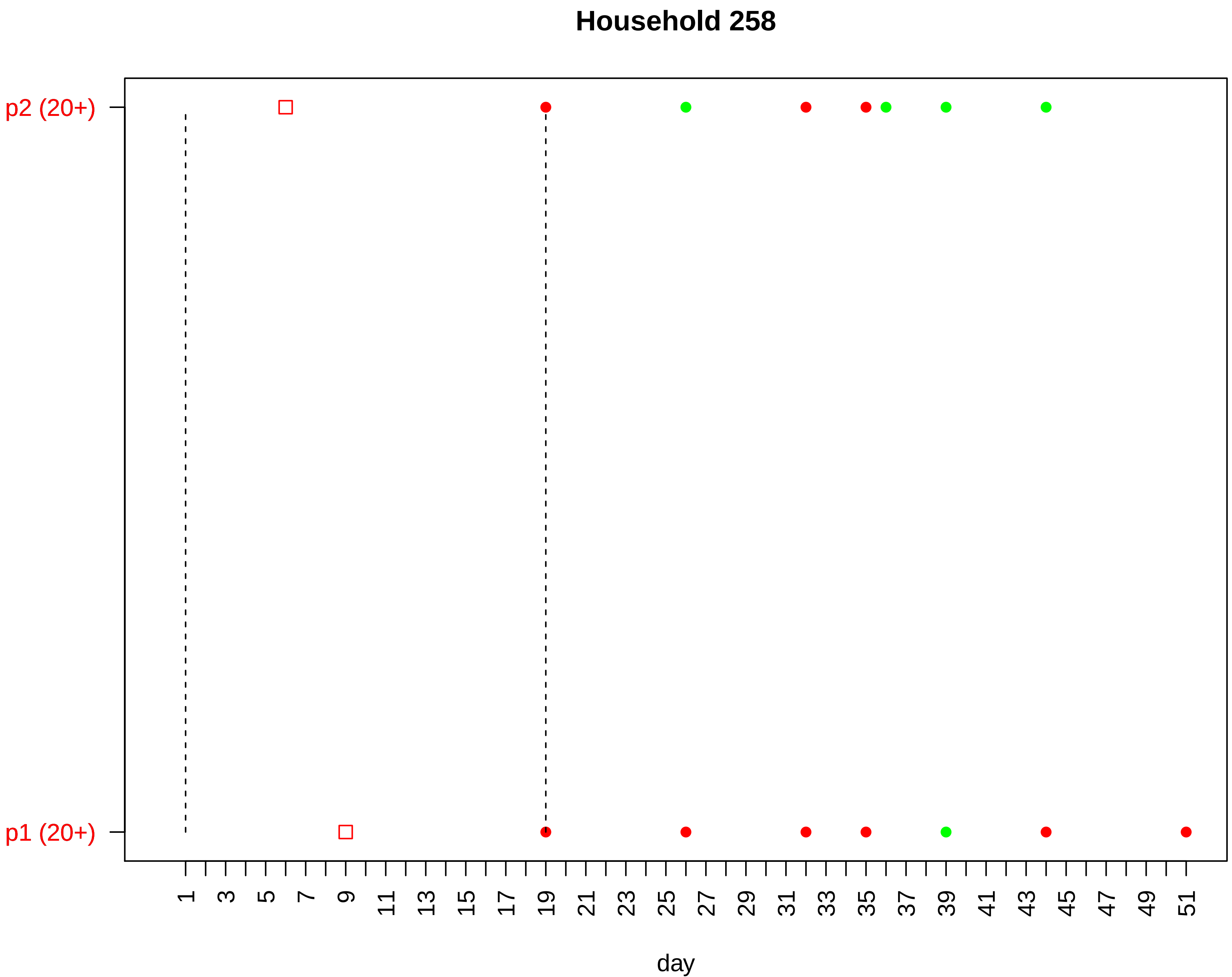

Household 258
p2 (20+)
p1 (20+)
1
3
5
7
9
11
13
15
17
19
21
23
25
27
29
31
33
35
37
39
41
43
45
47
49
51
day

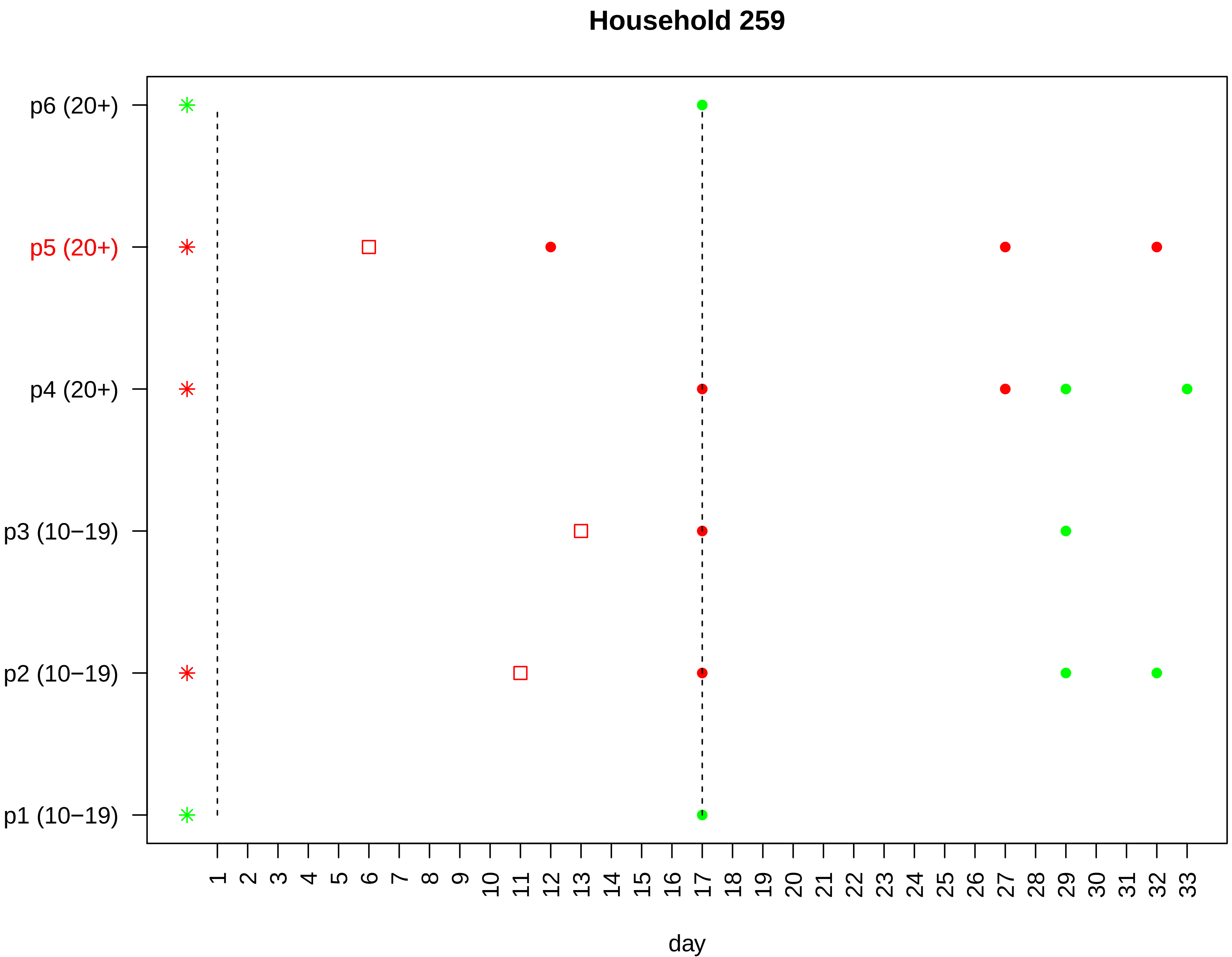

Household 259
p6 (20+)
p5 (20+)
p4 (20+)
p3 (10−19)
p2 (10−19)
p1 (10−19)
day

#### Household 260

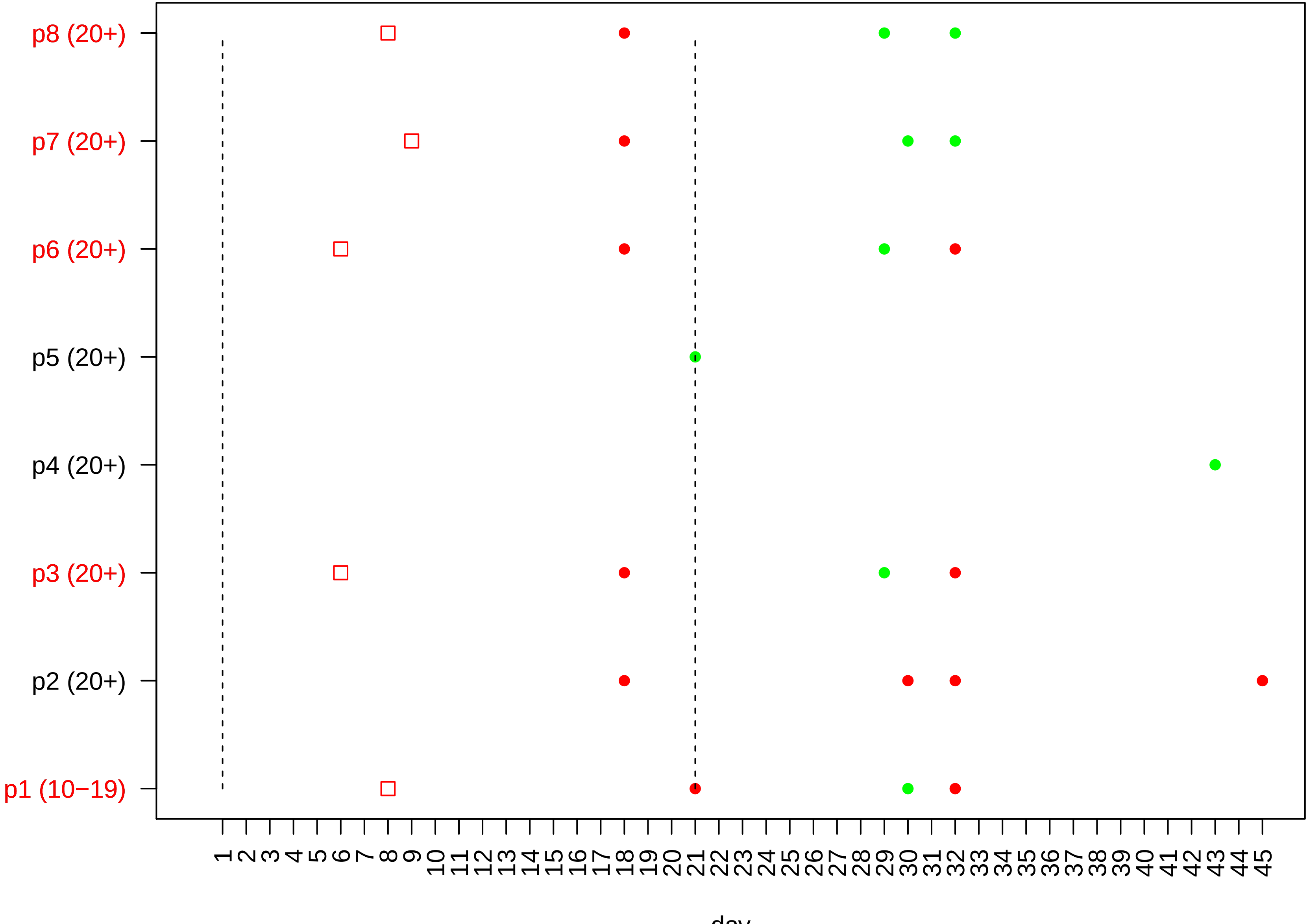

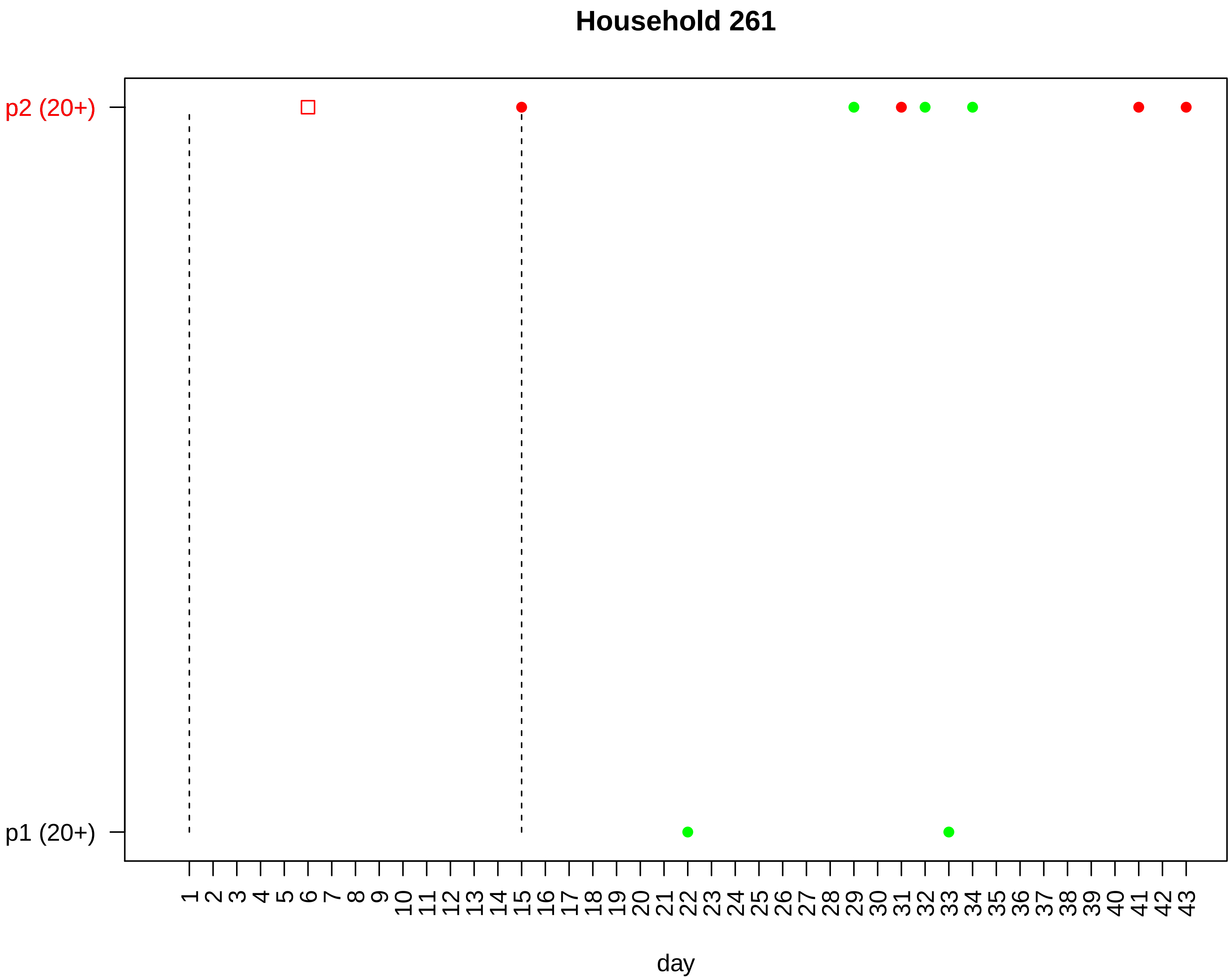

Household 261
p2 (20+)
p1 (20+)
day

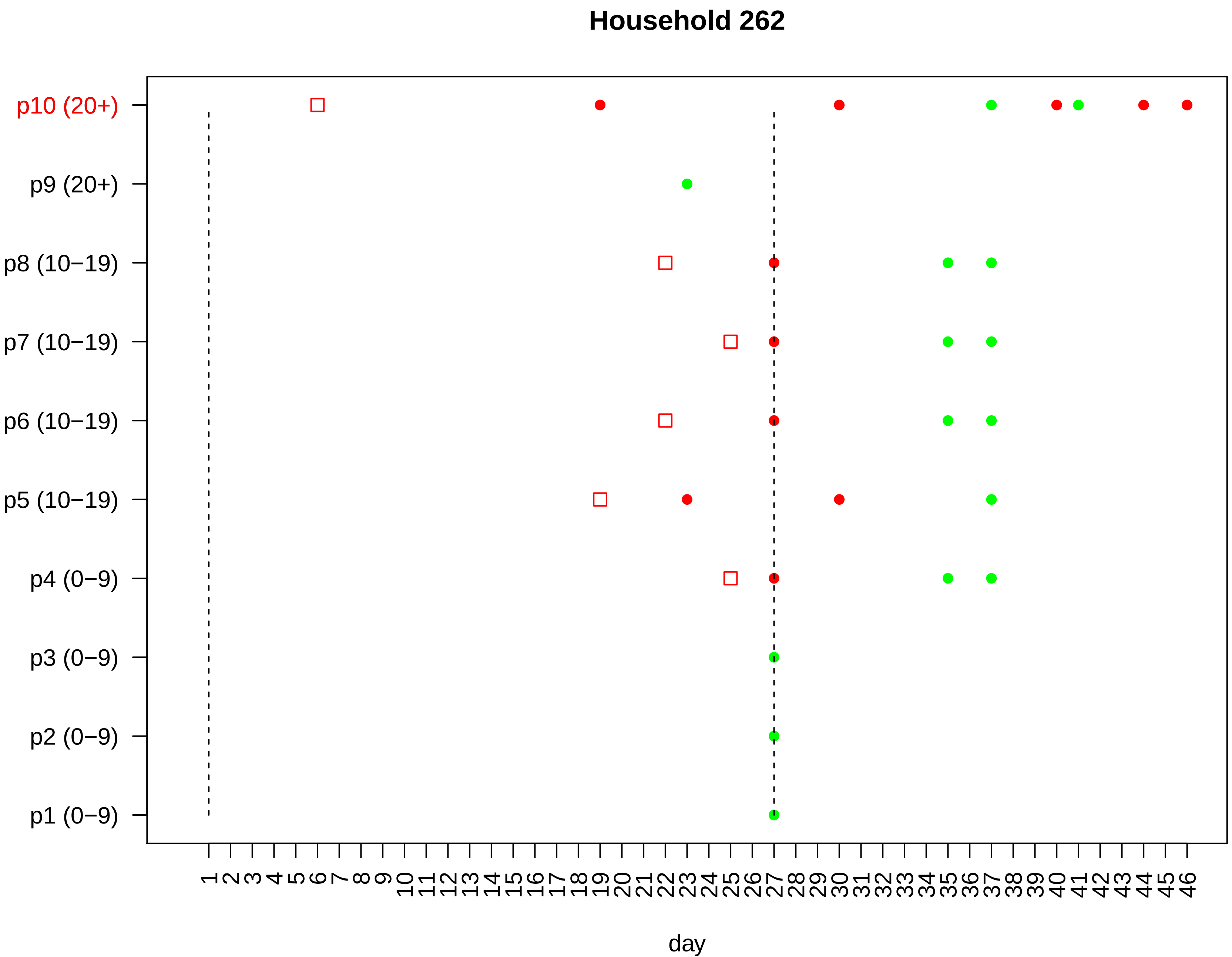

Household 262
p10 (20+)
p9 (20+)
p8 (10−19)
p7 (10−19)
p6 (10−19)
p5 (10−19)
p4 (0−9)
p3 (0−9)
p2 (0−9)
p1 (0−9)
1 2 3 4 5 6 7 8 9 10 11 12 13 14 15 16 17 18 19 20 21 22 23 24 25 26 27 28 29 30 31 32 33 34 35 36 37 38 39 40 41 42 43 44 45 46
day

#### Household 263

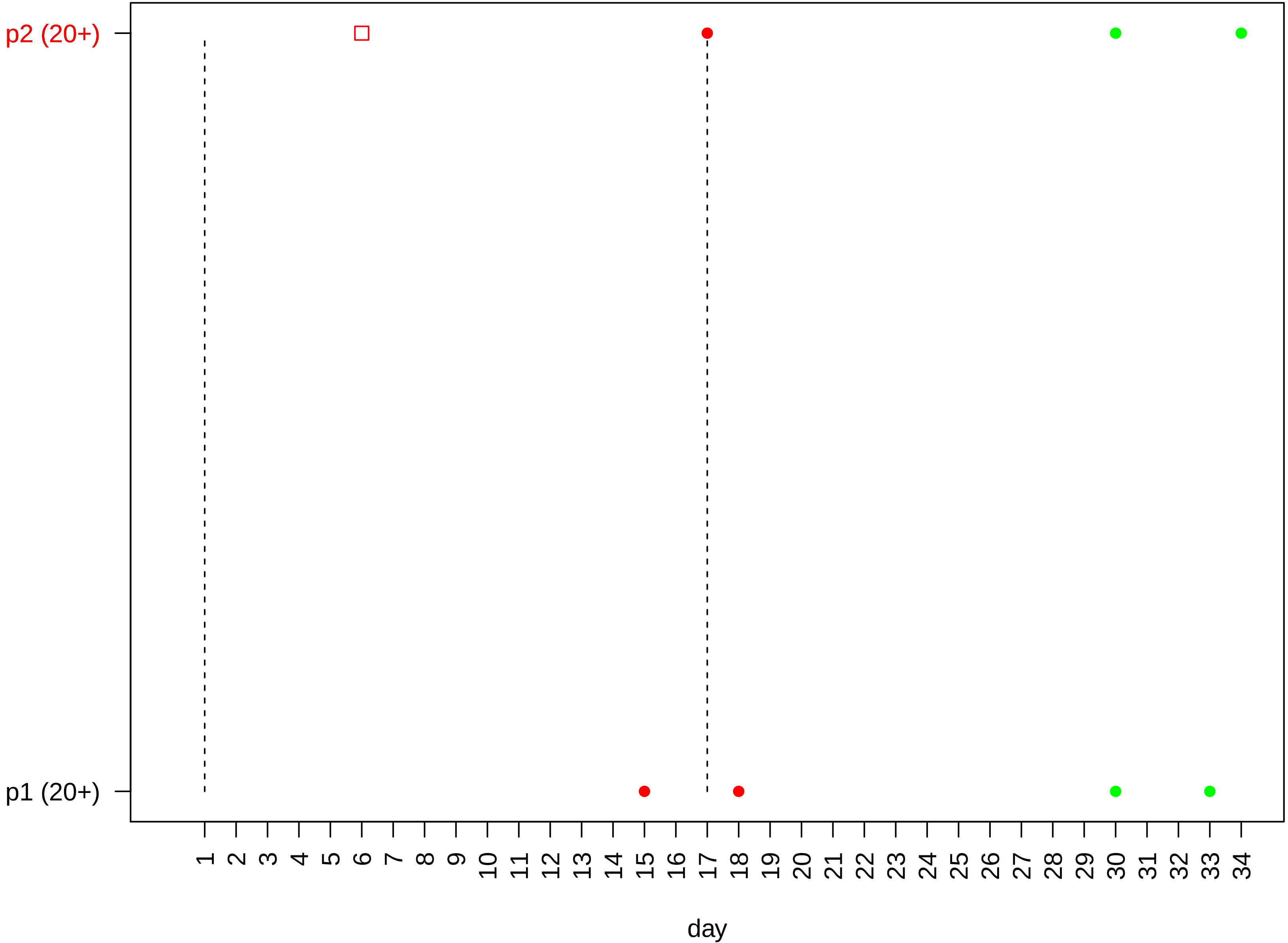

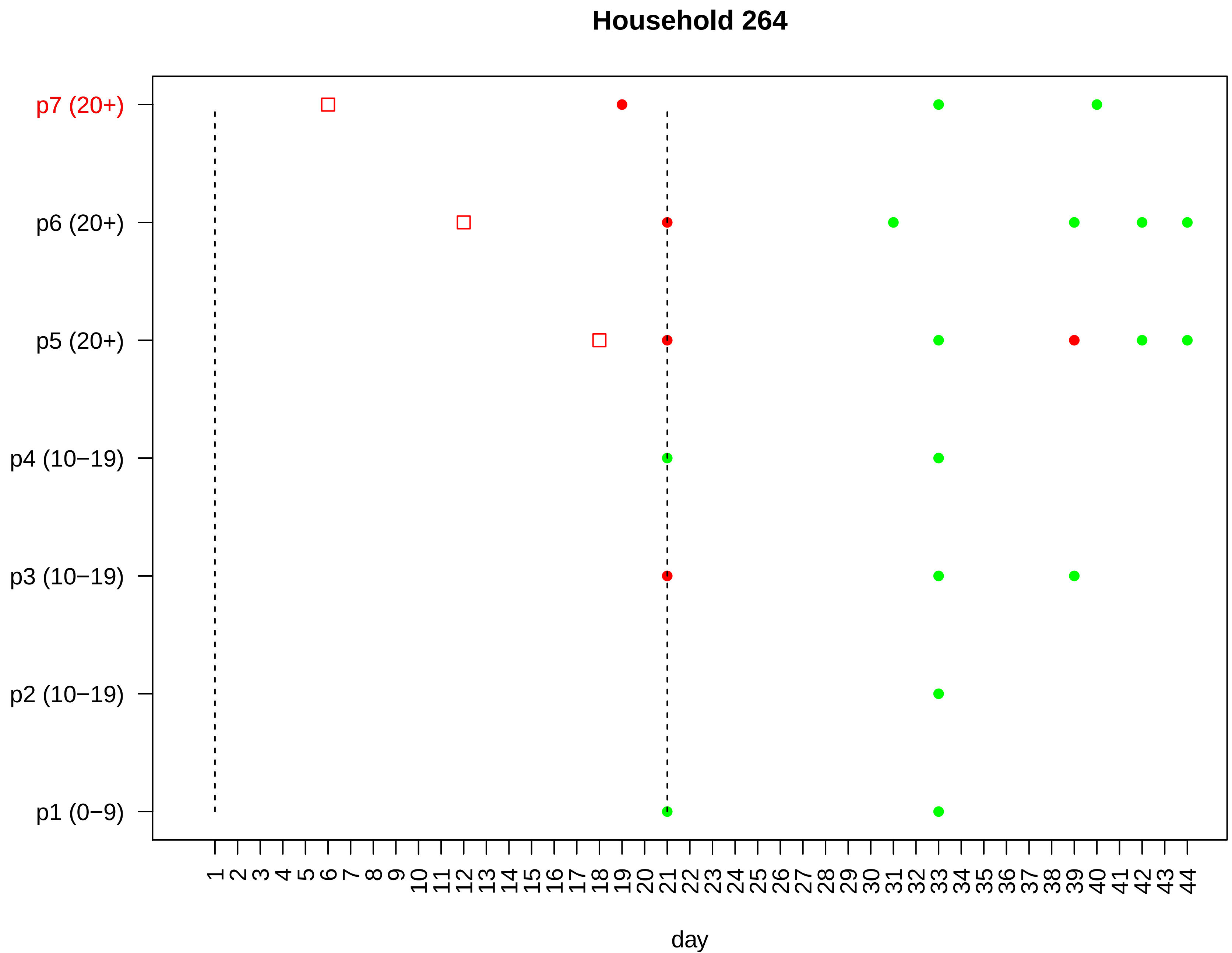

Household 264
p7 (20+)
p6 (20+)
p5 (20+)
p4 (10−19)
p3 (10−19)
p2 (10−19)
p1 (0−9)
1 2 3 4 5 6 7 8 9 10 11 12 13 14 15 16 17 18 19 20 21 22 23 24 25 26 27 28 29 30 31 32 33 34 35 36 37 38 39 40 41 42 43 44
day

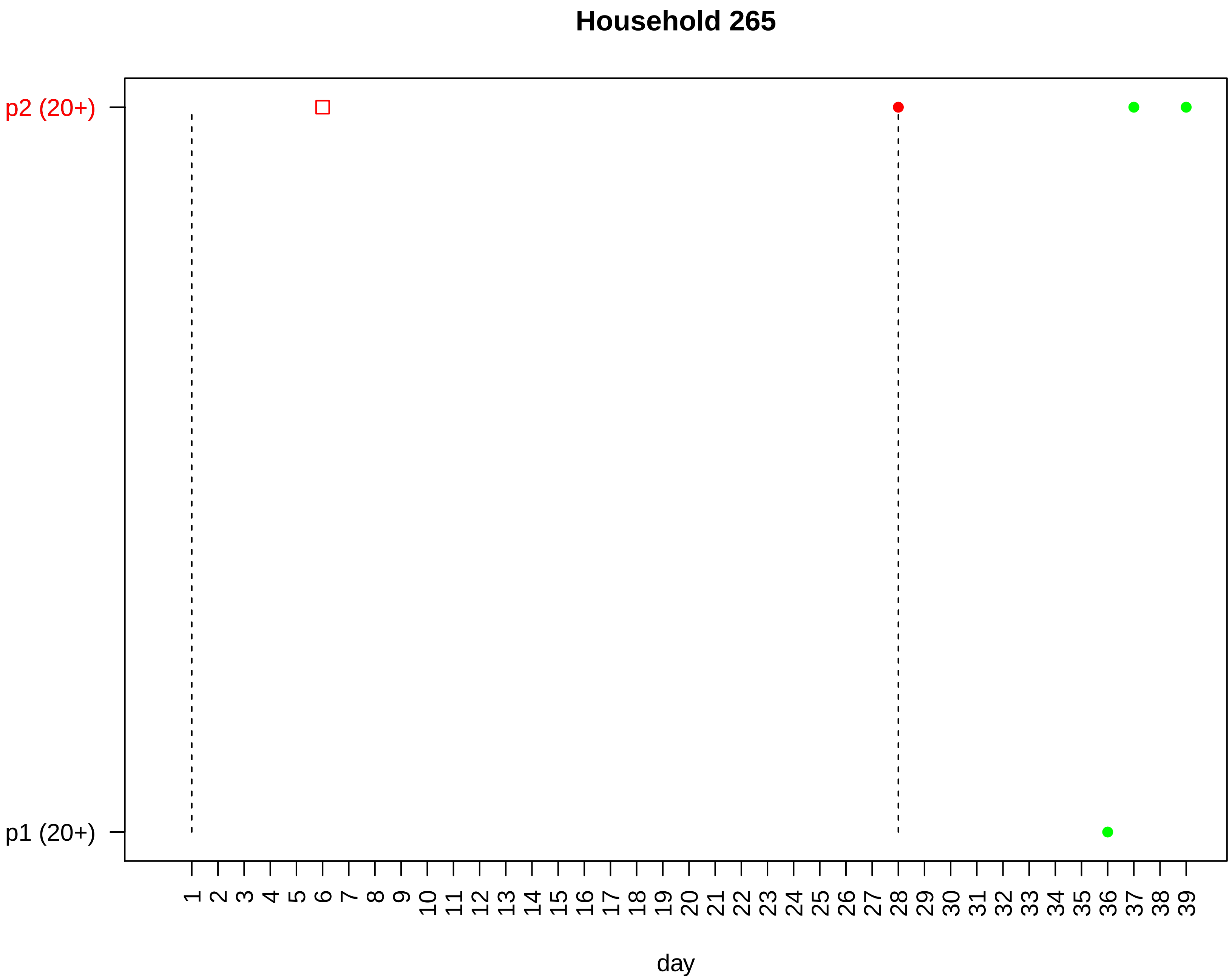

Household 265
p2 (20+)
p1 (20+)
1 2 3 4 5 6 7 8 9 10 11 12 13 14 15 16 17 18 19 20 21 22 23 24 25 26 27 28 29 30 31 32 33 34 35 36 37 38 39
day

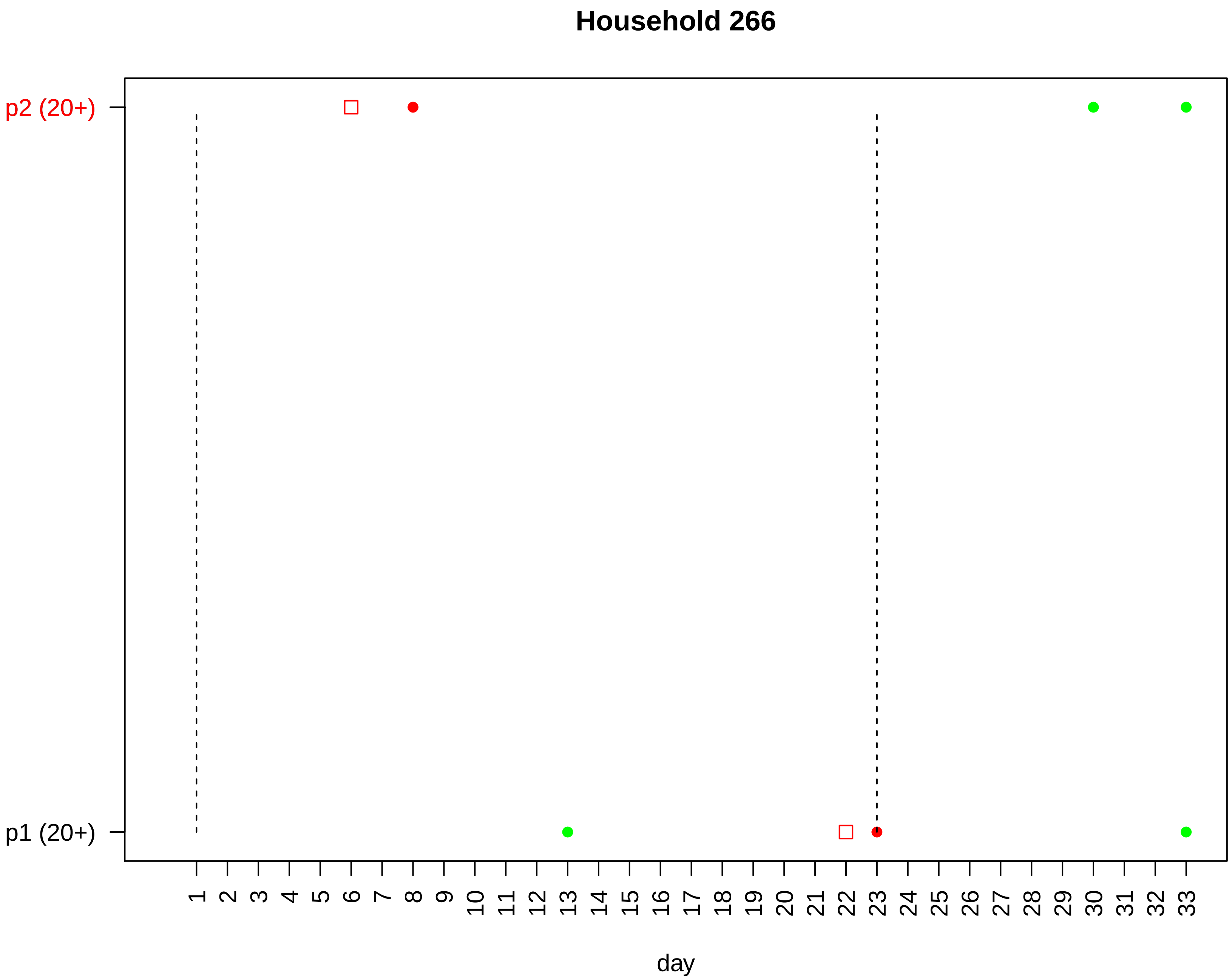

Household 266
p2 (20+)
p1 (20+)
day

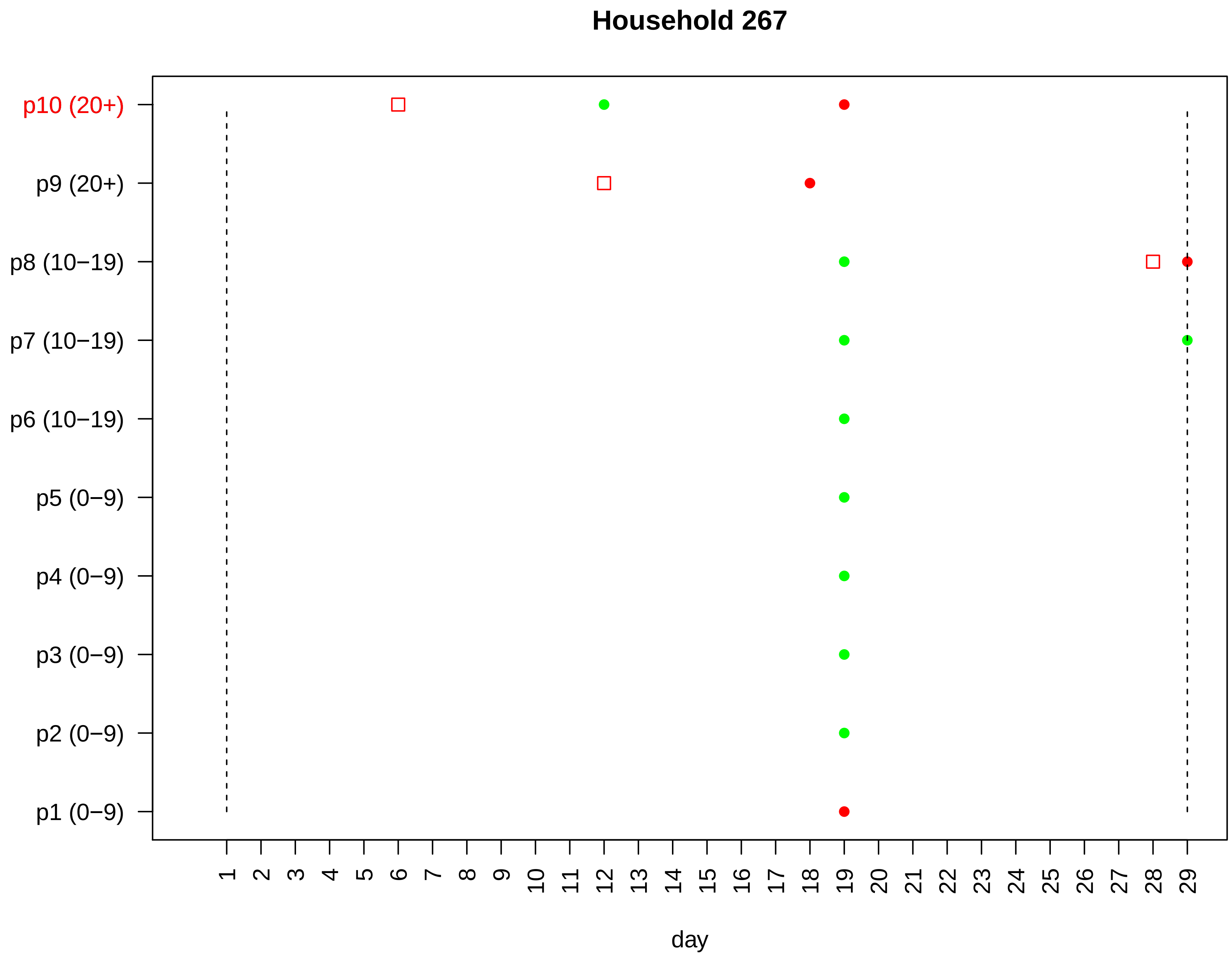

Household 267
p10 (20+)
p9 (20+)
p8 (10−19)
p7 (10−19)
p6 (10−19)
p5 (0−9)
p4 (0−9)
p3 (0−9)
p2 (0−9)
p1 (0−9)
1 2 3 4 5 6 7 8 9 10 11 12 13 14 15 16 17 18 19 20 21 22 23 24 25 26 27 28 29
day

#### Household 268

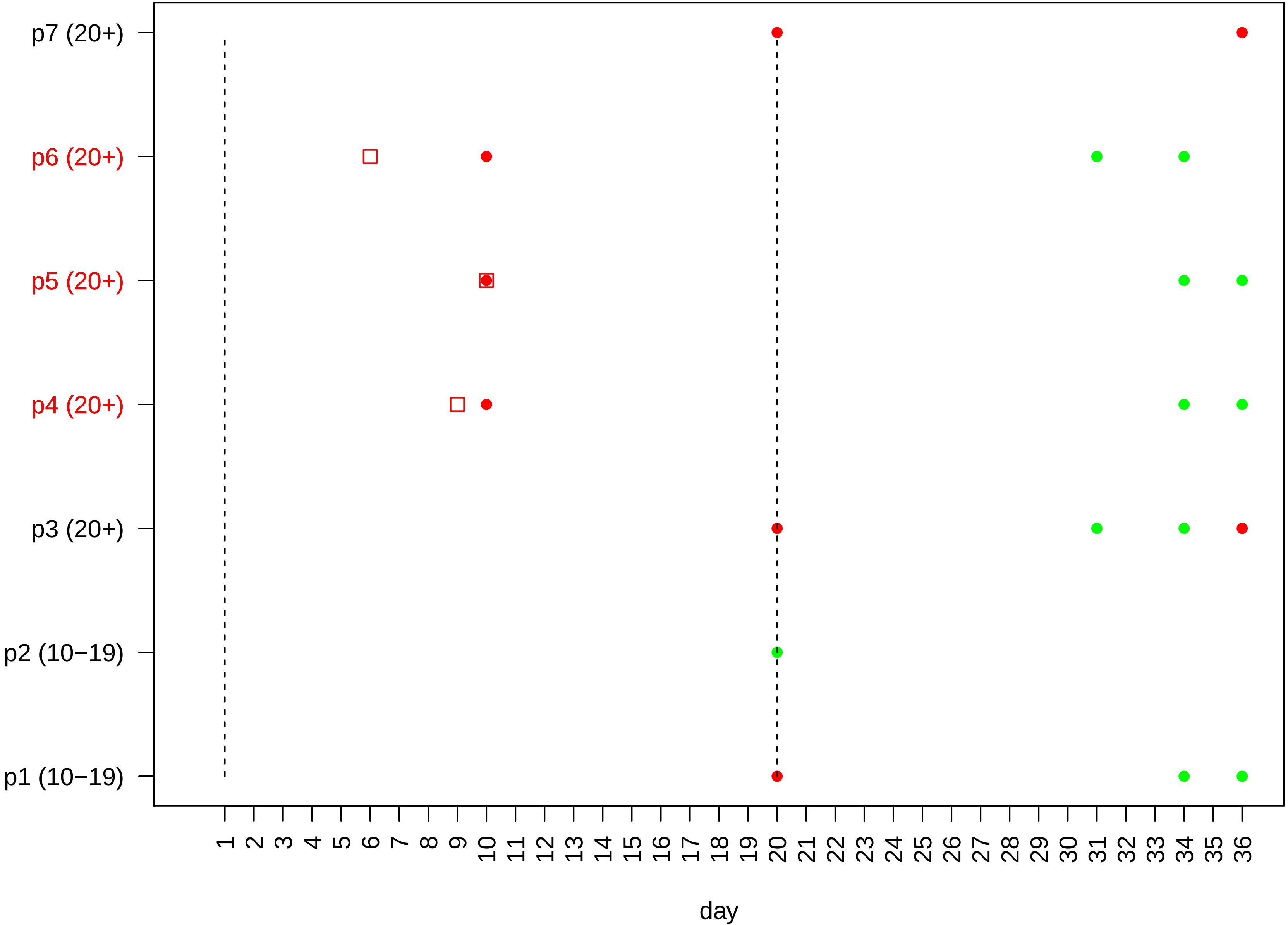

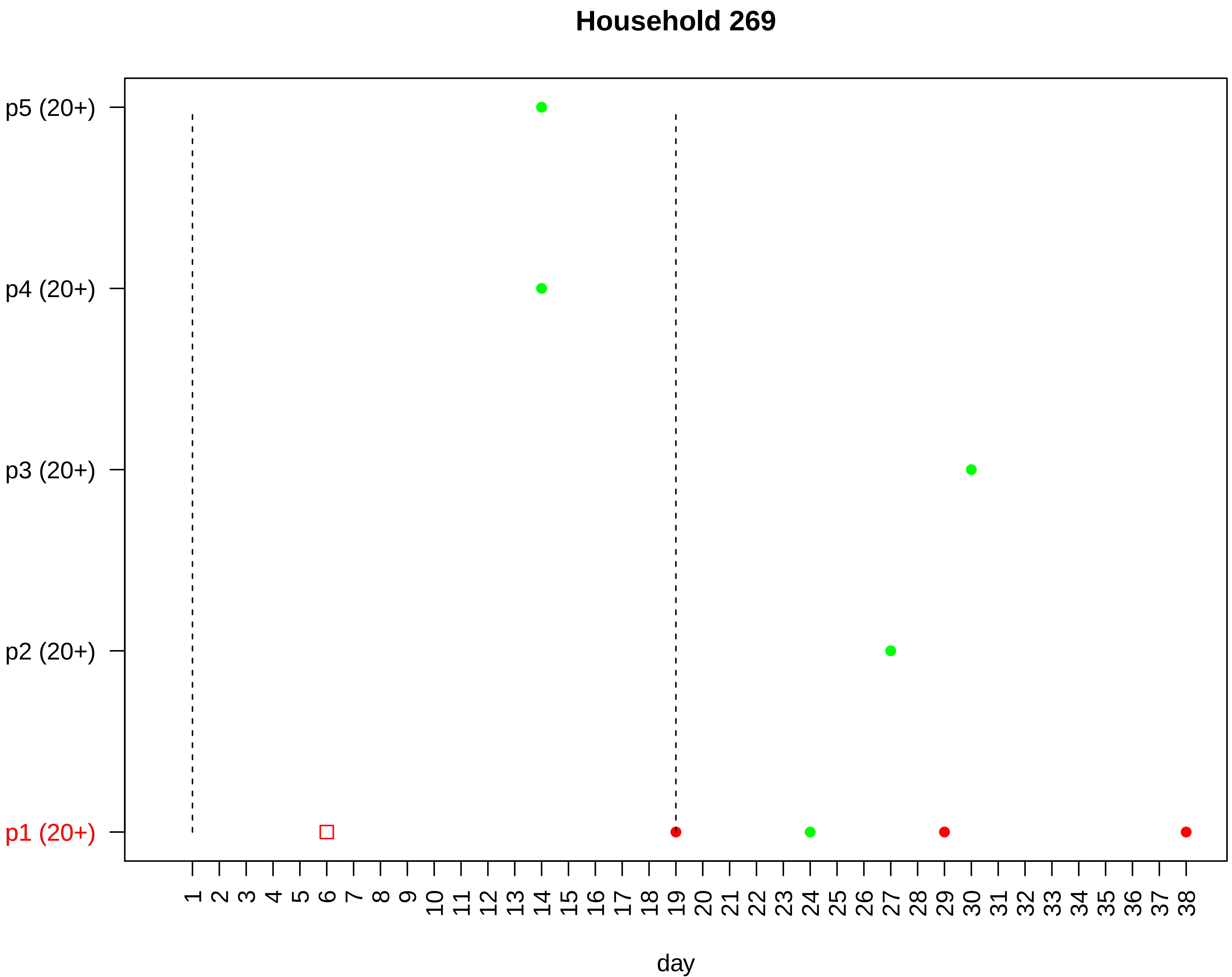

Household 269
p5 (20+)
p4 (20+)
p3 (20+)
p2 (20+)
p1 (20+)
1 2 3 4 5 6 7 8 9 10 11 12 13 14 15 16 17 18 19 20 21 22 23 24 25 26 27 28 29 30 31 32 33 34 35 36 37 38
day

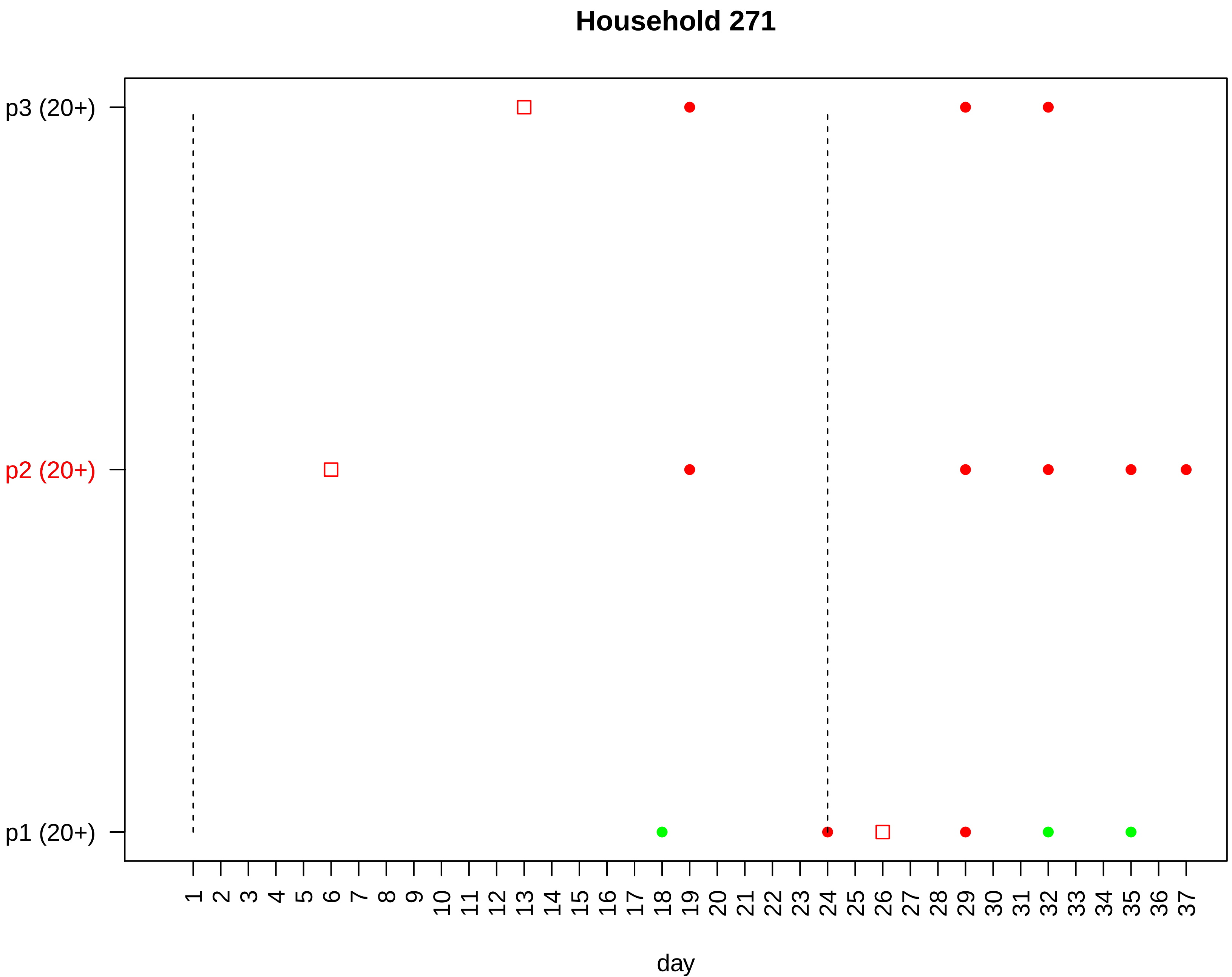

Household 271
p3 (20+)
p2 (20+)
p1 (20+)
day

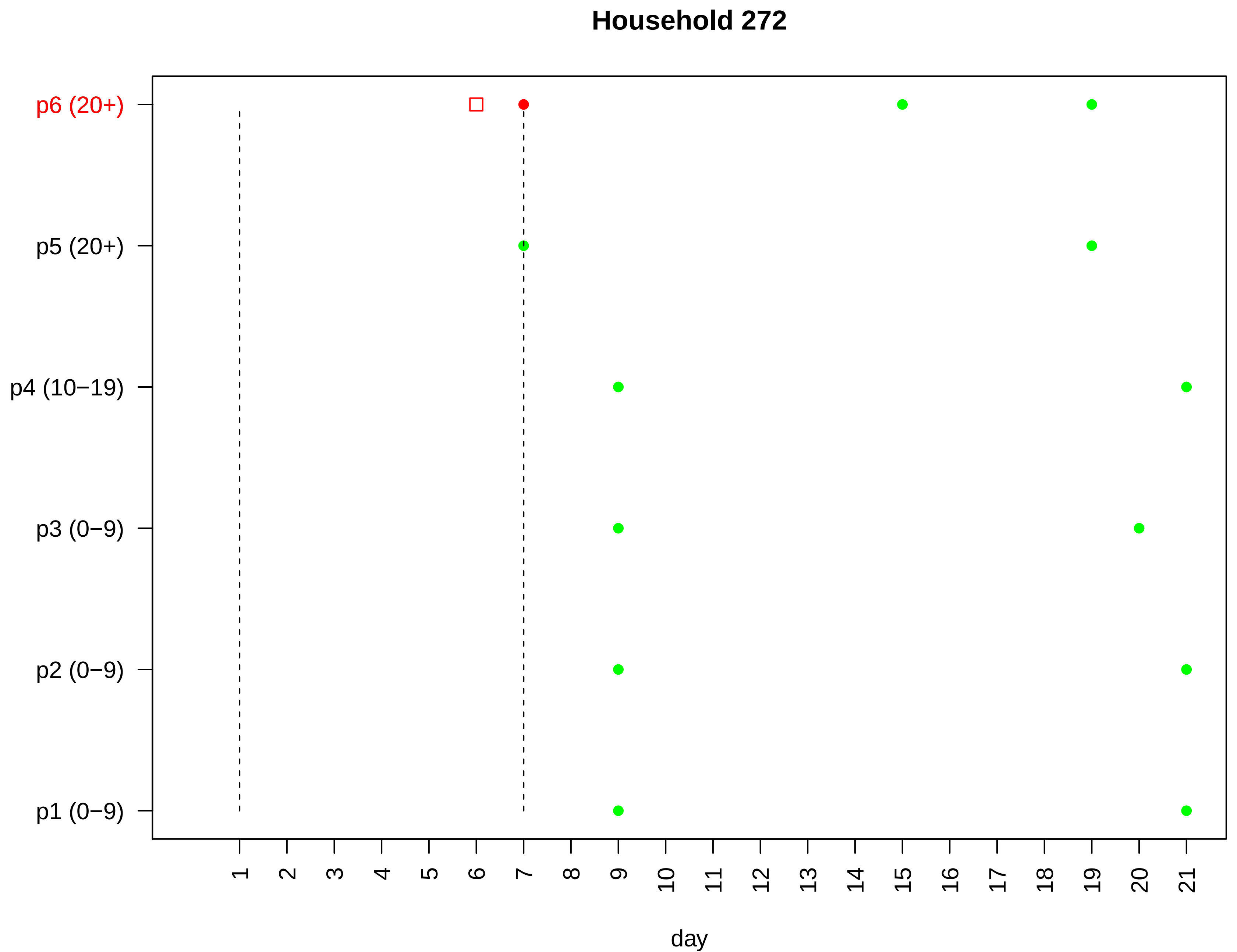

Household 272
p6 (20+)
p5 (20+)
p4 (10−19)
p3 (0−9)
p2 (0−9)
p1 (0−9)
day

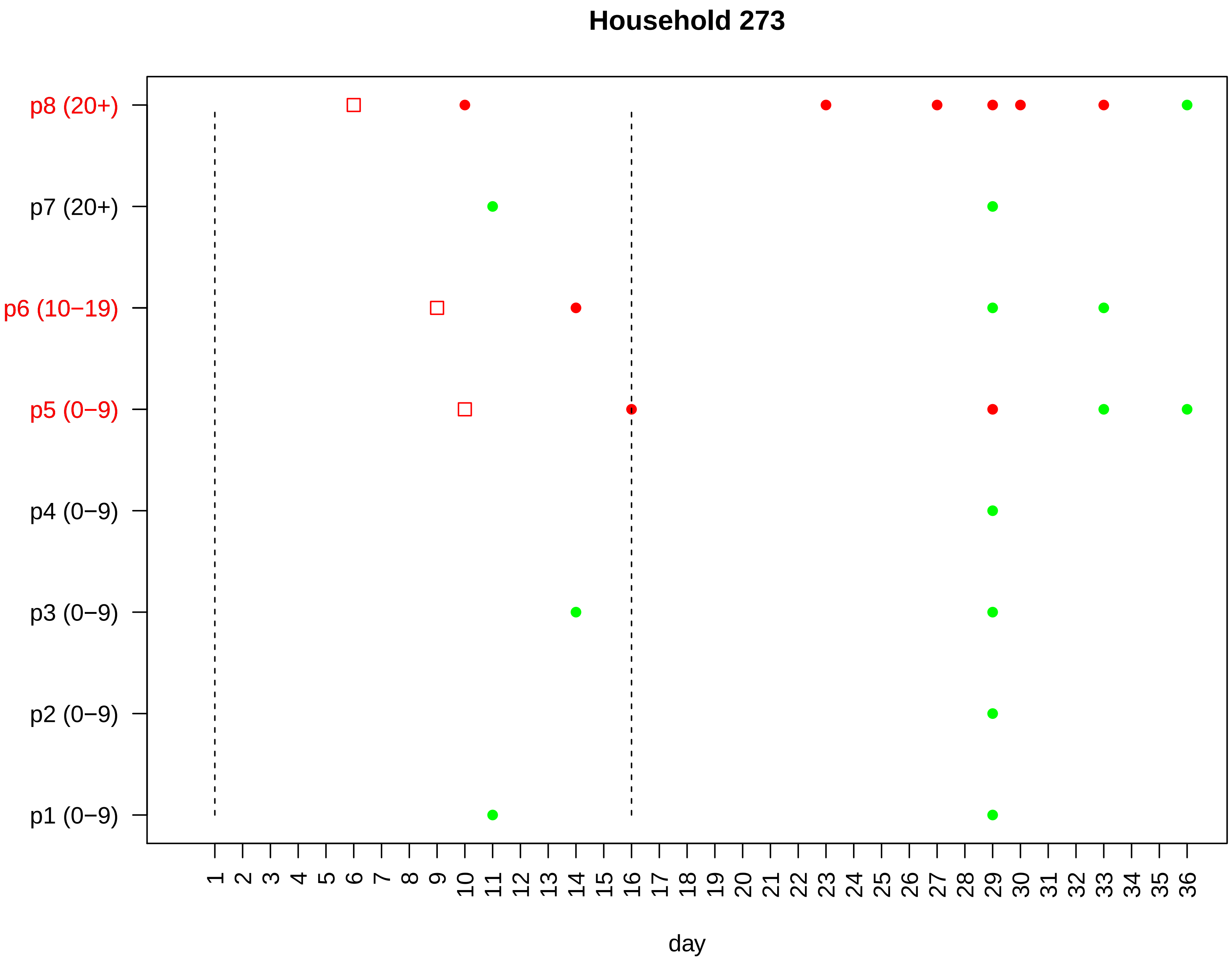

Household 273
p8 (20+)
p7 (20+)
p6 (10−19)
p5 (0−9)
p4 (0−9)
p3 (0−9)
p2 (0−9)
p1 (0−9)
1 2 3 4 5 6 7 8 9 10 11 12 13 14 15 16 17 18 19 20 21 22 23 24 25 26 27 28 29 30 31 32 33 34 35 36
day

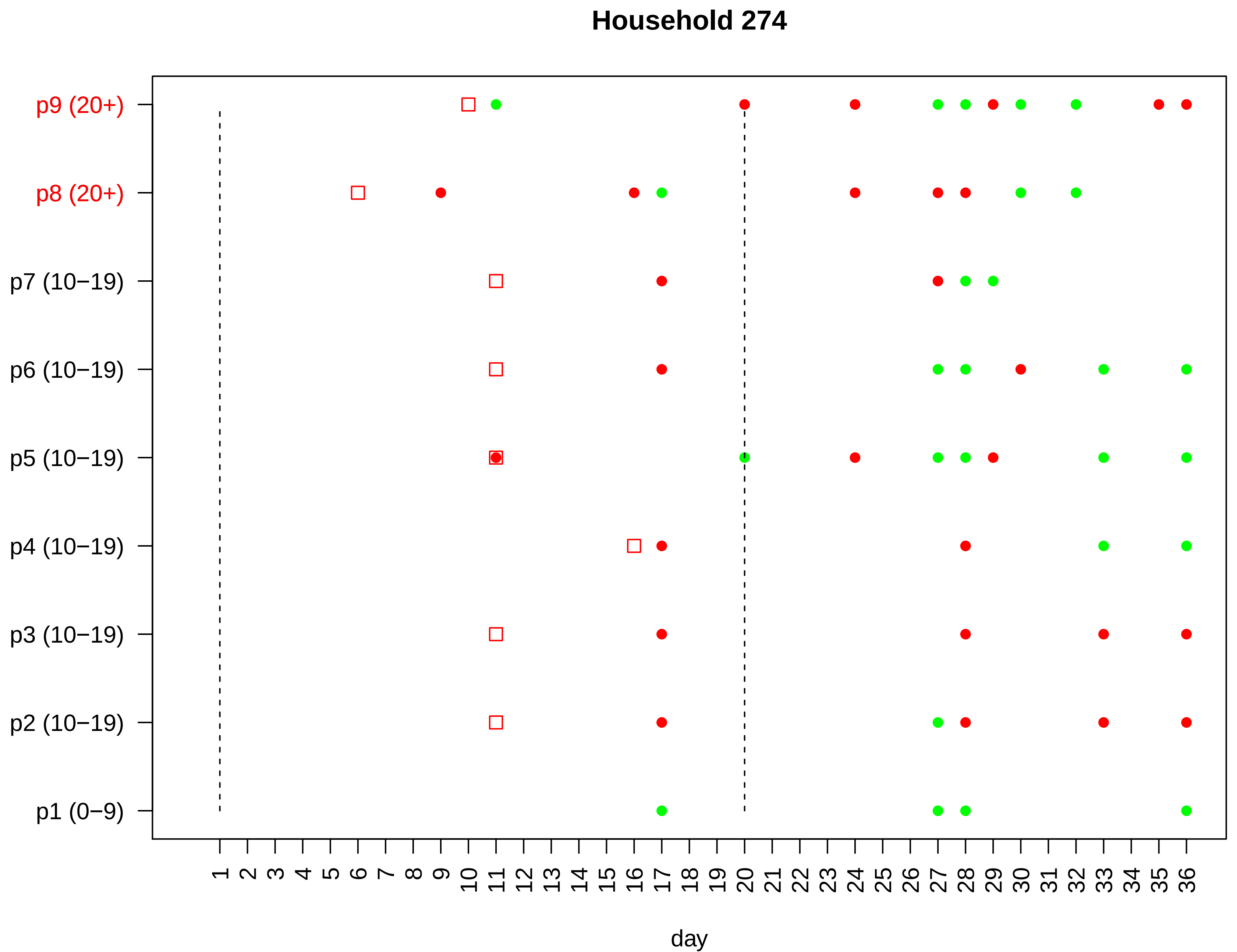

Household 274
p9 (20+)
p8 (20+)
p7 (10−19)
p6 (10−19)
p5 (10−19)
p4 (10−19)
p3 (10−19)
p2 (10−19)
p1 (0−9)
1 2 3 4 5 6 7 8 9 10 11 12 13 14 15 16 17 18 19 20 21 22 23 24 25 26 27 28 29 30 31 32 33 34 35 36
day

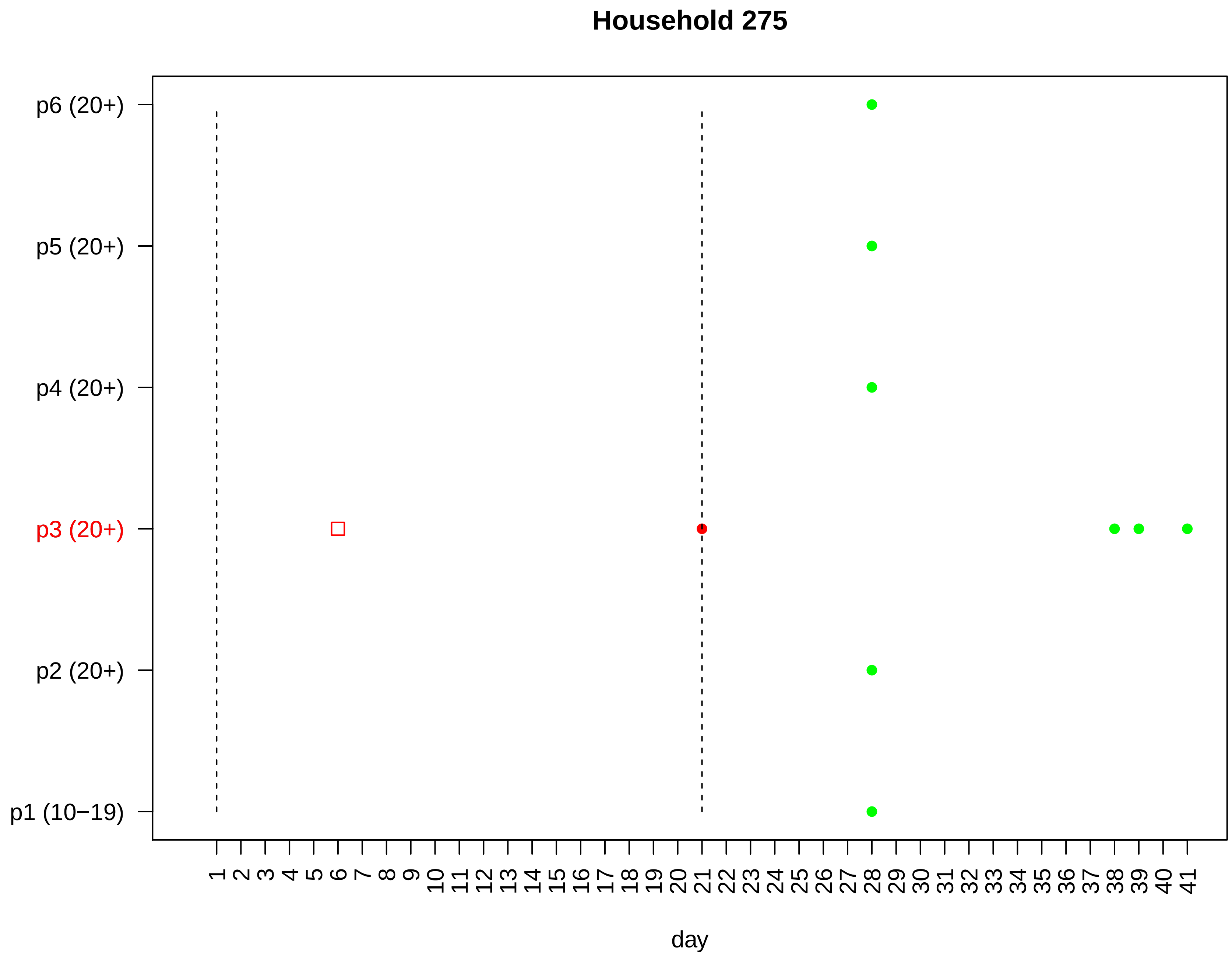

Household 275
p6 (20+)
p5 (20+)
p4 (20+)
p3 (20+)
p2 (20+)
p1 (10−19)
1 2 3 4 5 6 7 8 9 10 11 12 13 14 15 16 17 18 19 20 21 22 23 24 25 26 27 28 29 30 31 32 33 34 35 36 37 38 39 40 41
day

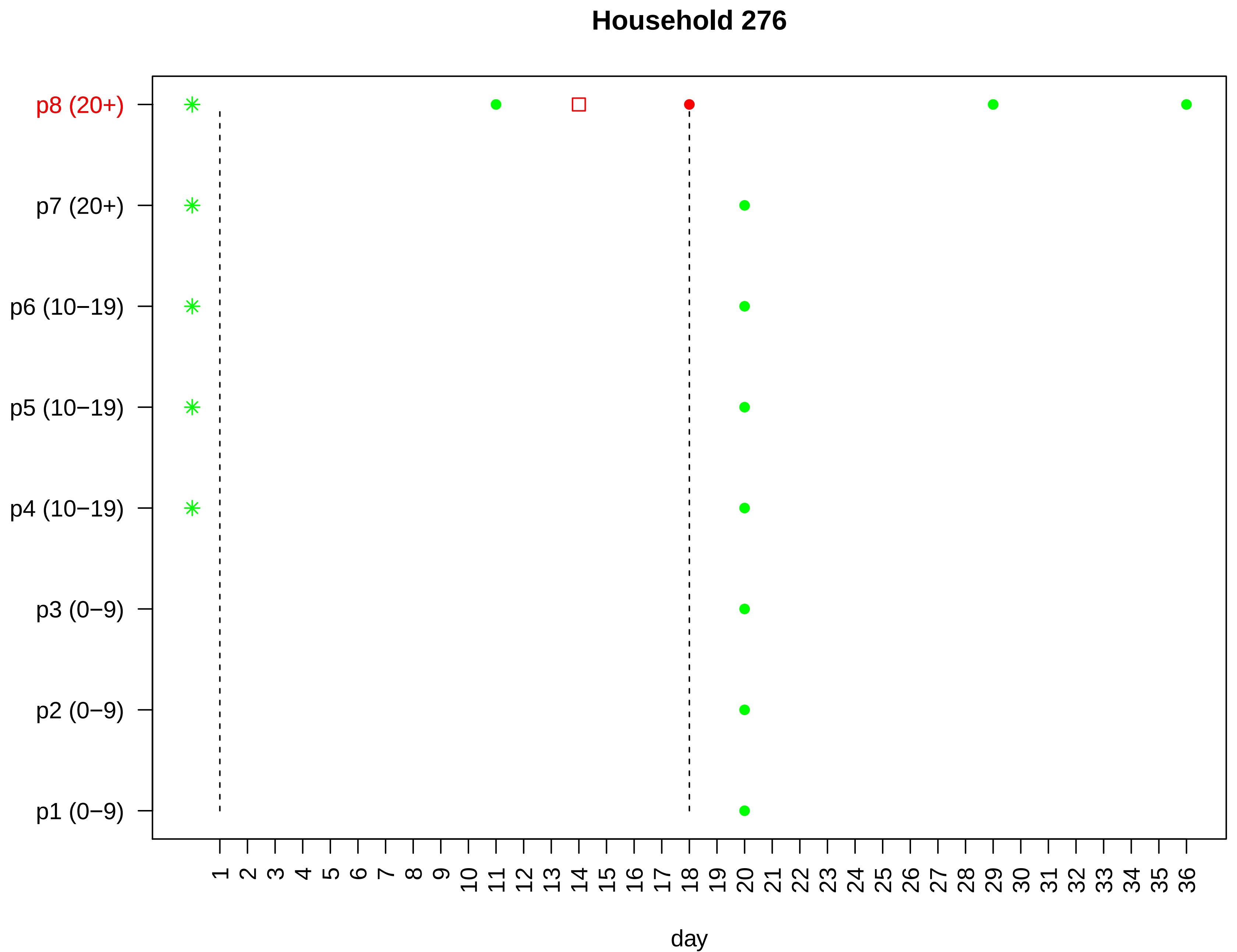

Household 276
p8 (20+)
p7 (20+)
p6 (10−19)
p5 (10−19)
p4 (10−19)
p3 (0−9)
p2 (0−9)
p1 (0−9)
1 2 3 4 5 6 7 8 9 10 11 12 13 14 15 16 17 18 19 20 21 22 23 24 25 26 27 28 29 30 31 32 33 34 35 36
day

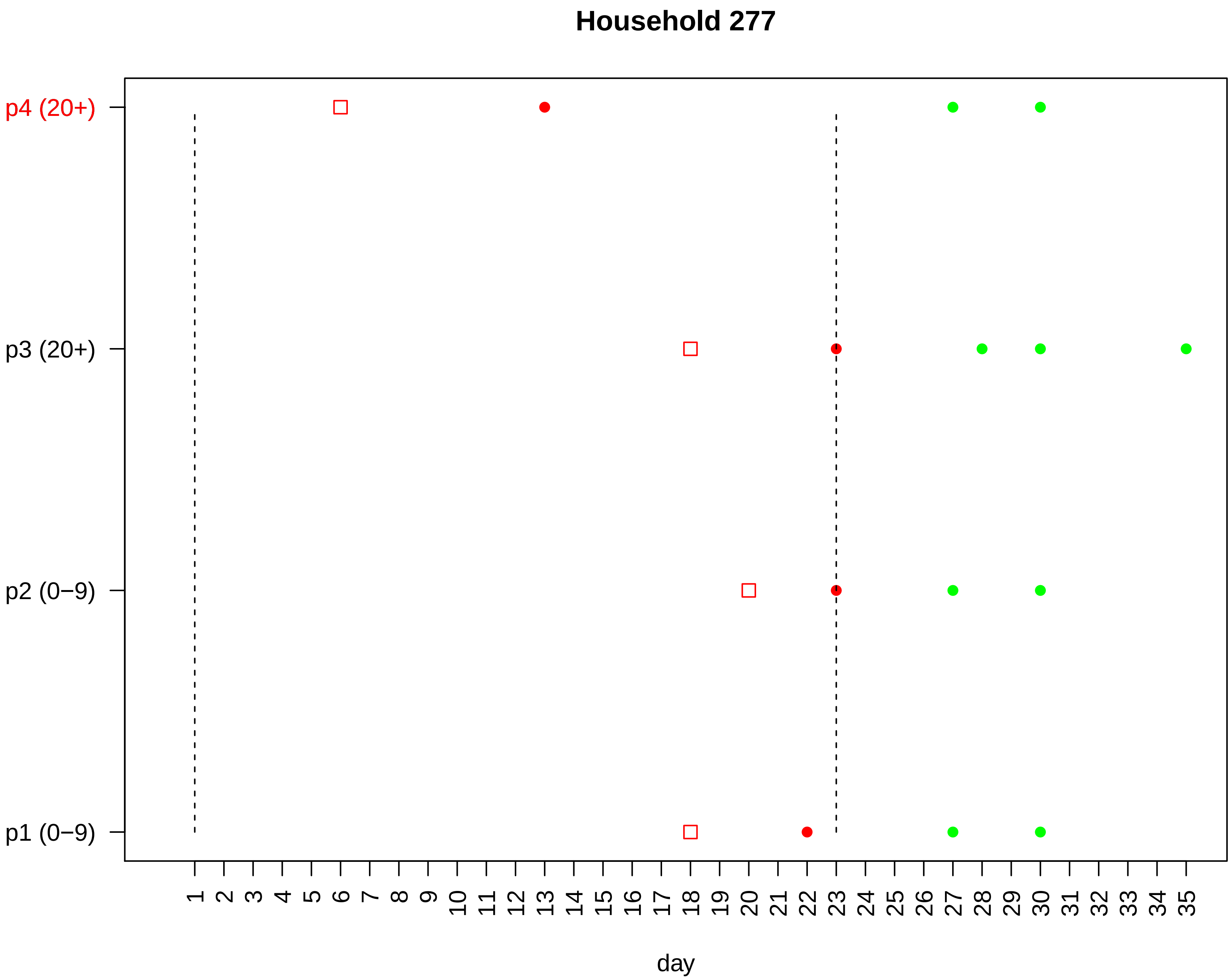

Household 277
p4 (20+)
p3 (20+)
p2 (0−9)
p1 (0−9)
1 2 3 4 5 6 7 8 9 10 11 12 13 14 15 16 17 18 19 20 21 22 23 24 25 26 27 28 29 30 31 32 33 34 35
day

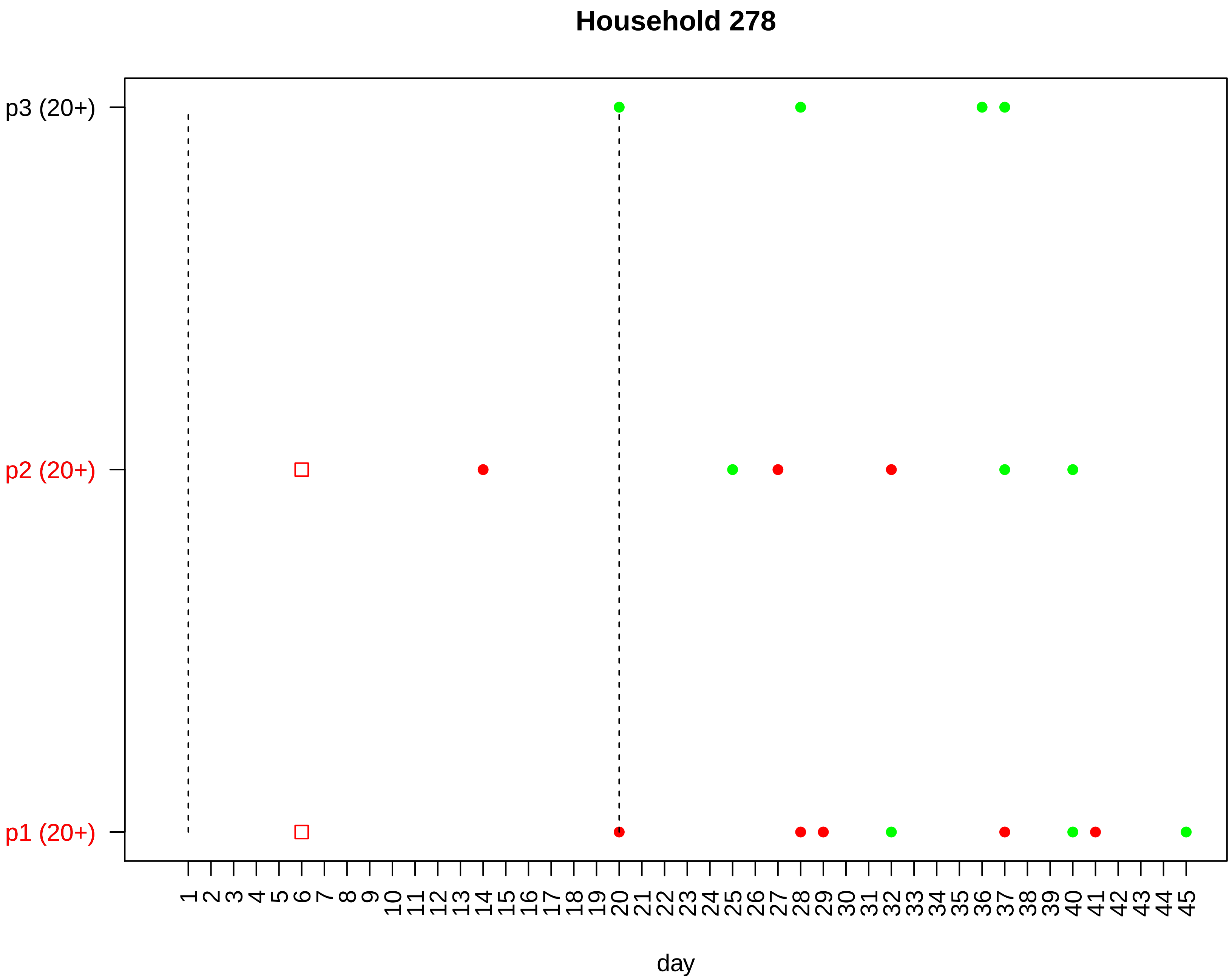

Household 278
p3 (20+)
p2 (20+)
p1 (20+)
1 2 3 4 5 6 7 8 9 10 11 12 13 14 15 16 17 18 19 20 21 22 23 24 25 26 27 28 29 30 31 32 33 34 35 36 37 38 39 40 41 42 43 44 45
day

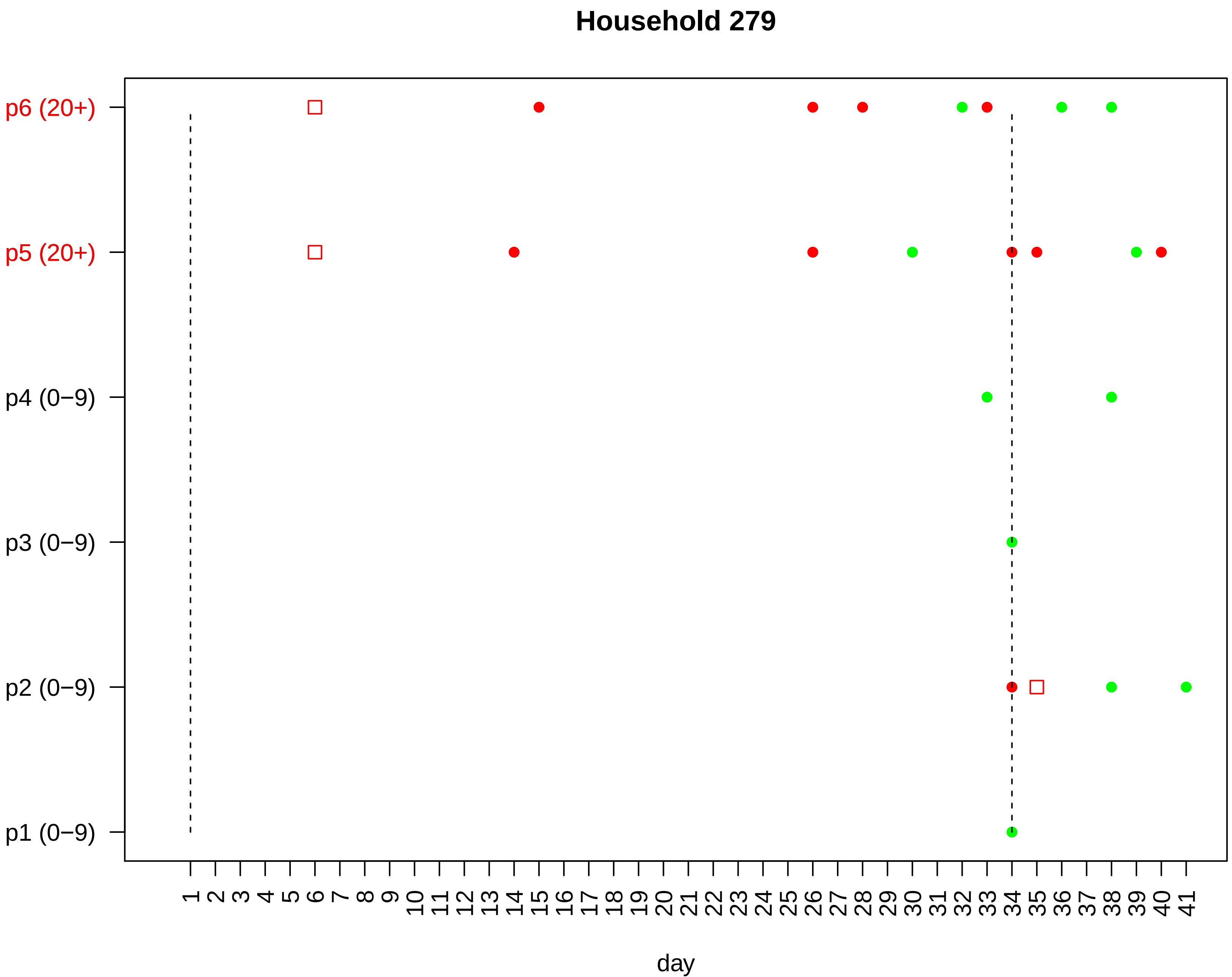
Household 279
p6 (20+)
p5 (20+)
p4 (0−9)
p3 (0−9)
p2 (0−9)
p1 (0−9)
1 2 3 4 5 6 7 8 9 10 11 12 13 14 15 16 17 18 19 20 21 22 23 24 25 26 27 28 29 30 31 32 33 34 35 36 37 38 39 40 41
day

### Household 280

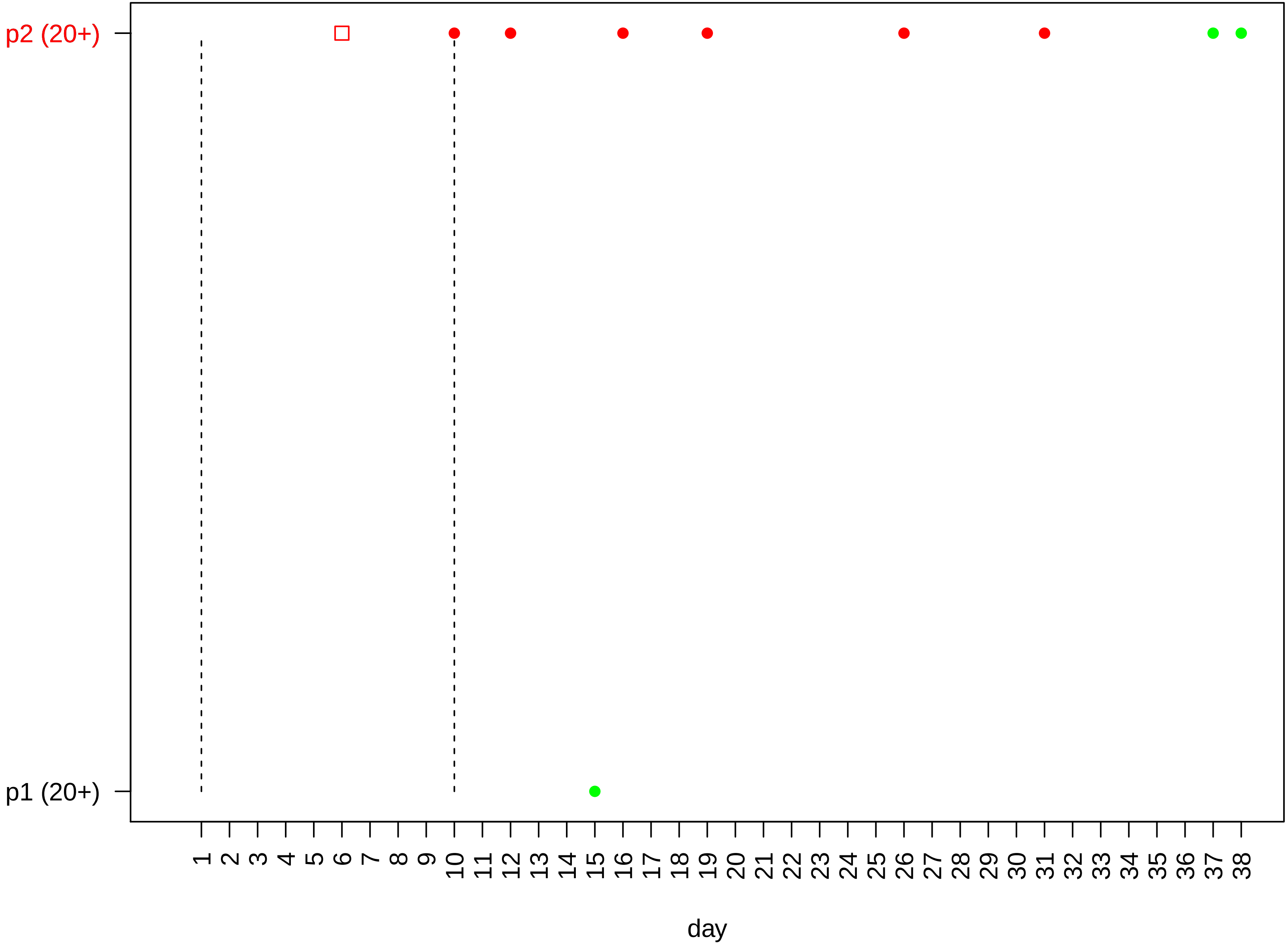

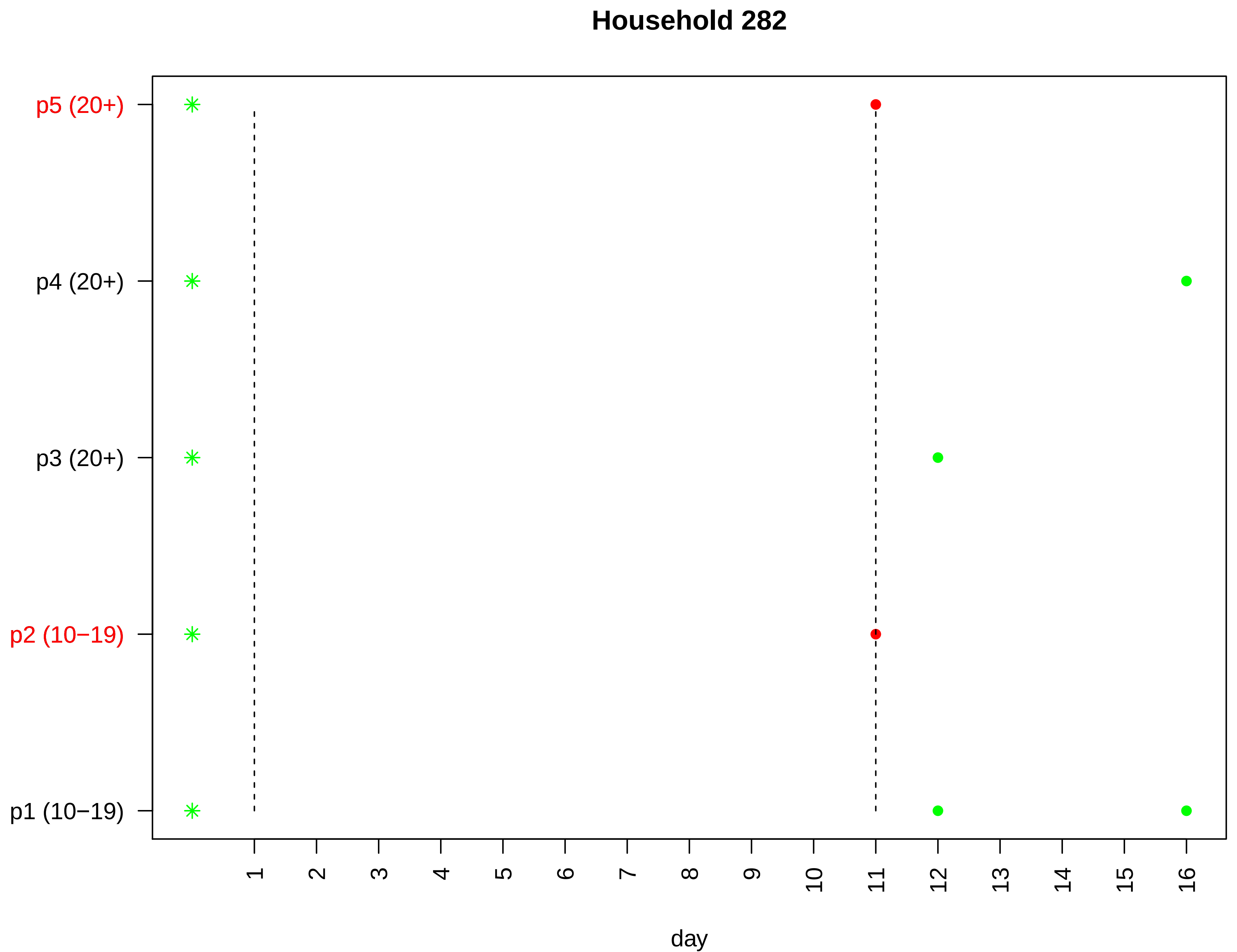
Household 282
p5 (20+)
p4 (20+)
p3 (20+)
p2 (10−19)
p1 (10−19)
day

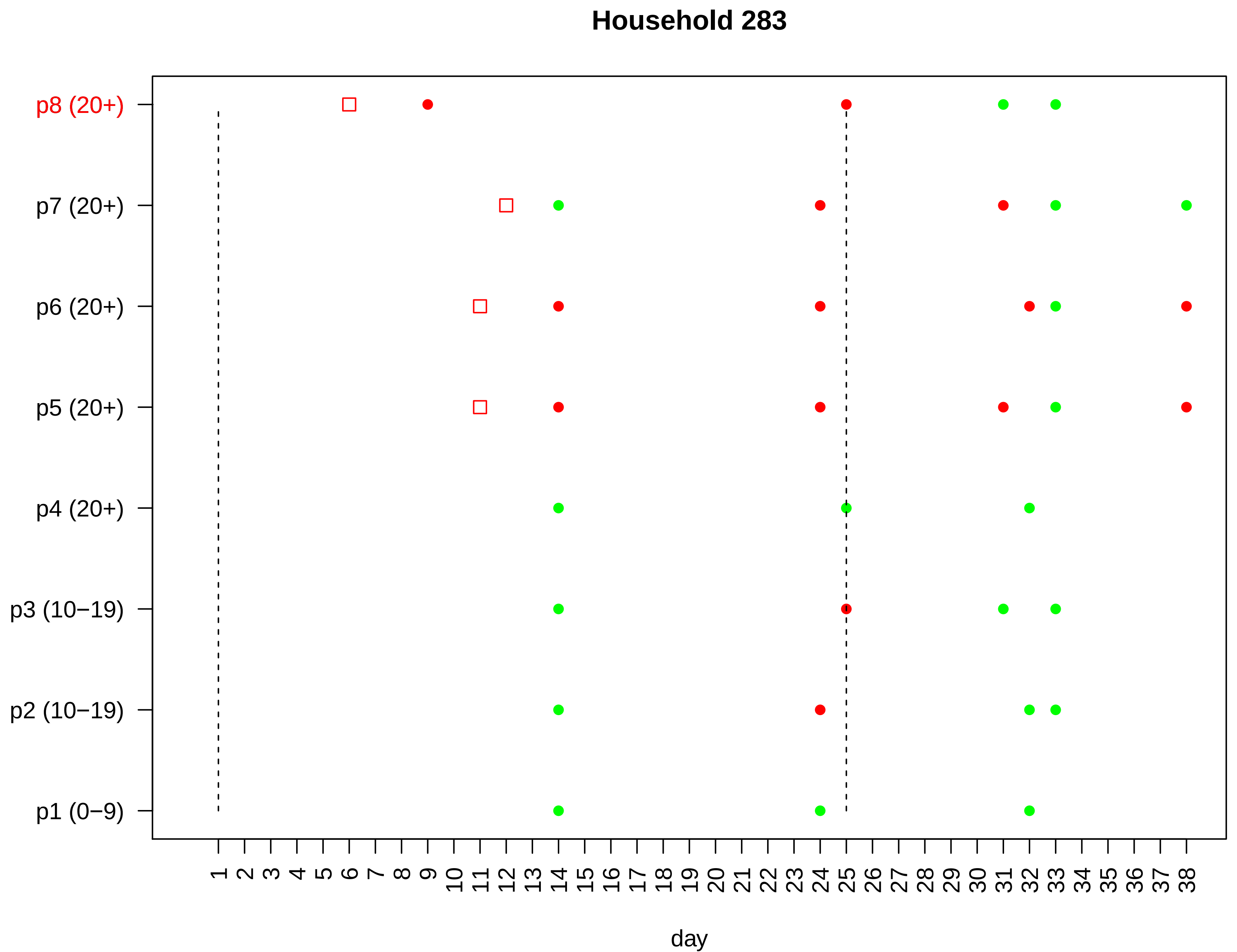

Household 283
p8 (20+)
p7 (20+)
p6 (20+)
p5 (20+)
p4 (20+)
p3 (10−19)
p2 (10−19)
p1 (0−9)
1 2 3 4 5 6 7 8 9 10 11 12 13 14 15 16 17 18 19 20 21 22 23 24 25 26 27 28 29 30 31 32 33 34 35 36 37 38
day

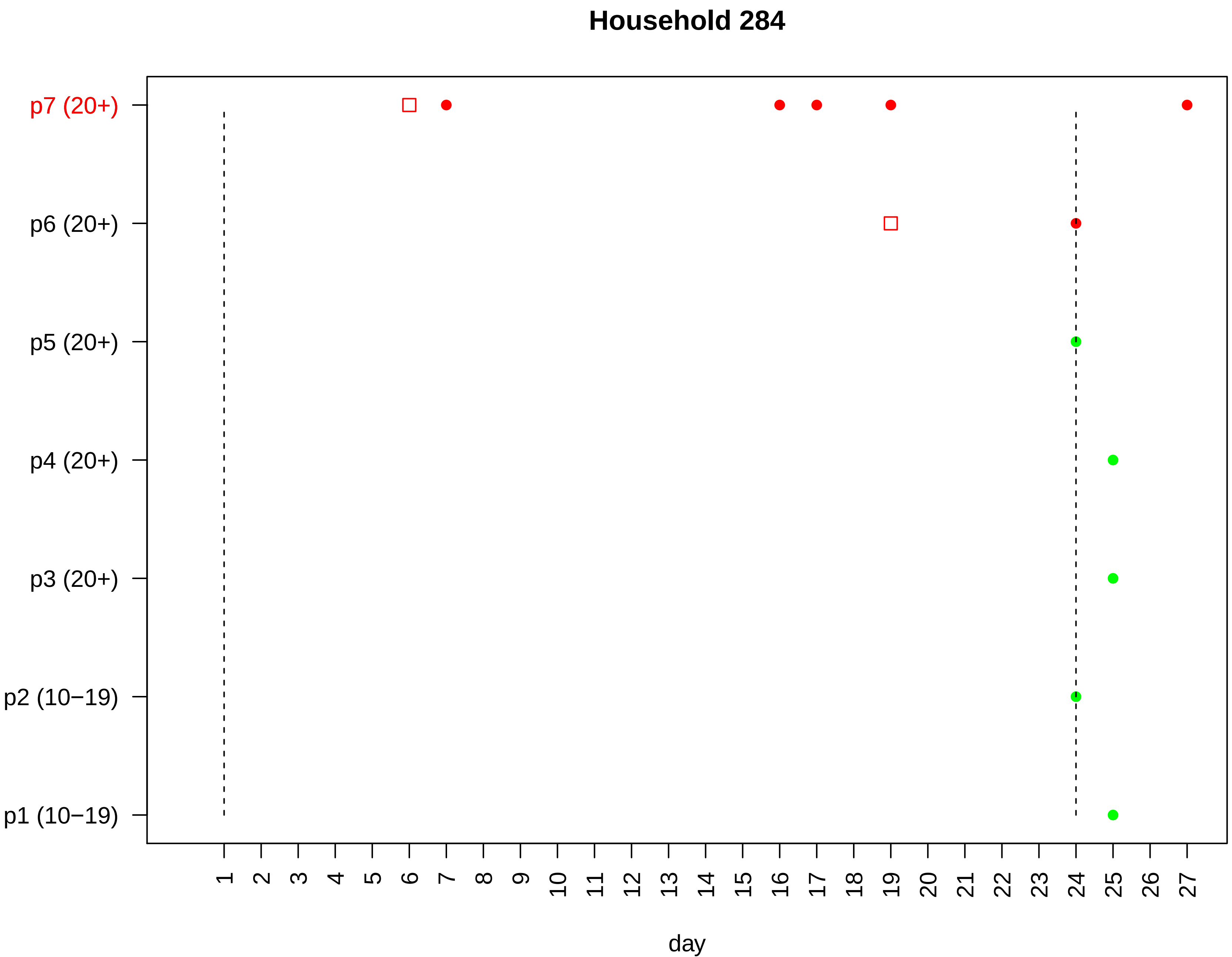

Household 284
p7 (20+)
p6 (20+)
p5 (20+)
p4 (20+)
p3 (20+)
p2 (10−19)
p1 (10−19)
1 2 3 4 5 6 7 8 9 10 11 12 13 14 15 16 17 18 19 20 21 22 23 24 25 26 27
day

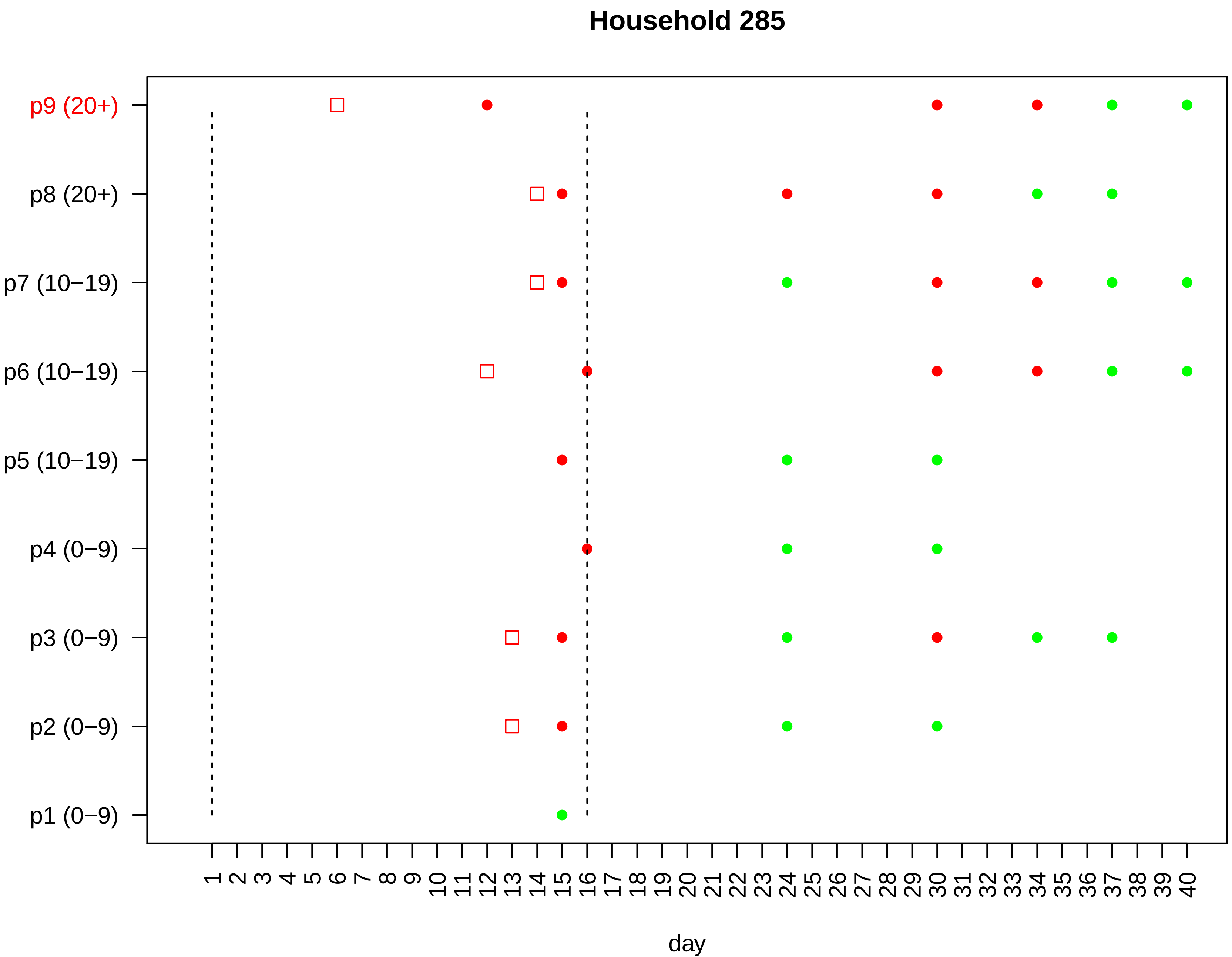

Household 285
p9 (20+)
p8 (20+)
p7 (10−19)
p6 (10−19)
p5 (10−19)
p4 (0−9)
p3 (0−9)
p2 (0−9)
p1 (0−9)
1 2 3 4 5 6 7 8 9 10 11 12 13 14 15 16 17 18 19 20 21 22 23 24 25 26 27 28 29 30 31 32 33 34 35 36 37 38 39 40
day

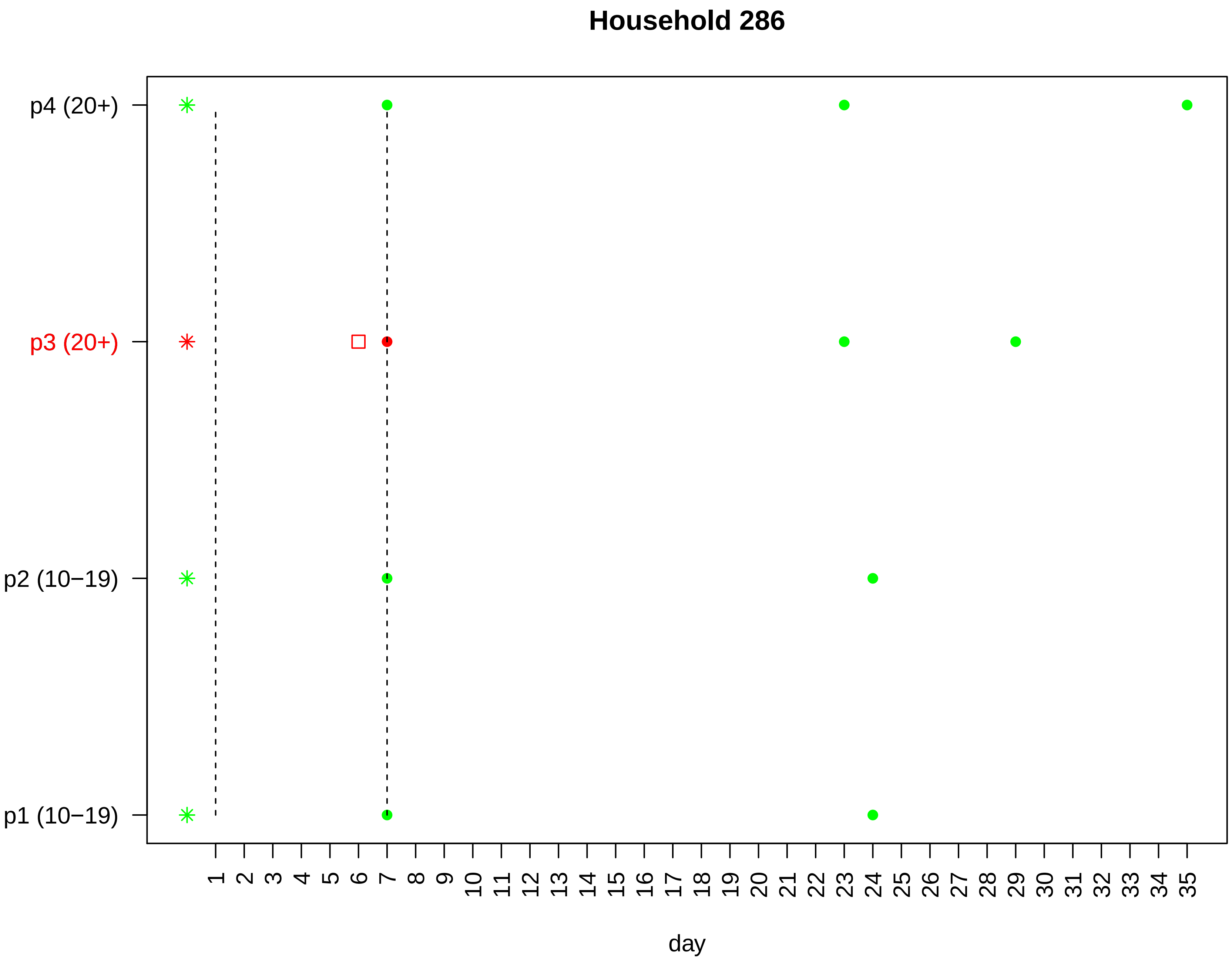

Household 286
p4 (20+)
p3 (20+)
p2 (10−19)
p1 (10−19)
day

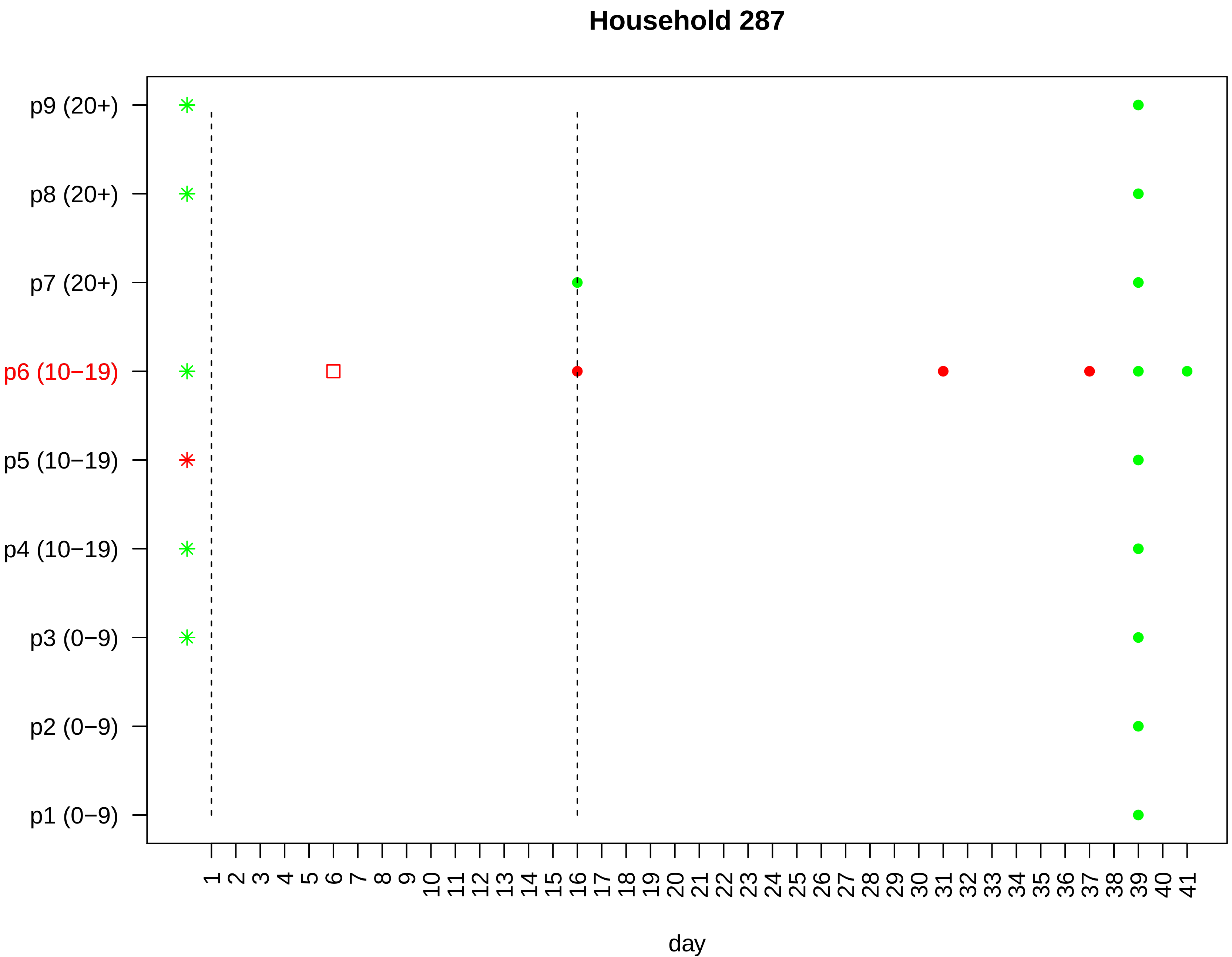

Household 287
p9 (20+)
p8 (20+)
p7 (20+)
p6 (10−19)
p5 (10−19)
p4 (10−19)
p3 (0−9)
p2 (0−9)
p1 (0−9)
1 2 3 4 5 6 7 8 9 10 11 12 13 14 15 16 17 18 19 20 21 22 23 24 25 26 27 28 29 30 31 32 33 34 35 36 37 38 39 40 41
day

### Household 288

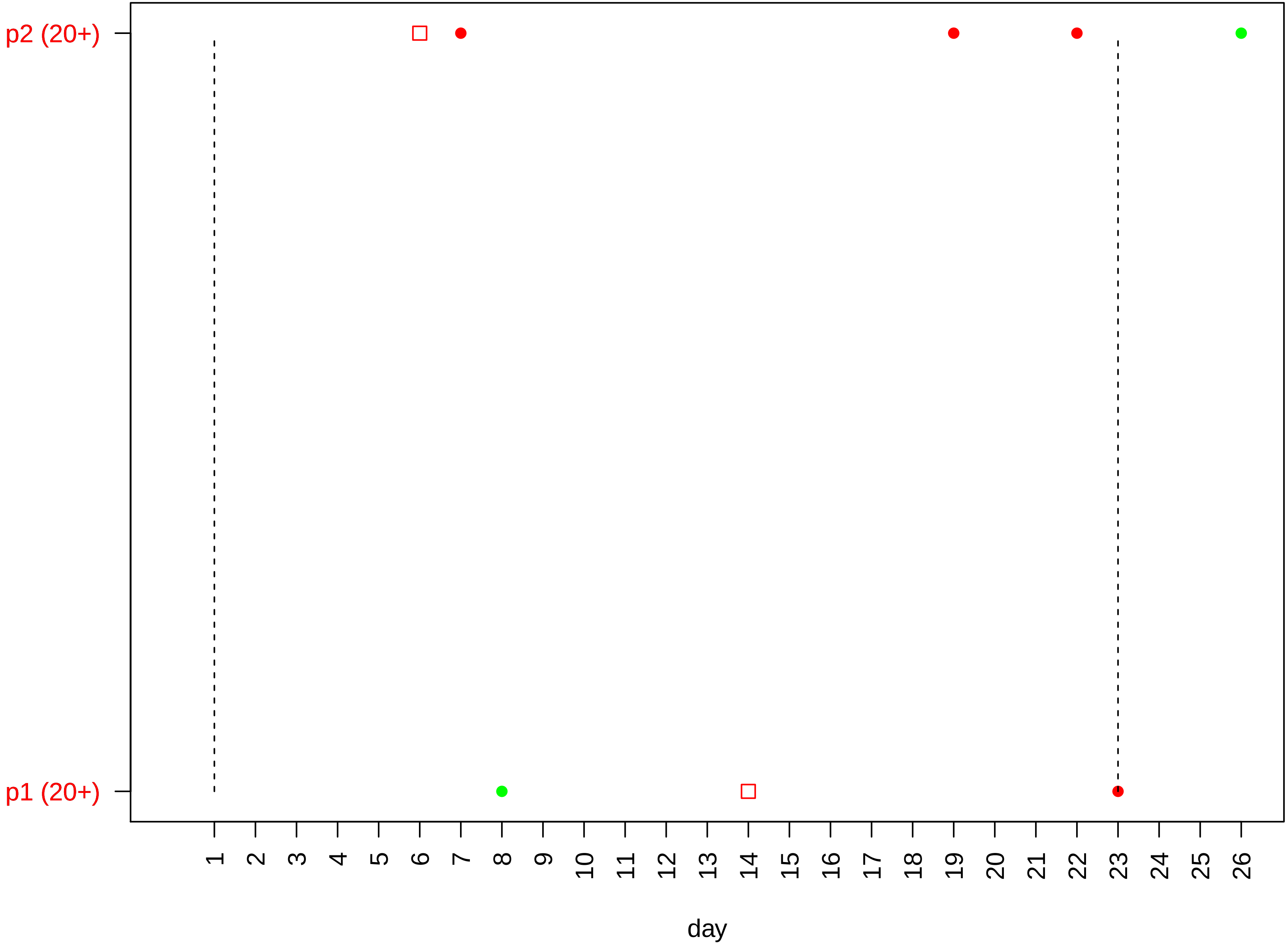

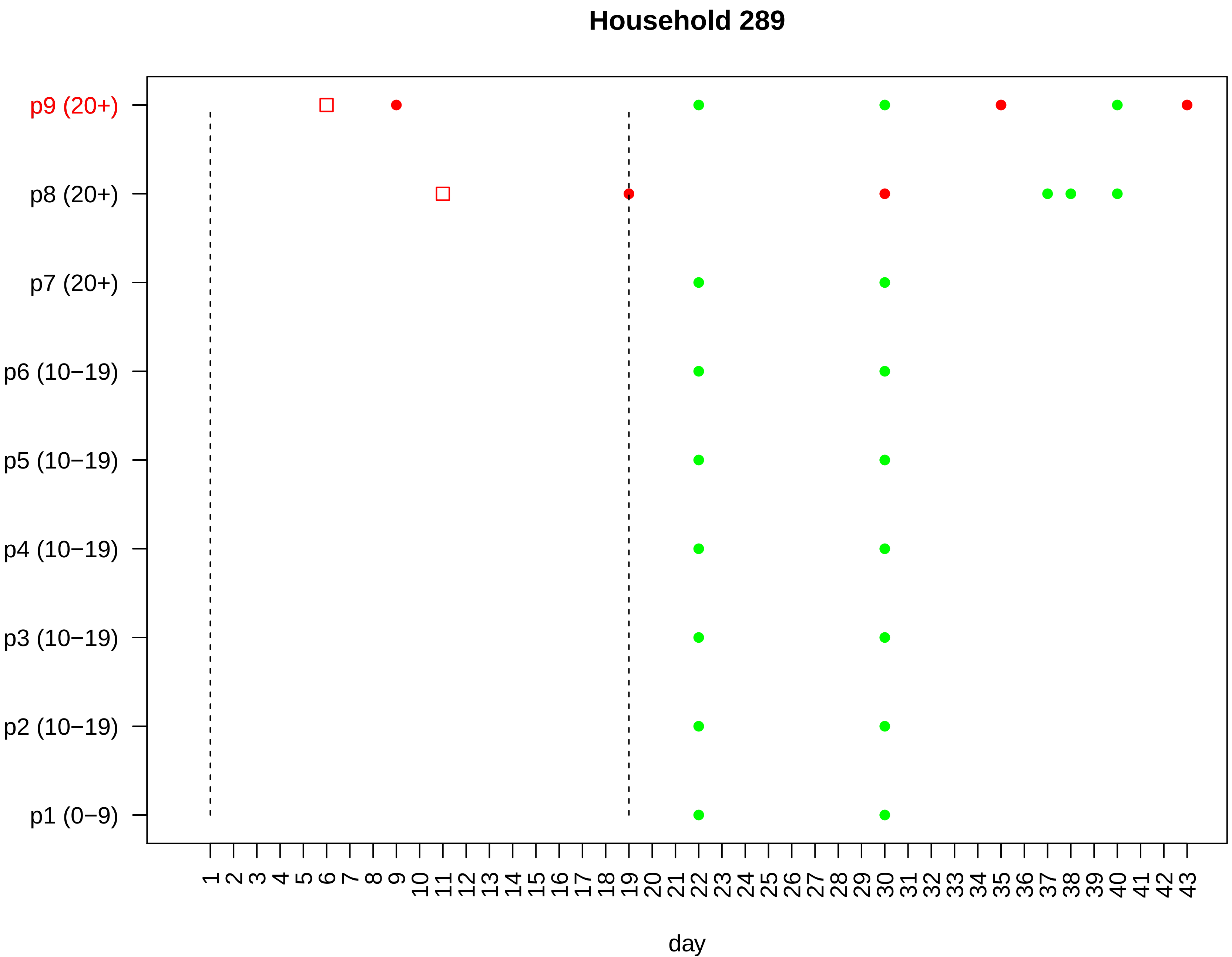

Household 289
p9 (20+)
p8 (20+)
p7 (20+)
p6 (10−19)
p5 (10−19)
p4 (10−19)
p3 (10−19)
p2 (10−19)
p1 (0−9)
1 2 3 4 5 6 7 8 9 10 11 12 13 14 15 16 17 18 19 20 21 22 23 24 25 26 27 28 29 30 31 32 33 34 35 36 37 38 39 40 41 42 43
day

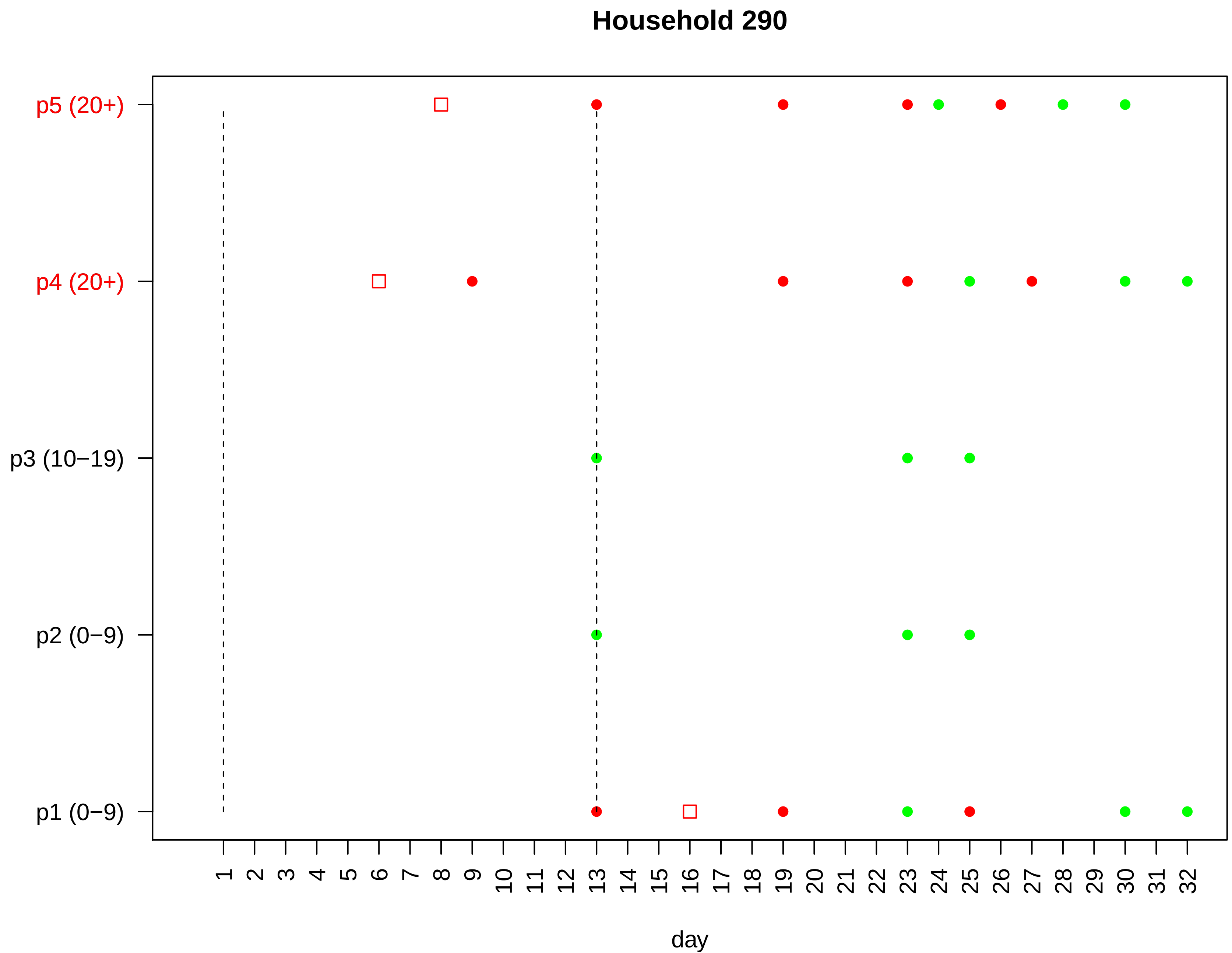

Household 290
p5 (20+)
p4 (20+)
p3 (10−19)
p2 (0−9)
p1 (0−9)
day

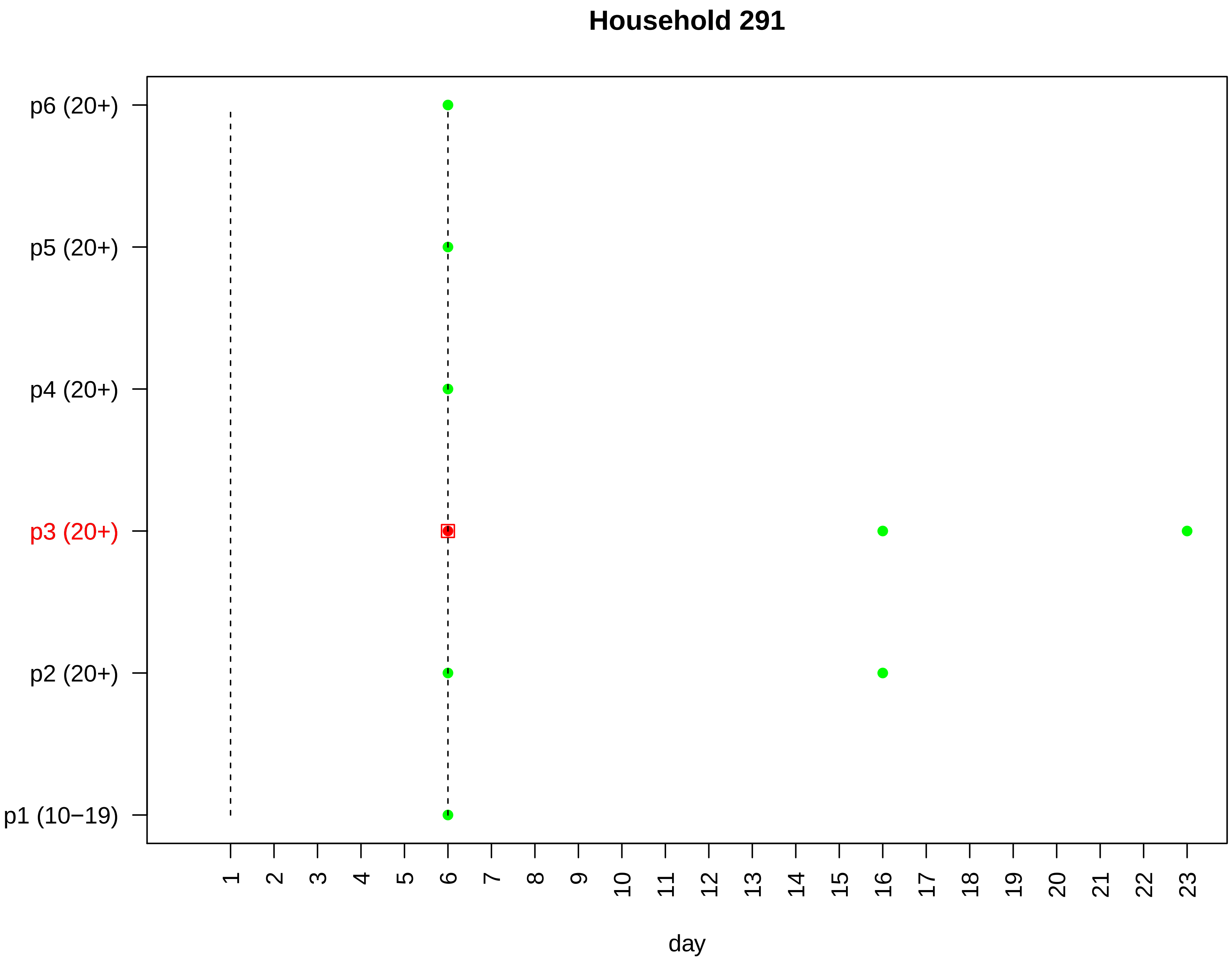
Household 291
p6 (20+)
p5 (20+)
p4 (20+)
p3 (20+)
p2 (20+)
p1 (10−19)
1 2 3 4 5 6 7 8 9 10 11 12 13 14 15 16 17 18 19 20 21 22 23
day

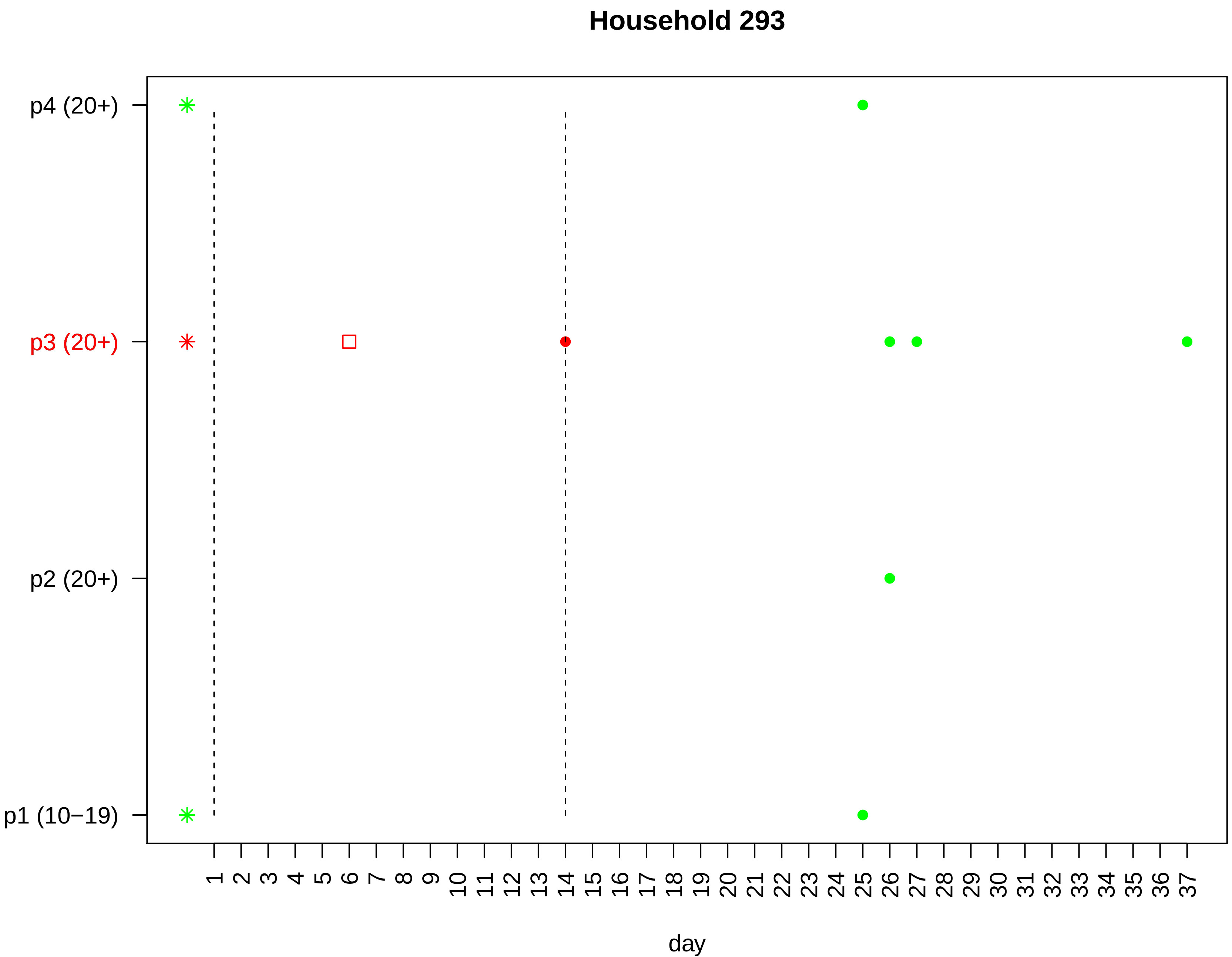
Household 293
p4 (20+)
p3 (20+)
p2 (20+)
p1 (10−19)
1 2 3 4 5 6 7 8 9 10 11 12 13 14 15 16 17 18 19 20 21 22 23 24 25 26 27 28 29 30 31 32 33 34 35 36 37
day

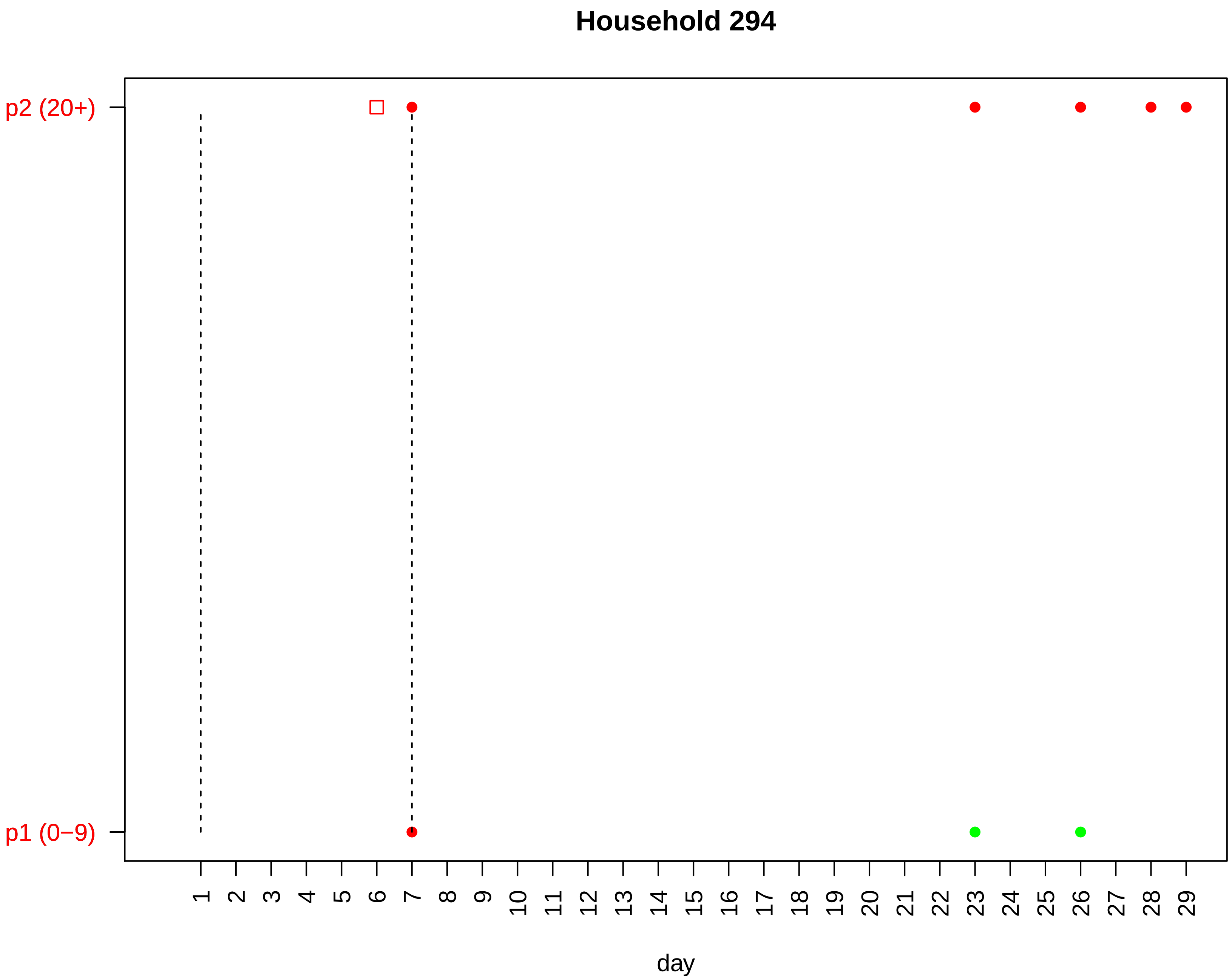

Household 294
p2 (20+)
p1 (0−9)
1 2 3 4 5 6 7 8 9 10 11 12 13 14 15 16 17 18 19 20 21 22 23 24 25 26 27 28 29
day

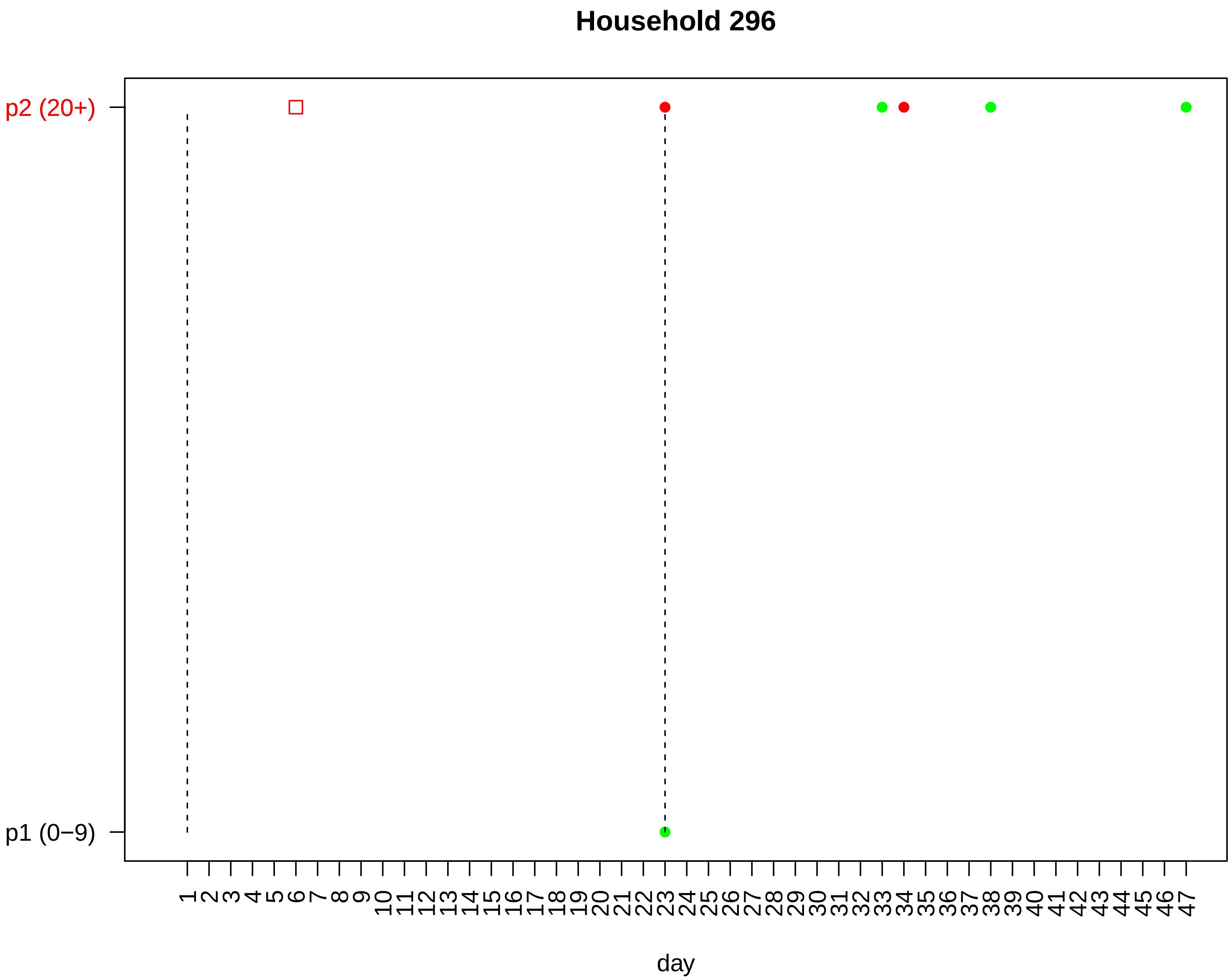

Household 296
p2 (20+)
p1 (0−9)
day

#### Household 297

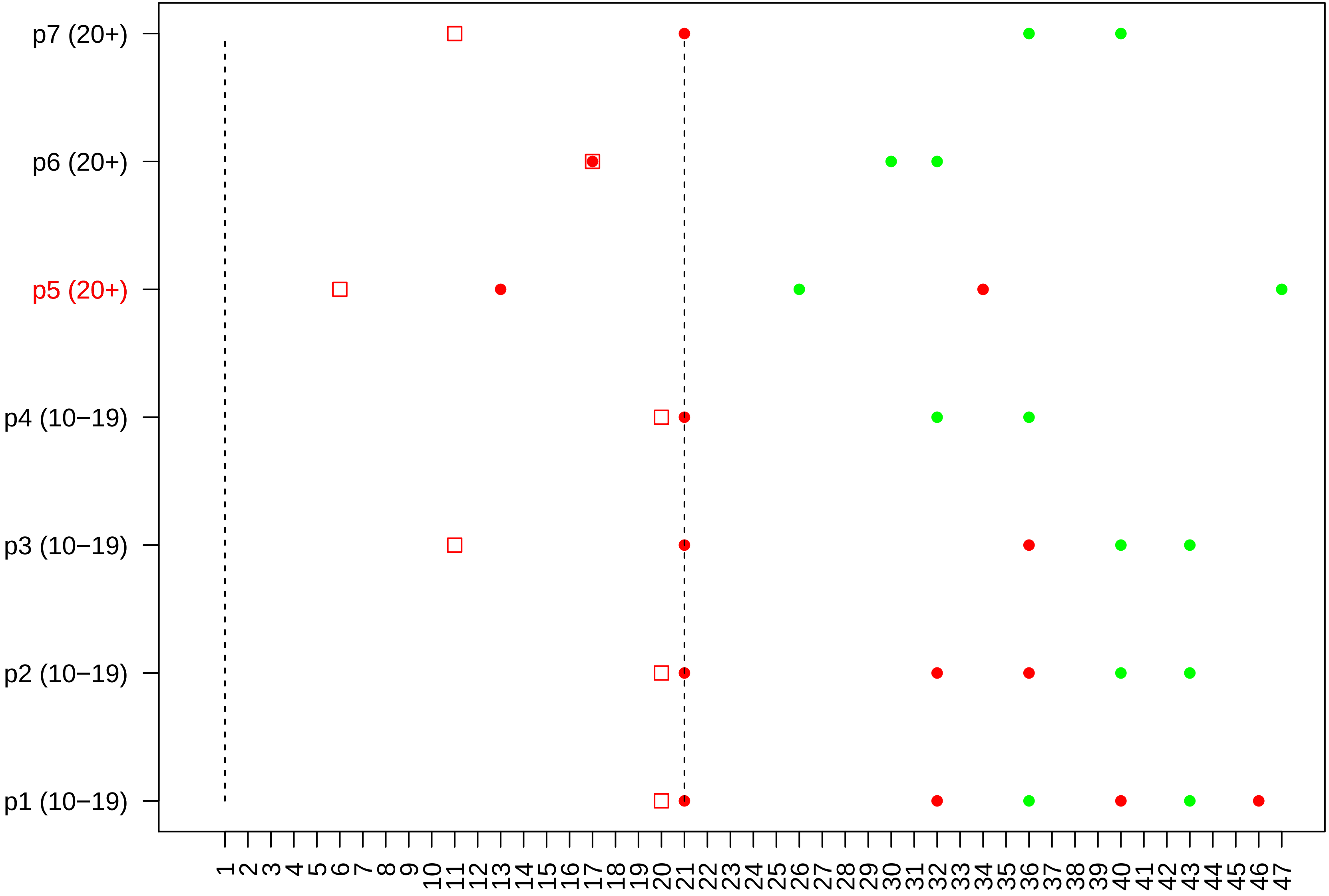

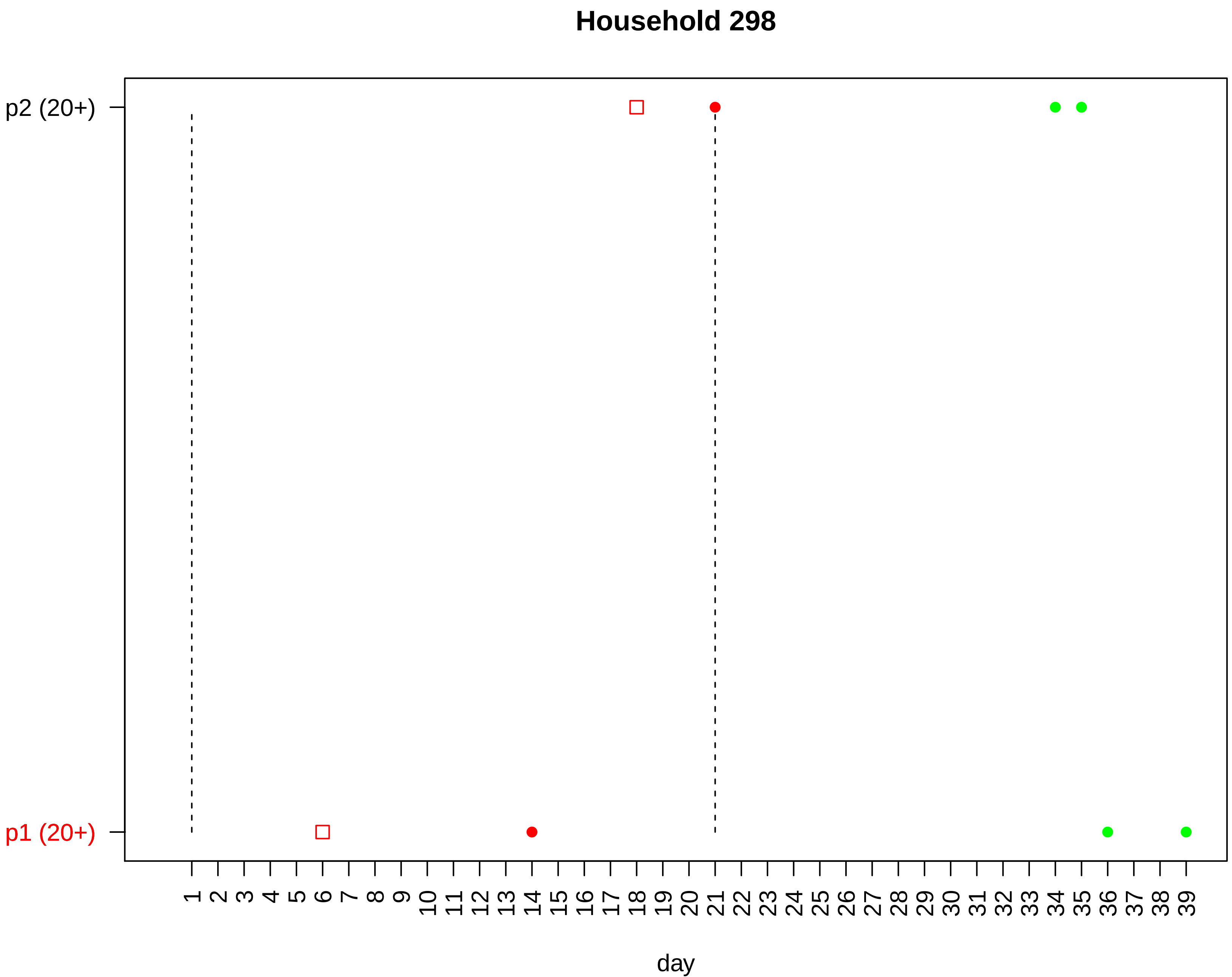

Household 298
p2 (20+)
p1 (20+)
1 2 3 4 5 6 7 8 9 10 11 12 13 14 15 16 17 18 19 20 21 22 23 24 25 26 27 28 29 30 31 32 33 34 35 36 37 38 39
day

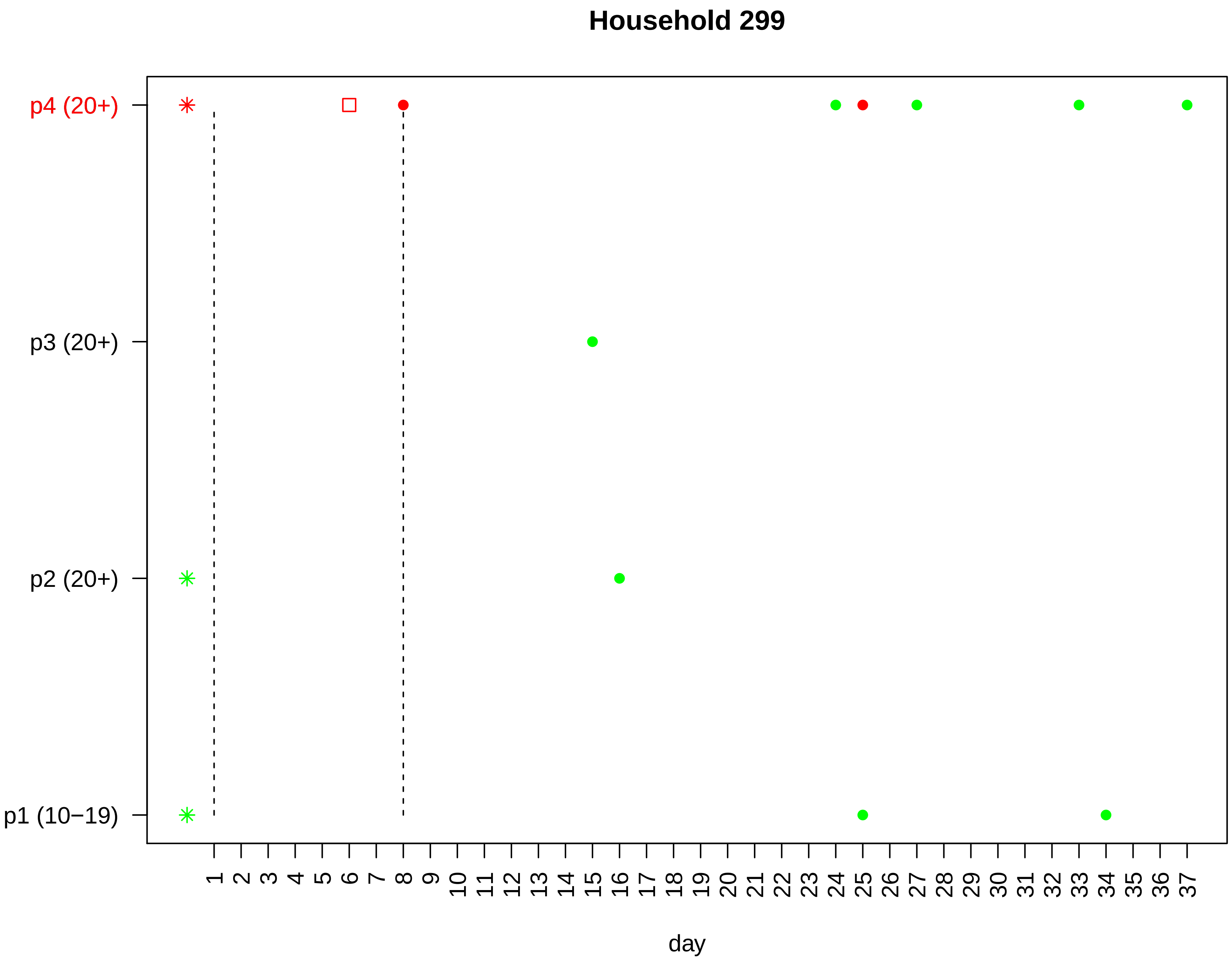

Household 299
p4 (20+)
p3 (20+)
p2 (20+)
p1 (10−19)
1 2 3 4 5 6 7 8 9 10 11 12 13 14 15 16 17 18 19 20 21 22 23 24 25 26 27 28 29 30 31 32 33 34 35 36 37
day

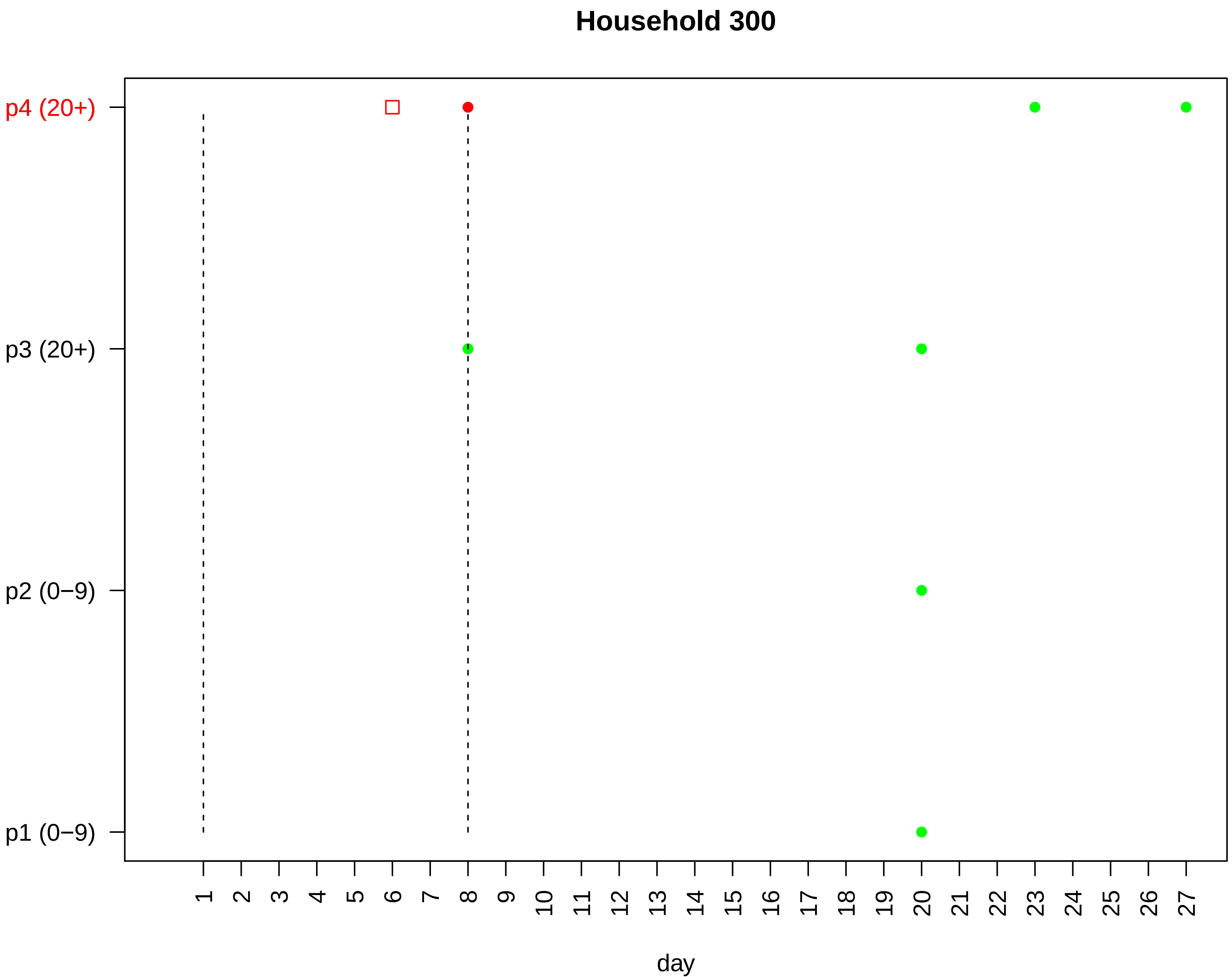

Household 300
p4 (20+)
p3 (20+)
p2 (0−9)
p1 (0−9)
1 2 3 4 5 6 7 8 9 10 11 12 13 14 15 16 17 18 19 20 21 22 23 24 25 26 27
day

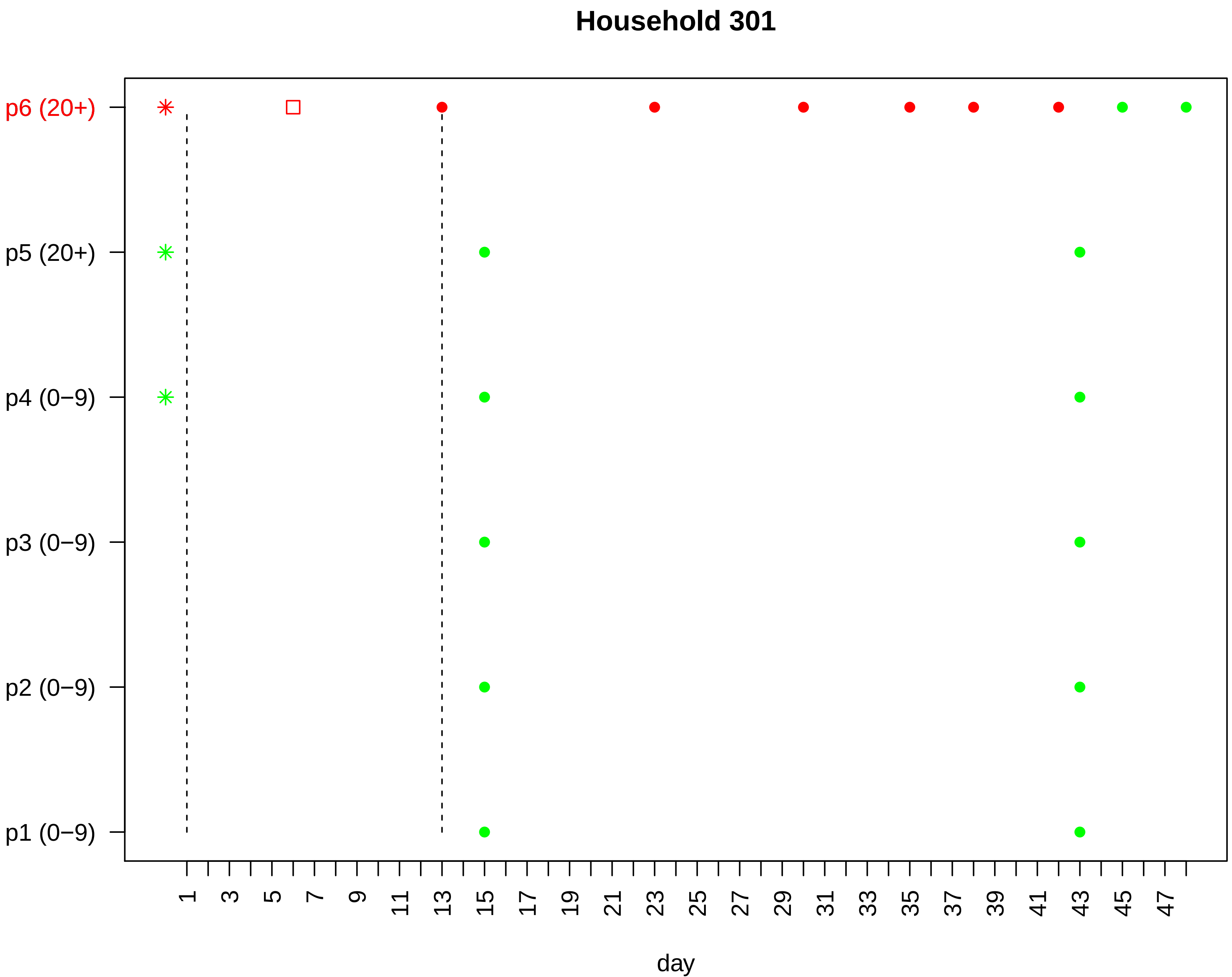

Household 301
p6 (20+)
p5 (20+)
p4 (0−9)
p3 (0−9)
p2 (0−9)
p1 (0−9)
1
3
5
7
9
11
13
15
17
19
21
23
25
27
29
31
33
35
37
39
41
43
45
47
day

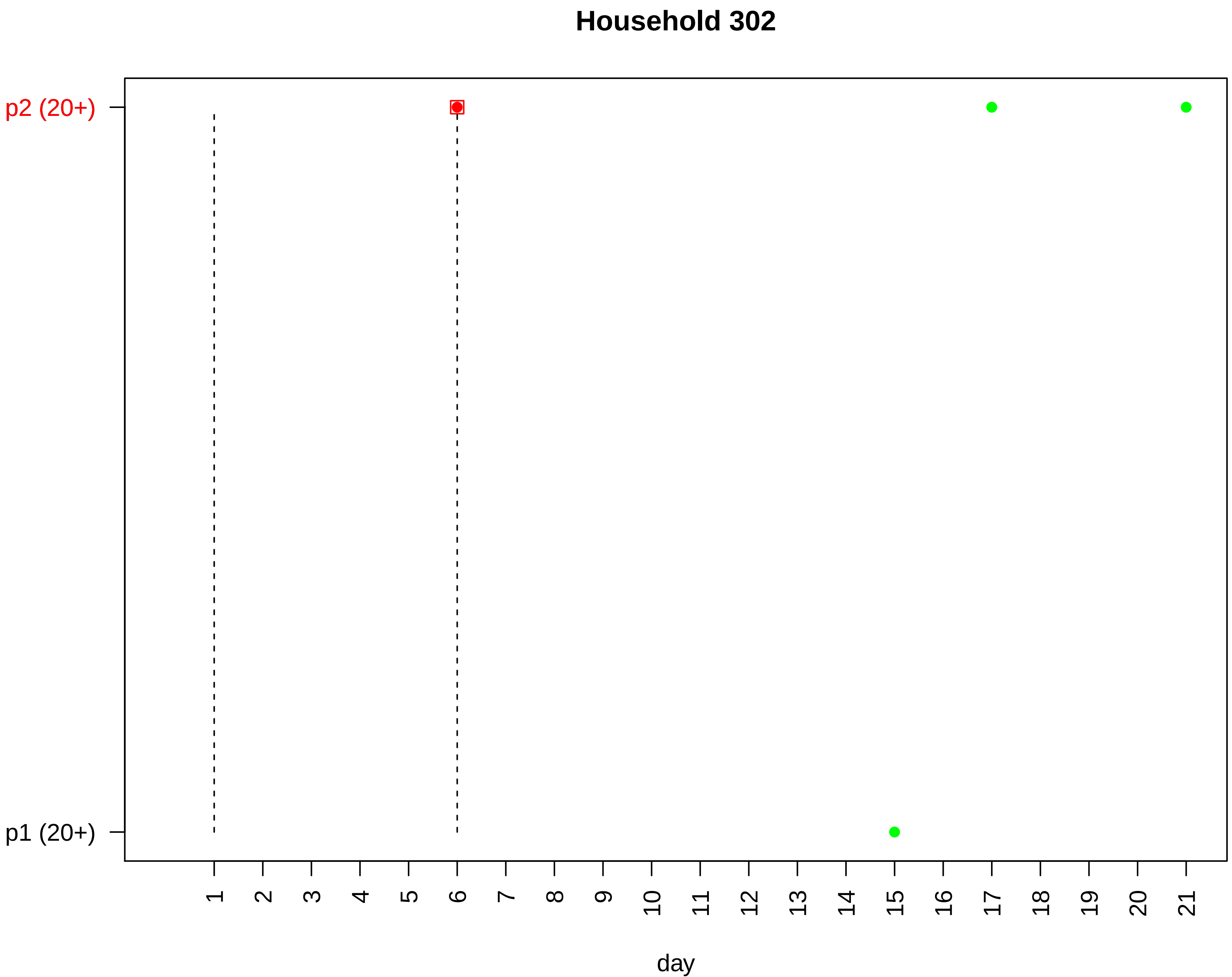

Household 302
p2 (20+)
p1 (20+)
day

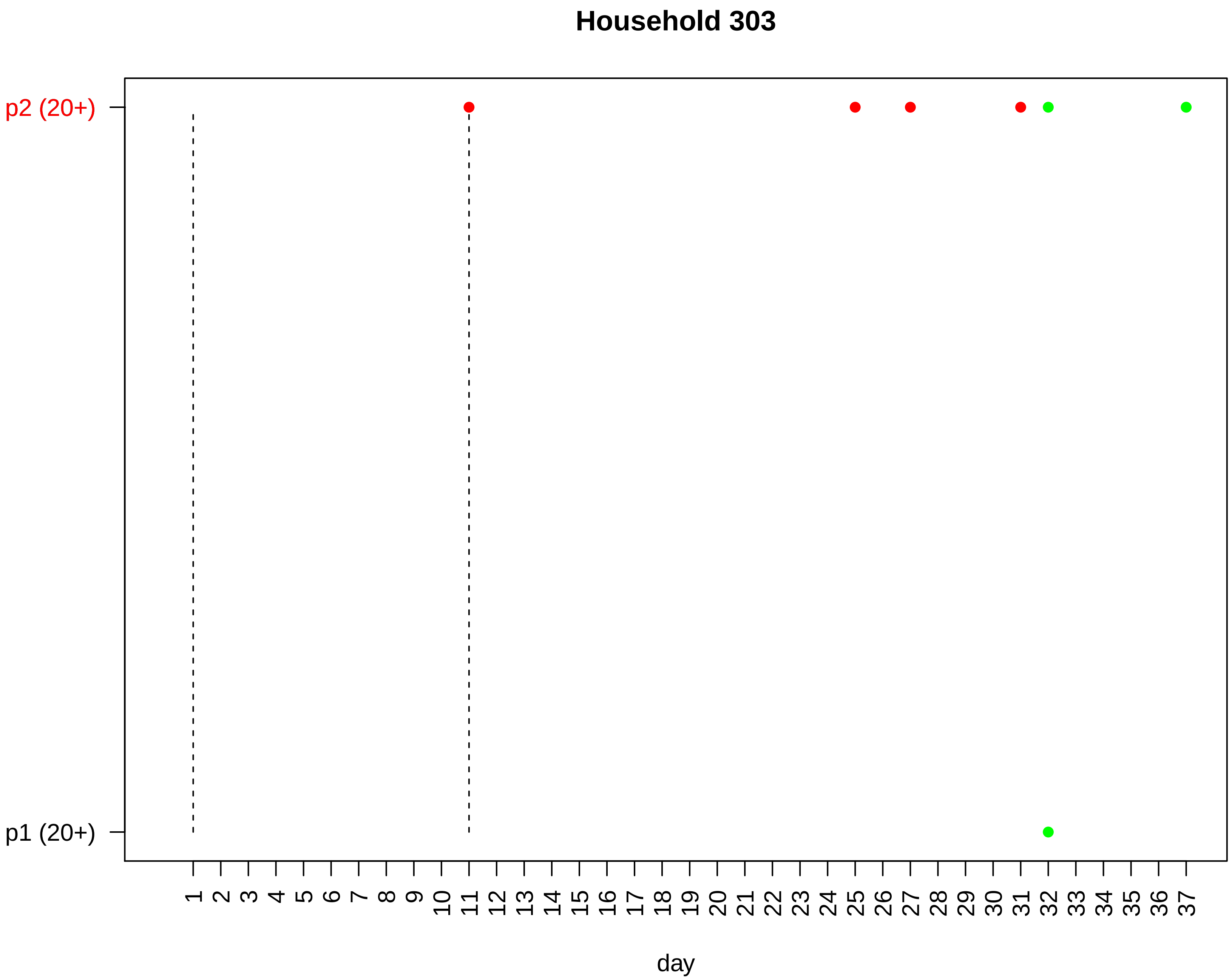
Household 303
p2 (20+)
p1 (20+)
day

#### Household 304

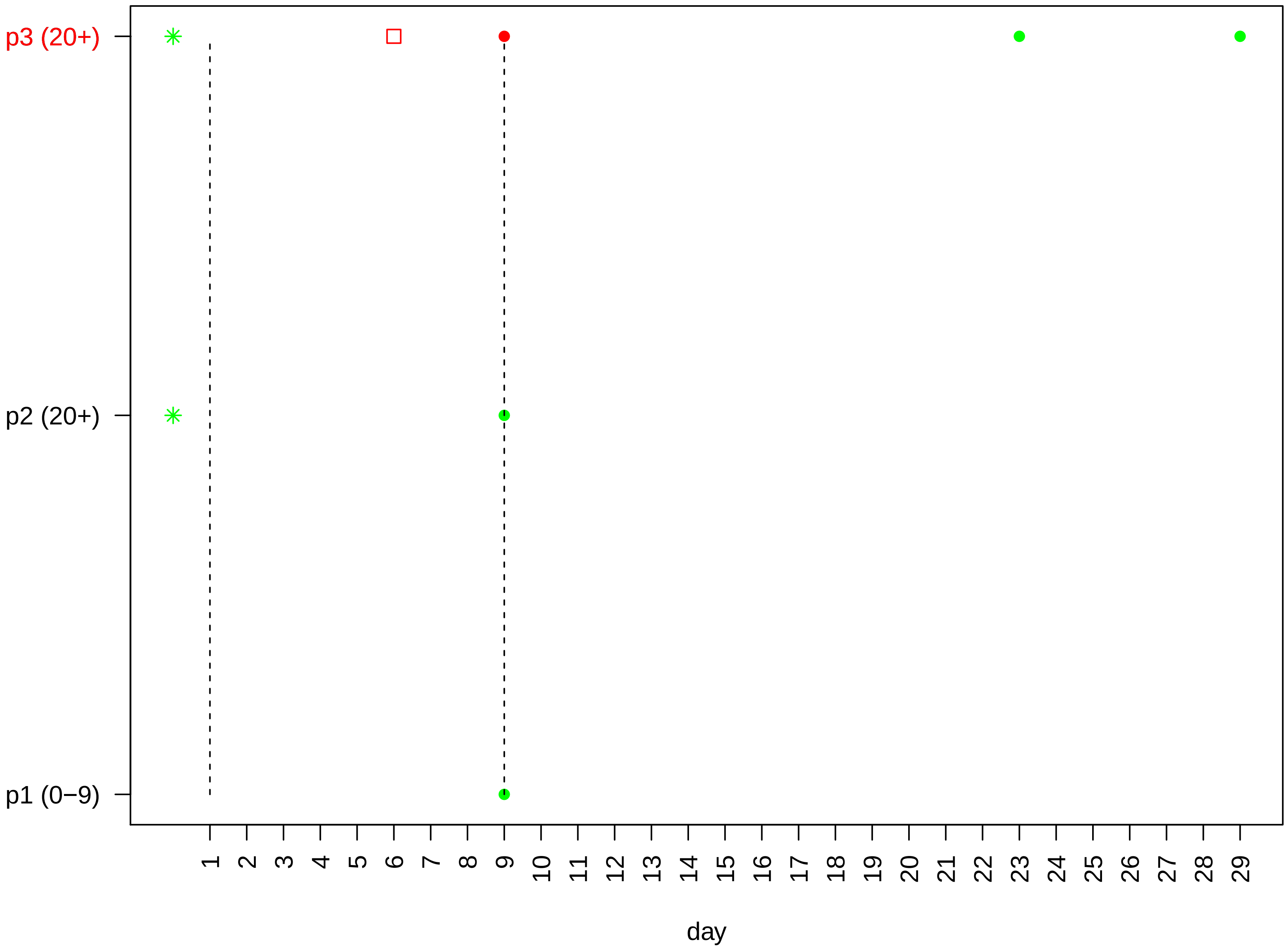

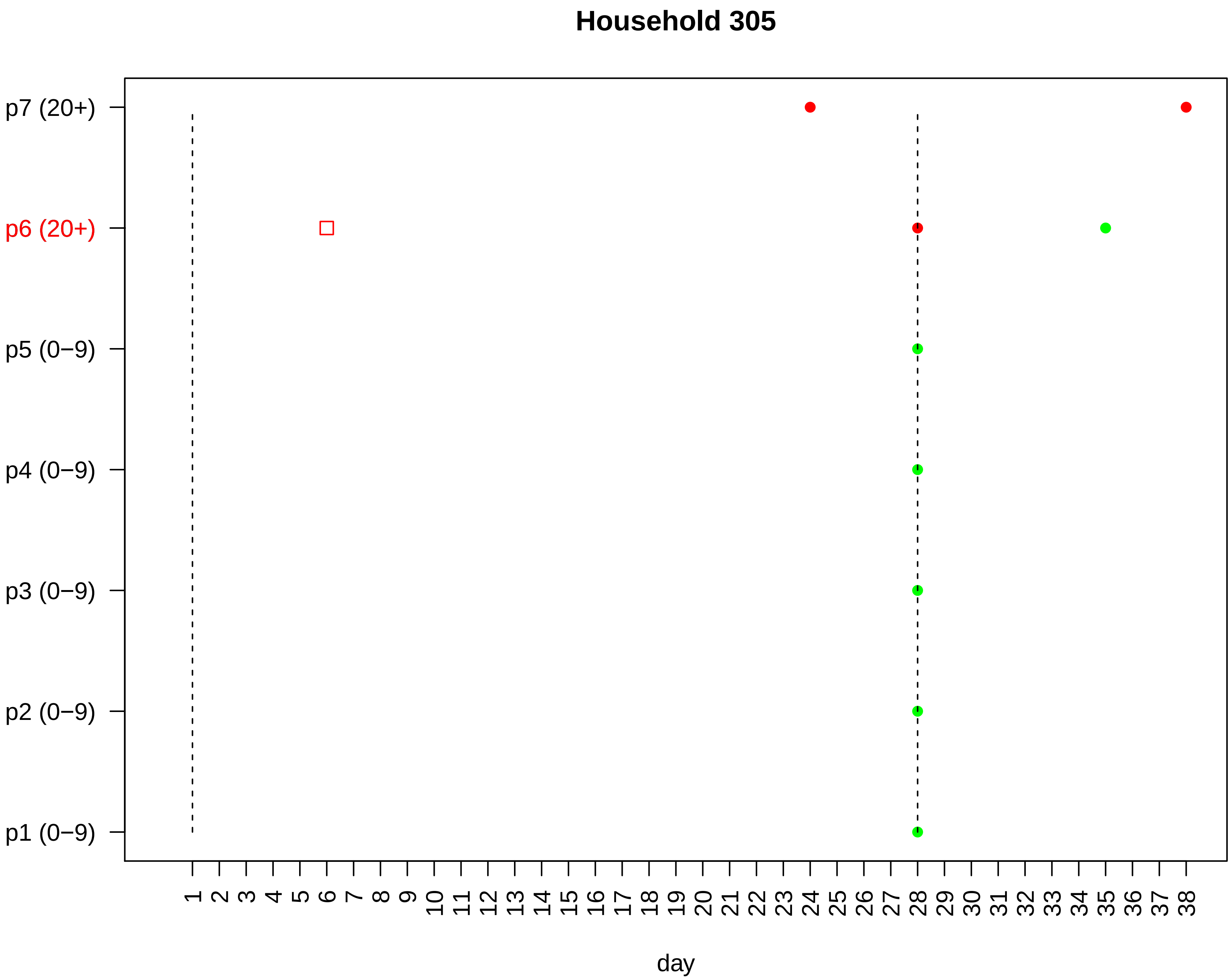

Household 305
p7 (20+)
p6 (20+)
p5 (0−9)
p4 (0−9)
p3 (0−9)
p2 (0−9)
p1 (0−9)
1 2 3 4 5 6 7 8 9 10 11 12 13 14 15 16 17 18 19 20 21 22 23 24 25 26 27 28 29 30 31 32 33 34 35 36 37 38
day

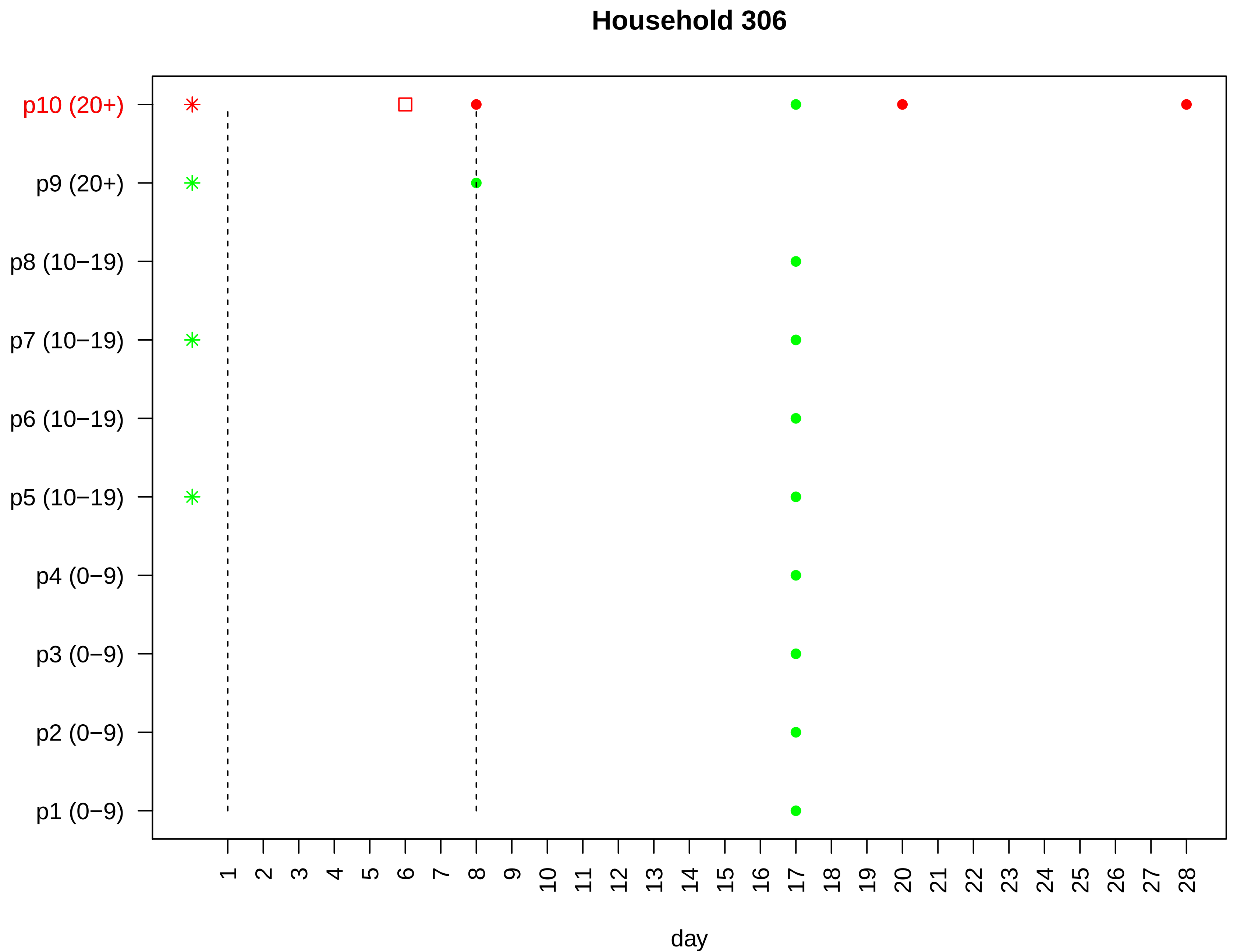

Household 306
p10 (20+)
p9 (20+)
p8 (10−19)
p7 (10−19)
p6 (10−19)
p5 (10−19)
p4 (0−9)
p3 (0−9)
p2 (0−9)
p1 (0−9)
1 2 3 4 5 6 7 8 9 10 11 12 13 14 15 16 17 18 19 20 21 22 23 24 25 26 27 28
day

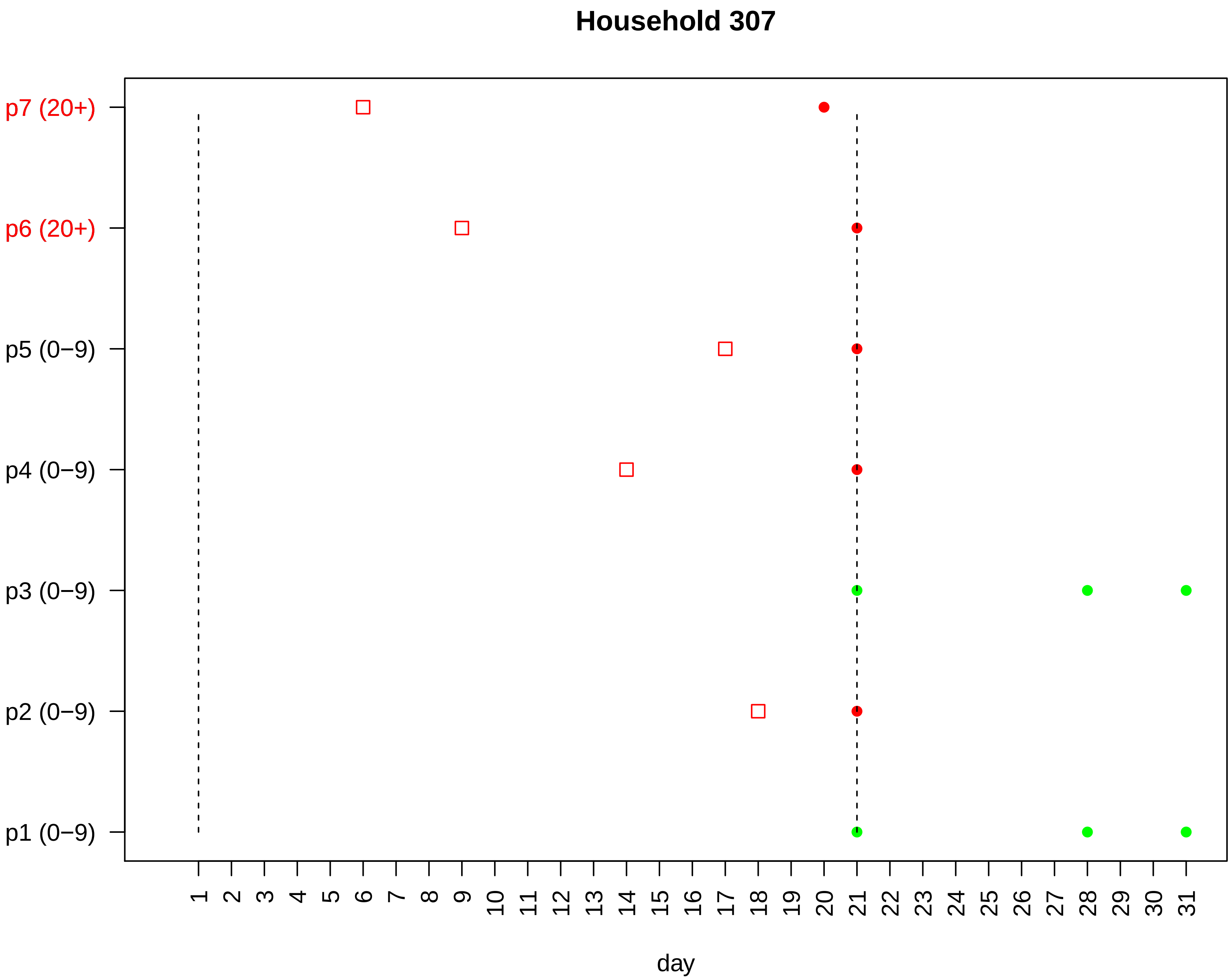

Household 307
p7 (20+)
p6 (20+)
p5 (0−9)
p4 (0−9)
p3 (0−9)
p2 (0−9)
p1 (0−9)
1 2 3 4 5 6 7 8 9 10 11 12 13 14 15 16 17 18 19 20 21 22 23 24 25 26 27 28 29 30 31
day

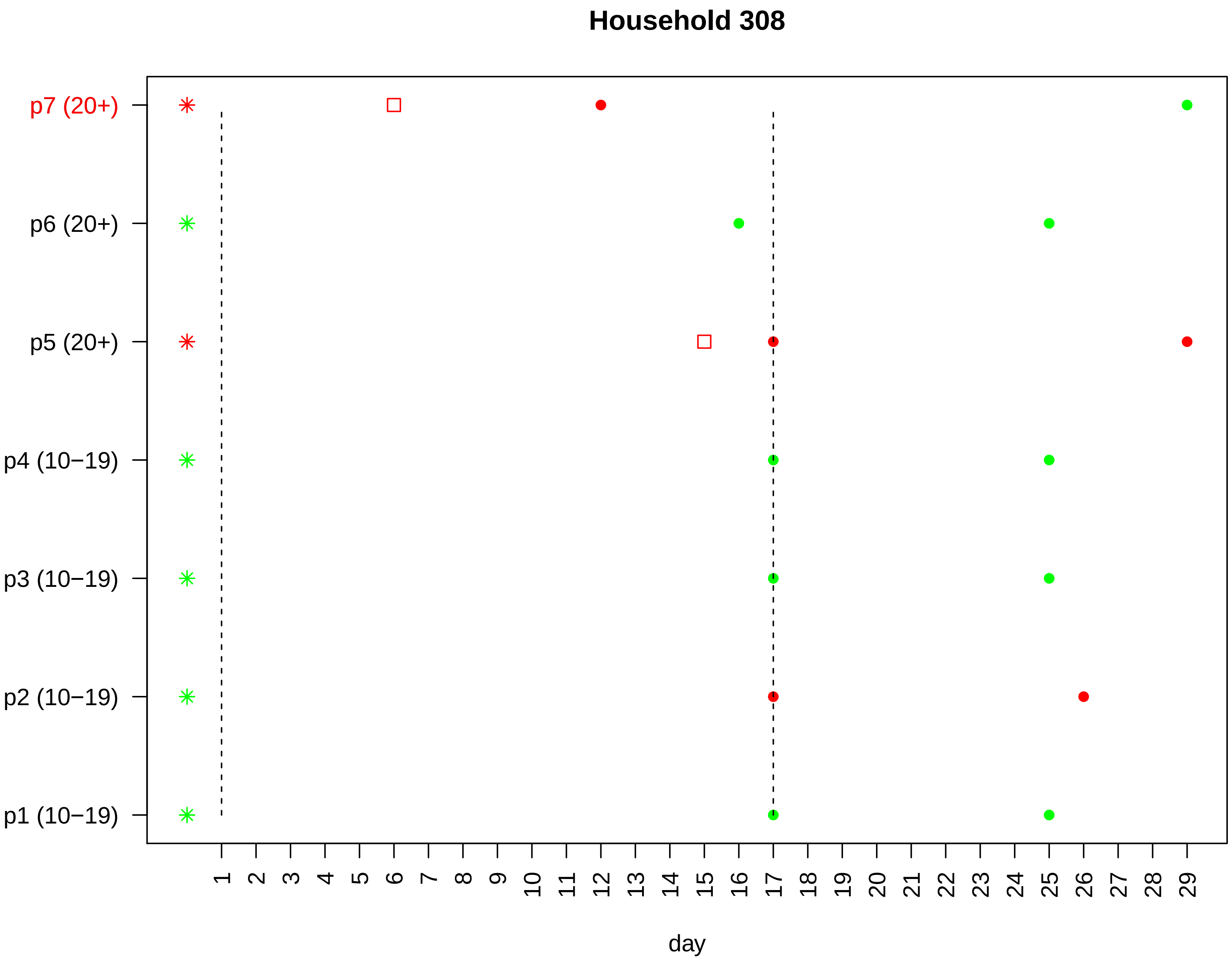
Household 308
p7 (20+)
p6 (20+)
p5 (20+)
p4 (10−19)
p3 (10−19)
p2 (10−19)
p1 (10−19)
1 2 3 4 5 6 7 8 9 10 11 12 13 14 15 16 17 18 19 20 21 22 23 24 25 26 27 28 29
day

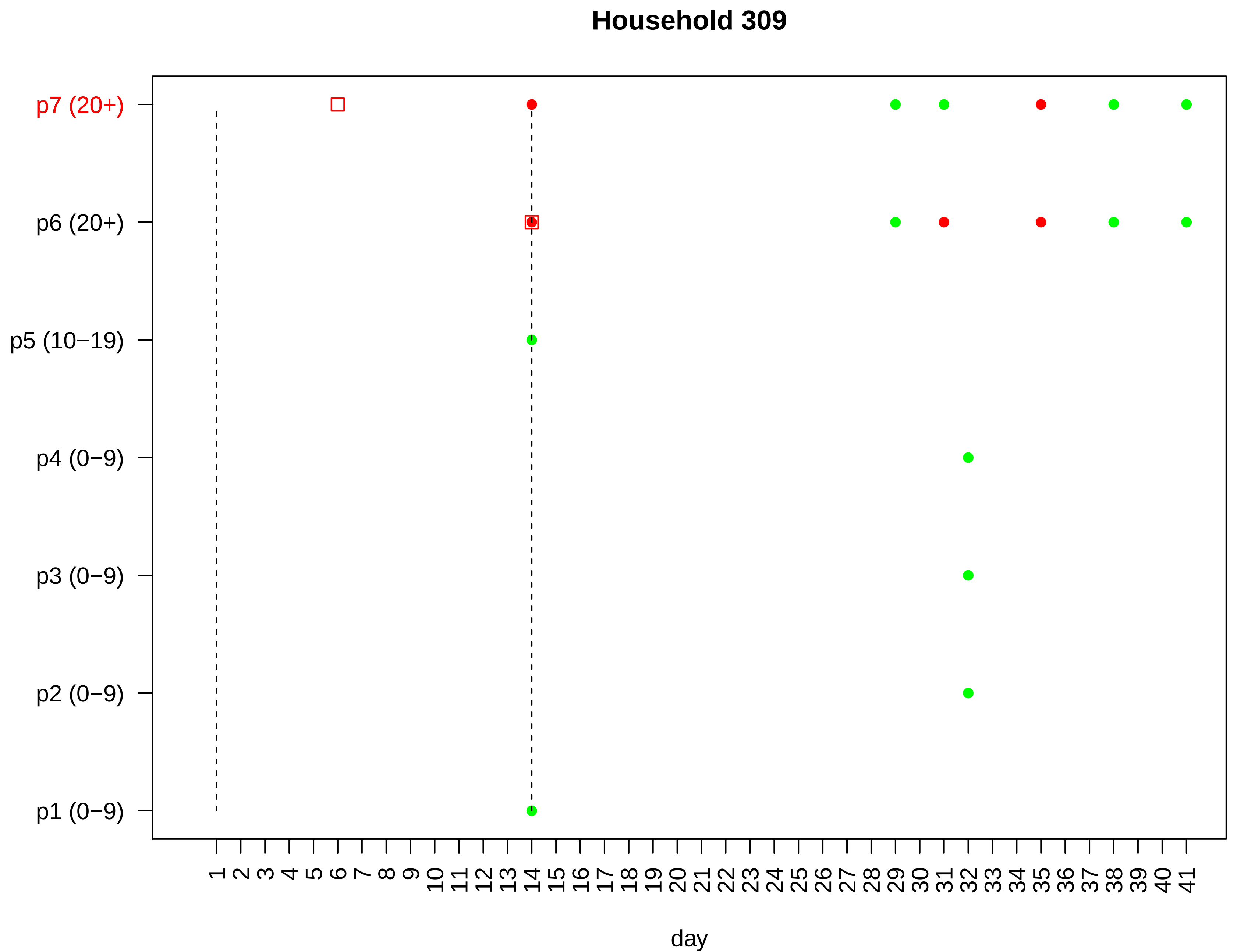

Household 309
p7 (20+)
p6 (20+)
p5 (10−19)
p4 (0−9)
p3 (0−9)
p2 (0−9)
p1 (0−9)
1 2 3 4 5 6 7 8 9 10 11 12 13 14 15 16 17 18 19 20 21 22 23 24 25 26 27 28 29 30 31 32 33 34 35 36 37 38 39 40 41
day

#### Household 310

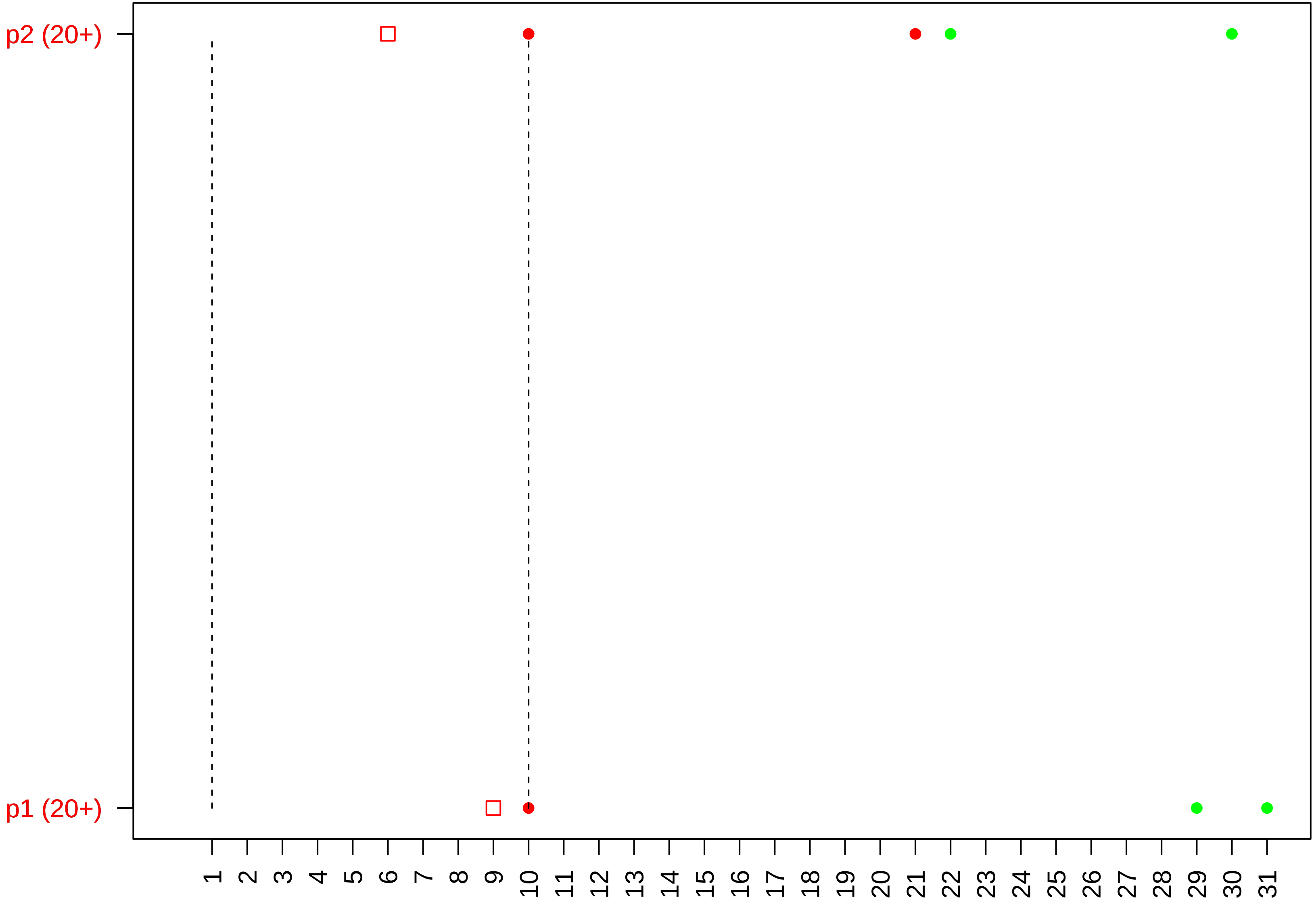

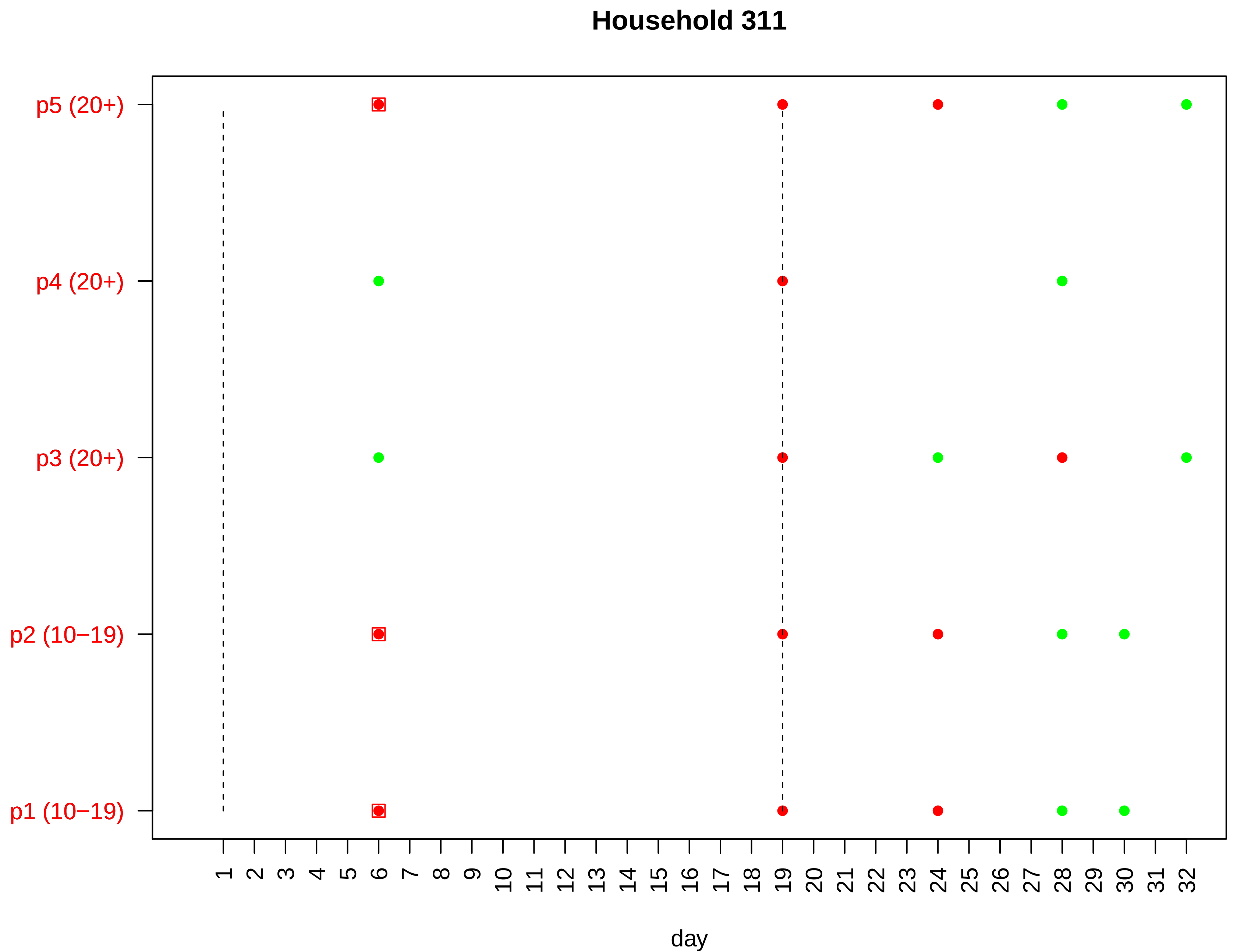

Household 311
p5 (20+)
p4 (20+)
p3 (20+)
p2 (10−19)
p1 (10−19)
day

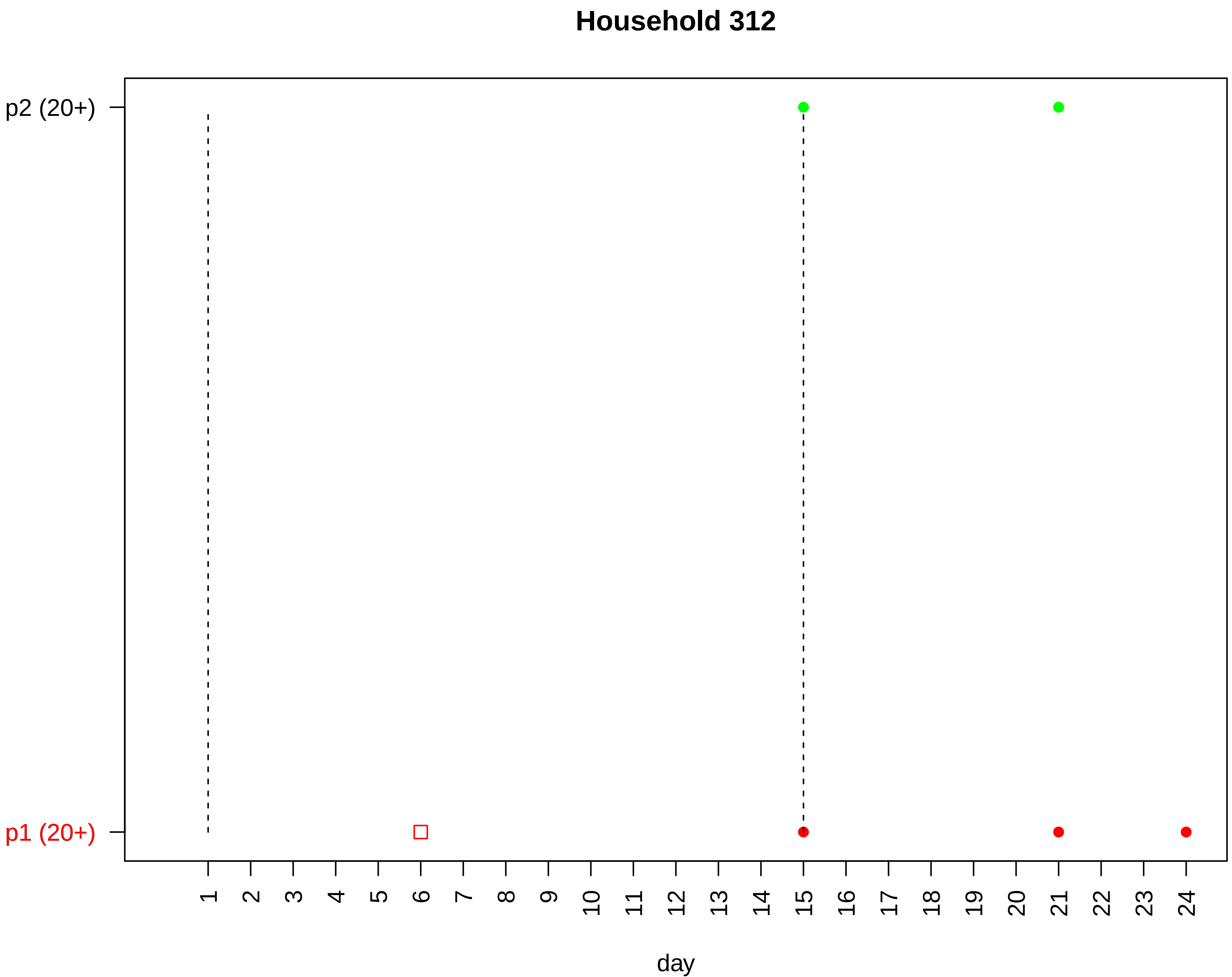
Household 312
p2 (20+)
p1 (20+)
day

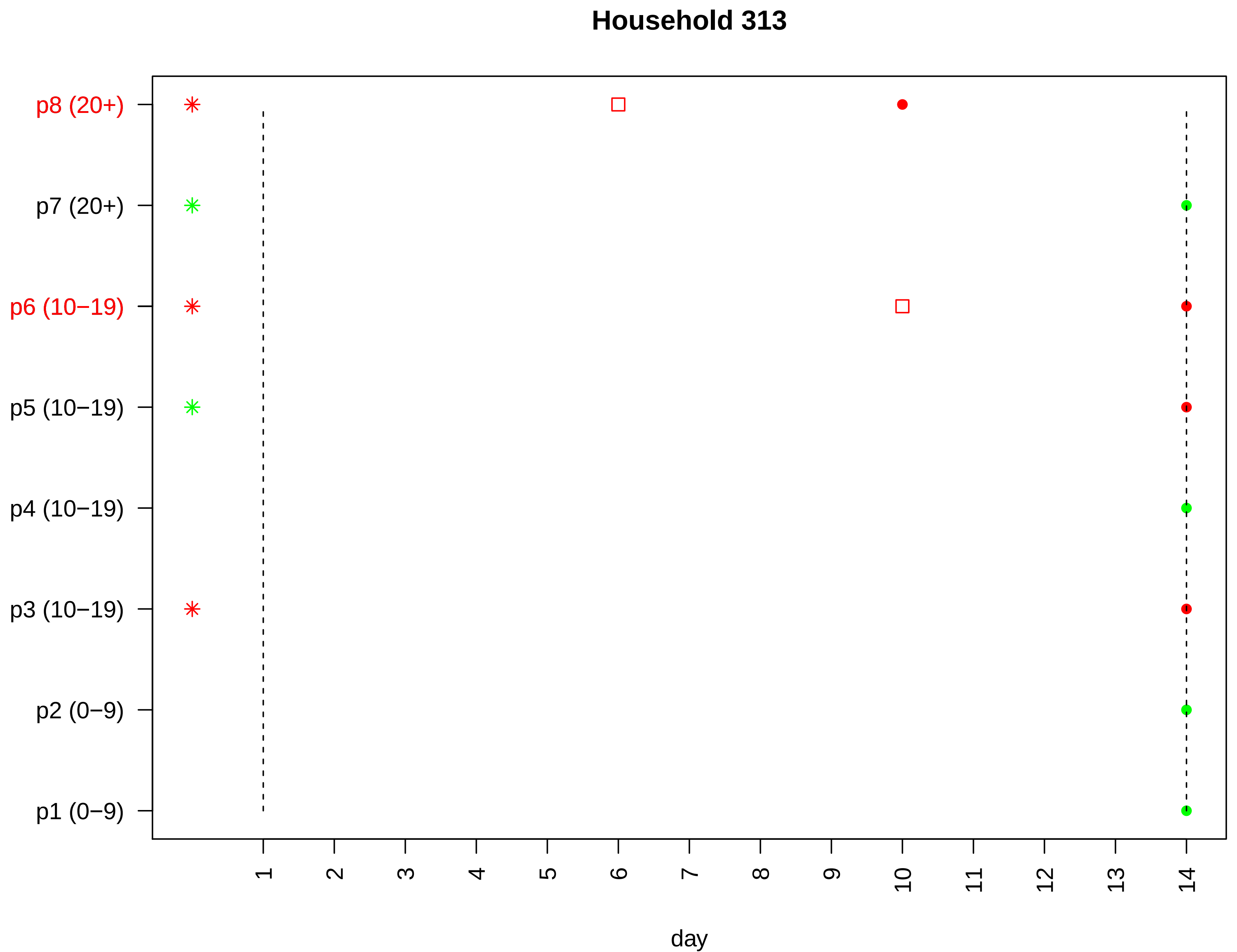

Household 313
p8 (20+)
p7 (20+)
p6 (10−19)
p5 (10−19)
p4 (10−19)
p3 (10−19)
p2 (0−9)
p1 (0−9)
1
2
3
4
5
6
7
8
9
10
11
12
13
14
day

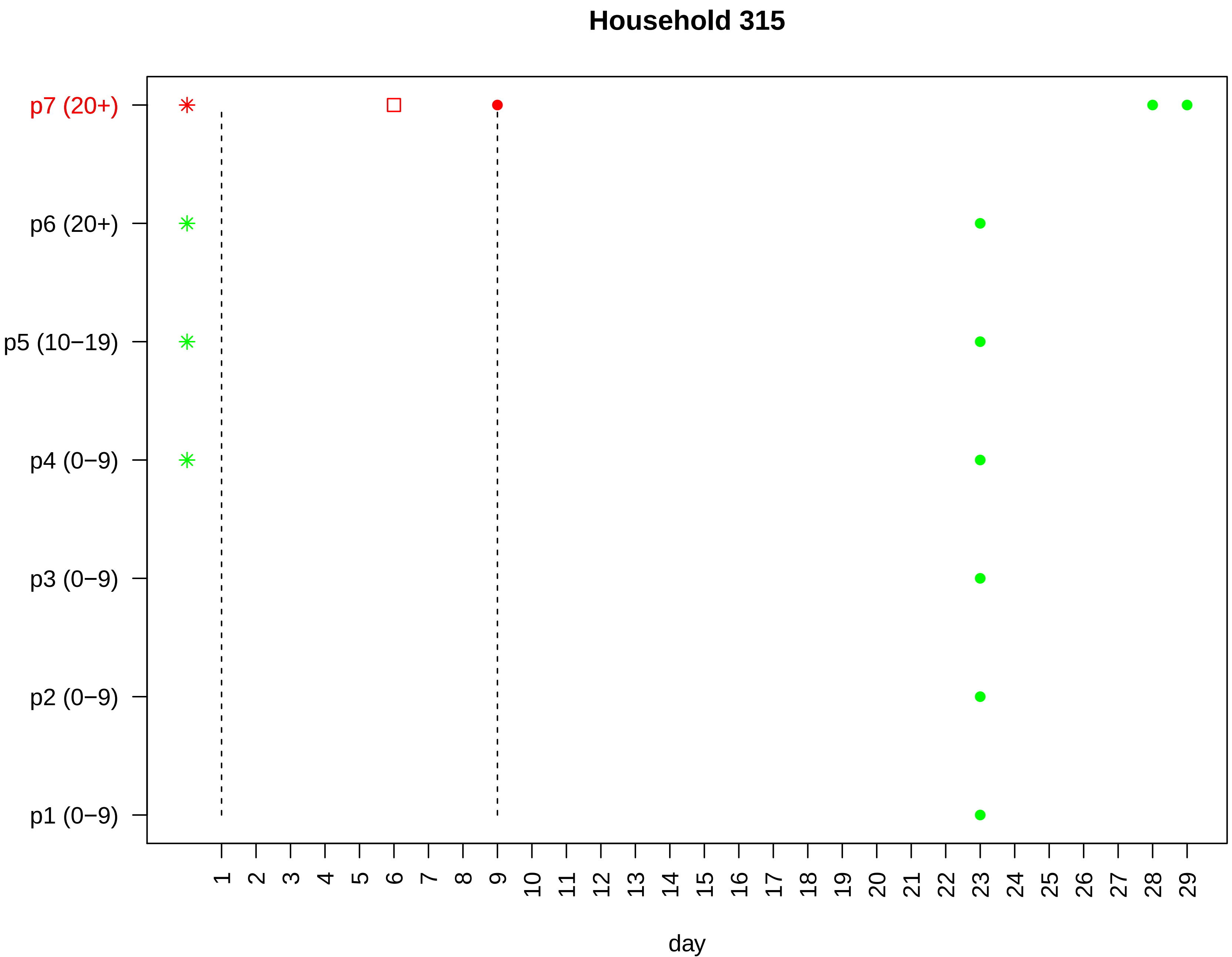

Household 315
p7 (20+)
p6 (20+)
p5 (10−19)
p4 (0−9)
p3 (0−9)
p2 (0−9)
p1 (0−9)
1 2 3 4 5 6 7 8 9 10 11 12 13 14 15 16 17 18 19 20 21 22 23 24 25 26 27 28 29
day

### Household 316

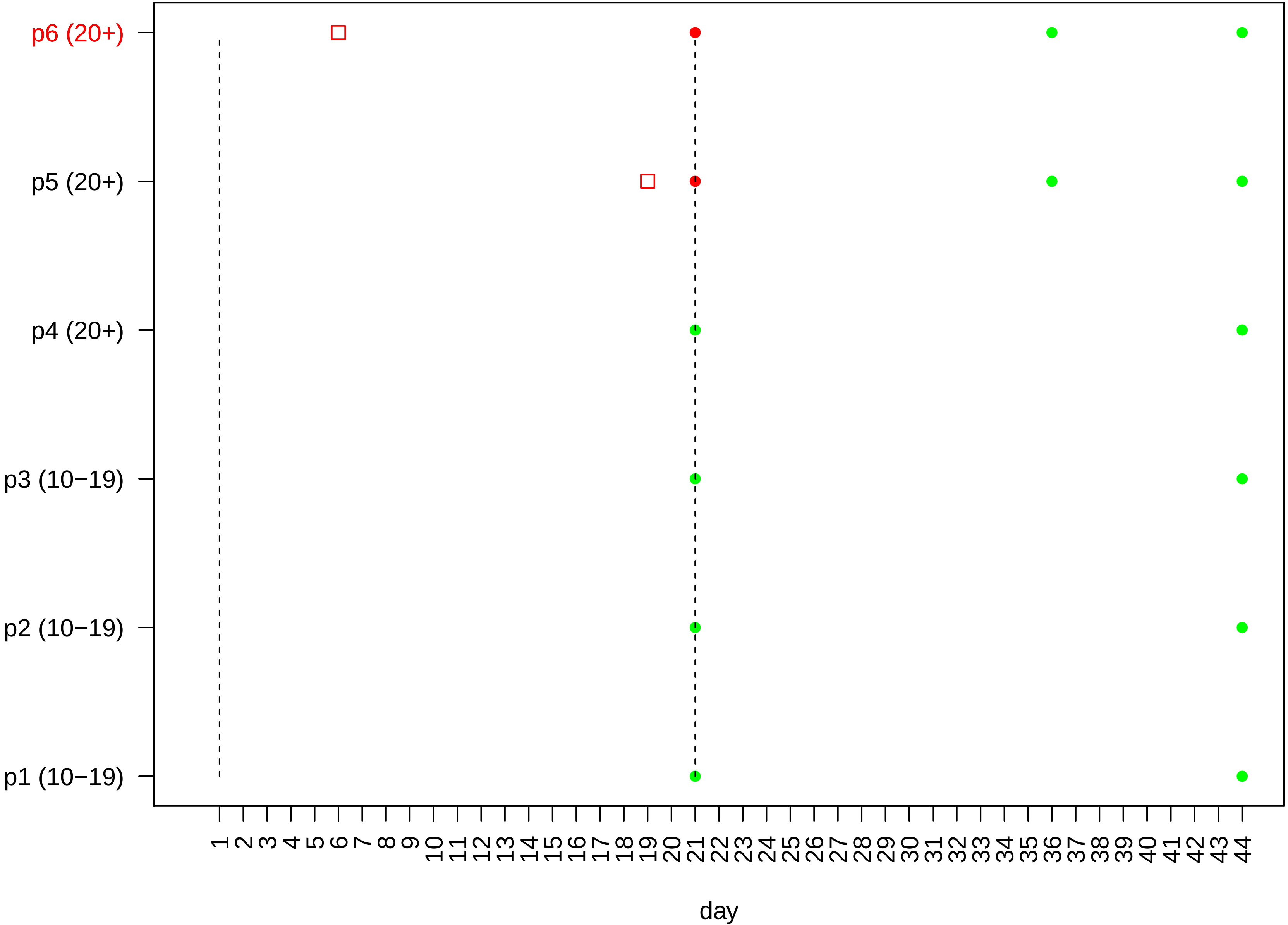

#### Household 317

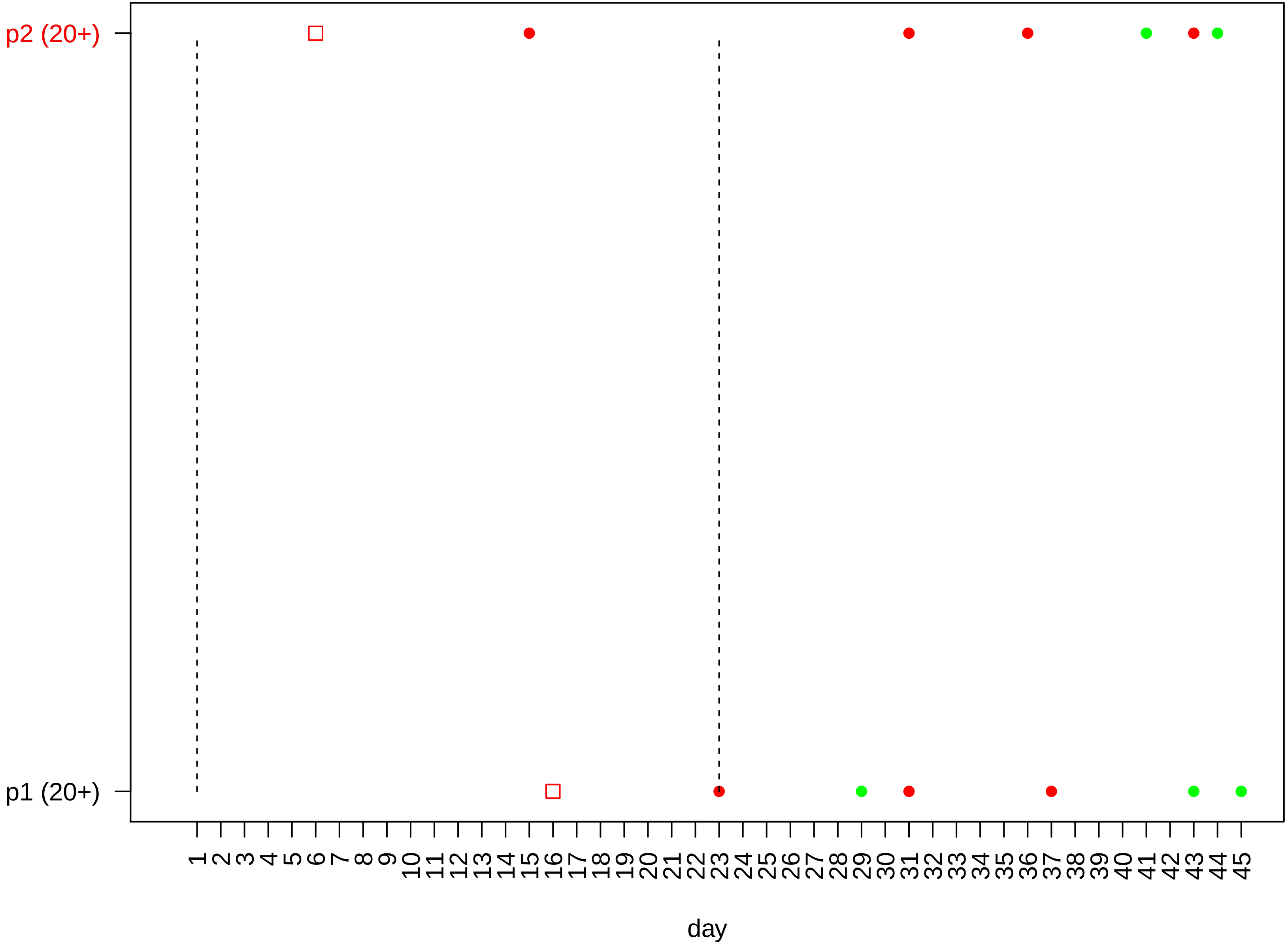

#### Household 318

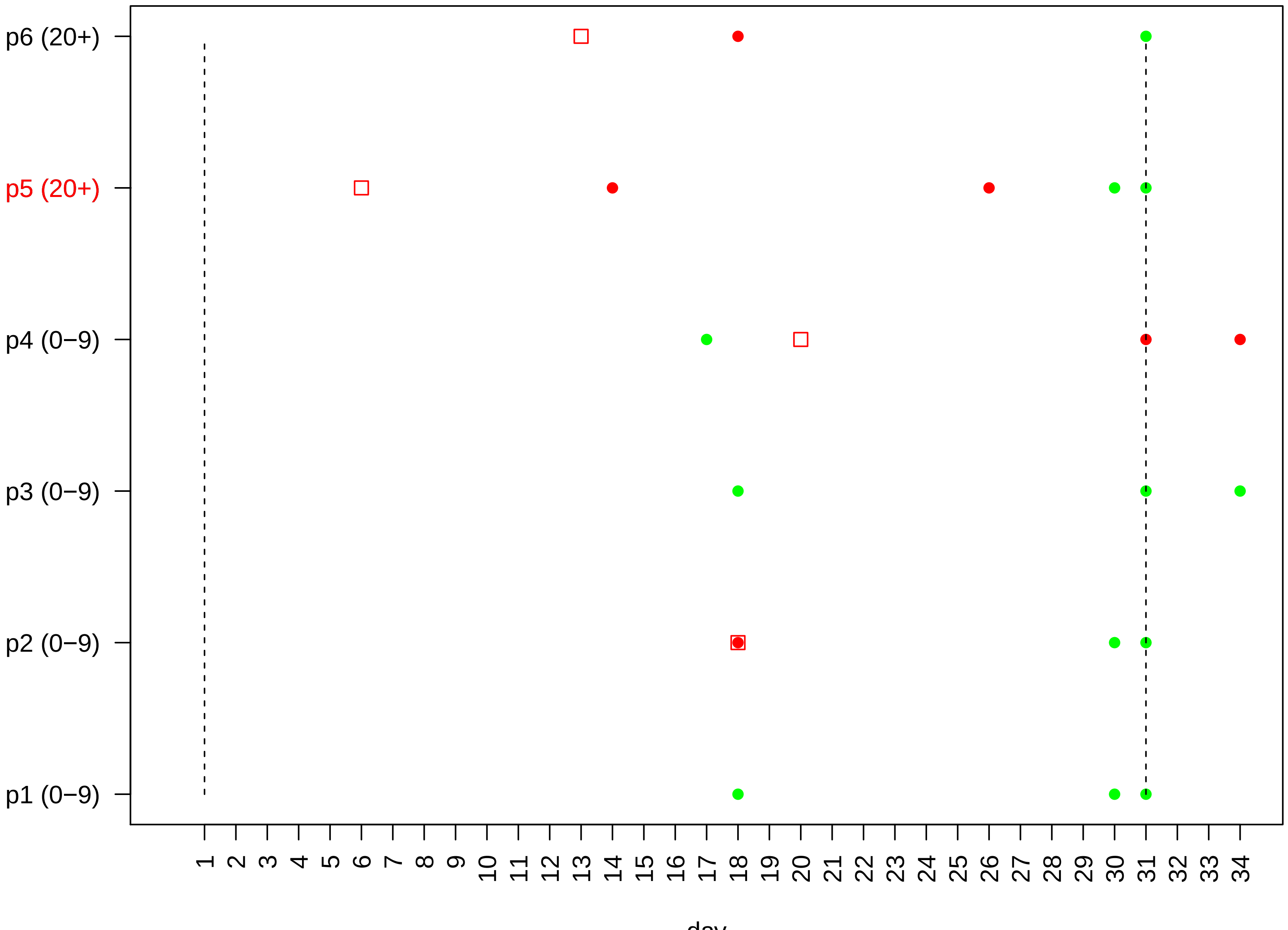

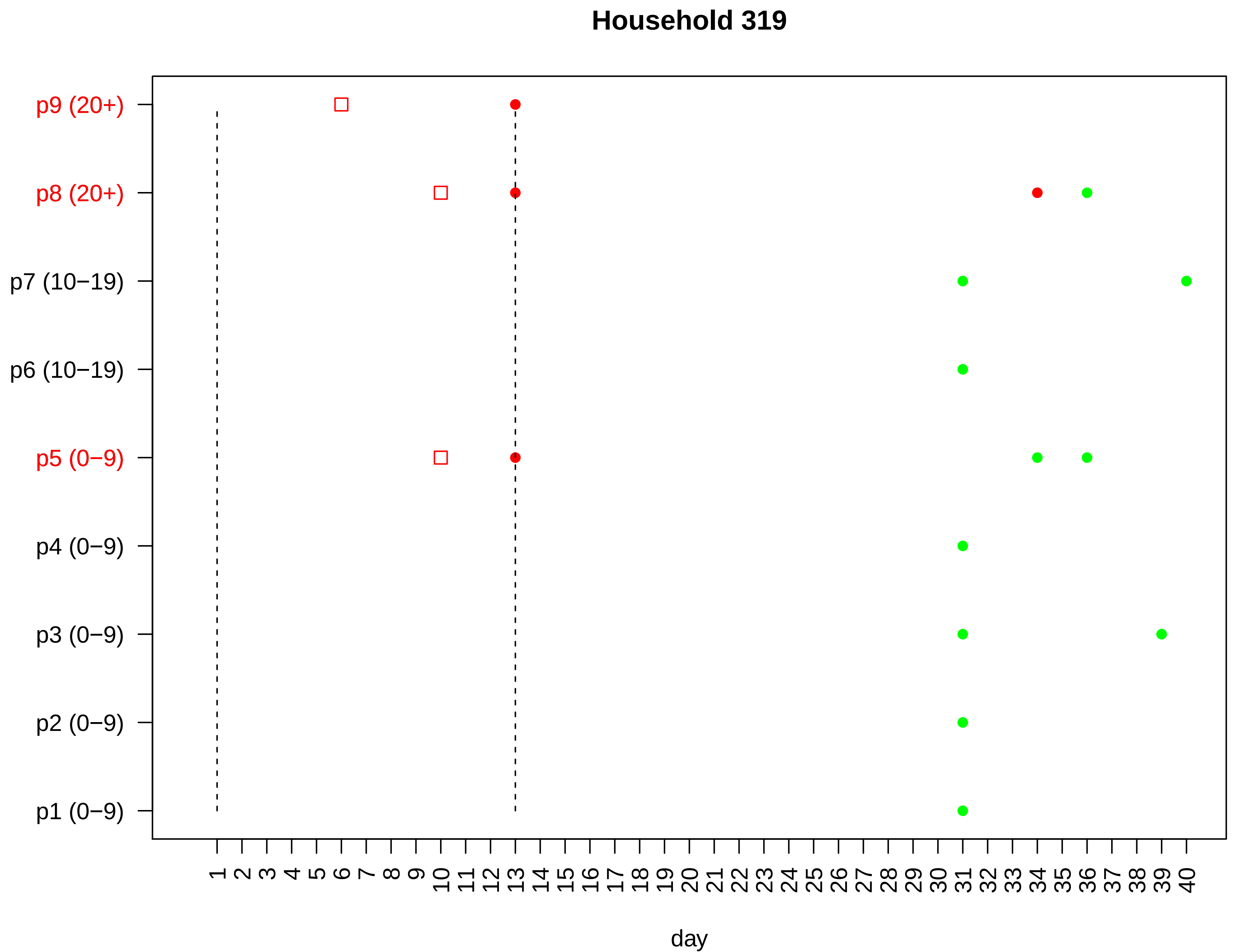

Household 319
p9 (20+)
p8 (20+)
p7 (10−19)
p6 (10−19)
p5 (0−9)
p4 (0−9)
p3 (0−9)
p2 (0−9)
p1 (0−9)
1 2 3 4 5 6 7 8 9 10 11 12 13 14 15 16 17 18 19 20 21 22 23 24 25 26 27 28 29 30 31 32 33 34 35 36 37 38 39 40
day

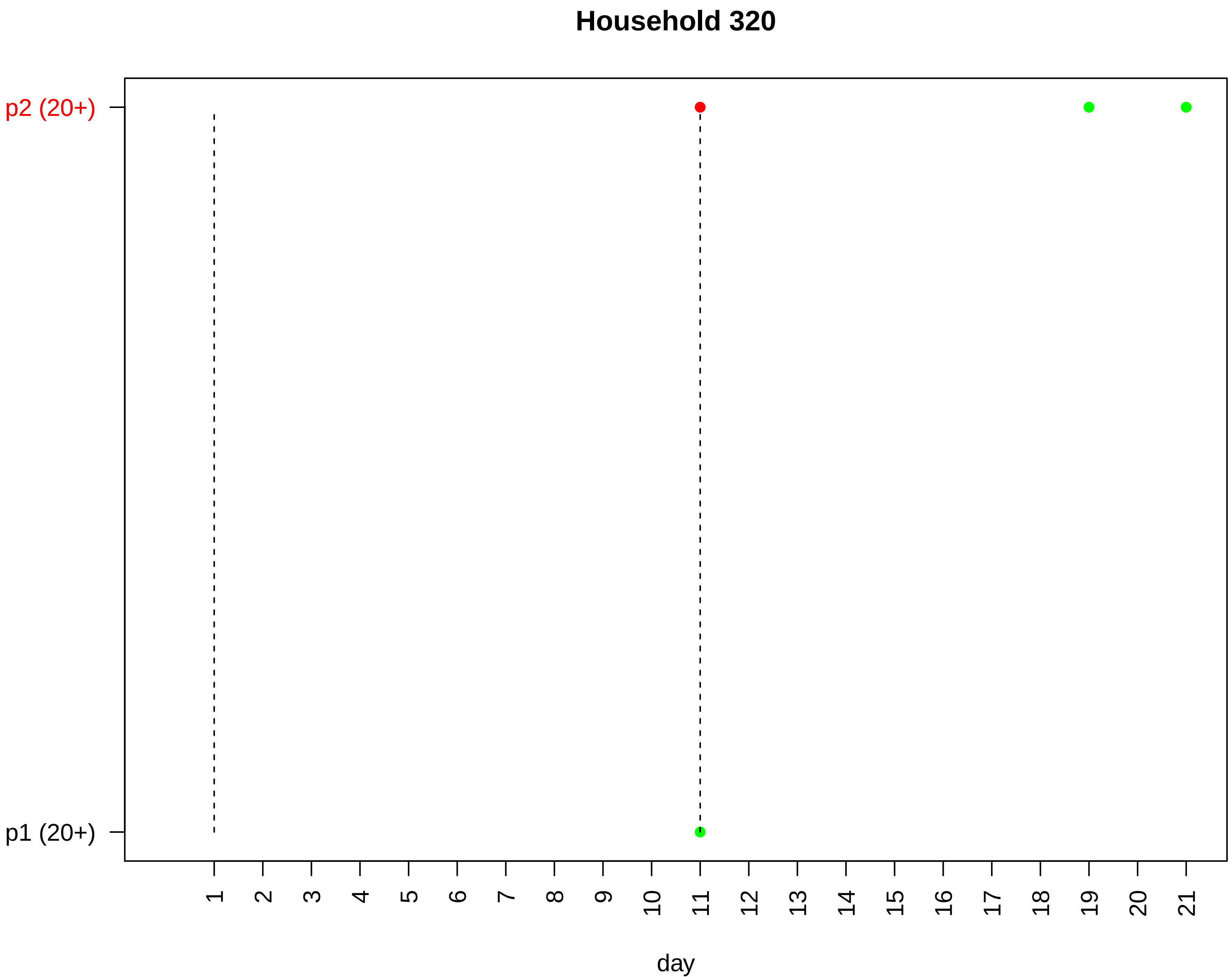
Household 320
p2 (20+)
p1 (20+)
day

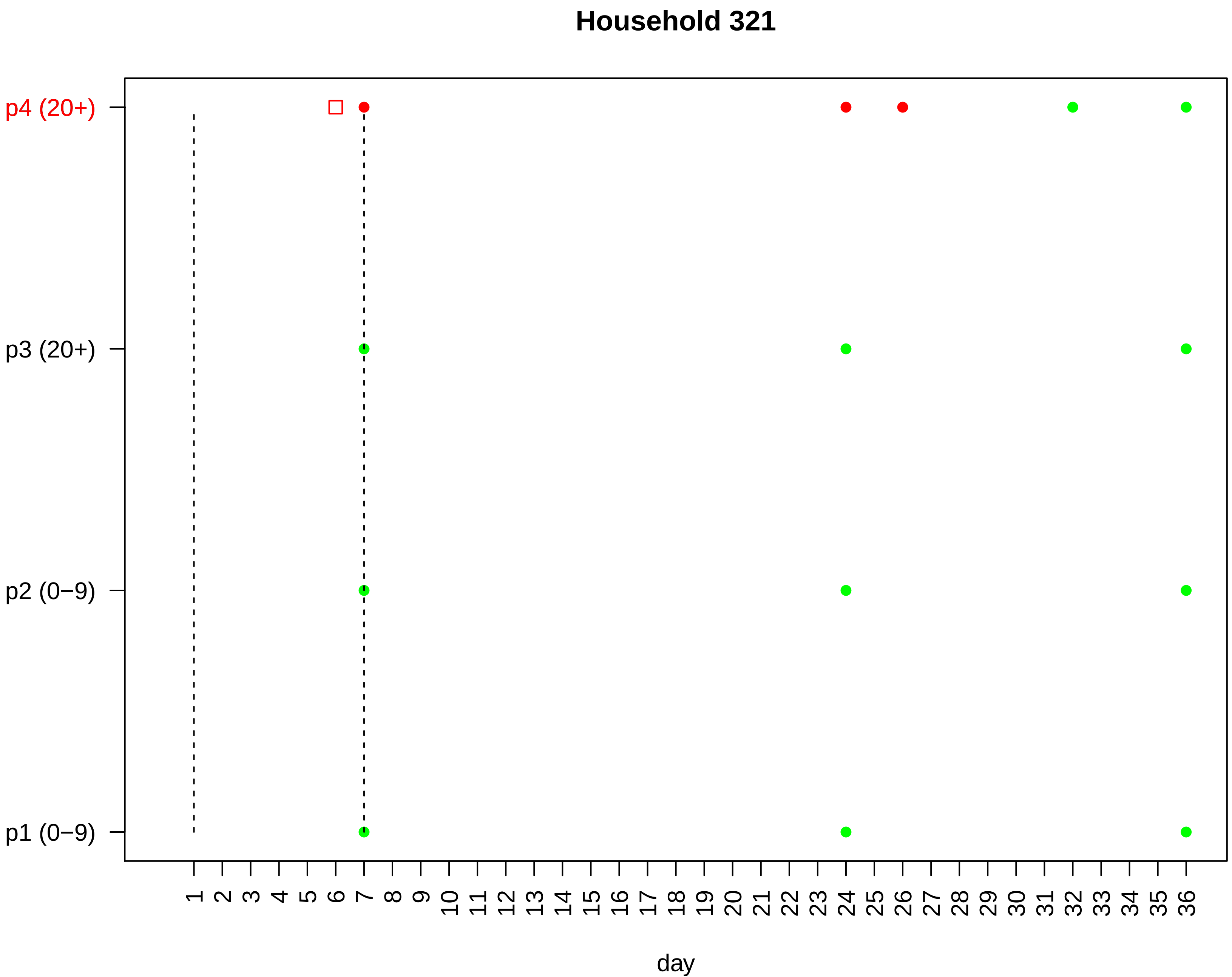
Household 321
p4 (20+)
p3 (20+)
p2 (0−9)
p1 (0−9)
day

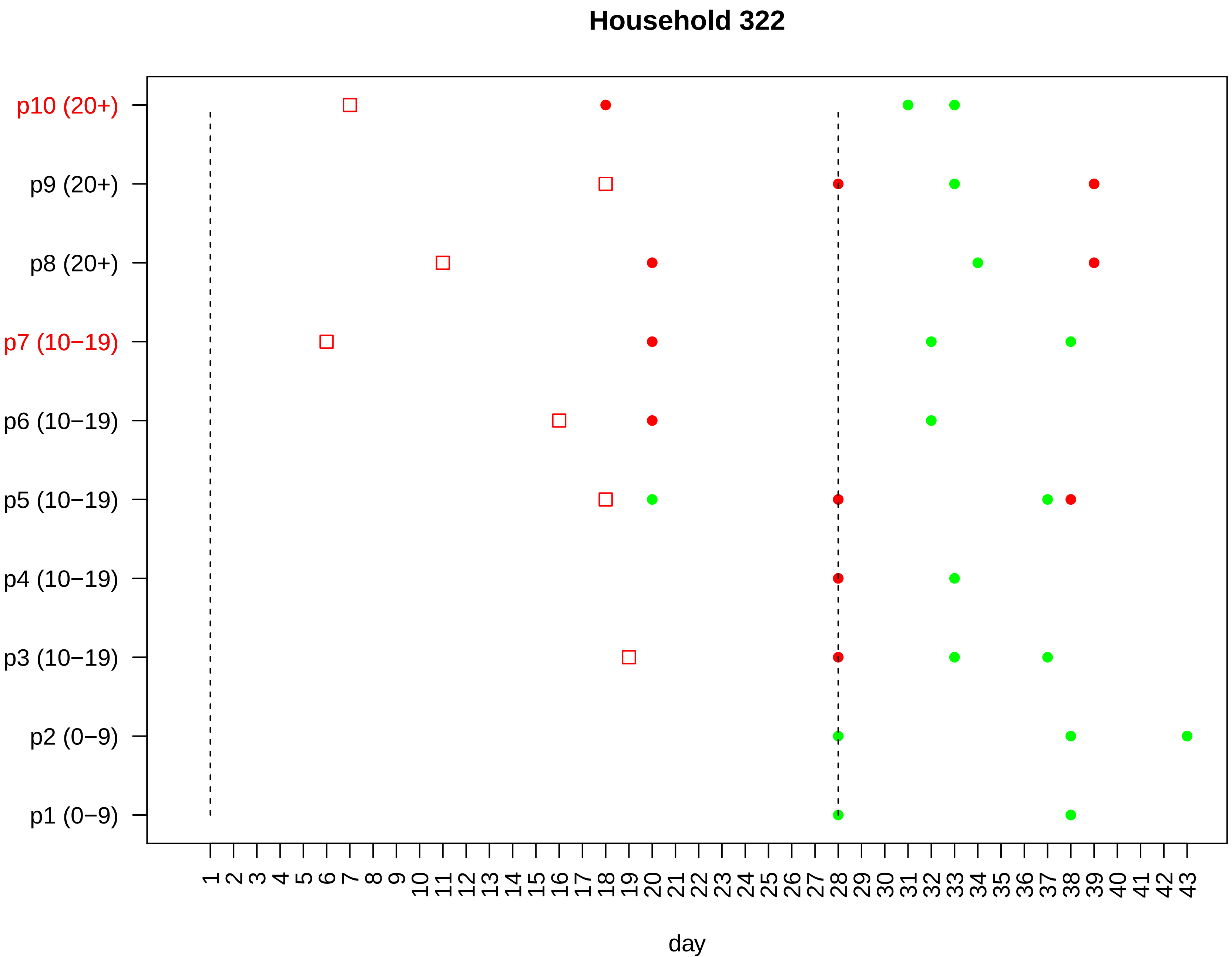

Household 322
p10 (20+)
p9 (20+)
p8 (20+)
p7 (10−19)
p6 (10−19)
p5 (10−19)
p4 (10−19)
p3 (10−19)
p2 (0−9)
p1 (0−9)
1 2 3 4 5 6 7 8 9 10 11 12 13 14 15 16 17 18 19 20 21 22 23 24 25 26 27 28 29 30 31 32 33 34 35 36 37 38 39 40 41 42 43
day

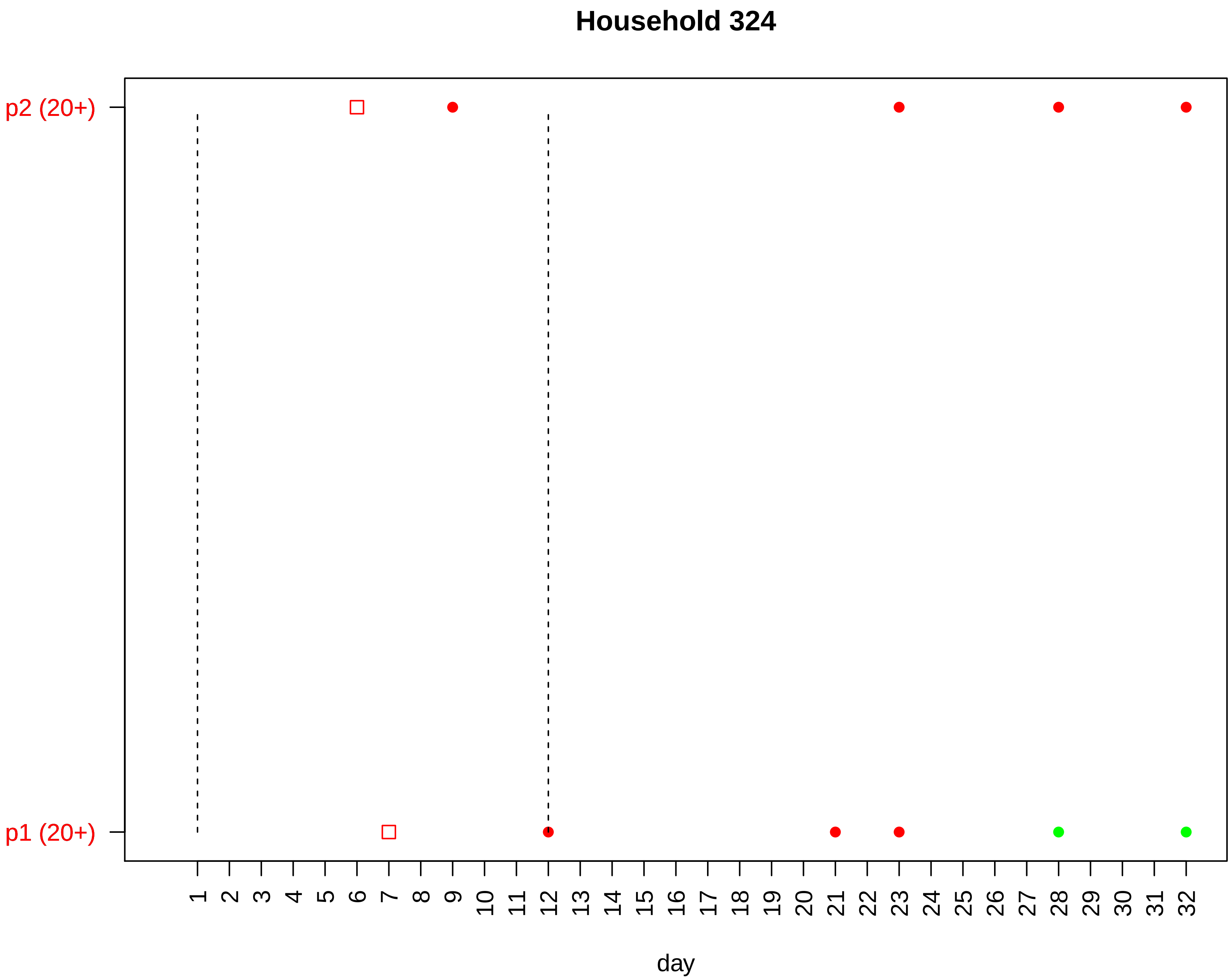

Household 324
p2 (20+)
p1 (20+)
1 2 3 4 5 6 7 8 9 10 11 12 13 14 15 16 17 18 19 20 21 22 23 24 25 26 27 28 29 30 31 32
day

### Household 326

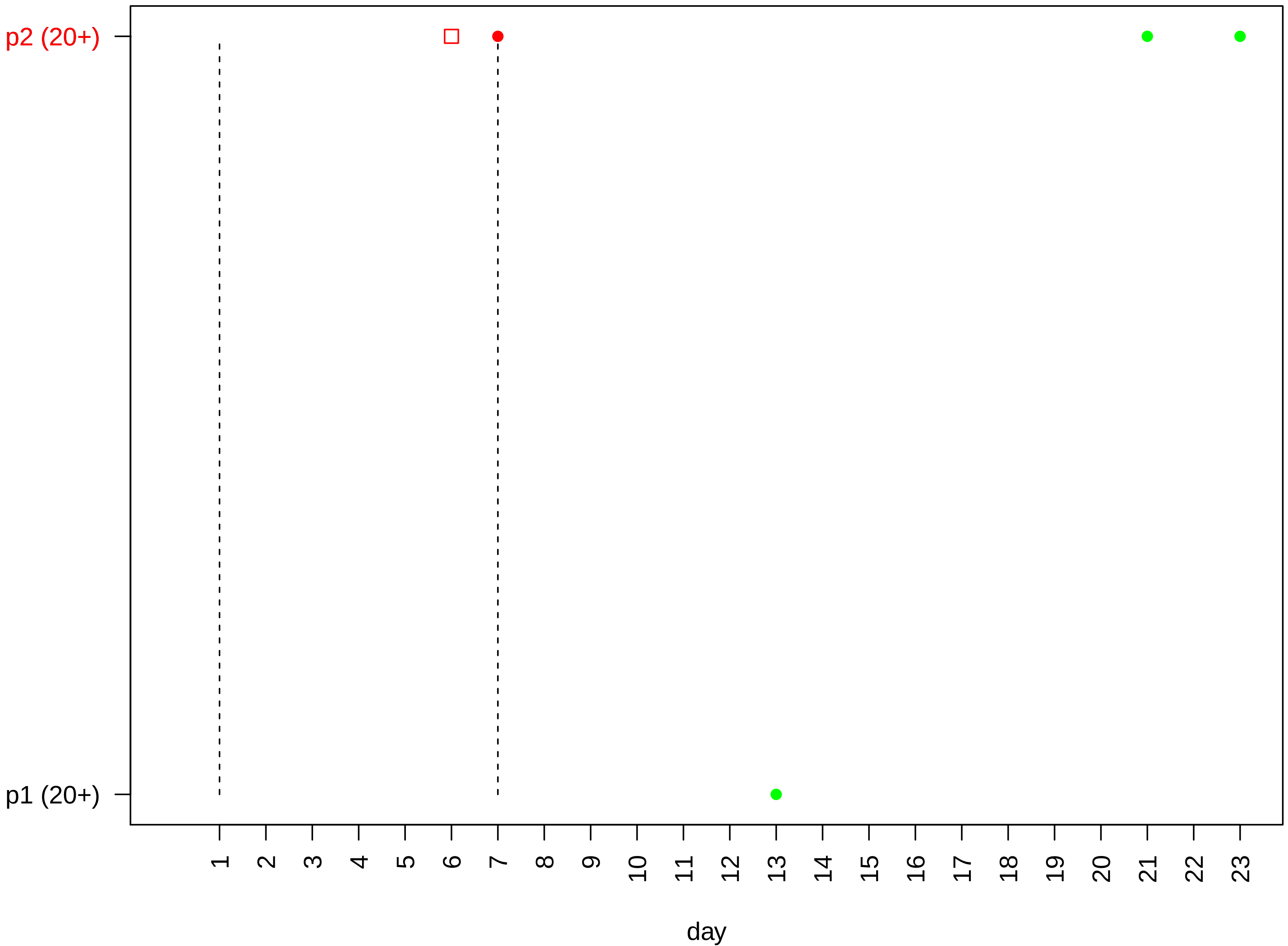

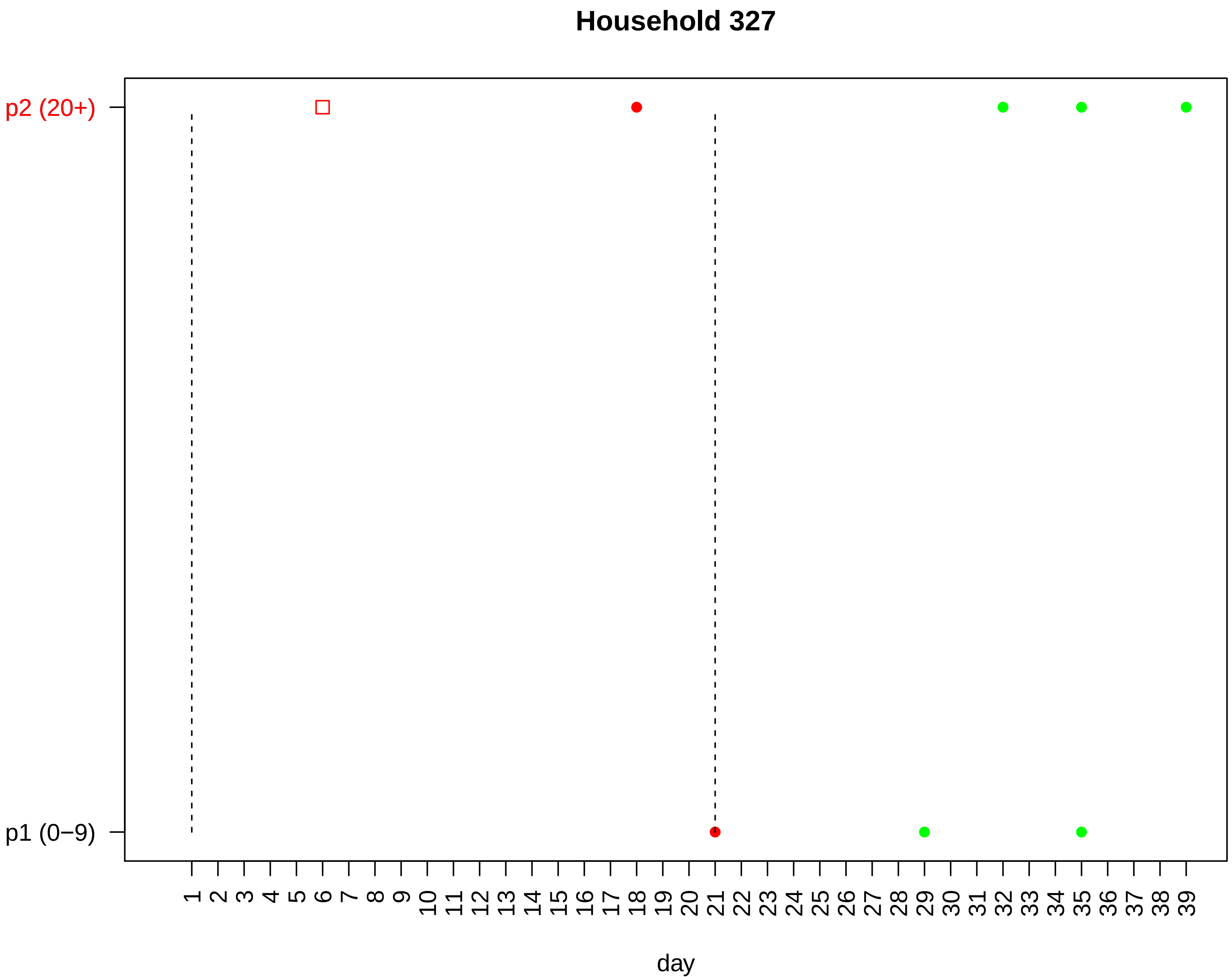

Household 327
p2 (20+)
p1 (0−9)
1 2 3 4 5 6 7 8 9 10 11 12 13 14 15 16 17 18 19 20 21 22 23 24 25 26 27 28 29 30 31 32 33 34 35 36 37 38 39
day

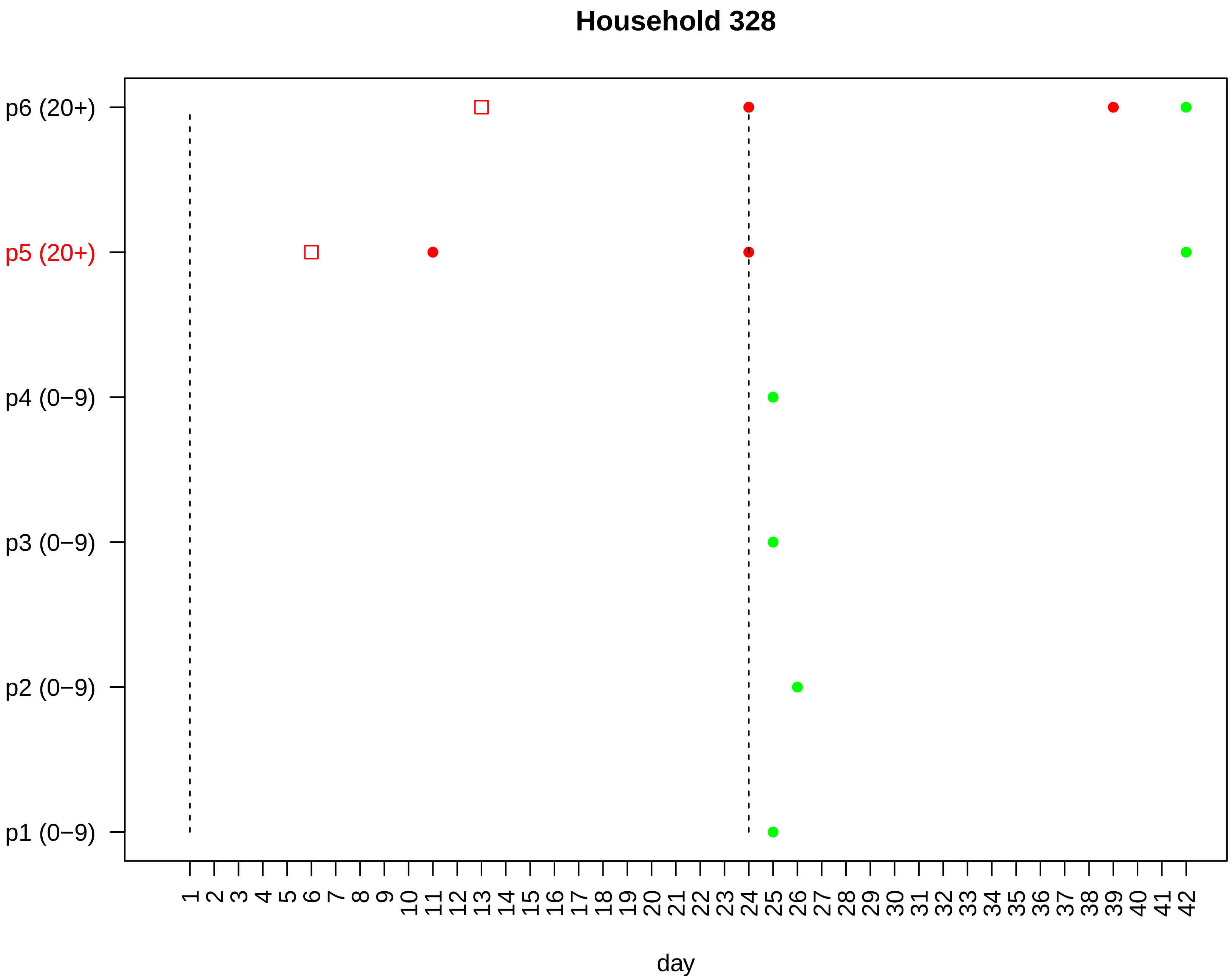

Household 328
p6 (20+)
p5 (20+)
p4 (0−9)
p3 (0−9)
p2 (0−9)
p1 (0−9)
1 2 3 4 5 6 7 8 9 10 11 12 13 14 15 16 17 18 19 20 21 22 23 24 25 26 27 28 29 30 31 32 33 34 35 36 37 38 39 40 41 42
day

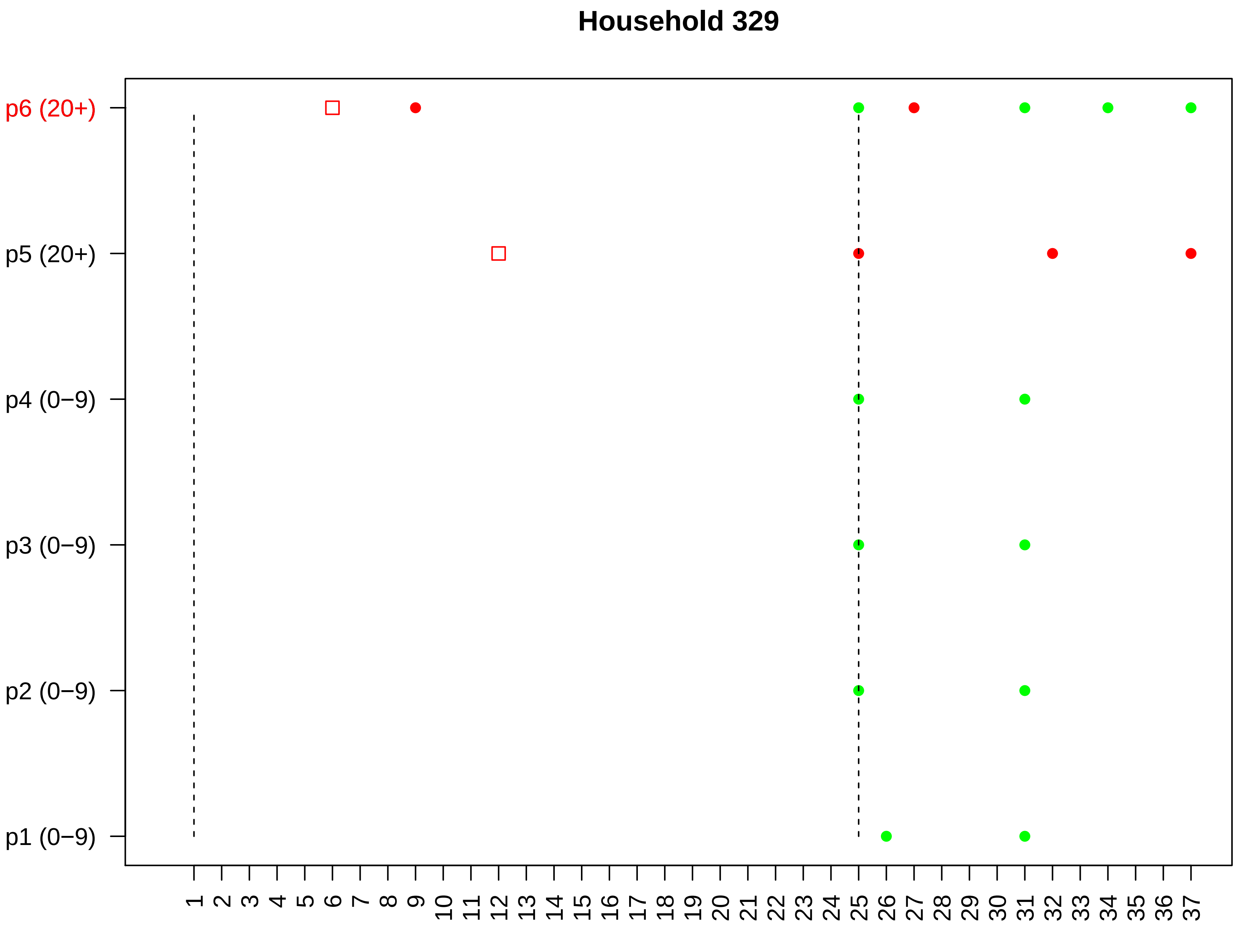

Household 329
p6 (20+)
p5 (20+)
p4 (0−9)
p3 (0−9)
p2 (0−9)
p1 (0−9)
day

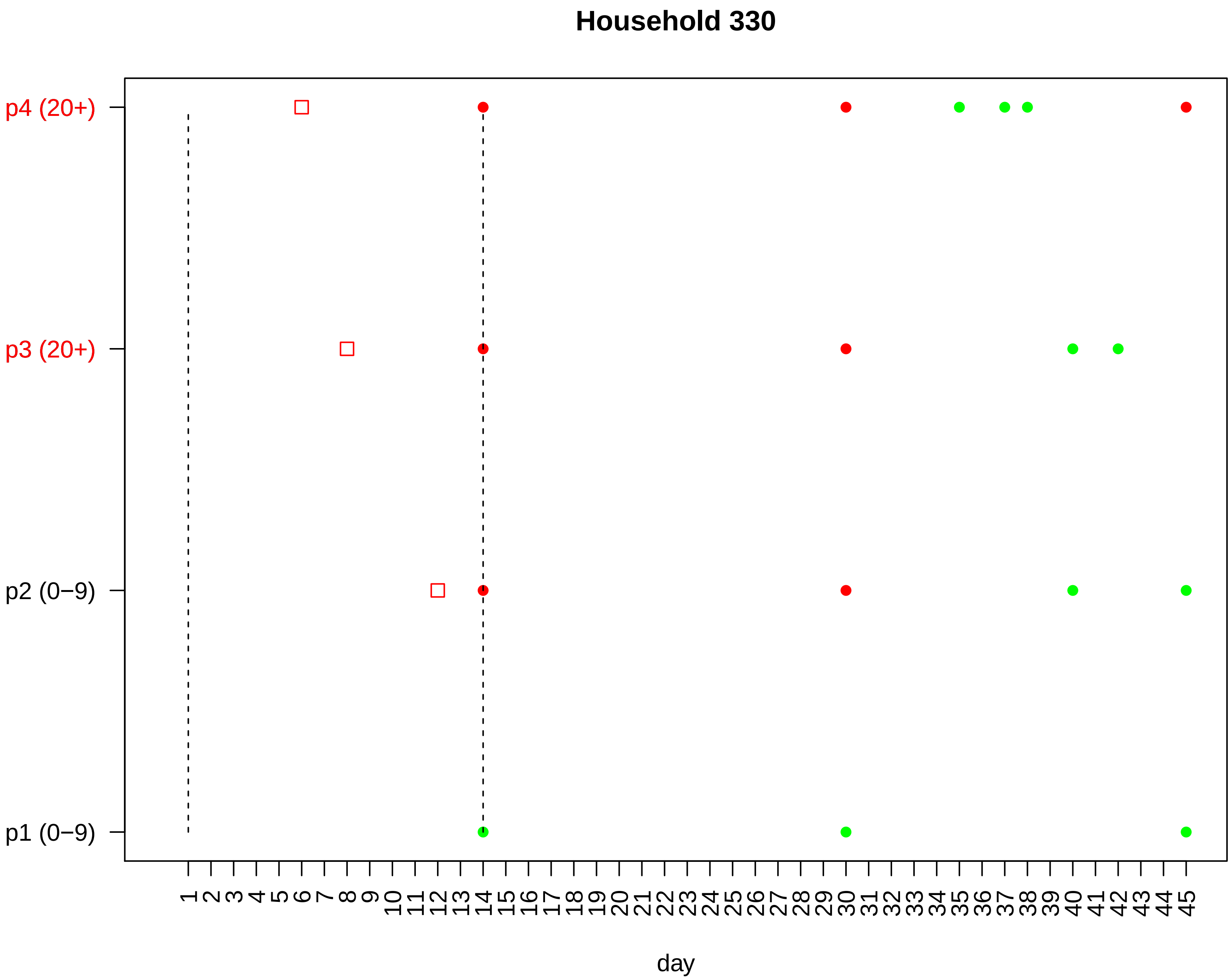

Household 330
p4 (20+)
p3 (20+)
p2 (0−9)
p1 (0−9)
1 2 3 4 5 6 7 8 9 10 11 12 13 14 15 16 17 18 19 20 21 22 23 24 25 26 27 28 29 30 31 32 33 34 35 36 37 38 39 40 41 42 43 44 45
day

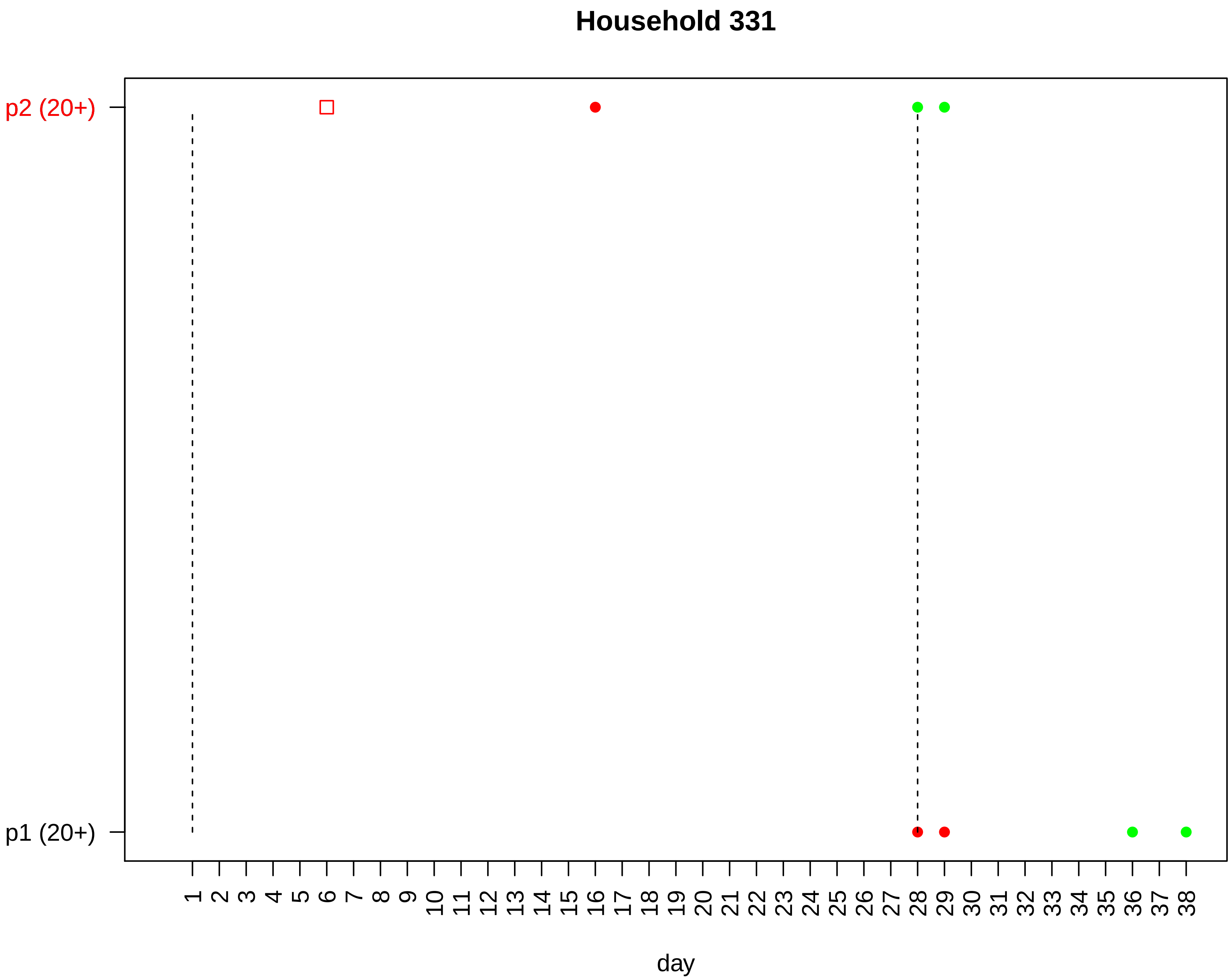
Household 331
p2 (20+)
p1 (20+)
day

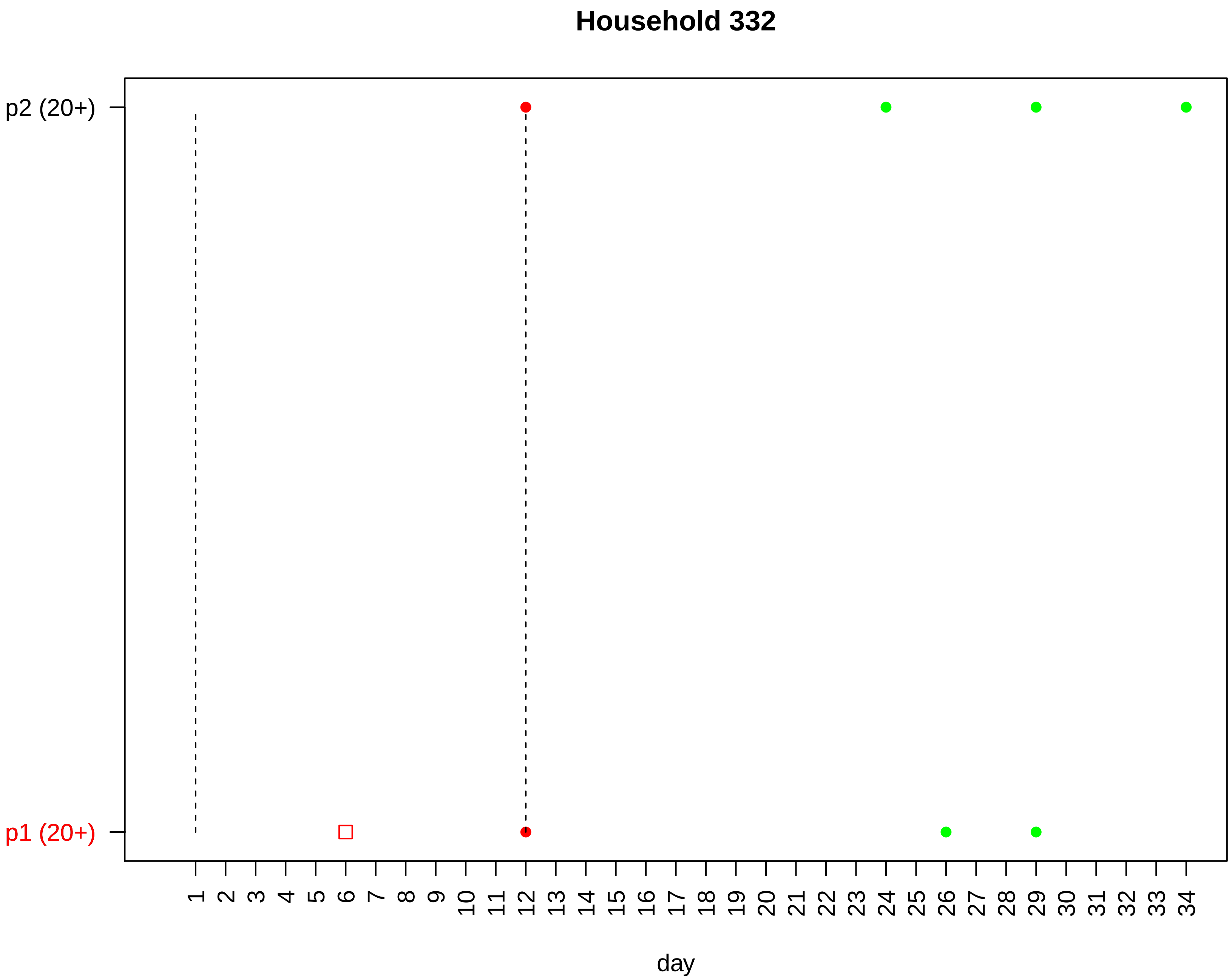

Household 332
p2 (20+)
p1 (20+)
day

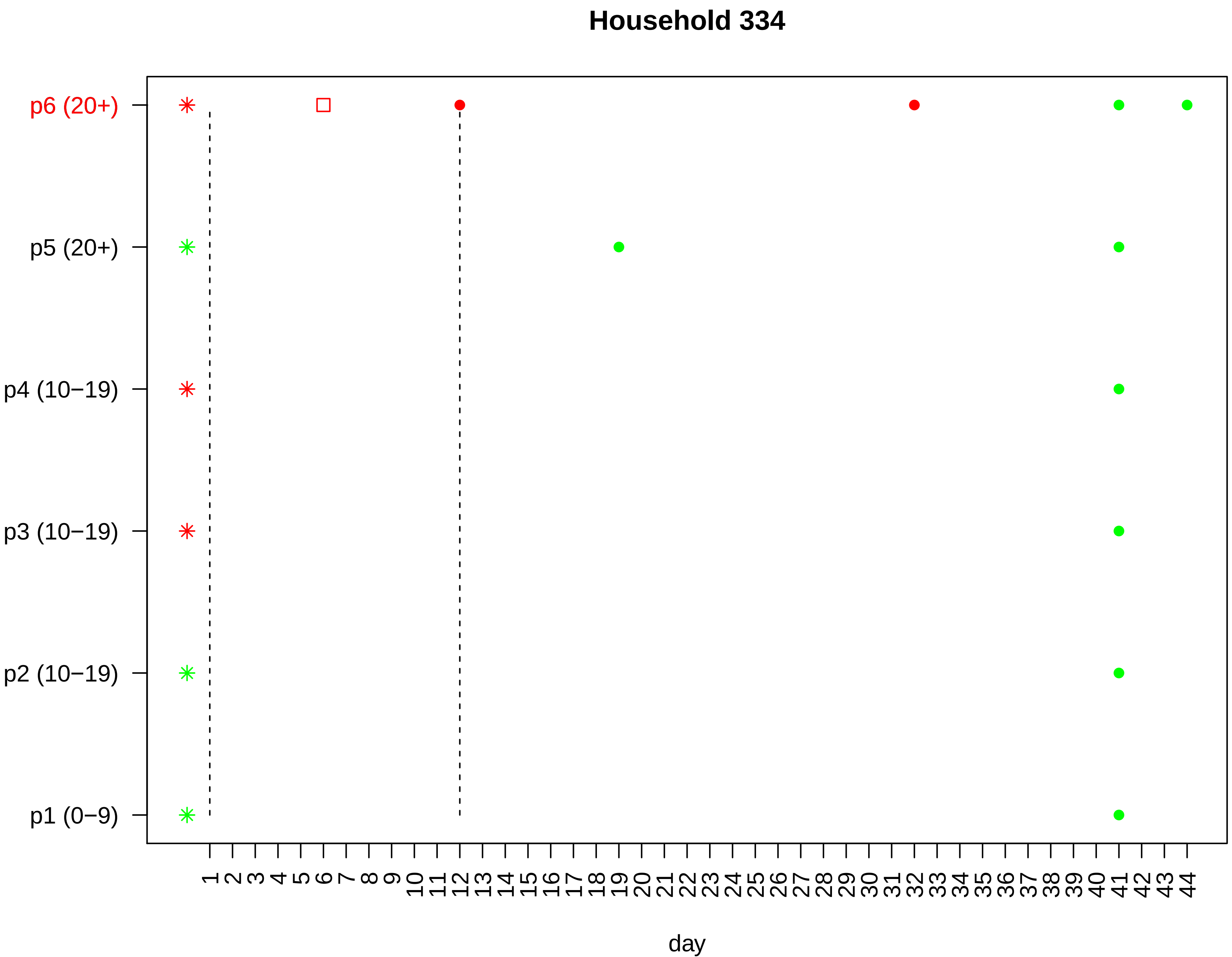

Household 334
p6 (20+)
p5 (20+)
p4 (10−19)
p3 (10−19)
p2 (10−19)
p1 (0−9)
1 2 3 4 5 6 7 8 9 10 11 12 13 14 15 16 17 18 19 20 21 22 23 24 25 26 27 28 29 30 31 32 33 34 35 36 37 38 39 40 41 42 43 44
day

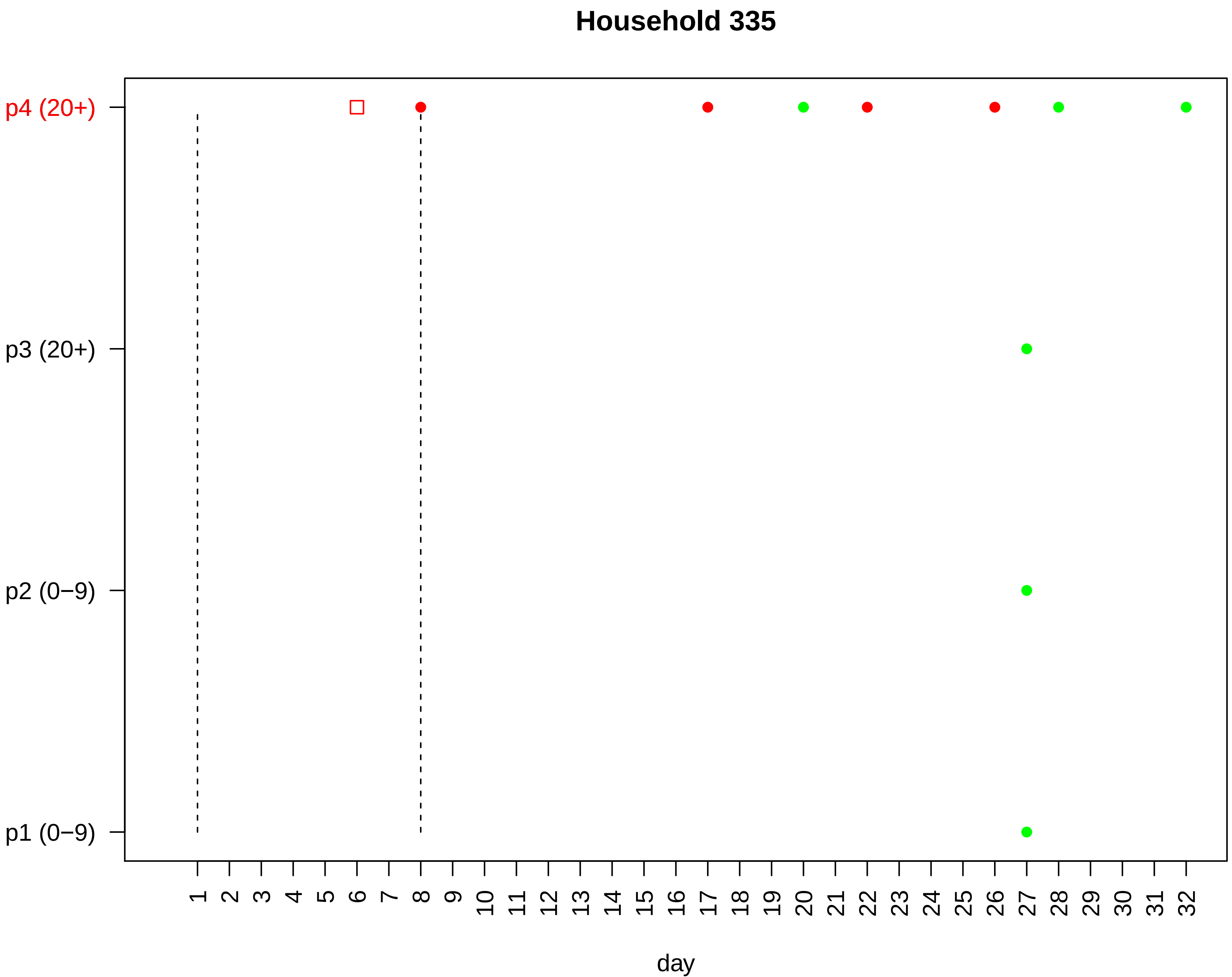

Household 335
p4 (20+)
p3 (20+)
p2 (0−9)
p1 (0−9)
1 2 3 4 5 6 7 8 9 10 11 12 13 14 15 16 17 18 19 20 21 22 23 24 25 26 27 28 29 30 31 32
day

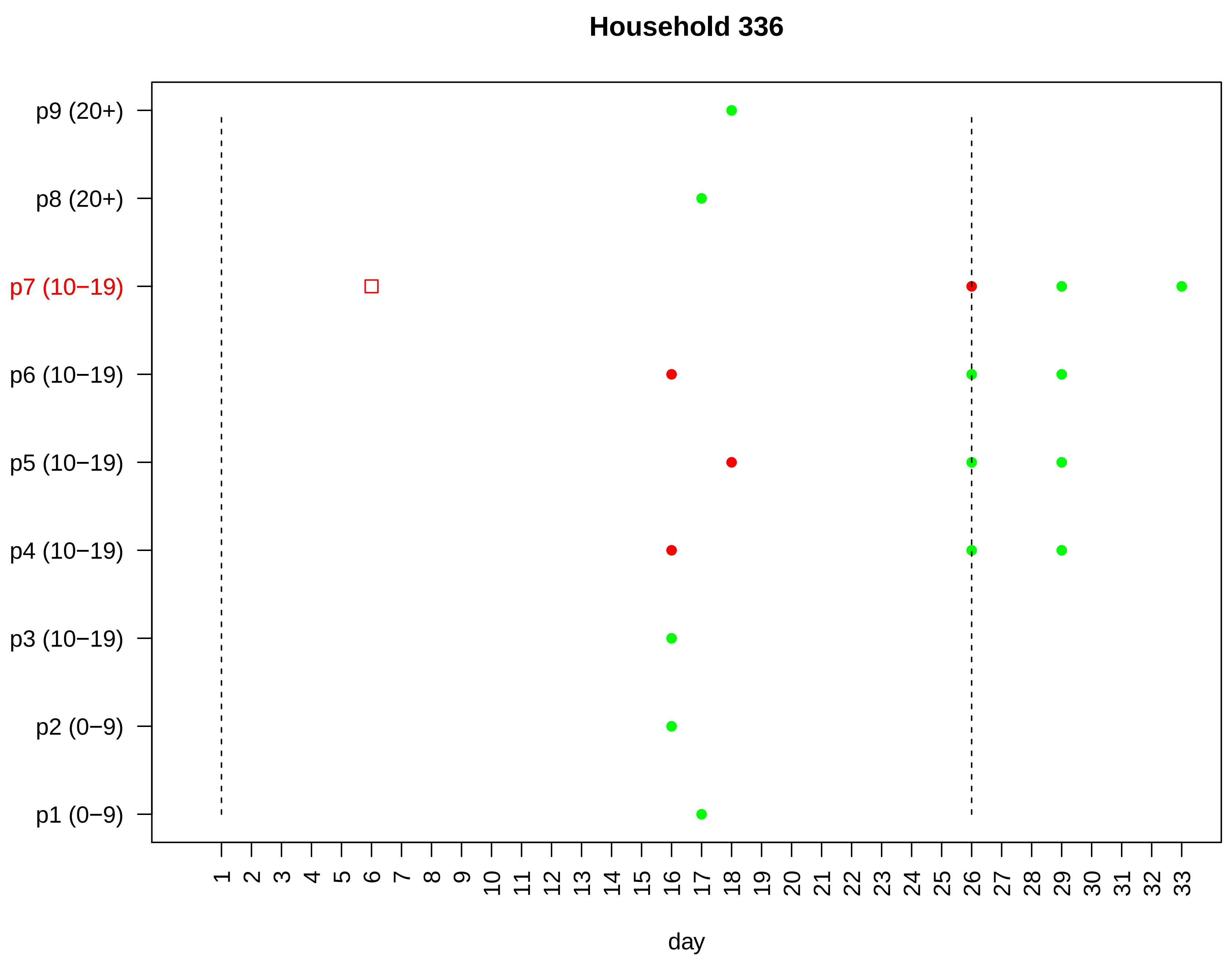
Household 336
p9 (20+)
p8 (20+)
p7 (10−19)
p6 (10−19)
p5 (10−19)
p4 (10−19)
p3 (10−19)
p2 (0−9)
p1 (0−9)
1 2 3 4 5 6 7 8 9 10 11 12 13 14 15 16 17 18 19 20 21 22 23 24 25 26 27 28 29 30 31 32 33
day

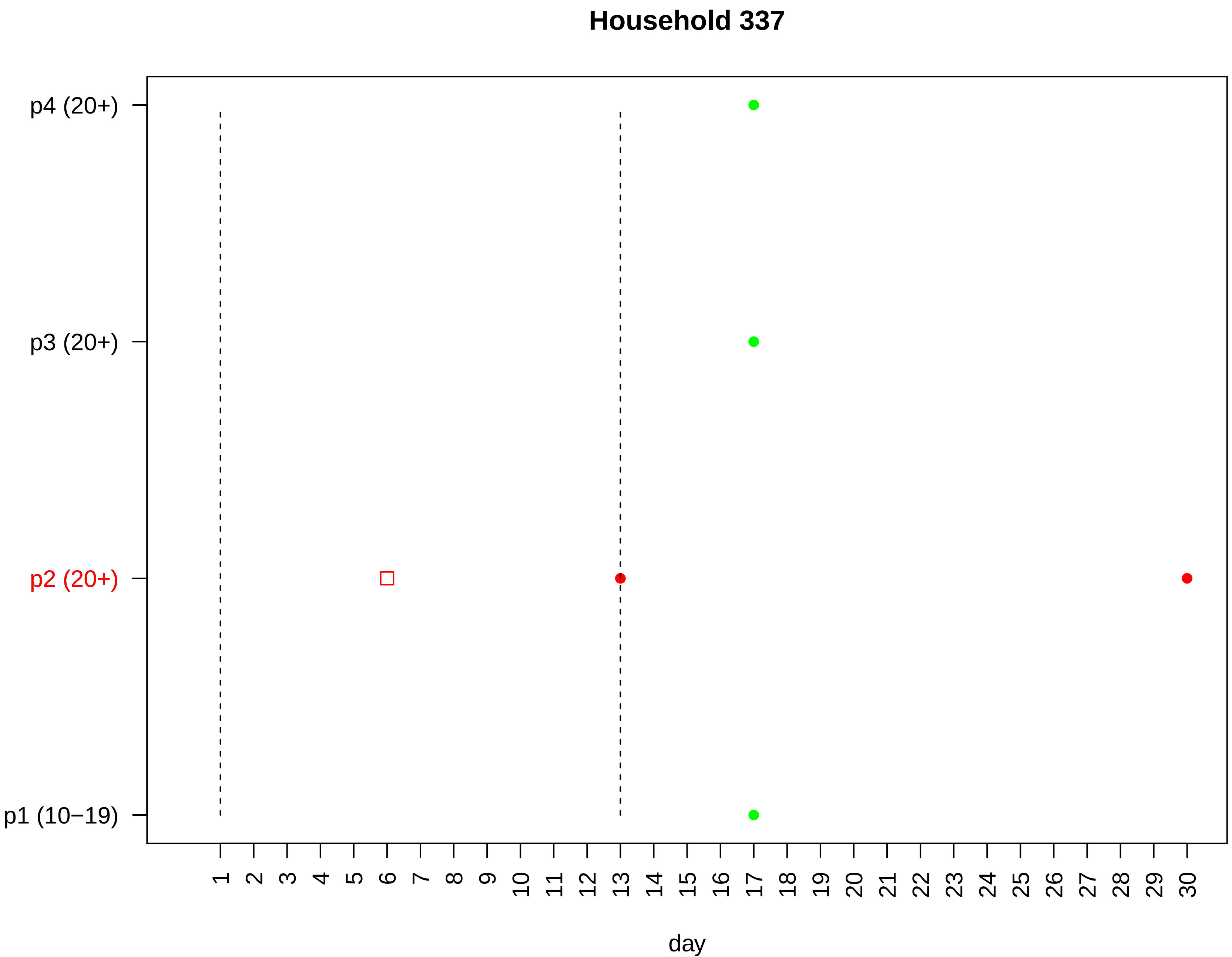

Household 337
p4 (20+)
p3 (20+)
p2 (20+)
p1 (10−19)
1 2 3 4 5 6 7 8 9 10 11 12 13 14 15 16 17 18 19 20 21 22 23 24 25 26 27 28 29 30
day

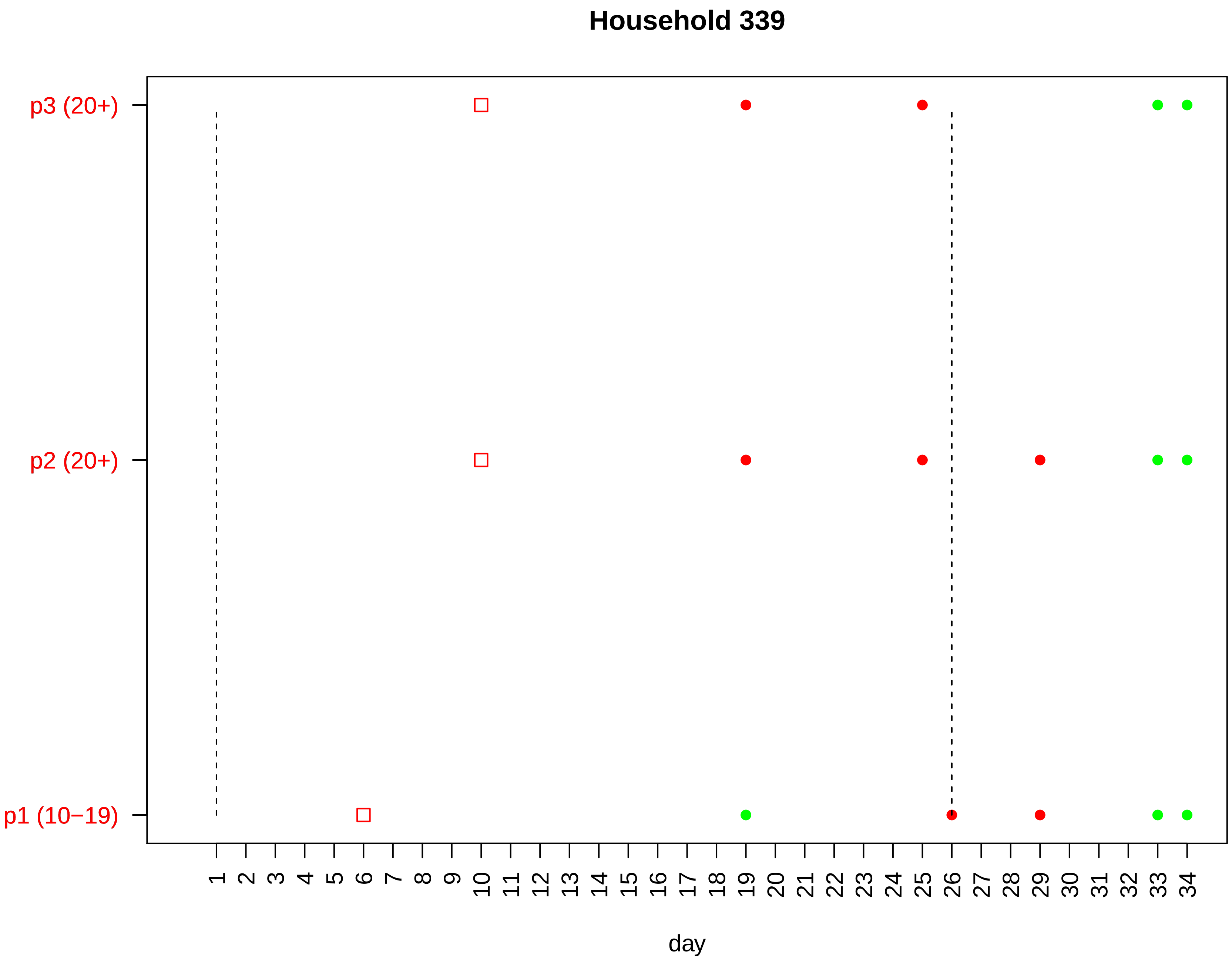

Household 339
p3 (20+)
p2 (20+)
p1 (10−19)
1 2 3 4 5 6 7 8 9 10 11 12 13 14 15 16 17 18 19 20 21 22 23 24 25 26 27 28 29 30 31 32 33 34
day

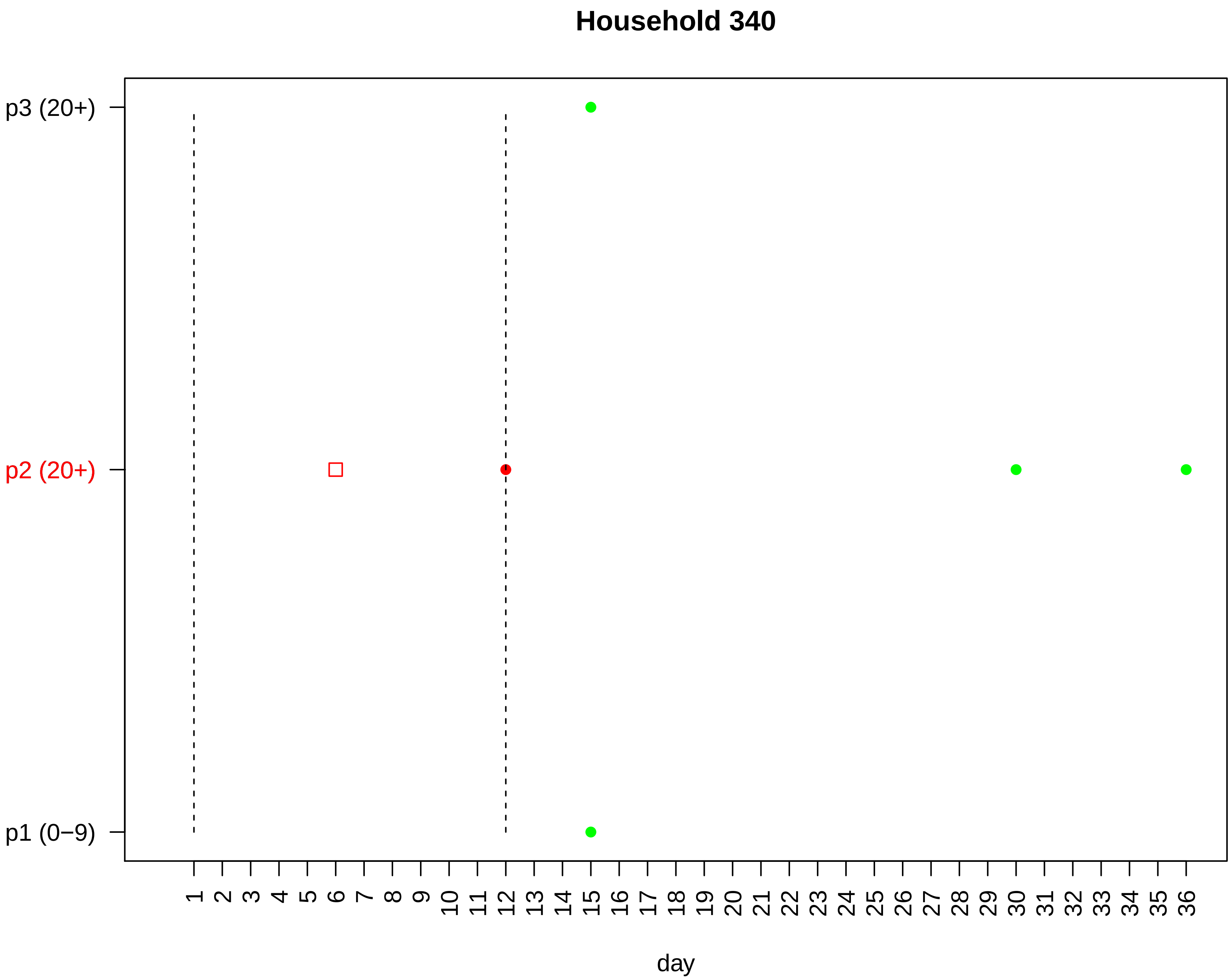

Household 340
p3 (20+)
p2 (20+)
p1 (0−9)
1 2 3 4 5 6 7 8 9 10 11 12 13 14 15 16 17 18 19 20 21 22 23 24 25 26 27 28 29 30 31 32 33 34 35 36
day

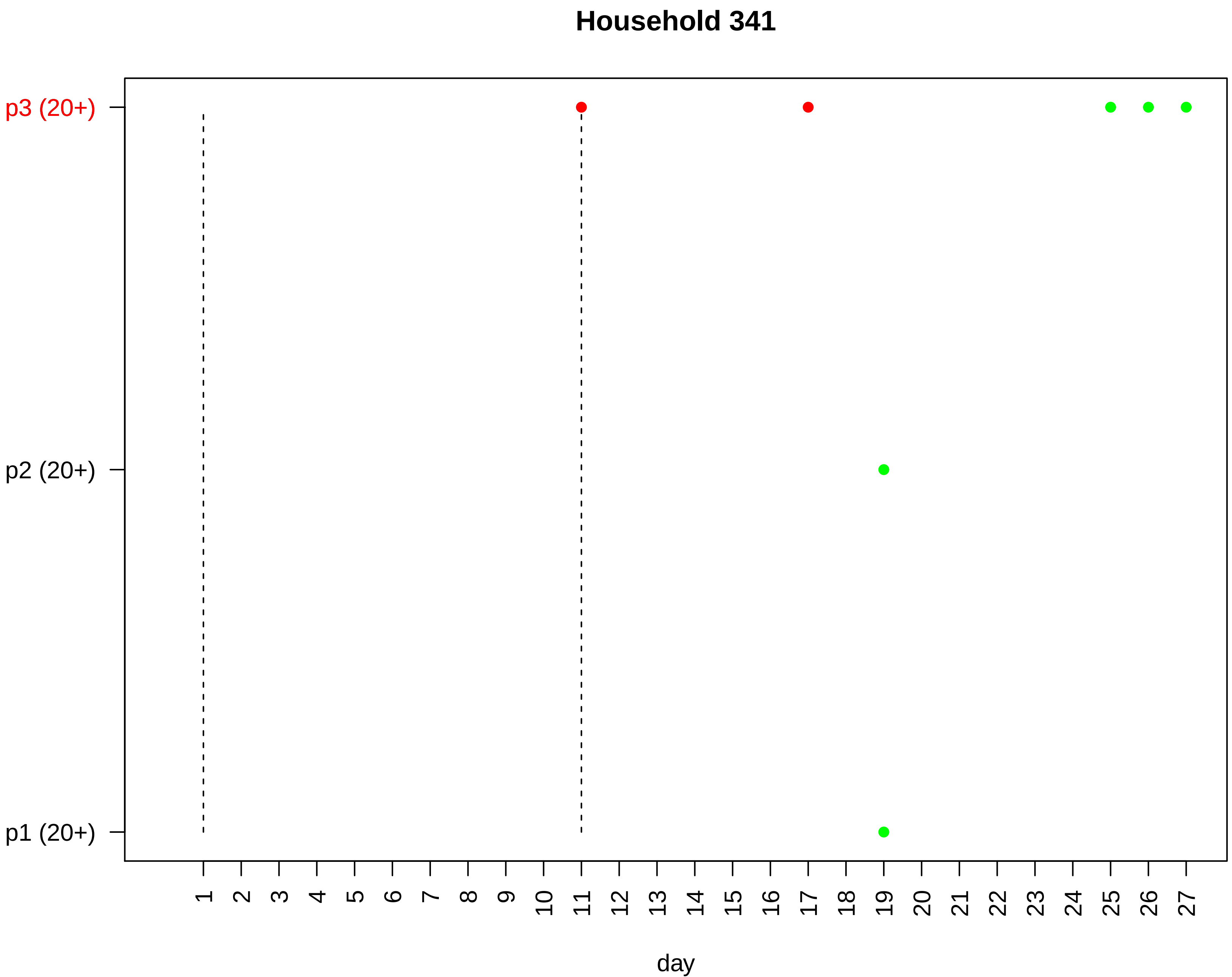
Household 341
p3 (20+)
p2 (20+)
p1 (20+)
day

### Household 342

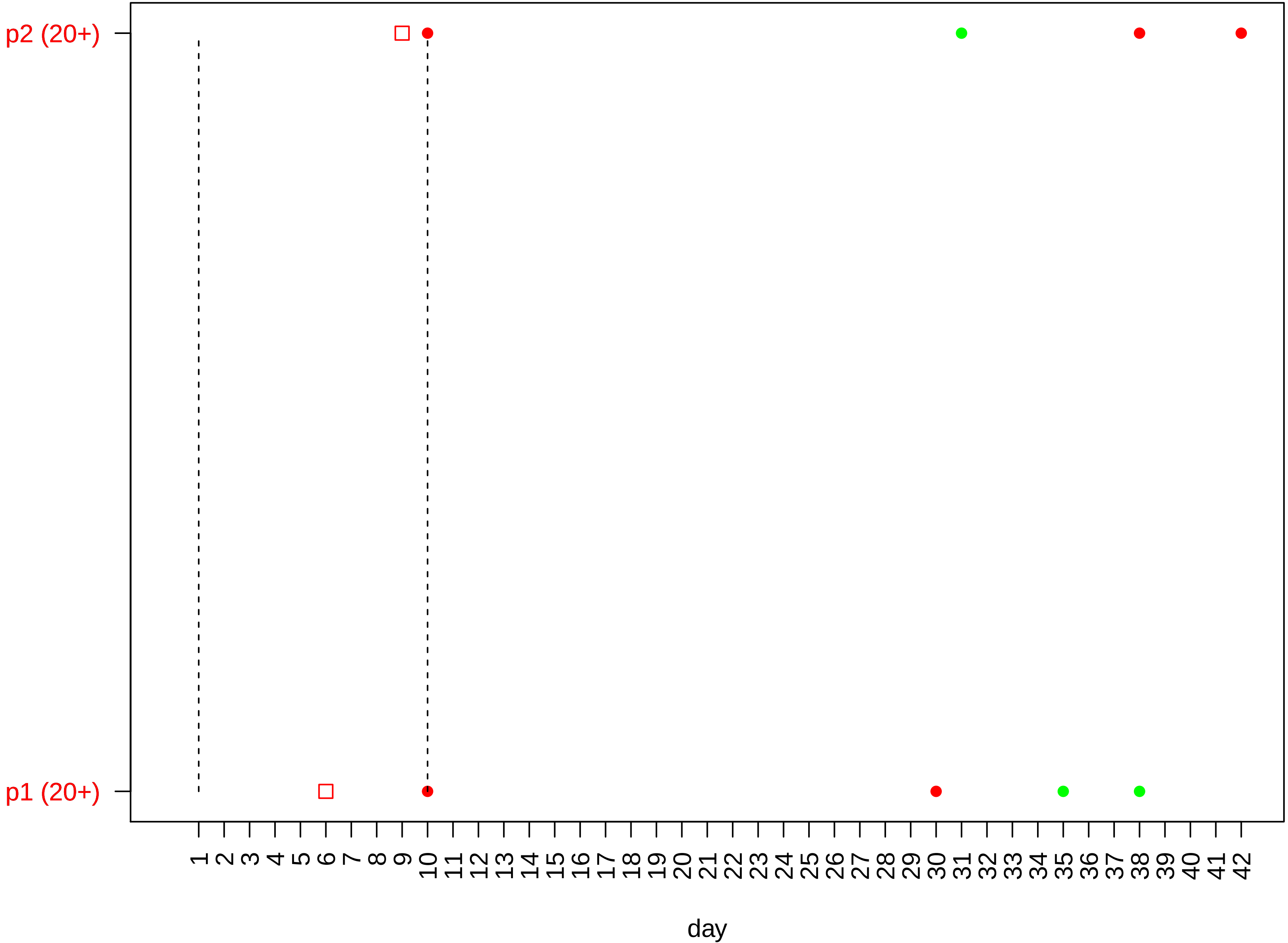

#### Household 343

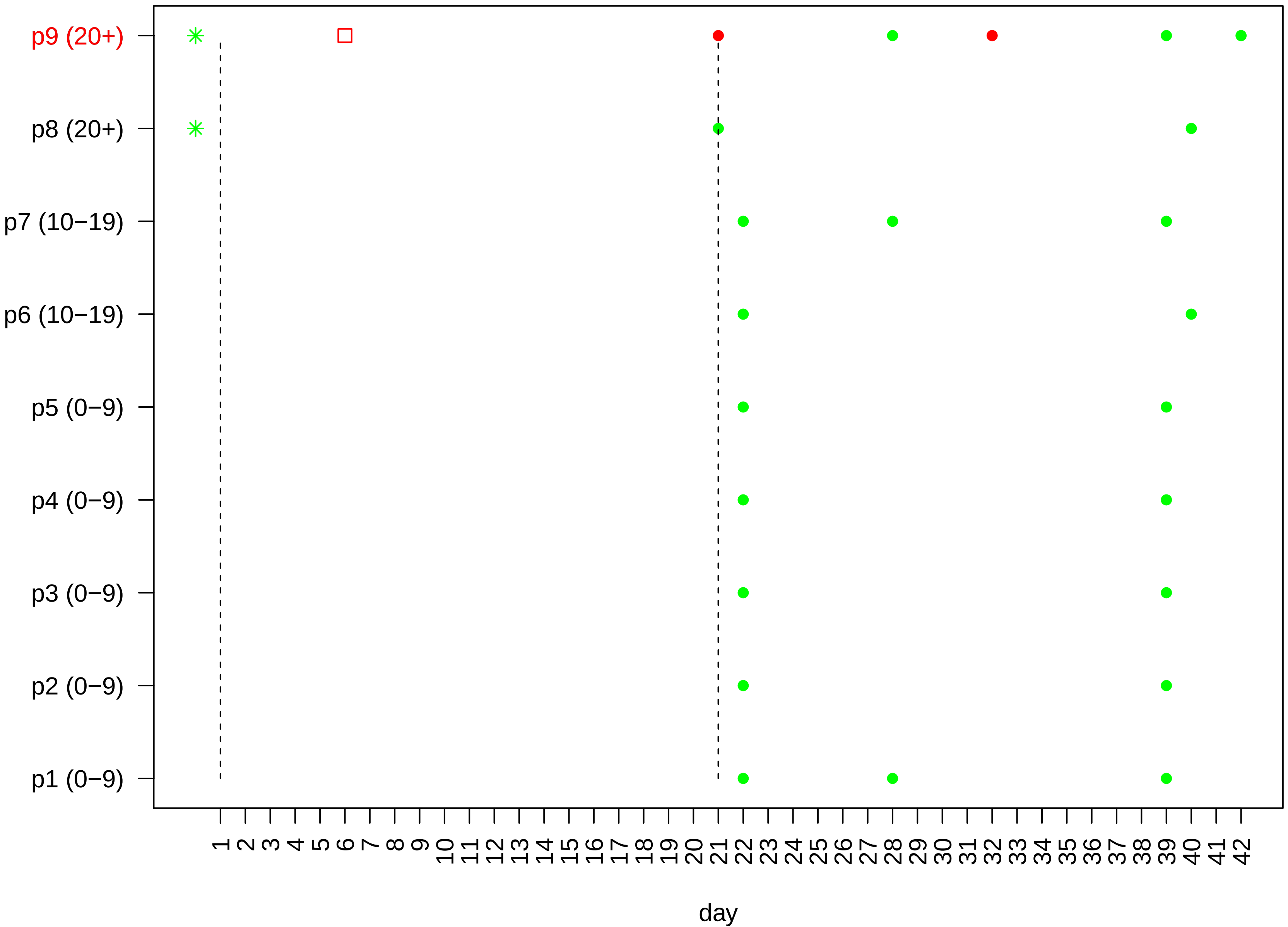

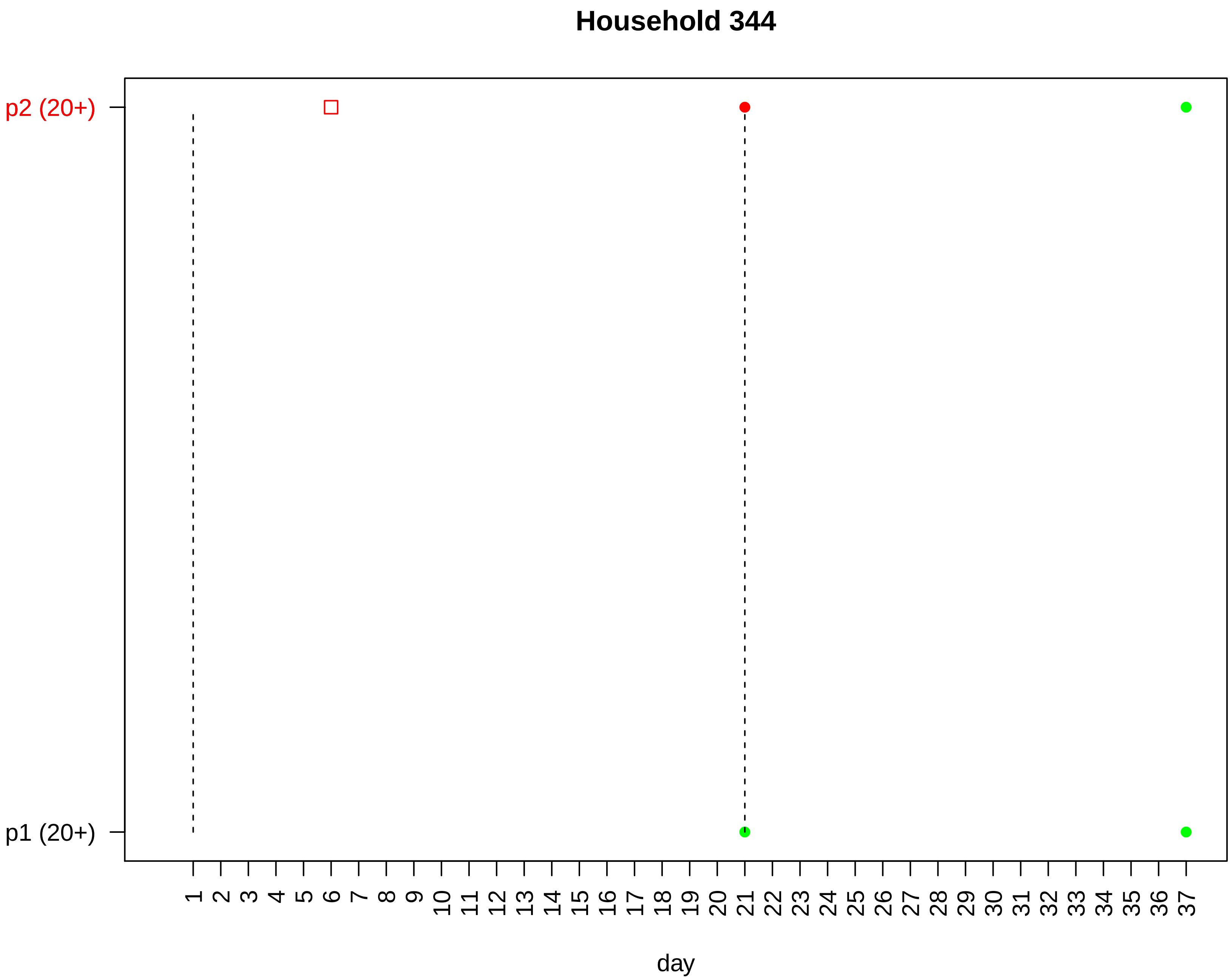

Household 344
p2 (20+)
p1 (20+)
day

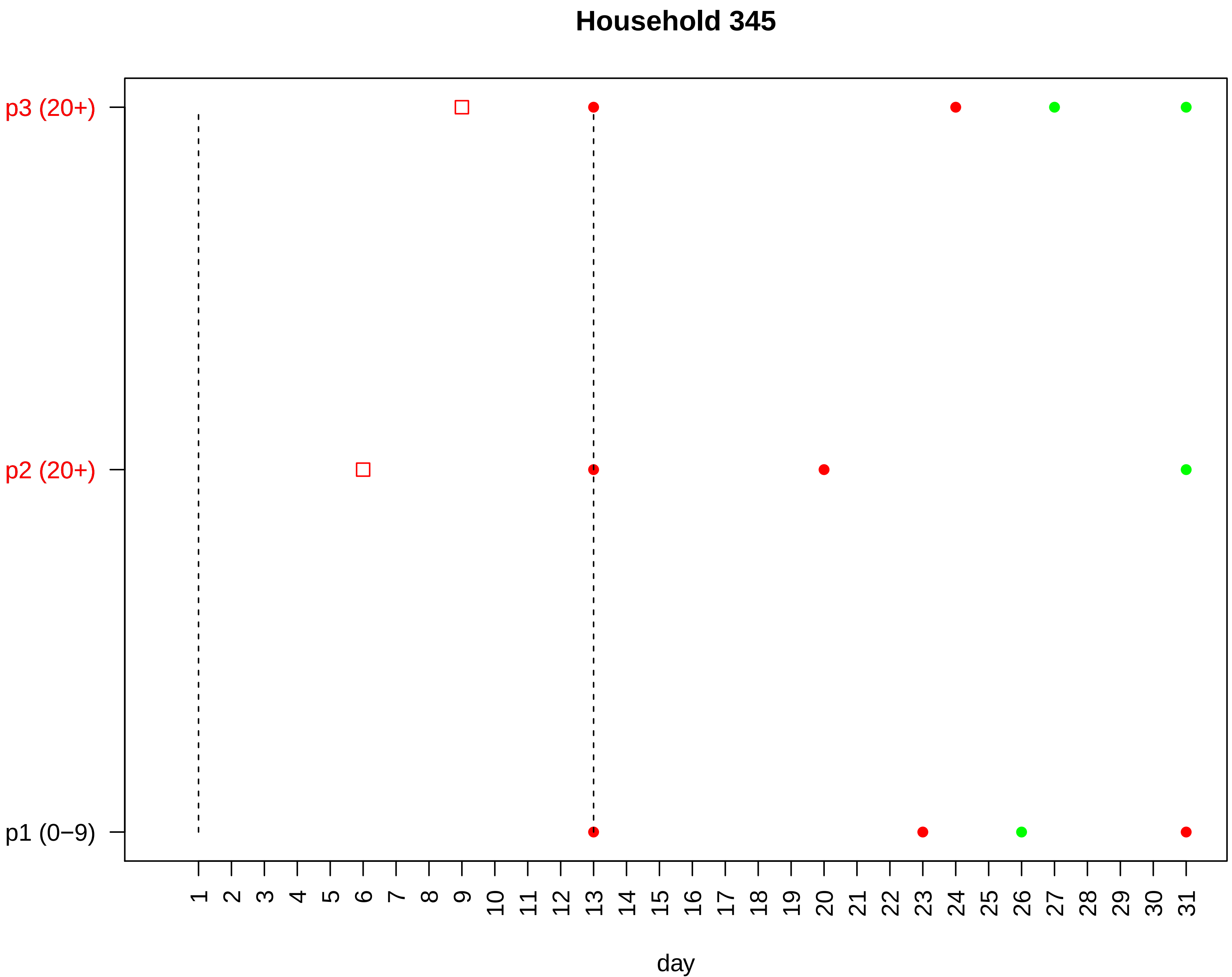

Household 345
p3 (20+)
p2 (20+)
p1 (0−9)
1 2 3 4 5 6 7 8 9 10 11 12 13 14 15 16 17 18 19 20 21 22 23 24 25 26 27 28 29 30 31
day

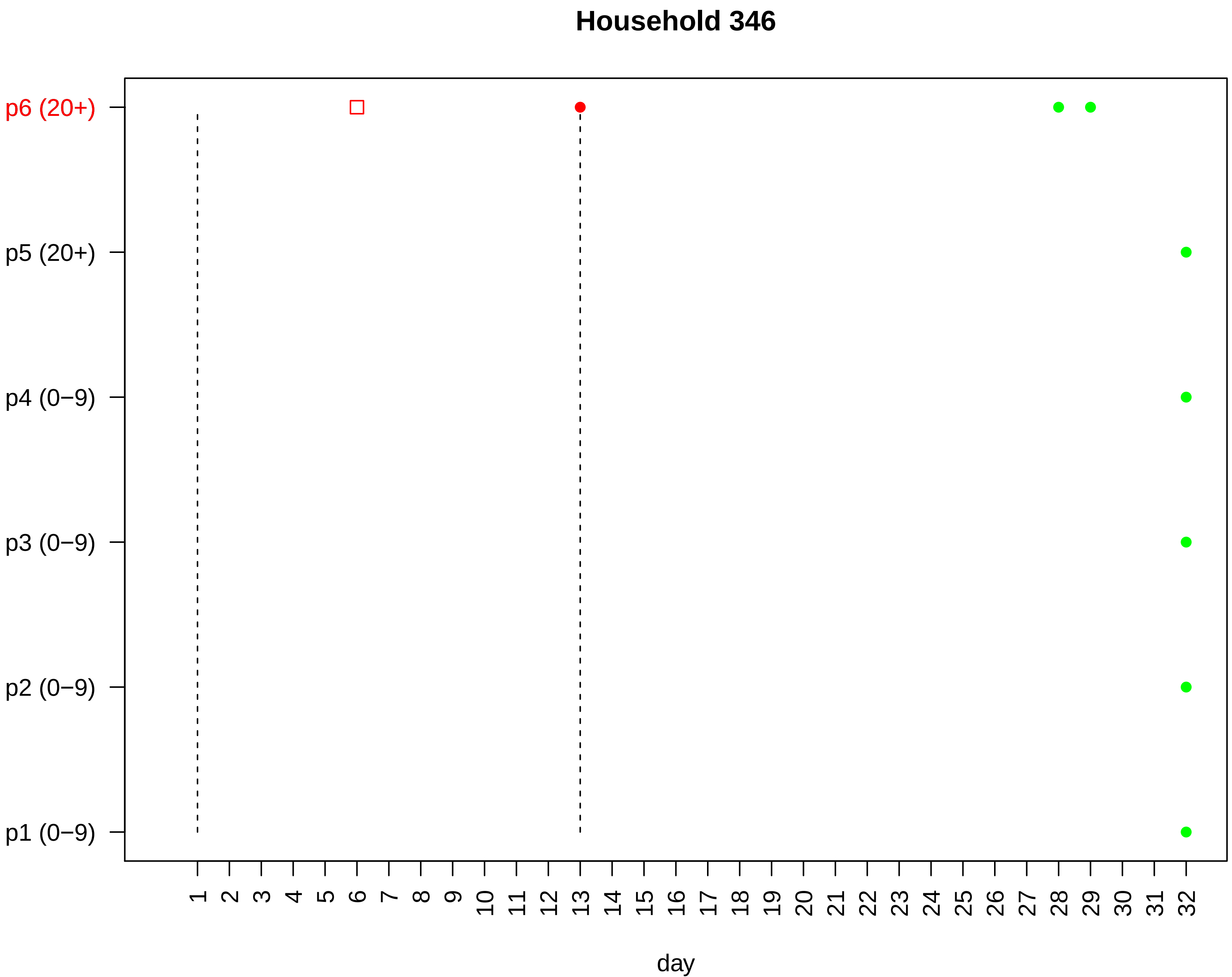
Household 346
p6 (20+)
p5 (20+)
p4 (0−9)
p3 (0−9)
p2 (0−9)
p1 (0−9)
1 2 3 4 5 6 7 8 9 10 11 12 13 14 15 16 17 18 19 20 21 22 23 24 25 26 27 28 29 30 31 32
day

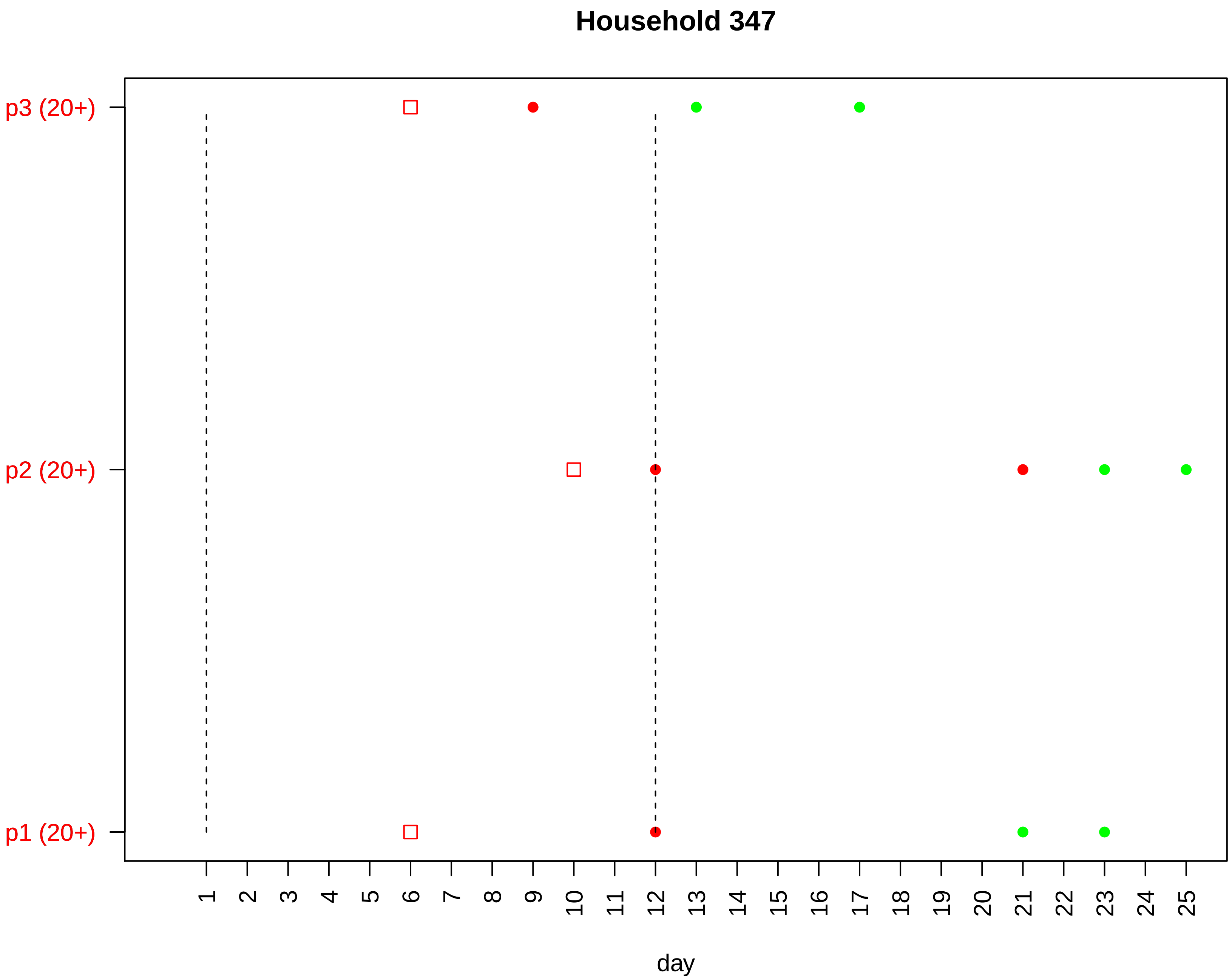

Household 347
p3 (20+)
p2 (20+)
p1 (20+)
day

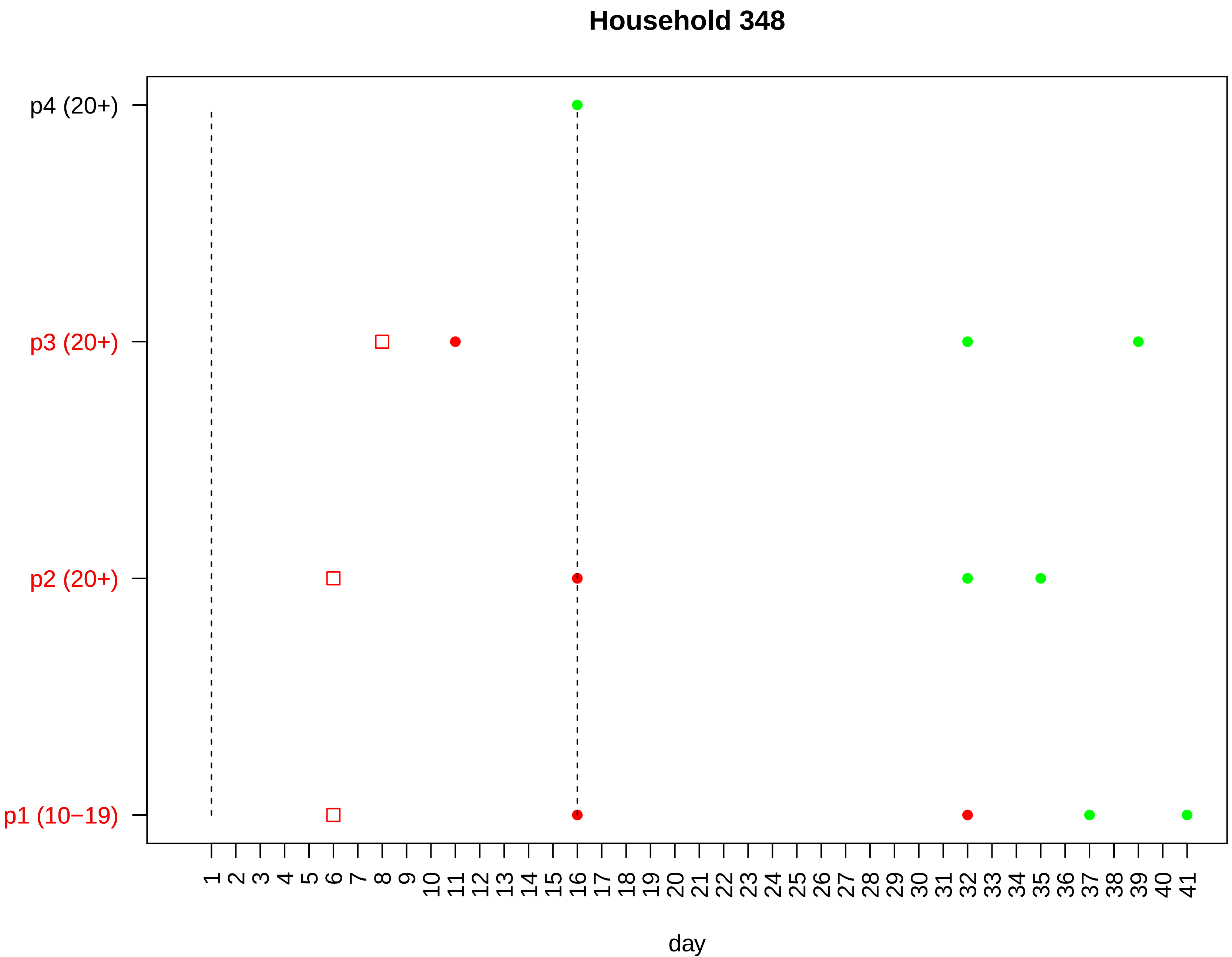
Household 348
p4 (20+)
p3 (20+)
p2 (20+)
p1 (10−19)
1 2 3 4 5 6 7 8 9 10 11 12 13 14 15 16 17 18 19 20 21 22 23 24 25 26 27 28 29 30 31 32 33 34 35 36 37 38 39 40 41
day

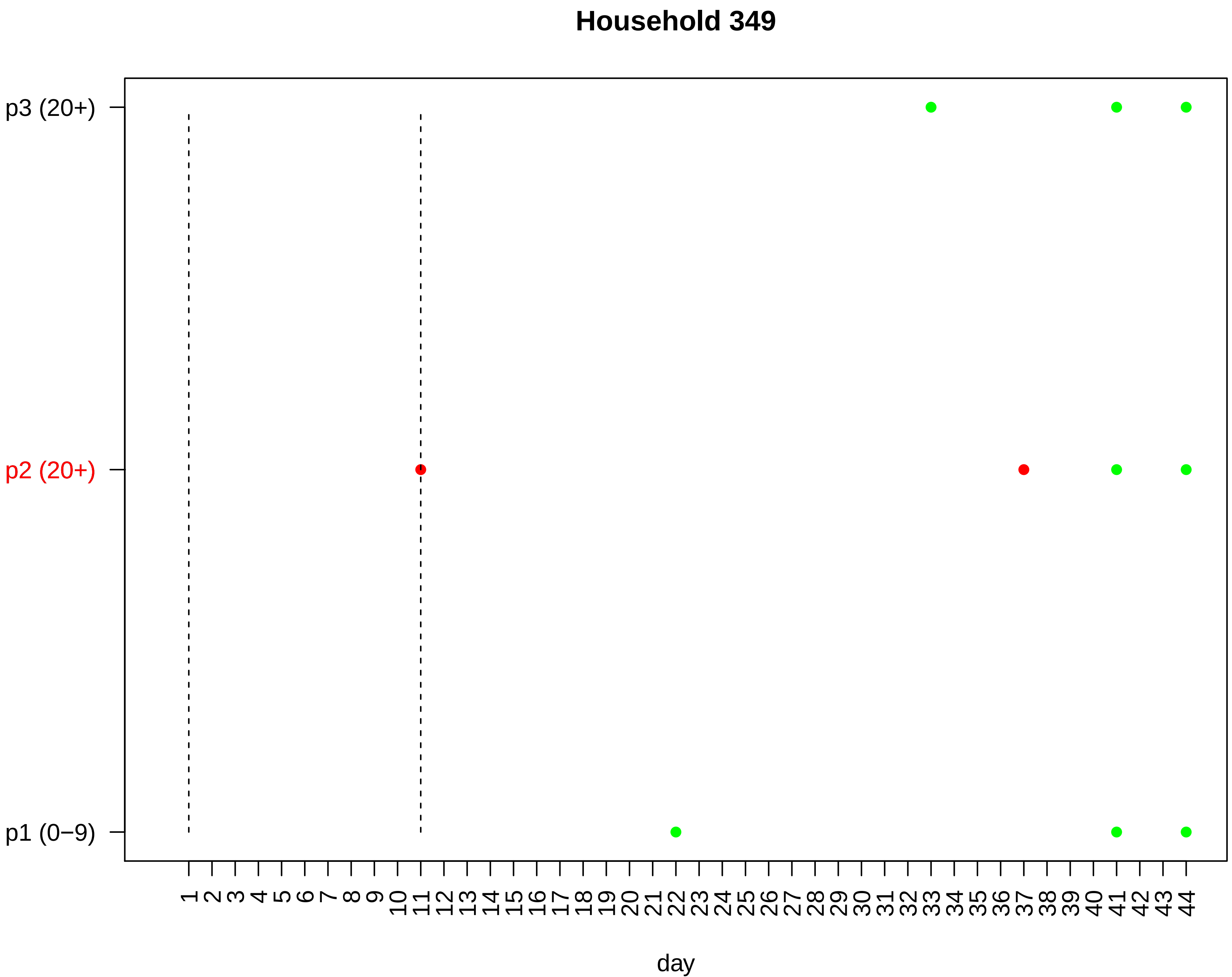

Household 349
p3 (20+)
p2 (20+)
p1 (0−9)
1 2 3 4 5 6 7 8 9 10 11 12 13 14 15 16 17 18 19 20 21 22 23 24 25 26 27 28 29 30 31 32 33 34 35 36 37 38 39 40 41 42 43 44
day

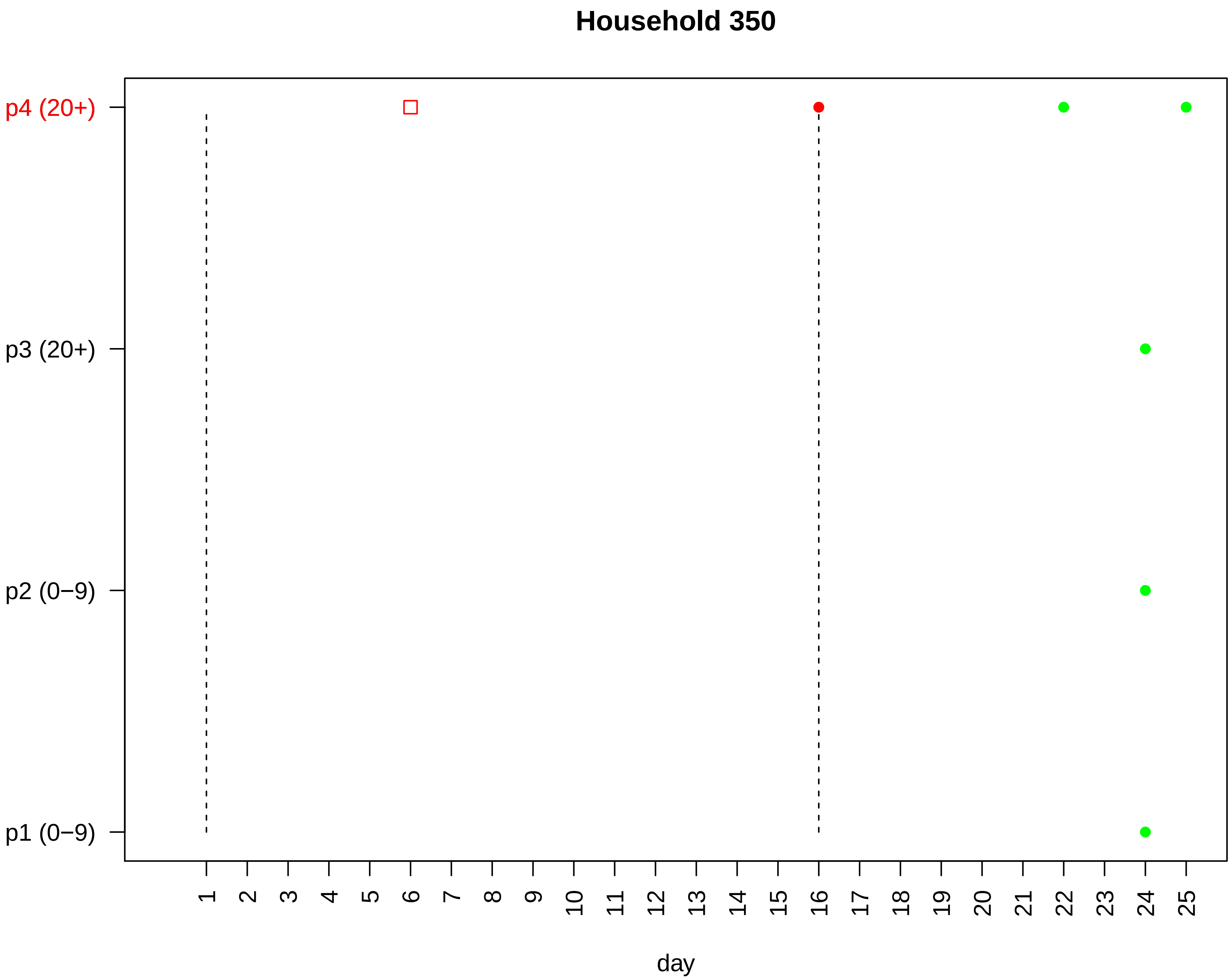

Household 350
p4 (20+)
p3 (20+)
p2 (0−9)
p1 (0−9)
day

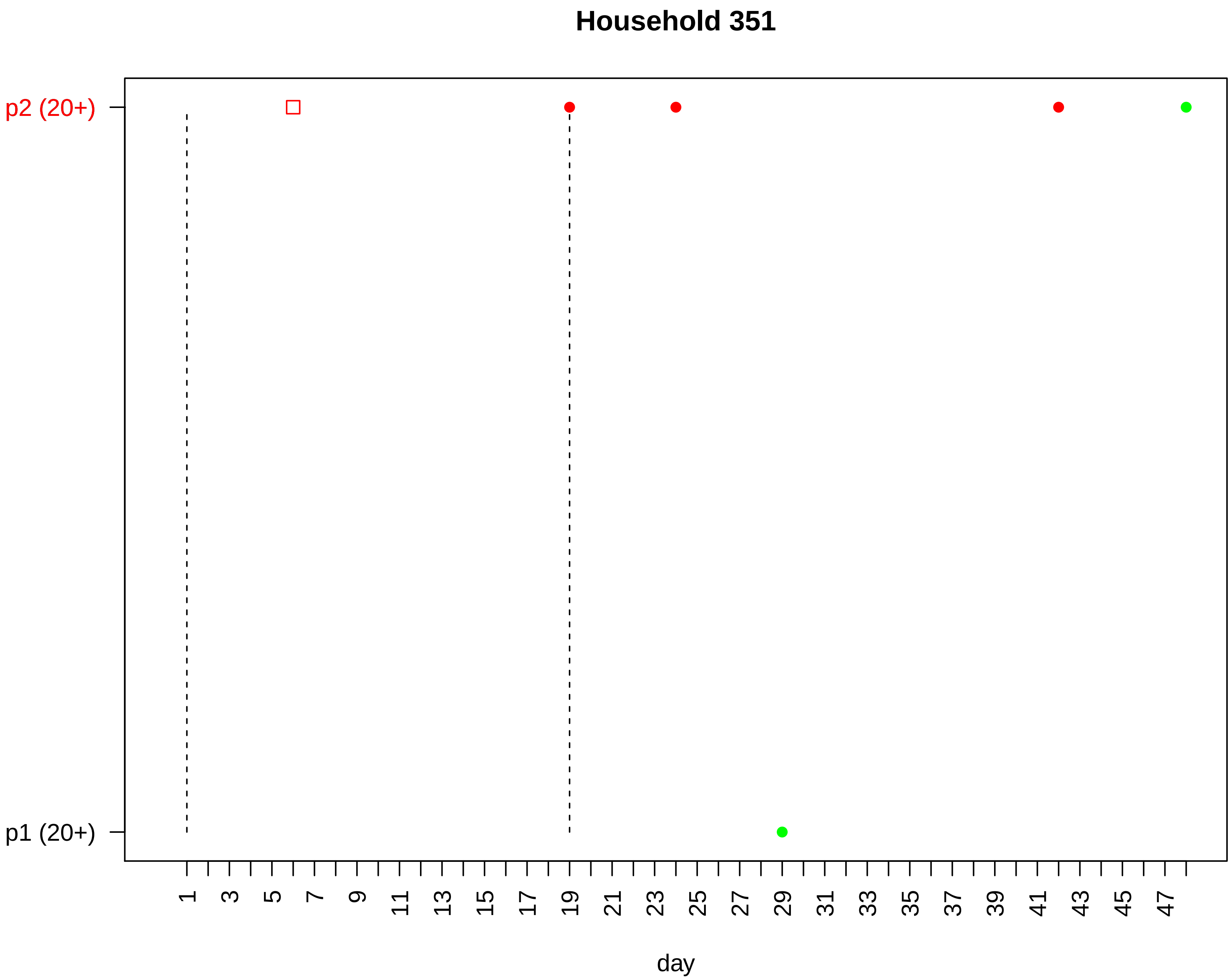

Household 351
p2 (20+)
p1 (20+)
1
3
5
7
9
11
13
15
17
19
21
23
25
27
29
31
33
35
37
39
41
43
45
47
day

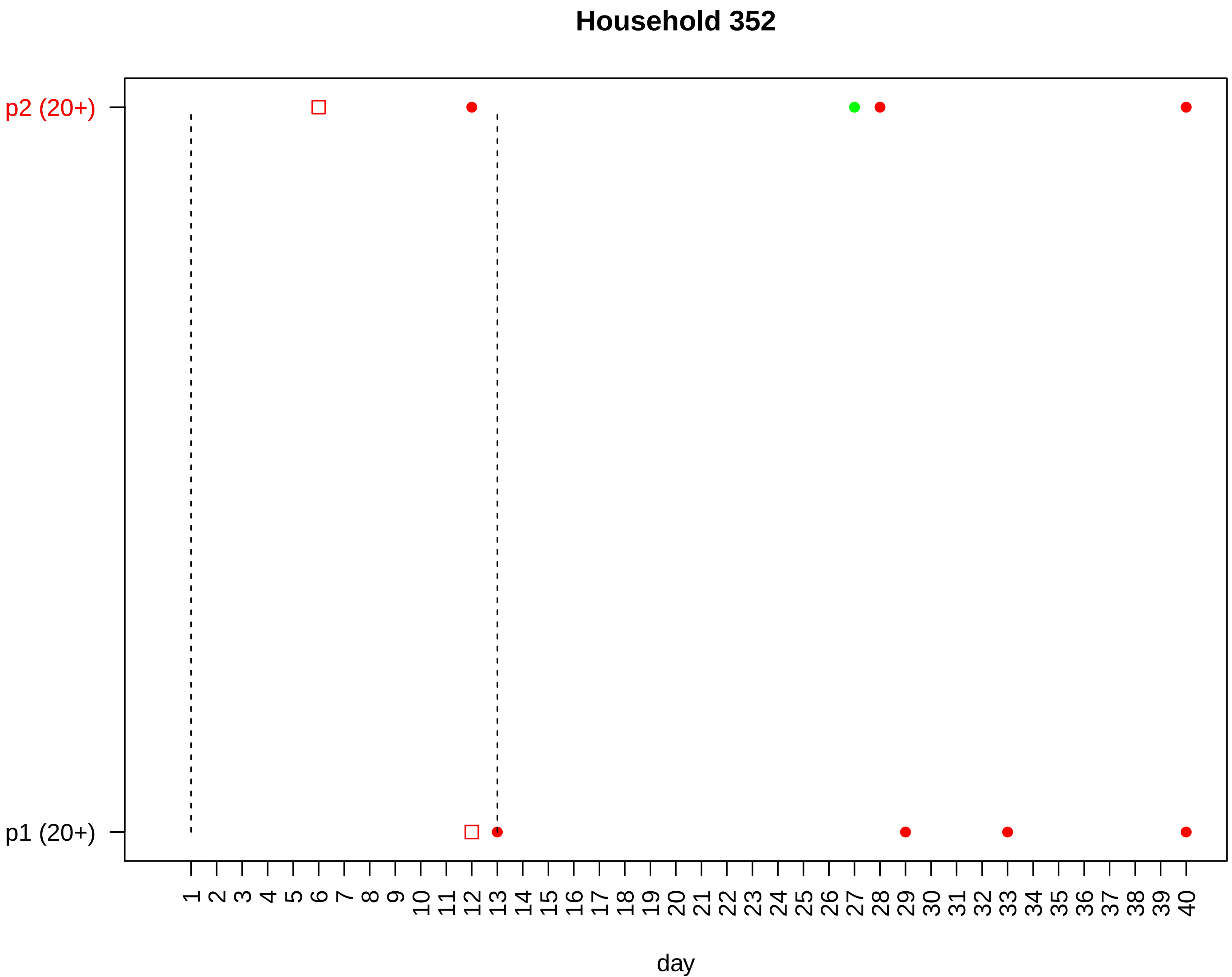

Household 352
p2 (20+)
p1 (20+)
1 2 3 4 5 6 7 8 9 10 11 12 13 14 15 16 17 18 19 20 21 22 23 24 25 26 27 28 29 30 31 32 33 34 35 36 37 38 39 40
day

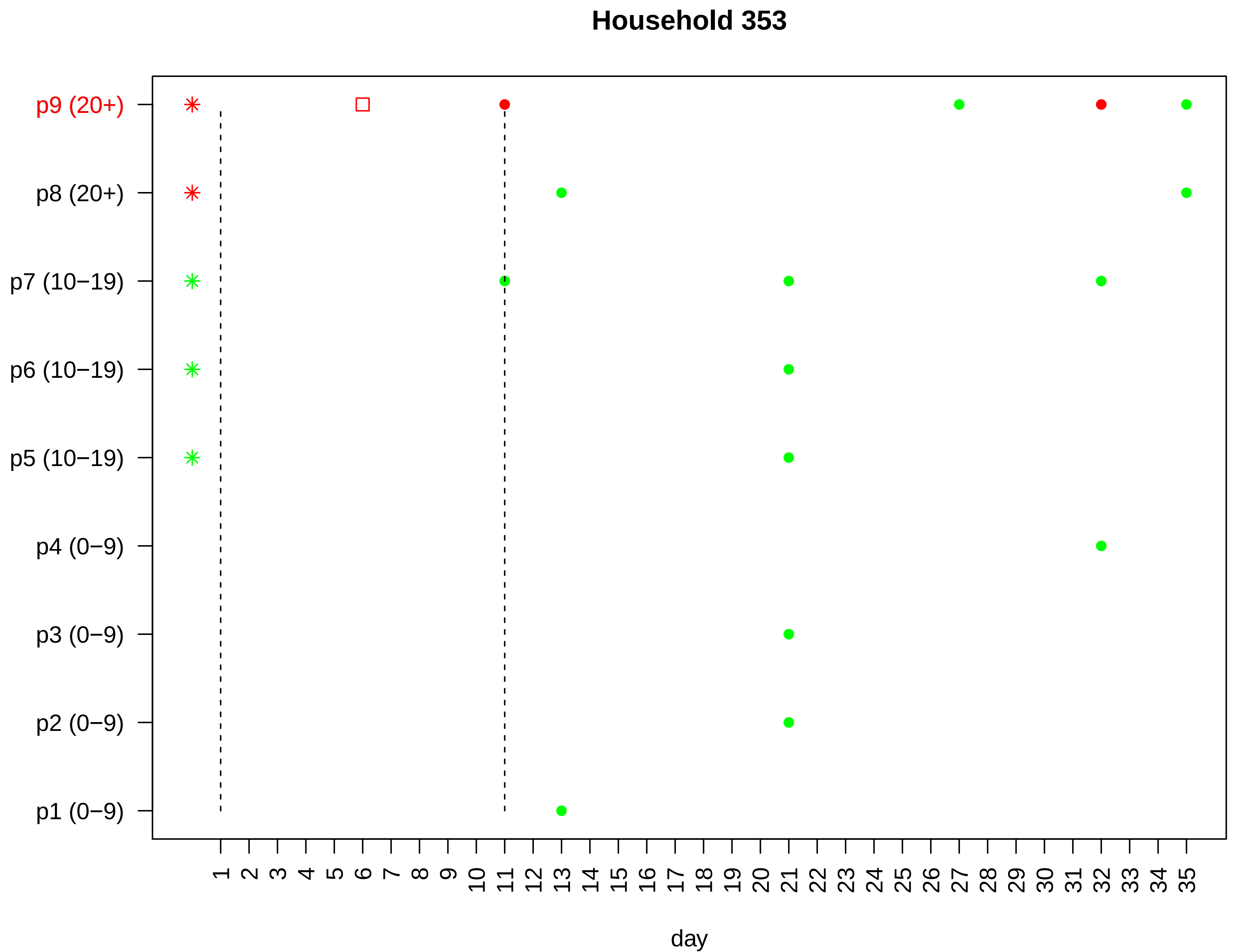

Household 353
p9 (20+)
p8 (20+)
p7 (10−19)
p6 (10−19)
p5 (10−19)
p4 (0−9)
p3 (0−9)
p2 (0−9)
p1 (0−9)
1 2 3 4 5 6 7 8 9 10 11 12 13 14 15 16 17 18 19 20 21 22 23 24 25 26 27 28 29 30 31 32 33 34 35
day

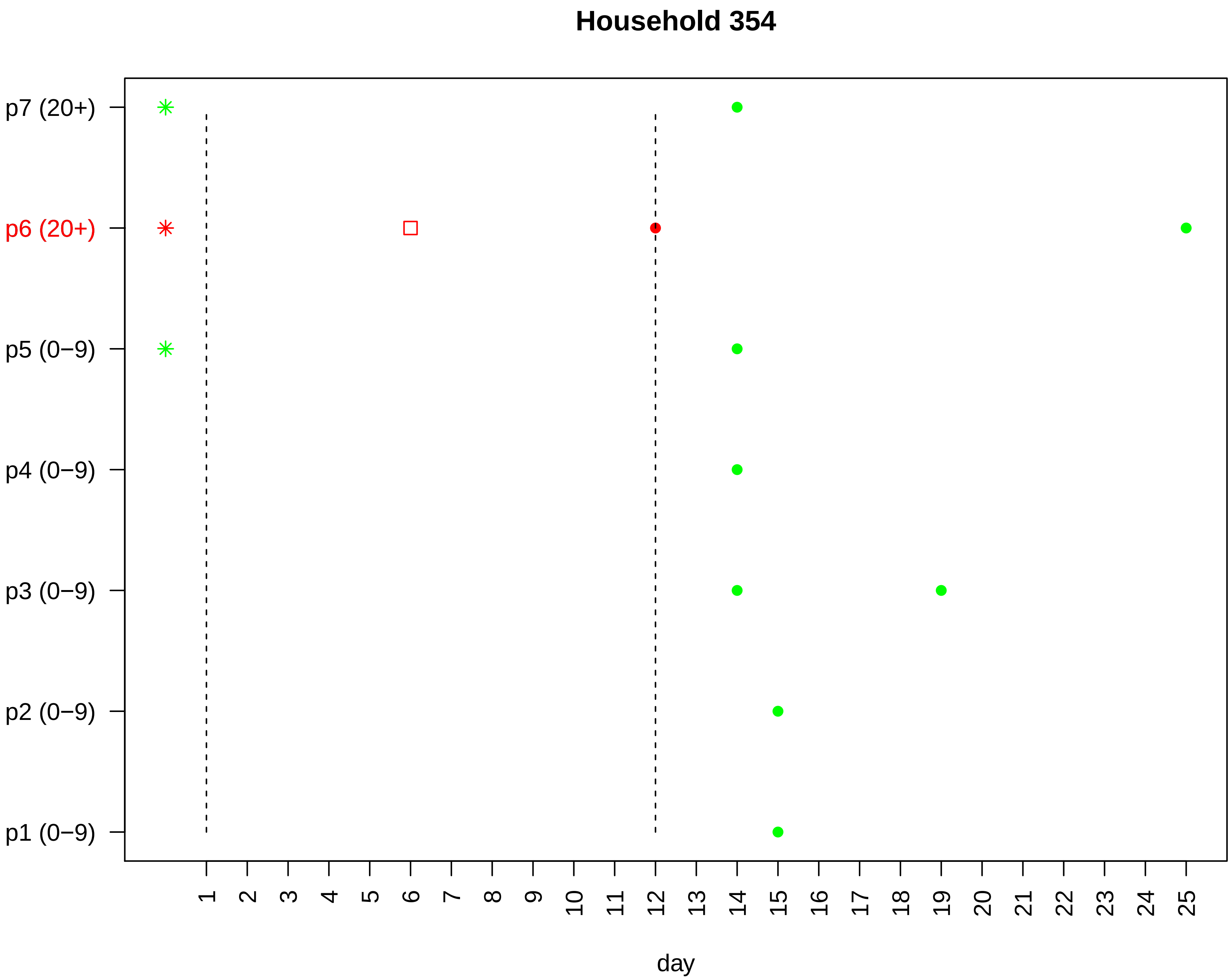

Household 354
p7 (20+)
p6 (20+)
p5 (0−9)
p4 (0−9)
p3 (0−9)
p2 (0−9)
p1 (0−9)
1 2 3 4 5 6 7 8 9 10 11 12 13 14 15 16 17 18 19 20 21 22 23 24 25
day

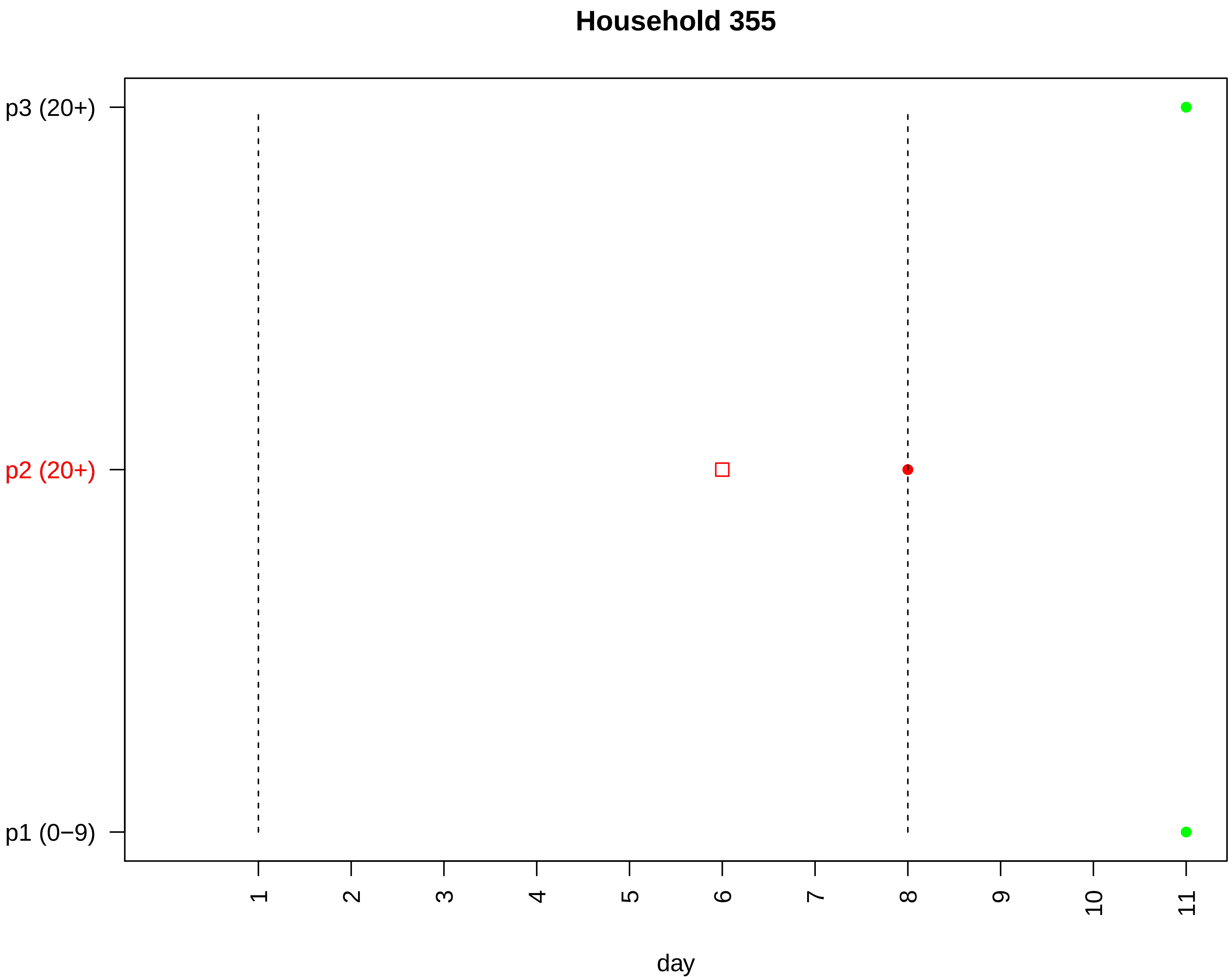

Household 355
p3 (20+)
p2 (20+)
p1 (0−9)
1
2
3
4
5
6
7
8
9
10
11
day

#### Household 356

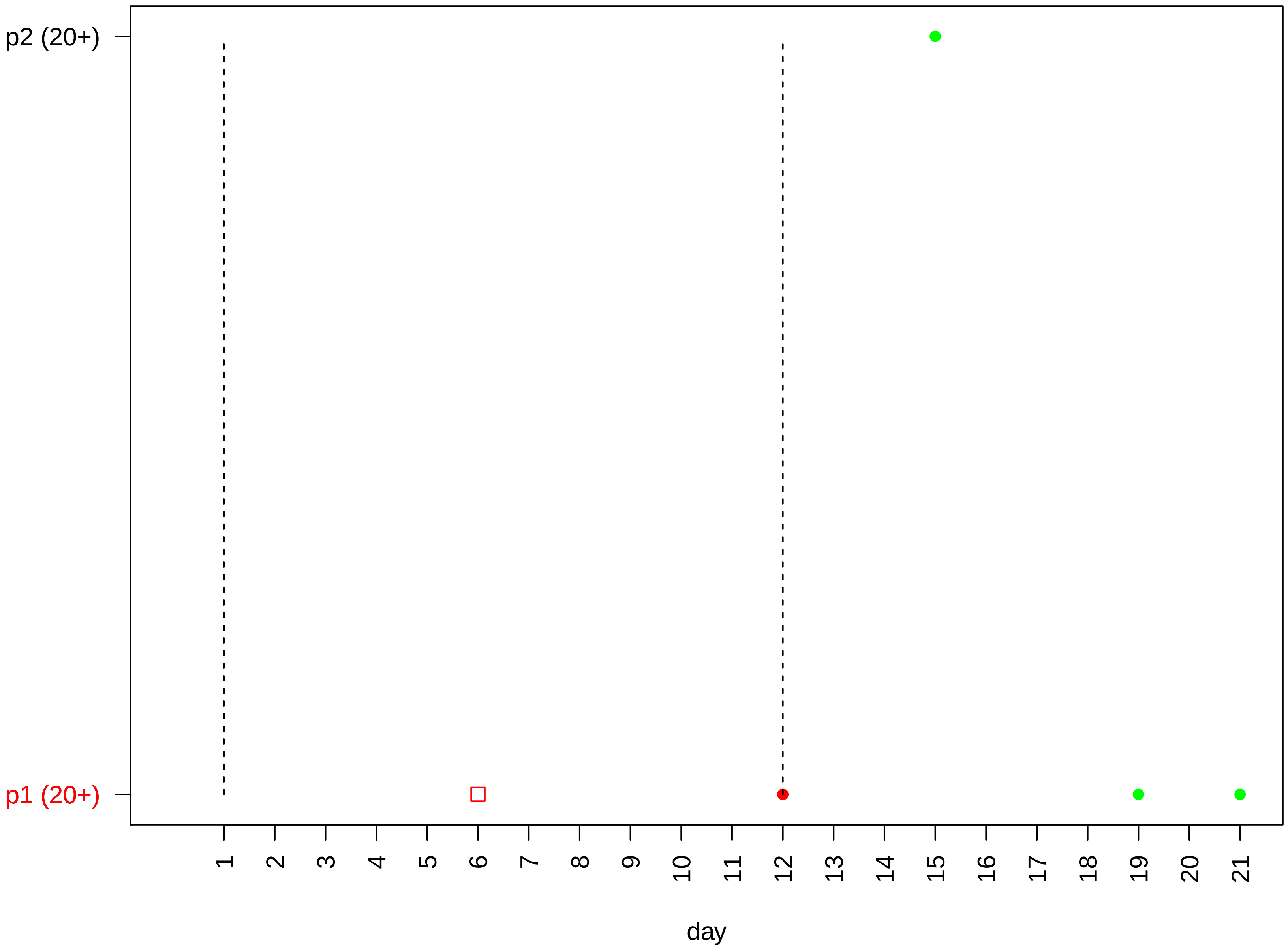

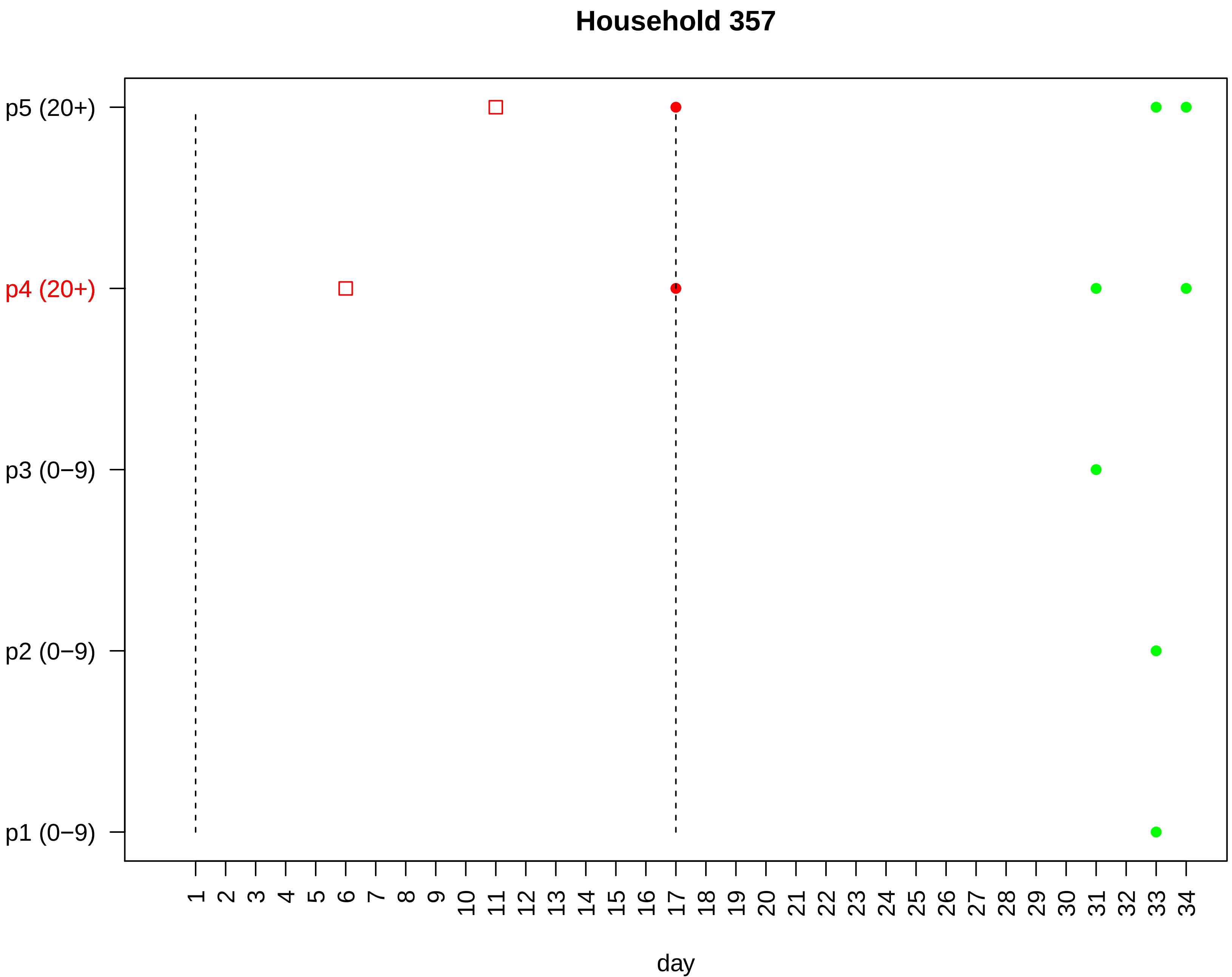

Household 357
p5 (20+)
p4 (20+)
p3 (0−9)
p2 (0−9)
p1 (0−9)
day

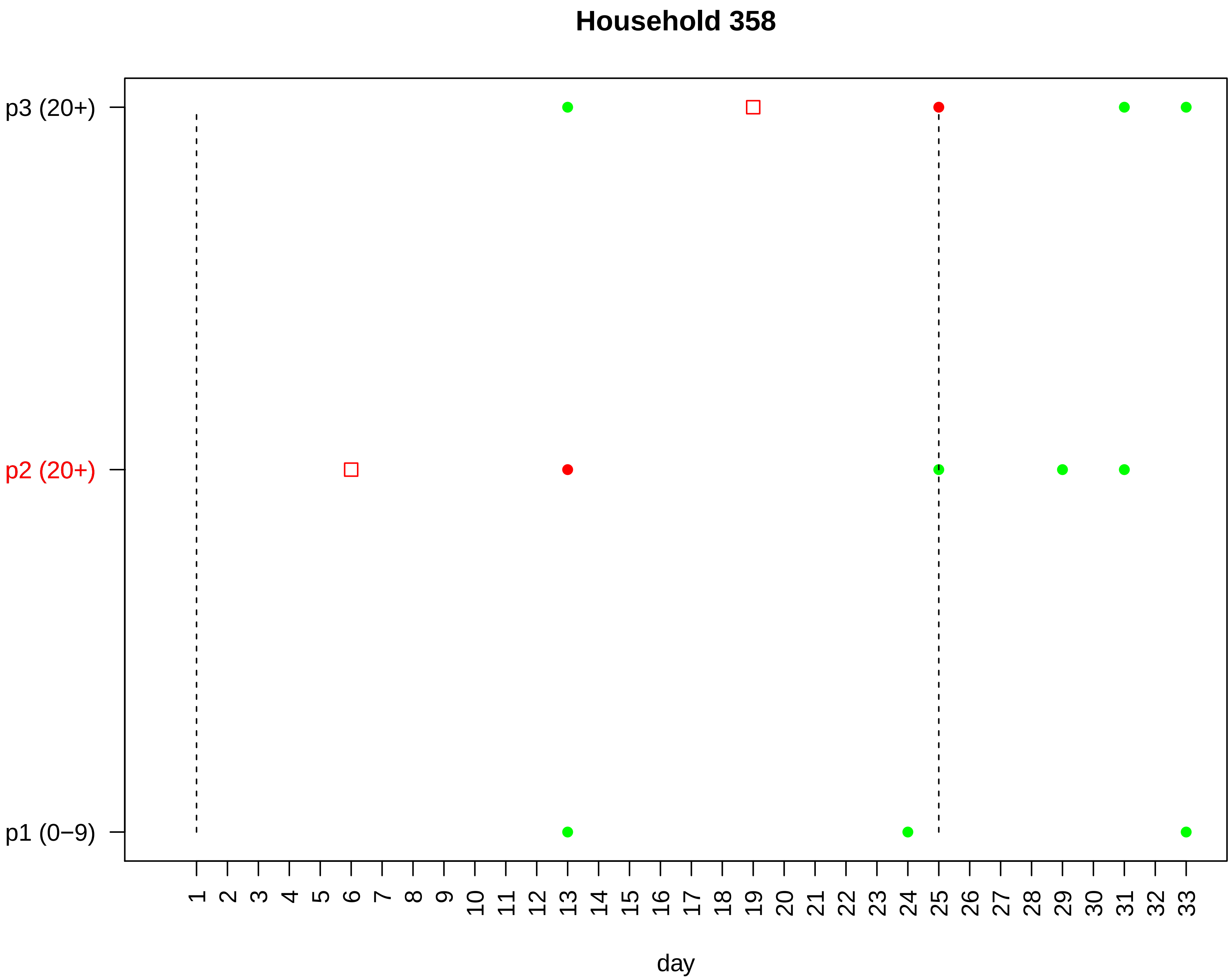
Household 358
p3 (20+)
p2 (20+)
p1 (0−9)
1 2 3 4 5 6 7 8 9 10 11 12 13 14 15 16 17 18 19 20 21 22 23 24 25 26 27 28 29 30 31 32 33
day

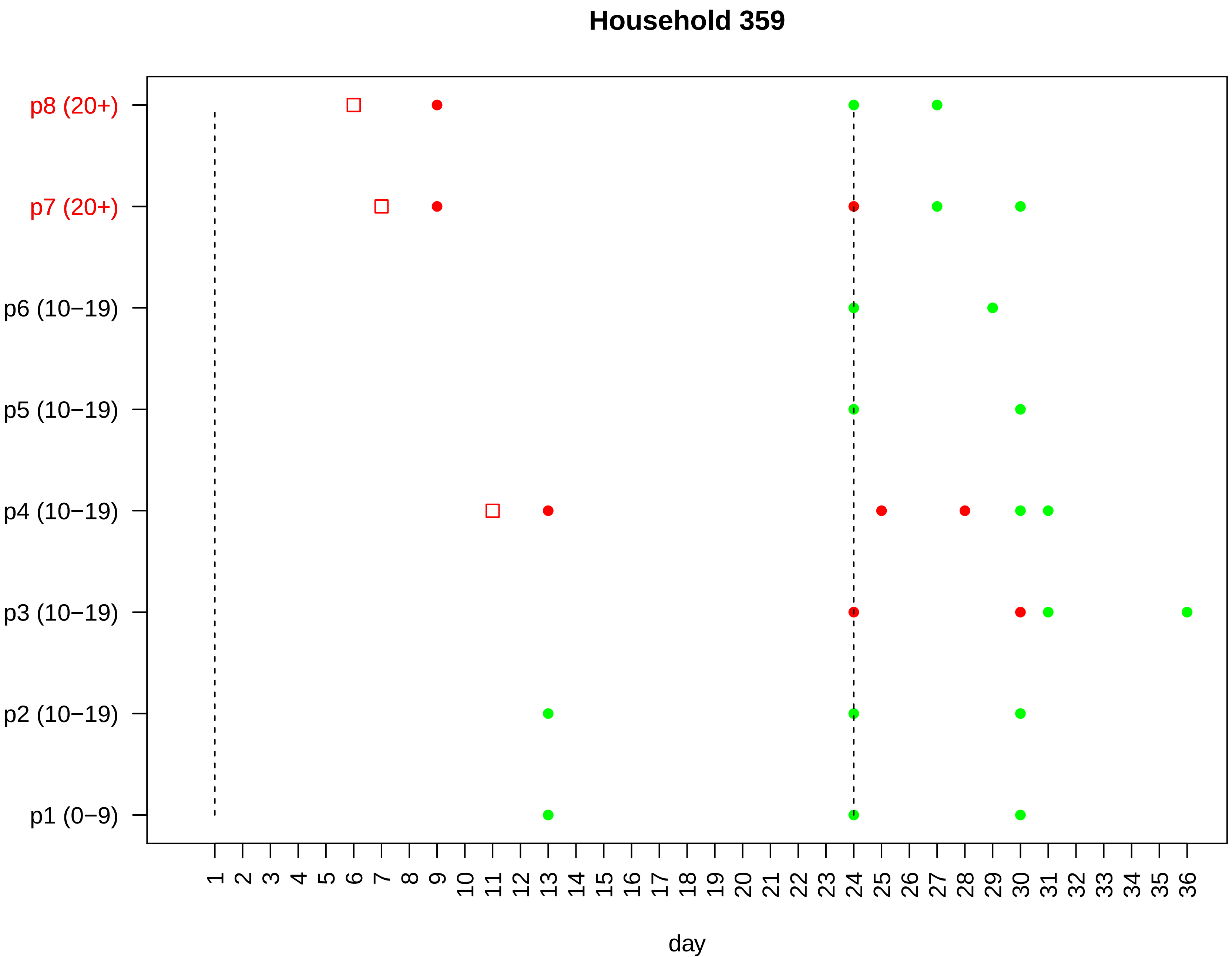

Household 359
p8 (20+)
p7 (20+)
p6 (10−19)
p5 (10−19)
p4 (10−19)
p3 (10−19)
p2 (10−19)
p1 (0−9)
1 2 3 4 5 6 7 8 9 10 11 12 13 14 15 16 17 18 19 20 21 22 23 24 25 26 27 28 29 30 31 32 33 34 35 36
day

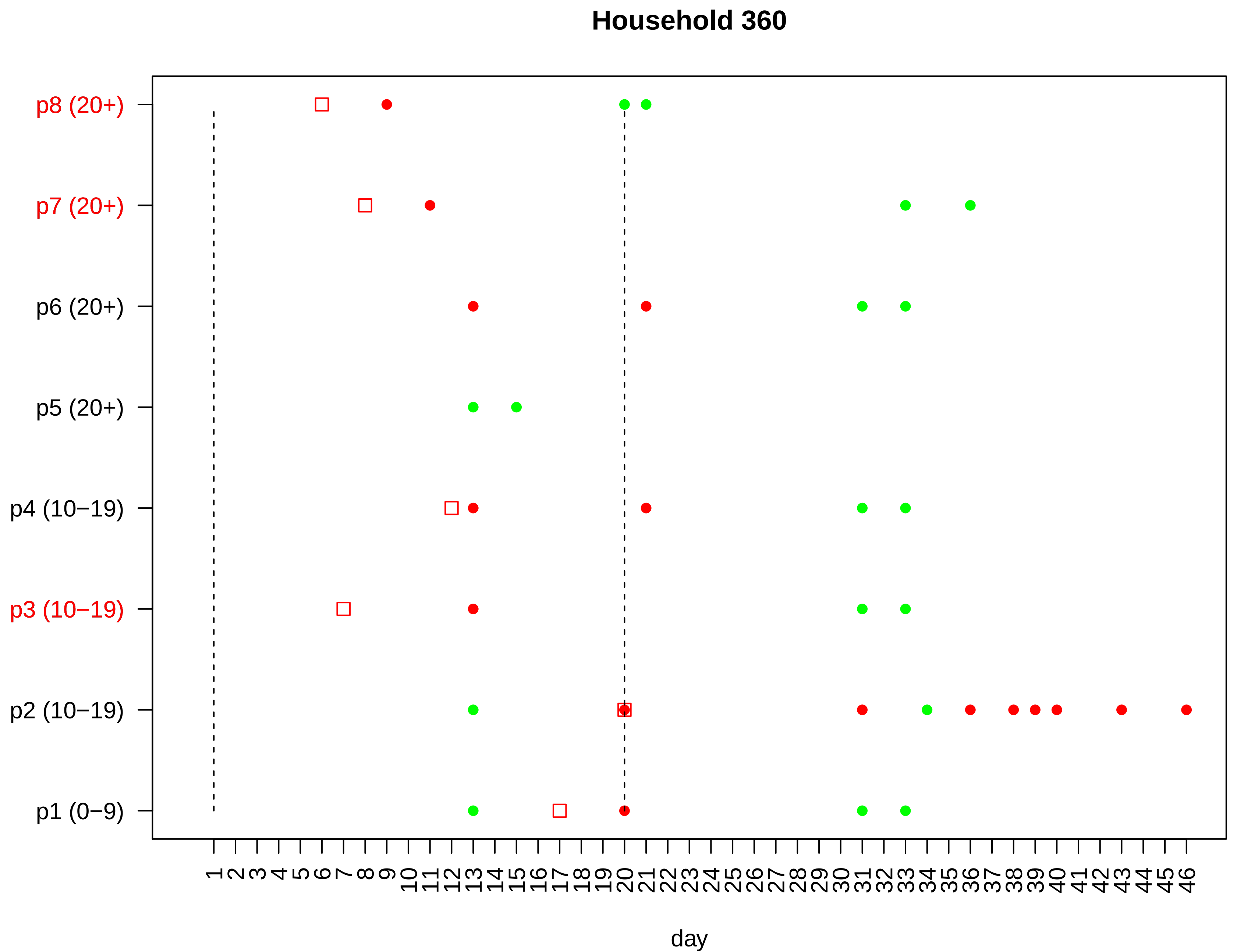

Household 360
p8 (20+)
p7 (20+)
p6 (20+)
p5 (20+)
p4 (10−19)
p3 (10−19)
p2 (10−19)
p1 (0−9)
1 2 3 4 5 6 7 8 9 10 11 12 13 14 15 16 17 18 19 20 21 22 23 24 25 26 27 28 29 30 31 32 33 34 35 36 37 38 39 40 41 42 43 44 45 46
day

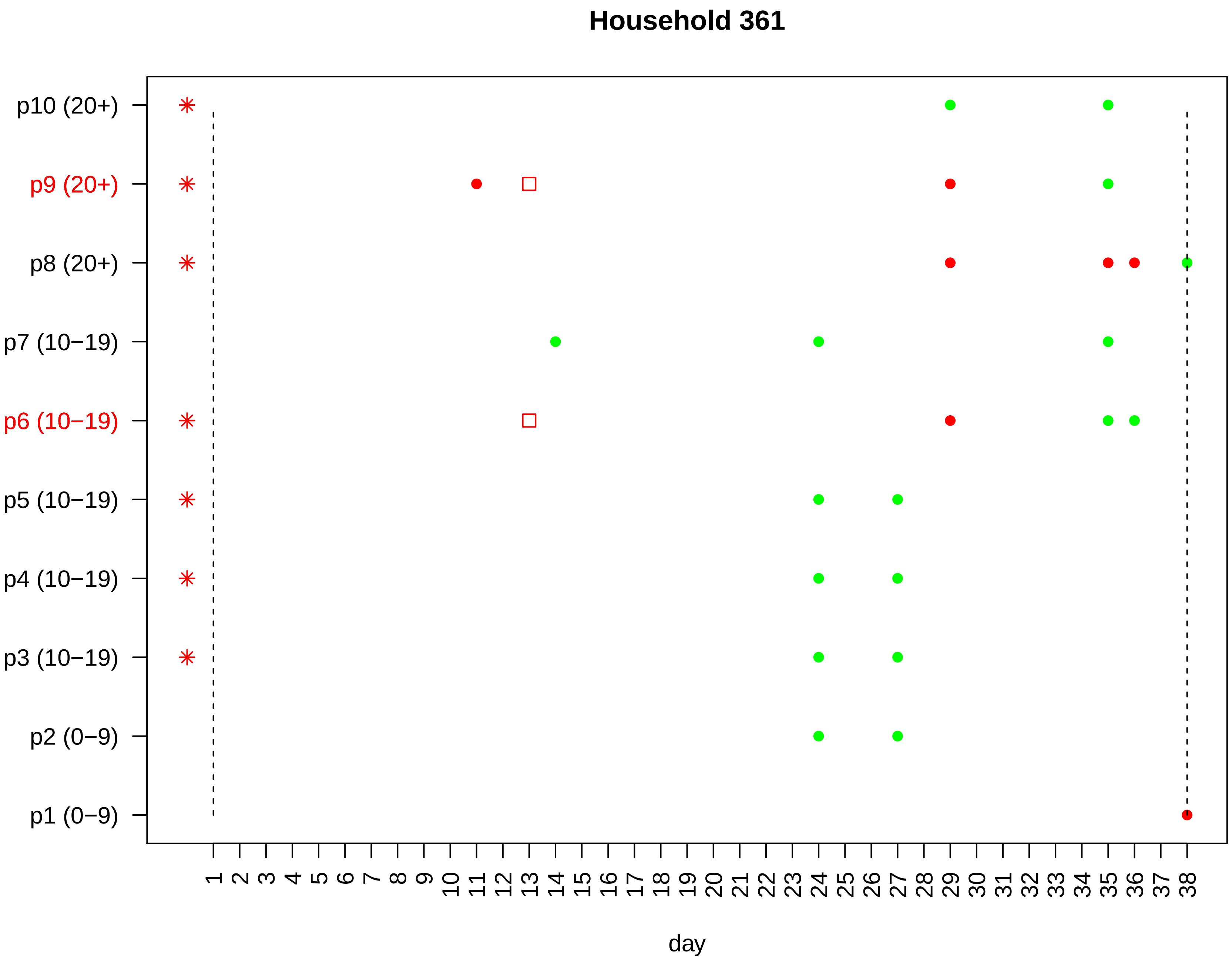
Household 361
p10 (20+)
p9 (20+)
p8 (20+)
p7 (10−19)
p6 (10−19)
p5 (10−19)
p4 (10−19)
p3 (10−19)
p2 (0−9)
p1 (0−9)
1 2 3 4 5 6 7 8 9 10 11 12 13 14 15 16 17 18 19 20 21 22 23 24 25 26 27 28 29 30 31 32 33 34 35 36 37 38
day

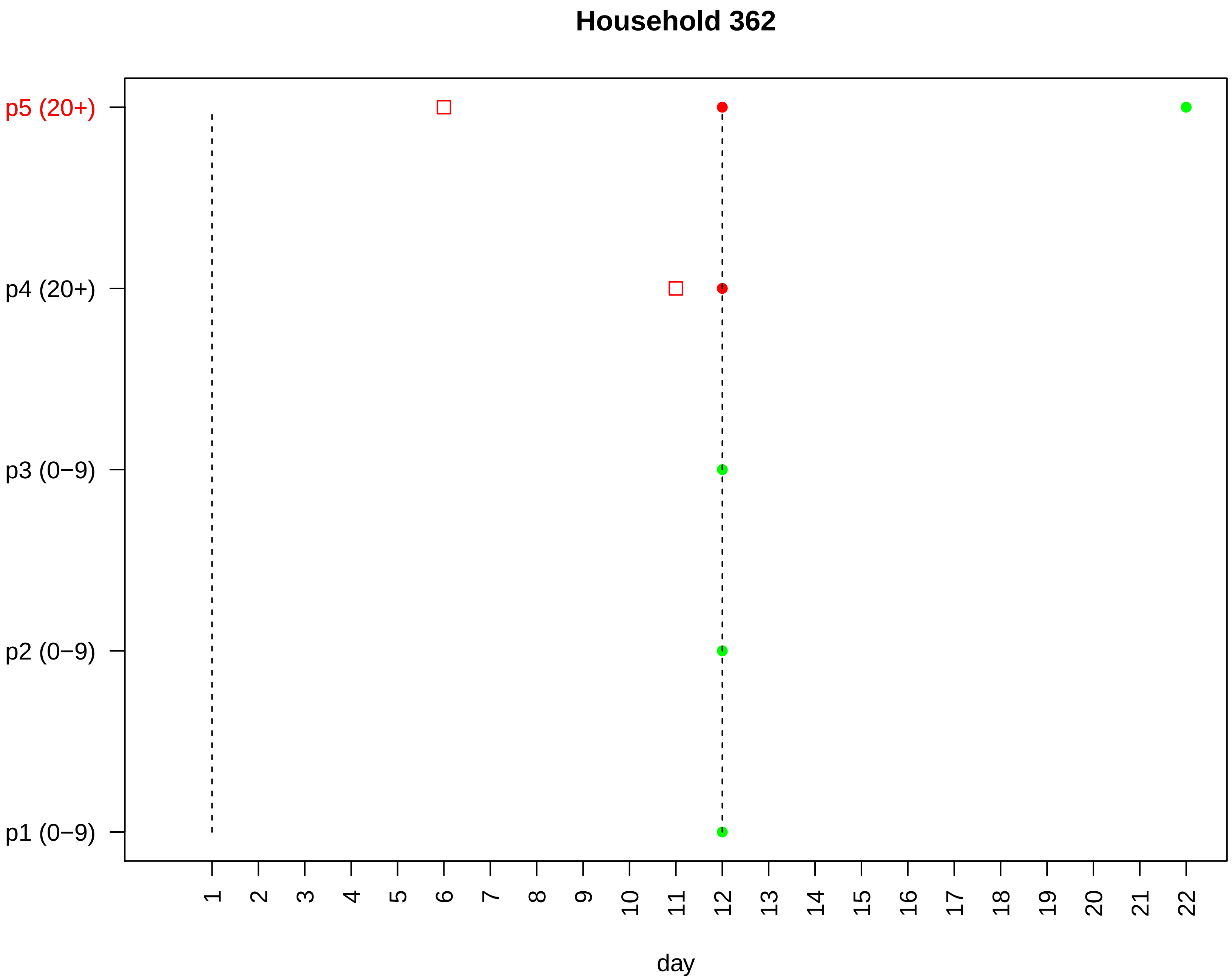

Household 362
p5 (20+)
p4 (20+)
p3 (0−9)
p2 (0−9)
p1 (0−9)
day

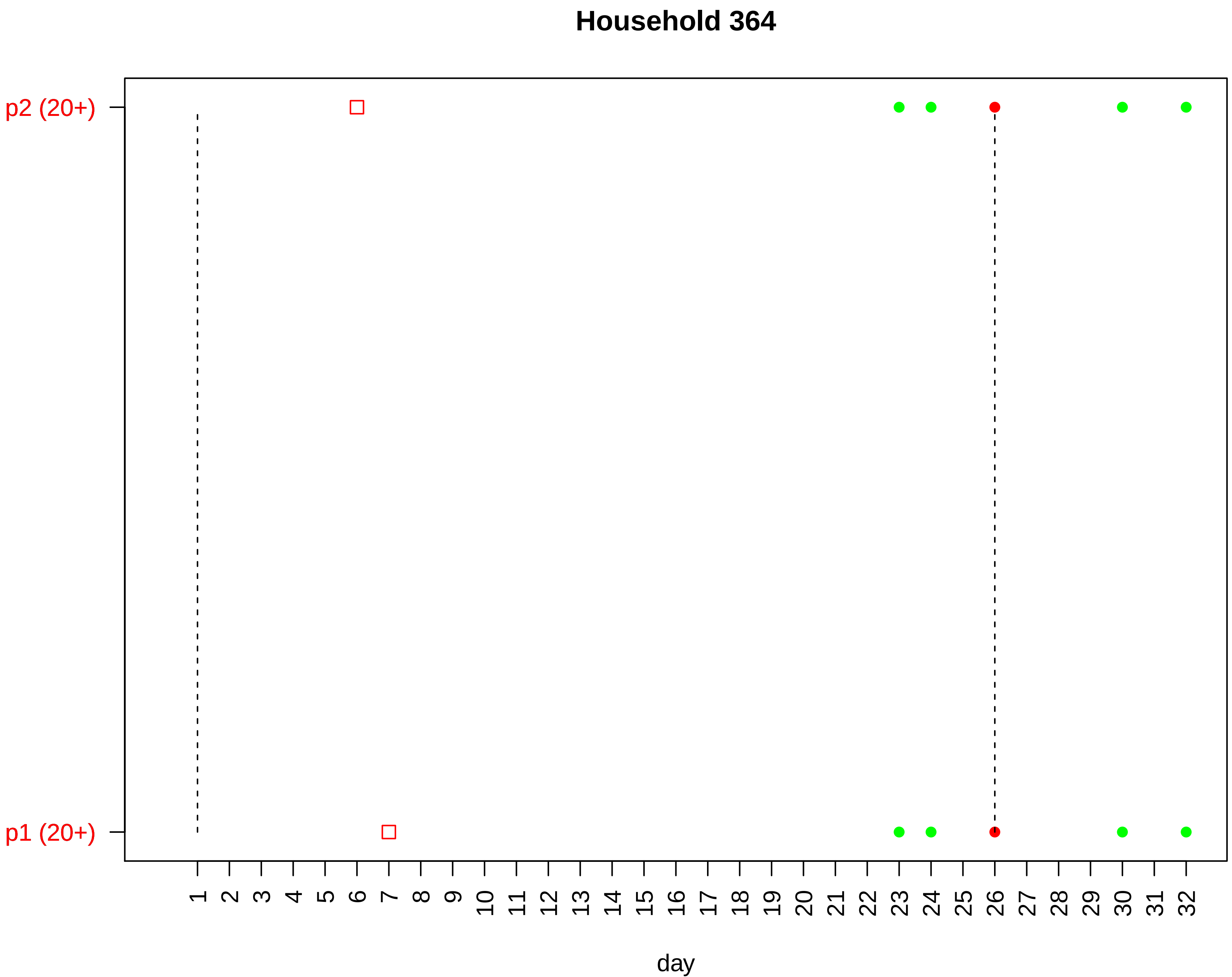

Household 364
p2 (20+)
p1 (20+)
day

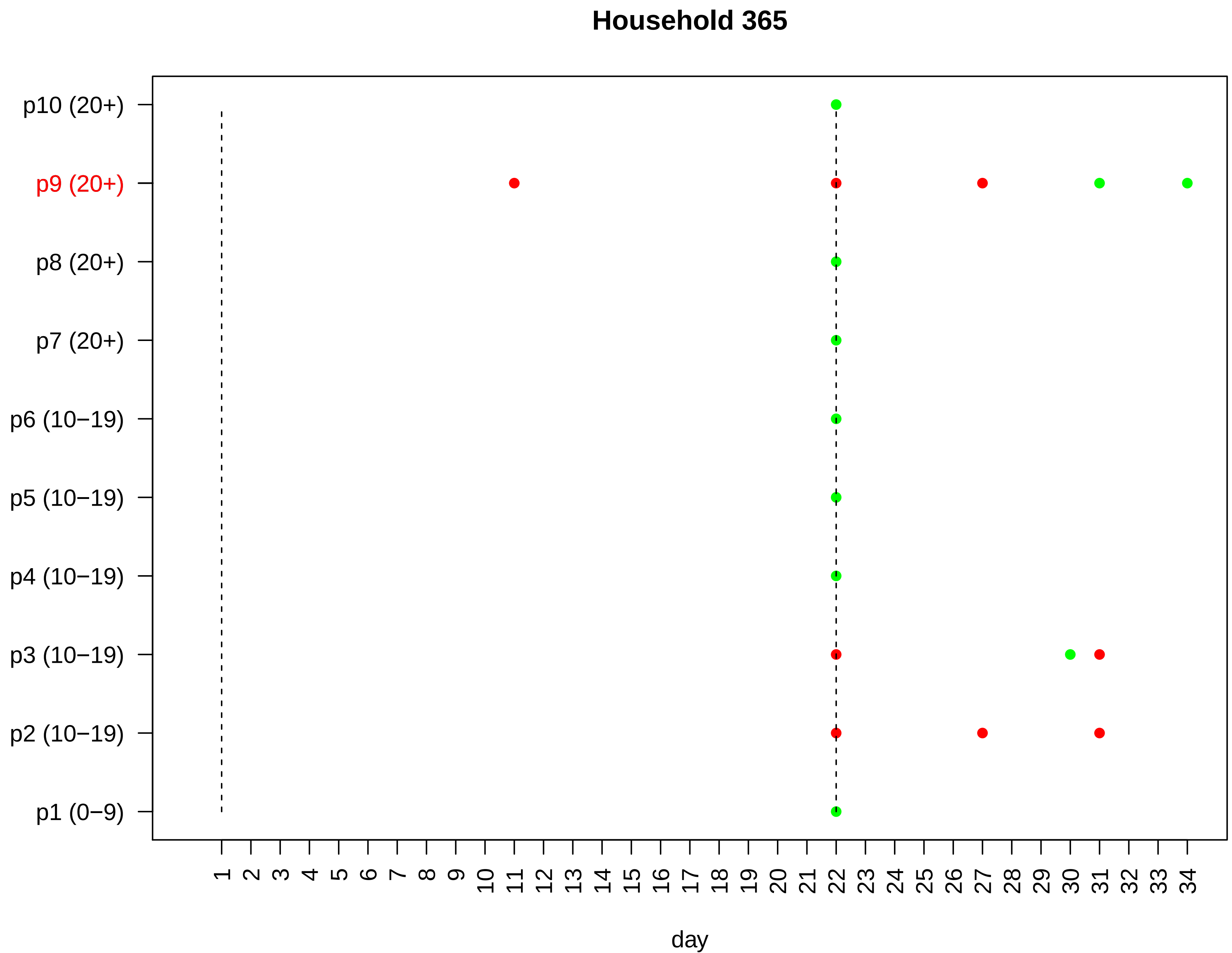

Household 365
p10 (20+)
p9 (20+)
p8 (20+)
p7 (20+)
p6 (10−19)
p5 (10−19)
p4 (10−19)
p3 (10−19)
p2 (10−19)
p1 (0−9)
1 2 3 4 5 6 7 8 9 10 11 12 13 14 15 16 17 18 19 20 21 22 23 24 25 26 27 28 29 30 31 32 33 34
day

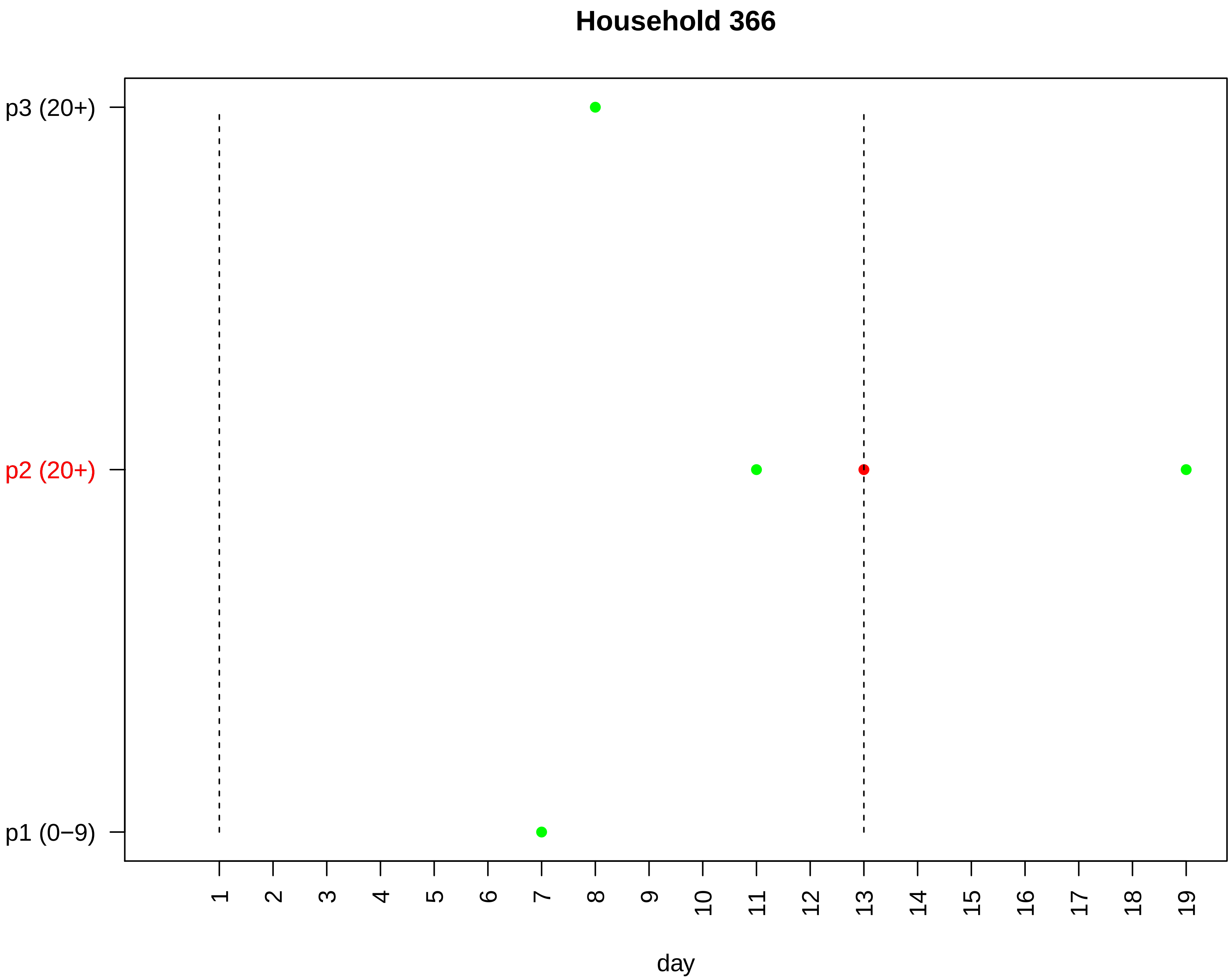
Household 366
p3 (20+)
p2 (20+)
p1 (0−9)
day

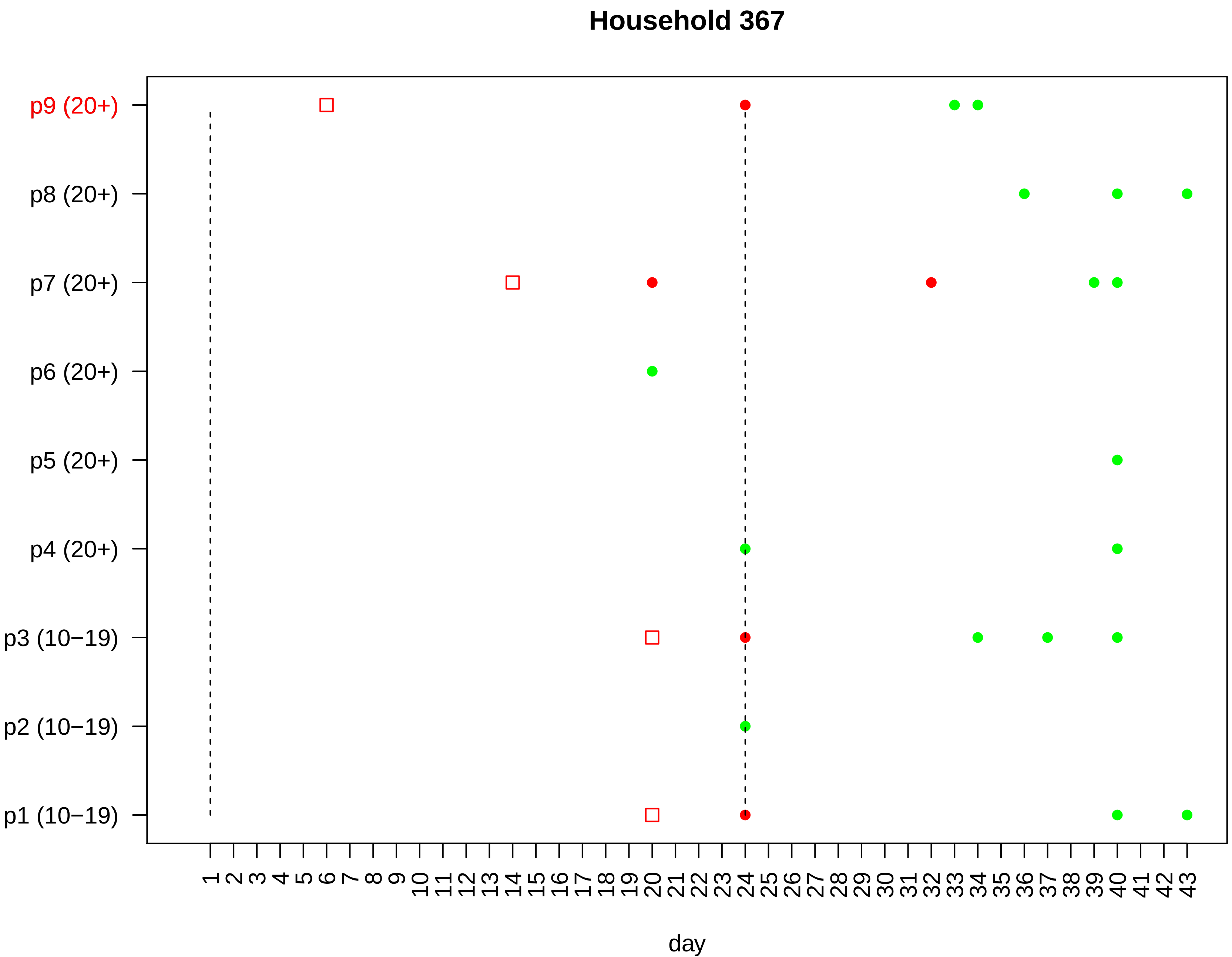

Household 367
p9 (20+)
p8 (20+)
p7 (20+)
p6 (20+)
p5 (20+)
p4 (20+)
p3 (10−19)
p2 (10−19)
p1 (10−19)
1 2 3 4 5 6 7 8 9 10 11 12 13 14 15 16 17 18 19 20 21 22 23 24 25 26 27 28 29 30 31 32 33 34 35 36 37 38 39 40 41 42 43
day

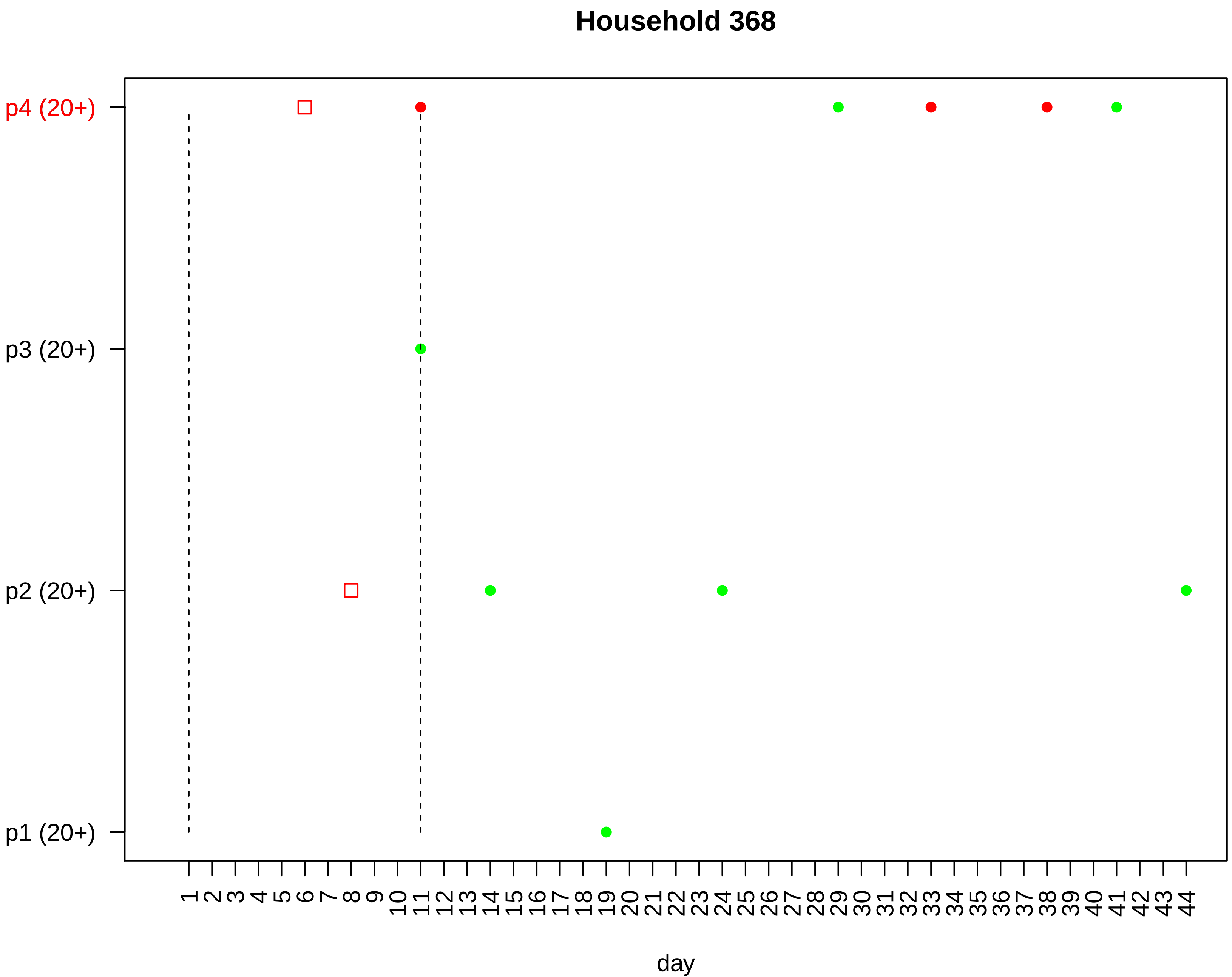

Household 368
p4 (20+)
p3 (20+)
p2 (20+)
p1 (20+)
1 2 3 4 5 6 7 8 9 10 11 12 13 14 15 16 17 18 19 20 21 22 23 24 25 26 27 28 29 30 31 32 33 34 35 36 37 38 39 40 41 42 43 44
day

#### Household 369

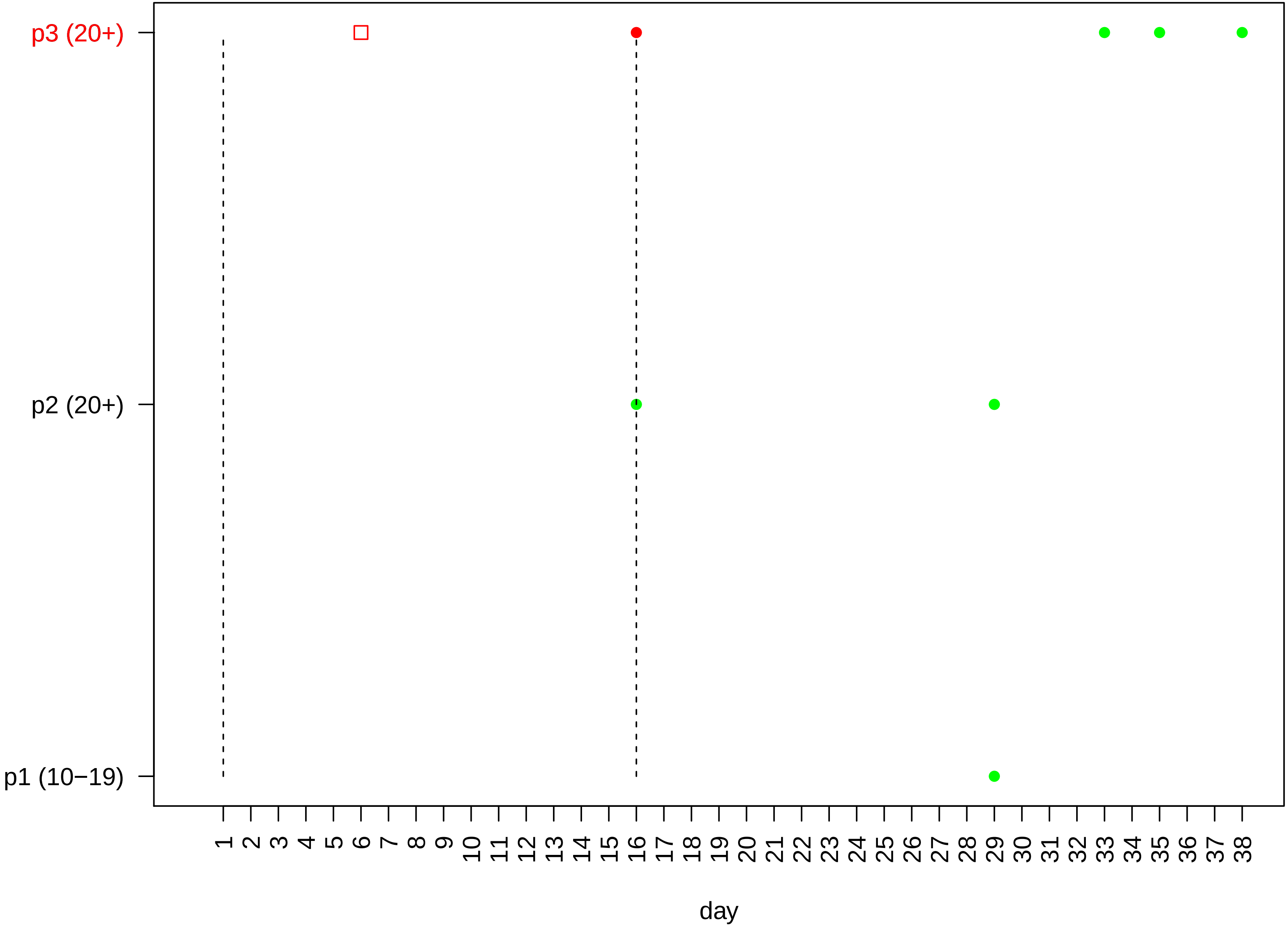

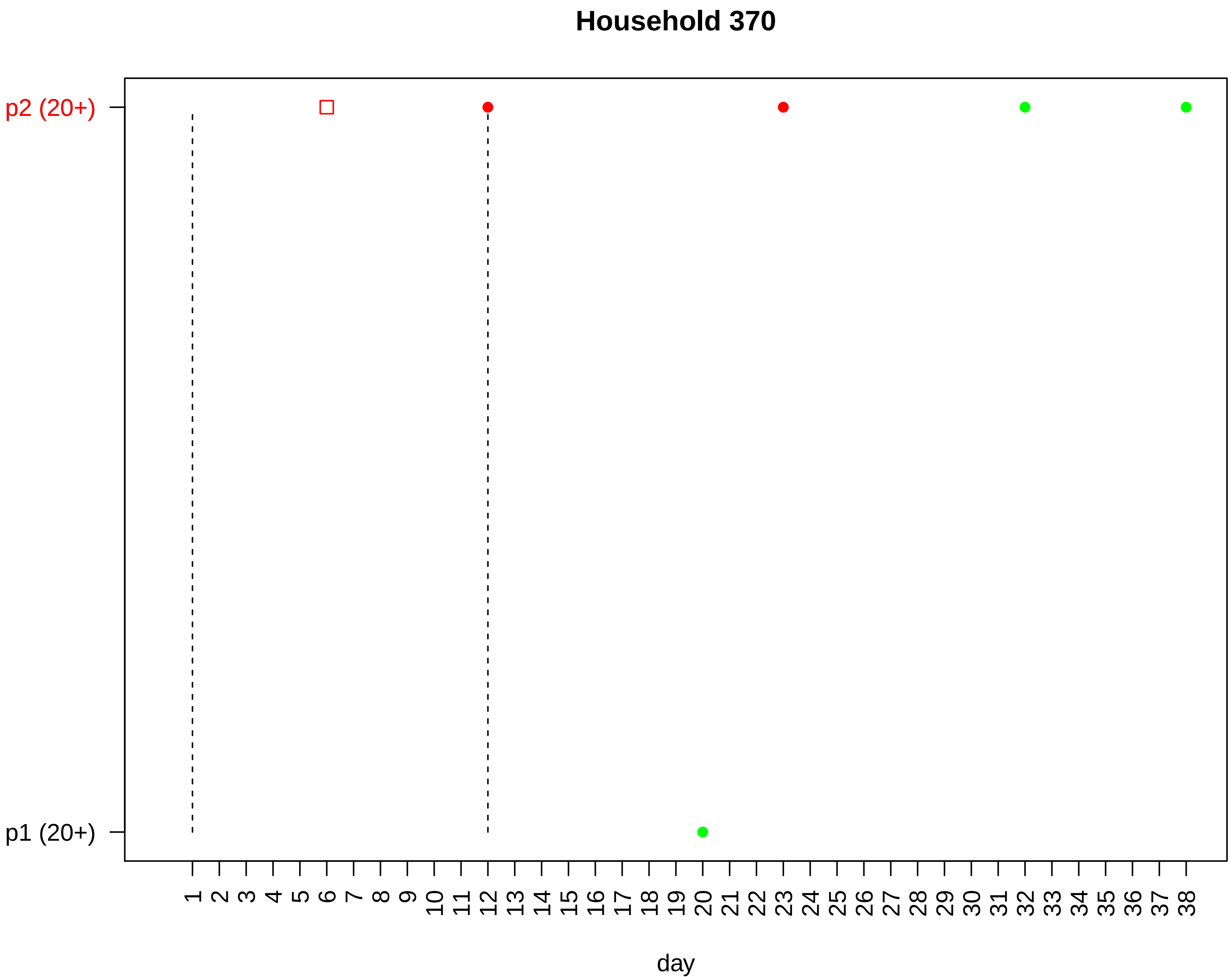
Household 370
p2 (20+)
p1 (20+)
1 2 3 4 5 6 7 8 9 10 11 12 13 14 15 16 17 18 19 20 21 22 23 24 25 26 27 28 29 30 31 32 33 34 35 36 37 38
day

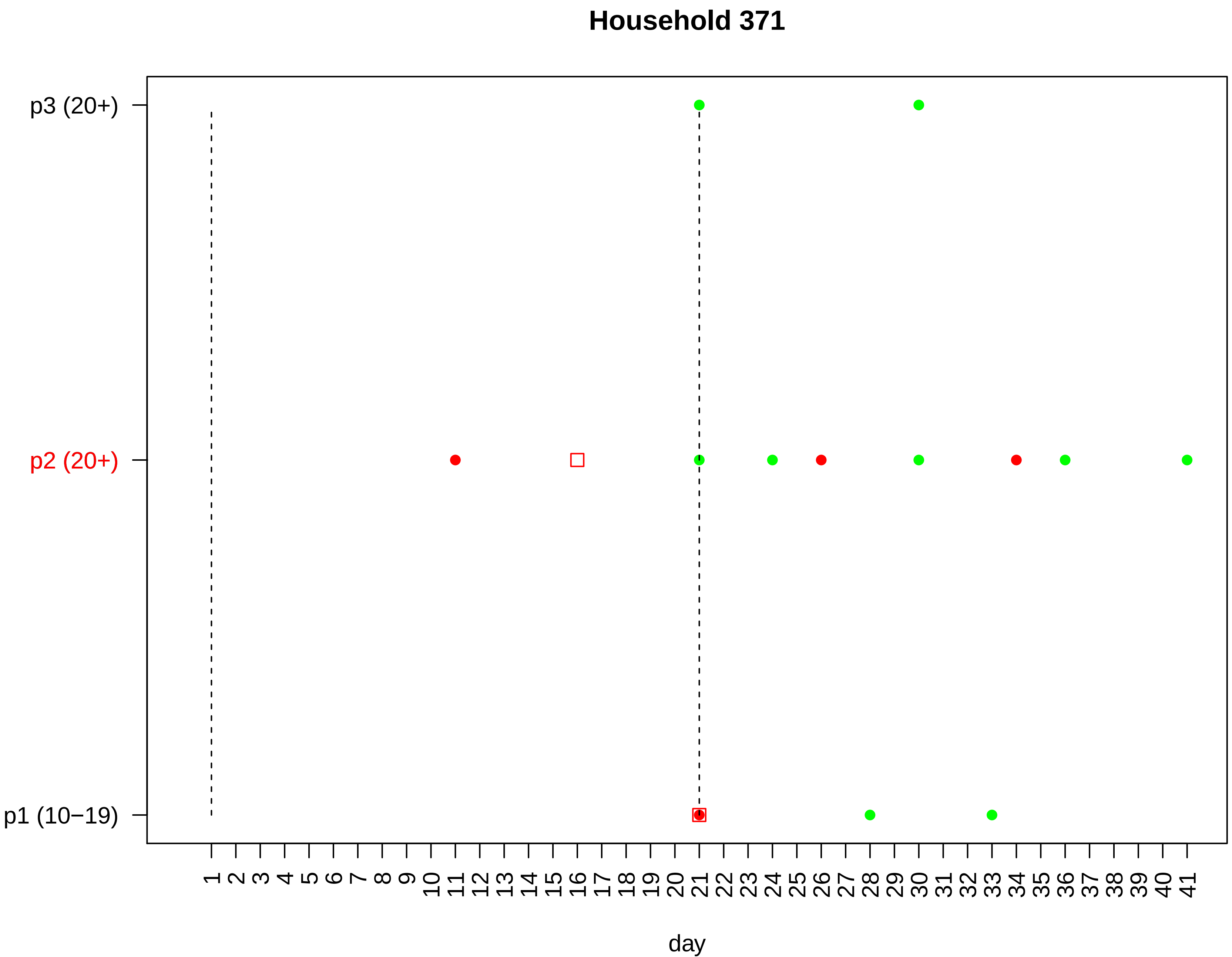
Household 371
p3 (20+)
p2 (20+)
p1 (10−19)
1 2 3 4 5 6 7 8 9 10 11 12 13 14 15 16 17 18 19 20 21 22 23 24 25 26 27 28 29 30 31 32 33 34 35 36 37 38 39 40 41
day

#### Household 372

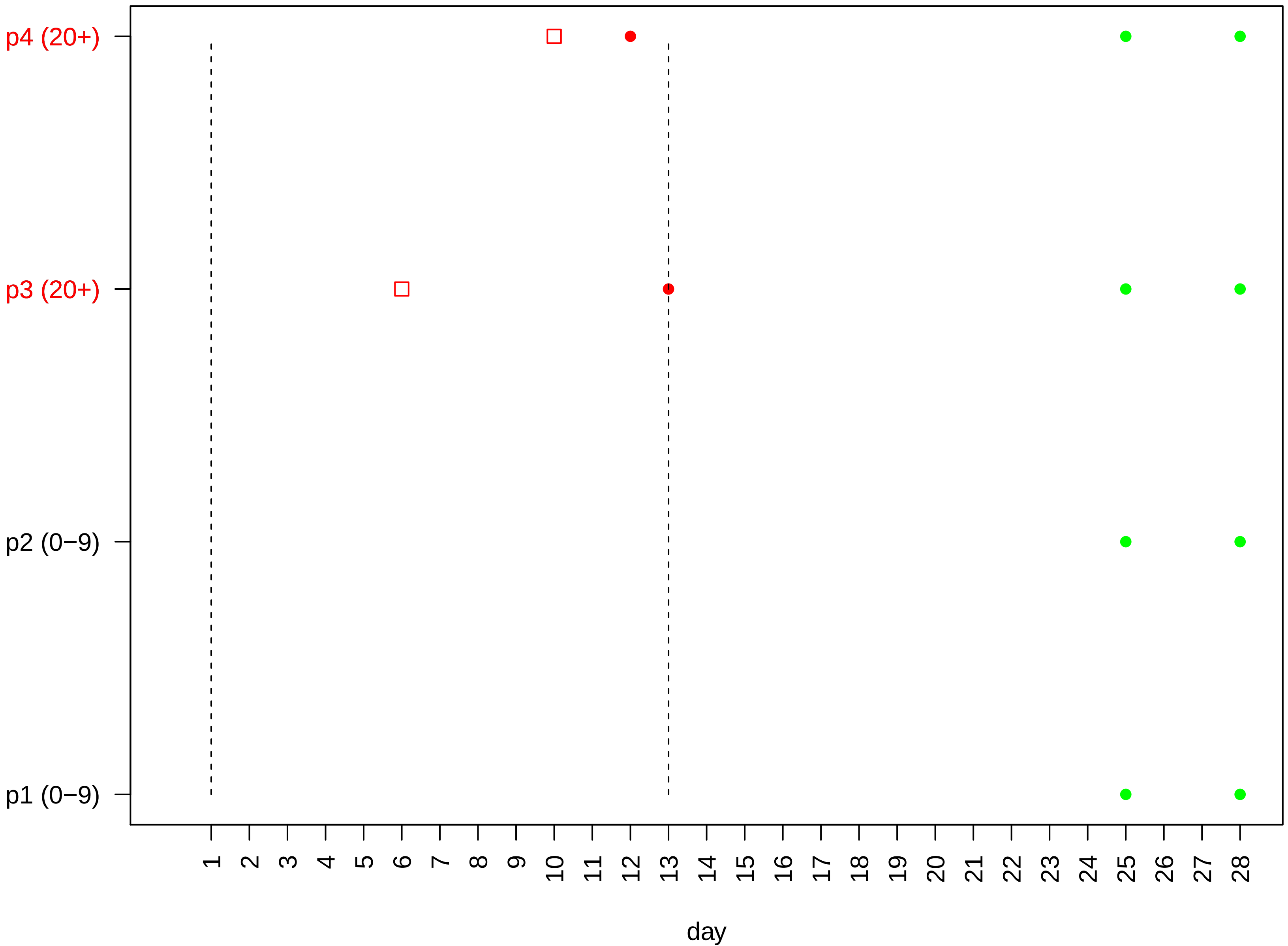

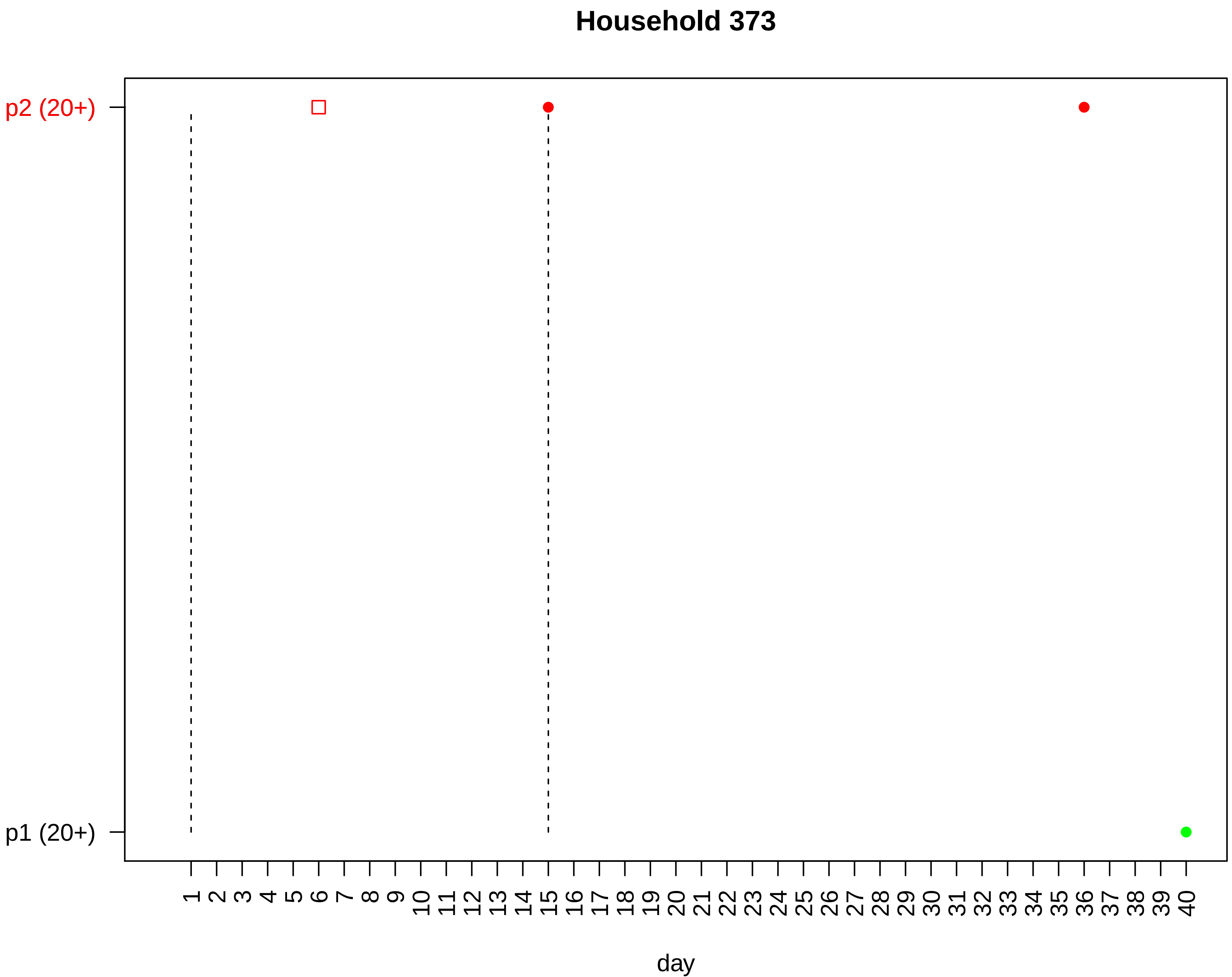
Household 373
p2 (20+)
p1 (20+)
1 2 3 4 5 6 7 8 9 10 11 12 13 14 15 16 17 18 19 20 21 22 23 24 25 26 27 28 29 30 31 32 33 34 35 36 37 38 39 40
day

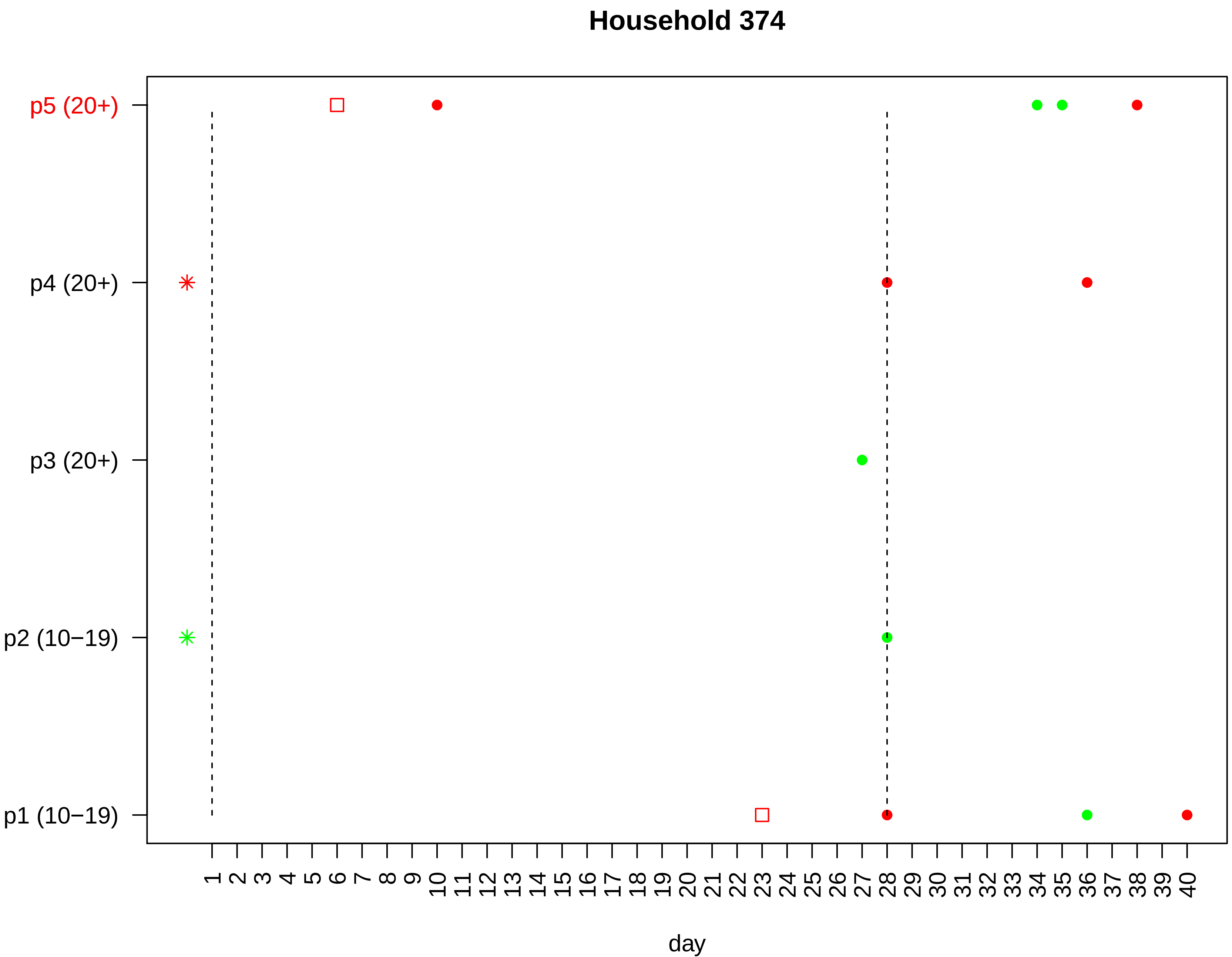
Household 374
p5 (20+)
p4 (20+)
p3 (20+)
p2 (10−19)
p1 (10−19)
1 2 3 4 5 6 7 8 9 10 11 12 13 14 15 16 17 18 19 20 21 22 23 24 25 26 27 28 29 30 31 32 33 34 35 36 37 38 39 40
day

Household 375

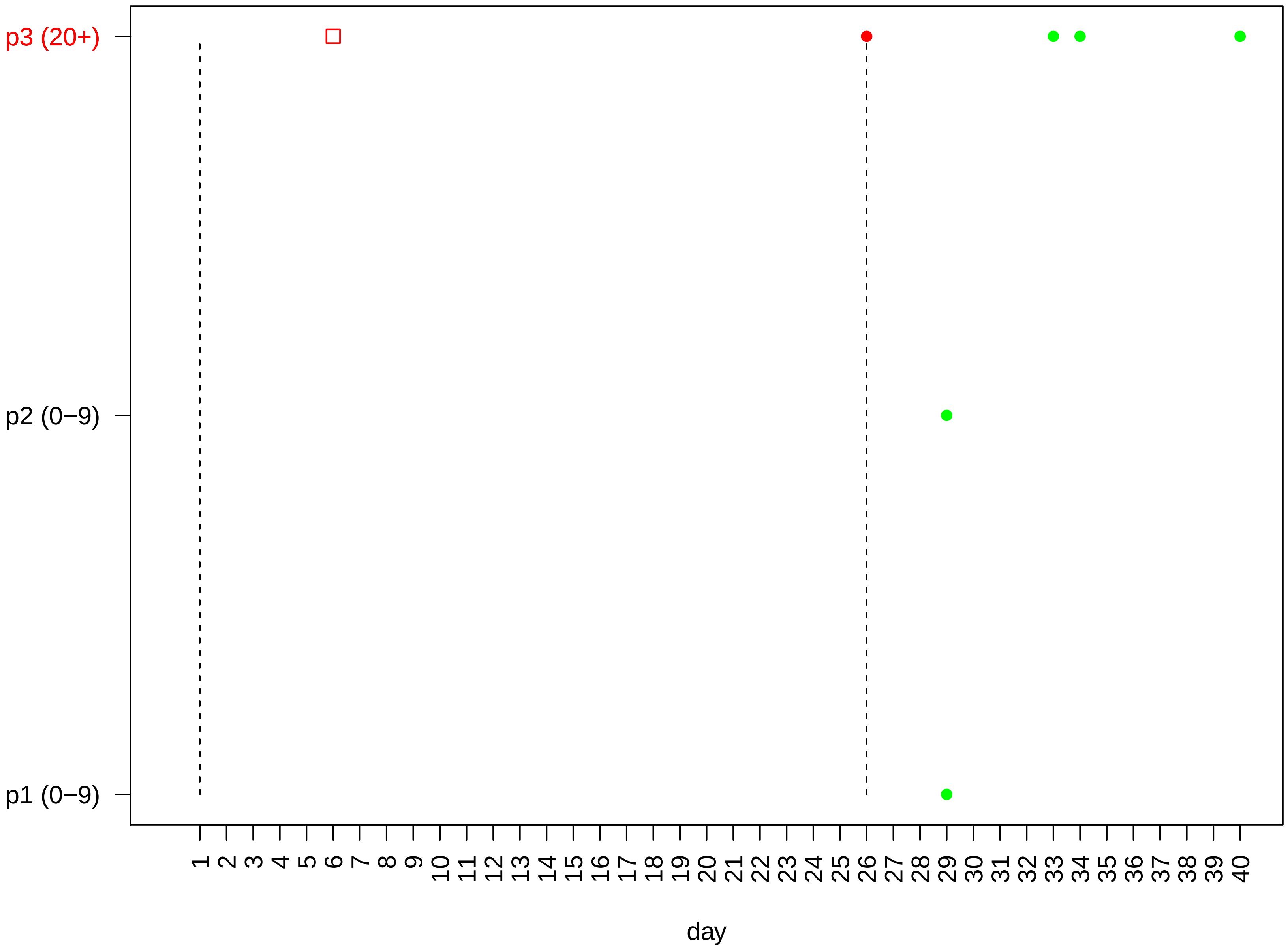

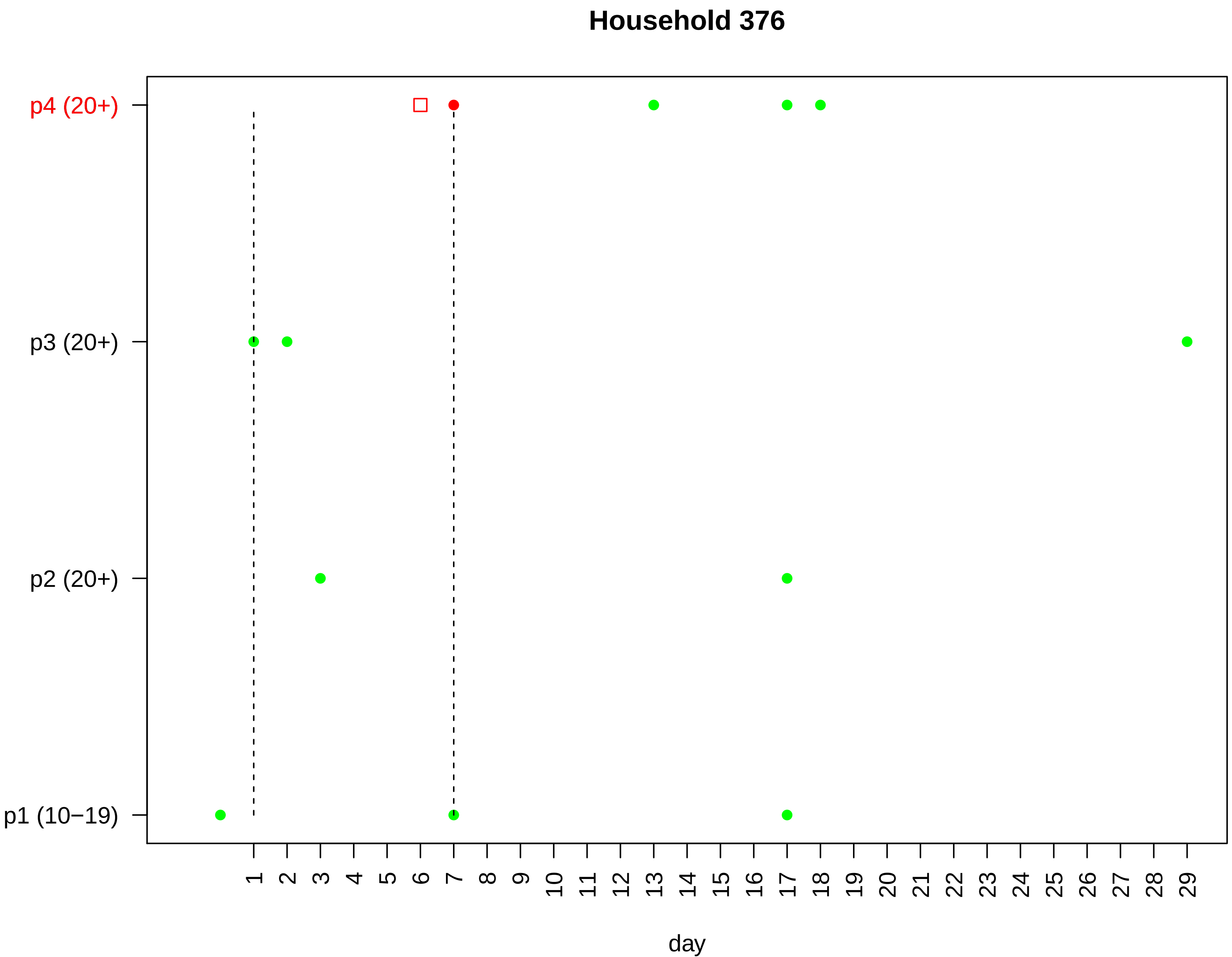
Household 376
p4 (20+)
p3 (20+)
p2 (20+)
p1 (10−19)
1 2 3 4 5 6 7 8 9 10 11 12 13 14 15 16 17 18 19 20 21 22 23 24 25 26 27 28 29
day

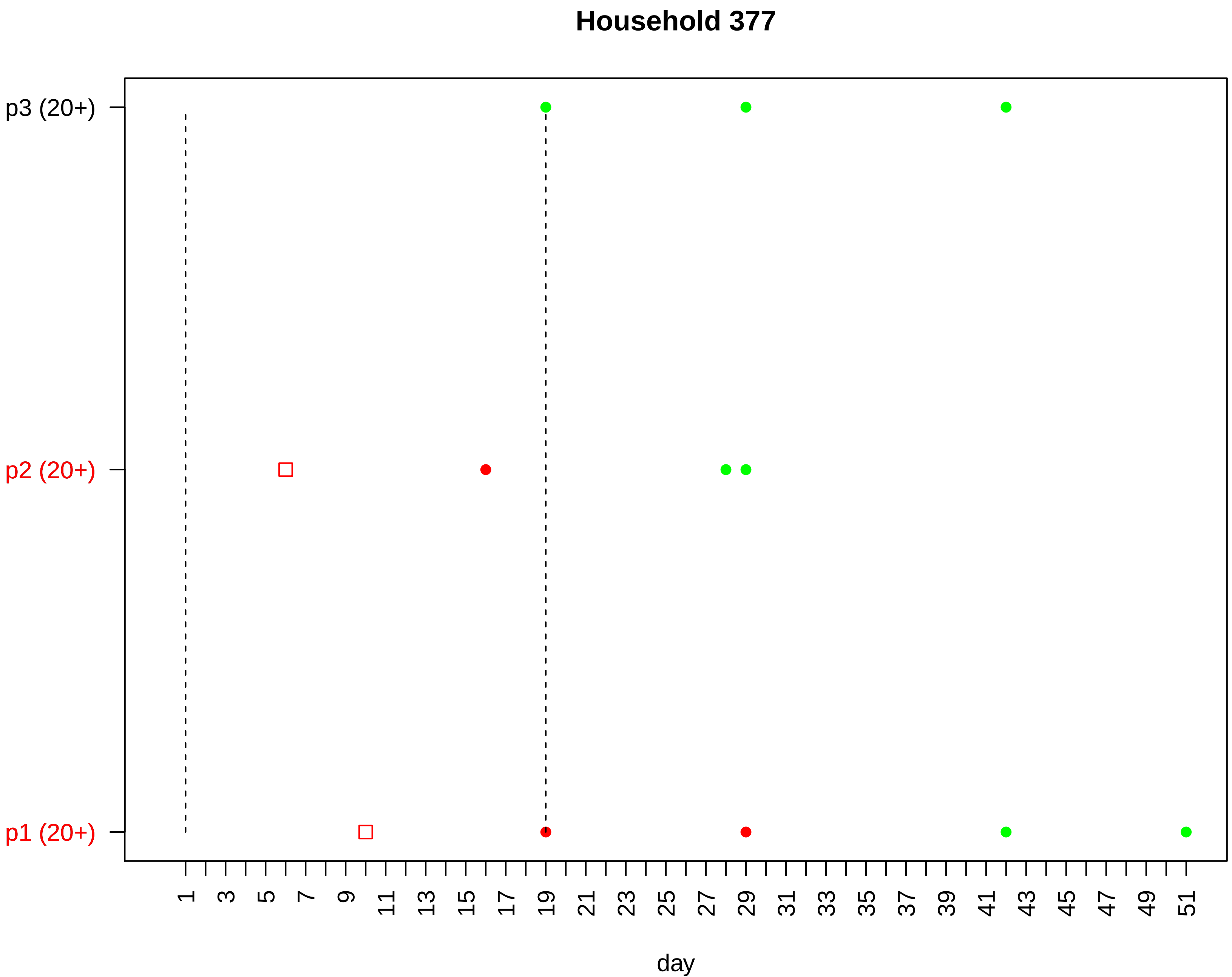

Household 377
p3 (20+)
p2 (20+)
p1 (20+)
1
3
5
7
9
11
13
15
17
19
21
23
25
27
29
31
33
35
37
39
41
43
45
47
49
51
day

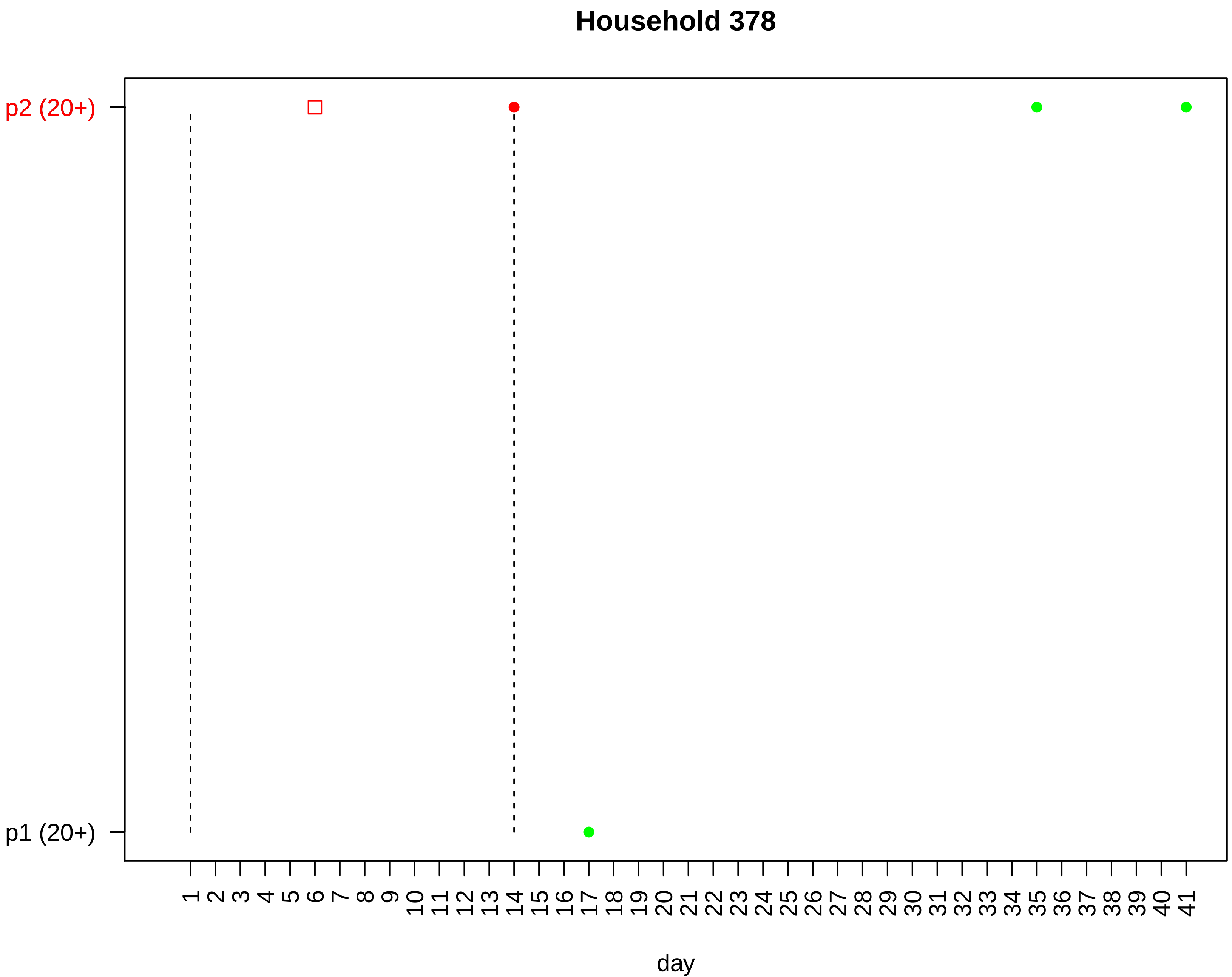

Household 378
p2 (20+)
p1 (20+)
1 2 3 4 5 6 7 8 9 10 11 12 13 14 15 16 17 18 19 20 21 22 23 24 25 26 27 28 29 30 31 32 33 34 35 36 37 38 39 40 41
day

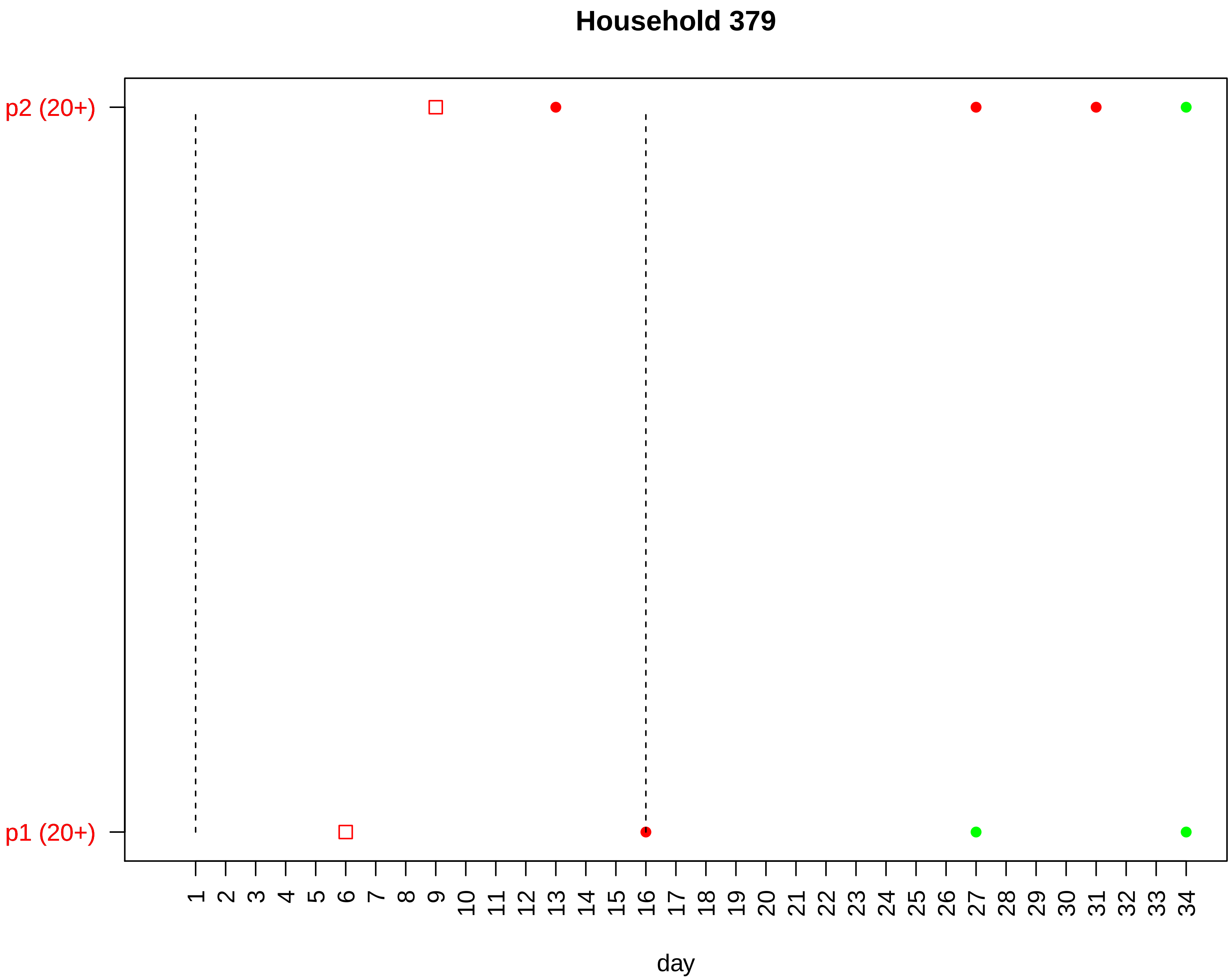
Household 379
p2 (20+)
p1 (20+)
1 2 3 4 5 6 7 8 9 10 11 12 13 14 15 16 17 18 19 20 21 22 23 24 25 26 27 28 29 30 31 32 33 34
day

#### Household 380

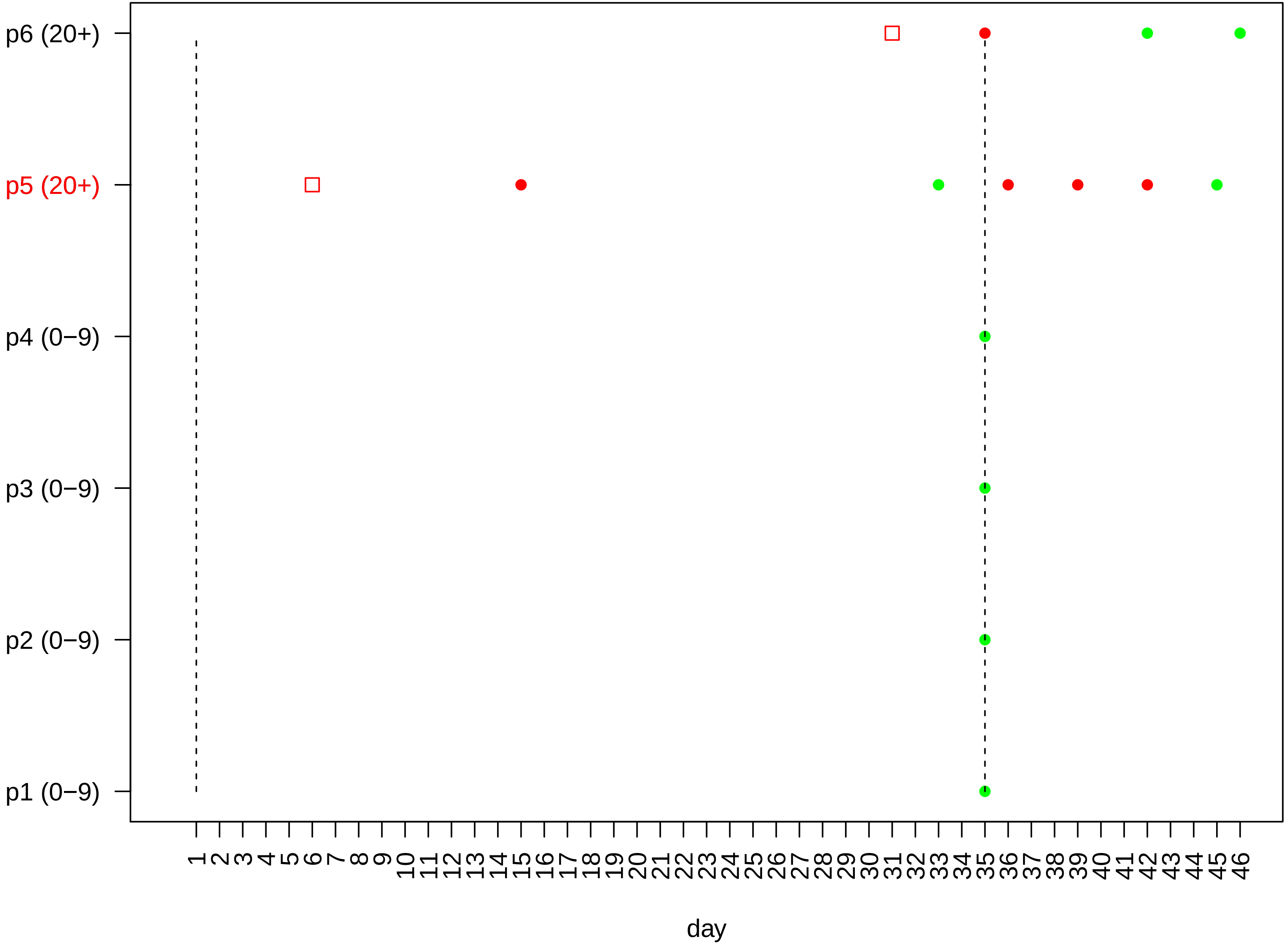

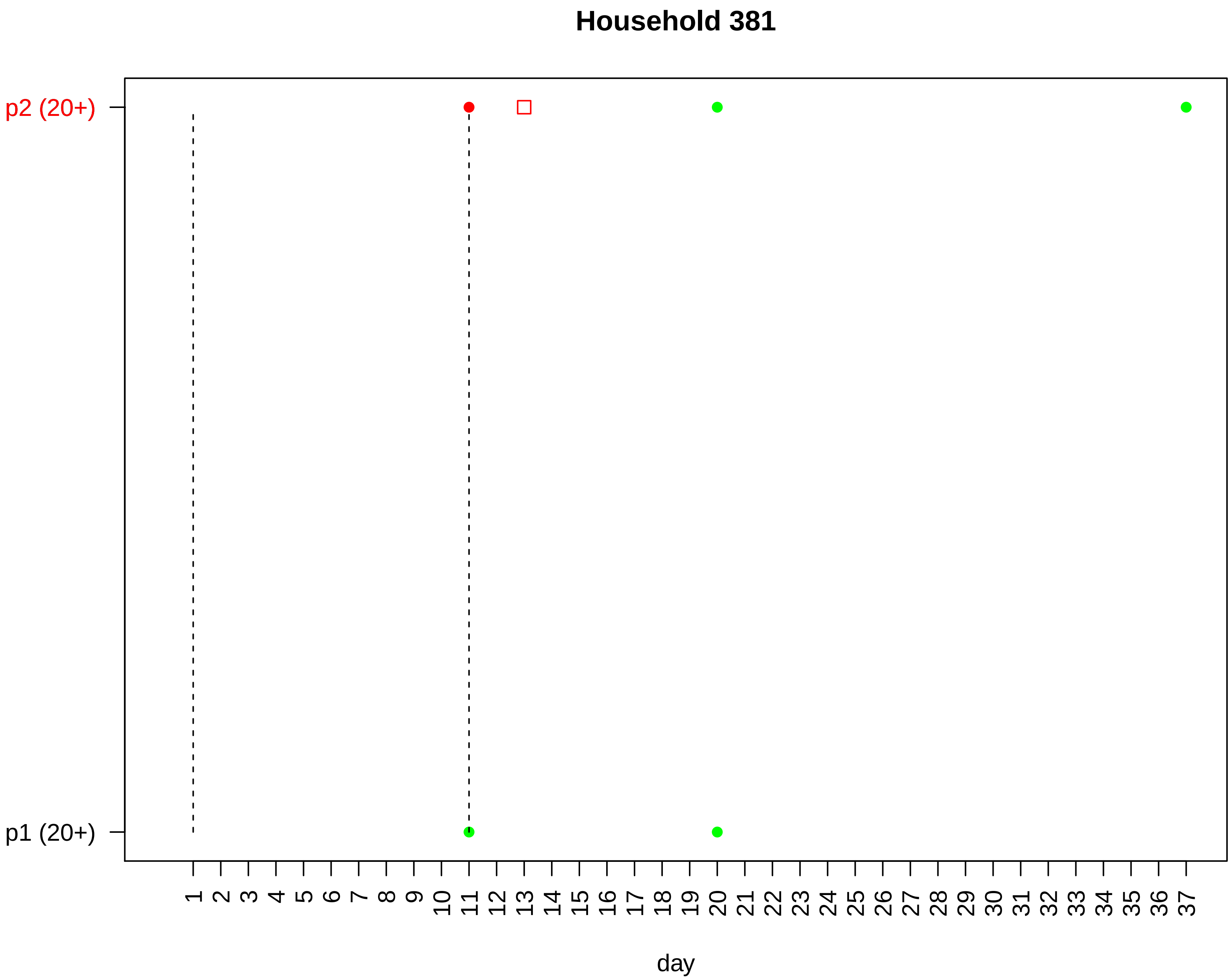
Household 381
p2 (20+)
p1 (20+)
day

### Household 382

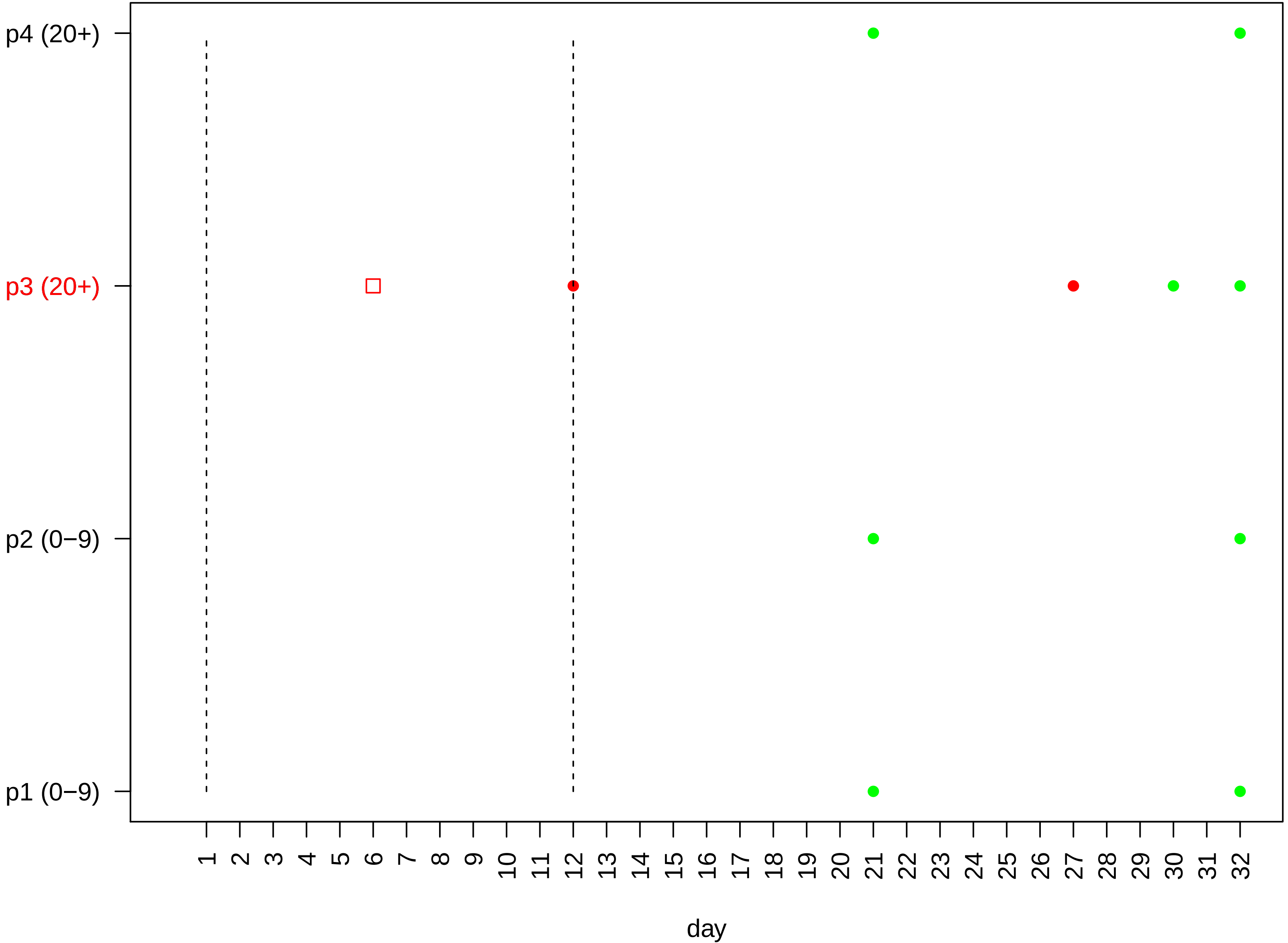

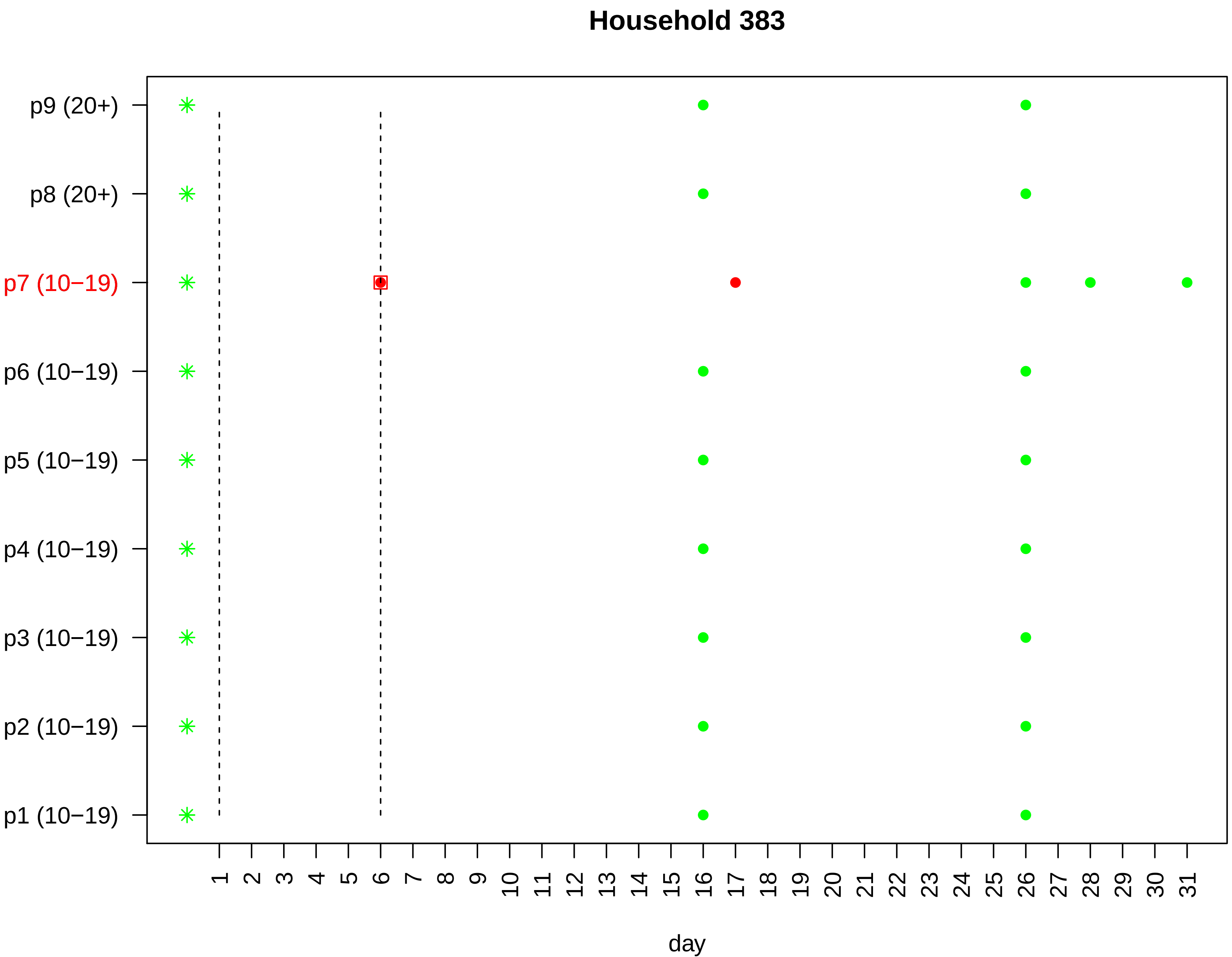

Household 383
p9 (20+)
p8 (20+)
p7 (10−19)
p6 (10−19)
p5 (10−19)
p4 (10−19)
p3 (10−19)
p2 (10−19)
p1 (10−19)
1 2 3 4 5 6 7 8 9 10 11 12 13 14 15 16 17 18 19 20 21 22 23 24 25 26 27 28 29 30 31
day

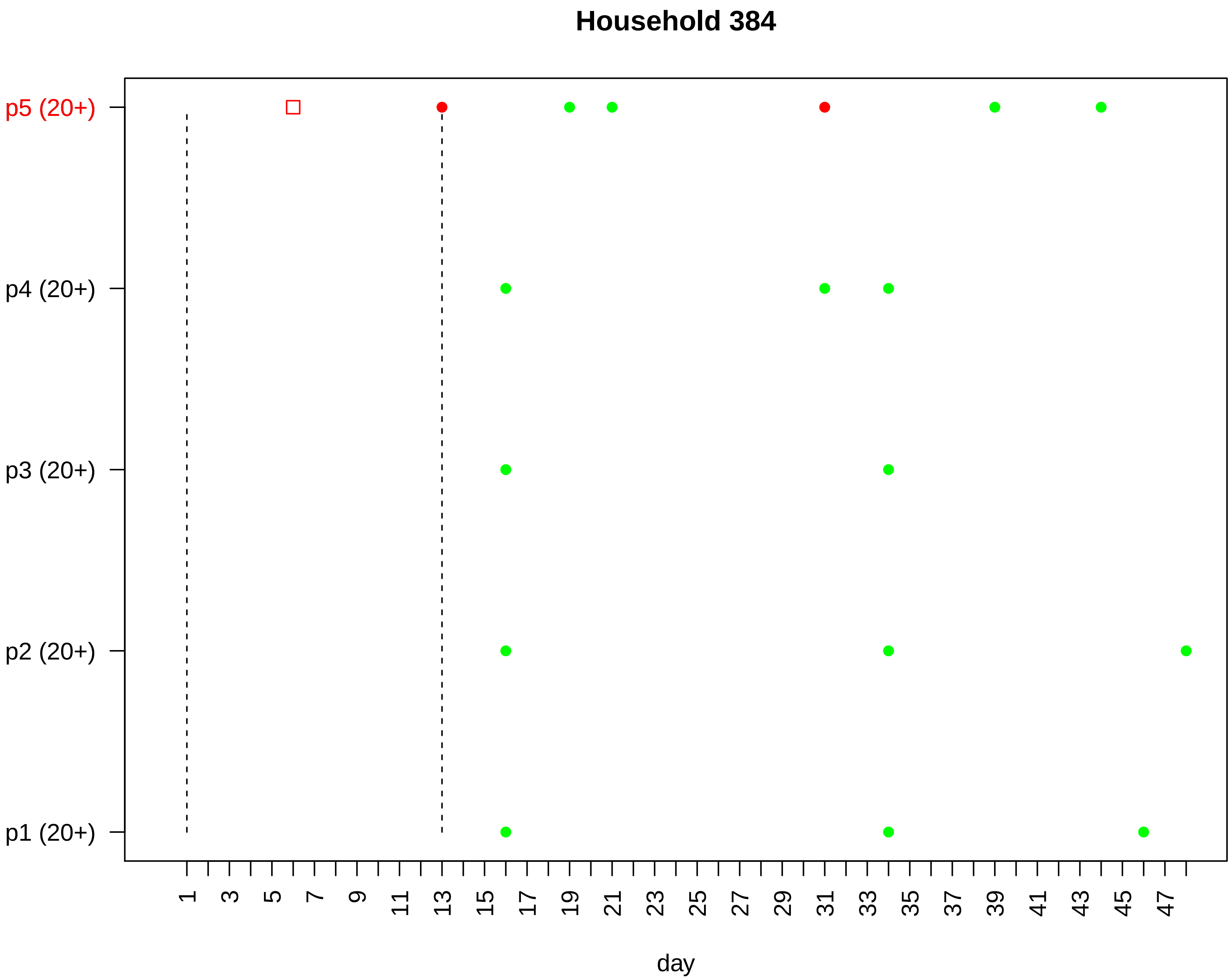

Household 384
p5 (20+)
p4 (20+)
p3 (20+)
p2 (20+)
p1 (20+)
1
3
5
7
9
11
13
15
17
19
21
23
25
27
29
31
33
35
37
39
41
43
45
47
day

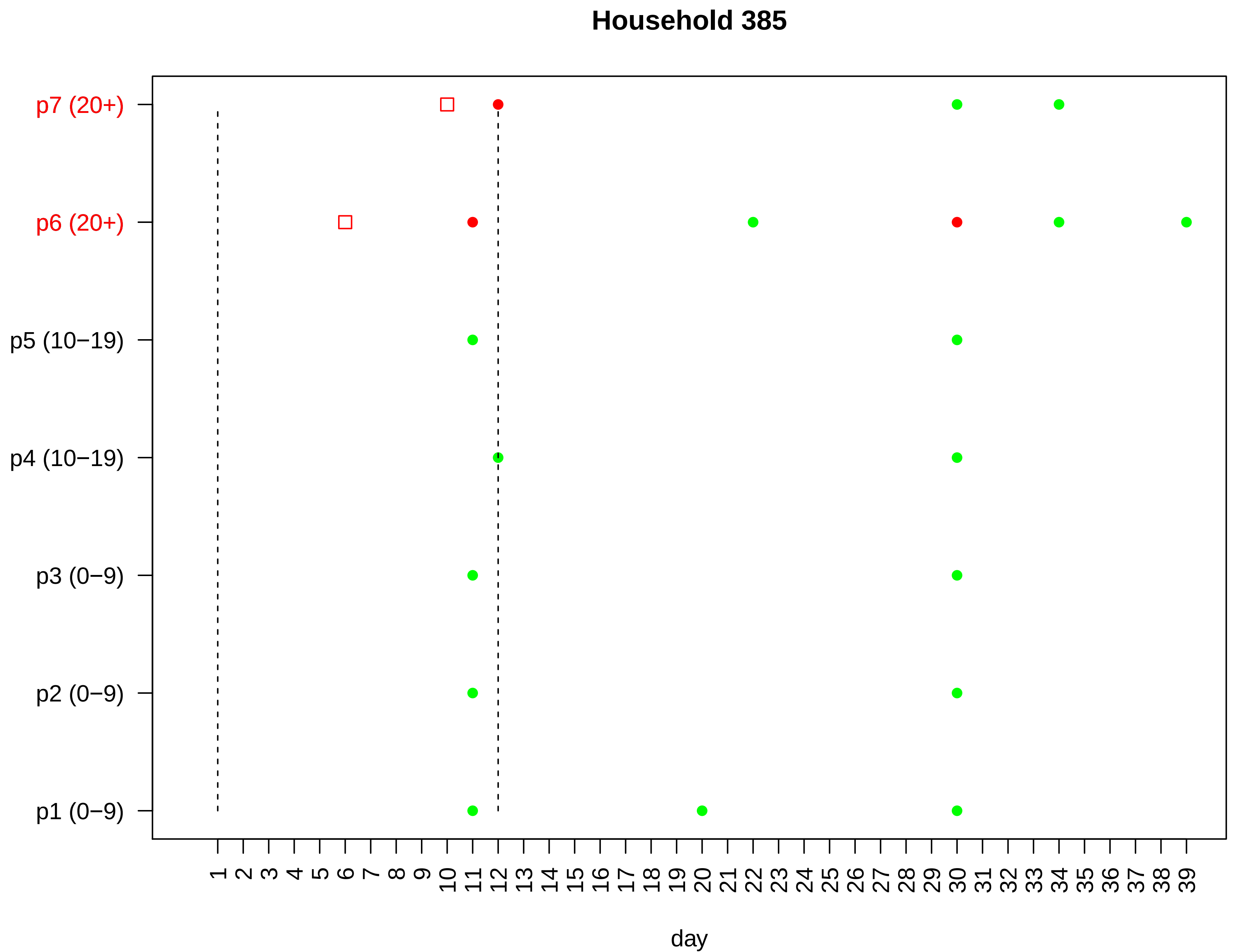

Household 385
p7 (20+)
p6 (20+)
p5 (10−19)
p4 (10−19)
p3 (0−9)
p2 (0−9)
p1 (0−9)
1 2 3 4 5 6 7 8 9 10 11 12 13 14 15 16 17 18 19 20 21 22 23 24 25 26 27 28 29 30 31 32 33 34 35 36 37 38 39
day

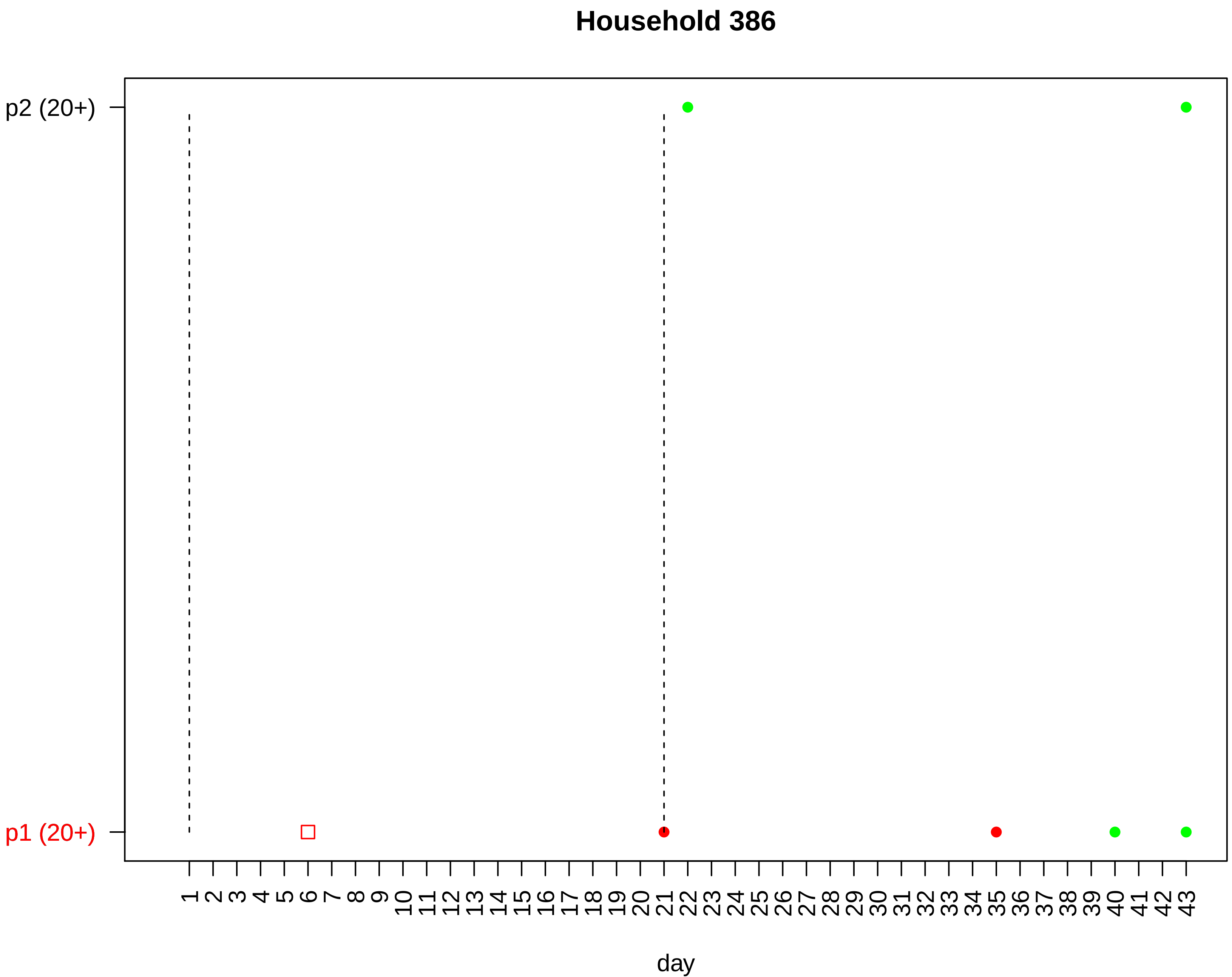

Household 386
p2 (20+)
p1 (20+)
1 2 3 4 5 6 7 8 9 10 11 12 13 14 15 16 17 18 19 20 21 22 23 24 25 26 27 28 29 30 31 32 33 34 35 36 37 38 39 40 41 42 43
day

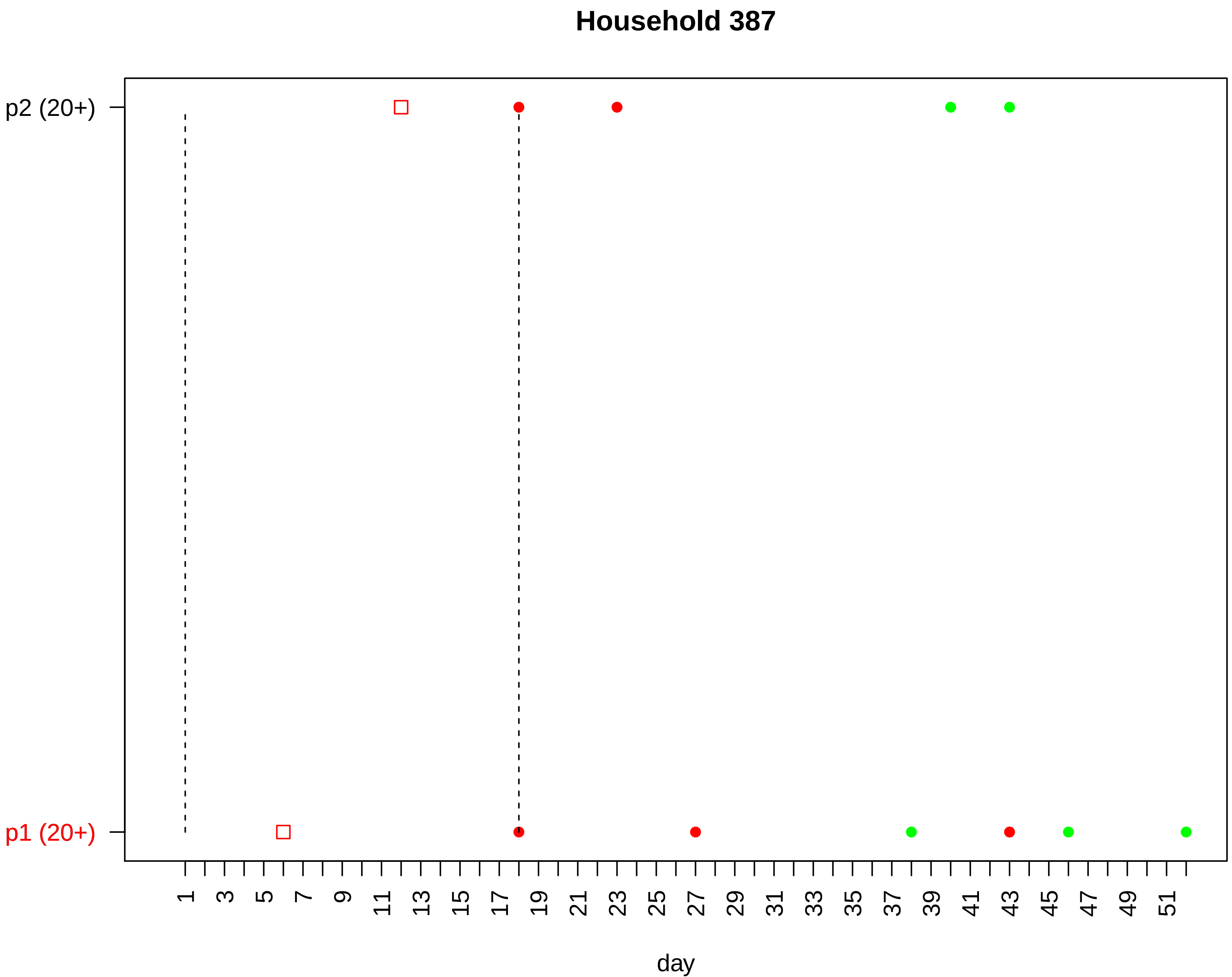
Household 387
p2 (20+)
p1 (20+)
1
3
5
7
9
11
13
15
17
19
21
23
25
27
29
31
33
35
37
39
41
43
45
47
49
51
day

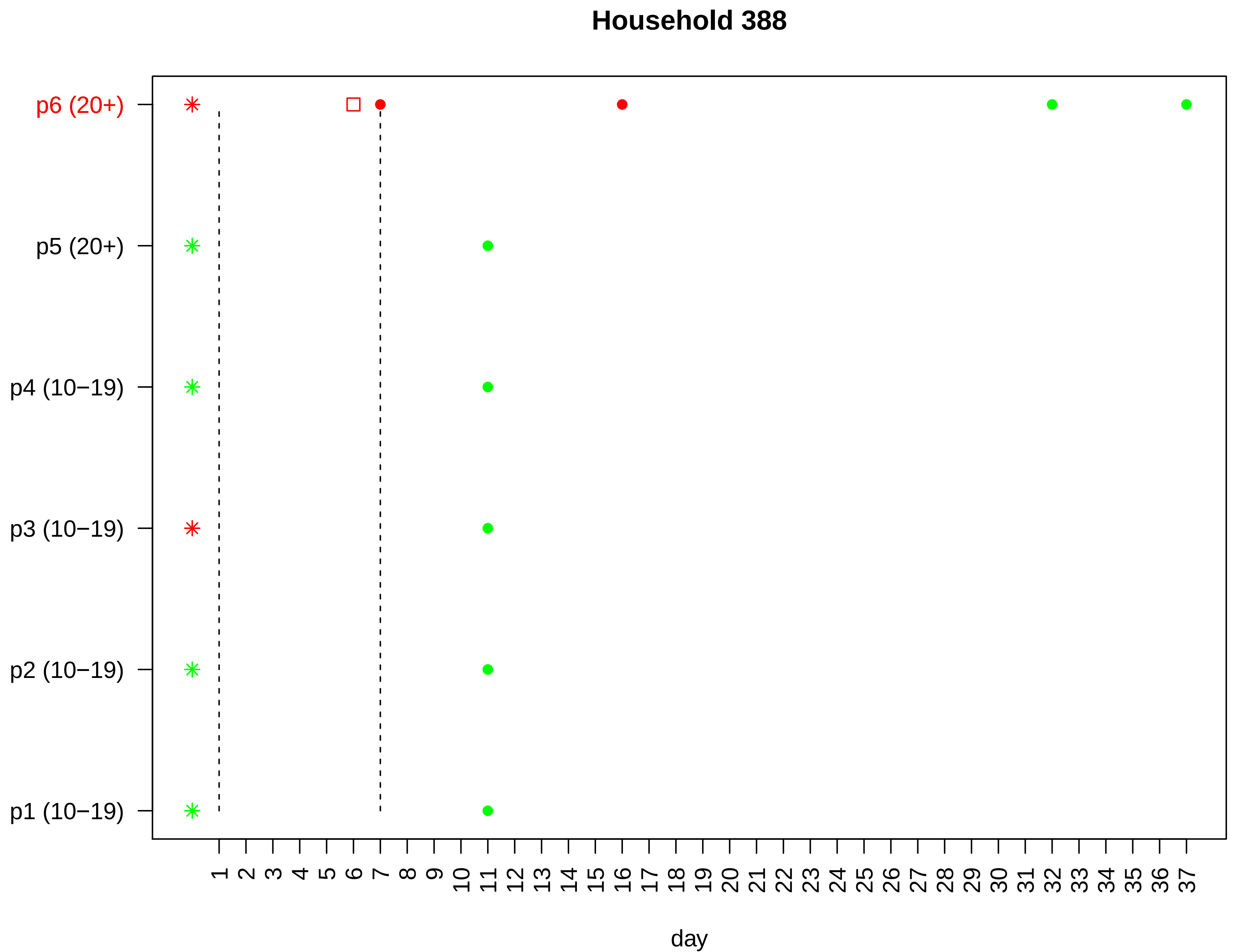

Household 388
p6 (20+)
p5 (20+)
p4 (10−19)
p3 (10−19)
p2 (10−19)
p1 (10−19)
1 2 3 4 5 6 7 8 9 10 11 12 13 14 15 16 17 18 19 20 21 22 23 24 25 26 27 28 29 30 31 32 33 34 35 36 37
day

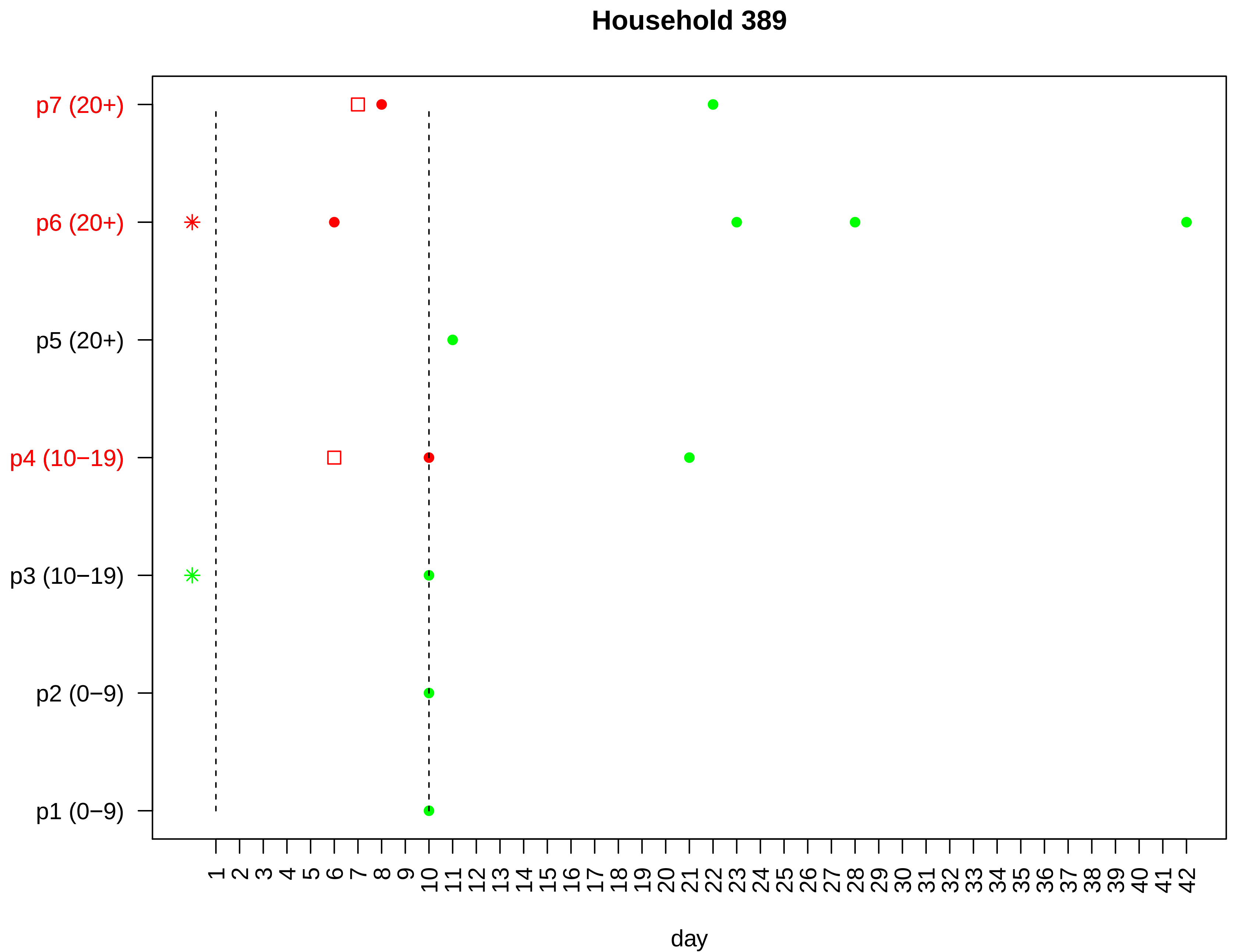

Household 389
p7 (20+)
p6 (20+)
p5 (20+)
p4 (10−19)
p3 (10−19)
p2 (0−9)
p1 (0−9)
1 2 3 4 5 6 7 8 9 10 11 12 13 14 15 16 17 18 19 20 21 22 23 24 25 26 27 28 29 30 31 32 33 34 35 36 37 38 39 40 41 42
day

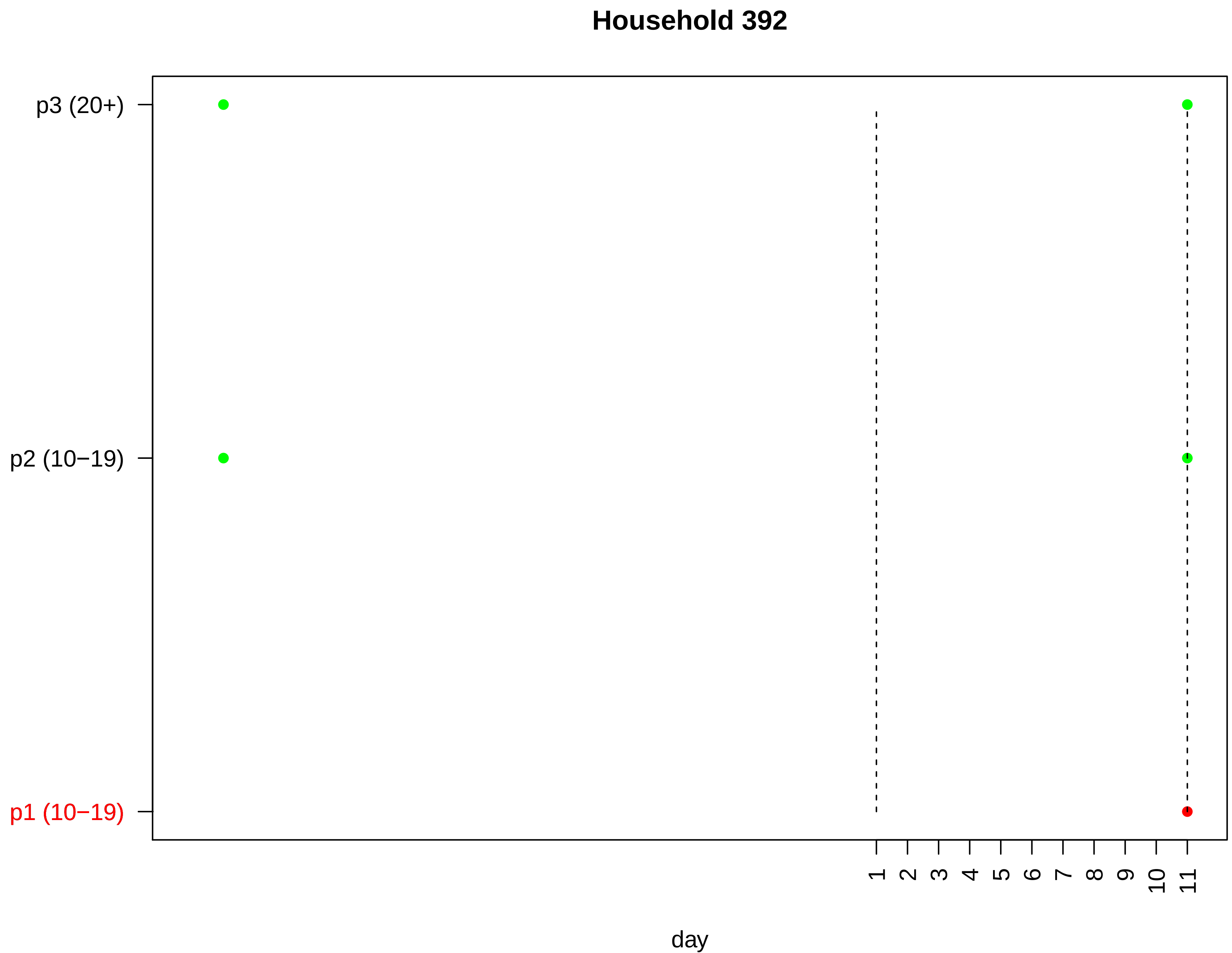
Household 392
p3 (20+)
p2 (10−19)
p1 (10−19)
1
2
3
4
5
6
7
8
9
10
11
day

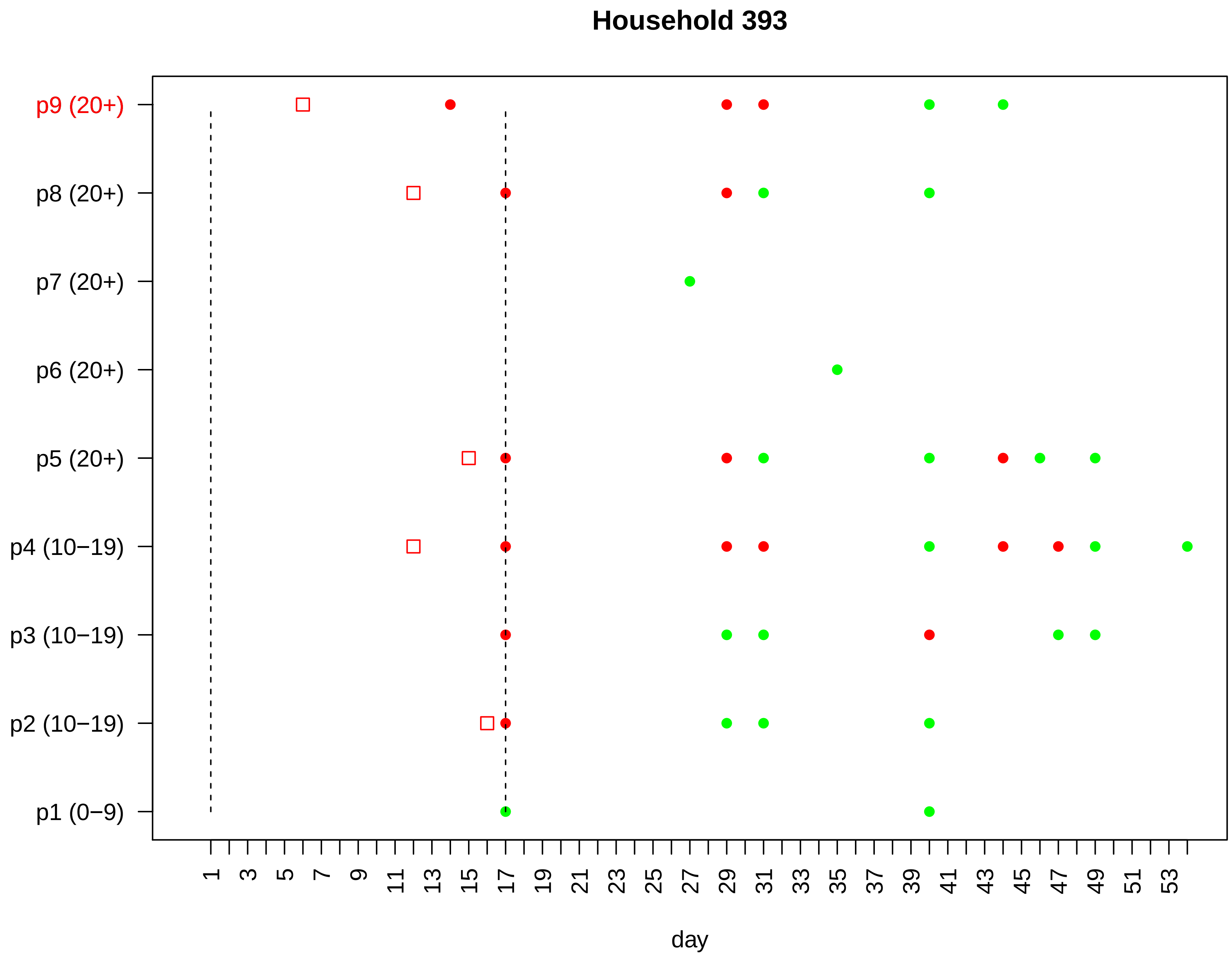

Household 393
p9 (20+)
p8 (20+)
p7 (20+)
p6 (20+)
p5 (20+)
p4 (10−19)
p3 (10−19)
p2 (10−19)
p1 (0−9)
1 3 5 7 9 11 13 15 17 19 21 23 25 27 29 31 33 35 37 39 41 43 45 47 49 51 53
day

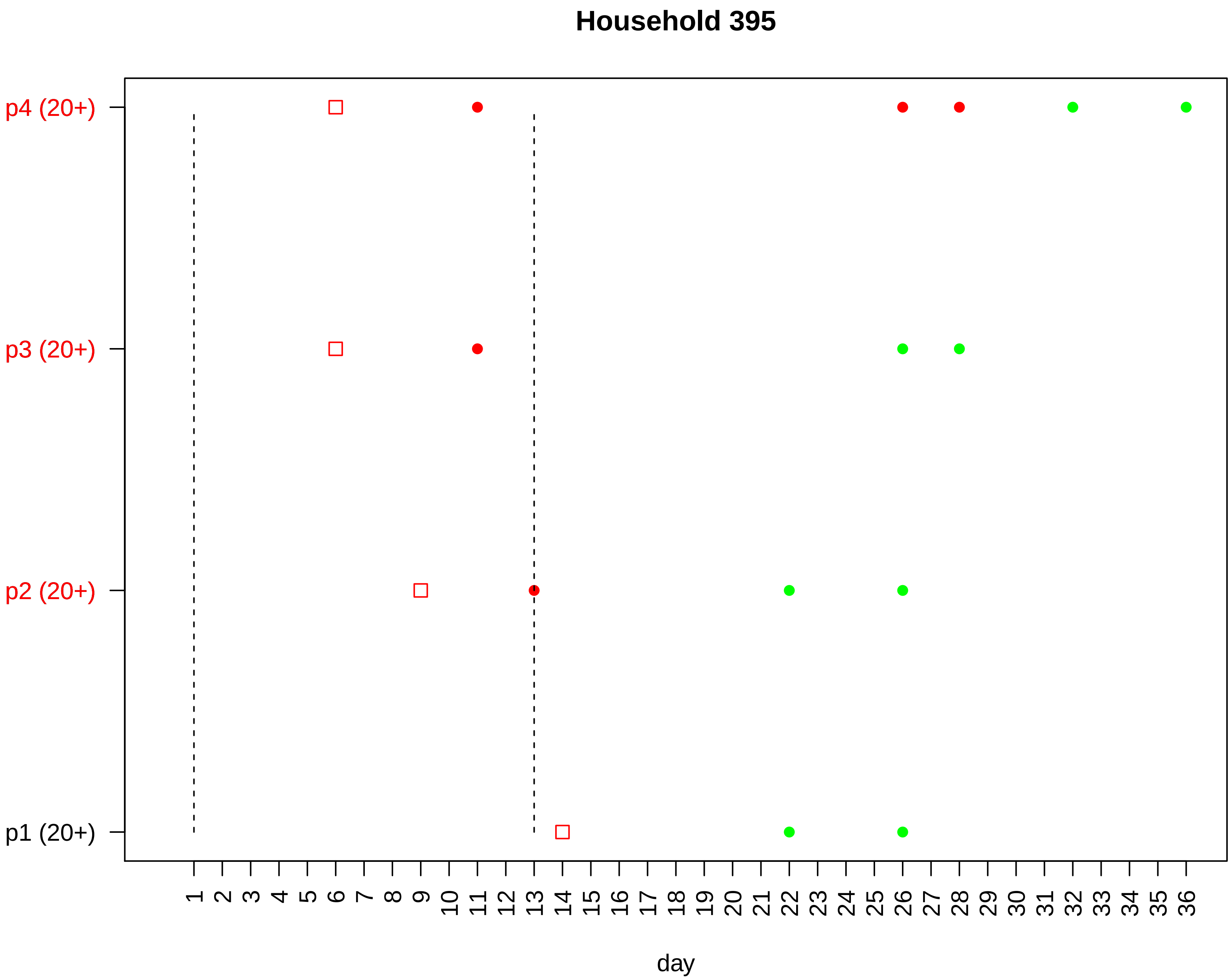

Household 395
p4 (20+)
p3 (20+)
p2 (20+)
p1 (20+)
1 2 3 4 5 6 7 8 9 10 11 12 13 14 15 16 17 18 19 20 21 22 23 24 25 26 27 28 29 30 31 32 33 34 35 36
day

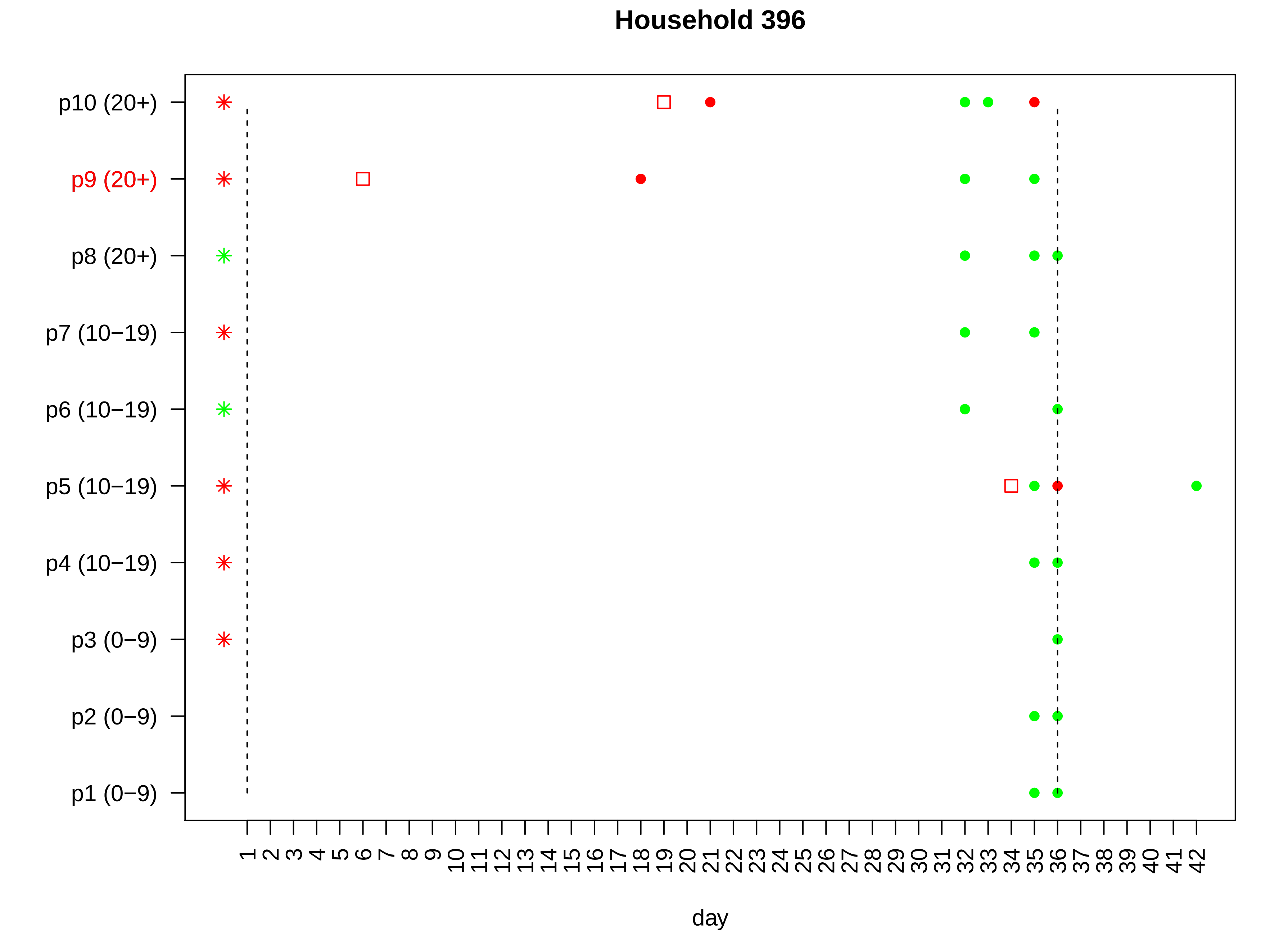

Household 396
p10 (20+)
p9 (20+)
p8 (20+)
p7 (10−19)
p6 (10−19)
p5 (10−19)
p4 (10−19)
p3 (0−9)
p2 (0−9)
p1 (0−9)
1 2 3 4 5 6 7 8 9 10 11 12 13 14 15 16 17 18 19 20 21 22 23 24 25 26 27 28 29 30 31 32 33 34 35 36 37 38 39 40 41 42
day

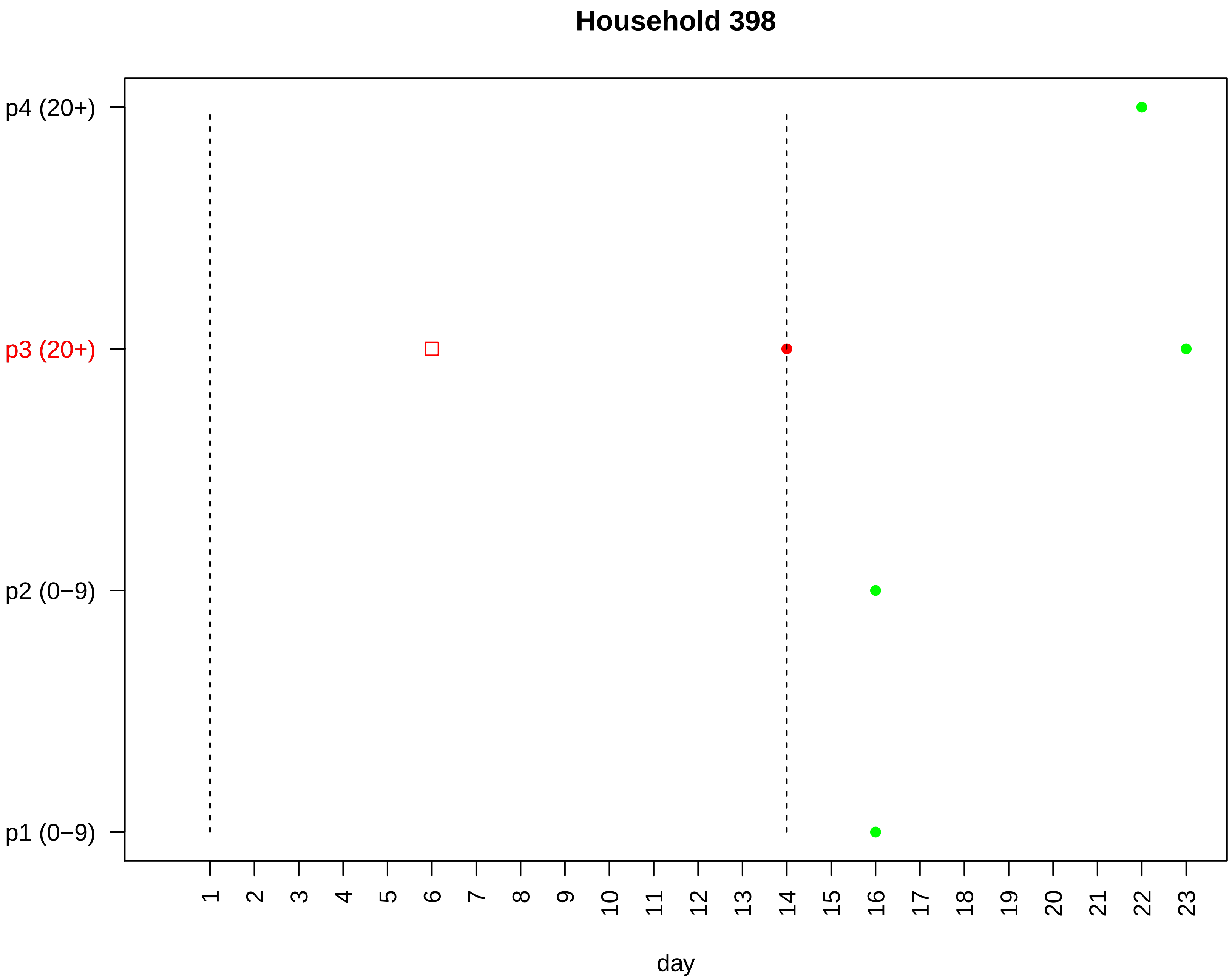
Household 398
p4 (20+)
p3 (20+)
p2 (0−9)
p1 (0−9)
day

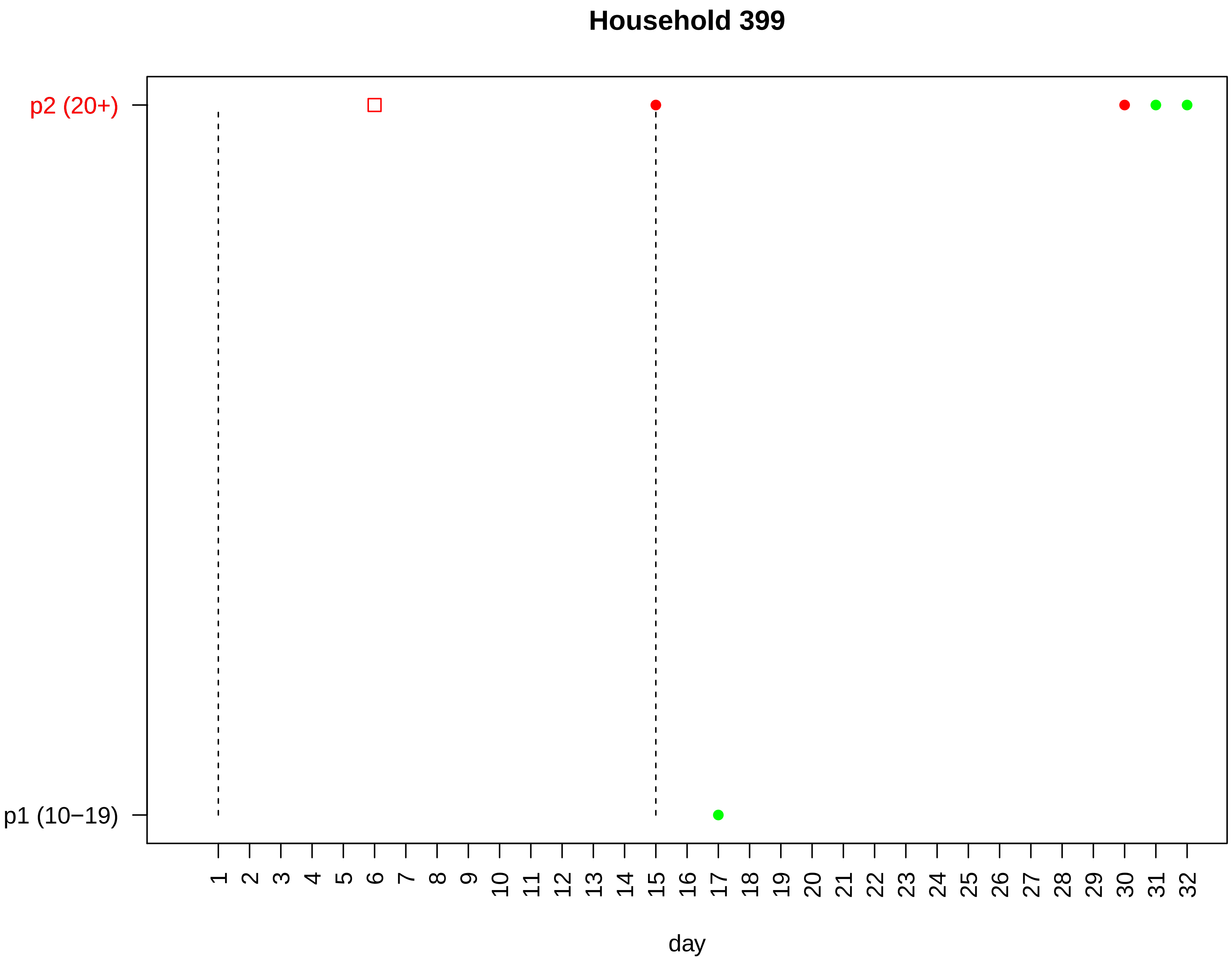

Household 399
p2 (20+)
p1 (10−19)
1 2 3 4 5 6 7 8 9 10 11 12 13 14 15 16 17 18 19 20 21 22 23 24 25 26 27 28 29 30 31 32
day

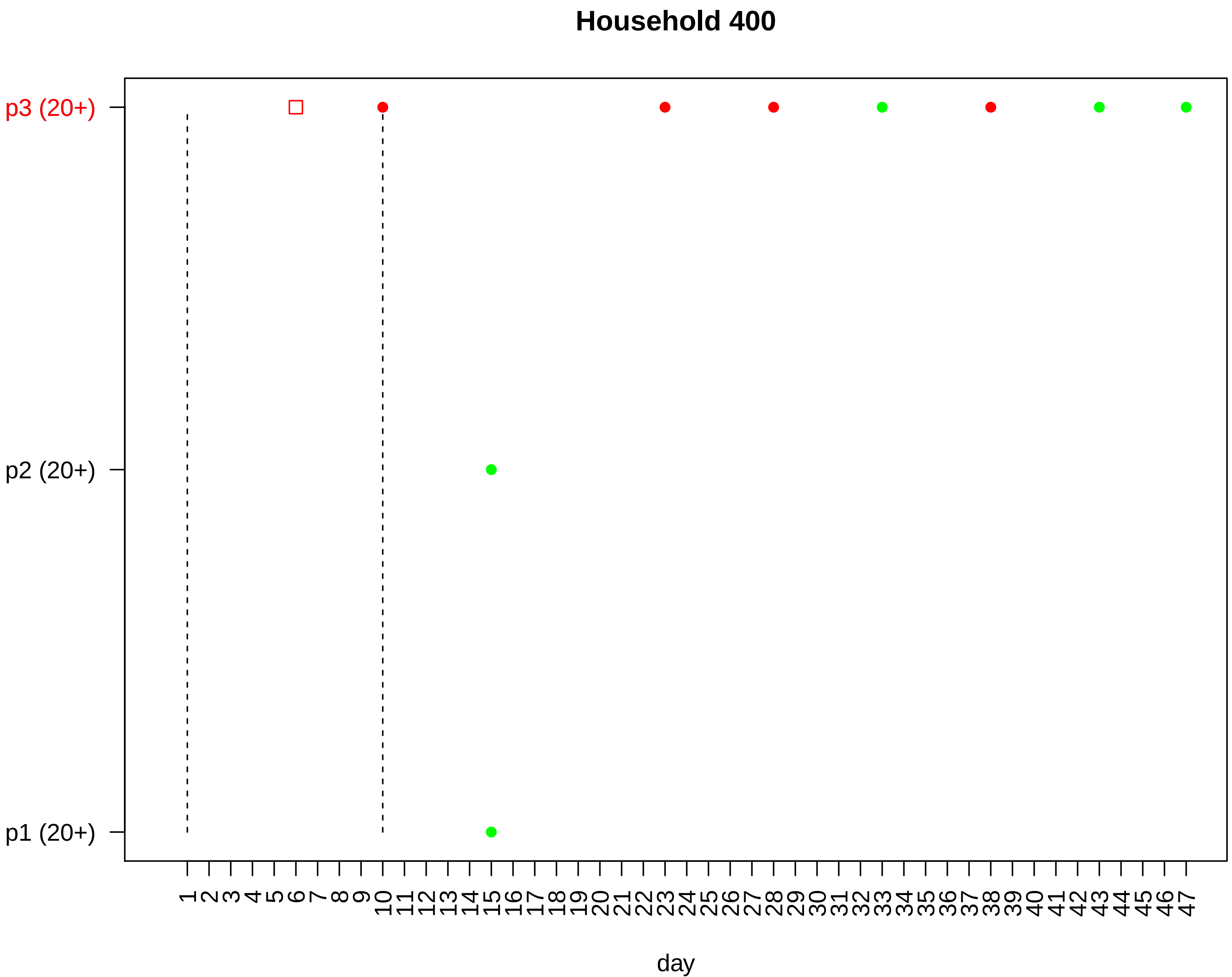
Household 400
p3 (20+)
p2 (20+)
p1 (20+)
1 2 3 4 5 6 7 8 9 10 11 12 13 14 15 16 17 18 19 20 21 22 23 24 25 26 27 28 29 30 31 32 33 34 35 36 37 38 39 40 41 42 43 44 45 46 47
day

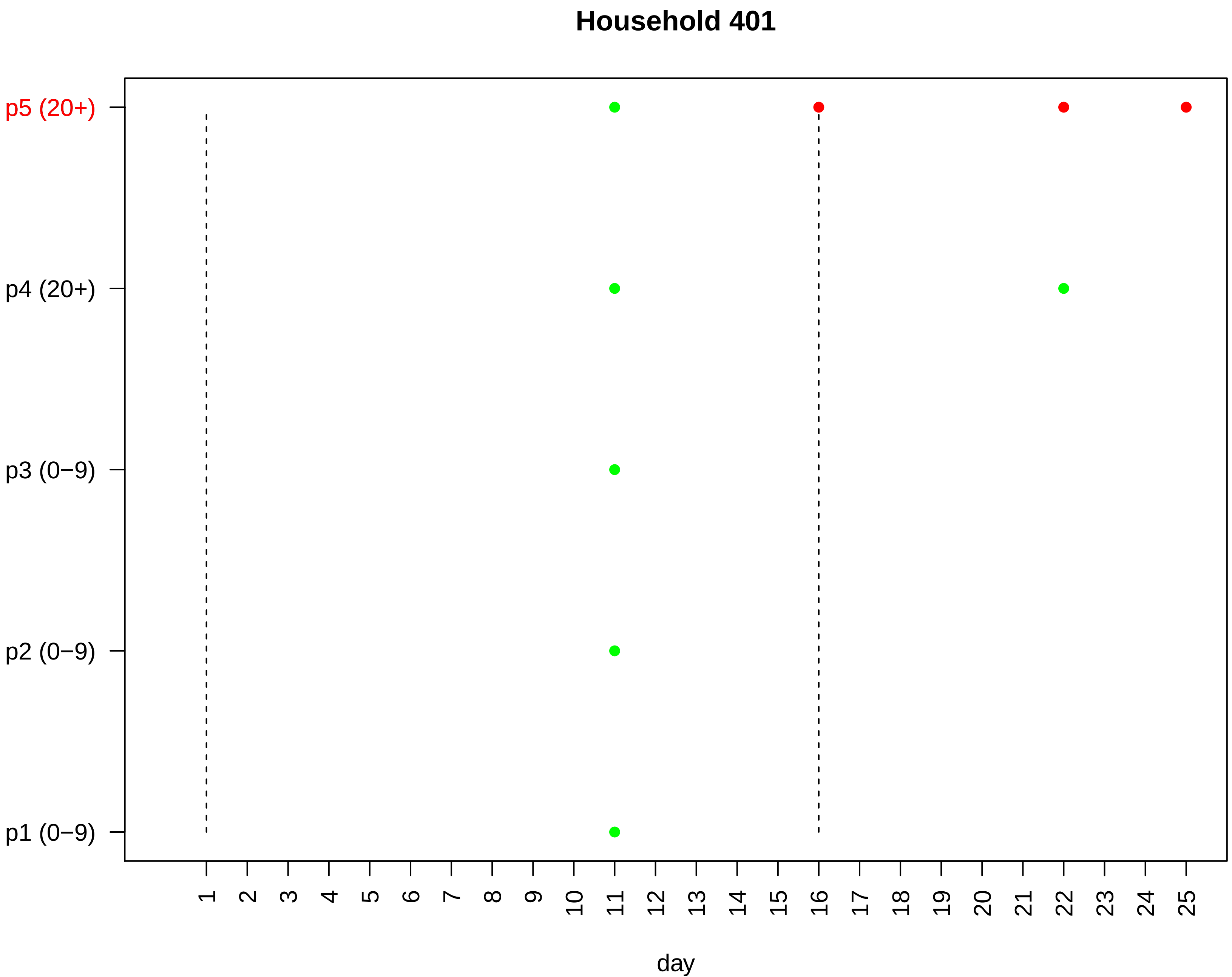

Household 401
p5 (20+)
p4 (20+)
p3 (0−9)
p2 (0−9)
p1 (0−9)
1 2 3 4 5 6 7 8 9 10 11 12 13 14 15 16 17 18 19 20 21 22 23 24 25
day

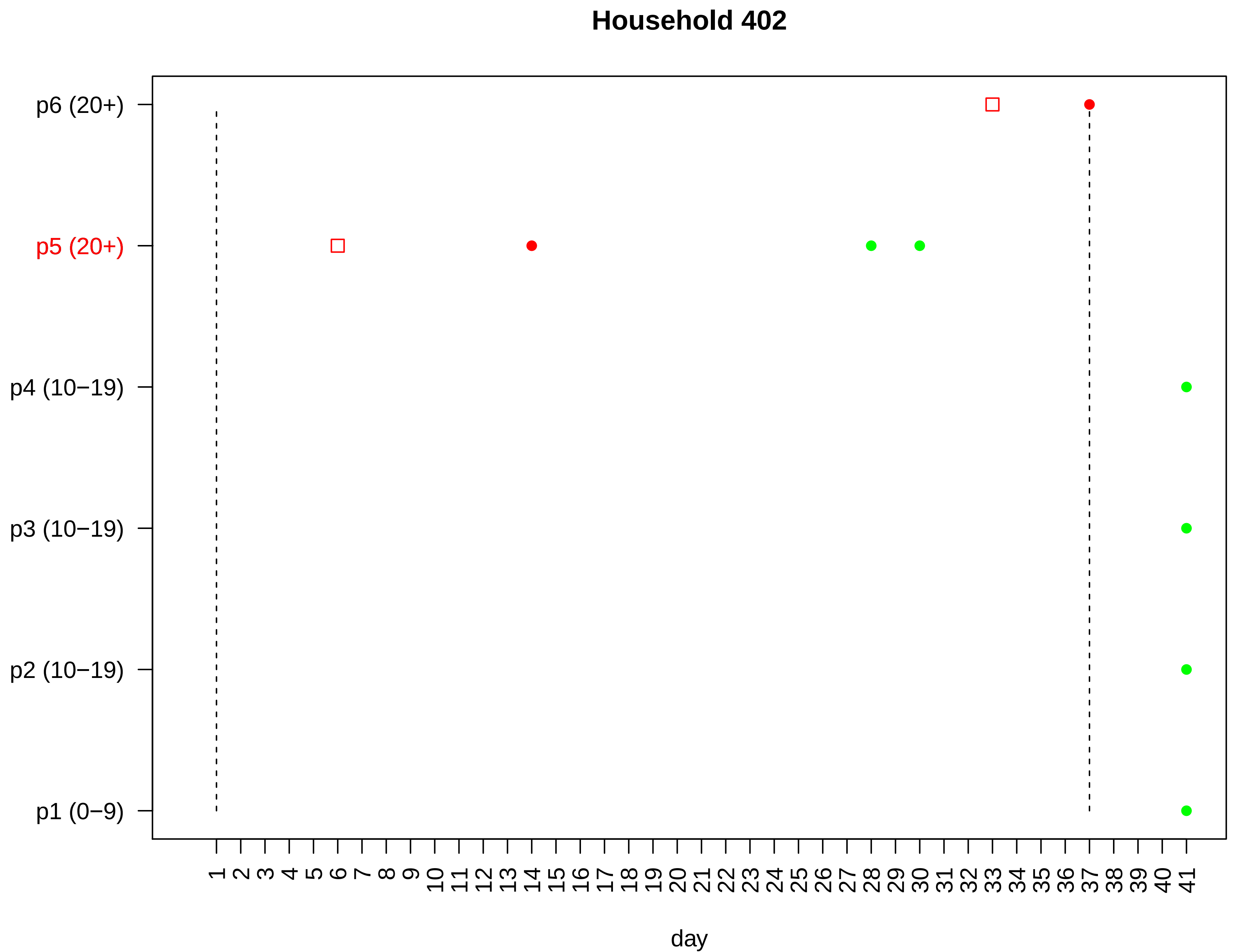

Household 402
p6 (20+)
p5 (20+)
p4 (10−19)
p3 (10−19)
p2 (10−19)
p1 (0−9)
1 2 3 4 5 6 7 8 9 10 11 12 13 14 15 16 17 18 19 20 21 22 23 24 25 26 27 28 29 30 31 32 33 34 35 36 37 38 39 40 41
day

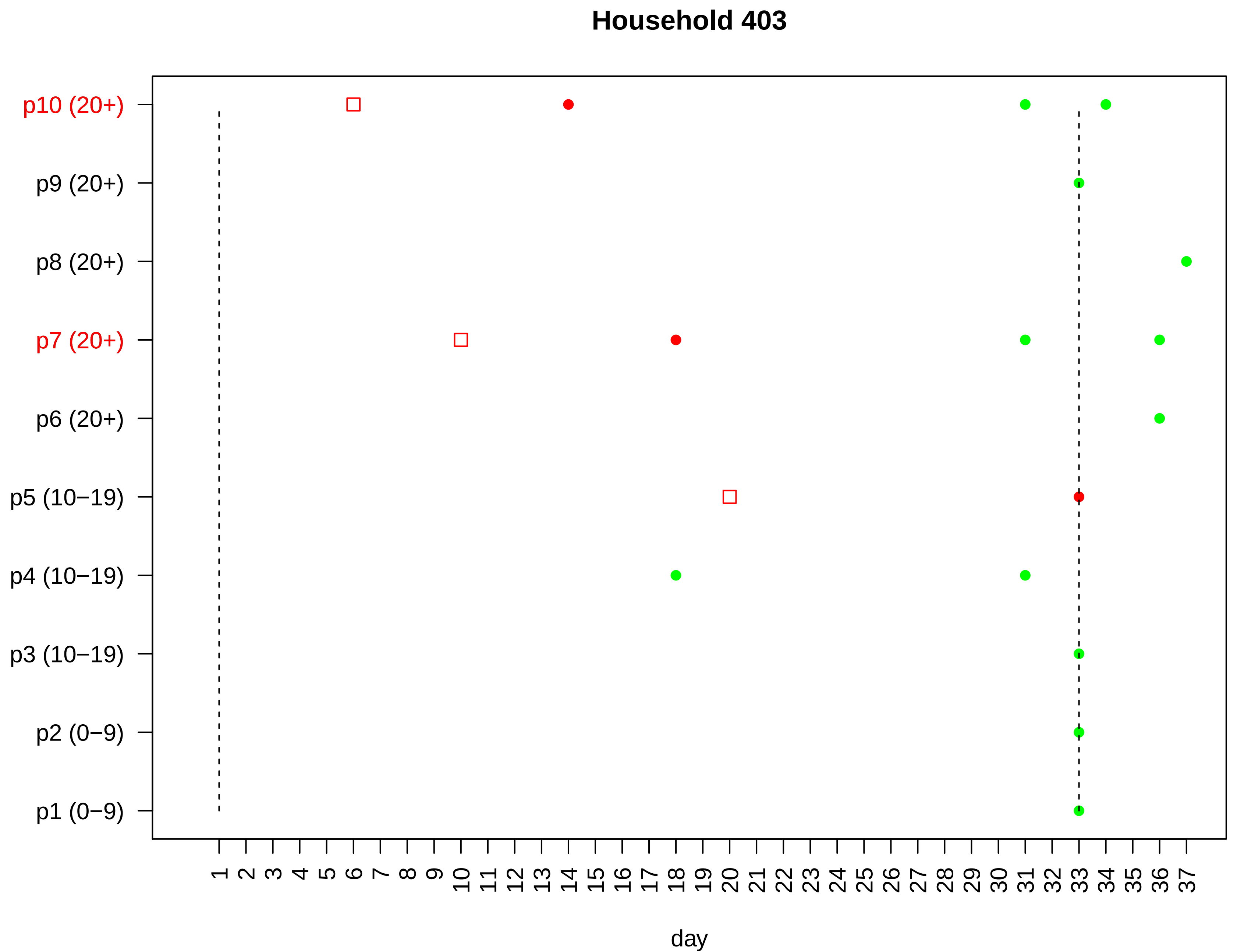

Household 403
p10 (20+)
p9 (20+)
p8 (20+)
p7 (20+)
p6 (20+)
p5 (10−19)
p4 (10−19)
p3 (10−19)
p2 (0−9)
p1 (0−9)
1 2 3 4 5 6 7 8 9 10 11 12 13 14 15 16 17 18 19 20 21 22 23 24 25 26 27 28 29 30 31 32 33 34 35 36 37
day

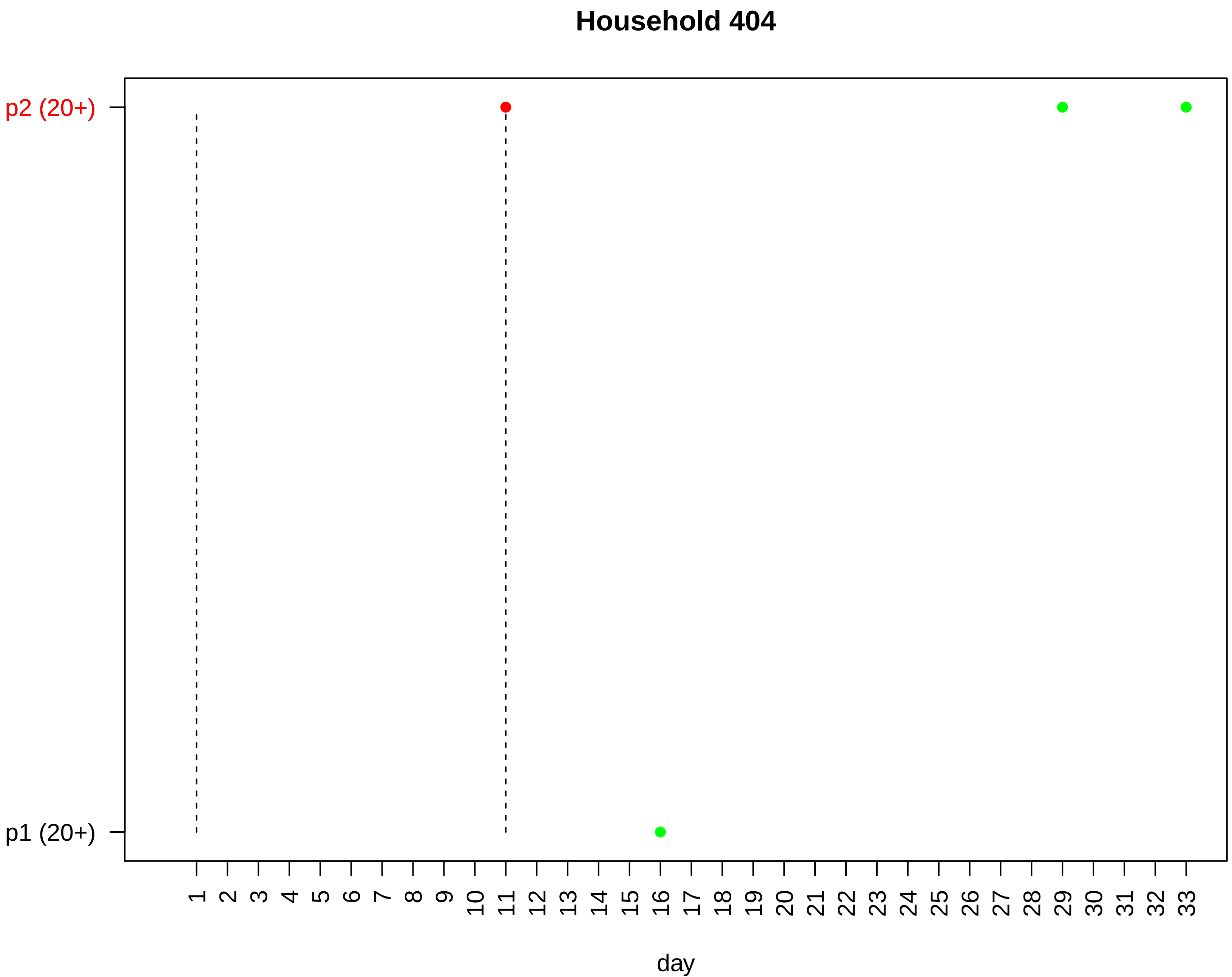

Household 404
p2 (20+)
p1 (20+)
day

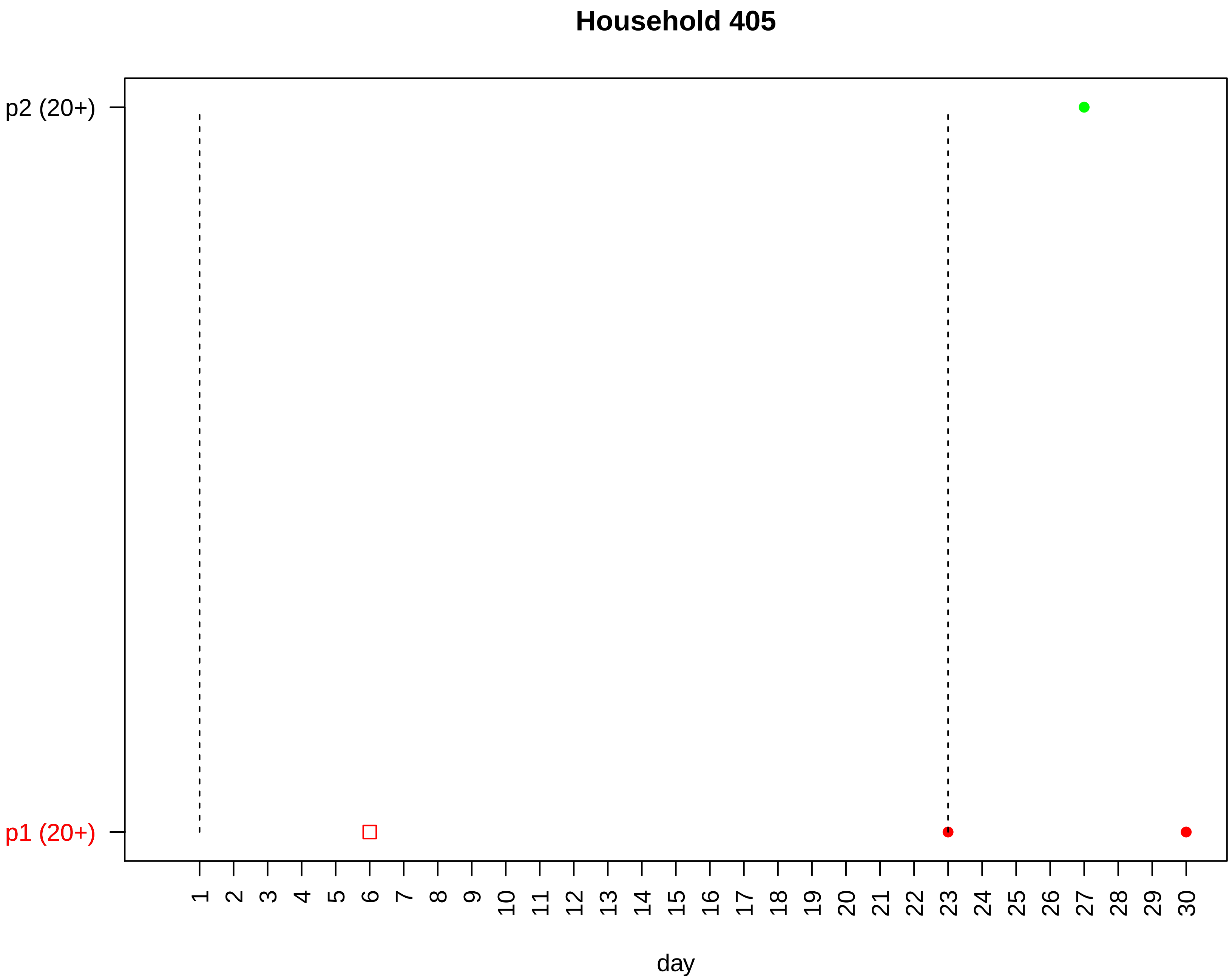
Household 405
p2 (20+)
p1 (20+)
1 2 3 4 5 6 7 8 9 10 11 12 13 14 15 16 17 18 19 20 21 22 23 24 25 26 27 28 29 30
day

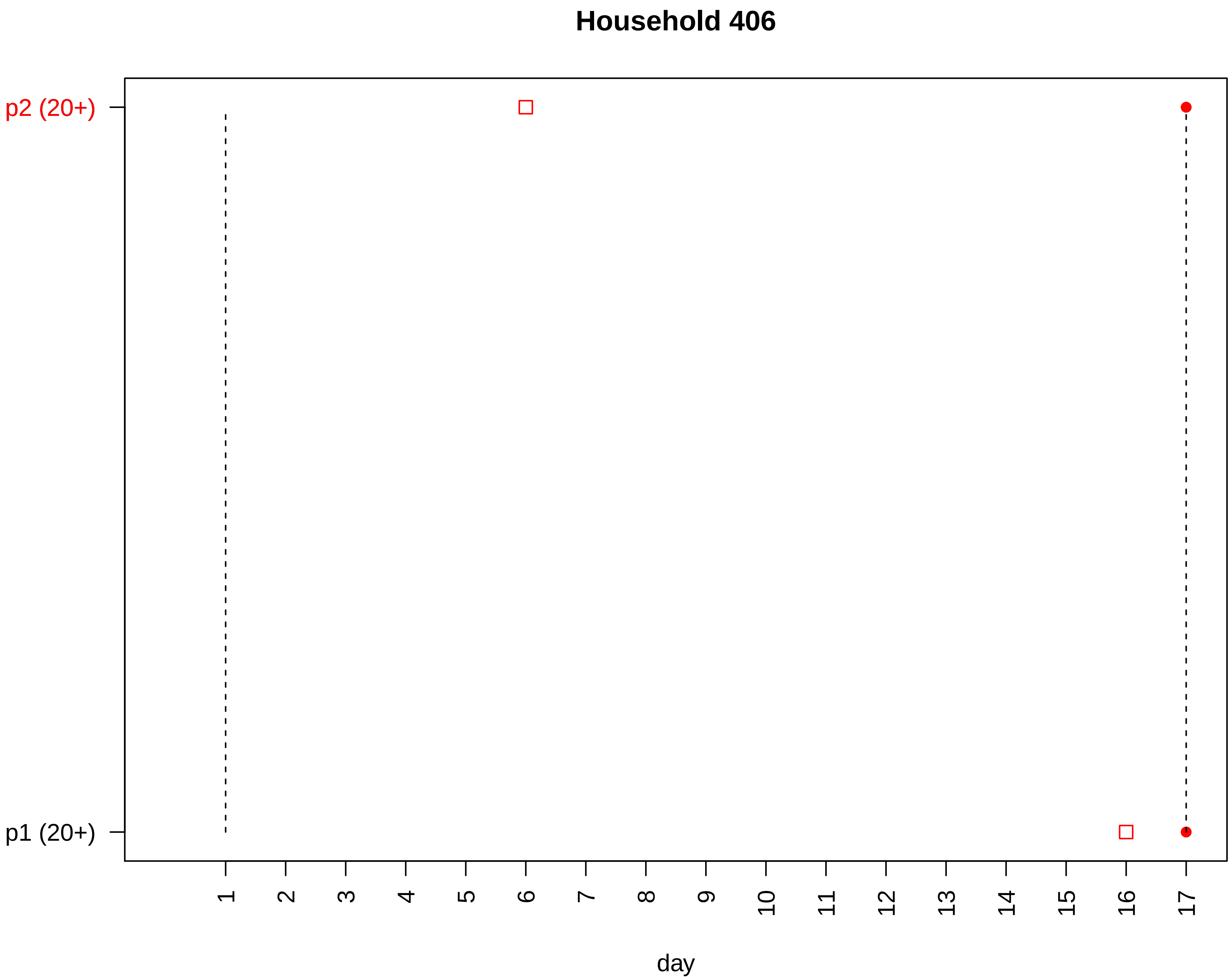
Household 406
p2 (20+)
p1 (20+)
day

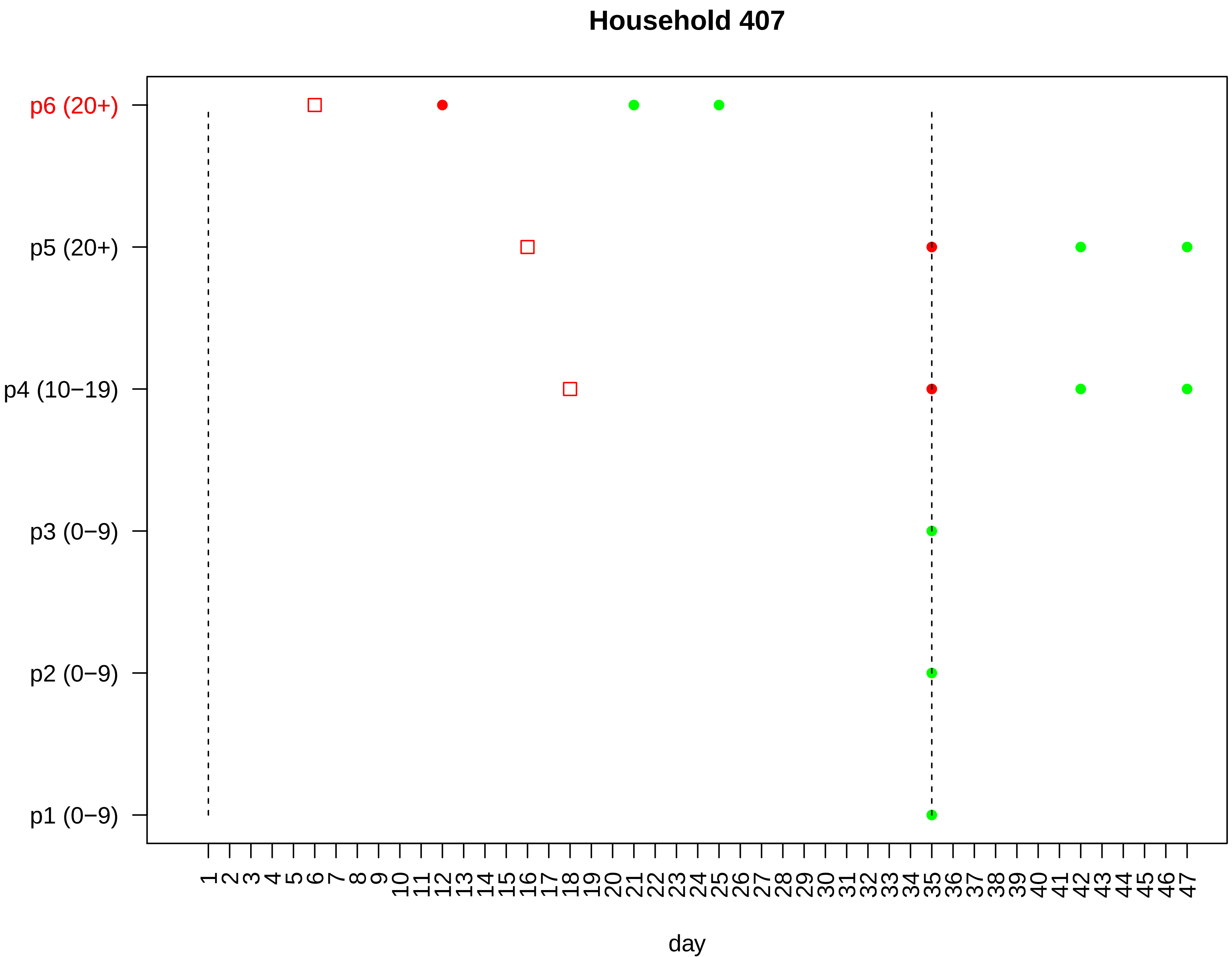

Household 407
p6 (20+)
p5 (20+)
p4 (10−19)
p3 (0−9)
p2 (0−9)
p1 (0−9)
1 2 3 4 5 6 7 8 9 10 11 12 13 14 15 16 17 18 19 20 21 22 23 24 25 26 27 28 29 30 31 32 33 34 35 36 37 38 39 40 41 42 43 44 45 46 47
day

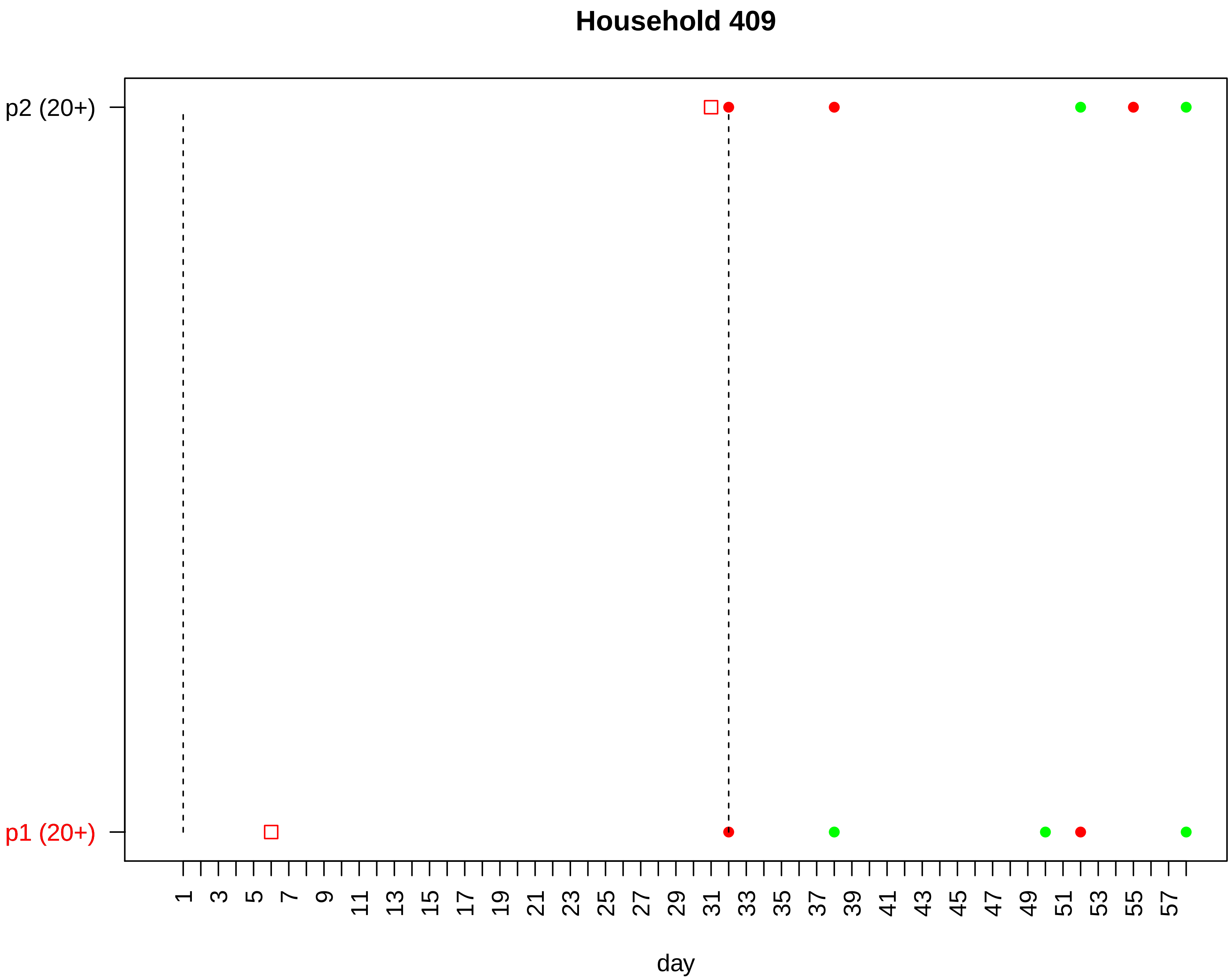
Household 409
p2 (20+)
p1 (20+)
1
3
5
7
9
11
13
15
17
19
21
23
25
27
29
31
33
35
37
39
41
43
45
47
49
51
53
55
57
day

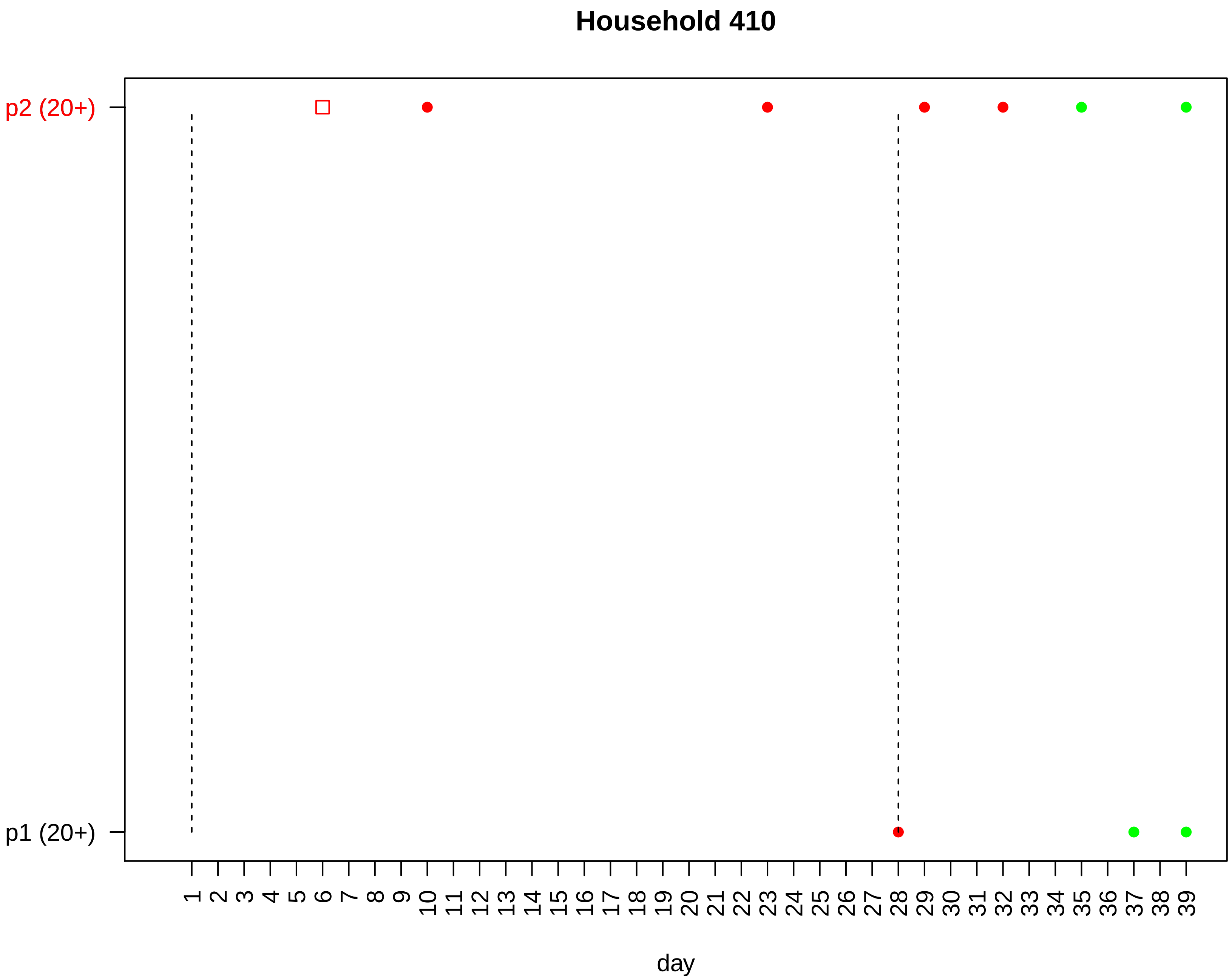

Household 410
p2 (20+)
p1 (20+)
day

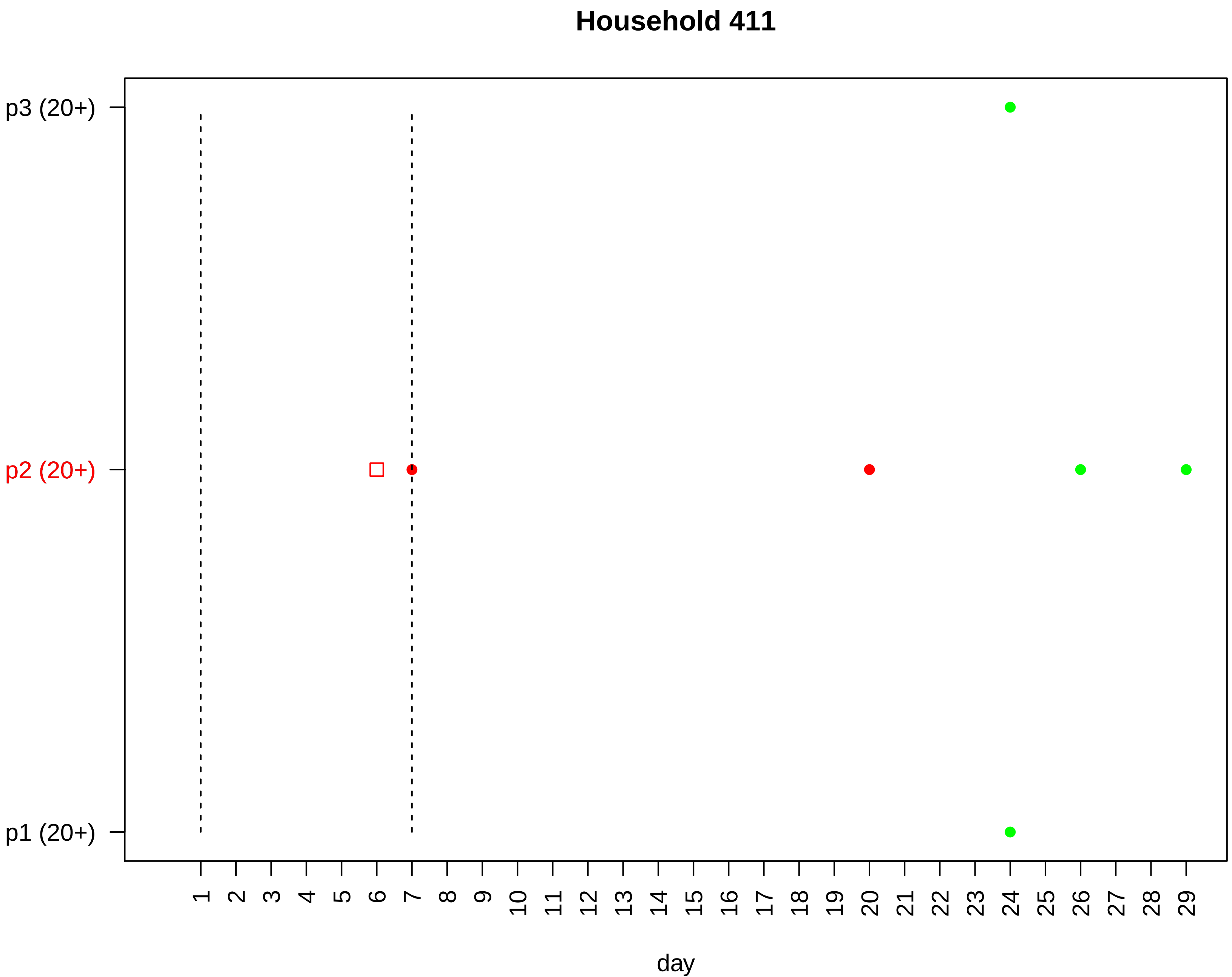

Household 411
p3 (20+)
p2 (20+)
p1 (20+)
day

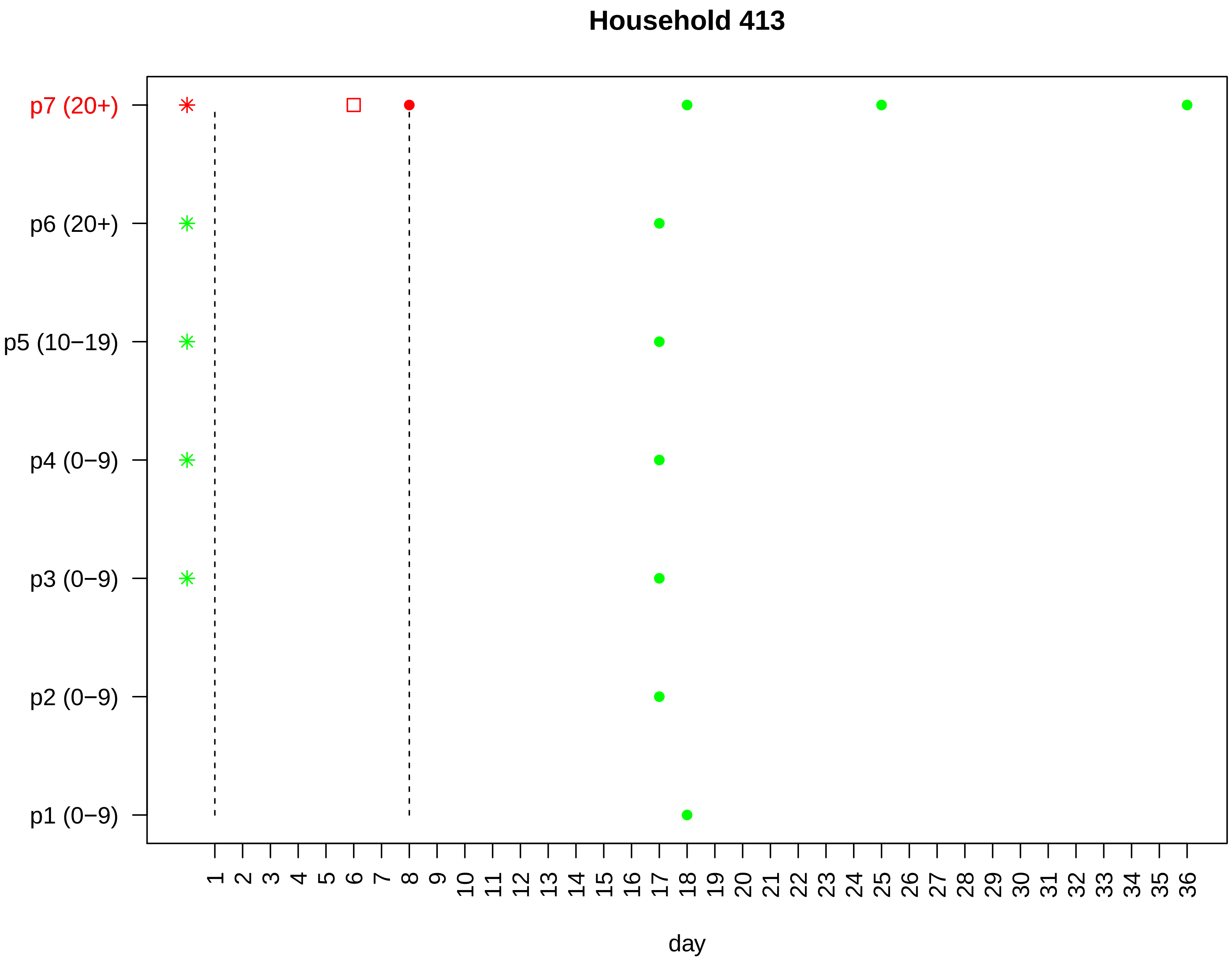

Household 413
p7 (20+)
p6 (20+)
p5 (10−19)
p4 (0−9)
p3 (0−9)
p2 (0−9)
p1 (0−9)
1 2 3 4 5 6 7 8 9 10 11 12 13 14 15 16 17 18 19 20 21 22 23 24 25 26 27 28 29 30 31 32 33 34 35 36
day

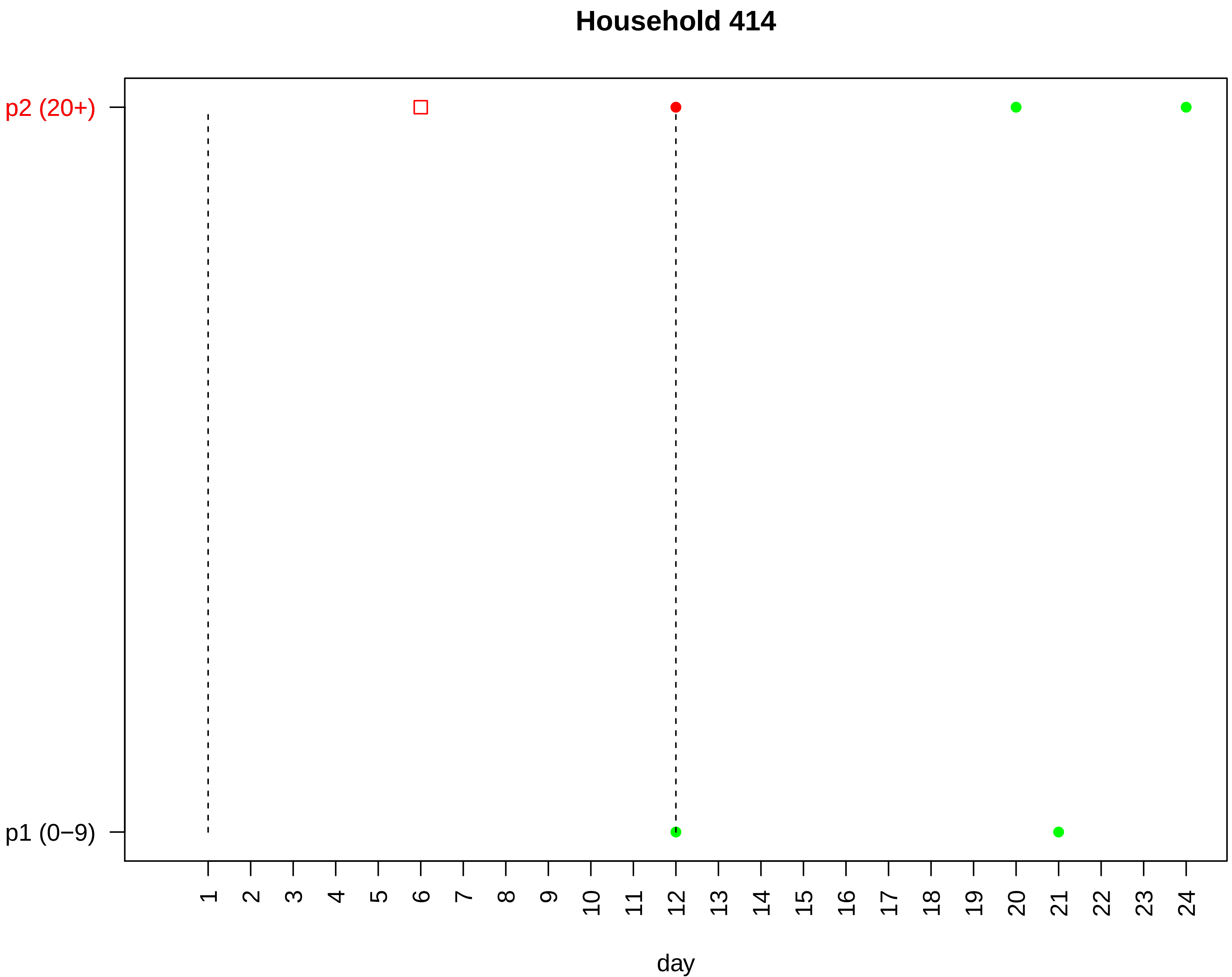

Household 414
p2 (20+)
p1 (0−9)
day

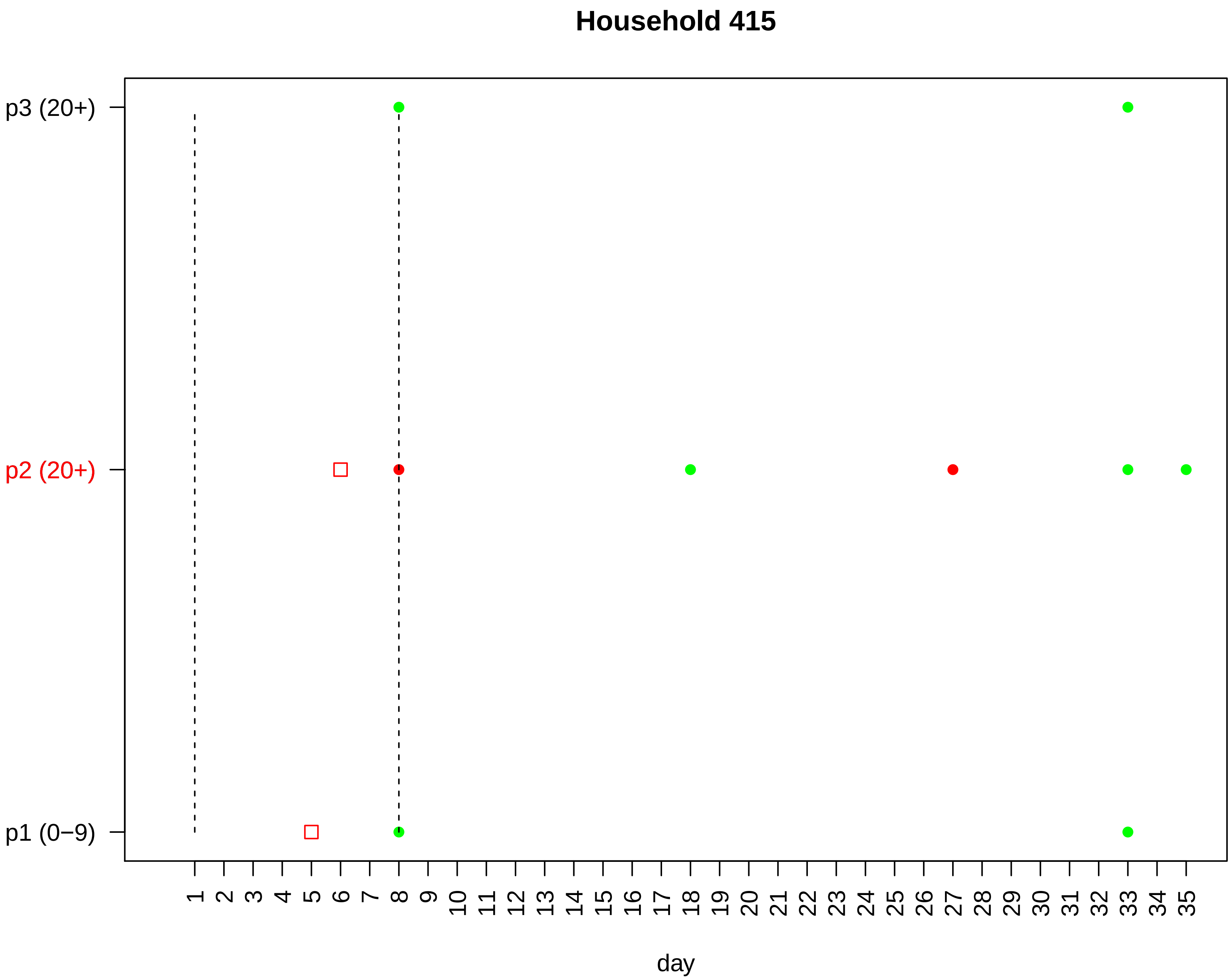
Household 415
p3 (20+)
p2 (20+)
p1 (0−9)
1 2 3 4 5 6 7 8 9 10 11 12 13 14 15 16 17 18 19 20 21 22 23 24 25 26 27 28 29 30 31 32 33 34 35
day

### Household 416

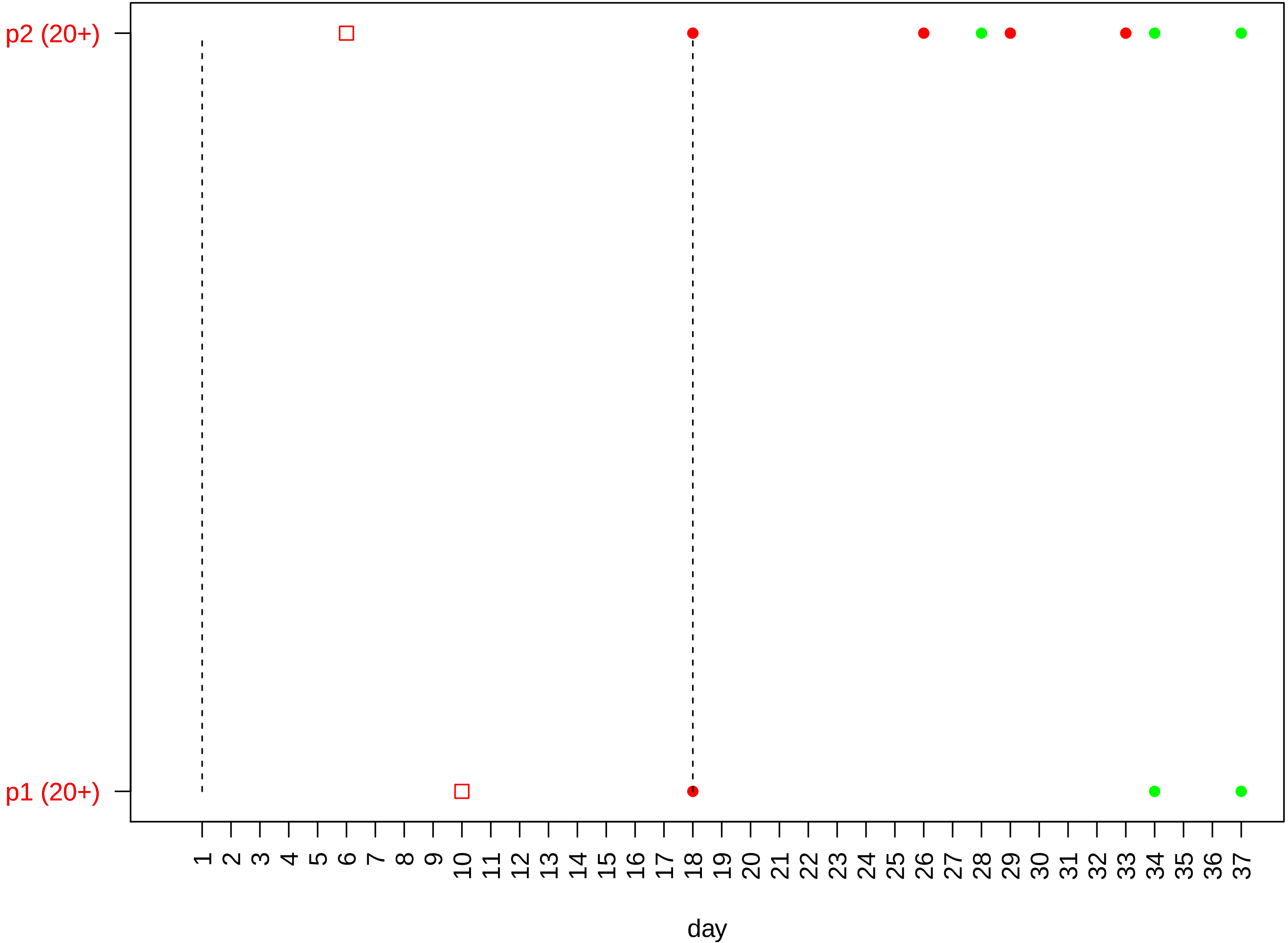

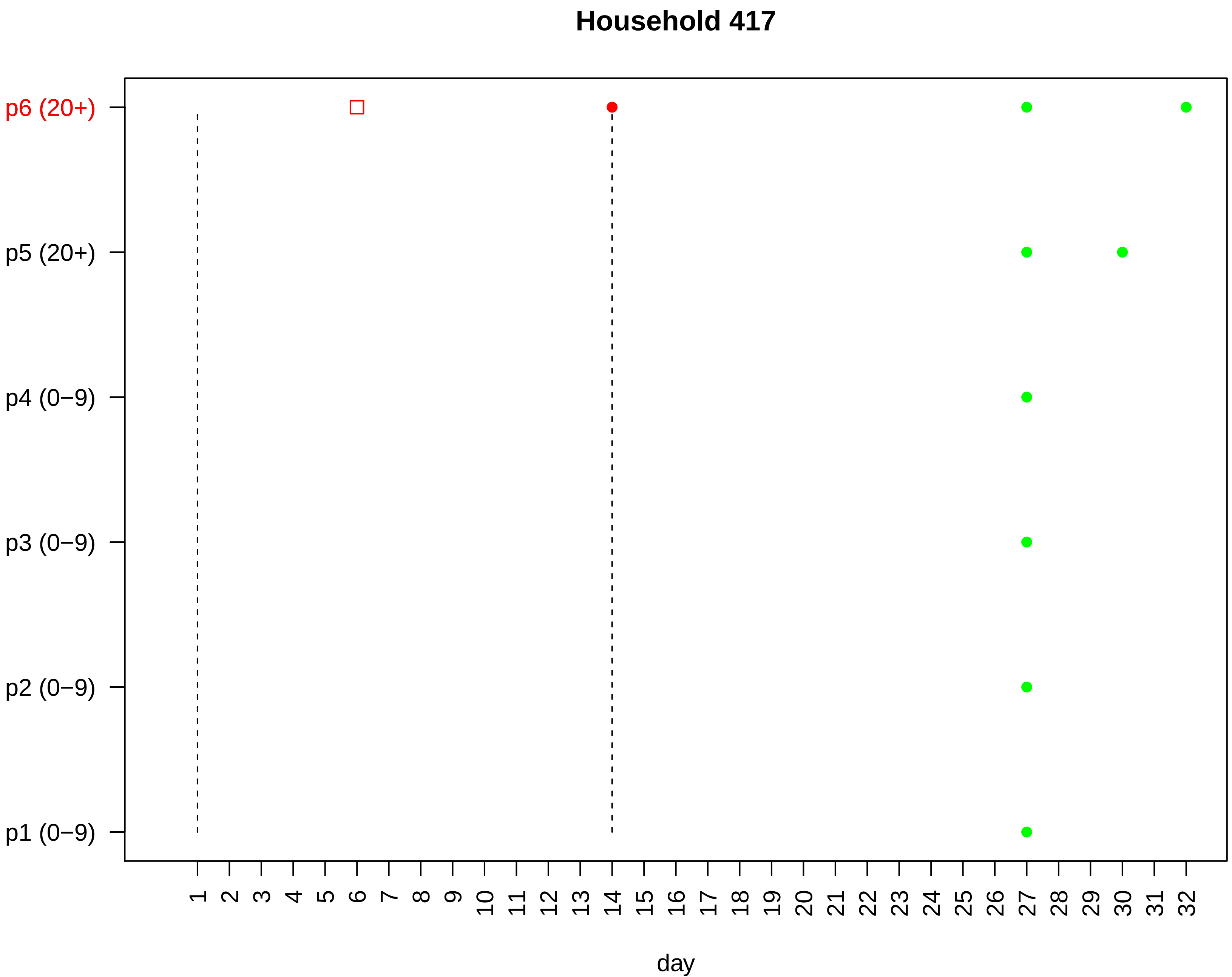

Household 417
p6 (20+)
p5 (20+)
p4 (0−9)
p3 (0−9)
p2 (0−9)
p1 (0−9)
1 2 3 4 5 6 7 8 9 10 11 12 13 14 15 16 17 18 19 20 21 22 23 24 25 26 27 28 29 30 31 32
day

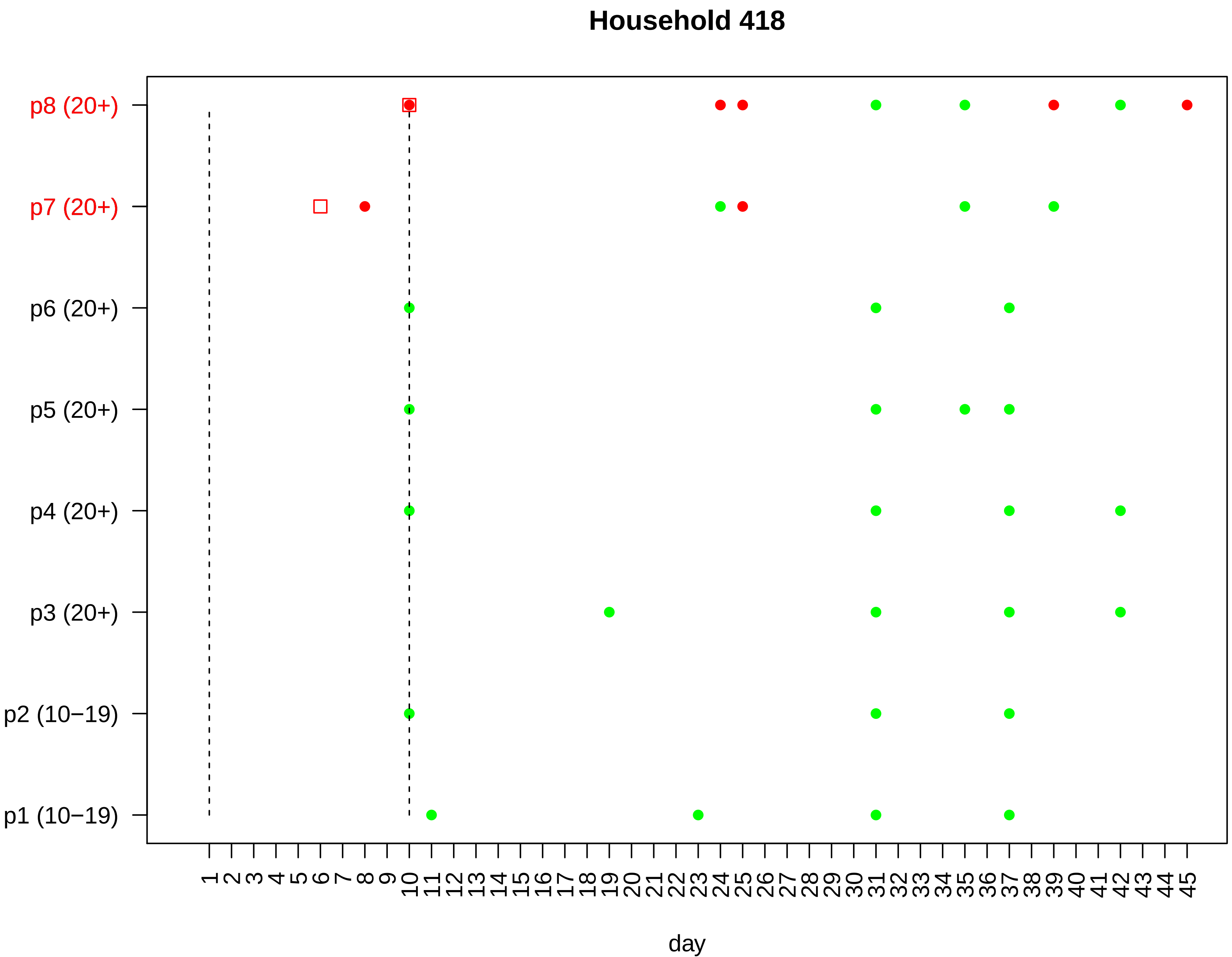

Household 418
p8 (20+)
p7 (20+)
p6 (20+)
p5 (20+)
p4 (20+)
p3 (20+)
p2 (10−19)
p1 (10−19)
1 2 3 4 5 6 7 8 9 10 11 12 13 14 15 16 17 18 19 20 21 22 23 24 25 26 27 28 29 30 31 32 33 34 35 36 37 38 39 40 41 42 43 44 45
day

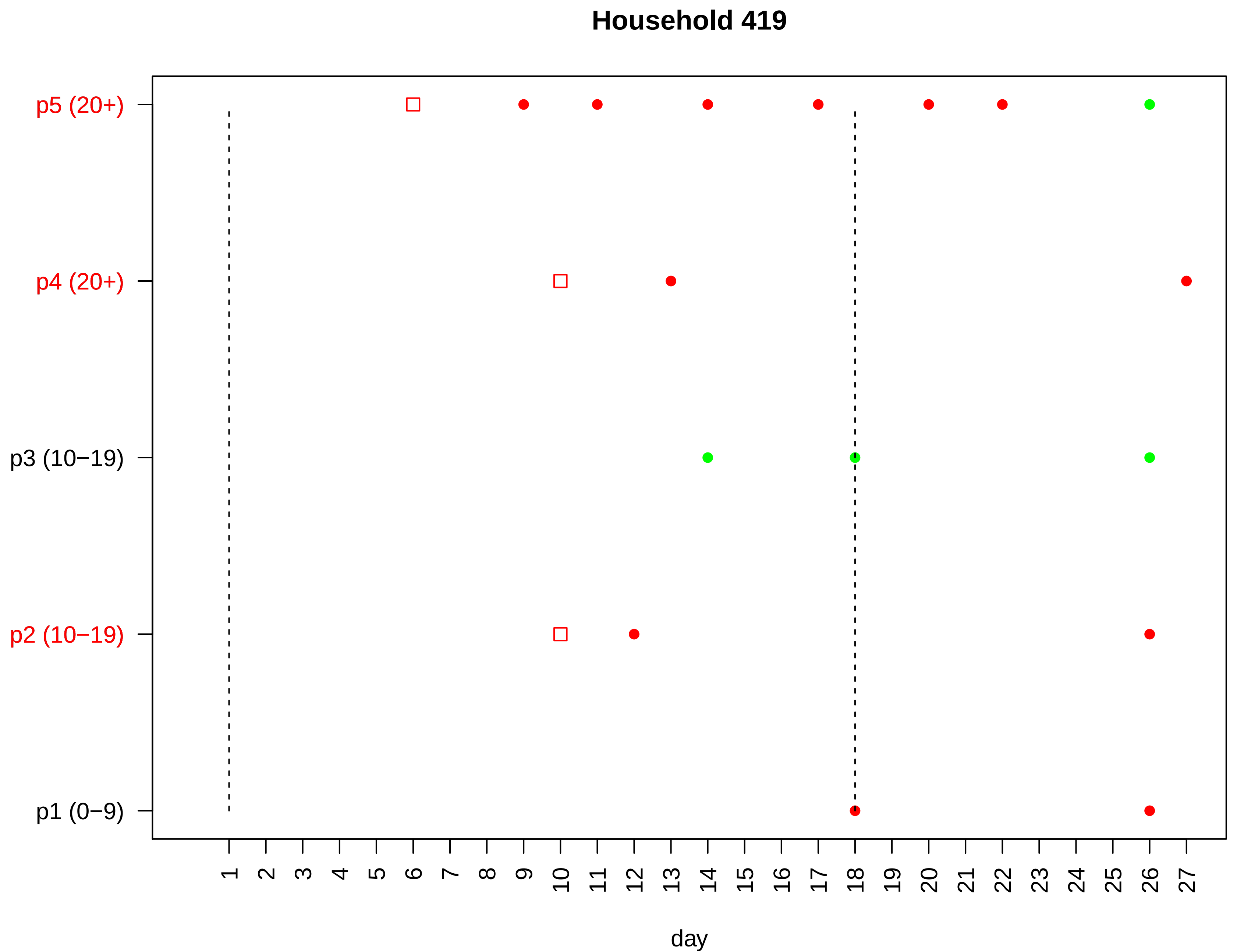

Household 419
p5 (20+)
p4 (20+)
p3 (10−19)
p2 (10−19)
p1 (0−9)
1 2 3 4 5 6 7 8 9 10 11 12 13 14 15 16 17 18 19 20 21 22 23 24 25 26 27
day

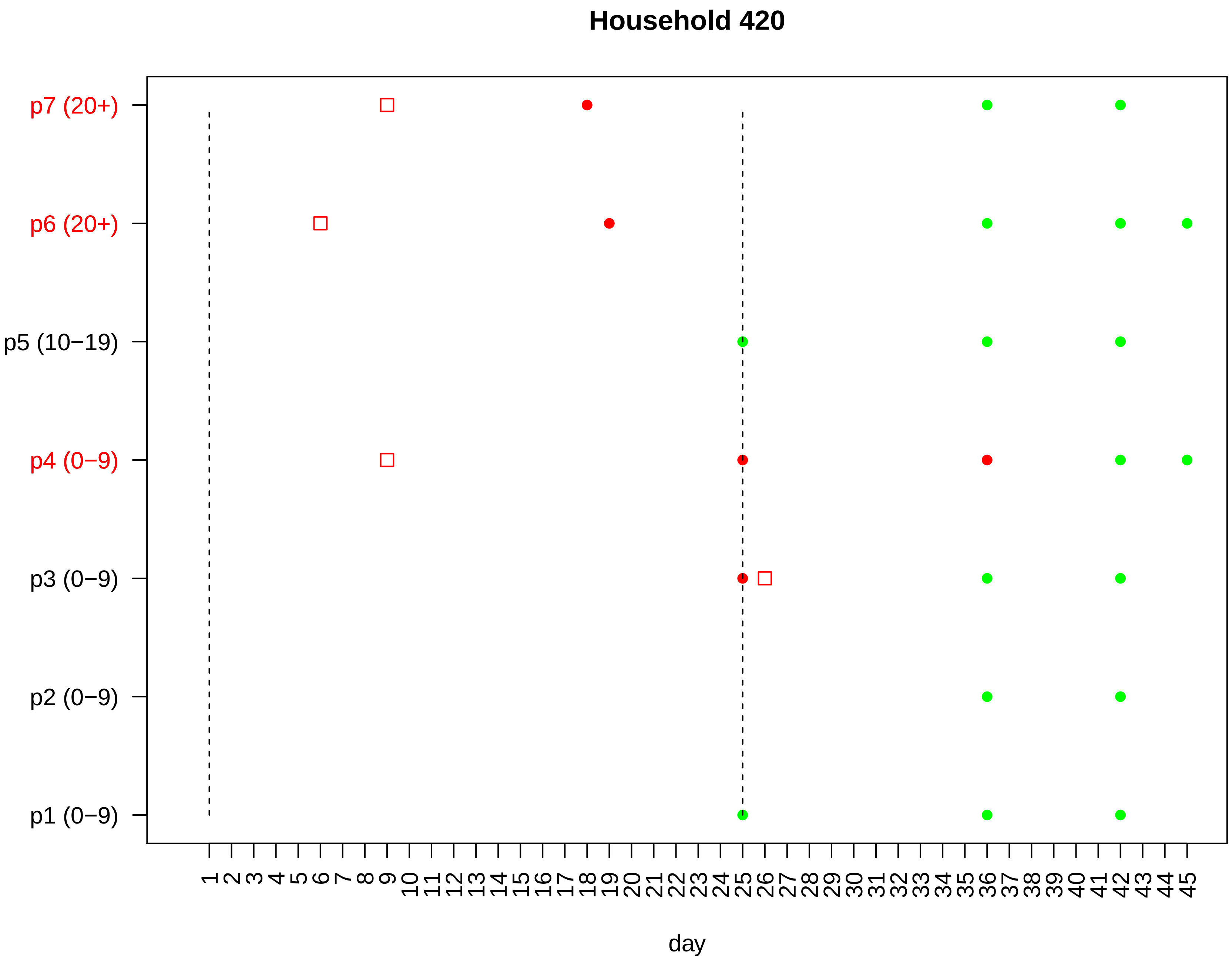

Household 420
p7 (20+)
p6 (20+)
p5 (10−19)
p4 (0−9)
p3 (0−9)
p2 (0−9)
p1 (0−9)
1 2 3 4 5 6 7 8 9 10 11 12 13 14 15 16 17 18 19 20 21 22 23 24 25 26 27 28 29 30 31 32 33 34 35 36 37 38 39 40 41 42 43 44 45
day

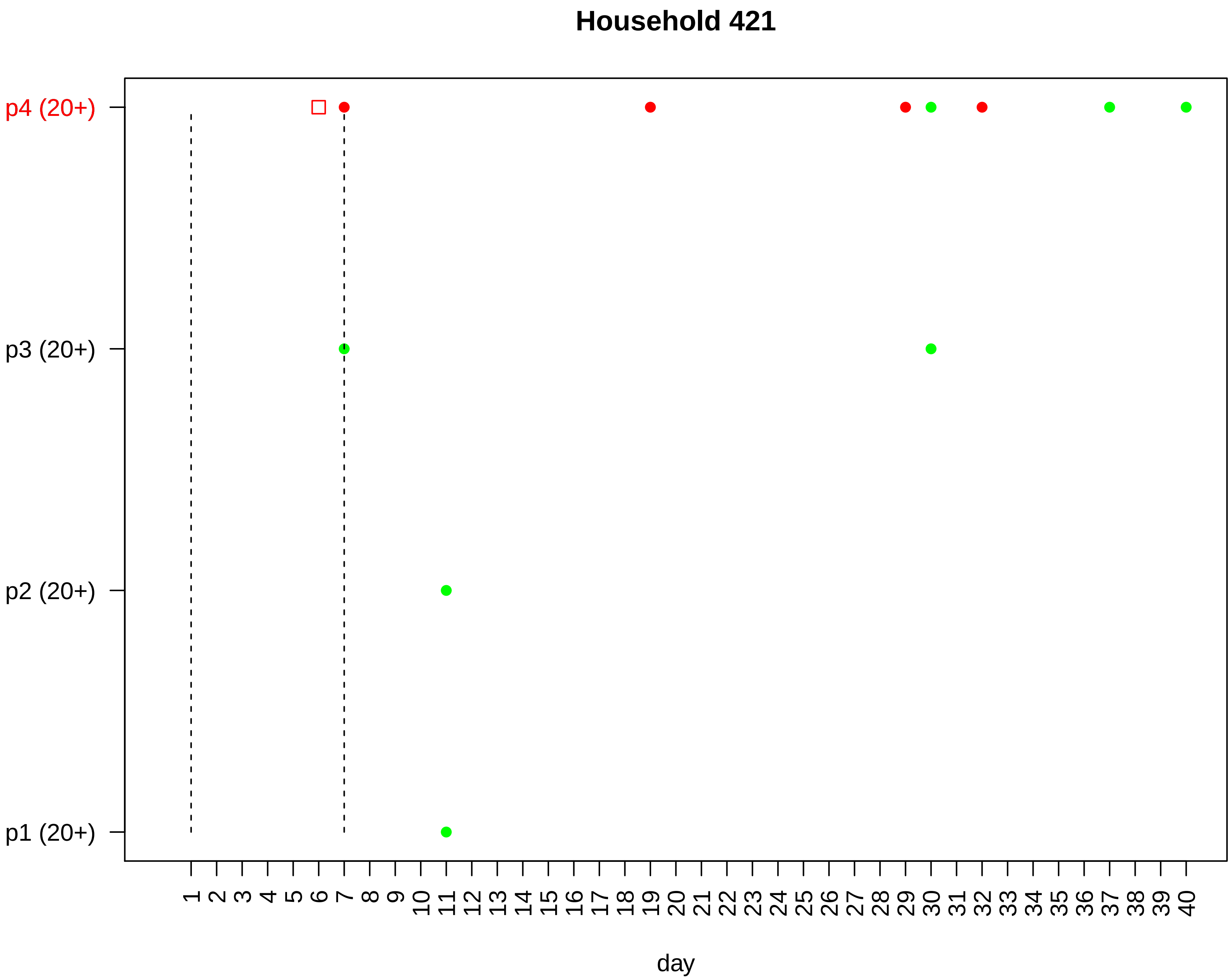
Household 421
p4 (20+)
p3 (20+)
p2 (20+)
p1 (20+)
1 2 3 4 5 6 7 8 9 10 11 12 13 14 15 16 17 18 19 20 21 22 23 24 25 26 27 28 29 30 31 32 33 34 35 36 37 38 39 40
day

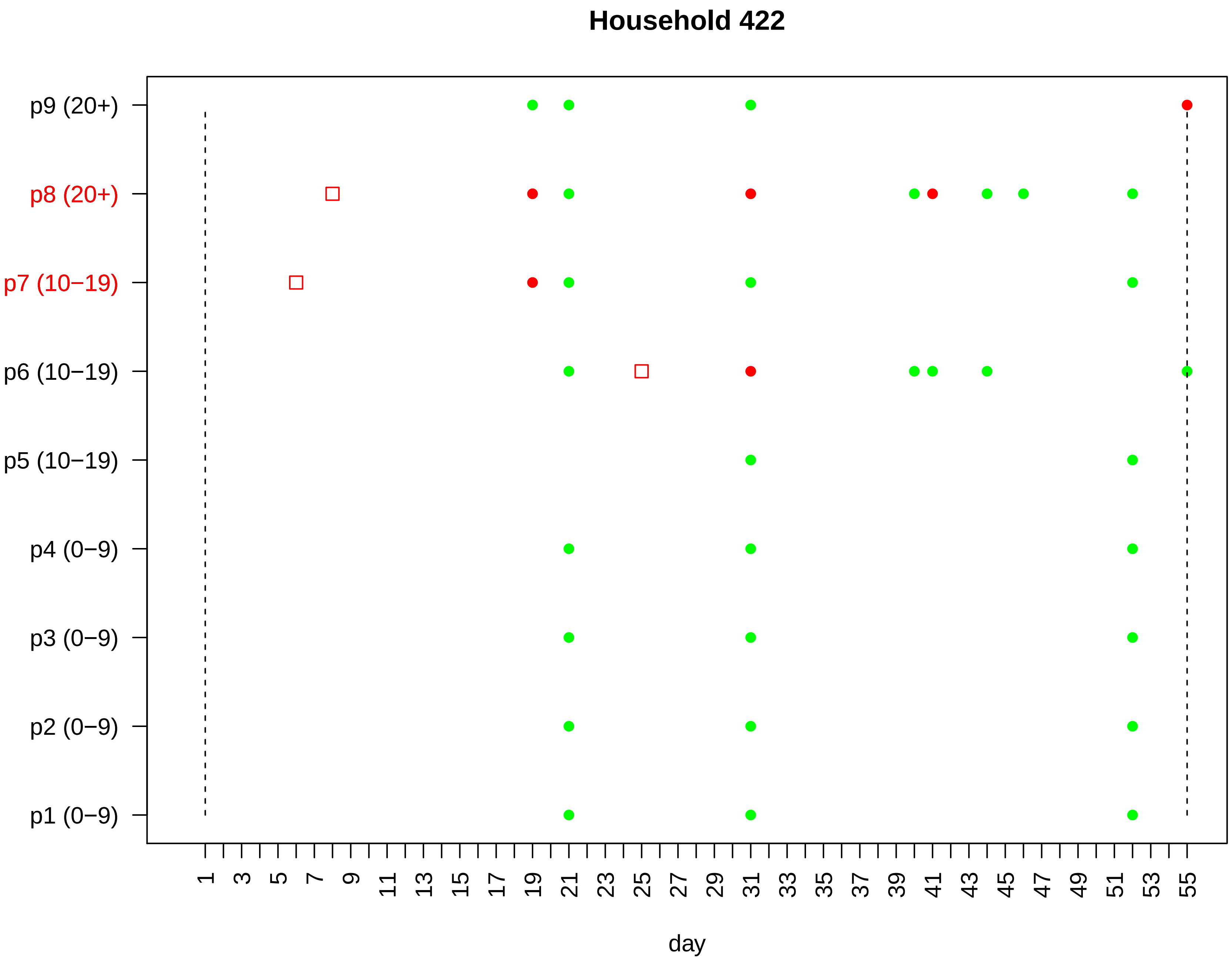

Household 422
p9 (20+)
p8 (20+)
p7 (10−19)
p6 (10−19)
p5 (10−19)
p4 (0−9)
p3 (0−9)
p2 (0−9)
p1 (0−9)
1 3 5 7 9 11 13 15 17 19 21 23 25 27 29 31 33 35 37 39 41 43 45 47 49 51 53 55
day

#### Household 423

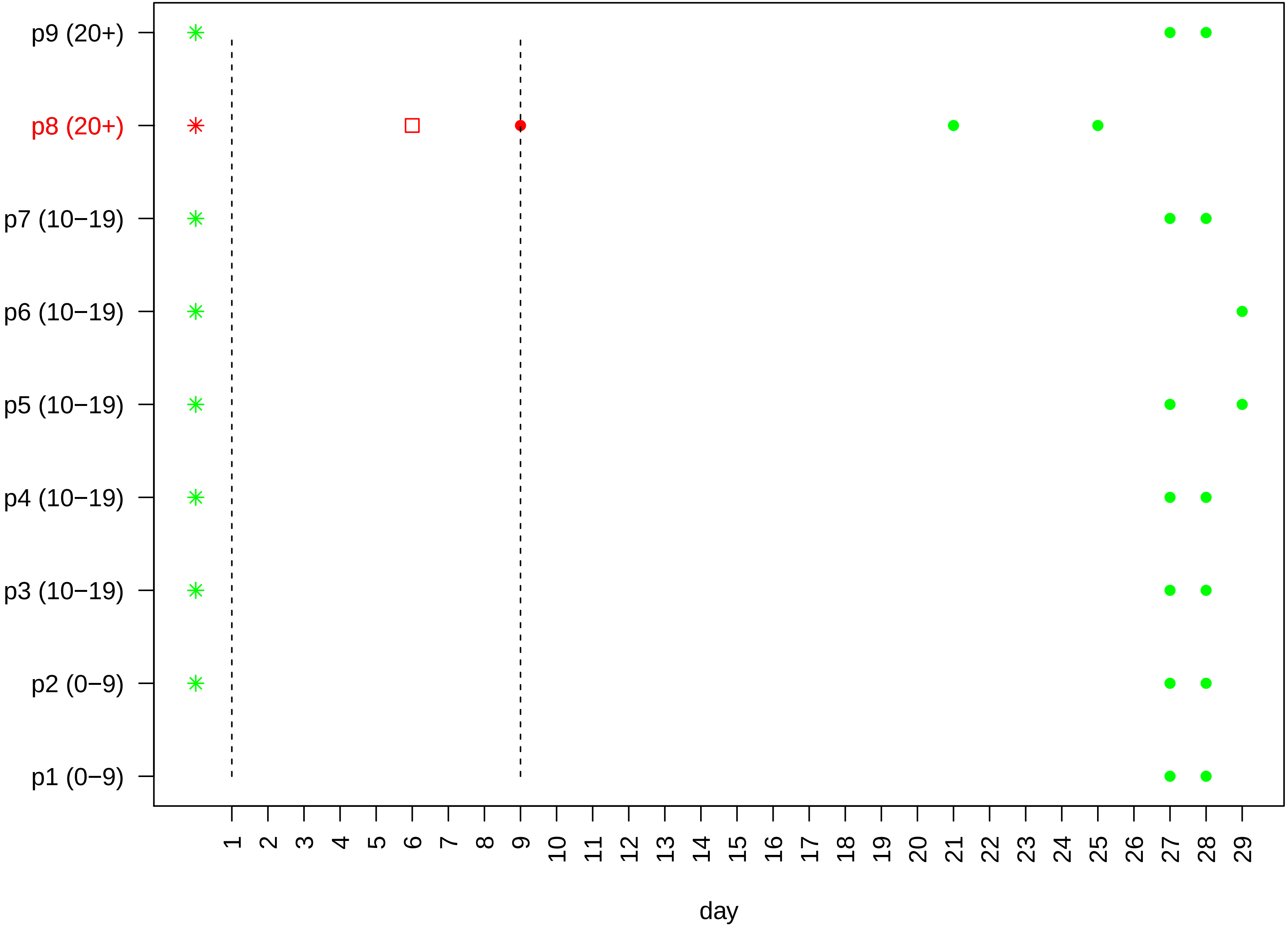

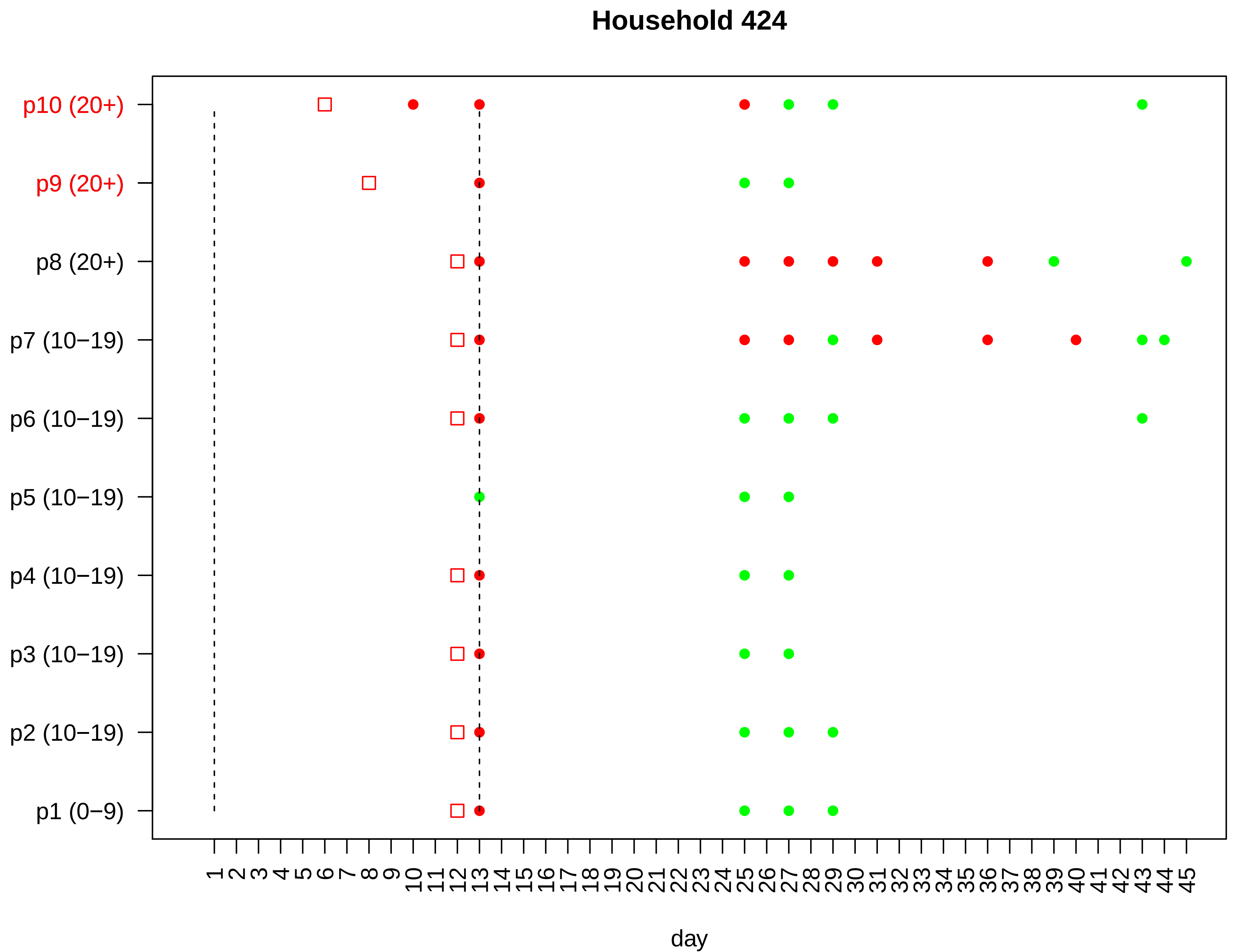

Household 424
p10 (20+)
p9 (20+)
p8 (20+)
p7 (10−19)
p6 (10−19)
p5 (10−19)
p4 (10−19)
p3 (10−19)
p2 (10−19)
p1 (0−9)
1 2 3 4 5 6 7 8 9 10 11 12 13 14 15 16 17 18 19 20 21 22 23 24 25 26 27 28 29 30 31 32 33 34 35 36 37 38 39 40 41 42 43 44 45
day

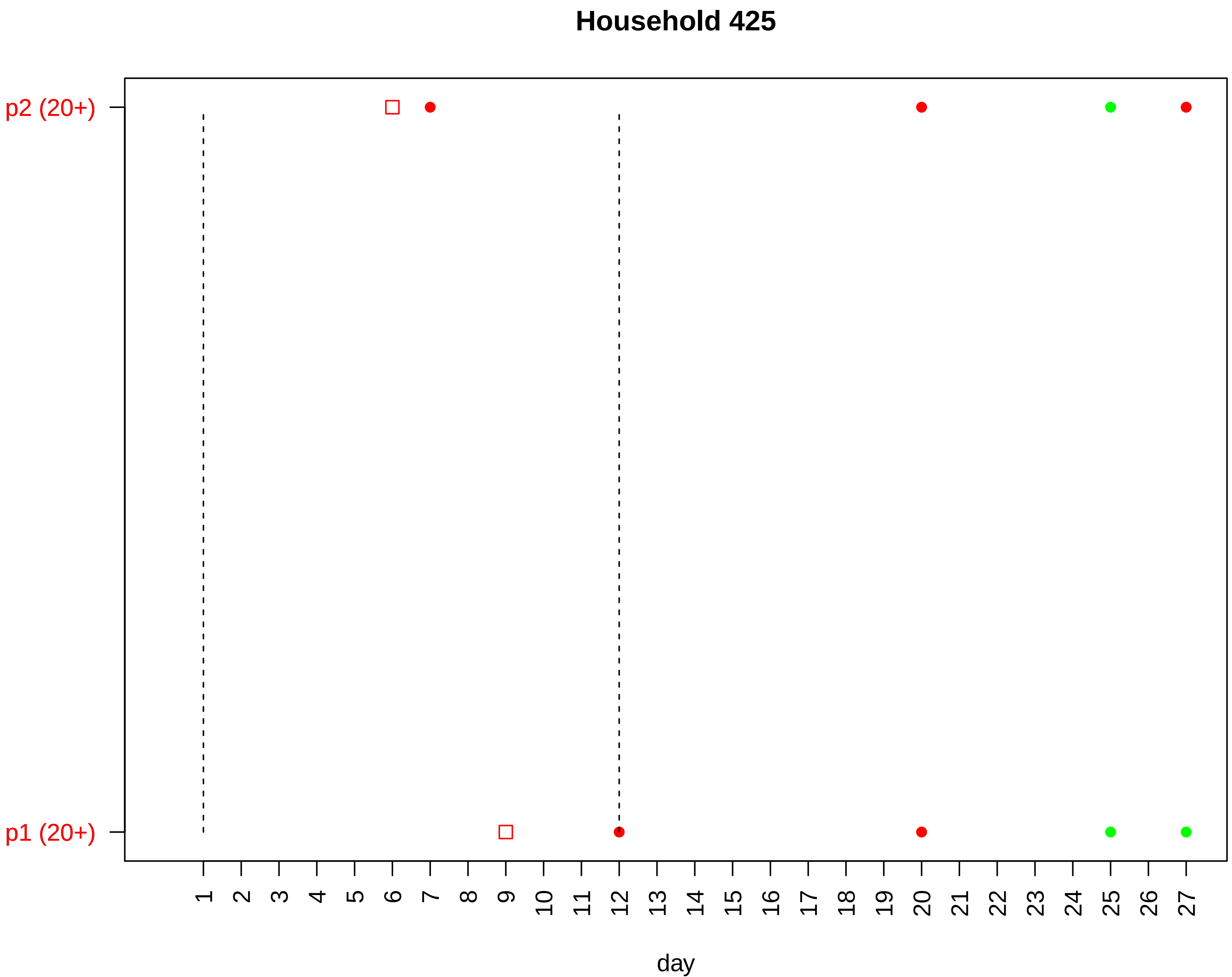

Household 425
p2 (20+)
p1 (20+)
day

#### Household 426

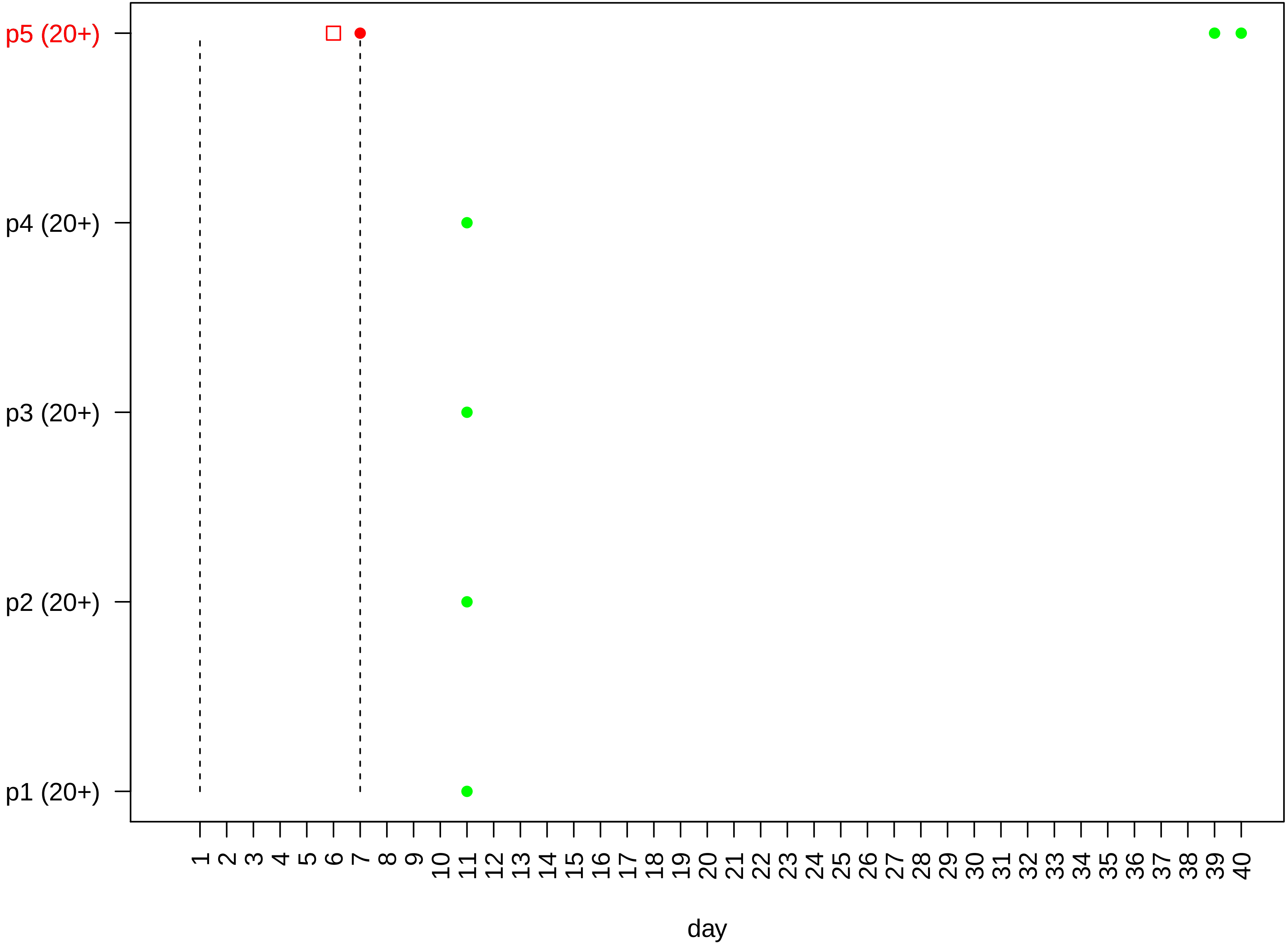

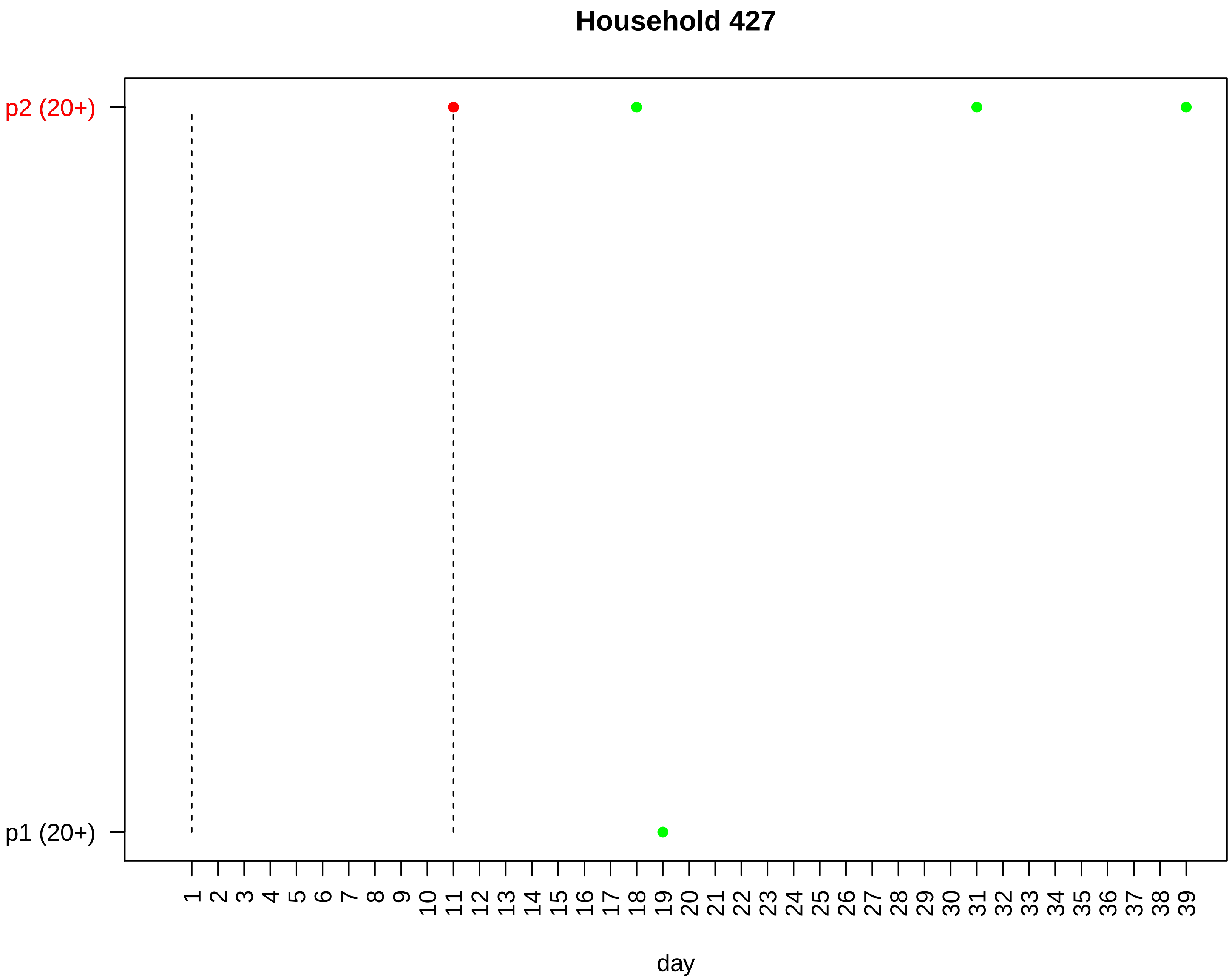

Household 427
p2 (20+)
p1 (20+)
1 2 3 4 5 6 7 8 9 10 11 12 13 14 15 16 17 18 19 20 21 22 23 24 25 26 27 28 29 30 31 32 33 34 35 36 37 38 39
day

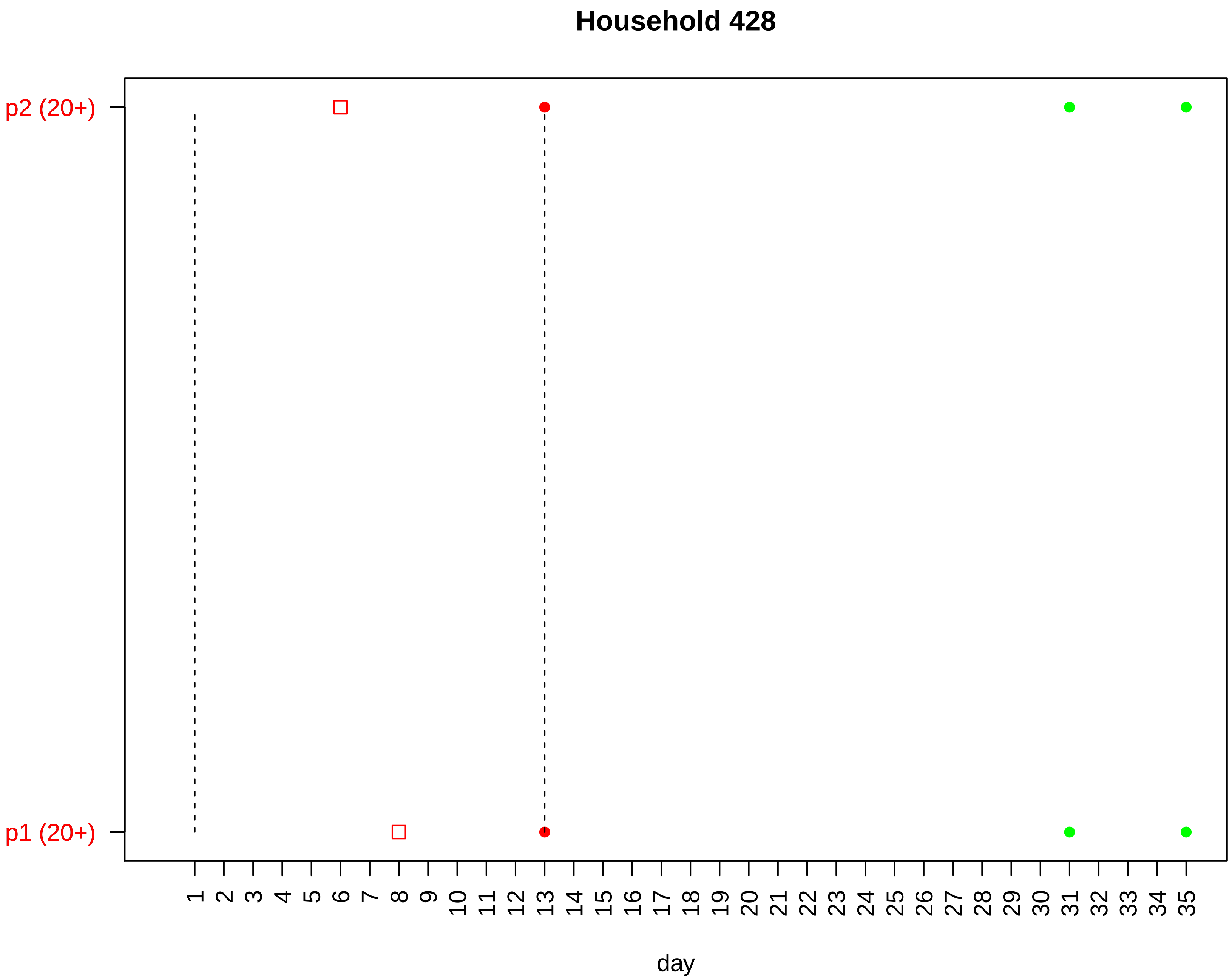

Household 428
p2 (20+)
p1 (20+)
1 2 3 4 5 6 7 8 9 10 11 12 13 14 15 16 17 18 19 20 21 22 23 24 25 26 27 28 29 30 31 32 33 34 35
day

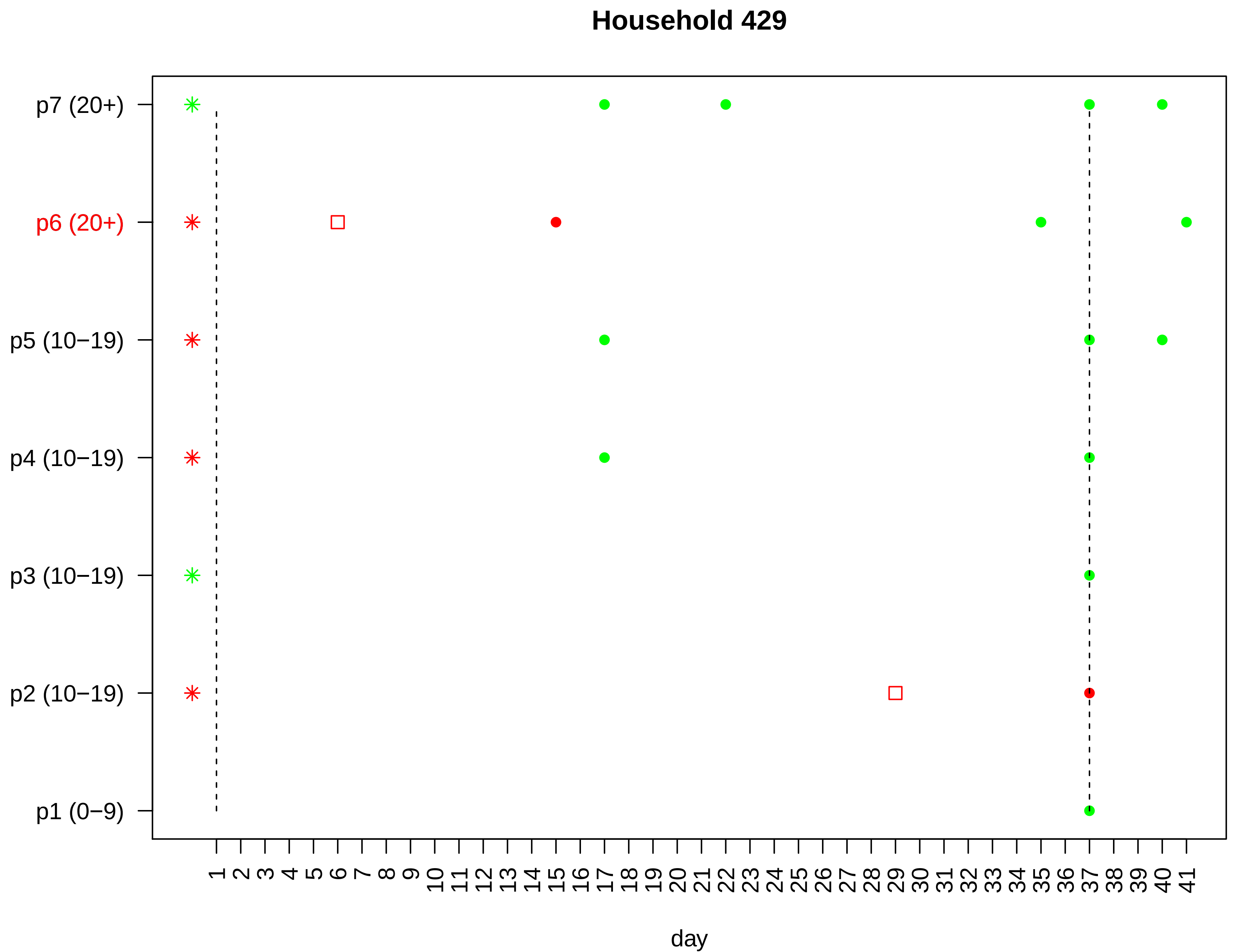

Household 429
p7 (20+)
p6 (20+)
p5 (10−19)
p4 (10−19)
p3 (10−19)
p2 (10−19)
p1 (0−9)
1 2 3 4 5 6 7 8 9 10 11 12 13 14 15 16 17 18 19 20 21 22 23 24 25 26 27 28 29 30 31 32 33 34 35 36 37 38 39 40 41
day

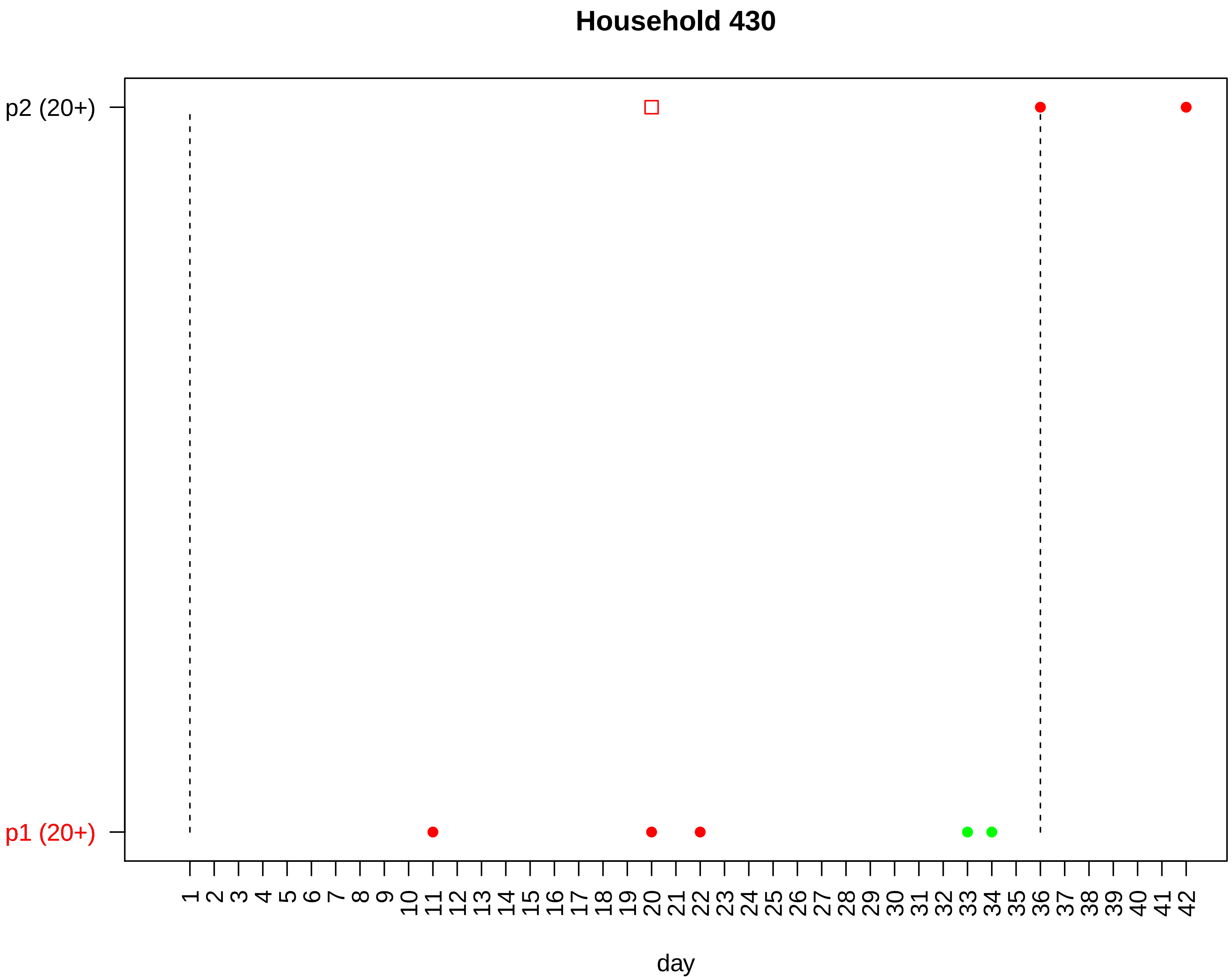

Household 430
p2 (20+)
p1 (20+)
1 2 3 4 5 6 7 8 9 10 11 12 13 14 15 16 17 18 19 20 21 22 23 24 25 26 27 28 29 30 31 32 33 34 35 36 37 38 39 40 41 42
day

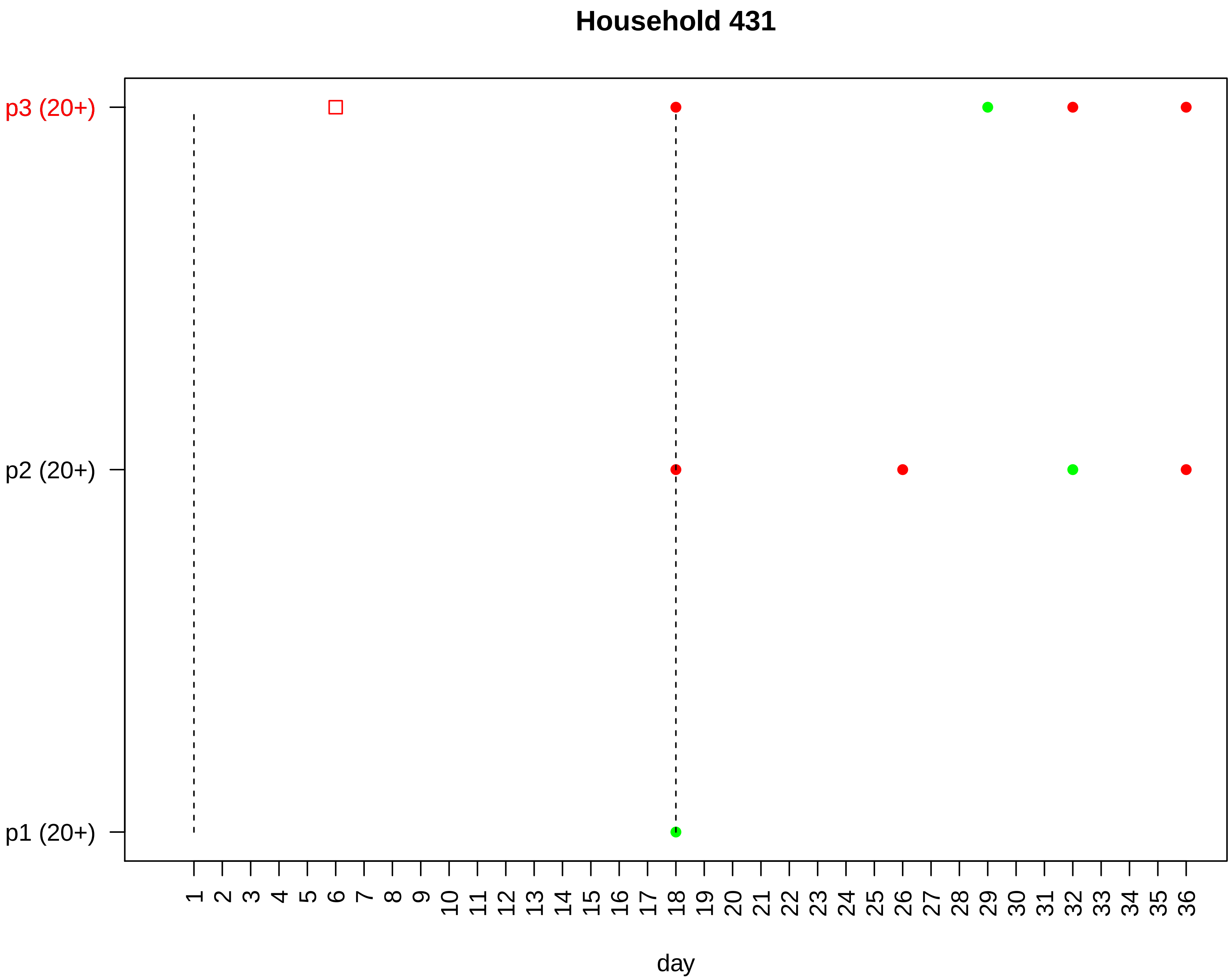
Household 431
p3 (20+)
p2 (20+)
p1 (20+)
day

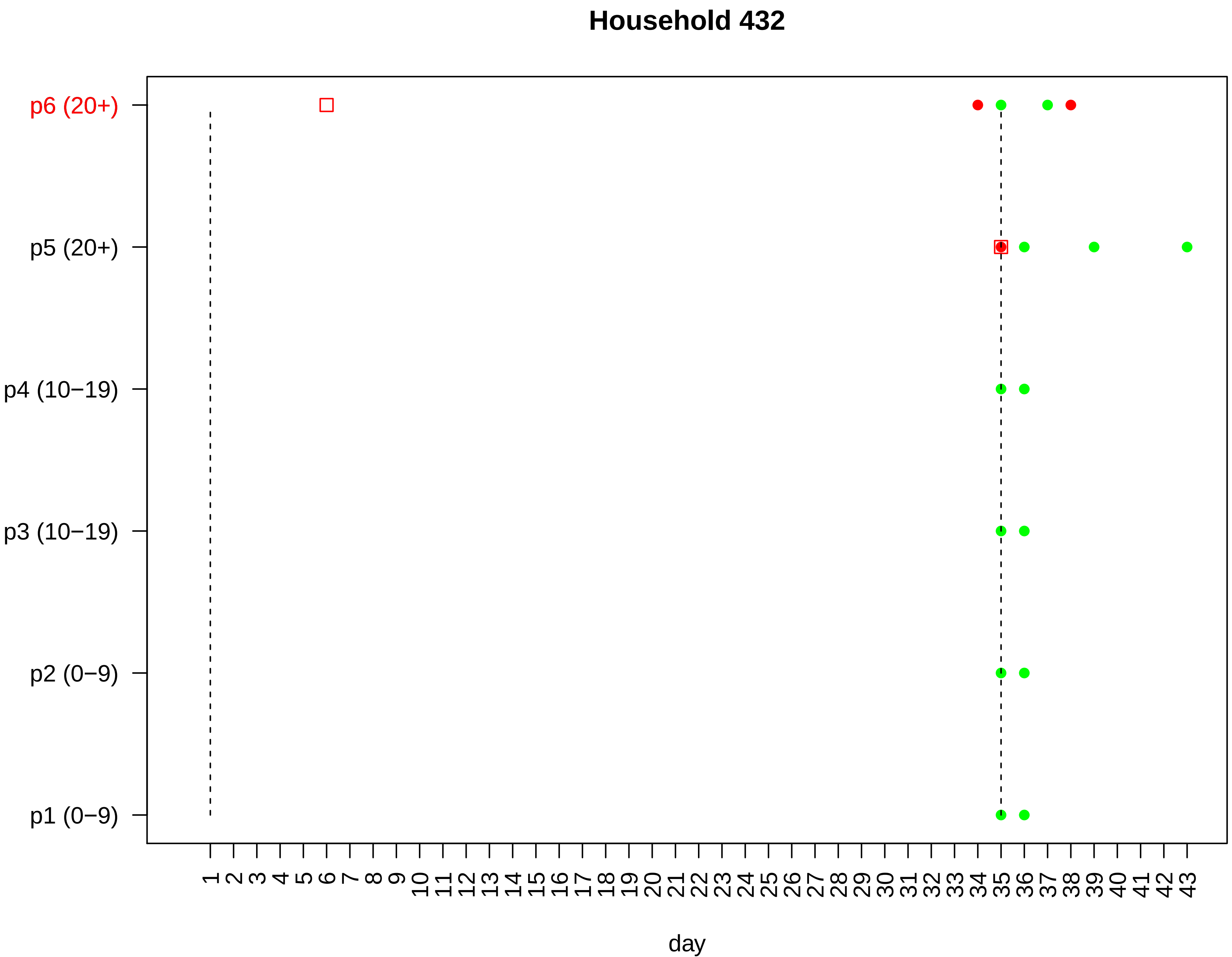
Household 432
p6 (20+)
p5 (20+)
p4 (10−19)
p3 (10−19)
p2 (0−9)
p1 (0−9)
1 2 3 4 5 6 7 8 9 10 11 12 13 14 15 16 17 18 19 20 21 22 23 24 25 26 27 28 29 30 31 32 33 34 35 36 37 38 39 40 41 42 43
day

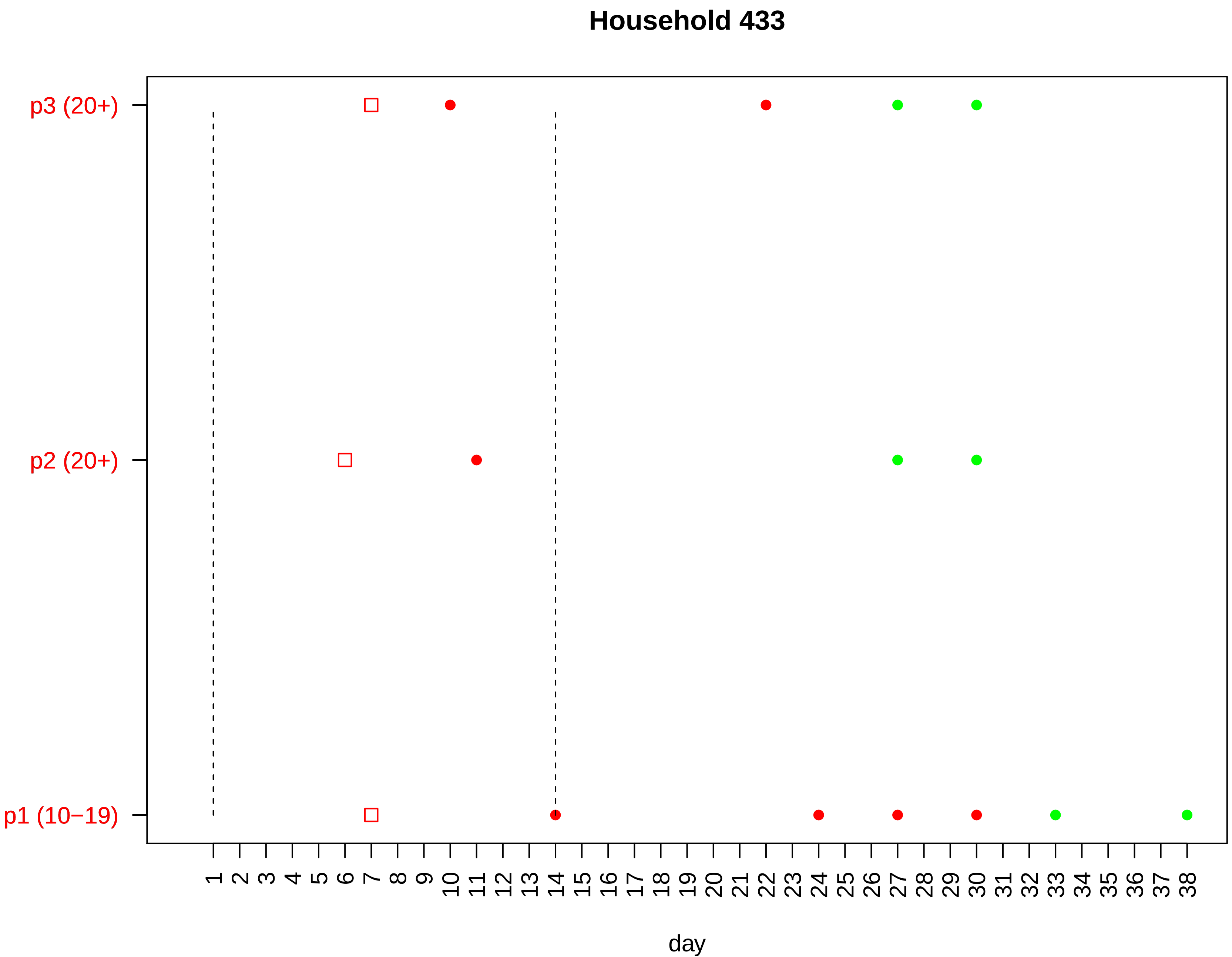

Household 433
p3 (20+)
p2 (20+)
p1 (10−19)
1 2 3 4 5 6 7 8 9 10 11 12 13 14 15 16 17 18 19 20 21 22 23 24 25 26 27 28 29 30 31 32 33 34 35 36 37 38
day

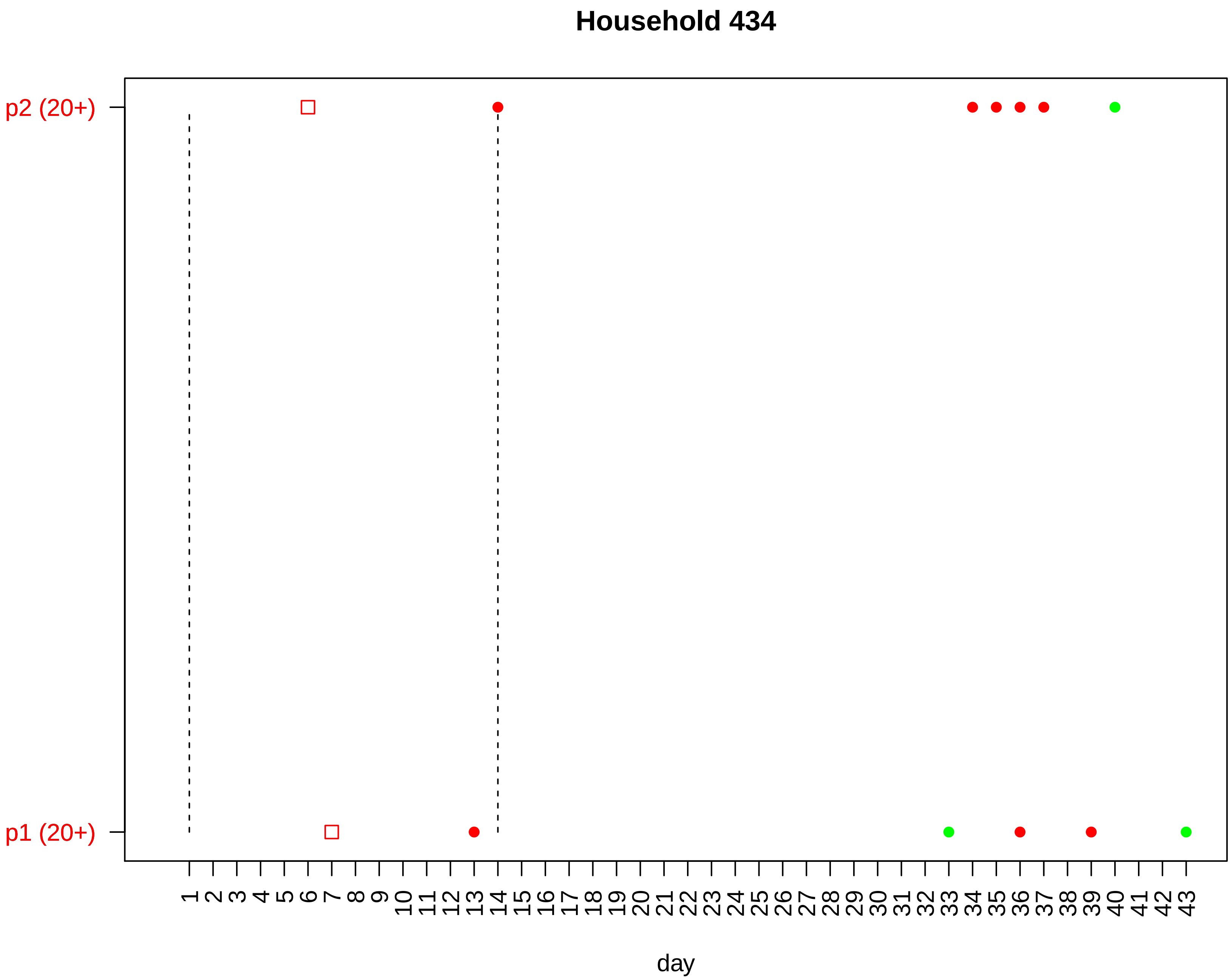

Household 434
p2 (20+)
p1 (20+)
day

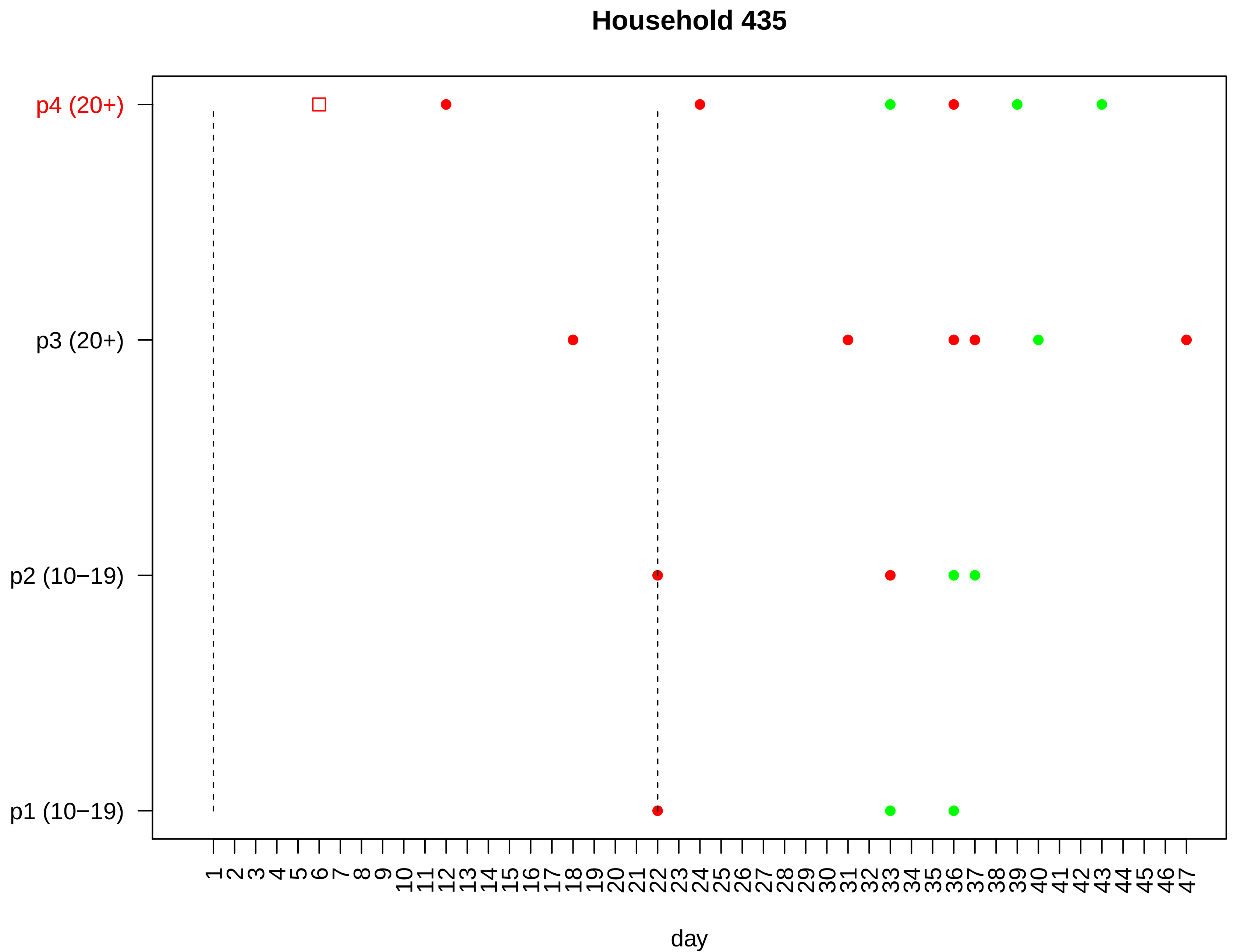
Household 435
p4 (20+)
p3 (20+)
p2 (10−19)
p1 (10−19)
1 2 3 4 5 6 7 8 9 10 11 12 13 14 15 16 17 18 19 20 21 22 23 24 25 26 27 28 29 30 31 32 33 34 35 36 37 38 39 40 41 42 43 44 45 46 47
day

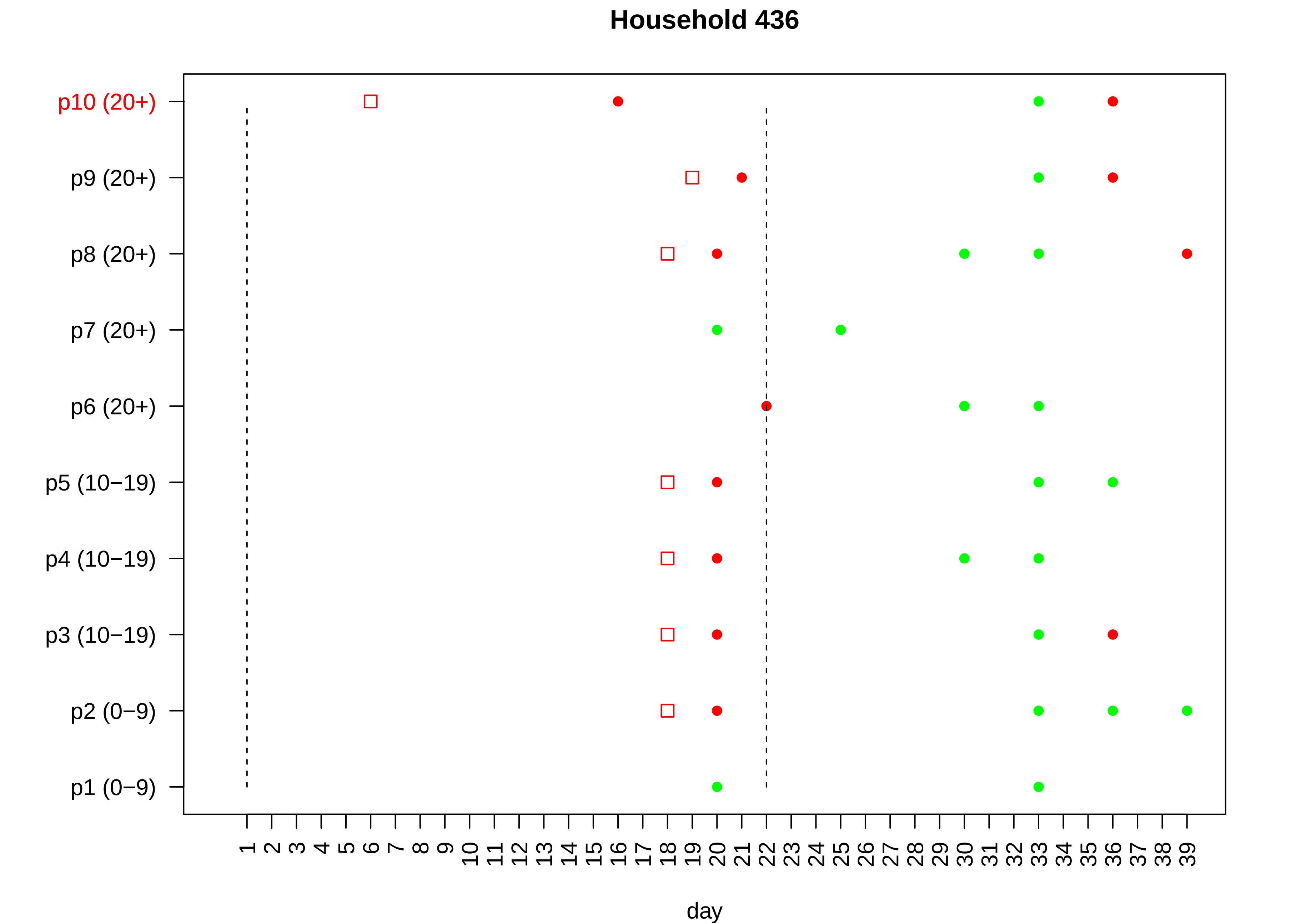

Household 436
p10 (20+)
p9 (20+)
p8 (20+)
p7 (20+)
p6 (20+)
p5 (10−19)
p4 (10−19)
p3 (10−19)
p2 (0−9)
p1 (0−9)
1 2 3 4 5 6 7 8 9 10 11 12 13 14 15 16 17 18 19 20 21 22 23 24 25 26 27 28 29 30 31 32 33 34 35 36 37 38 39
day

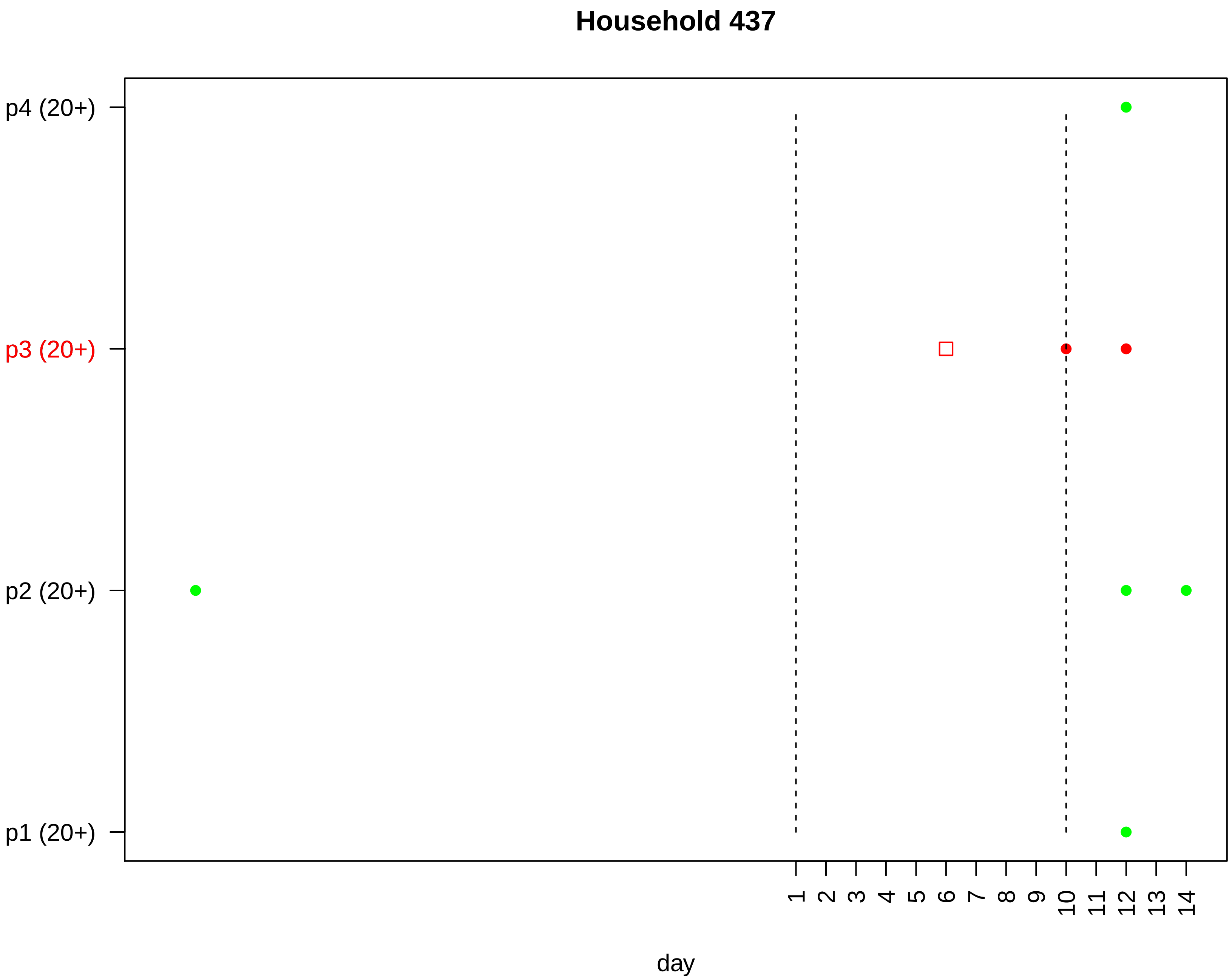

Household 437
p4 (20+)
p3 (20+)
p2 (20+)
p1 (20+)
1
2
3
4
5
6
7
8
9
10
11
12
13
14
day

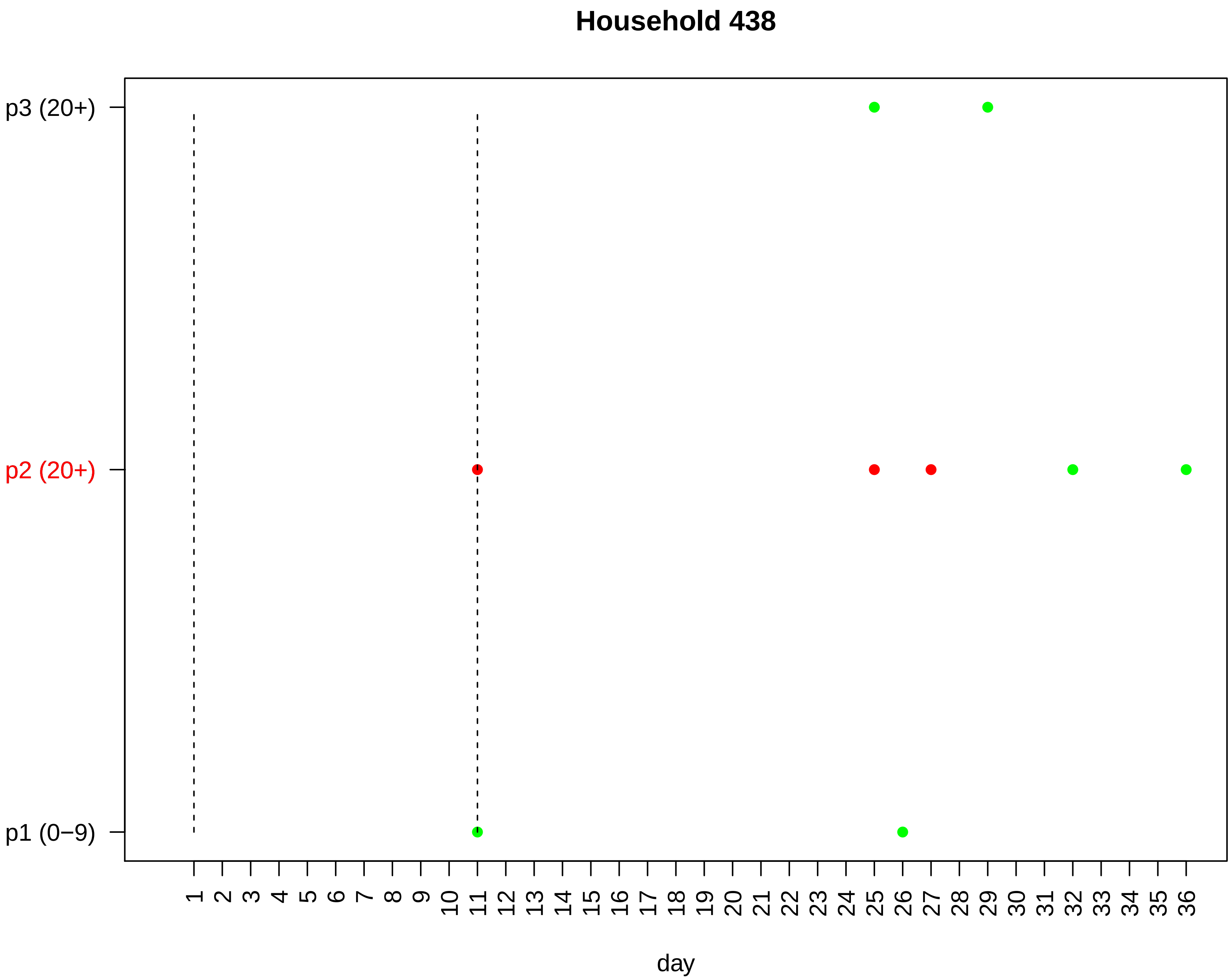
Household 438
p3 (20+)
p2 (20+)
p1 (0−9)
1 2 3 4 5 6 7 8 9 10 11 12 13 14 15 16 17 18 19 20 21 22 23 24 25 26 27 28 29 30 31 32 33 34 35 36
day

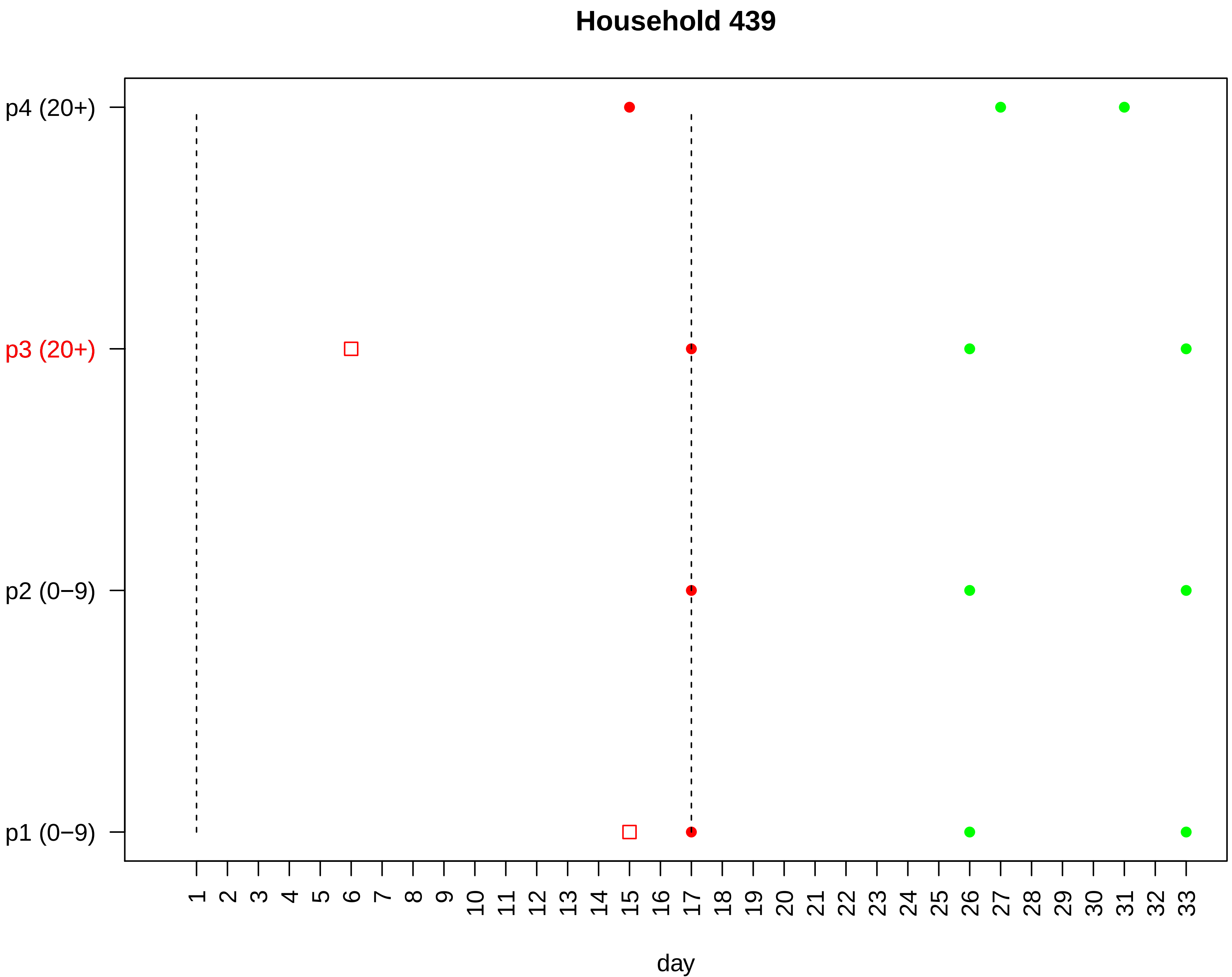

Household 439
p4 (20+)
p3 (20+)
p2 (0−9)
p1 (0−9)
1 2 3 4 5 6 7 8 9 10 11 12 13 14 15 16 17 18 19 20 21 22 23 24 25 26 27 28 29 30 31 32 33
day

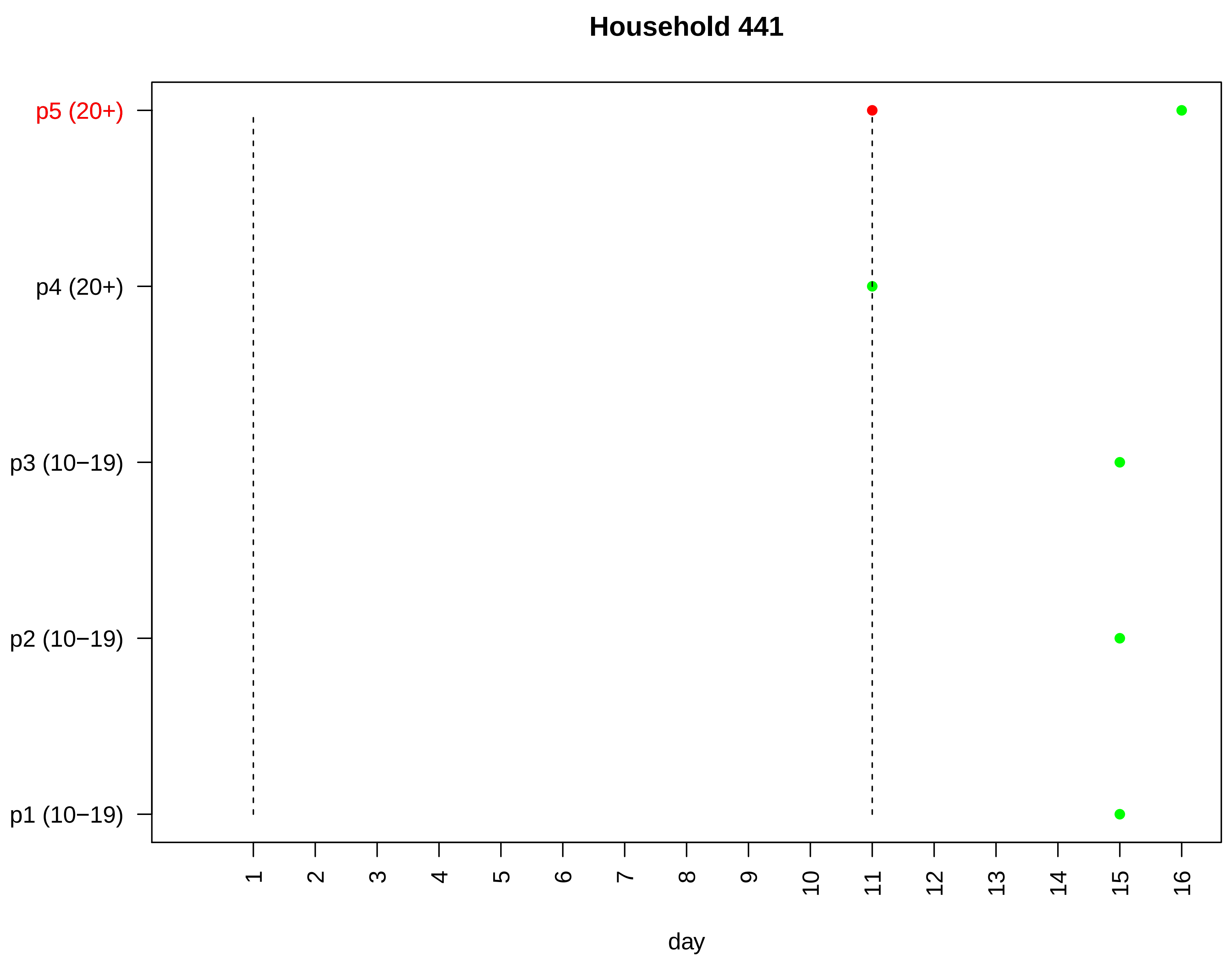

Household 441
p5 (20+)
p4 (20+)
p3 (10−19)
p2 (10−19)
p1 (10−19)
day

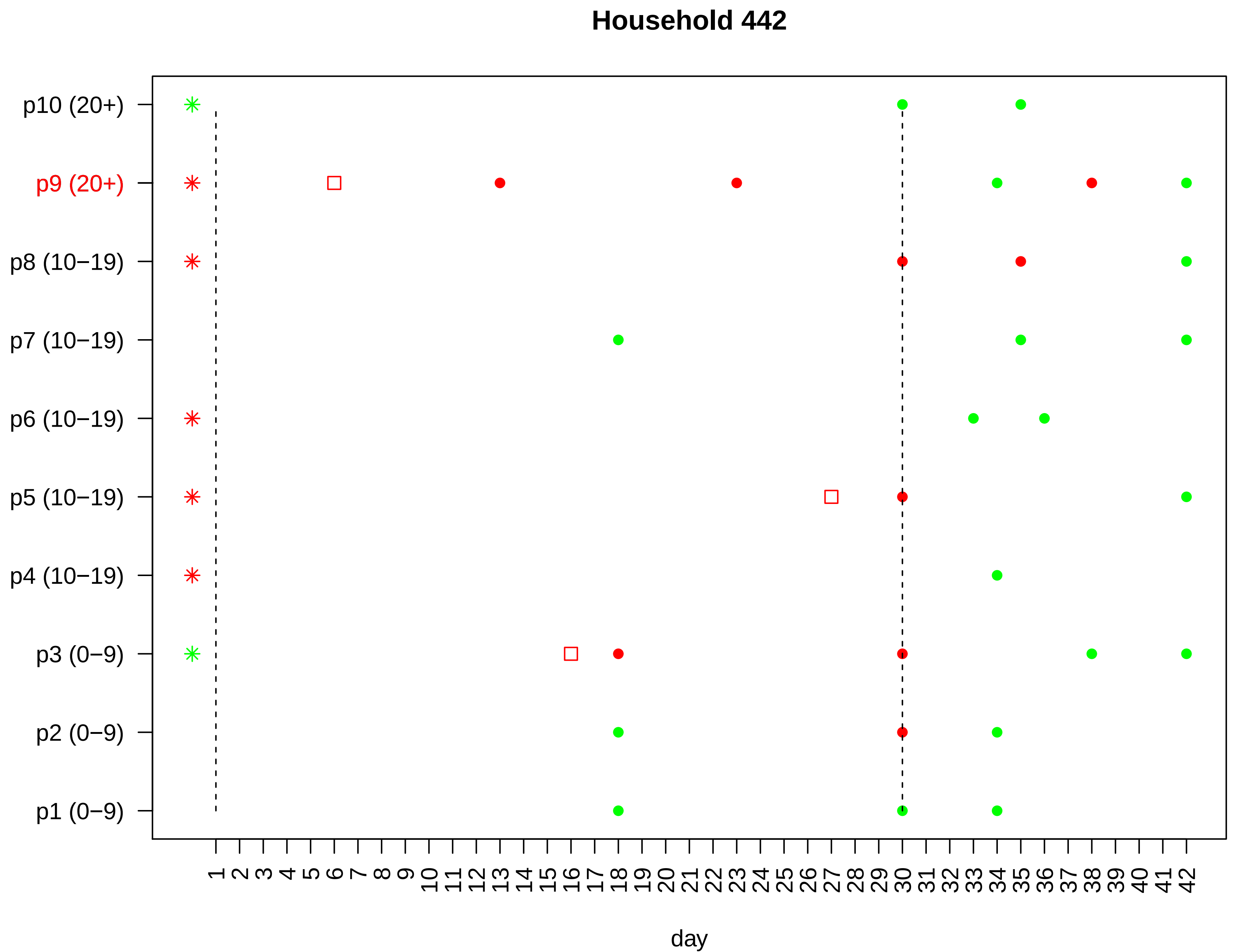
Household 442
p10 (20+)
p9 (20+)
p8 (10−19)
p7 (10−19)
p6 (10−19)
p5 (10−19)
p4 (10−19)
p3 (0−9)
p2 (0−9)
p1 (0−9)
1 2 3 4 5 6 7 8 9 10 11 12 13 14 15 16 17 18 19 20 21 22 23 24 25 26 27 28 29 30 31 32 33 34 35 36 37 38 39 40 41 42
day

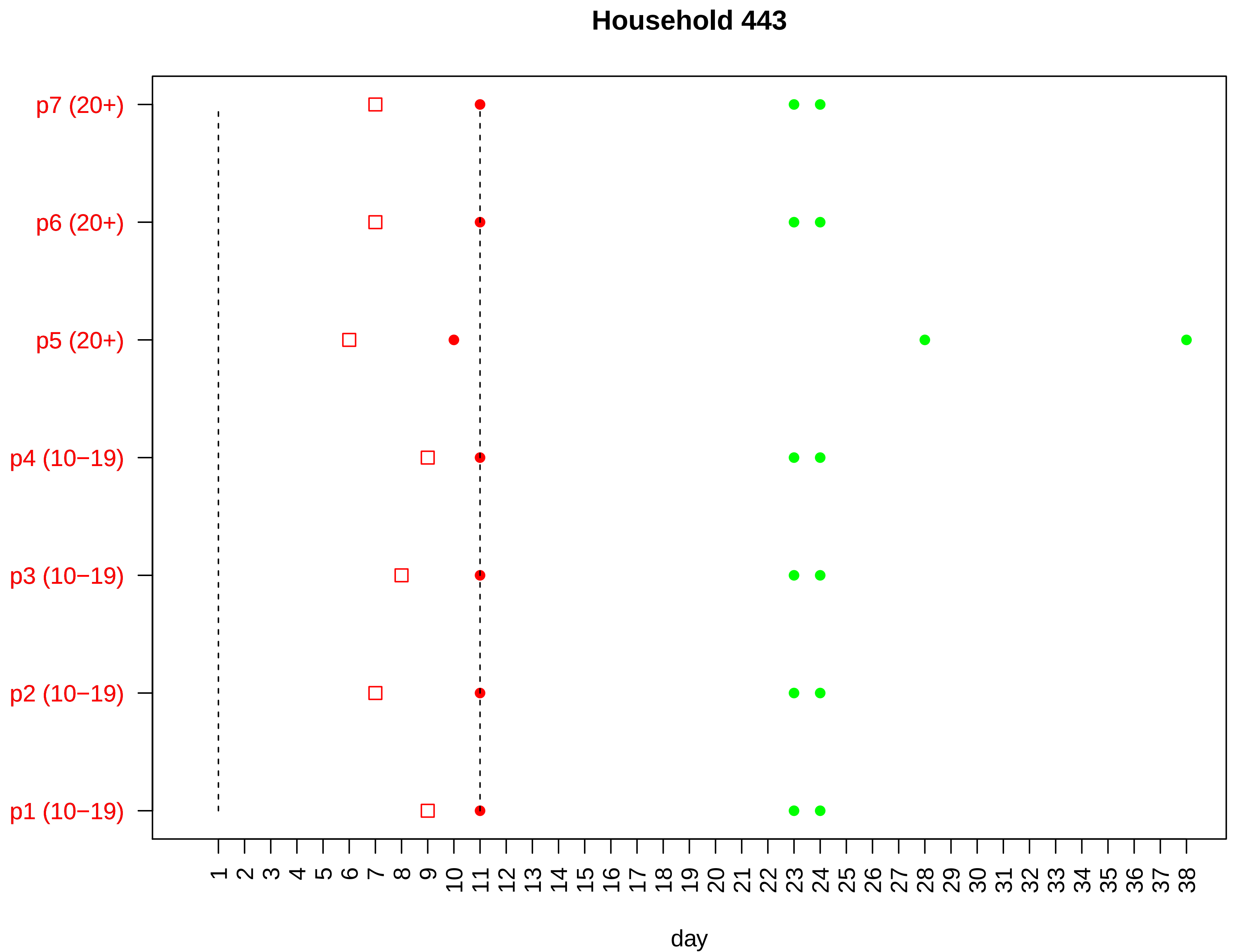

Household 443
p7 (20+)
p6 (20+)
p5 (20+)
p4 (10−19)
p3 (10−19)
p2 (10−19)
p1 (10−19)
1 2 3 4 5 6 7 8 9 10 11 12 13 14 15 16 17 18 19 20 21 22 23 24 25 26 27 28 29 30 31 32 33 34 35 36 37 38
day

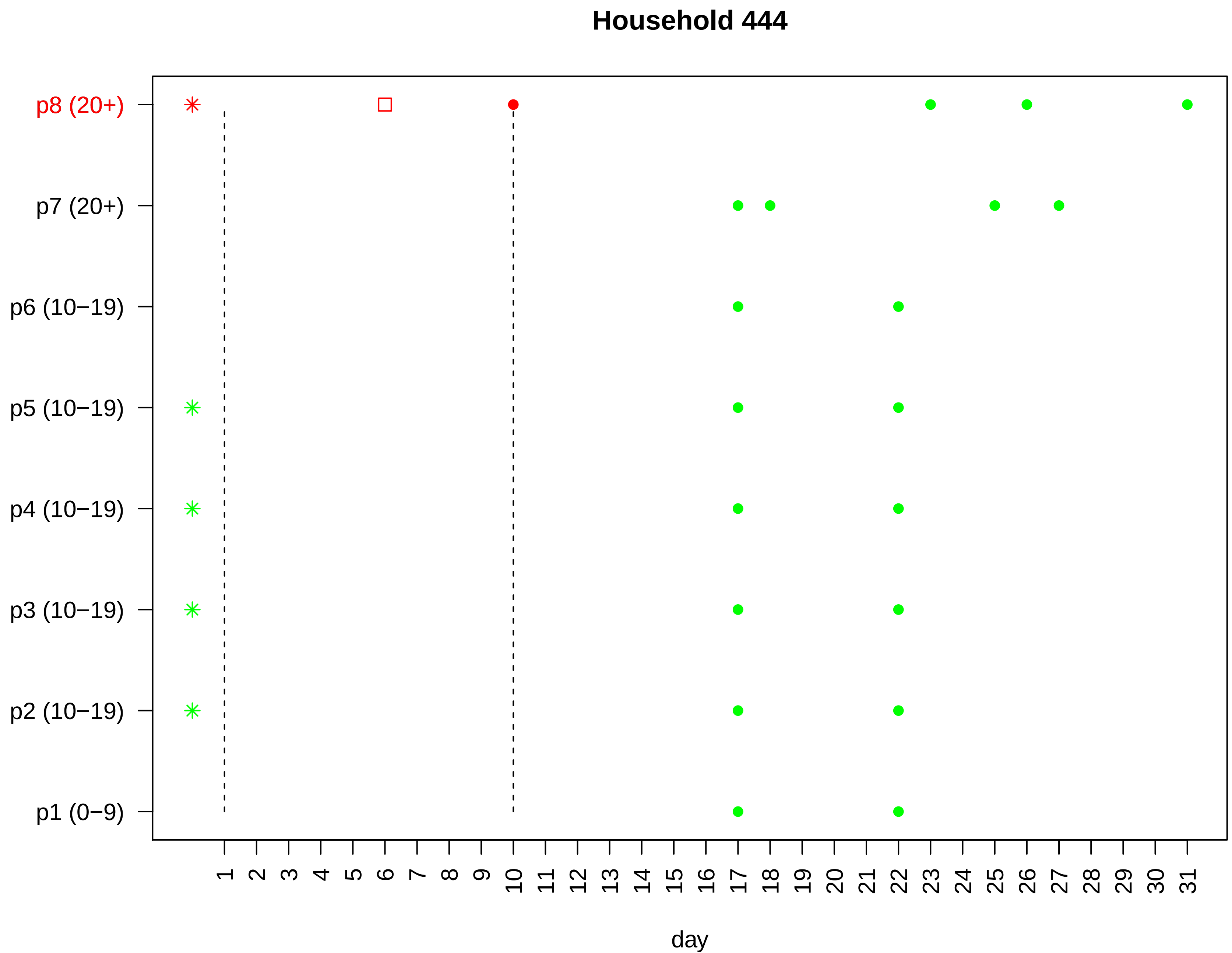

Household 444
p8 (20+)
p7 (20+)
p6 (10−19)
p5 (10−19)
p4 (10−19)
p3 (10−19)
p2 (10−19)
p1 (0−9)
1 2 3 4 5 6 7 8 9 10 11 12 13 14 15 16 17 18 19 20 21 22 23 24 25 26 27 28 29 30 31
day

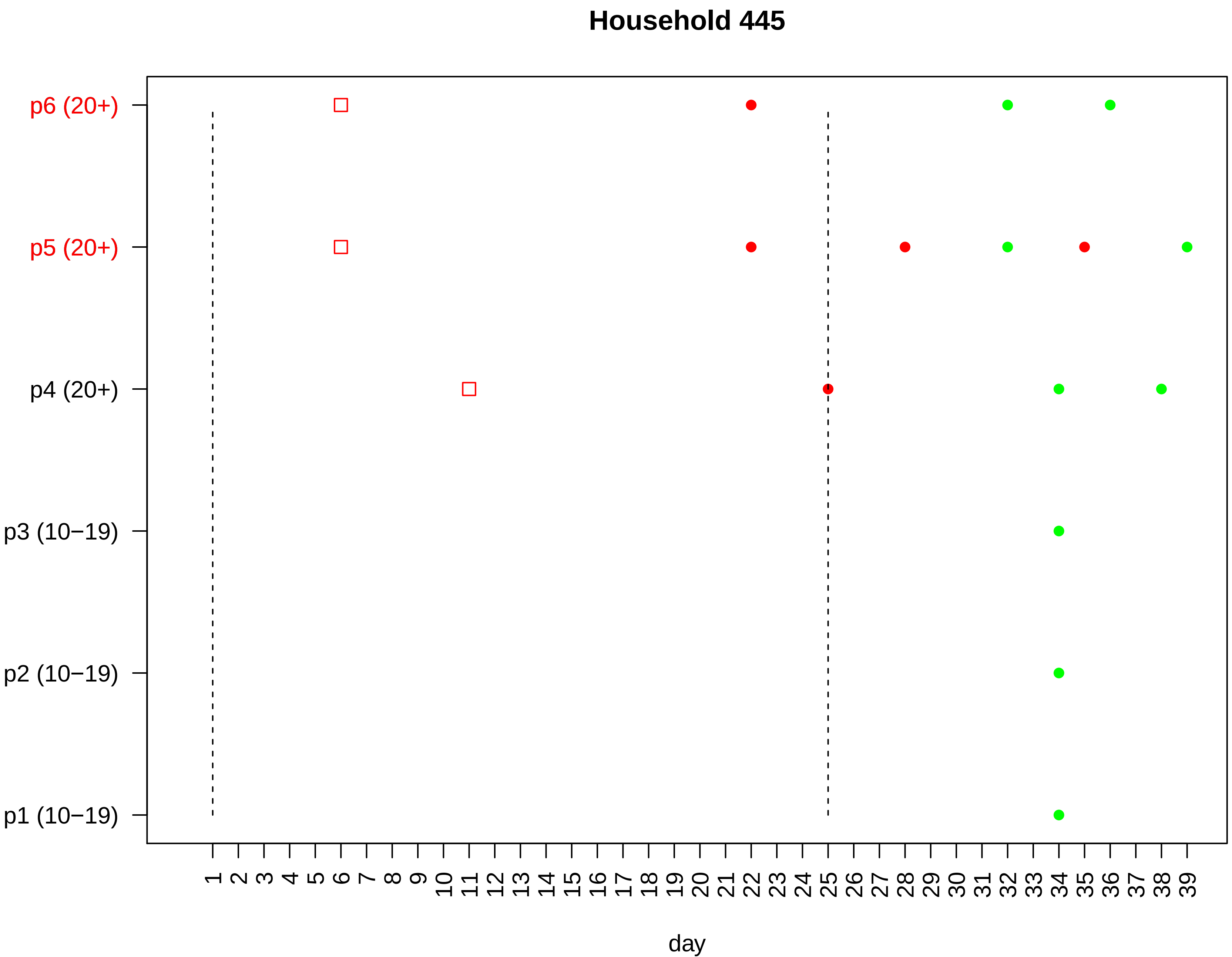
Household 445
p6 (20+)
p5 (20+)
p4 (20+)
p3 (10−19)
p2 (10−19)
p1 (10−19)
1 2 3 4 5 6 7 8 9 10 11 12 13 14 15 16 17 18 19 20 21 22 23 24 25 26 27 28 29 30 31 32 33 34 35 36 37 38 39
day

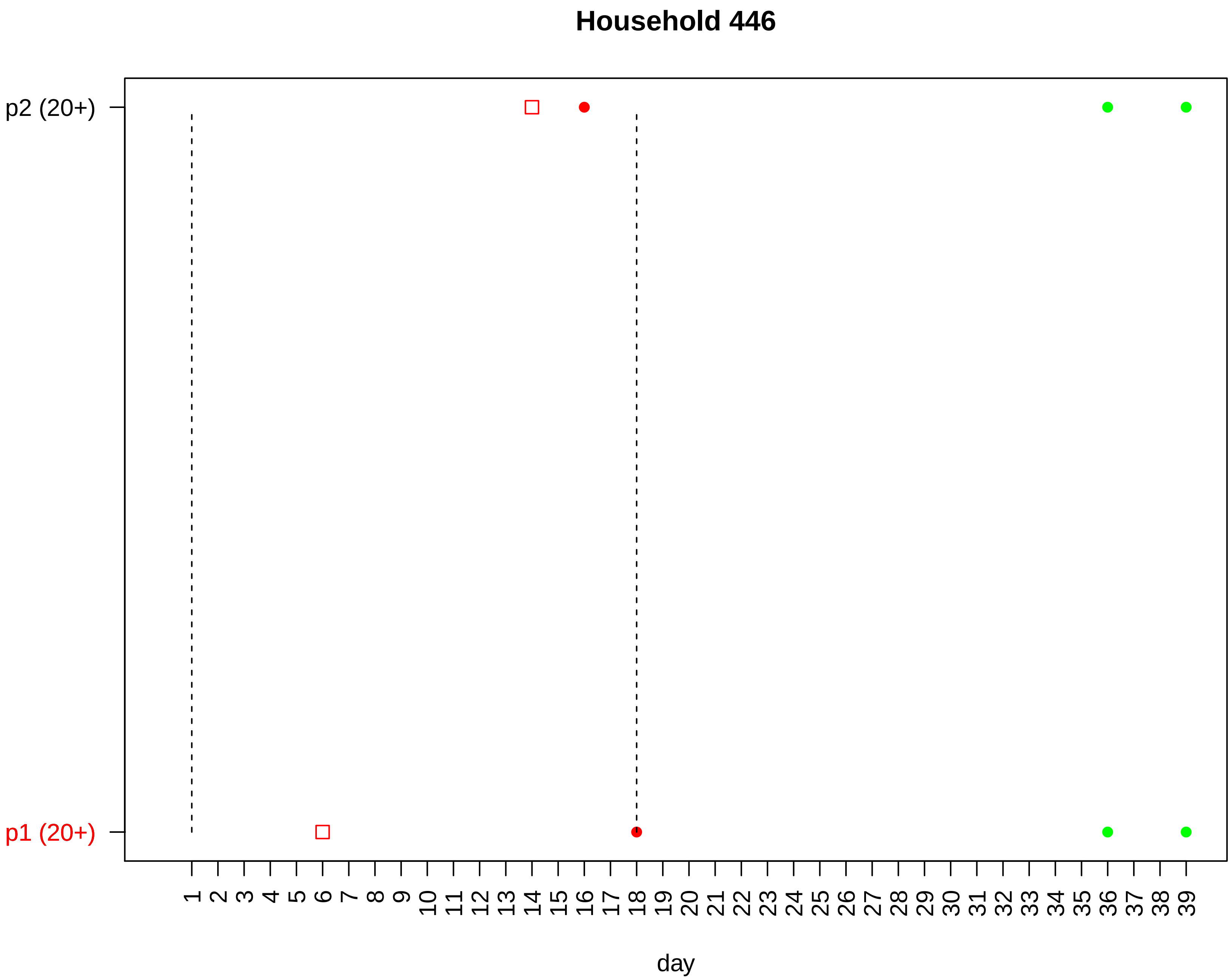
Household 446
p2 (20+)
p1 (20+)
day

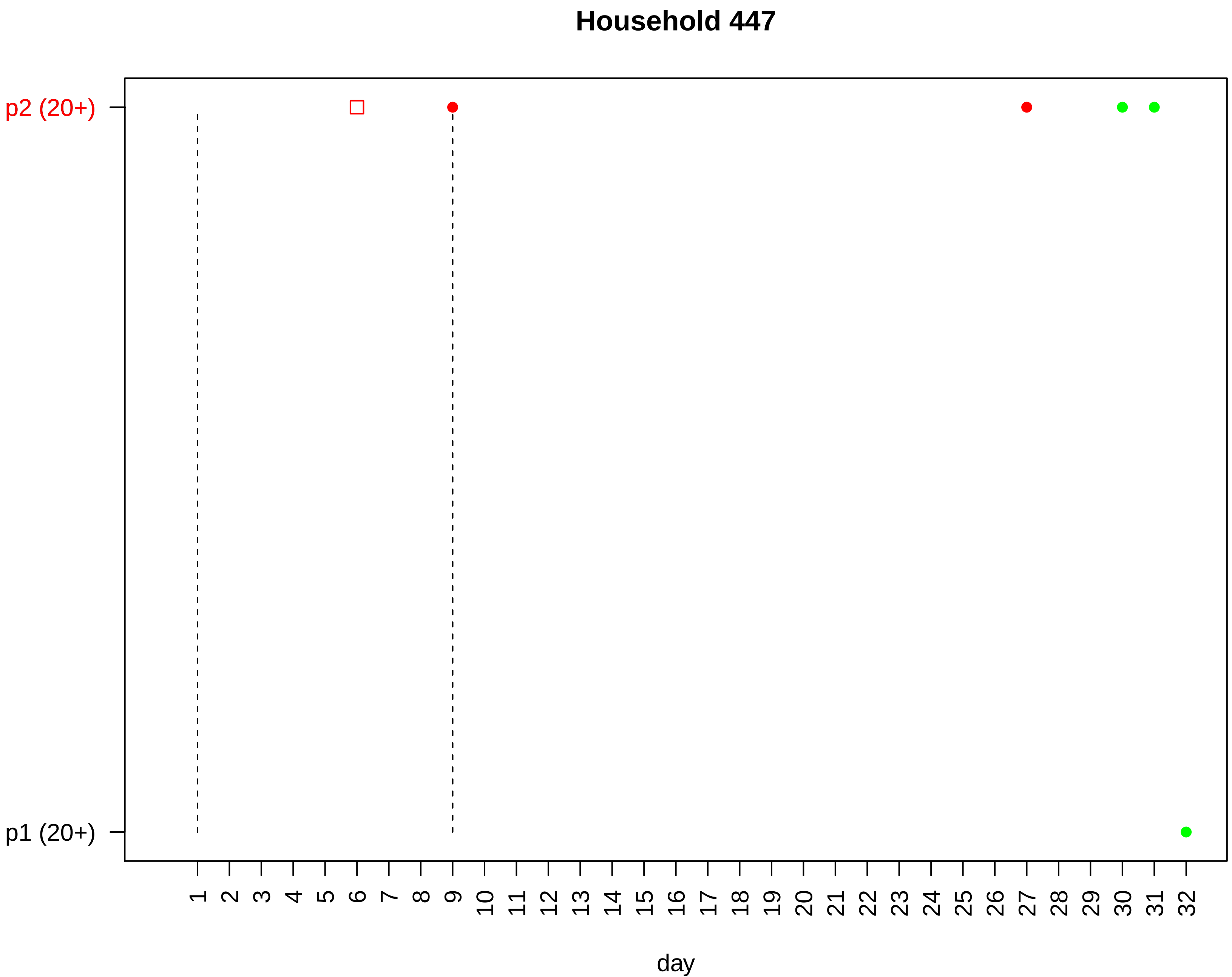
Household 447
p2 (20+)
p1 (20+)
1 2 3 4 5 6 7 8 9 10 11 12 13 14 15 16 17 18 19 20 21 22 23 24 25 26 27 28 29 30 31 32
day

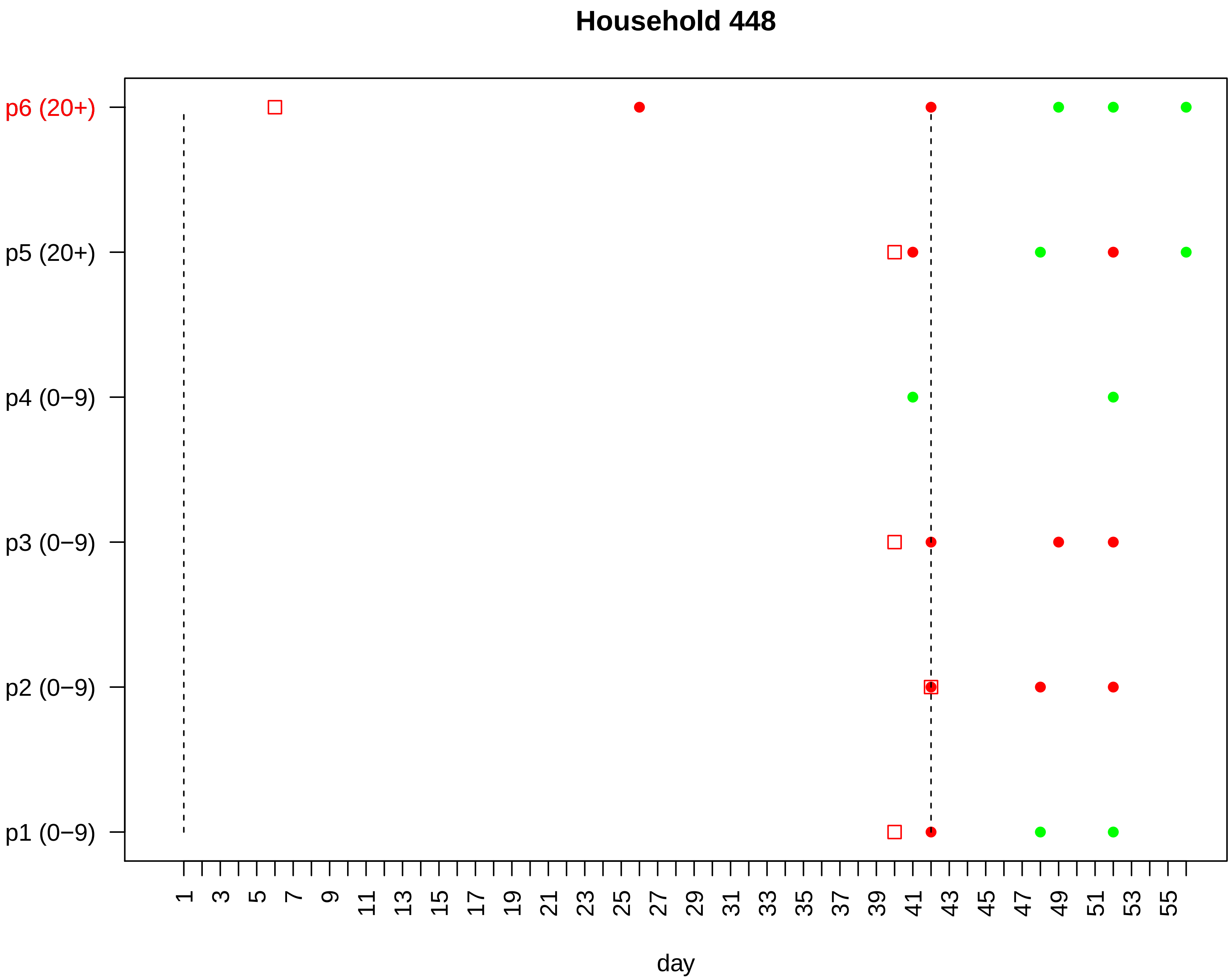

Household 448
p6 (20+)
p5 (20+)
p4 (0−9)
p3 (0−9)
p2 (0−9)
p1 (0−9)
1 3 5 7 9 11 13 15 17 19 21 23 25 27 29 31 33 35 37 39 41 43 45 47 49 51 53 55
day

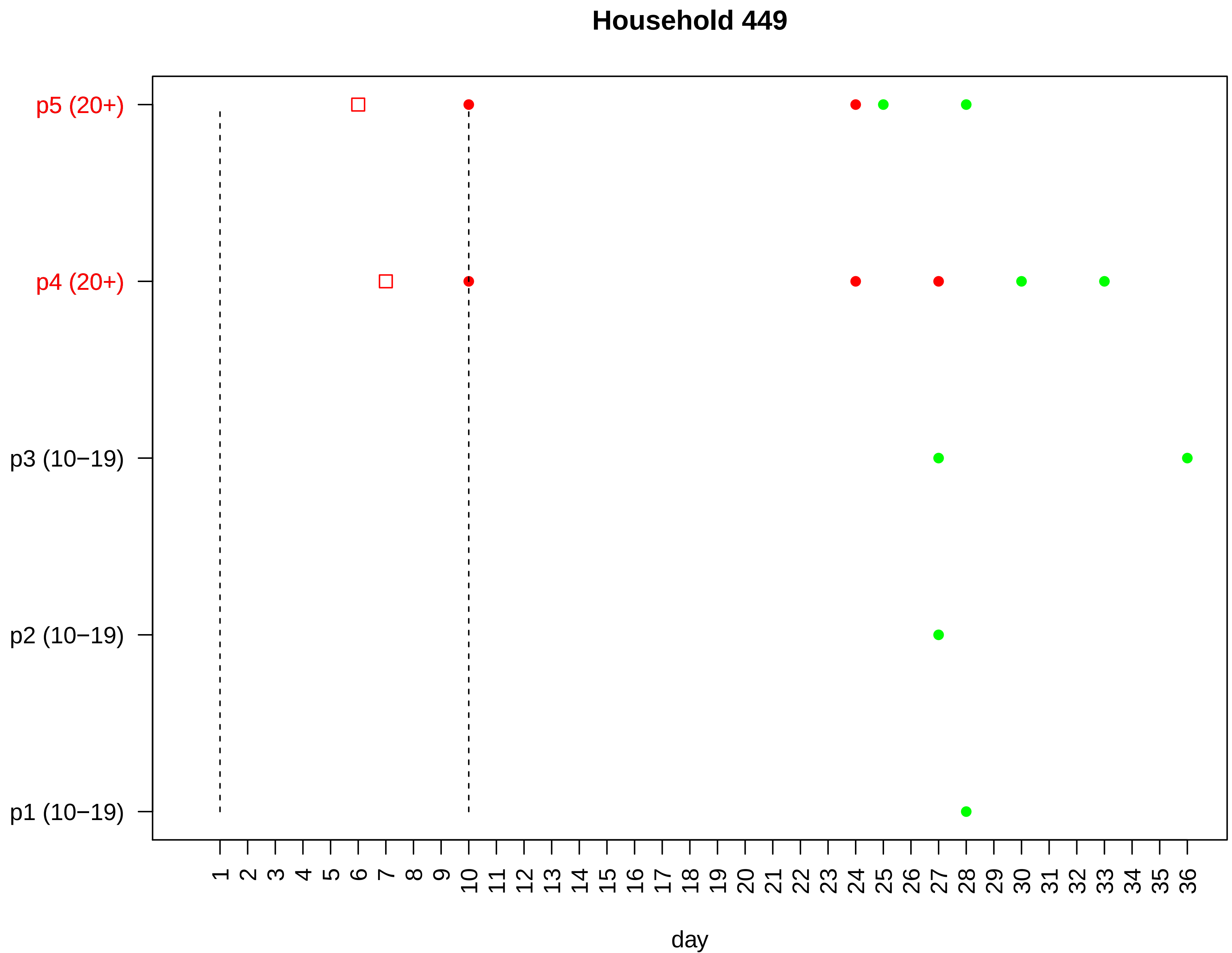

Household 449
p5 (20+)
p4 (20+)
p3 (10−19)
p2 (10−19)
p1 (10−19)
1 2 3 4 5 6 7 8 9 10 11 12 13 14 15 16 17 18 19 20 21 22 23 24 25 26 27 28 29 30 31 32 33 34 35 36
day

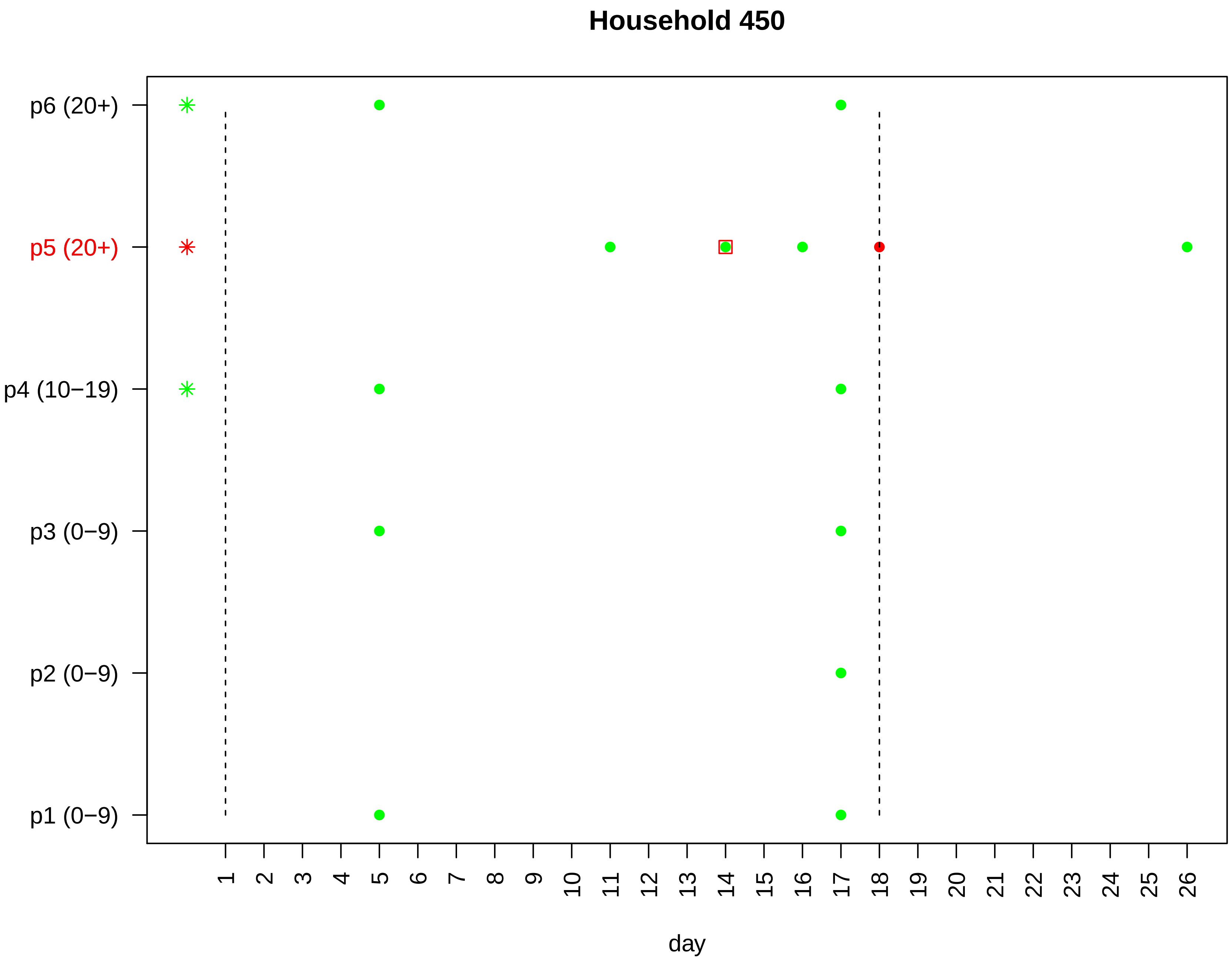

Household 450
p6 (20+)
p5 (20+)
p4 (10−19)
p3 (0−9)
p2 (0−9)
p1 (0−9)
1 2 3 4 5 6 7 8 9 10 11 12 13 14 15 16 17 18 19 20 21 22 23 24 25 26
day

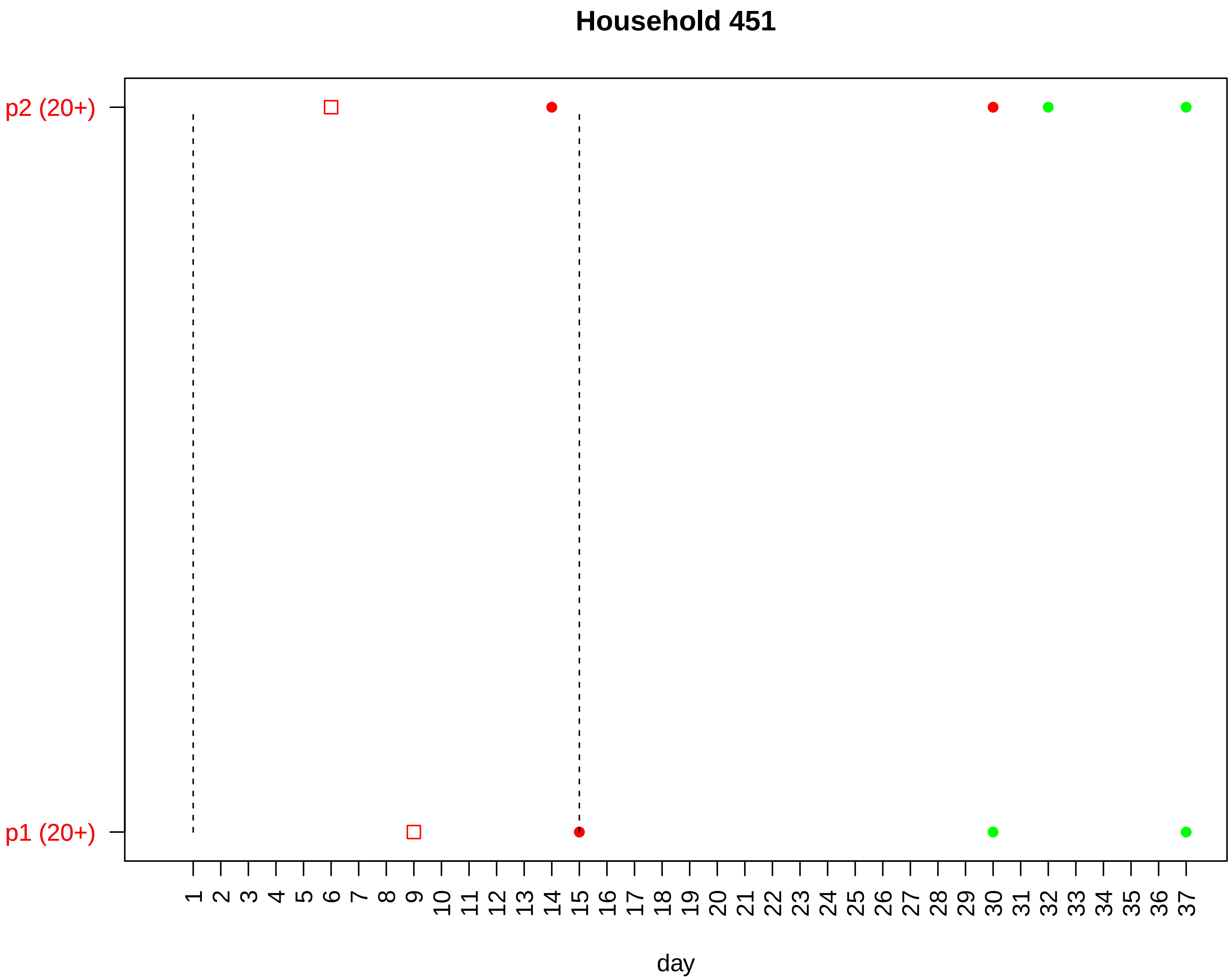

Household 451
p2 (20+)
p1 (20+)
1 2 3 4 5 6 7 8 9 10 11 12 13 14 15 16 17 18 19 20 21 22 23 24 25 26 27 28 29 30 31 32 33 34 35 36 37
day

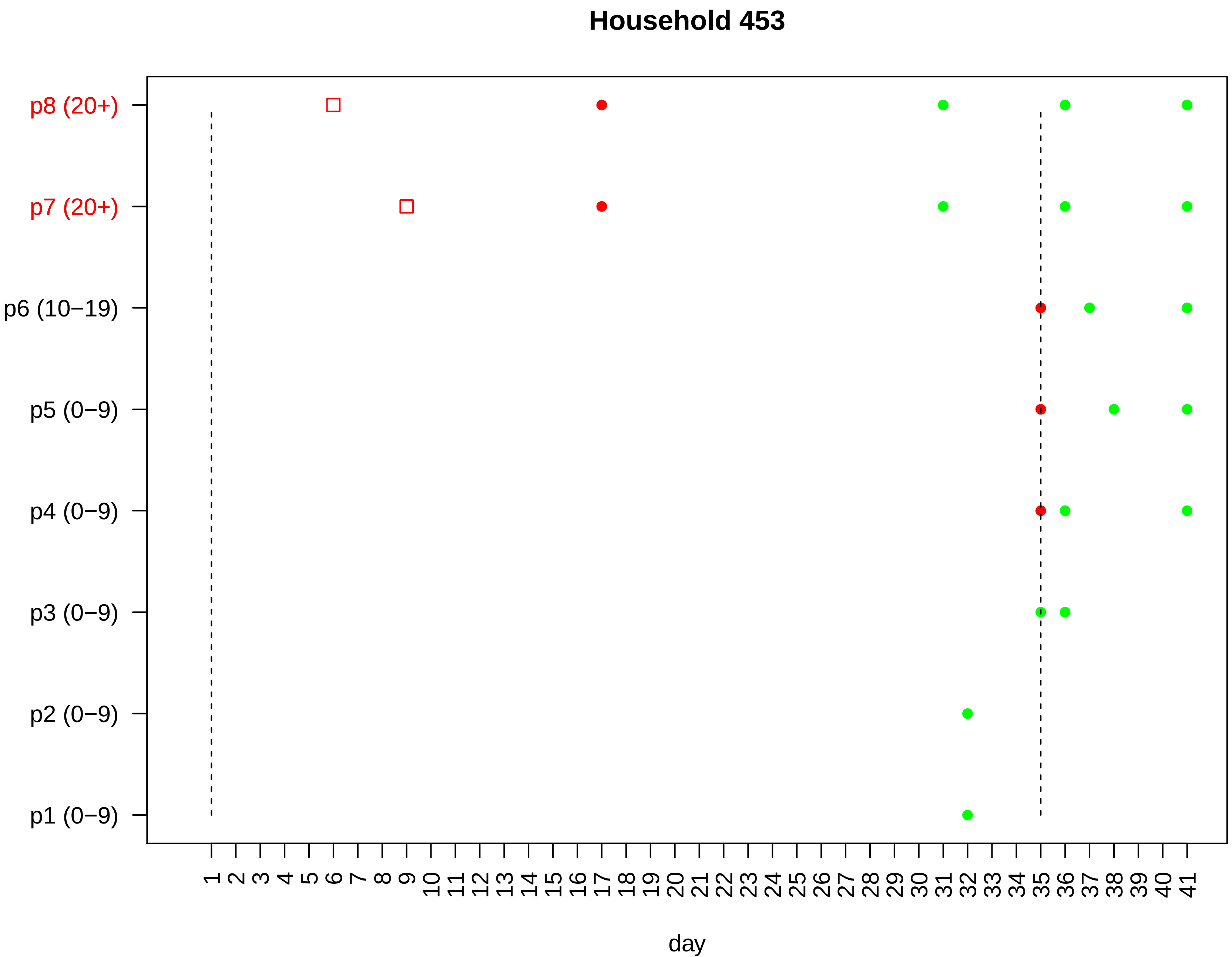
Household 453
p8 (20+)
p7 (20+)
p6 (10−19)
p5 (0−9)
p4 (0−9)
p3 (0−9)
p2 (0−9)
p1 (0−9)
1 2 3 4 5 6 7 8 9 10 11 12 13 14 15 16 17 18 19 20 21 22 23 24 25 26 27 28 29 30 31 32 33 34 35 36 37 38 39 40 41
day

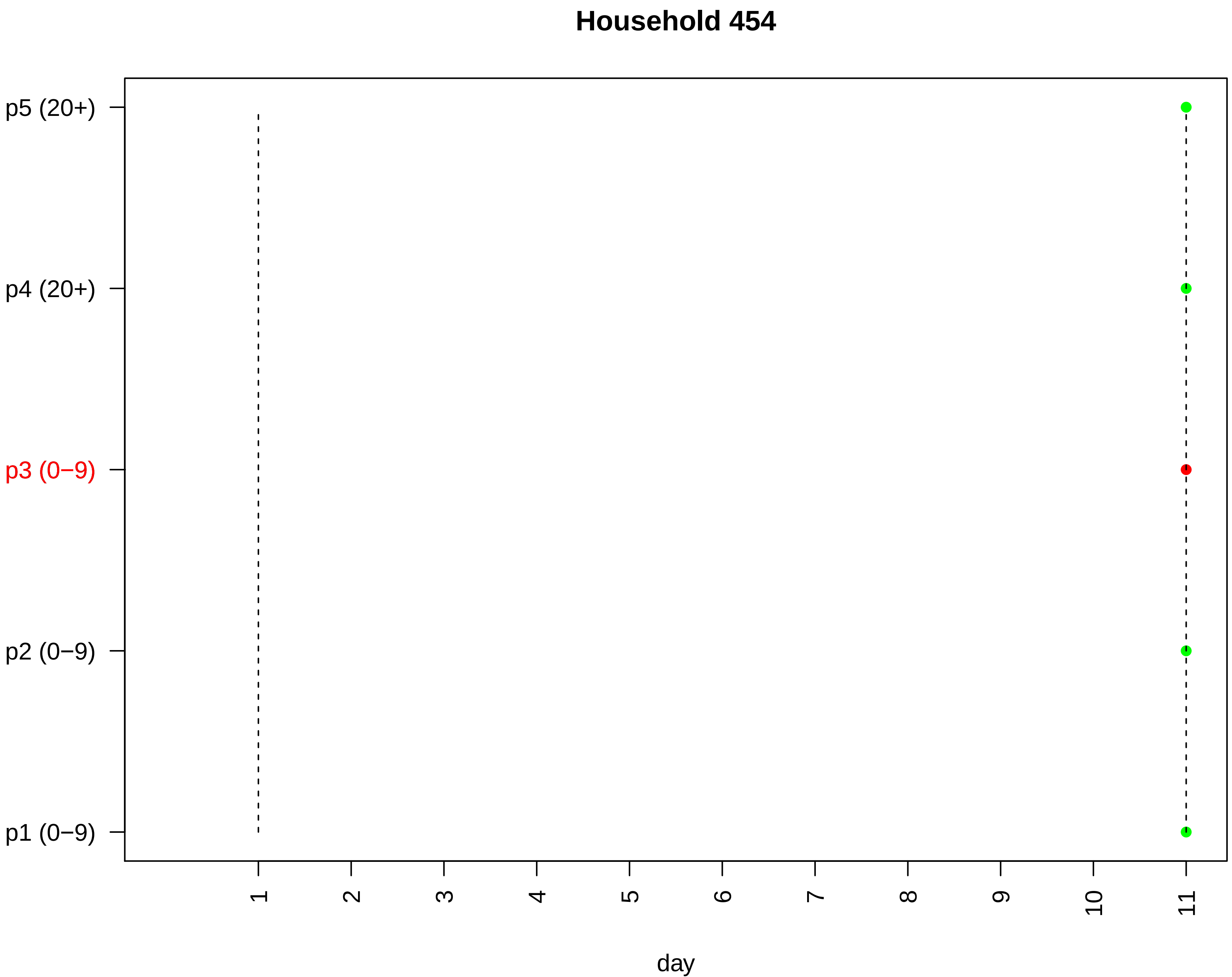
Household 454
p5 (20+)
p4 (20+)
p3 (0−9)
p2 (0−9)
p1 (0−9)
1
2
3
4
5
6
7
8
9
10
11
day

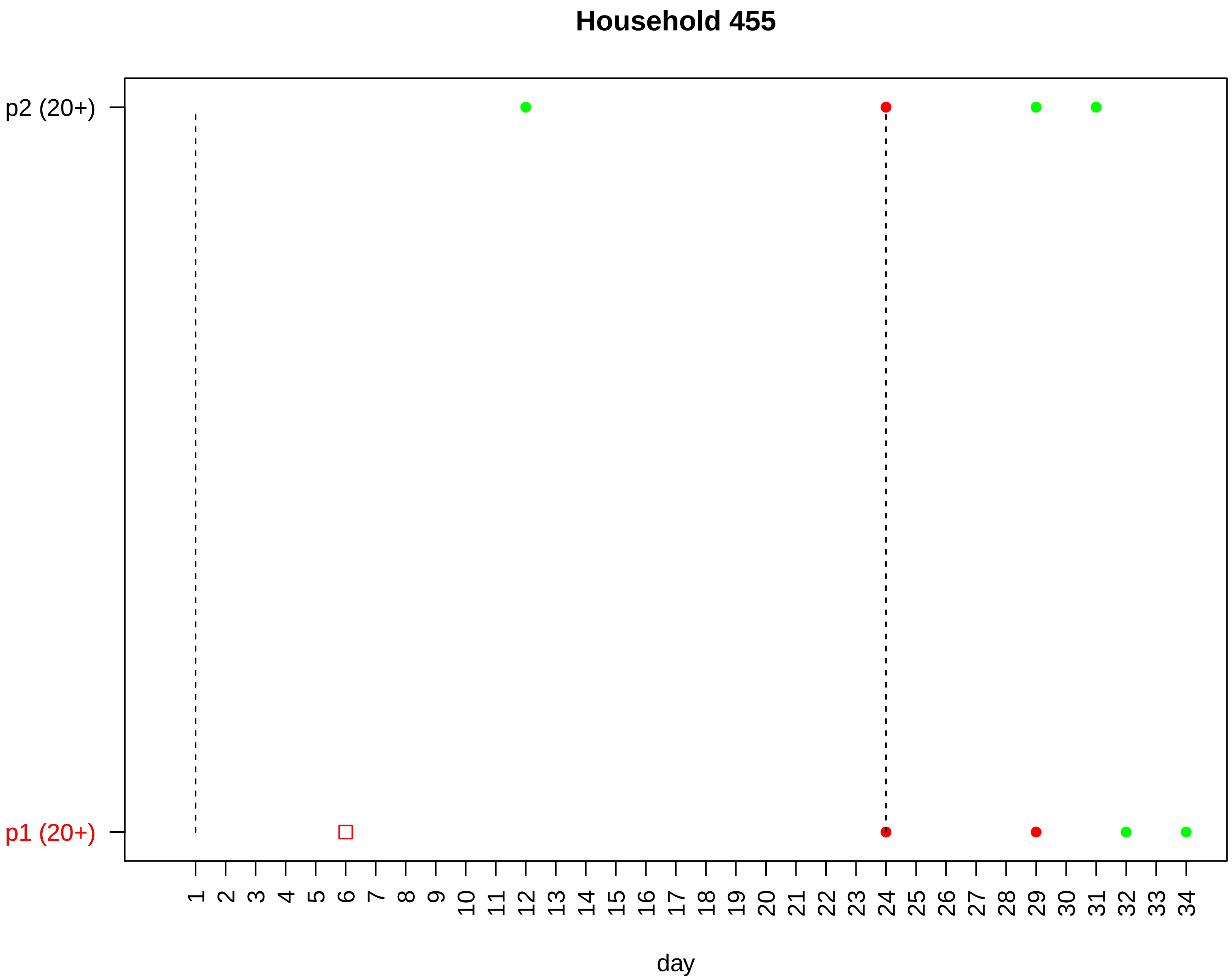

Household 455
p2 (20+)
p1 (20+)
day

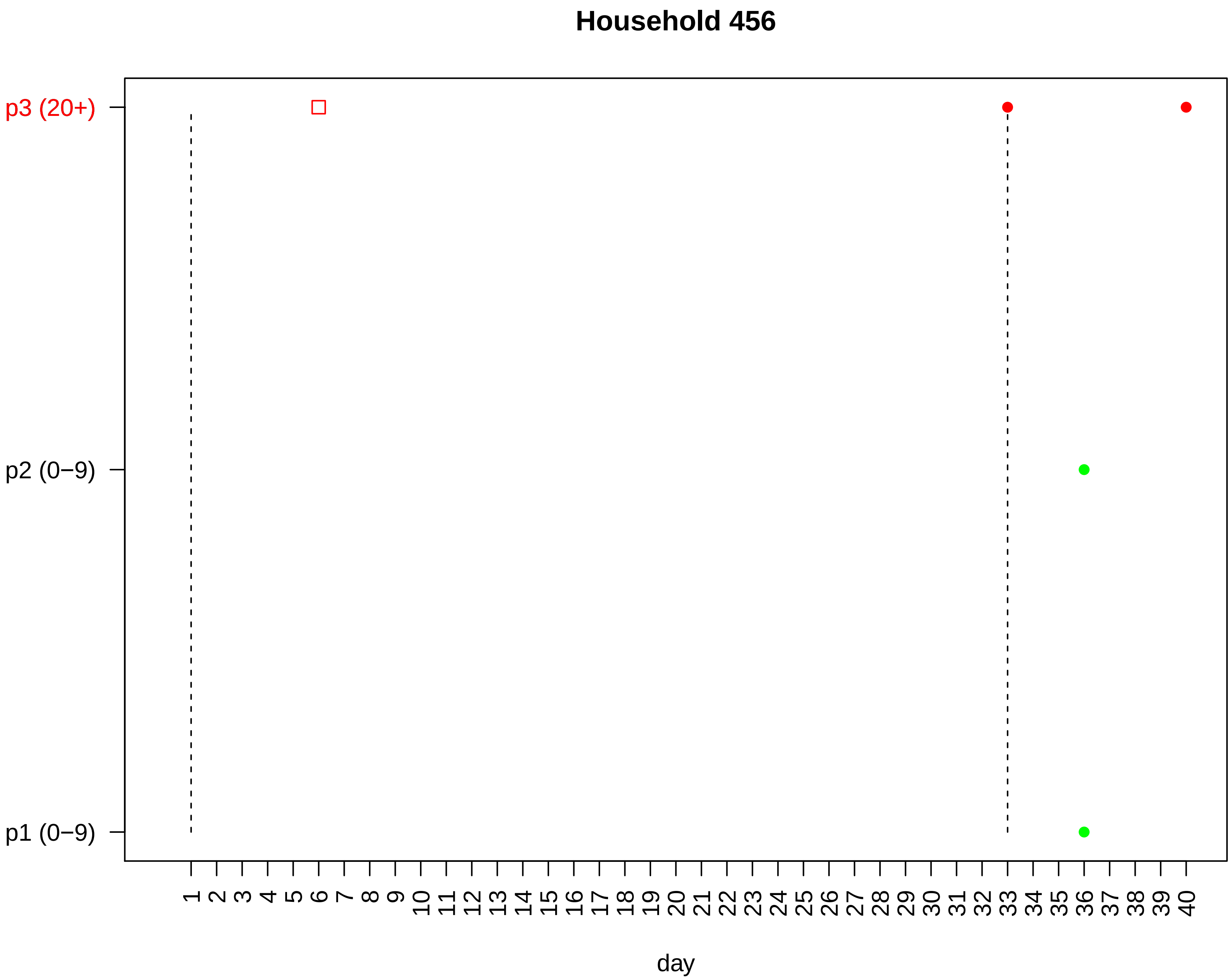

Household 456
p3 (20+)
p2 (0−9)
p1 (0−9)
1 2 3 4 5 6 7 8 9 10 11 12 13 14 15 16 17 18 19 20 21 22 23 24 25 26 27 28 29 30 31 32 33 34 35 36 37 38 39 40
day

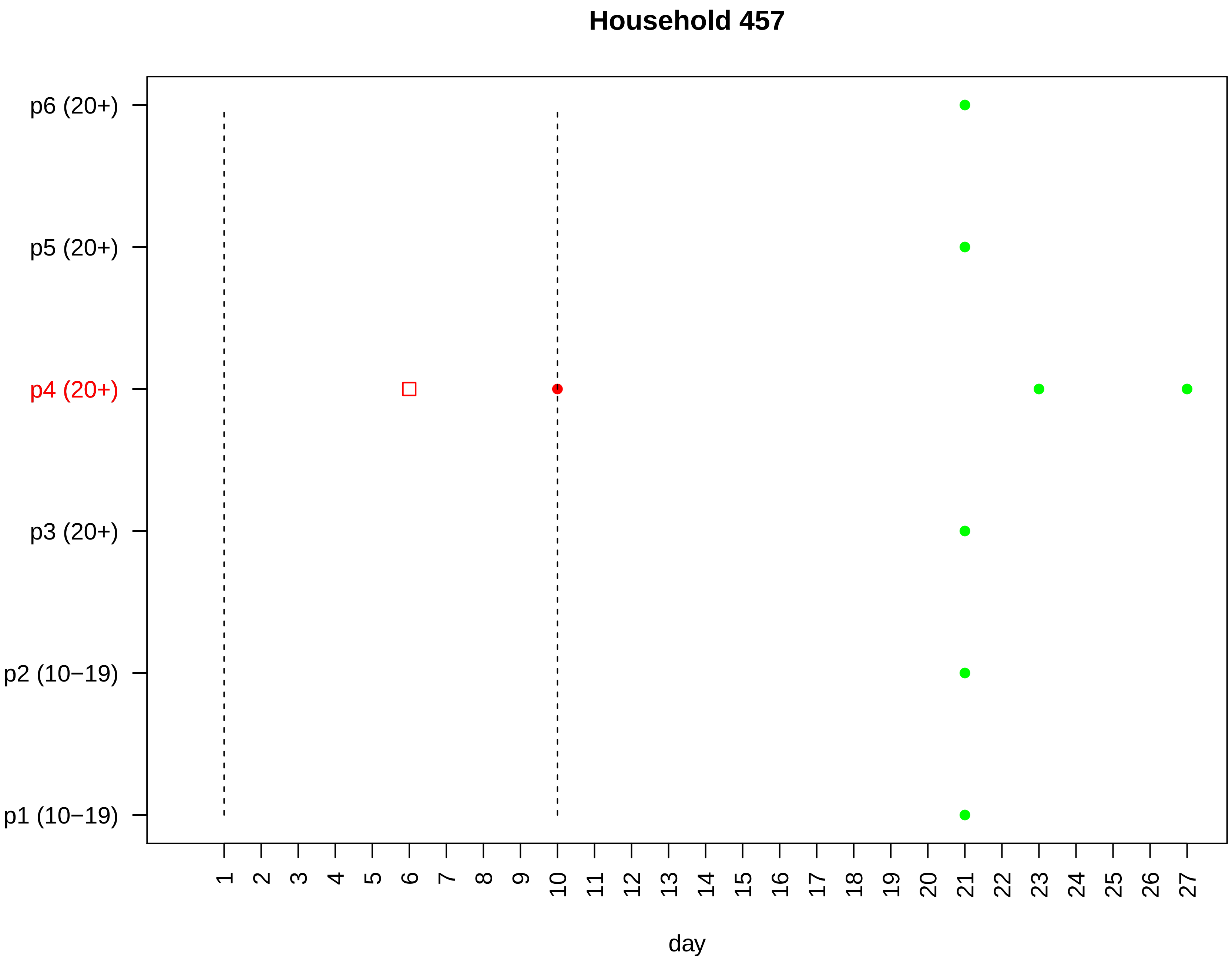
Household 457
p6 (20+)
p5 (20+)
p4 (20+)
p3 (20+)
p2 (10−19)
p1 (10−19)
1 2 3 4 5 6 7 8 9 10 11 12 13 14 15 16 17 18 19 20 21 22 23 24 25 26 27
day

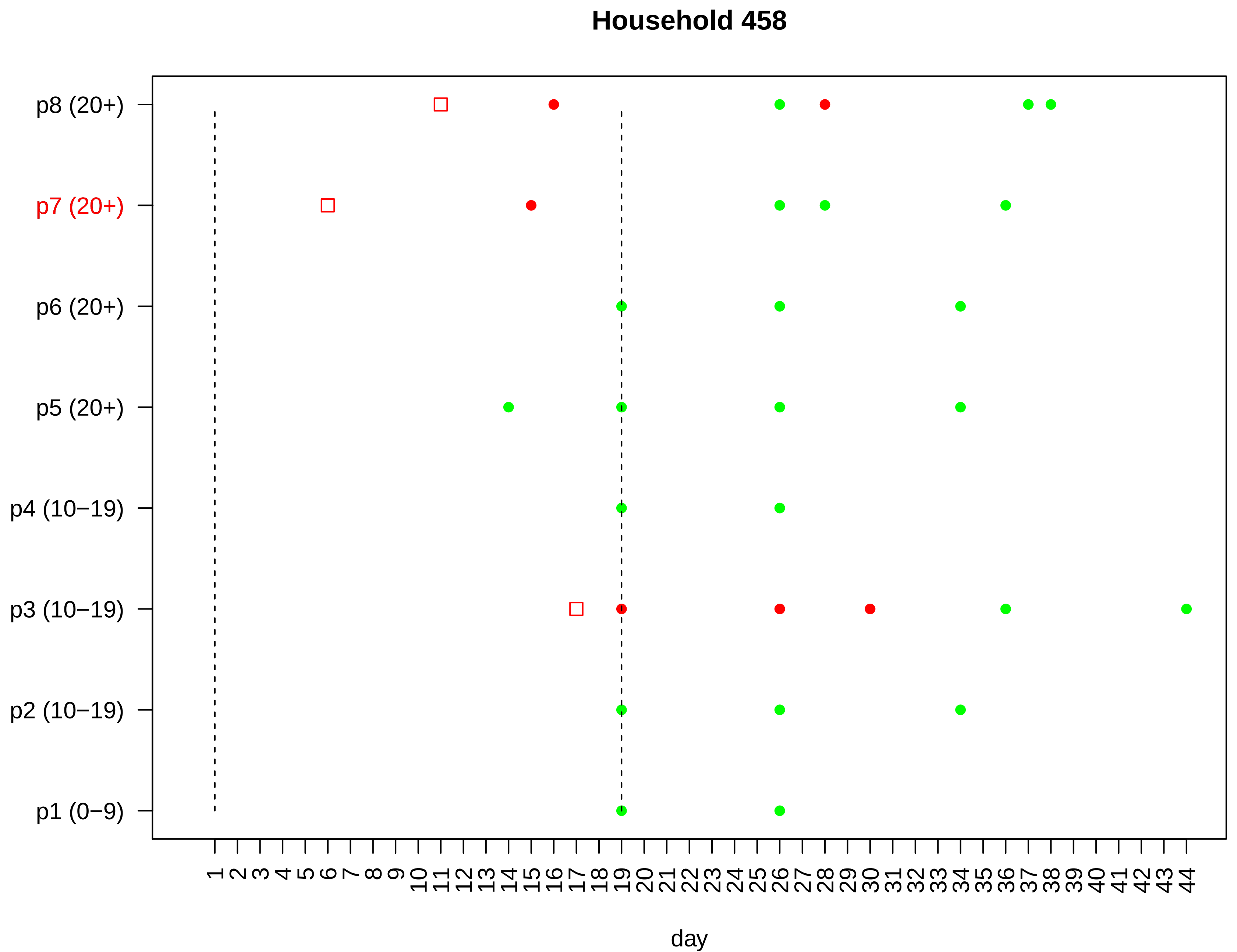

Household 458
p8 (20+)
p7 (20+)
p6 (20+)
p5 (20+)
p4 (10−19)
p3 (10−19)
p2 (10−19)
p1 (0−9)
1 2 3 4 5 6 7 8 9 10 11 12 13 14 15 16 17 18 19 20 21 22 23 24 25 26 27 28 29 30 31 32 33 34 35 36 37 38 39 40 41 42 43 44
day

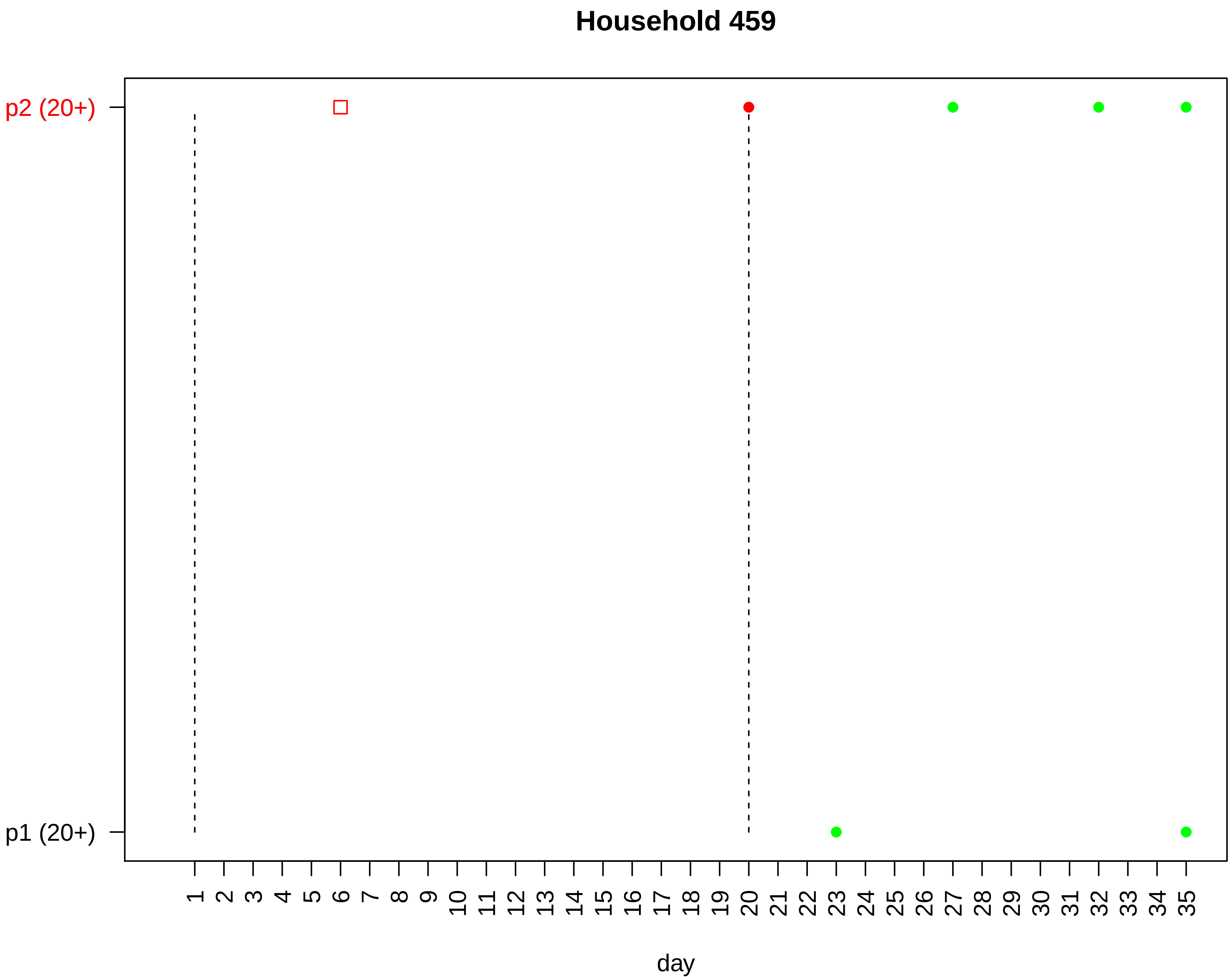

Household 459
p2 (20+)
p1 (20+)
day

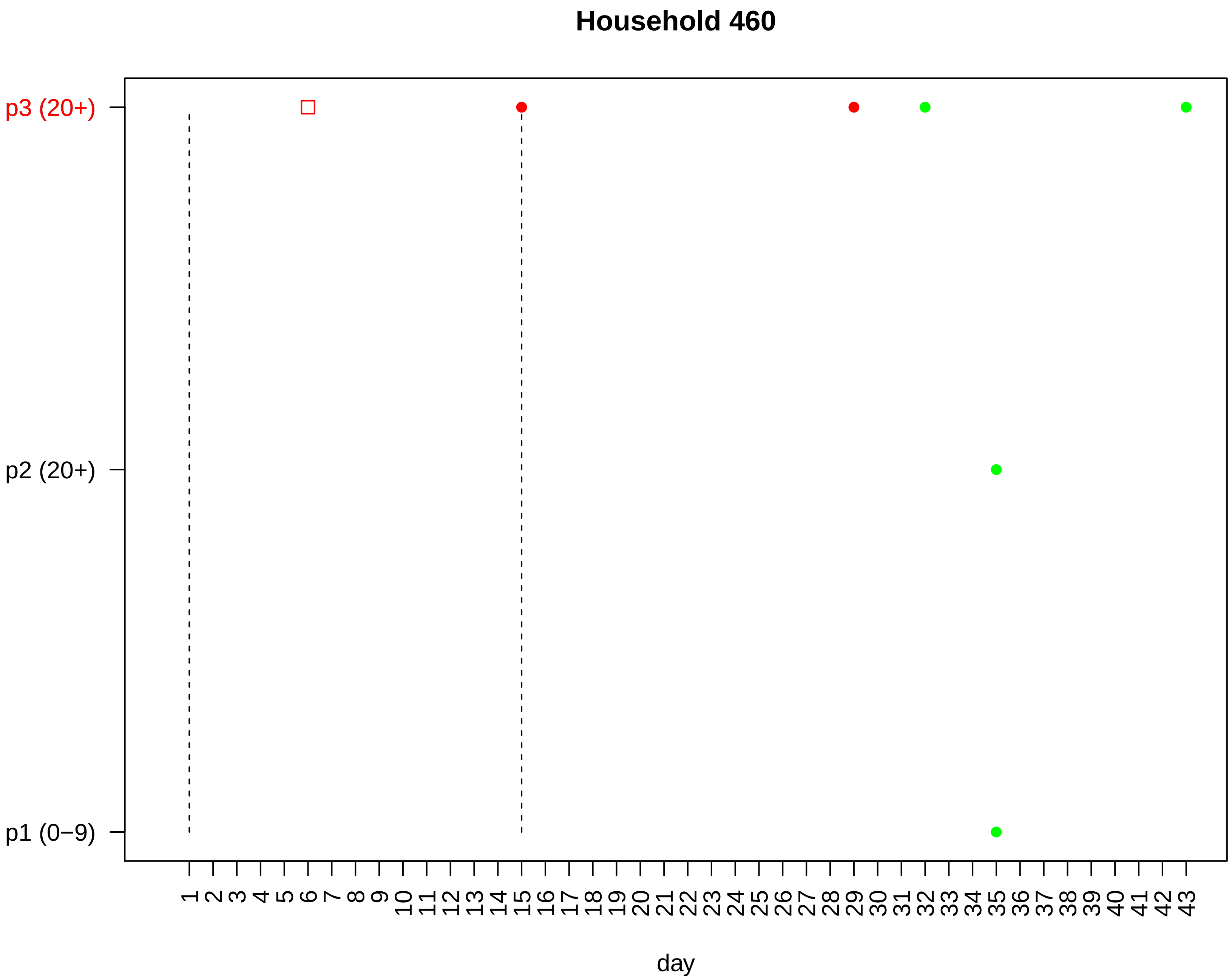

Household 460
p3 (20+)
p2 (20+)
p1 (0−9)
1 2 3 4 5 6 7 8 9 10 11 12 13 14 15 16 17 18 19 20 21 22 23 24 25 26 27 28 29 30 31 32 33 34 35 36 37 38 39 40 41 42 43
day

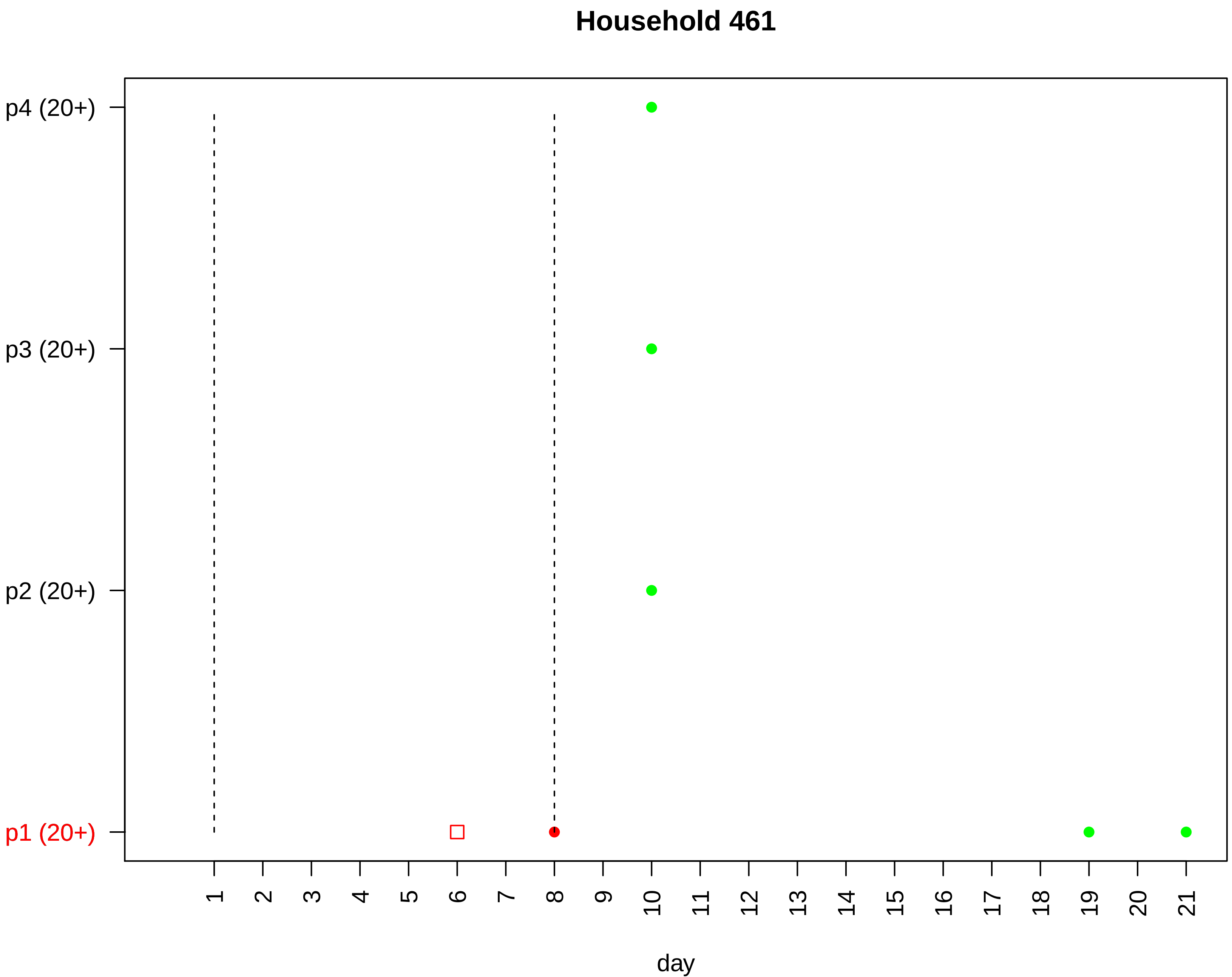

Household 461
p4 (20+)
p3 (20+)
p2 (20+)
p1 (20+)
day

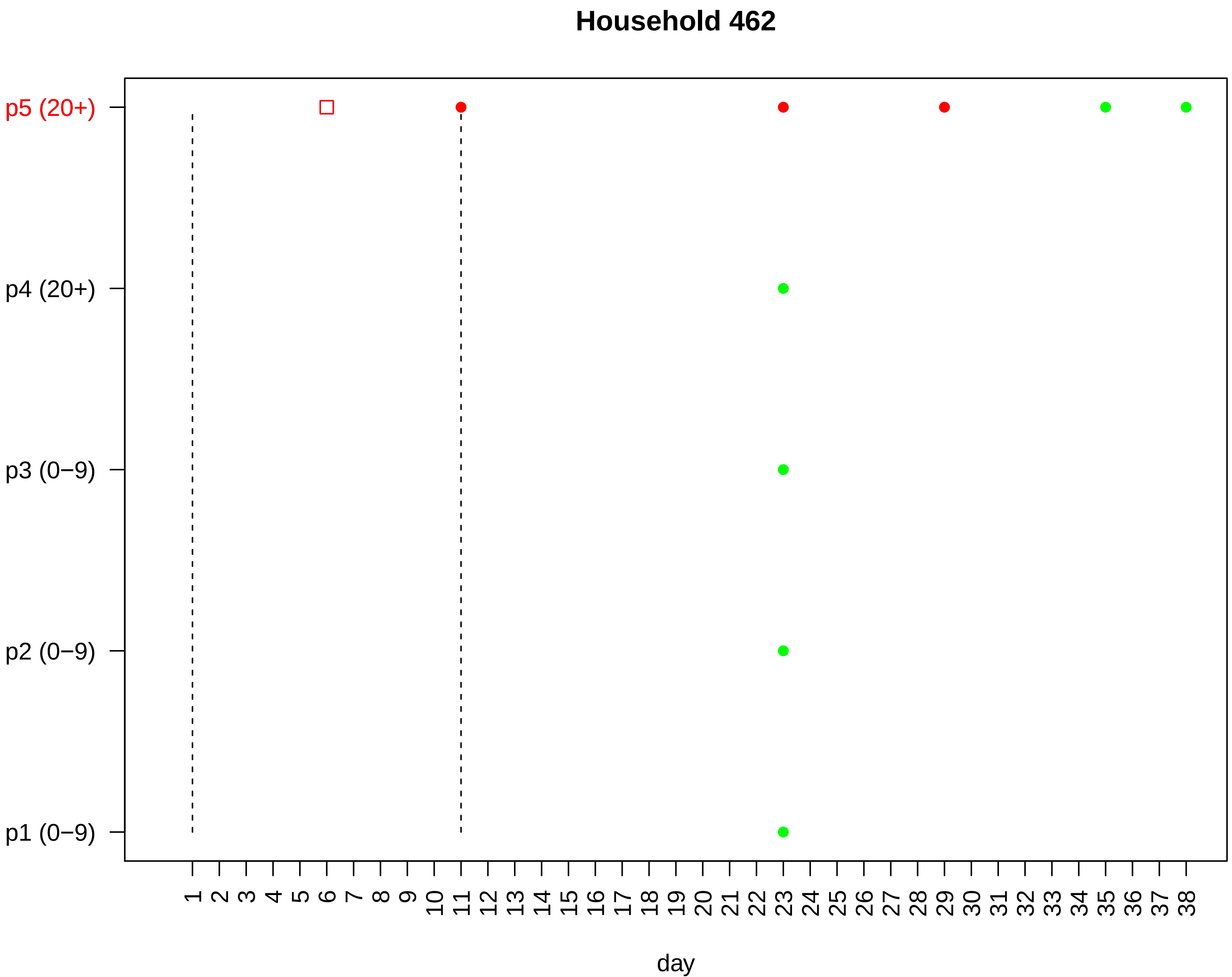

Household 462
p5 (20+)
p4 (20+)
p3 (0−9)
p2 (0−9)
p1 (0−9)
1 2 3 4 5 6 7 8 9 10 11 12 13 14 15 16 17 18 19 20 21 22 23 24 25 26 27 28 29 30 31 32 33 34 35 36 37 38
day

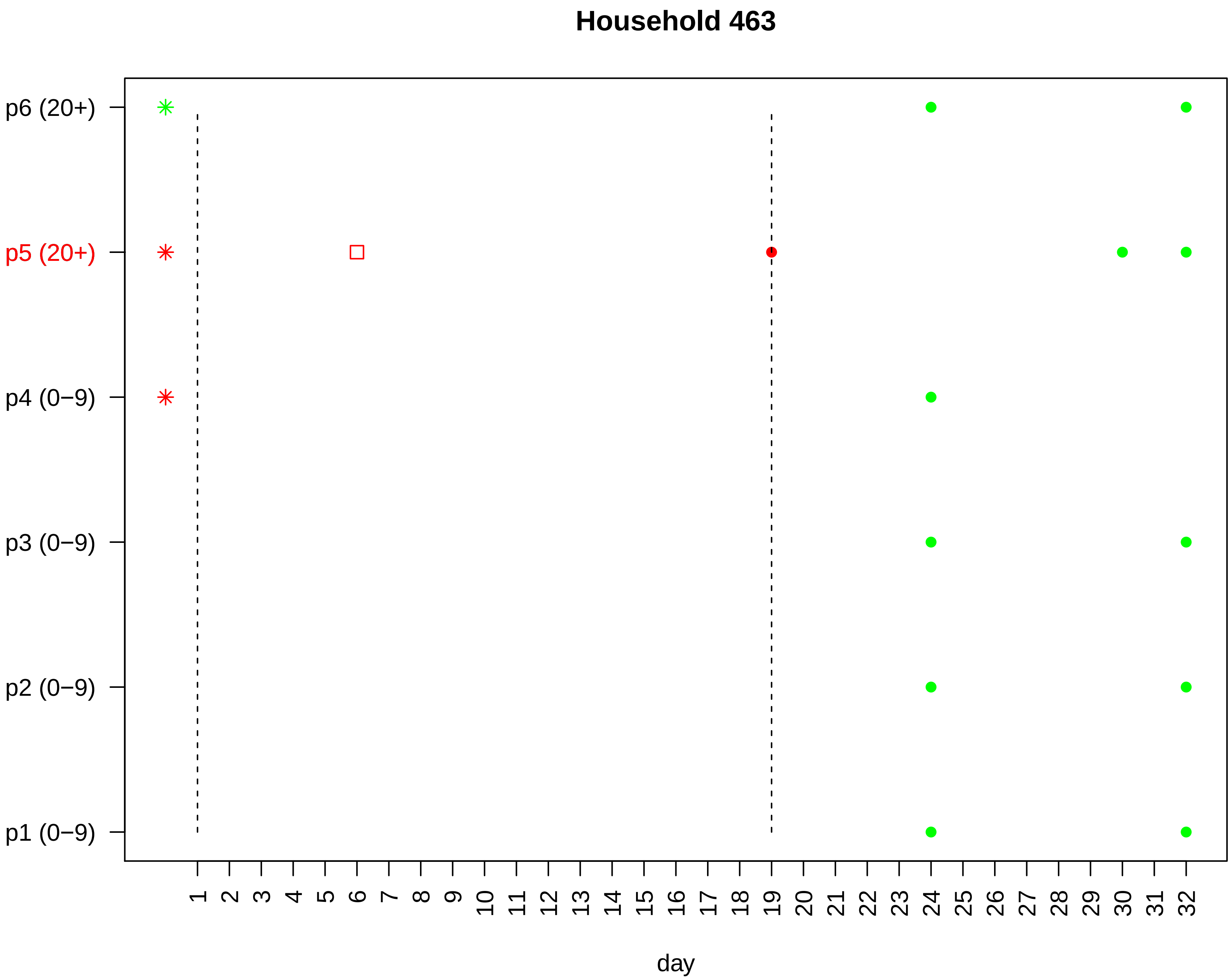

Household 463
p6 (20+)
p5 (20+)
p4 (0−9)
p3 (0−9)
p2 (0−9)
p1 (0−9)
1 2 3 4 5 6 7 8 9 10 11 12 13 14 15 16 17 18 19 20 21 22 23 24 25 26 27 28 29 30 31 32
day

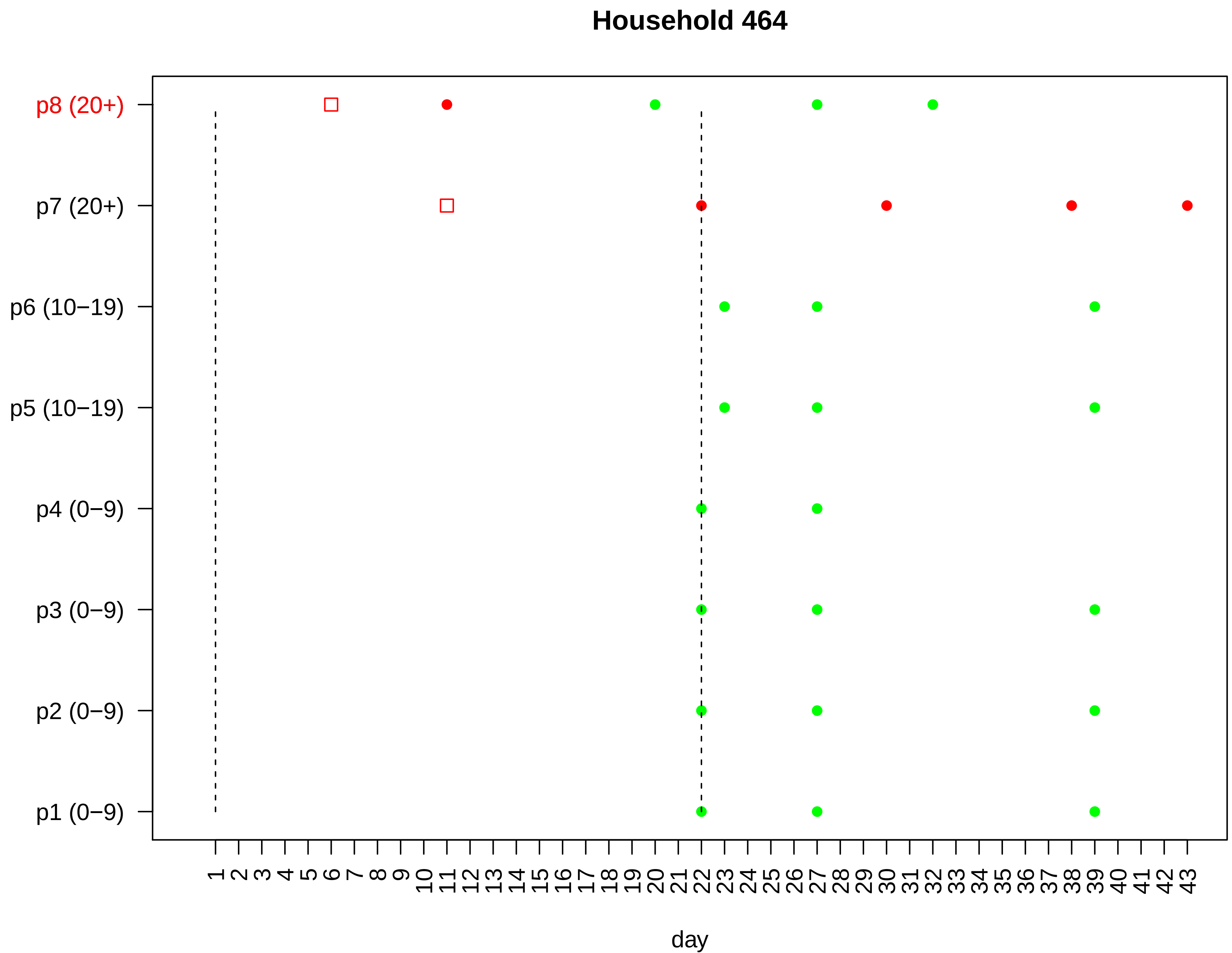
Household 464
p8 (20+)
p7 (20+)
p6 (10−19)
p5 (10−19)
p4 (0−9)
p3 (0−9)
p2 (0−9)
p1 (0−9)
1 2 3 4 5 6 7 8 9 10 11 12 13 14 15 16 17 18 19 20 21 22 23 24 25 26 27 28 29 30 31 32 33 34 35 36 37 38 39 40 41 42 43
day

### Household 465

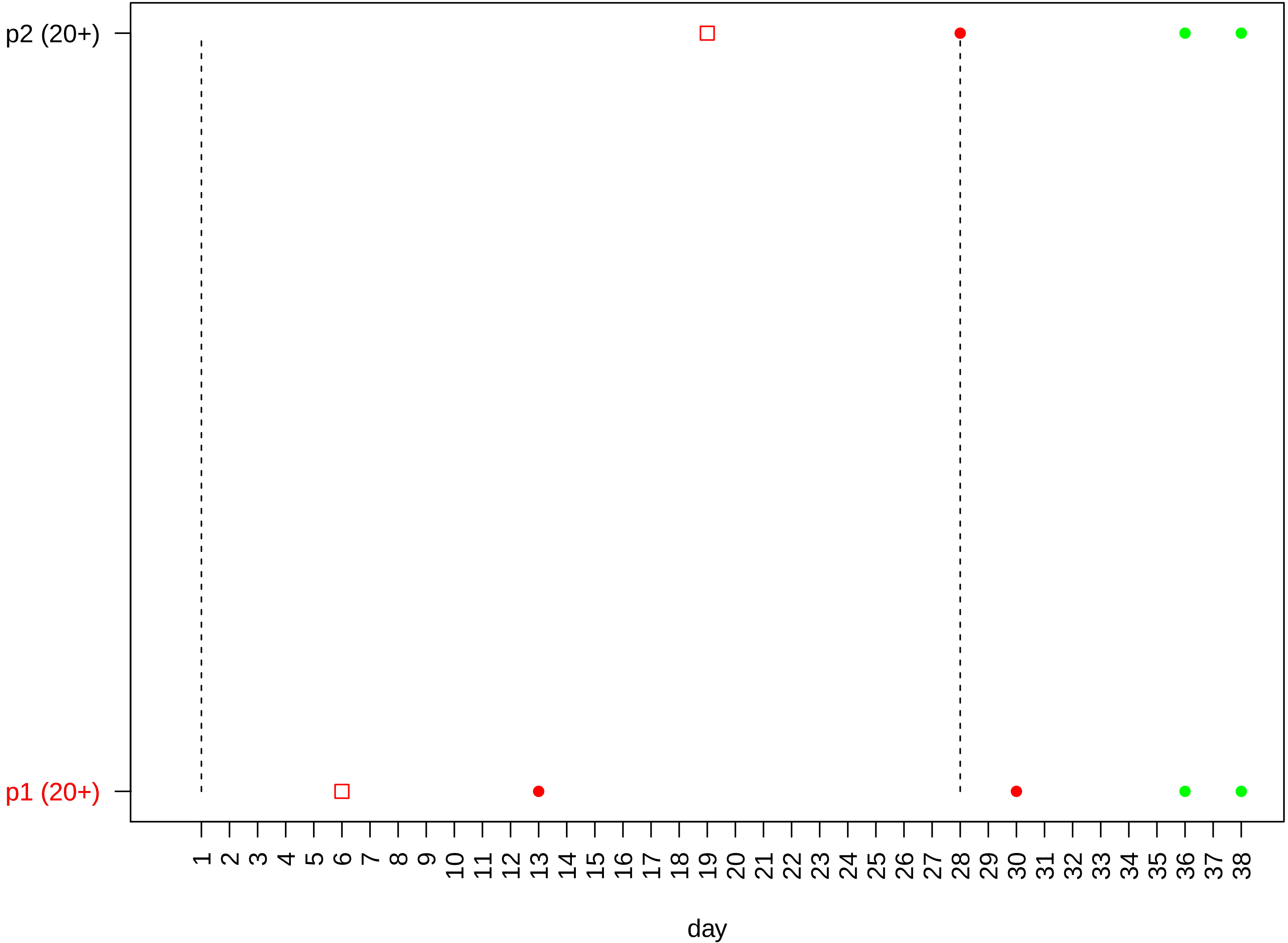

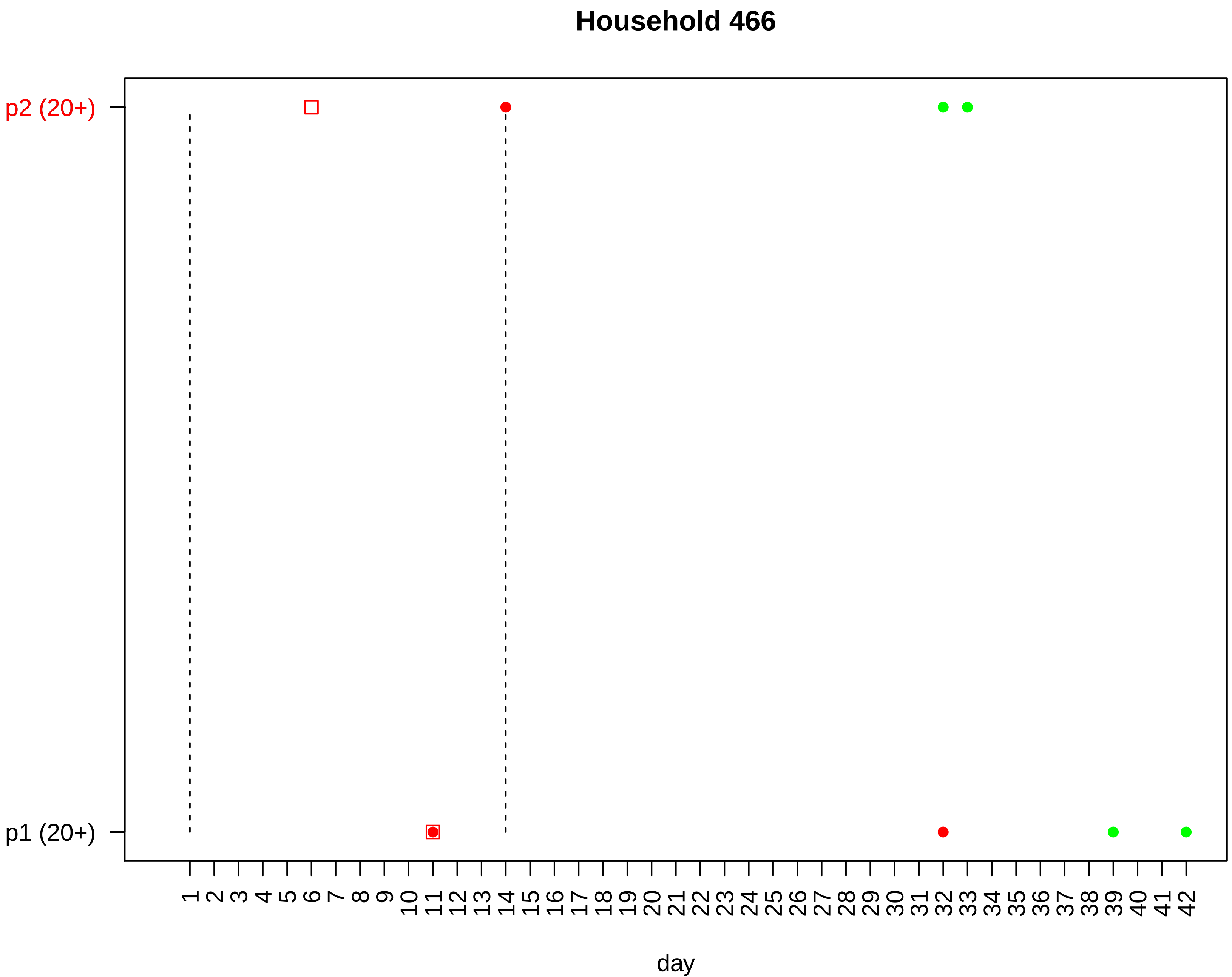

Household 466
p2 (20+)
p1 (20+)
1 2 3 4 5 6 7 8 9 10 11 12 13 14 15 16 17 18 19 20 21 22 23 24 25 26 27 28 29 30 31 32 33 34 35 36 37 38 39 40 41 42
day

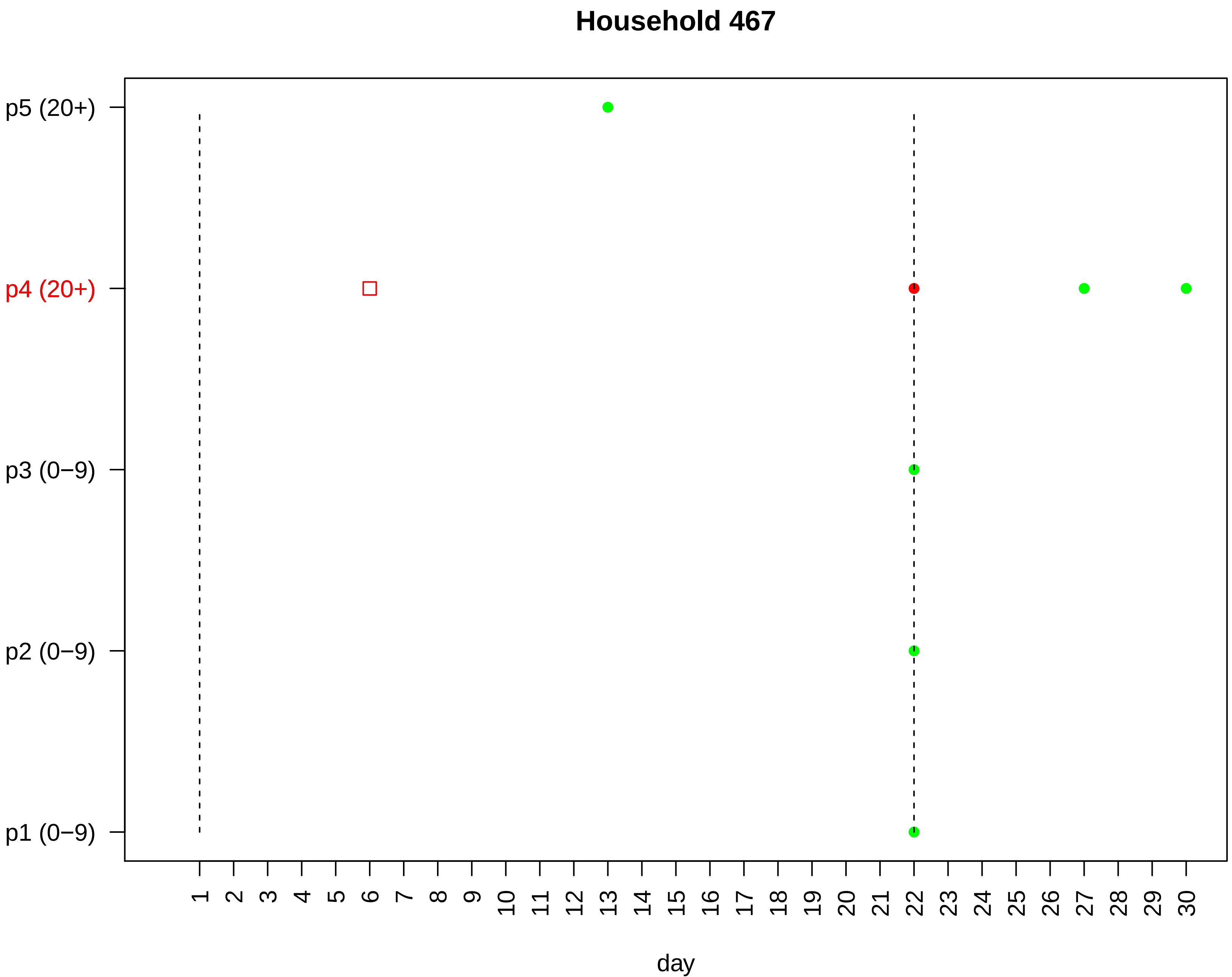

Household 467
p5 (20+)
p4 (20+)
p3 (0−9)
p2 (0−9)
p1 (0−9)
day

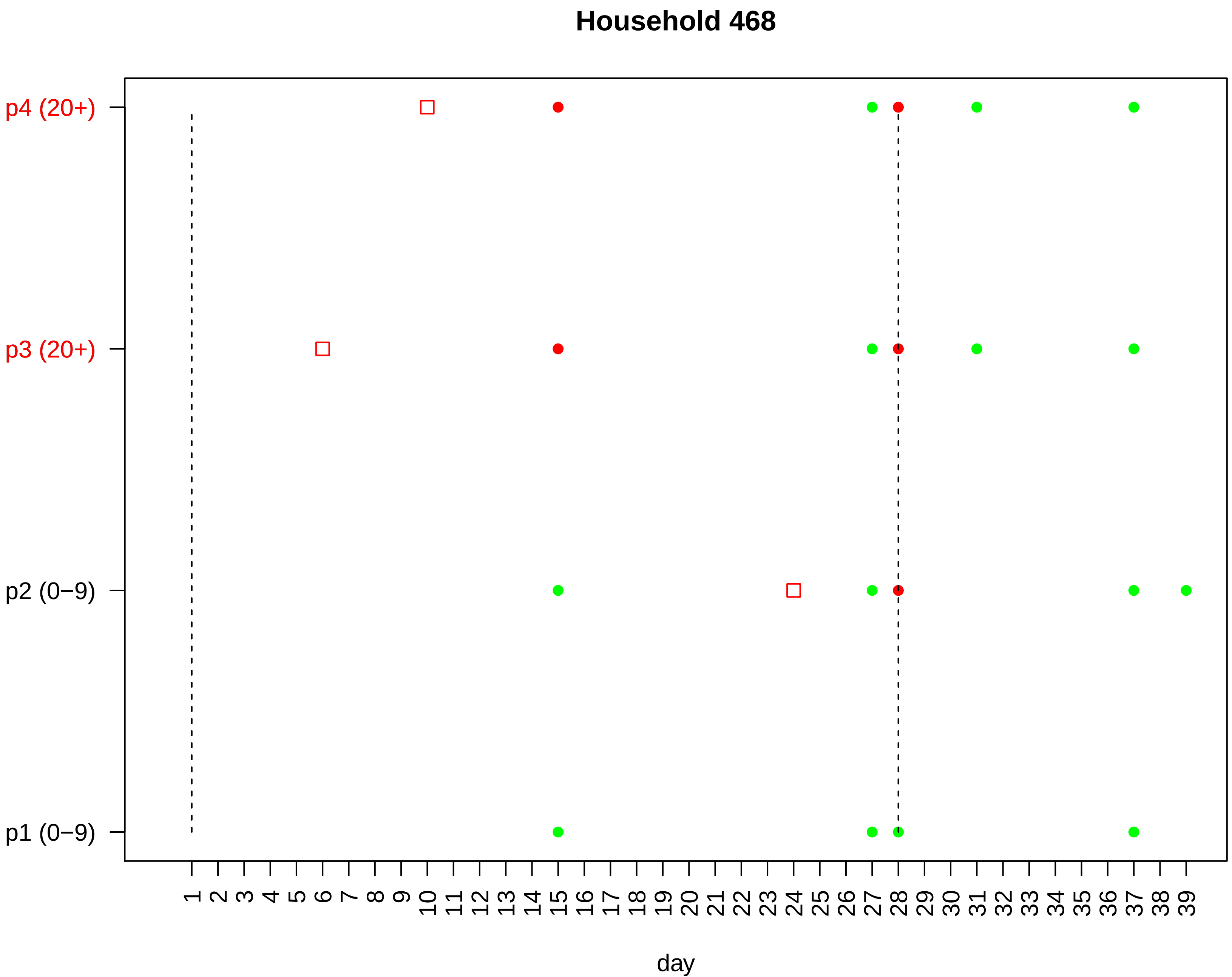
Household 468
p4 (20+)
p3 (20+)
p2 (0−9)
p1 (0−9)
1 2 3 4 5 6 7 8 9 10 11 12 13 14 15 16 17 18 19 20 21 22 23 24 25 26 27 28 29 30 31 32 33 34 35 36 37 38 39
day

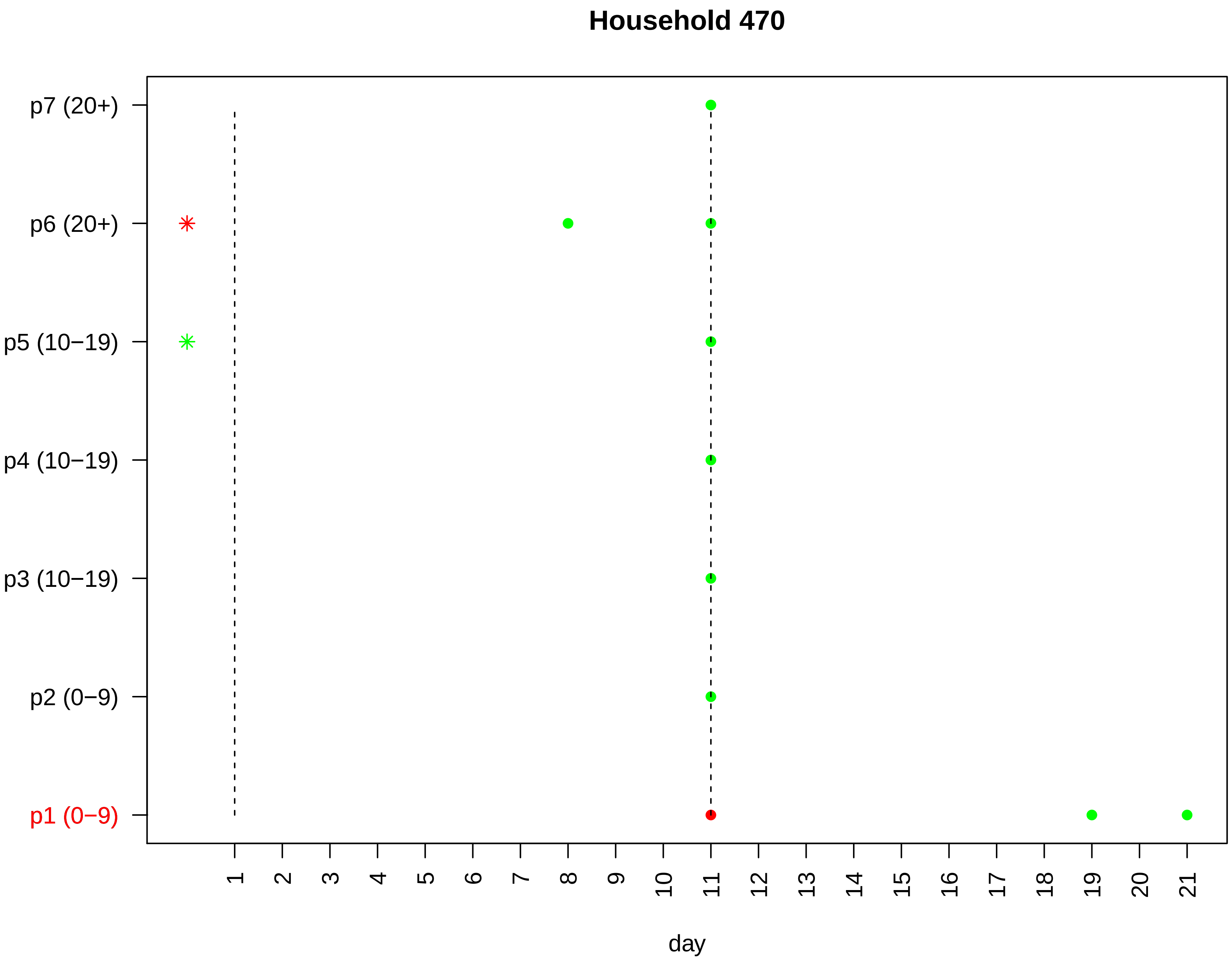

Household 470
p7 (20+)
p6 (20+)
p5 (10−19)
p4 (10−19)
p3 (10−19)
p2 (0−9)
p1 (0−9)
day

#### Household 471

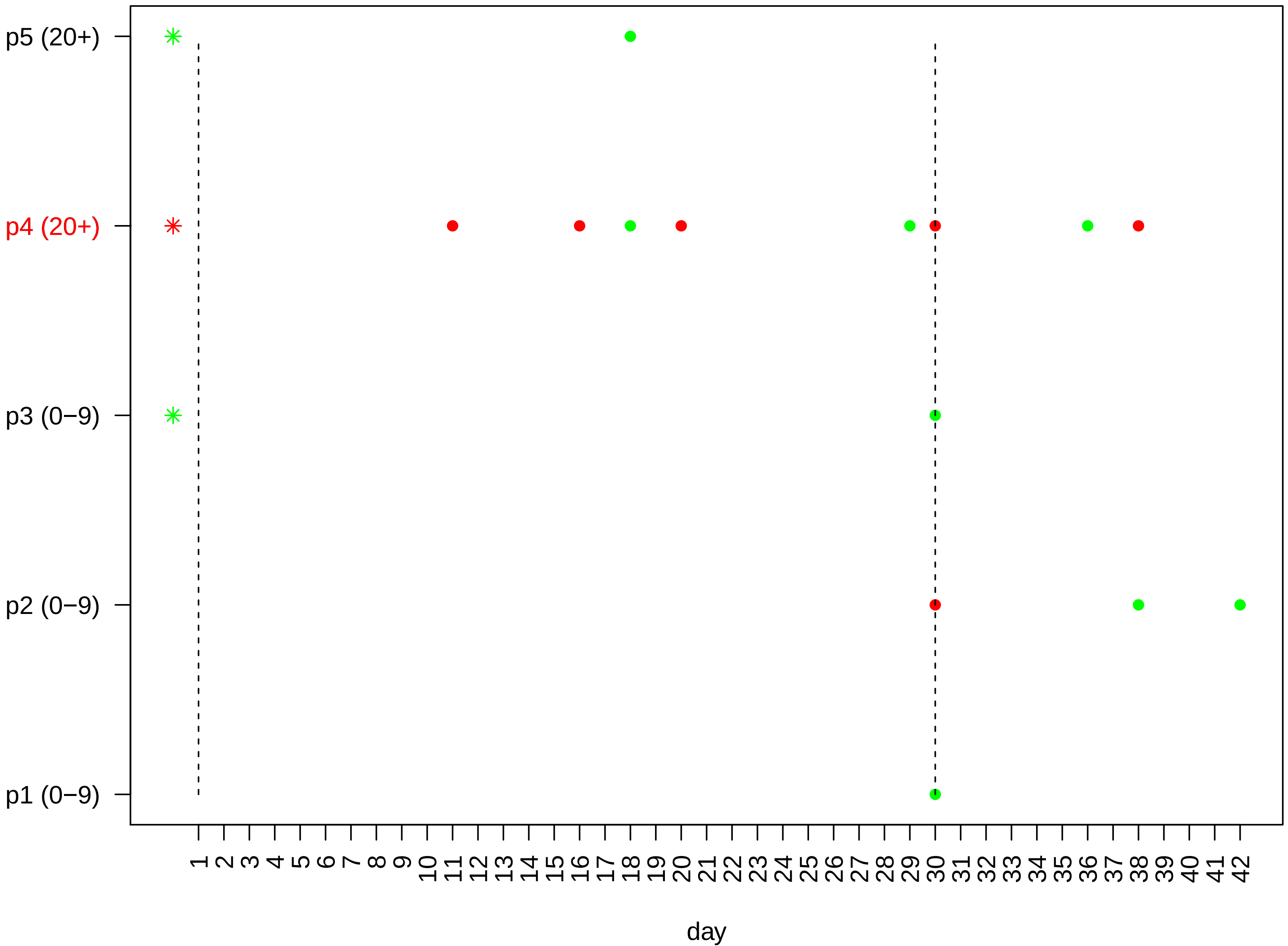

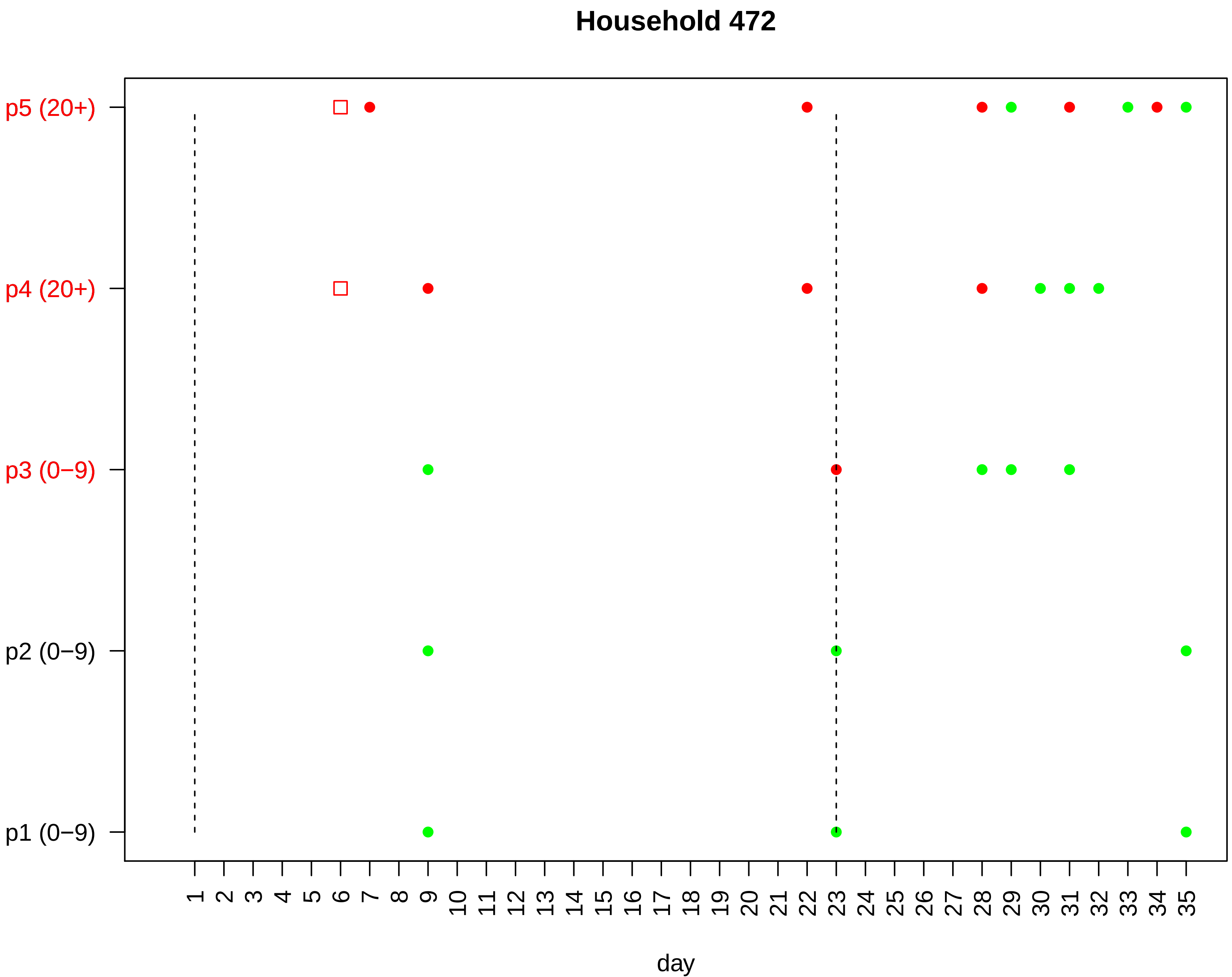

Household 472
p5 (20+)
p4 (20+)
p3 (0−9)
p2 (0−9)
p1 (0−9)
1 2 3 4 5 6 7 8 9 10 11 12 13 14 15 16 17 18 19 20 21 22 23 24 25 26 27 28 29 30 31 32 33 34 35
day

### Household 473

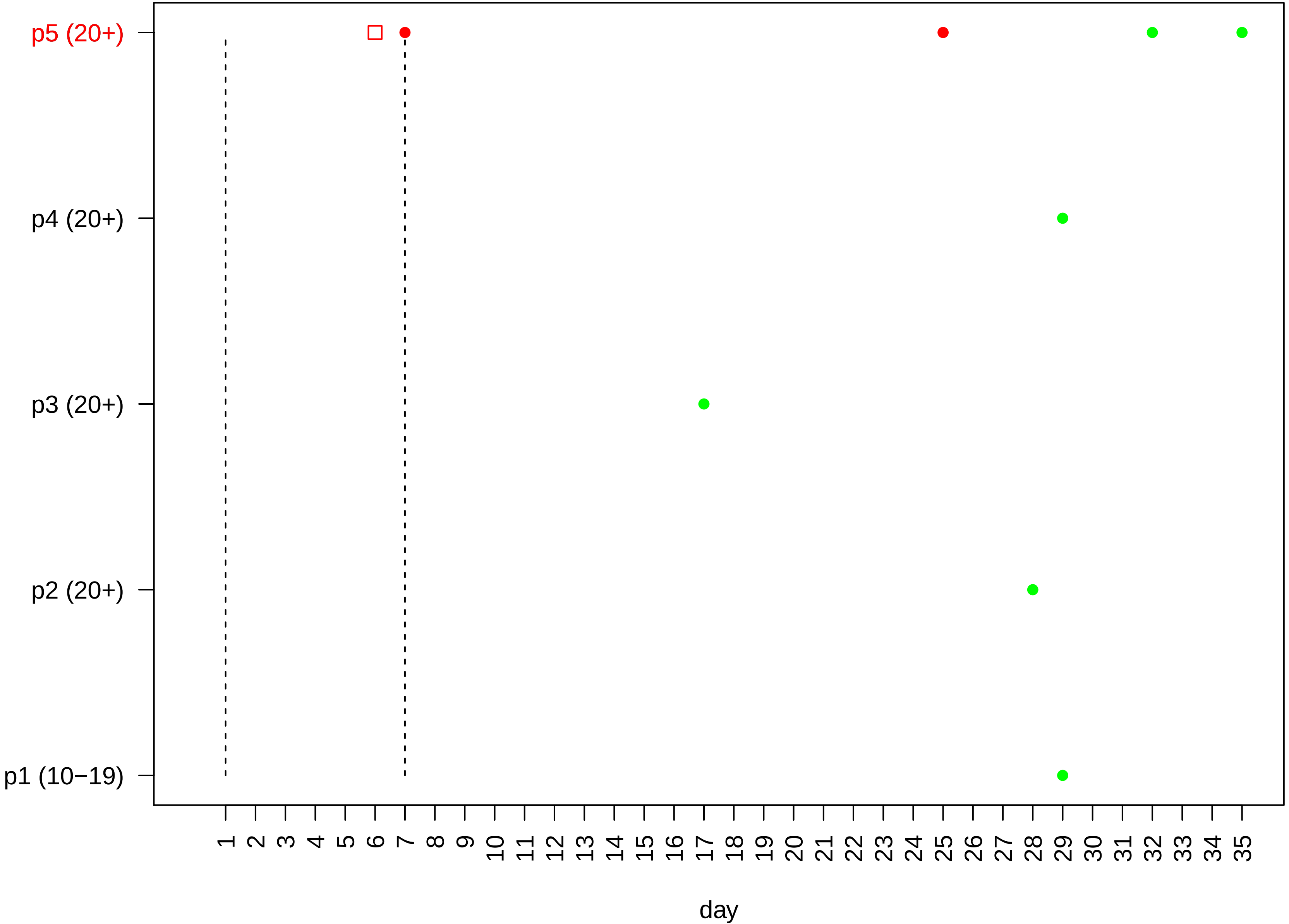

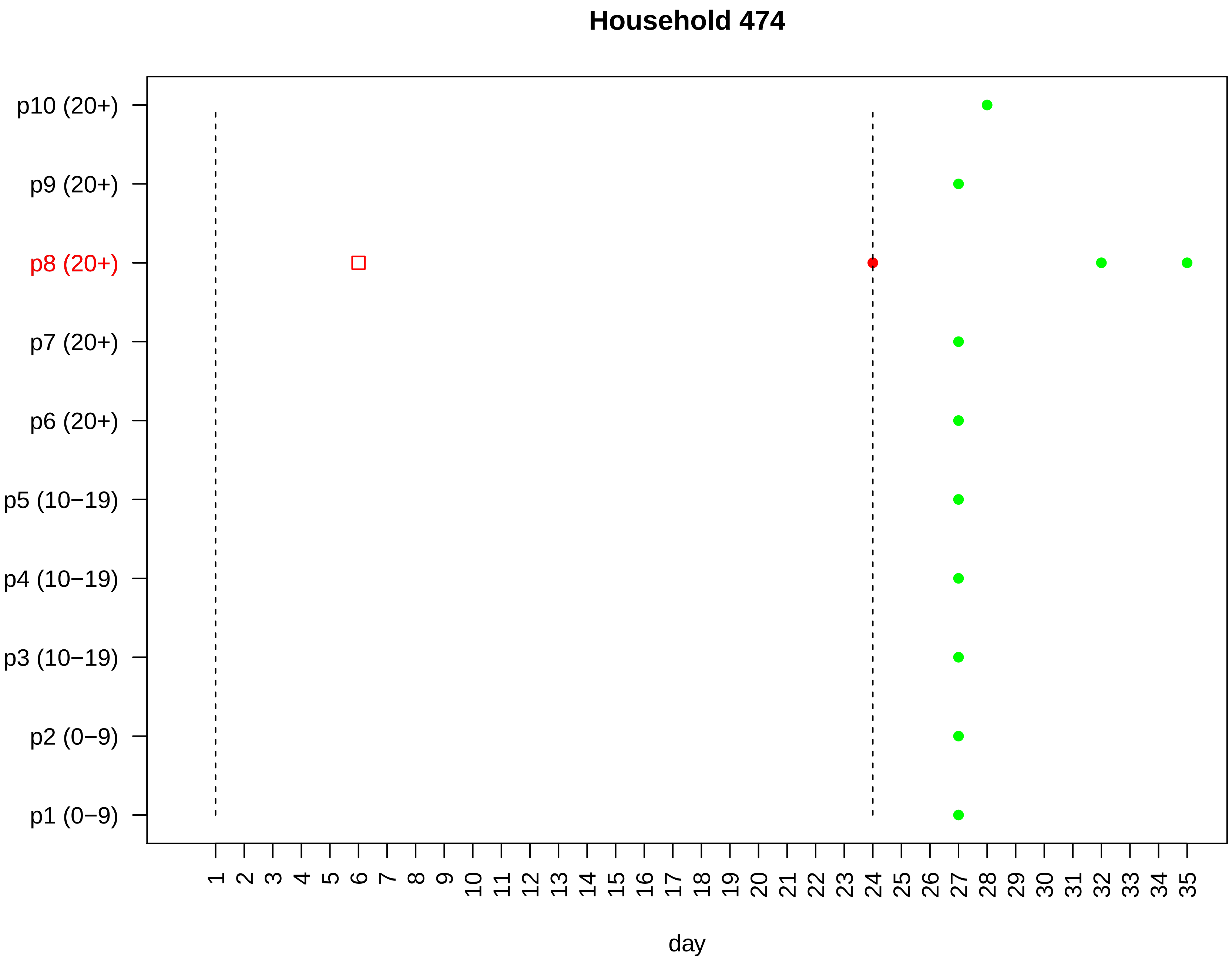

Household 474
p10 (20+)
p9 (20+)
p8 (20+)
p7 (20+)
p6 (20+)
p5 (10−19)
p4 (10−19)
p3 (10−19)
p2 (0−9)
p1 (0−9)
1 2 3 4 5 6 7 8 9 10 11 12 13 14 15 16 17 18 19 20 21 22 23 24 25 26 27 28 29 30 31 32 33 34 35
day

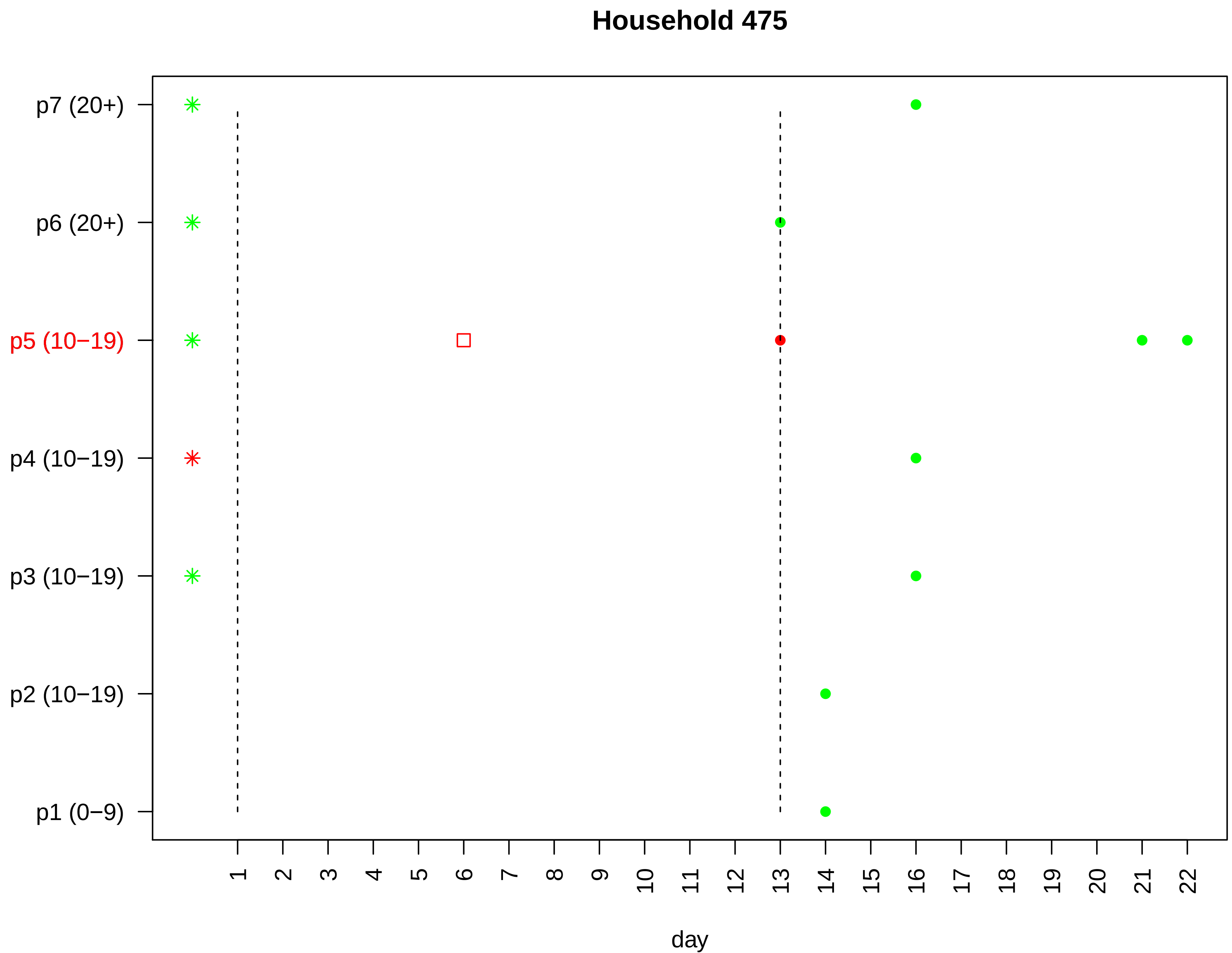

Household 475
p7 (20+)
p6 (20+)
p5 (10−19)
p4 (10−19)
p3 (10−19)
p2 (10−19)
p1 (0−9)
day

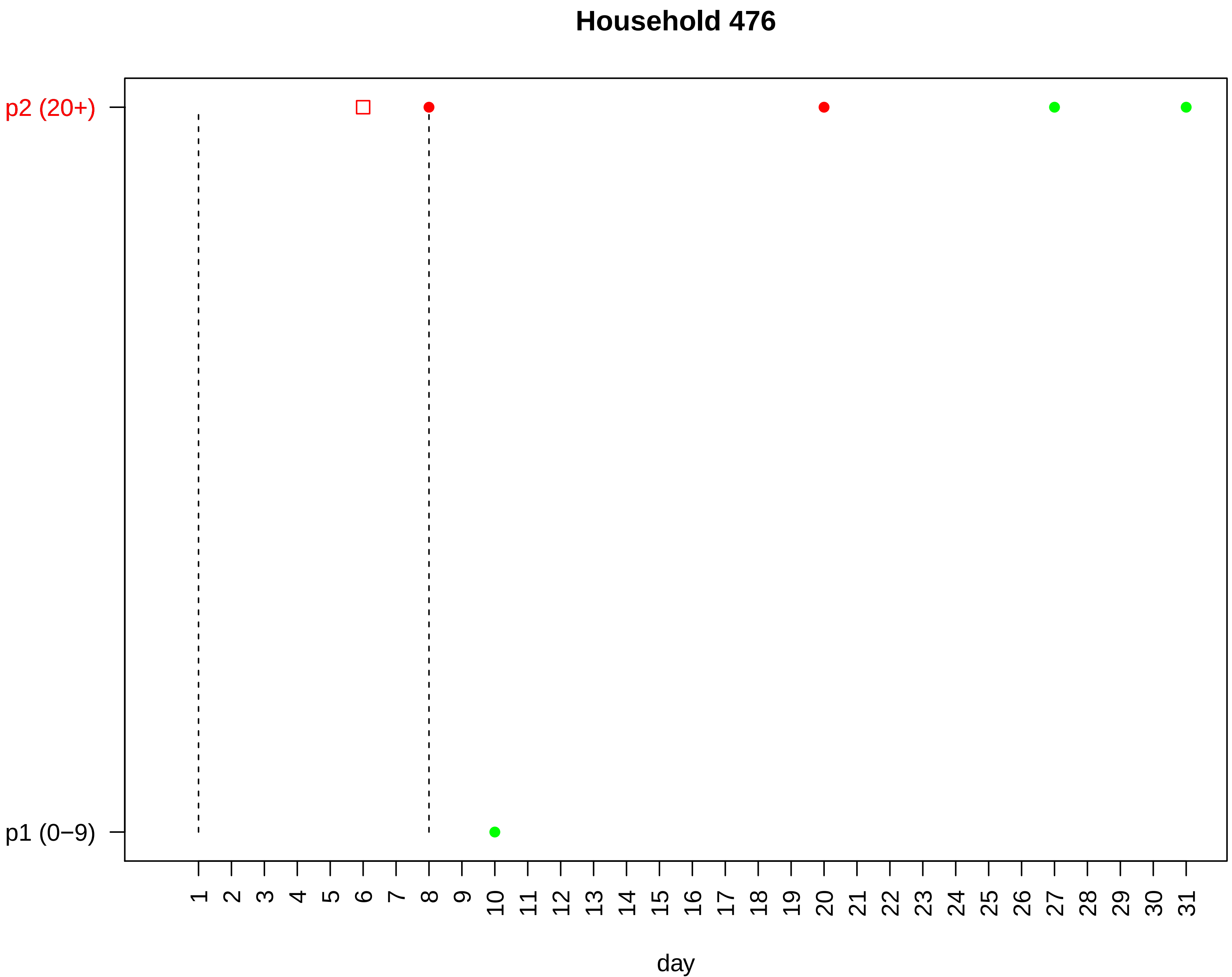

Household 476
p2 (20+)
p1 (0−9)
day

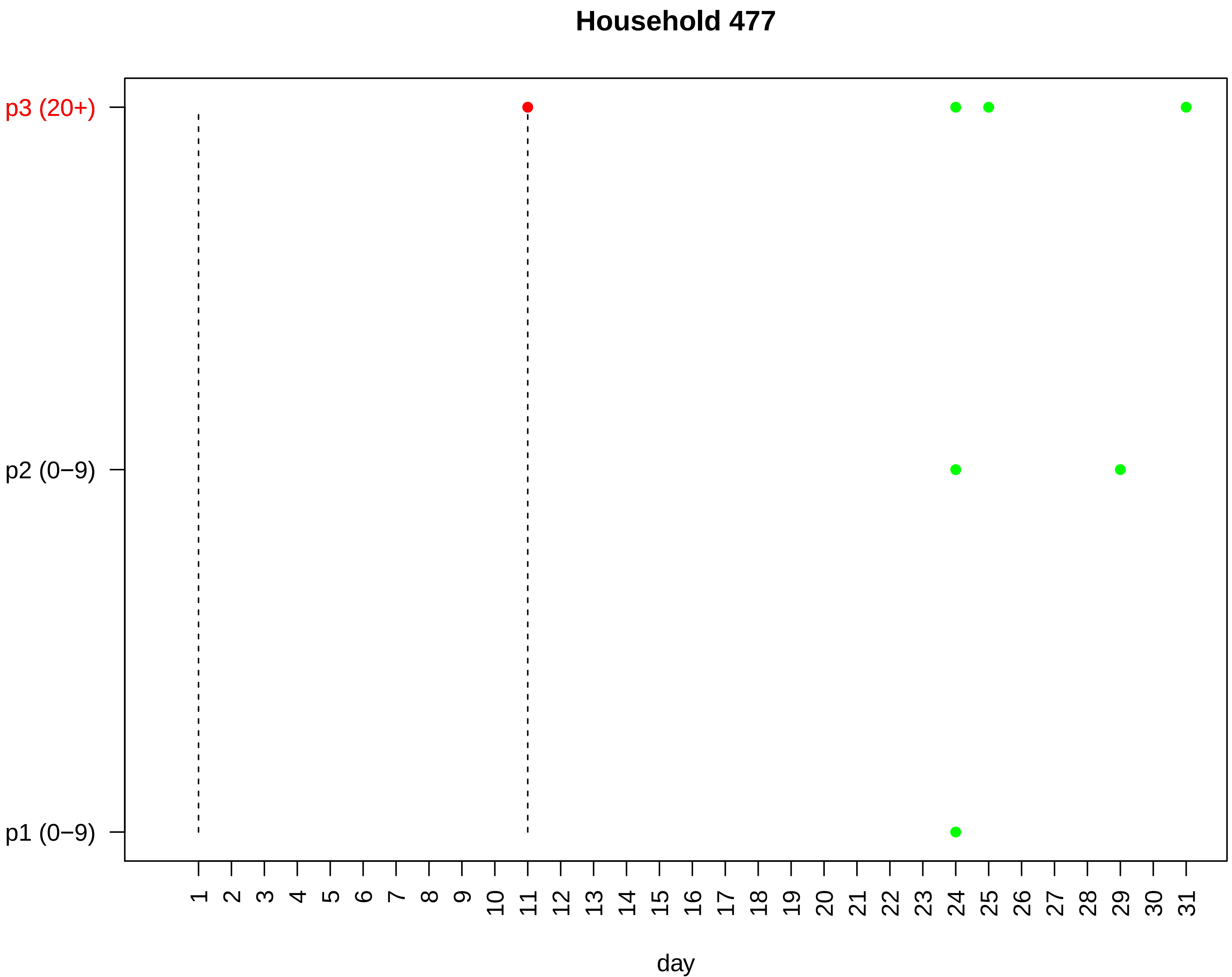
Household 477
p3 (20+)
p2 (0−9)
p1 (0−9)
day

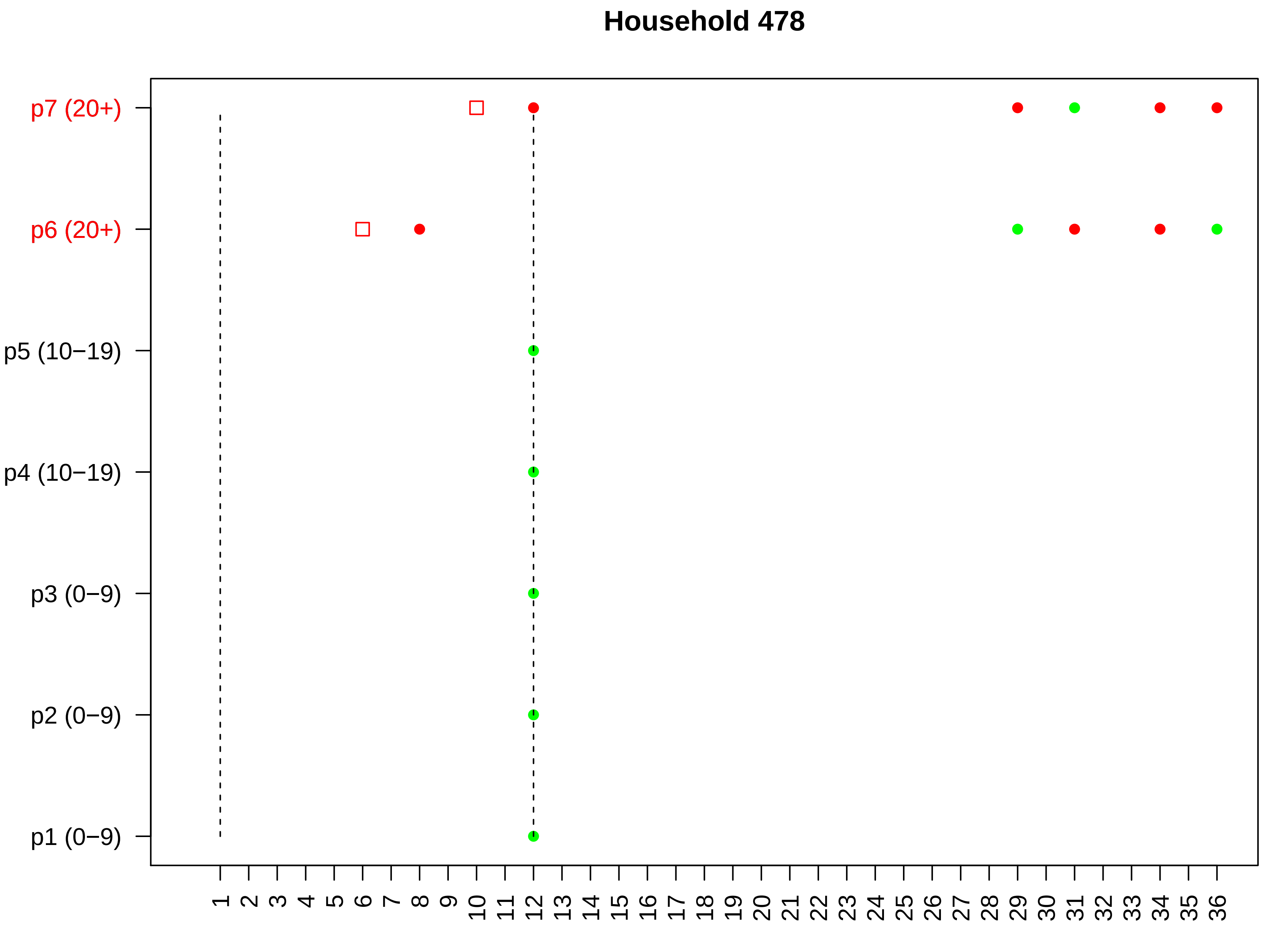

Household 478
p7 (20+)
p6 (20+)
p5 (10−19)
p4 (10−19)
p3 (0−9)
p2 (0−9)
p1 (0−9)
1 2 3 4 5 6 7 8 9 10 11 12 13 14 15 16 17 18 19 20 21 22 23 24 25 26 27 28 29 30 31 32 33 34 35 36
day

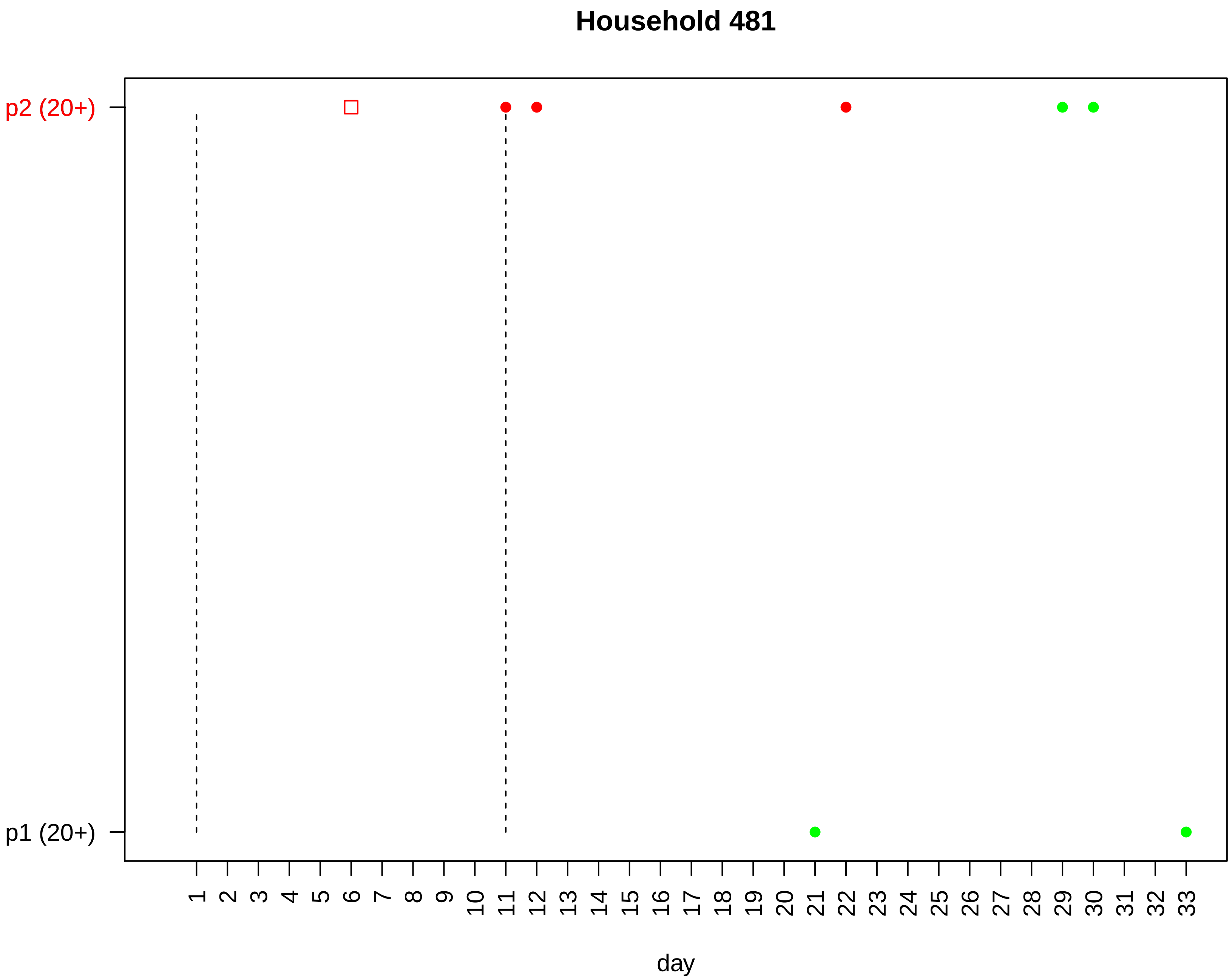

Household 481
p2 (20+)
p1 (20+)
day

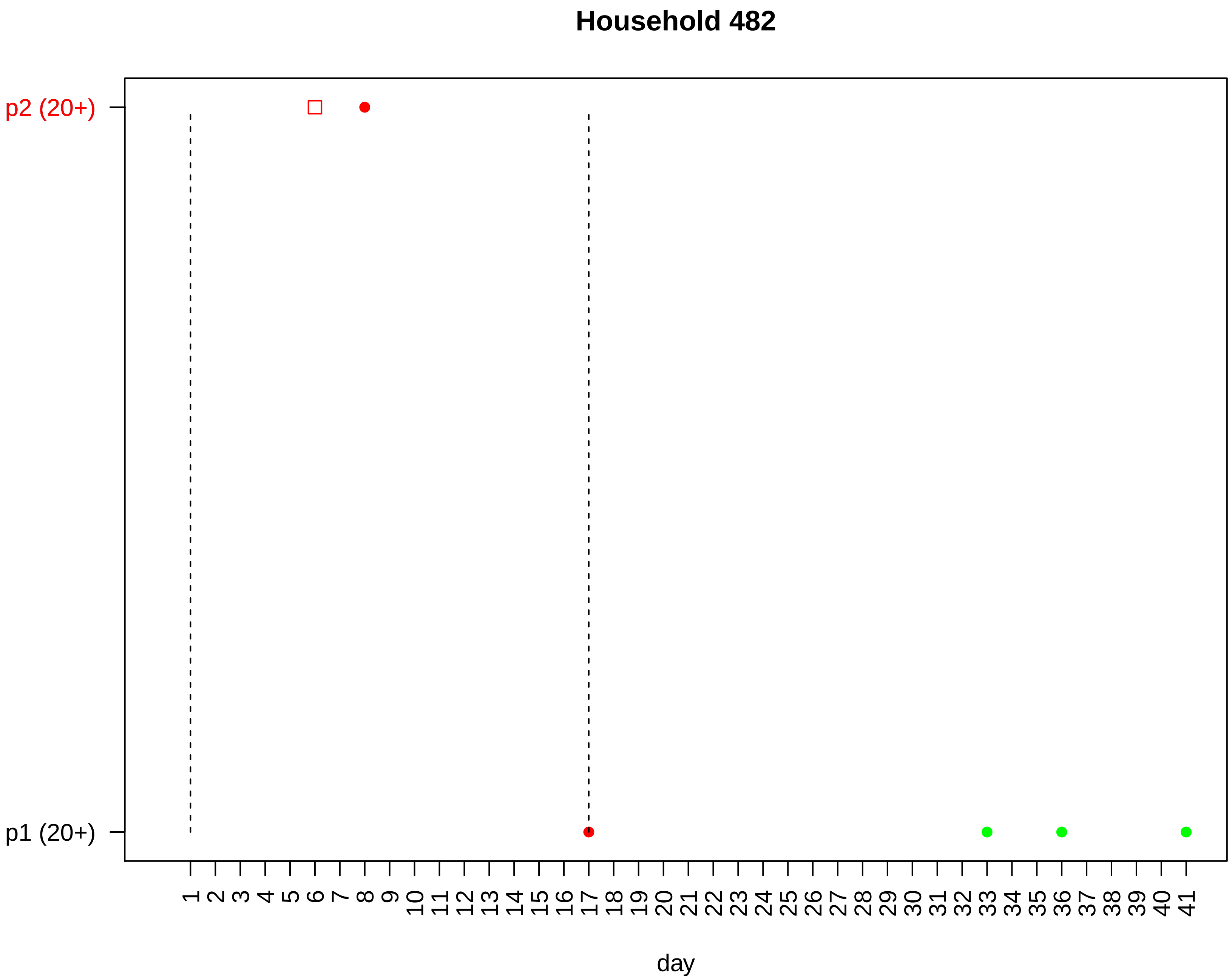

Household 482
p2 (20+)
p1 (20+)
day

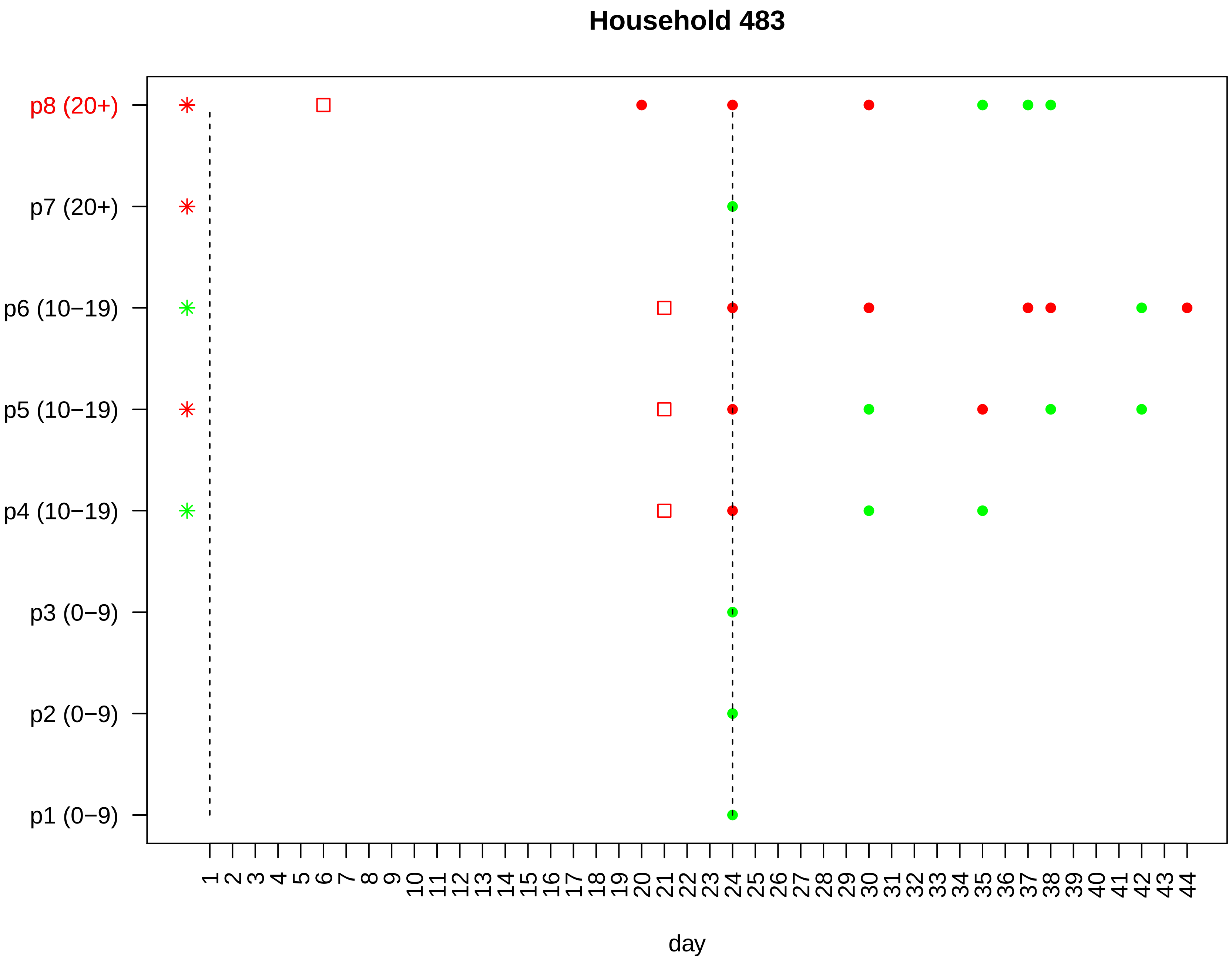
Household 483
p8 (20+)
p7 (20+)
p6 (10−19)
p5 (10−19)
p4 (10−19)
p3 (0−9)
p2 (0−9)
p1 (0−9)
1 2 3 4 5 6 7 8 9 10 11 12 13 14 15 16 17 18 19 20 21 22 23 24 25 26 27 28 29 30 31 32 33 34 35 36 37 38 39 40 41 42 43 44
day

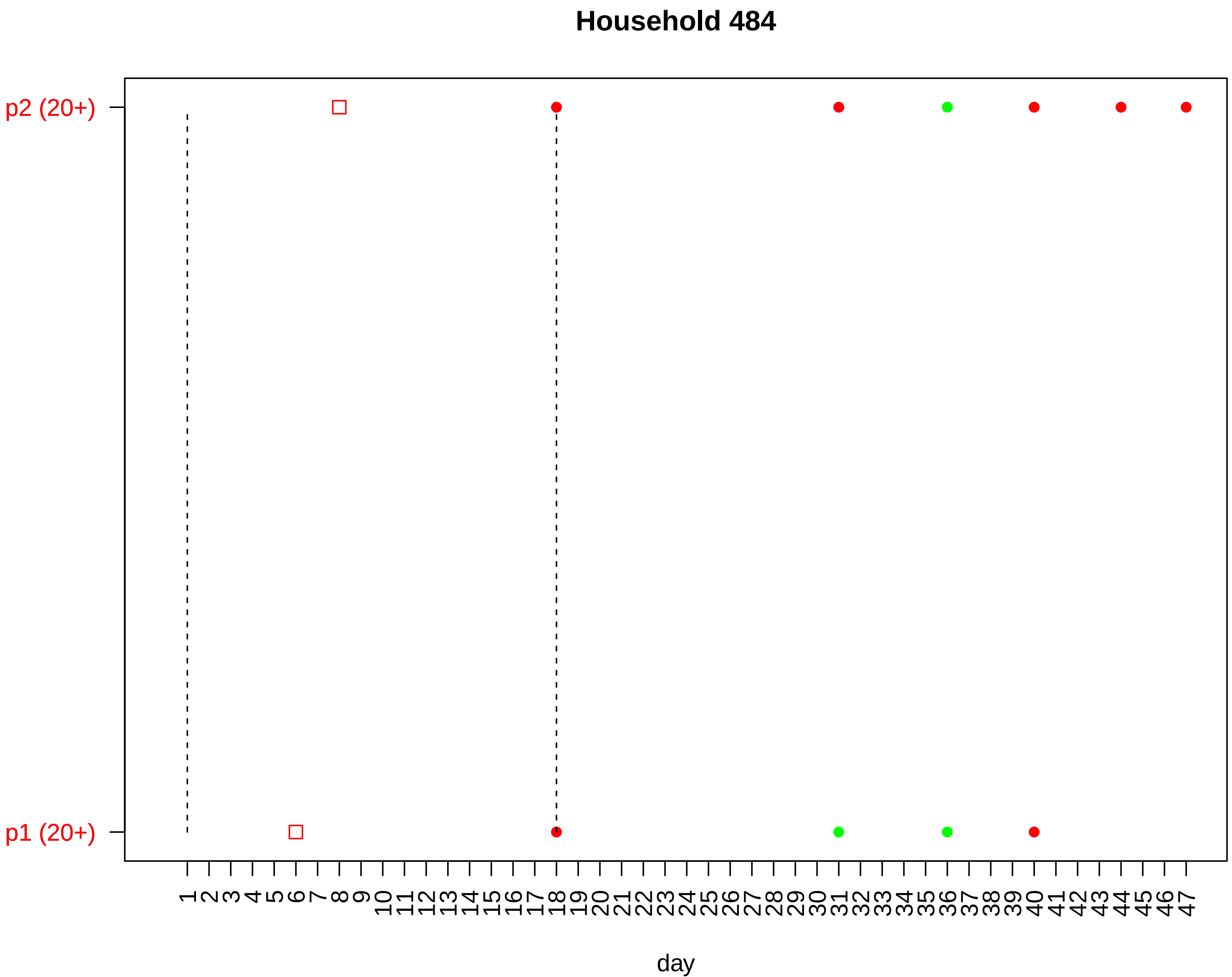

Household 484
p2 (20+)
p1 (20+)
day

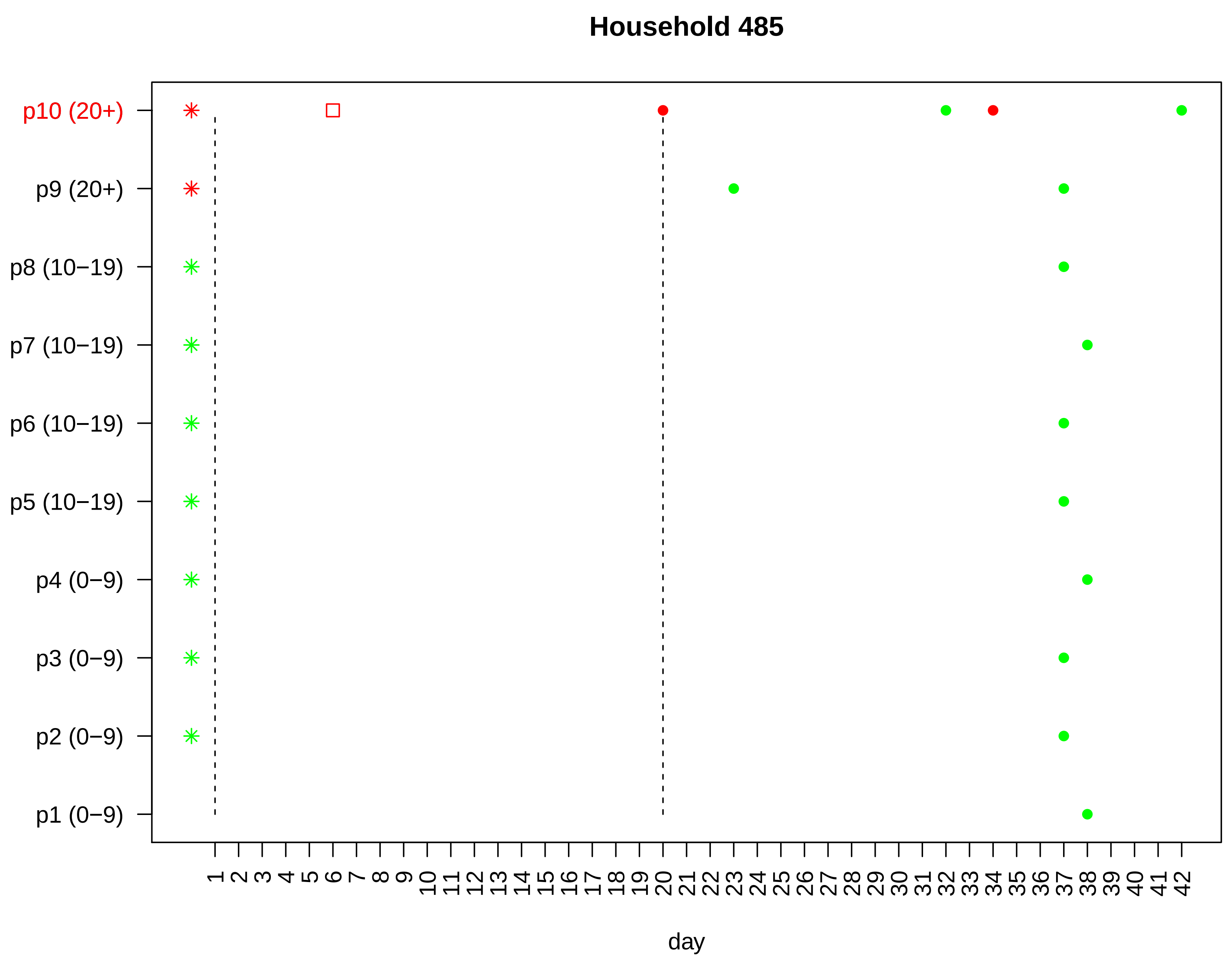

Household 485
p10 (20+)
p9 (20+)
p8 (10−19)
p7 (10−19)
p6 (10−19)
p5 (10−19)
p4 (0−9)
p3 (0−9)
p2 (0−9)
p1 (0−9)
1 2 3 4 5 6 7 8 9 10 11 12 13 14 15 16 17 18 19 20 21 22 23 24 25 26 27 28 29 30 31 32 33 34 35 36 37 38 39 40 41 42
day

#### Household 486

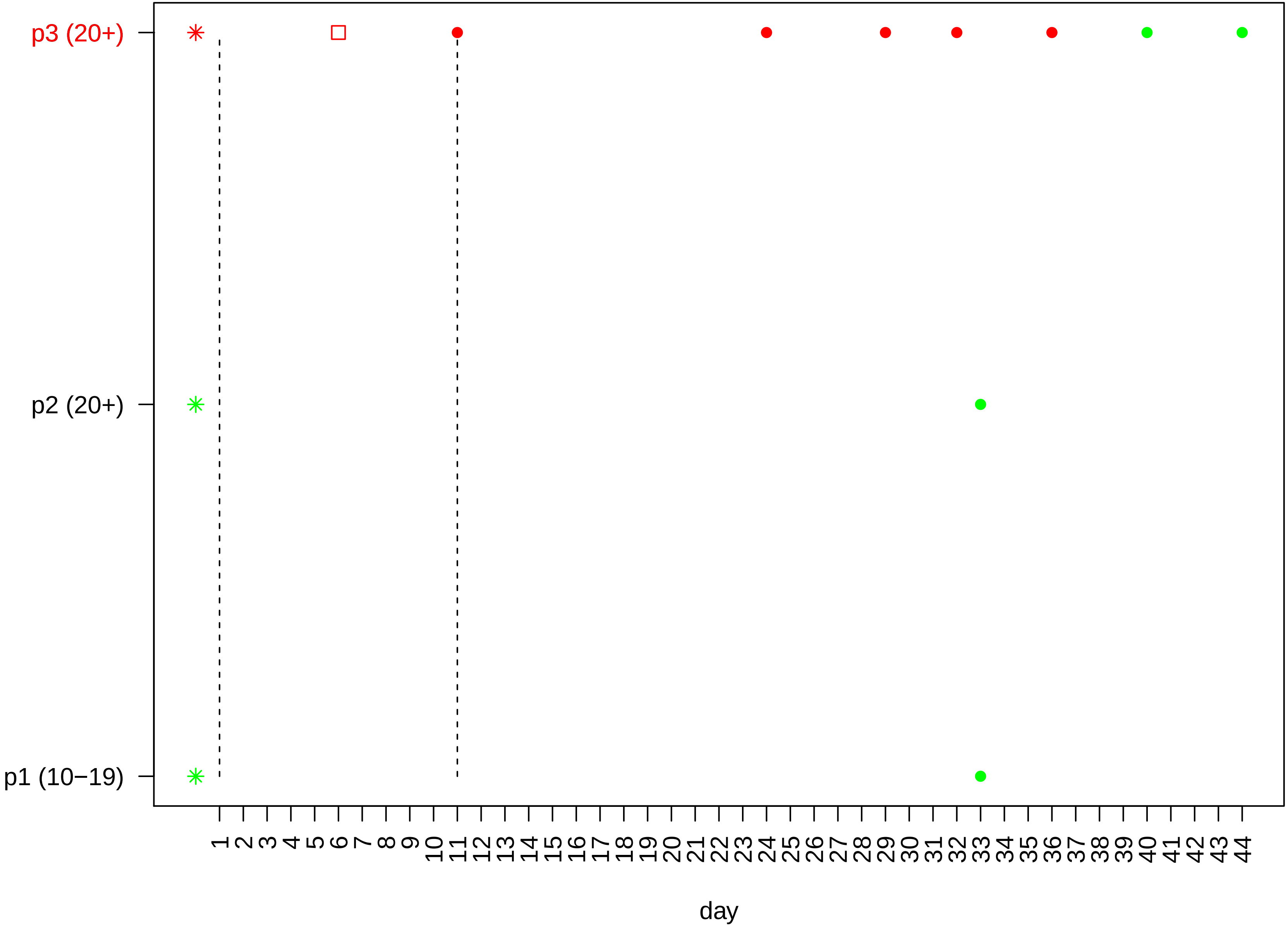

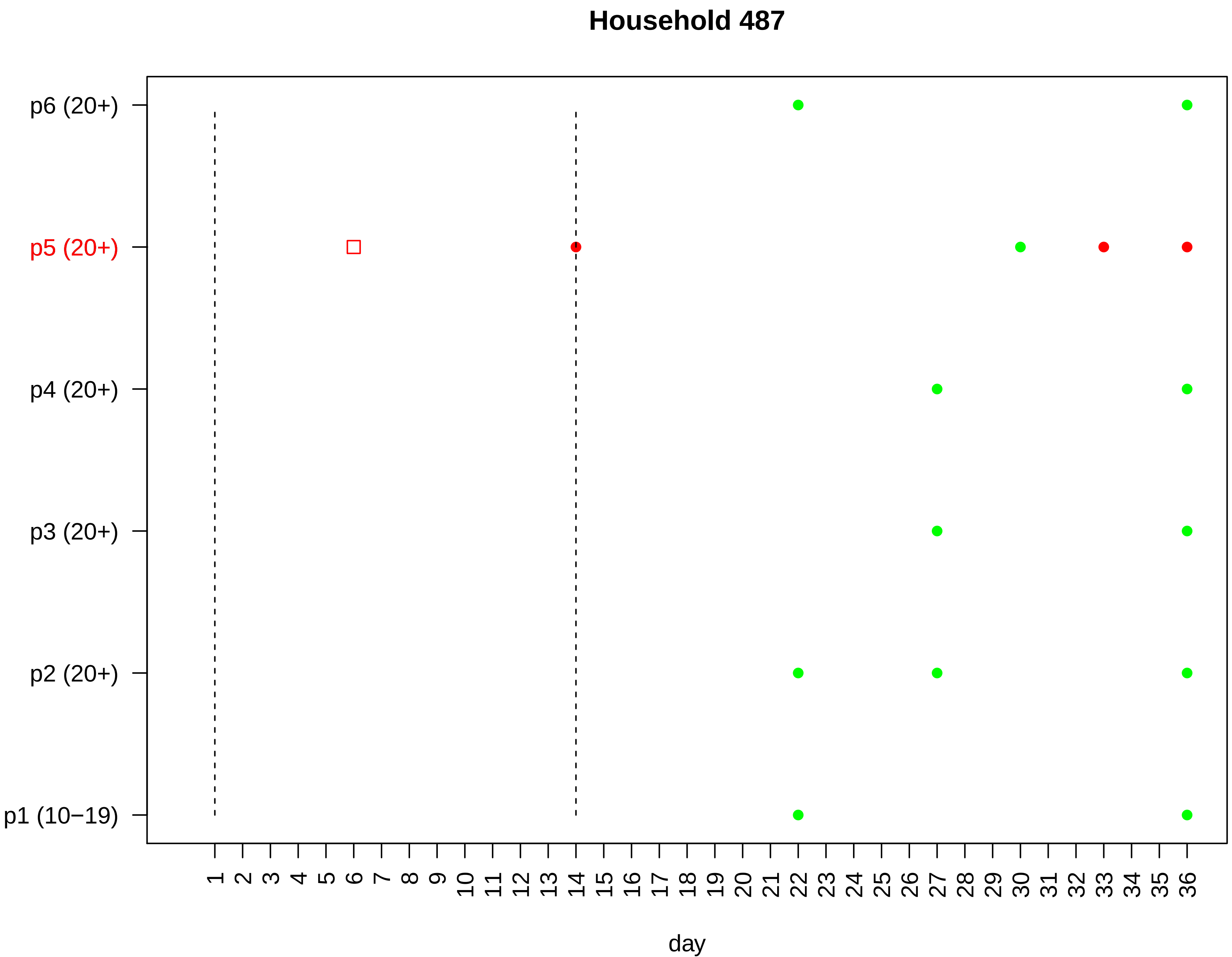

Household 487
p6 (20+)
p5 (20+)
p4 (20+)
p3 (20+)
p2 (20+)
p1 (10−19)
1 2 3 4 5 6 7 8 9 10 11 12 13 14 15 16 17 18 19 20 21 22 23 24 25 26 27 28 29 30 31 32 33 34 35 36
day

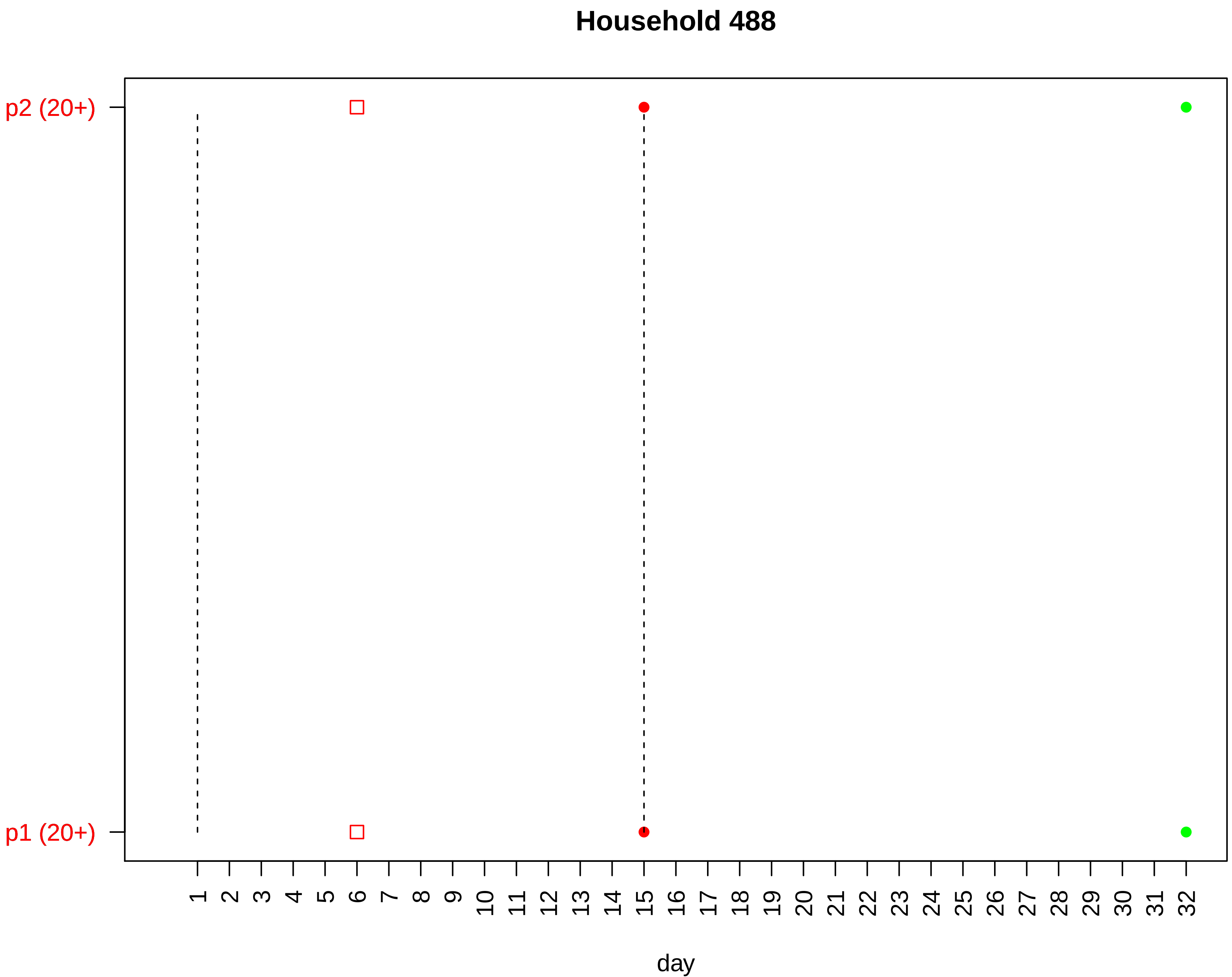
Household 488
p2 (20+)
p1 (20+)
day

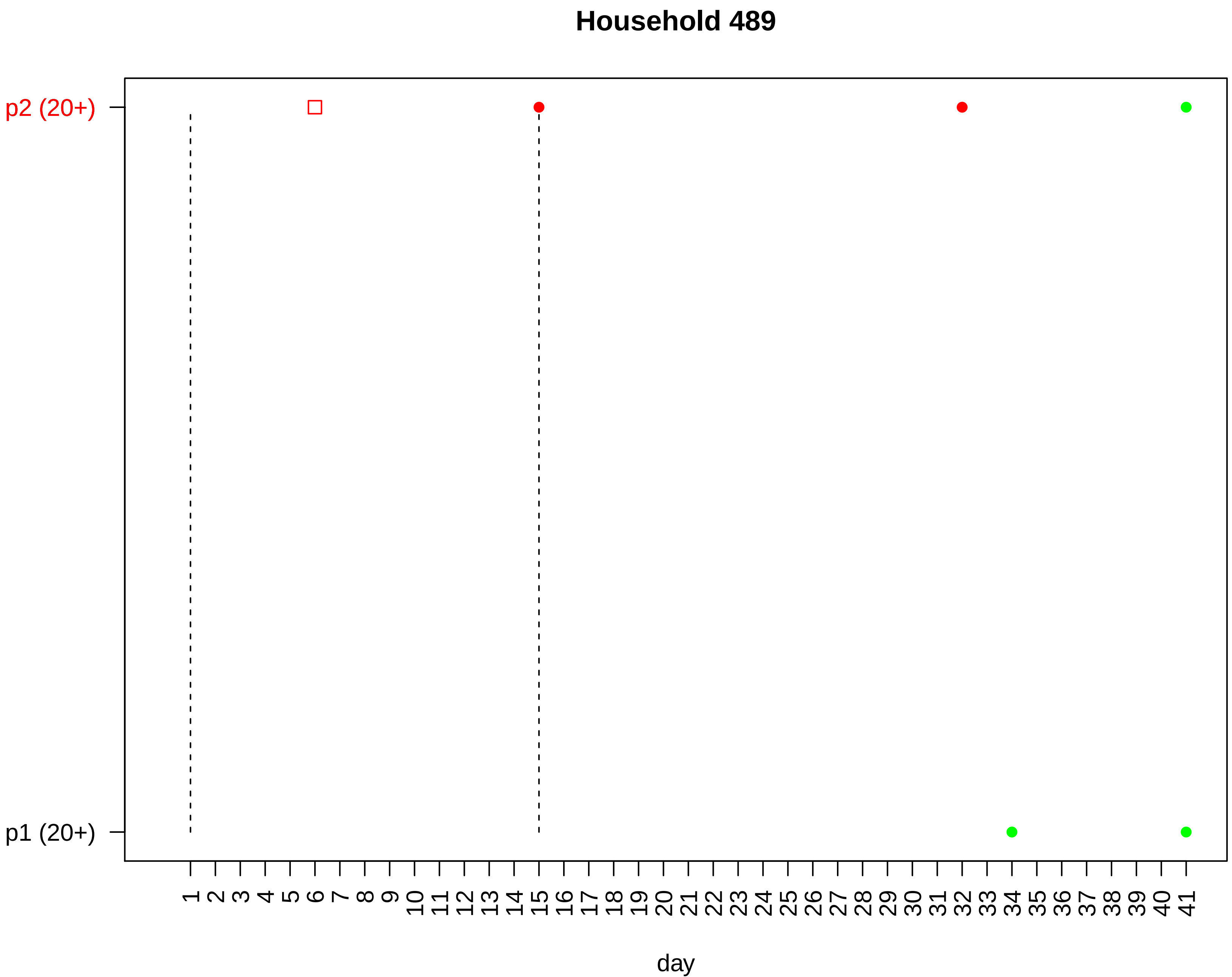
Household 489
p2 (20+)
p1 (20+)
day

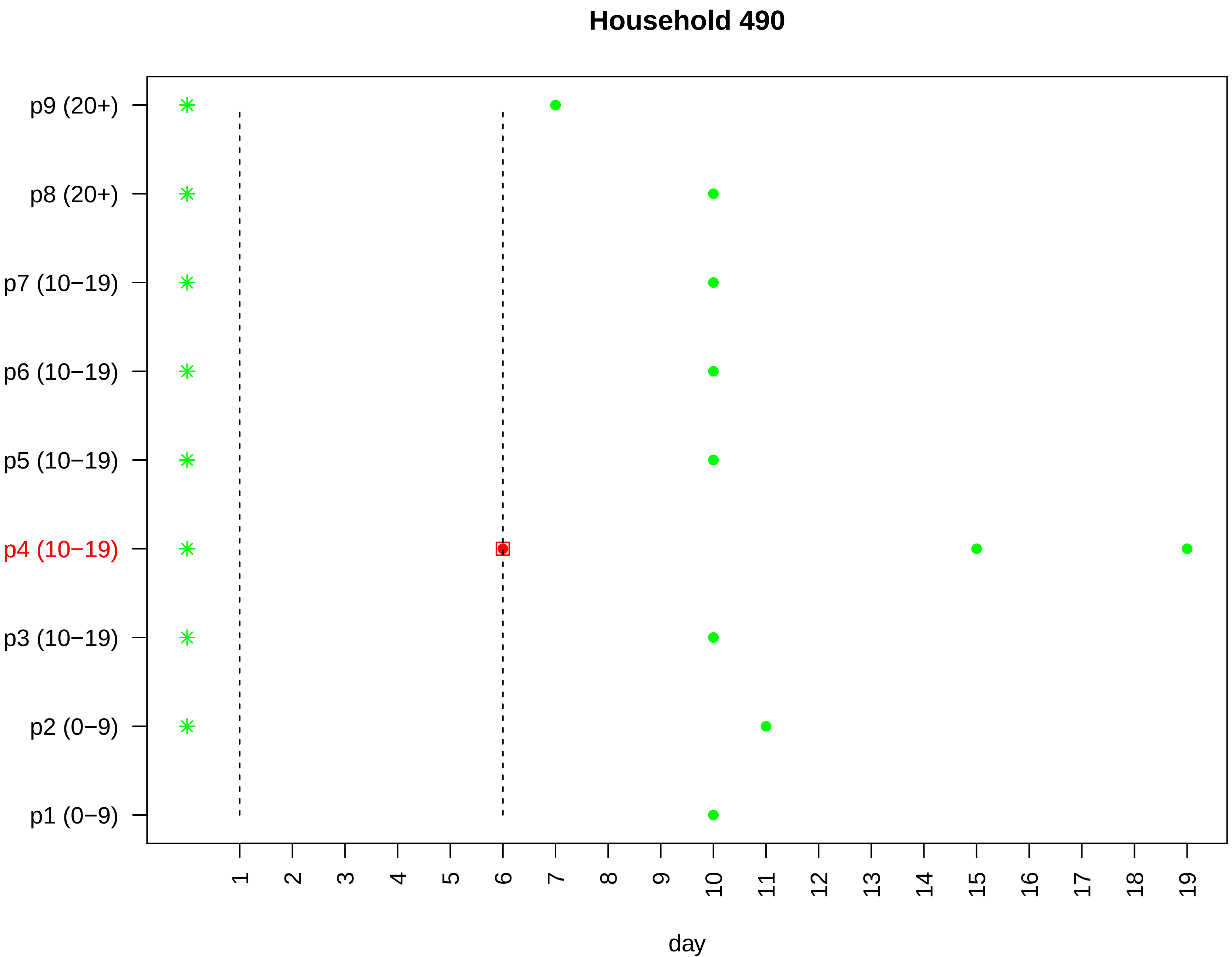

Household 490
p9 (20+)
p8 (20+)
p7 (10−19)
p6 (10−19)
p5 (10−19)
p4 (10−19)
p3 (10−19)
p2 (0−9)
p1 (0−9)
1 2 3 4 5 6 7 8 9 10 11 12 13 14 15 16 17 18 19
day

#### Household 492

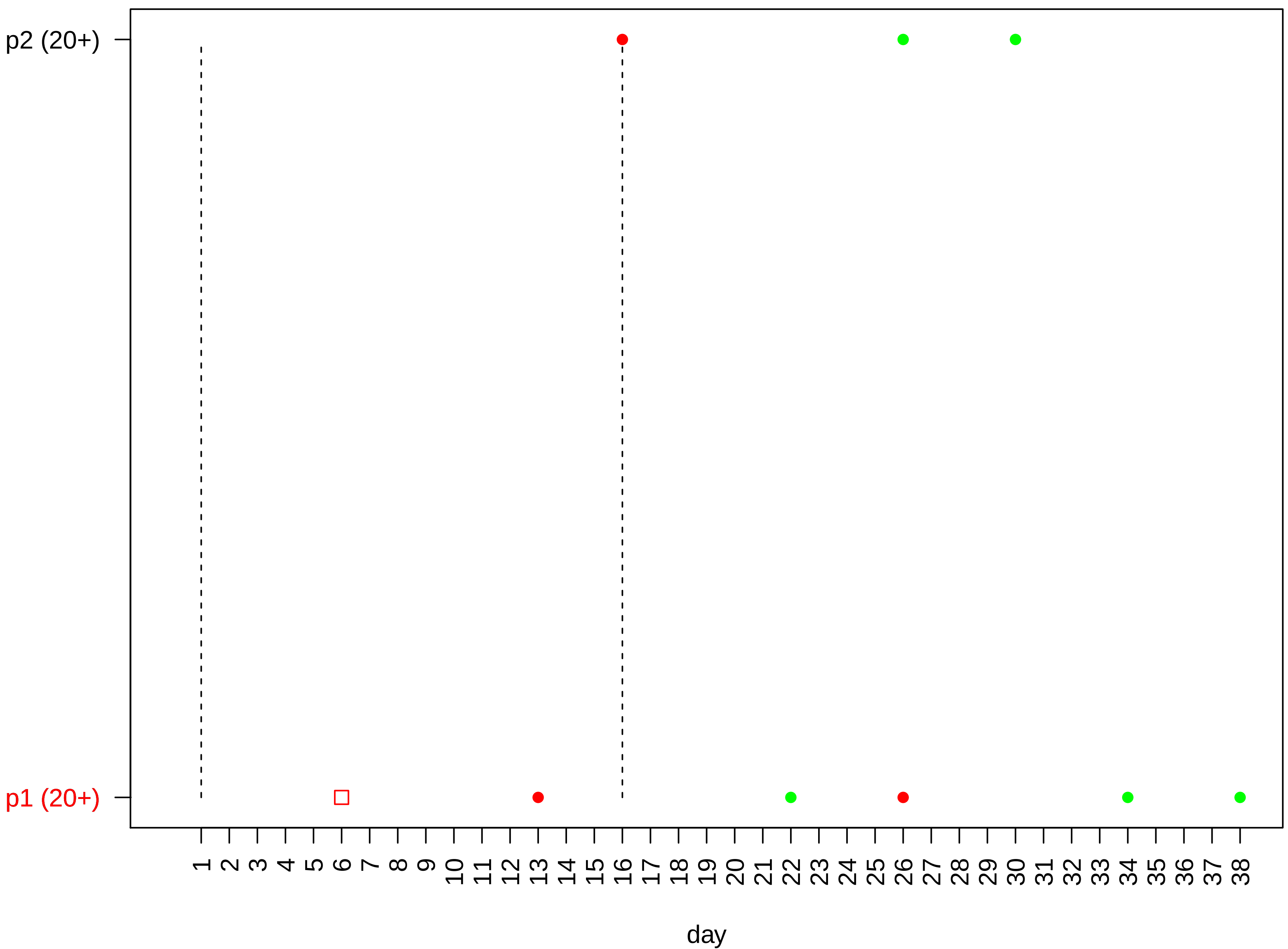

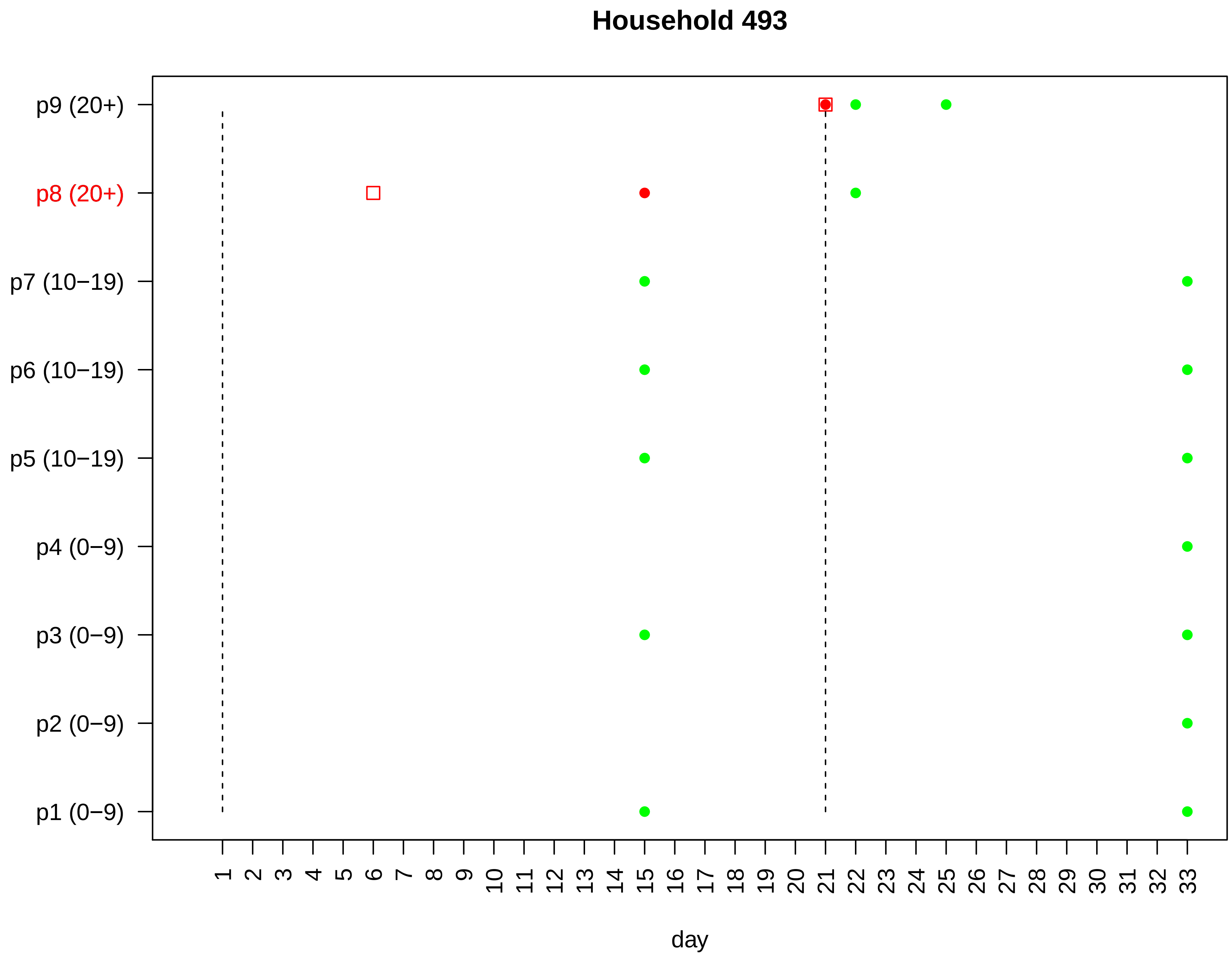
Household 493
p9 (20+)
p8 (20+)
p7 (10−19)
p6 (10−19)
p5 (10−19)
p4 (0−9)
p3 (0−9)
p2 (0−9)
p1 (0−9)
1 2 3 4 5 6 7 8 9 10 11 12 13 14 15 16 17 18 19 20 21 22 23 24 25 26 27 28 29 30 31 32 33
day

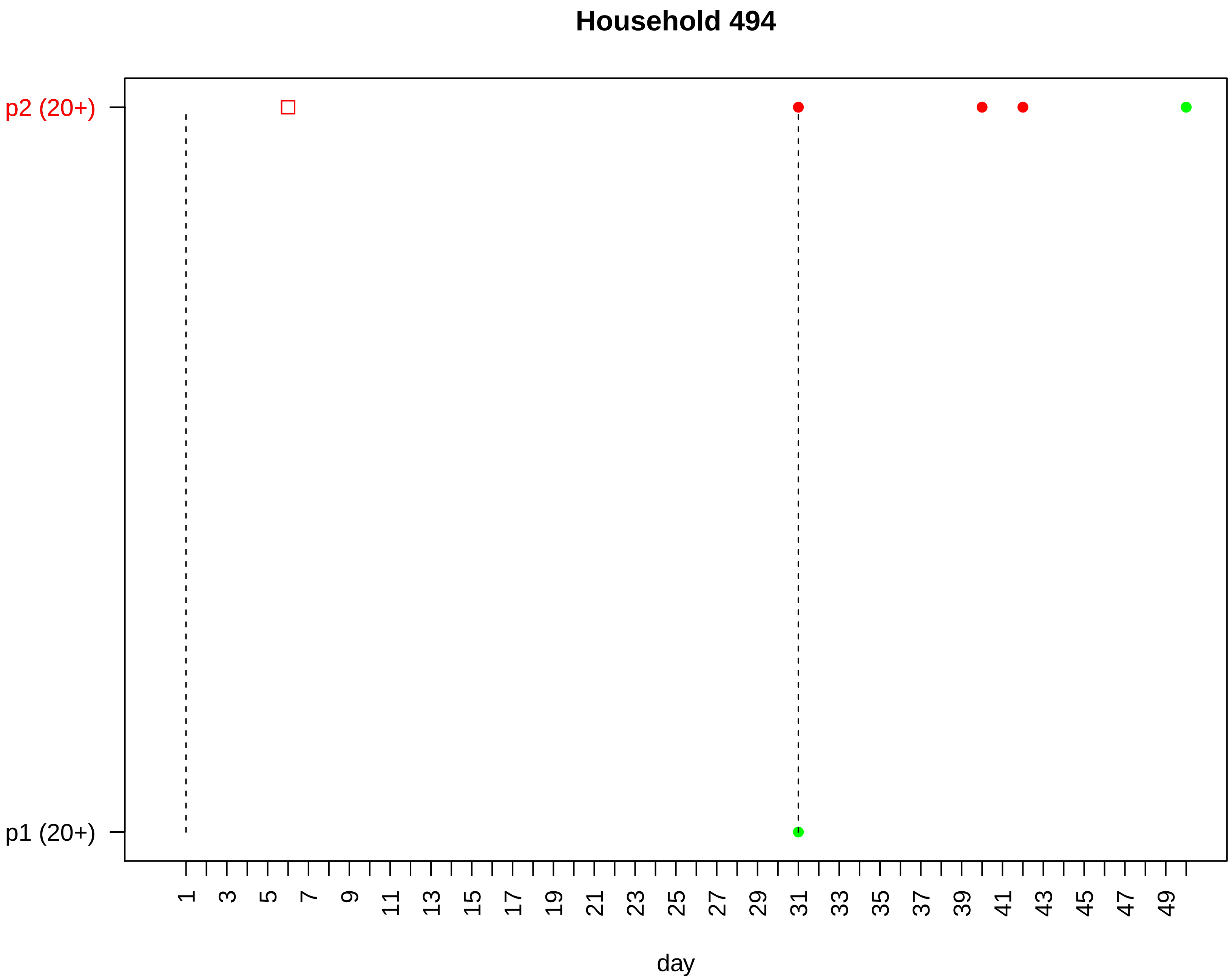

Household 494
p2 (20+)
p1 (20+)
1
3
5
7
9
11
13
15
17
19
21
23
25
27
29
31
33
35
37
39
41
43
45
47
49
day

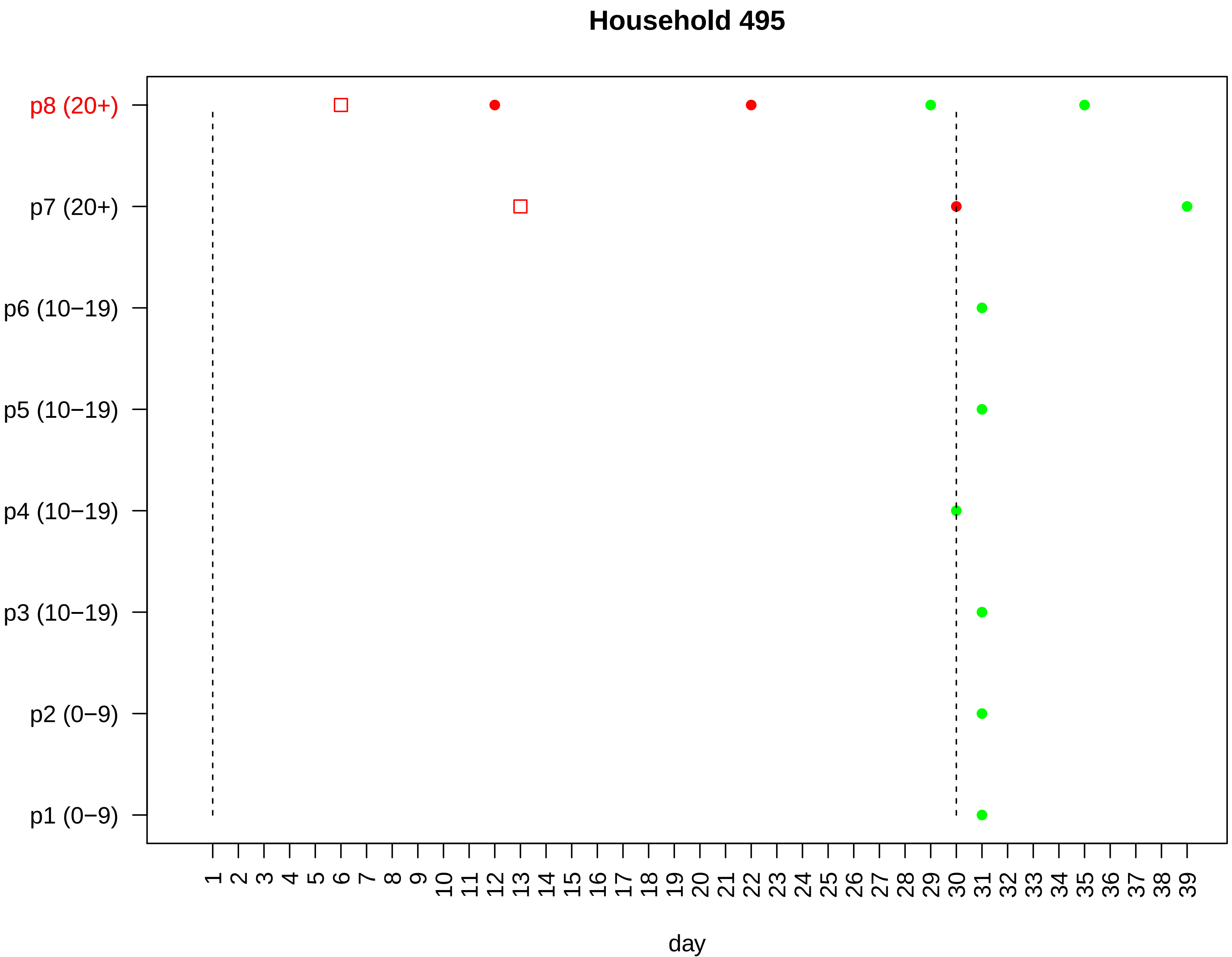

Household 495
p8 (20+)
p7 (20+)
p6 (10−19)
p5 (10−19)
p4 (10−19)
p3 (10−19)
p2 (0−9)
p1 (0−9)
1 2 3 4 5 6 7 8 9 10 11 12 13 14 15 16 17 18 19 20 21 22 23 24 25 26 27 28 29 30 31 32 33 34 35 36 37 38 39
day

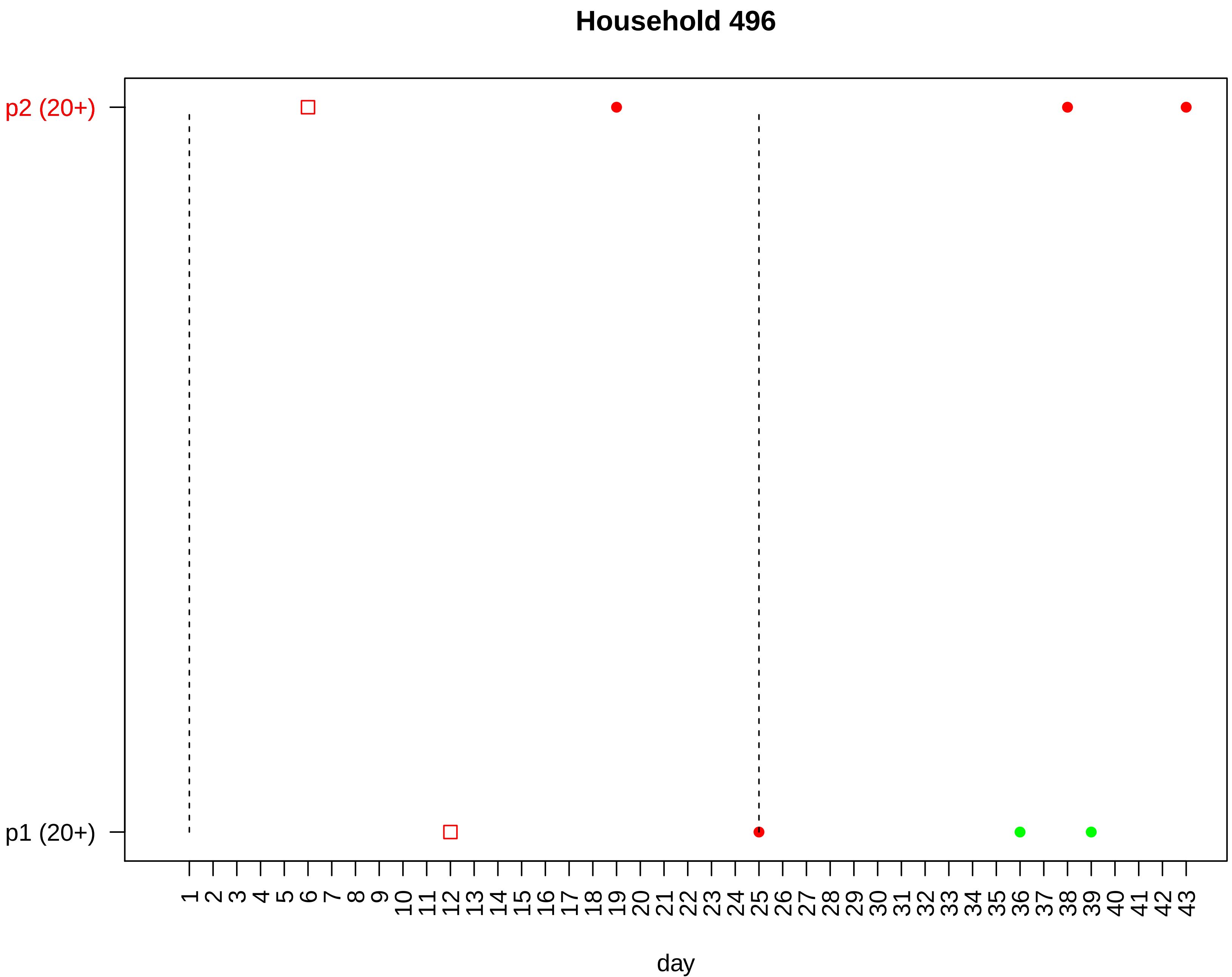

Household 496
p2 (20+)
p1 (20+)
1 2 3 4 5 6 7 8 9 10 11 12 13 14 15 16 17 18 19 20 21 22 23 24 25 26 27 28 29 30 31 32 33 34 35 36 37 38 39 40 41 42 43
day

#### Household 497

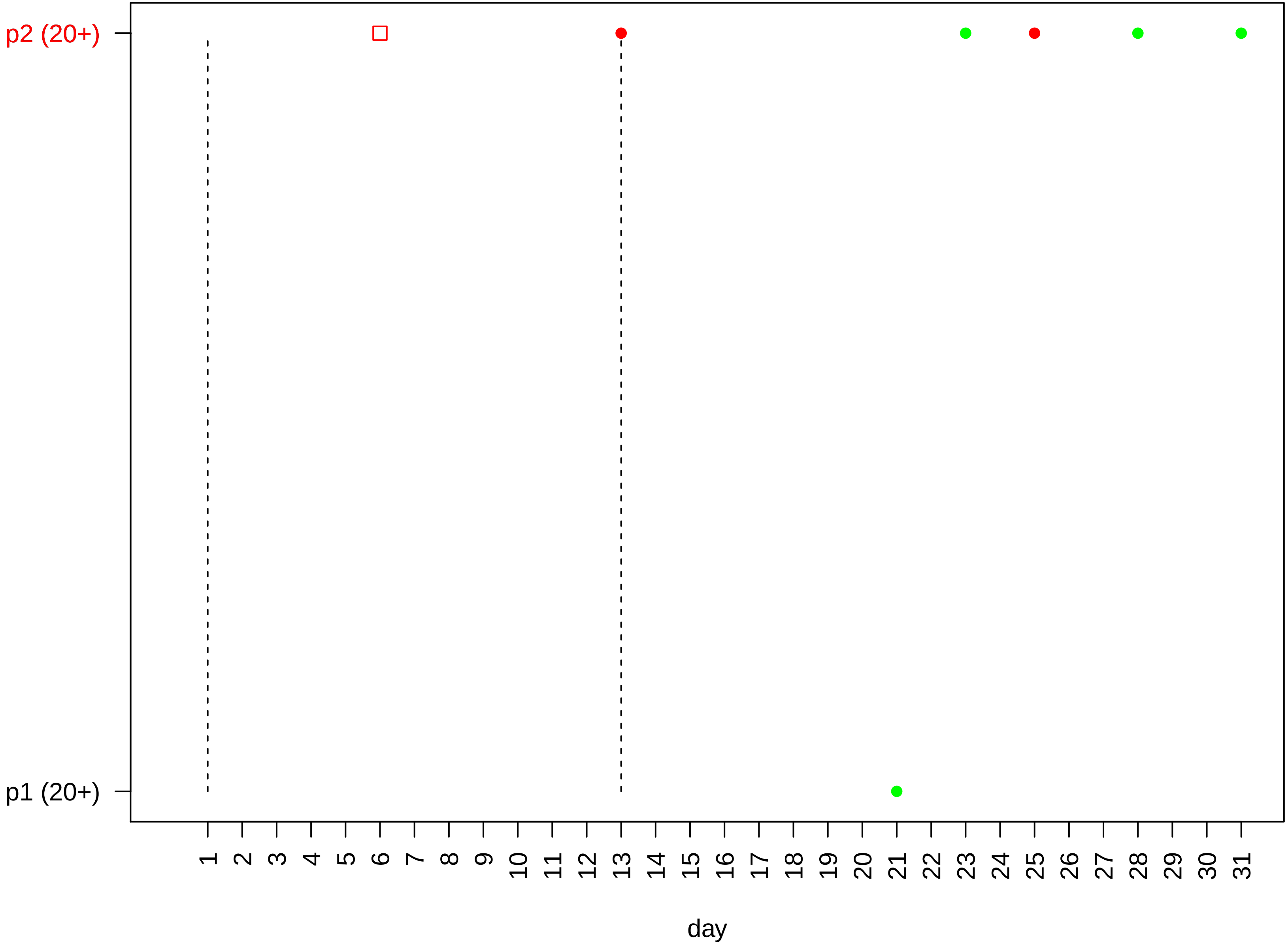

#### Household 498

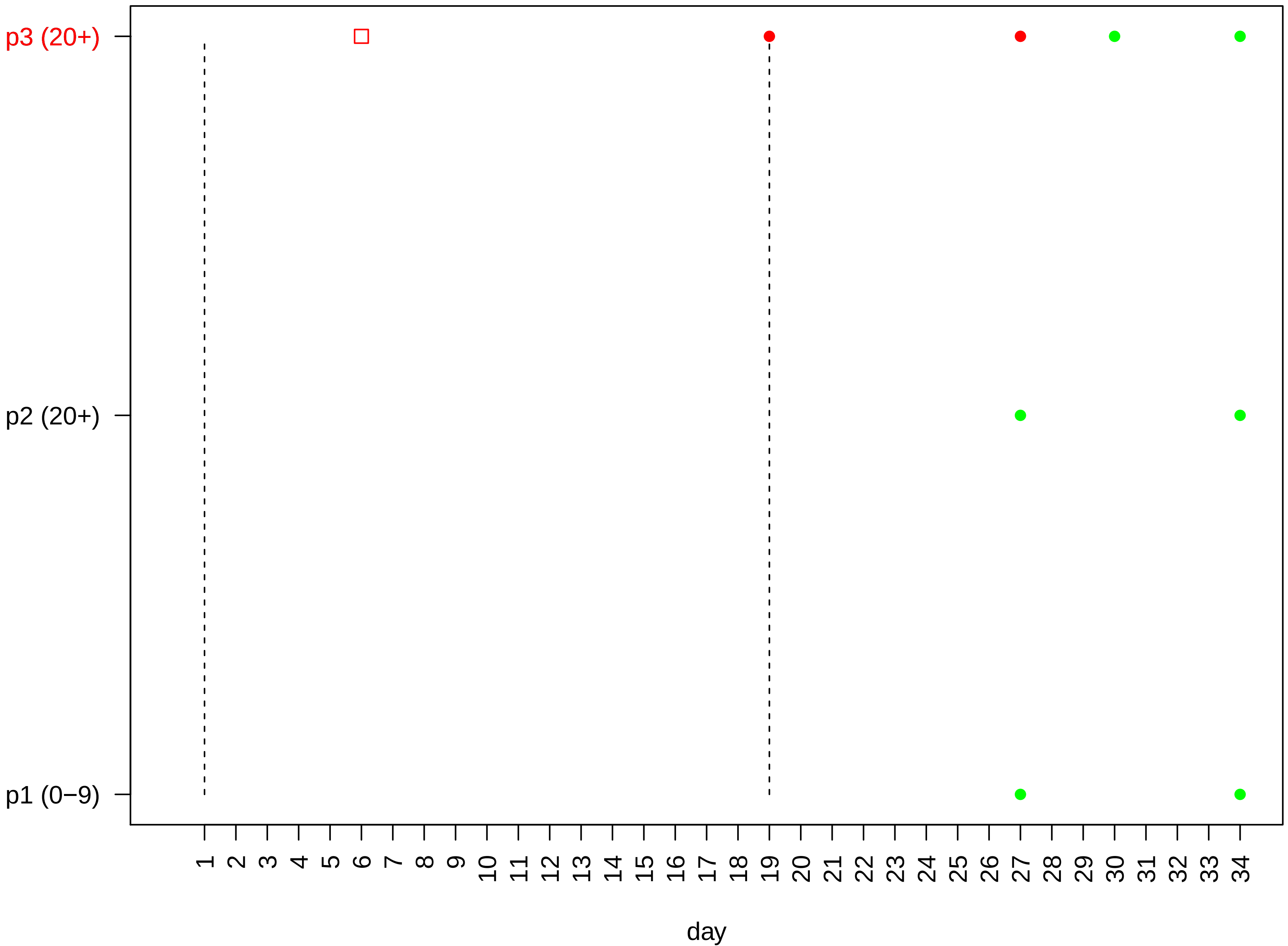

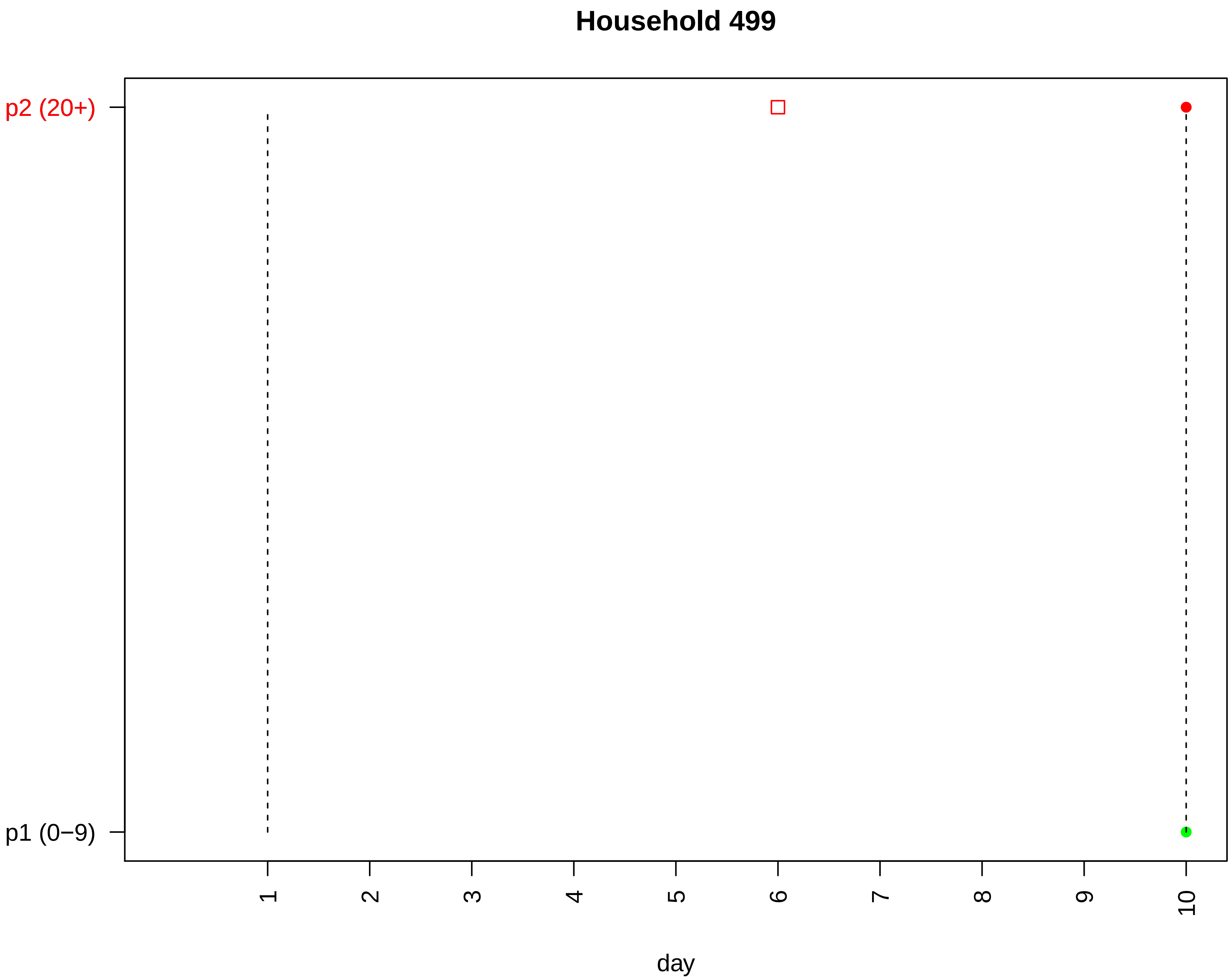

Household 499
p2 (20+)
p1 (0−9)
1
2
3
4
5
6
7
8
9
10
day

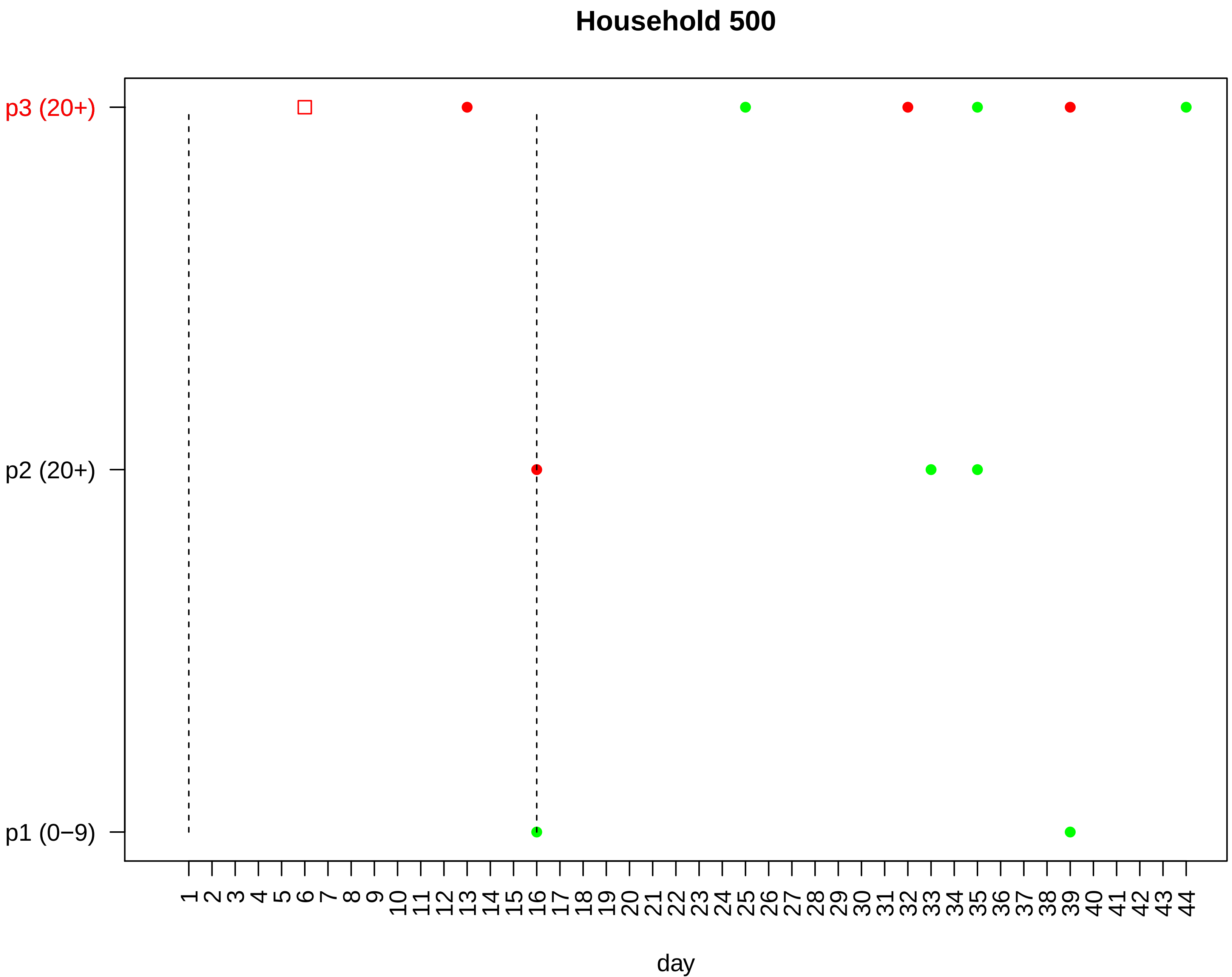
Household 500
p3 (20+)
p2 (20+)
p1 (0−9)
1 2 3 4 5 6 7 8 9 10 11 12 13 14 15 16 17 18 19 20 21 22 23 24 25 26 27 28 29 30 31 32 33 34 35 36 37 38 39 40 41 42 43 44
day

#### Household 501

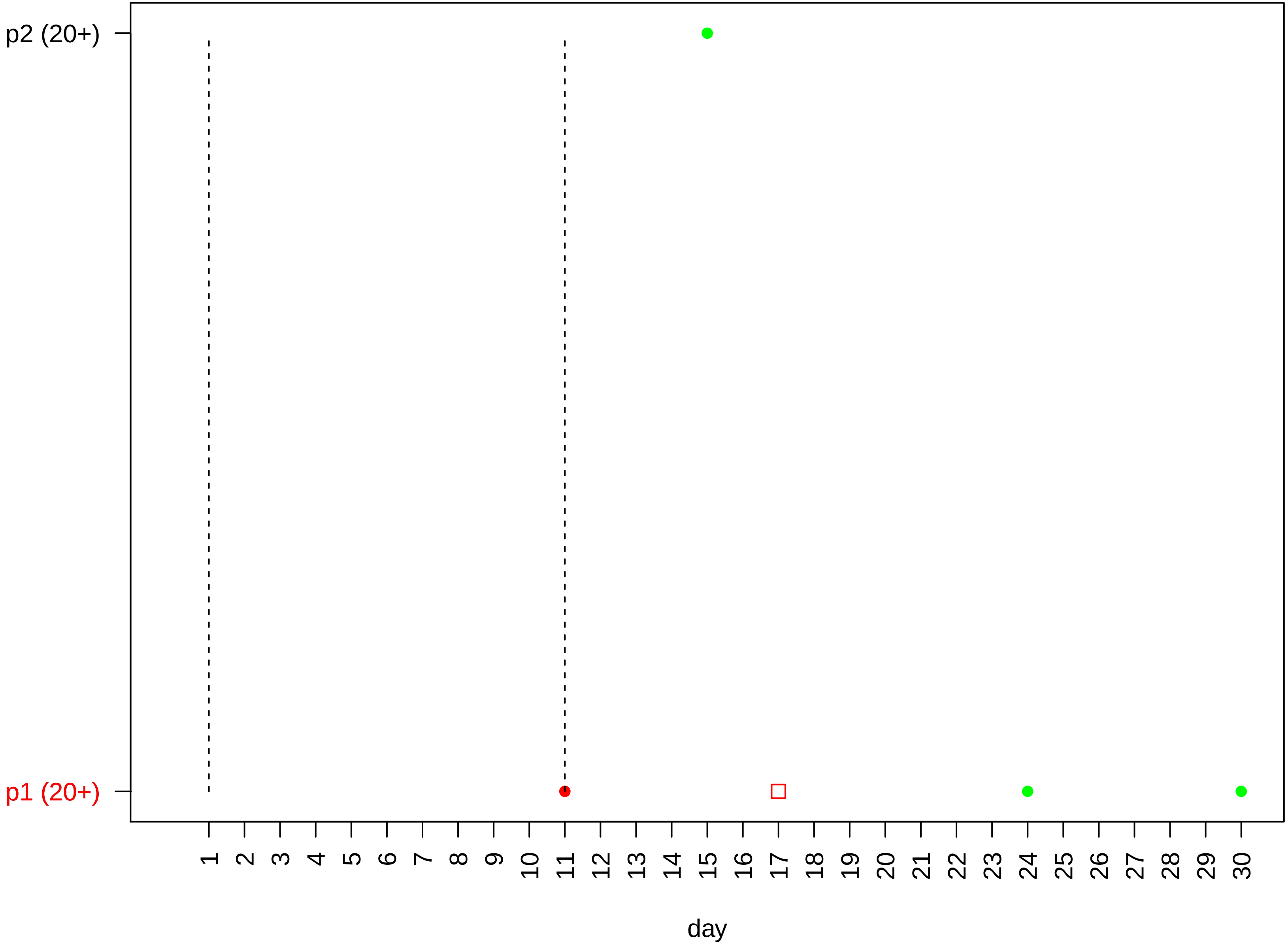

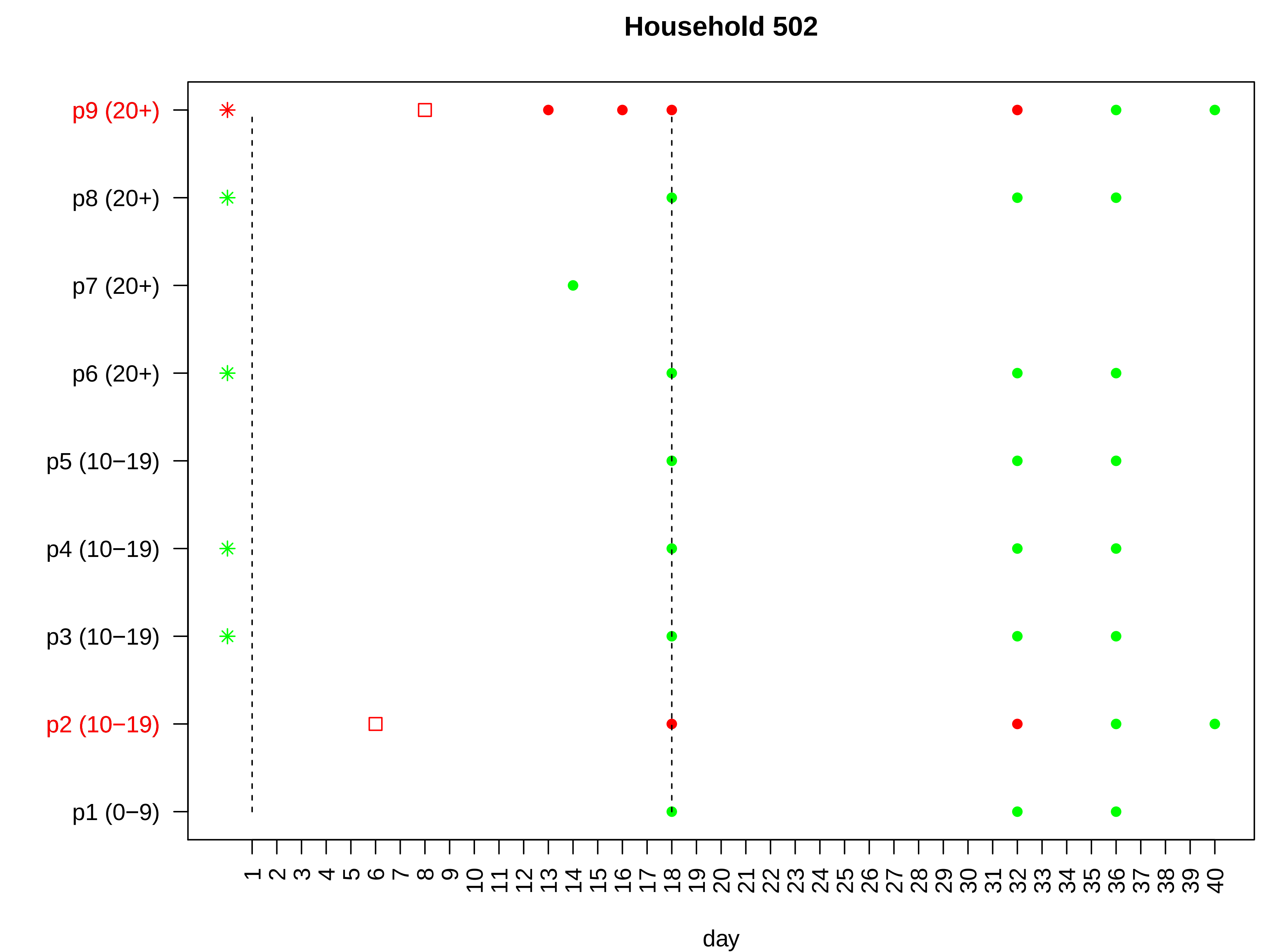

Household 502
p9 (20+)
p8 (20+)
p7 (20+)
p6 (20+)
p5 (10−19)
p4 (10−19)
p3 (10−19)
p2 (10−19)
p1 (0−9)
1 2 3 4 5 6 7 8 9 10 11 12 13 14 15 16 17 18 19 20 21 22 23 24 25 26 27 28 29 30 31 32 33 34 35 36 37 38 39 40
day

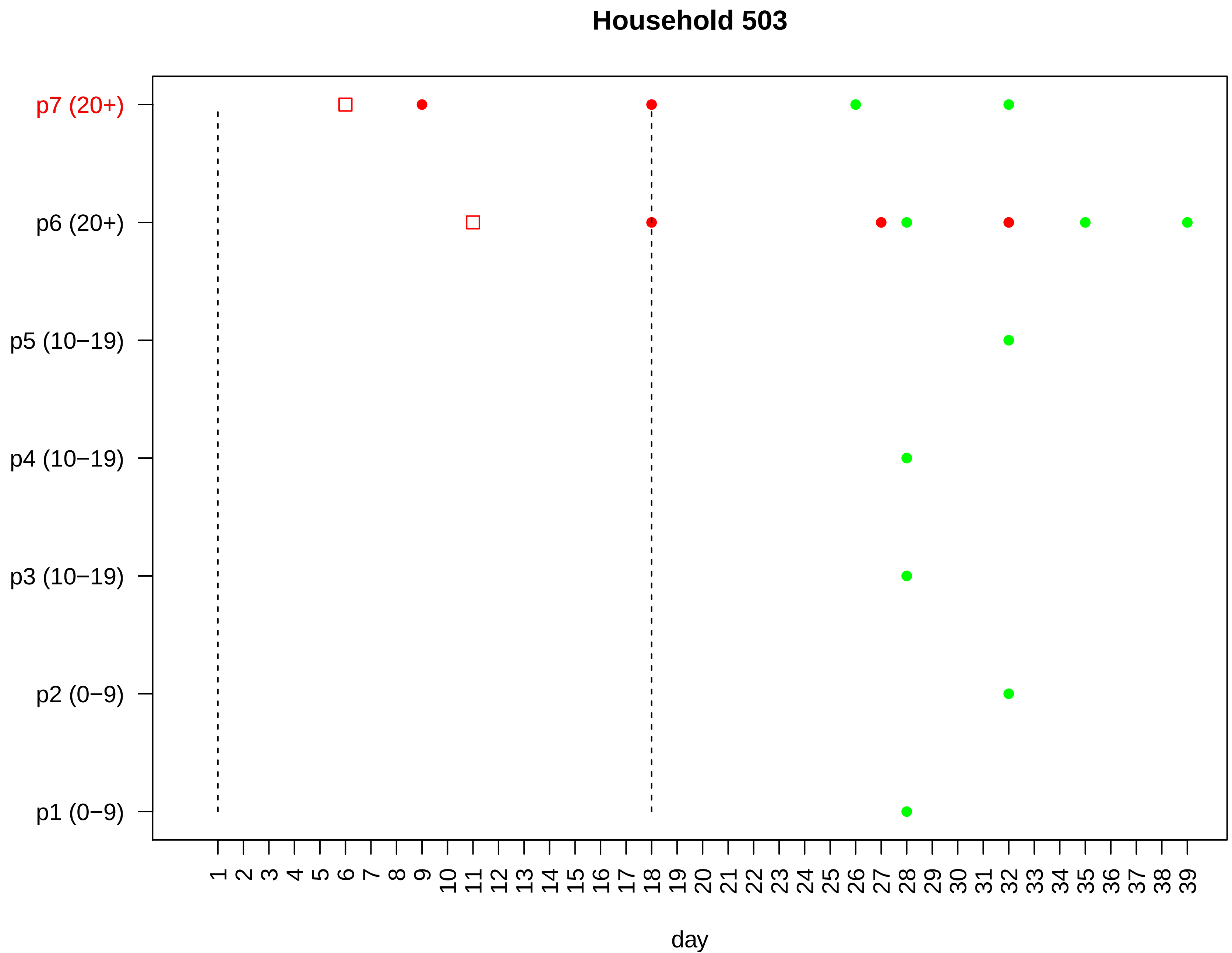

Household 503
p7 (20+)
p6 (20+)
p5 (10−19)
p4 (10−19)
p3 (10−19)
p2 (0−9)
p1 (0−9)
1 2 3 4 5 6 7 8 9 10 11 12 13 14 15 16 17 18 19 20 21 22 23 24 25 26 27 28 29 30 31 32 33 34 35 36 37 38 39
day

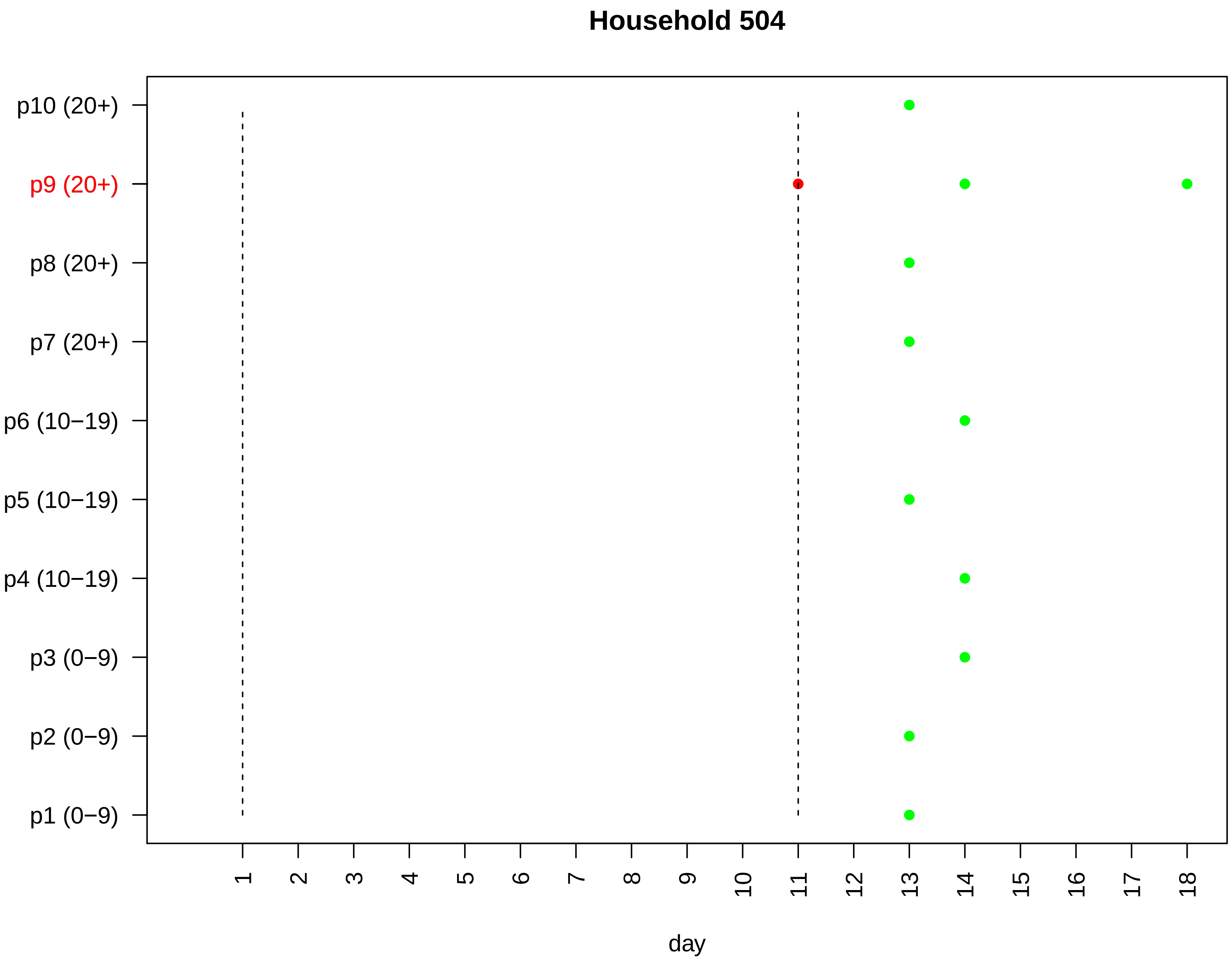

Household 504
p10 (20+)
p9 (20+)
p8 (20+)
p7 (20+)
p6 (10−19)
p5 (10−19)
p4 (10−19)
p3 (0−9)
p2 (0−9)
p1 (0−9)
day

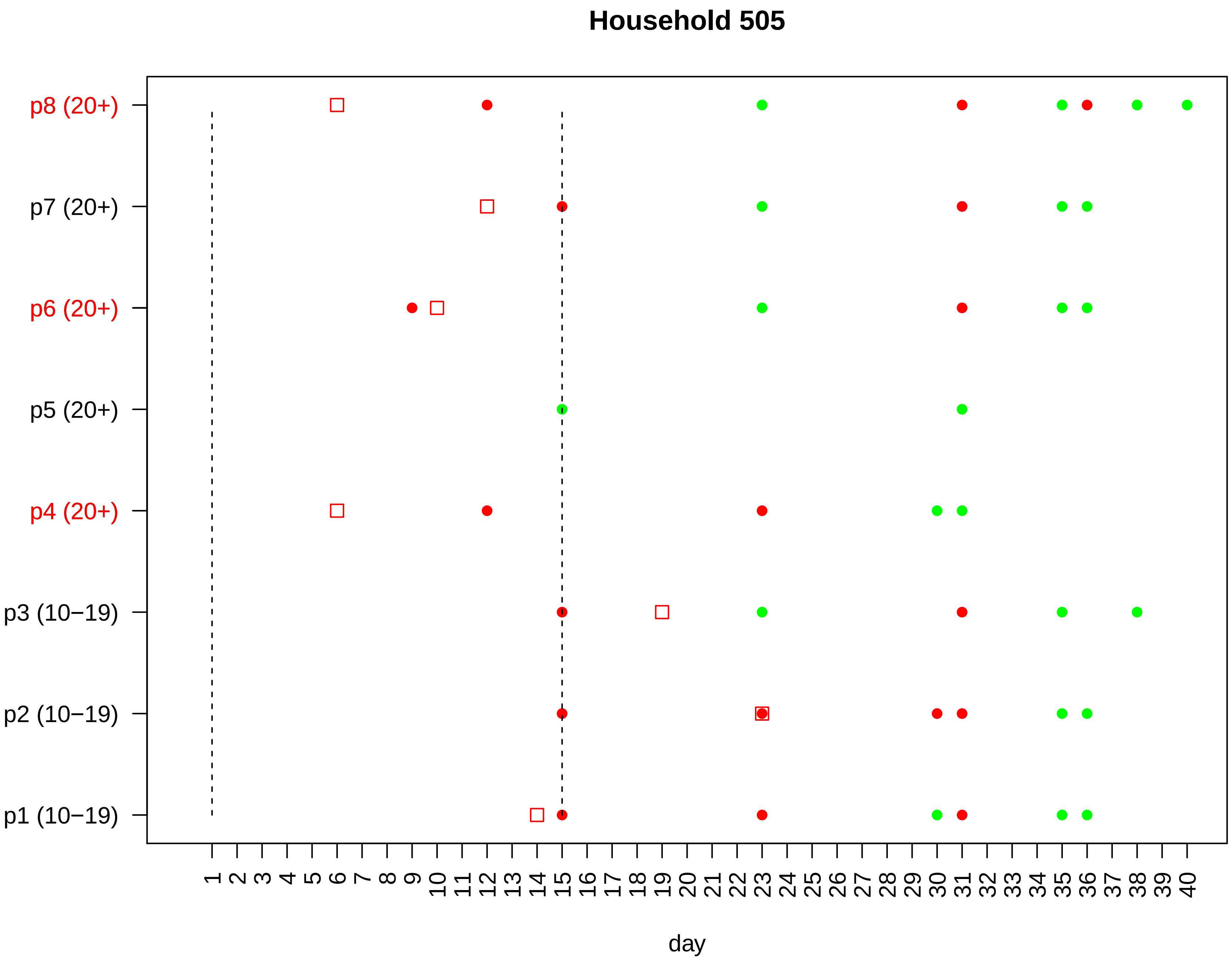

Household 505
p8 (20+)
p7 (20+)
p6 (20+)
p5 (20+)
p4 (20+)
p3 (10−19)
p2 (10−19)
p1 (10−19)
1 2 3 4 5 6 7 8 9 10 11 12 13 14 15 16 17 18 19 20 21 22 23 24 25 26 27 28 29 30 31 32 33 34 35 36 37 38 39 40
day

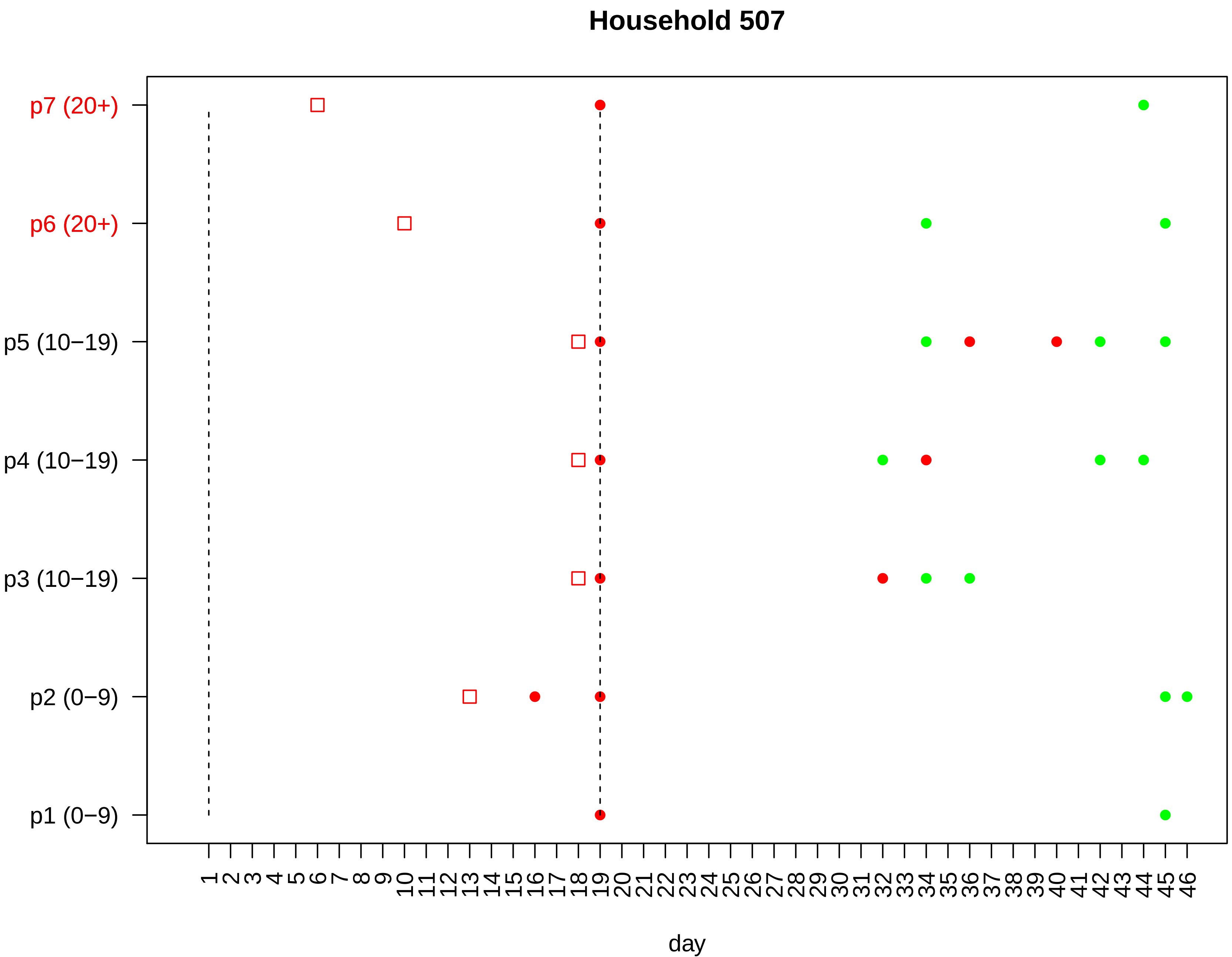

Household 507
p7 (20+)
p6 (20+)
p5 (10−19)
p4 (10−19)
p3 (10−19)
p2 (0−9)
p1 (0−9)
1 2 3 4 5 6 7 8 9 10 11 12 13 14 15 16 17 18 19 20 21 22 23 24 25 26 27 28 29 30 31 32 33 34 35 36 37 38 39 40 41 42 43 44 45 46
day

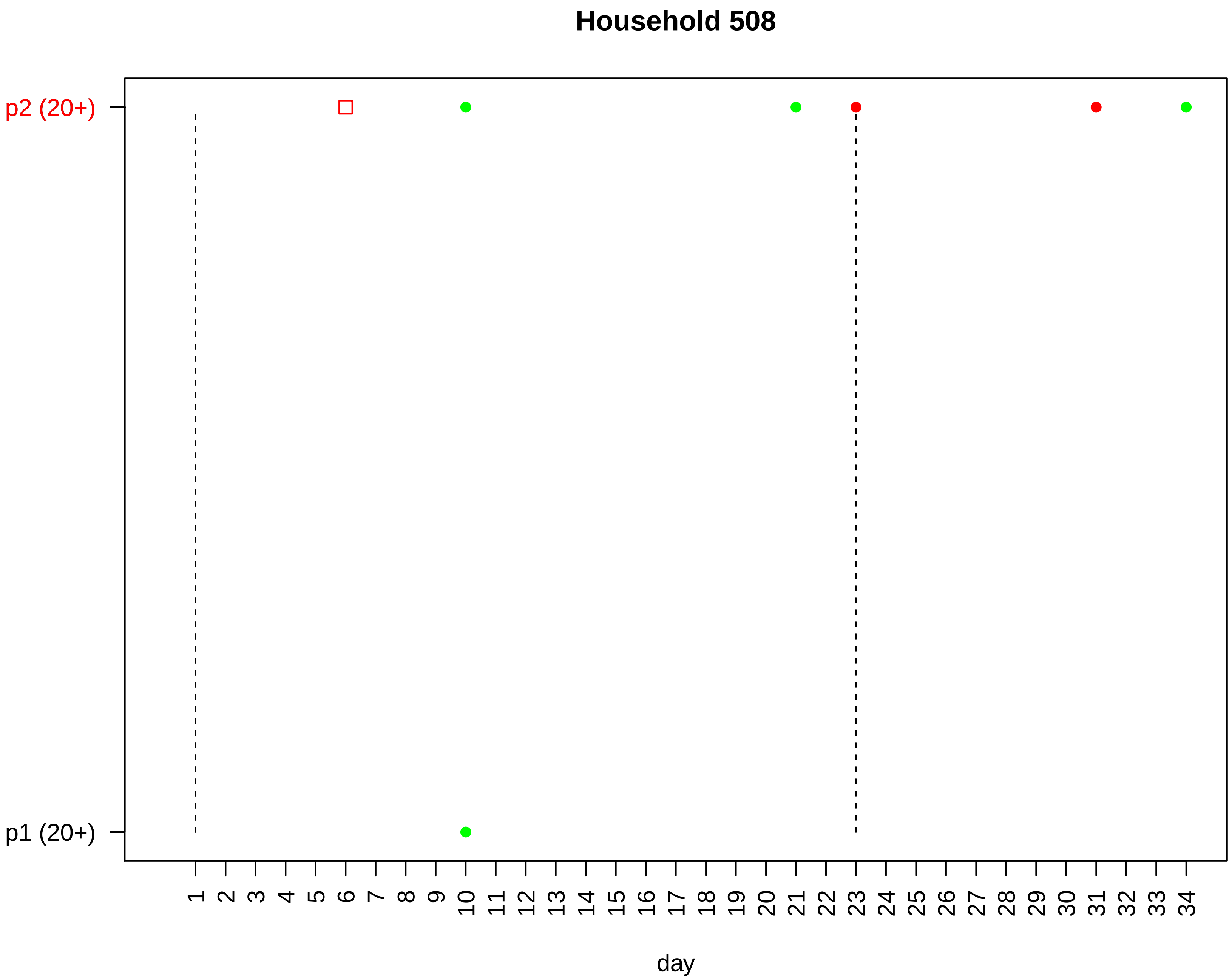
Household 508
p2 (20+)
p1 (20+)
1 2 3 4 5 6 7 8 9 10 11 12 13 14 15 16 17 18 19 20 21 22 23 24 25 26 27 28 29 30 31 32 33 34
day

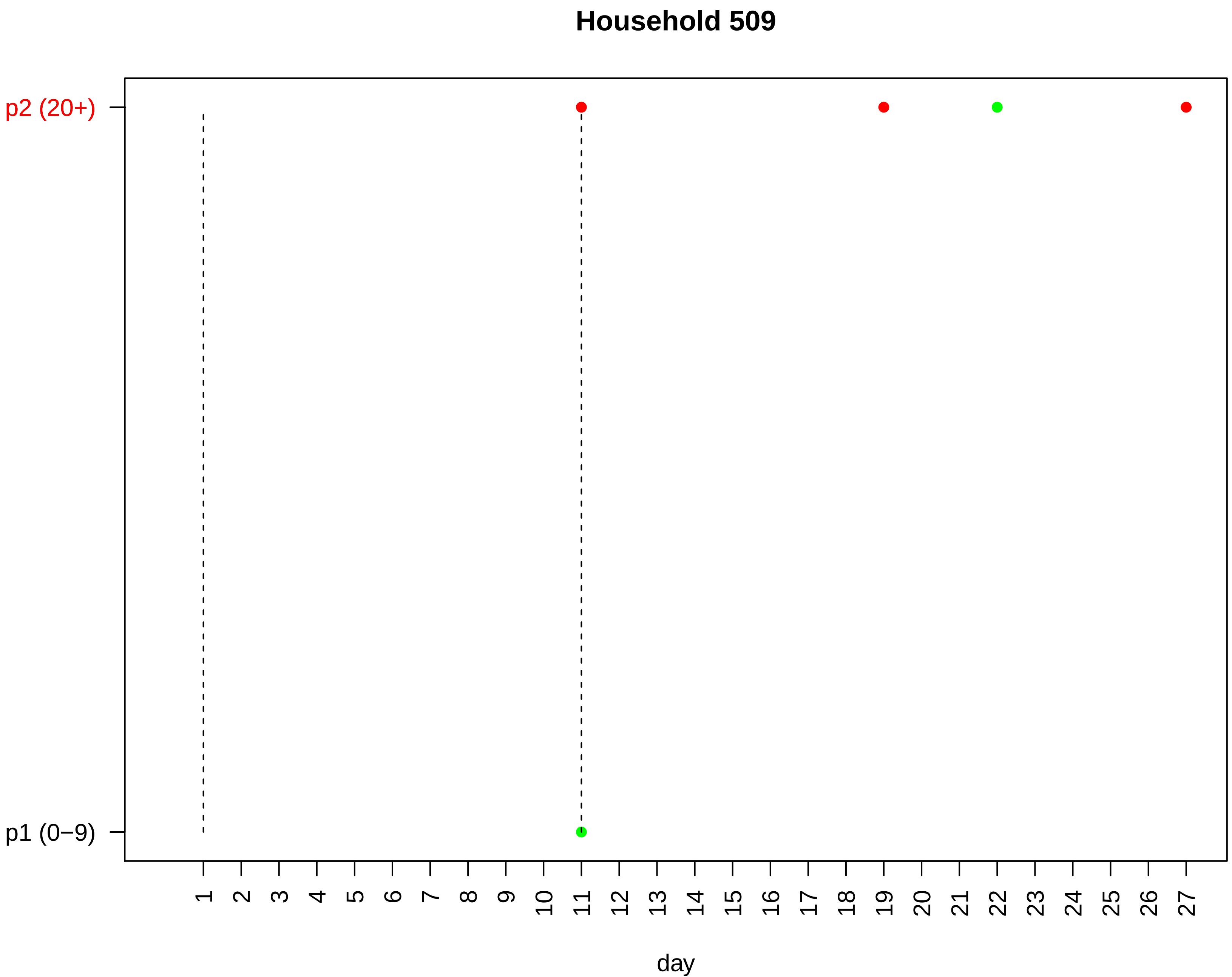
Household 509
p2 (20+)
p1 (0−9)
day

#### Household 510

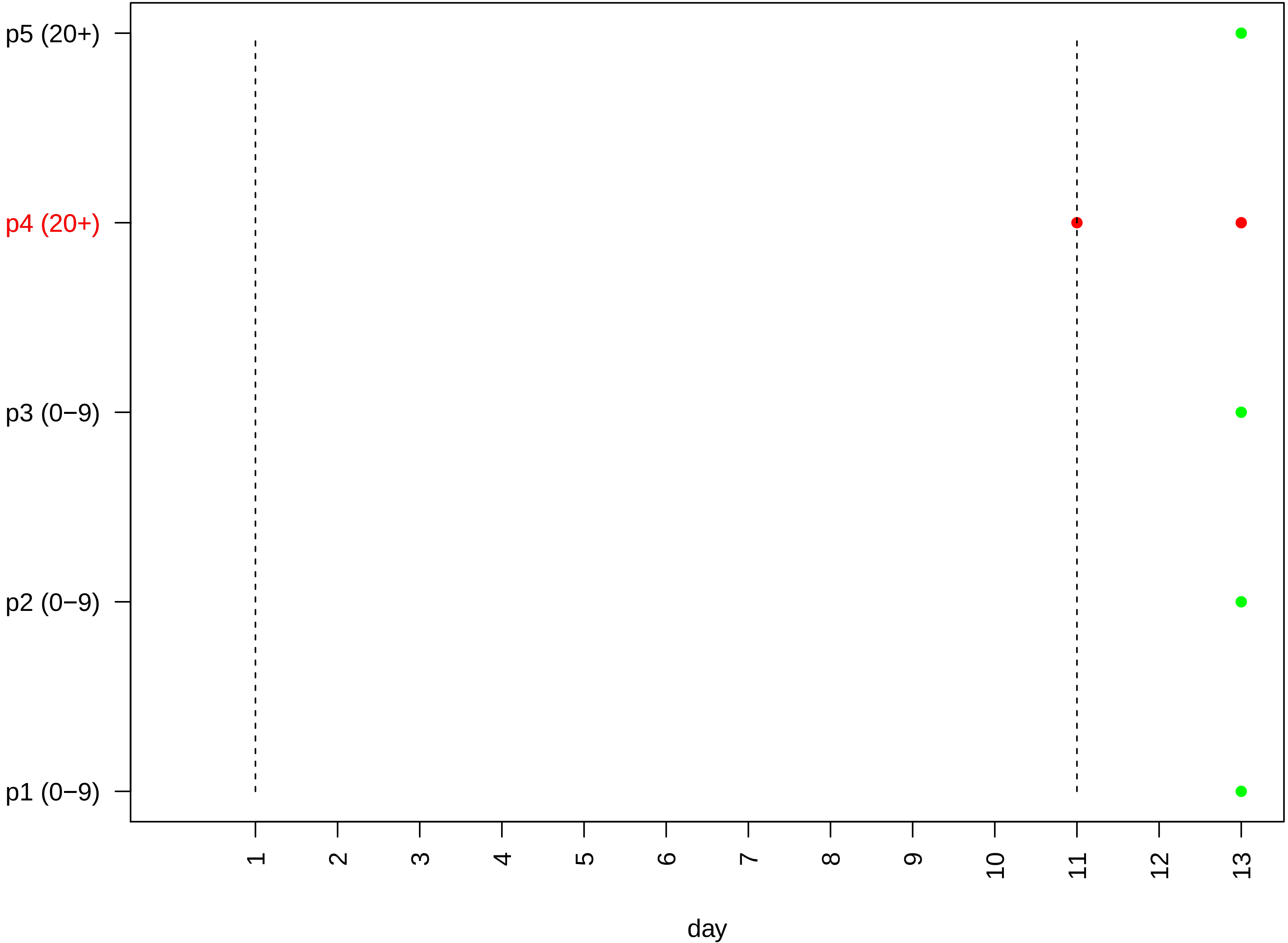

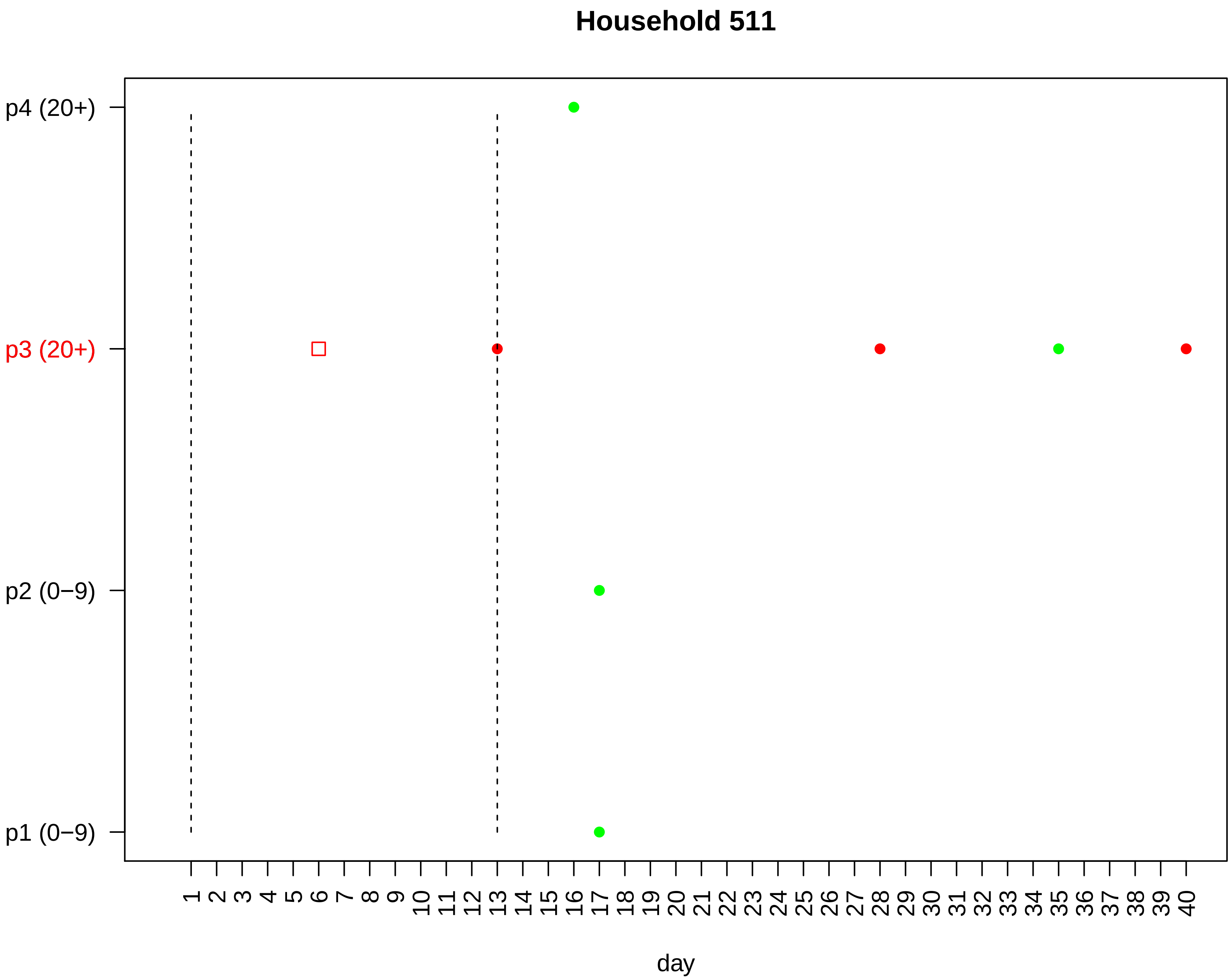

Household 511
p4 (20+)
p3 (20+)
p2 (0−9)
p1 (0−9)
1 2 3 4 5 6 7 8 9 10 11 12 13 14 15 16 17 18 19 20 21 22 23 24 25 26 27 28 29 30 31 32 33 34 35 36 37 38 39 40
day

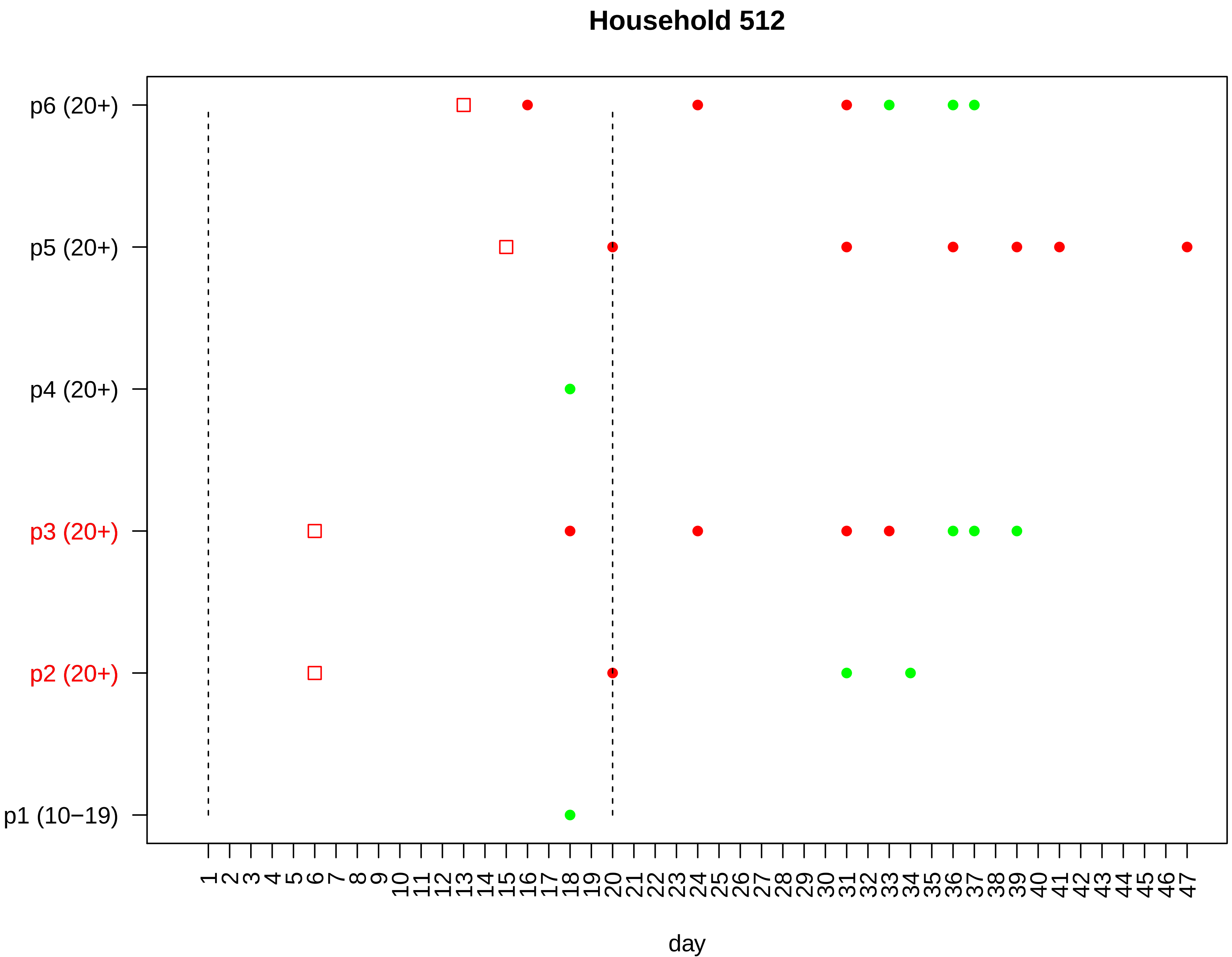

Household 512
p6 (20+)
p5 (20+)
p4 (20+)
p3 (20+)
p2 (20+)
p1 (10−19)
1 2 3 4 5 6 7 8 9 10 11 12 13 14 15 16 17 18 19 20 21 22 23 24 25 26 27 28 29 30 31 32 33 34 35 36 37 38 39 40 41 42 43 44 45 46 47
day

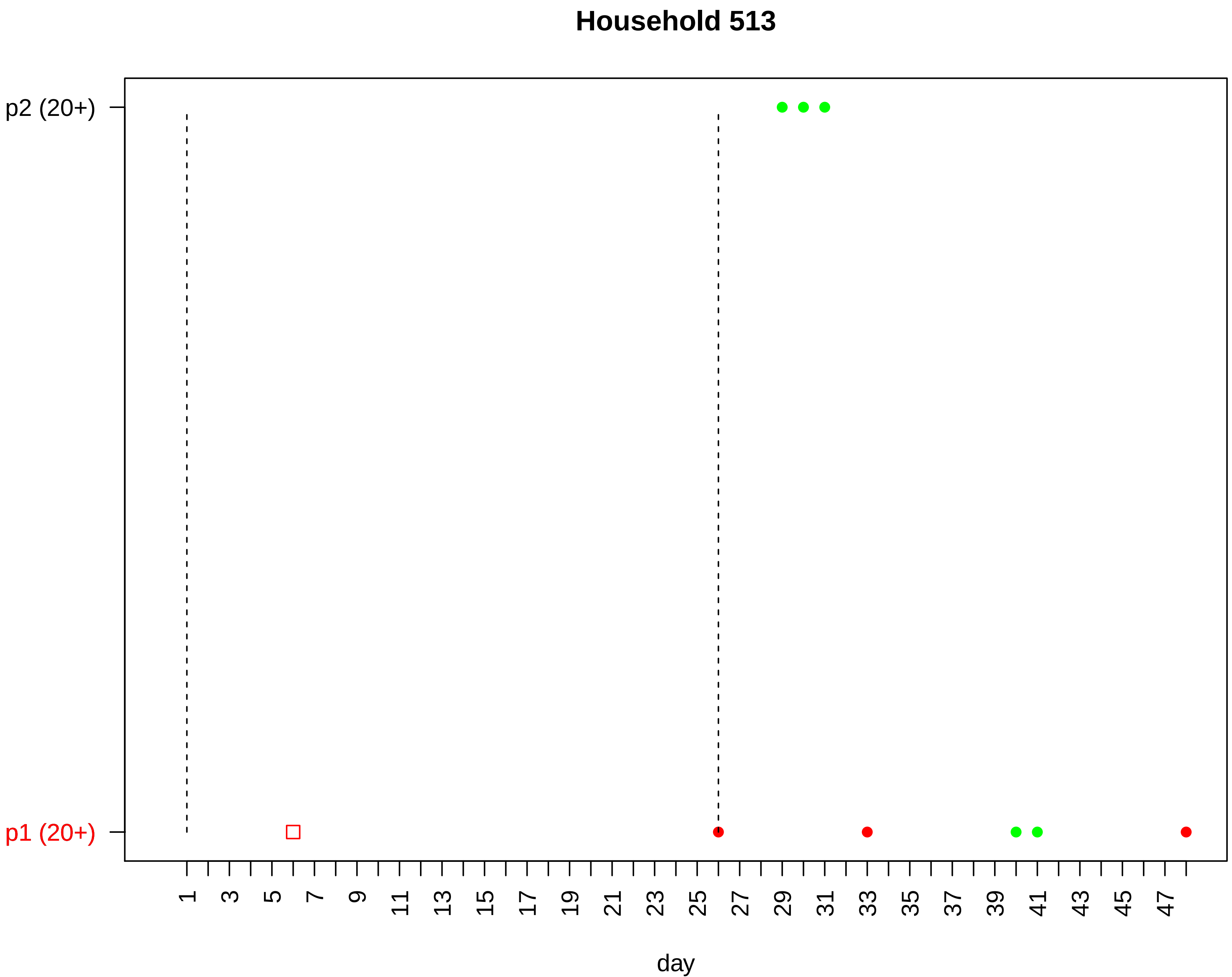

Household 513
p2 (20+)
p1 (20+)
1
3
5
7
9
11
13
15
17
19
21
23
25
27
29
31
33
35
37
39
41
43
45
47
day

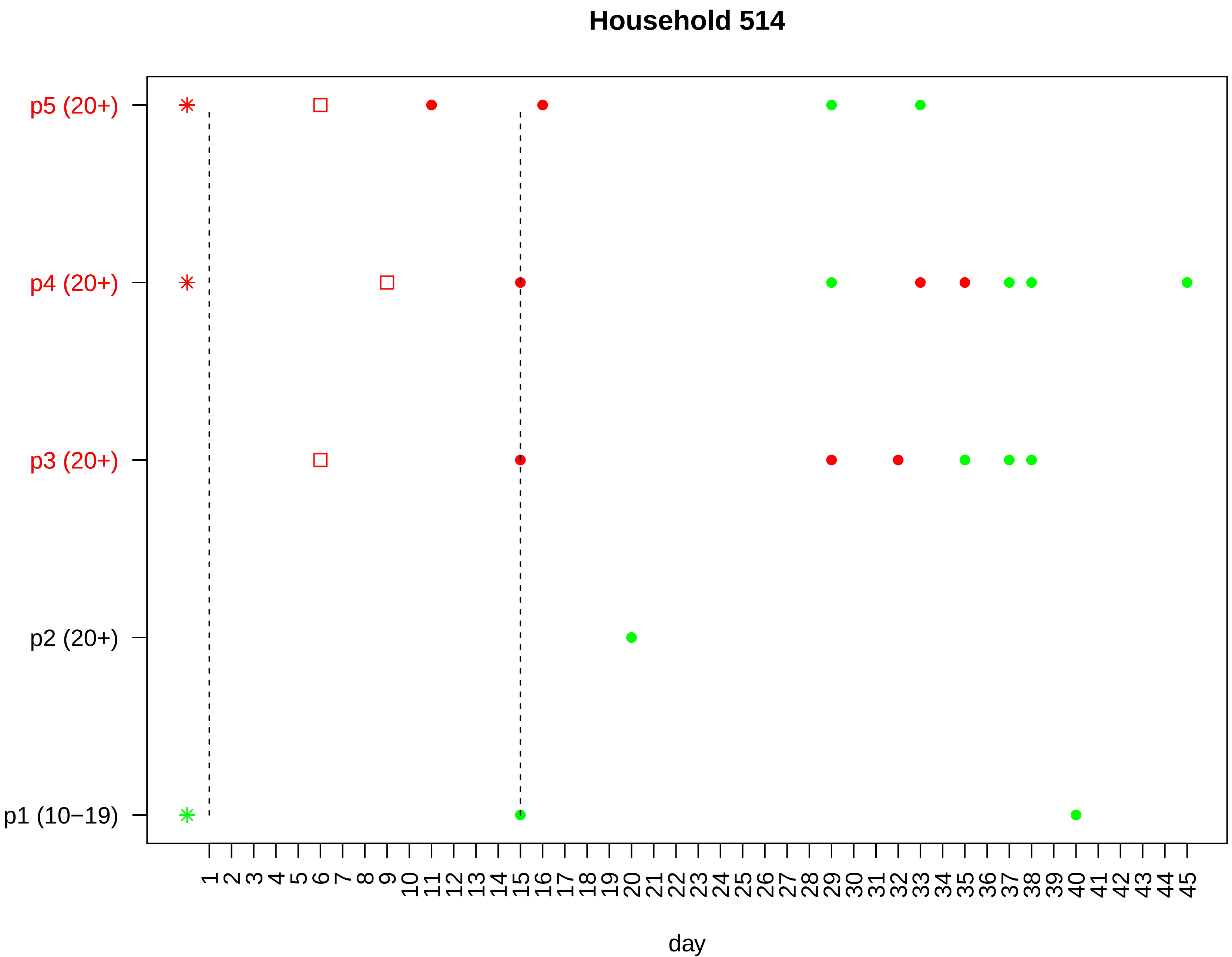
Household 514
p5 (20+)
p4 (20+)
p3 (20+)
p2 (20+)
p1 (10−19)
1 2 3 4 5 6 7 8 9 10 11 12 13 14 15 16 17 18 19 20 21 22 23 24 25 26 27 28 29 30 31 32 33 34 35 36 37 38 39 40 41 42 43 44 45
day

#### Household 515

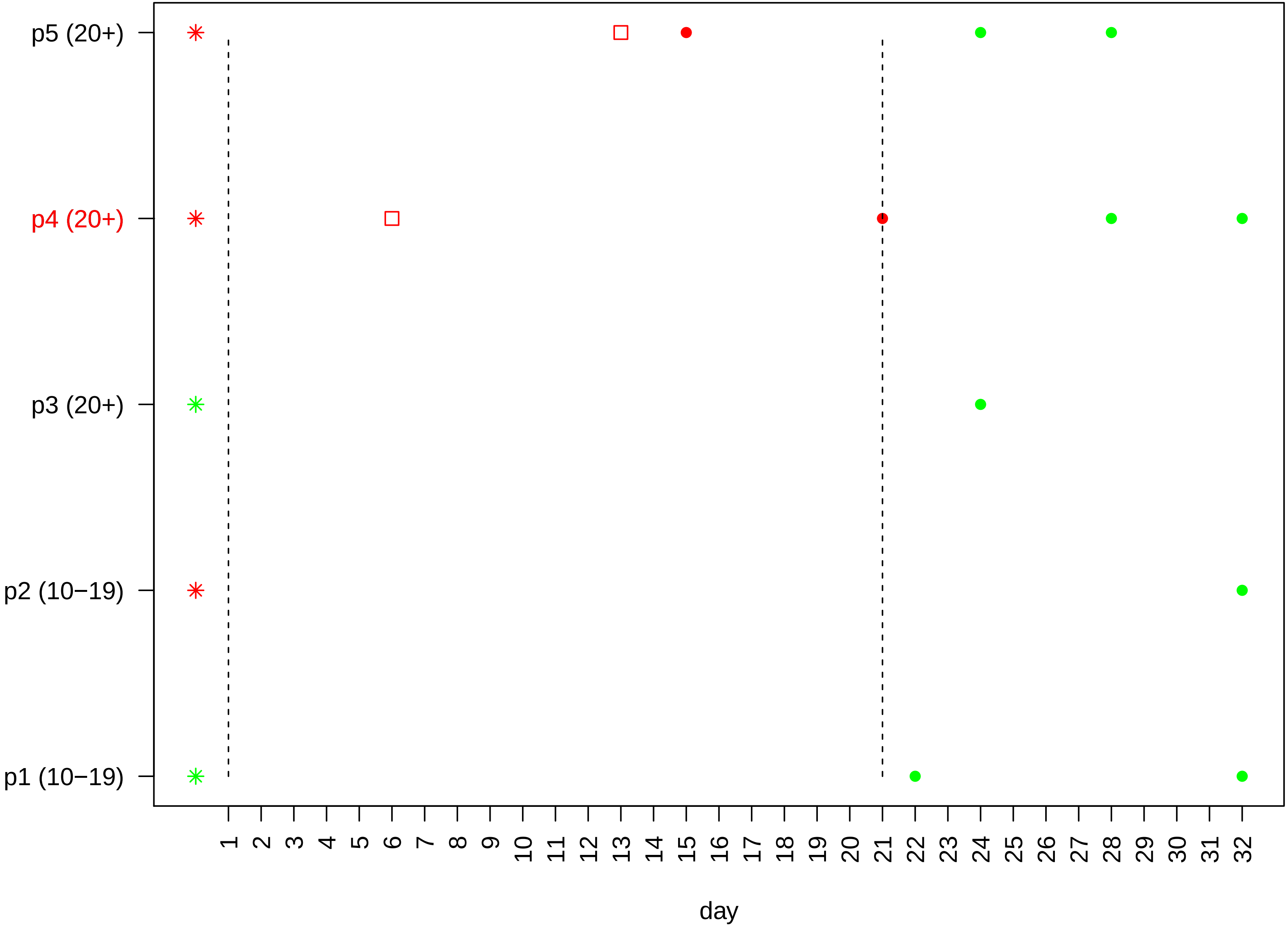

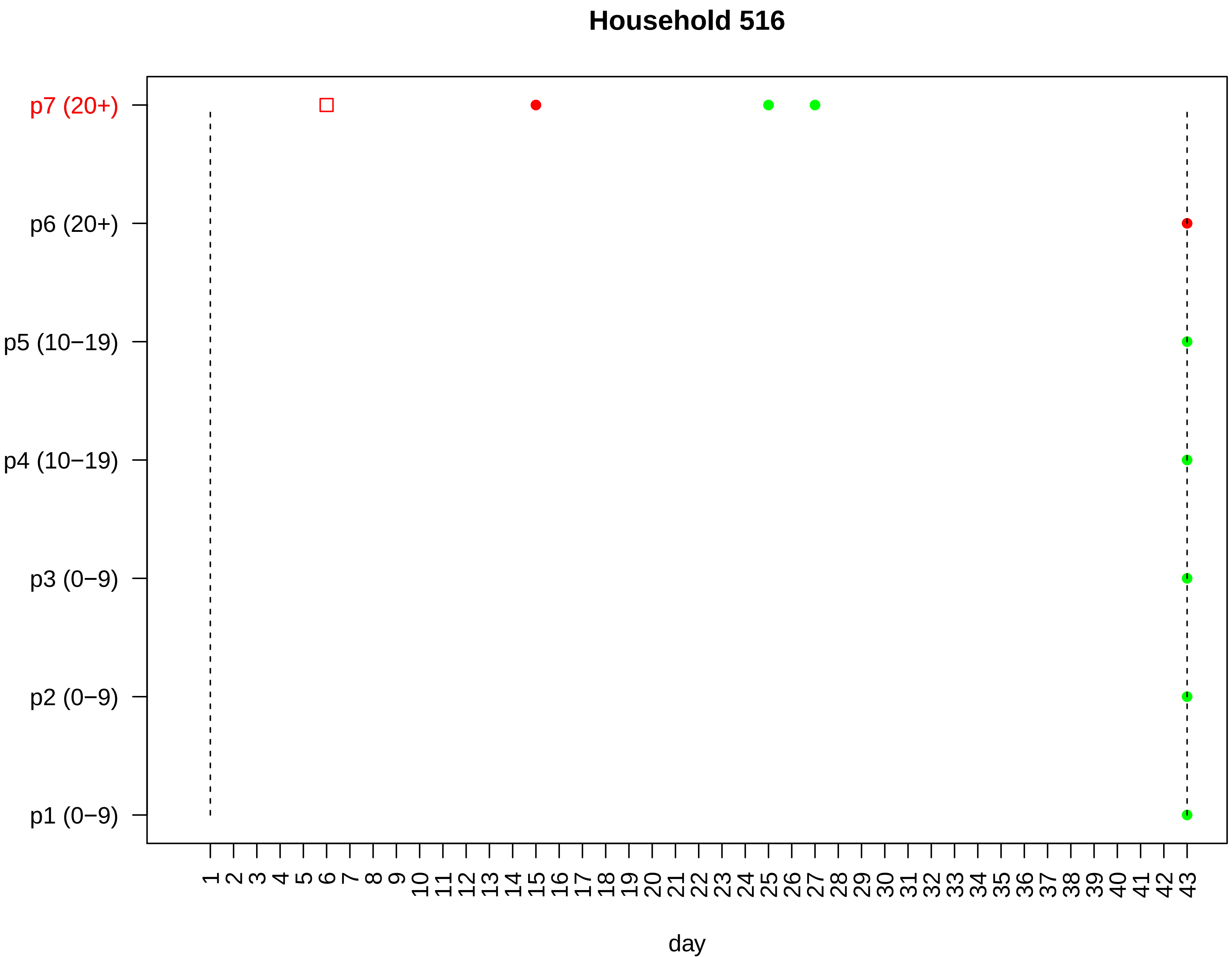

Household 516
p7 (20+)
p6 (20+)
p5 (10−19)
p4 (10−19)
p3 (0−9)
p2 (0−9)
p1 (0−9)
1 2 3 4 5 6 7 8 9 10 11 12 13 14 15 16 17 18 19 20 21 22 23 24 25 26 27 28 29 30 31 32 33 34 35 36 37 38 39 40 41 42 43
day

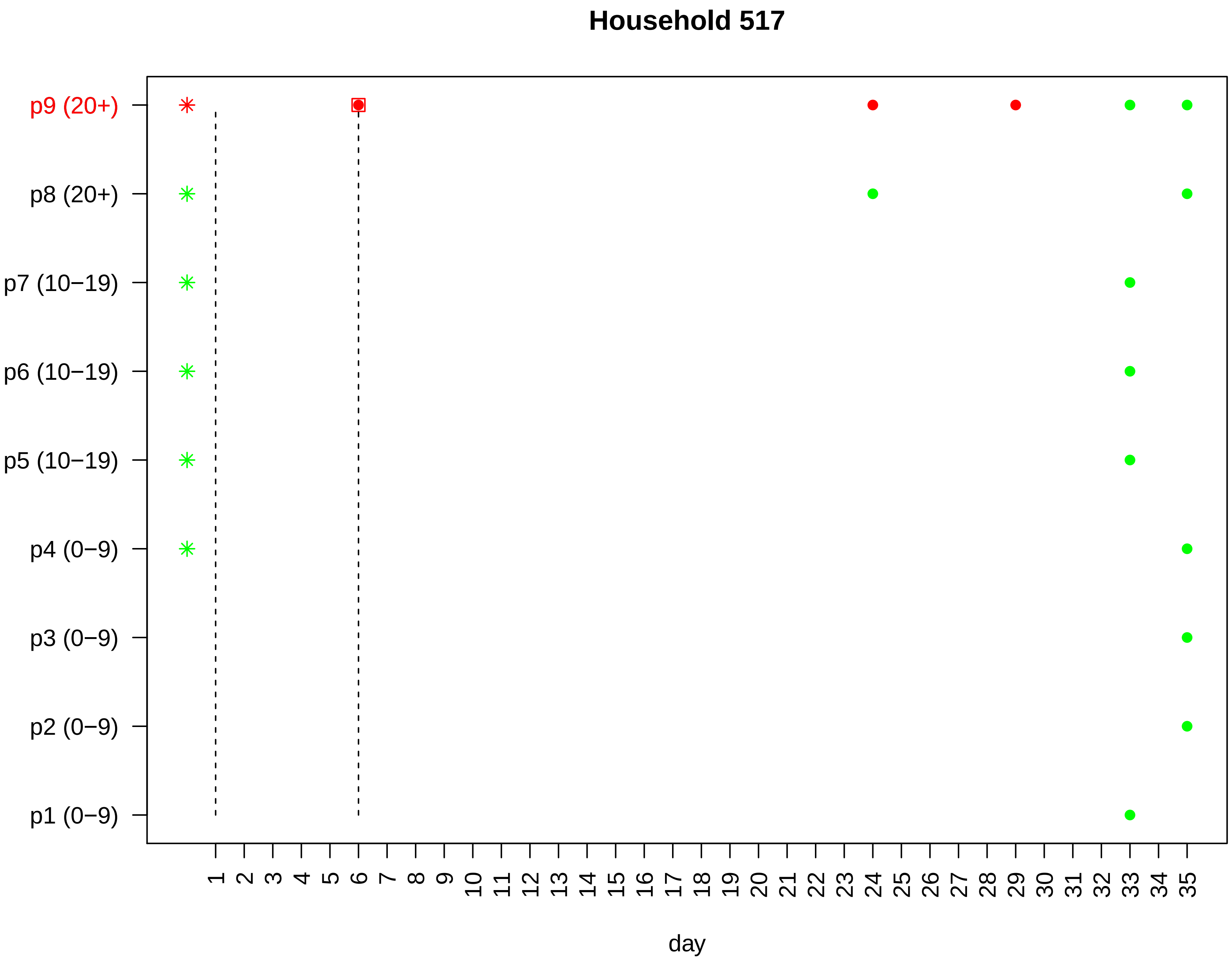

Household 517
p9 (20+)
p8 (20+)
p7 (10−19)
p6 (10−19)
p5 (10−19)
p4 (0−9)
p3 (0−9)
p2 (0−9)
p1 (0−9)
1 2 3 4 5 6 7 8 9 10 11 12 13 14 15 16 17 18 19 20 21 22 23 24 25 26 27 28 29 30 31 32 33 34 35
day

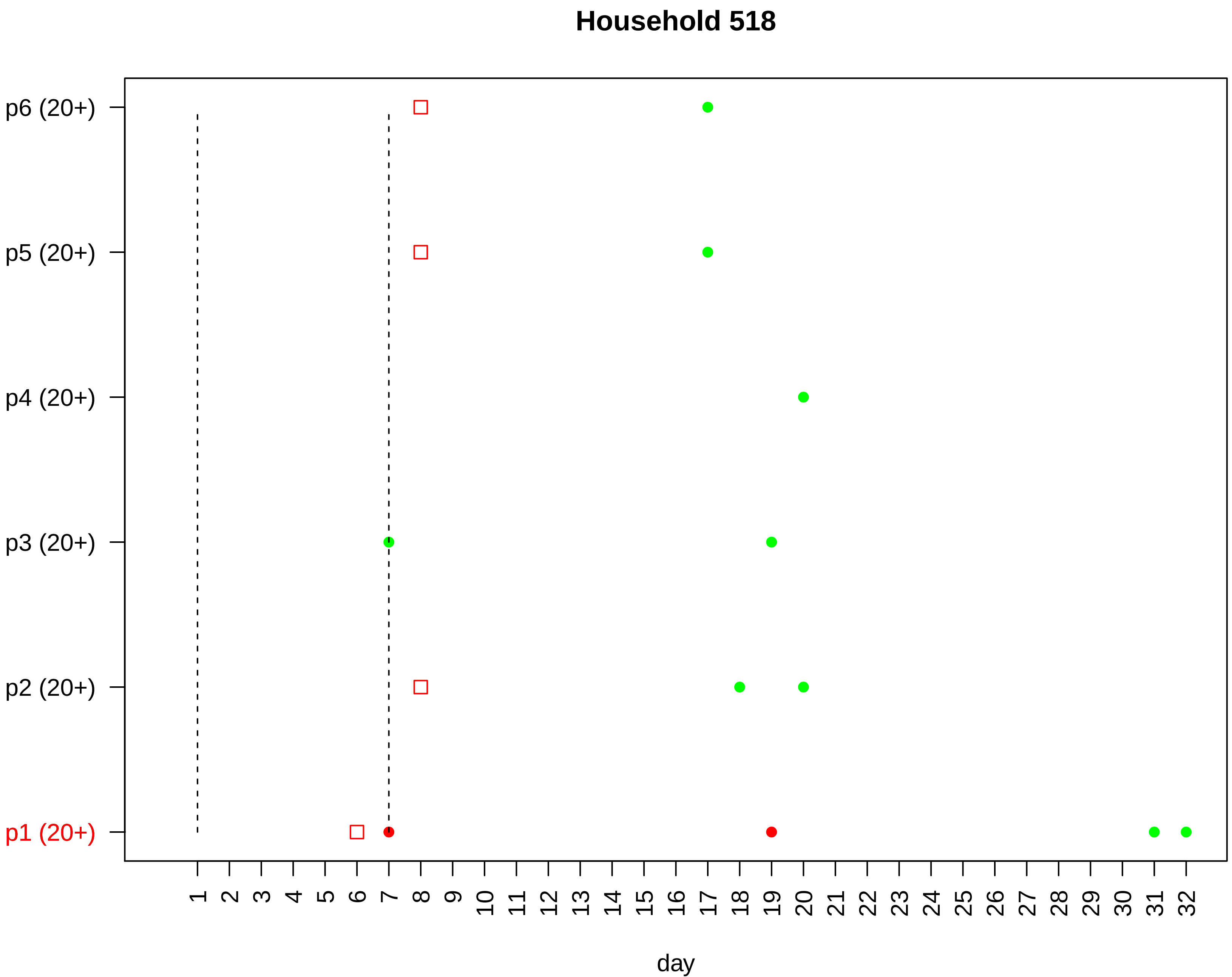

Household 518
p6 (20+)
p5 (20+)
p4 (20+)
p3 (20+)
p2 (20+)
p1 (20+)
1 2 3 4 5 6 7 8 9 10 11 12 13 14 15 16 17 18 19 20 21 22 23 24 25 26 27 28 29 30 31 32
day

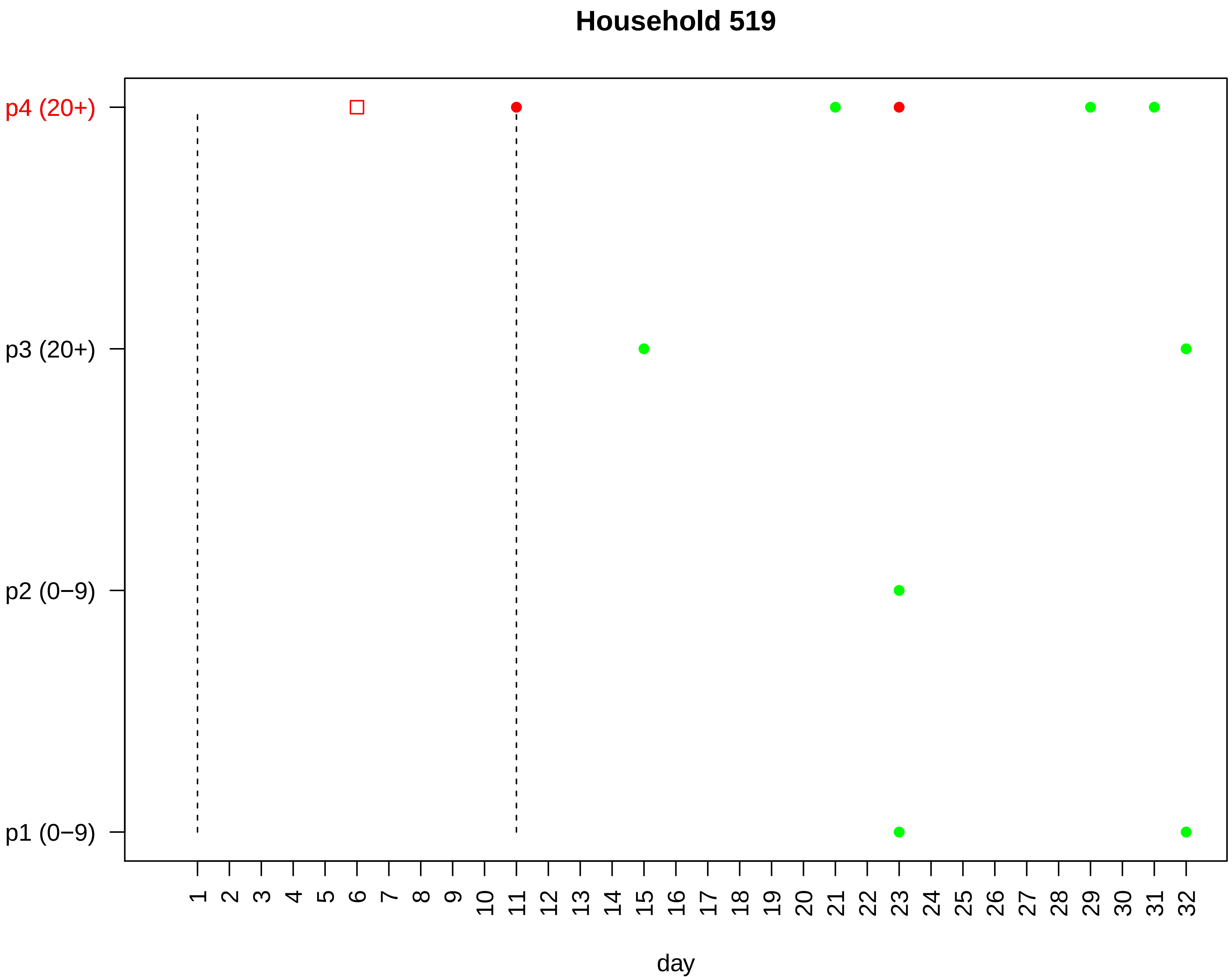

Household 519
p4 (20+)
p3 (20+)
p2 (0−9)
p1 (0−9)
day

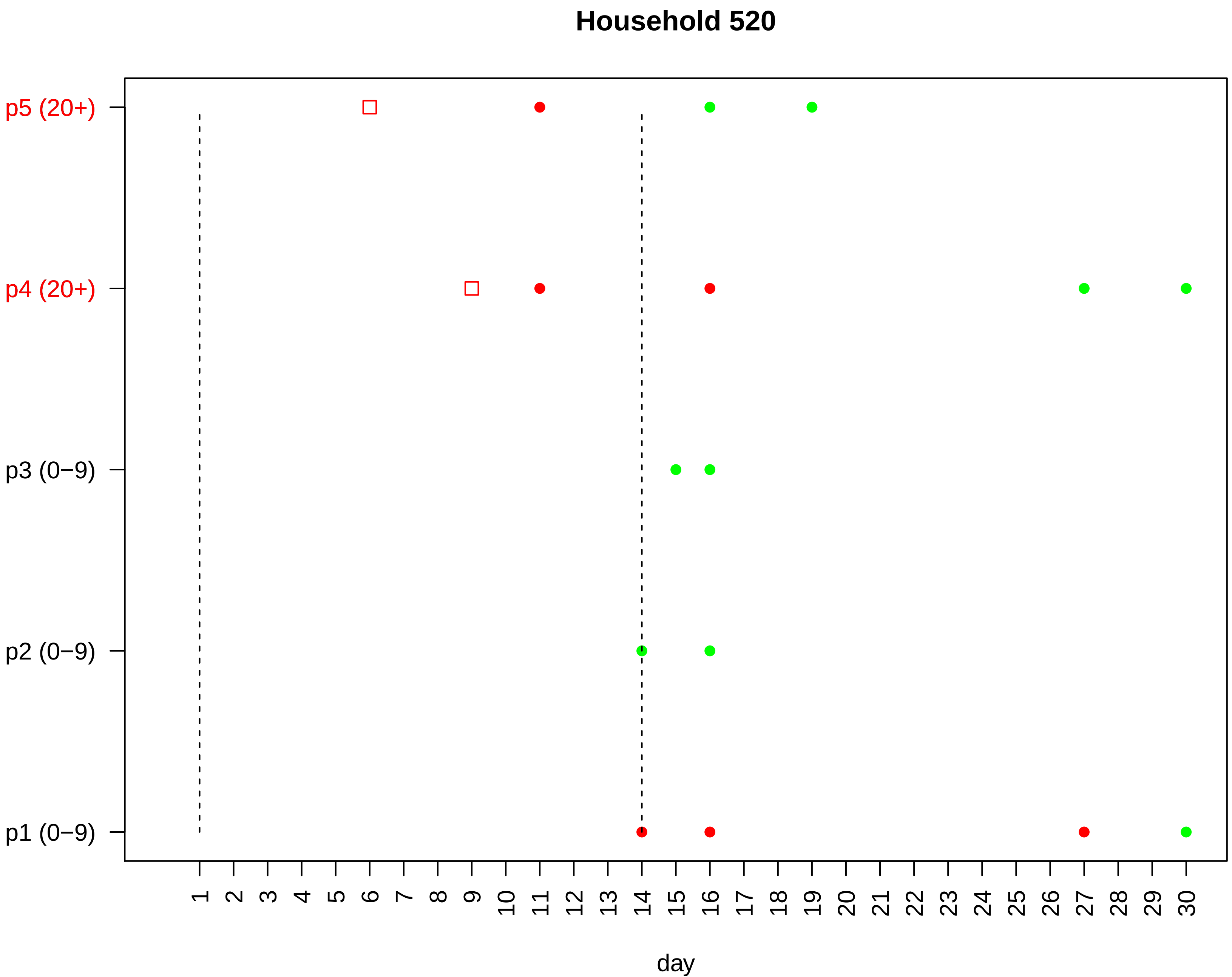
Household 520
p5 (20+)
p4 (20+)
p3 (0−9)
p2 (0−9)
p1 (0−9)
day

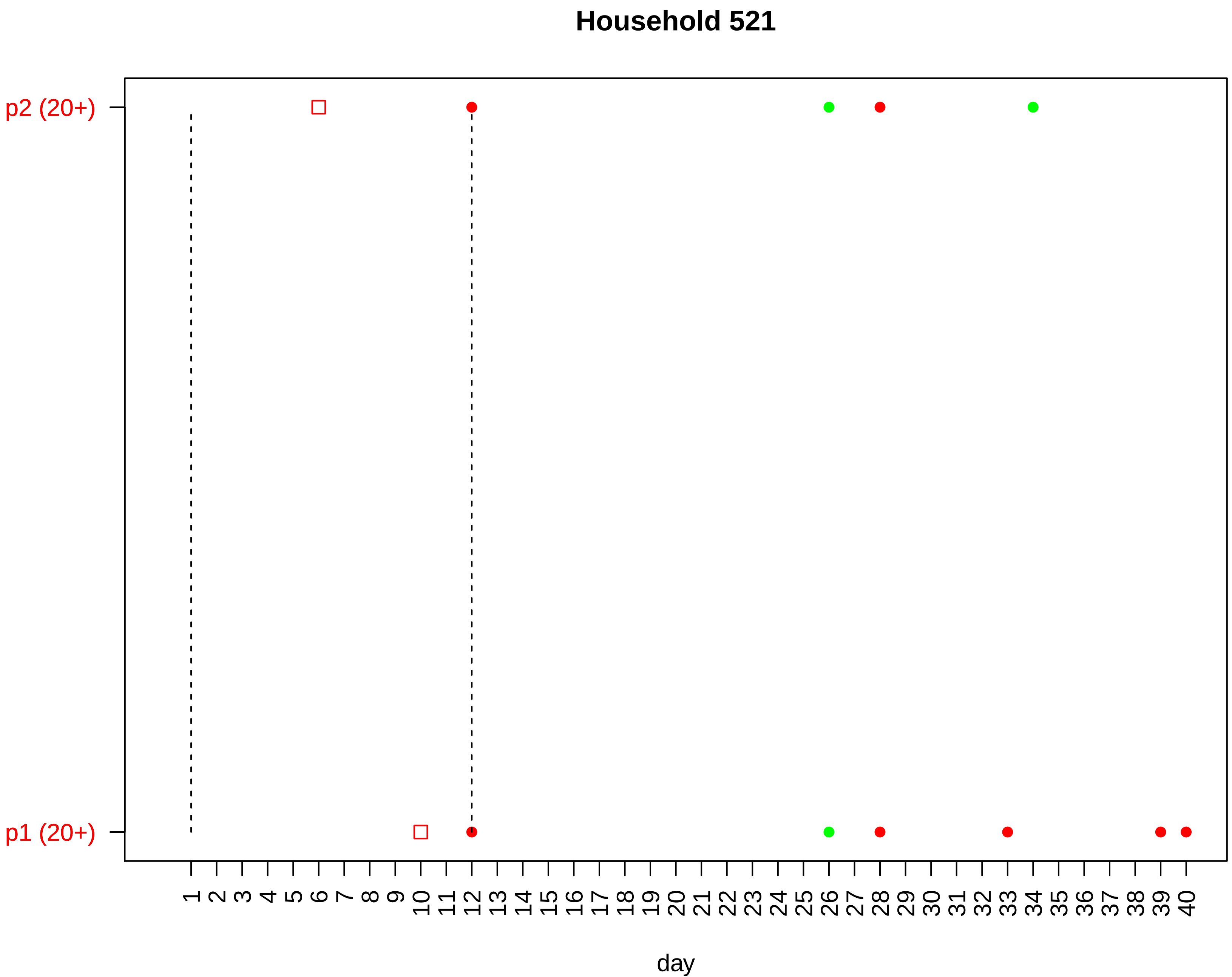

Household 521
p2 (20+)
p1 (20+)
1 2 3 4 5 6 7 8 9 10 11 12 13 14 15 16 17 18 19 20 21 22 23 24 25 26 27 28 29 30 31 32 33 34 35 36 37 38 39 40
day

#### Household 522

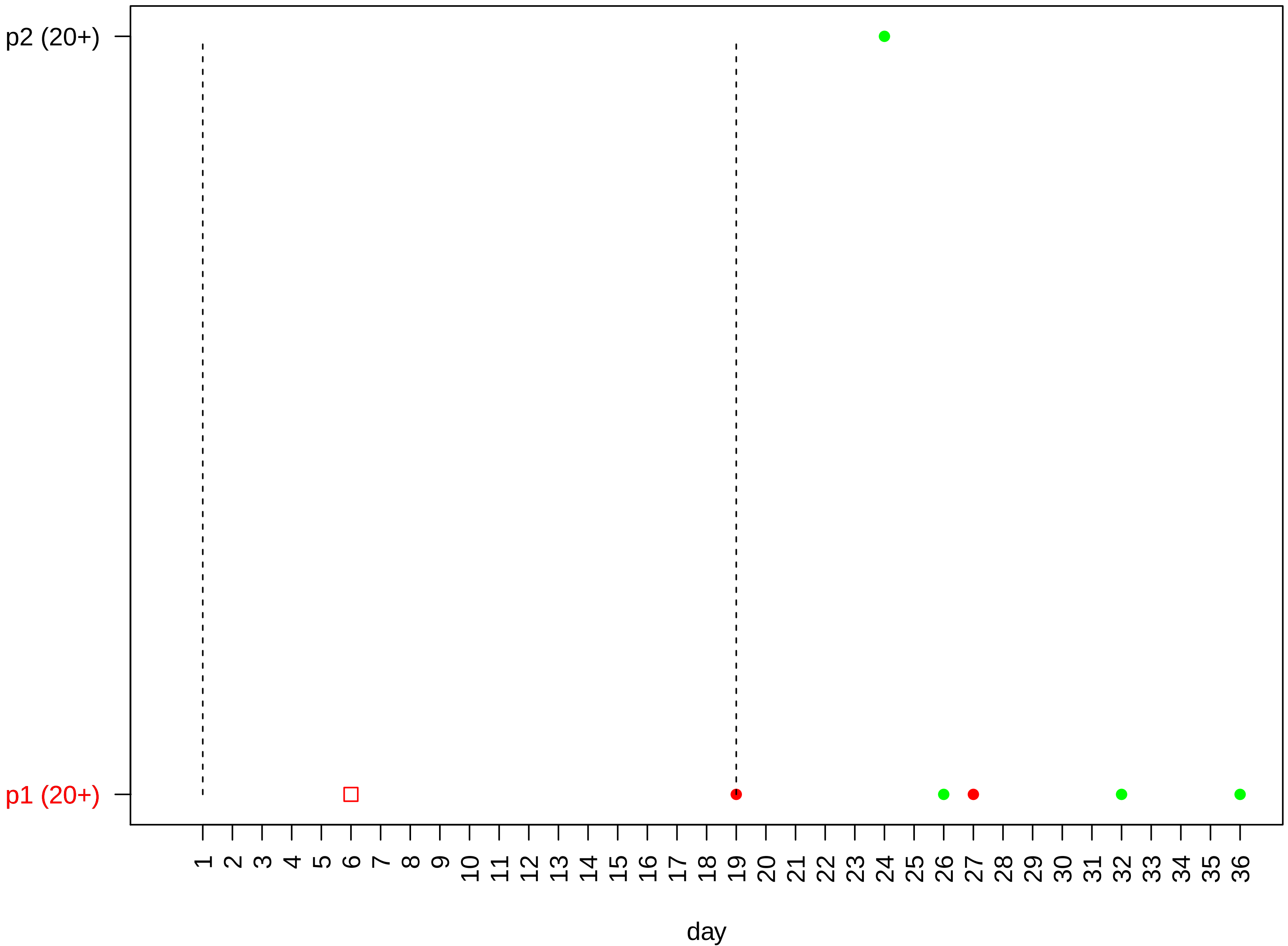

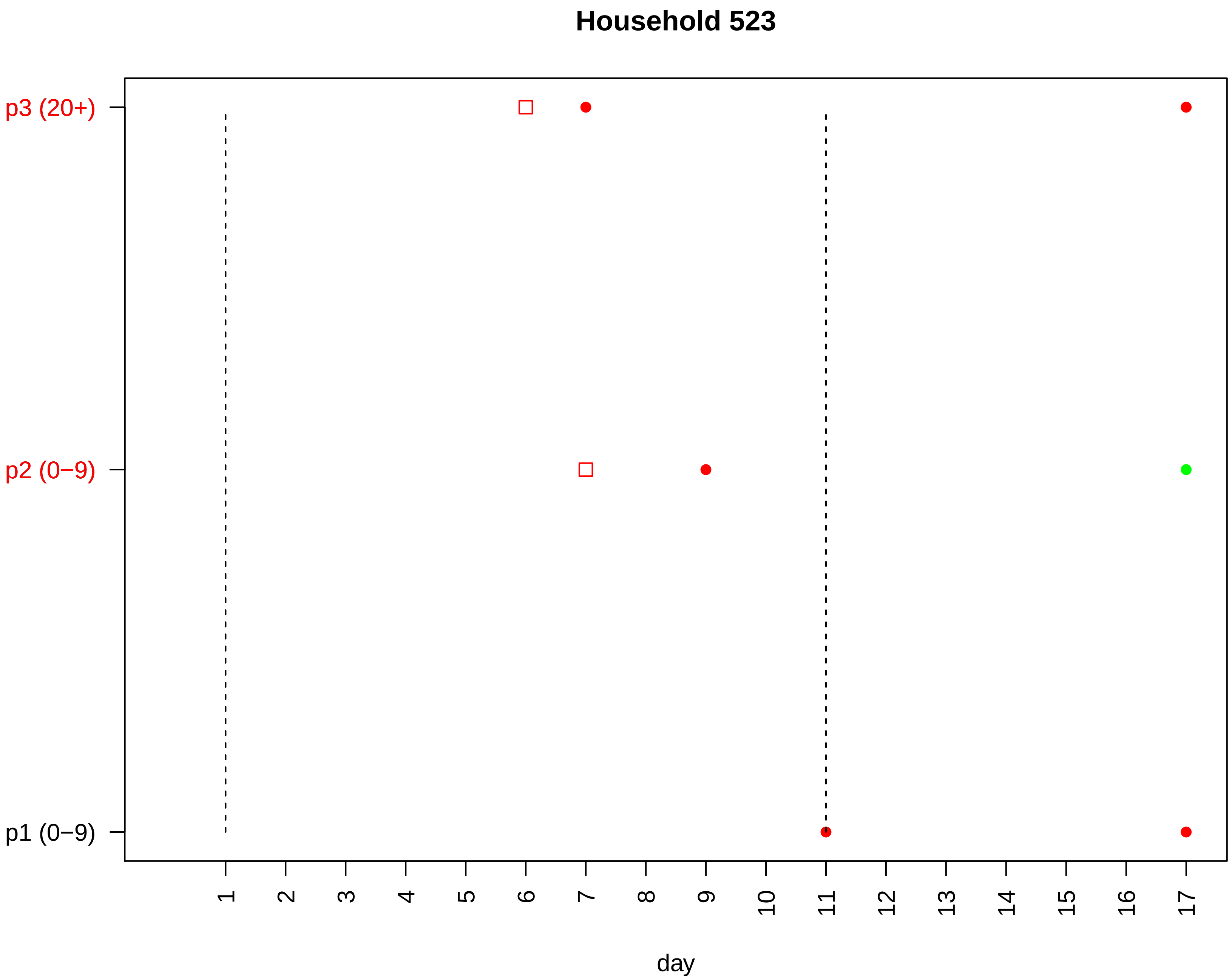

Household 523
p3 (20+)
p2 (0−9)
p1 (0−9)
day

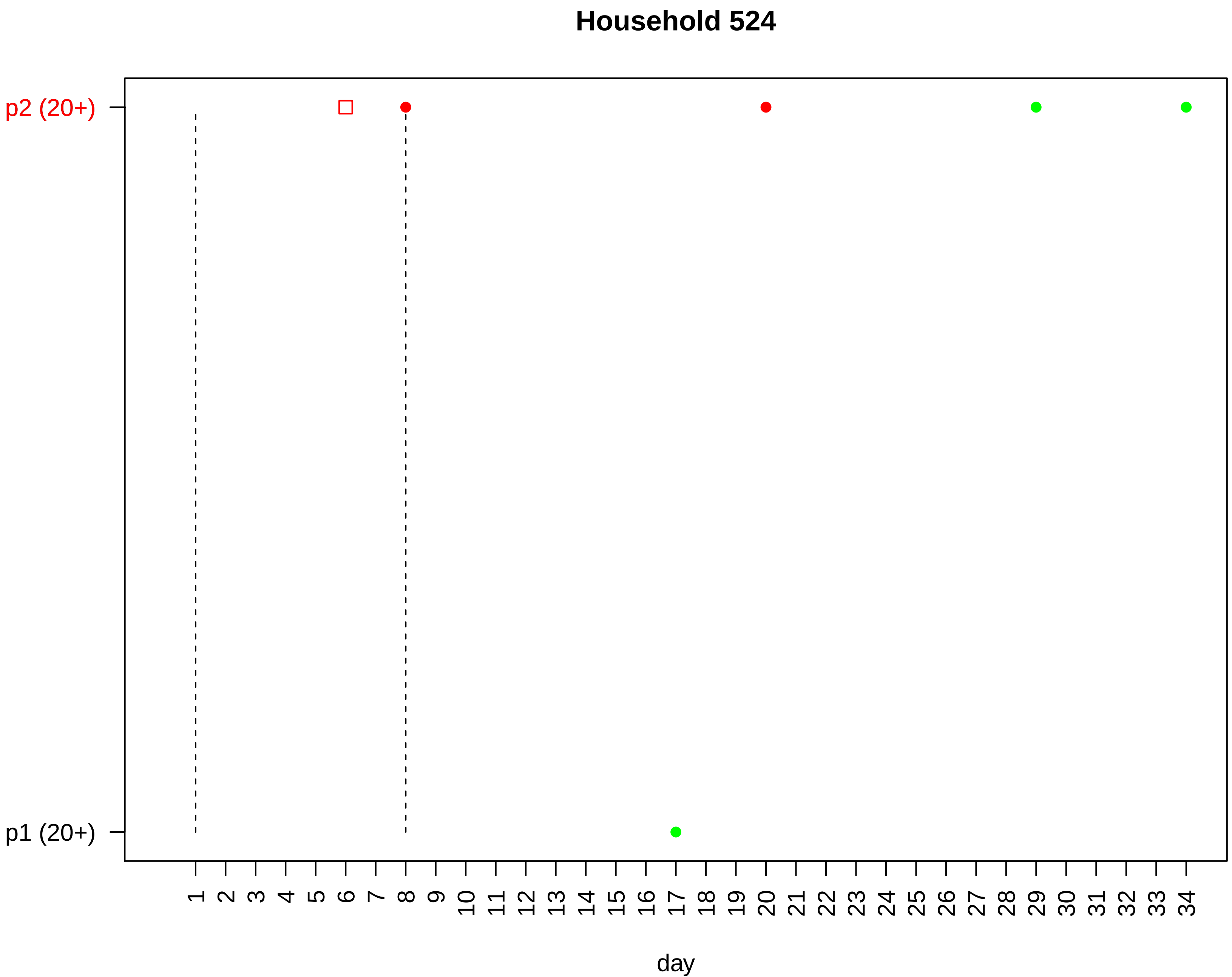

Household 524
p2 (20+)
p1 (20+)
day

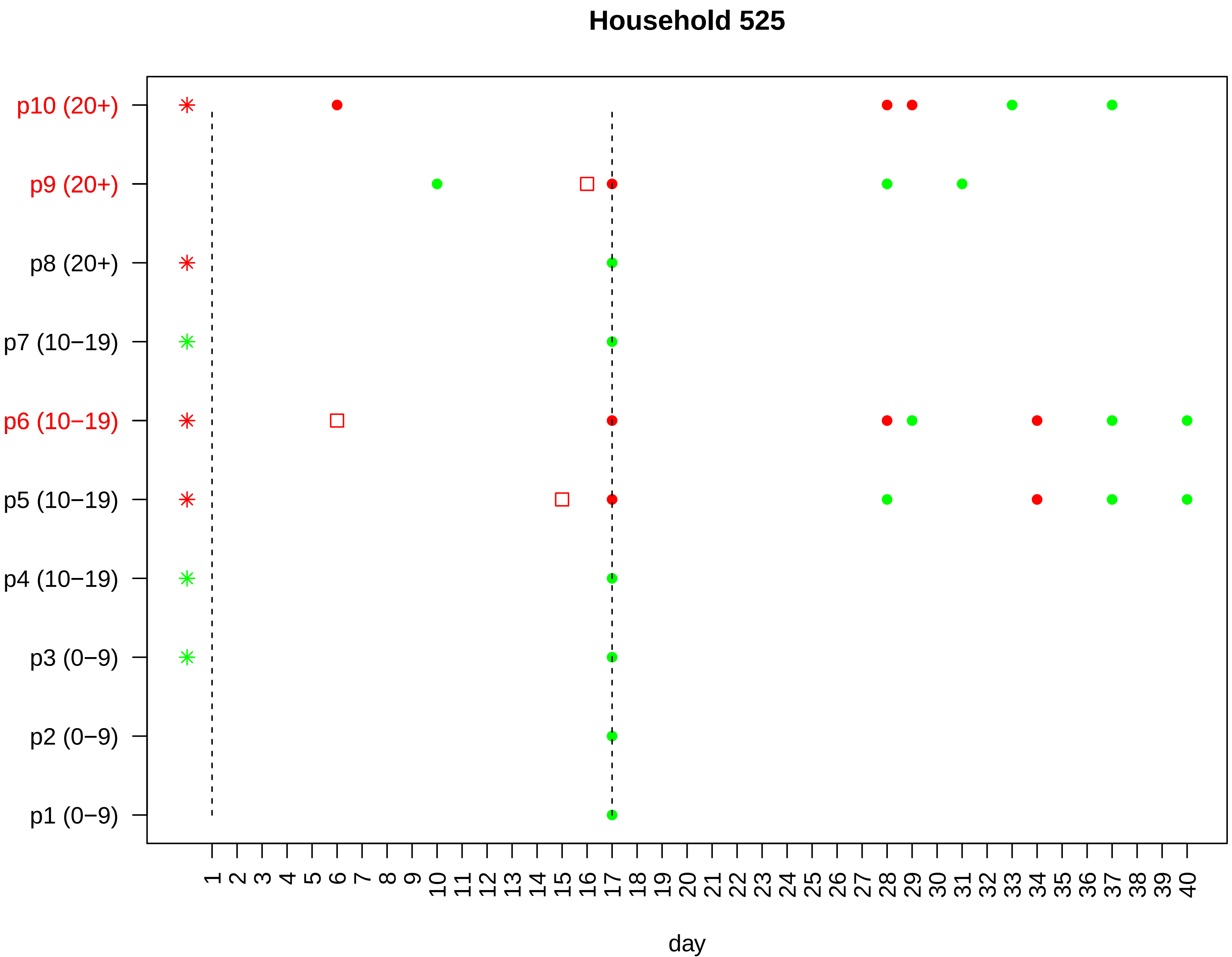

Household 525
p10 (20+)
p9 (20+)
p8 (20+)
p7 (10−19)
p6 (10−19)
p5 (10−19)
p4 (10−19)
p3 (0−9)
p2 (0−9)
p1 (0−9)
1 2 3 4 5 6 7 8 9 10 11 12 13 14 15 16 17 18 19 20 21 22 23 24 25 26 27 28 29 30 31 32 33 34 35 36 37 38 39 40
day

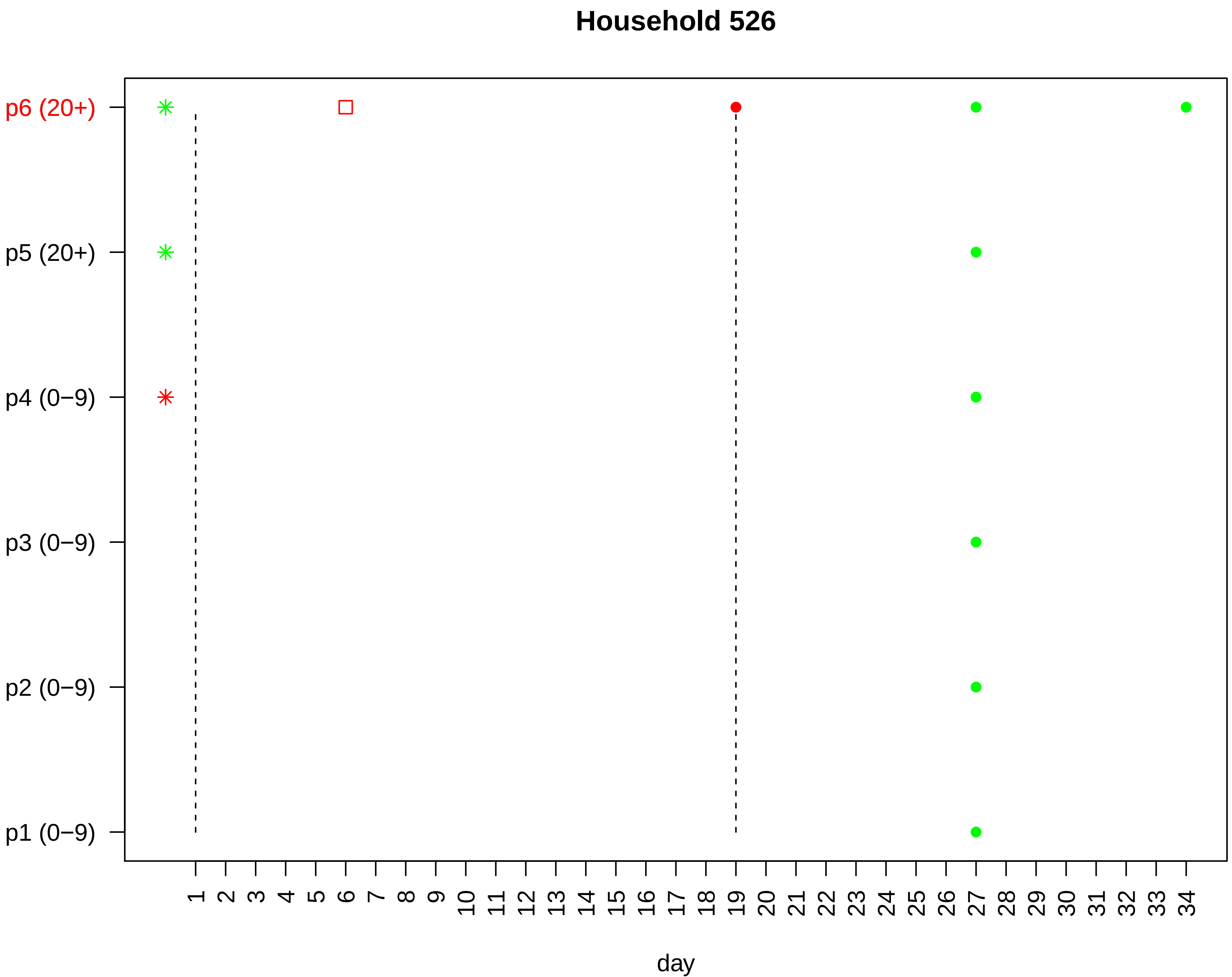

Household 526
p6 (20+)
p5 (20+)
p4 (0−9)
p3 (0−9)
p2 (0−9)
p1 (0−9)
1 2 3 4 5 6 7 8 9 10 11 12 13 14 15 16 17 18 19 20 21 22 23 24 25 26 27 28 29 30 31 32 33 34
day

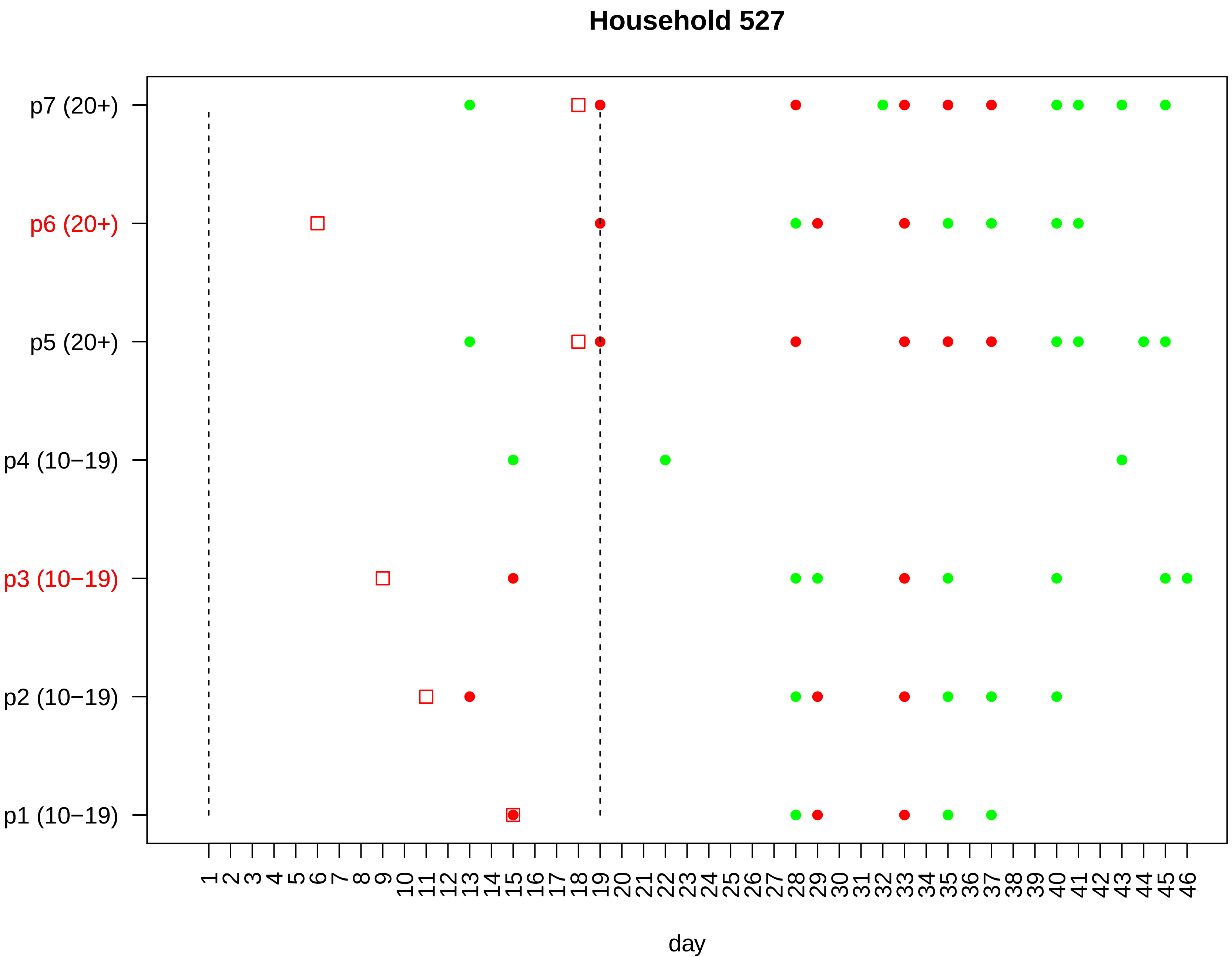

Household 527
p7 (20+)
p6 (20+)
p5 (20+)
p4 (10−19)
p3 (10−19)
p2 (10−19)
p1 (10−19)
1 2 3 4 5 6 7 8 9 10 11 12 13 14 15 16 17 18 19 20 21 22 23 24 25 26 27 28 29 30 31 32 33 34 35 36 37 38 39 40 41 42 43 44 45 46
day

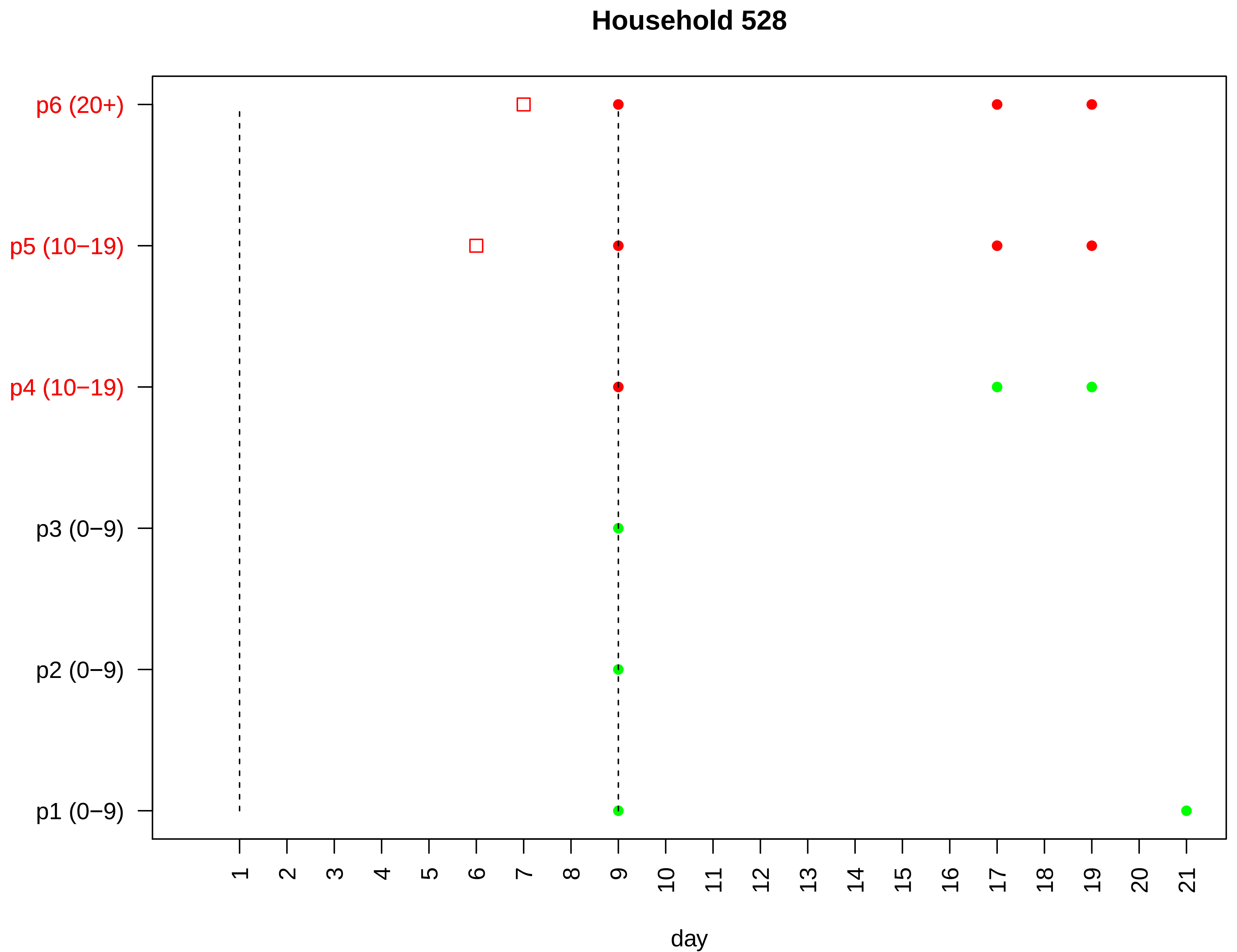
Household 528
p6 (20+)
p5 (10−19)
p4 (10−19)
p3 (0−9)
p2 (0−9)
p1 (0−9)
day

Household 529

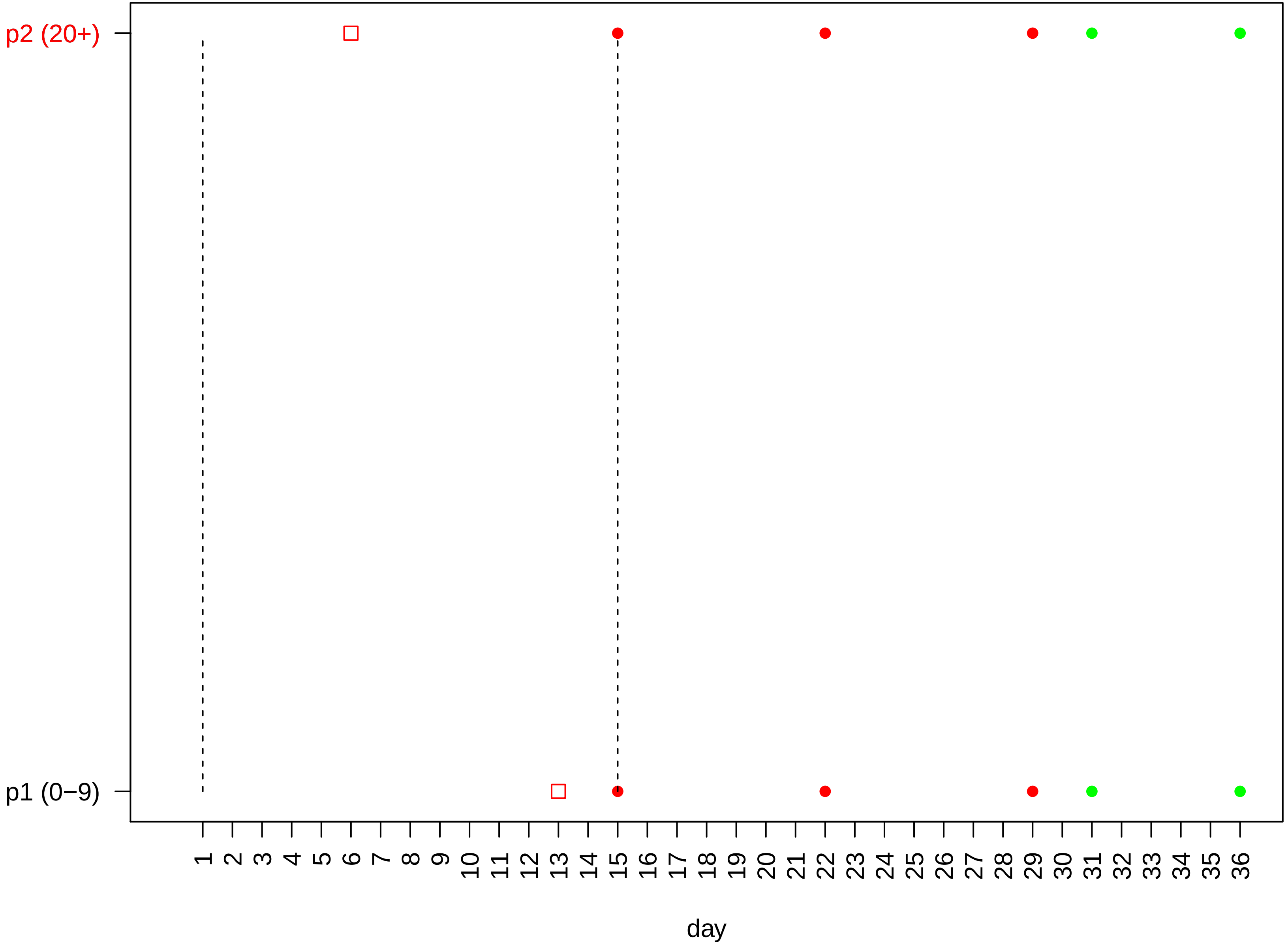

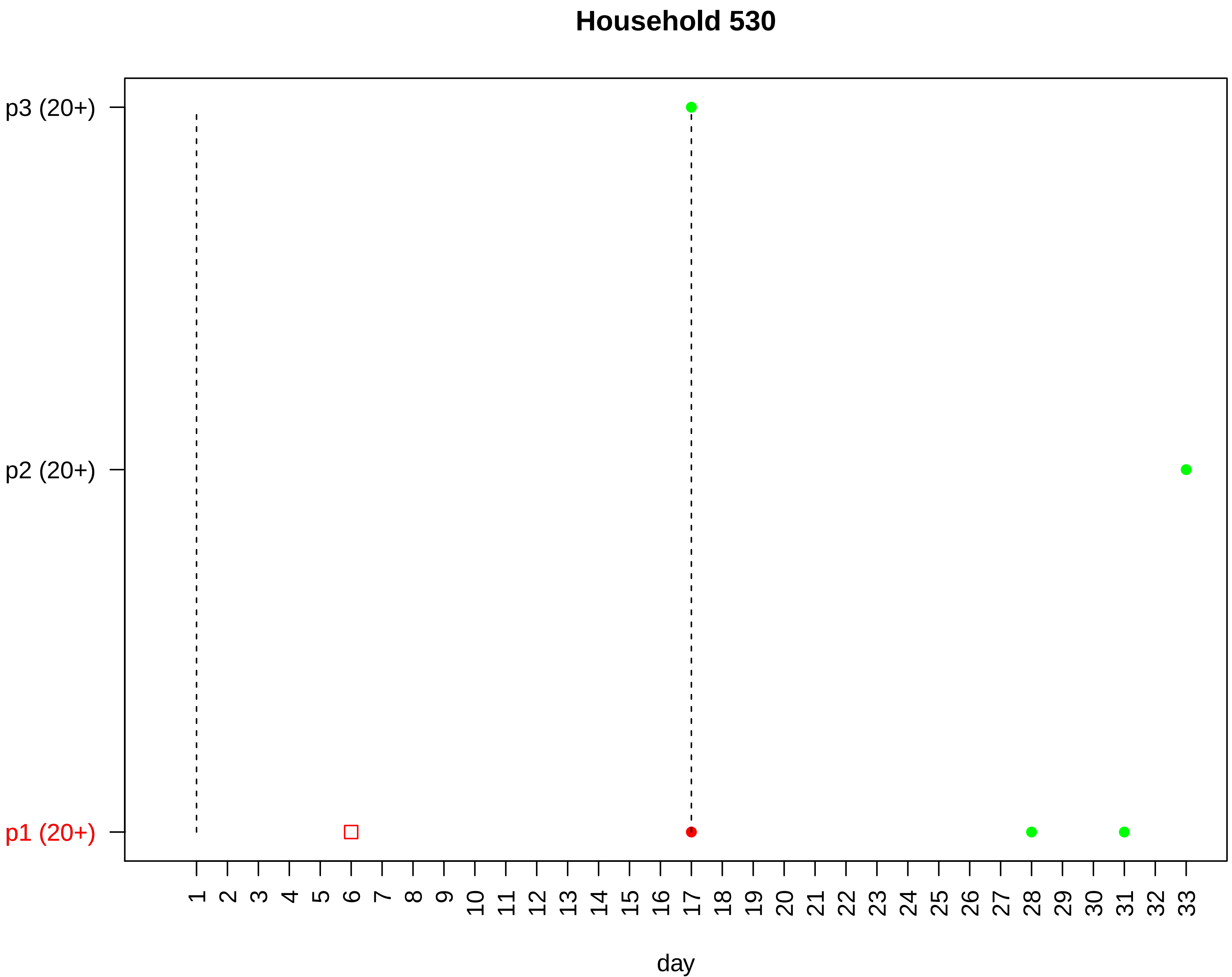

Household 530
p3 (20+)
p2 (20+)
p1 (20+)
day

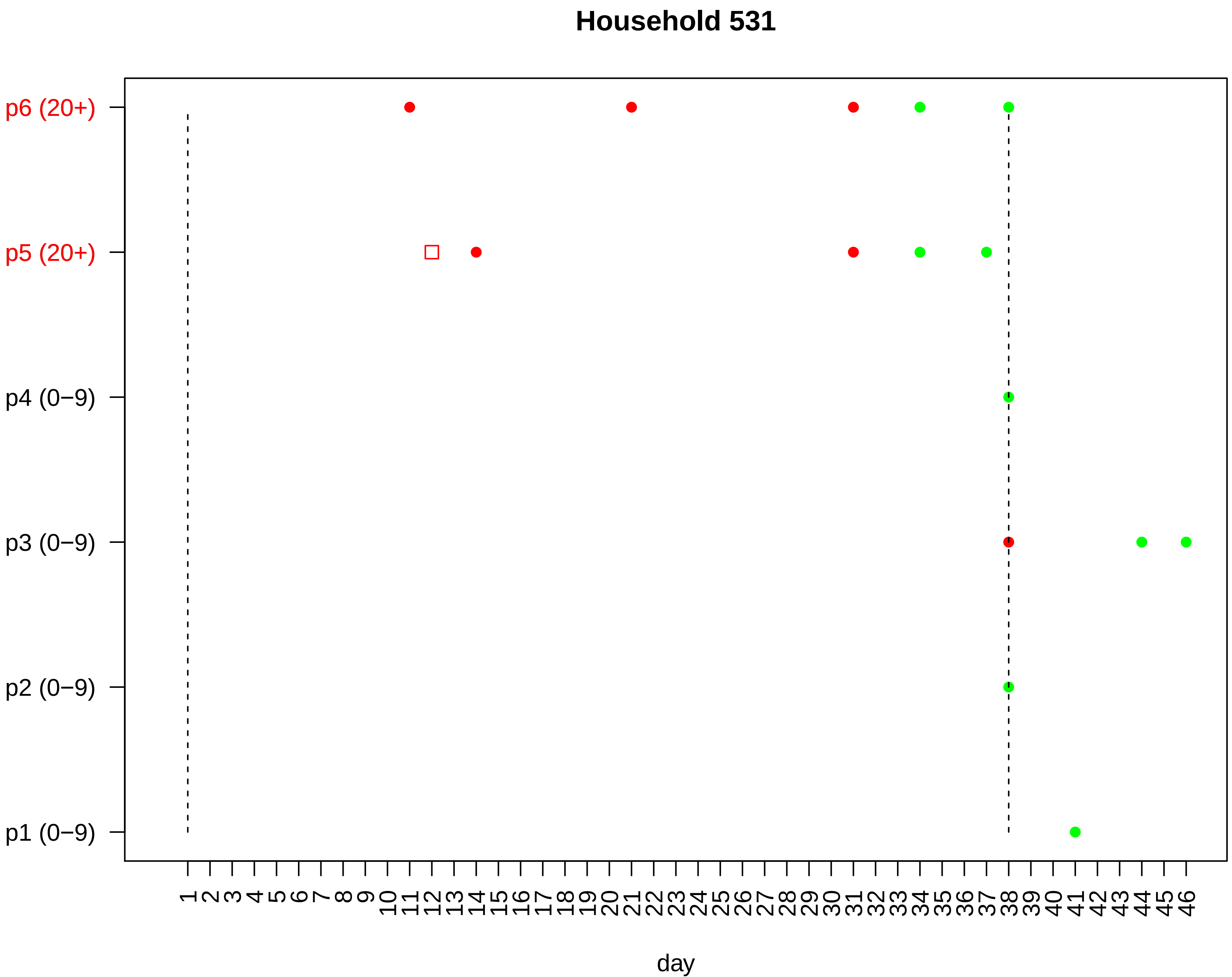

Household 531
p6 (20+)
p5 (20+)
p4 (0−9)
p3 (0−9)
p2 (0−9)
p1 (0−9)
1 2 3 4 5 6 7 8 9 10 11 12 13 14 15 16 17 18 19 20 21 22 23 24 25 26 27 28 29 30 31 32 33 34 35 36 37 38 39 40 41 42 43 44 45 46
day

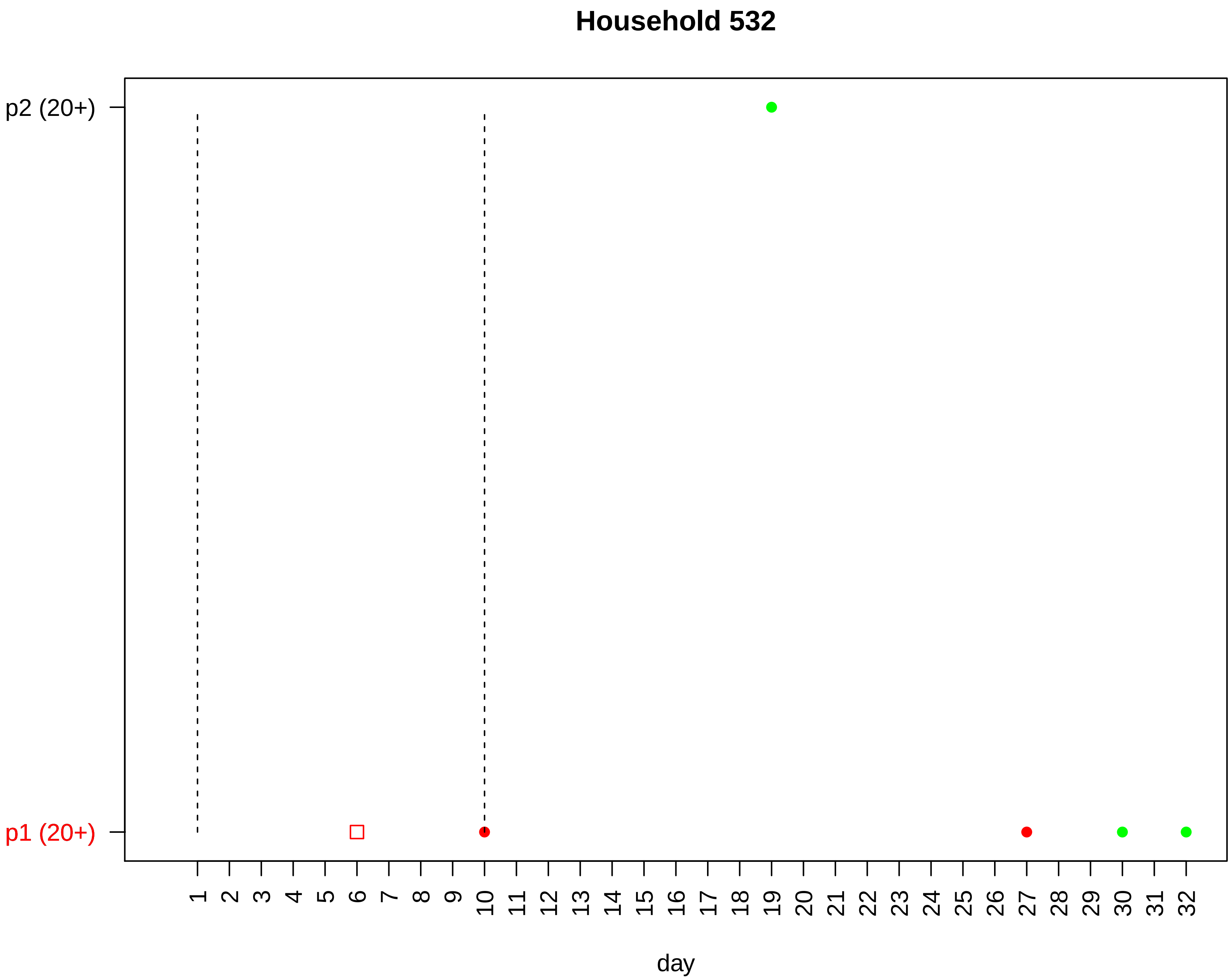

Household 532
p2 (20+)
p1 (20+)
day

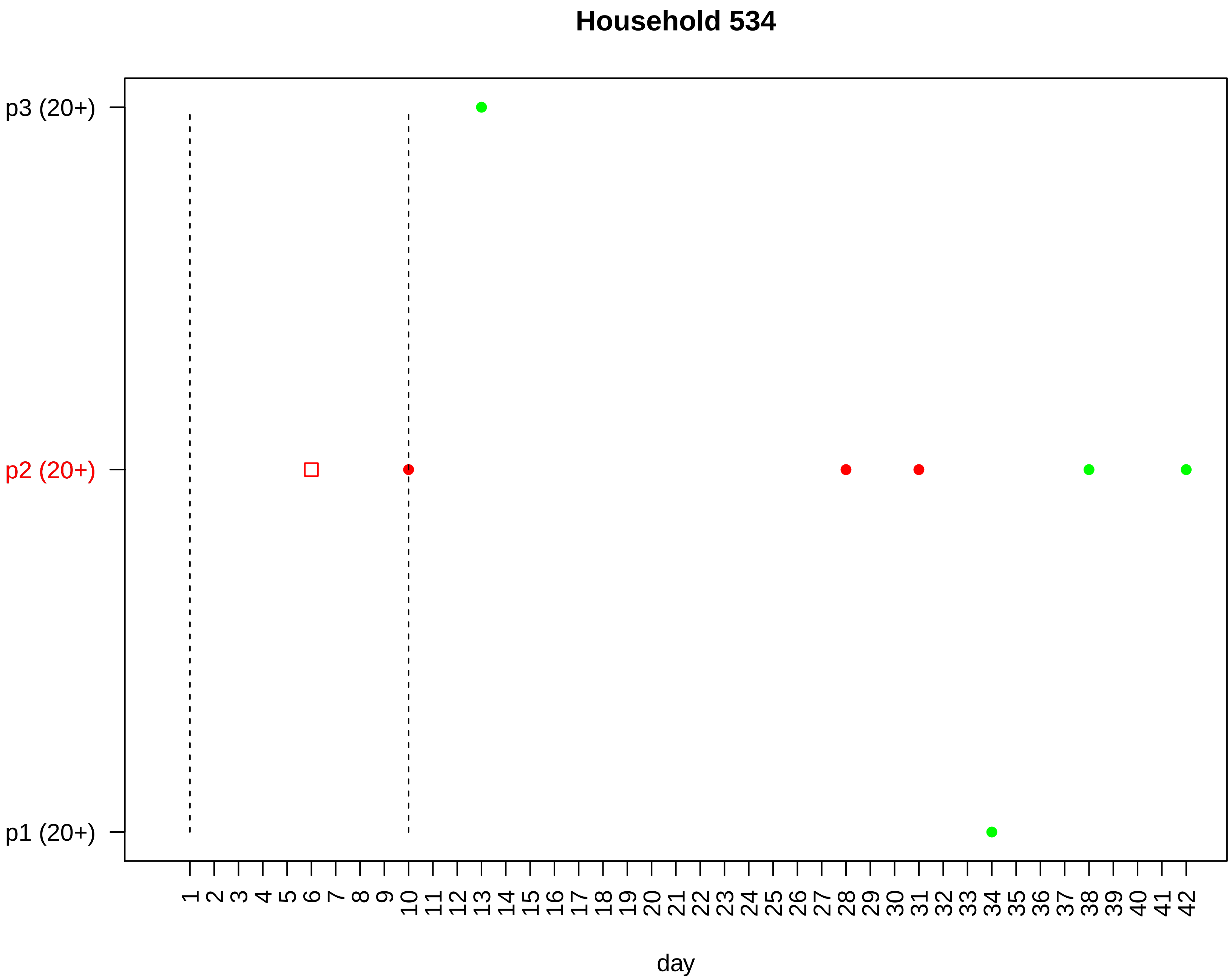

Household 534
p3 (20+)
p2 (20+)
p1 (20+)
1 2 3 4 5 6 7 8 9 10 11 12 13 14 15 16 17 18 19 20 21 22 23 24 25 26 27 28 29 30 31 32 33 34 35 36 37 38 39 40 41 42
day

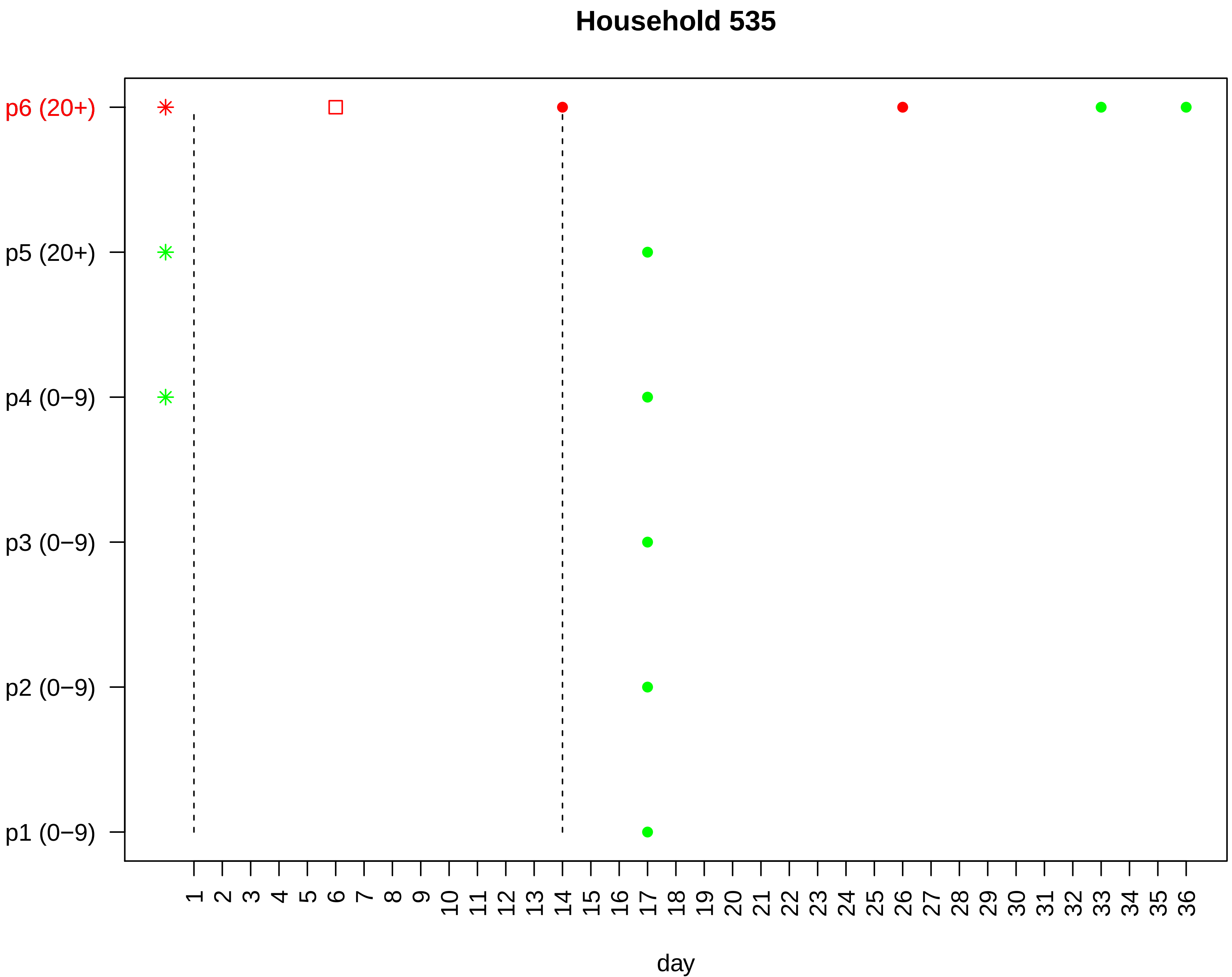
Household 535
p6 (20+)
p5 (20+)
p4 (0−9)
p3 (0−9)
p2 (0−9)
p1 (0−9)
1 2 3 4 5 6 7 8 9 10 11 12 13 14 15 16 17 18 19 20 21 22 23 24 25 26 27 28 29 30 31 32 33 34 35 36
day

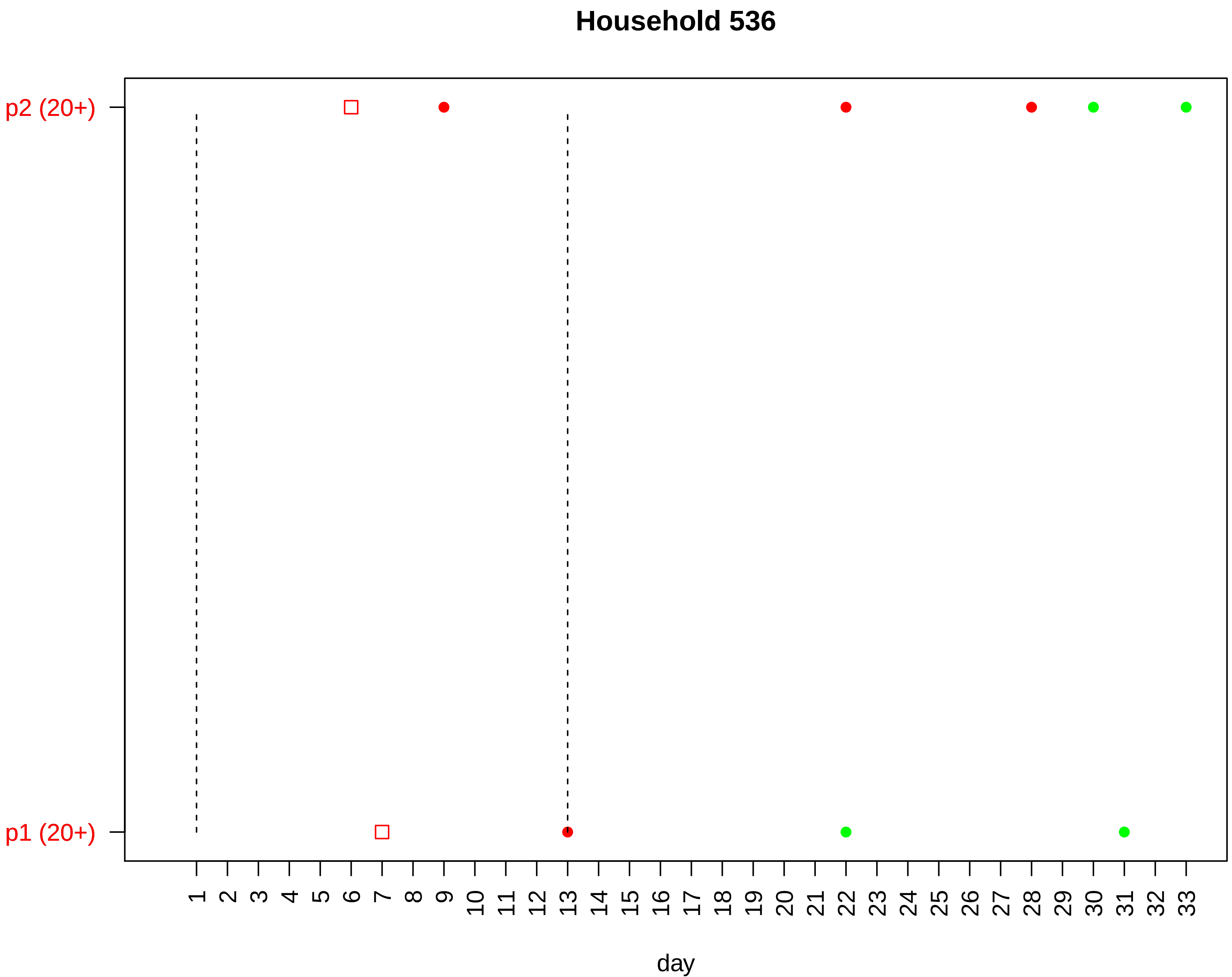

Household 536
p2 (20+)
p1 (20+)
day

### Household 537

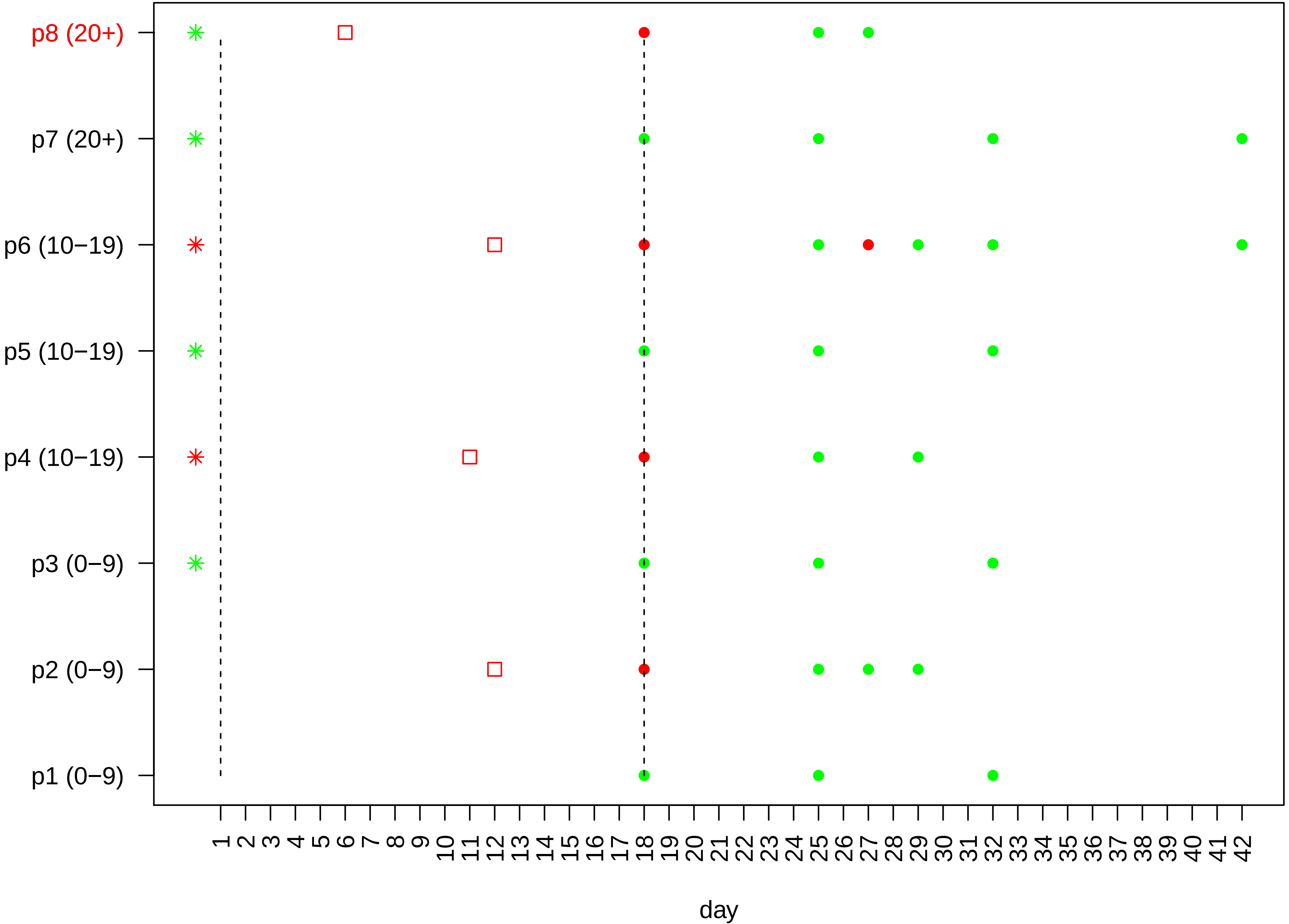

#### Household 538

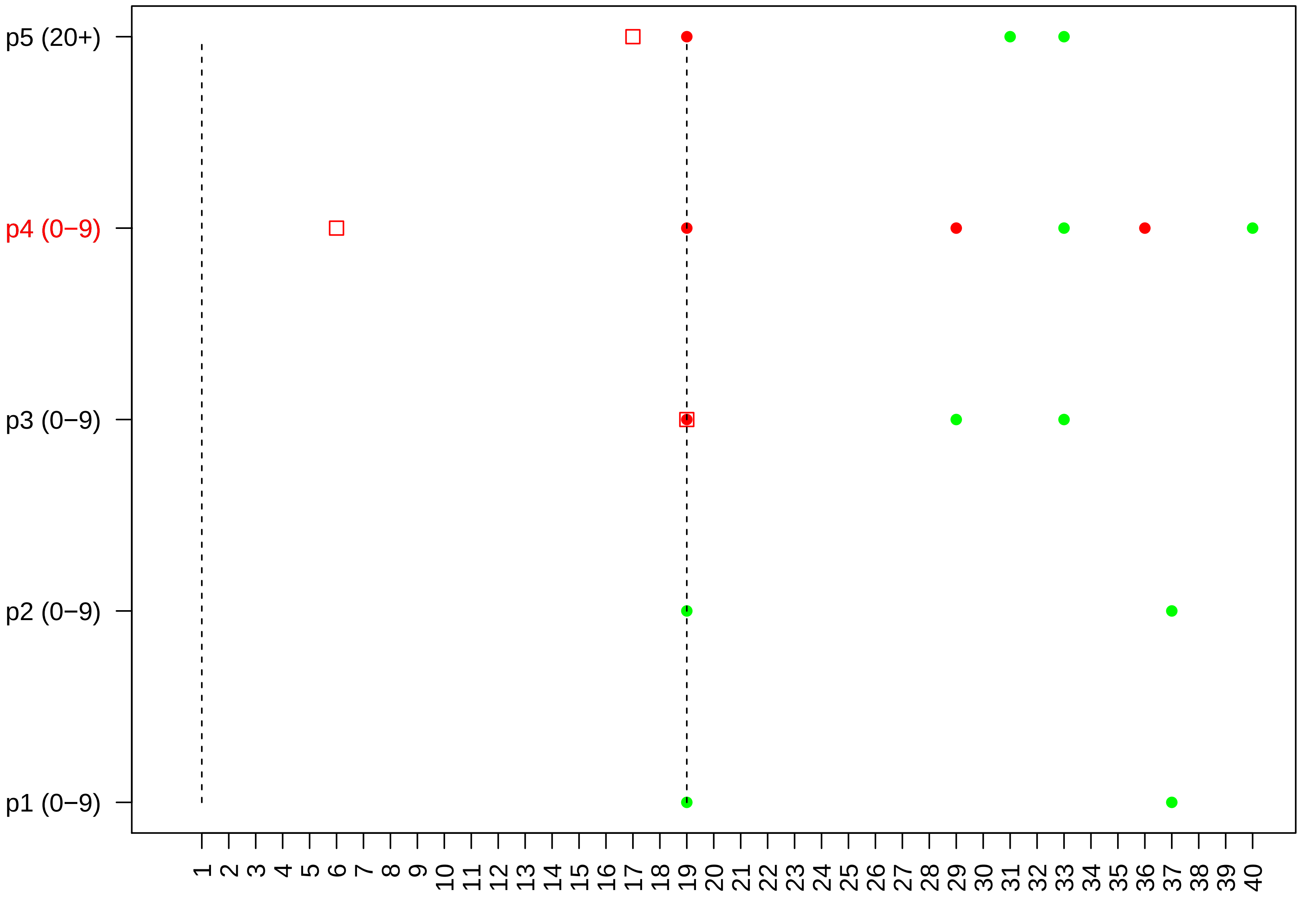

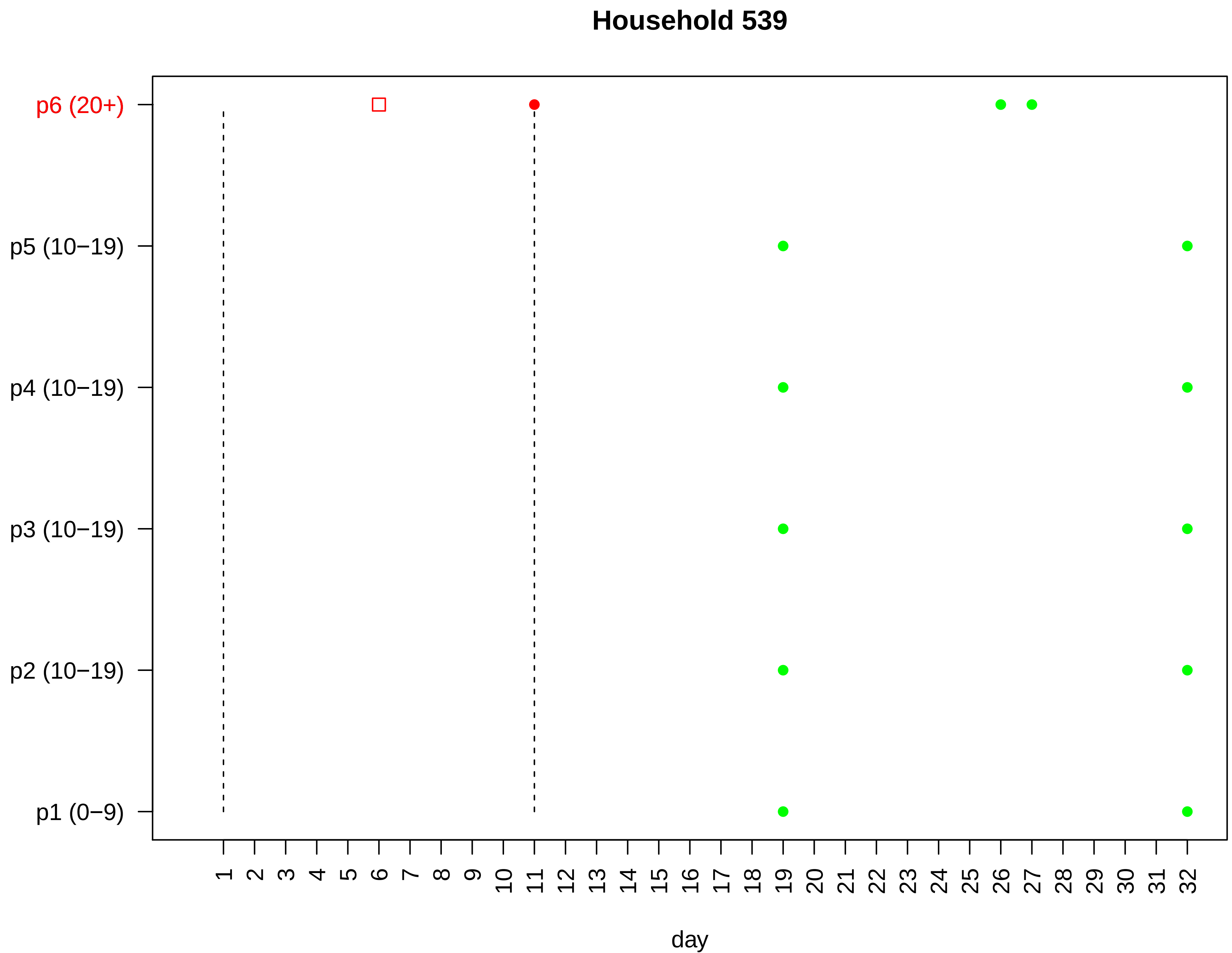

Household 539
p6 (20+)
p5 (10−19)
p4 (10−19)
p3 (10−19)
p2 (10−19)
p1 (0−9)
1 2 3 4 5 6 7 8 9 10 11 12 13 14 15 16 17 18 19 20 21 22 23 24 25 26 27 28 29 30 31 32
day

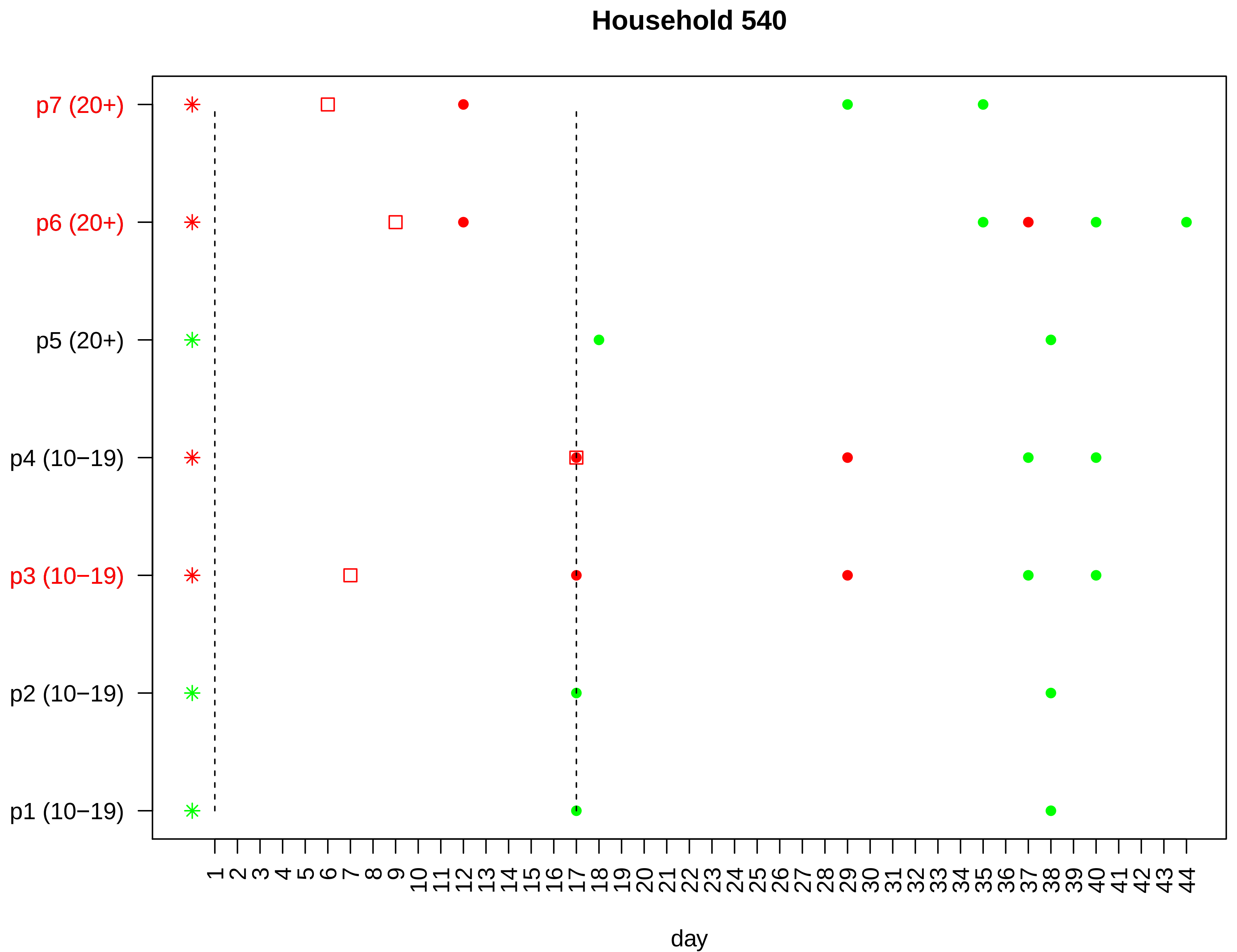

Household 540
p7 (20+)
p6 (20+)
p5 (20+)
p4 (10−19)
p3 (10−19)
p2 (10−19)
p1 (10−19)
1 2 3 4 5 6 7 8 9 10 11 12 13 14 15 16 17 18 19 20 21 22 23 24 25 26 27 28 29 30 31 32 33 34 35 36 37 38 39 40 41 42 43 44
day

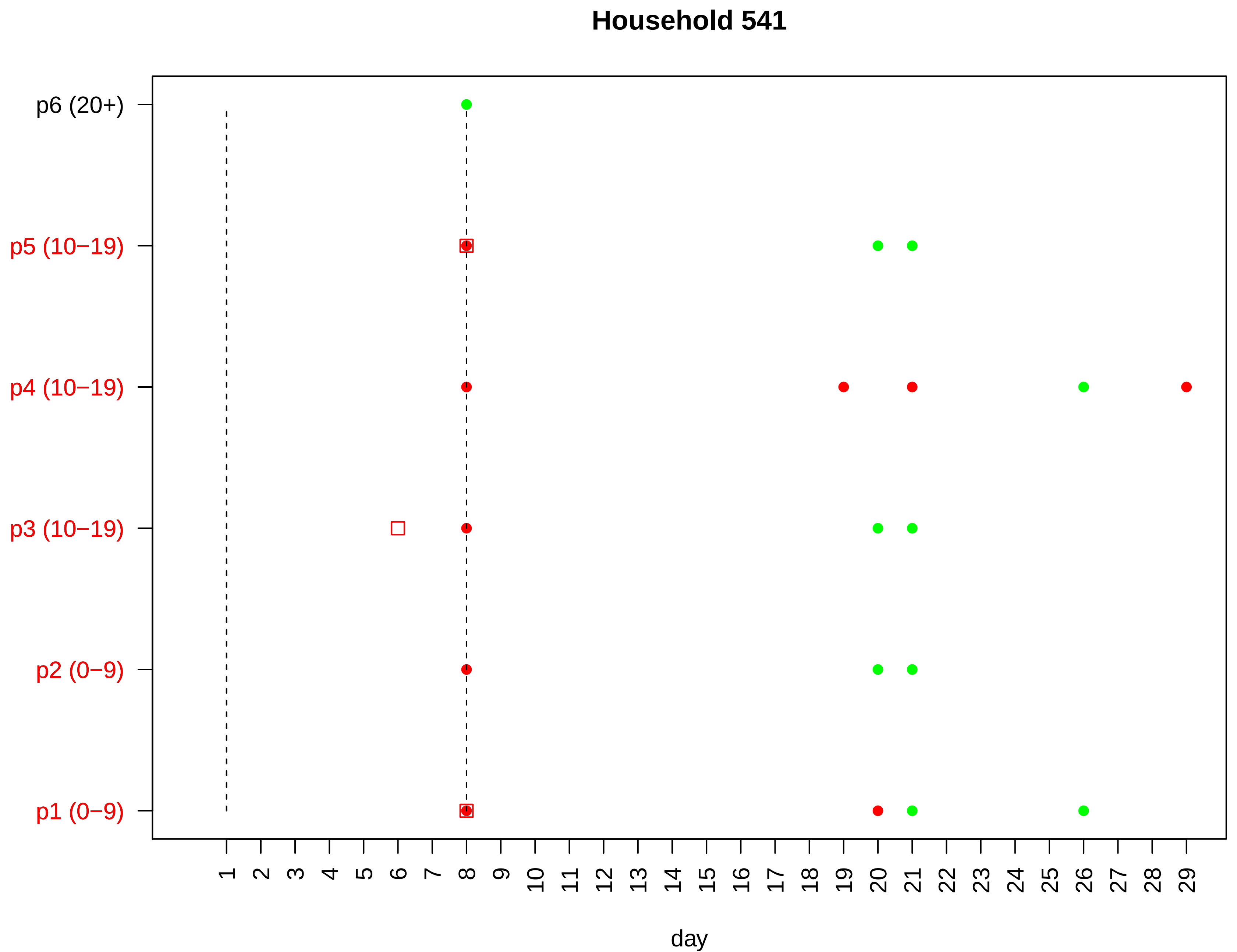

Household 541
p6 (20+)
p5 (10−19)
p4 (10−19)
p3 (10−19)
p2 (0−9)
p1 (0−9)
1 2 3 4 5 6 7 8 9 10 11 12 13 14 15 16 17 18 19 20 21 22 23 24 25 26 27 28 29
day

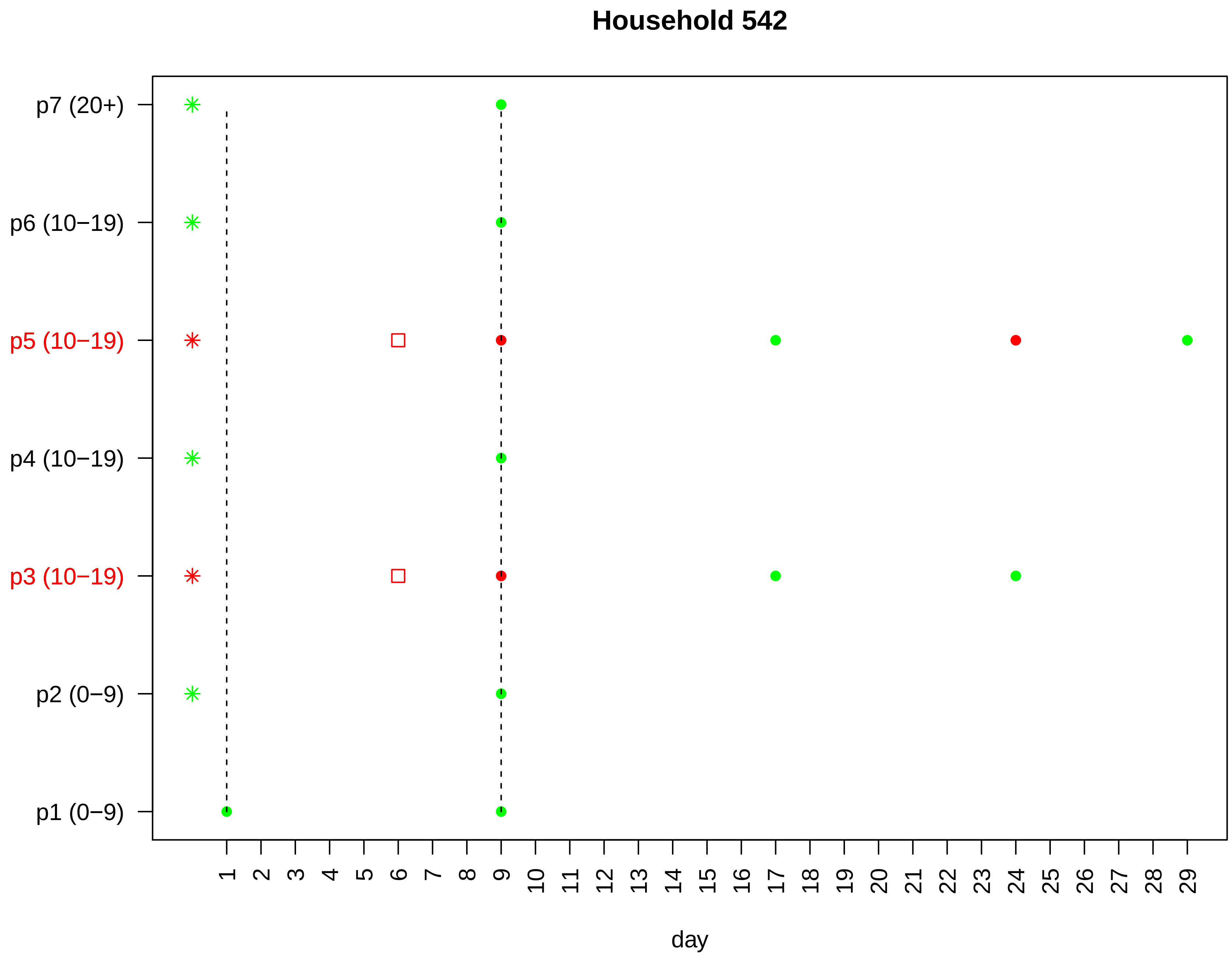

Household 542
p7 (20+)
p6 (10−19)
p5 (10−19)
p4 (10−19)
p3 (10−19)
p2 (0−9)
p1 (0−9)
day

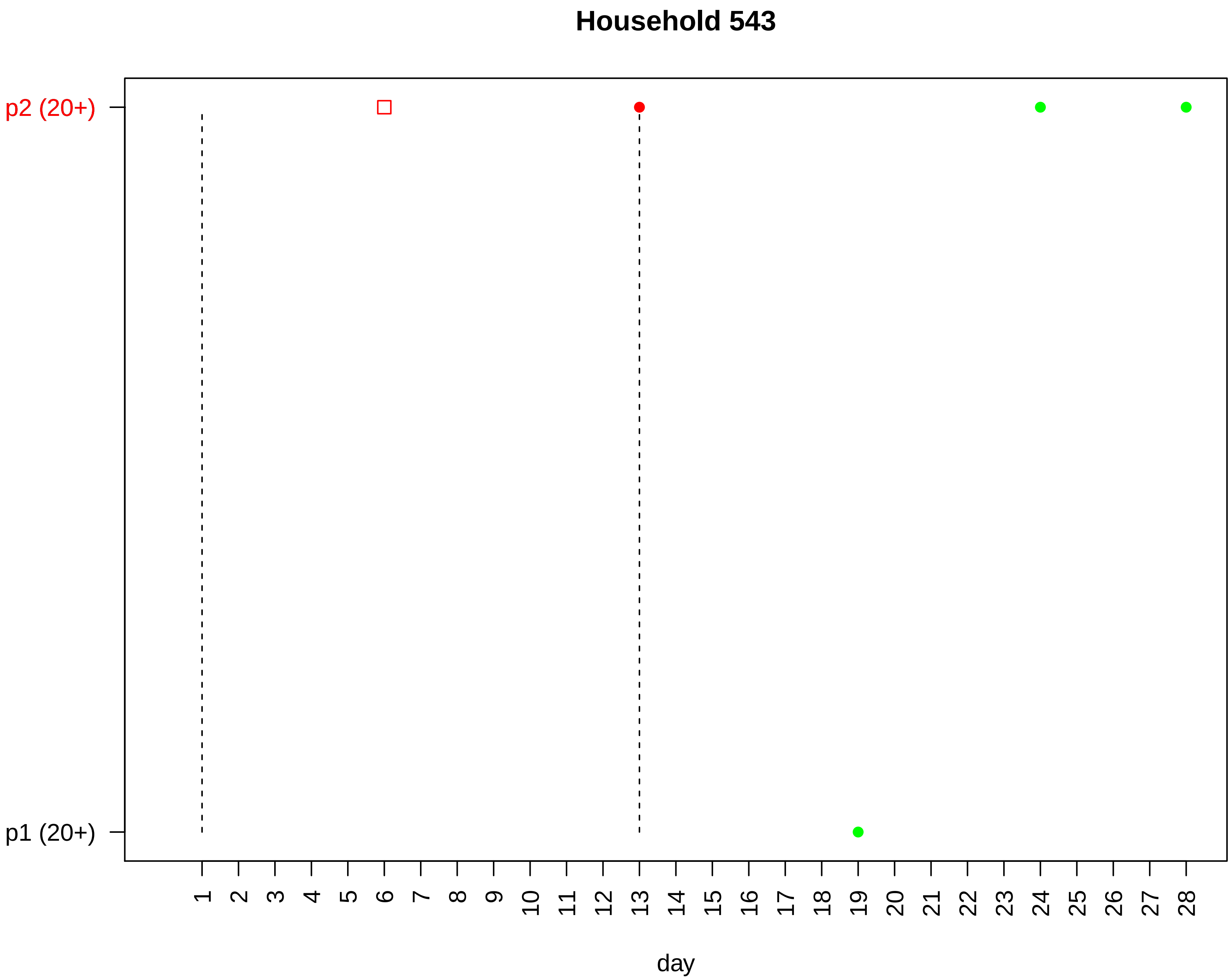
Household 543
p2 (20+)
p1 (20+)
1 2 3 4 5 6 7 8 9 10 11 12 13 14 15 16 17 18 19 20 21 22 23 24 25 26 27 28
day

### Household 544

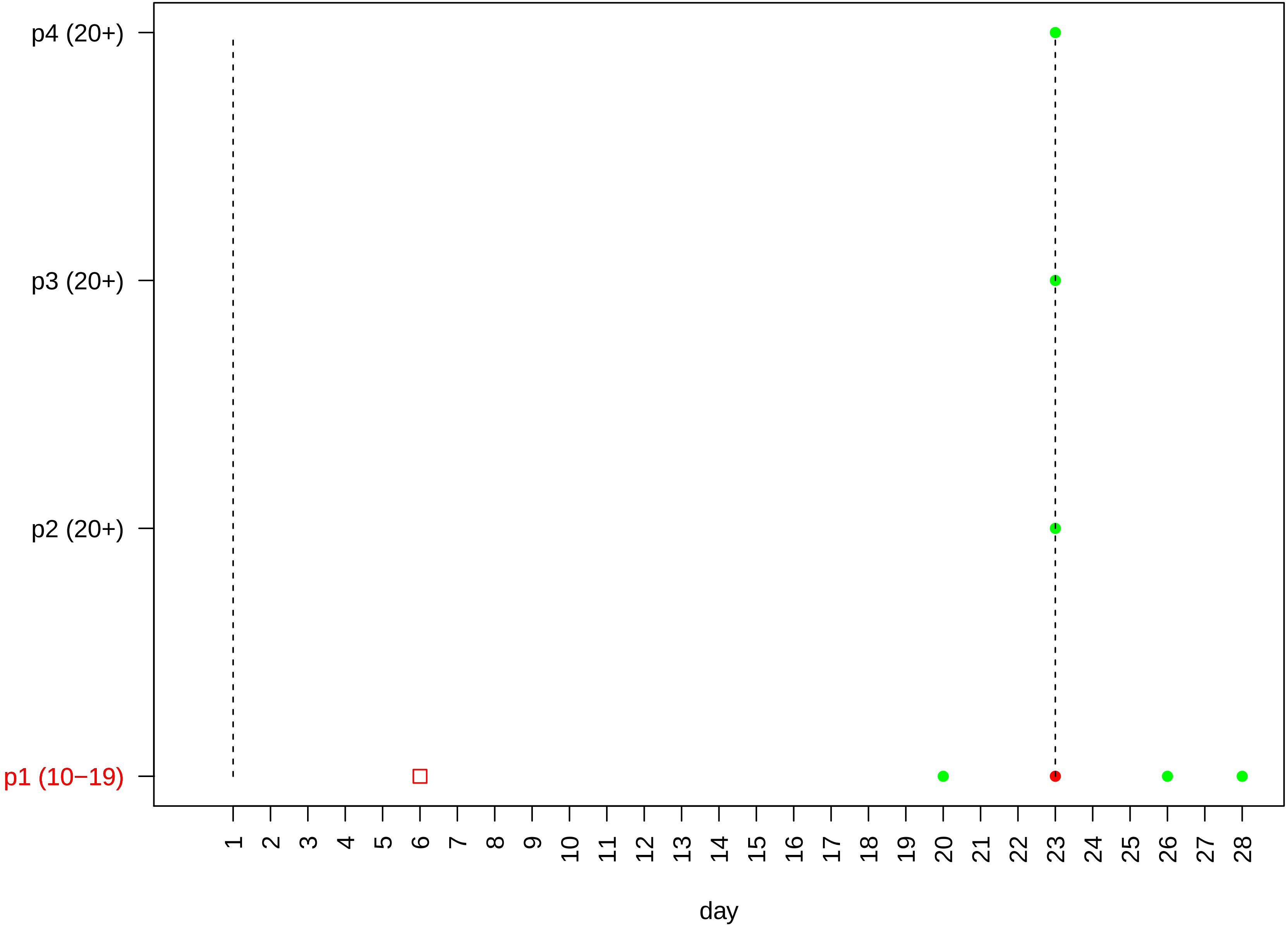

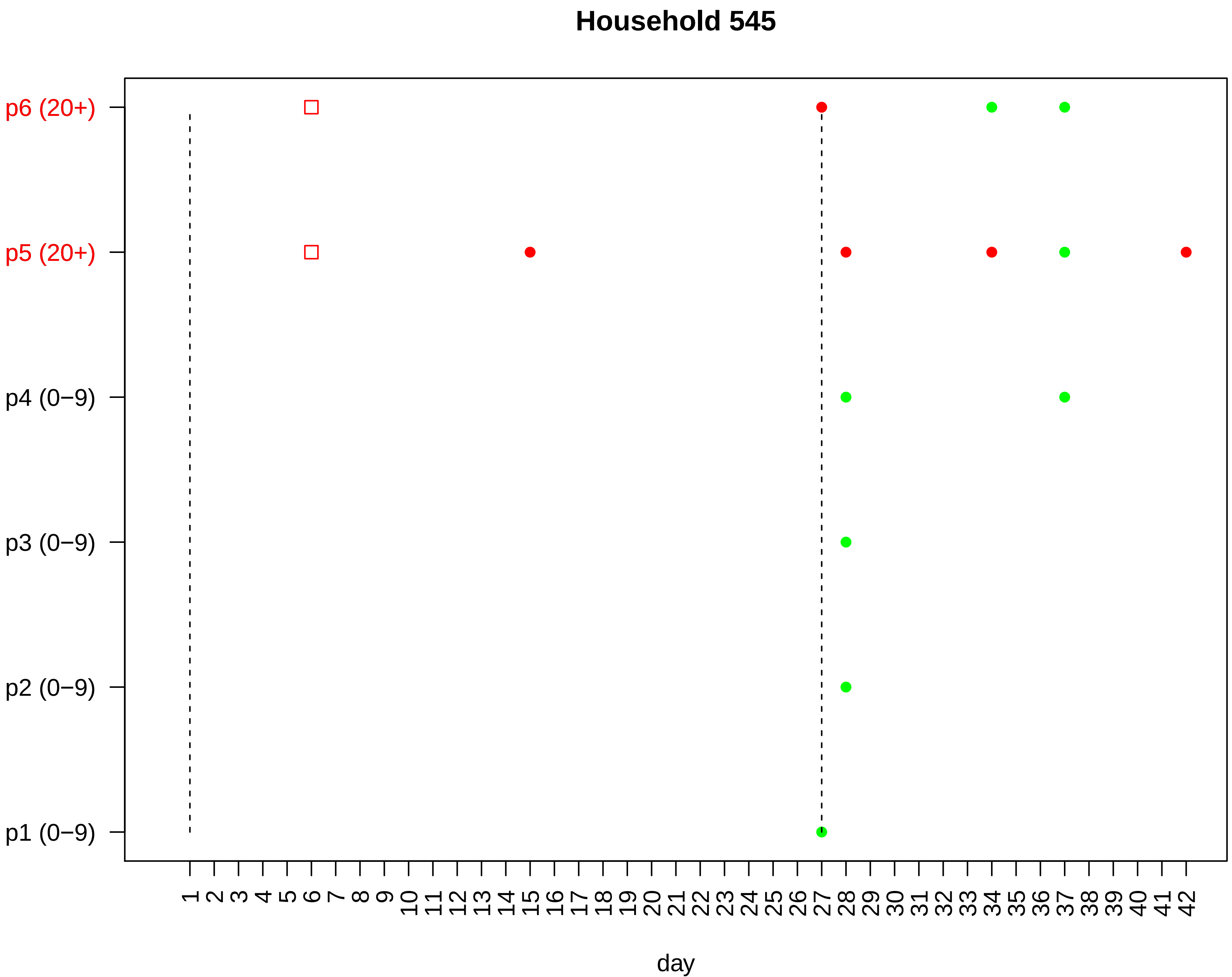

Household 545
p6 (20+)
p5 (20+)
p4 (0−9)
p3 (0−9)
p2 (0−9)
p1 (0−9)
1 2 3 4 5 6 7 8 9 10 11 12 13 14 15 16 17 18 19 20 21 22 23 24 25 26 27 28 29 30 31 32 33 34 35 36 37 38 39 40 41 42
day

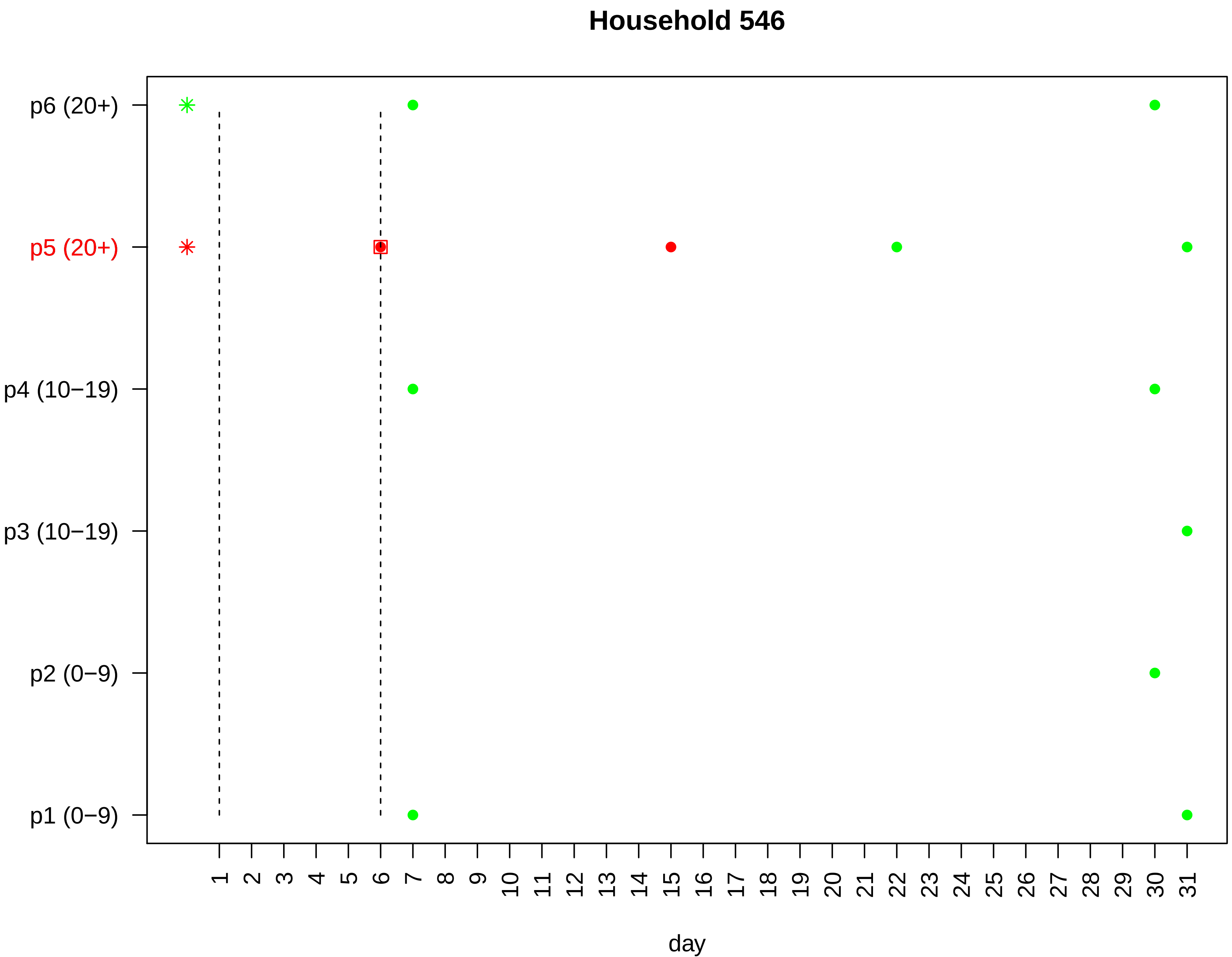

Household 546
p6 (20+)
p5 (20+)
p4 (10−19)
p3 (10−19)
p2 (0−9)
p1 (0−9)
1 2 3 4 5 6 7 8 9 10 11 12 13 14 15 16 17 18 19 20 21 22 23 24 25 26 27 28 29 30 31
day

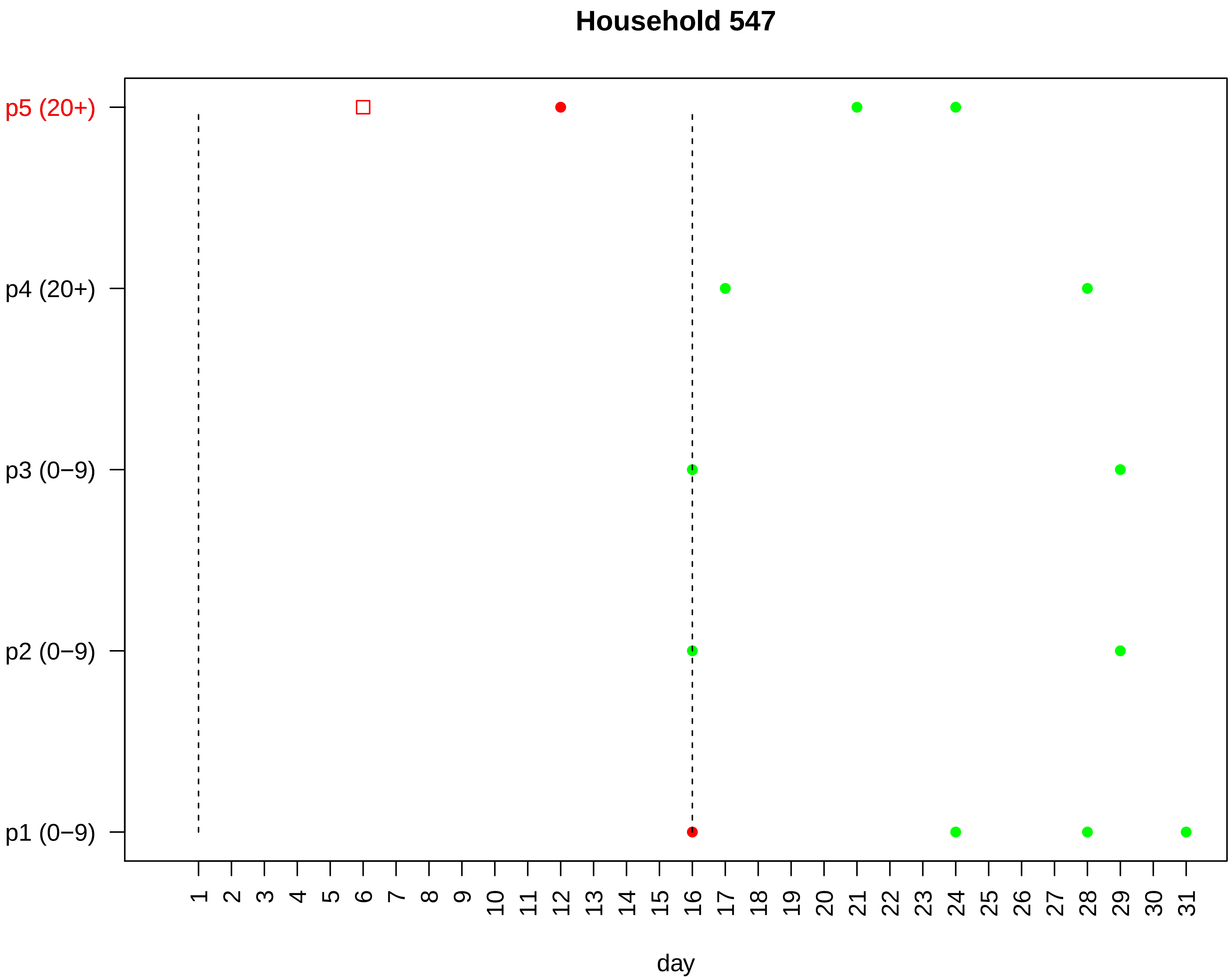

Household 547
p5 (20+)
p4 (20+)
p3 (0−9)
p2 (0−9)
p1 (0−9)
1 2 3 4 5 6 7 8 9 10 11 12 13 14 15 16 17 18 19 20 21 22 23 24 25 26 27 28 29 30 31
day

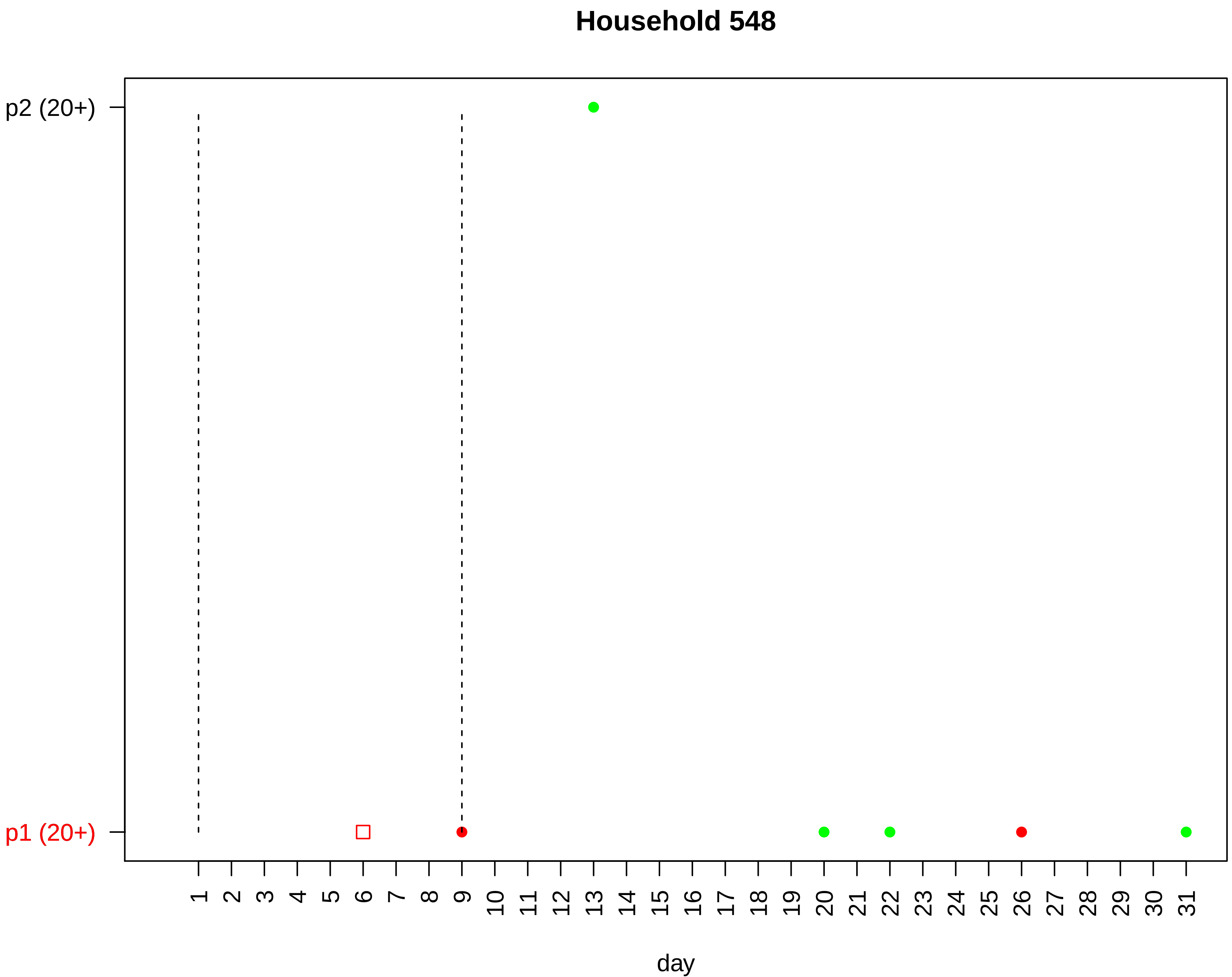
Household 548
p2 (20+)
p1 (20+)
day

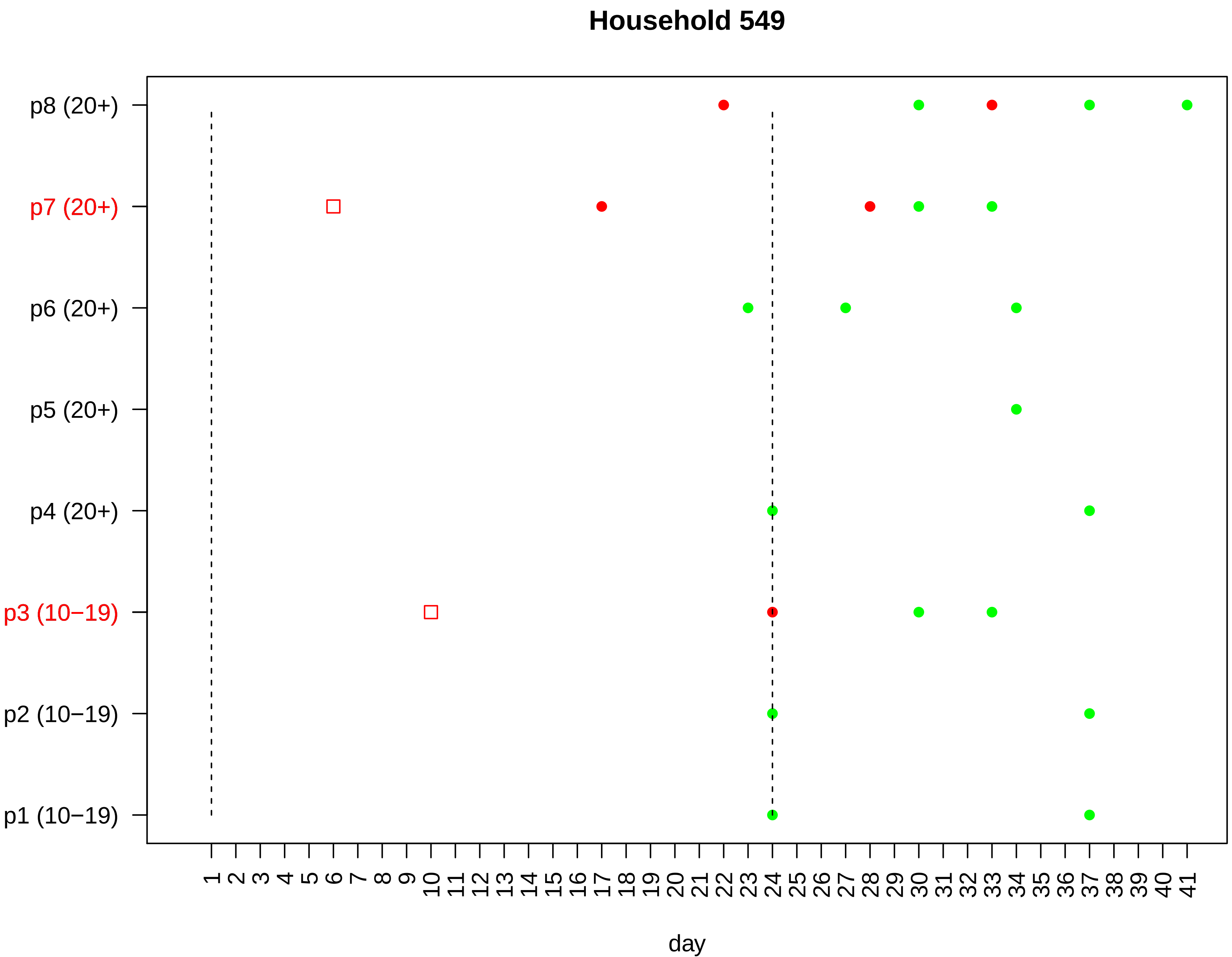

Household 549
p8 (20+)
p7 (20+)
p6 (20+)
p5 (20+)
p4 (20+)
p3 (10−19)
p2 (10−19)
p1 (10−19)
1 2 3 4 5 6 7 8 9 10 11 12 13 14 15 16 17 18 19 20 21 22 23 24 25 26 27 28 29 30 31 32 33 34 35 36 37 38 39 40 41
day

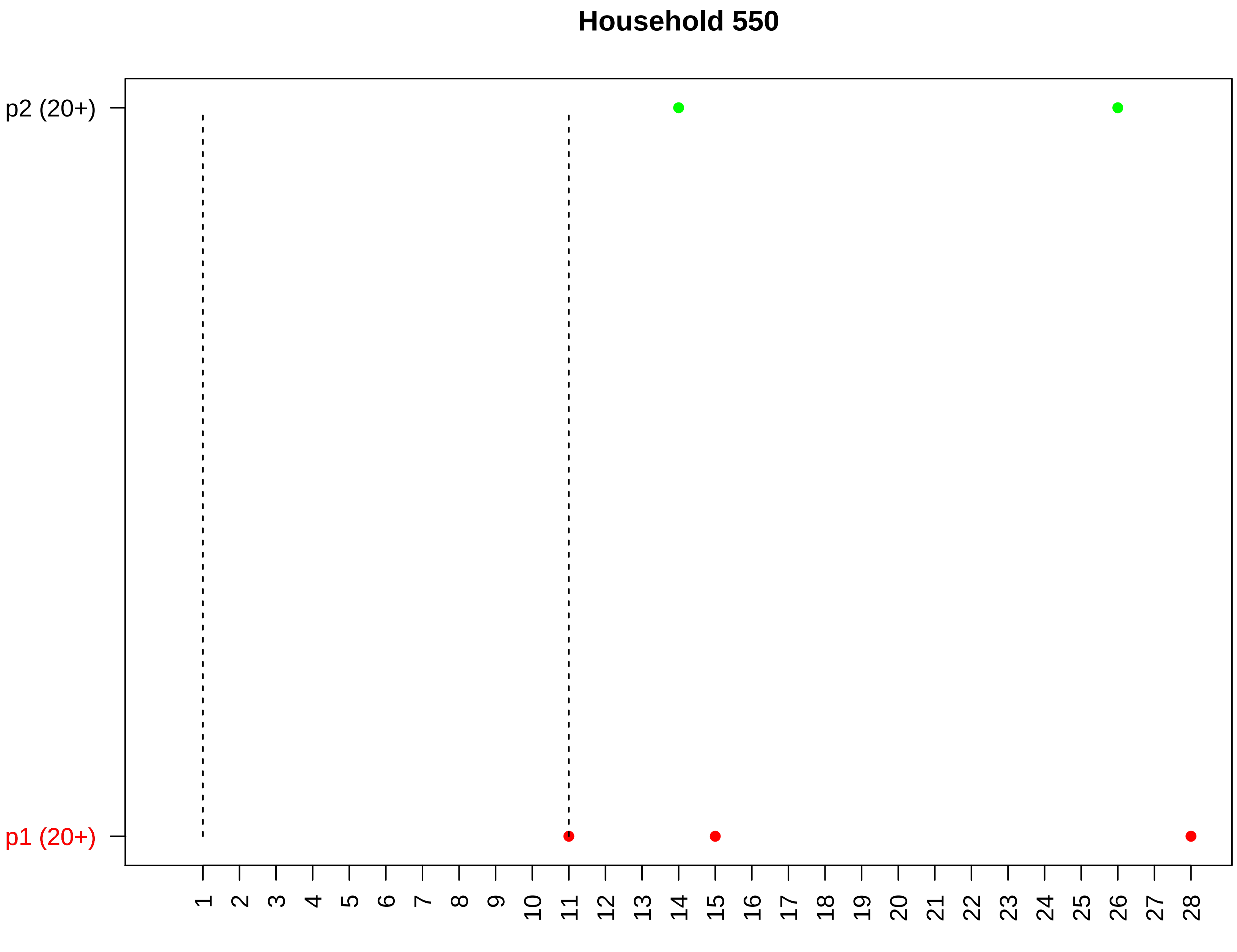

Household 550
p2 (20+)
p1 (20+)
day

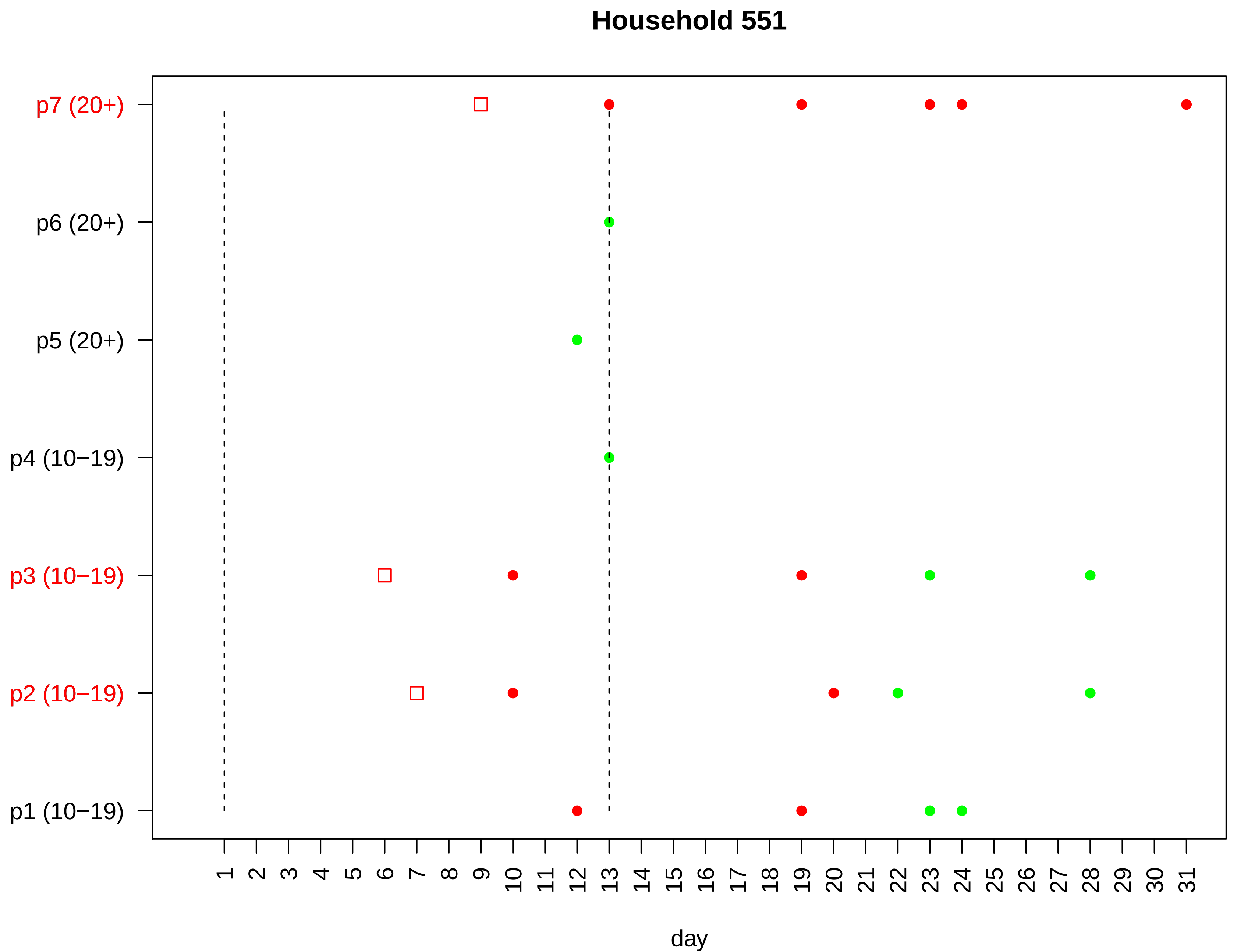

Household 551
p7 (20+)
p6 (20+)
p5 (20+)
p4 (10−19)
p3 (10−19)
p2 (10−19)
p1 (10−19)
1 2 3 4 5 6 7 8 9 10 11 12 13 14 15 16 17 18 19 20 21 22 23 24 25 26 27 28 29 30 31
day

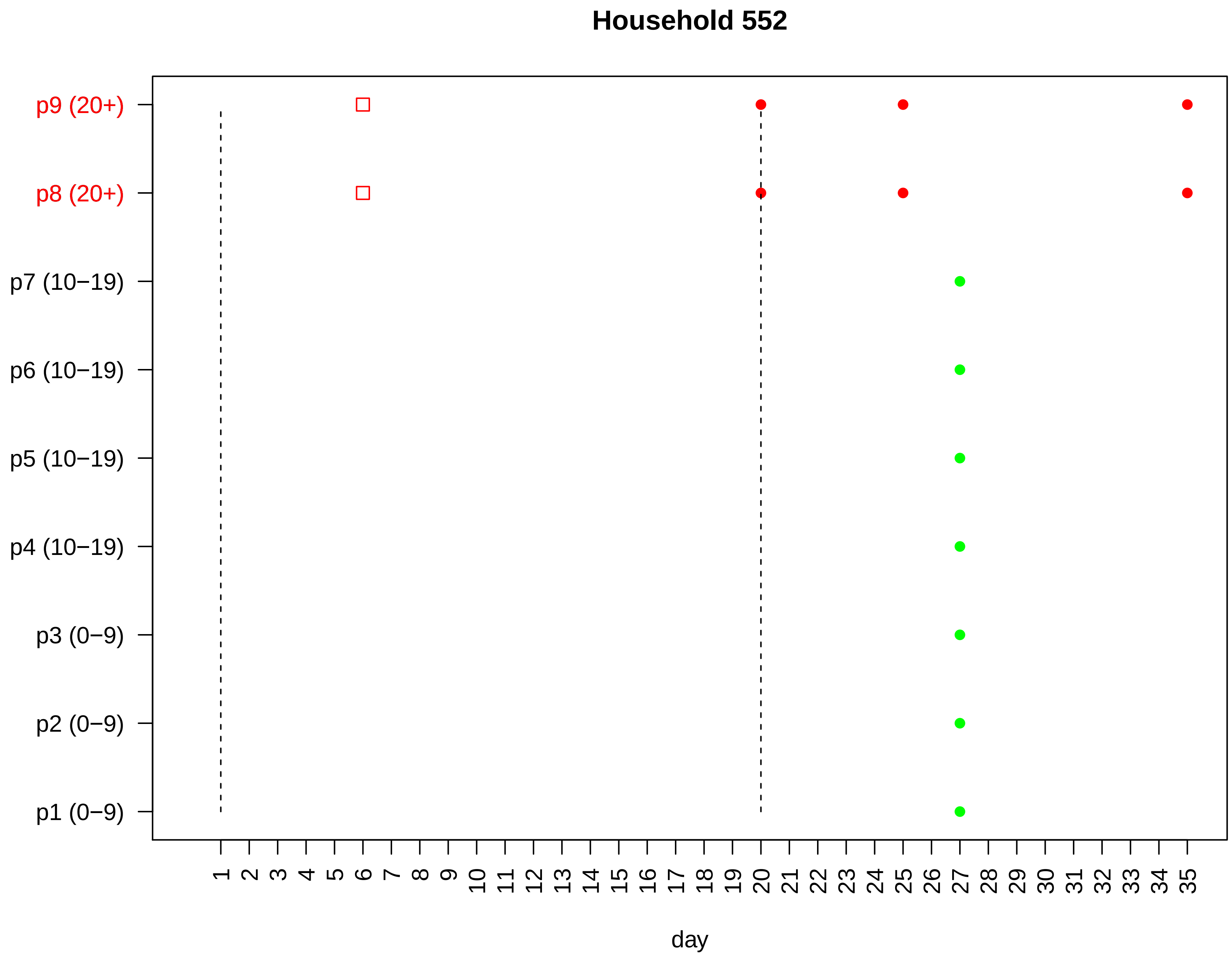

Household 552
p9 (20+)
p8 (20+)
p7 (10−19)
p6 (10−19)
p5 (10−19)
p4 (10−19)
p3 (0−9)
p2 (0−9)
p1 (0−9)
1 2 3 4 5 6 7 8 9 10 11 12 13 14 15 16 17 18 19 20 21 22 23 24 25 26 27 28 29 30 31 32 33 34 35
day

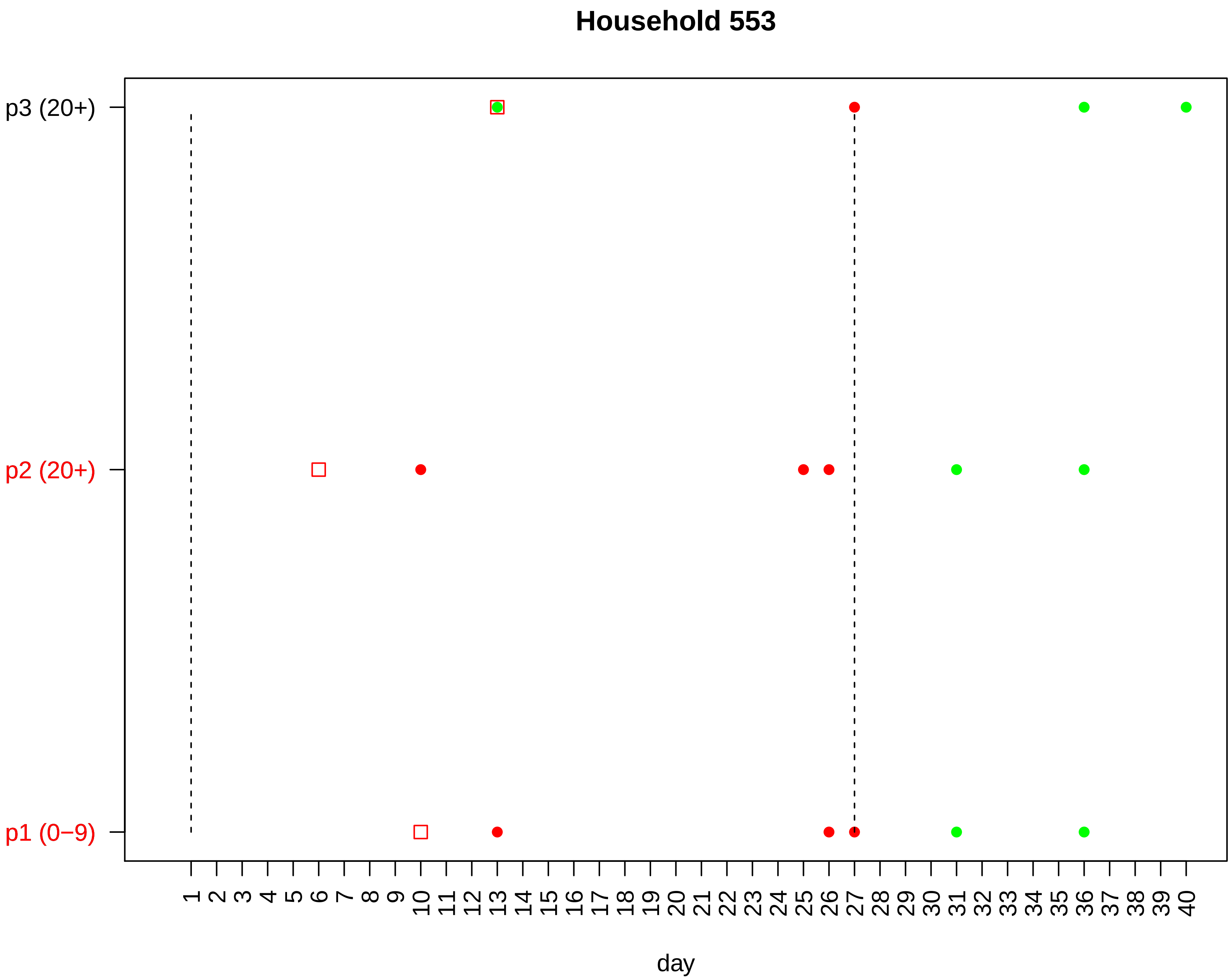

Household 553
p3 (20+)
p2 (20+)
p1 (0−9)
1 2 3 4 5 6 7 8 9 10 11 12 13 14 15 16 17 18 19 20 21 22 23 24 25 26 27 28 29 30 31 32 33 34 35 36 37 38 39 40
day

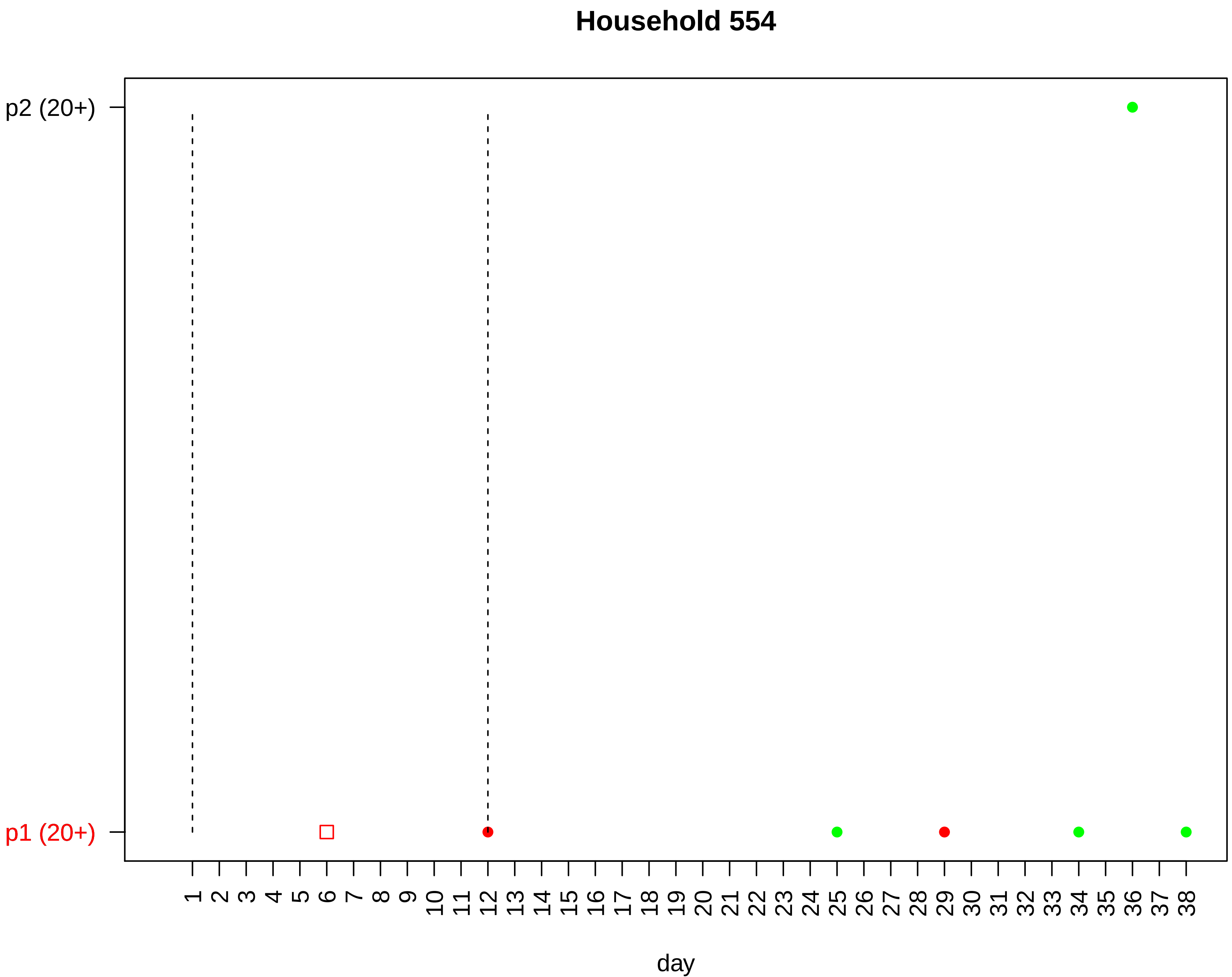
Household 554
p2 (20+)
p1 (20+)
1 2 3 4 5 6 7 8 9 10 11 12 13 14 15 16 17 18 19 20 21 22 23 24 25 26 27 28 29 30 31 32 33 34 35 36 37 38
day

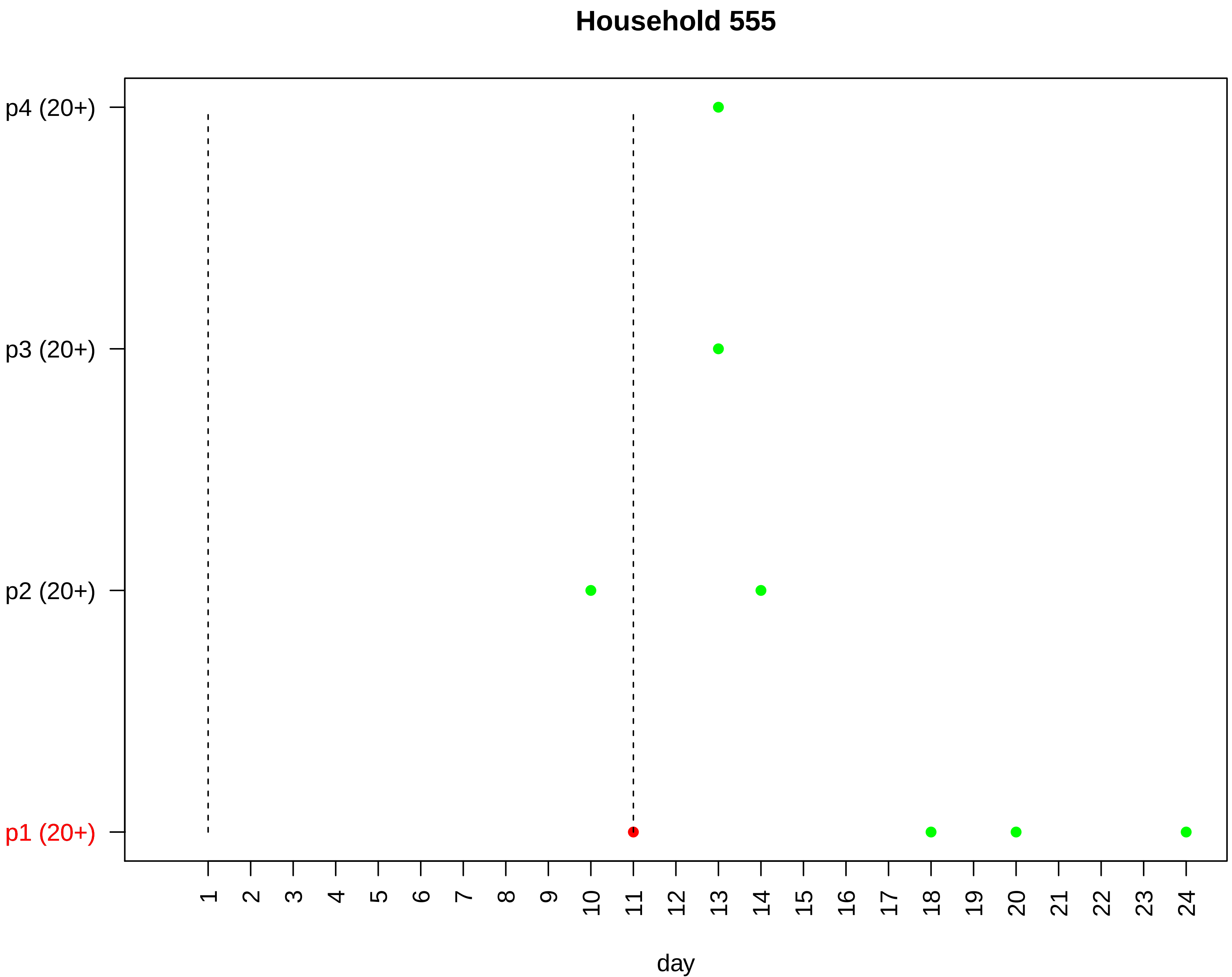
Household 555
p4 (20+)
p3 (20+)
p2 (20+)
p1 (20+)
day

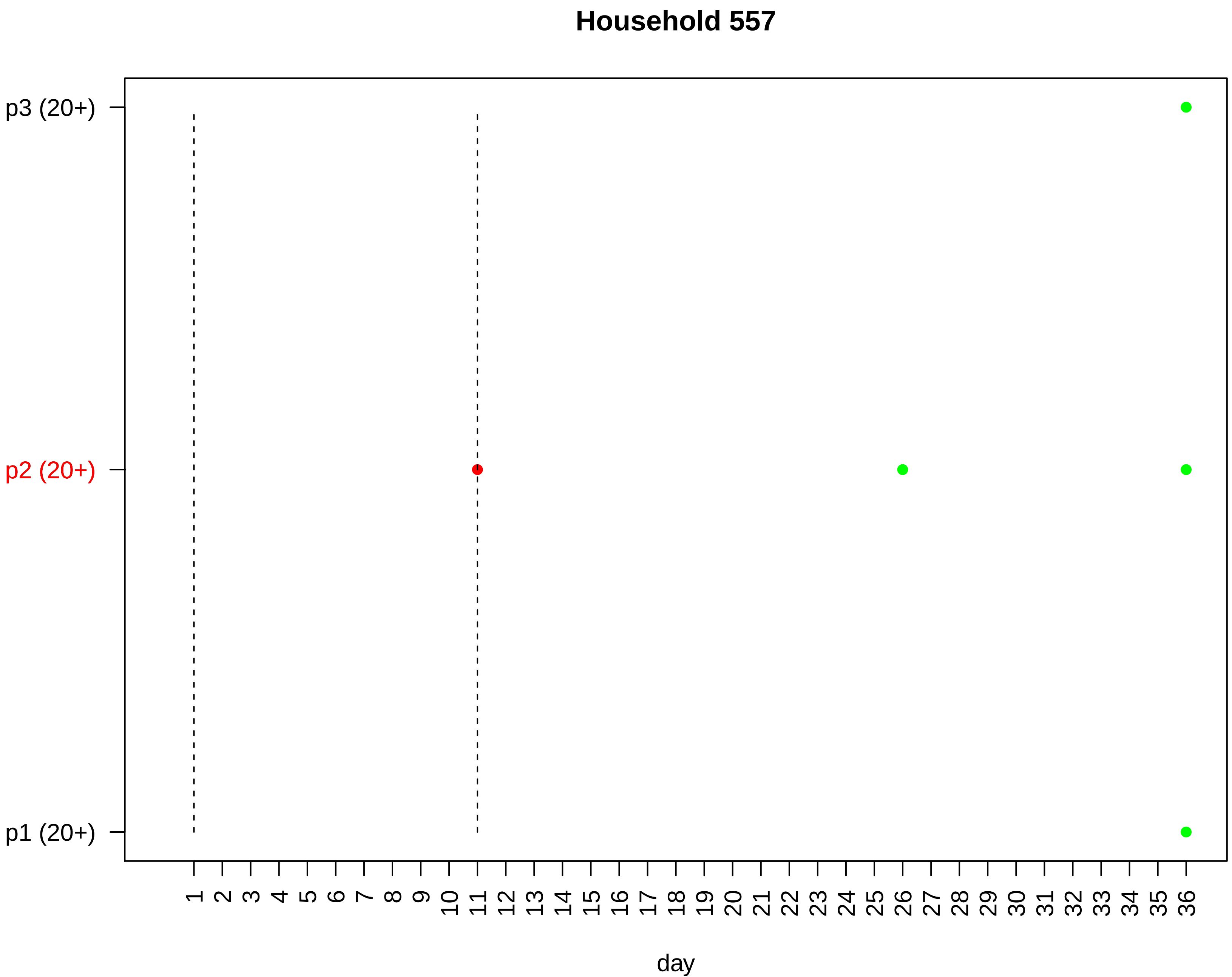
Household 557
p3 (20+)
p2 (20+)
p1 (20+)
1 2 3 4 5 6 7 8 9 10 11 12 13 14 15 16 17 18 19 20 21 22 23 24 25 26 27 28 29 30 31 32 33 34 35 36
day

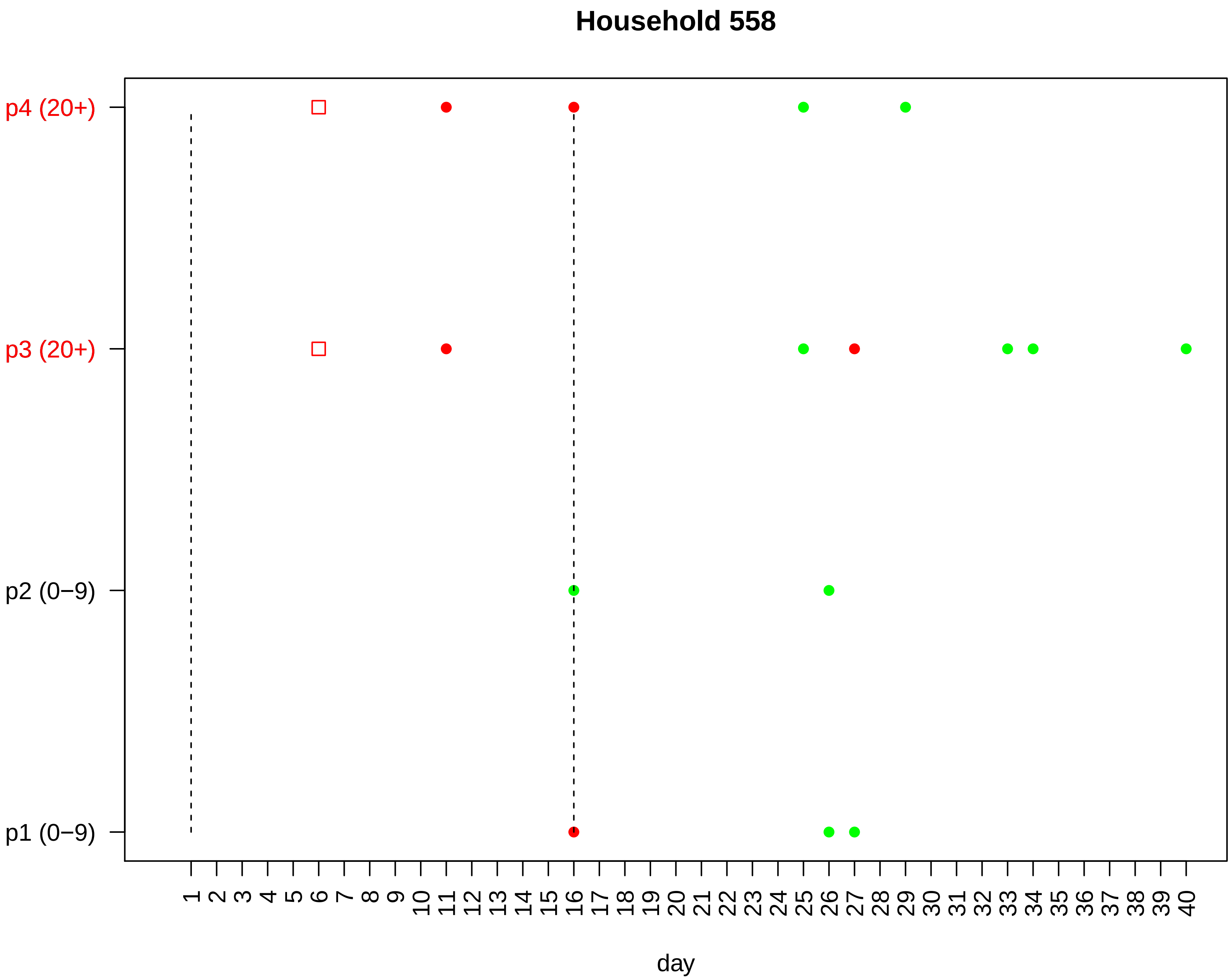
Household 558
p4 (20+)
p3 (20+)
p2 (0−9)
p1 (0−9)
1 2 3 4 5 6 7 8 9 10 11 12 13 14 15 16 17 18 19 20 21 22 23 24 25 26 27 28 29 30 31 32 33 34 35 36 37 38 39 40
day

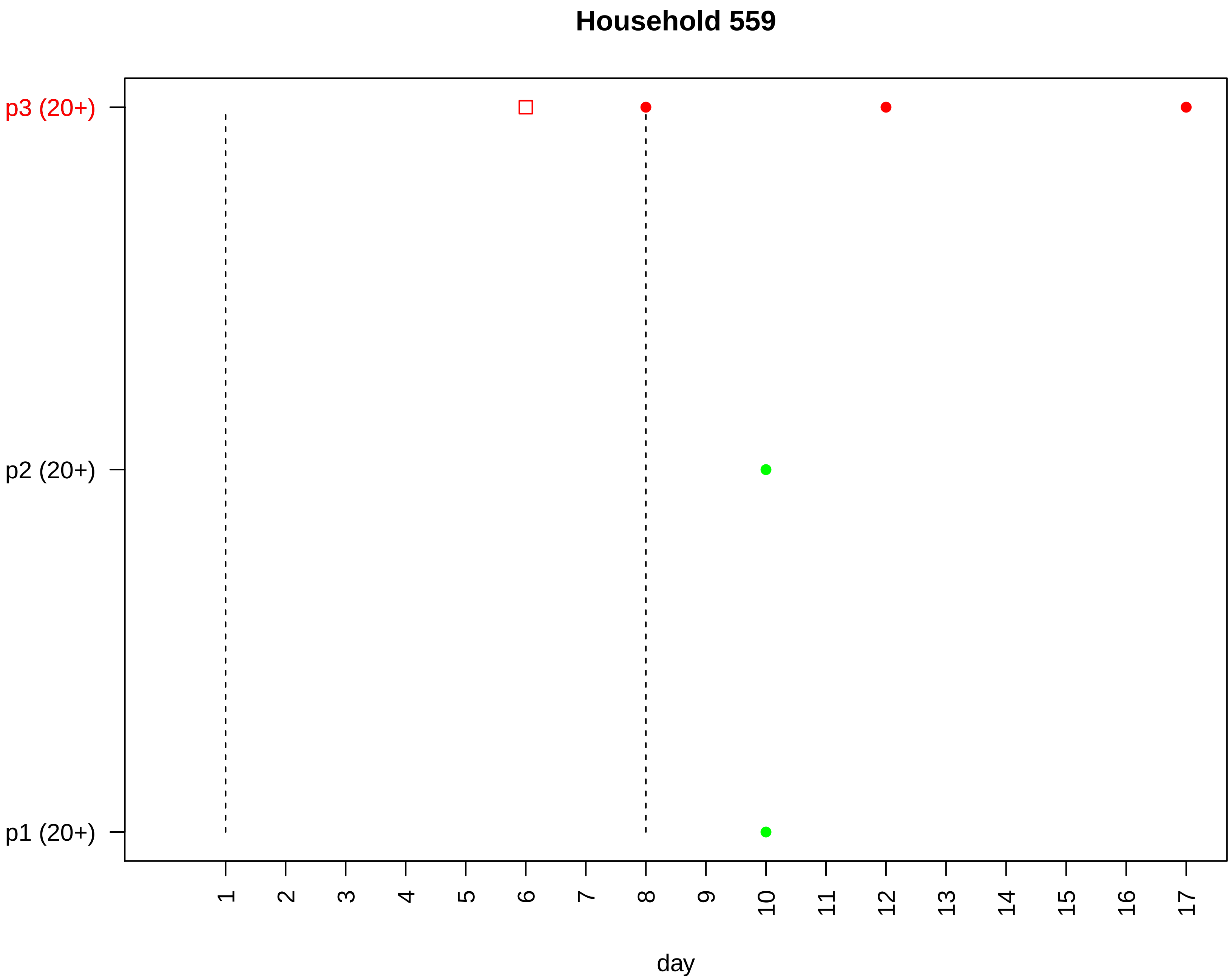

Household 559
p3 (20+)
p2 (20+)
p1 (20+)
day

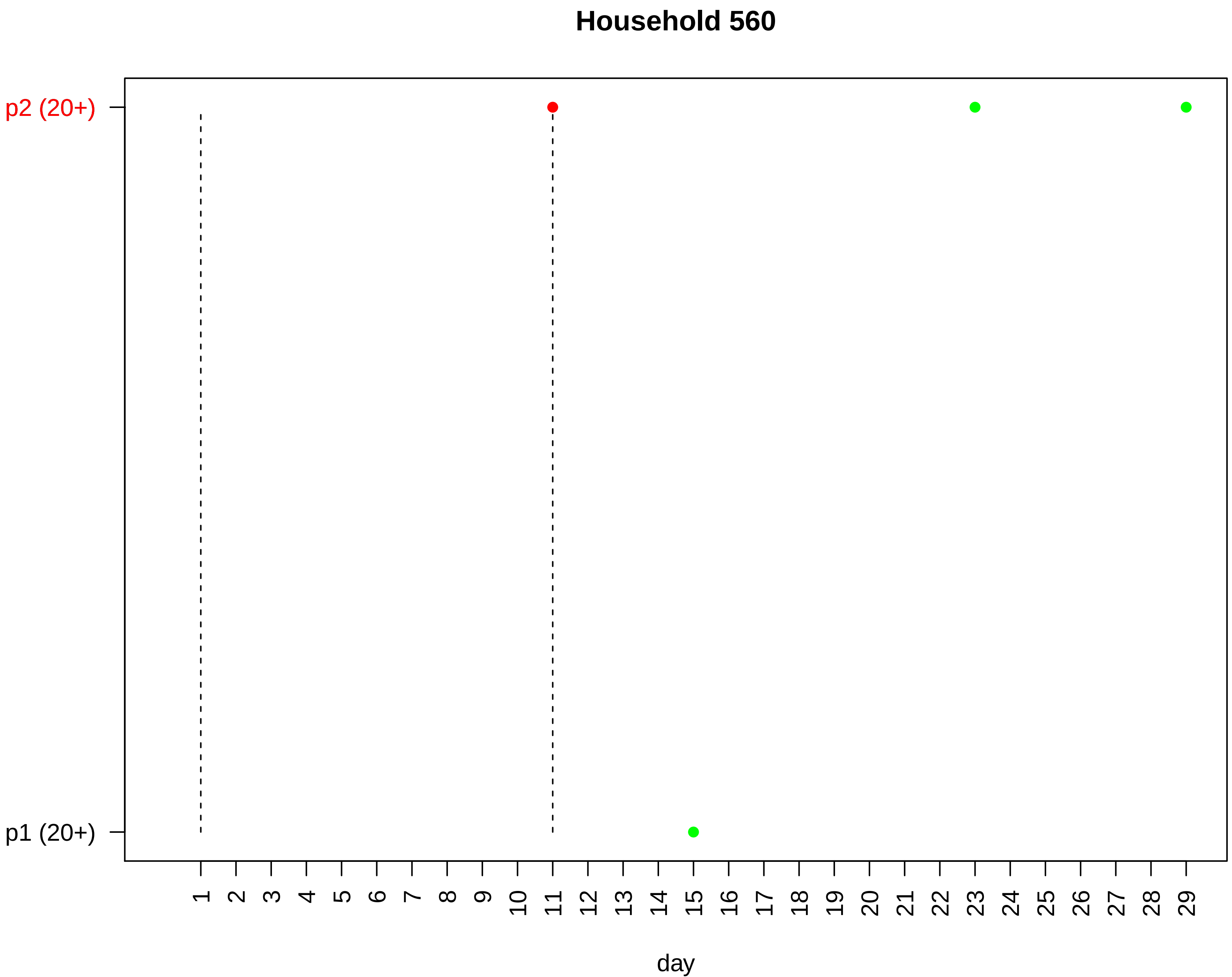

Household 560
p2 (20+)
p1 (20+)
day

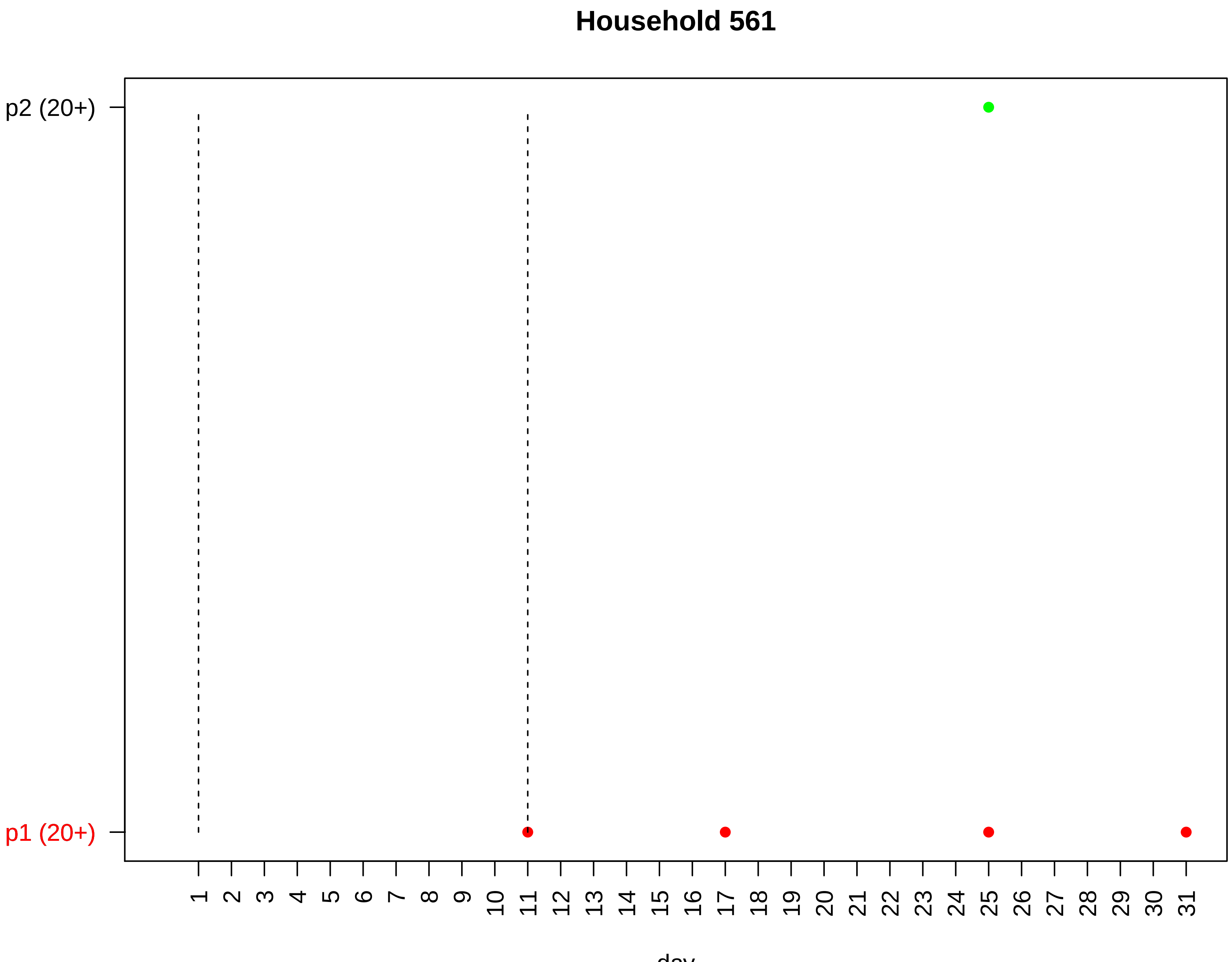
Household 561
p2 (20+)
p1 (20+)
day

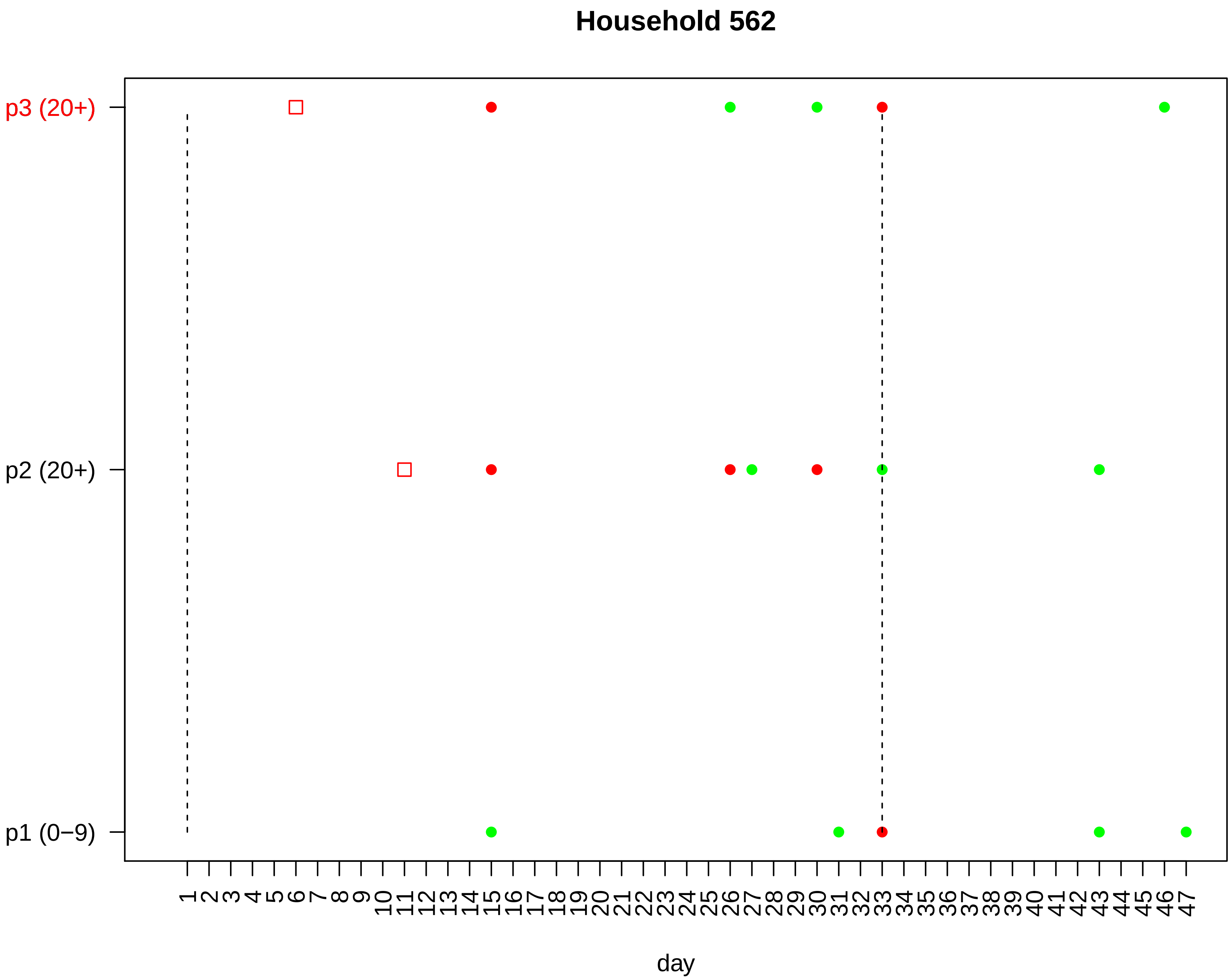
Household 562
p3 (20+)
p2 (20+)
p1 (0−9)
1 2 3 4 5 6 7 8 9 10 11 12 13 14 15 16 17 18 19 20 21 22 23 24 25 26 27 28 29 30 31 32 33 34 35 36 37 38 39 40 41 42 43 44 45 46 47
day

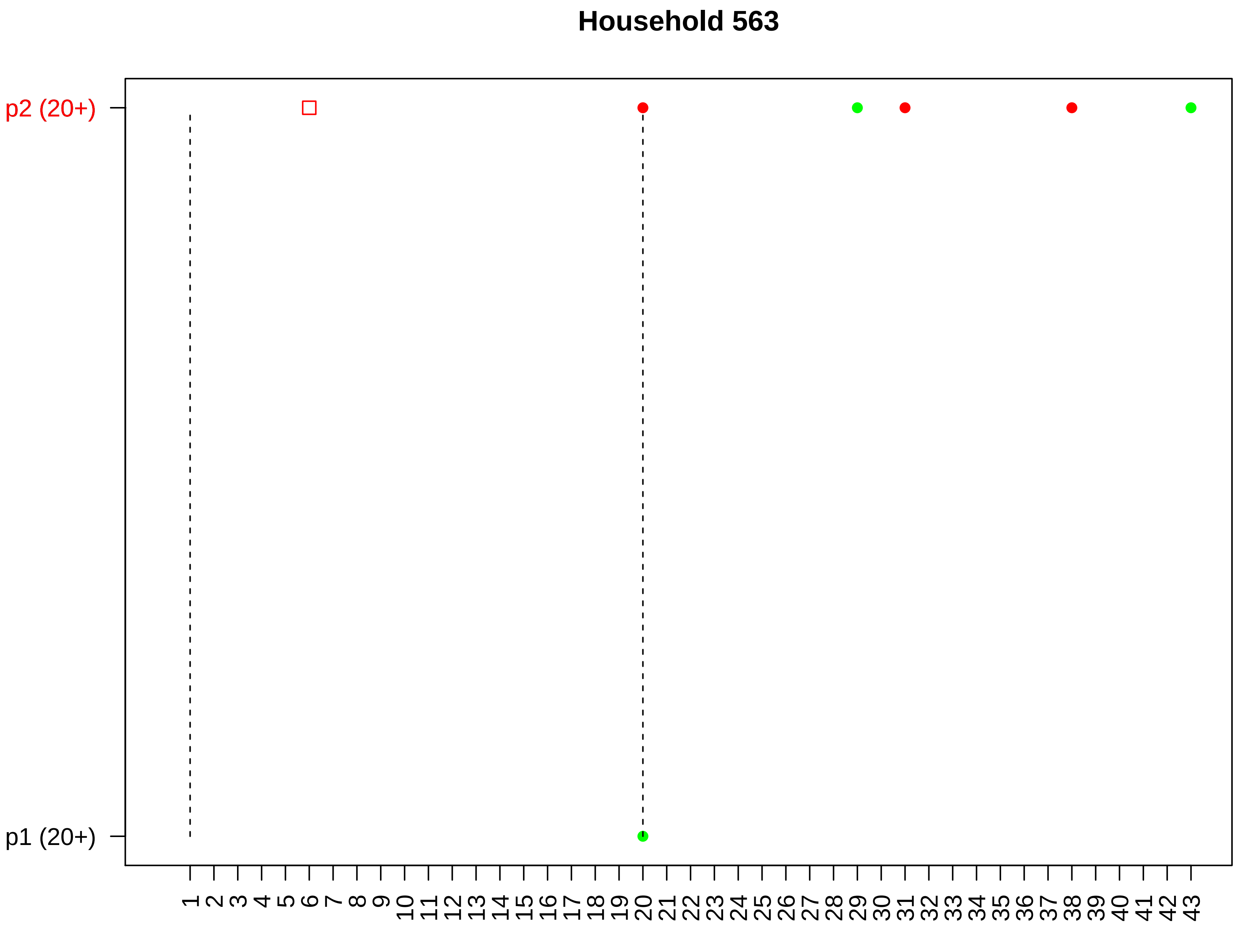

Household 563
p2 (20+)
p1 (20+)
day

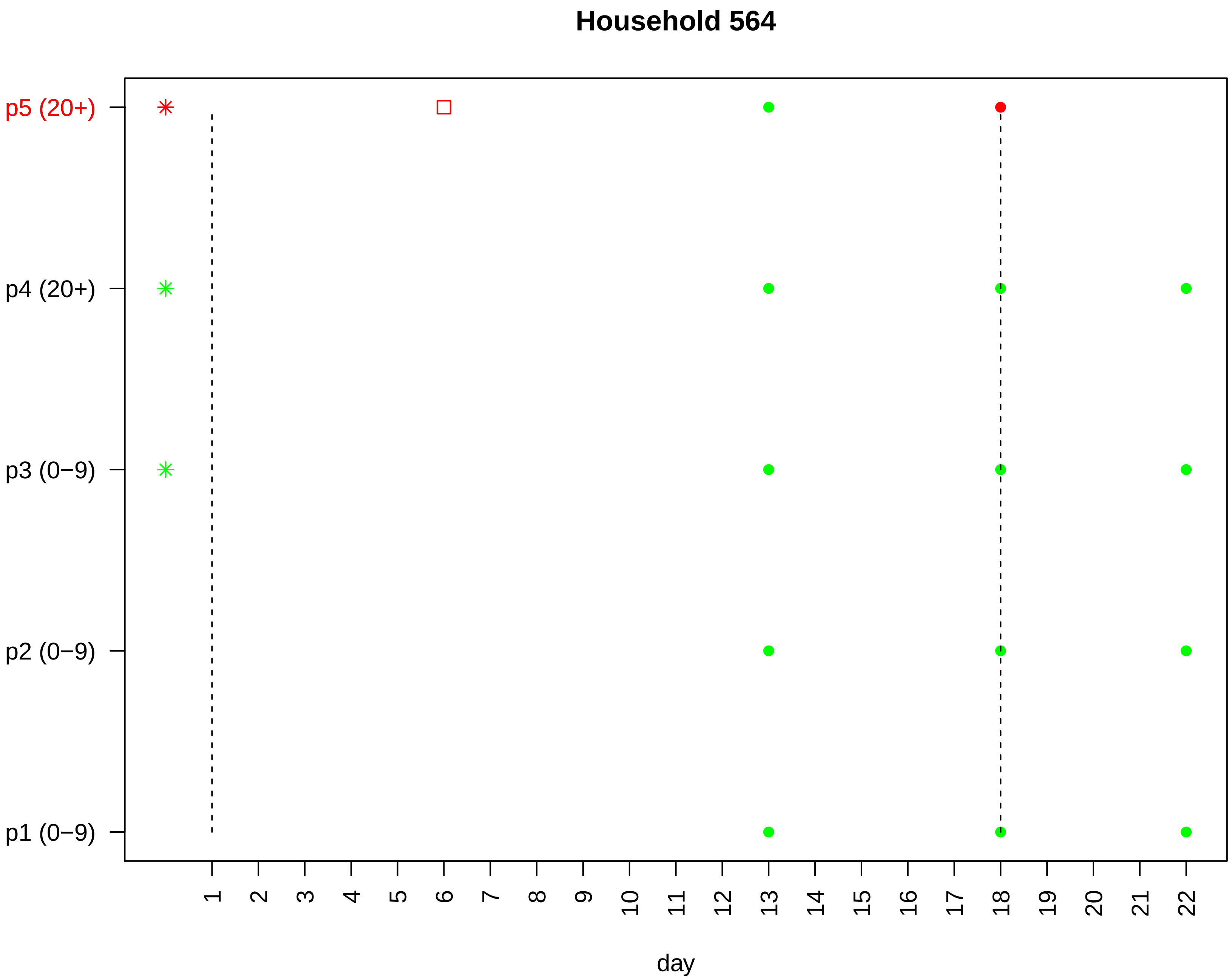

Household 564
p5 (20+)
p4 (20+)
p3 (0−9)
p2 (0−9)
p1 (0−9)
day

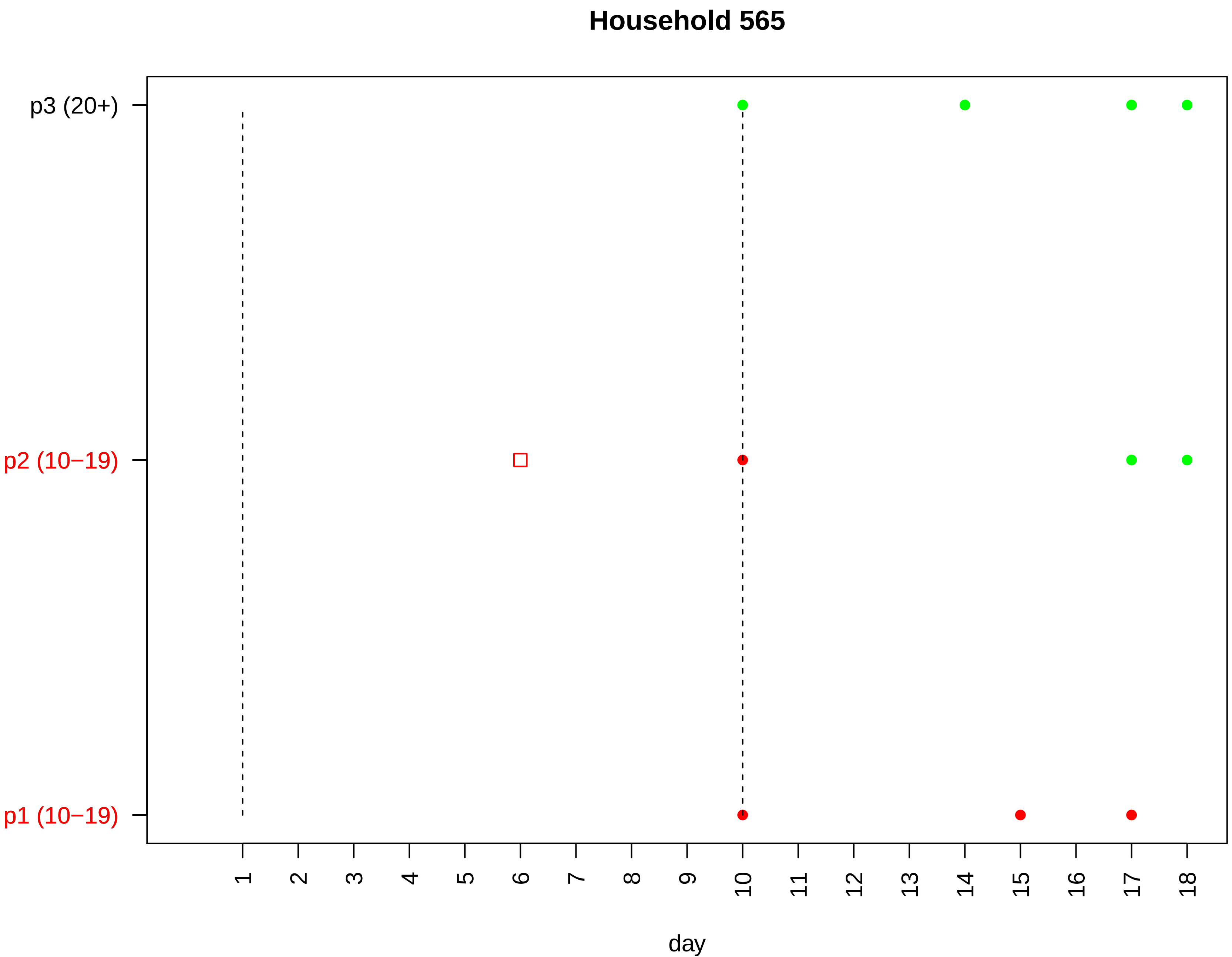

Household 565
p3 (20+)
p2 (10−19)
p1 (10−19)
day

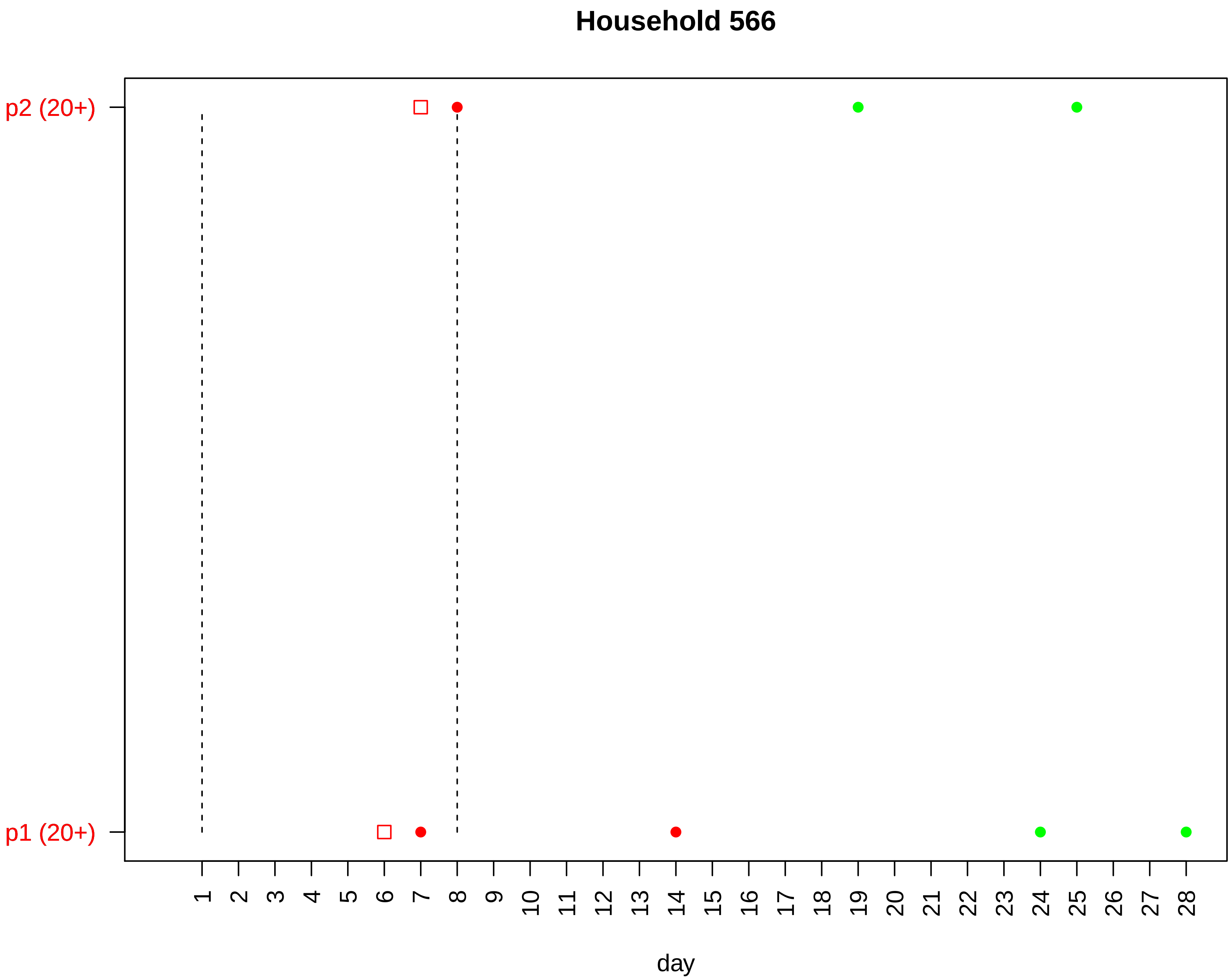

Household 566
p2 (20+)
p1 (20+)
day

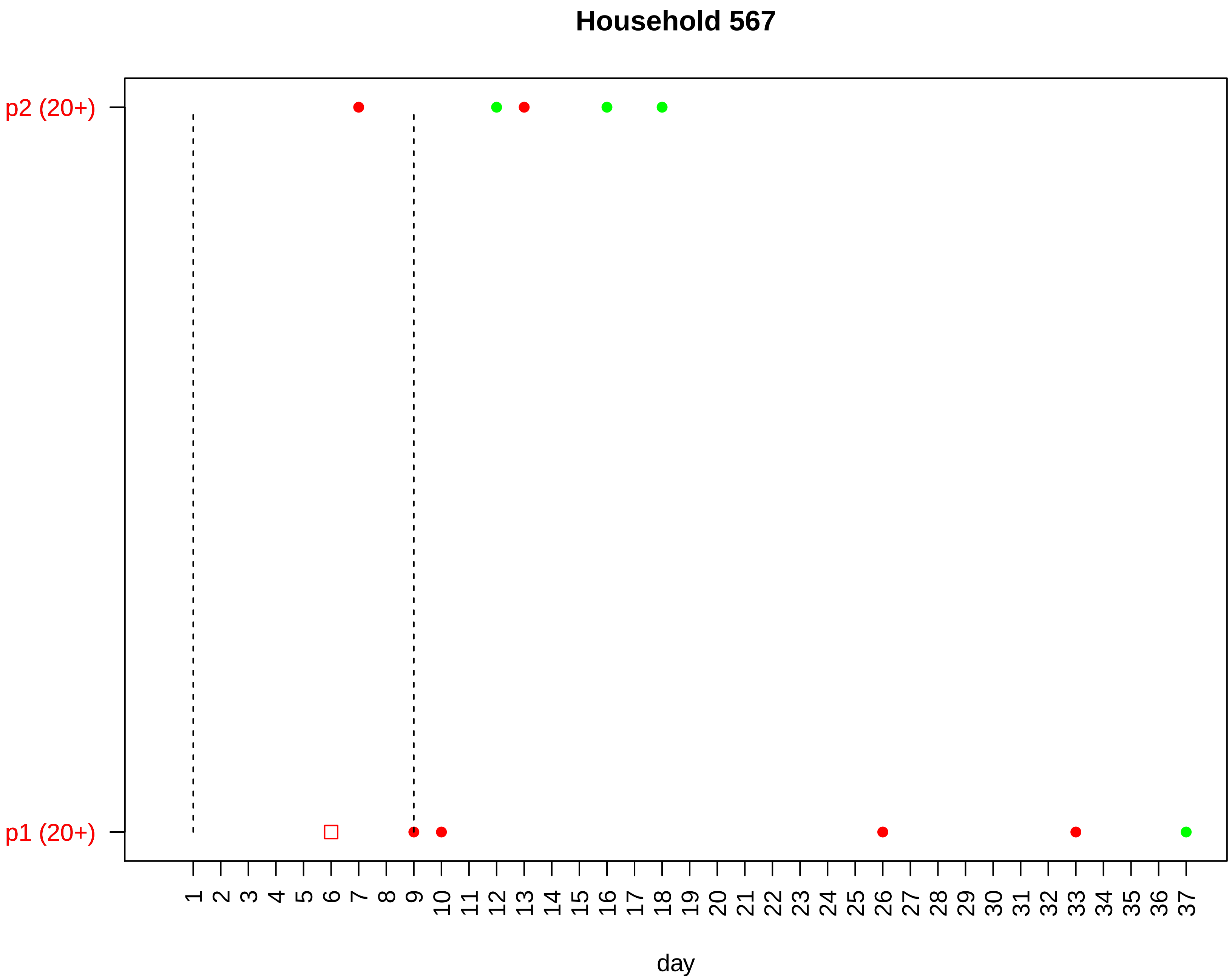

Household 567
p2 (20+)
p1 (20+)
1 2 3 4 5 6 7 8 9 10 11 12 13 14 15 16 17 18 19 20 21 22 23 24 25 26 27 28 29 30 31 32 33 34 35 36 37
day

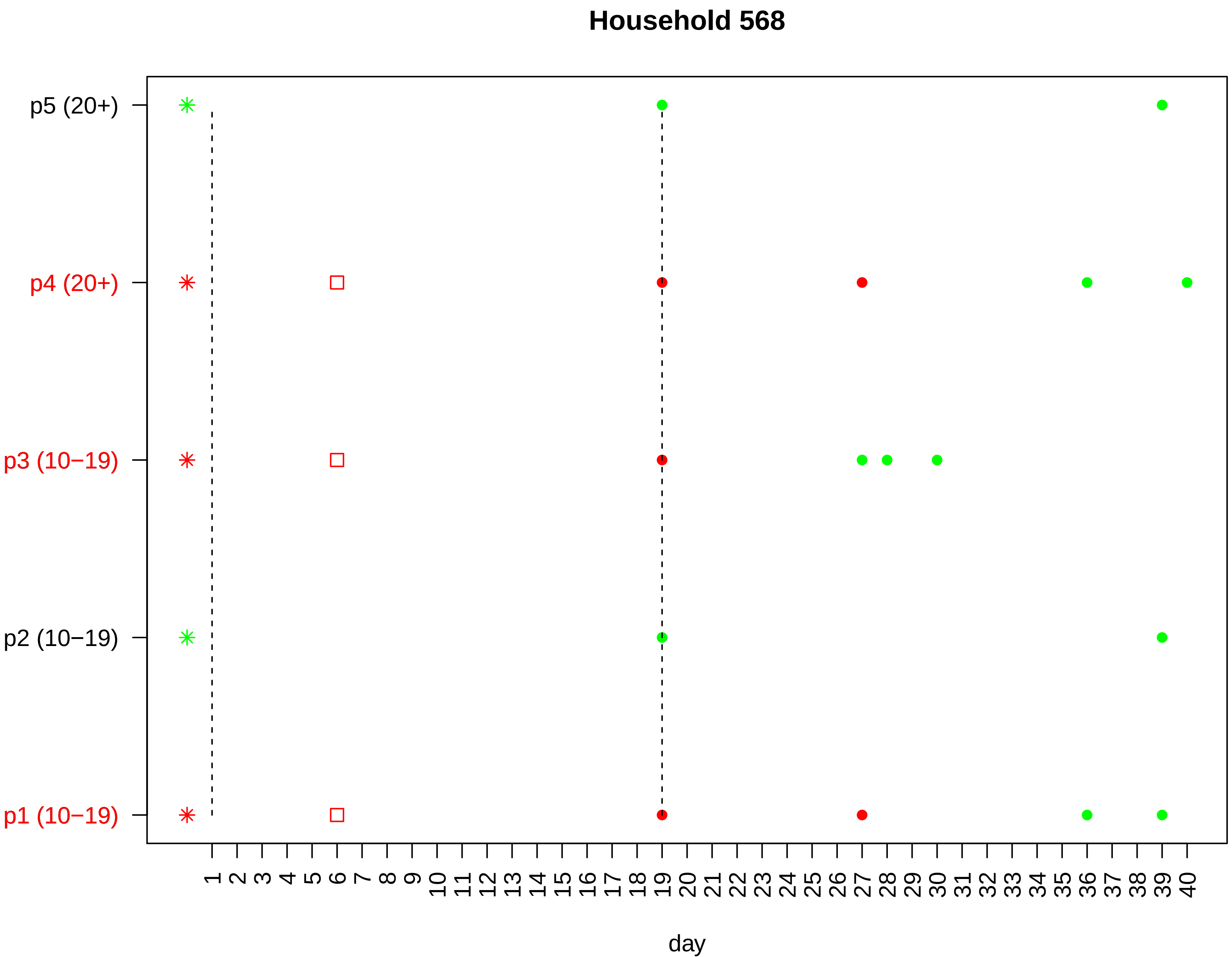

Household 568
p5 (20+)
p4 (20+)
p3 (10−19)
p2 (10−19)
p1 (10−19)
1 2 3 4 5 6 7 8 9 10 11 12 13 14 15 16 17 18 19 20 21 22 23 24 25 26 27 28 29 30 31 32 33 34 35 36 37 38 39 40
day

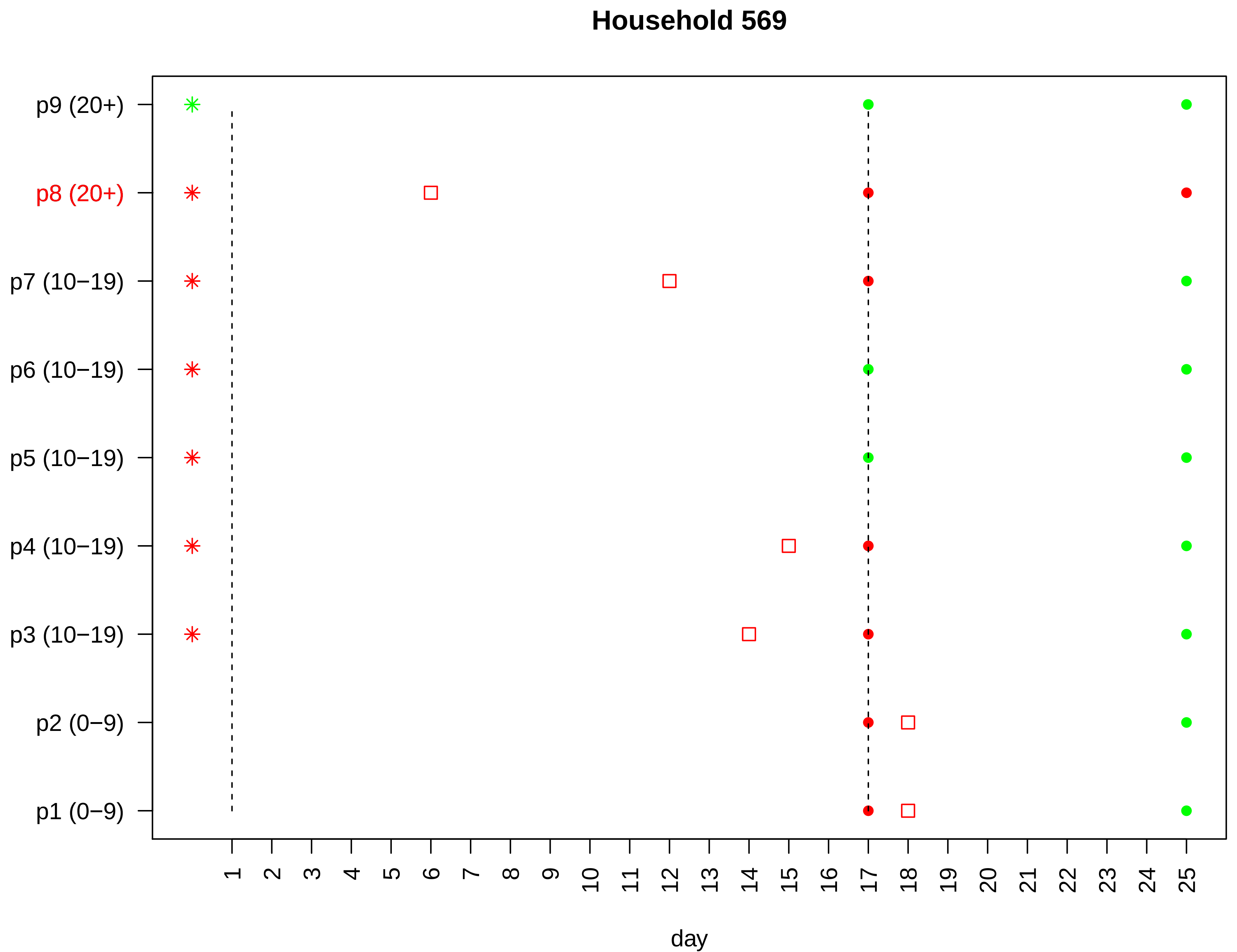

Household 569
p9 (20+)
p8 (20+)
p7 (10−19)
p6 (10−19)
p5 (10−19)
p4 (10−19)
p3 (10−19)
p2 (0−9)
p1 (0−9)
1 2 3 4 5 6 7 8 9 10 11 12 13 14 15 16 17 18 19 20 21 22 23 24 25
day

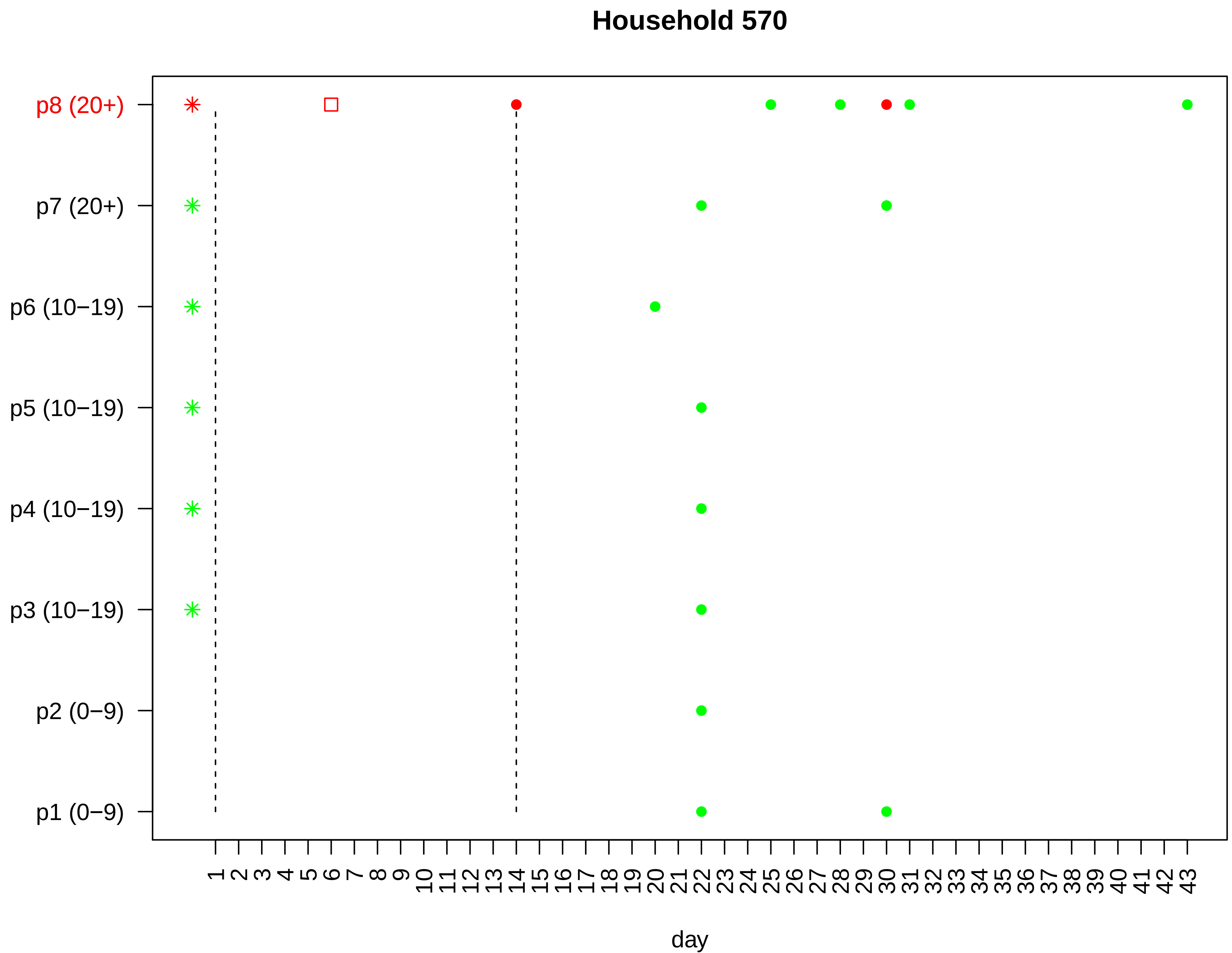
Household 570
p8 (20+)
p7 (20+)
p6 (10−19)
p5 (10−19)
p4 (10−19)
p3 (10−19)
p2 (0−9)
p1 (0−9)
1 2 3 4 5 6 7 8 9 10 11 12 13 14 15 16 17 18 19 20 21 22 23 24 25 26 27 28 29 30 31 32 33 34 35 36 37 38 39 40 41 42 43
day

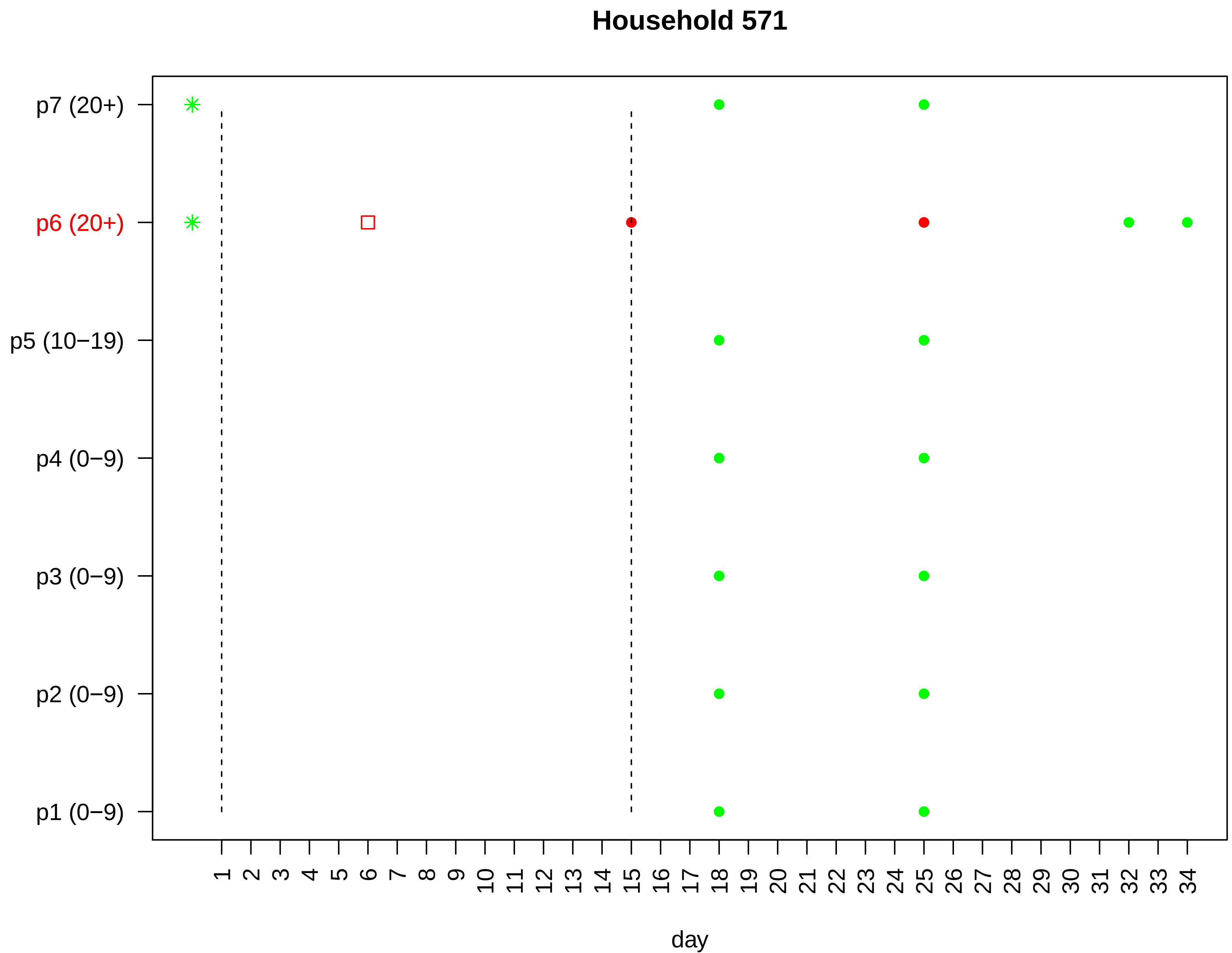
Household 571
p7 (20+)
p6 (20+)
p5 (10−19)
p4 (0−9)
p3 (0−9)
p2 (0−9)
p1 (0−9)
1 2 3 4 5 6 7 8 9 10 11 12 13 14 15 16 17 18 19 20 21 22 23 24 25 26 27 28 29 30 31 32 33 34
day

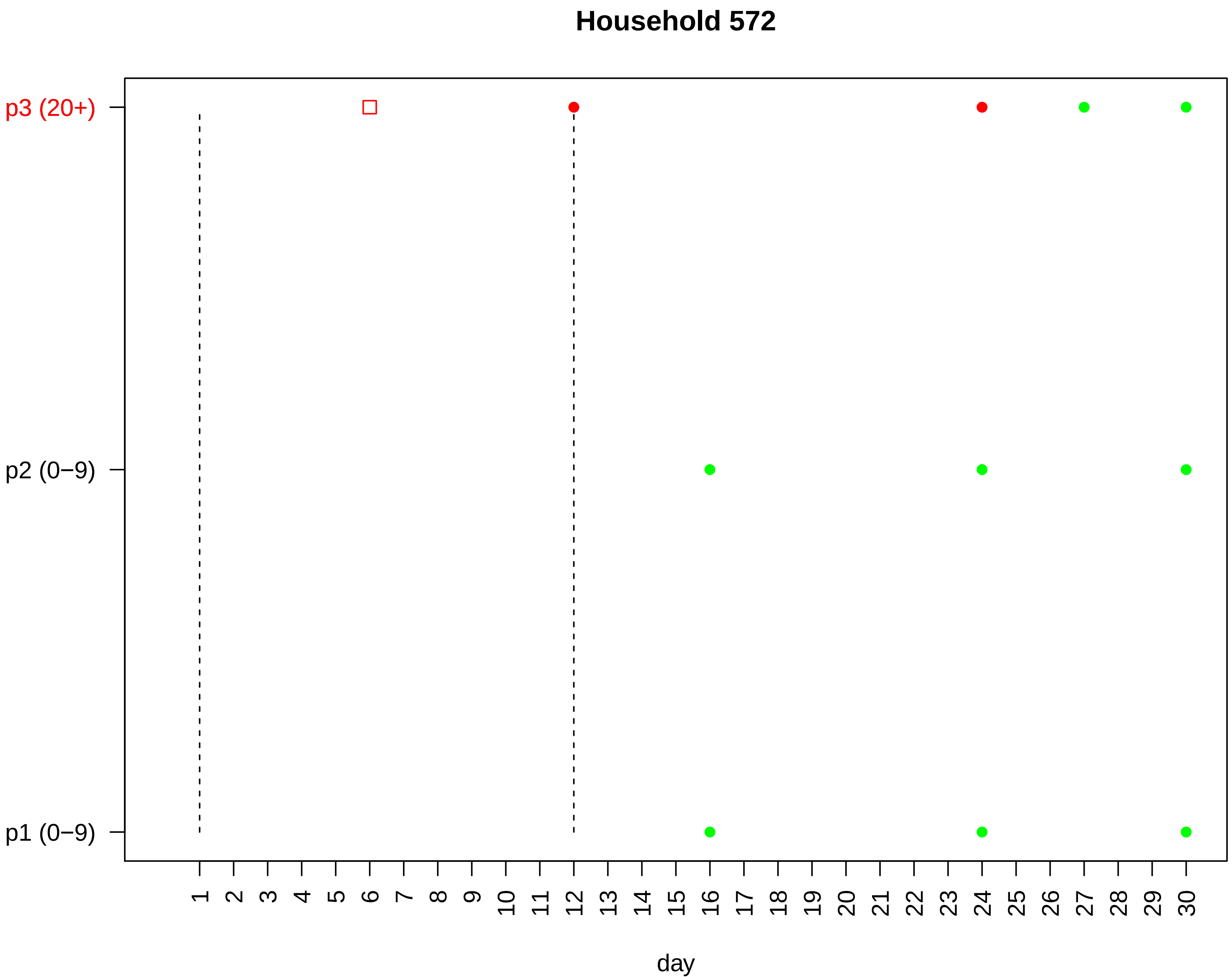
Household 572
p3 (20+)
p2 (0−9)
p1 (0−9)
1 2 3 4 5 6 7 8 9 10 11 12 13 14 15 16 17 18 19 20 21 22 23 24 25 26 27 28 29 30
day

#### Household 573

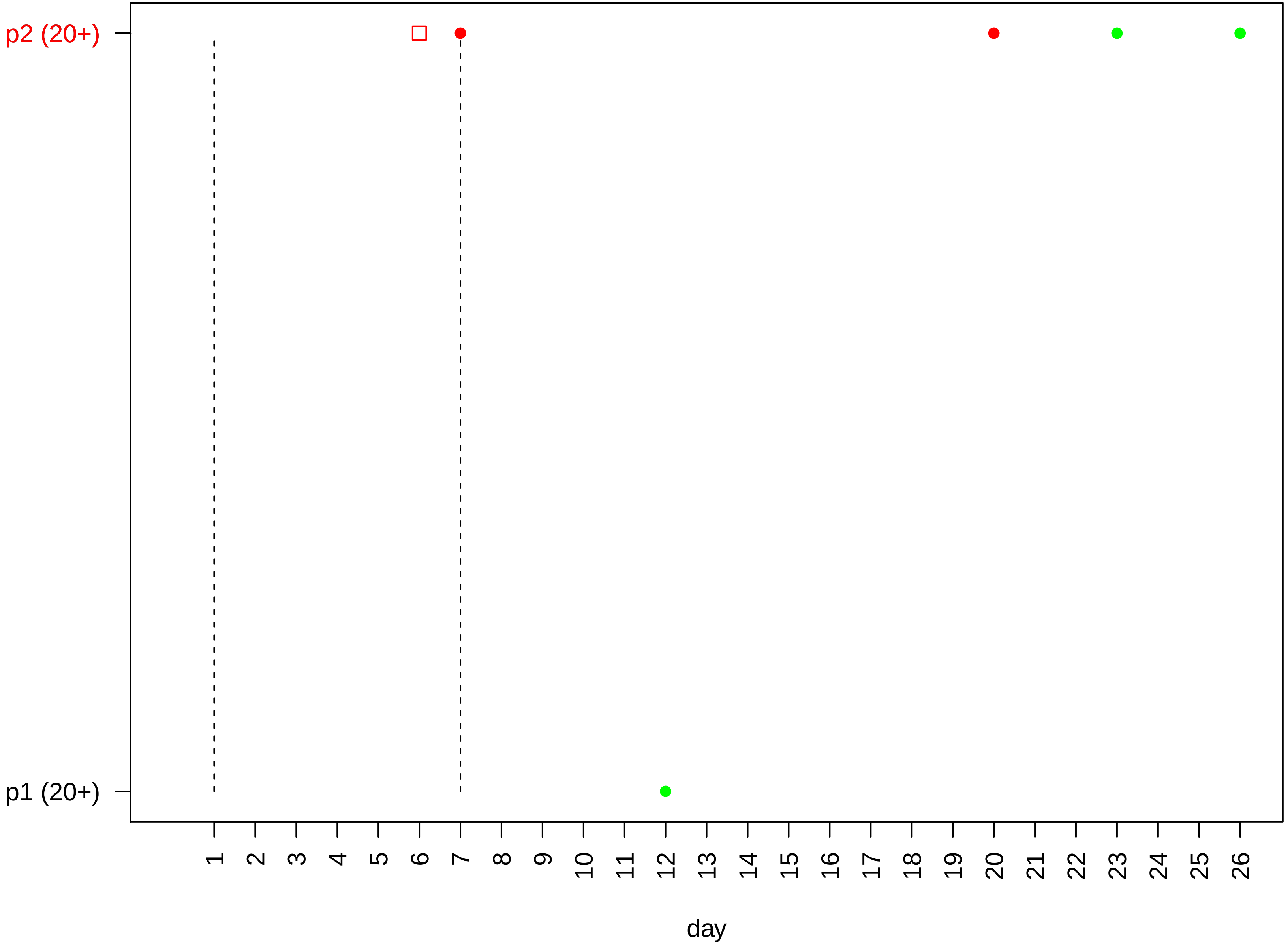

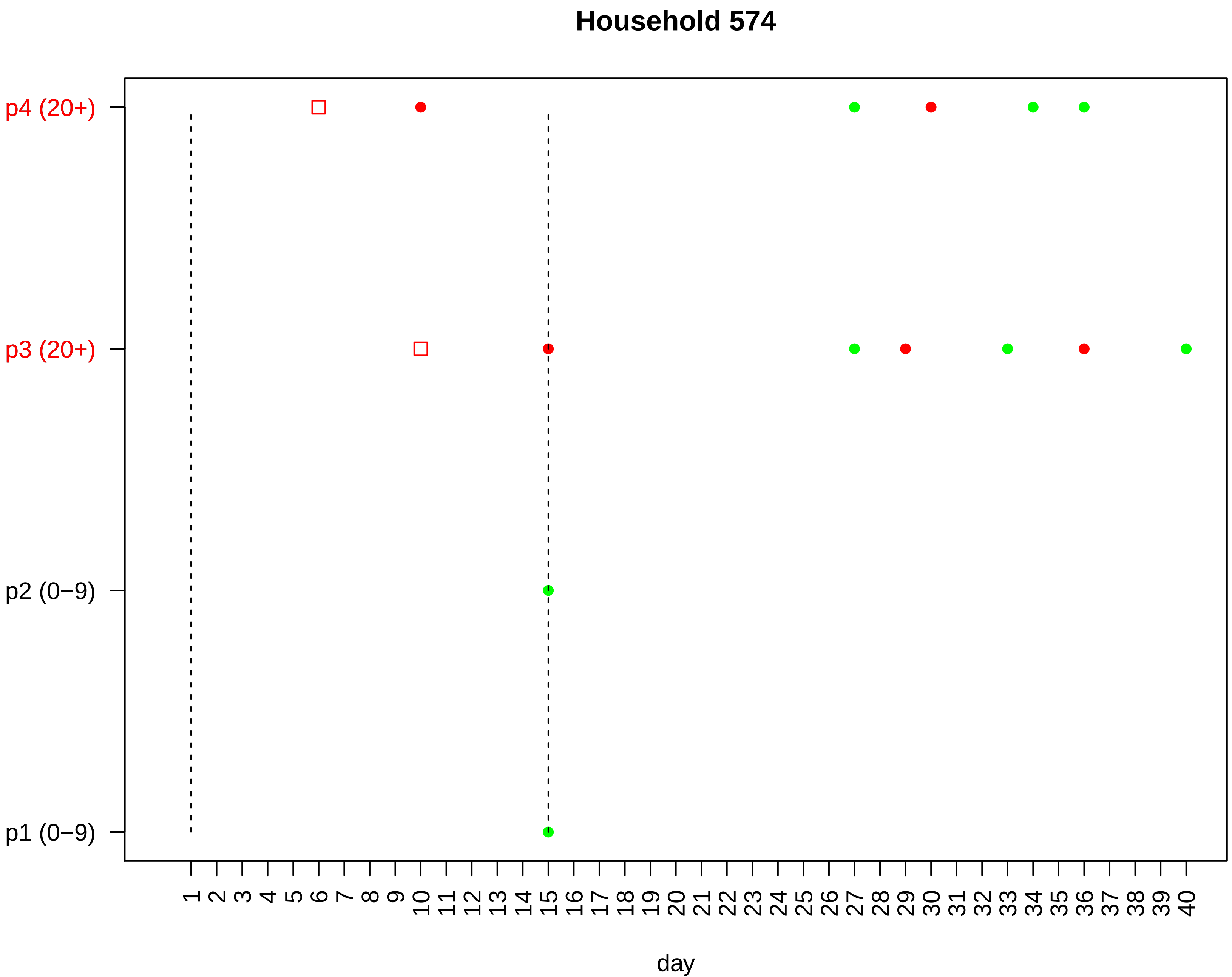
Household 574
p4 (20+)
p3 (20+)
p2 (0−9)
p1 (0−9)
1 2 3 4 5 6 7 8 9 10 11 12 13 14 15 16 17 18 19 20 21 22 23 24 25 26 27 28 29 30 31 32 33 34 35 36 37 38 39 40
day

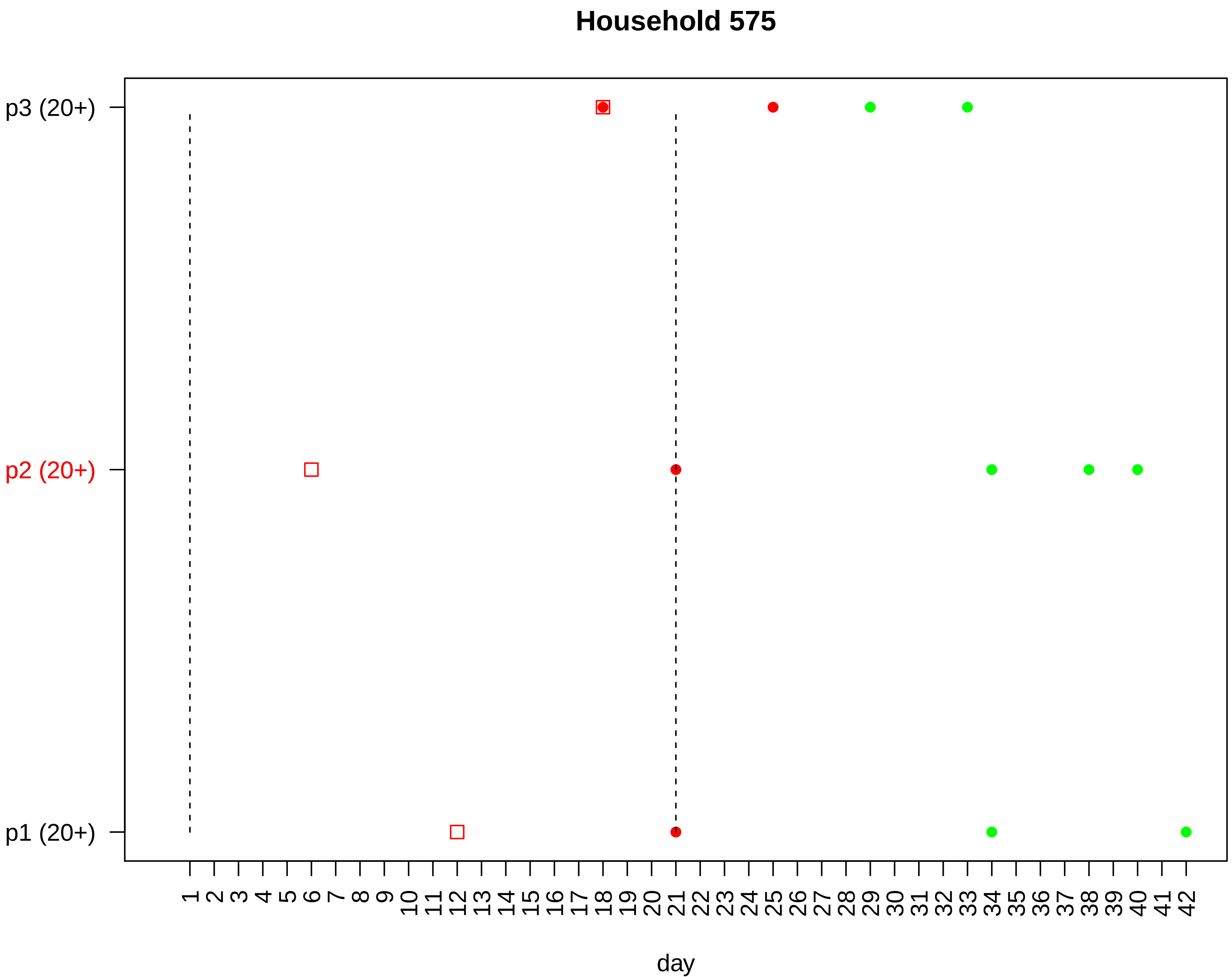

Household 575
p3 (20+)
p2 (20+)
p1 (20+)
1 2 3 4 5 6 7 8 9 10 11 12 13 14 15 16 17 18 19 20 21 22 23 24 25 26 27 28 29 30 31 32 33 34 35 36 37 38 39 40 41 42
day

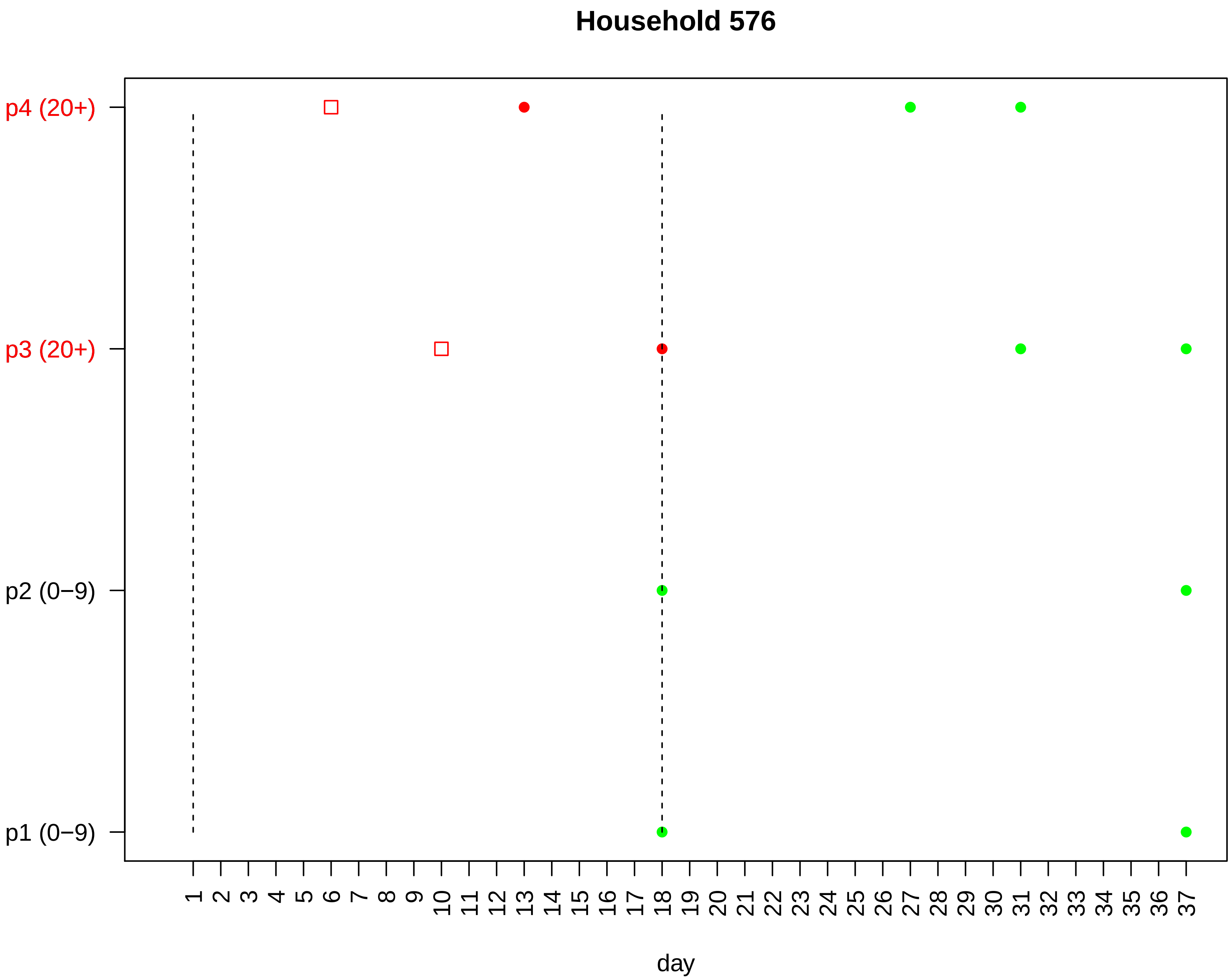

Household 576
p4 (20+)
p3 (20+)
p2 (0−9)
p1 (0−9)
1 2 3 4 5 6 7 8 9 10 11 12 13 14 15 16 17 18 19 20 21 22 23 24 25 26 27 28 29 30 31 32 33 34 35 36 37
day

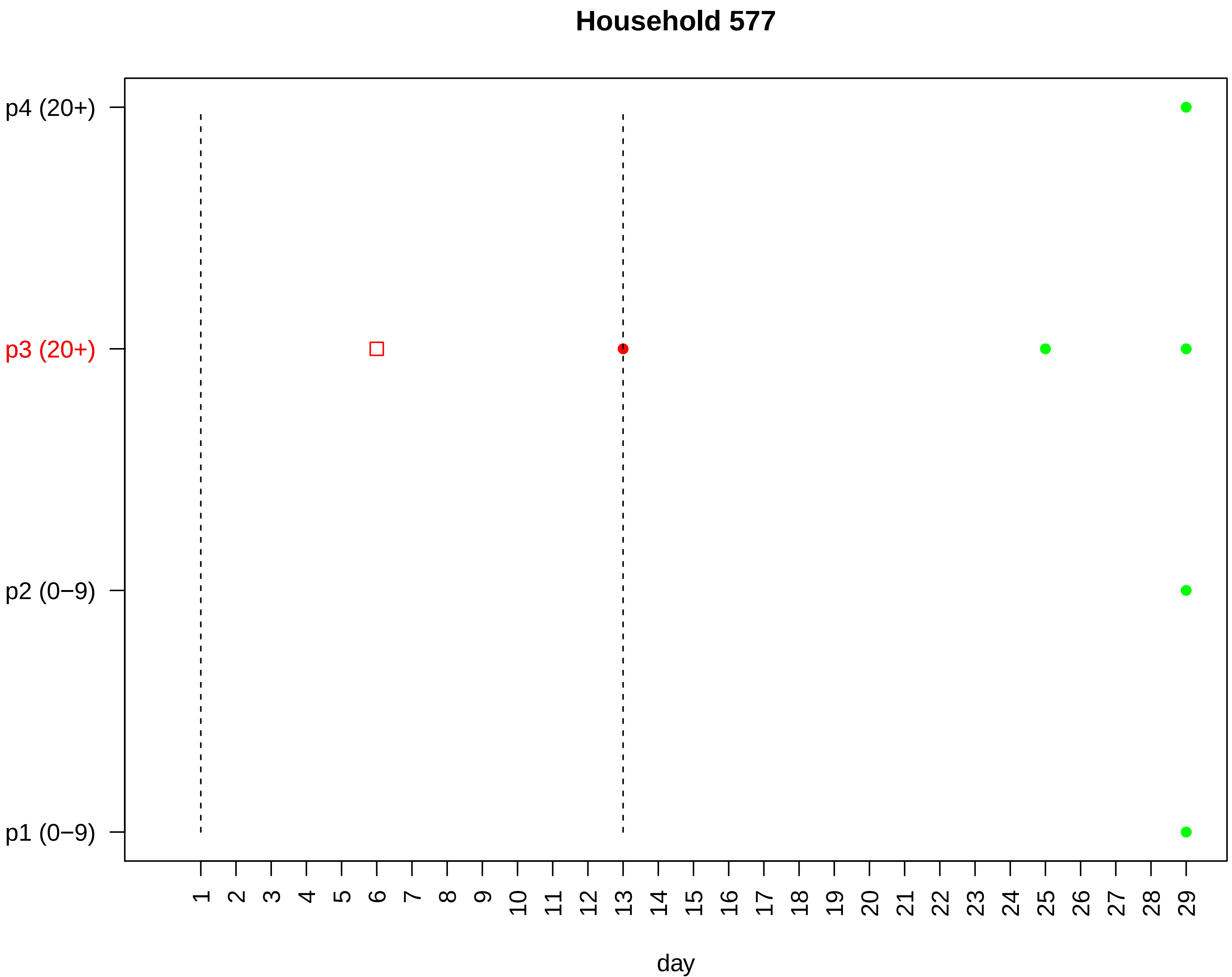

Household 577
p4 (20+)
p3 (20+)
p2 (0−9)
p1 (0−9)
day

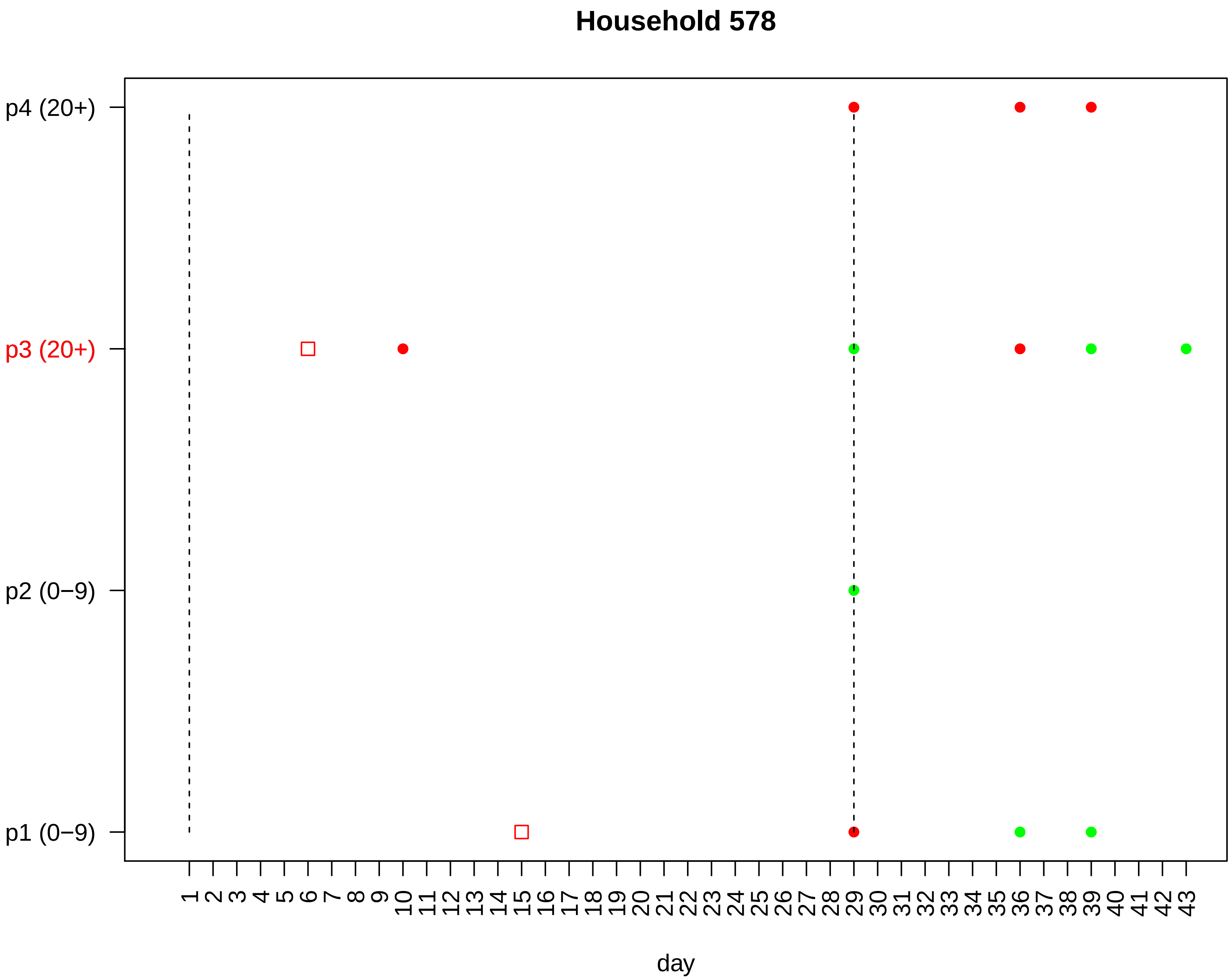

Household 578
p4 (20+)
p3 (20+)
p2 (0−9)
p1 (0−9)
1 2 3 4 5 6 7 8 9 10 11 12 13 14 15 16 17 18 19 20 21 22 23 24 25 26 27 28 29 30 31 32 33 34 35 36 37 38 39 40 41 42 43
day

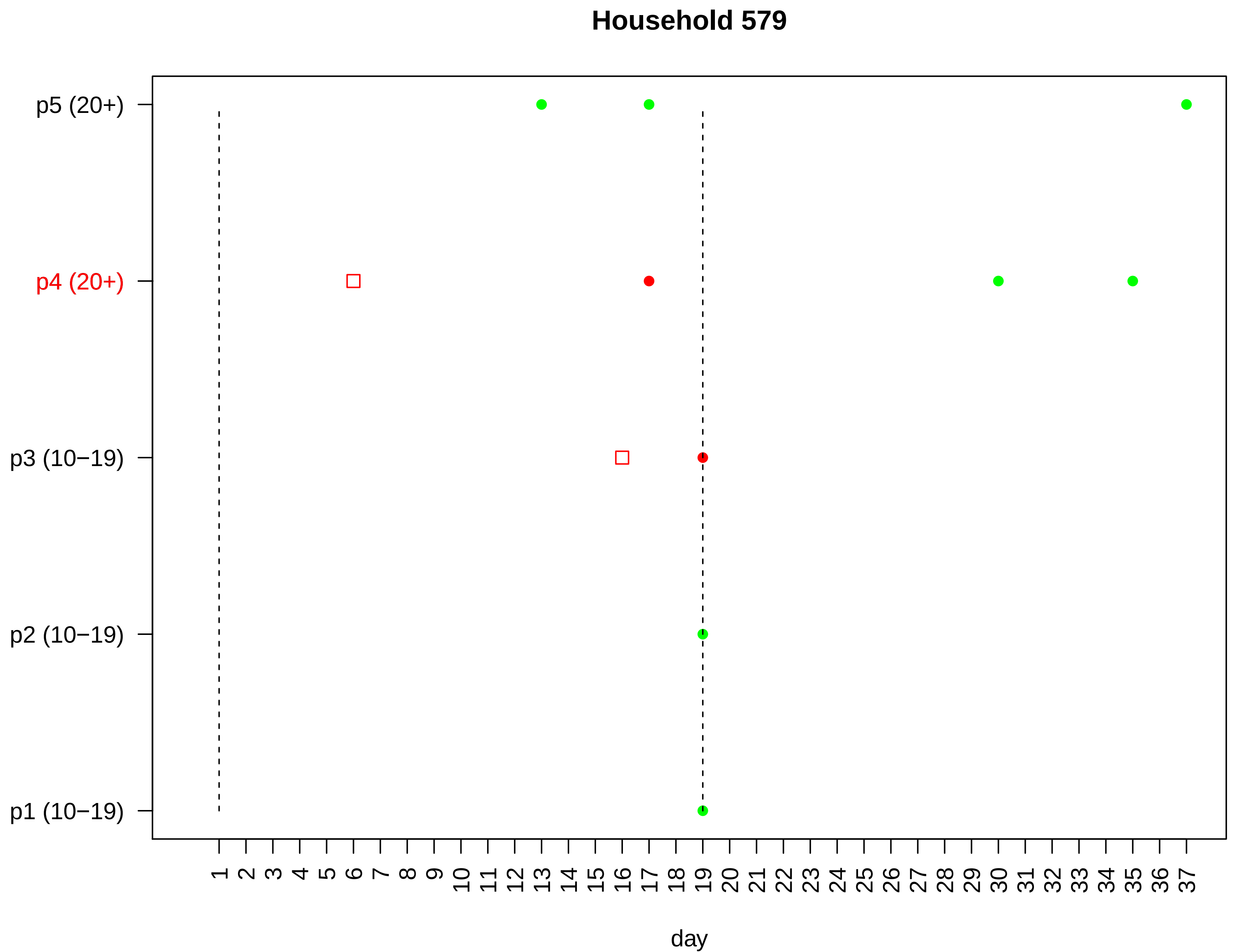

Household 579
p5 (20+)
p4 (20+)
p3 (10−19)
p2 (10−19)
p1 (10−19)
1 2 3 4 5 6 7 8 9 10 11 12 13 14 15 16 17 18 19 20 21 22 23 24 25 26 27 28 29 30 31 32 33 34 35 36 37
day

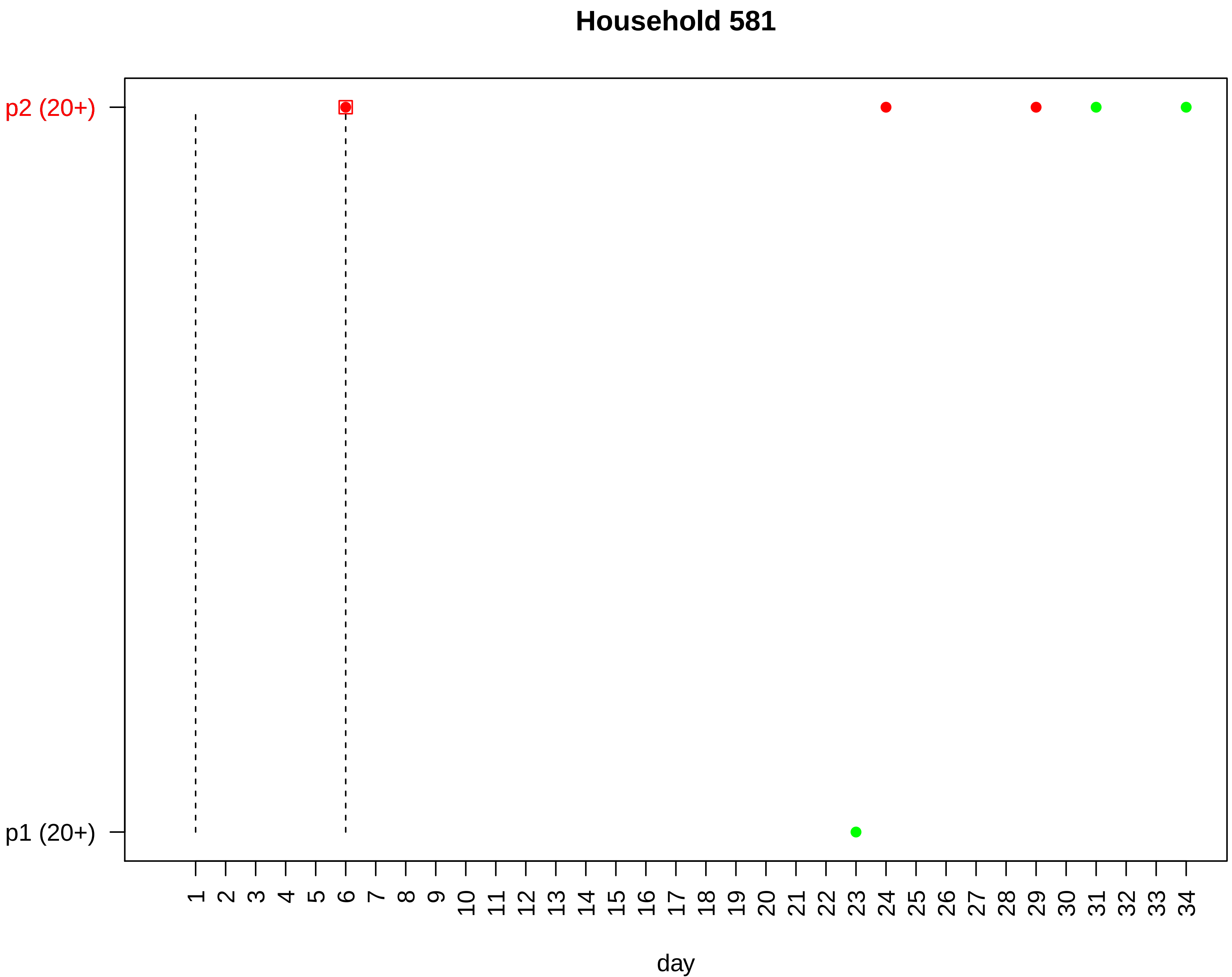

Household 581
p2 (20+)
p1 (20+)
day

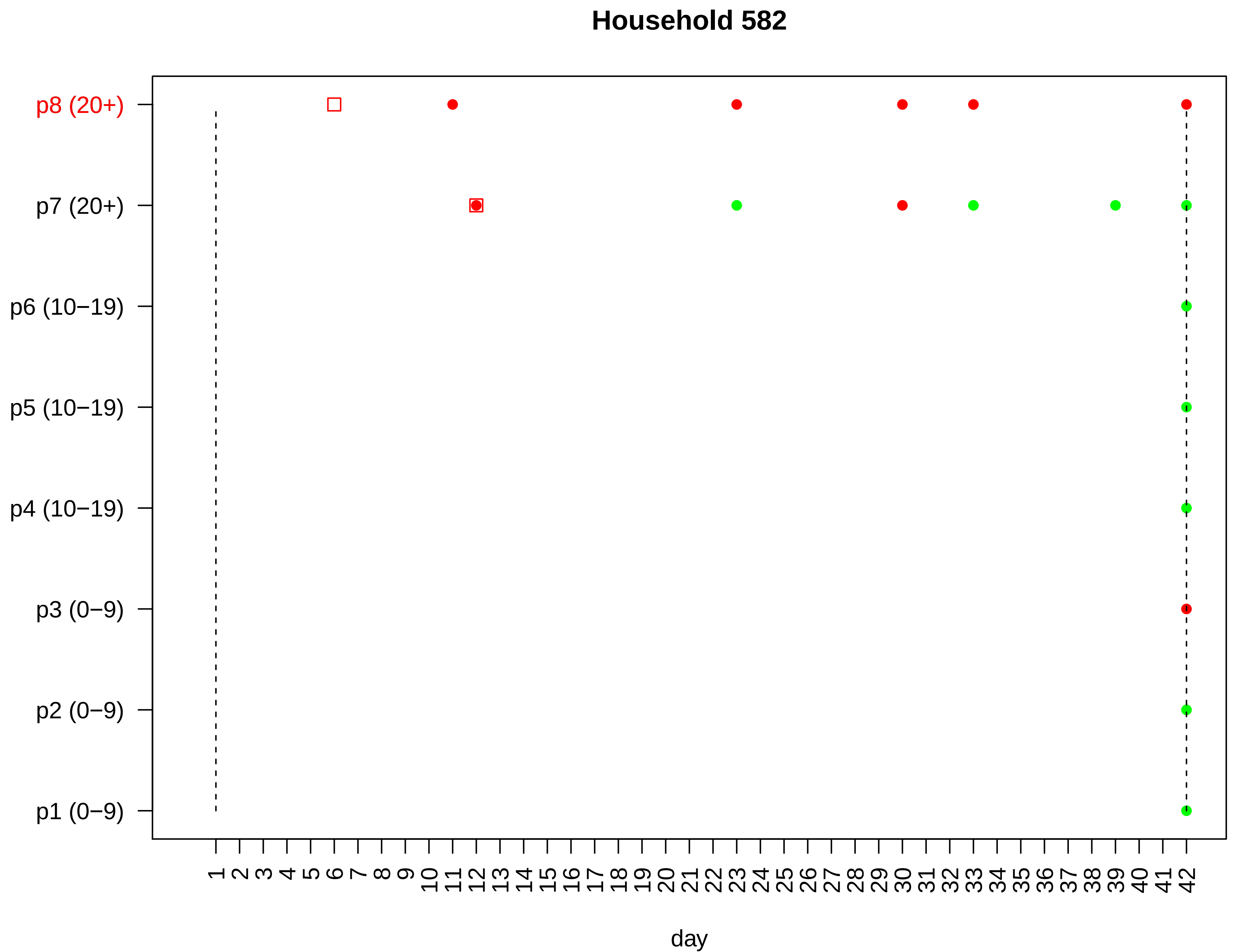
Household 582
p8 (20+)
p7 (20+)
p6 (10−19)
p5 (10−19)
p4 (10−19)
p3 (0−9)
p2 (0−9)
p1 (0−9)
1 2 3 4 5 6 7 8 9 10 11 12 13 14 15 16 17 18 19 20 21 22 23 24 25 26 27 28 29 30 31 32 33 34 35 36 37 38 39 40 41 42
day

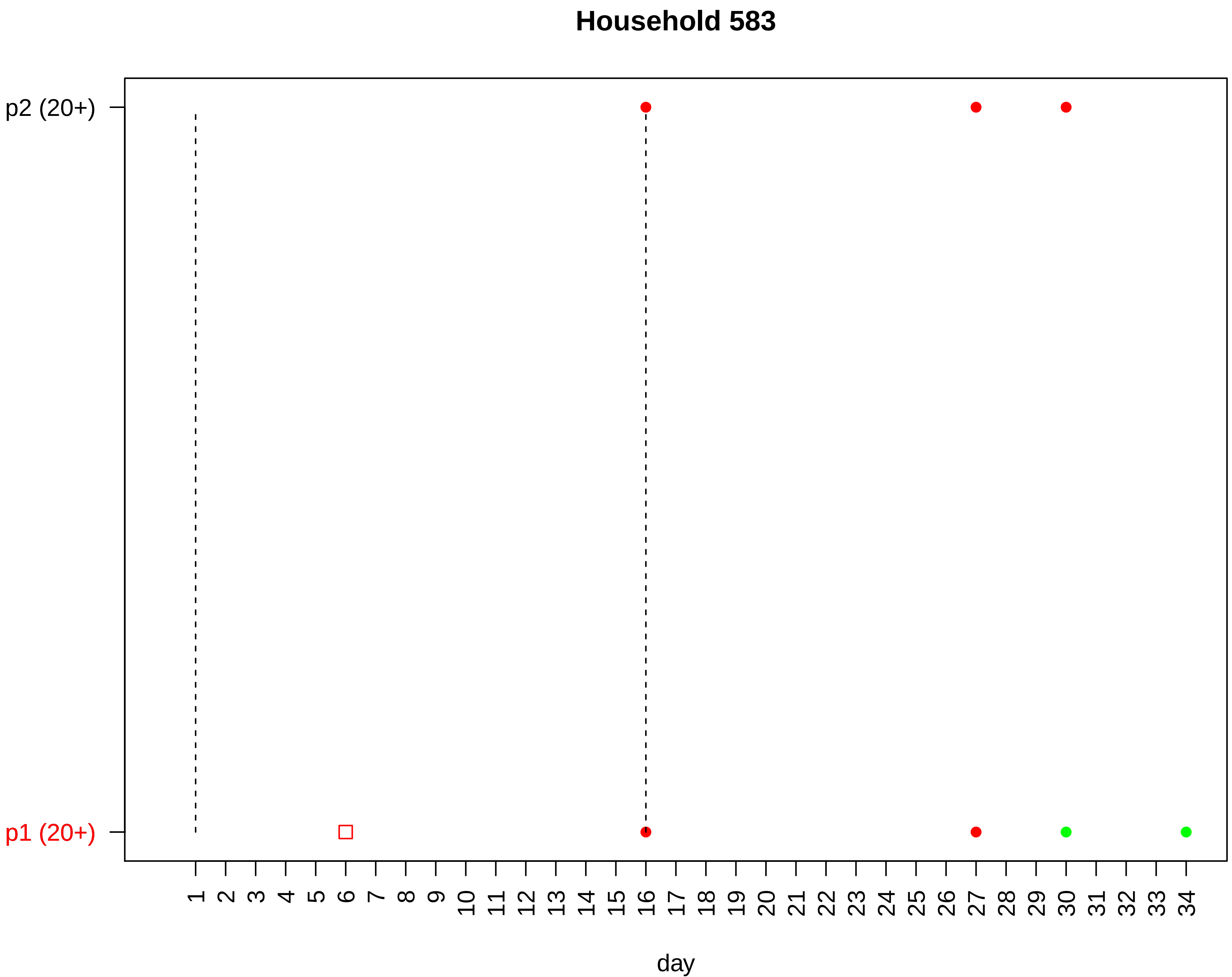
Household 583
p2 (20+)
p1 (20+)
day

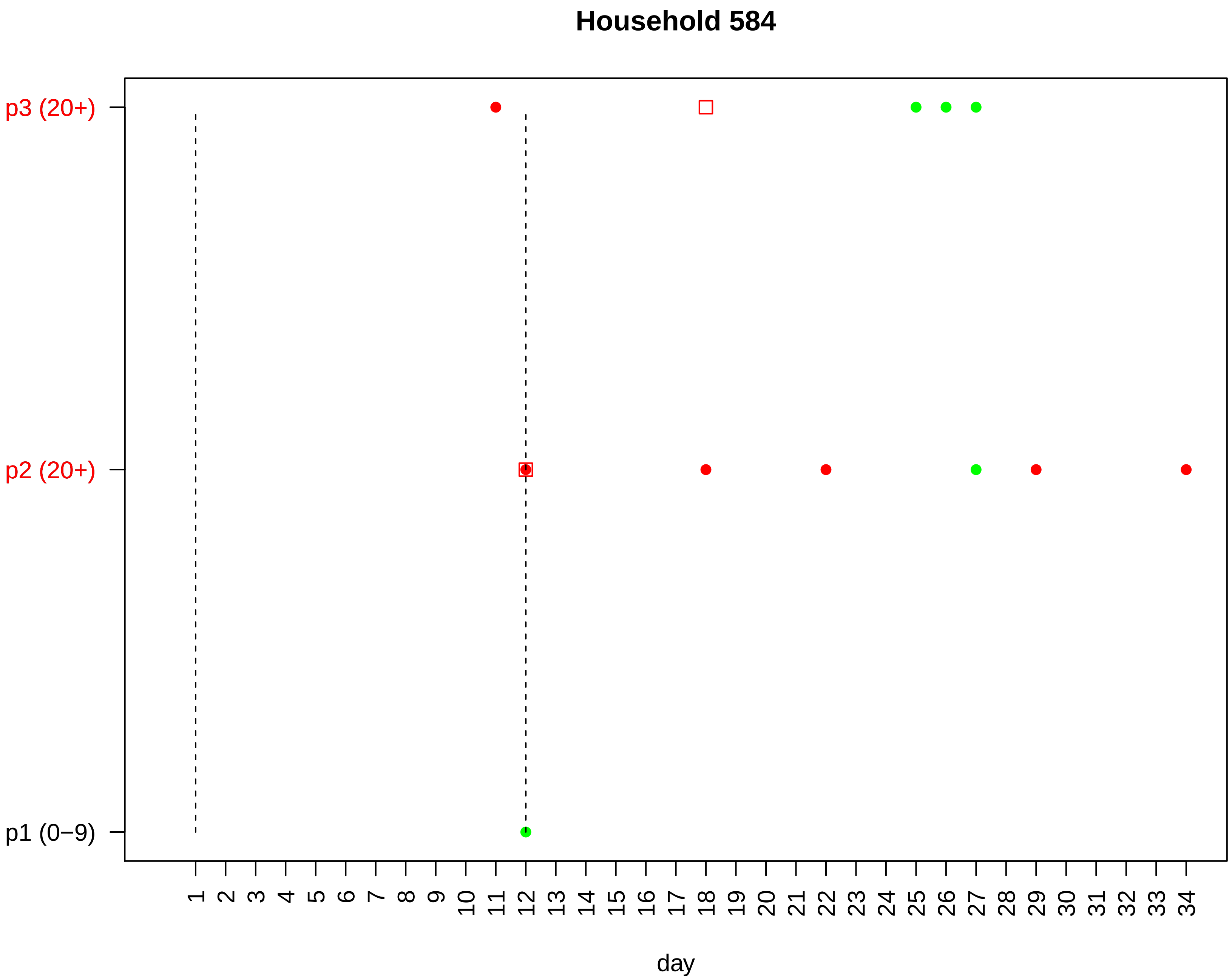

Household 584
p3 (20+)
p2 (20+)
p1 (0−9)
1 2 3 4 5 6 7 8 9 10 11 12 13 14 15 16 17 18 19 20 21 22 23 24 25 26 27 28 29 30 31 32 33 34
day

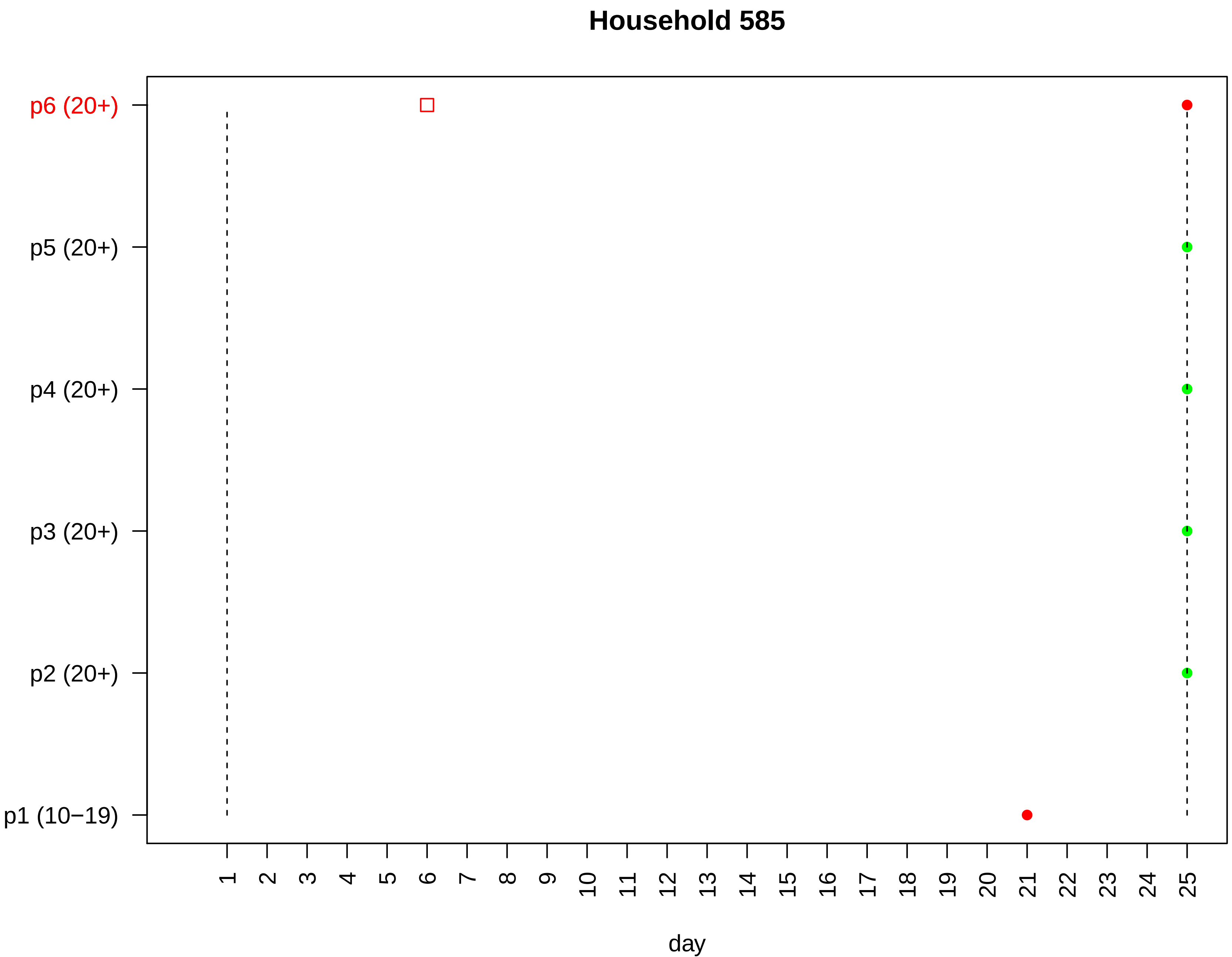

Household 585
p6 (20+)
p5 (20+)
p4 (20+)
p3 (20+)
p2 (20+)
p1 (10−19)
1 2 3 4 5 6 7 8 9 10 11 12 13 14 15 16 17 18 19 20 21 22 23 24 25
day

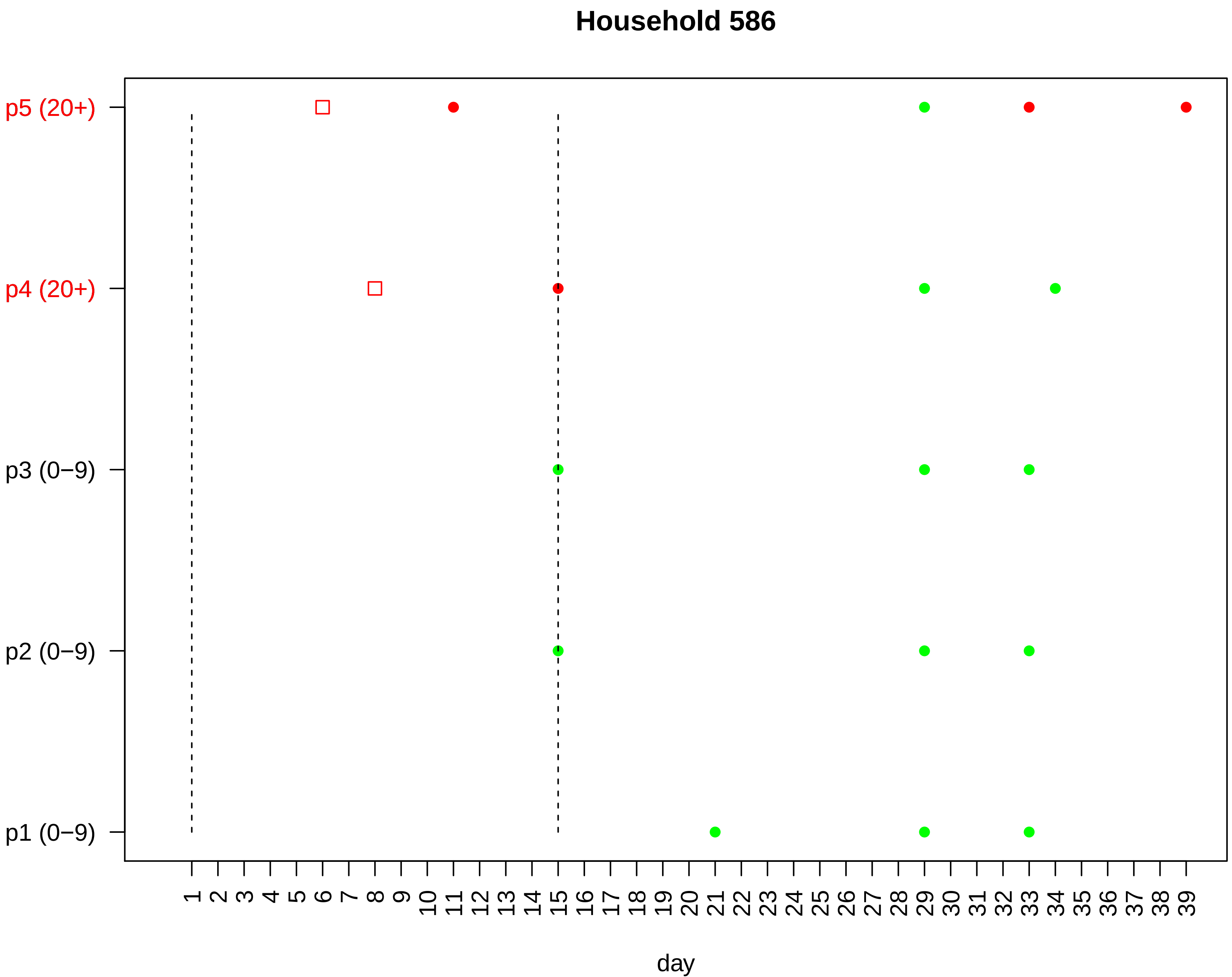

Household 586
p5 (20+)
p4 (20+)
p3 (0−9)
p2 (0−9)
p1 (0−9)
1 2 3 4 5 6 7 8 9 10 11 12 13 14 15 16 17 18 19 20 21 22 23 24 25 26 27 28 29 30 31 32 33 34 35 36 37 38 39
day

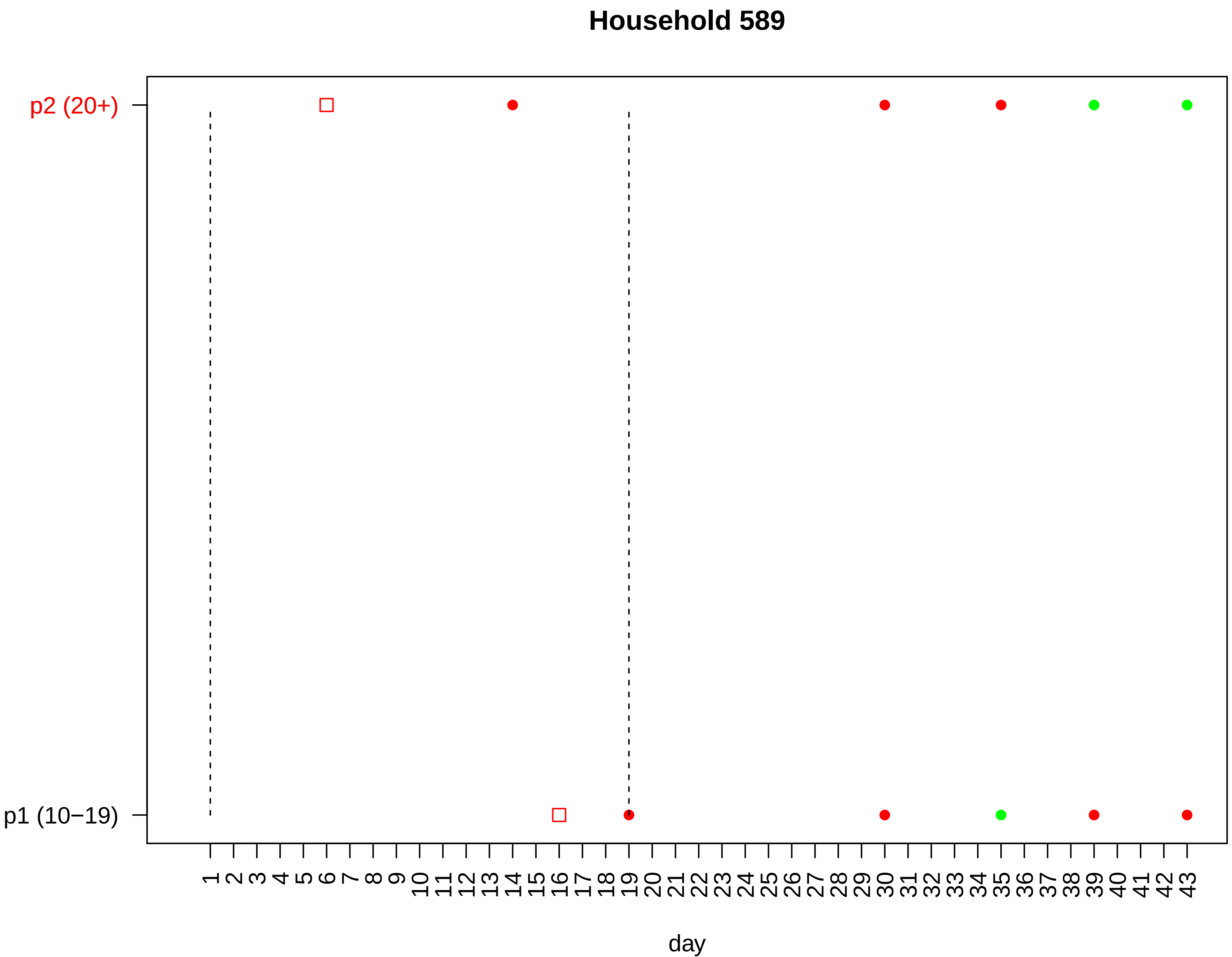

Household 589
p2 (20+)
p1 (10−19)
day

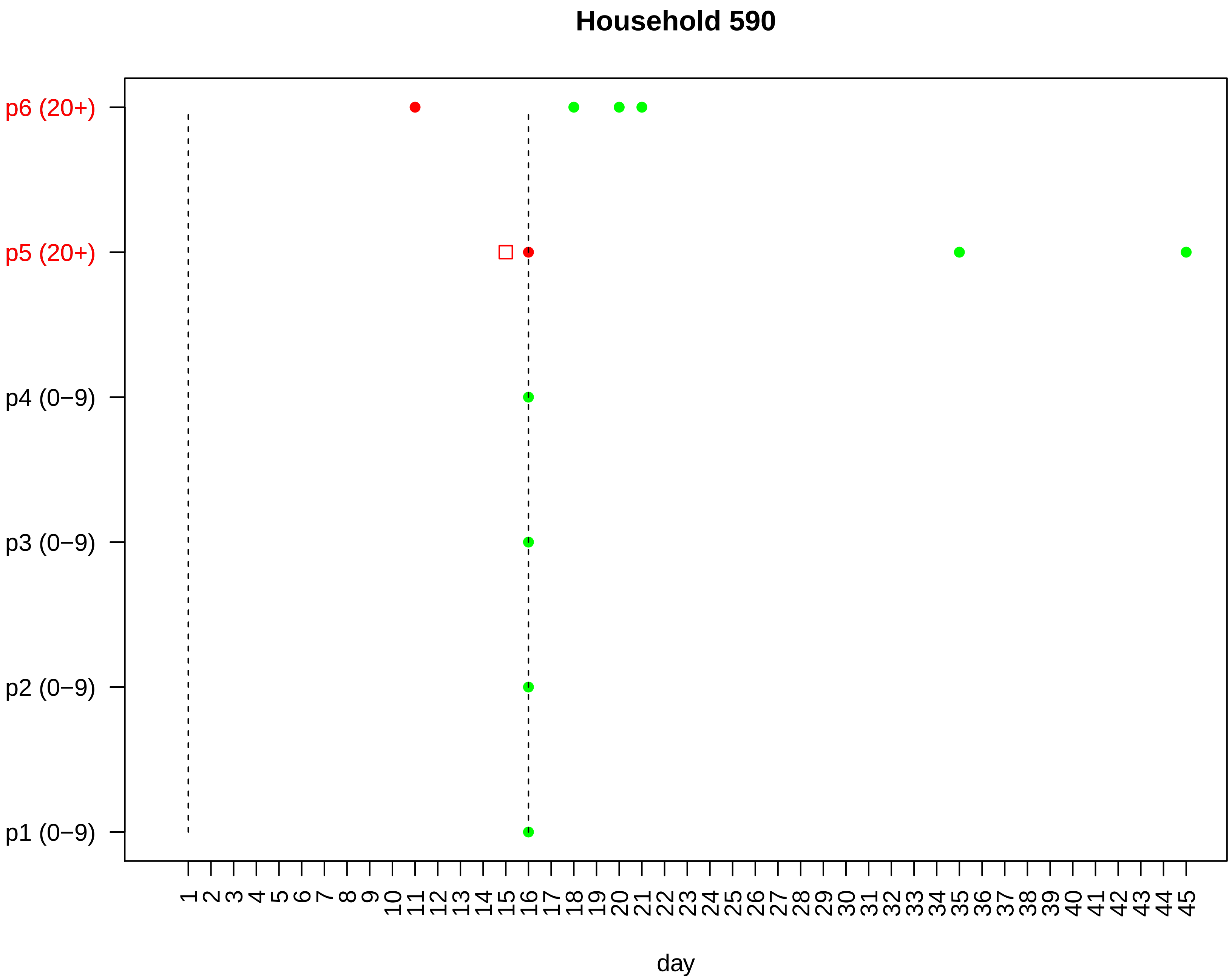

Household 590
p6 (20+)
p5 (20+)
p4 (0−9)
p3 (0−9)
p2 (0−9)
p1 (0−9)
1 2 3 4 5 6 7 8 9 10 11 12 13 14 15 16 17 18 19 20 21 22 23 24 25 26 27 28 29 30 31 32 33 34 35 36 37 38 39 40 41 42 43 44 45
day

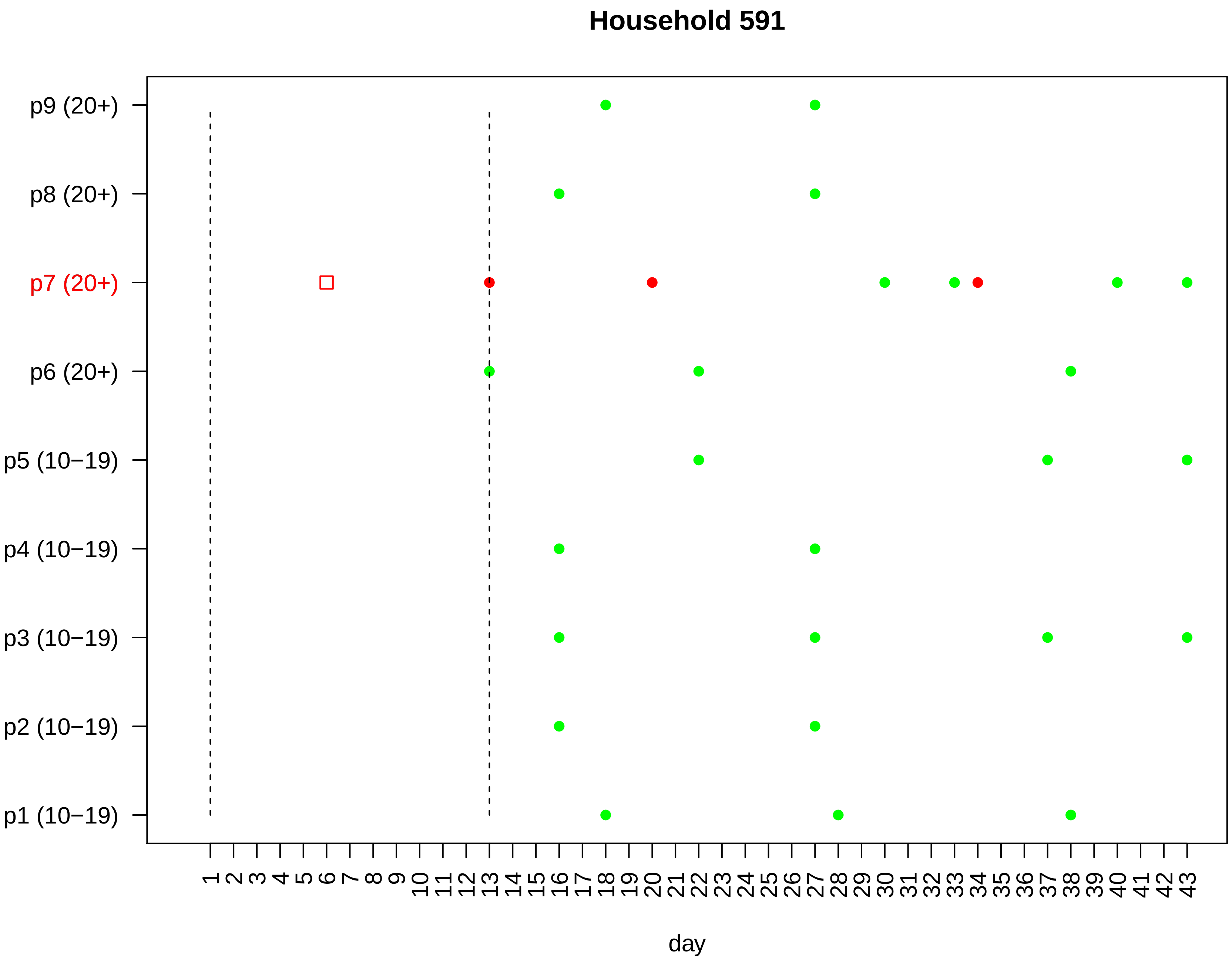

Household 591
p9 (20+)
p8 (20+)
p7 (20+)
p6 (20+)
p5 (10−19)
p4 (10−19)
p3 (10−19)
p2 (10−19)
p1 (10−19)
1 2 3 4 5 6 7 8 9 10 11 12 13 14 15 16 17 18 19 20 21 22 23 24 25 26 27 28 29 30 31 32 33 34 35 36 37 38 39 40 41 42 43
day

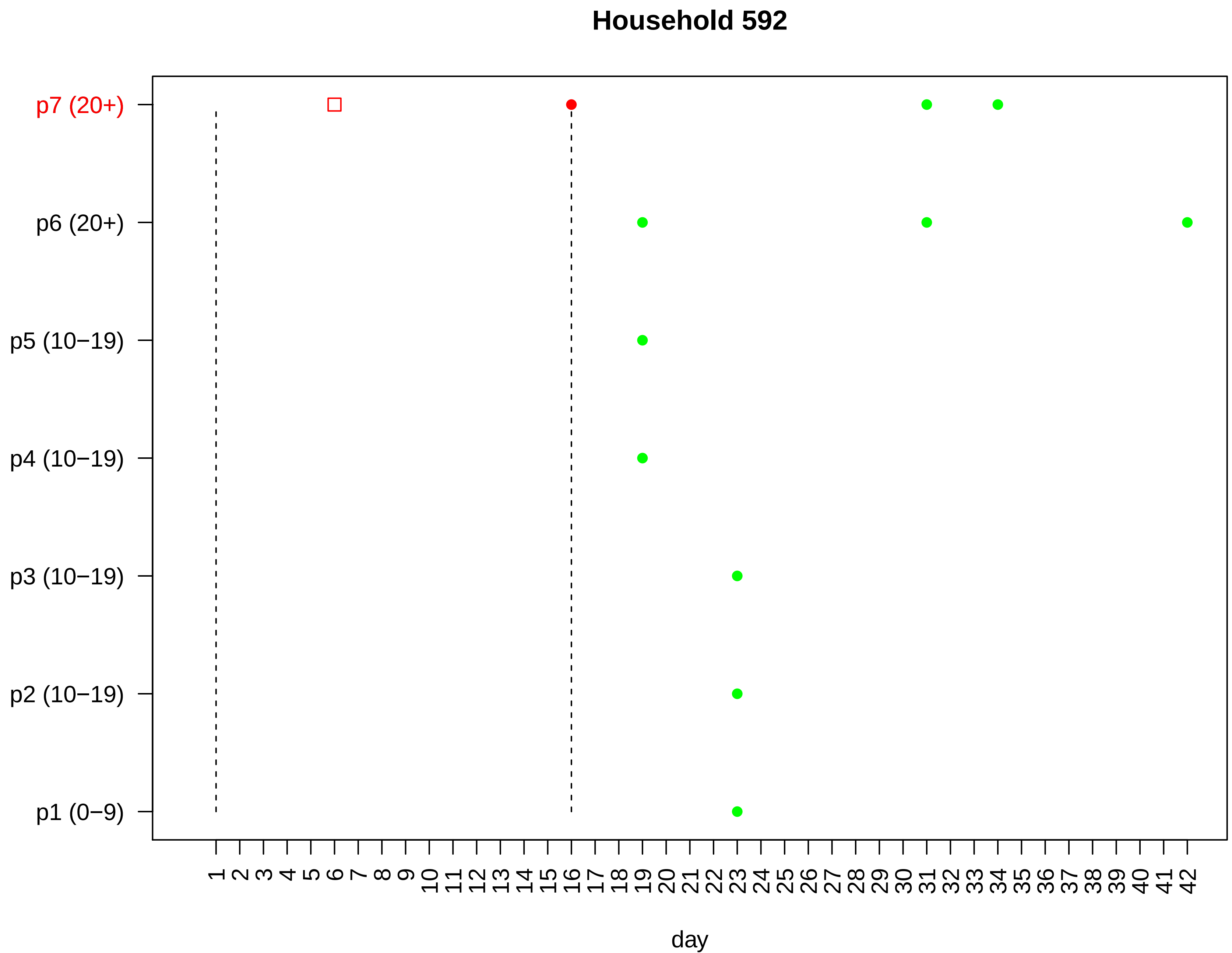

Household 592
p7 (20+)
p6 (20+)
p5 (10−19)
p4 (10−19)
p3 (10−19)
p2 (10−19)
p1 (0−9)
1 2 3 4 5 6 7 8 9 10 11 12 13 14 15 16 17 18 19 20 21 22 23 24 25 26 27 28 29 30 31 32 33 34 35 36 37 38 39 40 41 42
day

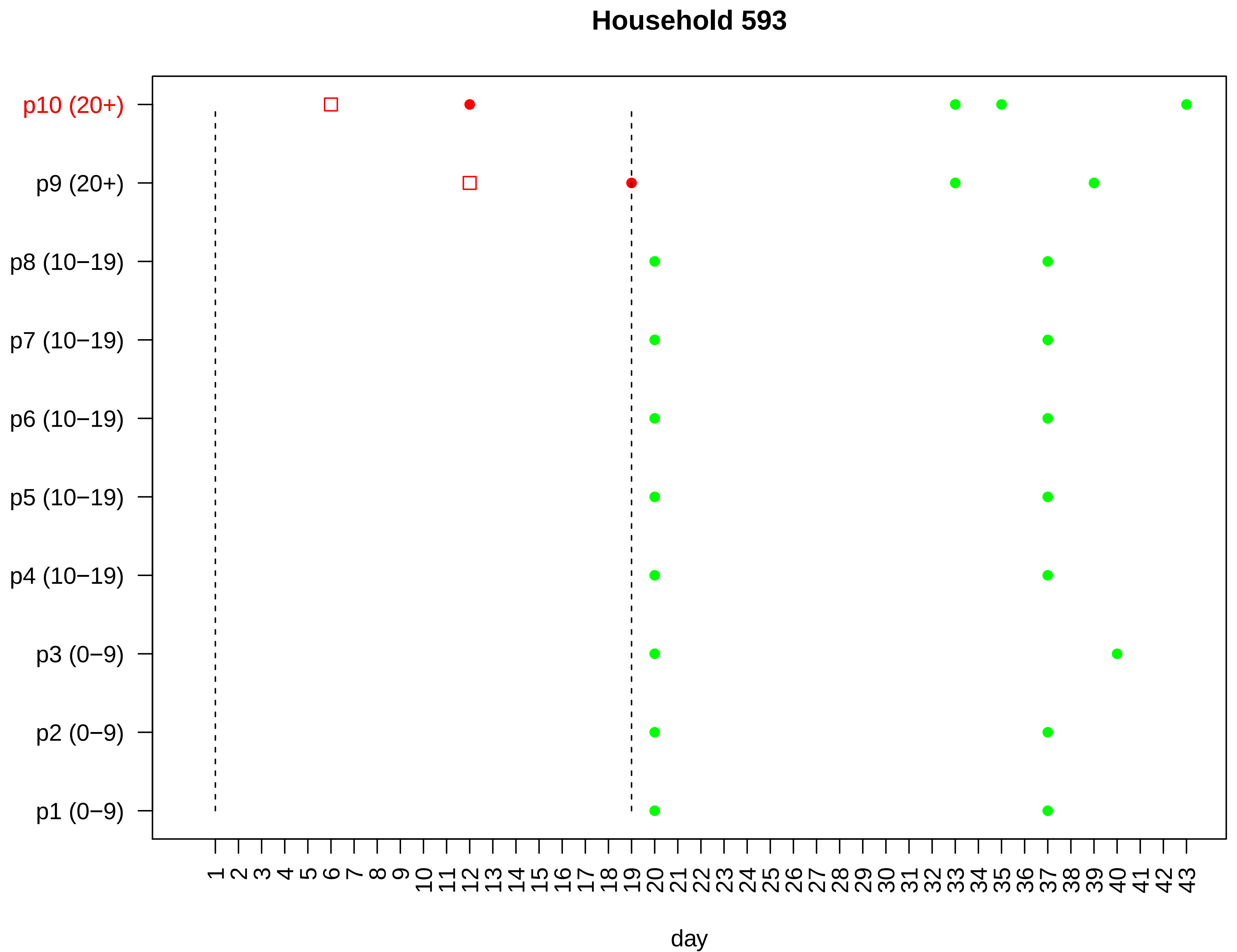

Household 593
p10 (20+)
p9 (20+)
p8 (10−19)
p7 (10−19)
p6 (10−19)
p5 (10−19)
p4 (10−19)
p3 (0−9)
p2 (0−9)
p1 (0−9)
1 2 3 4 5 6 7 8 9 10 11 12 13 14 15 16 17 18 19 20 21 22 23 24 25 26 27 28 29 30 31 32 33 34 35 36 37 38 39 40 41 42 43
day

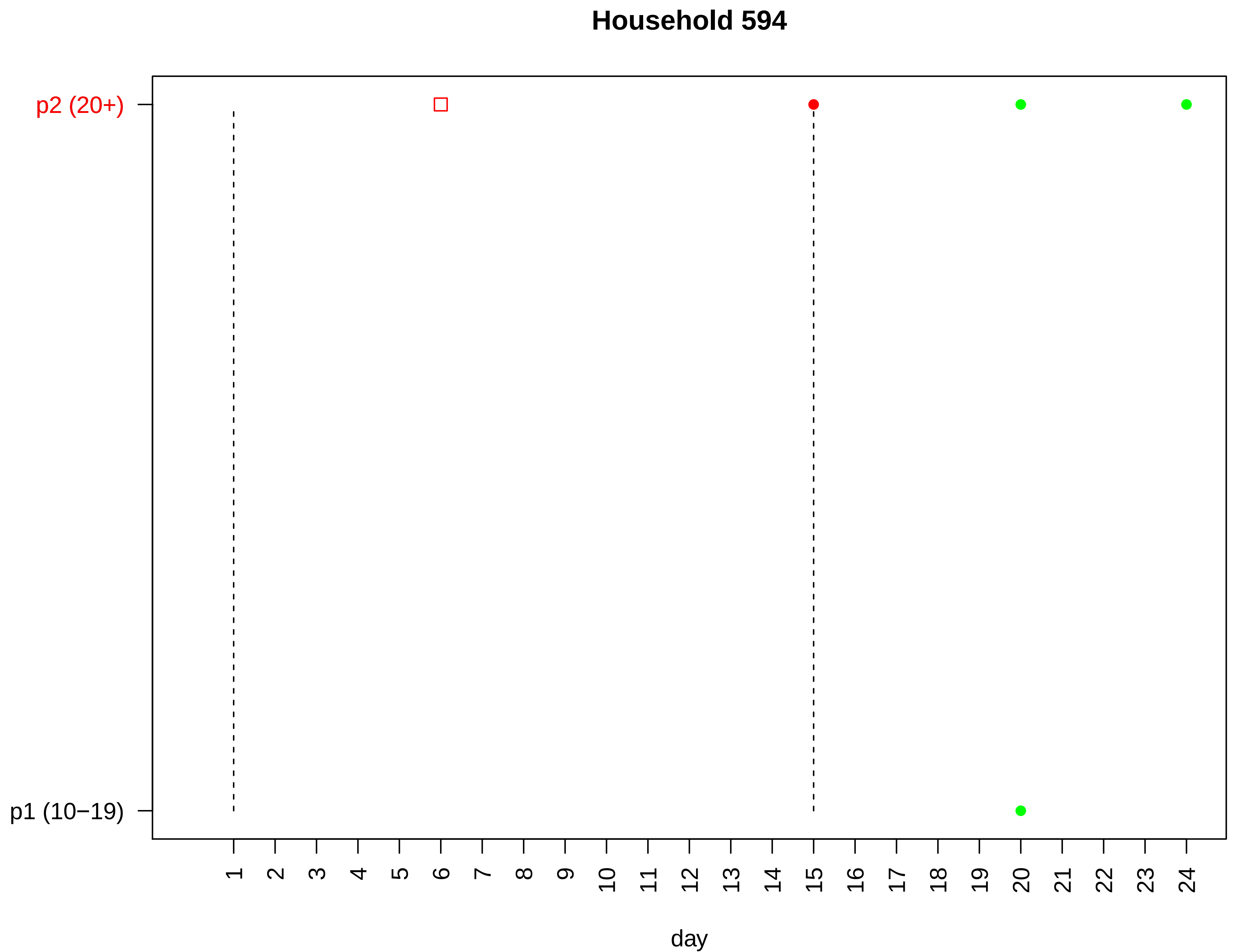
Household 594
p2 (20+)
p1 (10−19)
day

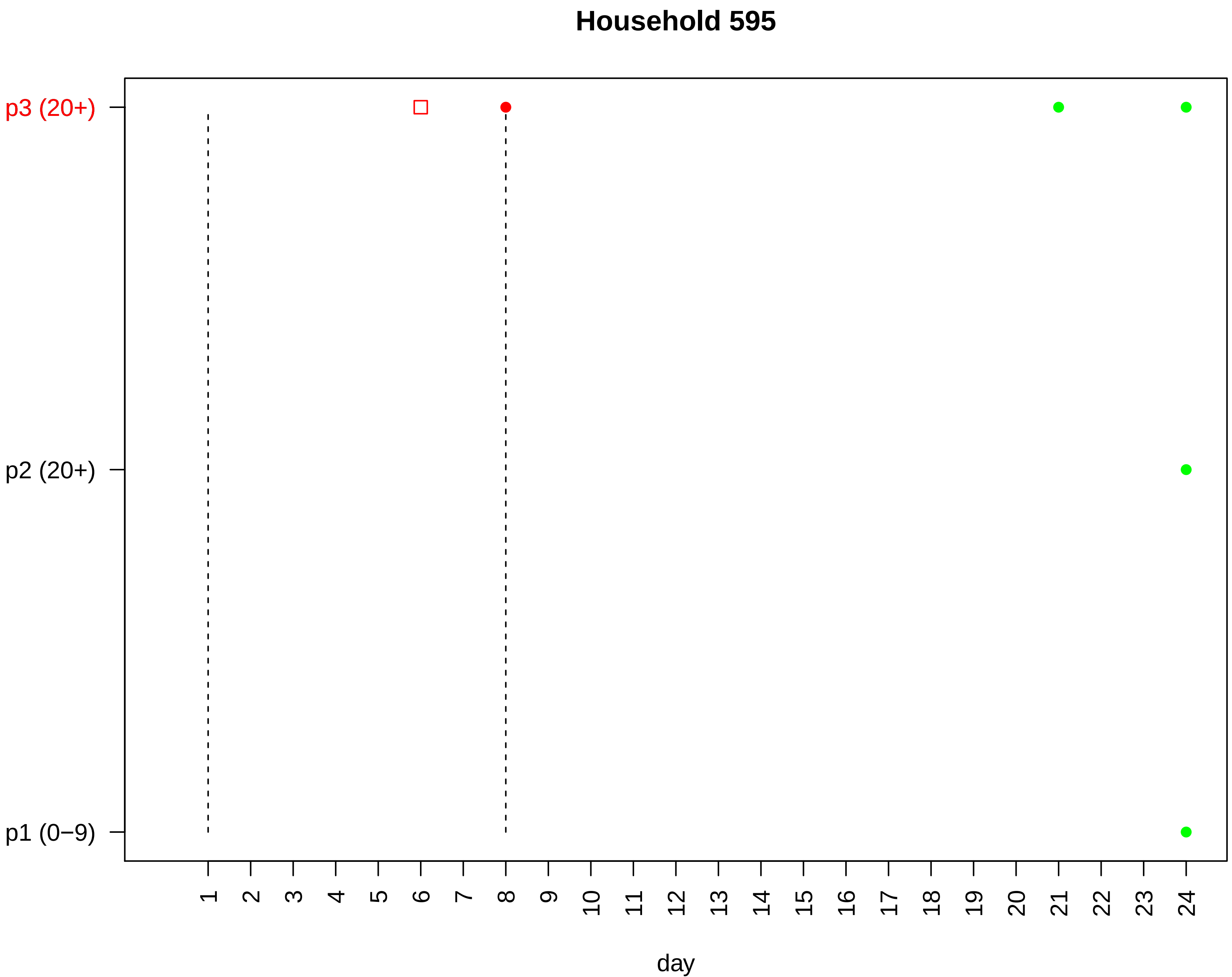

Household 595
p3 (20+)
p2 (20+)
p1 (0−9)
day

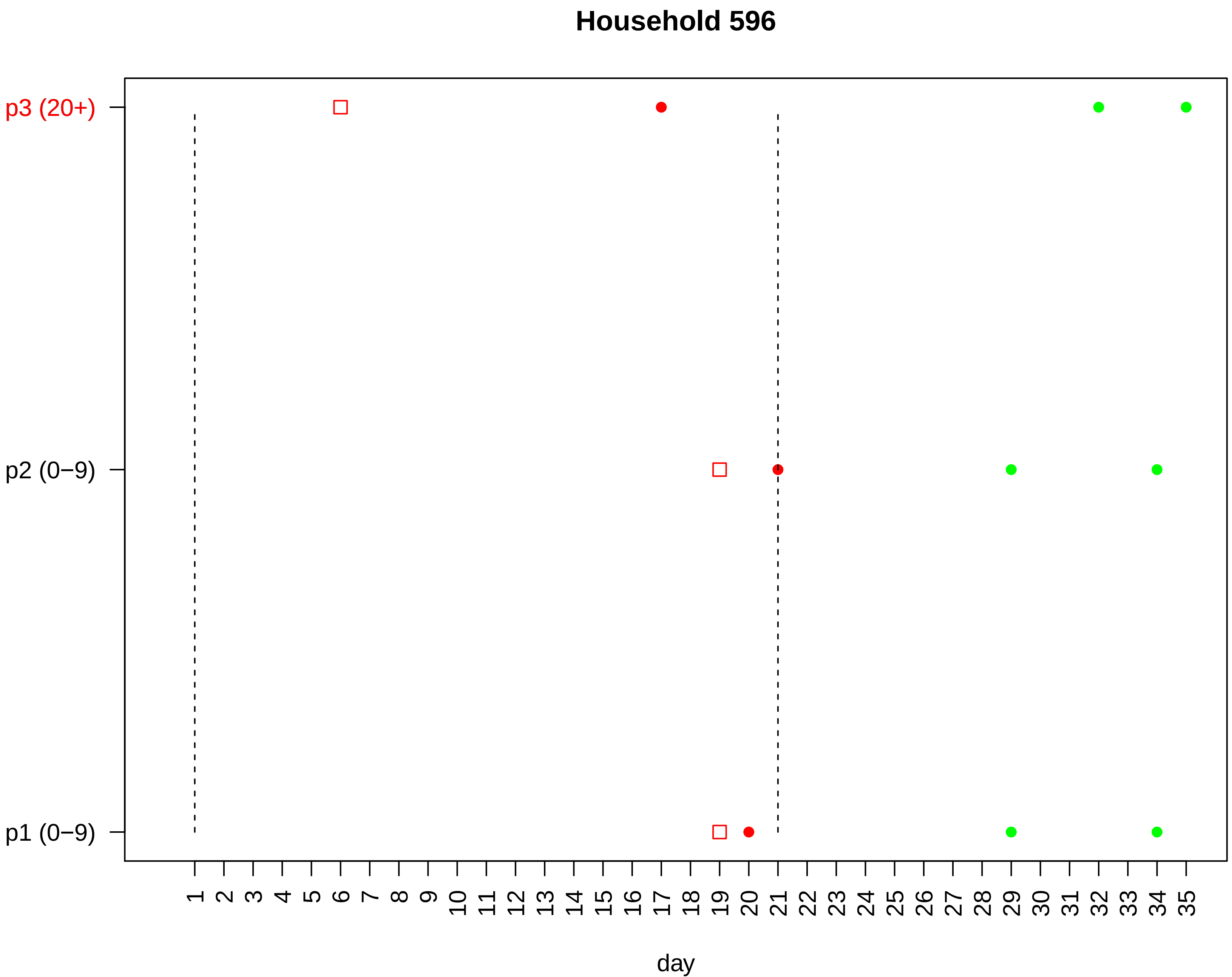

Household 596
p3 (20+)
p2 (0−9)
p1 (0−9)
1 2 3 4 5 6 7 8 9 10 11 12 13 14 15 16 17 18 19 20 21 22 23 24 25 26 27 28 29 30 31 32 33 34 35
day

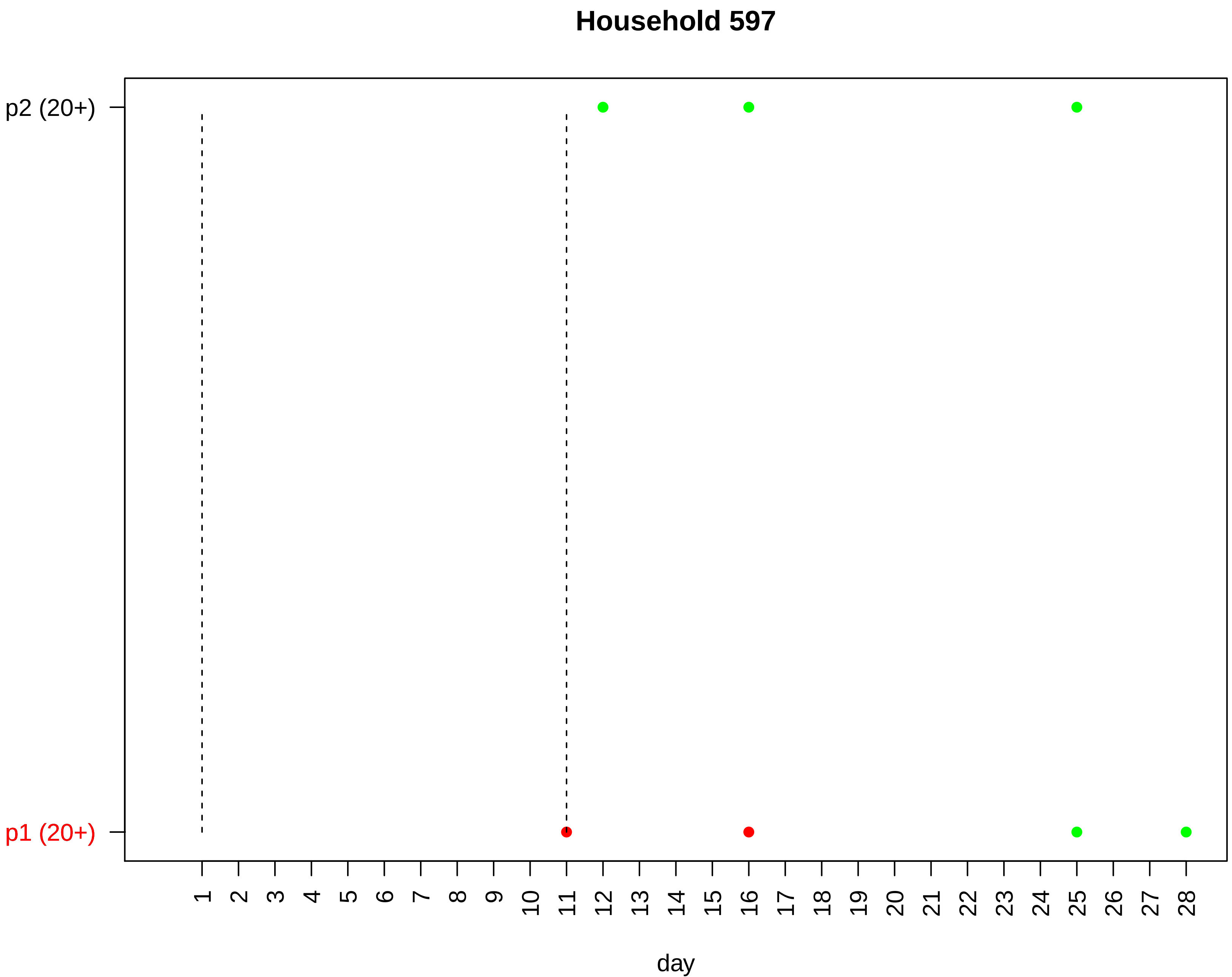

Household 597
p2 (20+)
p1 (20+)
day

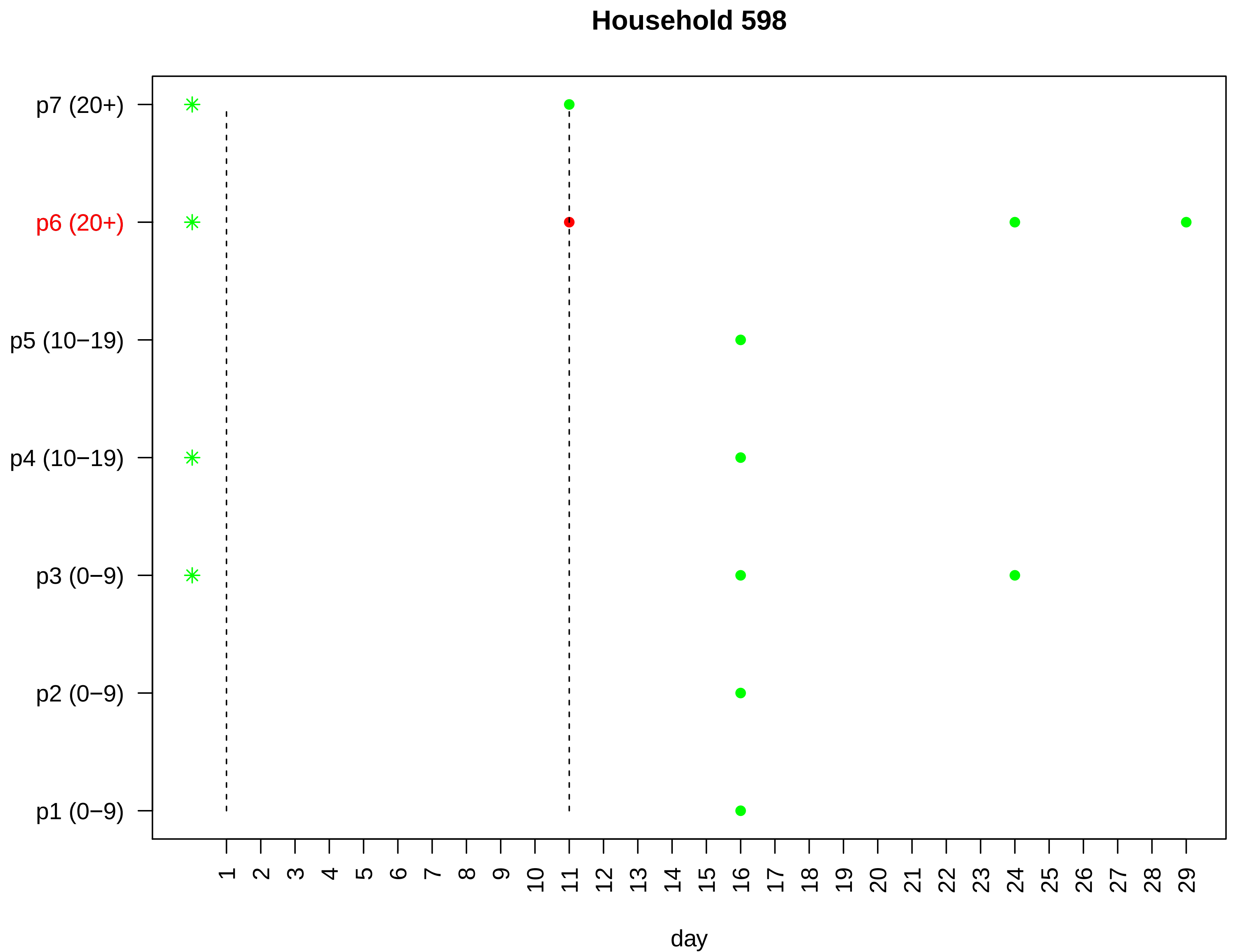

Household 598
p7 (20+)
p6 (20+)
p5 (10−19)
p4 (10−19)
p3 (0−9)
p2 (0−9)
p1 (0−9)
1 2 3 4 5 6 7 8 9 10 11 12 13 14 15 16 17 18 19 20 21 22 23 24 25 26 27 28 29
day

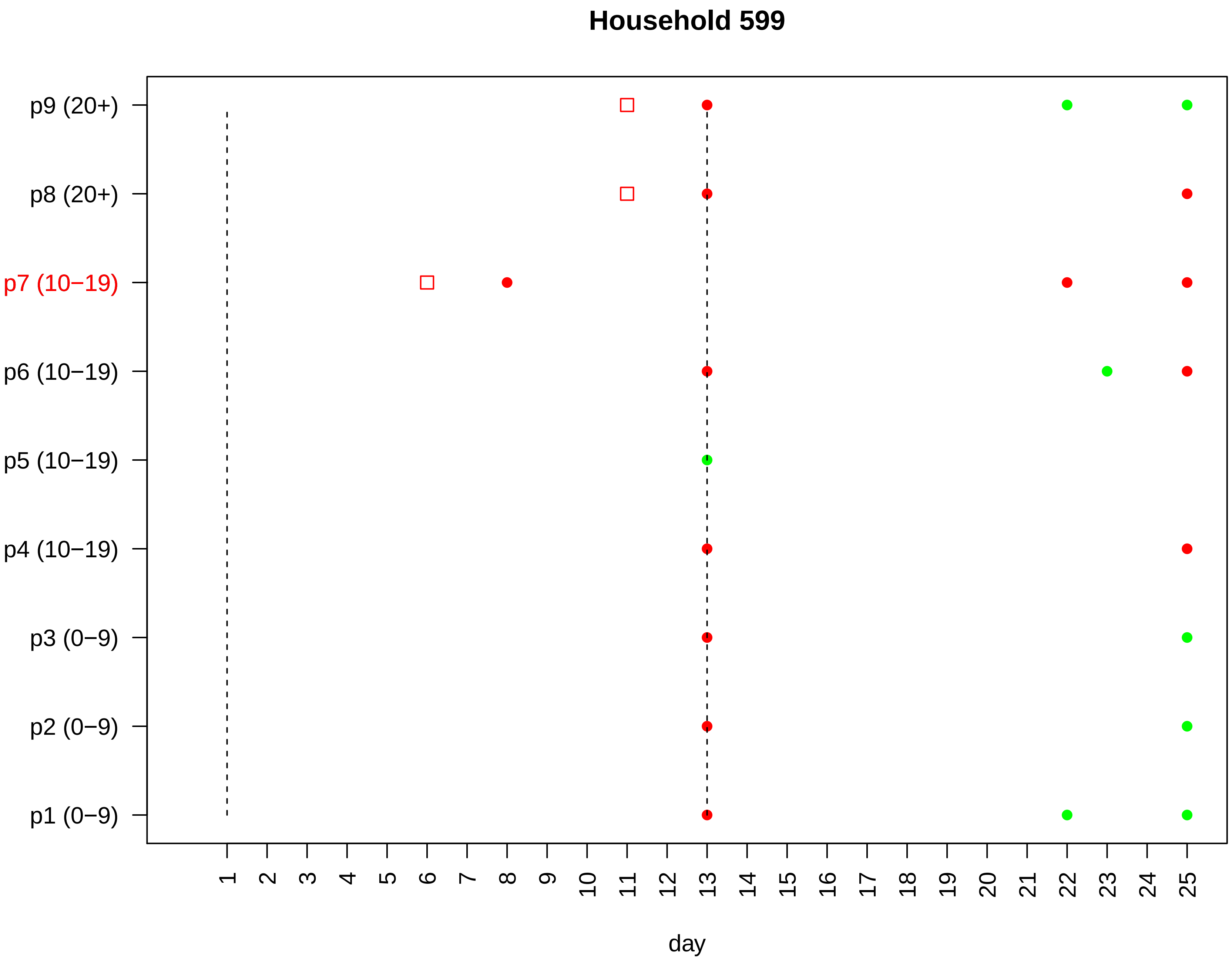

Household 599
p9 (20+)
p8 (20+)
p7 (10−19)
p6 (10−19)
p5 (10−19)
p4 (10−19)
p3 (0−9)
p2 (0−9)
p1 (0−9)
1 2 3 4 5 6 7 8 9 10 11 12 13 14 15 16 17 18 19 20 21 22 23 24 25
day

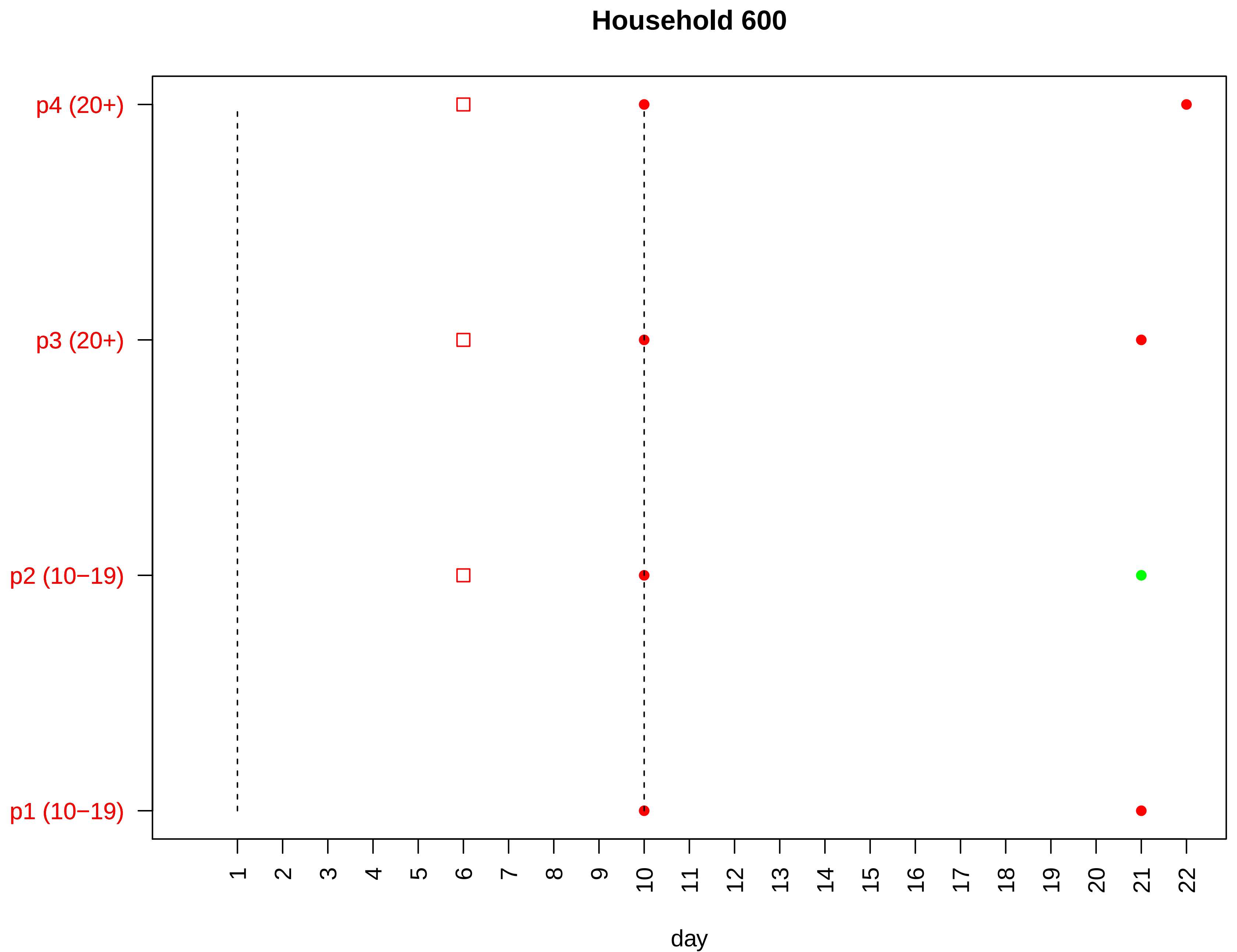
Household 600
p4 (20+)
p3 (20+)
p2 (10−19)
p1 (10−19)
day

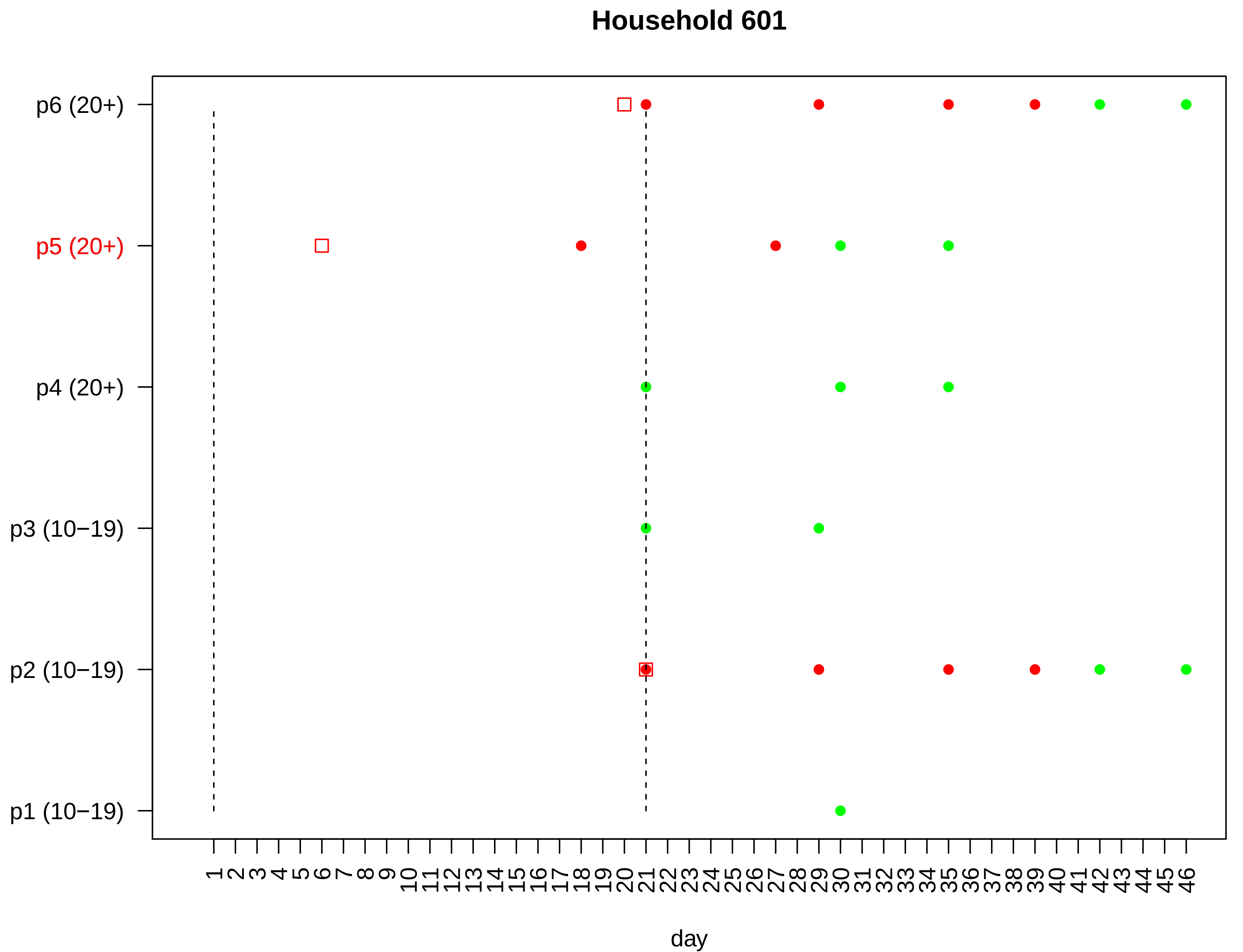
Household 601
p6 (20+)
p5 (20+)
p4 (20+)
p3 (10−19)
p2 (10−19)
p1 (10−19)
1 2 3 4 5 6 7 8 9 10 11 12 13 14 15 16 17 18 19 20 21 22 23 24 25 26 27 28 29 30 31 32 33 34 35 36 37 38 39 40 41 42 43 44 45 46
day

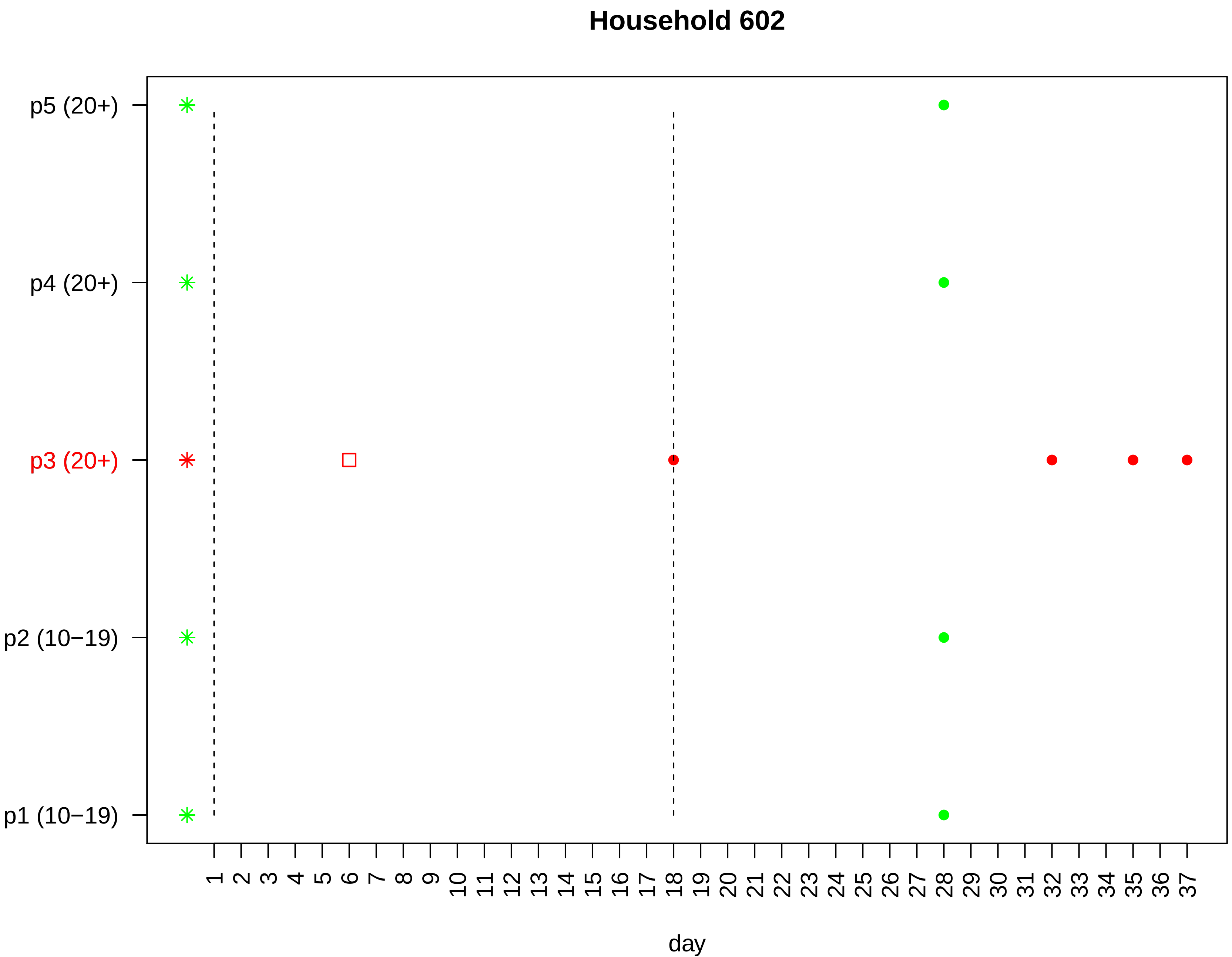

Household 602
p5 (20+)
p4 (20+)
p3 (20+)
p2 (10−19)
p1 (10−19)
1 2 3 4 5 6 7 8 9 10 11 12 13 14 15 16 17 18 19 20 21 22 23 24 25 26 27 28 29 30 31 32 33 34 35 36 37
day

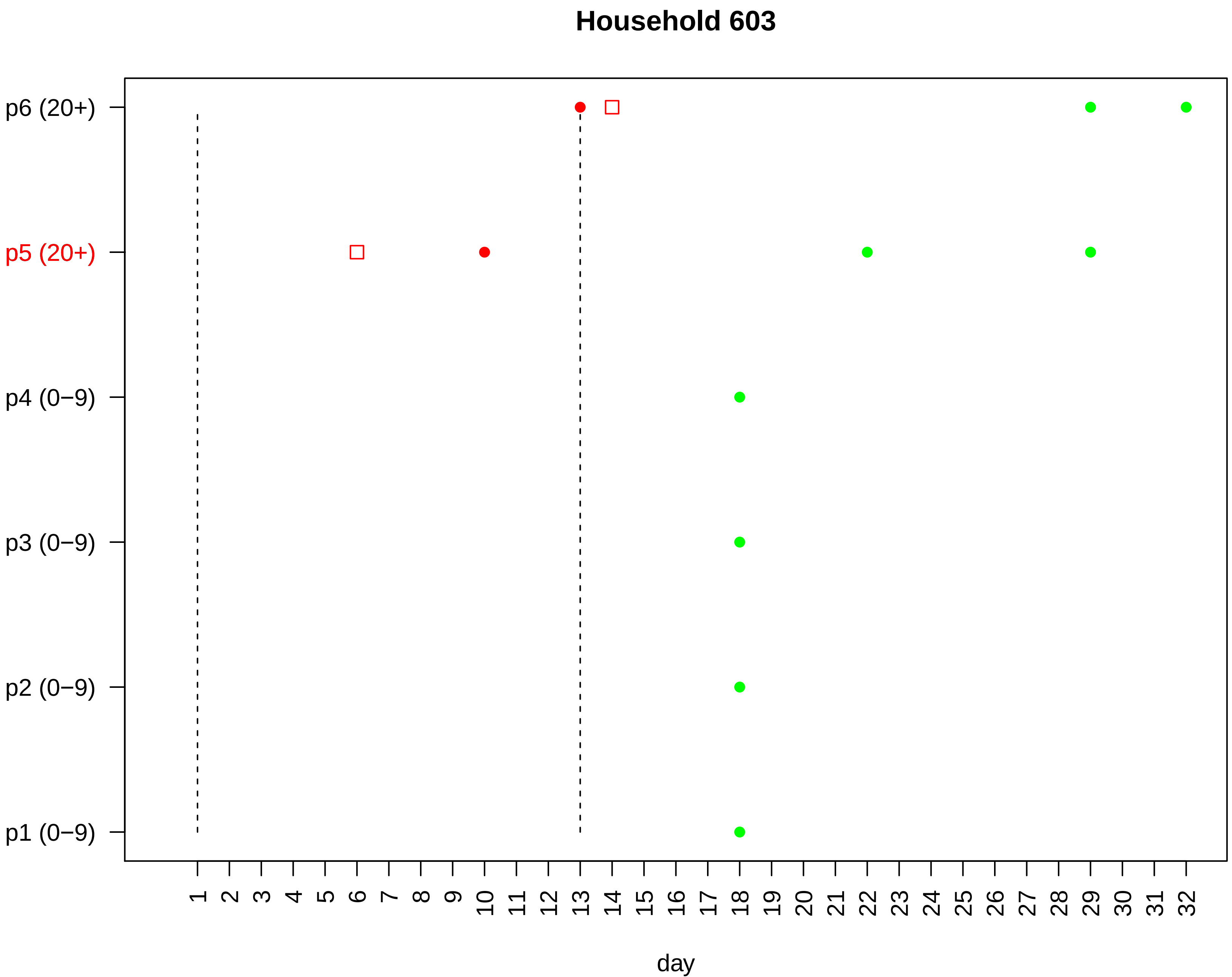
Household 603
p6 (20+)
p5 (20+)
p4 (0−9)
p3 (0−9)
p2 (0−9)
p1 (0−9)
1 2 3 4 5 6 7 8 9 10 11 12 13 14 15 16 17 18 19 20 21 22 23 24 25 26 27 28 29 30 31 32
day

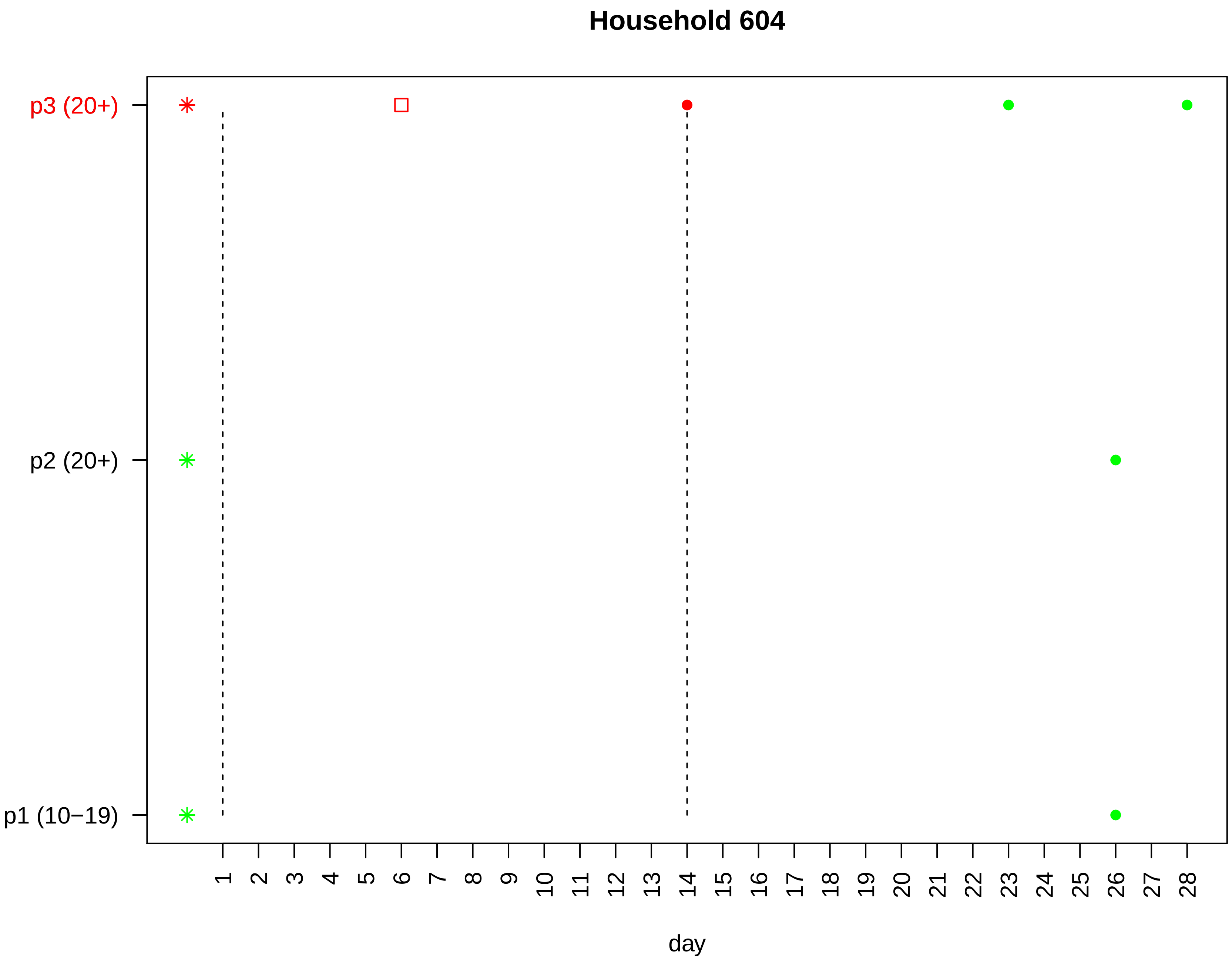

Household 604
p3 (20+)
p2 (20+)
p1 (10−19)
1 2 3 4 5 6 7 8 9 10 11 12 13 14 15 16 17 18 19 20 21 22 23 24 25 26 27 28
day

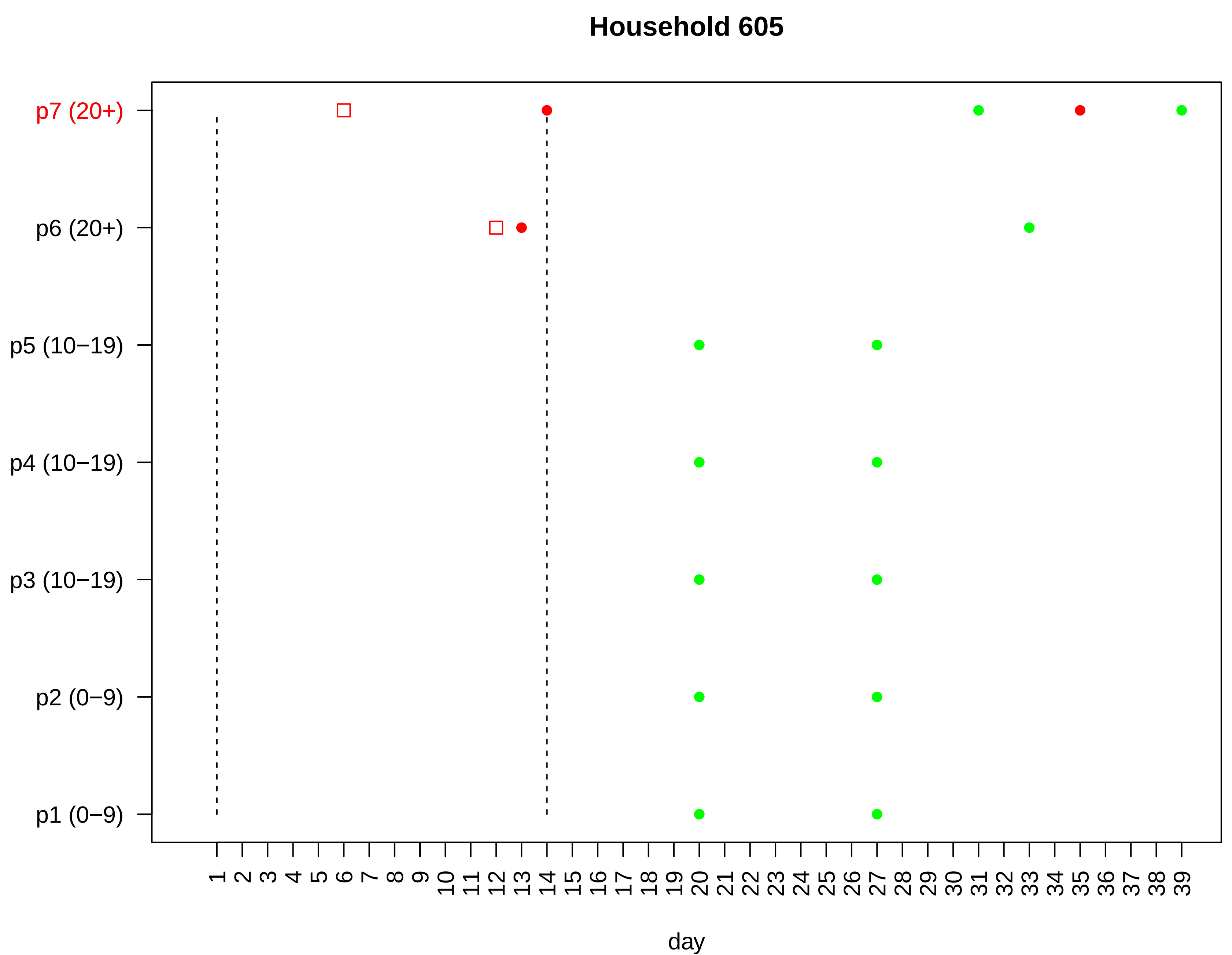

Household 605
p7 (20+)
p6 (20+)
p5 (10−19)
p4 (10−19)
p3 (10−19)
p2 (0−9)
p1 (0−9)
1 2 3 4 5 6 7 8 9 10 11 12 13 14 15 16 17 18 19 20 21 22 23 24 25 26 27 28 29 30 31 32 33 34 35 36 37 38 39
day

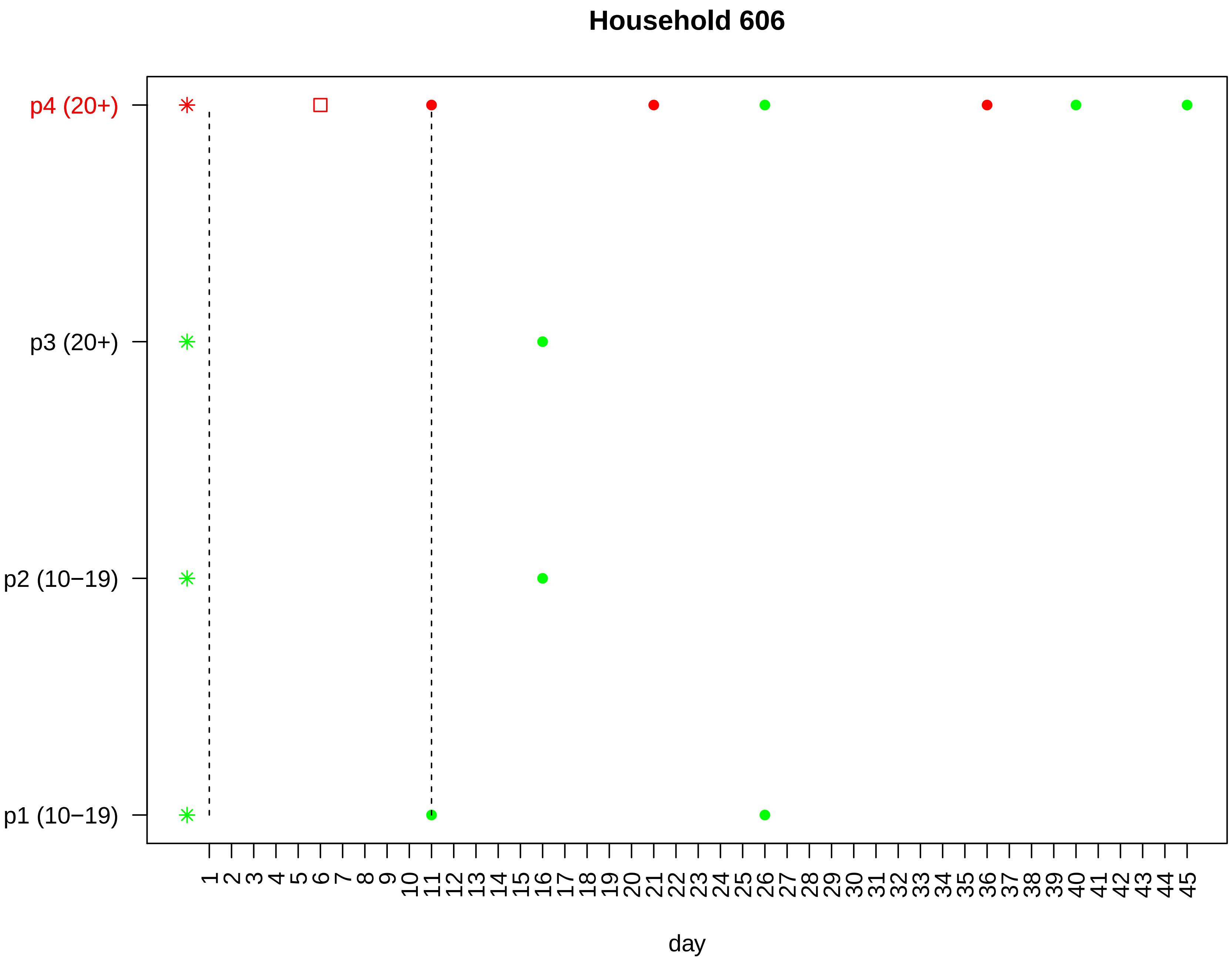

Household 606
p4 (20+)
p3 (20+)
p2 (10−19)
p1 (10−19)
1 2 3 4 5 6 7 8 9 10 11 12 13 14 15 16 17 18 19 20 21 22 23 24 25 26 27 28 29 30 31 32 33 34 35 36 37 38 39 40 41 42 43 44 45
day

### Household 607

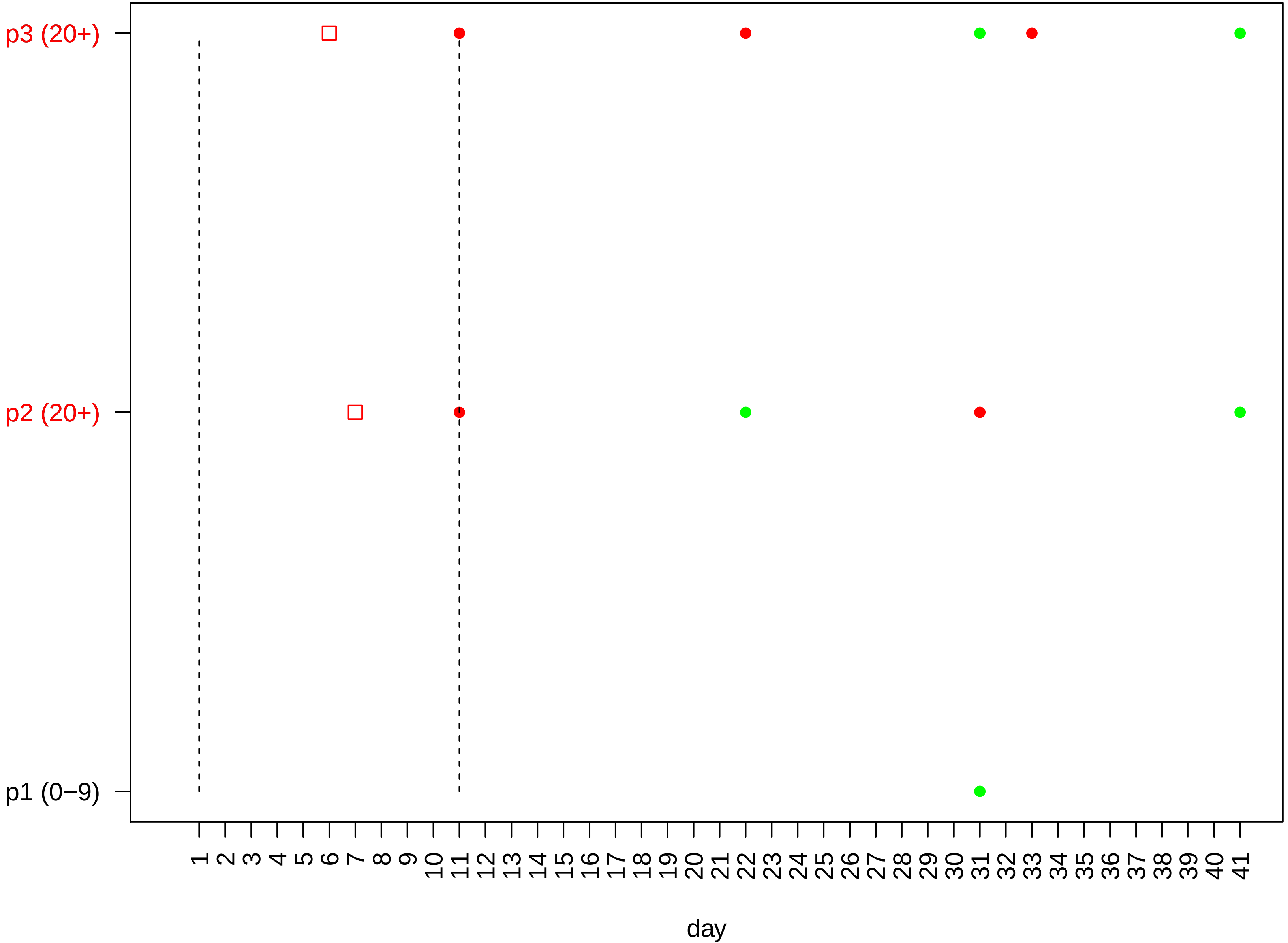

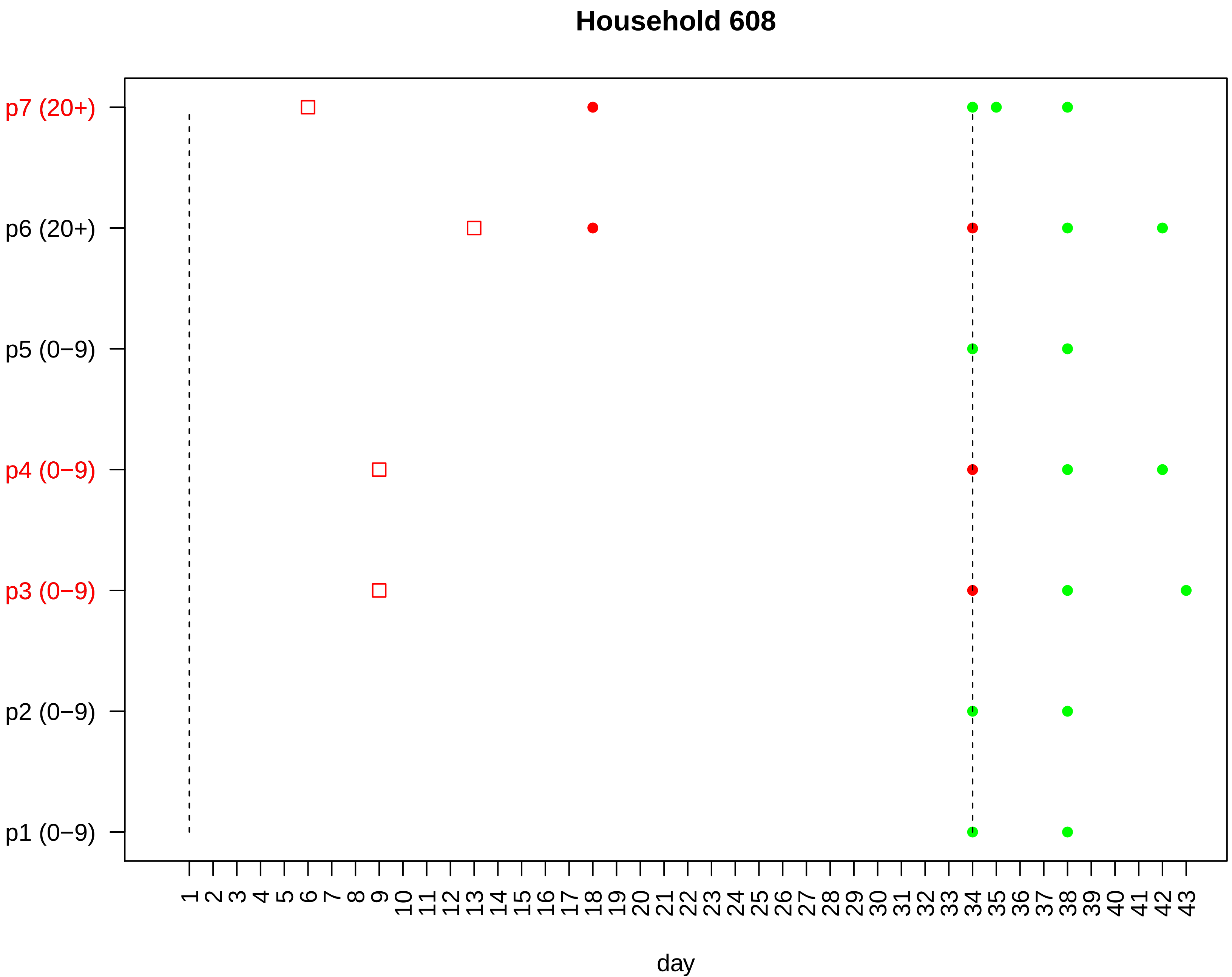

Household 608
p7 (20+)
p6 (20+)
p5 (0−9)
p4 (0−9)
p3 (0−9)
p2 (0−9)
p1 (0−9)
1 2 3 4 5 6 7 8 9 10 11 12 13 14 15 16 17 18 19 20 21 22 23 24 25 26 27 28 29 30 31 32 33 34 35 36 37 38 39 40 41 42 43
day

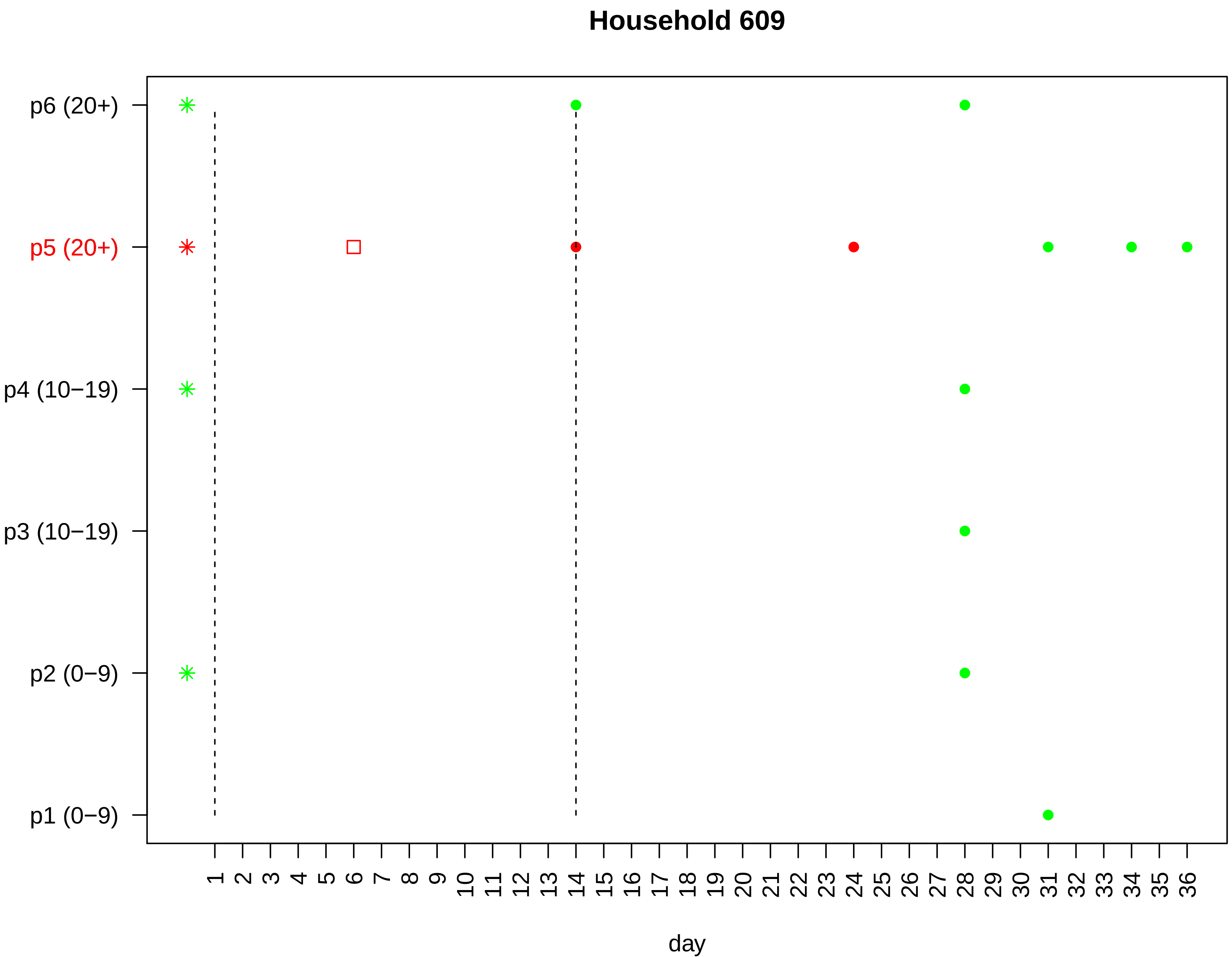

Household 609
p6 (20+)
p5 (20+)
p4 (10−19)
p3 (10−19)
p2 (0−9)
p1 (0−9)
1 2 3 4 5 6 7 8 9 10 11 12 13 14 15 16 17 18 19 20 21 22 23 24 25 26 27 28 29 30 31 32 33 34 35 36
day

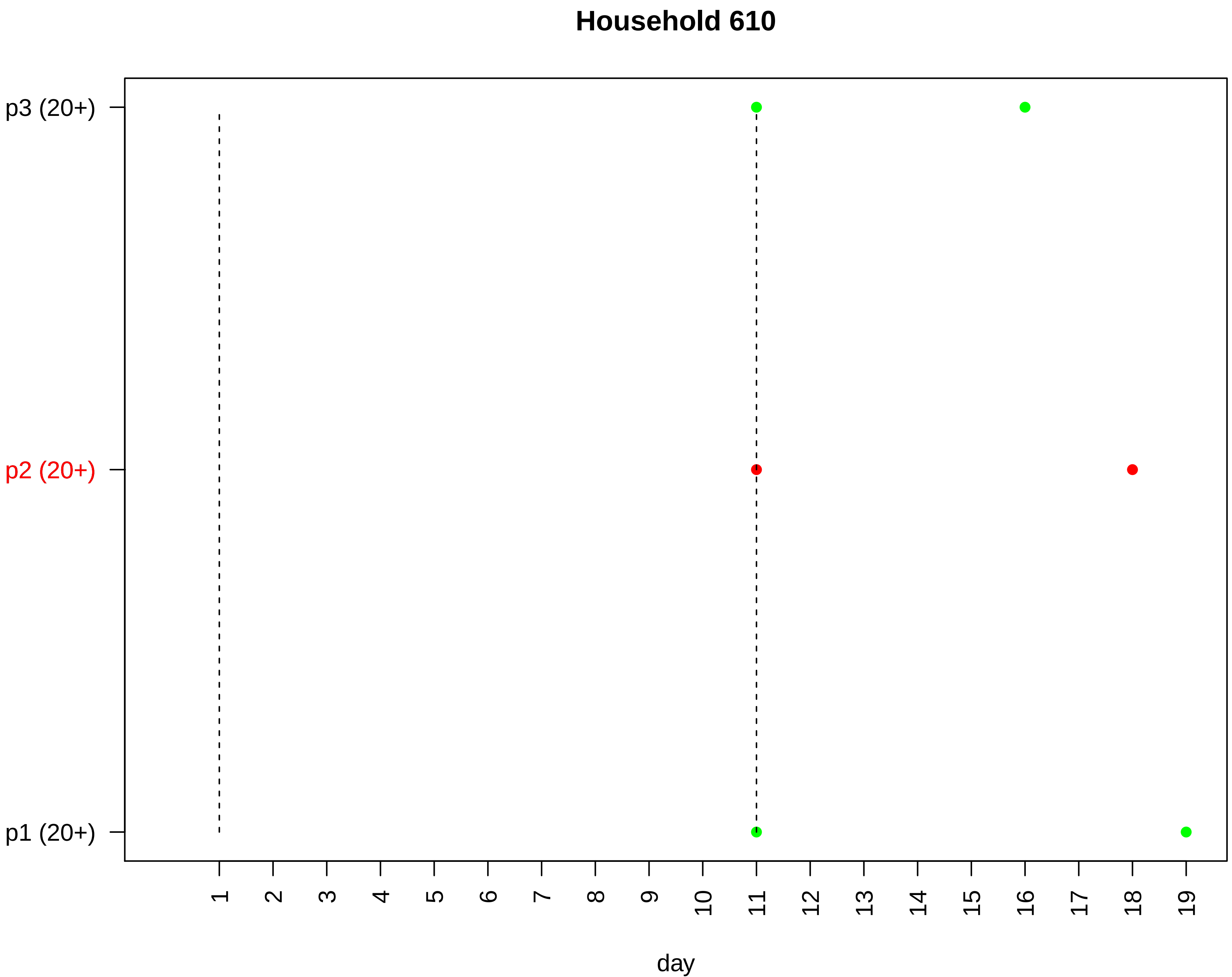

Household 610
p3 (20+)
p2 (20+)
p1 (20+)
day

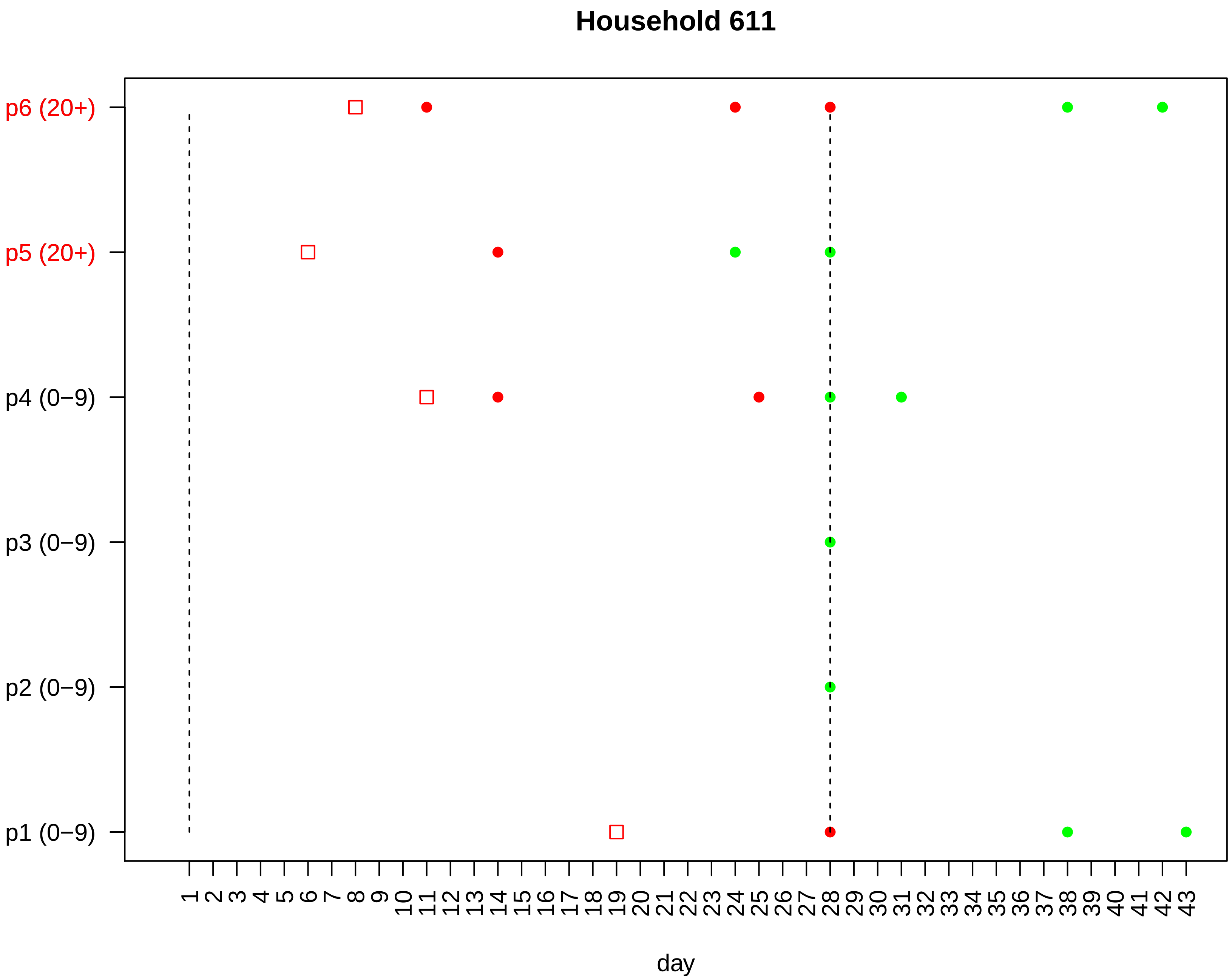

Household 611
p6 (20+)
p5 (20+)
p4 (0−9)
p3 (0−9)
p2 (0−9)
p1 (0−9)
day

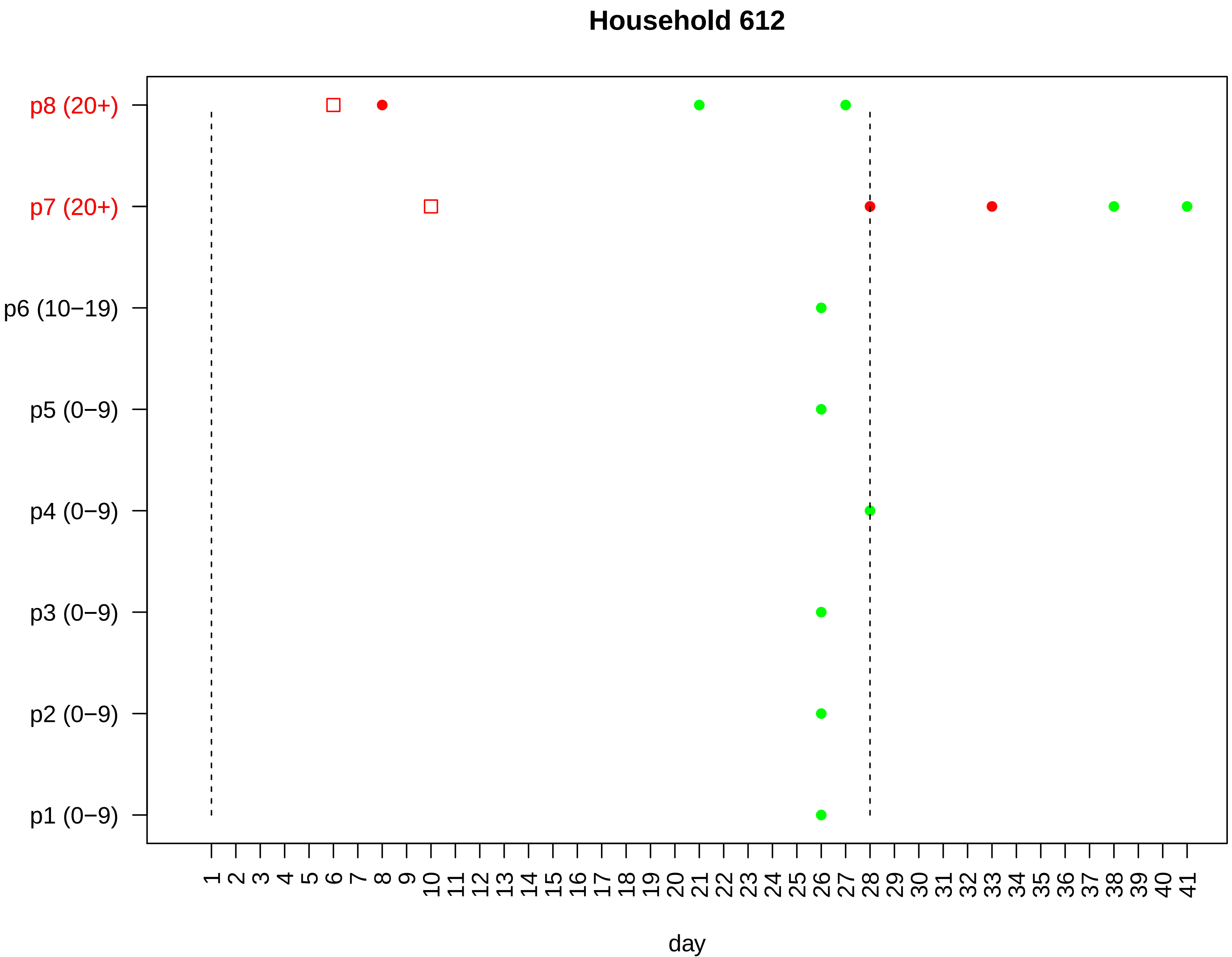
Household 612
p8 (20+)
p7 (20+)
p6 (10−19)
p5 (0−9)
p4 (0−9)
p3 (0−9)
p2 (0−9)
p1 (0−9)
1 2 3 4 5 6 7 8 9 10 11 12 13 14 15 16 17 18 19 20 21 22 23 24 25 26 27 28 29 30 31 32 33 34 35 36 37 38 39 40 41
day

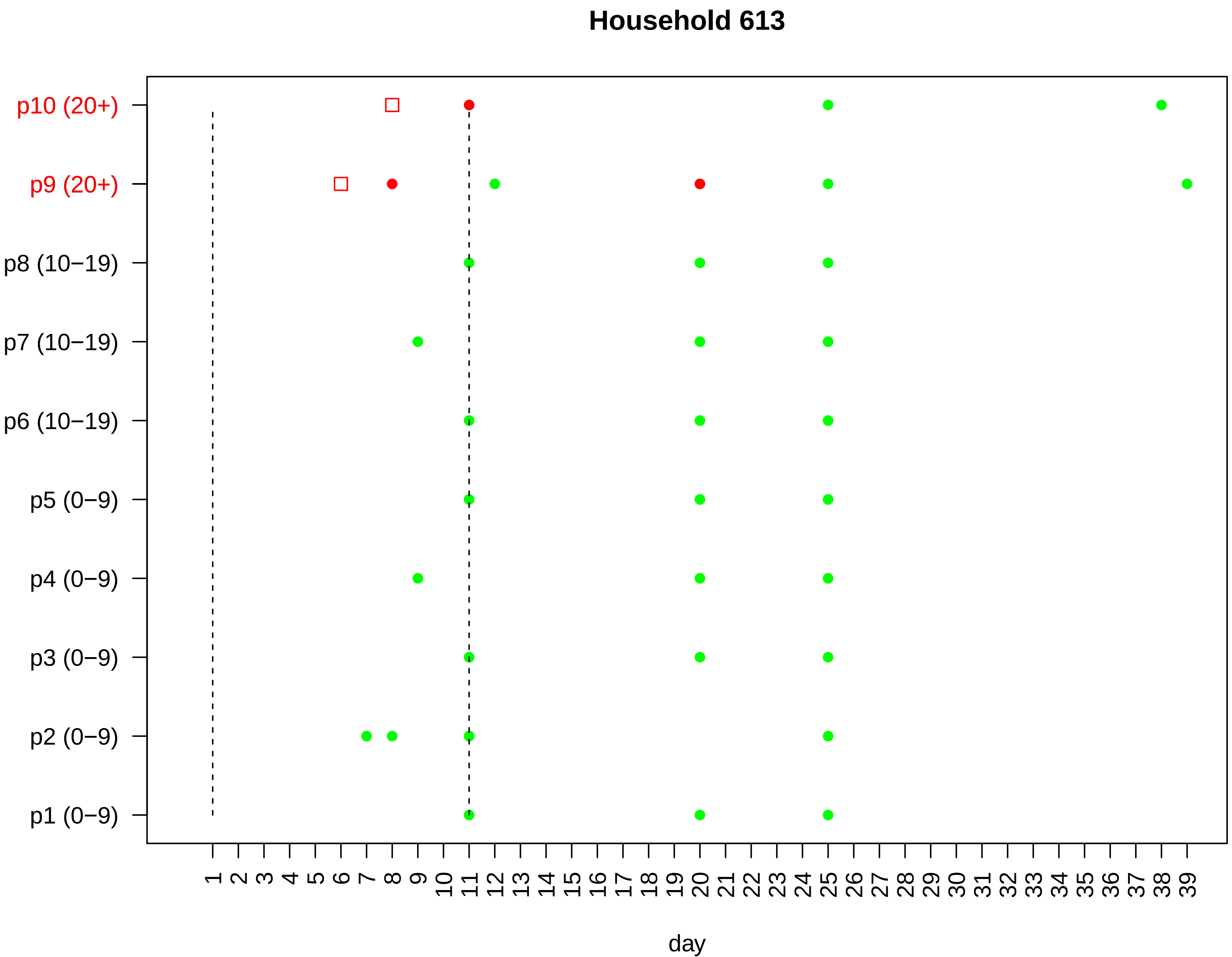
Household 613
p10 (20+)
p9 (20+)
p8 (10−19)
p7 (10−19)
p6 (10−19)
p5 (0−9)
p4 (0−9)
p3 (0−9)
p2 (0−9)
p1 (0−9)
1 2 3 4 5 6 7 8 9 10 11 12 13 14 15 16 17 18 19 20 21 22 23 24 25 26 27 28 29 30 31 32 33 34 35 36 37 38 39
day

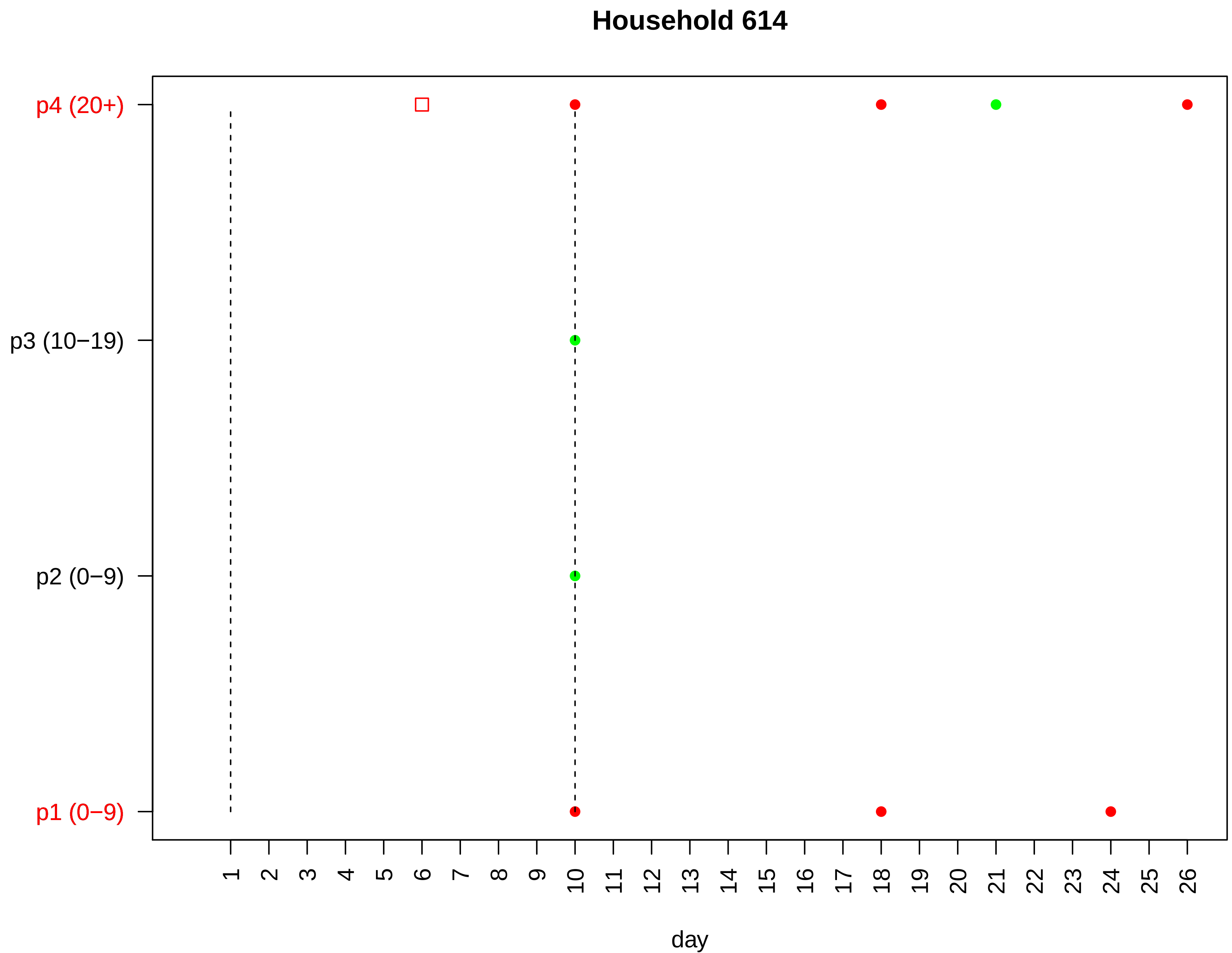

Household 614
p4 (20+)
p3 (10−19)
p2 (0−9)
p1 (0−9)
day

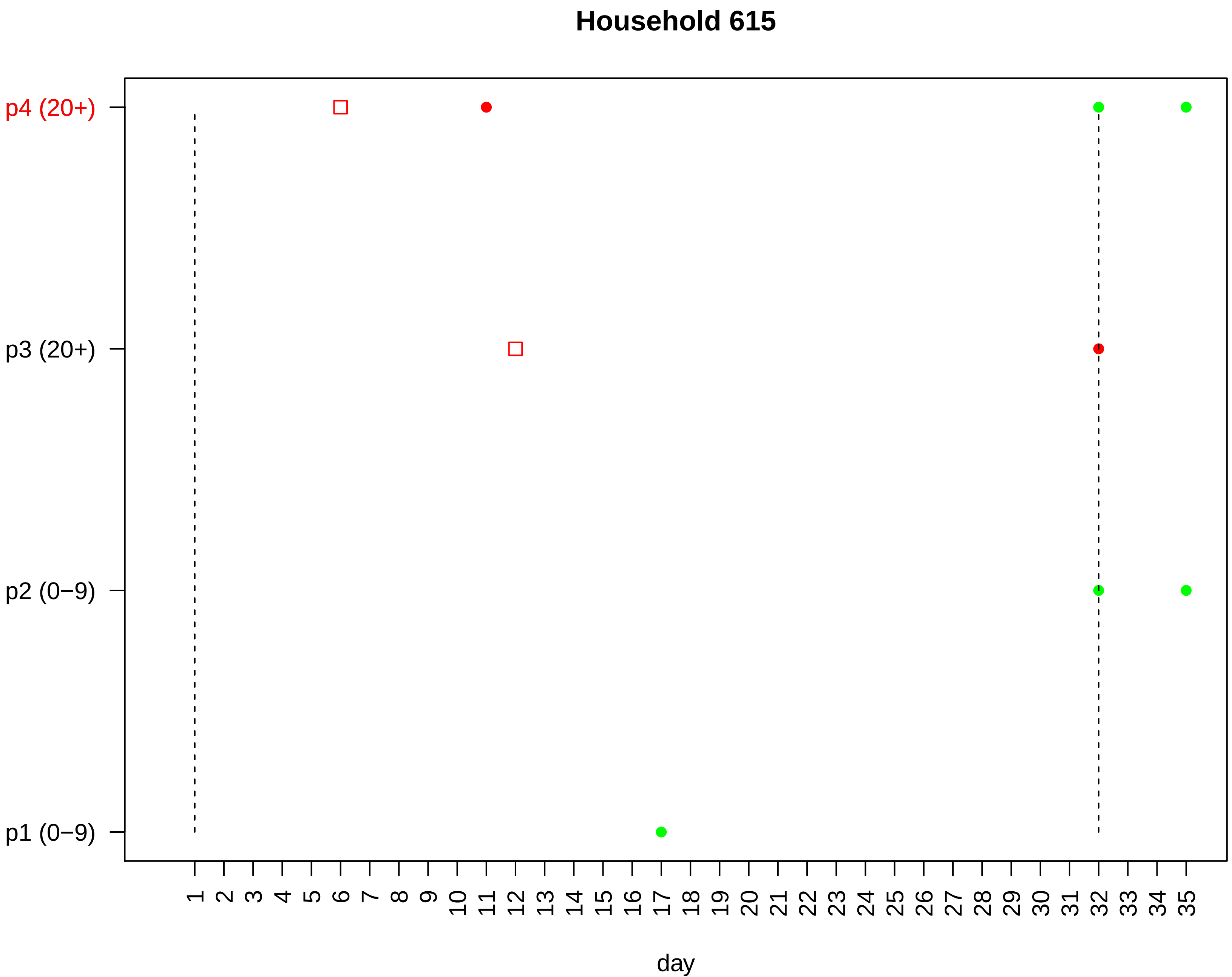

Household 615
p4 (20+)
p3 (20+)
p2 (0−9)
p1 (0−9)
1 2 3 4 5 6 7 8 9 10 11 12 13 14 15 16 17 18 19 20 21 22 23 24 25 26 27 28 29 30 31 32 33 34 35
day

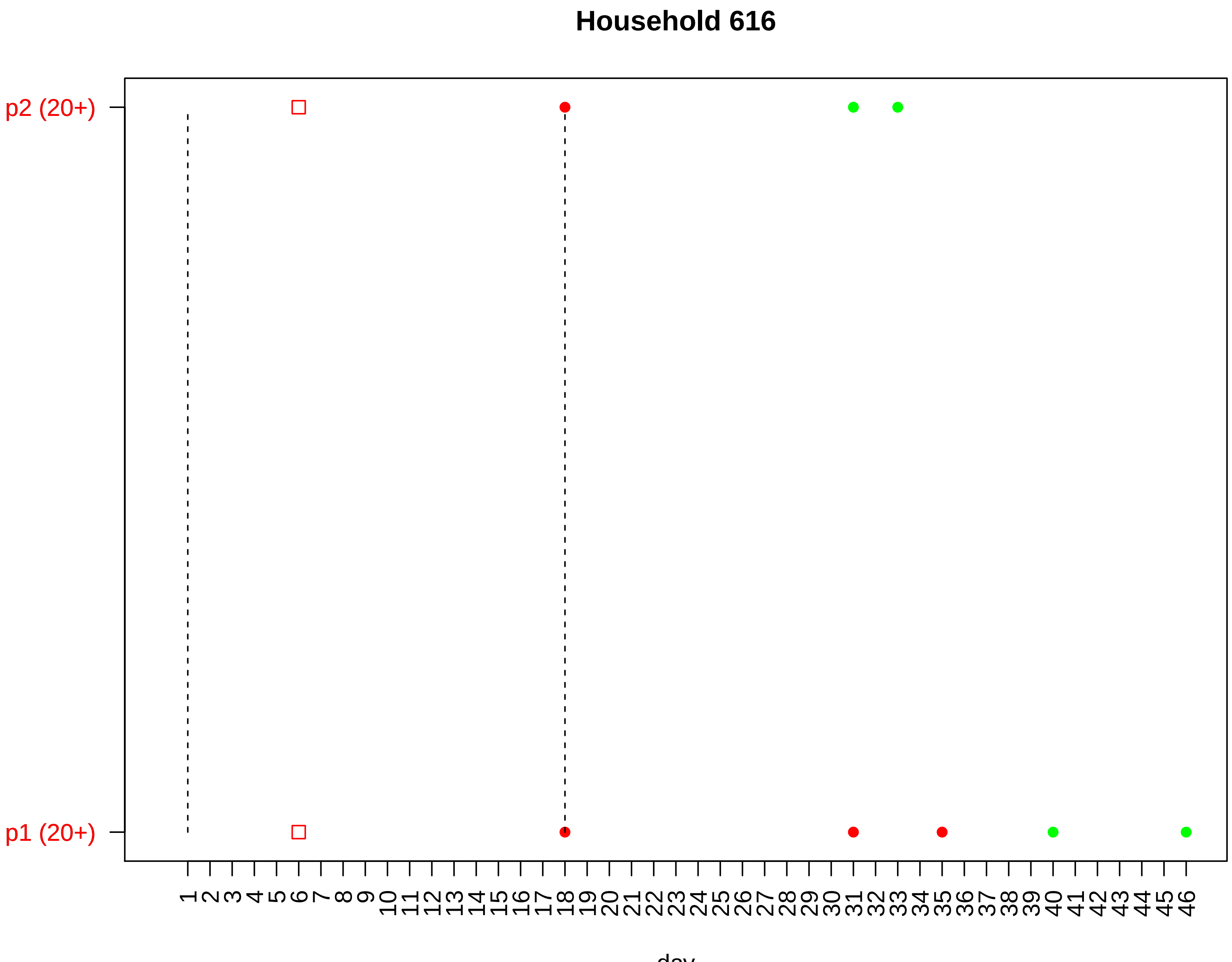

Household 616
p2 (20+)
p1 (20+)
1 2 3 4 5 6 7 8 9 10 11 12 13 14 15 16 17 18 19 20 21 22 23 24 25 26 27 28 29 30 31 32 33 34 35 36 37 38 39 40 41 42 43 44 45 46
day

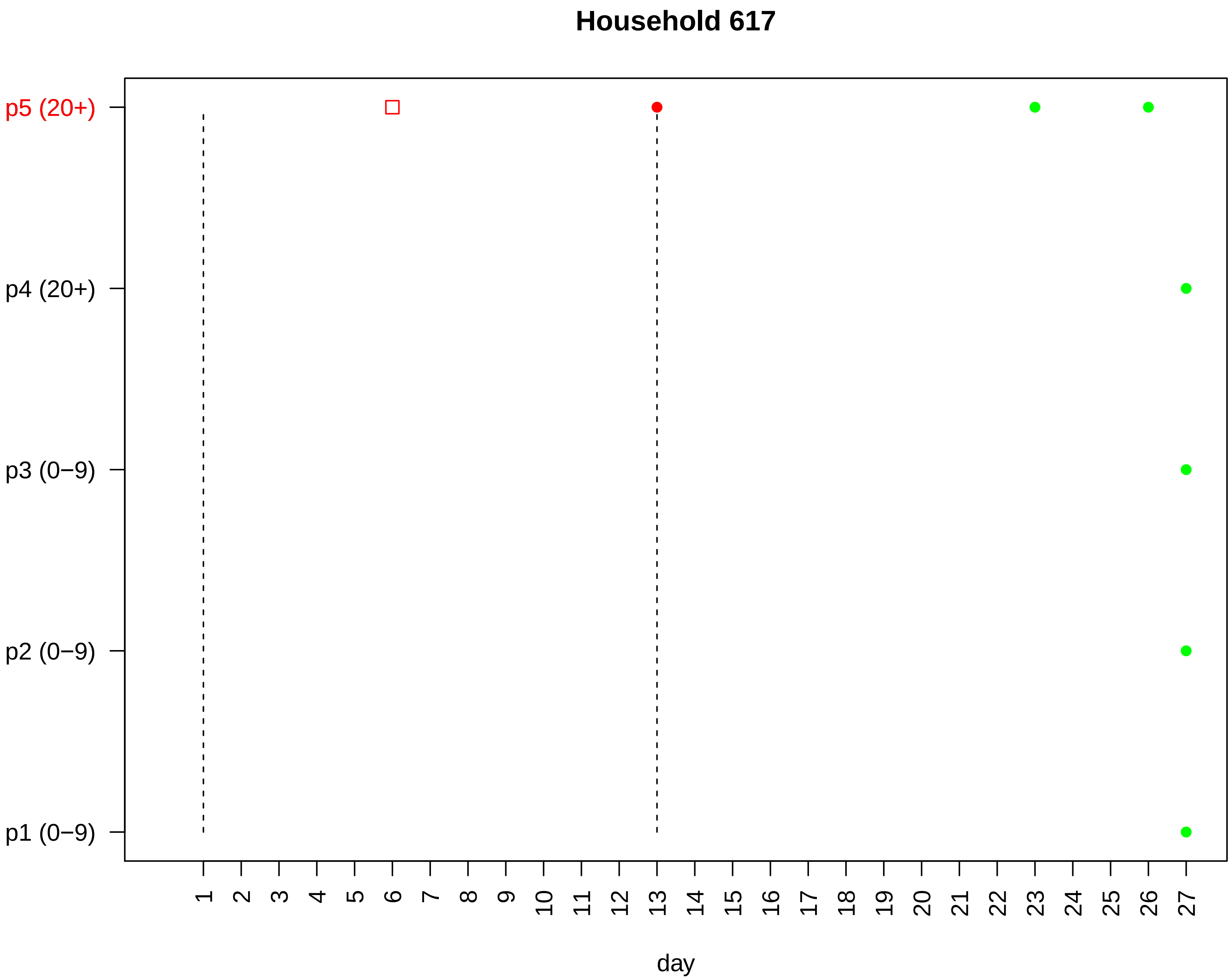

Household 617
p5 (20+)
p4 (20+)
p3 (0−9)
p2 (0−9)
p1 (0−9)
day

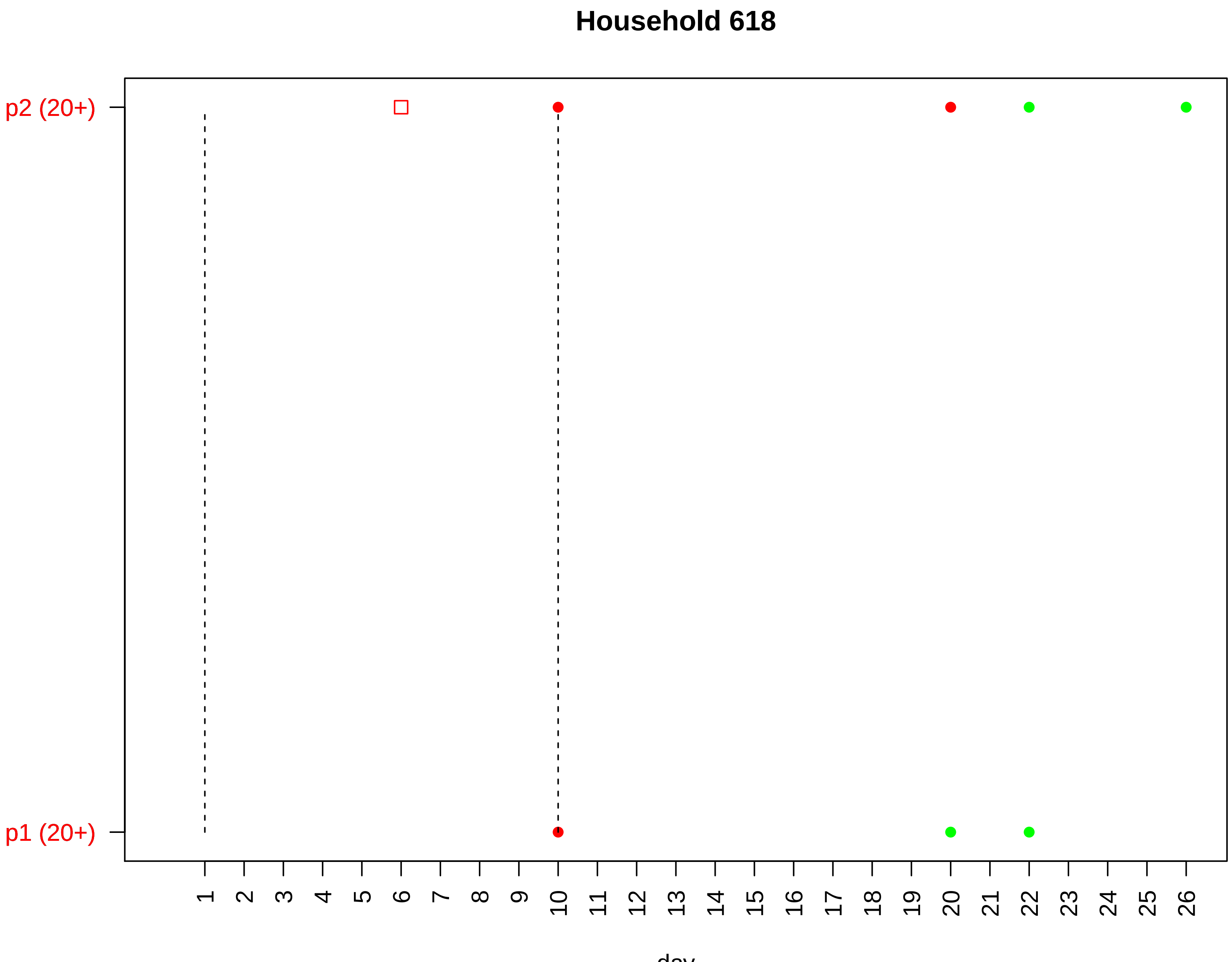

Household 618
p2 (20+)
p1 (20+)
1 2 3 4 5 6 7 8 9 10 11 12 13 14 15 16 17 18 19 20 21 22 23 24 25 26
day

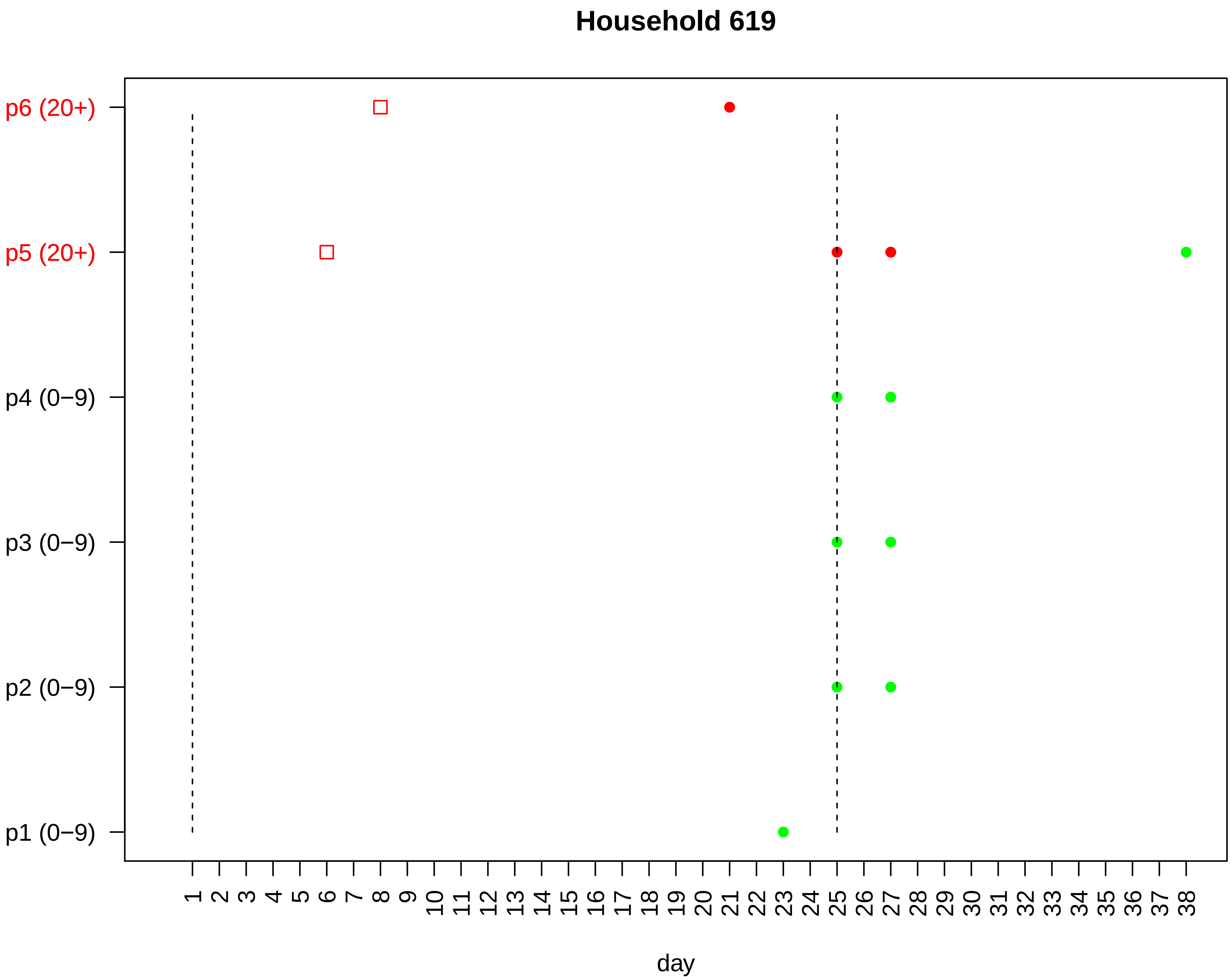
Household 619
p6 (20+)
p5 (20+)
p4 (0−9)
p3 (0−9)
p2 (0−9)
p1 (0−9)
1 2 3 4 5 6 7 8 9 10 11 12 13 14 15 16 17 18 19 20 21 22 23 24 25 26 27 28 29 30 31 32 33 34 35 36 37 38
day

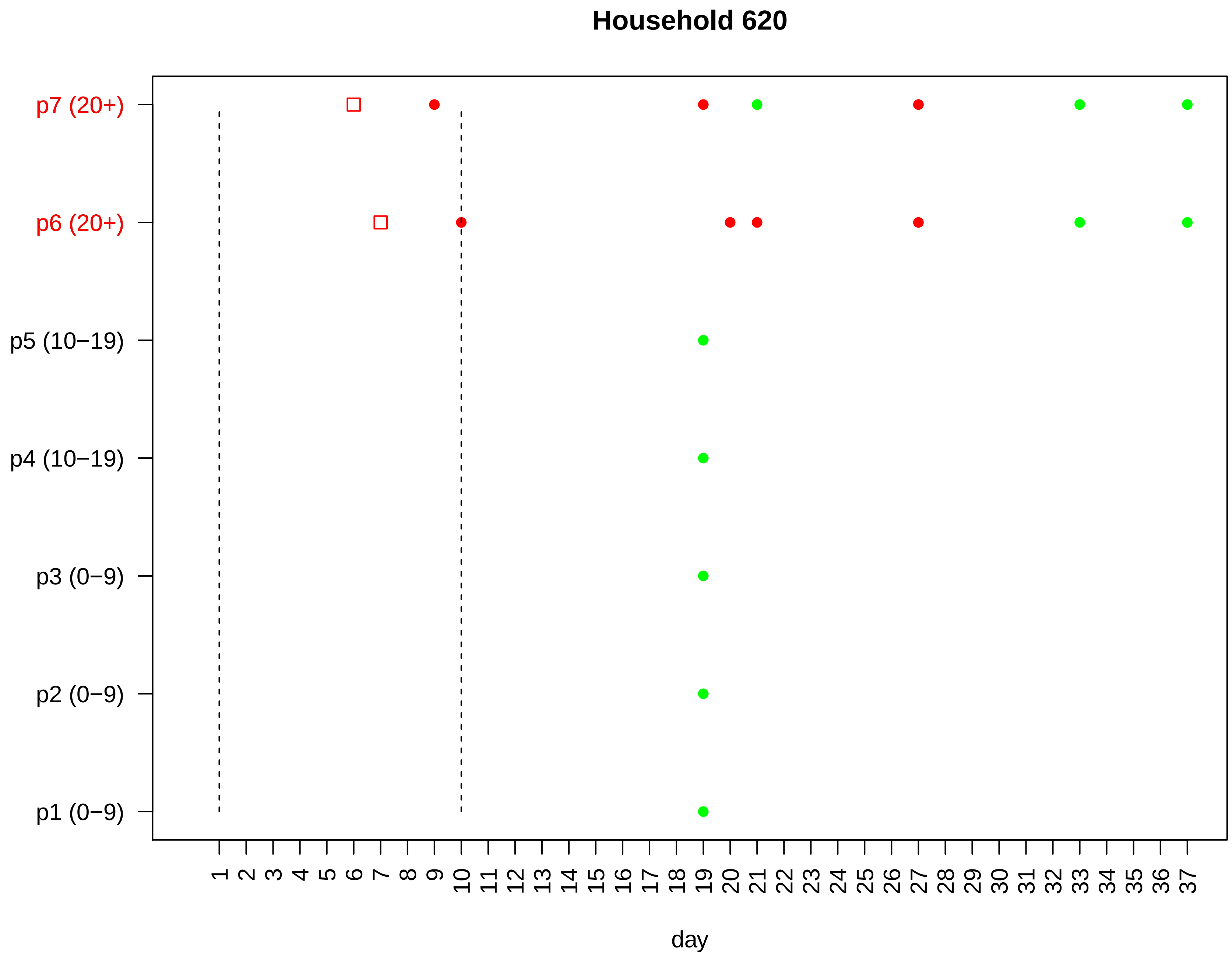

Household 620
p7 (20+)
p6 (20+)
p5 (10−19)
p4 (10−19)
p3 (0−9)
p2 (0−9)
p1 (0−9)
1 2 3 4 5 6 7 8 9 10 11 12 13 14 15 16 17 18 19 20 21 22 23 24 25 26 27 28 29 30 31 32 33 34 35 36 37
day

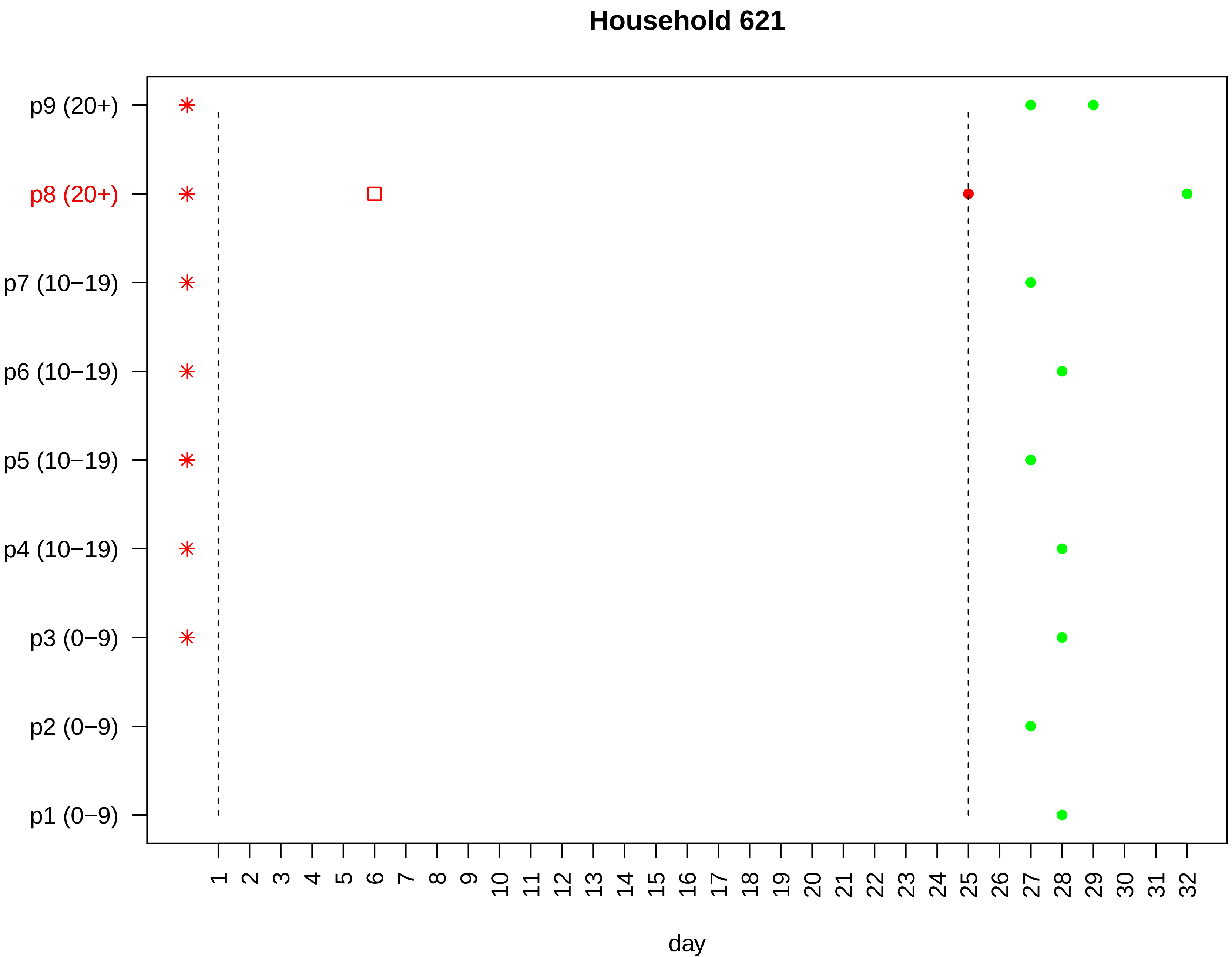

Household 621
p9 (20+)
p8 (20+)
p7 (10−19)
p6 (10−19)
p5 (10−19)
p4 (10−19)
p3 (0−9)
p2 (0−9)
p1 (0−9)
1 2 3 4 5 6 7 8 9 10 11 12 13 14 15 16 17 18 19 20 21 22 23 24 25 26 27 28 29 30 31 32
day

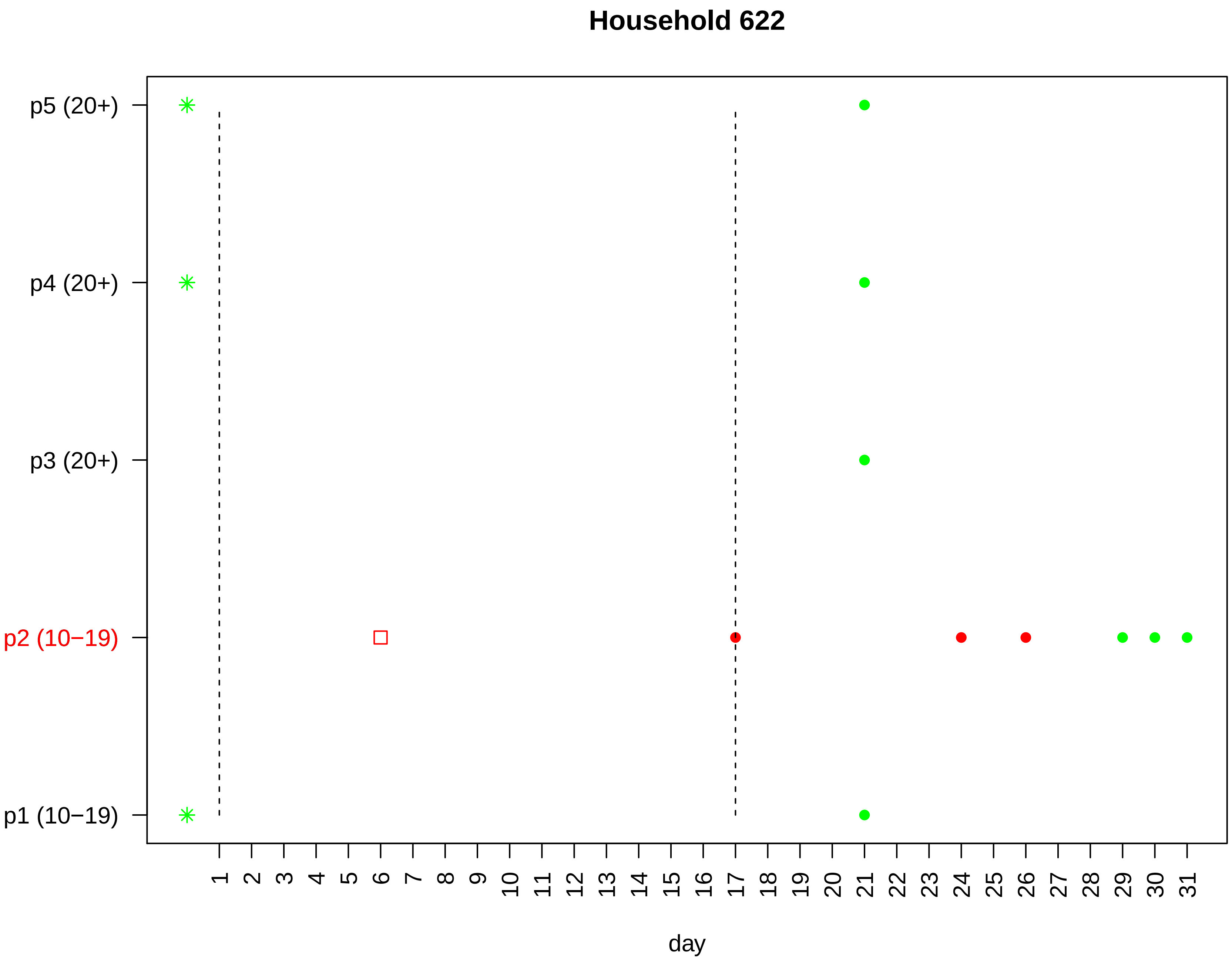

Household 622
p5 (20+)
p4 (20+)
p3 (20+)
p2 (10−19)
p1 (10−19)
day

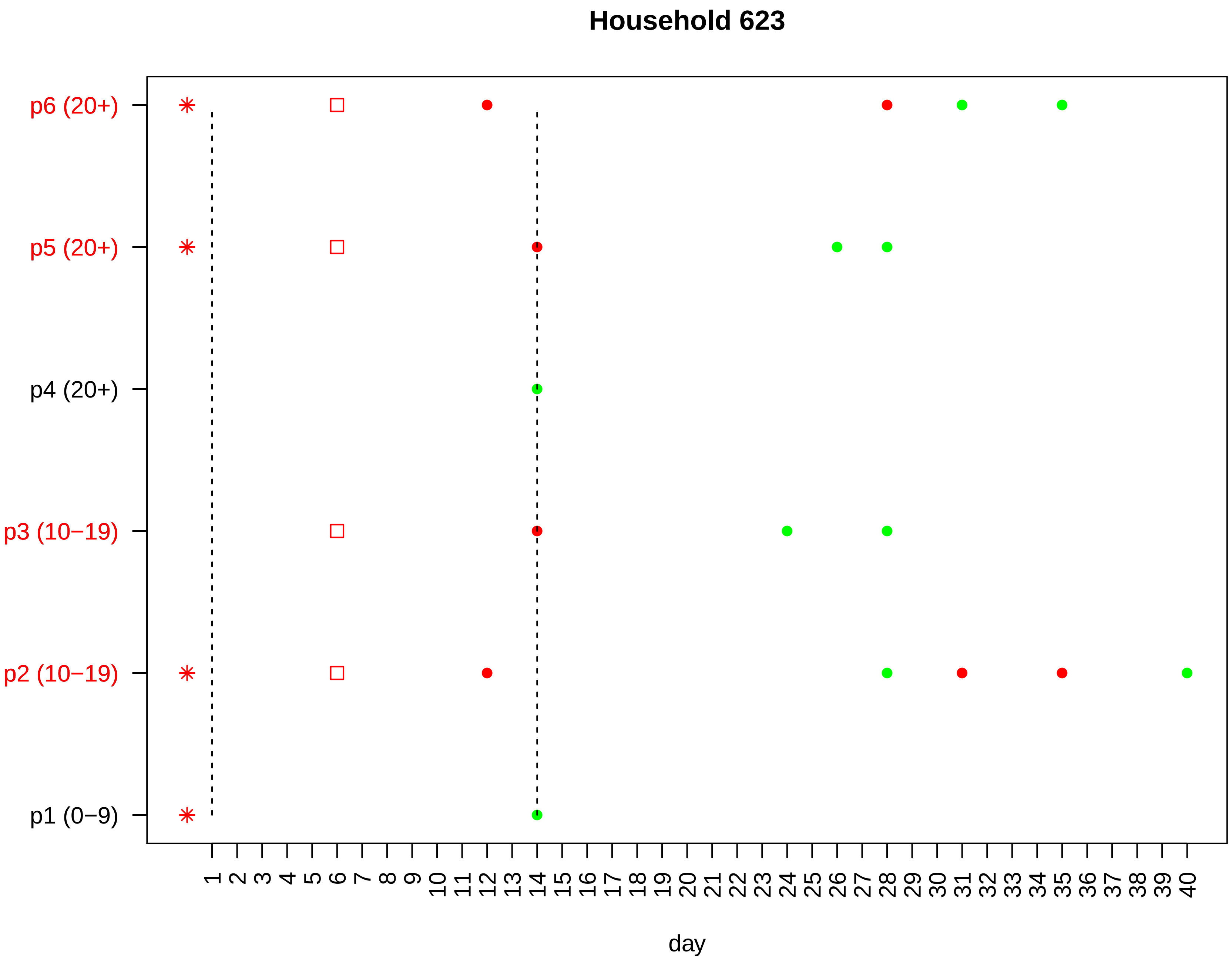

Household 623
p6 (20+)
p5 (20+)
p4 (20+)
p3 (10−19)
p2 (10−19)
p1 (0−9)
1 2 3 4 5 6 7 8 9 10 11 12 13 14 15 16 17 18 19 20 21 22 23 24 25 26 27 28 29 30 31 32 33 34 35 36 37 38 39 40
day

#### Household 624

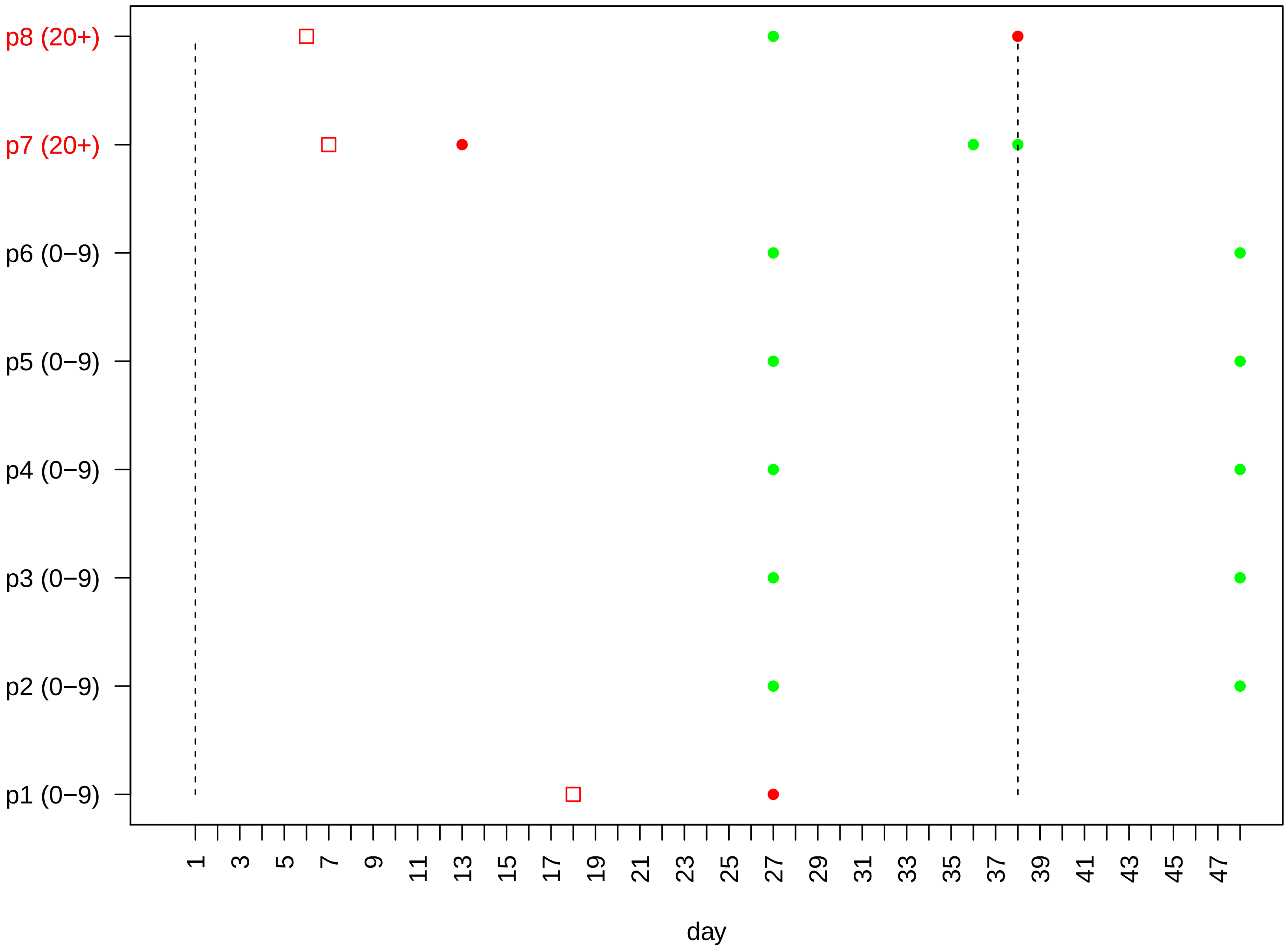

### Household 625

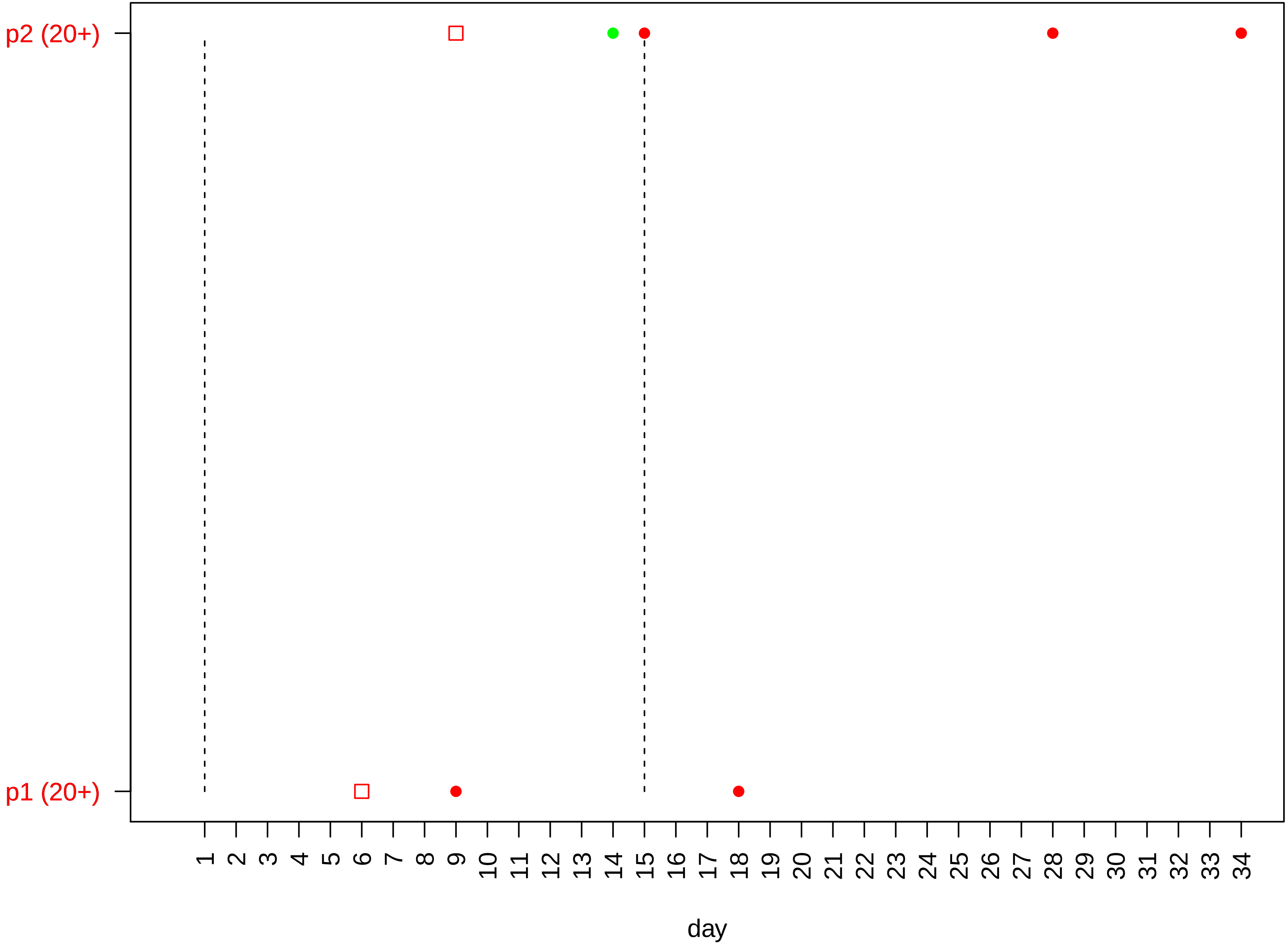

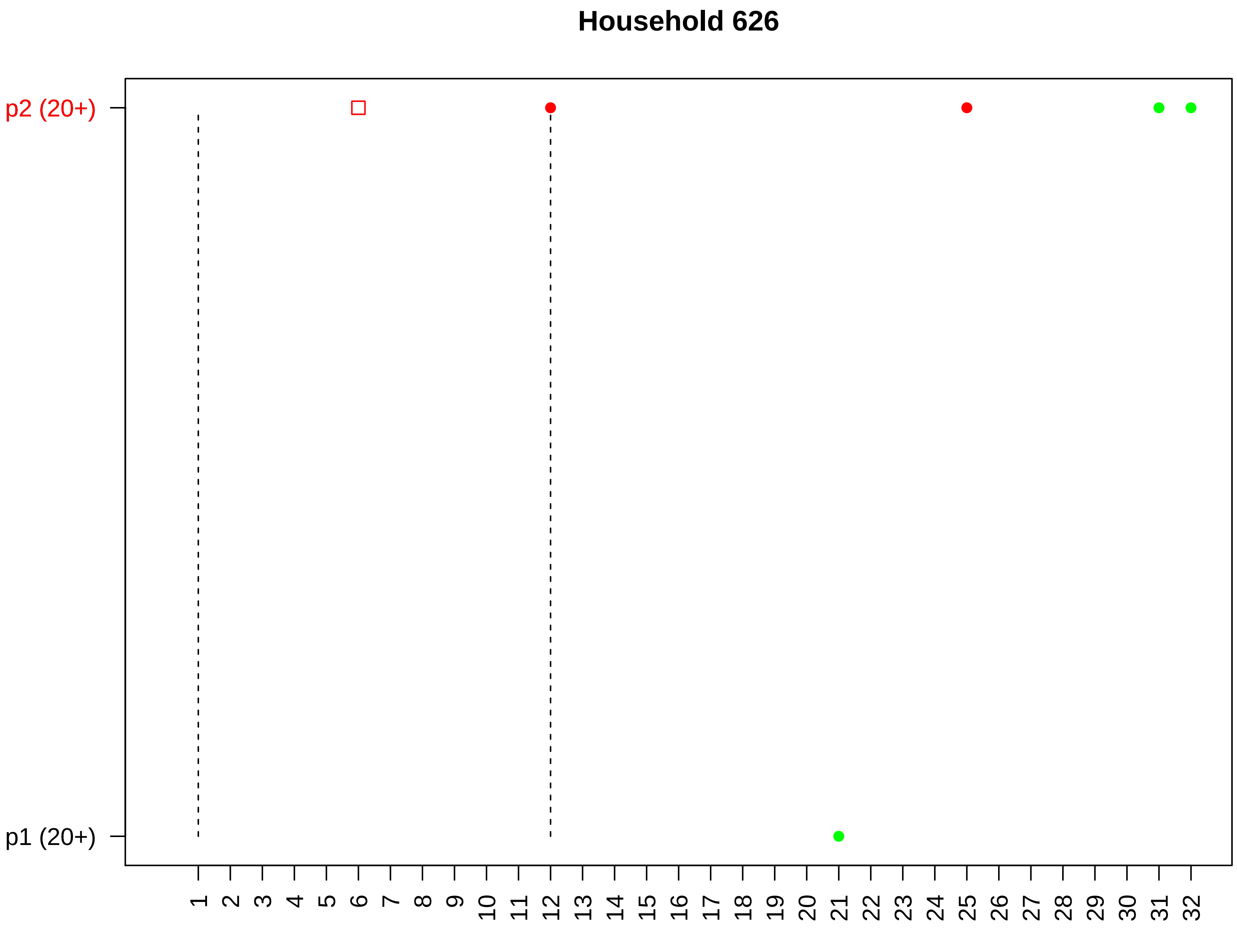

Household 626
p2 (20+)
p1 (20+)
1 2 3 4 5 6 7 8 9 10 11 12 13 14 15 16 17 18 19 20 21 22 23 24 25 26 27 28 29 30 31 32
day

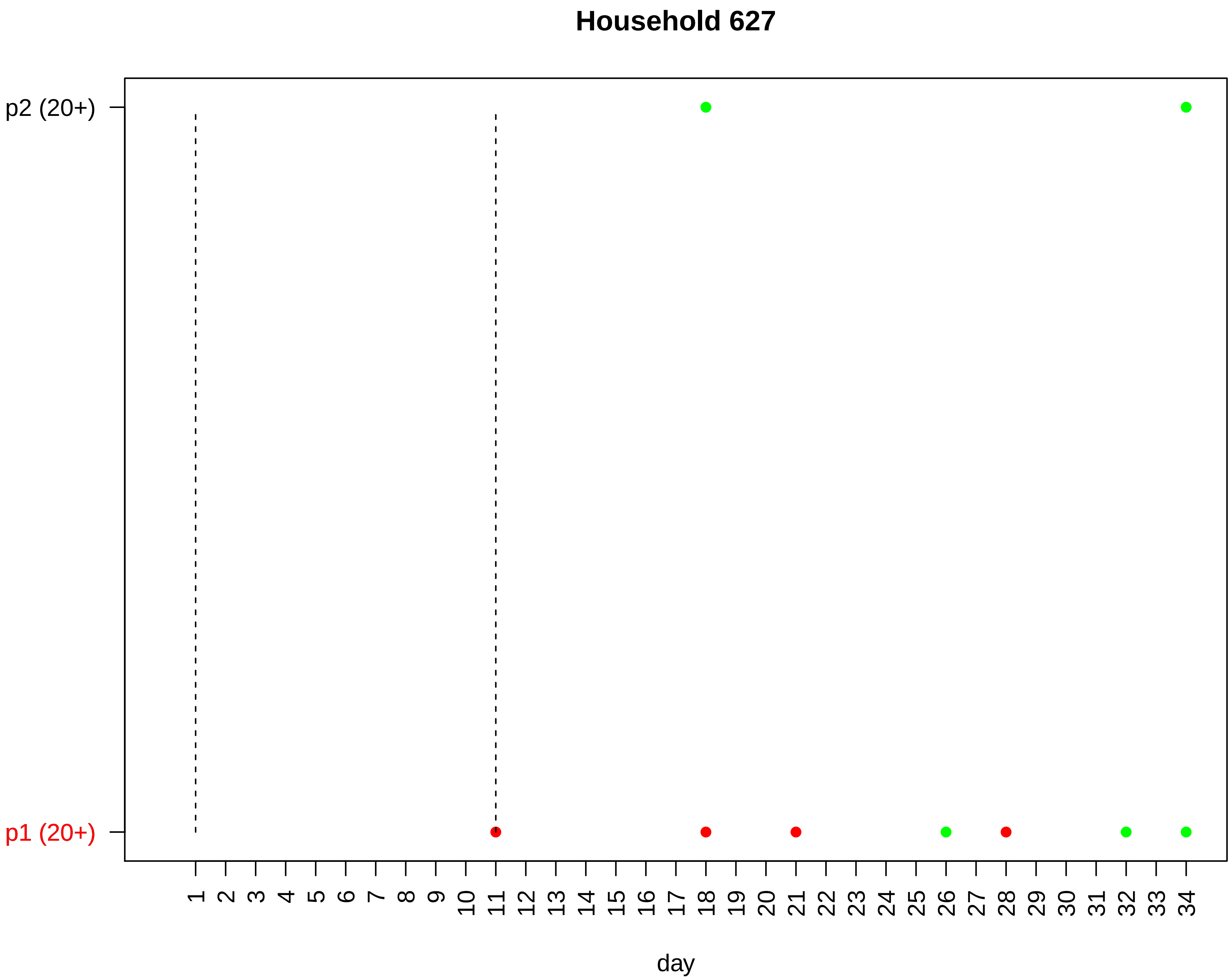
Household 627
p2 (20+)
p1 (20+)
1 2 3 4 5 6 7 8 9 10 11 12 13 14 15 16 17 18 19 20 21 22 23 24 25 26 27 28 29 30 31 32 33 34
day

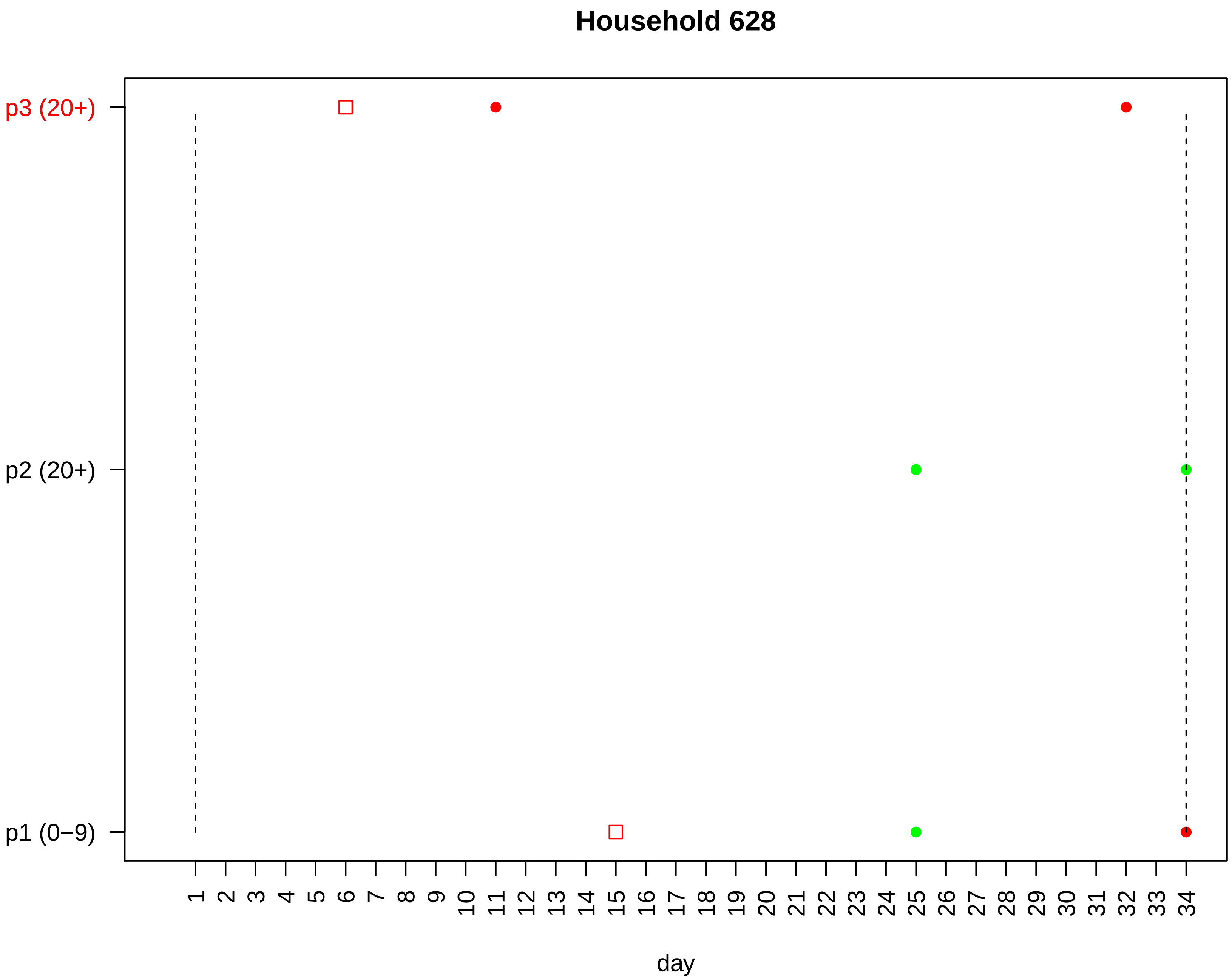
Household 628
p3 (20+)
p2 (20+)
p1 (0−9)
1 2 3 4 5 6 7 8 9 10 11 12 13 14 15 16 17 18 19 20 21 22 23 24 25 26 27 28 29 30 31 32 33 34
day

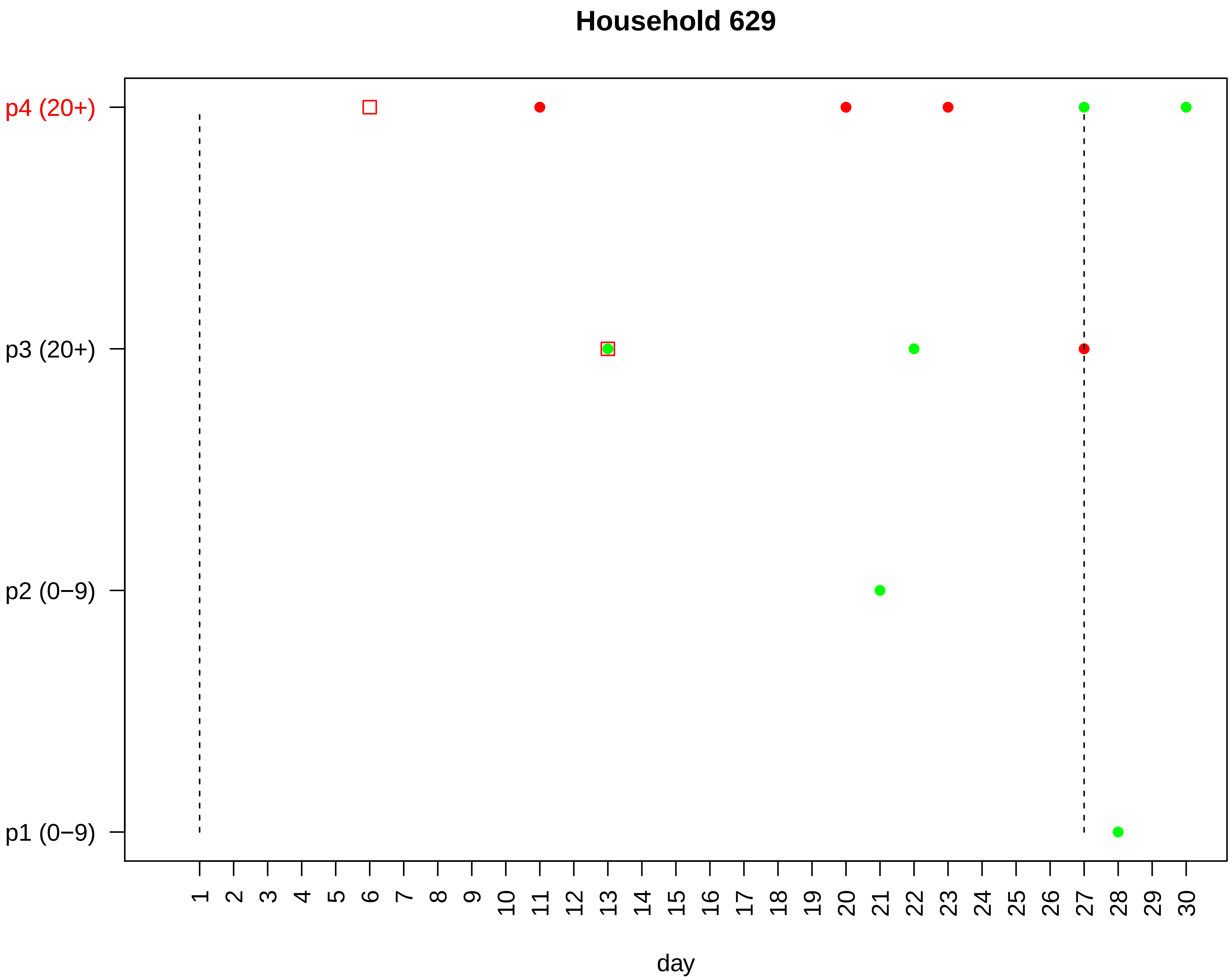
Household 629
p4 (20+)
p3 (20+)
p2 (0−9)
p1 (0−9)
1 2 3 4 5 6 7 8 9 10 11 12 13 14 15 16 17 18 19 20 21 22 23 24 25 26 27 28 29 30
day

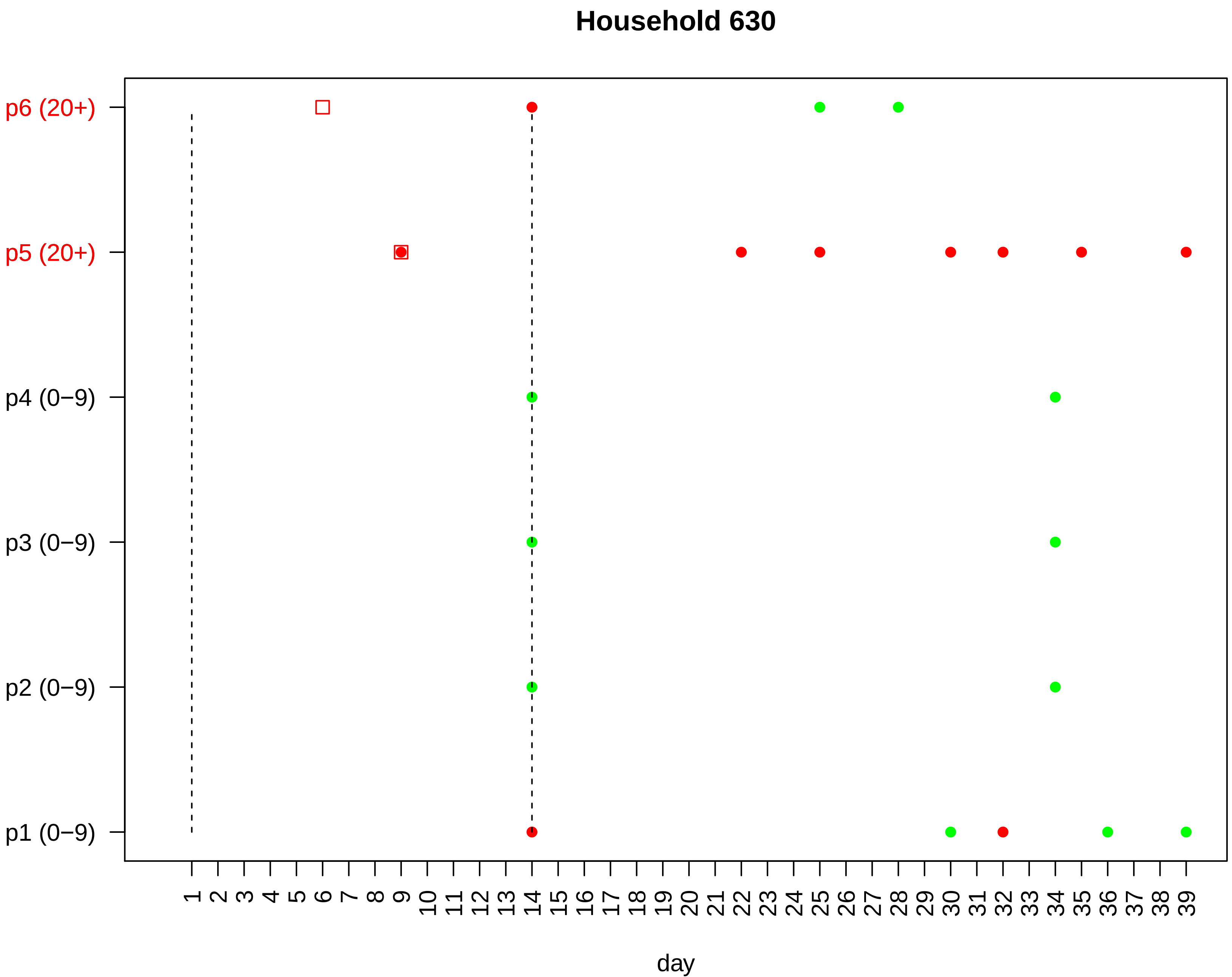
Household 630
p6 (20+)
p5 (20+)
p4 (0−9)
p3 (0−9)
p2 (0−9)
p1 (0−9)
1 2 3 4 5 6 7 8 9 10 11 12 13 14 15 16 17 18 19 20 21 22 23 24 25 26 27 28 29 30 31 32 33 34 35 36 37 38 39
day

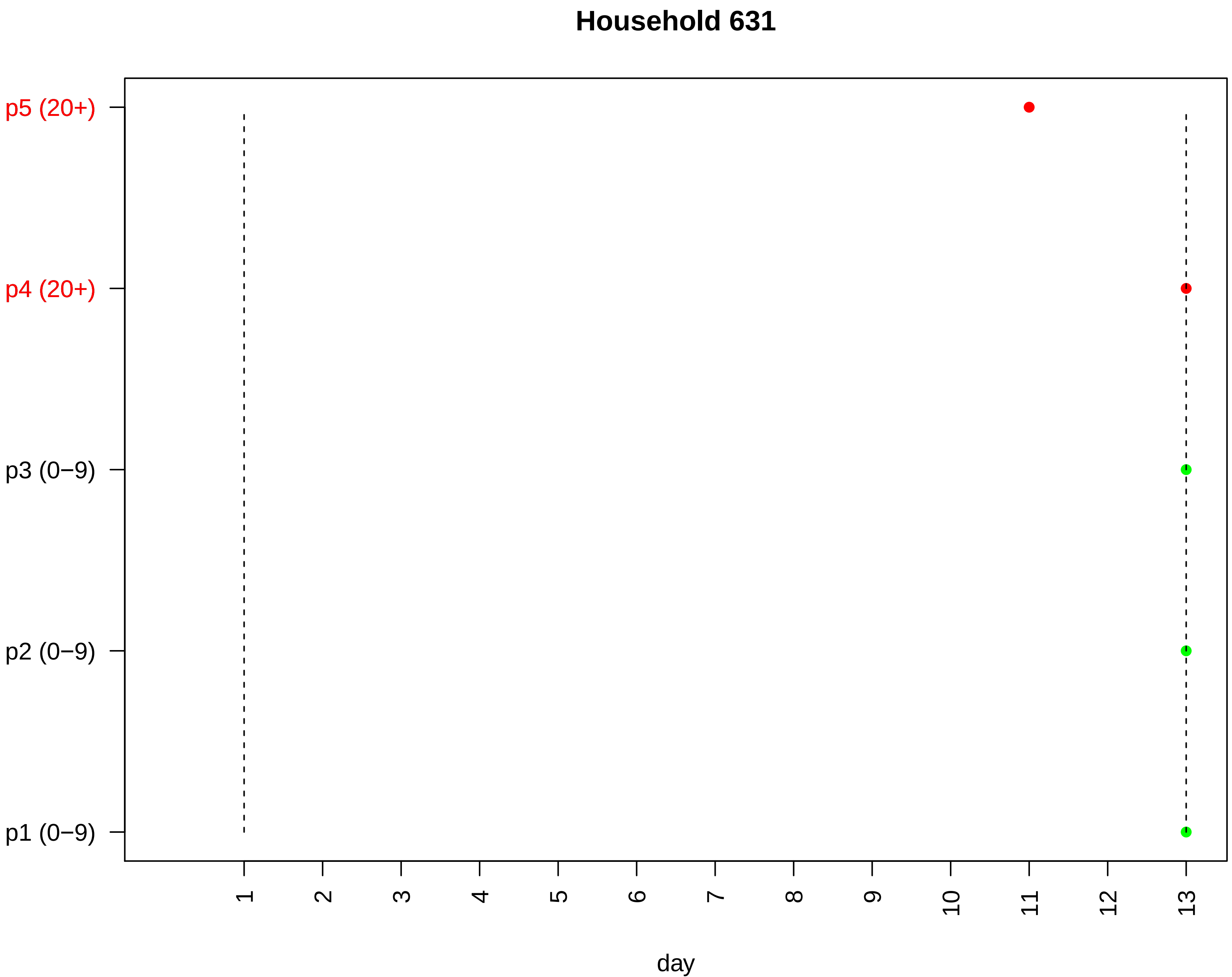
Household 631
p5 (20+)
p4 (20+)
p3 (0−9)
p2 (0−9)
p1 (0−9)
1
2
3
4
5
6
7
8
9
10
11
12
13
day

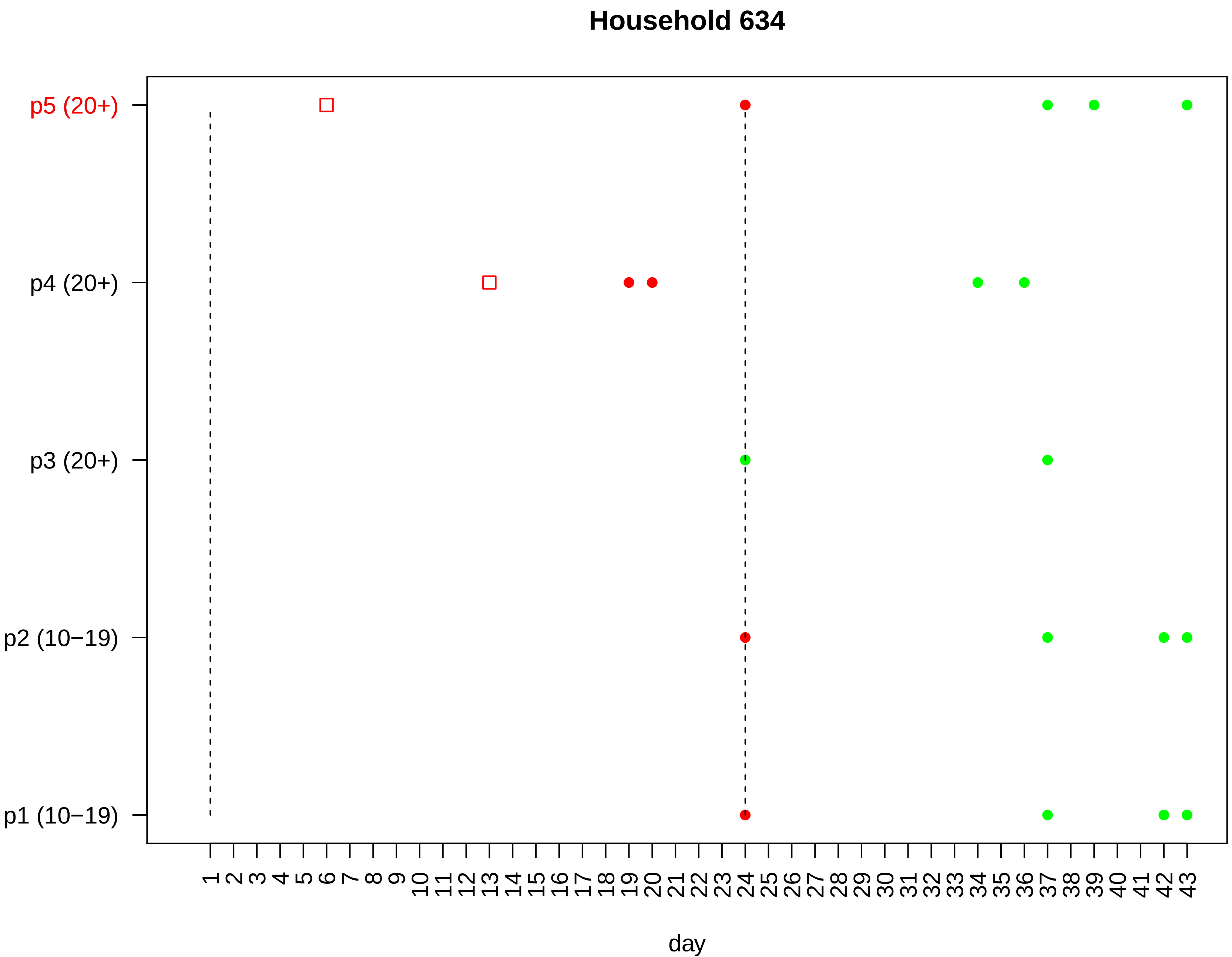

Household 634
p5 (20+)
p4 (20+)
p3 (20+)
p2 (10−19)
p1 (10−19)
1 2 3 4 5 6 7 8 9 10 11 12 13 14 15 16 17 18 19 20 21 22 23 24 25 26 27 28 29 30 31 32 33 34 35 36 37 38 39 40 41 42 43
day

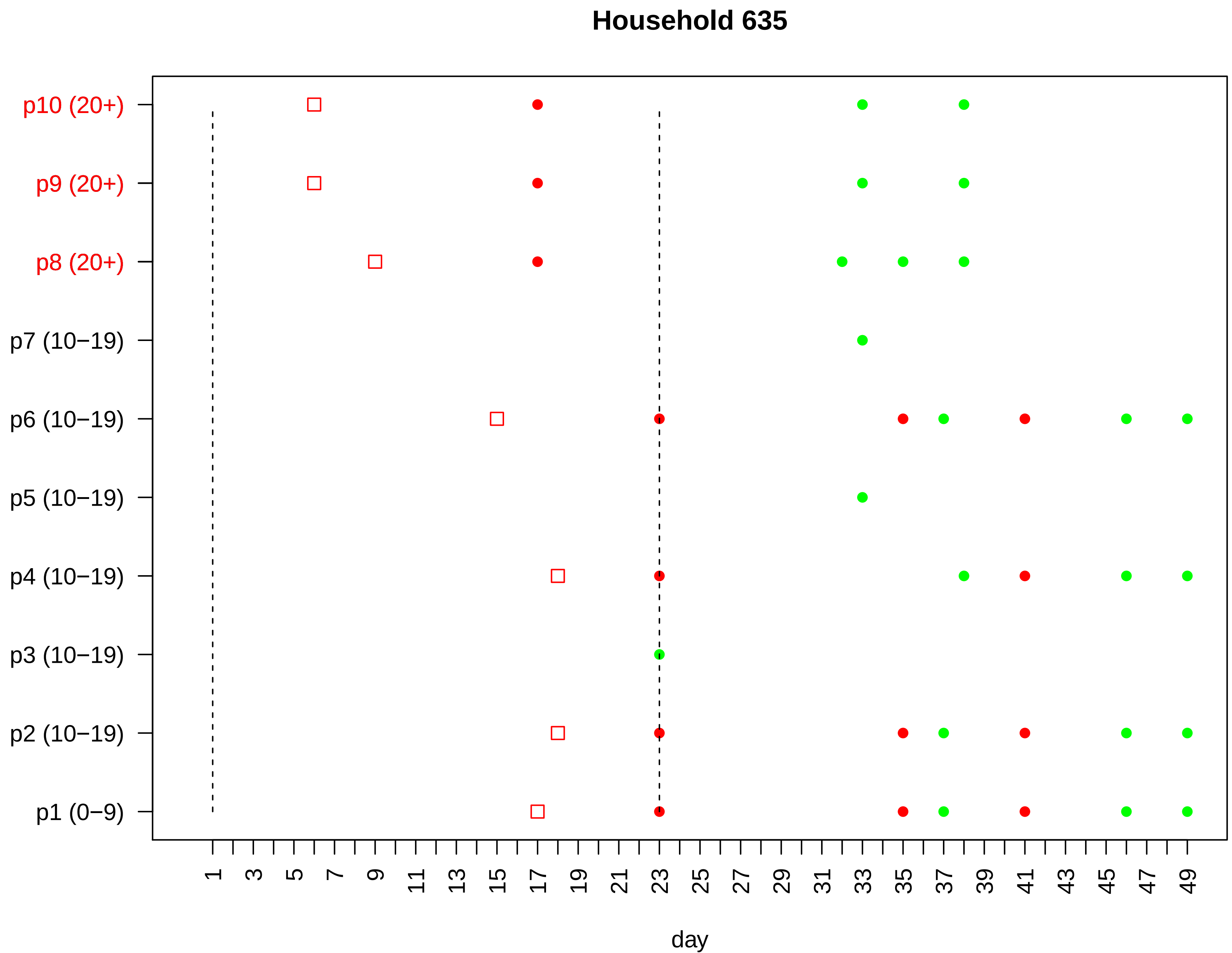

Household 635
p10 (20+)
p9 (20+)
p8 (20+)
p7 (10−19)
p6 (10−19)
p5 (10−19)
p4 (10−19)
p3 (10−19)
p2 (10−19)
p1 (0−9)
1 3 5 7 9 11 13 15 17 19 21 23 25 27 29 31 33 35 37 39 41 43 45 47 49
day

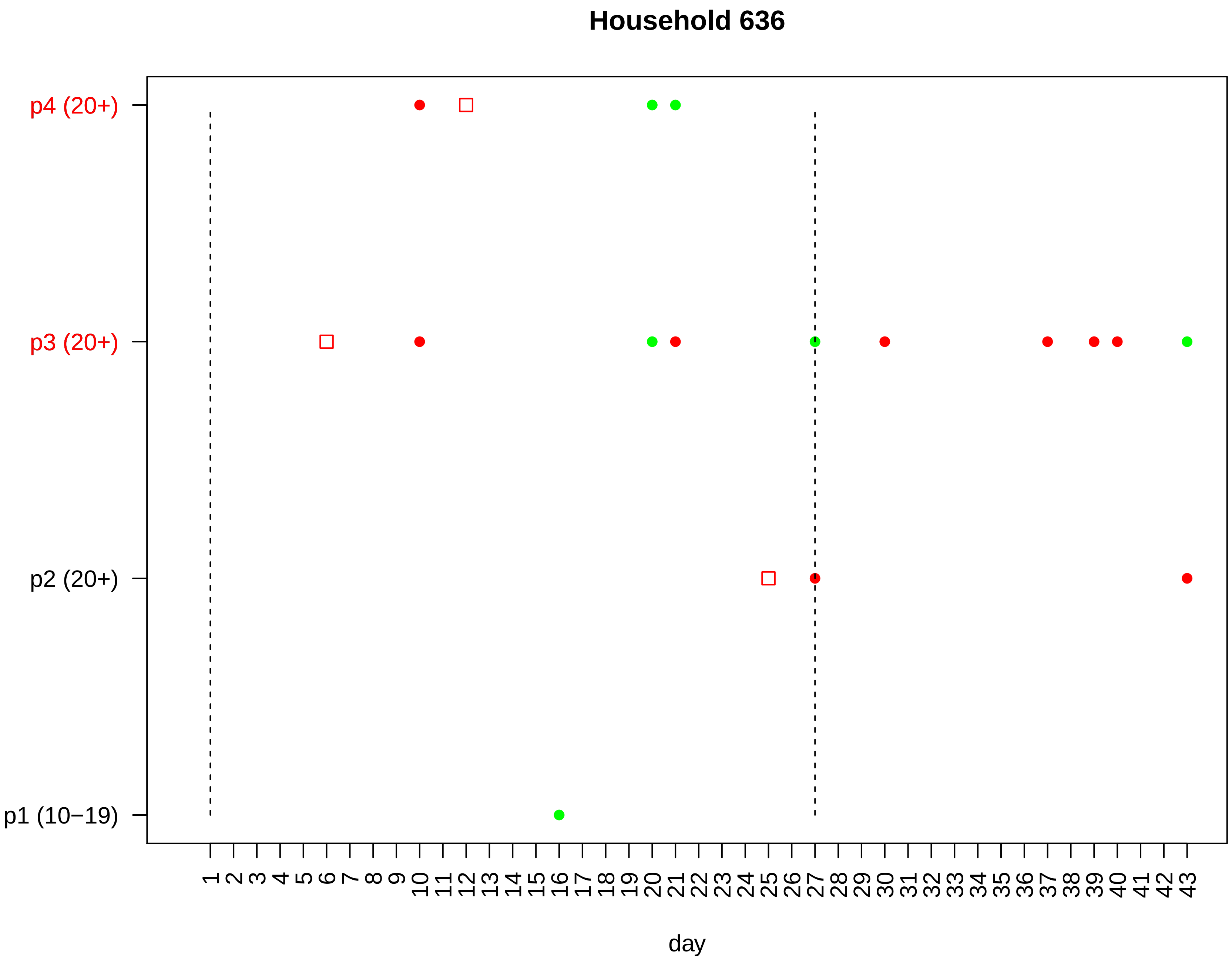

Household 636
p4 (20+)
p3 (20+)
p2 (20+)
p1 (10−19)
1 2 3 4 5 6 7 8 9 10 11 12 13 14 15 16 17 18 19 20 21 22 23 24 25 26 27 28 29 30 31 32 33 34 35 36 37 38 39 40 41 42 43
day

### Household 637

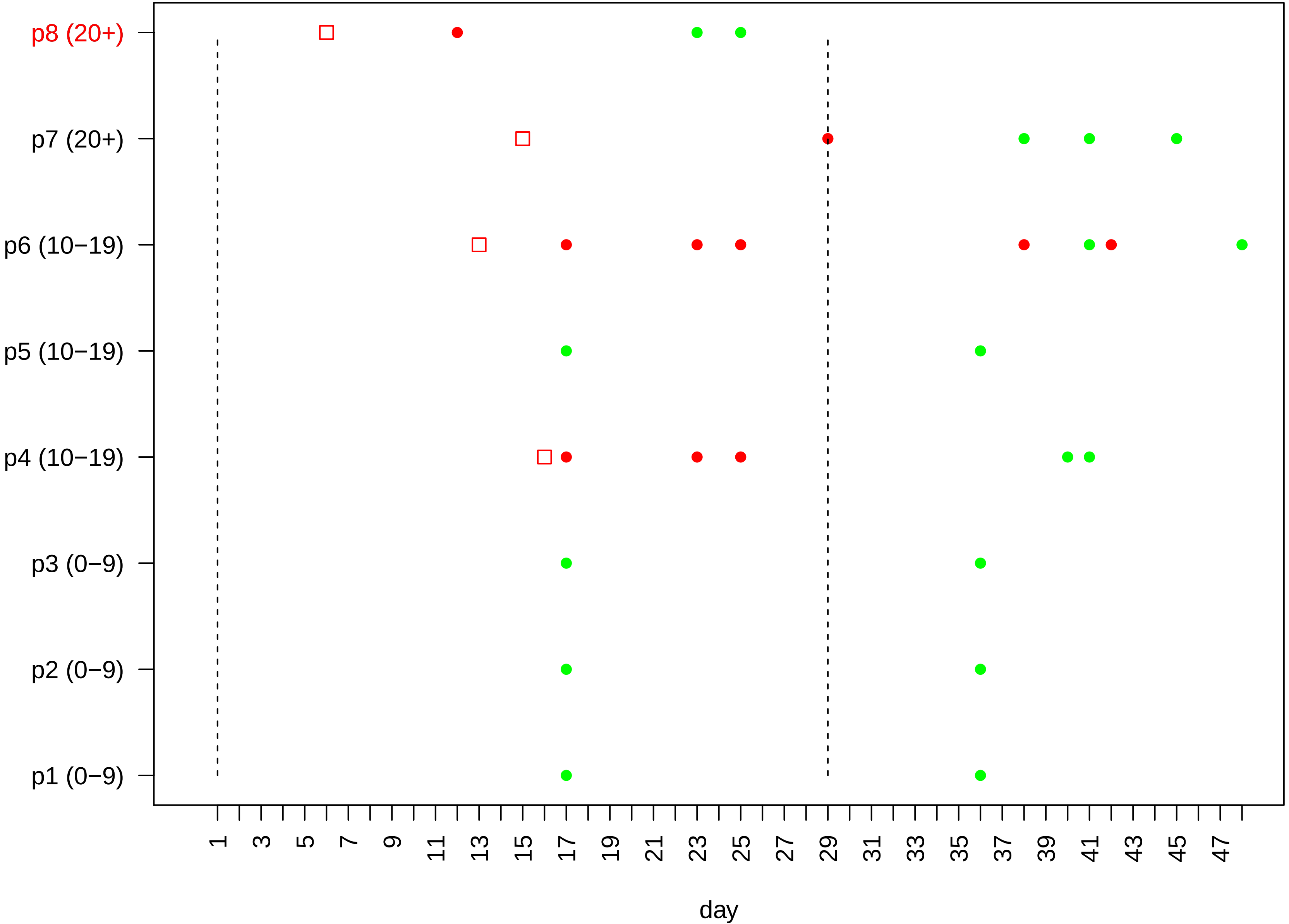
